# Sugar sweetened and artificially sweetened beverages, fruit and vegetable juices and cancer risk: a World Cancer Research Fund International Global Cancer Update Programme (CUP Global) systematic literature review and meta-analysis

**DOI:** 10.64898/2026.06.22.26355879

**Authors:** Ahmad Jayedi, Georgios Markozannes, Sayada Zartasha Kazmi, Margarita Cariolou, Rita Vieira, Sonia Kiss, Katia Balducci, Eirini Pagkalidou, Sofia Cividini, Dagfinn Aune, Darren C Greenwood, Laure Dossus, Emma Fontvieille, Nahid Ahmadi, Yahya Mahamat-Saleh, Amanda J. Cross, Marc J. Gunter, Shalini Jayasekar Zürn, Christian C. Abnet, Pietro Ferrari, Vanessa L.Z. Gordon-Dseagu, Kristy Maskell, Christelle Clary, Helen Croker, Panagiota Mitrou, Elio Riboli, Monica Baskin, Rajiv Chowdhury, Mia Gaudet, Edward Giovannucci, Ellen Kampman, Sarah J. Lewis, Anne M. May, Yikyung Park, Tobias Pischon, Gianluca Severi, Lynette Hill, Matty P. Weijenberg, John Krebs, Konstantinos K Tsilidis, Doris S.M. Chan

**Affiliations:** Department of Epidemiology and Biostatistics, School of Public Health, Imperial College London, London; Department of Hygiene and Epidemiology, University of Ioannina Medical School, Ioannina, Greece; Department of Nutrition, Oslo New University College, Oslo; Department of Research, The Cancer Registry of Norway, Oslo, Norway; Leeds Institute for Data Analytics, University of Leeds, Leeds, UK; Nutrition and Metabolism Branch, International Agency for Research on Cancer, Lyon, France; Department of Surgery and Cancer, Imperial College London, London, UK; Cancer Epidemiology and Prevention Research Unit, School of Public Health, Imperial College London, London, UK; Knowledge, Advocacy and Policy, Union for International Cancer Control, Geneva, Switzerland; Metabolic Epidemiology Branch (MEB), Division of Cancer Epidemiology and Genetics, National Cancer Institute, National Institutes of Health, Rockville, MD 20850, United States; Head, Nutrition and Metabolism Branch, International Agency for Research on Cancer, Lyon, France; World Cancer Research Fund International, London, UK; Massey Comprehensive Cancer Center, USA; Department of Global Health, Florida International University, Miami, Florida, USA; Trans-Divisional Research Program (TDRP), Division of Cancer Epidemiology Genetics (DCEG), National Cancer Institute (NCI), National Institutes of Health, Rockville, MD 20850, United States; Department of Epidemiology, Harvard T.H. Chan School of Public Health, Boston, MA, USA; Division of Human Nutrition and Health, Wageningen University & Research, Wageningen, The Netherlands; Population Health Sciences, Bristol Medical School, University of Bristol, Bristol, UK; University Medical Centre Utrecht, Julius Centre for Health Sciences and Primary Care, Utrecht, The Netherlands; Netherlands Cancer Institute, Division of Psychosocial Research & Epidemiology, PO Box 90203, 1006 BE Amsterdam, The Netherlands |; Division of Public Health Sciences, Washington University School of Medicine, Washington University in St.Louis, St Louis, USA; Molecular Epidemiology Research Group, Max Delbrück Center for Molecular Medicine, Berlin, Germany; CESP U1018, U.Paris-Saclay, UVSQ, INSERM, France; Public Representative; Department of Epidemiology, Maastricht University, Maastricht, The Netherlands; Department of Biology, University of Oxford, Oxford, UK

**Author notes:** Corresponding authors: Konstantinos K Tsilidis, Doris S.M. Chan Department of Epidemiology and Biostatistics, School of Public Health, Imperial College London, London.

## Abstract

**Background:** Sugar sweetened beverages (SSBs), artificially sweetened beverages (ASBs), and fruit and vegetable juices are consumed worldwide, yet their associations with cancer remain unclear.

**Methods:** Within World Cancer Research Fund International’s Global Cancer Update Programme (CUP Global), we conducted a systematic review by searching PubMed and Embase until September 2024 for cohort studies of SSBs, ASBs, and juices and cancer risk. Meta-analyses were conducted to calculate the relative risks (RRs) and 95% CIs per 1 serving/day (355 mL for SSBs/ASBs; 177 mL for juices). Evidence was graded by the CUP Global Expert Panel. The CUP Global standard protocol was registered at: https://osf.io/7utbm/.

**Findings:** We identified 158 publications from 51 cohorts. There was evidence of a probable causal positive association for SSBs, including carbonated SSBs, with pancreatic cancer incidence (RR 1.09 [95% CI 1.01–1.16]; I^2^=8%, n=18 studies), and for SSBs with colorectal cancer incidence (RR 1.07 [95% CI 1.00–1.14]; I^2^=41%, n=13). Limited suggestive evidence supported positive associations of SSBs with ovarian (RR 1.61 [95%CI 1.03–2.53]; I^2^=0%, n=2), endometrial (RR 1.21 [95%CI 1.03-1.42]; I^2^=0%, n=3), and postmenopausal breast cancer incidence (RR 1.05 [95%CI 1.00-1.10] ; I^2^=0%, n=6), and of carbonated ASBs with leukaemia incidence (RR 1.29 [95%CI 1.01-1.64]; I^2^=0%, n=2). There was evidence of a probable causal positive association for orange juice with different types of skin cancer including melanoma (RR 1.21 [95%CI 1.08-1.34]; I^2^=0%, n=4), and basal (RR 1.12 [95%CI 1.06-1.17]; I^2^=46%, n=2) and squamous cell carcinoma (RR 1.13 [95%CI 1.04-1.24]; I^2^=0%, n=2). An interactive evidence platform is available at: https://teacup.cc.ic.ac.uk/soft-drinks-cancer.html.

**Interpretation:** This review provides evidence supporting probable causal associations of SSBs with pancreatic and colorectal cancers, and of orange juice with skin cancers, with additional suggestive evidence for SSBs with other obesity-related cancers, extending concerns about sugary drink consumption beyond cardiometabolic health to cancer risk.

**Funding:** World Cancer Research Fund network of charities (American Institute for Cancer Research; World Cancer Research Fund; Wereld Kanker Onderzoek Fonds).

## Introduction

The intake of sugar sweetened beverages (SSBs) has increased worldwide over recent decades.^1^ Between 1990 and 2018, global consumption of SSBs rose by 0.68 servings/week among children and adolescents^2^ and by 0.37 servings/week in adults,^3^ resulting in an average weekly intake of 3.6 servings for children and adolescents and 2.7 servings for adults in 2018. These beverages represent the predominant source of added free sugars in the diet while providing minimal nutritional value.^4^

A growing body of evidence suggests that high consumption of added free sugars from SSBs contributes to weight gain, promotes de novo lipogenesis and ectopic fat accumulation, and exacerbates insulin resistance and low-grade systemic inflammation^5^ and is therefore linked to higher risks of type 2 diabetes ^6^ and heart disease.^7^ These concerns may encourage consumers to seek alternatives perceived as healthier.^8^ Artificially sweetened beverages (ASBs) and fruit juices are commonly consumed as substitutes for SSBs.^9^ However, potential adverse effects of ASBs on the gut microbiome, appetite regulation, sweet taste perception, and fat storage have been raised.^10^ In addition, commercially produced juice products (e.g., juice drinks and nectars) may contain added free sugars, fruit juice concentrates or other sweetening ingredients, further increasing their overall free sugar content. ^11^ Pure juices contain certain nutrients and are included in dietary guidelines in several countries, where modest consumption (e.g., half or one glass of unsweetened 100% fruit or vegetable juice) contributes to recommended fruit and vegetable intake.^12^ However, even 100% juices are low in fibre and relatively high in free sugars and energy-dense, and excessive consumption may promote weight gain.^13^

Epidemiological evidence linking SSBs consumption to cancer risk remains inconsistent and inconclusive ^14–16^. For ASBs, a meta-analysis of cohort studies suggested a positive association for leukaemia,^16^ while a recent umbrella review found no association for gastrointestinal cancers and total cancer mortality.^17^ Evidence for fruit juice is also inconsistent. ^15,16^ These findings, however, are limited by the inclusion of case-control studies and by heterogeneous exposure definitions, including the combination of different beverage types and lack of distinction between pure and commercially produced juices. Such heterogeneity complicates interpretation and limits the ability to draw conclusions about specific beverage types.

In 2018, the World Cancer Research Fund/American Institute for Cancer Research (WCRF/AICR) Third Expert Report^18^ issued 10 evidence-based recommendations for cancer prevention, including limiting SSBs intake, largely based on evidence linking SSBs to weight gain. At that time, however, epidemiologic evidence directly linking SSBs, ASBs, and fruit and vegetable juices to cancer risk was limited. To address these gaps, WCRF International’s Global Cancer Update Programme (CUP Global) prioritised ^19^ SSBs, ASBs, and fruit and vegetable juices for an updated systematic literature review to complement the conclusions of the Third Expert Report. This work presents the results of a systematic literature review examining the associations between different types of soft drinks and juices and the risks of site-specific cancer incidence in adults, and alongside a companion review on sedentary behaviour and cancer risk (Markozannes et al, unpublished) contributes to a series of CUP Global reviews examining lifestyle-related exposures and cancer risk that will inform the Fourth Expert Report which will be developed by WCRF International in the coming years.

## Methods

The present systematic review and meta-analysis have been conducted in accordance with a pre-registered CUP Global standard protocol (https://osf.io/7utbm/) and is reported in line with the Preferred Reporting Items for Systematic Reviews and Meta-Analyses (PRISMA) checklist (**Supplementary Table 1**).^20^

### Search strategy and selection criteria

We searched PubMed and Embase until 1^st^ September 2024 using a prespecified standard CUP Global strategy (**Supplementary Text 1**), supplemented by reference screening. In brief, eligible studies included randomised controlled trials (RCTs), cohort studies, Mendelian randomisation (MR) studies, and pooled analyses of such studies, on soft drinks or juices intake and adult cancer incidence or mortality that reported multivariable adjusted risk estimates (more information provided in **Supplementary Text 2)**.

We classified beverages into four main groups including 1) SSBs; 2) ASBs; 3) juices, and 4) soft drinks (unspecified non-alcoholic sweetened drinks, assumed to be a combination of SSBs and ASBs) (**Supplementary Text 3).** Risk of bias was assessed using a modified Risk of Bias for Nutrition Observational Studies (RoB-NObs) tool,^21^ originally developed by the U.S Department of Agriculture (USDA) Nutrition Evidence Systematic Review based on Cochrane’s Risk of bias In Non-randomised Studies of Interventions (ROBINS-I),^22^ and further adapted and validated by the CUP Global team at Imperial College London for lifestyle-disease associations (**Supplementary Text 4**). The RoB domains and criteria to rate studies in relation to each domain are listed in **Supplementary Tables 3-4**.

The characteristics and the results of the studies were extracted into the CUP Global database. Whenever possible, we preferentially extracted multivariable adjusted models excluding adiposity measures to avoid overadjustment for sugary beverages (SSBs, juices and soft drinks), while adiposity-adjusted models were used for ASBs (**Supplementary Text 5)**. The list of information extracted from MR studies is provided in **Supplementary Text 6**. Article selection, data extraction, and risk of bias assessment were double-checked in at least 10% of the records by a second reviewer. Any disagreement between reviewers was resolved with the review coordinator.

### Data analysis

Our primary analyses were linear dose-response meta-analyses when two or more cohort studies provided sufficient information. When dose-response meta-analysis was not possible or when more than a third of the included studies did not have sufficient information for inclusion in a dose-response meta-analysis, a supporting categorical high versus low analysis was conducted. When a meta-analysis was not possible, the studies were descriptively reviewed. Non-linear dose-response meta-analyses were conducted when at least five primary studies provided sufficient information (**Supplementary Text 7**). Meta-analyses were conducted separately for incidence and mortality, with additional combined analyses for high-fatality cancers (pancreas, liver, lung, and gallbladder). Hamling’s method was used to pool results reported by cancer subtypes (e.g., colon and rectal cancer) to account for common denominators in risk estimates.^23^ Each reported exposure was reviewed and analysed only once, however, we conducted additional analyses combining studies which reported on all SSBs with studies which reported only on carbonated SSBs, as carbonated SSBs constitute the predominant component of SSBs and this approach would increase statistical power. A similar approach was decided for carbonated ASBs and ASBs. We also combined orange juice with overall citrus fruit juice since orange juice is a good representative for citrus juice. A similar approach was used for pure fruit juice and fruit juice (**Supplementary Text 7**).

A DerSimonian-Laird random effects model^24^ was used for linear dose-response and categorical meta-analyses. The increment units for the linear dose-response meta-analyses were 355 mL (12 US fluid oz)/day for SSBs, ASBs, and soft drinks,^25^ and 177 ml (6 US fluid oz)/day for fruit or vegetable juices.^26^ The proportion of total variability in effect estimates attributable to heterogeneity between studies was assessed by the I^2^ metric.^27^ Potential sources of heterogeneity were explored by pre-specified subgroup analyses (**Supplementary Text 7**), when at least two studies reported the results for the same subgroup. Leave-one-out sensitivity analyses were conducted to assess the impact of individual studies on the summary estimate.^28^ Non-linear dose-response meta-analyses were conducted by one-stage mixed effects models ^29,30^ using restricted cubic splines with three knots placed at fixed percentiles (10th, 50th, and 90th) of the exposure distribution. ^31^ The potential for small study effects, including publication bias, was evaluated using the Egger’s test^32^ and by visually inspecting the funnel plots for asymmetry, when at least 10 studies were available.

Analyses were conducted in Stata version 16.1. We also assessed MR studies for completeness of evidence and reviewed them descriptively, but it is likely that these results are not fully credible, as the biological relevance of the genetic variants associated with these exposures is unclear.^33^

### Grading the evidence

An Expert Panel convened by WCRF International interpreted and graded the quality of the evidence as *convincing*, *probable, limited suggestive, limited no conclusion*, or *substantial effect on risk unlikely*, following predefined evidence-grading criteria designed to evaluate the likelihood of causality **(Supplementary Table 5)**. Evidence of biological plausibility in humans was also considered by the Expert Panel and assessed separately using a structured framework (REF) and reviewed by CUP Global collaborators at International Agency for Research on Cancer (IARC).

## Results

The systematic search identified 17 309 unique records, of which 633 were selected for full text review. Subsequently, 457 publications were excluded, resulting in 176 publications that were deemed potentially eligible (**Figure 1**). We additionally excluded 17 publications based on the reasons presented in **Supplementary Table 6** and added two additional publications found in a complementary search. Finally, 161 publications met the eligibility criteria, including 158 from 51 prospective observational cohort studies **(Supplementary Tables 7-10)** ^26,34–190^ and three MR studies ^191–193^, with no RCTs identified. A detailed list of exposure-outcome pairs assessed in each of the 51 cohort studies included in the systematic review is presented in (**excel file: Soft drinks and juices_Cohorts**).

**Figure 1.**
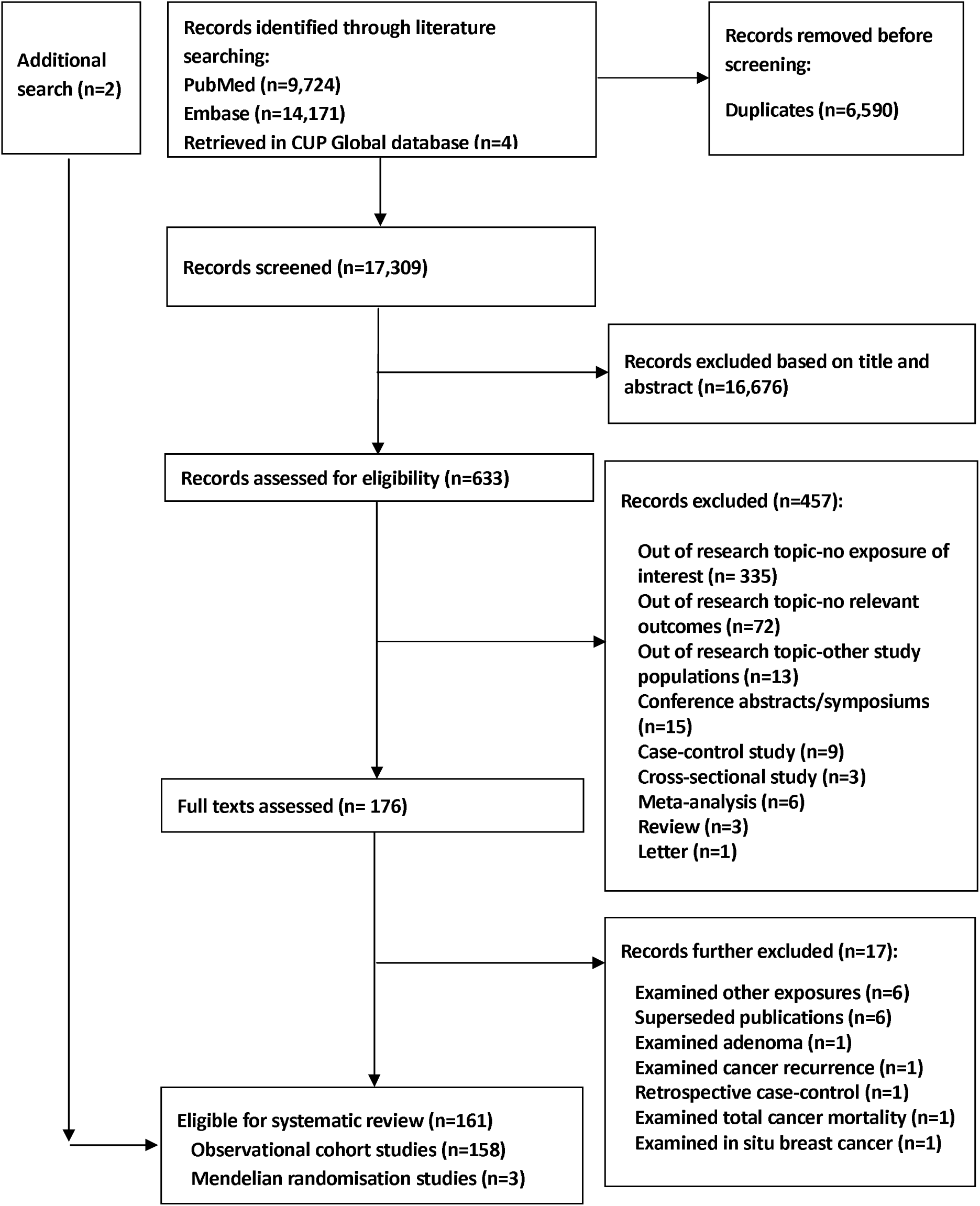
Flowchart of study selection process.

Detailed characteristics of the cohort studies on SSBs, ASBs, juices, and soft drinks are presented in **Supplementary Tables 11-17**. In brief, 20 studies (38%) (87 publications) were from North America,^34,37–39,41,42,46,48,50–52,54–60,62,63,71,72,74,75,77,79–82,86–89,93,94,98,102,105,108,109,113–116,119–123,128–131,135,138,141,143–145,148,150,151,153,157,158,160–163,168,170–175,177,178,183–190^ 16 studies (30%) (45 publications) were from Europe,^35,36,40,43,44,47,49,53,61,65–69,78,83,90,95,96,99,104,107,110–112,118,124,126,127,133,136,137,142,146,147,152,156,159,164–167,179–181^ eight (15%) (16 publications) were from Southeast/East Asia,^45,76,84,85,97,100,101,103,106,125,132,134,149,154,169,176^ two were from Australia,^70,73^ and eight pooled analyses from a combination of the above-mentioned regions.^26,64,91,92,117,139,155,182^

A summary of associations that received an evidence grading is presented in **Figures 2-4**. Summaries of all associations investigated across various exposure groups, including results for biological (e.g., molecular, histological, and clinical) subtypes and anatomical subsites and cancer mortality, are presented in **Supplementary Figures 1-7**, and summaries of associations across major cancer sites are presented in **Supplementary Figures 8-17**. Supporting results showing the categorical highest versus lowest comparisons are presented in **Supplementary Figures 18-69**. A brief overview of the results is provided below, with all estimates relating to cancer incidence, unless otherwise stated, and expressed per 1 serving/day (355 mL for SSBs and ASBs; 177 mL for juices). Results for cancer mortality and descriptive synthesis are detailed in **Supplementary Texts 8**-**16**, and single-study associations in **Supplementary Tables 18-21**.

**Figure 2.**
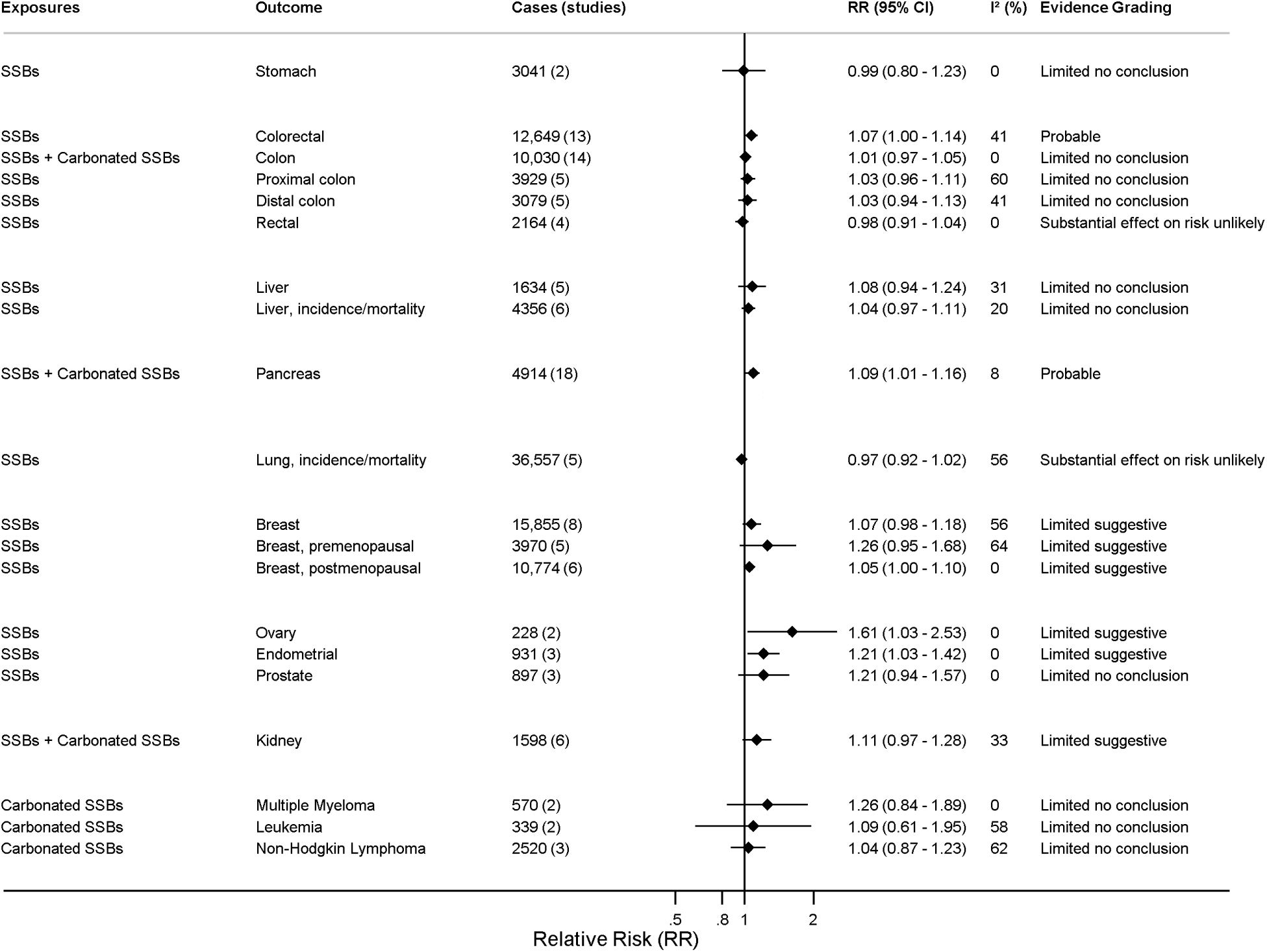
Summary forest plot showing the associations between sugar sweetened beverages (defined as liquids that are sweetened by adding free sugars, such as sucrose, high fructose corn syrup and sugars naturally present in honey, or syrups) (per 1 serving [355 mL]/day) and cancer risk (outcomes are incidence, unless otherwise stated). The forest plot shows the associations that received grading. Evidence grades were classified as convincing, probable, limited suggestive, limited no conclusion, or substantial effect on risk unlikely (indicating the likelihood of causality).

Thirty-nine studies (61 publications, including 10 pooled analyses) examined SSBs and cancer risk across 18 anatomical sites, encompassing 89 exposure-outcome pairs (64 incidence, 25 mortality) (Supplementary Tables 11).^34,36–40,42,44–47,53,64,66,70,72,74,75,78–82,84,85,90,97–101,105,106,108,109,113,114,122,126–131,135,145,146,150,157,168,169,172,173,177,178,180,182,184,186–188^ Forest plots indicating meta-analyses are indicated in **Supplementary Figures 70-104**, a summary of all associations in **Supplementary Figures 2-3**, and a summary of the associations that received grading in **Figure 2**.

There was a positive association between SSBs and ovarian (RR 1.61 [95%CI 1.03-2.53]; I^2^=0%) (2 studies, 228 cases)^38,70^ (**Supplementary Figure 70**) and endometrial cancer risk (RR 1.21 [95%CI 1.03-1.42]; I^2^=0%) (3 studies, 931 cases)^38,70,74^ (**Supplementary Figure 71**), as well as between SSBs and carbonated SSBs combined and pancreatic cancer risk (RR 1.09 [95%CI 1.01-1.16]; I^2^=8%) (18 studies, 5 publications including one pooled analysis of 14 cohorts, 4914 cases)^39,45,64,127,131^ (**Supplementary Figures 72-75).** There was also a positive association between SSBs and colorectal cancer risk (RR 1.07 [95%CI 1.00-1.14]; I^2^=41%) (13 studies, 11 publications, 12,649 cases)^38,40,44,70,81,97,101,108,129,135,168^ (**Supplementary Figure 76**), with little evidence of association for colorectal cancer anatomical subsites (**Supplementary Figures 77-82**). Of note, substituting SSBs with ASBs (per 1 serving/day) was associated with a lower risk of proximal colon cancer risk (RR 0.84 [95%CI 0.77-0.95]) (2 studies, 1 publication, 1275 cases)^178^ (**Supplementary** Figure 83).

There was also an indication of a positive association between SSBs and breast cancer risk (RR 1.07 [95%CI 0.98-1.18]; I^2^=56%) (8 studies, 7 publications, 15 855 cases)^38,44,97,108,128,145,146^ (**Supplementary Figure 84**) and premenopausal breast cancer risk (RR 1.26 [95%CI 0.95-1.68]; I^2^=64%) (5 studies, 4 publications, 3970 cases),^38,44,128,145^ although CIs included the null. The association was more evident for postmenopausal breast cancer risk (RR 1.05 [95%CI 1.00-1.10]; I^2^=0%) (6 studies, 5 publications, 10 774 cases)^38,44,70,130,145^ (**Supplementary Figure 85**). A positive association but with a CI crossing the null value was also observed between SSBs and carbonated SSBs combined and kidney cancer risk (RR 1.11 [95%CI 0.97-1.28]; I^2^=33%) (6 studies, 5 publications, 1598 cases)^66,70,98,100,173^ (**Supplementary Figures 87-88).** Little evidence of an association was observed for other investigated exposure-outcome pairs (**Supplementary Figures 89-104)**. Non-linear dose-response meta-analysis of colorectal, breast, and postmenopausal breast cancers indicated no evidence of departure from linearity (**Supplementary Figures 105-107**).

Subgroup analyses by study and participant’s characteristics are presented in **Supplementary Table 19** and **Supplementary Figures 108-154**. Of note, there was no heterogeneity in the subgroup analyses by potential presence of biases due to confounding and deviation from intended exposure and by frequency of dietary assessment (baseline vs repeated). The positive association between carbonated SSBs and pancreatic cancer was consistent across subgroups defined by BMI, alcohol consumption, and physical activity (**Supplementary Figure 139**). There were indications of heterogeneity by geographical location, where studies conducted in Europe indicated positive associations between SSBs and colorectal (RR 1.82 [95%CI 1.04-3.16]; I^2^=0%; n=2) (**Supplementary Figure 109**) and breast cancer risk (RR 2.07 [95%CI 1.27-3.38]; I^2^=0%; n=2) (**Supplementary Figure 140**), while studies conducted in the North America and East/Southeast Asia showed weaker or no association (P group differences = 0.018 and 0.021, respectively) (**Supplementary Table 19)**. Sensitivity analyses showed largely stable results (**Supplementary Figures 155-168**). Test for publication bias was possible for colorectal cancer, which indicated little evidence of small-study effects (P Egger’s test = 0.13) (**Supplementary Figure 169**).

Fifteen studies (26 publications, including seven pooled analyses) examined ASBs and cancer risk across 18 anatomical sites, covering 66 exposure-outcome pairs (47 incidence, 19 mortality) (Supplementary Tables 12).^39,44,49,66,70,72,74,75,78,79,98,109,113,114,126,127,140,144,145,150,151,168,177,180,186,187^ Linear dose-response meta-analyses are indicated in **Supplementary Figures 191-209**, a summary of all associations in **Supplementary Figure 4,** and a summary of the associations that received grading in **Figure 3**. A positive association was seen between carbonated ASBs and leukaemia risk (RR 1.29 [95%CI 1.01-1.64]; I^2^=0%) (2 studies, 1 publication, 339 cases)^150^ (**Supplementary Figure 191**), and between ASBs (RR 1.05 [95%CI 1.00-1.10]; I^2^=23%) (3 studies, 10 896 cases)^39,113,127^ and ASBs and carbonated ASBs combined (RR 1.03 [95%CI 1.01-1.05]; I^2^=0%) (5 studies, 4 publications, 11,275 cases)^39,113,127,151^ and pancreatic cancer incidence/mortality (**Supplementary Figures 192-194**). Little evidence of an association was observed for other investigated exposure-outcome pairs (**Supplementary Figures 195-209)**. There was no evidence of heterogeneity in the subgroup analyses (**Supplementary Table 20**, **Supplementary Figures 210-220**). The main results did not change materially in the influence analyses (**Supplementary Figures 221-228**).

**Figure 3.**
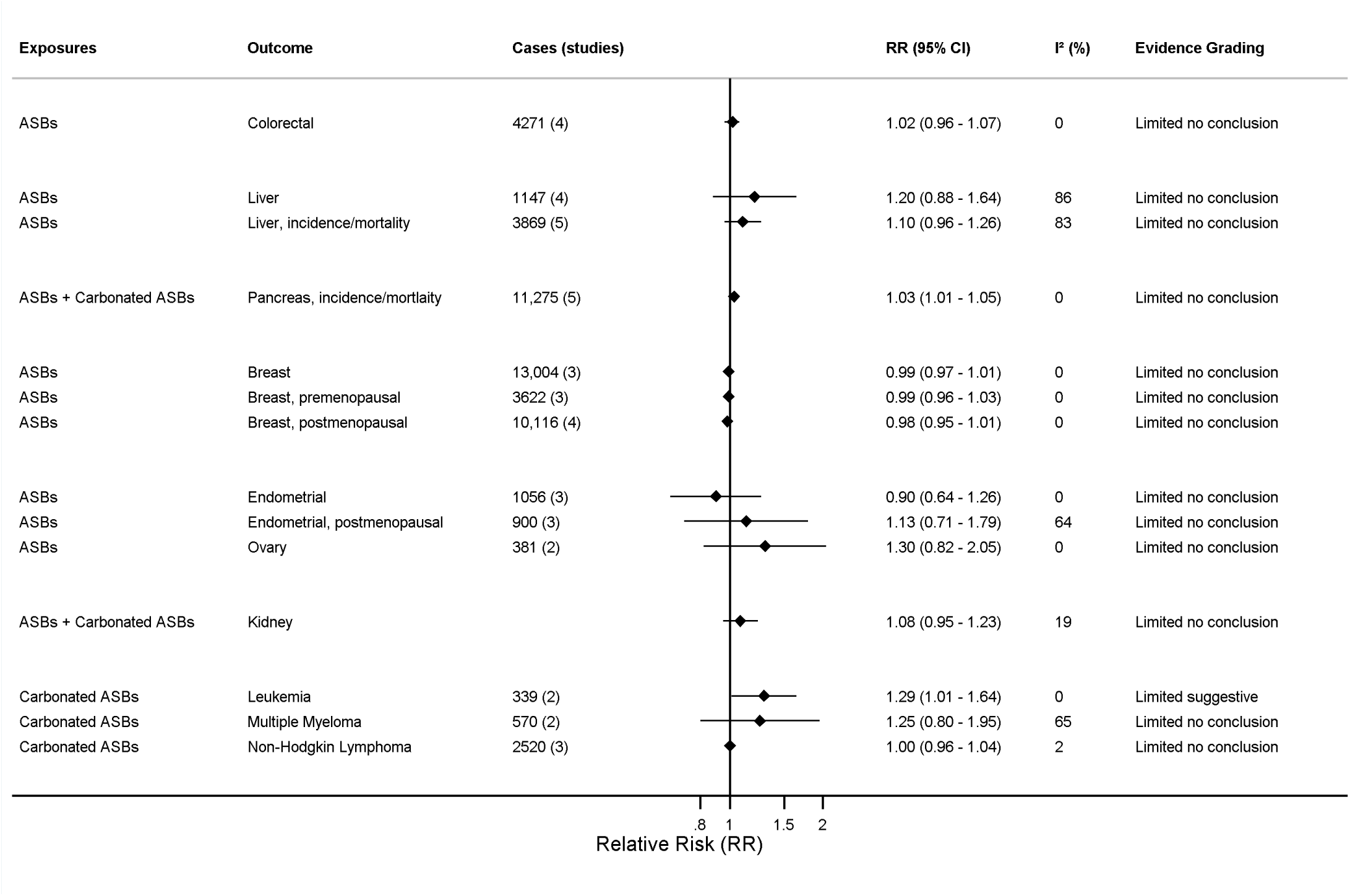
Summary forest plot showing the associations between artificially sweetened beverages (defined as low calorie or non-caloric beverages sweetened with artificial sweeteners, such as aspartame, acesulfame K, saccharin, sucralose, or neotame) (per 1 serving [355 mL]/day) and cancer risk (outcomes are incidence, unless otherwise stated). The forest plot shows the associations that received grading. Evidence grades were classified as convincing, probable, limited suggestive, limited no conclusion, or substantial effect on risk unlikely (indicating the likelihood of causality).

Forty studies (93 publications, including 13 pooled analyses) examined juices and cancer risk across 18 anatomical sites, encompassing 187 exposure-outcome pairs (167 incidence, 20 mortality) (Supplementary Tables 13).^26,36,38,41,43,44,47,49,51,52,54–60,62,63,65,66,69,72,74,76–79,81,83,84,86–94,98,103,104,107,108,110–112,115–118,120,121,123–125,127,131,133,134,136,138,139,141,142,147–149,152–156,158–162,164–167,170,171,174–176,179,181,183,185,188^ Forest plots presenting meta-analyses are indicated in **Supplementary Figures 242-291**, a summary of all associations in **Supplementary Figures 5-7,** and a summary of the associations that received an Expert Panel grading is shown in **Figure 4**.

**Figure 4.**
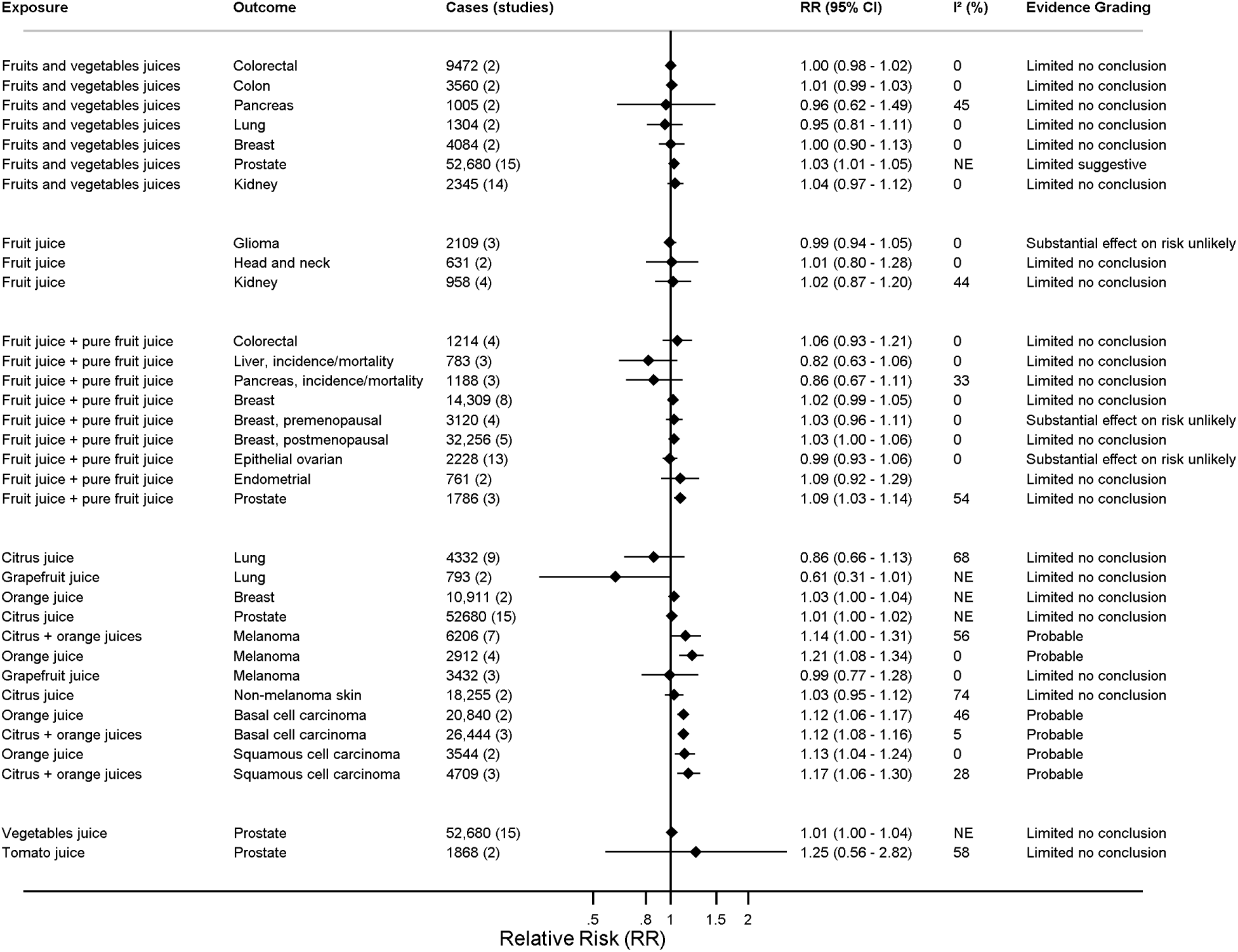
Summary forest plot showing the associations between juices (per 1 serving [177 mL]/day) and cancer risk (outcomes are incidence, unless otherwise stated). Juice exposures reflect definitions used in the individual studies and could include pure and commercially produced juices; where specific juice subtypes were reported separately (e.g., pure fruit juices), these were combined where appropriate for grading. The forest plot shows the associations that received grading. Evidence grades were classified as convincing, probable, limited suggestive, limited no conclusion, or substantial effect on risk unlikely (indicating the likelihood of causality).

In general, there was little evidence of an association between fruit juices and cancer incidence, except for positive associations between fruit juice (RR 1.09 [95%CI 1.03-1.14]; I^2^=75%) (2 studies, 1495 cases)^89,108^ and fruit juice and pure fruit juice combined (RR 1.09 [95%CI 1.03-1.14]; I^2^=54%) (3 studies, 1786 cases)^44,89,108^ and prostate cancer risk (**Supplementary Figures 242-246**). Based on a pooled analysis of 15 studies with 52 680 cases, there was a positive association between fruit and vegetable juice and risk of prostate cancer (RR 1.03 [95%CI=1.01-1.05]) and its subtypes including localised, high grade, and especially advanced restricted prostate cancer (RR 1.11 [95%CI=1.02-1.21]) (**Supplementary Figure 247**).^139^

Orange juice was associated with a higher risk of melanoma (RR 1.21 [95%CI 1.08-1.34]; I^2^=0%) (4 studies, 3 publications, 2912 cases)^54,110,171^ (**Supplementary Figure 269**) and skin basal cell (RR 1.12 [95%CI 1.06-1.17]; I^2^=46%) (2 studies, 1 publication, 20 840 cases)^170^ (**Supplementary Figure 270**) and squamous cell carcinoma (RR 1.13 [95%CI 1.04-1.24]; I^2^=0%) (2 studies, 1 publication, 3544 cases)^170^ (**Supplementary Figure 271**). Additional analyses combining studies reporting on citrus juice with those reporting on orange juice showed similar positive associations (**Supplementary Figures 272-274**). Other results indicated evidence of no association between grapefruit juice and risks of skin cancers, citrus juice and vegetables juice and prostate cancer, and fruit and vegetable juice and various cancers (**Supplementary Figures 275-291).** A limited number of subgroup analyses were possible which indicated no evidence of heterogeneity (**Supplementary Table 21**, **Supplementary Figures 292-299**). The main results did not change materially in influence analyses (**Supplementary Figures 300-307**).

Twenty-six studies reported on total soft drinks (**Supplementary Tables 14**).^26,35,36,39,48–50,61,65–68,71,73,79,85,95,96,102,117–119,125–127,132,136,137,140,143,147,159,163,180,183,189^ Meta-analyses were possible for 13 cancers **(Supplementary Figure 1)**, suggesting no clear associations between soft drinks and cancer incidence (**Supplementary Figure 309-325**, **Supplementary Table 22**, **Supplementary Texts 17-19**).

Risk of bias assessments are summarised in **Supplementary Figure 326** and detailed in **Supplementary Text 20**. Most publications (81%) had moderate, 8% serious, and 11% critical risk of bias due to confounding, reflecting incomplete adjustment for key factors such as smoking intensity (for smoking-related cancers), alcohol intake, reproductive factors, race, and skin cancer-related variables. Bias due to departure from intended exposures was frequently rated as critical (74% of publications), primarily due to reliance on single baseline dietary assessments. However, subgroup analyses by assessment frequency and risk of bias did not show important difference between subgroups. Other domains, including exposure classification, missing data, outcome measurement, and selective reporting, were generally of lower concern. Evidence from three MR studies ^191–193^ on genetically predicted self-reported intake of sweet beverages, ^192^ SSBs, ^191,192^ASBs, ^191^ and artificial sweeteners added to coffee or tea ^193^ and various cancer sites are presented in **Supplementary Table 23** and reviewed in **Supplementary Text 21.**

Based on IARC’s review findings, the MEC did not identify strong evidence of biological mechanisms linking soft drinks and cancer through insulin- and glucose-related IPs (Fontvieille et al., unpublished). The MEC also acknowledged that furocoumarins may be implicated in biological mechanisms linking citrus fruit juices (including orange juice) to skin cancer; however, further research is required to fully characterise these mechanisms (see **Text Box** (**Panel) 1** for more information).

A summary of the evidence grading agreed by the Expert Panel is presented in the **Text Box** (**Panel) 2**. The evidence was graded as probable, indicating the likelihood of causality, for a positive association between SSBs and pancreatic and colorectal cancers, and limited suggestive for ovarian, endometrial, kidney, and breast cancer overall and by menopausal status. The evidence was also graded probable for the positive associations between orange juice and citrus juice and orange juice combined and risks of melanoma and skin basal cell and squamous cell carcinomas, and lower evidence grades were concluded for other cancers. An online interactive tool to help explore the evidence can be found at: https://teacup.cc.ic.ac.uk/soft-drinks-cancer.html.

## Discussion

In this systematic literature review of 158 publications from 51 cohorts, we assessed associations between SSBs, ASBs, and fruits and vegetable juices and 19 cancer sites. Higher intake of SSBs was positively associated with the incidence of pancreatic, colorectal, ovarian, endometrial, kidney, and breast cancers, although the evidence supporting a causal association varied across cancers, with probable evidence for pancreatic and colorectal cancers, and limited suggestive evidence for ovarian, endometrial, kidney, and breast cancer overall and by menopausal status. ASBs showed mostly null associations, except for limited suggestive evidence supporting a positive association with leukaemia incidence. Among juices, orange juices and orange and citrus juices combined were positively associated with the incidence of melanoma and skin basal and squamous cell carcinomas (probable evidence), and fruit and vegetable juice with prostate cancer (limited suggestive evidence). Most other associations were null.

For SSBs, the results were indicative of positive associations with several obesity-related cancers, including pancreatic, colorectal, ovarian, endometrial, kidney, and breast cancers, suggesting that high intake of SSBs, partly through their contribution to weight gain, is associated with cancer risk. The evidence was graded as probable for pancreatic cancer, based on consistent findings from many cohort studies evaluating SSBs and carbonated SSBs. Notably, the positive association between carbonated SSBs and pancreatic cancer persisted across several lower-risk subgroups, including individuals with normal weight, non-alcohol consumers, and those with higher physical activity levels. This consistency strengthens the robustness of the association, reduces concerns about confounding by alcohol or physical inactivity, and suggests potential mechanisms beyond adiposity alone, including inflammation and insulin resistance, which are implicated in pancreatic carcinogenesis. ^194^ However, mechanistic evaluation within CUP Global did not identify strong evidence supporting insulin- and glucose-related pathways linking SSBs consumption to cancer, reflecting limitations in the available literature. From a public health perspective, excessive intake of SSBs remains a well-established contributor to weight gain, overweight, and obesity, which are recognised risk factors for multiple cancers, as outlined in the WCRF/AICR Third Expert Report in 2018. ^18^

Probable evidence was also observed for SSBs and colorectal cancer overall, based on 13 studies with low heterogeneity and risk of bias, supported by a positive association with colorectal cancer mortality. Subgroup analyses by using single versus repeated dietary assessments showed no heterogeneity, supporting the robustness of the findings. In contrast, evidence for colorectal cancer anatomical subsites was largely null, with limited no conclusion for colon cancer and strong evidence of no association for rectal cancer. These discrepancies likely reflect differences in data availability, as colorectal cancer analyses included more recent cohorts or updated dietary data, better capturing SSBs intake given its recent global increase in consumption. ^2,3^ In contrast, colon cancer estimates relied largely on an older pooled analysis and rectal cancer analysis was based on limited case numbers, underscoring the need for additional contemporary high-quality data.

Evidence for ovarian and endometrial cancers, two other obesity-related cancers,^195^ was graded as limited suggestive, based on a small number of studies with limited case numbers, underscoring the need for additional larger studies to strengthen the evidence base and improve certainty. Combined evidence from studies of SSBs and carbonated SSBs were suggestive of a positive association with kidney cancer, supported by a positive association between SSBs and kidney cancer mortality. Associations with breast cancer were consistently positive overall and by menopausal status, although the association was more evident for postmenopausal breast cancer, which is an established obesity-related cancer.^195^ Whilst the wide CIs limited statistical evidence for kidney, breast, and premenopausal breast cancers, the expert panel concluded that the magnitude of the point estimates and consistency in the direction of the associations across included studies supported limited suggestive evidence of a positive association.

There was limited suggestive evidence of a positive association between carbonated ASBs and leukaemia, based on a pooled analysis of two cohorts of health professionals with repeated dietary measurements and low risk of bias due to confounding, with stratified analyses indicating no effect modification by BMI.^150^ Experimental evidence in animals^196,197^ and re-analysis of long-term cancer studies conducted on rats suggest potential carcinogenicity of aspartame,^198^ whereas short-term experimental studies in animals largely support its safety.^199^ However, long-term health effects at intakes below established acceptable daily intake remain uncertain.^200^ Another possible explanation could involve the shared factors between SSBs and ASBs, such as the additional ingredients found in sodas or the materials used for soda packaging.^201,202^ There was also a positive association between ASBs (including carbonated ASBs) and pancreatic cancer risk; however, the evidence was graded as limited no conclusion. This was because more than 95% of the analytical weight was contributed by a single large cohort in the US.^113^ In addition, concerns remain regarding confounding by health status (e.g., pre-existing diabetes) and potential reverse causation, whereby individuals may have switched from SSBs to ASBs in response to recent weight gain or metabolic risk. These concerns are further amplified by the reliance of most studies on a single baseline dietary and confounders assessment, limiting the ability to capture changes in beverage consumption over time. Overall, the limited number of studies and inconsistent directions of associations across cancer sites preclude firm conclusions and underscore the need for further research.

Higher intake of orange juice and combined orange/citrus juices was consistently associated with higher risks of melanoma and basal and squamous cell carcinomas of the skin. Low between-study heterogeneity and low risk of bias for confounding and exposure misclassification, together with plausible biological mechanisms reviewed by the MEC, supported probable evidence gradings for skin cancers. Citrus furocoumarins/psoralens absorb ultraviolet radiation and can induce phototoxic skin reactions ^203,204^ which have lethal, mutagenic, and clastogenic effects.^205^ In two pooled US health professionals cohorts, the positive association between orange juice intake and squamous cell carcinoma was restricted to tumours at chronically sun-exposed sites, including the head, neck and extremities, with no association for tumours at truncal sites. ^170^ Similarly, in the UK Biobank, citrus fruits intake was positively associated with melanoma among individuals with fair skin but inversely associated among those with olive skin, supporting potential effect modification by sun exposure and sensitivity.^110^ Whilst the MEC identified the interaction between furocoumarins and sun exposure as a plausible biological mechanism, further research is needed to fully characterise these pathways.

Associations for fruit juice and pure fruit juice were largely null, although limited data was available for pure fruit juices. Based on a pooled analysis of 15 cohorts, ^139^ limited suggestive evidence was found for a positive association between intake of fruit and vegetable juice, including both pure and commercially produced ones, and prostate cancer incidence; the cautious grading from the expert panel for this association is due to a concern of detection bias due to greater health-seeking behaviour and prostate cancer screening among higher consumers. Positive associations between fruit juice and fruit juice and pure fruit juice combined and prostate cancer were also reported, although this finding was largely driven by an observational analysis within a large screening trial in the US,^89^ resulting in limited no conclusion grading and highlighting the need for further research.

Several limitations of the available evidence should be considered. Most included cohorts relied on a single baseline dietary assessment, introducing potential exposure misclassification, which is likely non-differential and may have attenuated true associations. In addition, most studies (82%) adjusted for adiposity, a potential mediator of SSBs-cancer associations, which may have further attenuated estimates, as suggested by modestly weaker associations in BMI-adjusted models.

Specified substitution analyses were rarely identified as such, although many models in the primary studies adjusted for total energy intake and may partially reflect unspecified isocaloric substitution; ^206,207^ however, those that performed specified substitution analyses suggested an inverse association for proximal colon cancer when substituting SSBs with ASBs. Nevertheless, further research is needed to evaluate substitution with lower-risk alternatives, particularly water. For ASBs, confounding by weight or underlying health status remains an important consideration to address, and where possible, rule out, as individuals with obesity or type 2 diabetes may preferentially consume these beverages; however, the few studies that examined associations stratified by BMI or diabetes status did not indicate heterogeneity across these subgroups. For juices, data on pure fruit juice were limited, despite its inclusion in dietary recommendations in some countries, underscoring the need for more robust evidence on its potential health effects. ^12^ In addition, most studies did not distinguish between pure and commercially produced juices, which differ in nutrient composition, particularly in added sugars, thereby limiting definitive conclusions.

This comprehensive evaluation of the evidence provides timely and policy-relevant insights for cancer prevention. Key strengths include the large evidence base, rigorous and harmonised exposure classification that avoided combining heterogeneous beverage types, and the use of multiple complementary analytical approaches, including dose-response meta-analyses, high-versus-low comparisons, and descriptive synthesis where data were limited. Extensive subgroup and sensitivity analyses supported the robustness of the findings. The integration of mechanistic evidence and the use of predefined criteria by an independent expert panel to grade the strength of evidence further strengthen the validity and interpretability of the conclusions. The findings strengthen the rationale for limiting SSBs consumption, extending concerns beyond obesity and cardiometabolic disease to site-specific cancer risks. Given the global rise in SSBs intake, ^2,3^ these results support population-level interventions, such as taxation policies, reformulation strategies, and clearer front-of-pack labelling, to reduce consumption. Results from intervention studies suggest that SSBs reduction leads to weight loss.^208^ Importantly, our findings also indicate that commonly used substitutes, including ASBs and certain juices, are not necessarily risk-free, underscoring the need for nuanced dietary guidance that differentiates between beverage types rather than promoting simple substitution. The evidence generated in the present review provides a robust foundation to inform future cancer prevention recommendations and highlights the importance of integrating beverage-specific guidance into national and international dietary policies. Specifically, this evidence will form part of the Fourth Expert Report which will be developed by WCRF International in the coming years.

In summary, the present SLR found evidence for a probable causal positive association between SSBs consumption and risk of colorectal and pancreatic cancers, and limited suggestive evidence for ovarian, endometrial, kidney, and breast cancer overall and by menopausal status. Evidence for ASBs was less clear, with limited suggestive evidence of a positive association for leukaemia. For juices, most associations were null, expect for evidence supporting probable causal positive associations between orange and citrus juices and melanoma and skin basal and squamous cell carcinomas. Future studies should focus on repeated measurements during the follow-up, more clear classification of dietary exposures (e.g., pure vs commercially produced fruit or vegetable juices or SSBs vs ASBs), exposure substitution models, and research on less-investigated cancers to generate more specific evidence-based guidelines. Future studies should also prioritise mechanistic research with longitudinal designs and standardised intermediate phenotypes measurement to clarify biological pathways.

## Research in context

### Evidence before this study

At the time of the WCRF/AICR Third Expert Report (2018), epidemiological evidence linking soft drinks and juices to cancer risk was limited. To prioritise future systematic review topics, CUP Global conducted a data-scanning and prioritisation exercise^19^ and searched PubMed (January 2019-February 2024) for meta-analyses, pooled analyses, randomised trials, Mendelian randomisation studies, and large cohort studies using standard CUP Global search strategy and identified soft drinks and juices as one of the prioritised topics for an updated systematic review.

Previous reviews found in the prioritisation exercise reported inconsistent associations for SSBs and inconclusive evidence for ASBs and juices, largely due to inclusion of case-control studies and heterogeneous exposure definitions, including combining SSBs with soft drinks (unspecified sweetened drinks, likely the combination of SSBs and ASBs) or grouping pure and commercially produced juices. Consequently, evidence for specific beverage and juice types remained unclear.

### Added value of this study

We performed a cancer-wide systematic review and dose-response meta-analysis of 158 publications from 51 cohort studies, encompassing 281,560 incident cancer cases across 19 major cancer sites. Using harmonised exposure definitions and pre-specified analytic strategies, we quantified associations for SSBs, ASBs, and fruit and vegetable juices overall and by types. We found that higher intake of SSBs is associated with higher risks of pancreatic, colorectal, ovarian, endometrial, kidney, and breast cancer overall and by menopausal status. We also found a positive association between carbonated ASBs and leukaemia incidence. Orange juice and citrus and orange juices combined were associated with higher risks of melanoma and skin basal cell and squamous cell carcinomas. There was also evidence of a positive association between fruit and vegetable juice and prostate cancer incidence.

### Implications of all the available evidence

Taken together, the evidence supports limiting consumption of SSBs as part of cancer prevention strategies, extending concerns beyond weight gain to site-specific cancer risk. The findings also highlight that not all beverage substitutes are necessarily risk-free, with emerging evidence for ASBs and citrus juices warranting caution. These results reinforce existing public health policies targeting sugary drink consumption and underscore the need for clearer dietary guidance that differentiates between beverage and juice types.

### Text Box (Panel) 1. Mechanistic review

Evidence of biological plausibility in humans was also considered by the Expert Panel and assessed separately using a structured framework. This first involved expert input and use of an automated tool (https://www.temmpo.org.uk/) to identify intermediate phenotypes of potential interest. Insulin-and glucose-related biomarkers and, in the case of citrus fruit juices and skin cancer, furocoumarins were identified as intermediate phenotypes of interest. CUP Global collaborators at International Agency for Research on Cancer (IARC) then reviewed the evidence linking exposures, intermediate phenotypes, and outcomes. Finally, the CUP Global Expert Committee on Cancer Mechanisms (MEC) evaluated the review findings using a decision-support tool and agreed on biological plausibility statements.

Based on IARC’s review findings, the MEC did not identify strong evidence of biological mechanisms linking soft drinks and cancer through insulin- and glucose-related intermediate phenotypes (Fontvieille et al., unpublished). This was due to several limitations in the current literature including a general lack of longitudinal study designs, standardised definitions and measurements of (subtypes of) exposures and intermediate phenotypes, and adequate control for key confounders. The MEC also acknowledged that furocoumarins may be implicated in biological mechanisms linking citrus fruit juices (including orange juice) to skin cancer; however, further research is required to fully characterise these mechanisms and potential interaction between furocoumarins and sun exposure. Of note, furocoumarin concentrations are higher in grapefruit than in oranges. The lack of association for grapefruit juice may reflect low consumption and a high proportion of non-consumers. Furocoumarins are concentrated in the citrus peel, and levels may therefore differ between homemade juices, which exclude the peel, and industrially processed juices, which may incorporate it. Further research is needed to characterise regional variation in intake, processing effects, the stability of furocoumarins during heat treatment, and the associations of other furocoumarin-containing foods such as parsnips and celery.

### Text Box (Panel) 2: Grading the quality of evidence

The CUP Global Expert Panel interpreted and graded the quality of the evidence following pre-defined CUP Global evidence grading criteria designed to evaluate the likelihood of causality (**Supplementary Table 5)**. Evidence conclusions were made based on aspects related to the quantity, consistency, magnitude and precision of the summary estimates, presence of a dose-response relationship, study design and risk of bias, generalisability and mechanistic plausibility of the results from observational studies. Evidence grades were classified as convincing, probable, limited suggestive, limited no conclusion, or substantial effect on risk unlikely (indicating the likelihood of causality).

Based on evaluation by the CUP Global Expert Panel, there was evidence of a probable causal positive association between SSBs intake and pancreatic and colorectal cancers, and between citrus and orange juice and melanoma, as well as basal and squamous cell carcinomas of the skin, supported by consistent findings across studies, study size, risk of bias, and plausible biological mechanisms. Positive associations between SSBs and ovarian and endometrial cancers were observed but graded as limited suggestive owing to small numbers of studies and cases. For postmenopausal breast cancer, there was consistent evidence of a positive association from large cohort studies, but the CIs were wide and crossed the null value; thus, the expert panel concluded limited suggestive evidence. Evidence for other cancer sites was graded as limited no conclusion and/or substantial effects on risk unlikely, reflecting null findings, heterogeneity, small study numbers, risk of bias, and limited mechanistic support. Evidence based on a single study was classified as insufficient evidence to draw a conclusion For ASBs, most associations were graded as limited no conclusion, reflecting small numbers of studies or cases, evidence of no association, imprecise estimates, between-study heterogeneity, and concerns about bias, particularly reverse causation related to diabetes, adiposity, prior SSBs intake, or recent weight gain. Evidence for the positive associations between carbonated ASBs and leukaemia was graded as limited suggestive, based on the relatively strong magnitude of associations observed in large cohort studies with repeated dietary assessments and low risk of bias, thereby reducing concerns about reverse causation.

For juices, a pooled analysis of 15 cohorts showed a positive association between fruit and vegetable juices and prostate cancer; however, the evidence was graded as limited suggestive due to potential detection bias and limited mechanistic support. Most other associations for juices were graded as limited no conclusion reflecting small numbers of studies or cases, imprecise estimates, between-study heterogeneity, and risk of bias. An online interactive tool to help explore the evidence can be found at: https://teacup.cc.ic.ac.uk/soft-drinks-cancer.html. Evidence presented in this systematic literature review will form part of the Fourth Expert Report which will be developed by WCRF International in the coming years.

## Data Availability

Data availability statement: Only publicly available data were used in our study. Data sources and handling of these data are described in the materials and methods section. Data are extracted into the CUP Global database and further details are available from the corresponding author upon request.

## Acknowledgments

We thank Teresa Norat for leading the WCRF/AICR Continuous Update Project (CUP) as principal investigator from 2007 to 2020 and for developing the original review protocols. We thank the current and past WCRF International CUP/CUP Global Imperial team members for their contribution to the literature search, study selection, and/or data extraction; Lam Teng and past CUP/CUP Global Imperial database managers for implementing and updating the CUP Global database; and project manager: Eduardo Seleiro for organising the references, proofreading the manuscript and coordinating the work of CUP Global at Imperial College London. We also thank the CUP Global Expert Committee on Cancer Mechanisms (MEC) for evaluating and reviewing the findings of the mechanistic review.

## IARC disclaimer

Where authors are identified as personnel of the International Agency for Research on Cancer/World Health Organization, the authors alone are responsible for the views expressed in this article and they do not necessarily represent the decisions, policy, or views of the International Agency for Research on Cancer/World Health Organization.

## Data availability statement

Only publicly available data were used in our study. Data sources and handling of these data are described in the materials and methods section. Data are extracted into the CUP Global database and further details are available from the corresponding author upon request.

## Ethics statement

Ethical approval was not required, since this work was based solely on the analysis of published data without collection of new data or participant involvement.

