## Supplementary Texts 1-21; Supplementary Tables 1-24; Supplementary Figures 1-326 for "Sugar sweetened and artificially sweetened beverages, fruit and vegetable juices and cancer risk: a World Cancer Research Fund International Global Cancer Update Programme (CUP Global) systematic literature review and meta-analysis"

#### Supplementary materials

**Table of text**

**Table of tables**

Supplementary Table 24. List of exposure-outcome pairs included in the meta-analyses and corresponding evidence gradings (the table will be replaced by another table that will be prepared by WCRF). .... **Error! Bookmark not defined.**

**Table of figures**

Supplementary Figure 21. Highest versus the lowest category results from the individual studies on soft drinks and sugar sweetened beverages and colorectal cancer incidence.. 227

Supplementary Figure 22. Highest versus the lowest category results from the individual studies on soft drinks and sugar sweetened beverages and colorectal cancer mortality. .. 228

Supplementary Figure 53. Highest versus the lowest category results from individual studies on artificially sweetened beverages and hematological cancers incidence and mortality... 259

Supplementary Figure 120. Linear dose-response meta-analysis of sugar sweetened beverages and proximal colon cancer incidence by smoking status. 326

Supplementary Figure 121. Linear dose-response meta-analysis of sugar sweetened beverages and proximal colon cancer incidence by alcohol drinking status. 327

Supplementary Figure 122. Linear dose-response meta-analysis of sugar sweetened beverages and proximal colon cancer incidence by physical activity status. 328

Supplementary Figure 123. Linear dose-response meta-analysis of sugar sweetened beverages and proximal colon cancer incidence by body mass index. 329

Supplementary Figure 124. Linear dose-response meta-analysis of carbonated sugar sweetened beverages and colon cancer incidence by follow-up duration. 330

Supplementary Figure 125. Linear dose-response meta-analysis of carbonated sugar sweetened beverages and colon cancer incidence by geographical region. 331

Supplementary Figure 126. Linear dose-response meta-analysis of carbonated sugar sweetened beverages and colon cancer incidence by frequency of dietary assessment. 332

Supplementary Figure 127. Linear dose-response meta-analysis of carbonated sugar sweetened beverages and colon cancer incidence by various factors. 333

Supplementary Figure 128. Linear dose-response meta-analysis of sugar sweetened beverages and distal colon cancer incidence by frequency of dietary assessment. 334

Supplementary Figure 129. Linear dose-response meta-analysis of sugar sweetened beverages and rectal cancer incidence by frequency of dietary assessment. 335

Supplementary Figure 130. Linear dose-response meta-analysis of sugar sweetened beverages and liver cancer by outcome type. 336

Supplementary Figure 131. Linear dose-response meta-analysis of sugar sweetened beverages and liver cancer incidence and mortality combined by adjustment for body mass index. 337

Supplementary Figure 132. Linear dose-response meta-analysis of sugar sweetened beverages and liver cancer incidence by exclusion of people with a history of diabetes. 338

Supplementary Figure 133. Linear dose-response meta-analysis of sugar sweetened beverages and pancreatic cancer by outcome type. 339

Supplementary Figure 134. Linear dose-response meta-analysis of sugar sweetened beverages and pancreatic cancer incidence and mortality combined by body mass index. 340

Supplementary Figure 135. Linear dose-response meta-analysis of sugar sweetened beverages and pancreatic cancer incidence and mortality combined by sex. 341

Supplementary Figure 136. Linear dose-response meta-analysis of sugar sweetened beverages and pancreatic cancer incidence by smoking status. 342

Supplementary Figure 137. Linear dose-response meta-analysis of sugar sweetened beverages and pancreatic cancer incidence and mortality combined by adjustment for body mass index. 343

Supplementary Figure 138. Linear dose-response meta-analysis of sugar sweetened beverages and pancreatic cancer incidence by exclusion of people with diabetes at baseline of the study. 344

Supplementary Figure 139. Linear dose-response meta-analysis of carbonated sugar sweetened beverages and pancreatic cancer incidence, by various factors. 345

Supplementary Figure 140. Linear dose-response meta-analysis of sugar sweetened beverages and breast cancer incidence by geographical region. 346

Supplementary Figure 146. Linear dose-response meta-analysis of sugar sweetened beverages and postmenopausal breast cancer incidence by bias related to confounding. 352

Supplementary Figure 153. Linear dose-response meta-analysis of sugar sweetened beverages and kidney cancer incidence by exclusion of people with diabetes at baseline.359

Supplementary Figure 161. Linear dose-response meta-analysis of sugar sweetened beverages and liver cancer incidence, leave-one-out analysis. 367

Supplementary Figure 162. Linear dose-response meta-analysis of sugar sweetened beverages and pancreatic cancer incidence, leave-one-out analysis. 368

Supplementary Figure 163. Linear dose-response meta-analysis of sugar sweetened beverages and breast cancer incidence, leave-one-out analysis. 369

Supplementary Figure 164. Linear dose-response meta-analysis of sugar sweetened beverages and endometrial cancer incidence, leave-one-out analysis. 370

Supplementary Figure 165. Linear dose-response meta-analysis of sugar sweetened beverages and prostate cancer incidence, leave-one-out analysis. 371

Supplementary Figure 166. Linear dose-response meta-analysis of sugar sweetened beverages and kidney cancer incidence, leave-one-out analysis. 372

Supplementary Figure 167. Linear dose-response meta-analysis of carbonated sugar sweetened beverages and non-Hodgkin lymphoma incidence, leave-one-out analysis. 373

Supplementary Figure 168. Linear dose-response meta-analysis of sugar sweetened beverages and kidney cancer incidence, comparison of the models with and without adjustment for body mass index. 374

Supplementary Figure 169. Funnel plot of the association between sugar sweetened beverages, per 1 serving (355 mL)/day and colorectal cancer incidence. 375

Supplementary Figure 170. Linear dose-response meta-analysis of sugar sweetened beverages and colorectal cancer mortality. 376

Supplementary Figure 171. Linear dose-response meta-analysis of sugar sweetened beverages and colon and rectal cancer mortality. 377

Supplementary Figure 172. Linear dose-response meta-analysis of sugar sweetened beverages and kidney cancer mortality. 378

Supplementary Figure 173. Linear dose-response meta-analysis of sugar sweetened beverages and lung cancer mortality. 379

Supplementary Figure 174. Linear dose-response meta-analysis of sugar sweetened beverages and breast cancer mortality. 380

Supplementary Figure 175. Linear dose-response meta-analysis of sugar sweetened beverages and prostate cancer mortality. 381

Supplementary Figure 176. Linear dose-response meta-analysis of sugar sweetened beverages and kidney cancer mortality by sex. 382

Supplementary Figure 177. Linear dose-response meta-analysis of sugar sweetened beverages and colorectal cancer mortality by frequency of dietary assessment. 383

Supplementary Figure 178. Linear dose-response meta-analysis of sugar sweetened beverages and colorectal cancer mortality by geographical region. 384

Supplementary Figure 179. Linear dose-response meta-analysis of sugar sweetened beverages and colorectal cancer mortality by bias related to confounding. 385

Supplementary Figure 180. Linear dose-response meta-analysis of sugar sweetened beverages and colorectal cancer mortality by bias related to departure from intended exposure. 386

Supplementary Figure 181. Linear dose-response meta-analysis of sugar sweetened beverages and breast cancer mortality by frequency of dietary assessment. 387

Supplementary Figure 182. Linear dose-response meta-analysis of sugar sweetened beverages and breast cancer mortality by exclusion of people with diabetes at baseline. . 388

Supplementary Figure 214. Linear dose-response meta-analysis of artificially sweetened beverages and pancreatic cancer incidence and mortality combined by body mass index.420

Supplementary Figure 227. Linear dose-response meta-analysis of artificially sweetened beverages and postmenopausal endometrial cancer incidence, leave-one-out analysis. .. 433

Supplementary Figure 312. Linear dose-response meta-analysis of soft drinks and lung cancer incidence. 518

Supplementary Figure 313. Linear dose-response meta-analysis of soft drinks and breast cancer incidence. 519

Supplementary Figure 314. Linear dose-response meta-analysis of soft drinks and endometrial cancer incidence. 520

Supplementary Figure 315. Linear dose-response meta-analysis of carbonated soft drinks and kidney cancer incidence. 521

Supplementary Figure 316. Linear dose-response meta-analysis of soft drinks and bladder cancer incidence. 522

Supplementary Figure 317. Linear dose-response meta-analysis of carbonated and caffeinated soft drinks and glioma incidence. 523

Supplementary Figure 318. Linear dose-response meta-analysis of carbonated and decaffeinated soft drinks and glioma incidence. 524

Supplementary Figure 319. Linear dose-response meta-analysis of soft drinks and pancreatic cancer incidence by geographical region. 525

Supplementary Figure 320. Linear dose-response meta-analysis of soft drinks and pancreatic cancer incidence by follow-up duration. 526

Supplementary Figure 321. Linear dose-response meta-analysis of soft drinks and liver cancer incidence by geographical region. 527

Supplementary Figure 322. Linear dose-response meta-analysis of carbonated soft drinks and kidney cancer incidence by sex. 528

Supplementary Figure 323. Linear dose-response meta-analysis of soft drinks and liver cancer incidence, leave-one-out analysis. 529

Supplementary Figure 324. Linear dose-response meta-analysis of soft drinks and pancreatic cancer incidence, leave-one-out analysis. 530

Supplementary Figure 325. Linear dose-response meta-analysis of carbonated soft drinks and kidney cancer incidence, leave-one-out analysis. 531

Supplementary Figure 326. Observational studies risk of bias assessment. 532

**Supplementary Table 1. PRISMA checklist.**

| Section and Topic | Item # | Checklist item | Location where item is reported (page #) |
|---|---|---|---|
| TITLE | | | |
| Title | 1 | Identify the report as a systematic review. | 1 |
| ABSTRACT | | | |
| Abstract | 2 | See the PRISMA 2020 for Abstracts checklist. | NA |
| INTRODUCTION | | | |
| Rationale | 3 | Describe the rationale for the review in the context of existing knowledge. | 6 |
| Objectives | 4 | Provide an explicit statement of the objective(s) or question(s) the review addresses. | 6 |
| METHODS | | | |
| Eligibility criteria | 5 | Specify the inclusion and exclusion criteria for the review and how studies were grouped for the syntheses. | 7 |
| Information sources | 6 | Specify all databases, registers, websites, organisations, reference lists and other sources searched or consulted to identify studies. Specify the date when each source was last searched or consulted. | 7 |
| Search strategy | 7 | Present the full search strategies for all databases, registers and websites, including any filters and limits used. | 7 |
| Selection process | 8 | Specify the methods used to decide whether a study met the inclusion criteria of the review, including how many reviewers screened each record and each report retrieved, whether they worked independently, and if applicable, details of automation tools used in the process. | 7 / 9 |
| Data collection process | 9 | Specify the methods used to collect data from reports, including how many reviewers collected data from each report, whether they worked independently, any processes for obtaining or confirming data from study investigators, and if applicable, details of automation tools used in the process. | 7 / 9 |
| Data items | 10a | List and define all outcomes for which data were sought. Specify whether all results that were compatible with each outcome domain in each study were sought (e.g. for all measures, time points, analyses), and if not, the methods used to decide which results to collect. | 7 / Supplementary Table 2 |

| Section and Topic | Item # | Checklist item | Location where item is reported (page #) |
|---|---|---|---|
| | 10b | List and define all other variables for which data were sought (e.g. participant and intervention characteristics, funding sources). Describe any assumptions made about any missing or unclear information. | 7 / Supplementary tables 11-14 |
| Study risk of bias assessment | 11 | Specify the methods used to assess risk of bias in the included studies, including details of the tool(s) used, how many reviewers assessed each study and whether they worked independently, and if applicable, details of automation tools used in the process. | 7-8 |
| Effect measures | 12 | Specify for each outcome the effect measure(s) (e.g. risk ratio, mean difference) used in the synthesis or presentation of results. | 8-9 |
| Synthesis methods | 13a | Describe the processes used to decide which studies were eligible for each synthesis (e.g. tabulating the study intervention characteristics and comparing against the planned groups for each synthesis (item #5)). | 8-9 |
| | 13b | Describe any methods required to prepare the data for presentation or synthesis, such as handling of missing summary statistics, or data conversions. | 8-9 |
| | 13c | Describe any methods used to tabulate or visually display results of individual studies and syntheses. | 9 |
| | 13d | Describe any methods used to synthesize results and provide a rationale for the choice(s). If meta-analysis was performed, describe the model(s), method(s) to identify the presence and extent of statistical heterogeneity, and software package(s) used. | 8-9 |
| | 13e | Describe any methods used to explore possible causes of heterogeneity among study results (e.g. subgroup analysis, meta-regression). | 8-9 |
| | 13f | Describe any sensitivity analyses conducted to assess robustness of the synthesized results. | 8-9 |
| Reporting bias assessment | 14 | Describe any methods used to assess risk of bias due to missing results in a synthesis (arising from reporting biases). | 9 |
| Certainty assessment | 15 | Describe any methods used to assess certainty (or confidence) in the body of evidence for an outcome. | 9 |
| RESULTS | | | |
| Study selection | 16a | Describe the results of the search and selection process, from the number of records identified in the search to the number of studies included in the review, ideally using a flow diagram. | 9 |
| | 16b | Cite studies that might appear to meet the inclusion criteria, but which were excluded, and explain why they were excluded. | 9 |
| Study characteristics | 17 | Cite each included study and present its characteristics. | 10 / 16-17 |

| Section and Topic | Item # | Checklist item | Location where item is reported (page #) |
|---|---|---|---|
| Risk of bias in studies | 18 | Present assessments of risk of bias for each included study. | 16 |
| Results of individual studies | 19 | For all outcomes, present, for each study: (a) summary statistics for each group (where appropriate) and (b) an effect estimates and its precision (e.g. confidence/credible interval), ideally using structured tables or plots. | 10-17 |
| Results of syntheses | 20a | For each synthesis, briefly summarise the characteristics and risk of bias among contributing studies. | 10-17 |
| | 20b | Present results of all statistical syntheses conducted. If meta-analysis was done, present for each the summary estimate and its precision (e.g. confidence/credible interval) and measures of statistical heterogeneity. If comparing groups, describe the direction of the effect. | 10-17 |
| | 20c | Present results of all investigations of possible causes of heterogeneity among study results. | 10-17 |
| | 20d | Present results of all sensitivity analyses conducted to assess the robustness of the synthesized results. | 10-17 |
| Reporting biases | 21 | Present assessments of risk of bias due to missing results (arising from reporting biases) for each synthesis assessed. | 16 |
| Certainty of evidence | 22 | Present assessments of certainty (or confidence) in the body of evidence for each outcome assessed. | To add |
| DISCUSSION | | | |
| Discussion | 23a | Provide a general interpretation of the results in the context of other evidence. | 17-20 |
| | 23b | Discuss any limitations of the evidence included in the review. | 18-19 |
| | 23c | Discuss any limitations of the review processes used. | 18-19 |
| | 23d | Discuss implications of the results for practice, policy, and future research. | 19-20 |
| OTHER INFORMATION | | | |
| Registration and protocol | 24a | Provide registration information for the review, including register name and registration number, or state that the review was not registered. | 7 |
| | 24b | Indicate where the review protocol can be accessed, or state that a protocol was not prepared. | 7 |
| | 24c | Describe and explain any amendments to information provided at registration or in the protocol. | |

| Section and Topic | Item # | Checklist item | Location where item is reported (page #) |
| --- | --- | --- | --- |
| Support | 25 | Describe sources of financial or non-financial support for the review, and the role of the funders or sponsors in the review. | |
| Competing interests | 26 | Declare any competing interests of review authors. | |
| Availability of data, code and other materials | 27 | Report which of the following are publicly available and where they can be found: template data collection forms; data extracted from included studies; data used for all analyses; analytic code; any other materials used in the review. | |

**Supplementary Text 1. Search strategy to find potential eligible studies.**

#### A. PubMed

**a. Searching for studies on cancer**

| | |
|---|---|
| 1 | "Neoplasms"[MeSH Terms] |
| 2 | Neoplasm*[Title/Abstract] OR cancer*[Title/Abstract] OR carcinoma*[Title/Abstract] OR tumour*[Title/Abstract] OR tumor*[Title/Abstract] OR malignancy[Title/Abstract] OR malignant[Title/Abstract] OR adenocarcinoma[Title/Abstract] |
| 3 | leukemia[Title/Abstract] OR lymphoma[Title/Abstract] OR myeloma[Title/Abstract] |
| 4 | melanoma[Title/Abstract] |
| 5 | glioma[Title/Abstract] |
| 6 | #1 OR #2 OR #3 OR #4 OR #5 |

**b. Searching for studies on soft drinks***

| | |
|---|---|
| 7 | "Carbonated Beverages"[MeSH Terms] OR "Artificially Sweetened Beverages"[MeSH Terms] OR "Sugar-Sweetened Beverages"[MeSH Terms] OR "Fruit and Vegetable Juices"[MeSH Terms] OR "beverage*"[Text Word] OR "sugar* drink*"[Text Word] OR "sugar containing drink*"[Text Word] OR "carbonated drink*"[Text Word] OR "soft drink*"[Text Word] OR "energy drink*"[Text Word] OR "sweet* drink*"[Text Word] OR "nonalcoholic drink*"[Text Word] OR "non-alcoholic drink*"[Text Word] OR "Soda"[Text Word] OR "juice*"[Text Word] OR "sugar* food*"[Text Word] OR "Added sugar"[Text Word] OR "fruit drink*"[Text Word] |
| 8 | "Dietary Approaches To Stop Hypertension"[MeSH Terms] OR "food, processed"[MeSH Terms] OR "diabetes risk reduction diet*"[Text Word] OR "type 2 diabetes prevention diet*"[Text Word] OR "healthy eating index"[Text Word] OR "plant based diet* index"[Text Word] OR "ultra processed food*"[Text Word] OR "ultraprocessed food*"[Text Word] OR "world cancer research fund international"[Text Word] OR "WCRF"[Text Word] OR "WCRF/AICR"[Text Word] OR "recommended food score"[Text Word] OR "diet* index*"[Text Word] |
| 9 | #7 OR #8 |

*As a main exposure or a component within a dietary pattern.

**c. Combining the searches**

| | |
|---|---|
| 10 | #6 AND #9 |

**d. Limiting to human studies**

| | |
|---|---|
| 11 | animal [MeSH Terms] NOT human [MeSH Terms] |
| 12 | #10 NOT #11 |

#### B. Embase (Ovid)

**a. Searching for studies on cancer**

| | |
|---|---|
| 1 | exp neoplasm/ |
| 2 | (Cancer* or carcinoma* or tumour* or tumor* or malignancy or malignant or adenocarcinoma or leukemia or lymphoma or myeloma or melanoma or glioma).ab,ti. |
| 3 | 1 or 2 |

**b. Searching for studies on soft drinks***

| | |
|---|---|
| 4 | exp soft drink/ or exp artificially sweetened beverage/ or exp carbonated beverage/ or exp fruit juice/ or exp sweetened beverage/ or exp energy drink/ |
| 5 | sweet drink.mp. or soda.mp. |
| 6 | exp mango juice/ or exp fruit juice/ or exp orange juice/ or exp pear juice/ or juice.mp. or exp berry juice/ or exp grape juice/ or exp apple juice/ or exp lemon juice/ |

| | |
|---|---|
| 7 | ("Beverage*" or "sugar* drink*" or "carbonated drink*" or "soft drink*" or "energy drink*" or "sweet* drink*" or "nonalcoholic drink*" or "non-alcoholic drink*" or soda or "juice*" or "sugar* food*" or "added sugar" or "fruit drink*").tw. |
| 8 | exp DASH diet/ or exp ultra-processed food/ |
| 9 | ("Dietary Approaches to Stop Hypertension" or "DASH diet*" or "ultra-processed food*" or "ultraprocessed food*" or "diabetes risk reduction diet" or "type 2 diabetes prevention diet" or "healthy eating index" or "world cancer research fund international" or "WCRF" or "WCRF/AICR" or "plant-based diet* index" or "recommended food score" or "diet* index").tw. |
| 10 | 4 or 5 or 6 or 7 or 8 or 9 |

*As a main exposure or a component within a dietary pattern.

**c. Combining the searches**

| | |
|---|---|
| 11 | 3 and 10 |

**d. Limiting to human studies**

| | |
|---|---|
| 12 | exp animal/ not exp human/ |
| 13 | 11 not 12 |

**e. Limiting to certain publication types**

| | |
|---|---|
| 14 | limit 13 to (article or article in press or conference paper or "conference review" or editorial or erratum or "review" or tombstone) |

#### Supplementary Text 2. Systematic search and study selection criteria

A systematic literature search was performed in PubMed and Embase until 1st September 2024, using a pre-specified search strategy (**Supplementary Text 1**). We also checked the reference lists of all relevant original cohorts and related reviews. Selection criteria included the following: 1) randomised controlled trials (RCTs), observational cohort studies, or pooled analyses of such studies or Mendelian randomisation (MR) studies; 2) studies involving individuals not diagnosed with cancer (excluding non-melanoma skin cancer) at the time of intervention or exposure evaluation; 3) studies assessing the intake of soft drinks or juices as an exposure; 4) studies reporting on the incidence or mortality of primary cancer at any anatomical site during adulthood (a non-exhaustive list of cancer outcomes considered eligible for the present SLR is provided in **Supplementary Table 2**); and 5) studies providing multivariable adjusted estimates expressed as hazard ratios, risk ratios, relative risks (RR), or odds ratios. For publications that reported results on the same exposure-outcome associations from the same cohort, we selected those with sufficient information for inclusion in the dose-response meta-analyses, otherwise, we selected those with the highest number of cancer cases. Pooled analyses were preferred over the individual articles published by their component studies. If available, we used study-specific estimates included in the pooled analyses.

#### Supplementary Text 3. Definition and classification of soft drinks used in the present systematic review.

The included studies investigated a wide variety of soft drinks and juices. We systematically reviewed and meta-analysed comparable exposures based on information provided in the studies, including 1) soft drinks (unspecified sweetened drinks, likely a combination of sugar sweetened beverages (SSBs) and artificially sweetened beverages (ASBs)); 2) SSBs; 3) ASBs; 4) fruit and vegetable juices; 5) fruit juices; and 6) vegetable juices. Specific types of soft drinks, such as carbonated or non-carbonated, sugar sweetened or artificially sweetened, soda or fruit drinks, pure fruit juices, or specific fruit juices, were analysed separately. Each reported exposure was reviewed only once, however, additional analyses were conducted combining SSBs with carbonated SSBs, ASBs with carbonated ASBs, fruit juice with pure fruit juice, and citrus juices with orange juice, as these constitute the predominant components of their respective beverage categories.

#### Soft drinks

Non-alcoholic sweetened beverages that include various types of both SSBs and ASBs. Studies that reported on unspecified soft drinks and did not differentiate between SSBs and ASBs and so likely included both were classified into this group. In addition, studies that reported on soft or sweetened drinks and did not report additional information regarding the type of sweeteners were classified into this category. Based on information provided in the included studies, beverages in this category were classified as 1) soft drinks, 2) carbonated soft drinks (e.g., soda), 3) caffeinated carbonated soft drinks, 4) decaffeinated carbonated soft drinks, and 5) cola.

##### Sugar sweetened beverages

Liquids that are sweetened by adding free sugars, such as sucrose, high fructose corn syrup and sugars naturally present in honey, syrups, fruit juices and fruit juice concentrate. This includes, among others, sodas/carbonated drinks, sports drinks, energy drinks, sweetened waters, cordials, barley water, and coffee- and tea-based beverages with sugars or syrups added. This does not include versions of these drinks which are ‘sugar-free’ or sweetened only with artificial sweeteners. Based on information provided in the included studies, beverages in this category were classified as 1) SSBs (that could be with or without fruit juices), 2) carbonated SSBs, 3) non-carbonated SSBs, 4) sugar-sweetened cola, 5) fruit drinks, and 6) carbonated fruit drinks.

##### Artificially sweetened beverages

Low calorie or non-caloric beverages including among others, sodas/carbonated drinks, sports drinks, energy drinks, sweetened waters, cordials, barley water, and coffee- and tea-based beverages sweetened with artificial sweeteners, such as aspartame, acesulfame K, saccharin, sucralose, or neotame. These drinks are often called ‘low sugar’, ‘sugar-free’ or ‘diet’ drinks. Based on information provided in the included studies, beverages in this category were classified as 1) ASBs, 2) carbonated ASBs, 3) non-carbonated ASBs, 4) artificially sweetened cola, and 5) artificially sweetened fruit drinks.

##### Fruit or vegetable juices

Fruit or vegetable juices are liquids expressed from materials of a fruit or vegetable plant by crushing, comminuting and pressing. It can be clear, cloudy or pulpy from a single or various plants. 100% pure juices do not have additives, such as sugars, sweeteners, preservatives, flavourings, or colourings. Nectars, among other ingredients such as water and sugar, contain about 25%-50% fruit juice or pulp by weight; juice drinks, beverages, or cocktails contain about 10%-20% juice; and juice punches contain about 1% juice. Based on information provided in the included studies, beverages in this category were classified as 1) fruit and vegetable juices, 2) fruit juices, 3) 100% pure fruit juices, 4) apple juice, 5) citrus fruit juice, 6) orange juice, 7) grapefruit juice, 8) vegetable juices, 9) tomato juice, and 10) carrot juice. Studies that expressed the exposure as juice and did not report additional information regarding the type of juice, were classified as fruits and vegetables juices. For fruit juices to be classified as pure, studies must have provided a clear indication that the juices are labelled as pure, fresh, 100%, or natural. In the absence of such explicit statements, these juices were categorised as fruit juices, which includes a mixture of pure and commercially produced fruit juices.

**Supplementary Text 4. Risk of bias assessment of observational cohort studies.**

We used a modified version of the Risk of Bias for Nutrition Observational Studies (RoB-NObs) tool to assess the risk of bias of the studies included in the meta-analyses[21]. The U.S Department of Agriculture (USDA) Nutrition Evidence Systematic Review initially developed RoB-NObs based on a modification of the Cochrane's Risk of bias In Non-randomised Studies of Interventions (ROBINS-I)[22]. The CUP Global team at Imperial College London further adapted and tested the tool, incorporating additional prompting questions and guidance to assess risk of bias in observational studies investigating lifestyle-disease associations focusing on adiposity, physical activity, sedentary behaviour, and dietary/nutritional exposures. The RoB domains and criteria to rate studies in relation to each domain are listed in **Supplementary Tables 3-4**.

#### Supplementary Text 5. Detailed information for selection of statistical models for inclusion in the meta-analysis.

For inclusion in the meta-analyses, a model was considered multivariable if it adjusted for at least two variables in addition to the exposure of interest. Models adjusted for age or sex only were not considered multivariable. We aimed to include the estimated from multivariable maximally adjusted models in the meta-analyses. However, in the associations between SSBs and cancer risk, body mass index (BMI) and other adiposity measures (e.g., waist circumference, waist-to-hip ratio) were considered mediators in the causal pathway between exposures and cancer incidence. Since the handling of adiposity measures varied across studies, we adopted a structured approach to model selection and sensitivity analyses to address potential mediator adjustment bias.

Primary studies were categorised based on how they handled adiposity measures in their reported multivariable models: (1) studies could report a single multivariable-adjusted model that excluded adiposity measures. These estimates were considered the most appropriate for inclusion in the primary meta-analysis, as these models avoided adjusting for mediators and captured the total effect of the exposure; (2) studies could report only a multivariable-adjusted model that included adiposity measures. These results were also included in the meta-analysis when no alternative multivariable estimates were available. Although adjusting for mediators (e.g., BMI) may attenuate the observed associations, these studies were retained to avoid omission of relevant studies; and (3) for studies presenting both types of models, we prioritised the mediator-unadjusted estimates (i.e., models without adiposity adjustment) as the most appropriate for inclusion in the primary meta-analysis. A similar approach was applied for ASBs, but we included multivariable-adjusted models that included adiposity measures in the model as there is evidence of a reverse causality bias in the association between ASBs and health outcomes. For soft drinks that includes SSBs, ASBs, and juices, we selected the models that did not control for adiposity measures as the major components of soft drinks are SSBs+juices. However, few studies reported data on both adiposity-adjusted and unadjusted models for soft drinks.

Additionally, we compared the adiposity-unadjusted and adiposity-adjusted estimates, focusing on the magnitude of the estimates and the statistical significance of the associations. This approach aimed to evaluate the potential bias introduced by adjusting for mediator variables. In cases where large qualitative differences were observed (e.g., changes in statistical significance between the two models), sensitivity meta-analyses were performed using the adiposity-adjusted estimates.

| Multivariable-adjusted model(s) reported in the studies | | SSBs/Juices | ASBs |
|---|---|---|---|
| One model (without adiposity measures) | | ✓ | ✓* |
| One model (with adiposity measures) | | ✓* | ✓ |
| Two models** | Without adiposity measures | ✓ | × |
| | With adiposity measures | × | ✓ |

*To avoid omission of studies in the analysis.

**Subgroup and sensitivity analyses are conducted to compare the results between the models.

**Supplementary Text 6. Details of information extracted from Mendelian randomisation studies.**

Mendelian randomisation studies using genetic variants derived from genome-wide association studies (GWAS) as instrumental variables to assess associations of genetically predicted self-reported consumption of soft drinks and juices and cancer risk were reviewed. The following information was extracted: the study design (one- or two-sample MR), exposure and outcome GWAS data sources, population ancestries and sample sizes, information on genetic instruments including number of single nucleotide polymorphisms (SNPs) associated with soft drinks or juices utilised in the analyses and variance explained ($R^2$) by the instrument, primary analysis (e.g., fixed or random effects inverse-variance weighted [IVW] method), sensitivity MR analyses (e.g., MR-Egger, weighted median, MR-PRESSO), and any multivariable MR analyses. When multiple analyses on overlapping exposure and outcome populations from different publications were identified, the analysis utilising the largest outcome sample size (reflecting the largest power) was considered as the primary MR analysis, while the rest were also kept and checked for consistency of results against the main analysis. Results from the main MR analyses were compared with the sensitivity analyses to assess concordance and robustness.

#### Supplementary Text 7. Detailed statistical methods used for dose-response meta-analysis.

Our primary analyses were linear dose-response meta-analyses when at least two primary studies with per unit exposure increment or three or more exposure categories provided sufficient information. Non-linear dose-response meta-analyses were conducted when at least five primary studies provided sufficient information. Hazard ratios, RRs, and odds ratios were considered equivalent due to the low absolute risk of cancer and were combined into a summary RR. When studies reported results by subpopulations (e.g., males and females) but not overall, a fixed effect model was used to estimate an overall association before inclusion of the study in the meta-analysis. Hamling's method was used to pool results reported by cancer subtypes (e.g., colon and rectal cancer) to account for common denominators in risk estimates[23]. When dose-response meta-analysis was not possible or when more than a third of the included studies did not have sufficient information for inclusion in a dose-response meta-analysis, a supporting categorical high versus low analysis was conducted. When a meta-analysis was not possible or for individual studies that could not be meta-analysed, the studies were descriptively reviewed. This approach allows a comprehensive summation of study results while minimising the number of categorical meta-analysis that may pool results for different exposure comparisons. Separate meta-analyses were conducted for incidence (primary) and mortality (secondary) outcomes. For cancers with high mortality rate including pancreas, liver, lung, and gallbladder cancers, we combined incidence and mortality outcomes in an additional analysis. For descriptive synthesis, the results for each exposure-outcome association reported in the individual studies were presented in supplementary tables and reviewed in the text.

Linear dose-response meta-analyses were conducted when there were at least two independent studies reporting results on the same exposure-outcome association. Both fixed effect and inverse variance DerSimonian-Laird random-effects were conducted to summarise the results from the included studies. Important differences, for instance influence from small studies on the summary risk estimates from the two models, were investigated and reported appropriately; otherwise, summary risk estimate from the random-effects model were reported only.

Linear dose-response meta-analysis were conducted to allow expressing the results of each study in the same increment unit for a given exposure. An increment unit consistent with the previous CUP SLRs were used; otherwise, an increment unit that reflects the usual unit of exposure (e.g., measures that used in most of the included studies) were used, including:

- 355 mL (12 US fluid oz)/day for soft drinks, SSBs, and ASBs[25]
- 177 ml (6 US fluid oz)/day for fruit or vegetable juices[26]

The measures of association, including hazard ratio, risk ratio, and odds ratio, derived from the individual studies were aggregated and referred to collectively as the summary relative risk. These measures were roughly equivalent in research examining the current topic, based on the assumption that cancer cases are infrequent, and the absolute risks of cancer are minimal. To be included in a linear dose-response meta-analysis, studies must have reported the following data:

- For studies investigating cancer risk associated with a continuous exposure variable: relative risk and its measure of variance (standard errors (SE), 95% confidence intervals (CIs) or P-value) per unit increment in exposure that can be readily converted or re-scaled if needed. In cases where the units of measurement differed between results the units were converted, where possible, such that all results used the same measurement.

- For studies investigating cancer risk associated with a categorical comparison of exposure variable: relative risk and its measure of variance (as above), number of cases, person-years or number of non-cases, exposure categories (cut-offs, mean or median) for at least three exposure level comparisons. For studies reporting intakes in unit/1000 kcal/day, the intake in unit/day were estimated using the average energy intake for each exposure category, or if not provided, the average energy intake overall reported in the article. For each exposure category, we assigned the midpoint of the lower and upper boundaries. For open-ended highest categories, we assumed that the category width was the same as that of the adjacent lower category and assigned the midpoint accordingly. For open-ended lowest categories, we assigned the midpoint using the available boundary and the width of the adjacent category; where the lower bound was not reported, it was assumed to be zero. Standard approaches were used to derive missing data (e.g., number of participants or cases across categories) for meta-analysis [209,210].

The dose-response slope of the association estimated only from a categorical analysis in a study were derived using generalised least-squares for trend estimation. When the mean or median intake of the reference intake category (relative risk = 1) was not zero, all the association estimates assigned to exposure categories were rescaled so that the reference point was always zero. Studies of both sexes combined or of single sex only were first be meta-analysed together, follows by subgroup analysis by sex. When a study did not report an overall result but results of the sub-populations (e.g. males and females, smokers and non-smokers) only, a fixed-effect model was used to estimate an overall result before pooling with the other studies in the meta-analysis. Similarly, studies reporting results only by cancer subtype (e.g., colon and rectal) were first pooled before meta-analysing with the other studies. In this case, the Hamling's method that accounts for the common denominators used in calculating the risk estimates for the outcomes was used[23]. When results from multiple studies are reported in the same publication, e.g. in a pooled analysis, the results of each component study were included separately if they were provided. The pooled result was not used directly, if possible, to maintain the independence of observations included and to allow exploration of heterogeneity across study results.

Pre-specified subgroup analyses included participant's characteristics (sex, race/ethnicity, BMI status, physical activity level, smoking status, alcohol drinking status, and inclusion or exclusion of people with a history of diabetes at baseline) and study characteristics (geographical location, publication year, study size, length of follow-up, frequency of dietary assessment and risk of bias domains). In case where primary studies reported both adiposity adjusted and adiposity unadjusted estimates that resulted in differences in the inference, we conducted a sensitivity analysis by replacing the adiposity-unadjusted estimate with the adiposity-adjusted one, to assess its impact on the summary estimate.

We conducted additional analyses combining SSBs with carbonated SSBs, and ASBs with carbonated ASBs, as these constitute the predominant components of their respective beverage categories and this approach would increase statistical power. We also combined orange juice with overall citrus fruit juice since orange juice is a good surrogate for citrus juice. We did not combine grapefruit juice with citrus juice due to generally low grapefruit intake. A similar approach was used for pure fruit juice and fruit juice. For studies that provided estimates for both overall SSBs and carbonated SSBs, only the overall SSBs estimate was used to avoid double counting of study participants. A similar approach was used when combining other exposure groups. Therefore, additional analyses included:

- Combining SSBs and carbonated SSBs
- Combining ASBs and carbonated ASBs
- Combining fruit juice and pure fruit juice
- Combining citrus juice and orange juice

Non-linear dose-response meta-analyses were conducted in a one-stage mixed effects models[29] using restricted cubic splines with three knots placed at fixed percentiles (10th, 50th, and 90th) of the exposure distribution[31]. The number of studies was chosen as few data points may drive the estimation of the non-linear curve with wide 95% confidence interval.

#### Supplementary Text 8. Descriptive review of sugar sweetened beverages and cancer incidence.

Forest plots showing study-specific results for the highest versus lowest exposure level comparisons were presented in **Supplementary Figures 18-41.** One study that could not be included in the highest versus lowest category meta-analysis of bladder cancer reported little evidence of an association between SSBs and fruit juice consumption and bladder cancer in males and females per each 2 points increase in the Alternative Healthy Eating Index-2010 component score[82]. One pooled analysis of NHS and HPFS reported that substituting 1 serving of SSBs per day with an equivalent amount of ASBs was not associated with distal colon or rectal cancers[178]. Overall, there were 11 publications that reported on SSBs, carbonated SSBs or fruit drinks and colorectal cancer (n=2), colon cancer (n=1), liver cancer (n=2), pancreatic cancer (n=2), lung cancer (n=1), breast cancer (n=1), postmenopausal breast cancer (n=2), premenopausal breast cancer (n=1), and prostate cancer (n=1) that could not be included in the meta-analyses because of insufficient data format[37,47,78,80,105,122,130,146,169,187,188]. In general, the results were null or borderline significant, except for SUN cohort study[146] that reported an increased risk of postmenopausal breast cancer incidence with higher intake of SSBs ($RR_{0.05 \text{ vs } 0 \text{ serving/day:}}$ 2.12, 95%CI: 1.01-4.41) (**Supplementary Figure 32**).

Twenty-four exposure-outcome associations were only reported by one study[34,36,47,70,72,74,75,84,90,128,135,172,186,211] (**Supplementary Table 16**). There was a positive association between SSB and early-onset colorectal cancer in the NHS II ($RR_{\geq 2 \text{ servings/day vs } <0.9 \text{ serving/week}}$: 2.18, 95%CI: 1.10-4.35)[72]. Of note, substitution of SSBs with ASBs was associated with a lower risk of early-onset colorectal cancer in the NHS II ($RR_{\text{per 1 serving/day}}$: 0.83, 95%CI: 0.69-0.99), but substitution of SSBs with fruit juice was not[72]. In addition, lower intake of SSBs, as part of the WCRF/AICR cancer prevention recommendation score, was associated with a lower risk of ER+/PR+ breast cancer in the BWHS[128], whereas the rest showed null associations between SSBs, fruit drinks or sugar sweetened cola and various cancers. One publication from the NHS II suggested that SSB intake during adolescence (13-18 years old) may be associated with a higher risk of early-onset colorectal cancer ($RR_{\geq 2 \text{ servings/day vs } <0.9 \text{ serving/week}}$: 3.41, 95%CI: 1.08-10.80)[72]. Intake of SSBs was also associated with a higher risk of symptomatic prostate cancer incidence in Malmo Diet and Cancer Study (MDCS) ($RR_{Q5 \text{ vs } Q1}$: 1.38, 95%CI: 1.04-1.84) [47], but not with low- or high-risk prostate cancer incidence. The Melbourne Collaborative Cohort Study also indicated little evidence of an association between SSBs and aggressive prostate cancer incidence[70].

#### Supplementary Text 9. Meta-analyses of sugar sweetened beverages and cancer mortality.

There was a positive association between SSBs and colorectal cancer (RR: 1.02, 95%CI=1.00-1.04; $I^2$=0) (4 studies, 3 publication, 16,855 cases)[113,126,178] **(Supplementary Figure 170),** proximal colon cancer (RR: 1.38, 95%CI=1.11-1.71; $I^2$=0) (2 studies, 1 publication, 409 cases)[178] (**Supplementary Figure 171**) and kidney cancer (RR: 1.06, 95%CI=1.02-1.10; $I^2$=0) (2 studies, 2 publications, 3394 cases)[66,113] (**Supplementary Figure 172**). Substitution of one serving/day of SSBs with ASBs was associated with a lower risk of proximal colon cancer (RR: 0.80, 95%CI=0.71-0.95) (2 studies, 1 publication, 409 cases)[178] (**Supplementary Figure 83**). Little evidence of an association was observed for lung, breast, and prostate cancer mortality (**Supplementary Figures 173-175).**

There was no evidence of subgroup differences in the small number of associations that could be examined (**Supplementary Figure 176-186**). In the influence analyses, the main results did not change excluding in turn each individual study (**Supplementary Figures 187-189**). The exception was SSBs and breast cancer which became evident after exclusion of the Cancer Prevention Study II that accounted for ~54% of the meta-analysis weight[113] (RR: 1.10, 95%CI=1.02-1.19) (**Supplementary Figure 187**). Forest plots showing study-specific results for the highest versus lowest exposure level comparisons were presented in **Supplementary Figures 18-41,** with descriptive synthesis and single-study associations detailed in **Supplementary Text 10** and **Supplementary Table 16,** respectively**.**

**Supplementary Text 10. Descriptive review of sugar sweetened beverages and cancer mortality.**

Forest plots showing study-specific results for the highest versus lowest exposure level comparisons were presented in **Supplementary Figures 18-41.** There was one publication from NIH-AARP that could not be included in the meta-analysis of prostate cancer mortality and reported little evidence of an association with SSBs, as part of the AHEI-2010 score[42]. One publication from CPS II reported on SSBs and 12 cancer mortality outcomes for which only one association estimate was available (**Supplementary Table 16**). There was a positive association between and SSBs and non-Hodgkin lymphoma mortality ($RR_{\geq 2 \text{ vs } 0\text{-}0.9 \text{ drinks/day}}$: 1.16, 95%CI: 1.05-1.28) (**Supplementary Figure 41**), while the rest showed no association.

#### Supplementary Text 11. Descriptive review of artificially sweetened beverages and cancer incidence.

Forest plots showing study-specific results for the highest versus lowest exposure level comparison are presented in **Supplementary Figures 42-54.** There was a positive association between carbonated ASBs and multiple myeloma and non-Hodgkin lymphoma in the HPFS[150] (**Supplementary Figure 53**), however, the linear dose-response meta-analyses indicated null associations. Other results were generally null and consistent with the meta-analyses. Two studies reported on prostate cancer[44,70], which could not be combined since one study reported an implausible association estimate[44]. Neither of the studies found an association with prostate cancer. The Malborne Collaborative Cohort Study (MCCS)[70] reported no association for the highest versus the lowest category of ASBs intake and aggressive prostate cancer incidence (RR: 0.81, 95%CI: 0.49-1.33), and the NutriNet-Santé study[44] in France reported an inverse association with 95%CI crossing the null value ($RR_{\text{per 10 mL/day}}$: 0.58, 95%CI: 0.28-1.18). Overall, there were three publications on ASBs and colorectal cancer (n=2), liver (n=1) and breast (n=1) cancers that could not be included in the meta-analyses because of insufficient data format[44,78,187]. The results of these studies were all null. Twenty-three exposure-outcome associations were only reported by one study (**Supplementary Table 17**)[70,72,75,78,140,144,177,180]. There was an inverse association between diet soft drinks and lung cancer in females in PLCO[177]. There was also a positive association between ASBs and papillary thyroid cancer in the EPIC study[180], and a borderline positive association between artificially sweetened decaffeinated cola and invasive breast cancer in the Women's Health Study (WHS)[75]. The rest showed null associations between ASBs, caffeinated or decaffeinated ASBs, or artificially sweetened cola and various cancers.

#### Supplementary Text 12. Meta-analyses of artificially sweetened beverages and cancer mortality.

There was an inverse association between ASBs intake and lung cancer mortality (RR: 0.97, 95%CI=0.96-0.99; $I^2$=0) (3 studies, 2 publications, 34,381 cases)[109,113] (**Supplementary Figure 229**), with ~96% of the meta-analytical weight from one large cohort study. The rest of the results were null (**Supplementary Figures 230-233**). A few subgroup analyses were possible, with no evidence of heterogeneity between subgroups (**Supplementary Figures 234-239**). The main results did not change materially in the influence analyses (**Supplementary Figures 240-241**). Forest plots showing study-specific results for the highest versus lowest exposure level comparison are presented in **Supplementary Figures 42-54,** and information on descriptive synthesis is provided in **Supplementary Text 13**.

#### Supplementary Text 13. Descriptive review of artificially sweetened beverages and cancer mortality.

Forest plots showing study-specific results for the highest versus lowest exposure level comparison are presented in **Supplementary Figures 42-54.** The results were generally null, except for a positive association between ASBs and pancreas cancer mortality (**Supplementary Figure 45**) and an inverse association with lung cancer mortality (**Supplementary Figure 46**) in the CPS II[113], which the latter was consistent the linear dose-response meta-analyses of lung cancer mortality. Thirteen exposure-outcome associations were only reported by CPS II[113] (**Supplementary Table 17**). The results were mixed of positive or inverse associations with various cancers, with 95%CIs crossing the null for all outcomes.

#### Supplementary Text 14. Descriptive review of juices and cancer incidence.

Forest plots showing study-specific results for the highest versus lowest exposure level comparison were presented in **Supplementary Figures 55-69.** Highest versus the lowest results were consistent the positive associations observed between fruit and vegetable juice and distal colon cancer incidence and orange juice and different types of skin cancer. Two studies reported on fruit juice and liver cancer, which could not be combined. The MEC study indicated little evidence of an association between fruit juice, per each one-point increase in the component score of the total, healthful and unhealthful plant-based diet, and hepatocellular carcinoma[88], and the Women's Health Initiative Observational Study (WHI-OS) indicated an inverse association but with 95%CI crossing the null value with liver cancer per each one serving per day[183]. Two other studies reported on tomato juice and bladder cancer, which could not be combined. The PLCO study reported no association for bladder cancer incidence across categories of tomato juice and per each one SD increase per day[175], and the HPFS reported a null association without reporting an effect estimate[120]. Overall, there were five publications on fruit juice and colorectal (n=2), pancreas (n=1) and prostate (n=2) cancers that could not be included in the meta-analyses because of insufficient data format[47,78,87,161,188]. There was a positive association with pancreatic cancer in PLCO[188] and a borderline inverse association with colorectal cancer in MEC[87]. The rest showed null associations. The NHS II investigated the association between fruit juice including total fruit juice and apple, orange, and grapefruit juices during early adulthood (aged 27-44 years) and breast cancer incidence later in life which suggested a borderline positive association for total fruit juice[51]. In that study, intake of fruit juices during adolescence (aged 13-18 years) was not associated with breast cancer risk. Similarly, there was no association between incidence of breast cancer later in life and orange juice during adolescence (aged 12-18 years) in the NHS[57] and during childhood (3-5 years) in the NHS and NHS II[121]. The NIH-AARP study reported on fruit juice at 12-13 years of age and indicated a positive association with non-advanced prostate incidence and inverse associations with advanced prostate cancer incidence and prostate cancer mortality later in life[94].

A total of 88 exposure-outcome associations were only reported by one study[38,43,44,47,49,51,58-60,63,72,74,77,78,81,84,86,89,94,107,111,117,118,120,123,124,133,138,141,147,153,156,159-162,165-167,174,176,179,181,185] (**Supplementary Table 18**). There was an inverse association between fruit and vegetable juice and rectal cancer in the MEC study[81], and between apple juice and non-Hodgkin lymphoma in the Iowa Women's Health Study (IWHS)[162].There were also positive associations between fruit juice and polycythaemia vera in the NIH-AARP study[141], pure fruit juice and endometrial cancer in a Canadian cohort[38], fruit and vegetable juice and urothelial cancer in the EPIC study[147], and citrus juice and papillary thyroid cancer in females in NIH-AARP study[174]. Of note, the EPIC study indicated positive associations between citrus juice and basal cell carcinoma and squamous cell carcinoma[107], which was consistent with the linear dose-response meta-analyses of citrus juice and melanoma incidence and orange juice and melanoma, basal cell carcinoma and squamous cell carcinoma incidence. The rest showed null or borderline associations for various juices.

#### Supplementary Text 15. Meta-analyses of juices and cancer mortality.

A pooled analysis of 15 cohorts with 3205 cases[139] reported results from three linear dose-response meta-analyses for citrus juice, vegetable juice and fruit and vegetable juice and prostate cancer mortality, which indicated little evidence of an association (**Supplementary Figure 308**). Detailed information on descriptive synthesis and single-study associations is provided in **Supplementary Text 16** and **Supplementary Table 18,** respectively**.**

**Supplementary Text 16. Descriptive review of juices and cancer mortality.**

A total of 16 exposure-outcome associations were only reported by one study[66,76,77,134,149] (**Supplementary Table 18**). Pure fruit juice was associated with a higher risk of liver cancer mortality in males and, in contrast, was associated with a lower risk of bladder cancer mortality in males in the Japan Collaborative Cohort Study for Evaluation of Cancer (JACC)[76]. Other results were mixed of positive or inverse associations between fruit juice, pure fruit juice, fruit and vegetable juice and tomato juice and various cancer mortality outcomes with 95%CIs crossing the null value in all association estimates.

#### Supplementary Text 17. Meta-analyses of soft drinks.

Twenty-six studies (36 publications, including six pooled analyses) reported on soft drinks and cancer across 18 anatomical sites, encompassing 58 exposure–outcome pairs (51 incidence, seven mortality) with 46,842 incident cases and 5,453 cancer deaths (**Supplementary Table 11**)[26,35,36,39,48-50,61,65-68,71,73,79,85,95,96,102,117-119,125-127,132,136,137,140,143,147,159,163,180,183,189]. Meta-analyses were possible for 13 cancer incidence (**Supplementary Figure 1**). Linear dose-response meta-analyses showed no clear associations between soft drinks and cancer incidence (**Supplementary Figures 309-318**). Subgroup analyses indicated little heterogeneity (**Supplementary Table 22, Supplementary Figures 319-322**), and results remained consistently null after sequential study exclusion (**Supplementary Figures 323-325**). Publication bias was not assessed due to the low number of studies (n<10). Forest plots for highest versus lowest exposure comparisons are shown in **Supplementary Figures 18–41**, with descriptive synthesis detailed in **Supplementary Texts 18–19**; 31 exposure-outcome associations reported by single studies are summarised in **Supplementary Table 15**.

**Supplementary Text 18. Descriptive review of soft drinks and cancer incidence.**

Forest plots showing the study-specific results for the highest versus the lowest exposure level comparisons are presented in **Supplementary Figures 18-41.** The European Prospective Investigation into Cancer and Nutrition (EPIC)[159] indicated a positive association between soft drinks and hepatocellular carcinoma (**Supplementary Figure 26**), and the Swedish Mammography Cohort (SMC) and Cohort of Swedish Men (COSM)[96] indicated a positive association with gallbladder and extrahepatic biliary tract cancers (**Supplementary Figure 27)**, however, linear dose-response meta-analyses including these two publications indicated null associations. Three studies reported on soft drinks and colorectal cancer which could not be meta-analysed. The EPIC study reported a positive association between soft drinks and colorectal cancer incidence[136], with further analyses by anatomical subsite suggesting a positive association with colon cancer, especially distal colon cancer, and a borderline association with rectal cancer[136]. Other results for colorectal cancer were null, suggesting an inverse association but with 95%CI crossing the null in the Netherlands Cohort Study (NLCS)[136] and EPIC-Italy[35]. Two studies reported on soft drinks and kidney cancer, of which the Prostate, Lung, Colorectal and Ovarian (PLCO) Cancer Screening Trial reported a positive association with renal cell cancer[102], and the EPIC study reported no association in either sex[66].

Overall, there were three publications on pancreas (n=1), endometrial (n=1), and lung (n=1) cancers which could not be included in the meta-analyses because of insufficient data format[61,85,177]. The results of these studies were generally null and consistent with the main meta-analyses, except for PLCO[177] that reported an inverse association between soft drinks and lung cancer in females which was not observed in males but with overlapping CIs between sexes (**Supplementary Figure 29**, **see footnote**). Thirty-seven exposure-outcome associations were only reported by one study (**Supplementary Table 15**)[48-50,73,118,125,137,140,143,159,163,180]. There was a positive association between carbonated soft drinks and pancreas adenocarcinoma in the Singapore Chinese Health Study (SCHS)[125], and between soft drinks and differentiated thyroid cancer, especially papillary thyroid cancer, in the EPIC study[180], whereas the rest showed a null association between total, carbonated, caffeinated, and decaffeinated soft drinks and cola and cancers of the head and neck, oral cavity, stomach, oesophagus, colon, pancreas, breast, ovary, prostate, brain, and haematological system.

**Supplementary Text 19. Descriptive review of soft drinks and cancer mortality.**

Two studies reported on soft drinks and colorectal cancer mortality which could not be meta-analysed. The EPIC study reported a positive association with colorectal cancer mortality[126], while the Hokkaido study in Japan indicated no association[85]. Two studies reported on soft drinks and kidney cancer mortality, of which the PLCO study reported a positive association but with 95%CI crossing the null with renal cell cancer mortality[102], and the EPIC study reported a borderline positive association in females and no association in males[66]. Six exposure-outcome associations were only reported by one study[85,126] (**Supplementary Table 15**) indicating no association between soft drinks or cola and colorectal, lung, breast, and prostate cancers mortality.

#### Supplementary Text 20. Risk of bias assessment.

The risk of bias assessment of 99 publications (38 studies) across seven domains are presented in **Supplementary Figure 326**. Regarding confounding, most publications (81%) had moderate, 8% serious, and 11% critical risk of bias. This was mainly because publications did not include all critically important confounding variables **(Supplementary Table 4),** including smoking intensity (n=5), alcohol drinking (n=5), reproductive factors (n=4), race (n=3), and skin cancer-related risk factors (n=1) in their multivariable analyses. Regarding selection of participants, 99% of the publications had moderate risk of bias, and the remaining 1% had serious risk. Regarding exposure classification, 28% of the publications had low, 62% moderate, and 10% serious risk of bias, with no study at critical risk. Publications rated as serious risk provided insufficient information regarding the composition of exposures (e.g., juices) and did not report detailed information on validity estimates of dietary assessment tool to measure that specific exposure. Bias due to departures from intended exposures was rated as critical in 74% of publications, serious in 5%, and moderate in 21% of publications. Changes in exposure status may have occurred after baseline exposure assessment but repeated measurements were limited to 22 publications from eight studies (22%) including NHS, NHS II and HPFS (every 2-4 years), CPS II (baseline assessment and then an update after 4 years), BWHS (baseline assessment and then an update after 6 years), UK biobank (baseline assessment and then 4 rounds during the next 2 years), SU.VI.MAX (a 24-hour dietary record every 2 months, altogether, six times per year), and NutriNet-Santé (≥2 assessments every six months), while the others used a single baseline dietary assessment. However, subgroup analyses by dietary assessment frequency (single baseline vs repeated measurement) and by bias due to departure from intended exposure (moderate vs critical bias) did not indicate any notable difference between the subgroups for colorectal cancer incidence and mortality, colon, proximal and distal colon cancer incidence, breast, pre and postmenopausal breast cancer incidence, breast cancer mortality, prostate cancer incidence and mortality, and non-Hodgkin lymphoma incidence. In bias due to missing data, 47% of publications had low, 5% moderate, and 11% serious risk, while 36% lacked sufficient information for assessment. Bias in the measurement of outcomes was not a concern, as almost all publications (99%) were rated as low risk. Bias in the selection of reported results was moderate in 95% of publications, while 5% were classified as low.

#### Supplementary Text 21. Mendelian randomisation studies.

Three MR studies were identified reporting results on 175 MR analyses of various soft drinks with cancer, using two-sample summary-level information from large GWAS studies or consortia of European ancestry. The studies investigated genetically predicted self-reported intake of sweet beverages (21 SNPs)[191], SSBs (23 SNPs[191], or 12 SNPs)[192], ASBs (16 SNPs)[192], and artificial sweeteners added to coffee (12 SNPs) or tea (15 SNPs)[193]. The genetic instruments included 12-23 SNPs (**Supplementary table 23**).

One MR study indicated no association between genetically predicted sweet beverages intake (including SSBs, ASBs, pure non-grapefruit juices, flavoured milk, and hot chocolate) and risk of colon or rectal malignant neoplasms[191]. The study also indicated a positive association between genetically predicted SSBs intake and risk of colonic malignant neoplasms (OR 1.013, 95%CI: 1.001-1.026), but not with rectal malignant neoplasms[191]. Another MR study investigating genetically predicted SSBs consumption and 13 obesity-related cancers reported a positive association with breast cancer (OR per 1 SD: 1.888, 95% CI: 1.115-3.196), but not with other cancers, including colorectal cancer[192]. As for genetically predicted ASBs consumption, the study reported a positive association with colorectal (OR 6.88, 95%CI: 1.55-30.51) and breast cancer (OR: 3.88, 95%CI: 2.02-9.78)[192]. Further two-step MR suggested a potential mediation by BMI in the association of ASBs with colorectal cancer. No association was found between ASBs and other cancers including oesophageal, stomach, liver, pancreatic, endometrial, ovarian, prostate, kidney and thyroid cancers[192]. The last MR study investigated genetically predicted artificial sweeteners added to coffee and tea in relation to 67 and 71 cancer outcomes of different grades and histology, and suggested that genetically predicted artificial sweeteners added to coffee was associated with a higher risk of high- and low-grade ovarian cancer, and that artificial sweeteners added to tea was associated with higher risk of cancers of the oral cavity and pharynx[193]. Of note, inverse associations were found between artificial sweeteners added to tea and cancers of the bronchus and lung. No other association was found between artificial sweeteners added to tea or coffee and cancer outcomes[193].

**Supplementary Table 2. Eligibility criteria for cancer outcomes commonly investigated in the epidemiological studies[a]**

| Cancer sites (incidence or mortality) | Inclusion criteria | Exclusion criteria |
|---|---|---|
| Bladder cancer | Bladder cancer; transitional cell carcinoma (urothelial carcinoma), squamous cell carcinoma, adenocarcinoma; histologically defined carcinoma in situ; and imprecise anatomical definitions of cancer sites that include bladder cancer, such as urological tract cancer, provided that they are of invasive carcinoma. | Pre-malignant bladder cancer other than histologically defined carcinoma in situ. |
| Brain and other central nervous system cancers | Brain cancer; glioma, glioblastoma; grade III meningioma; spinal cord cancer; brain combined with meninges and spinal cord cancers. | Benign brain tumours; benign spinal cord tumours; metastatic spinal cancer. |
| Breast cancer | Invasive breast cancer; premenopausal or postmenopausal breast cancer; any molecular, genetic, or histopathological subtypes. | In situ breast cancer. |
| Cervical cancer | Invasive or microinvasive cervical cancer; squamous cell carcinoma (SCC), epidermoid carcinoma (large cell or small cell, keratinizing or nonkeratinizing), or adenocarcinoma; cervical intraepithelial neoplasia III (CIN 3)/carcinoma in situ (CIS). | Pre-invasive neoplastic lesions (cervical dysplasia, cervical intraepithelial neoplasia (CIN I or II), squamous intraepithelial lesion (SIL, different grades). |
| Colorectal cancer | Colorectal cancer; (distal, proximal) colon cancer, rectal cancer; any molecular, genetic, or histopathological subtypes; Lynch syndrome colorectal cancer, early onset colorectal cancer. | Pre-malignant colorectal conditions, e.g. colorectal adenomas or polyps. |
| Endometrial cancer | Endometrial cancer; type I endometrial cancer (endometrioid adenocarcinoma), type II endometrial cancer (serous, clear cell, carcinosarcoma and undifferentiated/dedifferentiated cancers) | ---- |
| Gallbladder cancer | Gallbladder cancer; imprecise anatomical definitions of cancer sites that include gallbladder cancer, such as biliary tract cancer. | ---- |
| Kidney cancer | Kidney cancer; (clear cell, non-clear cell) renal cell carcinoma (RCC), renal parenchyma cancer; renal pelvis cancer, transitional cell carcinoma (TCC) (urothelial carcinoma). | Wilm's tumour (more common in children) or cancer related to schistosomiasis infection. |
| Liver cancer | Liver cancer; hepatocellular carcinoma (HCC). | ---- |
| Lung cancer | Lung cancer; non-small cell lung cancer (NSCLC), adenocarcinoma, squamous cell carcinoma, large cell carcinoma; small cell lung cancer (SCLC). | ---- |
| Lymphoid and hematopoietic cancers | Leukaemia; acute lymphoblastic/lymphocytic leukaemia (ALL), acute myeloid leukaemia (AML), chronic lymphocytic leukaemia (CLL)[b], chronic myeloid leukaemia (CML), chronic myelomonocytic leukaemia (CMML). | ---- |

| Cancer sites (incidence or mortality) | Inclusion criteria | Exclusion criteria |
| --- | --- | --- |
| | Lymphoma; non-Hodgkin lymphoma, Hodgkin lymphoma, diffuse large B-cell lymphoma, follicular lymphoma, small lymphocytic lymphoma (SLL)[b]<br><br>Multiple myeloma. | |
| Mouth, pharyngeal, and laryngeal cancer | Cancer of the mouth, pharynx or larynx as separate outcomes, as combinations of these cancers, or for all these cancers combined; head and neck cancer; upper aerodigestive cancer; and other cancer groups that explicitly include mouth, pharynx, or larynx cancers. | ---- |
| Nasopharyngeal cancer | Nasopharyngeal cancer. | Cancer combinations that include nasopharyngeal cancer. |
| Oesophageal cancer | Oesophageal cancer; gastro-oesophageal cancer; squamous cell carcinoma, adenocarcinoma. | Upper aerodigestive cancer that includes oesophageal cancer that is evaluated under mouth, pharyngeal, and laryngeal cancers. |
| Ovarian cancer | Ovarian cancer; epithelial ovarian carcinoma; serous carcinomas. | ---- |
| Pancreatic cancer | Pancreatic cancer. | ---- |
| Prostate cancer | Prostate cancer; advanced/aggressive prostate cancer (see (a)-(h) below), early localised/low grade prostate cancer; imprecise anatomical definitions of cancer sites that include prostate cancer, such as urogenital cancer.<br><br>(a) stage 3-4 on the AJCC 1992 classification<br>(b) advanced cancer<br>(c) advanced or metastatic cancer<br>(d) metastatic cancer<br>(e) stage C or D on the Whitemore/Jewertt scale<br>(f) fatal cancer<br>(g) high stage or grade<br>(h) Gleason grade >= 7 | Pre-malignant prostate conditions, e.g. high-grade prostate intra epithelial neoplasia. |
| Skin cancer | Skin cancer, including basal cell carcinoma (BCC), basal cell epithelioma; squamous cell carcinoma (SCC) of skin, squamous cell epithelioma; (invasive or in situ) melanoma, cutaneous melanoma; non-melanoma skin cancer (NMSC). | Skin cancer in patients with AIDS (e.g. Kaposi's sarcoma and AIDS). |
| Small intestine cancer | Small intestine cancer. | Small intestine polyps. |
| Stomach cancer | Gastric (stomach) cancer; cardia or non-cardia gastric cancers. | Anatomical definitions of cancer sites that include gastric cancer, e.g. gastrointestinal cancer; gastro-oesophageal cancer that is evaluated under oesophageal cancer. |
| Thyroid cancer | Thyroid cancer; differentiated thyroid cancer (papillary, follicular, and oncocytic carcinoma). | ---- |

[a]All listed cancers were reviewed in the TER, apart from brain and other central nervous system cancers, lymphoid and hematopoietic cancers, small intestine cancer, and thyroid cancer. The list is

not meant to be exhaustive. Any additional cancers or cancer subtypes identified in eligible studies during the literature search will be included in the present SLRs.

[b]CLL is the same disease as SLL.

**Supplementary Table 3. Risk of Bias for Nutrition Observational Studies (RoB-NObs) tool for evaluation of cancer incidence studies (modified version dated 29/01/2025).**

Assess for each exposure-outcome association for publications included in the meta-analyses or narrative reviews.

Some sections can be assessed for the overall publication (1, 2, 5, 7) but exposure and outcome-related domains should be assessed separately (3, 4, 6). We consider lumping together similar exposures (e.g., dietary exposures) or outcomes (e.g., mortality outcomes).

The levels of judgement are low, moderate, serious, critical, and no information for each of the seven domains. There is no overall risk of bias.

Abbreviations:
Y: Yes
PY: Probably yes
N: No
PN: Probably no
NI: No information

#### 1. Bias due to confounding.

| Question | Issues/ Remarks | Prompting questions/ indicators |
|---|---|---|
| 1.1 Is there potential for confounding of the effect of exposure in this study?<br>**If N or PN**: skip all remaining questions (1.2 to 1.8) and go to Bias due to confounding: Risk of bias judgement; the study can be considered to be at low risk of bias due to confounding and no further signalling questions need be considered.<br>**If Y or PY**: answer question 1.2 to determine whether there is a need to assess time-varying confounding. | Issues:<br>- Confounding cannot be ruled out in observational studies. | This question should be always answered as **PY** in observational studies of the areas we work on. |

| Question | Issues/ Remarks | Prompting questions/ indicators |
|---|---|---|
| 1.2. **If Y or PY to 1.1**: Was the analysis based on splitting follow-up time according to exposure received?<br>**If N or PN:** skip 1.3 and answer questions 1.4 to 1.6, which relate to baseline confounding. If Y or PY, go to question 1.3. | Issues:<br>- Exposures may change across time and are subject to time-varying confounding.<br>Remarks:<br>- This question asks whether a time-varying exposure analysis was conducted (which also requires to adjust for time-varying confounders). | Consider whether the study has evaluated the exposure at multiple time points. This question should be always answered as either **Y** or **N.**<br><br>-**Y** The study has evaluated the exposure at multiple time points and the investigators have performed time-varying exposure/ confounder analyses.<br>-**N** The study has not evaluated the exposure at multiple time points OR the investigators have not performed time-varying exposure/ confounder analyses. |
| 1.3. **If Y or PY to 1.2**: Were exposure discontinuations or switches likely to be related to factors that are prognostic for the outcome?<br>**If N or PN:** answer questions 1.4 to 1.6, which relate to baseline confounding only. Do not answer 1.7 and 1.8, which relate to both baseline and time-varying confounding.<br>**If Y or PY**: skip questions 1.4 to 1.6, and answer questions 1.7 and 1.8, which relate to both baseline confounding and time-varying confounding. | Issues:<br>- How to tell whether the changes are related to prognostic factors for the outcome?<br><u>Remarks:</u><br>- If exposure switches are unrelated to the outcome, for example when the outcome is an unexpected harm, then time-varying confounding will not be present and only control for baseline confounding is required.<br>- This is unlikely on the areas and outcomes we work on. | This question should be always answered as **PY** in observational studies of the areas we work on. |
| Questions relating to baseline confounding | | |
| 1.4. **If N or PN to 1.2 or 1.3**: Did the authors use an appropriate analysis method that adjusted for all the critically important confounding variables at baseline?<br>Go to 1.5 | Issues:<br>- Which method of adjustments can be considered as adequate?<br>Remarks:<br>- Any method to adjust for measured confounders should be acceptable e.g., stratification, regression, matching, | -**N** Not adjusted at least for all the required confounding variables.<br>-**PY** Adjusted at least for all the required confounders.<br>-**Y** Adjusted at least for all the required plus the desired confounding variables.<br>Please consult the external list of important confounding variables below. |

| Question | Issues/ Remarks | Prompting questions/ indicators |
|---|---|---|
| | restriction, standardization, and inverse probability weighting, multivariable analysis.<br>- Consider the association as "adjusted" when the required factors (as they appear in the Prompting questions/ indicators) were used in matching between those with or without a study outcome or stratification in analysis.<br>- Consider the association as "adjusted" when the required factors (as they appear in the Prompting questions/ indicators) were tested but not included in the final statistical model due to lack of statistical significance or small change in risk estimate.<br>E.g., age was tested univariably and was found to be not significant or the estimates did not change by more than 10%. | |
| 1.5. Were confounders that were adjusted for measured validly and reliably by the variables available in this study?<br>Go to 1.6 | Issues:<br>- Residual confounding is possible if covariates are not measured validly and reliably.<br>Remarks:<br>- For some topics, a list of valid and reliable measures of confounding domains will be specified in the review protocol but for others such a list may not be available.<br>- Study authors may cite references to support the use of a particular measure.<br>- If authors control for confounding variables with no indication of their validity or reliability pay attention to the subjectivity of the measure. | -**Y** Adiposity (any index or measure) was measured. Dietary factors and physical activity were self-reported but based on a validated questionnaire, or their assessment was validated against another assessment method (within the same study or externally in another study reported in the paper). Comorbidities (if relevant) were ascertained by a physician, medical records, registries.<br>-**PY** Adiposity (any index or measure) was self-reported. Dietary factors and physical activity were self-reported but based on a validated questionnaire, or their assessment was validated against another assessment method (within the same study or externally in another study reported in the paper). Comorbidities (if relevant) were self-reported but further ascertained by a physician, medical records, registries. |

| Question | Issues/ Remarks | Prompting questions/ indicators |
|---|---|---|
| | - Subjective measures (e.g. based on self-report) may have lower validity and reliability than objective measures such as lab findings. | -**N/ PN** Adiposity (any index or measure) was self-reported. Dietary factors and physical activity were self-reported but were not based on a validated questionnaire, or their assessment was not validated against another assessment method. Comorbidities (if relevant) are self-reported and not further ascertained.<br>**-NI** Information was not reported in detail/ separately for each critical confounder<br>When two or more different confounders lead to different judgement then go with the worst-case scenario. |
| 1.6. Did the authors avoid adjusting for post-exposure variables?<br><br>Skip to Bias due to confounding: Risk of bias judgement. | Issues:<br>- Controlling for post-exposure variables that are affected by the exposure is not appropriate.<br>- Controlling for mediating variables estimates the direct effect of the exposure and may introduce bias.<br>- Controlling for common effects of the exposure and outcome introduces bias.<br>Remarks:<br>- Confounders should be measured at or before (ideally) the time that exposure was assessed and not after (a few days/ weeks is ok but definitively not longer). | This question should be always answered as either **PY** or **PN**.<br><br>-**PY** The study has not adjusted for post-exposure variables affected by the exposure, mediators, colliders (common effects of the exposure and outcome) AND the adjustment variables should be measured at or before the time of exposure assessment.<br>-**PN** All other cases. |
| Questions relating to baseline and time-varying confounding | | |
| 1.7. **If Y or PY to 1.3**: Did the authors use an appropriate analysis method that adjusted for all the critically important confounding variables, including baseline and time-varying confounding?<br>**If N or PN to 1.7**: skip to Bias due to confounding: Risk of bias judgement. | Issues:<br>- Exposures and confounders may change across time.<br>Remarks:<br>- Any method to adjust for baseline and time-varying confounding should be considered appropriate (e.g., regular | Please consult the external list of important confounding variables.<br>Time-fixed confounders<br>-**PY** Age, sex (if relevant), race (if relevant), reproductive factors (if relevant), comorbidities (if relevant).<br>Time-varying confounders |

| Question | Issues/ Remarks | Prompting questions/ indicators |
|---|---|---|
| **If Y or PY to 1.7**, answer question 1.8. | regression, methods based on inverse probability weighting, marginal structural models). | **-Y** Adiposity, smoking, dietary factors, physical activity, comorbidities (if relevant).<br>**-PN** All other cases. |
| 1.8 **If Y or PY to 1.3 and Y or PY to 1.7**: Were confounders that were adjusted for measured validly and reliably by the variables available in this study?<br><br>Skip to Bias due to confounding: Risk of bias judgement. | | **-Y** Adiposity (any index or measure) was measured. Dietary factors and physical activity were self-reported but based on a validated questionnaire, or their assessment was validated against another assessment method (within the same study or externally in another study reported in the paper). Comorbidities (if relevant) were ascertained by a physician, medical records, registries.<br>**-PY** Adiposity (any index or measure) was self-reported. Dietary factors and physical activity were self-reported but based on a validated questionnaire, or their assessment was validated against another assessment method (within the same study or externally in another study reported in the paper). Comorbidities (if relevant) were self-reported but further ascertained by a physician, medical records, registries.<br>**-N/ PN** Adiposity (any index or measure) was self-reported. Dietary factors and physical activity were self-reported but were not based on a validated questionnaire, or their assessment was not validated against another assessment method. Comorbidities (if relevant) are self-reported and not further ascertained.<br>**-NI** Information was not reported in detail/ separately for each critical confounder<br>When two or more different confounders lead to different judgement then go with the worst-case scenario. |
| RoB judgement | | |
| Low risk of bias | No confounding expected. Observational studies should never have low RoB in this domain. | |

| Question | Issues/ Remarks | Prompting questions/ indicators |
|---|---|---|
| The study is comparable to a well-performed randomized trial with regard to this domain. | | |
| Moderate risk of bias<br>The study is sound for an observational study with regard to this domain but cannot be considered comparable to a well-performed randomized trial. | (i) Confounding expected, all known important confounding domains appropriately measured and controlled for;<br>and<br>(ii) Reliability and validity of measurement of important domains were sufficient, such that we do not expect serious residual confounding. | |
| Serious risk of bias<br>The study has some important problems. | (i) At least one key confounder was not appropriately measured, or not controlled for;<br>or<br>(ii) Reliability or validity of measurement of a key confounder **could not be evaluated or** was low enough that we expect serious residual confounding. | |
| Critical risk of bias<br>The study is too problematic to provide any useful evidence. | (i) Confounding is inherently not controllable;<br>or<br>(ii) The use of negative controls strongly suggests unmeasured confounding. | |

#### 2. Bias in selection of participants into the study.

| Question | Issues/ Remarks | Prompting questions/ indicators |
|---|---|---|
| 2.1. Was selection of participants into the study or into the analysis based on participant characteristics observed after the start of the exposure assessment?<br>If **N or PN:** go to 2.4 (skip 2.2 and 2.3).<br>If **Y or PY**: go to 2.2 and 2.3. | Issues:<br>- Systematic difference in risk factors and risk of the outcome between study participants and non-participants. Can arise from the initial self-selection of study participants into the study. | Expect to answer this question almost always as **Y/ PY**.<br><br>-**N/ PN** Study population was a random sample of the target population or was based on a national registry.<br>-**Y/ PY** If participant contact or consent is required, selection bias may occur. |
| 2.2. **If Y or PY to 2.1**: Were the post-exposure variables that influenced selection of participants (into the study or analysis) associated with exposure?<br>Go to 2.3 | Issues:<br>- Post-exposure variables may influence study selection which can lead to unrepresentative exposure groups.<br>Remarks: | -**Y** An analysis has been performed and there is a difference (10% or more) in the exposure value/ rate between the groups.<br>-**PY** An analysis has not been performed.<br>-**PY** An analysis has been performed and there is a difference (5-10%) in the exposure value/ rate between the groups. |

| Question | Issues/ Remarks | Prompting questions/ indicators |
|---|---|---|
| | - The focus of this question should only be on the exposure and not on other variables.<br>- Consider whether an analysis (by exposure status) comparing participants and non-participants has been performed.<br>- Consider whether the characteristics (age, sex [when appropriate], BMI or other exposures e.g., diet, physical activity [when appropriate]) between the participants and non-participants at baseline (at the time of exposure) are comparable.<br>- Keep in mind that sometimes studies compare baseline characteristics of included vs. excluded individuals based on a certain exclusion criterion, but we need to evaluate here whether they have also performed something similar for individuals that never participated [volunteered] in the study. | -**PN** An analysis has been performed and there is a difference (<5%) in the exposure value/ rate between the groups.<br>-**N** An analysis has been performed and there is no difference in the exposure value/ rate between the groups OR there is a statement in the manuscript that potential selection of participants was not associated with exposure. |
| 2.3. **If Y or PY to 2.1**: Were the post-exposure variables that influenced selection of participants (into the study or analysis) associated with the outcome?<br>Go to 2.4 | Issues:<br>- Post-exposure variables may influence study selection leading to unrepresentative outcome ascertainment.<br>- In cohort studies (of cancer incidence), any self-selection of participants should not be associated with the outcome that occurs years later. | *Expect to answer this question almost always as **N/PN** when cohort studies are evaluated.* |
| 2.4. Do start of follow-up and start of exposure assessment coincide for most participants?<br>If **N or PN**: answer 2.5. | Remarks:<br>- The time of follow-up start should coincide with the time of exposure assessment. | -**Y** Follow-up for outcome assessment started at the time of exposure assessment (or within 3 months maximum).<br>**-PN** The study has mentioned that the start of follow-up for outcome assessment and the time of |

| Question | Issues/ Remarks | Prompting questions/ indicators |
|---|---|---|
| If **Y or PY**: go to Bias in selection of participants into the study: Risk of bias judgement. | | exposure assessment did not coincide (the exposure was assessed after the start of follow-up) but there was no information on the timing between the two measurements.<br>**-N** The start of follow-up for outcome assessment only started after the time of exposure assessment (more than 3 months).<br>**-NI** The study has not mentioned anything about the start of follow-up for outcome assessment and the time of exposure assessment. |
| 2.5 **If Y or PY to 2.2 and 2.3, or N or PN to 2.4**: Were adjustment techniques that were likely to correct for the presence of selection biases used?<br>Go to Bias in selection of participants into the study: Risk of bias judgement. | Remarks:<br>- It is in principle possible to correct for selection biases, for example by using inverse probability weights to create a pseudo-population in which the selection bias has been removed, or by modelling the distributions of the missing participants or follow up times and outcome events and including them using missing data methodology.<br>- Check whether any adjustment techniques (e.g., inverse probability weighting, g-formula, marginal structural models) were used for accounting for selection bias either as a main analysis or a sensitivity analysis. | **-Y/ PY** Such an analysis was performed.<br>**-N/ PN** Such an analysis was not performed. |
| RoB judgement | | |
| Low risk of bias<br>The study is comparable to a well-performed randomized trial with regard to this domain. | (i) All participants who would have been eligible for inclusion, were included in the study;<br>and<br>(ii) For each participant, start of follow up and start of exposure coincided. | |
| Moderate risk of bias<br>The study is sound for an observational study with regard to this domain but cannot be | (i) All participants who would have been eligible for inclusion, were included in the study;<br>and<br>There was no information on whether start of follow up and start of exposure coincided for all participants. | |

| Question | Issues/ Remarks | Prompting questions/ indicators |
|---|---|---|
| considered comparable to a well-performed randomized trial. | or<br>(ii) Selection into the study may have been related to exposure and outcome;<br>and<br>The authors used appropriate methods to adjust for the selection bias;<br>or<br>(iii) Start of follow up and start of exposure do not coincide for all participants;<br>and<br>(a) the proportion of participants for which this was the case was too low to induce important bias;<br>or<br>(b) the authors used appropriate methods to adjust for the selection bias;<br>or<br>(c) the review authors are confident that the rate (hazard) ratio for the effect of exposure remains constant over time. | |
| Serious risk of bias<br>The study has some important problems. | (i) Selection into the study was related (but not very strongly) to exposure and outcome;<br>and<br>This could not be adjusted for in analyses;<br>or<br>(ii) Start of follow up and start of exposure do not coincide;<br>and<br>A potentially important amount of follow-up time is missing from analyses;<br>and<br>The rate ratio is not constant over time | |
| Critical risk of bias<br>The study is too problematic to provide any useful evidence. | (i) Selection into the study was very strongly related to exposure and outcome;<br>and<br>This could not be adjusted for in analyses;<br>or<br>(ii) A substantial amount of follow-up time is likely to be missing from analyses;<br>and<br>The rate ratio is not constant over time. | |

##### 3. Bias in classification of exposures.

| Question | Issues/ Remarks | Prompting questions/ indicators |
|---|---|---|
| 3.1. Is the exposure that was assessed clearly defined? | Issues:<br>- Ambiguity in the definition may lead to bias in the classification of participants.<br>Remarks:<br>- Example where exposure may not be clearly defined may be e.g., a study has an unclear definition of dietary and supplement intake of vitamins and ends up with a mixture of info for the different participants in the study. | Expect to answer this question most often as **Y/ PY**.<br><br>-**Y/ PY** The study provided details for the definition of the exposure.<br>-**N/ PN** The study did not provide sufficient details for the definition of the exposure.<br>E.g., The study aim is to assess vitamin D, but did not define what is considered as vitamin D. |
| 3.2. Does the exposure that was assessed represent the exposure of interest? | | Expect to answer this question most often as **Y/ PY**.<br><br>-**Y/ PY** The assessed exposure directly represents the exposure of interest.<br>-**N/ PN** The assessed exposure does not directly represent the exposure of interest.<br>E.g., The study aim is to assess total physical activity, but data are only on recreational physical activity. |
| 3.3. Were the methods used to assess the exposure clearly described? | | Expect to answer this question most often as **Y/ PY**.<br><br>-**Y/ PY** The study provided details on the assessment methods of the exposure of interest.<br>-**N/ PN** The study did not provide sufficient details on the assessment methods of the exposure of interest. |
| 3.4. Were the methods used to measure the exposure valid and/ or reliable? | | -**Y** Assessment was based on any objective measure of assessing the exposure (measured adiposity, measured physical activity, measured sedentary behaviour, diet biomarkers).<br>-**Y** Exposure was self-reported and assessment was validated against another exposure assessment method, in whole or subset of study populations AND |

| Question | Issues/ Remarks | Prompting questions/ indicators |
|---|---|---|
| | | replicated with the same exposure method at a different time point.<br>-**PY** Assessment was based on self-reported weight/ height/ BMI.<br>-**PY** Exposure was self-reported, and assessment was validated against another exposure assessment method, in whole or subset of study populations OR replicated with the same exposure method at a different time point.<br>-**PY** Exposure was self-reported, and assessment was validated against another exposure assessment method AND replicated with the same exposure method at a different time point, but these were performed in another study cohort (also check discussion section).<br>-**PN** Exposure was self-reported, and assessment was validated against another exposure assessment method OR replicated with the same exposure method at a different time point, but these were performed in another study cohort (also check discussion section).<br>-**PN** Assessment was based on retrospective assessment or chart review of exposures.<br>-**N** Exposure was self-reported and assessment was not validated against another exposure assessment method OR not replicated with the same exposure method at a different time point.<br>-**NI** No data on validation AND replication of the self-reported measurement. |
| 3.5. Were the same methods used to assess the exposure status for all participants/ groups? | | -**PY/ Y** Assessment method and timeframe were the same regardless of exposure status.<br>-**PN/ N** Assessment method and timeframe differed in terms of exposure status. |
| 3.6. Were the methods used to define exposure status for participants/ groups clearly described? | Remarks:<br>- A study may use different definitions of exposure, an exposure may be measured | -**Y/ PY** The study provided details on how the exposure status was defined. |

| Question | Issues/ Remarks | Prompting questions/ indicators |
|---|---|---|
| | based on a score, or a continuously measured exposure may be used as categories. The definition of the variable used for the exposure should be clearly defined. | -**N/ PN** The study did not provide sufficient details on how the exposure status was defined. |
| 3.7. Were the methods used to define exposure status for participants/ groups likely to result in minimal random or systematic exposure misclassification? | Remarks:<br>- Misclassification exists in self-reported diet, adiposity, and physical activity assessments.<br>- The question is whether this misclassification is different in those with high vs. low levels of exposure, which might be the case.<br>- It is expected to see different statistics evaluating validity/ reliability (correlation coefficient, sensitivity, specificity, kappa, etc.).<br>- Please use common sense (or check for established thresholds for the specific exposure) about what an acceptable level for the specific statistic is, but as guidance: a correlation coefficient of 0.4 or larger can be considered acceptable for nutritional exposures but not acceptable for BMI for which we would expect a correlation coefficient of 0.7 or larger. | -**Y** Exposure assessment is based on objective measures (measured adiposity, measured physical activity, diet biomarkers) or based on self-reported anthropometric factors AND a validity/ reliability statistic is presented and is at an acceptable level.<br>-**PY** Exposure assessment is based on objective measures (measured adiposity, measured physical activity, diet biomarkers) or based on self-reported anthropometric factors AND a validity/ reliability statistic is not presented.<br>-**PY** Exposure assessment is based on any self-reported measure (except for anthropometric factors) AND a validity/ reliability statistic is presented and is at an acceptable level<br>-**PN** Exposure assessment is based on any self-reported measure (except for anthropometric factors) AND a validity/ reliability statistic is not presented.<br>-**N** A validity/ reliability statistic is presented, either for objective or the self-reported measures and is not at an acceptable level. |
| 3.8. Could classification of exposure status been affected by the presence of the outcome, knowledge of the outcome or risk of the outcome?<br>**If Y or PY**, there may be serious risk of bias. Go to Bias in classification of exposures: Risk of bias judgement. | Issues:<br>- There is the possibility that the exposure of interest may be influenced by undiagnosed disease, thus resulting in misclassification. | -**Y/ PY** The study included participants for which assessment of the exposure was performed at least 1 or 2 years before diagnosis and there was no mention about performing some type of lag analysis (e.g., the study excluded events that happen within 1 or 2 years of exposure assessment in a sensitivity analysis).<br>-**N/ PN** All other cases. |

| Question | Issues/ Remarks | Prompting questions/ indicators |
|---|---|---|
| RoB judgement | | |
| Low risk of bias<br>The study is comparable to a well-performed randomized trial with regard to this domain. | (i) The exposure and the methods used to assess the exposure were well defined and represent the exposure of interest;<br>and<br>(ii) Methods were valid, reliable, the same across groups, and likely to result in minimal random or systematic exposure misclassification;<br>and<br>(iii) Exposure status was not affected by the presence of the outcome, knowledge of the outcome or risk of the outcome. | |
| Moderate risk of bias<br>The study is sound for an observational study with regard to this domain but cannot be considered comparable to a well-performed randomized trial. | (i) The exposure and the methods used to assess the exposure are defined and represent the exposure of interest;<br>and<br>(ii) Methods were valid, reliable, the same across groups, and likely to result in minimal random or systematic exposure misclassification;<br>or<br>Exposure status was not affected by the presence of the outcome, knowledge of the outcome or risk of the outcome. | |
| Serious risk of bias<br>The study has some important problems. | (i) Exposure status or the methods used to assess the exposure are not well defined or do not represent the exposure of interest;<br>and<br>(ii) Methods were not valid and reliable, the same across groups, or were likely to result in some degree of random or systematic exposure misclassification;<br>or<br>Exposure status was affected by the presence of the outcome, knowledge of the outcome or risk of the outcome. | |
| Critical risk of bias<br>The study is too problematic to provide any useful evidence. | (i) Exposure status and the methods used to assess the exposure are not well defined or do not represent the exposure of interest;<br>and<br>(ii) Methods were not valid and reliable, were not the same across groups, and were likely to result in substantial random or systematic exposure misclassification;<br>and | |

| Question | Issues/ Remarks | Prompting questions/ indicators |
|---|---|---|
| | (iii) Exposure status was affected by the presence of the outcome, knowledge of the outcome or risk of the outcome. | |

**4. Bias due to departures from intended exposures.**

| Question | Issues/ Remarks | Prompting questions/ indicators |
|---|---|---|
| 4.1. Is there concern that changes in exposure status occurred among participants that were unbalanced across groups and likely to impact the outcome? | Issues:<br>- Changes of exposure status may happen after baseline exposure assessment | This question should be always answered as **PY** in observational studies of the areas we work on. |
| 4.2. Were any critical co-exposures that occurred unbalanced between exposure groups and likely to impact the outcome? | | This question should be always answered as **PY** in observational studies of the areas we work on. |
| 4.3. **If Y or PY to 4.1, or 4.2**: Were adjustment techniques that are likely to correct for these issues (i.e., changes in exposure status and/ or unbalanced co-exposures) used?<br><br>Go to Bias due to departures from intended exposures: Risk of bias judgement. | Remarks:<br>- Check whether there were repeated exposure assessments and whether these were used in time-varying models.<br>- Check whether there was any type of sensitivity analysis.<br>E.g., the study calculated an intraclass correlation coefficient (ICC) of exposure assessment with time and the ICC is high. | -**Y** The study has measured exposure and important confounders more than once and performed a time-varying exposure (and confounder) analysis.<br>-**PY** The study has measured exposure more than once in a subset of the cohort and either performed a time-varying exposure (and confounder) analysis in that subset or estimated a correlation between the several exposure (and confounder) measures and the correlation was high.<br>-**PN** The study has measured only exposure more than once and either performed a time-varying exposure but not confounder analysis in the total sample or in a subset or estimated a correlation between the exposure measures and the correlation was adequate.<br>-**N** The study has not performed a time-varying exposure analysis. |
| RoB judgement | | |

| Question | Issues/ Remarks | Prompting questions/ indicators |
|---|---|---|
| Low risk of bias<br>The study is comparable to a well-performed randomized trial with regard to this domain. | There were no changes in the exposure status that were likely to impact the outcome, and any important co-exposures were balanced across exposure groups. ***No study should have low RoB in this domain.*** | |
| Moderate risk of bias<br>The study is sound for an observational study with regard to this domain but cannot be considered comparable to a well-performed randomized trial. | (i) There were changes in exposures status or important coexposures were not balanced across groups;<br>and<br>(ii) the impact on the outcome is expected to be slight or measurement and/or adjustment techniques were used to correct for the issues. | |
| Serious risk of bias<br>The study has some important problems. | (i) There were changes in exposure status or important co-exposures were not balanced across groups that were likely to impact the outcome;<br>and<br>(ii) No or inappropriate measurement and/or adjustment techniques were used to correct for the issues. | |
| Critical risk of bias<br>The study is too problematic to provide any useful evidence. | (i) There were substantial changes in exposures status, or important co-exposures were not balanced across groups, that were likely to impact the outcome;<br>and<br>(ii) No or inappropriate measurement and/or adjustment techniques were used to correct for the issues. | |

#### 5. Bias due to missing data.

| Question | Issues/ Remarks | Prompting questions/ indicators |
|---|---|---|
| 5.1. Were there missing outcome data? | Issues:<br>- Missing outcome measurements due to loss to follow-up or any missed outcome assessment may lead to bias in the results. | -**N/PN** The participants were completely followed-up and assessed for outcomes or the percent of missingness was <10%.<br>-**N/PN** No mention of missingness but outcomes were ascertained via record linkages.<br>-**Y/PY** ≥10% of the participants missing an outcome.<br>-**NI** There was no information about missing outcome data. |
| 5.2. Were participants excluded due to missing data on exposure status? | Issues: | -**N/PN** The participants were completely assessed for exposures or the percent of missingness was <10%. |

| Question | Issues/ Remarks | Prompting questions/ indicators |
| --- | --- | --- |
| | - Differential inclusion of participants in the analysis may lead to bias in the results. | -**Y/PY** ≥10% of the participants missing the exposure.<br>**-Y/PY** Study mentions that only participants with complete data on exposure were included.<br>**-NI** There was no information about missing data on exposure status. |
| 5.3. Were participants excluded due to missing data on other variables (besides outcome data and exposure status) needed for the analysis? | Issues:<br>- Missing covariates needed in the analysis may lead to incomplete adjustment or residual confounding.<br>Remarks:<br>- Please consult the external list of important confounding variables. | -**N/PN** The participants were completely assessed for important confounders or the percent of missingness (cumulatively) was <10%.<br>**-Y/PY** ≥10% of the participants (cumulatively) missing important confounders.<br>**-Y/PY** Study mentions that only participants with complete data on confounders were included.<br>**-NI** There was no information about missing data on confounders. |
| 5.4. **If Y or PY to 5.1, 5.2 or 5.3**: Are the proportion of participants and reasons for missing data similar across exposure groups? | Issues:<br>- Differential rates of missing data between the exposure groups may lead to bias in the results.<br>Remarks:<br>- To answer, the study has to provide the information, such as, in a sub-analysis comparing the proportion (and reason) of participants with missing outcome (or confounder) data across exposure groups. | **-Y** The proportions of missing outcome (or confounder) data were found to be the same or similar (<10%) across exposure groups.<br>**-Y** The proportions of missing data were dissimilar (≥10% difference) BUT reasons for missingness did not differ across exposure groups.<br>**-PY** The proportions of missing data were slightly different (10-<15%) AND reasons for missingness were either not presented or differ across exposure groups.<br>**-PN** The proportions of missing data were substantially different (15-<20%) AND reasons for missingness were either not presented or differ across exposure groups.<br>**-N** The proportions of missing data were critically different (≥20%) AND reasons for missingness were either not presented or differ across exposure groups.<br>-**NI** No sub-analysis performed. Studies with missing exposure data are highly unlikely to have conducted such a sensitivity analysis. |

| Question | Issues/ Remarks | Prompting questions/ indicators |
| --- | --- | --- |
| 5.5. **If Y or PY to 5.1, 5.2 or 5.3**: Were appropriate statistical methods used to account for missing data?<br><br>Go to Bias due to missing data: Risk of bias judgement. | Remarks:<br>- The use of appropriate statistical methods would remove/minimise any risk of bias. | -**Y/ PY** The study performed a multiple imputation analysis, inverse probability weighting, g-formula, marginal structural model (or any other equivalent method) for missing data.<br>-**PN** The study performed a simple imputation analysis for missing data.<br>-**PN** The study used a missing-indicator for categorical variables.<br>**-PN** The study performed a complete case analysis and a sensitivity analysis for missing data using a simple imputation method and found no materially different results.<br>-**N** The study did not account for missing data using any statistical methods.<br>-**N** The study had only performed a complete case analysis. |
| RoB judgement | | |
| Low risk of bias<br>The study is comparable to a well-performed randomized trial with regard to this domain. | (i) Data were reasonably complete;<br>or<br>(ii) Proportions of and reasons for missing participants were similar across exposure groups;<br>or<br>(iii) The analysis addressed missing data and is likely to have removed any risk of bias. | |
| Moderate risk of bias<br>The study is sound for an observational study with regard to this domain but cannot be considered comparable to a well-performed randomized trial. | (i) Proportions of and reasons for missing participants differ slightly across exposure groups;<br>and<br>(ii) The analysis is unlikely to have removed the risk of bias arising from the missing data. | |
| Serious risk of bias<br>The study has some important problems. | (i) Proportions of missing participants differ substantially across exposures;<br>or<br>Reasons for missingness differ substantially across exposures;<br>and<br>(ii) The analysis is unlikely to have removed the risk of bias arising from the missing data;<br>or | |

| Question | Issues/ Remarks | Prompting questions/ indicators |
|---|---|---|
| | Missing data were addressed inappropriately in the analysis;<br>or<br>The nature of the missing data means that the risk of bias cannot be removed through appropriate analysis. | |
| Critical risk of bias<br>The study is too problematic to provide any useful evidence. | (i) (Unusual) There were critical differences between exposures in participants with missing data; and<br>(ii) Missing data were not, or could not, be addressed through appropriate analysis. | |
| **No information** on which to base a judgement about risk of bias for this domain. | There is too little information to make a judgement (for example if only an abstract is available for the study). | |

#### 6. Bias in measurement of outcomes.

| Question | Issues/ Remarks | Prompting questions/ indicators |
|---|---|---|
| 6.1. Could the outcome measure have been influenced by knowledge of the exposure received? | Remarks:<br>- Outcome measures such as cancer incidence and mortality involve negligible assessor judgment. Risk of bias due to measurement of these outcomes would be expected to be low. | -**N** Mortality (all-cause mortality, cause-specific mortality) or any primary cancer outcomes were ascertained by record linkages to death registries, cancer registries, or hospital records.<br>-**N** Mortality outcomes were reported by their proxies and verified by record linkages to death registries, cancer registries, or hospital records.<br>-**N** Incidence outcomes were based on biopsy/ measures of laboratory testing.<br>-**PN** Mortality outcomes were reported by their proxies, and it has not been verified or verification not reported.<br>-**PN** Outcomes were self-reported by the participants or their proxies, with an objective verification.<br>-**PY** Outcomes were self-reported by the participants or their proxies, without any objective verification. |

| **Question** | **Issues/ Remarks** | **Prompting questions/ indicators** |
|---|---|---|
| 6.2. Were outcome assessors aware of the exposure received by study participants? | Remarks:<br>- "Outcome assessors" here is interpreted as "the researchers who processed the data".<br>- For self-reported measures, "outcome assessors" are "the participants" who reported the data. | -**N** Outcomes were ascertained by record linkages to death registries, cancer registries, hospital records, or laboratory measurements.<br>-**N** Outcomes were ascertained by assessors "blinded" to the exposure status of the participants.<br>-**PN** Outcomes were ascertained by independent assessors without "blinding" or "blinding" not reported.<br>-**PY** Any outcomes self-reported by the participants or their proxies, without mention of any further ascertainment.<br>-**NI** No information was reposted about outcome ascertainment. |
| 6.3. Were the methods of outcome assessment the same across exposure groups? | Issues:<br>Outcome assessment methods (detection methods and threshold, time point, definition, and measurements) should be compatible across exposure groups.<br>Remarks:<br>- Studies normally only describe one outcome assessment method that was applied to all participants, any difference would likely be described explicitly. | -**Y/ PY** Same outcome detection methods, thresholds, time point (ended on the date of event or end of study follow-up, whichever came first), same definition, same measurements, across exposure groups.<br>-**PY** It was not explicitly reported whether outcome detection methods were the same across exposure groups.<br>-**PN** The authors have explicitly described some differences of outcome ascertainment e.g., definition, detection methods, threshold, and measurement across exposure groups.<br>-**N** The authors have explicitly described very different outcome assessment, e.g., definition, detection methods, threshold, and measurement across exposure groups. |
| 6.4. Were any systematic errors during measurement of the outcome related to exposure received?<br><br>Go to Bias in measurement of outcomes: Risk of bias judgement. | Issues:<br>- This question refers to differential misclassification of outcomes.<br>- Systematic errors in measuring the outcome, if present, could cause bias if they | -**PY** The outcome was self-reported by the participants.<br>-**PY** The authors have explicitly described different outcome assessment methods across exposure groups.<br>-**N/ PN** All other instances. |

| Question | Issues/ Remarks | Prompting questions/ indicators |
|---|---|---|
| | are related to exposure (or to a confounder of the exposure-outcome association).<br>Remarks:<br>Example: Outcome was detected by health screening/ examination and there was differential attendance related to the lifestyle of the participants. | |
| RoB judgement | | |
| Low risk of bias<br>The study is comparable to a well-performed randomized trial with regard to this domain. | (i) The methods of outcome assessment were comparable across exposure groups;<br>and<br>(ii) The outcome measure was unlikely to be influenced by knowledge of the exposure received by study participants (i.e. is objective) or the outcome assessors were unaware of the exposure received by study participants ***or there was no information provided about outcome ascertainment***;<br>and<br>(iii) Any error in measuring the outcome is unrelated to exposure status. | |
| Moderate risk of bias<br>The study is sound for an observational study with regard to this domain but cannot be considered comparable to a well-performed randomized trial. | (i) The methods of outcome assessment were comparable across exposure groups;<br>and<br>(ii) The outcome measure is only minimally influenced by knowledge of the exposure received by study participants ***or there was no information provided about outcome ascertainment***;<br>and<br>(iii) Any error in measuring the outcome is only minimally related to exposure status. | |
| Serious risk of bias<br>The study has some important problems. | (i) The methods of outcome assessment were not comparable across exposure groups;<br>or<br>(ii) The outcome measure was subjective (i.e. vulnerable to influence by knowledge of the exposure received by study participants);<br>and<br>The outcome was assessed by assessors aware of the exposure received by study participants;<br>or<br>(iii) Error in measuring the outcome was related to exposure status. | |

| Question | Issues/ Remarks | Prompting questions/ indicators |
|---|---|---|
| Critical risk of bias<br>The study is too problematic to provide any useful evidence. | The methods of outcome assessment were so different that they cannot reasonably be compared across exposure groups. | |

##### 7. Bias in selection of reported result.

| Question | Issues/ Remarks | Prompting questions/ indicators |
|---|---|---|
| 7.1. Is the reported effect estimate likely to be selected on the basis of the results from multiple outcome measurements within the outcome domain? | Issues:<br>- It is possible to generate multiple effect estimates for different measurements of the same or related outcome/s.<br>- Studies may selectively report a subset of results for the pre-specified outcomes depending on their clinical importance (but without having stated this in the methods/protocol) or statistical significance, magnitude, or direction of the findings, which may threaten the validity of the review.<br>Remarks:<br>- Research protocol and statistical analysis plan is often unclear in observational studies. This question has to be determined from the study aim and methods and their consistency against the results reported. | -**N** All findings for the pre-specified outcomes (primary or secondary) as stated in the methods or the available protocol were reported fully in the results.<br>-**PN** Some of the findings for the pre-specified outcomes (primary or secondary) as stated in the methods or the available protocol were reported briefly as null (data not shown or with a P-value only) because of not achieving statistical significance.<br>-**Y/ PY** Not all findings for the pre-specified outcomes were reported or only statistically significant findings were reported, without mentioning of the statistical significance of the non-reported findings. |
| 7.2. Is the reported effect estimate likely to be selected on the basis of the results from multiple analyses of the exposure-outcome relationship? | Issues:<br>- Studies may analyse an exposure-outcome association in a number of pre-specified manners. Application of such methods generates multiple effect estimates.<br>- If the analyst does not pre-specify the methods to be applied, and multiple estimates are generated but only one or a subset is | -**N** All findings for the pre-specified analyses (different adjustments, different transformation of variables, different analysis techniques) as stated in the methods or the available protocol were reported fully in the results.<br>-**PN** Some of the findings for the pre-specified analyses (different adjustments, different transformation of variables, different analysis |

| Question | Issues/ Remarks | Prompting questions/ indicators |
|---|---|---|
| | reported (e.g., depending on their nature or significance of the findings), there is a risk of selective reporting on the basis of results.<br>Remarks:<br>Examples include unadjusted and adjusted models; use of final value vs change from baseline vs analysis of covariance; different transformations of variables; a continuously scaled outcome converted to categorical data with different cut-points; different sets of covariates used for adjustment; and different analytic strategies for dealing with missing data. | techniques) as stated in the methods or the available protocol were reported briefly/ partially as null (data not shown or with a P-value only) because of not achieving statistical significance.<br>-**Y/ PY** Not all findings for the pre-specified analyses (different adjustments, different transformation of variables, different analysis techniques) were reported or only statistically significant findings were reported, without mentioning of the statistical significance of the non-reported findings. |
| 7.3. Is the reported effect estimate likely to be selected on the basis of the results from different subgroups?<br><br>Go to Bias in selection of reported result: Risk of bias judgement. | Issues:<br>- Particularly with large cohorts often available from routine data sources, it is possible to generate multiple effect estimates for different subgroups or simply to omit varying proportions of the original cohort that is not part of the design. If multiple estimates are generated but only one or a subset is reported, there is a risk of selective reporting on the basis of results. | -**N** All findings for the pre-specified subgroup analyses (exposure and cancer subtypes, population subgroups) as stated in the methods or the available protocol were reported fully in the results.<br>-**PN** Some of the findings for the pre-specified subgroup analyses (exposure and cancer subtypes, population subgroups) as stated in the methods or the available protocol were reported briefly/ partially as null (data not shown or with a P-value only) because of not achieving statistical significance.<br>-**Y/ PY** Not all findings for the pre-specified subgroup analyses (exposure and cancer subtypes, population subgroups) were reported or only statistically significant findings were reported, without mentioning of the statistical significance of the non-reported findings. |
| RoB judgement | | |
| Low risk of bias<br>The study is comparable to a well-performed randomized trial with regard to this domain. | There is clear evidence (usually through examination of a pre-registered protocol or statistical analysis plan) that all reported results correspond to all intended outcomes, analyses and sub-cohorts. | |

| **Question** | **Issues/ Remarks** | **Prompting questions/ indicators** |
|---|---|---|
| Moderate risk of bias<br>The study is sound for an observational study with regard to this domain but cannot be considered comparable to a well-performed randomized trial. | (i) The outcome measurements and analyses are consistent with an a priori plan; or are clearly defined and both internally and externally consistent;<br>and<br>(ii) There is no indication of selection of the reported analysis from among multiple analyses;<br>and<br>(iii) There is no indication of selection of the cohort or subgroups for analysis and reporting on the basis of the results. | |
| Serious risk of bias<br>The study has some important problems. | (i) Outcomes are defined in different ways in the methods and results sections, or in different publications of the study;<br>or<br>(ii) There is a high risk of selective reporting from among multiple analyses;<br>or<br>(iii) The cohort or subgroup is selected from a larger study for analysis and appears to be reported on the basis of the results. | |
| Critical risk of bias<br>The study is too problematic to provide any useful evidence. | (i) There is evidence or strong suspicion of selective reporting of results;<br>and<br>(ii) The unreported results are likely to be substantially different from the reported results. | |

**Supplementary Table  4. List of important confounding variables.**

| Cancer | Confounder* | | Mediator variable(s)** |
|---|---|---|---|
| Bladder | Required | Age, sex, smoking | Body fatness |
| | Desirable | --- | |
| Brain and spinal cord | Required | Age, sex | --- |
| | Desirable | --- | |
| Colorectal | Required | Age, sex | Body fatness |
| | Desirable | Smoking, alcohol, physical activity, red/processed meat | |
| Gallbladder/bile ducts | Required | Age, sex, smoking | Body fatness |
| | Desirable | Gallstones/gallbladder disease/cholecystitis | |
| Kidney | Required | Age, sex, alcohol | Body fatness |
| | Desirable | Physical activity | |
| Liver | Required | Age, sex, alcohol | Body fatness |
| | Desirable | Smoking, race, physical activity, one of: HBV, HCV, liver cirrhosis | |
| Lung | Required | Age, sex, smoking | --- |
| | Desirable | Physical activity, race, environmental tobacco smoke | |
| Lymphoid and hematopoietic | Required | Age, sex | --- |
| | Desirable | --- | |
| Mouth, pharynx, and larynx | Required | Age, sex, smoking, alcohol | Body fatness |
| | Desirable | Race, environmental tobacco smoke | |

| **Cancer** | **Confounder*** | | **Mediator variable(s)**** |
| --- | --- | --- | --- |
| Nasopharynx | Required<br>Desirable | Age, sex, smoking<br>--- | Body fatness |
| Oesophageal adenocarcinoma | Required<br>Desirable | Age, sex<br>Smoking, alcohol, race, physical activity | Body fatness |
| Oesophageal squamous cell carcinoma | Required<br>Desirable | Age, sex, smoking, alcohol<br>Race, physical activity | Body fatness |
| Pancreas | Required<br>Desirable | Age, sex, smoking<br>Pancreatitis, alcohol | Body fatness |
| Melanoma | Required<br>Desirable | Age, sex, at least one of the following: UV light -sun exposure/ indoor tanning, sunscreen use, moles, fair skin, red/blond hair, blue/green eyes, freckles<br>Race | --- |
| Basal cell skin cancer | Required<br>Desirable | Age, sex, at least one of the following: UV light - sun exposure/ indoor tanning, sunscreen use, radiation therapy, immune-suppressing drugs<br>Race | --- |
| Small intestine | Required<br>Desirable | Age, sex<br>Inflammatory Bowel Disease | --- |
| Stomach | Required<br>Desirable | Age, sex, smoking, alcohol<br>Salt/preserved foods, race, Helicobacter Pylori | Body fatness |

| **Cancer** | **Confounder*** | | **Mediator variable(s)**** |
|---|---|---|---|
| Thyroid | Required<br>Desirable | Age, sex<br>One of: Benign thyroid nodules, goiter, thyroiditis | --- |
| Sex-specific cancers | | | |
| Breast - premenopausal | Required<br>Desirable | Age, reproductive factor(s)<br>Alcohol, smoking, HRT/ oral contraceptive use, physical activity | Body fatness |
| Breast - postmenopausal | Required<br>Desirable | Age, reproductive factor(s)<br>Alcohol, smoking, HRT, physical activity | Body fatness |
| Cervix | Required<br>Desirable | Age, HPV, smoking<br>Physical activity | Body fatness |
| Endometrium | Required<br>Desirable | Age, race<br>HRT/ oral contraceptive use, physical activity | Body fatness |
| Ovary | Required<br>Desirable | Age, reproductive factor(s)<br>HRT/ oral contraceptive use | Body fatness |
| Prostate | Required<br>Desirable | Age, race<br>Physical activity, smoking | --- |

***Smoking:** For cancers not related very strongly to smoking such as prostate, breast or colorectal, status is ok. For smoking-related cancers, adjustments should also include duration and intensity.

** **Mediator variable(s)** should be considered as an intermediate factor(s) between soft drinks, sugar sweetened beverages, juices, ultraprocessed foods, and sedentary behaviour and cancer and should not be adjusted for when investigating these associations (please see Supplementary Text 3 for more information).

**Reproductive factor(s):** At least one of the following: age at menarche, age at menopause, parity, lactation.

**Supplementary Table 5. WCRF grading criteria for evidence on diet, nutrition, physical activity and cancer incidence.**

The CUP Global Grading Criteria summary table below provides a summary of the conditions required to reach each conclusion, along with the list of the possible upgrading and downgrading factors.

##### CUP Global Grading Criteria summary table

| Evidence grades | GRADING CRITERIA FOR EVIDENCE ON DIET, NUTRITION, PHYSICAL ACTIVITY AND CANCER INCIDENCE |
|---|---|
| **Convincing** | A combination of all the following points<br><br>• **Volume and type of evidence**: Convincing epidemiological evidence from several well-conducted, independent cohort studies and, when available, supporting evidence from pertinent RCTs<br>• **Consistency of evidence**: This includes absence of unexplained heterogeneity between studies<br>• **Quality of evidence**: This excludes with confidence the possibility that the observed association results from systematic error, including confounding, measurement error, and selection bias<br>• **Level of statistical significance and precision**: When available, summary risk estimates from meta-analyses with sufficiently narrow 95%CI, clearly excluding no association |
| **Probable** | A combination of all the following points<br><br>• **Volume and type of evidence**: Sufficient epidemiological evidence from at least two well-conducted, independent cohort studies<br>• **Consistency of evidence**: This includes absence of unexplained heterogeneity between studies<br>• **Quality of evidence**: This excludes with confidence the possibility that the observed association results from systematic error, including confounding, measurement error, and selection bias<br>• **Level of statistical significance and precision**: When available, summary risk estimates from meta-analyses indicating an association with 95%CI excluding the null |
| **Limited suggestive** | A combination of both of the following points<br><br>• **Volume and type of evidence**: Evidence from at least two independent cohort studies<br>• **Consistency of evidence**: Some unexplained heterogeneity may be present<br>• **Quality of Evidence:** Not able to rule out the possibility that the observed association may primarily result from systematic error, including confounding and selection bias<br>• **Level of statistical significance and precision:** When available, summary risk estimates from meta-analyses suggesting an association, but with 95% CI that may include the null |
| **Limited – no conclusion** | Any of the following reasons – or a combination of them:<br><br>• **Volume and type of evidence**: Too few studies available. Note there may be instances where the Panel considers an individual study warrants a *Limited-no conclusion* judgement<br>• **Consistency of evidence**: Inconsistency of direction of effect across different studies<br>• **Quality of the evidence**: Poor quality of studies (for example, lack of adjustment for known confounders)<br>• **Level of statistical significance and precision:** Too imprecise risk estimate(s) |
| **Substantial effect on risk unlikely** | A combination of all the following points<br><br>• **Volume and type of evidence**: Convincing epidemiological evidence from several well-conducted, independent cohort studies of an absence of effect and, when available, supporting evidence of no effect from pertinent RCTs<br>• **Consistency of evidence**: This includes absence of unexplained heterogeneity between studies<br>• **Quality of evidence**: This excludes with confidence the possibility that the observed association results from systematic error, including confounding, measurement error, and selection bias<br>• **Level of statistical significance and precision**: When available, summary risk estimates from meta-analyses close to 1.0, with sufficiently narrow 95%CI clearly indicating that an association is unlikely |

Note that there are exposures for which there is insufficient data to make a judgement (typically when only a single cohort study, of moderate quality, is available for a specific exposure-outcome pair of interest). These do not qualify for any of the evidence grades above.

In addition, there are associations for which the expert panel did not grade the quality of evidence. These include:

1- Combined exposures (e.g., SSBs + Carbonated SSBs; ASBs and Carbonated ASBs; Fruit juice + Pure Fruit juice) or outcome types (Incidence + Mortality): the approach was not grading the underlying single exposures (SSBs, Carbonated SSBs, ASBs, Carbonated ASBs, Fruit juice, Pure fruit juice) or outcome types (Incidence).

2- Cancer subsites or subtypes (ER+ or ER- breast cancer; localised or advanced prostate cancer; distal and proximal colon; distal stomach): The numbers of studies for these cancer subtypes/subsites are often too low to inform evidence grading, unlike for colon or rectum cancers and cancers among pre- or postmenopausal women. For the latter, overall cancers (colorectal cancer, breast cancer) are also graded, utilising all available evidence.

3- Exposure components (Fruit drinks; Sugar-sweetened cola; Apple juice): the approach was not grading the individual components as we have grading for the major exposure (e.g., fruit drinks and sugar-sweetened cola are components of SSBs). For each cancer, the evidence was graded utilising all available evidence.

4- Cancer mortality: The evidence for cancer mortality outcome was not graded and considered as supportive evidence.

**Upgrading factors**

- Presence of a **plausible gradient (‘dose-response’)** in the association. Such a gradient need not be linear or even in the same direction across the different levels of exposure, so long as this can be explained plausibly.
- **Strong and plausible mechanistic evidence**, either from human studies or relevant animal models, that typical human exposures can lead to relevant cancer outcomes.
- A particularly **large summary effect size** (an odds ratio or relative risk of 2.0 or more - depending on the unit of exposure - with a 95%CI excluding the null), after appropriate control for confounders.
- Evidence from well-conducted, pertinent (in terms of exposure, dosage, and duration of intervention and follow-up period) **randomised controlled trials in humans supporting** the epidemiological evidence.
- Additional consideration regarding **Mendelian randomisation (MR) studies:**

There is currently no formal method for assessing the quality of MR studies in a rapidly evolving field or for determining how well these studies apply to specific nutritional exposures. Therefore, MR studies are not formally listed as an upgrading factor. However, all MR studies that appear potentially relevant will be presented to the Panel, who will decide, case by case, whether to incorporate that evidence into its judgment and, if so, how.

**Downgrading factors**

- Evidence from well-conducted, pertinent (in terms of exposure, dosage, and duration of intervention and follow-up period) **randomised controlled trials in humans unsupportive** of the epidemiological evidence
- Evidence of **publication bias**

**Supplementary Table 6. Studies excluded and reasons of exclusion from the potentially eligible studies after further assessment.**

| Reason of exclusion | Number of studies | Reference |
|---|---|---|
| **17 publications excluded after further inspection** | | |
| Examined in situ breast cancer | 1 | [211] |
| Examined other exposures (e.g., black tea, green tea, mixed beverages, aspartame, sugar from sugary beverages, fruit plus fruit juices, and dietary indexes) | 6 | [212-217] |
| Superseded publications | 6 | [218-223] |
| Examined adenoma | 1 | [224] |
| Examined cancer recurrence | 1 | [225] |
| Retrospective case-control | 1 | [226] |
| Examined total cancer mortality | 1 | [227] |

**Supplementary Table 7. List of studies included in the systematic review on sugar sweetened beverages and studies included and not included in the dose-response meta-analysis (cancers are ordered by ICD-10 codes).**

| Cancer | Exposure | Outcome | Publications identified | Publications included in various meta-analyses | Publications included in the descriptive synthesis only | Superseded |
|---|---|---|---|---|---|---|
| Head and neck | Sugar sweetened beverages | Incidence | 1 publication[172] | N/A | 1 publication[172] | NA |
| Larynx, oral cavity, and pharynx | Sugar sweetened beverages | Mortality | 1 publication[113] | N/A | 1 publication[113] | NA |
| Oral cavity, larynx, pharynx, and oesophagus | Red fruit drinks | Incidence | 1 publication[90] | N/A | 1 publication[90] | NA |
| Oral cavity, larynx, pharynx, and oesophagus | Lemonade | Incidence | 1 publication[90] | N/A | 1 publication[90] | NA |
| Oesophageal | Sugar sweetened beverages | Mortality | 1 publication[113] | N/A | 1 publication[113] | NA |
| Small intestine | Sugar sweetened beverages | Mortality | 1 publication[113] | N/A | 1 publication[113] | NA |
| Stomach | Sugar sweetened beverages | Incidence | 2 publications[84,97] | 2 publications[84,97] | N/A | NA |
| Stomach cardia | Sugar sweetened beverages | Incidence | 2 publications[70,84] | 2 publications[70,84] | N/A | NA |
| Stomach non-cardia | Sugar sweetened beverages | Incidence | 1 publication[84] | N/A | 1 publication[84] | NA |
| Stomach distal | Sugar sweetened beverages | Incidence | 1 publication[34] | N/A | 1 publication[34] | NA |
| Stomach | Sugar sweetened beverages | Mortality | 1 publication[113] | N/A | 1 publication[113] | NA |
| Colorectal | Sugar sweetened beverages | Incidence | 16 publications[38,40,44,70,78,81,97,101,108,122,129,135,168,178,219,223] | 12 publications[38,40,44,70,81,97,101,108,129,135,168,178] | 2 publications[78,122] Insufficient data format | 2 publications[219,223] |
| Colorectal | Sugar sweetened beverages | Mortality | 3 publications[113,126,178] | 3 publications[113,126,178] | N/A | |
| Early onset colorectal | Sugar sweetened beverages, adulthood | Incidence | 1 publication[72] | N/A | 1 publication[72] | N/A |
| Early onset colorectal | Substitution of sugar sweetened beverages with artificially sweetened beverages | Incidence | 1 publication[72] | N/A | 1 publication[72] | N/A |
| Early onset colorectal | Substitution of sugar sweetened beverages with fruit juice | Incidence | 1 publication[72] | N/A | 1 publication[72] | N/A |

| Cancer | Exposure | Outcome | Publications identified | Publications included in various meta-analyses | Publications included in the descriptive synthesis only | Superseded |
|---|---|---|---|---|---|---|
| Early onset colorectal | Sugar sweetened beverages, adolescence | Incidence | 1 publication[72] | N/A | 1 publication[72] | N/A |
| Colorectal | Sweetened bottled water or tea | Incidence | 1 publication[135] | N/A | 1 publication[135] | N/A |
| Colorectal | Fruit drinks | Incidence | 1 publication[135] | N/A | 1 publication[135] | N/A |
| Colorectal | Sugar sweetened beverages without fruit drinks | Incidence | 1 publication[135] | N/A | 1 publication[135] | N/A |
| Colon | Sugar sweetened beverages | Incidence | 6 publications[38,78,81,101,129,178] | 5 publications[38,81,101,129,178] | 1 publication[78] Insufficient data format | NA |
| Colon | Carbonated sugar sweetened beverages | Incidence | 1 publication[182] | 1 publication[182] | N/A | N/A |
| Colon | Sugar sweetened beverages | Mortality | 1 publication[109] | 1 publication[109] | N/A | N/A |
| Proximal colon | Sugar sweetened beverages | Incidence | 4 publications[81,101,135,178] | 4 publications[81,101,135,178] | N/A | N/A |
| Proximal colon | Substitution of sugar sweetened beverages with artificially sweetened beverages | Incidence | 1 publication[178] | 1 publication[178] | N/A | N/A |
| Proximal colon | Substitution of sugar sweetened beverages with artificially sweetened beverages | Mortality | 1 publication[178] | 1 publication[178] | N/A | N/A |
| Distal colon | Sugar sweetened beverages | Incidence | 4 publications[81,101,135,178] | 4 publications[81,101,135,178] | N/A | NA |
| Distal colon | Substitution of sugar sweetened beverages with artificially sweetened beverages | Incidence | 1 publication[178] | N/A | 1 publication[178] No risk estimate | N/A |
| Distal colon | Sugar sweetened beverages | Mortality | 1 publication[178] | 1 publication[178] | N/A | N/A |
| Distal colon | Substitution of sugar sweetened beverages with | Mortality | 1 publication[178] | N/A | 1 publication[178] No risk estimate | N/A |

| Cancer | Exposure | Outcome | Publications identified | Publications included in various meta-analyses | Publications included in the descriptive synthesis only | Superseded |
|---|---|---|---|---|---|---|
| | artificially sweetened beverages | | | | | |
| Rectal | Sugar sweetened beverages | Incidence | 4 publications[78,81,101,178] | 3 publications[81,101,178] | 1 publication[78]<br>Insufficient data format | N/A |
| Rectal | Substitution of sugar sweetened beverages with artificially sweetened beverages | Incidence | 1 publication[178] | N/A | 1 publication[178]<br>No risk estimate | N/A |
| Rectal | Sugar sweetened beverages | Mortality | 1 publication[178] | 1 publication[178] | N/A | N/A |
| Rectal | Substitution of sugar sweetened beverages with artificially sweetened beverages | Mortality | 1 publication[178] | N/A | 1 publication[178]<br>No risk estimate | N/A |
| Liver | Sugar sweetened beverages | Incidence/mortality | 7 publications[79,105,106,113,159,186,222] | 5 publications[79,106,113,159,186] | 1 publication[105]<br>Insufficient data format | 1 publication[222] |
| Liver | Carbonated sugar sweetened beverages | Incidence | 2 publications[79,187] | 1 publication[79] | 1 publication[187]<br>Insufficient data format | N/A |
| Liver | Sugar sweetened beverages without fruit drinks | Incidence | 1 publication[186] | N/A | 1 publication[186] | NA |
| Liver | Fruit drinks | Incidence | 3 publications[79,186,187] | 2 publications[79,186] | 1 publication[187]<br>Insufficient data format | NA |
| Liver | Sugar sweetened fruit drinks | Incidence | 1 publication[79] | 1 publication[79] | N/A | N/A |
| Gallbladder | Sugar sweetened beverages | Mortality | 1 publication[113] | N/A | 1 publication[113] | NA |
| Pancreas | Sugar sweetened beverages | Incidence/mortality | 7 publications[39,45,80,113,127,184,188] | 5 publications[39,45,113,127,184] | 2 publications[80,188]<br>Insufficient data format | N/A |
| Pancreas | Carbonated sugar sweetened beverages | Incidence/mortality | 2 publications[64,131] | 2 publications[64,131] | N/A | NA |
| Pancreas | Sugar sweetened cola | Incidence/mortality | 1 publication[151] | 1 publication[151] | N/A | NA |
| Pancreatic adenocarcinoma | Carbonated sugar sweetened beverages | Incidence | 1 publication[64] | 1 publication[64] | N/A | NA |

| Cancer | Exposure | Outcome | Publications identified | Publications included in various meta-analyses | Publications included in the descriptive synthesis only | Superseded |
|---|---|---|---|---|---|---|
| Lung | Sugar sweetened beverages | Incidence | 3 publications[37,97,190] | 2 publications[97,190] | 1 publication[37] Insufficient data format | NA |
| Lung | Sugar sweetened beverages | Mortality | 2 publications[109,113] | 2 publications[109,113] | N/A | NA |
| Melanoma | Sugar sweetened beverages | Mortality | 1 publication[113] | N/A | 1 publication[113] | NA |
| Breast | Sugar sweetened beverages | Incidence | 10 publications[38,44,97,108,128,145,146,169,220,223] | 7 publications[38,44,97,108,128,145,146] | 1 publication[169] Insufficient data format | 2 publications[220,223] |
| Breast | Caffeinated sugar sweetened cola | Incidence | 1 publication[75] | N/A | 1 publication[75] | N/A |
| Breast | Decaffeinated sugar sweetened cola | Incidence | 1 publication[75] | N/A | 1 publication[75] | N/A |
| Breast | Sugar sweetened beverages | Mortality | 3 publications[109,113,126] | 3 publications[109,113,126] | N/A | N/A |
| Postmenopausal breast | Sugar sweetened beverages | Incidence | 7 publications[38,44,70,128,130,145,146] | 5 publications[38,44,70,128,145] | 2 publications[130,146] Insufficient data format | N/A |
| Premenopausal breast | Sugar sweetened beverages | Incidence | 5 publications[38,44,128,145,146] | 4 publications[38,44,128,145] | 1 publication[146] Insufficient data format | NA |
| Basal like breast | Sugar sweetened beverages | Incidence | 1 publication[145] | 1 publication[145] | N/A | NA |
| Luminal A breast | Sugar sweetened beverages | Incidence | 1 publication[145] | 1 publication[145] | N/A | NA |
| Luminal B breast | Sugar sweetened beverages | Incidence | 1 publication[145] | 1 publication[145] | N/A | NA |
| HER2 breast | Sugar sweetened beverages | Incidence | 1 publication[145] | 1 publication[145] | N/A | NA |
| ER- breast | Sugar sweetened beverages | Incidence | 1 publication[145] | 1 publication[145] | N/A | NA |
| ER+ breast | Sugar sweetened beverages | Incidence | 1 publication[145] | 1 publication[145] | N/A | NA |
| ER-/PR- breast | Sugar sweetened beverages | Incidence | 1 publication[128] | N/A | 1 publication[128] | NA |
| ER+/PR+ breast | Sugar sweetened beverages | Incidence | 1 publication[128] | N/A | 1 publication[128] | NA |
| ER+ or PR+ breast | Sugar sweetened beverages | Incidence | 1 publication[128] | N/A | 1 publication[128] | NA |
| Endometrial | Sugar sweetened beverages | Incidence | 3 publications[38,70,74] | 3 publications[38,70,74] | N/A | NA |
| Type I endometrial | Sugar sweetened beverages | Incidence | 2 publications[38,74] | 2 publications[38,74] | N/A | NA |
| Type II endometrial | Sugar sweetened beverages | Incidence | 1 publication[74] | N/A | 1 publication[74] | NA |
| Endometrial | Sugar sweetened beverages | Mortality | 1 publication[113] | N/A | 1 publication[113] | NA |

| Cancer | Exposure | Outcome | Publications identified | Publications included in various meta-analyses | Publications included in the descriptive synthesis only | Superseded |
|---|---|---|---|---|---|---|
| Ovarian | Sugar sweetened beverages | Incidence | 2 publications[38,70] | 2 publications[38,70] | N/A | NA |
| Ovarian | Sugar sweetened beverages | Mortality | 1 publication[113] | N/A | 1 publication[113] | NA |
| Prostate | Sugar sweetened beverages | Incidence | 6 publications[44,47,70,97,108,223] | 4 publications[44,70,97,108] | 1 publication[47]<br>Insufficient data format | 1 publication[223] |
| Prostate | Sugar sweetened beverages | Mortality | 4 publications[42,109,113,126] | 3 publications[109,113,126] | 1 publication[42]<br>Insufficient data format | NA |
| Kidney | Sugar sweetened beverages | Incidence | 4 publications[66,70,100,173] | 4 publications[66,70,100,173] | N/A | NA |
| Kidney | Carbonated sugar sweetened beverages | Incidence | 1 publication[98] | 1 publication[98] | | NA |
| Kidney | Fruit drinks | Incidence | 1 publication[36] | N/A | 1 publication[36] | NA |
| Kidney | Sugar sweetened beverages | Mortality | 2 publications[66,113] | 2 publications[66,113] | N/A | NA |
| Bladder | Sugar sweetened beverages | Incidence | 3 publications[46,82,100] | N/A | 3 publications[46,82,100]<br>Insufficient data format | NA |
| Bladder | Sugar sweetened beverages | Mortality | 1 publication[113] | N/A | 1 publication[113] | NA |
| Brain | Sugar sweetened beverages | Mortality | 1 publication[113] | N/A | 1 publication[113] | NA |
| Thyroid | Sugar sweetened beverages | Incidence | 2 publications[53,180] | N/A | 2 publications[53,180]<br>Insufficient data format | NA |
| Papillary thyroid | Sugar sweetened beverages | Incidence | 1 publication[180] | N/A | 1 publication[180]<br>Insufficient data format | NA |
| Follicular thyroid | Sugar sweetened beverages | Incidence | 1 publication[180] | N/A | 1 publication[180]<br>Insufficient data format | NA |
| Leukaemia | Sugar sweetened beverages | Mortality | 1 publication[113] | N/A | 1 publication[113] | NA |
| Multiple Myeloma | Sugar sweetened beverages | Mortality | 1 publication[113] | N/A | 1 publication[113] | NA |
| Non-Hodgkin lymphoma | Sugar sweetened beverages | Mortality | 1 publication[113] | N/A | 1 publication[113] | NA |
| Leukaemia | Carbonated sugar sweetened beverages | Incidence | 1 publication[150] | 1 publication[150] | N/A | NA |
| Multiple myeloma | Carbonated sugar sweetened beverages | Incidence | 1 publication[150] | 1 publication[150] | N/A | NA |
| Non-Hodgkin Lymphoma | Carbonated sugar sweetened beverages | Incidence | 2 publications[114,150] | 2 publications[114,150] | N/A | NA |

**Supplementary Table 8. List of studies included in the systematic review on artificially sweetened drinks and studies included and not included in the dose-response meta-analysis (cancers are ordered by ICD-10 codes).**

| Cancer | Exposure | Outcome | Publications identified | Publications included in various meta-analyses | Publications included in the descriptive synthesis only | Superseded |
|---|---|---|---|---|---|---|
| Larynx/oral cavity/pharynx | Artificially sweetened beverages | Mortality | 1 publication[113] | N/A | 1 publication[113] | N/A |
| Oesophageal | Artificially sweetened beverages | Mortality | 1 publication[113] | N/A | 1 publication[113] | N/A |
| Gastric cardia | Artificially sweetened beverages | Incidence | 1 publication[70] | N/A | 1 publication[70] | N/A |
| Stomach | Artificially sweetened beverages | Mortality | 1 publication[113] | N/A | 1 publication[113] | N/A |
| Small intestinal | Artificially sweetened beverages | Mortality | 1 publication[113] | N/A | 1 publication[113] | N/A |
| Colorectal | Artificially sweetened beverages | Incidence | 4 publications[44,70,78,168] | 2 publications[70,168] | 2 publications: [44], Implausible effect size per 10 g/day; [78], Insufficient data format | N/A |
| Early onset colorectal | Artificially sweetened beverages | Incidence | 1 publication[72] | N/A | 1 publication[72] | N/A |
| Rectal | Artificially sweetened beverages | Incidence | 1 publication[78] | N/A | 1 publication[78] | N/A |
| Colorectal | Artificially sweetened beverages | Mortality | 2 publications[113,126] | 2 publications[113,126] | N/A | N/A |
| Colon | Artificially sweetened beverages | Mortality | 1 publication[109] | 1 publication[109] | N/A | N/A |
| Liver | Artificially sweetened beverages | Incidence/mortality | 5 publications[79,113,159,186,187] | 4 publications[79,113,159,186] | 1 publication[187] Insufficient data format | N/A |
| Liver | Carbonated artificially sweetened beverages | Incidence/mortality | 2 publications[79,187] | 1 publication[79] | I publication[187] | N/A |
| Liver | Artificially sweetened fruit drinks | Incidence/mortality | 1 publication[79] | 1 publication[79] | N/A | N/A |
| Gallbladder | Artificially sweetened beverages | Mortality | 1 publication[113] | N/A | 1 publication[113] | N/A |
| Pancreas | Artificially sweetened beverages | Incidence/mortality | 3 publications[39,113,127] | 3 publications[39,113,127] | N/A | N/A |
| Pancreas | Carbonated artificially sweetened beverages | Incidence/mortality | 1 publication[151] | 1 publication[151] | N/A | N/A |
| Pancreas | Artificially sweetened cola | Incidence/mortality | 1 publication[151] | 1 publication[151] | N/A | N/A |
| Lung | Artificially sweetened beverages | Incidence | 1 publication[177] | N/A | 1 publication[177] | N/A |
| Lung | Artificially sweetened beverages | Mortality | 2 publications[109,113] | 2 publications[109,113] | N/A | N/A |
| Melanoma | Artificially sweetened beverages | Mortality | 1 publication[113] | N/A | 1 publication[113] | N/A |
| Breast | Artificially sweetened beverages | Incidence | 3 publications[44,49,145] | 2 publications[49,145] | 1 publication[44] | N/A |

| Cancer | Exposure | Outcome | Publications identified | Publications included in various meta-analyses | Publications included in the descriptive synthesis only | Superseded |
|---|---|---|---|---|---|---|
| | | | | | Implausible effect size per 10 g/day | |
| Breast | Artificially sweetened caffeinated cola | Incidence | 1 publication[75] | N/A | 1 publication[75] | N/A |
| Breast | Artificially sweetened decaffeinated cola | Incidence | 1 publication[75] | N/A | 1 publication[75] | N/A |
| Postmenopausal | Artificially sweetened beverages | Incidence | 4 publications[44,49,70,145] | 3 publications[49,70,145] | 1 publication[44] Implausible effect size per 10 g/day | N/A |
| Premenopausal | Artificially sweetened beverages | Incidence | 3 publications[44,49,145] | 2 publications[49,145] | 1 publication[44] Implausible effect size per 10 g/day | N/A |
| Luminal A | Artificially sweetened beverages | Incidence | 1 publication[145] | 1 publication[145] | N/A | N/A |
| Luminal B | Artificially sweetened beverages | Incidence | 1 publication[145] | 1 publication[145] | N/A | N/A |
| Basal like | Artificially sweetened beverages | Incidence | 1 publication[145] | 1 publication[145] | N/A | N/A |
| HER2 | Artificially sweetened beverages | Incidence | 1 publication[145] | 1 publication[145] | N/A | N/A |
| RE- breast | Artificially sweetened beverages | Incidence | 1 publication[145] | 1 publication[145] | N/A | N/A |
| RE+ breast | Artificially sweetened beverages | Incidence | 1 publication[145] | 1 publication[145] | N/A | N/A |
| Breast | Artificially sweetened beverages | Mortality | 3 publications[109,113,126] | 3 publications[109,113,126] | N/A | N/A |
| Endometrial | Artificially sweetened beverages | Incidence | 3 publications[49,70,74] | 3 publications[49,70,74] | N/A | N/A |
| Postmenopausal endometrial | Artificially sweetened beverages | Incidence | 3 publications[49,70,74] | 3 publications[49,70,74] | N/A | N/A |
| Endometrial | Artificially sweetened beverages | Mortality | 1 publication[113] | N/A | 1 publication[113] | N/A |
| Ovarian | Artificially sweetened beverages | Incidence | 2 publications[49,70] | 2 publications[49,70] | N/A | N/A |
| Ovarian | Artificially sweetened beverages | Mortality | 1 publication[113] | N/A | 1 publication[113] | N/A |
| Prostate | Artificially sweetened beverages | Incidence | 2 publications[44,70] | N/A | 2 publications[44,70] Implausible effect size per 10 g/day in one study | N/A |
| Prostate | Artificially sweetened beverages | Mortality | 3 publications[109,113,126] | 3 publications[109,113,126] | N/A | N/A |
| Kidney | Artificially sweetened beverages | Incidence | 3 publications[66,70,144] | 3 publications[66,70,144] | N/A | N/A |
| Kidney | Carbonated artificially sweetened beverages | Incidence | 1 publication[98] | 1 publication[98] | N/A | N/A |
| Kidney | Artificially sweetened beverages | Mortality | 2 publications[66,113] | 2 publications[66,113] | N/A | N/A |
| Bladder | Artificially sweetened beverages | Incidence | 1 publication[144] | N/A | 1 publication[144] | N/A |
| Bladder | Artificially sweetened beverages | Mortality | 1 publication[113] | N/A | 1 publication[113] | N/A |
| Brain | Artificially sweetened beverages | Mortality | 1 publication[113] | N/A | 1 publication[113] | N/A |
| Thyroid | Artificially sweetened beverages | Incidence | 1 publication[180] | N/A | 1 publication[180] | N/A |
| Papillary thyroid | Artificially sweetened beverages | Incidence | 1 publication[180] | N/A | 1 publication[180] | N/A |
| Follicular thyroid | Artificially sweetened beverages | Incidence | 1 publication[180] | N/A | 1 publication[180] | N/A |
| Leukaemia | Carbonated artificially sweetened beverages | Incidence | 1 publication[150] | 1 publication[150] | N/A | N/A |
| Multiple Myeloma | Carbonated artificially sweetened beverages | Incidence | 1 publication[150] | 1 publication[150] | N/A | N/A |
| Non-Hodgkin Lymphoma | Carbonated artificially sweetened beverages | Incidence | 1 publication[150] | 1 publication[150] | N/A | N/A |

| Cancer | Exposure | Outcome | Publications identified | Publications included in various meta-analyses | Publications included in the descriptive synthesis only | Superseded |
|---|---|---|---|---|---|---|
| Leukaemia | Artificially sweetened beverages | Mortality | 1 publication[113] | N/A | 1 publication[113] | N/A |
| Multiple Myeloma | Artificially sweetened beverages | Mortality | 1 publication[113] | N/A | 1 publication[113] | N/A |
| Non-Hodgkin Lymphoma | Artificially sweetened beverages | Mortality | 1 publication[113] | N/A | 1 publication[113] | N/A |
| Myeloproliferative neoplasms | Caffeinated diet soft drinks | Incidence | 1 publication[140] | NA | 1 publication[140] | NA |
| Polycythemia vera | Caffeinated diet soft drinks | Incidence | 1 publication[140] | NA | 1 publication[140] | NA |
| Essential thrombocythemia | Caffeinated diet soft drinks | Incidence | 1 publication[140] | NA | 1 publication[140] | NA |
| Myeloproliferative neoplasms | Decaffeinated diet soft drinks | Incidence | 1 publication[140] | NA | 1 publication[140] | NA |
| Polycythemia vera | Decaffeinated diet soft drinks | Incidence | 1 publication[140] | NA | 1 publication[140] | NA |
| Essential thrombocythemia | Decaffeinated diet soft drinks | Incidence | 1 publication[140] | NA | 1 publication[140] | NA |

**Supplementary Table 9. List of studies included in the systematic review on juices and studies included and not included in the dose-response meta-analysis (cancers are ordered by ICD-10 codes).**

| Cancer | Exposure | Outcome | Publications identified | Publications included in various meta-analyses | Publications included in the descriptive synthesis only | Superseded |
|---|---|---|---|---|---|---|
| Head and neck | Fruit juice | Incidence | 2 publications[59,90] | 2 publications[59,90] | N/A | N/A |
| Oral squamous cell | Fruit juice | Incidence | 1 publication[59] | N/A | 1 publication[59] | N/A |
| Oropharynx and hypopharynx squamous cell | Fruit juice | Incidence | 1 publication[59] | N/A | 1 publication[59] | N/A |
| Oesophageal adenocarcinoma | Fruit juice | Incidence | 1 publication[59] | N/A | 1 publication[59] | N/A |
| Oesophageal squamous cell | Fruit juice | Incidence | 1 publication[59] | N/A | 1 publication[59] | N/A |
| Oesophageal | Pure fruit juice | Mortality | 1 publication[76] | N/A | 1 publication[76] | N/A |
| Stomach | Fruit juice | Incidence | 1 publication[59] | N/A | 1 publication[59] | N/A |
| Stomach | Citrus juice | Incidence | 1 publication[43] | N/A | 1 publication[43] | N/A |
| Stomach | Pure orange juice | Incidence | 1 publication[84] | N/A | 1 publication[84] | N/A |
| Stomach | Pure fruit juice | Mortality | 1 publication[76] | N/A | 1 publication[76] | N/A |
| Colorectal | Fruit juice | Incidence | 4 publications[78,87,93,108] | 2 publications[93,108] | 2 publications[78,87]<br>Insufficient data format | N/A |
| Colorectal | Pure fruit juice | Incidence | 2 publications[38,44] | 2 publications[38,44] | N/A | N/A |
| Colorectal | Fruit and vegetable juice | Incidence | 2 publications[81,136] | 2 publications[81,136] | N/A | N/A |
| Colorectal | Tomato juice | Incidence | 1 publication[77] | N/A | 1 publication[77] | N/A |
| Early onset colorectal | Fruit juice | Incidence | 1 publication[72] | N/A | 1 publication[72] | N/A |
| Colorectal | Tomato juice | Mortality | 1 publication[77] | N/A | 1 publication[77] | N/A |
| Colon | Fruit juice | Incidence | 1 publication[78] | N/A | 1 publication[78] | N/A |
| Colon | Pure fruit juice | Incidence | 1 publication[38] | N/A | 1 publication[38] | N/A |
| Colon | Citrus juice | Incidence | 1 publication[165] | N/A | 1 publication[165] | N/A |
| Colon | Other fruit juices (not citrus) | Incidence | 1 publication[165] | N/A | 1 publication[165] | N/A |
| Colon | Fruit and vegetable juice | Incidence | 2 publications[81,158] | 2 publications[81,158] | N/A | N/A |
| Colon | Pure fruit juice | Mortality | 1 publication[76] | N/A | 1 publication[76] | N/A |
| Proximal colon | Fruit and vegetable juice | Incidence | 2 publications[81,158] | 2 publications[81,158] | N/A | N/A |
| Distal colon | Fruit and vegetable juice | Incidence | 2 publications[81,158] | 2 publications[81,158] | N/A | N/A |
| Rectal | Fruit juice | Incidence | 1 publication[78] | N/A | 1 publication[78] | N/A |
| Rectal | Citrus juice | Incidence | 1 publication[165] | N/A | 1 publication[165] | N/A |
| Rectal | Other fruit juices (not citrus) | Incidence | 1 publication[165] | N/A | 1 publication[165] | N/A |
| Rectal | Fruit and vegetable juice | Incidence | 1 publication[81] | N/A | 1 publication[81] | N/A |
| Rectal | Pure fruit juice | Mortality | 1 publication[76] | N/A | 1 publication[76] | N/A |
| Liver | Fruit juice | Incidence/mortality | 2 publications[88,183] | N/A | 2 publications[88,183] | N/A |
| Liver | Pure fruit juice | Mortality | 1 publication[76] | N/A | 1 publication[76] | N/A |
| Liver | Citrus | Incidence | 1 publication[79] | 1 publication[79] | N/A | N/A |
| Liver | Other fruit juice (not citrus) | Incidence | 1 publication[185] | N/A | 1 publication[185] | N/A |
| Liver | Fruit and vegetable juice | Incidence | 1 publication[159] | N/A | 1 publication[159] | N/A |

| Cancer | Exposure | Outcome | Publications identified | Publications included in various meta-analyses | Publications included in the descriptive synthesis only | Superseded |
|---|---|---|---|---|---|---|
| Intrahepatic bile duct | Fruit and vegetable juice | Incidence | 1 publication[159] | N/A | 1 publication[159] | N/A |
| Extrahepatic bile duct | Fruit and vegetable juice | Incidence | 1 publication[159] | N/A | 1 publication[159] | N/A |
| Ampulla of Vater | Fruit and vegetable juice | Incidence | 1 publication[159] | N/A | 1 publication[159] | N/A |
| Liver | Vegetable juice | Incidence | 2 publications[79] | 2 publications[79] | N/A | N/A |
| Liver | Other juices (not citrus and vegetables) | Incidence | 2 publications[79] | 2 publications[79] | N/A | N/A |
| Gallbladder | Pure fruit juice | Mortality | 1 publication[76] | N/A | 1 publication[76] | N/A |
| Pancreas | Fruit juice | Incidence | 4 publications[41,131,167,188] | 3 publications[41,131,167] | 1 publication[188] Insufficient data format | N/A |
| Pancreas | Pure fruit juice | Incidence/mortality | 2 publications[103,154] | N/A | 2 publications[103,154] Insufficient data format | N/A |
| Pancreas | Pure apple juice | Incidence | 1 publication[176] | N/A | 1 publication[176] | N/A |
| Pancreas | Pure orange juice | Incidence | 1 publication[176] | N/A | 1 publication[176] | N/A |
| Pancreas | Citrus juice | Incidence | 1 publication[167] | N/A | 1 publication[167] | N/A |
| Pancreas | Fruit and vegetable juice | Incidence/mortality | 2 publications[125,127] | 2 publications[125,127] | N/A | N/A |
| Pancreas | Tomato juice | Incidence | 1 publication[176] | N/A | 1 publication[176] | N/A |
| Laryngeal squamous cell | Fruit juice | Incidence | 1 publication[59] | N/A | 1 publication[59] | N/A |
| Lung | Citrus juice | Incidence | 2 publications[104,155] | 2 publications[104,155] | N/A | N/A |
| Lung | Other fruit juice (not citrus) | Incidence | 1 publication[165] | N/A | 1 publication[165] | N/A |
| Lung | Grapefruit juice | Incidence | 1 publication[55] | 1 publication[55] | N/A | N/A |
| Lung | Fruit and vegetable juice | Incidence | 2 publications[65,104] | 2 publications[65,104] | N/A | N/A |
| Lung | Fruit juice | Mortality | 1 publication[134] | N/A | 1 publication[134] | N/A |
| Lung | Pure fruit juice | Mortality | 1 publication[76] | N/A | 1 publication[76] | N/A |
| Melanoma | Citrus juice | Incidence | 4 publications[107,110,115,116] | 4 publications[107,110,115,116] | N/A | N/A |
| Melanoma | Orange juice | Incidence | 3 publications[54,110,171] | 3 publications[54,110,171] | N/A | N/A |
| Melanoma | Grapefruit juice | Incidence | 2 publications[110,171] | 2 publications[110,171] | N/A | N/A |
| Non-melanoma skin | Citrus juice | Incidence | 2 publications[111,148] | 2 publications[111,148] | N/A | N/A |
| Non-melanoma skin | Orange juice | Incidence | 1 publication[111] | N/A | 1 publication[111] | N/A |
| Non-melanoma skin | Grapefruit juice | Incidence | 1 publication[111] | N/A | 1 publication[111] | N/A |
| Basal cell carcinoma | Citrus juice | Incidence | 1 publication[107] | N/A | 1 publication[107] | N/A |
| Basal cell carcinoma | Orange juice | Incidence | 1 publication[170] | 1 publication[170] | N/A | N/A |
| Basal cell carcinoma | Grapefruit juice | Incidence | 1 publication[170] | 1 publication[170] | N/A | N/A |
| Squamous cell carcinoma | Citrus juice | Incidence | 1 publication[107] | N/A | 1 publication[107] | N/A |
| Squamous cell carcinoma | Orange juice | Incidence | 1 publication[170] | 1 publication[170] | N/A | N/A |
| Squamous cell carcinoma | Grapefruit juice | Incidence | 1 publication[170] | 1 publication[170] | N/A | N/A |
| Squamous cell carcinoma on head, neck and extremities | Orange juice | Incidence | 1 publication[170] | 1 publication[170] | N/A | N/A |

| Cancer | Exposure | Outcome | Publications identified | Publications included in various meta-analyses | Publications included in the descriptive synthesis only | Superseded |
|---|---|---|---|---|---|---|
| Squamous cell carcinoma on truncal sites | Orange juice | Incidence | 1 publication[170] | 1 publication[170] | N/A | N/A |
| Squamous cell carcinoma on head, neck and extremities | Grapefruit juice | Incidence | 1 publication[170] | 1 publication[170] | N/A | N/A |
| Squamous cell carcinoma on truncal sites | Grapefruit juice | Incidence | 1 publication[170] | 1 publication[170] | N/A | N/A |
| Breast | Fruit juice | Incidence | 5 publications[52,69,108,112,156] | 5 publications[52,69,108,112,156] | N/A | N/A |
| Breast | Fruit juice, early adulthood | Incidence | 1 publication[51] | N/A | 1 publication[51] | N/A |
| Breast | Fruit juice, adolescence | Incidence | 1 publication[51] | N/A | 1 publication[51] | N/A |
| Breast | Pure fruit juice | Incidence | 2 publications[38,44] | 2 publications[38,44] | N/A | N/A |
| Breast | Citrus juice | Incidence | 1 publication[160] | N/A | 1 publication[160] | N/A |
| Breast | Apple juice | Incidence | 1 publication[52] | 1 publication[52] | N/A | N/A |
| Breast | Apple juice, early adulthood | Incidence | 1 publication[51] | N/A | 1 publication[51] | N/A |
| Breast | Apple juice, adolescence | Incidence | 1 publication[51] | N/A | 1 publication[51] | N/A |
| Breast | Orange juice | Incidence | 2 publications[51,52] | I publication[52] | 1 publication[51] In early adulthood | N/A |
| Breast | Orange juice, adolescence | Incidence | 2 publications[51,57] | N/A | 2 publications[51,57] In adolescence | N/A |
| Breast | Orange juice, children | Incidence | 1 publication[121] | N/A | 1 publication[121] | N/A |
| Breast | Grapefruit juice | Incidence | 2 publications[51,86] | 2 publications[51,86] | N/A | N/A |
| Breast | Other fruit juices (not orange and apple) | Incidence | 1 publication[52] | 1 publication[52] | N/A | N/A |
| Breast | Other fruit juices (not citrus and apple) | Incidence | 1 publication[51] | N/A | 1 publication[51] | N/A |
| Breast | Fruit and vegetable juice | Incidence | 2 publications[133,164] | 2 publications[133,164] | N/A | N/A |
| Breast | Pure fruit and vegetable juice | Incidence | 1 publication[49] | N/A | 1 publication[49] | N/A |
| Breast | Tomato juice | Incidence | 1 publication[153] | N/A | 1 publication[153] | N/A |
| Breast | Pure fruit juice | Mortality | 1 publication[76] | N/A | 1 publication[76] | N/A |
| Postmenopausal breast | Fruit juice | Incidence | 3 publications[51,52,83] | 2 publications[52,83] | 1 publication [51] In early adulthood | N/A |
| Postmenopausal breast | Pure fruit juice | Incidence | 2 publications[38,44] | 2 publications[38,44] | N/A | N/A |
| Postmenopausal breast | Citrus juice | Incidence | 1 publication[160] | N/A | 1 publication[160] | N/A |
| Postmenopausal breast | Grapefruit juice | Incidence | 1 publication[86] | N/A | 1 publication[86] | N/A |
| Postmenopausal breast | Pure fruit and vegetable juice | Incidence | 1 publication[49] | N/A | 1 publication[49] | N/A |
| Premenopausal breast | Fruit juice | Incidence | 2 publications[51,52] | 1 publication[52] | 1 publication [51] In early adulthood | N/A |
| Premenopausal breast | Pure fruit juice | Incidence | 2 publications[38,44] | 2 publications[38,44] | N/A | N/A |
| Premenopausal breast | Pure fruit and vegetable juice | Incidence | 1 publication[49] | N/A | 1 publication[49] | N/A |
| ER- breast | Fruit juice | Incidence | 1 publication[63] | N/A | 1 publication[63] | N/A |

| Cancer | Exposure | Outcome | Publications identified | Publications included in various meta-analyses | Publications included in the descriptive synthesis only | Superseded |
|---|---|---|---|---|---|---|
| ER- breast | Orange juice | Incidence | 1 publication[52] | 1 publication[52] | N/A | N/A |
| ER- breast | Apple juice | Incidence | 1 publication[52] | 1 publication[52] | N/A | N/A |
| ER- breast | Other fruit juice juices (not apple and orange) | Incidence | 1 publication | 1 publication[52] | N/A | N/A |
| ER- breast | Fruit and vegetable juice | Incidence | 1 publication[133] | N/A | 1 publication[133] | N/A |
| ER+ breast | Apple juice | Incidence | 1 publication[52] | 1 publication[52] | N/A | N/A |
| ER+ breast | Orange juice | Incidence | 1 publication[52] | 1 publication[52] | N/A | N/A |
| ER+ breast | Other fruit juice juices (not apple and orange) | Incidence | 1 publication[52] | 1 publication[52] | N/A | N/A |
| ER+ breast | Orange juice | Incidence | 1 publication[52] | 1 publication[52] | N/A | N/A |
| ER+ breast | Fruit and vegetable juice | Incidence | 1 publication[133] | N/A | 1 publication[133] | N/A |
| ERα- breast | Fruit juice | Incidence | 1 publication[156] | N/A | 1 publication[156] | N/A |
| ERα+ breast | Fruit juice | Incidence | 1 publication[156] | N/A | 1 publication[156] | N/A |
| ERα+/β- breast | Fruit juice | Incidence | 1 publication[156] | N/A | 1 publication[156] | N/A |
| ERα+/β+ breast | Fruit juice | Incidence | 1 publication[156] | N/A | 1 publication[156] | N/A |
| ERβ- breast | Fruit juice | Incidence | 1 publication[156] | N/A | 1 publication[156] | N/A |
| ERβ+ breast | Fruit juice | Incidence | 1 publication[156] | N/A | 1 publication[156] | N/A |
| Type I endometrial | Fruit juice | Incidence | 1 publication[74] | N/A | 1 publication [74] | N/A |
| Type II endometrial | Fruit juice | Incidence | 1 publication[74] | N/A | 1 publication [74] | N/A |
| Endometrial | Pure fruit juice | Incidence | 1 publication[38] | N/A | 1 publication[38] | N/A |
| Type I endometrial | Pure fruit juice | Incidence | 1 publication[38] | N/A | 1 publication[38] | N/A |
| Endometrial cancer | Fruit and vegetable juice | Incidence | 1 publication[117] | N/A | 1 publication[117] | N/A |
| Endometrial cancer | Pure fruit and vegetable juice | Incidence | 1 publication[49] | N/A | 1 publication[49] | N/A |
| Uterine cervical | Pure fruit juice | Mortality | 1 publication[76] | N/A | 1 publication[76] | N/A |
| Epithelial ovarian | Fruit juice | Incidence | 1 publication[91] | 1 publication[91] | N/A | N/A |
| Ovarian | Pure fruit juice | Incidence | 1 publication[38] | N/A | 1 publication[38] | N/A |
| Epithelial ovarian | Pure fruit juice | Incidence | 1 publication[38] | N/A | 1 publication[38] | N/A |
| Epithelial ovarian | Other fruit juices (not citrus) | Incidence | 1 publication[124] | N/A | 1 publication[124] | N/A |
| Epithelial ovarian | Fruit and vegetable juice | Incidence | 1 publication[118] | N/A | 1 publication[118] | N/A |
| Ovarian | Pure fruit and vegetable juice | Incidence | 1 publication[49] | N/A | 1 publication[49] | N/A |
| Ovarian | Fruit juice | Mortality | 1 publication[149] | N/A | 1 publication[149] | N/A |
| Prostate | Fruit juice | Incidence | 4 publications[47,89,94,108,161] | 2 publications[89,108] | 2 publications[47,161] Insufficient data format | N/A |
| Prostate | Fruit juice, adolescence | Incidence | 1 publication[94] | N/A | 1 publication[94] | N/A |
| Prostate | Pure fruit juice | Incidence | 1 publication[44] | N/A | 1 publication[44] | N/A |
| Prostate | Citrus juice | Incidence | 1 publication[139] | 1 publication[139] | N/A | N/A |
| Prostate | Fruit and vegetable juice | Incidence | 1 publication[139] | 1 publication[139] | N/A | N/A |
| Prostate | Vegetable juice | Incidence | 2 publications[139,221] | 1 publication[139] | N/A | 1 publication[221] |
| Prostate | Tomato juice | Incidence | 2 publications[56,152] | 2 publications[56,152] | N/A | N/A |
| Prostate | Fruit juice | Mortality | 1 publication[94] | N/A | 1 publication[94] | N/A |
| Prostate | Pure fruit juice | Mortality | 1 publication[76] | N/A | 1 publication[76] | N/A |
| Prostate | Citrus juice | Mortality | 1 publication[139] | 1 publication[139] | N/A | N/A |
| Prostate | Fruit and vegetable juice | Mortality | 1 publication[139] | 1 publication[139] | N/A | N/A |

| Cancer | Exposure | Outcome | Publications identified | Publications included in various meta-analyses | Publications included in the descriptive synthesis only | Superseded |
|---|---|---|---|---|---|---|
| Prostate | Vegetable juice | Mortality | 1 publication[139] | 1 publication[139] | N/A | N/A |
| Advanced prostate | Fruit juice in adolescent | Incidence | 1 publication[94] | N/A | 1 publication[94] | N/A |
| Advanced prostate | Citrus juice | Incidence | 1 publication[139] | 1 publication[139] | N/A | N/A |
| Advanced prostate | Fruit and vegetable juice | Incidence | 1 publication[139] | 1 publication[139] | N/A | N/A |
| Advanced prostate | Vegetable juice | Incidence | 1 publication[139] | 1 publication[139] | N/A | N/A |
| Advanced restricted | Citrus juice | Incidence | 1 publication[139] | 1 publication[139] | N/A | N/A |
| Advanced restricted | Fruit and vegetable juice | Incidence | 1 publication[139] | 1 publication[139] | N/A | N/A |
| Advanced restricted | Vegetable juice | Incidence | 1 publication[139] | 1 publication[139] | N/A | N/A |
| Non-advanced prostate | Fruit juice in adolescent | Incidence | 1 publication[94] | N/A | 1 publication[94] | N/A |
| Extraporastatic prostate | Fruit juice | Incidence | 1 publication[89] | N/A | 1 publication[89] | N/A |
| Aggressive prostate | Fruit juice | Incidence | 1 publication[89] | N/A | 1 publication[89] | NA |
| High grade prostate | Fruit juice | Incidence | 1 publication[161] | N/A | 1 publication[161] | N/A |
| High grade prostate | Citrus juice | Incidence | 1 publication[139] | 1 publication[139] | N/A | N/A |
| High grade prostate | Fruit and vegetable juice | Incidence | 1 publication[139] | 1 publication[139] | N/A | N/A |
| High grade prostate | Vegetable juice | Incidence | 1 publication[139] | 1 publication[139] | N/A | N/A |
| Low grade prostate | Citrus juice | Incidence | 1 publication[139] | 1 publication[139] | N/A | N/A |
| Low grade prostate | Fruit and vegetable juice | Incidence | 1 publication[139] | 1 publication[139] | N/A | N/A |
| Low grade prostate | Vegetable juice | Incidence | 1 publication[139] | 1 publication[139] | N/A | N/A |
| Localised prostate | Citrus juice | Incidence | 1 publication[139] | 1 publication[139] | N/A | N/A |
| Localised prostate | Fruit and vegetable juice | Incidence | 1 publication[139] | 1 publication[139] | N/A | N/A |
| Localised prostate | Vegetable juice | Incidence | 1 publication[139] | 1 publication[139] | N/A | N/A |
| High risk | Fruit juice | Incidence | 1 publication[47] | N/A | 1 publication[47] | N/A |
| Low risk | Fruit juice | Incidence | 1 publication[47] | N/A | 1 publication[47] | N/A |
| Symptomatic prostate | Fruit juice | Incidence | 1 publication[47] | N/A | 1 publication[47] | N/A |
| Kidney | Fruit juice | Incidence | 3 publications[36,98,142] | 3 publications[36,98,142] | N/A | N/A |
| Kidney | Fruit and vegetable juice | Incidence | 3 publications[26,66,218] | 2 publications[26,66] | N/A | 1 publication[218] |
| Kidney | Fruit juice | Mortality | 1 publication[76] | N/A | 1 publication[76] | N/A |
| Kidney | Fruit and vegetable juice | Mortality | 1 publication[66] | N/A | 1 publication[66] | N/A |
| Bladder | Fruit juice | Incidence | 1 publication[138] | N/A | 1 publication[138] | N/A |
| Bladder | Sweetened fruit juice | Incidence | 1 publication[123] | N/A | 1 publication[123] | N/A |
| Bladder | Apple juice | Incidence | 1 publication[120] | N/A | 1 publication[120] | N/A |
| Bladder | Citrus juice | Incidence | 1 publication[181] | N/A | 1 publication[181] | N/A |
| Bladder | Orange juice | Incidence | 1 publication[120] | N/A | 1 publication[120] | N/A |
| Bladder | Grapefruit juice | Incidence | 1 publication[120] | N/A | 1 publication[120] | N/A |
| Bladder | Other fruit juices (not citrus and apple) | Incidence | 1 publication[120] | N/A | 1 publication[120] | N/A |
| Urothelial | Fruit and vegetable juice | Incidence | 1 publication[147] | N/A | 1 publication[147] | N/A |
| High risk urothelial | Fruit and vegetable juice | Incidence | 1 publication[147] | N/A | 1 publication[147] | N/A |
| Low risk urothelial | Fruit and vegetable juice | Incidence | 1 publication[147] | N/A | 1 publication[147] | N/A |
| Bladder | Tomato | Incidence | 2 publications[120,175] | N/A | 2 publications[120,175] Insufficient data format | N/A |
| Bladder | Pure fruit juice | Mortality | 1 publication[76] | N/A | 1 publication[76] | N/A |
| Glioma | Fruit juice | Incidence | 1 publication[92] | 1 publication[92] | N/A | N/A |

| Cancer | Exposure | Outcome | Publications identified | Publications included in various meta-analyses | Publications included in the descriptive synthesis only | Superseded |
|---|---|---|---|---|---|---|
| Thyroid | Citrus juice | Incidence | 1 publication[174] | N/A | 1 publication[174] | N/A |
| Papillary thyroid | Citrus juice | Incidence | 1 publication[174] | N/A | 1 publication[174] | N/A |
| Differentiated thyroid | Fruit and vegetable juice | Incidence | 1 publication[179] | N/A | 1 publication[179] | N/A |
| Papillary thyroid | Fruit and vegetable juice | Incidence | 1 publication[179] | N/A | 1 publication[179] | N/A |
| Follicular thyroid | Fruit and vegetable juice | Incidence | 1 publication[179] | N/A | 1 publication[179] | N/A |
| Myeloproliferative neoplasms | Fruit juice | Incidence | 1 publication[141] | N/A | 1 publication[141] | N/A |
| Polycythaemia vera | Fruit juice | Incidence | 1 publication[141] | N/A | 1 publication[141] | N/A |
| Essential thrombocythemia | Fruit juice | Incidence | 1 publication[141] | N/A | 1 publication[141] | N/A |
| non-Hodgkin lymphoma | Apple juice | Incidence | 1 publication[162] | N/A | 1 publication[162] | N/A |
| Diffuse large B-cell lymphoma | Apple juice | Incidence | 1 publication[162] | N/A | 1 publication[162] | N/A |
| Follicular lymphoma | Apple juice | Incidence | 1 publication[162] | N/A | 1 publication[162] | N/A |

**Supplementary Table 10. List of studies included in the systematic review on soft drinks, and studies included and not included in the dose-response meta-analysis (cancers are ordered by ICD-10 codes).**

| Cancer | Exposure | Outcome | Publications identified | Publications included in various meta-analyses | Publications included in the descriptive synthesis only | Superseded |
|---|---|---|---|---|---|---|
| Pharynx | Carbonated soft drinks | Incidence | 1 publication[143] | N/A | 1 publication[143] | NA |
| Oral | Carbonated soft drinks | Incidence | 1 publication[143] | N/A | 1 publication[143] | NA |
| Oesophageal squamous cell | Carbonated soft drinks | Incidence | 1 publication[143] | N/A | 1 publication[143] | NA |
| Oesophageal adenocarcinoma | Carbonated soft drinks | Incidence | 1 publication[143] | N/A | 1 publication[143] | NA |
| Stomach cardia | Carbonated soft drinks | Incidence | 1 publication[143] | N/A | 1 publication[143] | NA |
| Stomach non-cardia | Carbonated soft drinks | Incidence | 1 publication[143] | N/A | 1 publication[143] | NA |
| Colorectal | Soft drinks | Incidence | 2 publications[35,136] | N/A | 2 publications[35,136] Insufficient data format | NA |
| Colorectal | Soft drinks | Mortality | 2 publications[85,126] | N/A | 2 publications[85,126] Insufficient data format | NA |
| Colorectal | Cola | Mortality | 1 publication[85] | N/A | 1 publication[85] | NA |
| Colon | Soft drinks | Incidence | 1 publication[136] | N/A | 1 publication[136] Insufficient data format | NA |
| Proximal colon | Soft drinks | Incidence | 1 publication[136] | N/A | 1 publication[136] Insufficient data format | NA |
| Distal colon | Soft drinks | Incidence | 1 publication[136] | N/A | 1 publication[136] Insufficient data format | NA |
| Rectal | Soft drinks | Incidence | 1 publication[136] | N/A | 1 publication[136] Insufficient data format | NA |
| Liver | Soft drinks | Incidence | 3 publications[79,159,183] | 3 publications[79,159,183] | N/A | NA |
| Liver | Carbonated soft drinks | Incidence | 1 publication[79] | 1 publication[79] | N/A | NA |
| Gallbladder | Soft drinks | Incidence | 2 publications[96,159] | 2 publications[96,159] | N/A | NA |
| Intrahepatic biliary tract | Soft drinks | Incidence | 2 publications[96,159] | 2 publications[96,159] | N/A | NA |
| Extrahepatic biliary tract | Soft drinks | Incidence | 2 publications[96,159] | 2 publications[96,159] | N/A | NA |
| Ampulla of Vater | Soft drinks | Incidence | 1 publication[159] | N/A | 1 publication[159] | NA |
| Pancreas | Soft drinks | Incidence | 5 publications[39,85,95,127,189] | 4 publications[39,85,95,127,189] | 1 publication[85] Insufficient data format | NA |
| Pancreatic adenocarcinoma | Carbonated soft drinks | Incidence | 1 publication[125] | N/A | 1 publication[125] | NA |
| Larynx | Carbonated soft drinks | Incidence | 1 publication[143] | N/A | 1 publication[143] | NA |
| Lung | Soft drinks | Incidence | 3 publications[65,67,177] | 2 publications[65,67] | 1 publication[177] Insufficient data format | NA |
| Lung | Soft drinks | Mortality | 1 publication[85] | N/A | 1 publication[85] | NA |
| Lung | Cola | Mortality | 1 publication[85] | N/A | 1 publication[85] | NA |
| Squamous cell | Soft drinks | Incidence | 1 publication[73] | N/A | 1 publication[73] | NA |
| Breast | Soft drinks | Incidence | 2 publications[49,68] | 2 publications[49,68] | NA | NA |
| Postmenopausal breast | Soft drinks | Incidence | 1 publication[49] | N/A | 1 publication[49] | NA |
| Premenopausal breast | Soft drinks | Incidence | 1 publication[49] | N/A | 1 publication[49] | NA |
| Breast | Soft drinks | Mortality | 1 publication[126] | N/A | 1 publication[126] | NA |

| Cancer | Exposure | Outcome | Publications identified | Publications included in various meta-analyses | Publications included in the descriptive synthesis only | Superseded |
|---|---|---|---|---|---|---|
| Endometrial | Soft drinks | Incidence | 3 publications[49,61,117] | 2 publications[49,117] | 1 publication[61]<br>Insufficient data format | NA |
| Ovarian | Soft drinks | Incidence | 1 publication[49] | N/A | 1 publication[49] | NA |
| Epithelial ovarian | Cola | Incidence | 1 publication[163] | N/A | 1 publication[163] | NA |
| Prostate | Soft drinks | Incidence | 1 publication[137] | NA | 1 publication[137] | NA |
| Prostate | Soft drinks | Mortality | 2 publications[126,137] | N/A | 1 publication[126] | 1 publication [137] |
| Prostate | Cola | Incidence/mortality | 1 publication[50] | N/A | 1 publication[50] | NA |
| Kidney | Soft drinks | Incidence | 2 publications[66,102] | N/A | 2 publications[66,102]<br>Insufficient data format | NA |
| Kidney | Carbonated soft drinks | Incidence | 2 publications[26,36] | 2 publications[26,36] | N/A | NA |
| Kidney | Soft drinks | Mortality | 2 publications[66,102] | N/A | 2 publications[66,102]<br>Insufficient data format | NA |
| Bladder | Soft drinks | Incidence | 2 publications[119,147] | 2 publications[119,147] | N/A | NA |
| Glioma | Carbonated soft drinks | Incidence | 1 publication[48] | N/A | 1 publication[48] | NA |
| Glioma | Carbonated and caffeinated soft drinks | Incidence | 2 publications[48,71] | 2 publications[48,71] | N/A | NA |
| Glioma | Carbonated and decaffeinated soft drinks | Incidence | 2 publications[48,71] | 2 publications[48,71] | N/A | NA |
| Thyroid | Soft drinks | Incidence | 1 publication[180] | N/A | 1 publication[180] | NA |
| Papillary thyroid | Soft drinks | Incidence | 1 publication[180] | N/A | 1 publication[180] | NA |
| Follicular thyroid | Soft drinks | Incidence | 1 publication[180] | N/A | 1 publication[180] | NA |
| Myeloproliferative neoplasms | Caffeinated soft drinks | Incidence | 1 publication[140] | NA | 1 publication[140] | NA |
| Polycythemia vera | Caffeinated soft drinks | Incidence | 1 publication[140] | NA | 1 publication[140] | NA |
| Essential thrombocythemia | Caffeinated soft drinks | Incidence | 1 publication[140] | NA | 1 publication[140] | NA |
| Myeloproliferative neoplasms | Decaffeinated soft drinks | Incidence | 1 publication[140] | NA | 1 publication[140] | NA |
| Polycythemia vera | Decaffeinated soft drinks | Incidence | 1 publication[140] | NA | 1 publication[140] | NA |
| Essential thrombocythemia | Decaffeinated soft drinks | Incidence | 1 publication[140] | NA | 1 publication[140] | NA |

#### Supplementary Table 11. Descriptive table of the included observational studies of sugar sweetened beverages and cancer incidence and mortality.

| Author, Year Country (PMID) | Study name, type Age, sex, ethnicity, characteristics of participants | Cases/ Study size Median follow-up (years) | Case ascertainment | Exposure Assessment, detail | Outcome Sub-population | Comparison | RR (95%CI) Ptrend | Adjustments | | | | | | | | | | | |
|---|---|---|---|---|---|---|---|---|---|---|---|---|---|---|---|---|---|---|---|
| | | | | | | | | Age | Sex | Smo | Alc | Obe | Phy | Diet | Med | Mens | Repr | Oth | Skin |
| **Mouth, pharynx, and larynx** | | | | | | | | | | | | | | | | | | | |
| Wu 2023 USA (37810918)[172] | Prostate, Lung, Colorectal and Ovarian Cancer Screening trial<br>Prospective Cohort<br>Range:55-74 years<br>M/F<br>Mixed | 279 / 101,755<br>8.84 years | Annual study update form, medical records, death certificates and family reports. | Questionnaire,<br>**SSB**:<br>Sugar-sweetened beverages | Incidence, Head and Neck cancer | ≥318.19 vs ≤31.95 g/day | 1.14 (0.82 - 1.58)<br>Ptrend:0.087 | ✓ | ✓ | ✓ | ✓ | ✓ | | ✓ E | D | | | ✓ | |
| McCullough 2022 USA (36107009)[113] | Cancer Prevention Study II<br>Prospective Cohort<br>M/F<br>Mainly white | 1,852 / 31,691<br>27.7 years | self-report & death certificates | Questionnaire,<br>**SSB**:<br>Regular colas and other nondiet soft drinks | Mortality, Larynx/oral cavity/pharynx | 2+ drinks/day vs never | 1.04 (0.87 - 1.24)<br>Ptrend:0.963 | ✓ | | ✓ | ✓ | | | ✓ * | | | | ✓ | |
| | | 1,852 / 31,691<br>27.7 years | | | | Per 1 drink day | 1.01 (0.97 - 1.06) | ✓ | | ✓ | ✓ | | | ✓ * | | | | ✓ | |
| Kjaerheim 1998 Norway (9486469)[90] | Norwegian Men UADT<br>Prospective Cohort<br>M | 63 / 10,900<br>25 years | population survey | FFQ,<br>Lemonade | Incidence, Oral cavity, larynx, pharynx, esophagus | 6+ times/month vs < monthly | 0.90 (0.50 - 1.60)<br>Ptrend:>0.5 | ✓ | | ✓ | ✓ | | | | | | | | |
| | | 65 / 10,900<br>25 years | | FFQ,<br>Red fruit drink | | 6+ times/month vs < monthly | 1.20 (0.60 - 2.20)<br>Ptrend:>0.5 | ✓ | | ✓ | ✓ | | | | | | | | |
| **Oesophageal** | | | | | | | | | | | | | | | | | | | |
| McCullough 2022 USA (36107009)[113] | Cancer Prevention Study II<br>Prospective Cohort<br>M/F<br>Mainly white | 2,727 / 934,777<br>27.7 years | self-report & death certificates | Questionnaire,<br>**SSB**:<br>Regular colas and other nondiet soft drinks. | Mortality, Oesophageal cancer | 2+ drinks/day vs never | 0.88 (0.76 - 1.03)<br>Ptrend:0.016 | ✓ | | ✓ | ✓ | | | ✓ * | | | | ✓ | |
| | | | | | | Per 1 drink day | 0.97 (0.92 - 1.01) | ✓ | | ✓ | ✓ | | | ✓ * | | | | ✓ | |
| **Stomach** | | | | | | | | | | | | | | | | | | | |
| Lee 2023 Korea (37536716)[97] | Health Examinees-Gem study<br>Prospective Cohort<br>Range:40-69 years<br>M/F | 498 / 36,266 | Korea Central Cancer Registry +Cancer Statistics in Korea | Semi-quantitative FFQ,<br>**SSB**:<br>Sugar-sweetened beverages | Incidence, Stomach cancer, male | 0 vs >250 g/day | 1.69 (0.83 - 3.46)[1] | | | ✓ | | | | ✓ E | | | | ✓ | |
| | | 402 / 68,130 | | | Incidence, Stomach cancer, female | 0 vs >250 g/day | 1.58 (0.59 - 4.22)[1] | | | ✓ | | | | ✓ E | | | | ✓ | |
| Khairan 2023 Japan (36167978)[84] | The Japan Public Health Center-based Prospective Study<br>Prospective Cohort<br>Range:40-59 years<br>M/F<br>Japanese | 676 / 39,353<br>16.7 years | medical and population cancer registries, death certificates | Questionnaire & FFQ,<br>**SSB**:<br>Beverages containing caloric sweeteners, which included β-carotene- and calcium fortified beverages, canned coffee, carbonated beverages, 100% fruit juice (apple and orange juice), lactic acid bacteria beverages, and vitamin fortified beverages | Incidence, Gastric cancer, females | >278.1 vs 0 - 0.04 ml/day | 1.03 (0.79 - 1.33)<br>Ptrend:0.47 | ✓ | | ✓ | ✓ | ✓ | ✓ | ✓ E | | | | ✓ | |

| Author, Year Country (PMID) | Study name, type Age, sex, ethnicity, characteristics of participants | Cases/ Study size Median follow-up (years) | Case ascertainment | Exposure Assessment, detail | Outcome Sub-population | Comparison | RR (95%CI) Ptrend | Adjustments Age | Sex | Smo | Alc | Obe | Phy | Diet | Med | Mens | Repr | Oth | Skin |
|---|---|---|---|---|---|---|---|---|---|---|---|---|---|---|---|---|---|---|---|
| | | 1,465 / 35,102<br>16.7 years | | | Incidence, Gastric cancer, males | >398.6 vs 0 - 30.5 ml/day | 0.98 (0.82 - 1.17)<br>Ptrend:0.48 | ✓ | | ✓ | ✓ | ✓ | ✓ | ✓<br>E | | | | ✓ | |
| McCullough 2022 USA (36107009)[113] | Cancer Prevention Study II<br>Prospective Cohort<br>M/F<br>Mainly white | 2,798 / 934,777<br>27.7 years | self-report & death certificates | Questionnaire,<br>**SSB**:<br>Regular colas and other nondiet soft drinks. | Mortality, Stomach cancer | 2+ drinks/day vs never | 1.08 (0.94 - 1.25)<br>Ptrend:0.128 | ✓ | | ✓ | ✓ | | | ✓<br>* | | | | ✓ | |
| | | | | | | Per 1 drink day | 1.03 (0.99 - 1.08) | ✓ | | ✓ | ✓ | | | ✓<br>* | | | | ✓ | |
| Khairan 2023 Japan (36167978)[84] | The Japan Public Health Center-based Prospective Study<br>Prospective Cohort<br>Range:40-59 years<br>M/F<br>Japanese | 26 / 39,353<br>16.7 years | medical and population cancer registries, death certificates | Questionnaire & FFQ,<br>**SSB**:<br>Beverages containing caloric sweeteners, which included β-carotene- and calcium fortified beverages, canned coffee, carbonated beverages, 100% fruit juice (apple and orange juice), lactic acid bacteria beverages, and vitamin fortified beverages | Incidence, Cardia cancer, females | >278.1 vs 0 - 0.04 ml/day | 1.28 (0.32 - 5.12)<br>Ptrend:0.53 | ✓ | | ✓ | ✓ | ✓ | ✓ | ✓<br>E | | | | ✓ | |
| | | 99 / 35,102<br>16.7 years | | | Incidence, Cardia cancer, males | >398.6 vs 0 - 30.5 ml/day | 0.48 (0.23 - 0.99)<br>Ptrend:0.03 | ✓ | | ✓ | ✓ | ✓ | ✓ | ✓<br>E | | | | ✓ | |
| Hodge 2018 Australia (29463332)[70] | Melbourne Collaborative Cohort Study<br>Prospective Cohort<br>M/F<br>Excluded participants with diabetes at baseline | 165 / 35,593<br>23 years | cancer registry/death records/National Death Index | Self-administered Questionnaire,<br>**SSB**:<br>Sugar sweetened soft drinks | Incidence, Gastric cardia adenocarcino ma | ≥1/day vs <1/month | 1.17 (0.73 - 1.89)<br>Ptrend:0.37 | ✓ | ✓ | ✓ | ✓ | | ✓ | ✓ | | | | ✓ | |
| | | | | | | Linear model | 1.24 (0.77 - 1.99) | ✓ | ✓ | ✓ | ✓ | | ✓ | ✓ | | | | ✓ | |
| Acuna 2023 USA (36789943)[34] | The Multiethnic cohort study<br>Prospective Cohort<br>M/F<br>Multi-ethnic | / 176,752<br>19.2 years | cancer registries | Quantitative FFQ,<br>**SSB**:<br>Sugar sweetened beverages and fruit juice | Incidence, Gastric distal adenocarcino ma | Per 1 point increase in AHEI-2010 component score | 0.99 (0.97 - 1.01) | ✓ | ✓ | ✓ | | | | ✓<br>E | | | | ✓ | |
| | | | | Quantitative FFQ,<br>**SSB**:<br>Sugar-sweetened beverages | Incidence, Gastric distal adenocarcino ma | Per 1 point increase in DASH score component score | 0.96 (0.91 - 1.01) | ✓ | ✓ | ✓ | | | | ✓<br>E | | | | ✓ | |
| Khairan 2023 Japan (36167978)[84] | The Japan Public Health Center-based Prospective Study<br>Prospective Cohort<br>Range:40-59 years<br>M/F<br>Japanese | 650 / 39,353<br>16.7 years | medical and population cancer registries, death certificates | Questionnaire & FFQ,<br>**SSB**:<br>Beverages containing caloric sweeteners, which included β-carotene- and calcium fortified beverages, canned coffee, carbonated beverages, 100% fruit juice (apple and orange juice), lactic acid bacteria beverages, and vitamin fortified beverages | Incidence, non-Cardia cancer, females | >278.1 vs 0 - 0.04 ml/day | 1.01 (0.78 - 1.32)<br>Ptrend:0.56 | ✓ | | ✓ | ✓ | ✓ | ✓ | ✓<br>E | | | | ✓ | |

| Author, Year Country (PMID) | Study name, type Age, sex, ethnicity, characteristics of participants | Cases/ Study size Median follow-up (years) | Case ascertainment | Exposure Assessment, detail | Outcome Sub-population | Comparison | RR (95%CI) Ptrend | Adjustments Age | Sex | Smo | Alc | Obe | Phy | Diet | Med | Mens | Repr | Oth | Skin |
|---|---|---|---|---|---|---|---|---|---|---|---|---|---|---|---|---|---|---|---|
| | | 1,366 / 35,102 16.7 years | | | Incidence, non-Cardia cancer, males | >398.6 vs 0 - 30.5 ml/day | 1.03 (0.86 - 1.24) Ptrend:0.88 | ✓ | | ✓ | ✓ | ✓ | ✓ | ✓ E | | | | ✓ | |
| **Small intestine** | | | | | | | | | | | | | | | | | | | |
| McCullough 2022 USA (36107009)[113] | Cancer Prevention Study II Prospective Cohort M/F Mainly white | 324 / 934,777 27.7 years | self-report & death certificates | Questionnaire, **SSB**: Regular colas and other nondiet soft drinks. | Mortality, Small intestinal cancer | 2+ drinks/day vs never | 1.26 (0.84 - 1.90) Ptrend:0.609 | ✓ | | ✓ | ✓ | | | ✓ * | | | | ✓ | |
| | | | | | | Per 1 drink day | 1.01 (0.89 - 1.14) | ✓ | | ✓ | ✓ | | | ✓ * | | | | ✓ | |
| **Colorectal** | | | | | | | | | | | | | | | | | | | |
| Kanehara 2024 USA (38795743)[81] | Multiethnic Cohort Prospective Cohort Range:45-75 years M/F Mixed | 4,403 / 192,651 19.3 years | cancer registries | Quantitative FFQ, **SSB**: Chocolate milk, cocoa or Ovaltine; milkshakes or malts; regular sodas; fruit drink (not 100% fruit juice); cappuccino, regular coffee, decaffeinated coffee, and black tea (if sugar or honey were added to them) | Incidence, Colorectal cancer | Median 351.2 vs 0 g/1000 kcal | 1.03 (0.93 - 1.15) Ptrend:0.29 | ✓ | ✓ | ✓ | ✓ | ✓ | ✓ | ✓ E | ✓ D | | ✓ | ✓ | |
| Lee 2023 Korea (37536716)[97] | Health Examinees-Gem (HEXA-G) study Prospective Cohort Range:40-69 years M/F | 397 / 36,266 | Korea Central Cancer Registry +Cancer Statistics in Korea | Semi-quantitative FFQ, **SSB**: Sugar-sweetened beverages | Incidence, Colorectal cancer, male | 0 vs >250 g/day | 1.05 (0.46 - 2.40)[1] | | | ✓ | | | | ✓ E | | | | ✓ | |
| | | 388 / 68,130 | | | Incidence, Colorectal cancer, female | 0 vs >250 g/day | 3.39 (0.76 - 15.25)[1] | | | ✓ | | | | ✓ E | | | | ✓ | |
| Wang 2022 USA (38752573)[168] | Nurses' Health Study, Nurses' Health Study II, Health Professionals Follow-up Study Pooled Analysis Cohorts M/F Mainly white | 1,294 / 46,341 28 years | self-report, blinded physicians reviewed medical records & pathology report, next of kin, national death index. | Semi-quantitative FFQ, **SSB**: Sugar- or artificially sweetened beverages (Caffeine-free Coke; Coke/Pepsi/Cola; dairy coffee drink; Hawaiian punch; Low-calorie soda, caffeine-free; low-calorie soda, Pepsi, 7-up; other carbon beverage; other low-calorie Cola with caffeine) | Incidence, colorectal cancer, male | Per 1 serving day | 1.28 (1.05 - 1.55) | ✓ | | ✓ | ✓ | | ✓ | ✓ E | ✓ | | | ✓ | |
| | | 1,922 / 159,907 28 years | | | Incidence, Colorectal cancer, female | Per 1 serving day | 1.09 (0.94 - 1.25) | ✓ | | ✓ | ✓ | | ✓ | ✓ E | ✓ | ✓ | ✓ | ✓ | |
| Arthur 2021 Canada (33220638)[38] | Canadian Study of Diet, Lifestyle and Health Case Cohort F | 247 / 2,633 15 years | Cancer registry and national death index | Self-administered Questionnaire, **SSB**: Coca Cola, Pepsi, other cola, soft drinks (not dietetic), orange/grapefruit juice, apple/grape juice, other fruit juices, fruit drink/lemonade and power-based drinks/iced tea | Incidence, Colorectal cancer | >9.5 vs ≤3.2 cups/week | 1.27 (0.89 - 1.82) Ptrend:0.175 | ✓ | | ✓ | ✓ | ✓ | ✓ | ✓ | | | | ✓ | |
| | | | | Self-administered Questionnaire, **SSB**: sugar sweetened beverages; without fruit juices | | >3.3 cups/week vs none | 1.12 (0.79 - 1.60) Ptrend:0.256 | ✓ | | ✓ | ✓ | ✓ | ✓ | ✓ | | | | ✓ | |
| Leung 2021 Japan(33563648)[101] | The Japan Public Health Center-based Prospective Study Prospective Cohort | 940 / 33,709 15 years | Cancer registry and death certificates | | Incidence, Colorectal cancer, | Per 100 ml day | 0.99 (0.97 - 1.01) | ✓ | | ✓ | ✓ | ✓ | ✓ | ✓ E | ✓ D | | | | |

| Author, Year Country (PMID) | Study name, type Age, sex, ethnicity, characteristics of participants | Cases/ Study size Median follow-up (years) | Case ascertainment | Exposure Assessment, detail | Outcome Sub-population | Comparison | RR (95%CI) Ptrend | Adjustments Age | Sex | Smo | Alc | Obe | Phy | Diet | Med | Mens | Repr | Oth | Skin |
|---|---|---|---|---|---|---|---|---|---|---|---|---|---|---|---|---|---|---|---|
| | Age:56.5 years<br>M/F<br>Japanese | | | Self-administered Questionnaire,<br>**SSB**:<br>Beta-carotene fortified beverages, calcium fortified beverages, canned coffee, carbonated beverages, 100% fruit juices, lactic acid bacteria beverages and vitamin fortified beverages | male | | | | | | | | | | | | | | |
| | | 708 / 40,361<br>15 years | | | Incidence, Colorectal cancer, female | Per 100 ml day | 1.00 (0.96 - 1.05) | ✓ | | ✓ | ✓ | ✓ | ✓ | ✓ E | ✓ D | | | | |
| Barrubés 2020 Spain (32340309)[40] | PREDIMED<br>Prospective Cohort<br>Age:67 years<br>M/F<br>High risk of cardiovascular disease | 97 / 7,216<br>6 years | Medical records + national death index | Semi-quantitative FFQ,<br>**SSB**:<br>Sugar-sweetened beverages | Incidence, Colorectal cancer | 0 vs ≥ 250 g/day | 0.42 (0.19 - 0.93)[1]<br>Ptrend:0.192 | ✓ | ✓ | ✓ | ✓ | ✓ | ✓ | ✓ E | D | | | ✓ | |
| Chazelas 2019 France (31292122)[44] | NutriNet-Santé study<br>Prospective Cohort<br>Age:42.2 years<br>M/F | 166 / 101,257<br>5.1 years | self-report, medical records reviewed by physicians, health insurance database, mortality registry | 24 Hour Diet Recall,<br>**SSB**:<br>Beverages with more than 5% simple carbohydrates and 100% fruit juices (no added sugar), soft drinks (carbonated or not), syrups, 100% juice, fruit drinks, sugar sweetened hot beverages, milk-based sugar sweetened beverages, sport drinks, and energy drinks | Incidence, Colorectal cancer | Per 100 ml day | 1.12 (0.84 - 1.50) | ✓ | ✓ | ✓ | ✓ | ✓ | ✓ | ✓ E | D | | | ✓ | |
| | | | | 24 Hour Diet Recall,<br>**SSB**:<br>Sugary drinks except 100% fruit juices | | Per 100 ml day | 1.13 (0.74 - 1.72) | ✓ | ✓ | ✓ | ✓ | ✓ | ✓ | ✓ E | D | | | ✓ | |
| Pacheco 2019 USA (31596902)[135] | California Teachers Study, 1995<br>Prospective Cohort<br>Age:52 years<br>F<br>Mostly white teachers | 1,318 / 99,798<br>20.1 years | Cancer registry and national death index | Self-administered Questionnaire,<br>**SSB**:<br>sweetened carbonated and noncarbonated beverages (caloric soft drinks, sweetened bottled waters and/or teas, and fruit drinks) | Incidence, Colorectal cancer | ≥1 serving/day vs rare or never | 1.15 (0.87 - 1.52)<br>Ptrend:0.259 | ✓ | | ✓ | ✓ | | | ✓ | ✓ | ✓ | ✓ | ✓ | |
| | | | | **Sweetened bottled water or tea** | Incidence, Colorectal cancer | ≥1 serving/day vs rare or never | 1.24 (0.94 - 1.62)<br>Ptrend:0.287 | ✓ | | ✓ | ✓ | | | ✓ | ✓ | ✓ | ✓ | ✓ | |
| | | | | **Fruit drinks** | Incidence, Colorectal cancer | ≥1 serving/day vs rare or never | 0.44 (0.11 - 1.77)<br>Ptrend:0.290 | ✓ | | ✓ | ✓ | | | ✓ | ✓ | ✓ | ✓ | ✓ | |
| | | | | **SSBs without fruit drinks** | Incidence, Colorectal cancer | ≥1 serving/day vs rare or never | 1.03 (0.70 - 1.51)<br>Ptrend:0.730 | ✓ | | ✓ | ✓ | | | ✓ | ✓ | ✓ | ✓ | ✓ | |
| Hodge 2018 Australia (29463332)[70] | Melbourne Collaborative Cohort Study<br>Prospective Cohort<br>M/F | 1,055 / 35,593<br>23 years | cancer registry/death | Self-administered Questionnaire,<br>**SSB**:<br>Sugar sweetened soft drinks | Incidence, Colorectal cancer | ≥1/day vs <1/month | 1.28 (1.04-1.57) | ✓ | ✓ | ✓ | ✓ | | ✓ | ✓ E | | | | ✓ | |

| Author, Year Country (PMID) | Study name, type Age, sex, ethnicity, characteristics of participants | Cases/ Study size Median follow-up (years) | Case ascertainment | Exposure Assessment, detail | Outcome Sub-population | Comparison | RR (95%CI) Ptrend | Age | Sex | Smo | Alc | Obe | Phy | Diet | Med | Mens | Repr | Oth | Skin |
|---|---|---|---|---|---|---|---|---|---|---|---|---|---|---|---|---|---|---|---|
| | | | records/National Death Index | | | | | | | | | | | | | | | | |
| | | | | | | Linear model | 1.17 (0.96 - 1.43) | | | | | | | | | | | | |
| Makarem 2018 USA (29674390)[108] | Framingham Offspring Cohort<br>Prospective Cohort<br>Age:55.4 years<br>M/F | 68 / 3,184<br>~10 years | medical examinations | Semi-quantitative FFQ,<br>**SSB**:<br>Sugar sweetened beverages and fruit juice. | Incidence, Colorectal cancer | >9.4 vs <4 serving/week | 1.39 (0.68 - 2.82) | ✓ | ✓ | ✓ | ✓ | ✓ | ✓ | ✓ E | D | | | ✓ | |
| | | | | Semi-quantitative FFQ,<br>**SSB**:<br>Regular sugar-sweetened soda and fruit drinks (lemonade and punch) | | >3.5 vs >0.5 serving/week | 0.96 (0.51 - 1.82) | ✓ | ✓ | ✓ | ✓ | ✓ | ✓ | ✓ E | D | | | ✓ | |
| Miller 2013 USA (23864539)[122] | NIH- AARP Diet and Health Study<br>Prospective Cohort<br>Range:50-70 years<br>M/F<br>White non-Hispanic | 1,749 /<br>293,248<br>11 years | Cancer and mortality registries | Semi-quantitative FFQ,<br>**SSB**:<br>Sugar-sweetened beverages | Incidence, Colorectal cancer, male | Quintile 5 vs Quintile 1 | 0.94 (0.85 - 1.03) | ✓ | | ✓ | ✓ | ✓ | ✓ | ✓ E | | | ✓ | ✓ | |
| | | 1,016 /<br>198,593<br>11 years | | | Incidence, Colorectal cancer, female | Quintile 5 vs Quintile 1 | 1.09 (0.93 - 1.26) | ✓ | | ✓ | ✓ | ✓ | ✓ | ✓ E | | | ✓ | ✓ | |
| Nomura 2016 USA (27220873)[129] | Black Women's Health Study, 1995<br>Prospective Cohort<br>Range:21-69 years<br>F<br>Black | 354 /<br>49,103<br>15.1 years | self-report, linkage to cancer registries, medical and pathology records | FFQ,<br>**SSB**:<br>Sugary drinks | Incidence, Colorectal cancer, baseline data | 0 vs ≥250 g/day | 1.29 (0.72 - 2.30)[1]<br>Ptrend:0.57 | ✓ | | ✓ | ✓ | ✓ | ✓ | ✓ E | ✓ D | ✓ | | ✓ | |
| | | | | | Time-varying data | | 1.24 (0.71-2.17)[1]<br>Ptrend:0.88 | ✓ | | ✓ | ✓ | ✓ | ✓ | ✓ E | ✓ D | ✓ | | ✓ | |
| Yuan 2022 USA (35470384)[178] | Nurses' Health Study + Health Professionals Follow-up Study<br>Prospective Cohort<br>M/F<br>White<br>Health professionals | 3,018 /<br>121,111 | self-report & biennial questionnaire & active follow-up for participant deaths | Validated FFQ,<br>**SSB**:<br>Sugar-sweetened carbonated (caffeinated colas, caffeine-free colas, other carbonated noncola), and noncarbonated (fruit punches, lemonades, or other fruit drinks) beverages | Incidence, Colorectal cancer | >1/day vs never | 1.04 (0.85 - 1.27) | ✓ | ✓ | ✓ | ✓ | ✓ | ✓ | ✓ E | ✓ D | ✓ | ✓ | ✓ | |
| | | | | | | Per 1 serving day | 1.03 (0.94 - 1.12) | ✓ | ✓ | ✓ | ✓ | ✓ | ✓ | ✓ E | ✓ D | ✓ | ✓ | ✓ | |
| Jin 2023 UK (38004195)[78] | UK Biobank<br>Prospective Cohort<br>Age:55.87 years<br>M/F<br>Mainly white | 1,466 /<br>118,210<br>12.8 years | hospital records, cancer registry, death registry | 24 Hour Diet Recall,<br>**SSB**:<br>Sugar-sweetened beverages & other sugary drinks | Incidence, Colorectal cancer | Per 1 SD day | 1.00 (0.95 - 1.05) | ✓ | ✓ | ✓ | | ✓ | ✓ | ✓ E | ✓ D | | | ✓ | |
| Yuan 2022 USA (35470384)[178] | Nurses' Health Study + Health Professionals Follow-up Study<br>Prospective Cohort<br>M/F<br>White<br>Health professionals | 1,038 /<br>121,111 | self-report & biennial questionnaire & active follow-up for participant deaths | Validated FFQ,<br>**SSB**:<br>Sugar-sweetened carbonated (caffeinated colas, caffeine-free colas, other carbonated noncola), and noncarbonated (fruit punches, | Mortality, Colorectal cancer | >1/day vs never | 1.18 (0.84 - 1.65) | ✓ | ✓ | ✓ | ✓ | ✓ | ✓ | ✓ E | ✓ D | ✓ | ✓ | ✓ | |

| Author, Year Country (PMID) | Study name, type Age, sex, ethnicity, characteristics of participants | Cases/ Study size Median follow-up (years) | Case ascertainment | Exposure Assessment, detail | Outcome Sub-population | Comparison | RR (95%CI) Ptrend | Adjustments Age | Sex | Smo | Alc | Obe | Phy | Diet | Med | Mens | Repr | Oth | Skin |
|---|---|---|---|---|---|---|---|---|---|---|---|---|---|---|---|---|---|---|---|
| | | | | lemonades, or other fruit drinks) beverages. | | Per 1 serving day | 1.12 (0.97 - 1.30) | ✓ | ✓ | ✓ | ✓ | ✓ | ✓ | ✓ E | ✓ D | ✓ | ✓ | ✓ | |
| McCullough 2022 USA (36107009)[113] | Cancer Prevention Study II Prospective Cohort M/F Mainly white | 13,752 / 934,777 27.7 years | self-report & death certificates | Questionnaire, **SSB**: Regular colas and other nondiet soft drinks | Mortality, Colorectal cancer | 2+ drinks/day vs never | 1.09 (1.02 - 1.17) Ptrend:0.011 | ✓ | | ✓ | ✓ | | | ✓ * | | | | ✓ | |
| | | | | | | Per 1 drink day | 1.02 (1.00 - 1.04) | ✓ | | ✓ | ✓ | | | ✓ * | | | | ✓ | |
| Mullee 2019 Denmark,France,Germany,Greece,Italy,Netherlands,Norway,Spain,Sweden,UK (31479109)[126] | European Prospective Investigation into Cancer and nutrition Prospective Cohort Age:50.8 years M/F | 2,095 / 451,743 16.4 years | active follow up, cancer registry, death certificate and National death index | Sema-quantitative FFQ, **SSB**: Sugar sweetened soft drinks | Mortality, Colorectal cancer | ≥1 vs <1 glasses/month | 1.10 (0.86 - 1.40) Ptrend:0.41 | ✓ | ✓ | ✓ | ✓ | ✓ | ✓ | ✓ E * | | ✓ | ✓ | ✓ | |
| Hur 2021 USA (33958435)[72] | Nurses' Health Study II Prospective Cohort F nurses | 109 / 95,464 24 years | Medical records and National Death Index | Semi-quantitative FFQ, **SSB**: Sugar-sweetened carbonated or noncarbonated beverages (soft drinks, fruit drinks, sports drinks, and sweetened tea beverages) | Incidence, Early-onset colorectal cancer, adulthood intake | ≥2 servings/d vs <1 serving/week | 1.97 (0.96 - 4.03) Ptrend:0.03 | | | ✓ | ✓ | ✓ | ✓ | ✓ * | ✓ | ✓ | ✓ | ✓ | |
| | | | | | | Per 1 serving day | 1.15 (0.97 - 1.35) | | | ✓ | ✓ | ✓ | ✓ | ✓ * | ✓ | ✓ | ✓ | ✓ | |
| | | 35 / 41,272 ~51 years | | | Incidence, Early-onset colorectal cancer, adolescence intake | ≥2 servings/d vs <1 serving/week | 3.41 (1.08 - 10.80) Ptrend:0.01 | ✓ | | ✓ | ✓ | ✓ | ✓ | ✓ E | | | | ✓ | |
| | | | | | | Per 1 serving day | 1.32 (1.00 - 1.75) | ✓ | | ✓ | ✓ | ✓ | ✓ | ✓ E | | | | ✓ | |
| | | 109 / 95,464 24 years | | Substitution of SSB with ASB | Incidence, Early-onset colorectal cancer, adulthood intake | Per 1 serving day | 0.83 (0.69 – 0.99) | ✓ | | ✓ | ✓ | ✓ | ✓ | ✓ E | | | | ✓ | |
| | | 109 / 95,464 24 years | | Substitution of SSB with fruit juice | | Per 1 serving day | 1.02 (0.61 – 1.69) | ✓ | | ✓ | ✓ | ✓ | ✓ | ✓ E | | | | ✓ | |
| Nomura 2016 USA (27220873)[129] | Black Women's Health Study, 1995 Prospective Cohort Range:21-69 years F Black | 277 / 49,103 15.1 years | self-report, linkage to cancer registries, medical and pathology records | FFQ, **SSB**: Sugary drinks | Incidence, Colon cancer, baseline data | 0 vs ≥250 g/day | 1.20 (0.62 - 2.31)[1] Ptrend:0.40 | ✓ | | ✓ | ✓ | ✓ | ✓ | ✓ E | ✓ D | ✓ | | ✓ | |
| | | | | | Time-varying data | 0 vs ≥250 g/day | 1.16 (0.61 – 2.21)[1] | ✓ | | ✓ | ✓ | ✓ | ✓ | ✓ E | ✓ D | ✓ | | ✓ | |
| Arthur 2021 Canada (33220638)[38] | Canadian Study of Diet, Lifestyle and Health Case Cohort F | 192 / 2,633 15 years | Cancer registry and national death index | Self-administered Questionnaire, **SSB**: Coca Cola, Pepsi, other cola, soft | Incidence, Colon cancer | >9.5 vs ≤3.2 cups/week | 1.41 (0.95 - 2.27) Ptrend:0.074 | ✓ | | ✓ | ✓ | ✓ | ✓ | ✓ | | | | ✓ | |

| Author, Year Country (PMID) | Study name, type Age, sex, ethnicity, characteristics of participants | Cases/ Study size Median follow-up (years) | Case ascertainment | Exposure Assessment, detail | Outcome Sub-population | Comparison | RR (95%CI) Ptrend | Adjustments Age | Sex | Smo | Alc | Obe | Phy | Diet | Med | Mens | Repr | Oth | Skin |
|---|---|---|---|---|---|---|---|---|---|---|---|---|---|---|---|---|---|---|---|
| | | | | drinks (not dietetic), orange/grapefruit juice, apple/grape juice, other fruit juices, fruit drink/lemonade and power-based drinks/iced tea | | | | | | | | | | | | | | | |
| Leung 2021 Japan (33563648)[101] | The Japan Public Health Center-based Prospective Study<br>Prospective Cohort<br>Age:56.5 years<br>M/F<br>Japanese | 606 / 33,709<br>15 years | Cancer registry and death certificates | Self-administered Questionnaire,<br>**SSB**:<br>Beta-carotene fortified beverages, calcium fortified beverages, canned coffee, carbonated beverages, 100% fruit juices, lactic acid bacteria beverages and vitamin fortified beverages | Incidence, Colon cancer, male | ≥254 ml/day vs non consumers | 0.80 (0.63 - 1.02)<br>Ptrend:0.11 | ✓ | | ✓ | ✓ | ✓ | ✓ | ✓ E | ✓ D | | | | |
| | | | | | | Per 100 ml day | 0.99 (0.96 - 1.02) | ✓ | | ✓ | ✓ | ✓ | ✓ | ✓ E | ✓ D | | | | |
| | | 515 / 40,361<br>15 years | Cancer registry and death certificates | Self-administered Questionnaire,<br>**SSB**:<br>Beta-carotene fortified beverages, calcium fortified beverages, canned coffee, carbonated beverages, 100% fruit juices, lactic acid bacteria beverages and vitamin fortified beverages | Incidence, Colon cancer, female | ≥254 ml/day vs non consumers | 1.36 (1.03 - 1.78)<br>Ptrend:0.04 | ✓ | | ✓ | ✓ | ✓ | ✓ | ✓ E | ✓ D | | | | |
| | | | | | | Per 100 ml day | 0.99 (0.93 - 1.04) | ✓ | | ✓ | ✓ | ✓ | ✓ | ✓ E | ✓ D | | | | |
| Kanehara 2024 USA (38795743)[81] | Multiethnic Cohort<br>Prospective Cohort<br>Range:45-75 years<br>M/F<br>Mixed | 3,351 / 192,651<br>19.3 years | cancer registries | Quantitative FFQ,<br>**SSB**:<br>Chocolate milk, cocoa or Ovaltine; milkshakes or malts; regular sodas; fruit drink (not 100% fruit juice); cappuccino, regular coffee, decaffeinated coffee, and black tea (if sugar or honey were added to them) | Incidence, Colon cancer | Median 351.2 vs 0 g/1000 kcal | 1.05 (0.93 - 1.18)<br>Ptrend:0.23 | ✓ | ✓ | ✓ | ✓ | ✓ | ✓ | ✓ E | ✓ D | | ✓ | ✓ | |
| Jin 2023 UK (38004195)[78] | UK Biobank<br>Prospective Cohort<br>Age:55.87 years<br>M/F<br>Mainly white | 842 / 118,210<br>12.8 years | hospital records, cancer registry, death registry | 24 Hour Diet Recall,<br>**SSB**:<br>Sugar-sweetened beverages & other sugary drinks. | Incidence, Colon cancer | Per 1 SD day | 1.01 (0.94 - 1.08) | ✓ | ✓ | ✓ | | ✓ | ✓ | ✓ E | ✓ D | | | ✓ | |
| Malik 2019 USA (30882235 2)[109] | Nurses' Health Study + Health Professionals Follow-up Study<br>Pooled Analysis Cohorts<br>M/F<br>Mostly white<br>Health professionals | NR / 118,363<br>34 years | Physician confirmation | FFQ,<br>**SSB**:<br>Sugar-sweetened caffeinated and noncaffeinated colas, other (noncola) sugar-sweetened carbonated beverages, and noncarbonated sugar-sweetened beverages (fruit punches, lemonades, or other fruit drinks) | Mortality, Colon cancer | Consumption ≥2/day vs <1/month | 1.38 (0.99 - 1.92)<br>Ptrend:0.07 | ✓ | | ✓ | ✓ | ✓ | ✓ | ✓ E * | D | | ✓ | ✓ | |
| | | | | | | Per 1 serving day | 1.09 (0.99 - 1.19) | ✓ | | ✓ | ✓ | ✓ | ✓ | ✓ E * | D | | ✓ | ✓ | |
| Pacheco 2019 USA (31596902)[135] | California Teachers Study, 1995<br>Prospective Cohort<br>Age:52 years<br>F | 718 / 99,798<br>20.1 years | Cancer registry and national death index | Self-administered Questionnaire,<br>**SSB**:<br>sweetened carbonated and | Incidence, Proximal colon cancer | ≥1 serving/day vs Rare or never | 1.08 (0.72 - 1.61)<br>Ptrend:0.998 | ✓ | | ✓ | ✓ | | | ✓ | ✓ | ✓ | ✓ | ✓ | |

| Author, Year Country (PMID) | Study name, type Age, sex, ethnicity, characteristics of participants | Cases/ Study size Median follow-up (years) | Case ascertainment | Exposure Assessment, detail | Outcome Sub-population | Comparison | RR (95%CI) Ptrend | Adjustments Age | Sex | Smo | Alc | Obe | Phy | Diet | Med | Mens | Repr | Oth | Skin |
|---|---|---|---|---|---|---|---|---|---|---|---|---|---|---|---|---|---|---|---|
| | Mostly white teachers | | | noncarbonated beverages (caloric soft drinks, sweetened bottled waters and/or teas, and fruit drinks) | | | | | | | | | | | | | | | |
| Yuan 2022 USA (35470384)[178] | Nurses' Health Study and Health Professionals Follow-up Study Prospective Cohort F/M White Health professionals | 470 / 46,384 28 years | self-report & biennial questionnaire & active follow-up for participant deaths | Validated FFQ, **SSB**: Sugar-sweetened carbonated (caffeinated colas, caffeine-free colas, other carbonated noncola), and noncarbonated (fruit punches, lemonades, or other fruit drinks) beverages. | Incidence, Proximal colon cancer, male | Per 1 serving day | 1.29 (1.05 - 1.48) | ✓ | | ✓ | ✓ | ✓ | ✓ | ✓ E | ✓ D | ✓ | ✓ | ✓ | |
| | | 805 / 74,727 30 years | | | Incidence, Proximal colon cancer, female | Per 1 serving day | 1.07 (0.89 - 1.29) | ✓ | | ✓ | ✓ | ✓ | ✓ | ✓ E | ✓ D | ✓ | ✓ | ✓ | |
| | | 168 | | | <60 years old at diagnosis | Per 1 serving day | 1.22 (0.93, 1.61) | ✓ | | ✓ | ✓ | ✓ | ✓ | ✓ E | ✓ D | ✓ | ✓ | ✓ | |
| | | 402 | | | 60 to 60 years old at diagnosis | Per 1 serving day | 1.27 (1.03, 1.57) | ✓ | | ✓ | ✓ | ✓ | ✓ | ✓ E | ✓ D | ✓ | ✓ | ✓ | |
| | | 705 | | | ≥70 years old at diagnosis | Per 1 serving day | 1.09 (0.87, 1.36) | ✓ | | ✓ | ✓ | ✓ | ✓ | ✓ E | ✓ D | ✓ | ✓ | ✓ | |
| | | 597 | | | BMI<25 kg/m$^2$ | Per 1 serving day | 1.11 (0.90 - 1.37) | ✓ | ✓ | ✓ | ✓ | | ✓ | ✓ E | ✓ D | ✓ | ✓ | ✓ | |
| | | 675 | | | BMI≥25 kg/m$^2$ | Per 1 serving day | 1.25 (1.05 - 1.48) | ✓ | ✓ | ✓ | ✓ | | ✓ | ✓ E | ✓ D | ✓ | ✓ | ✓ | |
| | | 750 | | | Ever smokers | Per 1 serving day | 1.09 (0.90 - 1.30) | ✓ | ✓ | | ✓ | ✓ | ✓ | ✓ E | ✓ D | ✓ | ✓ | ✓ | |
| | | 495 | | | Never smoker | Per 1 serving day | 1.28 (1.04 - 1.59) | ✓ | ✓ | | ✓ | ✓ | ✓ | ✓ E | ✓ D | ✓ | ✓ | ✓ | |
| | | 256 | | | Never alcohol drinker | Per 1 serving day | 1.17 (0.90 - 1.50) | ✓ | ✓ | ✓ | | ✓ | ✓ | ✓ E | ✓ D | ✓ | ✓ | ✓ | |
| | | 750 | | | Alcohol intake 0.1-14.9 g/day | Per 1 serving day | 1.15 (0.95 - 1.39) | ✓ | ✓ | ✓ | | ✓ | ✓ | ✓ E | ✓ D | ✓ | ✓ | ✓ | |
| | | 269 | | | Alcohol intake ≥15 g/d | Per 1 serving day | 1.22 (0.88 - 1.68) | ✓ | ✓ | ✓ | | ✓ | ✓ | ✓ E | ✓ D | ✓ | ✓ | ✓ | |
| | | 644 | | | Physical activity < median | Per 1 serving day | 1.23 (1.03 - 1.47) | ✓ | ✓ | ✓ | ✓ | ✓ | | ✓ E | ✓ D | ✓ | ✓ | ✓ | |
| | | 596 | | | Physical activity > median | Per 1 serving day | 1.07 (0.85 - 1.35) | ✓ | ✓ | ✓ | ✓ | ✓ | | ✓ E | ✓ D | ✓ | ✓ | ✓ | |
| | | 636 | | | Fruit intake≤1 serving/day | Per 1 serving day | 1.16 (0.98, 1.38) | ✓ | ✓ | ✓ | ✓ | ✓ | | ✓ E | ✓ D | ✓ | ✓ | ✓ | |
| | | 639 | | | Fruit intake>1 serving/day | Per 1 serving day | 1.24 (0.99, 1.56) | ✓ | ✓ | ✓ | ✓ | ✓ | | ✓ | ✓ | ✓ | ✓ | ✓ | |

| Author, Year Country (PMID) | Study name, type Age, sex, ethnicity, characteristics of participants | Cases/ Study size Median follow-up (years) | Case ascertainment | Exposure Assessment, detail | Outcome Sub-population | Comparison | RR (95%CI) Ptrend | Adjustments Age | Sex | Smo | Alc | Obe | Phy | Diet | Med | Mens | Repr | Oth | Skin |
|---|---|---|---|---|---|---|---|---|---|---|---|---|---|---|---|---|---|---|---|
| | | | | | | | | | | | | | | E | D | | | | |
| | | | | **Substituting SSB with ASB**: substituting 1 serving of SSBs per day with an equivalent amount of ASBs | Incidence, Proximal colon cancer | Per 1 serving day | 0.84 (0.77 – 0.95) | ✓ | ✓ | ✓ | ✓ | ✓ | | ✓ E | ✓ D | ✓ | ✓ | ✓ | |
| Leung 2021 Japan (33563648)[101] | The Japan Public Health Center-based Prospective Study Prospective Cohort Age:56.5 years M/F Japanese | 258 / 33,709 15 years | Cancer registry and death certificates | Self-administered Questionnaire, **SSB**: Beta-carotene fortified beverages, calcium fortified beverages, canned coffee, carbonated beverages, 100% fruit juices, lactic acid bacteria beverages and vitamin fortified beverages | Incidence, Proximal colon cancer, male | ≥254 ml/day vs Non consumers | 0.82 (0.57 - 1.19) Ptrend:0.17 | ✓ | | ✓ | ✓ | ✓ | ✓ | ✓ E | ✓ D | | | | |
| | | | | | | Per 100 ml day | 0.99 (0.95 - 1.04) | ✓ | | ✓ | ✓ | ✓ | ✓ | ✓ E | ✓ D | | | | |
| | | 322 / 40,361 15 years | | | Incidence, Proximal colon cancer, female | ≥254 ml/day vs Non consumers | 1.45 (1.01 - 2.09) Ptrend:0.22 | ✓ | | ✓ | ✓ | ✓ | ✓ | ✓ E | ✓ D | | | | |
| | | | | | | Per 100 ml day | 0.98 (0.92 - 1.06) | ✓ | | ✓ | ✓ | ✓ | ✓ | ✓ E | ✓ D | | | | |
| | | 175 | | | Male, BMI <25 kg/m$^2$ | Per 100 ml day | 1.02 (0.97 - 1.07) | ✓ | | ✓ | ✓ | | ✓ | ✓ E | ✓ D | | | | |
| | | 83 | | | Male, BMI ≥25 kg/m$^2$ | Per 100 ml day | 0.93 (0.84 - 1.02) | ✓ | | ✓ | ✓ | | ✓ | ✓ E | ✓ D | | | | |
| | | 226 | | | Female, BMI <25 kg/m$^2$ | Per 100 ml day | 0.99 (0.91 - 1.07) | ✓ | | ✓ | ✓ | | ✓ | ✓ E | ✓ D | | | | |
| | | 96 | | | Female, BMI ≥25 kg/m$^2$ | Per 100 ml day | 0.97 (0.85 - 1.12) | ✓ | | ✓ | ✓ | | ✓ | ✓ E | ✓ D | | | | |
| | | 158 | | | Male, ever smokers | Per 100 ml day | 1.02 (0.97 - 1.07) | ✓ | | | ✓ | ✓ | ✓ | ✓ E | ✓ D | | | | |
| | | 100 | | | Male, never smokers | Per 100 ml day | 0.93 (0.84 - 1.02) | ✓ | | | ✓ | ✓ | ✓ | ✓ E | ✓ D | | | | |
| | | 17 | | | Female, ever smokers | Per 100 ml day | 0.88 (0.65 - 1.19) | ✓ | | | ✓ | ✓ | ✓ | ✓ E | ✓ D | | | | |
| | | 305 | | | Female, never smokers | Per 100 ml day | 0.99 (0.92 - 1.07) | ✓ | | | ✓ | ✓ | ✓ | ✓ E | ✓ D | | | | |
| | | 185 | | | Male current drinker | Per 100 ml day | 0.97 (0.91 - 1.03) | ✓ | | ✓ | | ✓ | ✓ | ✓ E | ✓ D | | | | |
| | | 73 | | | Male, non-drinker | Per 100 ml day | 1.02 (0.95 - 1.09) | ✓ | | ✓ | | ✓ | ✓ | ✓ E | ✓ D | | | | |
| | | 51 | | | Female, current drinker | Per 100 ml day | 0.92 (0.75 – 1.11) | ✓ | | ✓ | | ✓ | ✓ | ✓ E | ✓ D | | | | |

| Author, Year Country (PMID) | Study name, type Age, sex, ethnicity, characteristics of participants | Cases/ Study size Median follow-up (years) | Case ascertainment | Exposure Assessment, detail | Outcome Sub-population | Comparison | RR (95%CI) Ptrend | Adjustments: Age | Sex | Smo | Alc | Obe | Phy | Diet | Med | Mens | Repr | Oth | Skin |
|---|---|---|---|---|---|---|---|---|---|---|---|---|---|---|---|---|---|---|---|
| | | 271 | | | Female, non-drinkers | Per 100 ml day | 1.00 (0.92 - 1.08) | ✓ | | ✓ | | ✓ | ✓ | ✓ E | ✓ D | | | | |
| | | 126 | | | Male, MET<31.85 | Per 100 ml day | 0.96 (0.90 - 1.04) | ✓ | | ✓ | ✓ | ✓ | | ✓ E | ✓ D | | | | |
| | | 132 | | | Male, MET ≥31.85 | Per 100 ml day | 1.01 (0.96 - 1.08) | ✓ | | ✓ | ✓ | ✓ | | ✓ E | ✓ D | | | | |
| | | 134 | | | Female, MET <31.85 | Per 100 ml day | 0.97 (0.87 - 1.09) | ✓ | | ✓ | ✓ | ✓ | | ✓ E | ✓ D | | | | |
| | | 134 | | | Female, MET ≥31.85 | Per 100 ml day | 0.99 (0.90 - 1.09) | ✓ | | ✓ | ✓ | ✓ | | ✓ E | ✓ D | | | | |
| Kanehara 2024 USA (38795743)[81] | Multiethnic Cohort Prospective Cohort Range:45-75 years M/F Mixed | 2,074 / 192,651 19.3 years | cancer registries | Quantitative FFQ, **SSB**: Chocolate milk, cocoa or Ovaltine; milkshakes or malts; regular sodas; fruit drink (not 100% fruit juice); cappuccino, regular coffee, decaffeinated coffee, and black tea (if sugar or honey were added to them). | Incidence, Proximal colon cancer | Median 351.2 vs 0 g/1000 kcal | 0.98 (0.84 - 1.14) Ptrend:0.93 | ✓ | ✓ | ✓ | ✓ | ✓ | ✓ | ✓ E | ✓ D | | ✓ | ✓ | |
| Yuan 2022 USA (35470384)[178] | Nurses' Health Study + Health Professionals Follow-up Study Prospective Cohort M/F White Health professionals | 158 / 48,384 28 years | self-report & biennial questionnaire & active follow-up for participant deaths | Validated FFQ, **SSB**: Sugar-sweetened carbonated (caffeinated colas, caffeine-free colas, other carbonated noncola), and noncarbonated (fruit punches, lemonades, or other fruit drinks) beverages. | Mortality, Proximal colon cancer, male | Per 1 serving day | 1.47 (1.07 - 2.02) | ✓ | | ✓ | ✓ | ✓ | ✓ | ✓ E | ✓ D | ✓ | ✓ | ✓ | |
| | | 251 / 74,727 30 years | | | Mortality, Proximal colon cancer, female | Per 1 serving day | 1.30 (0.97 - 1.75) | ✓ | | ✓ | ✓ | ✓ | ✓ | ✓ E | ✓ D | ✓ | ✓ | ✓ | |
| | | 40 | | | <60 years old at diagnosis | Per 1 serving day | 1.98 (1.32, 2.97) | ✓ | | ✓ | ✓ | ✓ | ✓ | ✓ E | ✓ D | ✓ | ✓ | ✓ | |
| | | 121 | | | 60 to 60 years old at diagnosis | Per 1 serving day | 1.26 (0.85, 1.86) | ✓ | | ✓ | ✓ | ✓ | ✓ | ✓ E | ✓ D | ✓ | ✓ | ✓ | |
| | | 248 | | | ≥70 years old at diagnosis | Per 1 serving day | 1.34 (0.96, 1.87) | ✓ | | ✓ | ✓ | ✓ | ✓ | ✓ E | ✓ D | ✓ | ✓ | ✓ | |
| | | 204 | | | BMI<25 $kg/m^2$ | Per 1 serving day | 1.42 (1.06, 1.89) | ✓ | ✓ | ✓ | ✓ | | ✓ | ✓ E | ✓ D | ✓ | ✓ | ✓ | |
| | | 204 | | | BMI≥25 $kg/m^2$ | Per 1 serving day | 1.39 (1.03, 1.87) | ✓ | ✓ | ✓ | ✓ | | ✓ | ✓ E | ✓ D | ✓ | ✓ | ✓ | |
| | | 258 | | | Ever smokers | Per 1 serving day | 1.29 (0.97, 1.72) | ✓ | ✓ | | ✓ | ✓ | ✓ | ✓ E | ✓ D | ✓ | ✓ | ✓ | |
| | | 148 | | | Never smoker | Per 1 serving day | 1.53 (1.11, 2.12) | ✓ | ✓ | | ✓ | ✓ | ✓ | ✓ E | ✓ D | ✓ | ✓ | ✓ | |
| | | 82 | | | Never alcohol drinker | Per 1 serving day | 1.27 (0.80, 2.01) | ✓ | ✓ | ✓ | | ✓ | ✓ | ✓ E | ✓ D | ✓ | ✓ | ✓ | |

| Author, Year Country (PMID) | Study name, type Age, sex, ethnicity, characteristics of participants | Cases/ Study size Median follow-up (years) | Case ascertainment | Exposure Assessment, detail | Outcome Sub-population | Comparison | RR (95%CI) Ptrend | Adjustments Age | Sex | Smo | Alc | Obe | Phy | Diet | Med | Mens | Repr | Oth | Skin |
|---|---|---|---|---|---|---|---|---|---|---|---|---|---|---|---|---|---|---|---|
| | | 239 | | | Alcohol intake 0.1-14.9 g/day | Per 1 serving day | 1.36 (1.03, 1.80) | ✓ | ✓ | ✓ | | ✓ | ✓ | ✓ E | ✓ D | ✓ | ✓ | ✓ | |
| | | 88 | | | Alcohol intake ≥15 g/d | Per 1 serving day | 1.95 (1.19, 3.21) | ✓ | ✓ | ✓ | | ✓ | ✓ | ✓ E | ✓ D | ✓ | ✓ | ✓ | |
| | | 222 | | | Physical activity < median | Per 1 serving day | 1.60 (1.25, 2.06) | ✓ | ✓ | ✓ | ✓ | ✓ | | ✓ E | ✓ D | ✓ | ✓ | ✓ | |
| | | 182 | | | Physical activity > median | Per 1 serving day | 1.17 (0.80, 1.72) | ✓ | ✓ | ✓ | ✓ | ✓ | | ✓ E | ✓ D | ✓ | ✓ | ✓ | |
| | | 200 | | | Fruit intake≤1 serving/day | Per 1 serving day | 1.32 (1.01, 1.73) | ✓ | ✓ | ✓ | ✓ | ✓ | | ✓ E | ✓ D | ✓ | ✓ | ✓ | |
| | | 209 | | | Fruit intake>1 serving/day | Per 1 serving day | 1.52 (1.08, 2.14) | ✓ | ✓ | ✓ | ✓ | ✓ | | ✓ E | ✓ D | ✓ | ✓ | ✓ | |
| | | 409 / 121,111 28-30 years | | **Substituting SSB with ASB**: substituting 1 serving of SSBs per day with an equivalent amount of ASBs | Mortality, Proximal colon cancer | Per 1 serving day | 0.80 (0.71 – 0.95) | ✓ | ✓ | ✓ | ✓ | ✓ | | ✓ E | ✓ D | ✓ | ✓ | ✓ | |
| Pacheco 2019 USA (31596902)[135] | California Teachers Study, 1995 Prospective Cohort Age:52 years F Mostly white teachers | 600 / 99,798 20.1 years | Cancer registry and national death index | Self-administered Questionnaire, **SSB**: sweetened carbonated and noncarbonated beverages (caloric soft drinks, sweetened bottled waters and/or teas, and fruit drinks) | Incidence, Distal colorectum cancer | ≥1 serving/day vs Rare or never | 1.22 (0.82 - 1.80) Ptrend:0.101 | ✓ | | ✓ | ✓ | | | ✓ | ✓ | ✓ | ✓ | ✓ | |
| Leung 2021 Japan (33563648)[101] | The Japan Public Health Center-based Prospective Study Prospective Cohort Age:56.5 years M/F Japanese | 311 / 33,709 15 years | Cancer registry and death certificates | Self-administered Questionnaire, **SSB**: Beta-carotene fortified beverages, calcium fortified beverages, canned coffee, carbonated beverages, 100% fruit juices, lactic acid bacteria beverages and vitamin fortified beverages | Incidence, Distal colon cancer, male | ≥254 ml/day vs Non consumers | 0.83 (0.60 - 1.15) Ptrend:0.42 | ✓ | | ✓ | ✓ | ✓ | ✓ | ✓ E | ✓ D | | | | |
| | | 311 / 33,709 15 years | | | | Per 100 ml day | 1.00 (0.95 - 1.04) | ✓ | | ✓ | ✓ | ✓ | ✓ | ✓ E | ✓ D | | | | |
| | | 160 / 40,361 years | | | Incidence, Distal colon cancer, female | ≥254 ml/day vs non consumers | 1.38 (0.86 - 2.22) Ptrend:0.05 | ✓ | | ✓ | ✓ | ✓ | ✓ | ✓ E | ✓ D | | | | |
| | | 160 / 40,361 15 years | | | | Per 100 ml day | 1.00 (0.91 - 1.10) | ✓ | | ✓ | ✓ | ✓ | ✓ | ✓ E | ✓ D | | | | |
| Kanehara 2024 USA (38795743)[81] | Multiethnic Cohort Prospective Cohort Range:45-75 years M/F Mixed | 1,173 / 192,651 19.3 years | cancer registries | Quantitative FFQ, **SSB**: Chocolate milk, cocoa or Ovaltine; milkshakes or malts; regular sodas; fruit drink (not 100% fruit juice); cappuccino, regular coffee, decaffeinated coffee, and black tea (if sugar or honey were added to them). | Incidence, Distal colon cancer | Median 351.2 vs 0 g/1000 kcal | 1.15 (0.94 - 1.41) Ptrend:0.12 | ✓ | ✓ | ✓ | ✓ | ✓ | ✓ | ✓ E | ✓ D | | ✓ | ✓ | |

| Author, Year Country (PMID) | Study name, type Age, sex, ethnicity, characteristics of participants | Cases/ Study size Median follow-up (years) | Case ascertainment | Exposure Assessment, detail | Outcome Sub-population | Comparison | RR (95%CI) Ptrend | Adjustments Age | Sex | Smo | Alc | Obe | Phy | Diet | Med | Mens | Repr | Oth | Skin |
|---|---|---|---|---|---|---|---|---|---|---|---|---|---|---|---|---|---|---|---|
| Yuan 2022 USA (35470384)[178] | Nurses' Health Study + Health Professionals Follow-up Study Prospective Cohort M/F White Health professionals | 835 / 121,111 | self-report & biennial questionnaire & active follow-up for participant deaths | Validated FFQ, **SSB**: Sugar-sweetened carbonated (caffeinated colas, caffeine-free colas, other carbonated noncola), and noncarbonated (fruit punches, lemonades, or other fruit drinks) beverages. | Incidence, Distal colon cancer | >1/day vs never | 0.81 (0.54 - 1.20) | ✓ | ✓ | ✓ | ✓ | ✓ | ✓ | ✓ E | ✓ D | ✓ | ✓ | ✓ | |
| | | 835 / 121,111 | | | | Per 1 serving day | 0.88 (0.73 - 1.06) | ✓ | ✓ | ✓ | ✓ | ✓ | ✓ | ✓ E | ✓ D | ✓ | ✓ | ✓ | |
| | | 835 / 121,111 | | **Substituting SSB with ASB**: substituting 1 serving of SSBs per day with an equivalent amount of ASBs | | Per 1 serving day | Not significant | ✓ | ✓ | ✓ | ✓ | ✓ | | ✓ E | ✓ D | ✓ | ✓ | ✓ | |
| | | 266 / 121,111 | | **SSB**: Sugar-sweetened carbonated (caffeinated colas, caffeine-free colas, other carbonated noncola), and noncarbonated (fruit punches, lemonades, or other fruit drinks) beverages | Mortality, Distal colon cancer | Per 1 serving day | 0.97 (0.70 - 1.34) | ✓ | ✓ | ✓ | ✓ | ✓ | ✓ | ✓ E | ✓ D | ✓ | ✓ | ✓ | |
| | | 835 / 121,111 | | **Substituting SSB with ASB**: substituting 1 serving of SSBs per day with an equivalent amount of ASBs | | Per 1 serving day | Not significant | ✓ | ✓ | ✓ | ✓ | ✓ | | ✓ E | ✓ D | ✓ | ✓ | ✓ | |
| Leung 2021 Japan (33563648)[101] | The Japan Public Health Center-based Prospective Study Prospective Cohort Age:56.5 years M/F Japanese | 334 / 33,709 15 years | Cancer registry and death certificates | Self-administered Questionnaire, **SSB**: Beta-carotene fortified beverages, calcium fortified beverages, canned coffee, carbonated beverages, 100% fruit juices, lactic acid bacteria beverages and vitamin fortified beverages | Incidence, Rectal cancer, male | ≥254 ml/day vs non consumers | 0.91 (0.66 - 1.26) Ptrend:0.94 | ✓ | | ✓ | ✓ | ✓ | ✓ | ✓ E | ✓ D | | | | |
| | | | | | | Per 100 ml day | 0.98 (0.94 - 1.02) | ✓ | | ✓ | ✓ | ✓ | ✓ | ✓ E | ✓ D | | | | |
| | | 193 / 40,361 15 years | | Self-administered Questionnaire, **SSB**: Beta-carotene fortified beverages, calcium fortified beverages, canned coffee, carbonated beverages, 100% fruit juices, lactic acid bacteria beverages and vitamin fortified beverages | Incidence, Rectal cancer, female | ≥254 ml/day vs non consumers | 0.91 (0.62 - 1.35) Ptrend:0.66 | ✓ | | ✓ | ✓ | ✓ | ✓ | ✓ E | ✓ D | | | | |
| | | | | | | Per 100 ml day | 1.04 (0.96 - 1.11) | ✓ | | ✓ | ✓ | ✓ | ✓ | ✓ E | ✓ D | | | | |
| Kanehara 2024 USA (38795743)[81] | Multiethnic Cohort Prospective Cohort Range:45-75 years M/F Mixed | 1,014 / 192,651 19.3 years | cancer registries | Quantitative FFQ, **SSB**: Chocolate milk, cocoa or Ovaltine; milkshakes or malts; regular sodas; fruit drink (not 100% fruit juice); cappuccino, regular coffee, decaffeinated coffee, and black tea (if sugar or honey were added to them). | Incidence, Rectal cancer | Median 351.2 vs 0 g/1000 kcal | 0.99 (0.80 - 1.23) Ptrend:0.93 | ✓ | ✓ | ✓ | ✓ | ✓ | ✓ | ✓ E | ✓ D | | ✓ | ✓ | |

| Author, Year Country (PMID) | Study name, type Age, sex, ethnicity, characteristics of participants | Cases/ Study size Median follow-up (years) | Case ascertainment | Exposure Assessment, detail | Outcome Sub-population | Comparison | RR (95%CI) Ptrend | Adjustments | | | | | | | | | | | |
|---|---|---|---|---|---|---|---|---|---|---|---|---|---|---|---|---|---|---|---|
| | | | | | | | | Age | Sex | Smo | Alc | Obe | Phy | Diet | Med | Mens | Repr | Oth | Skin |
| Yuan 2022 USA (35470384)[178] | Nurses' Health Study + Health Professionals Follow-up Study<br>Prospective Cohort<br>M/F<br>White<br>Health professionals | 623 / 121,111 | self-report & biennial questionnaire & active follow-up for participant deaths | Validated FFQ,<br>**SSB**:<br>Sugar-sweetened carbonated (caffeinated colas, caffeine-free colas, other carbonated noncola), and noncarbonated (fruit punches, lemonades, or other fruit drinks) beverages. | Incidence, Rectal cancer | >1/day vs never | 0.70 (0.44 - 1.12) | ✓ | ✓ | ✓ | ✓ | ✓ | ✓ | ✓ E | ✓ D | ✓ | ✓ | ✓ | |
| | | | | | | Per 1 serving day | 0.89 (0.72 - 1.09) | ✓ | ✓ | ✓ | ✓ | ✓ | ✓ | ✓ E | ✓ D | ✓ | ✓ | ✓ | |
| | | | | **Substituting SSB with ASB**:<br>substituting 1 serving of SSBs per day with an equivalent amount of ASBs | | Per 1 serving day | Not significant | ✓ | ✓ | ✓ | ✓ | ✓ | | ✓ E | ✓ D | ✓ | ✓ | ✓ | |
| Jin 2023 UK (38004195)[78] | UK Biobank<br>Prospective Cohort<br>Age:55.87 years<br>M/F<br>Mainly white | 359 / 118,210<br>12.8 years | hospital records, cancer registry, death registry | 24 Hour Diet Recall,<br>**SSB**:<br>Sugar-sweetened beverages & other sugary drinks. | Incidence, Rectal cancer | Per 1 SD day | 1.01 (0.90 - 1.12) | ✓ | ✓ | ✓ | | ✓ | ✓ | ✓ E | ✓ D | | | ✓ | |
| Yuan 2022 USA (35470384)[178] | Nurses' Health Study + Health Professionals Follow-up Study<br>Prospective Cohort<br>M/F<br>White<br>Health professionals | 226 / 121,111 | self-report & biennial questionnaire & active follow-up for participant deaths | Validated FFQ,<br>**SSB**:<br>Sugar-sweetened carbonated (caffeinated colas, caffeine-free colas, other carbonated noncola), and noncarbonated (fruit punches, lemonades, or other fruit drinks) beverages. | Mortality, Rectal cancer | Per 1 serving day | 0.92 (0.64 - 1.31) | ✓ | ✓ | ✓ | ✓ | ✓ | ✓ | ✓ E | ✓ D | ✓ | ✓ | ✓ | |
| | | | | **Substituting SSB with ASB**:<br>substituting 1 serving of SSBs per day with an equivalent amount of ASBs | | Per 1 serving day | Not significant | ✓ | ✓ | ✓ | ✓ | ✓ | | ✓ E | ✓ D | ✓ | ✓ | ✓ | |
| Zhang 2010 multi-national (20453203)[182] | Pooled analysis Cohort (n=13 cohorts)<br>M/F<br>6-20 years | 4895 / 731,441 | Self-report verified by medical record | FFQ,<br>**Carbonated SSB**:<br>Carbonated sugar sweetened soft drinks | Incidence, Coln cancer | Per 12-ounce day | 1.00 (0.91 - 1.10) | ✓ | | ✓ | ✓ | ✓ | ✓ | ✓ E | | | ✓ | ✓ | |
| | | 1477 / 239,193 | | | Male | Per 12-ounce day | 1.05 (0.91 - 1.22) | ✓ | | ✓ | ✓ | ✓ | ✓ | ✓ E | | | ✓ | ✓ | |
| | | 3418 / 492,248 | | | Female | Per 12-ounce day | 0.96 (0.84 - 1.09) | ✓ | | ✓ | ✓ | ✓ | ✓ | ✓ E | | | ✓ | ✓ | |
| | | 780 / 239,193 | | | Proximal colon, male | Per 12-ounce day | 1.13 (0.93 – 1.46) | ✓ | | ✓ | ✓ | ✓ | ✓ | ✓ E | | | ✓ | ✓ | |
| | | 1846 / 492,248 | | | Proximal colon, female | Per 12-ounce day | 0.91 (0.76 – 1.10) | ✓ | | ✓ | ✓ | ✓ | ✓ | ✓ E | | | | ✓ | |
| | | 656 / 239,193 | | | Distal colon, male | Per 12-ounce day | 0.97 (0.76 – 1.24) | ✓ | | ✓ | ✓ | ✓ | ✓ | ✓ E | | | | ✓ | |

| Author, Year Country (PMID) | Study name, type Age, sex, ethnicity, characteristics of participants | Cases/ Study size Median follow-up (years) | Case ascertainment | Exposure Assessment, detail | Outcome Sub-population | Comparison | RR (95%CI) Ptrend | Adjustments | | | | | | | | | | | |
|---|---|---|---|---|---|---|---|---|---|---|---|---|---|---|---|---|---|---|---|
| | | | | | | | | Age | Sex | Smo | Alc | Obe | Phy | Diet | Med | Mens | Repr | Oth | Skin |
| | | 1283 / 492,248 | | | Distal colon, female | Per 12-ounce day | 1.04 (0.86 – 1.25) | ✓ | | ✓ | ✓ | ✓ | ✓ | ✓ E | | | ✓ | ✓ | |
| | | 1972 | | | Never smokers | Per 12-ounce day | 0.98 (0.80 – 1.18) | ✓ | | ✓ | ✓ | ✓ | ✓ | ✓ E | | | ✓ | ✓ | |
| | | 1619 | | | Past smokers | Per 12-ounce day | 1.10 (0.93 0 1.30) | ✓ | | ✓ | ✓ | ✓ | ✓ | ✓ E | | | | ✓ | |
| | | 837 | | | Current smokers | Per 12-ounce day | 1.02 (0.89 – 1.21) | ✓ | | ✓ | ✓ | ✓ | ✓ | ✓ E | | | ✓ | ✓ | |
| | | 1813 | | | Non-drinkers | Per 12-ounce day | 1.07 (0.93 – 1.22) | ✓ | | ✓ | ✓ | ✓ | ✓ | ✓ E | | | | ✓ | |
| | | 2227 | | | Alcohol intake <15 g/day | Per 12-ounce day | 0.94 (0.80 – 1.10) | ✓ | | ✓ | ✓ | ✓ | ✓ | ✓ E | | | ✓ | ✓ | |
| | | 845 | | | Alcohol intake ≥15 g/day | Per 12-ounce day | 1.14 (0.89 – 1.47) | ✓ | | ✓ | ✓ | ✓ | ✓ | ✓ E | | | | ✓ | |
| | | 1926 | | | Low physical activity | Per 12-ounce day | 0.94 (0.82 – 1.09) | ✓ | | ✓ | ✓ | ✓ | ✓ | ✓ E | | | | ✓ | |
| | | 1482 | | | Median physical activity | Per 12-ounce day | 0.94 (0.75 – 1.19) | ✓ | | ✓ | ✓ | ✓ | ✓ | ✓ E | | | | ✓ | |
| | | 930 | | | High physical activity | Per 12-ounce day | 1.21 (0.99 – 1.48) | ✓ | | ✓ | ✓ | ✓ | ✓ | ✓ E | | | | ✓ | |
| | | 2254 | | | BMI<25 kg/m$^2$ | Per 12-ounce day | 0.94 (0.77 – 1.14) | ✓ | | ✓ | ✓ | ✓ | ✓ | ✓ E | | | | ✓ | |
| | | 2519 | | | BMI≥25 kg/m$^2$ | Per 12-ounce day | 1.03 (0.91 – 1.17) | ✓ | | ✓ | ✓ | ✓ | ✓ | ✓ E | | | | ✓ | |
| | | 1686 | | | Ever postmenopausal hormone use | Per 12-ounce day | 1.11 (0.94 – 1.31) | ✓ | | ✓ | ✓ | ✓ | ✓ | ✓ E | | | | ✓ | |
| | | 1001 | | | Never postmenopausal hormone use | Per 12-ounce day | 0.72 (0.48 – 1.08) | ✓ | | ✓ | ✓ | ✓ | ✓ | ✓ E | | | | ✓ | |
| **Liver** | | | | | | | | | | | | | | | | | | | |
| McCullough 2022 USA (36107009)[113] | Cancer Prevention Study II Prospective Cohort M/F Mainly white | 2,722 / 934,777 27.7 years | self-report & death certificates | Questionnaire, **SSB**: Regular colas and other nondiet soft drinks. | Mortality, Liver cancer | 2+ drinks/day vs never | 1.07 (0.92 - 1.24) Ptrend:0.648 | ✓ | | ✓ | ✓ | | | ✓ * | | | | ✓ | |
| | | | | | | Per 1 drink day | 1.02 (0.98 - 1.06) Ptrend:0.340 | ✓ | | ✓ | ✓ | | | ✓ * | | | | ✓ | |
| | | | | | | | 1.01 (0.97 - 1.06) Ptrend:0.522 | ✓ | | ✓ | ✓ | ✓ | | ✓ * | | | | ✓ | |
| | | | | | | Wit BMI adjustment | 1.01 (0.97 – 1.06) | | | | | | | | | | | | |
| Stepien 2016 multi-national (25528243)[159] | European Prospective Investigation into Cancer and Nutrition study Prospective Cohort Mainly white | 127 / 477,206 11.4 years | cancer registries | Semi-quantitative FFQ: SSB: Sugar sweetened soft drinks | Incidence, HCC | Per 1 serving week | 1.00 (0.95 - 1.06) | | | ✓ | ✓ | ✓ | ✓ | ✓ | D | | | ✓ | |

| Author, Year Country (PMID) | Study name, type Age, sex, ethnicity, characteristics of participants | Cases/ Study size Median follow-up (years) | Case ascertainment | Exposure Assessment, detail | Outcome Sub-population | Comparison | RR (95%CI) Ptrend | Adjustments Age | Sex | Smo | Alc | Obe | Phy | Diet | Med | Mens | Repr | Oth | Skin |
|---|---|---|---|---|---|---|---|---|---|---|---|---|---|---|---|---|---|---|---|
| Jones 2022 USA (35728406)[79] | NIH-AARP + PLCO Pooled Analysis Cohorts M/F Mainly white | 587 / 553,874 12 years | NIH-AARP: cancer reg, PLCO: self-report + medical record/cancer reg | Questionnaire, **SSB**: Soda and fruit punch. | Incidence, Liver cancer, no diabetes & ≤12 yrs follow-up | Per times day | 1.12 (0.99 - 1.28) | √ | √ | √ | √ | √ | | E √ E | | | | √ | |
| | | 252 / 553,874 12 years | | | Incidence, Liver cancer, no diabetes & >12 yrs follow-up | Per times day | 0.80 (0.61 - 1.05) | √ | √ | √ | √ | √ | | √ E | | | | √ | |
| Luu 2021 Singapore (33129230)[106] | Singapore Chinese Health Study Prospective Cohort Range:45-74 years M/F Chinese | 561 / 61,321 17.7 years | Singapore Cancer and Singapore Birth and Death Registry | Semi-quantitative FFQ, **SSB**: Sugar sweetened beverages and fruit juice | Incidence, Hepatocellular carcinoma | Per 1 serving day | 0.98 (0.78 - 1.23) | √ | √ | √ | √ | √ | | √ E * | D | | | √ | |
| Zhao 2023 USA (37552302)[186] | Women's Health Initiative Prospective Cohort Range:50-79 years F Mixed Postmenopausal | 207 / 98,786 20.9 | self-administered questionnaire, verified by pathology reports, physician/discharge, operative/radiology reports, tumour registry. | FFQ, **SSB**: Sugar-sweetened beverages | Incidence, Liver cancer | ≥1 serving/day vs never - ≤3 servings/month, adjusted for BMI | 1.85 (1.16 - 2.96) Ptrend:0.01 | √ | | √ | √ | √ | √ | √ E | D | | √ | √ | |
| | | | | | | Not adjusted for BMI | 1.77 (1.11 - 2.82) Ptrend:0.02 | √ | | √ | √ | | √ | √ E | | | √ | √ | |
| | | 182 | | FFQ, **SSB**: Regular soft drinks or fruit drinks (Tang, Kool-Aid, Hi-C, and other fruit drinks) | Incidence, Liver cancer, excluding diabetes at baseline | After exclusion of diabetes | 2.03 (1.25 - 3.30) Ptrend:0.005 | √ | | √ | √ | √ | √ | √ E | D | | √ | √ | |
| Ma 2018 USA (30506561)[222] | Health professionals follow up study & Nurses' Health Study Pooled Analysis Cohorts Range:30-75 years M/F Mostly white | 143 / 138,688 32 years | Self-report, medical records and National Death Index | Validated FFQ, **SSB**: Sugar sweetened beverages and fruit juice | Incidence / Mortality, Hepatocellular carcinoma | Per 1 point | 0.99 (0.93 - 1.05) | √ | √ | √ | | √ | √ | √ E | D | | | √ | |
| Luo 2019 USA (31688024)[105] | Health professionals follow up study & Nurses' Health Study Pooled Analysis Cohorts M/F Mostly white Health professionals | 75 / 49,665 32 years | medical records, cancer registries, national death index | Semi-quantitative FFQ, **SSB**: Sugar-sweetened beverages | Incidence, Hepatocellular carcinoma, male | Q4 vs Q1 | 1.27 (0.62 - 2.60) Ptrend:0.61 | √ | | √ | √ | √ | √ | √ E | | | | √ | |
| | | 85 / 87,943 32 years | | | Incidence, Hepatocellular carcinoma, female | | 1.22 (0.65 - 2.28) Ptrend:0.37 | √ | | √ | √ | √ | √ | √ E | | | | √ | |
| Zhao 2024 USA (39226717)[187] | Southern Community Cohort Study Prospective Cohort Range:40-79 years M/F White, black, other | 453 | National death index and cancer registry | Semi-quantitative FFQ, **Carbonated SSB**: Regular soda: Coke, Sprite. | Incidence, Liver cancer | ≥median vs non-consumer | 0.78 (0.58 - 1.05) Ptrend:0.02 | √ | √ | √ | √ | √ | √ | √ E | √ D | | | √ | |

| Author, Year Country (PMID) | Study name, type Age, sex, ethnicity, characteristics of participants | Cases/ Study size Median follow-up (years) | Case ascertainment | Exposure Assessment, detail | Outcome Sub-population | Comparison | RR (95%CI) Ptrend | Adjustments Age | Sex | Smo | Alc | Obe | Phy | Diet | Med | Mens | Repr | Oth | Skin |
|---|---|---|---|---|---|---|---|---|---|---|---|---|---|---|---|---|---|---|---|
| Jones 2022 USA (35728406)[79] | NIH-AARP + Prostate, Lung, Colorectal and Ovarian Cancer Screening trial Pooled Analysis Cohorts M/F Mainly white | 587 | NIH-AARP: cancer reg, PLCO: self-report + medical record/cancer reg | Questionnaire, **Carbonated SSB**: Regular soda | Incidence, Liver cancer, no diabetes & ≤12 yrs follow-up | Per time day | 1.18 (1.03 - 1.35) | ✓ | ✓ | ✓ | ✓ | ✓ | | ✓ E | | | | ✓ | |
| | | 252 | | | Incidence, Liver cancer, no diabetes & >12 yrs follow-up | Per time day | 0.84 (0.62 - 1.13) | ✓ | ✓ | ✓ | ✓ | ✓ | | ✓ E | | | | ✓ | |
| | | 587 | | Questionnaire, **Fruit drinks**: Fruit punch | Incidence, Liver cancer, no diabetes & ≤12 yrs follow-up | Per time day | 0.98 (0.78 - 1.24) | ✓ | ✓ | ✓ | ✓ | ✓ | | ✓ E | | | | ✓ | |
| | | 252 | | | Incidence, Liver cancer, no diabetes & >12 yrs follow-up | Per time day | 0.88 (0.59 - 1.31) | ✓ | ✓ | ✓ | ✓ | ✓ | | ✓ E | | | | ✓ | |
| Zhao 2024 USA (39226717)[187] | Southern Community Cohort Study Prospective Cohort Range:40-79 years M/F White, black, other | 453 / 73,119 13.9 years | National death index and cancer registry | Semi-quantitative FFQ, **Fruit drinks**: Sweetened fruit drinks: Kool-Aid, punch, Tang, lemonade, or sunny delight | Incidence, Liver cancer | ≥median vs non-consumer g/day | 1.11 (0.86 - 1.45) Ptrend:0.39 | ✓ | ✓ | ✓ | ✓ | ✓ | ✓ | ✓ E | ✓ D | | | ✓ | |
| Zhao 2023 USA (37552302)[186] | Women's Health Initiative Prospective Cohort Range:50-79 years F Mixed Postmenopausal | 207 / 98,786 20.9 | self-administered questionnaire, verified by pathology reports, physician/discharge, operative/radiology reports, tumour registry. | FFQ, **Fruit drinks**: Fruit drinks (Tang, Kool-Aid, Hi-C, and other fruit drinks not including fruit juice) | Incidence, Liver cancer | ≥1 serving/day vs never - ≤3 servings/month | 1.66 (0.73 - 3.80) Ptrend:0.14 | ✓ | | ✓ | ✓ | | ✓ | ✓ E | | | ✓ | ✓ | |
| | | | | FFQ, **SSB without fruit drinks**: | | ≥1 serving/day vs never - ≤3 servings/month | 1.66 (0.97 - 2.85) Ptrend:0.14 | ✓ | | ✓ | ✓ | ✓ | ✓ | ✓ E | D | | ✓ | ✓ | |
| Jones 2022 USA (35728406)[79] | NIH-AARP + Prostate, Lung, Colorectal and Ovarian Cancer Screening trial Pooled Analysis Cohorts M/F Mainly white | 587 / 506,389 | NIH-AARP: cancer reg, PLCO: self-report + medical record/cancer reg | Questionnaire, **Sugar-sweetened fruit drinks**: Regular fruit punch | Incidence, Liver cancer, no diabetes & ≤12 yrs follow-up | Per time day | 0.89 (0.62 - 1.29) | ✓ | ✓ | ✓ | ✓ | ✓ | | ✓ E | | | | ✓ | |
| | | 252 / 506,389 | | | Incidence, Liver cancer, no diabetes & >12 yrs follow-up | Per time day | 0.60 (0.29 - 1.27) | ✓ | ✓ | ✓ | ✓ | ✓ | | ✓ E | | | | ✓ | |
| **Gallbladder/bile ducts** | | | | | | | | | | | | | | | | | | | |
| McCullough 2022 USA (36107009)[113] | Cancer Prevention Study II Prospective Cohort M/F Mainly white | 1,024 / 934,777 27.7 years | self-report & death certificates | Questionnaire, **SSB**: Regular colas and other nondiet soft drinks. | Mortality, Gallbladder cancer | 2+ drinks/day vs never | 1.13 (0.87 - 1.47) Ptrend:0.060 | ✓ | | ✓ | ✓ | | | ✓ * | | | | ✓ | |
| | | | | | | Per 1 drink day | 1.03 (0.96 - 1.11) | ✓ | | ✓ | ✓ | | | ✓ * | | | | ✓ | |

| Author, Year Country (PMID) | Study name, type Age, sex, ethnicity, characteristics of participants | Cases/ Study size Median follow-up (years) | Case ascertainment | Exposure Assessment, detail | Outcome Sub-population | Comparison | RR (95%CI) Ptrend | Adjustments Age | Sex | Smo | Alc | Obe | Phy | Diet | Med | Mens | Repr | Oth | Skin |
|---|---|---|---|---|---|---|---|---|---|---|---|---|---|---|---|---|---|---|---|
| **Pancreas** | | | | | | | | | | | | | | | | | | | |
| McCullough 2022 USA (36107009)[113] | Cancer Prevention Study II Prospective Cohort M/F Mainly white | 9,196 / 934,777 27.7 years | self-report & death certificates | Questionnaire, **SSB**: Regular colas and other nondiet soft drinks. | Mortality, Pancreas cancer | 2+ drinks/day vs never | 1.02 (0.94 - 1.11) Ptrend:0.754 | ✓ | | ✓ | ✓ | | | ✓ * | | | | ✓ | |
| | | 9,196 | | | | Per 1 drink day | 1.00 (0.98 - 1.03) | ✓ | | ✓ | ✓ | | | ✓ * | | | | ✓ | |
| | | 9,196 | | | | With BMI adjustment | 1.00 (0.98 - 1.02) | ✓ | | ✓ | ✓ | ✓ | | ✓ * | | | | ✓ | |
| | | 4,339 | | | Mortality, Pancreas cancer, male | Per 1 drink day | 1.01 (0.98 - 1.04) | ✓ | | ✓ | ✓ | | | ✓ * | | | | ✓ | |
| | | 4,857 | | | Mortality, Pancreas cancer, female | Per 1 drink day | 0.99 (0.95 - 1.03) | ✓ | | ✓ | ✓ | | | ✓ * | | | | ✓ | |
| | | 3,955 | | | Mortality, Pancreas cancer, never smokers | Per 1 drink day | 1.01 (0.97 - 1.05) | ✓ | | | ✓ | | | ✓ * | | | | ✓ | |
| | | 4,234 | | | Mortality, Pancreas cancer, normal weight (BMI 18.5 to <25 kg/m$^2$) | Per 1 drink day | 1.01 (0.97 - 1.05) | ✓ | | ✓ | ✓ | | | ✓ * | | | | ✓ | |
| | | 3,625 | | | Mortality, Pancreas cancer, overweight (BMI 25 to <30 kg/m$^2$) | Per 1 drink day | 0.98 (0.94 - 1.02) | ✓ | | ✓ | ✓ | | | ✓ * | | | | ✓ | |
| | | 1,002 | | | Mortality, Pancreas cancer, Obese (≥30 kg/m$^2$) | Per 1 drink day | 1.03 (0.97 - 1.09) | ✓ | | ✓ | ✓ | | | ✓ * | | | | ✓ | |
| Navarrete-Muñoz 2016 multi-national (27510540)[127] | European Prospective Investigation into Cancer and nutrition Prospective Cohort Age:51 years M/F Mostly white | 686 / 477,199 11.6 years | cancer and pathology registry, active follow up, health insurance record, mortality registry and contact of participants or next-of-kin | Diet Questionnaire, **SSB**: Sugar-sweetened beverages | Incidence / Mortality, Pancreas cancer | >121.4 vs 0.1 - 4.5 g/day | 0.92 (0.67 - 1.28)[1] Ptrend:0.46 | ✓ | ✓ | ✓ | ✓ | | ✓ | ✓ * | | | | ✓ | |
| | | | | | | Per 100 g day | 1.02 (0.97 - 1.08) | ✓ | ✓ | ✓ | ✓ | | ✓ | ✓ * | | | | ✓ | |
| | | 313 | | | Incidence / Mortality, | Per 336 g day | 1.04 (0.82 - 1.32) | ✓ | ✓ | ✓ | ✓ | | ✓ | ✓ * | | | | ✓ | |

| Author, Year Country (PMID) | Study name, type Age, sex, ethnicity, characteristics of participants | Cases/ Study size Median follow-up (years) | Case ascertainment | Exposure Assessment, detail | Outcome Sub-population | Comparison | RR (95%CI) Ptrend | Age | Sex | Smo | Alc | Obe | Phy | Diet | Med | Mens | Repr | Oth | Skin |
|---|---|---|---|---|---|---|---|---|---|---|---|---|---|---|---|---|---|---|---|
| | | | | | Pancreas cancer, male | | | | | | | | | | | | | | |
| | | 373 | | | Incidence / Mortality, Pancreas cancer, female | Per 336 g | 1.14 (0.87 - 1.50) | ✓ | ✓ | ✓ | ✓ | | ✓ | ✓ * | | | | ✓ | |
| | | 213 | | | Incidence / Mortality, Pancreas cancer, Current smoker | Per 336 g | 1.12 (0.85 - 1.47) | ✓ | ✓ | | ✓ | | ✓ | ✓ * | | | | ✓ | |
| | | 201 | | | Incidence / Mortality, Pancreas cancer, Former smoker | Per 336 g | 1.18 (0.86 - 1.63) | ✓ | ✓ | | ✓ | | ✓ | ✓ * | | | | ✓ | |
| | | 257 | | | Incidence / Mortality, Pancreas cancer, Never smoked | Per 336 g | 0.92 (0.61 - 1.40) | ✓ | ✓ | | ✓ | | ✓ | ✓ * | | | | ✓ | |
| Bao 2008 USA (18689380)[39] | NIH- AARP Diet and Health Study Prospective Cohort Age:62 years M/F Mostly white Retired | 1,258 / 487,922 7.2 years | Cancer registry and national death index | Dietary Recall, **SSB**: Regular soft drinks and regular fruit drinks | Incidence / Mortality, Pancreatic Adenocarcinoma | Median intake 430.9 vs 0 g/day | 0.83 (0.67 - 1.04) Ptrend:0.10 | ✓ | ✓ | ✓ | ✓ | ✓ | ✓ | ✓ E | | | | ✓ | |
| | | 775 | | | Incidence / Mortality, Pancreatic Adenocarcinoma, Never smoker or quit 10+ yrs ago | | 1.10 (0.79 - 1.54) Ptrend:0.96 | ✓ | ✓ | ✓ | ✓ | ✓ | ✓ | ✓ E * | | | | ✓ | |
| | | 998 | | | Incidence / Mortality, Pancreatic Adenocarcinoma, BMI <30 $kg/m^2$ | | 0.91 (0.68 - 1.20) Ptrend:0.34 | ✓ | ✓ | ✓ | ✓ | ✓ | ✓ | ✓ E * | | | | ✓ | |
| | | 231 | | | Incidence / Mortality, Pancreatic Adenocarcinoma, BMI ≥30 $kg/m^2$ | | 1.26 (0.71 - 2.24) Ptrend:0.29 | ✓ | ✓ | ✓ | ✓ | ✓ | ✓ | ✓ E * | | | | ✓ | |

| Author, Year Country (PMID) | Study name, type Age, sex, ethnicity, characteristics of participants | Cases/ Study size Median follow-up (years) | Case ascertainment | Exposure Assessment, detail | Outcome Sub-population | Comparison | RR (95%CI) Ptrend | Adjustments Age | Sex | Smo | Alc | Obe | Phy | Diet | Med | Mens | Repr | Oth | Skin |
|---|---|---|---|---|---|---|---|---|---|---|---|---|---|---|---|---|---|---|---|
| Julián-Serrano 2022 USA (35474368)[80] | NIH- AARP Diet and Health Study<br>Prospective Cohort<br>Range:50-71 years<br>M/F | / 535,824<br>16 years | State Health Registry, National Death Index | Semi-quantitative FFQ,<br>**SSB**:<br>Sugar sweetened beverages and fruit juice | Incidence, Pancreatic ductal adenocarcinoma | Per 1 point increase in AHEI-2010 component score | 1.01 (1.00 - 1.02) | ✓ | ✓ | ✓ | ✓ | ✓ | | ✓ E | D | | | | |
| | | / 535,824<br>16 years | | | | Per 1 increase in DASH diet component score | 1.04 (1.01 - 1.06) | ✓ | ✓ | ✓ | | ✓ | | ✓ E | D | | | | |
| Zhang 2020 USA (32716590)[(179)] | Prostate, Lung, Colorectal and Ovarian Cancer Screening trial<br>Prospective Cohort<br>Range:55-74 years<br>M/F | 337 /<br>95,962<br>8.87 years | Self-report, medical records and National Death Index | Diet History Questionnaire,<br>**SSB**:<br>Sugar sweetened beverages and fruit juice | Incidence, Pancreatic cancer | Per 1 serving/day | 1.42 (1.00 – 2.03) | ✓ | ✓ | ✓ | ✓ | ✓ | ✓ | ✓ E * | D | | | ✓ | |
| Zhong 2023 USA (36863825)[188] | Prostate, Lung, Colorectal and Ovarian Cancer Screening trial<br>Prospective Cohort<br>M/F<br>Mostly white | 387 /<br>101,748<br>8.86 years | Annual study update form, medical records, death certificates and family reports. | FFQ,<br>**SSB**: | Incidence, Pancreas cancer | Q4 vs Q1 | 1.01 (0.76 - 1.33)<br>Ptrend:0.519 | ✓ | ✓ | ✓ | ✓ | ✓ | | ✓ * | D | | | ✓ | |
| Chen 2022 Taiwan (35463371)[45] | Taiwan MJ clinics<br>Prospective Cohort<br>Age:39.9 years<br>M/F<br>Asian | 489 /<br>491,929<br>15 years | Cancer registry and death registry | Self-administered Questionnaire,<br>**SSB**:<br>Caffeinated or decaffeinated cola, carbonated SSBs, and noncarbonated SSBs. Bubble milk tea, a Taiwan handmade specialty drink with high sugar content, was included. | Incidence, Pancreas cancer | ≥2 vs 0 - <0.5 serving/day | 1.55 (1.08 - 2.23)<br>Ptrend:0.233 | ✓ | ✓ | ✓ | ✓ | ✓ | ✓ | | D | | | ✓ | |
| Steel 2023 (36306381)[157] | Multiethnic Cohort Study<br>Prospective Cohort<br>Range:45-75 years | 1,779 /<br>177,313<br>19.3 years | State Health Registry, National Death Index | FFQ,<br>**Sugary drinks**:<br>Sugar sweetened beverages | Incidence, Pancreatic cancer | Quintile 5 vs Quintile 1 | 0.98 (0.84 - 1.21)<br>Ptrend:0.36 | ✓ | ✓ | ✓ | ✓ | ✓ | ✓ | ✓ E | D | | | ✓ | |
| Genkinger 2012 (22194529)[64] | Pooled Analysis Cohorts<br>M/F | 2,047 /<br>853,894 | Self-report, medical record, cancer registries and death registries | FFQ,<br>**Carbonated SSB**:<br>Sugar-sweetened Carbonated Soft Drinks; Caffeine-free or caffeinated colas and non-colas carbonated beverages. | Incidence, Pancreas cancer | Per 175 g day | 1.06 (1.02 - 1.12) | ✓ | | ✓ | ✓ | ✓ | | ✓ E | D | | | | |
| | | 1,556 /<br>853,894 | | | Incidence, Adenocarcinoma of pancreas | Per 175 g | 1.08 (1.03 - 1.14) | ✓ | | ✓ | ✓ | ✓ | | ✓ E | D | | | | |
| Nöthlings 2007 USA (17991664 )[131] | Multiethnic Cohort<br>Prospective Cohort<br>Age:60 years<br>M/F<br>Multi-ethnic | 434 /<br>162,150<br>8 years | Cancer registry and national death index | FFQ,<br>**Carbonated SSB**:<br>Nondiet sodas | Incidence, Exocrine cancer of the pancreas | ≥75.7 vs 0 g/1000 kcal/day | 1.07 (0.82 - 1.41)<br>Ptrend:0.54 | ✓ | ✓ | ✓ | | ✓ | | ✓ E | | | | ✓ | |
| Schernhammer 2005 USA (16172216)[151] | Health Professionals Follow-up Study<br>Prospective Cohort<br>Range:40-75 years<br>M<br>Health professionals | 174 /<br>49,364<br>14 years | Death register/ subject or family | FFQ,<br>Sugar-sweetened cola: | Incidence / Mortality, Pancreatic cancer, male | >3 vs <1 servings/month | 0.92 (0.50 - 1.69)<br>Ptrend:0.77 | ✓ | | ✓ | | ✓ | ✓ | ✓ E * | D | | | | |
| | | 205 /<br>88,794<br>20 years | | | Incidence / Mortality, Pancreatic cancer, female | >3 vs <1 servings/month | 1.25 (0.70 - 2.22)<br>Ptrend:0.36 | ✓ | | ✓ | | ✓ | ✓ | ✓ E * | D | | | | |
| **Lung** | | | | | | | | | | | | | | | | | | | |

| Author, Year Country (PMID) | Study name, type Age, sex, ethnicity, characteristics of participants | Cases/ Study size Median follow-up (years) | Case ascertainment | Exposure Assessment, detail | Outcome Sub-population | Comparison | RR (95%CI) Ptrend | Adjustments | | | | | | | | | | | |
|---|---|---|---|---|---|---|---|---|---|---|---|---|---|---|---|---|---|---|---|
| | | | | | | | | Age | Sex | Smo | Alc | Obe | Phy | Diet | Med | Mens | Repr | Oth | Skin |
| McCullough 2022 USA (36107009)[113] | Cancer Prevention Study II<br>Prospective Cohort<br>M/F<br>Mainly white | 34,381 / 934,777<br>27.7 years | self-report & death certificates | Questionnaire,<br>**SSB**:<br>Regular colas and other nondiet soft drinks. | Mortality, Lung cancer | 2+ drinks/day vs never | 0.91 (0.87 - 0.95)<br>Ptrend:<0.0001 | ✓ | | ✓ | ✓ | | | ✓<br>* | | | | ✓ | |
| | | | | | | Per 1 drinks day | 0.98 (0.96 - 0.99) | ✓ | | ✓ | ✓ | | | ✓<br>* | | | | ✓ | |
| Malik 2019 USA (30882235)[109] | Health Professionals Follow-up Study + Nurses' Health Study<br>Pooled Analysis Cohorts<br>M/F<br>Mostly white<br>Health professionals | / 118,363<br>34 years | Physician confirmation | FFQ,<br>**SSB**:<br>Sugar-sweetened caffeinated and noncaffeinated colas, other (noncola) sugar-sweetened carbonated beverages, and noncarbonated sugar-sweetened beverages (fruit punches, lemonades, or other fruit drinks). | Mortality, Lung cancer | Consumption ≥2/day vs <1/month | 1.07 (0.86 - 1.33)<br>Ptrend:0.73 | ✓ | | ✓ | ✓ | ✓ | ✓ | ✓<br>E<br>* | D | | ✓ | ✓ | |
| | | | | | | Per 1 serving day | 1.01 (0.94 - 1.07) | ✓ | | ✓ | ✓ | ✓ | ✓ | ✓<br>E<br>* | D | | ✓ | ✓ | |
| Khan, 2004 Japan (15075007)[85] | Japan, Hokkaido Cohort Study<br>Prospective Cohort<br>Age:58 years<br>M/F | / 1,524<br>14.8 years | Area residency lists | Dietary History Questionnaire,<br>**Cola**:<br>Cola | Mortality, Lung cancer, male | Everyday + several times/weeks vs never + several times/month + several times/year | 0.50 (0.10 - 2.20) | ✓ | | ✓ | | | | | | | | | |
| Anic 2016 USA (26264348)[37] | NIH- AARP Diet and Health Study<br>Prospective Cohort<br>Range:50-71 years<br>M/F<br>Retired | 5,856 / 277,174<br>10.5 years | cancer registries | FFQ,<br>**SSB**:<br>Sugar-sweetened beverages | Incidence, Lung cancer, male | Per 1 point increase in the AHEI-2010 component score | 1.01 (1.00 - 1.02) | ✓ | ✓ | ✓ | ✓ | ✓ | ✓ | ✓<br>E | | | | ✓ | |
| | | | | | | Per 1 point increase in the DASH component score | 1.03 (1.00 - 1.05) | ✓ | ✓ | ✓ | ✓ | ✓ | ✓ | ✓<br>E | | | | ✓ | |
| | | | | | | Per 1 point increase in the HEI-2010 component score | 1.00 (0.99 - 1.00) | ✓ | ✓ | ✓ | ✓ | ✓ | ✓ | ✓<br>E | | | | ✓ | |
| | | 3,416 / 183,596<br>10.5 years | | | Incidence, Lung cancer, female | Per 1 point increase in the DASH component | 1.01 (0.98 - 1.04) | ✓ | ✓ | ✓ | ✓ | ✓ | ✓ | ✓<br>E | | | | ✓ | |
| | | | | | | Per 1 point increase in the AHEI-2010 component score | 1.01 (1.00 - 1.02) | ✓ | ✓ | ✓ | ✓ | ✓ | ✓ | ✓<br>E | | | | ✓ | |
| Zhu 2023 USA (37255940)[190] | Prostate, Lung, Colorectal and Ovarian Cancer Screening Trial<br>Prospective Cohort | 1,642 / 98,459<br>8.84 years | active follow-up/self-report, death | FFQ,<br>**SSB**:<br>Sugar-sweetened beverages | Incidence, Lung cancer | >495.22 vs ≤130.55 g/day | 0.80 (0.69 - 0.91)<br>Ptrend:0.007 | ✓ | ✓ | ✓ | ✓ | ✓ | | ✓<br>E | | | | ✓ | |

| Author, Year Country (PMID) | Study name, type Age, sex, ethnicity, characteristics of participants | Cases/ Study size Median follow-up (years) | Case ascertainment | Exposure Assessment, detail | Outcome Sub-population | Comparison | RR (95%CI) Ptrend | Adjustments: Age | Sex | Smo | Alc | Obe | Phy | Diet | Med | Mens | Repr | Oth | Skin |
|---|---|---|---|---|---|---|---|---|---|---|---|---|---|---|---|---|---|---|---|
| | Age:65.52 years<br>M/F<br>Mainly white<br>Screening Program | | certificates, family reports, medical records to confirm | | | | | | | | | | | | | | | | |
| Lee 2023 Korea (37536716)[97] | Health Examinees-Gem (HEXA-G) study<br>Prospective Cohort<br>Range:40-69 years<br>M/F | 291 / 36,266 | Korea Central Cancer Registry +Cancer Statistics in Korea | Semi-quantitative FFQ,<br>**SSB**:<br>Sugar-sweetened beverages | Incidence, Lung cancer, male | 0 vs >250 g/day | 0.61 (0.19 - 1.99)[1] | | | ✓ | | | | ✓ E | | | | ✓ | |
| | | 243 / 68,130 | | | Incidence, Lung cancer, female | 0 vs >250 g/day | 0.20 (0.03 - 1.59)[1] | | | ✓ | | | | ✓ E | | | | ✓ | |
| **Skin** | | | | | | | | | | | | | | | | | | | |
| McCullough 2022 USA (36107009)[113] | Cancer Prevention Study II<br>Prospective Cohort<br>M/F<br>Mainly white | 2,130 / 934,777<br>27.7 years | self-report & death certificates | Questionnaire,<br>**SSB**:<br>Regular colas and other nondiet soft drinks. | Mortality, Melanoma | 2+ drinks/day vs never | 0.88 (0.74 - 1.06)<br>Ptrend:0.682 | ✓ | | ✓ | ✓ | | | ✓ * | | | | ✓ | |
| | | | | | | Per 1 drink day | 0.97 (0.92 - 1.02) | ✓ | | ✓ | ✓ | | | ✓ * | | | | ✓ | |
| Leone 2020 Spain (32492135)[99] | Seguimiento Universidad de Navarra (SUN) cohort<br>Nested Case Control<br>Range:40-50 years<br>M/F | 101 | self-report physician diagnosis validated by oncologist | Semi-quantitative FFQ,<br>**Carbonated SSB**:<br>Sweetened carbonated beverages | Incidence, Basal cell carcinoma | Quintile 5 vs Quintile 1 (median 99 vs 0 g/day) | 1.67 (0.69 - 4.02)<br>Ptrend:0.6 | ✓ | | ✓ | | | ✓ | ✓ E | | | | ✓ | ✓ |
| **Breast (female)** | | | | | | | | | | | | | | | | | | | |
| Nomura 2016 USA (27578546)[128] | Black Women's Health Study, 1995<br>Prospective Cohort<br>Age:37.8 years<br>F<br>African American<br>African American | 1,827 / 49,103<br>13.86 years | self-report + cancer registry | FFQ,<br>**SSB**:<br>Sugar-sweetened beverages | Incidence, Breast cancer | 0 vs ≥ 250 g/d, baseline data | 0.71 (0.50 - 1.02)[1]<br>Ptrend:0.70 | ✓ | | ✓ | ✓ | ✓ | ✓ | ✓ E | | ✓ | ✓ | ✓ | |
| | | | | | | 0 vs ≥ 250 g/d, time-varying data | 0.73 (0.54 - 1.00)[1] | | | | | | | | | | | | |
| Arthur 2021 Canada (33220638)[38] | Canadian Study of Diet, Lifestyle and Health<br>Case Cohort<br>F | 870 / 2,976<br>15 years | Cancer registry and national death index | Self-administered Questionnaire,<br>**SSB**:<br>Coca Cola, Pepsi, other cola, soft drinks (not dietetic), orange/grapefruit juice, apple/grape juice, other fruit juices, fruit drink/lemonade and power-based drinks/iced tea | Incidence, Breast cancer | >9.3 vs ≤2.9 cups/week | 1.13 (0.91 - 1.39)<br>Ptrend:0.279 | ✓ | | ✓ | ✓ | ✓ | ✓ | ✓ | ✓ | ✓ | ✓ | ✓ | |
| | | | | Self-administered Questionnaire,<br>**SSB**:<br>Coca Cola, Pepsi, other cola, soft drinks (not dietetic), fruit drink/lemonade and power-based drinks/iced tea; without fruit juices | | >3.1 cups/week vs none | 1.08 (0.89 - 1.32)<br>Ptrend:0.252 | ✓ | | ✓ | ✓ | ✓ | ✓ | ✓ | ✓ | ✓ | ✓ | ✓ | |
| Makarem 2018 USA (29674390)[108] | Framingham Offspring Cohort<br>Prospective Cohort<br>Age:55.4 years<br>F | 124 / 1,689 | medical examinations | Semi-quantitative FFQ,<br>**SSB**:<br>Sugar sweetened beverages and fruit juice. | Incidence, Breast cancer | >8.4 vs <3.5 servings/week | 1.00 (0.65 - 1.57) | ✓ | | ✓ | ✓ | ✓ | ✓ | ✓ E | D | ✓ | ✓ | ✓ | |
| | | | | Semi-quantitative FFQ,<br>**SSB**: | | >1 vs 0 servings/week | 1.04 (0.64 - 1.71) | ✓ | | ✓ | ✓ | ✓ | ✓ | ✓ | D | ✓ | ✓ | ✓ | |

| Author, Year Country (PMID) | Study name, type Age, sex, ethnicity, characteristics of participants | Cases/ Study size Median follow-up (years) | Case ascertainment | Exposure Assessment, detail | Outcome Sub-population | Comparison | RR (95%CI) Ptrend | Adjustments Age | Sex | Smo | Alc | Obe | Phy | Diet | Med | Mens | Repr | Oth | Skin |
|---|---|---|---|---|---|---|---|---|---|---|---|---|---|---|---|---|---|---|---|
| | | | | Regular sugar-sweetened soda and fruit drinks (lemonade and punch). | | | | | | | | | | E | | | | | |
| Lee 2023 Korea (37536716)[97] | Health Examinees-Gem (HEXA-G) study<br>Prospective Cohort<br>Range:40-69 years<br>M/F<br>women | 862 / 68,130 | Korea Central Cancer Registry +Cancer Statistics in Korea | Semi-quantitative FFQ,<br>**SSB**:<br>Sugar-sweetened beverages | Incidence, Breast cancer | 0 vs >250 g/day | 1.15 (0.59 - 2.25)[1] | | | ✓ | | | | ✓<br>E | | | | ✓ | |
| Wang 2022 Taiwan (35159055)[169] | National Health Interview Survey<br>Prospective Cohort<br>Age:49.74 years<br>F<br>Taiwanese | 112 / 5,879<br>13.25 years | National Health Insurance Research Database and death registries and a panel of disease experts verifies diagnosis | Interview,<br>**SSB**:<br>Sugary drinks | Incidence, Breast cancer | Intake vs no intake | 1.46 (0.96 - 2.21)<br>Ptrend:0.074 | ✓ | | | ✓ | ✓ | | | | | ✓ | | |
| Romanos-Nanclares 2019 Spain (30284064)[146] | Seguimiento Universidad de Navarra (SUN) cohort study<br>Prospective Cohort<br>Age:33 years<br>F<br>Spanish | 100 / 10,713<br>10 years | Medical records and National Death Index | Semi-quantitative FFQ,<br>**SSB**:<br>Sugar-sweetened beverages | Incidence, Breast cancer | >1/week vs Never/seldom serving | 1.36 (0.74 - 2.50)<br>Ptrend:0.312 | ✓ | | ✓ | ✓ | ✓ | ✓ | ✓<br>E | D | ✓ | ✓ | ✓ | |
| Chazelas 2019 France (31292122)[44] | NutriNet-Santé study<br>Prospective Cohort<br>Age:42.2 years<br>M/F<br>women | 693 / 79,724<br>5.1 years | self-report, medical records reviewed by physicians, health insurance database, mortality registry | 24 Hour Diet Recall,<br>**SSB**:<br>Beverages with more than 5% simple carbohydrates and 100% fruit juices (no added sugar), soft drinks (carbonated or not), syrups, 100% juice, fruit drinks, sugar sweetened hot beverages, milk-based sugar sweetened beverages, sport drinks, and energy drinks. | Incidence, Breast cancer | Per 100 ml day | 1.22 (1.06 - 1.40) | ✓ | | ✓ | ✓ | ✓ | ✓ | ✓<br>E | D | ✓ | ✓ | ✓ | |
| | | | | | | Per 100 ml day | 1.24 (1.02 - 1.50) | ✓ | | ✓ | ✓ | ✓ | ✓ | ✓<br>E | D | ✓ | ✓ | ✓ | |
| Romanos-Nanclares 2021 USA (34114021)[145] | Nurses' Health Study and Nurses' Health Study II<br>Pooled Analysis Cohorts<br>F<br>Health professionals | 7,495 / 82,713<br>36 years | Medical records and National Death Index | FFQ,<br>**SSB**:<br>Sugar-sweetened caffeinated and noncaffeinated colas, other (noncola) sugar-sweetened carbonated beverages, and noncarbonated sugar-sweetened beverages (e.g., punch, lemonade, fruit drink, or sugared iced tea). | Incidence, Breast cancer, NHS | ≥1day vs <1/month servings | 1.09 (0.98 - 1.21)<br>Ptrend:0.10 | ✓ | | | ✓ | ✓ | ✓ | ✓<br>E<br>* | ✓ | ✓ | ✓ | ✓ | |
| | | | | | | Per 1 serving day | 1.04 (0.99 - 1.09) | ✓ | | | ✓ | ✓ | ✓ | ✓<br>E<br>* | ✓ | ✓ | ✓ | ✓ | |
| | | 3,884 / 93,085<br>36 years | Medical records and National Death Index | FFQ,<br>**SSB**:<br>Sugar-sweetened caffeinated and noncaffeinated colas, other (noncola) sugar-sweetened carbonated beverages, and noncarbonated sugar-sweetened beverages (e.g., punch, lemonade, fruit drink, or sugared iced tea). | Incidence, Breast cancer, NHS II | ≥1day vs <1/month servings | 0.93 (0.82 - 1.07)<br>Ptrend:0.12 | ✓ | | | ✓ | ✓ | ✓ | ✓<br>E<br>* | ✓ | ✓ | ✓ | ✓ | |

| Author, Year Country (PMID) | Study name, type Age, sex, ethnicity, characteristics of participants | Cases/ Study size Median follow-up (years) | Case ascertainment | Exposure Assessment, detail | Outcome Sub-population | Comparison | RR (95%CI) Ptrend | Adjustments: Age | Sex | Smo | Alc | Obe | Phy | Diet | Med | Mens | Repr | Oth | Skin |
|---|---|---|---|---|---|---|---|---|---|---|---|---|---|---|---|---|---|---|---|
| Mullee 2019 Denmark,France,Germany,Greece,Italy,Netherlands,Norway,Spain,Sweden,UK (31479109)[126] | European Prospective Investigation into Cancer and nutrition<br>Prospective Cohort<br>Age:50.8 years<br>M/F<br>women | 1,402 / 321,081<br>16.4 years | active follow up, cancer registry, death certificate and National death index | Semi-quantitative FFQ,<br>**SSB**:<br>Sugar Sweetened Soft Drinks | Mortality, Breast cancer | Per 1 serving day | 0.99 (0.94 - 1.05) | ✓ | | | ✓ | ✓ | ✓ | ✓ E * | ✓ | ✓ | ✓ | ✓ | |
| | | | | | | ≥1 vs <1 glasses/month | 1.21 (0.91 - 1.62)<br>Ptrend:0.10 | ✓ | ✓ | ✓ | ✓ | ✓ | ✓ | ✓ E * | | ✓ | ✓ | ✓ | |
| Malik 2019 USA (30882235)[109] | Nurses' Health Study<br>Prospective Cohort<br>F<br>Mostly white<br>Health professionals | NR / 80,647<br>34 years | Physician confirmation | FFQ,<br>**SSB**:<br>Sugar-sweetened caffeinated and noncaffeinated colas, other (noncola) sugar-sweetened carbonated beverages, and noncarbonated sugar-sweetened beverages (fruit punches, lemonades, or other fruit drinks). | Mortality, Breast cancer | Consumption ≥2/day vs <1/month | 1.34 (1.00 - 1.80)<br>Ptrend:0.02 | ✓ | | ✓ | ✓ | ✓ | ✓ | ✓ E * | D | | ✓ | ✓ | |
| | | | | | | Per 1 serving day | 1.09 (1.00 - 1.18) | ✓ | | ✓ | ✓ | ✓ | ✓ | ✓ E * | D | | ✓ | ✓ | |
| McCullough 2022 USA (36107009)[113] | Cancer Prevention Study II<br>Prospective Cohort<br>F<br>Mainly white<br>Postmenopausal | 6,074 / 518,464<br>27.7 years | self-report & death certificates | Questionnaire,<br>**SSB**:<br>Regular colas and other nondiet soft drinks. | Mortality, Breast cancer | 2+ drinks/day vs never | 1.05 (0.93 - 1.19)<br>Ptrend:0.558 | ✓ | | ✓ | ✓ | | | ✓ * | | ✓ | ✓ | ✓ | |
| | | | | | | Per 1 drink day | 1.00 (0.96 - 1.03) | ✓ | | ✓ | ✓ | | | ✓ * | | ✓ | ✓ | ✓ | |
| Nomura 2016 USA (27578546)[128] | Black Women's Health Study, 1995<br>Prospective Cohort<br>Age:37.8 years<br>F<br>African American | 881 / 49,103<br>13.86 years | self-report + cancer registry | FFQ,<br>**SSB**:<br>Sugar-sweetened beverages | Incidence, Postmenopausal breast cancer | 0 vs ≥ 250 g/d, time-varying data | 0.90 (0.62 - 1.30)1<br>Ptrend:0.53 | ✓ | | ✓ | ✓ | ✓ | ✓ | ✓ E | | ✓ | ✓ | ✓ | |
| Arthur 2021 Canada (33220638)[38] | Canadian Study of Diet, Lifestyle and Health<br>Case Cohort<br>F | 397 / 2,976<br>15 years | Cancer registry and national death index | Self-administered Questionnaire,<br>**SSB**:<br>Coca Cola, Pepsi, other cola, soft drinks (not dietetic), orange/grapefruit juice, apple/grape juice, other fruit juices, fruit drink/lemonade and power-based drinks/iced tea | Incidence, Breast cancer, postmenopausal | >9.3 vs ≤2.9 cups/week | 1.17 (0.85 - 1.59)<br>Ptrend:0.410 | ✓ | | ✓ | ✓ | ✓ | ✓ | ✓ | ✓ | ✓ | ✓ | ✓ | |
| Nomura 2016 USA (26756307)[130] | Iowa Women's Health Study<br>Prospective Cohort<br>Range:55-69 years<br>F<br>Postmenopausal | 3,189 / 36,626<br>23 years | Cancer registry | FFQ,<br>**SSB**:<br>Sugar-sweetened beverages | Incidence, Postmenopausal breast cancer | 0 vs >250 g/day | Risk difference:<br>0.00 (-0.01 - 0.01) | ✓ | | ✓ | ✓ | ✓ | ✓ | | | ✓ | ✓ | ✓ | |
| Hodge 2018 Australia (29463332)[70] | Melbourne Collaborative Cohort Study<br>Prospective Cohort<br>M/F<br>women | 946 / 21,492<br>23 years | cancer registry/death records/National Death Index | Self-administered Questionnaire,<br>**SSB**:<br>Sugar Sweetened Soft Drinks | Incidence, Postmenopausal breast cancer | Consumption ≥1/day vs <1/month | 1.11 (0.85 - 1.45)<br>Ptrend:0.05 | ✓ | | ✓ | ✓ | | ✓ | ✓ | | | | ✓ | |
| | | | | | | Linear model | 1.26 (1.00 - 1.58) | | | ✓ | ✓ | | ✓ | ✓ | | | | ✓ | |
| Romanos-Nanclares 2021 USA (34114021)[145] | Nurses' Health Study and Nurses' Health Study II<br>Pooled Analysis Cohorts<br>F<br>Health professionals | 8,140 / 175,798<br>36 years | Medical records and National Death Index | FFQ,<br>**SSB** | Incidence, Postmenopausal breast cancer | ≥1day vs <1/month servings | 1.07 (0.97 - 1.19)<br>Ptrend:0.40 | ✓ | | | ✓ | ✓ | ✓ | ✓ E * | ✓ | ✓ | ✓ | ✓ | |
| | | | | | | Per 1 serving day | 1.04 (0.99 - 1.09) | ✓ | | | ✓ | ✓ | ✓ | ✓ E | ✓ | ✓ | ✓ | ✓ | |

| Author, Year Country (PMID) | Study name, type Age, sex, ethnicity, characteristics of participants | Cases/ Study size Median follow-up (years) | Case ascertainment | Exposure Assessment, detail | Outcome Sub-population | Comparison | RR (95%CI) Ptrend | Adjustments Age | Sex | Smo | Alc | Obe | Phy | Diet * | Med | Mens | Repr | Oth | Skin |
|---|---|---|---|---|---|---|---|---|---|---|---|---|---|---|---|---|---|---|---|
| Chazelas 2019 France (31292122)[44] | NutriNet-Santé study<br>Prospective Cohort<br>Age:42.2 years<br>M/F<br>women | 410 /<br>79,724<br>5.1 years | self-report, medical records reviewed by physicians, health insurance database, mortality registry | 24 Hour Diet Recall,<br>**SSB**:<br>Beverages with more than 5% simple carbohydrates and 100% fruit juices (no added sugar), soft drinks (carbonated or not), syrups, 100% juice, fruit drinks, sugar sweetened hot beverages, milk-based sugar sweetened beverages, sport drinks, and energy drinks | Incidence, Postmenopausal breast cancer | Per 100 ml day | 1.08 (0.88 - 1.33) | ✓ | | ✓ | ✓ | ✓ | ✓ | ✓<br>E | D | ✓ | ✓ | ✓ | |
| Romanos-Nanclares 2019 Spain (30284064)[146] | Seguimiento Universidad de Navarra (SUN) Project<br>Prospective Cohort<br>Age:33 years<br>F<br>Spanish | 34 / 10,713<br>10 years | Medical records and National Death Index | Semi-quantitative FFQ,<br>**SSB**:<br>Sugar-sweetened beverages | Incidence, Breast cancer, postmenopausal | Regular intake vs never/seldom | 2.12 (1.01 - 4.41)<br>Ptrend:0.045 | ✓ | | ✓ | ✓ | ✓ | ✓ | ✓<br>E | D | ✓ | ✓ | ✓ | |
| Nomura 2016 USA (27578546)[128] | Black Women's Health Study, 1995<br>Prospective Cohort<br>Age:37.8 years<br>F<br>African American | 711 /<br>49,103<br>13.86 years | self-report + cancer registry | FFQ,<br>**SSB**:<br>Sugar-sweetened beverages | Incidence, Premenopausal breast cancer | 0 vs ≥ 250 g/d, time-varying data | 0.58 (0.31 - 1.10)[1]<br>Ptrend:0.02 | ✓ | | ✓ | ✓ | ✓ | ✓ | ✓<br>E | | ✓ | ✓ | ✓ | |
| Arthur 2021 Canada (33220638)[38] | Canadian Study of Diet, Lifestyle and Health<br>Case Cohort<br>F | 473 / 2,976<br>15 years | Cancer registry and national death index | Self-administered Questionnaire,<br>**SSB**:<br>Coca Cola, Pepsi, other cola, soft drinks (not dietetic), orange/grapefruit juice, apple/grape juice, other fruit juices, fruit drink/lemonade and power-based drinks/iced tea | Incidence, Breast cancer, premenopausal | >9.3 vs ≤2.9 cups/week | 1.10 (0.81 - 1.49)<br>Ptrend:0.385 | ✓ | | ✓ | ✓ | ✓ | ✓ | ✓ | ✓ | ✓ | ✓ | ✓ | |
| Romanos-Nanclares 2021 USA (34114021)[145] | Nurses' Health Study and Nurses' Health Study II<br>Pooled Analysis Cohorts<br>F<br>Health professionals | 2,503 /<br>175,798<br>36 years | Medical records and National Death Index | FFQ,<br>**SSB:**<br>Caffeinated and noncaffeinated colas with sugar, other (noncola) carbonated beverages with sugar, and noncarbonated sweetened beverages (e.g., punch, lemonade, fruit drink, or sugared iced tea) | Incidence, Premenopausal breast cancer | ≥1day vs <1/month servings | 0.96 (0.82 - 1.12)<br>Ptrend:0.95 | ✓ | | | ✓ | ✓ | ✓ | ✓<br>E<br>* | ✓ | ✓ | ✓ | ✓ | |
| | | | | | | Per 1 serving day | 1.02 (0.95 - 1.08) | ✓ | | | ✓ | ✓ | ✓ | ✓<br>E<br>* | ✓ | ✓ | ✓ | ✓ | |
| Chazelas 2019 France (31292122)[44] | NutriNet-Santé study<br>Prospective Cohort<br>Age:42.2 years<br>M/F | 283 /<br>79,724<br>5.1 years | self-report, medical records reviewed by physicians, health insurance database, mortality registry | 24 Hour Diet Recall,<br>**SSB**:<br>Beverages with more than 5% simple carbohydrates and 100% fruit juices (no added sugar), soft drinks (carbonated or not), syrups, 100% juice, fruit drinks, sugar sweetened hot beverages, milk-based sugar sweetened beverages, sport drinks, and energy drinks. | Incidence, Premenopausal breast cancer | Per 100 ml day | 1.26 (1.03 - 1.54) | ✓ | | ✓ | ✓ | ✓ | ✓ | ✓<br>E | D | ✓ | ✓ | ✓ | |
| Romanos-Nanclares 2019 Spain (30284064)[146] | Seguimiento Universidad de Navarra (SUN) Project<br>Prospective Cohort<br>Age:33 years<br>F<br>Spanish | 57 / 10,713<br>10 years | Medical records and National Death Index | Semi-quantitative FFQ,<br>**SSB**:<br>Sugar-sweetened beverages | Incidence, Breast cancer, premenopausal | Regular intake vs never/seldom | 1.16 (0.66 - 2.07)<br>Ptrend:0.602 | ✓ | | ✓ | ✓ | ✓ | ✓ | ✓<br>E | D | ✓ | ✓ | ✓ | |

| Author, Year Country (PMID) | Study name, type Age, sex, ethnicity, characteristics of participants | Cases/ Study size Median follow-up (years) | Case ascertainment | Exposure Assessment, detail | Outcome Sub-population | Comparison | RR (95%CI) Ptrend | Adjustments | | | | | | | | | | | |
|---|---|---|---|---|---|---|---|---|---|---|---|---|---|---|---|---|---|---|---|
| | | | | | | | | Age | Sex | Smo | Alc | Obe | Phy | Diet | Med | Mens | Repr | Oth | Skin |
| Romanos-Nanclares 2021 USA (34114021)[145] | Nurses' Health Study and Nurses' Health Study II Pooled Analysis Cohorts F Health professionals | 7,301 / 175,798 36 years | Medical records and National Death Index | FFQ, **SSB:** Caffeinated and noncaffeinated colas with sugar, other (noncola) carbonated beverages with sugar, and noncarbonated sweetened beverages (e.g., punch, lemonade, fruit drink, or sugared iced tea) | Incidence, ER+ | Per 1 serving day | 1.03 (0.98 - 1.07) | √ | | | √ | √ | √ | √ E * | √ | √ | √ | √ | |
| | | 1,693 / 175,798 36 years | | | Incidence, ER- | Per 1 serving day | 0.97 (0.89 - 1.06) | √ | | | √ | √ | √ | √ E * | √ | √ | √ | √ | |
| | | 2,673 / 175,798 36 years | | | Incidence, Luminal A breast cancer | Per 1 serving day | 1.00 (0.93 - 1.08) | √ | | | √ | √ | √ | √ E * | √ | √ | √ | √ | |
| | | 1,147 / 175,798 36 years | | | Incidence, Luminal B breast cancer | Per 1 serving day | 0.99 (0.88 - 1.12) | √ | | | √ | √ | √ | √ E * | √ | √ | √ | √ | |
| | | 250 / 175,798 36 years | | | Incidence, HER2+ breast cancer | Per 1 serving day | 0.92 (0.71 - 1.19) | √ | | | √ | √ | √ | √ E * | √ | √ | √ | √ | |
| | | 303 / 175,798 36 years | | | Incidence, Basal-like breast cancer | Per 1 serving day | 0.84 (0.67 - 1.05) | √ | | | √ | √ | √ | √ E * | √ | √ | √ | √ | |
| Nomura 2016 USA (27578546)[128] | Black Women's Health Study, 1995 Prospective Cohort Age:37.8 years F African American | 205 / 49,103 13.86 years | self-report + cancer registry | FFQ, **SSB**: Sugar-sweetened beverages | Incidence, Breast cancer ER- and PR- | 0 vs ≥ 250 g/d | 0.83 (0.44 - 1.54)[1] Ptrend:0.37 | √ | | √ | √ | √ | √ | √ E | | √ | √ | √ | |
| | | 881 / 49,103 13.86 years | | | Incidence, Breast cancer ER+ & PR+ | 0 vs ≥ 250 g/d | 0.55 (0.32 - 0.93)[1] Ptrend:0.07 | √ | | √ | √ | √ | √ | √ E | | √ | √ | √ | |
| | | 205 / 49,103 13.86 years | | | Incidence, Breast cancer ER+ or PR+ | 0 vs ≥ 250 g/d | 0.95 (0.40 - 2.23)[1] Ptrend:0.98 | √ | | √ | √ | √ | √ | √ E | | √ | √ | √ | |
| | | 166 / 49,103 13.86 years | | | Incidence, Triple negative breast cancer | 0 vs ≥ 250 g/d | 0.76 (0.27 - 2.14)[1] Ptrend:0.76 | √ | | √ | √ | √ | √ | √ E | | √ | √ | √ | |
| Ishitani 2008 USA (18852405 1 [75] | Women's Health Study Prospective Cohort Age:55 years F | 1,181 / 38,432 10 years | self-reported/death certificate/ medical records | FFQ, **Caffeinated sugar-sweetened cola**: Caffeinated cola | Incidence / Mortality, Invasive breast cancer | ≥1 can/day vs almost never | 1.17 (0.87 - 1.57) Ptrend:0.40 | √ | | √ | √ | √ | √ | √ E | √ | √ | √ | √ | |
| | | 1,170 / 38,432 10 years | | FFQ, **Decaffeinated sugar-sweetened cola**: Decaffeinated cola | | ≥1 can/week vs almost never | 0.83 (0.62 - 1.11) Ptrend:0.15 | √ | | √ | √ | √ | √ | √ E | √ | √ | √ | √ | |
| **Endometrial** | | | | | | | | | | | | | | | | | | | |
| McCullough 2022 USA (36107009)[113] | Cancer Prevention Study II Prospective Cohort F Mainly white Postmenopausal | 1,693 / 518,464 27.7 years | self-report & death certificates | Questionnaire, **SSB**: Regular colas and other nondiet soft drinks. | Mortality, Uterus/Endometrium | 2+ drinks/day vs never | 1.17 (0.95 - 1.45) Ptrend:0.136 | √ | | √ | √ | | | √ * | | √ | √ | √ | |
| | | | | | | Per 1 drink day | 1.04 (0.98 - 1.11) | √ | | √ | √ | | | √ * | | √ | √ | √ | |

| Author, Year Country (PMID) | Study name, type Age, sex, ethnicity, characteristics of participants | Cases/ Study size Median follow-up (years) | Case ascertainment | Exposure Assessment, detail | Outcome Sub-population | Comparison | RR (95%CI) Ptrend | Adjustments | | | | | | | | | | | |
|---|---|---|---|---|---|---|---|---|---|---|---|---|---|---|---|---|---|---|---|
| | | | | | | | | Age | Sex | Smo | Alc | Obe | Phy | Diet | Med | Mens | Repr | Oth | Skin |
| Arthur 2021 Canada (33220638)[38] | Canadian Study of Diet, Lifestyle and Health<br>Case Cohort<br>F | 169 / 2,351<br>15 years | Cancer registry and national death index | Self-administered Questionnaire,<br>**SSB**:<br>Coca Cola, Pepsi, other cola, soft drinks (not dietetic), orange/grapefruit juice, apple/grape juice, other fruit juices, fruit drink/lemonade and power-based drinks/iced tea | Incidence, Endometrial cancer | >7.6 vs ≤4.4 cups/week | 1.50 (1.03 - 2.17)<br>Ptrend:0.02 | ✓ | | ✓ | ✓ | ✓ | ✓ | ✓ | | ✓ | ✓ | ✓ | |
| | | 125 / 2,351<br>15 years | | | Incidence, type 1 endometrial cancer | >7.6 vs ≤4.4 cups/week | 1.67 (1.08 - 2.59)<br>Ptrend:0.015 | ✓ | | ✓ | ✓ | ✓ | ✓ | ✓ | | ✓ | ✓ | ✓ | |
| Inoue-Choi 2013 USA (24273064)[74] | Iowa Women's Health Study<br>Prospective Cohort<br>Age:61.6 years<br>F<br>Postmenopausal | 506 /<br>23,039<br>24 years | State Health Registry, National Death Index | FFQ,<br>**SSB**:<br>Sugar-sweetened Coke, Pepsi, or other colas and noncaffeinated sugar-sweetened Coke, Pepsi, or other colas; and other sugar-sweetened carbonated beverages ( 7-Up); and Hawaiian Punch, lemonade, other non-carbonated fruit drinks, and fruit juices | Incidence, Type 1 endometrial cancer | 9.9 - 72.3 vs 0 - 1.3 servings/week | 1.38 (1.03 - 1.87)<br>Ptrend:0.02 | ✓ | | ✓ | ✓ | ✓ | ✓ | ✓ | D | ✓ | ✓ | | |
| | | 86 / 23,039<br>24 years | | | Incidence, Type 2 endometrial cancer | | 1.05 (0.53- 2.06)<br>Ptrend:0.89 | ✓ | | ✓ | ✓ | ✓ | ✓ | ✓ | D | ✓ | ✓ | | |
| Hodge 2018 Australia (29463332)[70] | Melbourne Collaborative Cohort Study<br>Prospective Cohort<br>M/F<br>women | 167 /<br>21,492<br>23 years | cancer registry/death records/National Death Index | Self-administered Questionnaire,<br>**SSB**:<br>Sugar Sweetened Soft Drinks | Incidence, Endometrial Cancer | Consumption ≥1/day vs <1/month | 1.02 (0.54 - 1.91)<br>Ptrend:0.88 | ✓ | | ✓ | ✓ | | ✓ | ✓ | | | | ✓ | |
| | | | | | | Linear model | 1.04 (0.60 - 1.82) | | | ✓ | ✓ | | ✓ | ✓ | | | | ✓ | |
| **Ovary** | | | | | | | | | | | | | | | | | | | |
| McCullough 2022 USA (36107009)[113] | Cancer Prevention Study II<br>Prospective Cohort<br>F<br>Mainly white<br>Postmenopausal | 3,225 /<br>518,464<br>27.7 years | self-report & death certificates | Questionnaire,<br>**SSB**:<br>Regular colas and other nondiet soft drinks. | Mortality, Ovarian cancer | 2+ drinks/day vs never | 0.99 (0.84 - 1.17)<br>Ptrend:0.761 | ✓ | | ✓ | ✓ | | | ✓<br>* | | ✓ | ✓ | ✓ | |
| | | | | | | Per 1 drink day | 1.01 (0.97 - 1.06) | ✓ | | ✓ | ✓ | | | ✓<br>* | | ✓ | ✓ | ✓ | |
| Hodge 2018 Australia (29463332)[70] | Melbourne Collaborative Cohort Study<br>Prospective Cohort<br>M/F | 130 /<br>21,492<br>23 years | cancer registry/death records/National Death Index | Self-administered Questionnaire,<br>**SSB**:<br>Sugar Sweetened Soft Drinks | Incidence, Ovarian cancer | Consumption ≥1/day vs <1/month | 1.35 (0.71 - 2.56)<br>Ptrend:0.36 | ✓ | | ✓ | ✓ | | ✓ | ✓ | | | | ✓ | |
| | | | | | | Linear model | 1.32 (0.73 - 2.41) | | | ✓ | ✓ | | ✓ | ✓ | | | | ✓ | |
| Arthur 2021 Canada (33220638)[38] | Canadian Study of Diet, Lifestyle and Health<br>Case Cohort<br>F | 98 / 2,554<br>15 years | Cancer registry and national death index | Self-administered Questionnaire,<br>**SSB**:<br>Coca Cola, Pepsi, other cola, soft drinks (not dietetic), orange/grapefruit juice, apple/grape juice, other fruit juices, fruit drink/lemonade and power-based drinks/iced tea | Incidence, Ovarian cancer | >7.6 vs ≤4.3 cups/week | 1.63 (1.02 - 2.60)<br>Ptrend:0.022 | ✓ | | ✓ | ✓ | ✓ | ✓ | ✓ | | ✓ | ✓ | ✓ | |
| | | 72 / 2,554<br>15 years | | | Incidence, Epithelial ovarian cancer | >7.6 vs ≤4.3 cups/week | 1.51 (0.88 - 2.59)<br>Ptrend:0.084 | ✓ | | ✓ | ✓ | ✓ | ✓ | ✓ | | ✓ | ✓ | ✓ | |
| **Prostate** | | | | | | | | | | | | | | | | | | | |
| McCullough 2022 USA (36107009)[113] | Cancer Prevention Study II<br>Prospective Cohort<br>M<br>Mainly white | 9,381 /<br>416,313<br>27.7 years | self-report & death certificates | Questionnaire,<br>**SSB**:<br>Regular colas and other nondiet soft drinks. | Mortality, Prostate cancer | 2+ drinks/day vs never | 1.01 (0.93 - 1.09)<br>Ptrend:0.570 | ✓ | | ✓ | ✓ | | | ✓<br>* | | | | ✓ | |

#### Supplementary Materials

| Author, Year Country (PMID) | Study name, type Age, sex, ethnicity, characteristics of participants | Cases/ Study size Median follow-up (years) | Case ascertainment | Exposure Assessment, detail | Outcome Sub-population | Comparison | RR (95%CI) Ptrend | Adjustments: Age | Sex | Smo | Alc | Obe | Phy | Diet | Med | Mens | Repr | Oth | Skin |
|---|---|---|---|---|---|---|---|---|---|---|---|---|---|---|---|---|---|---|---|
| Mullee 2019 Denmark,France,Germany,Greece,Italy,Netherlands,Norway,Spain,Sweden,UK (31479109)[126] | European Prospective Investigation into Cancer and nutrition Prospective Cohort Age:50.8 years M/F | 907 / 130,662 16.4 years | active follow up, cancer registry, death certificate and National death index | Semi-quantitative FFQ, **SSB**: Sugar sweetened soft drinks | Mortality, Prostate cancer | Per 1 drink day | 0.99 (0.97 - 1.01) | ✓ | | ✓ | ✓ | | | ✓ * | | | | ✓ | |
| | | | | | | ≥1 vs <1 glasses /month | 1.08 (0.77 - 1.51) Ptrend:0.76 | ✓ | ✓ | ✓ | ✓ | ✓ | ✓ | ✓ E * | | ✓ | ✓ | ✓ | |
| Malik 2019 USA (30882235)[109] | Health Professionals Follow-up Study Prospective Cohort M Mostly white Health professionals | NR / 37,716 28 years | Physician confirmation | FFQ, **SSB**: Sugar-sweetened caffeinated and noncaffeinated colas, other (noncola) sugar-sweetened carbonated beverages, and noncarbonated sugar-sweetened beverages (fruit punches, lemonades, or other fruit drinks). | Mortality, Prostate cancer | Consumption ≥2/day vs <1/month | 0.77 (0.39 - 1.52) Ptrend:0.43 | ✓ | | ✓ | ✓ | ✓ | ✓ | ✓ E * | | | | ✓ | |
| | | | | | | Per 1 serving day | 1.01 (0.86 - 1.19) | ✓ | | ✓ | ✓ | ✓ | ✓ | ✓ E * | | | | ✓ | |
| Bosire 2013 USA (23408548)[42] | NIH- AARP Diet and Health Study Prospective Cohort Range:50-71 years M Retired | 428 / 293,464 8.9 years | annual linkage to the Social Security Administration Death Master File, follow-up searches of the National Death Index, Social Security Administration Death Master File, | FFQ, **SSB**: Sugar-sweetened beverages | Mortality, Prostate cancer | Higher AHEI-2010 component score | 0.99 (0.96 - 1.02) | ✓ | | ✓ | ✓ | ✓ | ✓ | ✓ E | D | | | ✓ | |
| Makarem 2018 USA (29674390)[108] | Framingham Offspring Cohort Prospective Cohort Range:26-84 years M/F | 157 / 3,184 ~10 years | cancer registry and pathology reports | FFQ, **SSB**: Sugar sweetened beverages and fruit juice. | Incidence, Prostate cancer | >10.4 vs <5.5 serving/week | 1.36 (0.88 - 2.09) | ✓ | | ✓ | ✓ | | | ✓ E | | | | | |
| Lee 2023 Korea (37536716)[97] | Health Examinees-Gem study Prospective Cohort Range:40-69 years M/F | 449 / 36,266 | Korea Central Cancer Registry +Cancer Statistics in Korea | Semi-quantitative FFQ, **SSB**: Sugar-sweetened beverages | Incidence, Prostate cancer | 0 vs >250 g/day | 1.10 (0.45 - 2.67)[1] | | | ✓ | | | | ✓ E | | | | ✓ | |
| Chazelas 2019 France (31292122)[44] | NutriNet-Santé study Prospective Cohort Age:42.2 years M/F | 291 / 21,533 5.1 years | self-report, medical records reviewed by physicians, health insurance database, mortality registry | 24 Hour Diet Recall, **SSB**: Beverages with more than 5% simple carbohydrates and 100% fruit juices (no added sugar), soft drinks (carbonated or not), syrups, 100% juice, fruit drinks, sugar sweetened hot beverages, milk-based sugar sweetened beverages, sport drinks, and energy drinks. | Incidence, Prostate cancer | Per 100 ml day | 1.11 (0.91 - 1.36) | ✓ | | ✓ | ✓ | ✓ | ✓ | ✓ E | D | | | ✓ | |
| Hodge 2018 Australia (29463332)[70] | Melbourne Collaborative Cohort Study Prospective Cohort M/F | 433 / 14,101 23 years | cancer registry/death records/National Death Index | Self-administered Questionnaire, **SSB**: Sugar Sweetened Soft Drinks | Incidence, Aggressive prostate cancer | Consumption ≥1/day vs <1/month | 1.08 (0.78 - 1.50) Ptrend:0.47 | ✓ | | ✓ | ✓ | | ✓ | ✓ | | | | ✓ | |
| | | | | | | Linear model | 1.12 (0.83 - 1.51) | | | ✓ | ✓ | | ✓ | ✓ | | | | ✓ | |

| Author, Year Country (PMID) | Study name, type Age, sex, ethnicity, characteristics of participants | Cases/ Study size Median follow-up (years) | Case ascertainment | Exposure Assessment, detail | Outcome Sub-population | Comparison | RR (95%CI) Ptrend | Adjustments | | | | | | | | | | | |
|---|---|---|---|---|---|---|---|---|---|---|---|---|---|---|---|---|---|---|---|
| | | | | | | | | Age | Sex | Smo | Alc | Obe | Phy | Diet | Med | Mens | Repr | Oth | Skin |
| Drake 2012 Sweden (23134882)[47] | Malmo Diet and Cancer, 1991<br>Prospective Cohort<br>Range:45-73 years<br>M | 513 / 8,128<br>15 years | linkage of personal identification numbers with the Swedish Cancer Register | FFQ,<br>**SSB**:<br>Sugar-sweetened beverages | Incidence, Prostate cancer | Median intake 297.8 vs 0 g/day | 1.13 (0.92 - 1.38)<br>Ptrend:0.221 | ✓ | | ✓ | ✓ | ✓ | ✓ | ✓<br>E | | | | ✓ | |
| | | 332 / 8,128<br>15 years | | | Incidence, Low-risk prostate cancer | Tertile 3 vs 0 g/day | 1.06 (0.82 - 1.37)<br>Ptrend:0.661 | ✓ | | ✓ | ✓ | ✓ | ✓ | ✓<br>E | | | | ✓ | |
| | | 176 / 8,128<br>15 years | | | Incidence, High-risk prostate cancer | Tertile 3 vs 0 g/day | 1.28 (0.92 - 1.78)<br>Ptrend:0.103 | ✓ | | ✓ | ✓ | ✓ | ✓ | ✓<br>E | | | | ✓ | |
| | | 143 / 8,128<br>15 years | | | Incidence, Symptomatic prostate cancer | Tertile 3 vs 0 g/day | 1.01 (0.68 - 1.51)<br>Ptrend:0.597 | ✓ | | ✓ | ✓ | ✓ | ✓ | ✓<br>E | | | | ✓ | |
| **Kidney** | | | | | | | | | | | | | | | | | | | |
| Leung 2021 Japan (34737349)[100] | The Japan Public Health Center-based Prospective Study<br>Prospective Cohort<br>Range:40-69 years<br>M/F<br>Japanese | 169 / 73,024<br>15.8 years | Cancer registry and death certificates and participant contact | FFQ,<br>**SSB**:<br>Beta-carotene-fortified beverages, calcium-fortified beverages, canned coffee, carbonated beverages, 100% fruit juices (apple juice and orange juice), lactic acid bacteria beverages, and vitamin-fortified beverages. | Incidence, Kidney cancer | (male: ≥254 ml/day, female ≥134 ml/day) vs non-consumers | 0.92 (0.57 - 1.47)<br>Ptrend:0.93 | ✓ | | ✓ | ✓ | ✓ | ✓ | ✓<br>E | D | | | | |
| | | 169 / 73,024<br>15.8 years | | | | Per 100 ml day | 1.02 (0.96 - 1.08) | ✓ | | ✓ | ✓ | ✓ | ✓ | ✓<br>E | D | | | | |
| | | 113 / 33,094<br>15.8 years | | | Male | Per 100 ml day | 1.01 (0.95 - 1.08) | ✓ | | ✓ | ✓ | ✓ | ✓ | ✓<br>E | D | | | | |
| | | 56 / 39,930<br>15.8 years | | | Female | Per 100 ml day | 1.07 (0.95 - 1.21) | ✓ | | ✓ | ✓ | ✓ | ✓ | ✓<br>E | D | | | | |
| Hodge 2018 Australia (29463332)[70] | Melbourne Collaborative Cohort Study<br>Prospective Cohort<br>M/F | 146 / 35,593<br>23 years | cancer registry/death records/National Death Index | Self-administered Questionnaire,<br>**SSB**:<br>Sugar Sweetened Soft Drinks | Incidence, Kidney cancer | Consumption ≥1/day vs <1/month | 1.48 (0.87 - 2.53)<br>Ptrend:0.29 | ✓ | ✓ | ✓ | ✓ | | ✓ | ✓ | | | | ✓ | |
| | | | | | | Linear model | 1.32 (0.79 - 2.19) | | ✓ | ✓ | ✓ | | ✓ | ✓ | | | | ✓ | |
| Xiang 2023 USA (37784113)[173] | Prostate, Lung, Colorectal and Ovarian Cancer Screening Trial<br>Prospective Cohort<br>Age:65.5 years<br>M/F<br>Mainly white | 446 / 101,755<br>8.8 years | self-report, family members, death certificates, medical records to confirm | Questionnaire & FFQ,<br>**SSB**:<br>Sugar-sweetened beverages | Incidence, Renal cancer | Q4 vs Q1 (≥318.19 vs ≤31.95 g/day) | 1.46 (1.10 - 1.93)<br>Ptrend:0.004 | ✓ | ✓ | ✓ | ✓ | ✓ | | ✓<br>E | D | | | ✓ | |
| Heath 2021 multi-national (33849969)[66] | European Prospective Investigation into Cancer and nutrition<br>Prospective Cohort<br>Age:52 years<br>M/F | 589 / 389,220<br>15 years | population registries or active follow up | Diet Questionnaire,<br>**SSB**:<br>carbonated/soft/isotonic drinks and diluted syrups - sugar-sweetened and artificially sweetened beverages. | Incidence, Renal cell carcinoma | Per 100 g day | 0.99 (0.94 - 1.05) | ✓ | ✓ | ✓ | ✓ | ✓ | ✓ | ✓<br>E<br>* | | | | ✓ | |
| | | 347 | | | Male | Per 100 g | 0.98 (0.91 - 1.05) | ✓ | ✓ | ✓ | ✓ | ✓ | ✓ | ✓<br>E | | | | ✓ | |

| Author, Year Country (PMID) | Study name, type Age, sex, ethnicity, characteristics of participants | Cases/ Study size Median follow-up (years) | Case ascertainment | Exposure Assessment, detail | Outcome Sub-population | Comparison | RR (95%CI) Ptrend | Adjustments | | | | | | | | | | | |
|---|---|---|---|---|---|---|---|---|---|---|---|---|---|---|---|---|---|---|---|
| | | | | | | | | Age | Sex | Smo | Alc | Obe | Phy | Diet | Med | Mens | Repr | Oth | Skin |
| | | | | | | | | | | | | | | * | | | | | |
| | | 242 | | | Female | Per 100 g | 1.03 (0.94 - 1.13) | ✓ | ✓ | ✓ | ✓ | ✓ | ✓ | ✓ E * | | | | ✓ | |
| | | 265 / 389,220 15 years | | | Mortality, Renal cell carcinoma | Per 100 g | 0.97 (0.89 - 1.07) | ✓ | ✓ | ✓ | ✓ | ✓ | ✓ | ✓ E * | | | | ✓ | |
| | | 142 | | | Male | Per 100 g | 0.92 (0.81 - 1.05) | ✓ | ✓ | ✓ | ✓ | ✓ | ✓ | ✓ E * | | | | ✓ | |
| | | 123 | | | Female | Per 100 g | 1.05 (0.93 - 1.18) | ✓ | ✓ | ✓ | ✓ | ✓ | ✓ | ✓ E * | | | | ✓ | |
| McCullough 2022 USA (36107009)[113] | Cancer Prevention Study II Prospective Cohort M/F Mainly white | 3,129 / 934,777 27.7 years | self-report & death certificates | Questionnaire, **SSB**: Regular colas and other nondiet soft drinks. | Mortality, Kidney cancer | 2+ drinks/day vs never | 1.17 (1.03 - 1.34) Ptrend:0.056 | ✓ | | ✓ | ✓ | | | ✓ * | | | | ✓ | |
| | | | | | Mortality, Kidney cancer | Per 1 drinks day | 1.06 (1.02 - 1.10) | ✓ | | ✓ | ✓ | | | ✓ * | | | | ✓ | |
| | | 1,921 | | | Male | Per 1 drinks day | 1.05 (1.01 - 1.10) | ✓ | | ✓ | ✓ | | | ✓ * | | | | ✓ | |
| | | | | | Female | Per 1 drinks day | 1.09 (1.02 - 1.16) | ✓ | | ✓ | ✓ | | | ✓ * | | | | ✓ | |
| Lee 2006 USA (16775182)[98] | Nurses' Health Study and Health Professionals Follow-up Study Pooled Analysis Cohorts Range:30-75 years M/F | 248 / 136,587 | Self-report, medical records reviewed by physician, national death index, death report by family members in follow up questionnaires | Validated FFQ, **Carbonated SSB**: sugar sweetened soda | Incidence, Renal cell cancer | Per 1 serving day | 0.95 (0.69 - 1.31) | ✓ | | ✓ | ✓ | ✓ | | ✓ E | D | | ✓ | | |
| Allen 2011 UK (21407222)[36] | The Million Women Study, 1996 Prospective Cohort Range:50-64 years F | 588 / 779,369 5.2 years | National Health Service central registers | FFQ, **Fruit drinks**: Fruit squash | Incidence, Renal cell carcinoma | ≥2 vs 0 or<1 drinks/day | 1.10 (0.84 - 1.45) | | | ✓ | | ✓ | | | | | | ✓ | |
| | | | | | | Per 1 drink day | 1.03 (0.94 - 1.14) | | | ✓ | | ✓ | | | | | | ✓ | |
| **Bladder** | | | | | | | | | | | | | | | | | | | |
| Kang 2024 USA (38931318)[82] | Multiethnic Cohort Study Prospective Cohort Range:45-75 years M/F | 843 / 84,227 19.2 years | Cancer registry and natural death index | Quantitative FFQ, **SSB**: Sugar sweetened beverages and fruit juice | Incidence, Invasive bladder cancer, male | Per 2 points increase in AHEI-2010 component score | 1.03 (0.99 - 1.07) | ✓ | | | | | | ✓ E | | | | ✓ | |
| | | 309 / 102,752 19.2 years | | | Incidence, Invasive bladder cancer, female | Per 2 points increase in AHEI-2010 component score | 0.99 (0.93 - 1.05) | ✓ | | | | | | ✓ E | | | | ✓ | |
| Chen 2023 USA (37721479)[46] | Prostate, Lung, Colorectal and Ovarian Cancer Screening Trial Prospective Cohort Range:55-74 years M/F | 378 / 99,001 11.7 years | Self-report, medical record + pathology report | Questionnaire & FFQ, **SSB**: Sugar sweetened beverages and fruit juice. | Incidence, Bladder cancer | Q4 vs Q1 (274.67 vs 564.66 g/day) | 1.02 (0.96 - 1.08) | ✓ | ✓ | ✓ | ✓ | ✓ | | ✓ E | D | | | ✓ | |

| Author, Year Country (PMID) | Study name, type Age, sex, ethnicity, characteristics of participants | Cases/ Study size Median follow-up (years) | Case ascertainment | Exposure Assessment, detail | Outcome Sub-population | Comparison | RR (95%CI) Ptrend | Adjustments<br>Age | Sex | Smo | Alc | Obe | Phy | Diet | Med | Mens | Repr | Oth | Skin |
|---|---|---|---|---|---|---|---|---|---|---|---|---|---|---|---|---|---|---|---|
| | Mainly white<br>Screening Program | | | | | | | | | | | | | | | | | | |
| Leung 2021 Japan (34737349)[100] | The Japan Public Health Center-based Prospective Study<br>Prospective Cohort<br>Range:40-69 years<br>M/F<br>Japanese | 297 / 73,024<br>15.8 years | Cancer registry and death certificates and participant contact | ,<br>**SSB**:<br>Beta-carotene-fortified beverages, calcium-fortified beverages, canned coffee, carbonated beverages, 100% fruit juices (apple juice and orange juice), lactic acid bacteria beverages, and vitamin-fortified beverages. | Incidence, Bladder cancer | (male: ≥254 ml/day, female ≥134 ml/day) vs non-consumers | 0.79 (0.56 - 1.12) Ptrend:0.33 | ✓ | | ✓ | ✓ | ✓ | ✓ | ✓ E | D | | | | |
| | | | | | | Per 100 ml day | 1.01 (0.97 - 1.06) | ✓ | | ✓ | ✓ | ✓ | ✓ | ✓ E | D | | | | |
| McCullough 2022 USA (36107009)[113] | Cancer Prevention Study II<br>Prospective Cohort<br>M/F<br>Mainly white | 3,419 / 934,777<br>27.7 years | self-report & death certificates | Questionnaire,<br>**SSB**:<br>Regular colas and other nondiet soft drinks. | Mortality, Bladder cancer | 2+ drinks/day vs never | 1.11 (0.97 - 1.27) Ptrend:0.050 | ✓ | | ✓ | ✓ | | | ✓ * | | | | ✓ | |
| | | | | | | Per 1 drink day | 1.03 (0.99 - 1.07) | ✓ | | ✓ | ✓ | | | ✓ * | | | | ✓ | |
| **Brain and Spinal cord** | | | | | | | | | | | | | | | | | | | |
| McCullough 2022 USA (36107009)[113] | Cancer Prevention Study II<br>Prospective Cohort<br>M/F<br>Mainly white | 3,402 / 934,777<br>27.7 years | self-report & death certificates | Questionnaire,<br>**SSB**:<br>Regular colas and other nondiet soft drinks. | Mortality, Brain cancer | 2+ drinks/day vs never | 0.96 (0.83 - 1.11) Ptrend:0.905 | ✓ | | ✓ | ✓ | | | ✓ * | | | | ✓ | |
| | | | | | | Per 1 drink day | 0.99 (0.95 - 1.03) | ✓ | | ✓ | ✓ | | | ✓ * | | | | ✓ | |
| **Thyroid** | | | | | | | | | | | | | | | | | | | |
| Feng 2022 UK (36508215)[53] | UK Biobank<br>Prospective Cohort<br>Age:57 years<br>M/F | 423 / 264,956<br>11.1 years | Cancer registry | Questionnaire,<br>**SSB**:<br>Sugar-sweetened beverages | Incidence, Thyroid cancer | Intake vs no intake | 1.14 (0.88 - 1.49) | ✓ | ✓ | | | | | | | | | ✓ | |
| Zamora-Ros 2023 multi-national (35907037)[180] | European Prospective Investigation into Cancer and nutrition<br>Prospective Cohort<br>Age:51.1 years<br>M/F | 495 / 450,064<br>13.9 years | Cancer and mortality registries, pathology registries, active follow up, health insurance records | Questionnaire,<br>**SSB**:<br>Sugar-sweetened beverages | Incidence, Thyroid cancer | 85.8–4201.7 ml/day vs non-consumers | 0.91 (0.62 - 1.34) Ptrend:0.86 | ✓ | ✓ | ✓ | ✓ | ✓ | ✓ | ✓ E | | ✓ | ✓ | ✓ | |
| | | | | | | Per 100 ml day | 1.06 (0.98 - 1.15) | ✓ | ✓ | ✓ | ✓ | ✓ | ✓ | ✓ E | | ✓ | ✓ | ✓ | |
| | | 403 / 450,064<br>13.9 years | | | Incidence, Papillary thyroid cancer | 85.8 – 4 to201.7 ml/day vs non-consumers | 0.96 (0.62 - 1.50) Ptrend:0.61 | ✓ | ✓ | ✓ | ✓ | ✓ | ✓ | ✓ E | | ✓ | ✓ | ✓ | |
| | | | | | | Per 100 ml day | 1.08 (0.99 - 1.17) | ✓ | ✓ | ✓ | ✓ | ✓ | ✓ | ✓ E | | ✓ | ✓ | ✓ | |
| | | 73 / 450,064<br>13.9 years | | | Incidence, Follicular thyroid cancer | 85.8 – 4 to201.7 ml/day vs non-consumers | 0.81 (0.34 - 1.92) Ptrend:0.80 | ✓ | ✓ | ✓ | ✓ | ✓ | ✓ | ✓ E | | ✓ | ✓ | ✓ | |
| | | | | | | Per 100 ml day | 0.93 (0.72 - 1.21) | ✓ | ✓ | ✓ | ✓ | ✓ | ✓ | ✓ E | | ✓ | ✓ | ✓ | |
| **Lymphoid and hematopoietic** | | | | | | | | | | | | | | | | | | | |

| Author, Year Country (PMID) | Study name, type Age, sex, ethnicity, characteristics of participants | Cases/ Study size Median follow-up (years) | Case ascertainment | Exposure Assessment, detail | Outcome Sub-population | Comparison | RR (95%CI) Ptrend | Adjustments: Age | Sex | Smo | Alc | Obe | Phy | Diet | Med | Mens | Repr | Oth | Skin |
|---|---|---|---|---|---|---|---|---|---|---|---|---|---|---|---|---|---|---|---|
| McCullough 2022 USA (36107009)[113] | Cancer Prevention Study II<br>Prospective Cohort<br>M/F<br>Mainly white | 6,319 / 934,777<br>27.7 years | self-report & death certificates | Questionnaire,<br>**SSB**:<br>Regular colas and other nondiet soft drinks. | Mortality, Leukaemia | 2+ drinks/day vs never | 1.04 (0.94 - 1.15)<br>Ptrend:0.375 | ✓ | | ✓ | ✓ | | | ✓<br>* | | | | ✓ | |
| | | | | | | Per 1 drink day | 1.00 (0.97 - 1.03) | ✓ | | ✓ | ✓ | ✓ | | ✓<br>* | | | | ✓ | |
| | | 3,511 / 934,777<br>27.7 years | | | Mortality, multiple Myeloma | 2+ drinks/day vs never | 0.97 (0.84 - 1.18)<br>Ptrend:0.581 | ✓ | | ✓ | ✓ | | | ✓<br>* | | | | ✓ | |
| | | | | | | Per 1 drink day | 1.00 (0.96 - 1.04) | ✓ | | ✓ | ✓ | | | ✓<br>* | | | | ✓ | |
| | | 6,600 / 934,777<br>27.7 years | | | Mortality, Non-Hodgkin Lymphoma | 2+ drinks/day vs never | 1.16 (1.05 - 1.28)<br>Ptrend:0.001 | ✓ | | ✓ | ✓ | | | ✓<br>* | | | | ✓ | |
| | | | | | | Per 1 drink day | 1.04 (1.02 - 1.07) | ✓ | | ✓ | ✓ | | | ✓<br>* | | | | ✓ | |
| Schernhammer 2012 USA (23097267)[150] | Nurses' Health Study + Health Professionals Follow-up Study<br>Pooled Analysis Cohorts<br>M/F<br>Health professionals | 186 / 47,810<br>22 years | medical records, cancer registries, national death index | Semi-quantitative FFQ,<br>**Carbonated SSB**:<br>sugar-sweetened soda; Caffeine-free or caffeinated colas and other soda | Incidence, Leukaemia, male | ≥1 serving/day vs none | 0.92 (0.42 - 2.02)<br>Ptrend:0.61 | ✓ | | ✓ | ✓ | ✓ | ✓ | ✓<br>E<br>* | | | | ✓ | |
| | | 153 / 77,218<br>22 years | | | Incidence, Leukaemia, female | | 1.39 (0.47 - 4.07)<br>Ptrend:0.21 | ✓ | | ✓ | ✓ | ✓ | ✓ | ✓<br>E<br>* | | ✓ | ✓ | ✓ | |
| | | 131 / 47,810<br>22 years | | | Incidence, multiple Myeloma, male | | 1.76 (0.77 - 4.03)<br>Ptrend:0.37 | ✓ | | ✓ | ✓ | ✓ | ✓ | ✓<br>E<br>* | | | | ✓ | |
| | | 154 / 77,218<br>22 years | | | Incidence, multiple Myeloma, female | | 1.07 (0.36 - 3.16)<br>Ptrend:0.58 | ✓ | | ✓ | ✓ | ✓ | ✓ | ✓<br>E<br>* | | ✓ | ✓ | ✓ | |
| | | 571 / 47,810<br>22 years | | | Incidence, Non-Hodgkin Lymphoma, male | | 1.66 (1.10 - 2.51)<br>Ptrend:0.03 | ✓ | | ✓ | ✓ | ✓ | ✓ | ✓<br>E<br>* | | | | ✓ | |
| | | 753 / 77,218<br>22 years | | | Incidence, Non-Hodgkin Lymphoma, female | | 1.01 (0.63 - 1.62)<br>Ptrend:0.59 | ✓ | | ✓ | ✓ | ✓ | ✓ | ✓<br>E<br>* | | ✓ | ✓ | ✓ | |
| McCullough 2014 USA (25342696)[114] | Cancer Prevention Study II<br>Prospective Cohort<br>Age:69 years<br>M/F | 354 / 43,350<br>10 years | medical records and cancer registries | FFQ,<br>**Carbonated SSB**:<br>Sugar-sweetened carbonated beverage | Incidence, Non-Hodgkin Lymphoma, male | high past and high current intake vs no past and no current intake | 1.19 (0.64 - 2.22) | ✓ | ✓ | ✓ | | ✓ | | ✓<br>E<br>* | D | | | | |
| | | 300 / 57,092<br>10 years | | | Incidence, Non-Hodgkin Lymphoma, female | | 2.29 (1.21 - 4.32) | ✓ | ✓ | ✓ | | ✓ | | ✓<br>E<br>* | D | | | | |

**Age**: age; **Alc**: alcohol; **Diet**: dietary factors and energy intake (E), or * adjustment for other soft drinks not under investigation; **Med**: medical-related factors (e.g. infections, diseases in the breast, digestive tract, gallbladder, liver, or pancreas, cancer screening, precancerous conditions, or pre-existing skin cancer) or diabetes (D); **Mens**: menstrual characteristics (e.g. age at menarche, menopausal status/duration, age at menopause, or type of menopause); **Obe**: obesity (adiposity measures); **Oth**: others (e.g. family history/genetic factor, race/ethnicity, ancestry, socioeconomic factors, or marriage); **Phy**: physical activity; **Repr**: reproductive or hormonal factors (e.g. parity, age at first/last birth, postmenopausal hormone use, or oral contraceptive use); **Sex**: sex; **Skin**: skin-related factors (e.g. UV light/sun exposure, sunburns, time spent outdoors, sunscreen use, indoor tanning, moles, fair skin, red/blond hair, blue/green eyes, or freckles); **Smo**: smoking**. Abbreviations**: AHEI, Alternative healthy Eating Index; DASH, dietary Approaches to Stop Hypertension; SD, standard deviation; HEI, Healthy Eating Index.[1] Reported association estimates have been converted using the Hamling method for the analyses.

#### Supplementary Table 12. Descriptive table of the included observational studies of artificially sweetened beverages and cancer incidence and mortality.

| Author, Year Country (PMID) | Study name, type Age, sex, ethnicity, characteristics of participants | Cases/ Study size Median follow-up (years) | Case ascertainment | Exposure Assessment, detail | Outcome Sub-population | Comparison | RR (95%CI) Ptrend | Adjustments: Age | Sex | Smo | Alc | Obe | Phy | Diet | Med | Mens | Repr | Oth | Skin |
|---|---|---|---|---|---|---|---|---|---|---|---|---|---|---|---|---|---|---|---|
| **Mouth Cancer** | | | | | | | | | | | | | | | | | | | |
| McCullough 2022 USA (36107009)[113] | Cancer Prevention Study II Prospective Cohort M/F Mainly white | 1,852 / 31,691 27.7 years | self-report & death certificates | Questionnaire, **ASB**: Diet soda or diet iced teas | Mortality, Larynx/oral cavity/pharynx | 2+ drinks/day vs never | 1.13 (0.94 - 1.37) Ptrend:0.107 | ✓ | | ✓ | ✓ | ✓ | | ✓ * | | | | ✓ | |
| | | 1,852 | | | | Per 1 drink day | 1.04 (0.99 - 1.09) | ✓ | | ✓ | ✓ | ✓ | | ✓ * | | | | ✓ | |
| **Oesophageal cancer** | | | | | | | | | | | | | | | | | | | |
| McCullough 2022 USA (36107009)[113] | Cancer Prevention Study II Prospective Cohort M/F Mainly white | 2,727 / 934,777 27.7 years | self-report & death certificates | Questionnaire, **ASB**: Diet soda or diet iced teas | Mortality, Oesophageal cancer | 2+ drinks/day vs never | 0.92 (0.78 - 1.08) Ptrend:0.155 | ✓ | | ✓ | ✓ | ✓ | | ✓ * | | | | ✓ | |
| | | 2,727 | | | | Per 1 drink day | 0.99 (0.94 - 1.03) | ✓ | | ✓ | ✓ | ✓ | | ✓ * | | | | ✓ | |
| **Stomach cancer** | | | | | | | | | | | | | | | | | | | |
| McCullough 2022 USA (36107009)[113] | Cancer Prevention Study II Prospective Cohort M/F Mainly white | 2,727 / 934,777 27.7 years | self-report & death certificates | Questionnaire, **ASB**: Diet soda or diet iced teas | Mortality, Stomach cancer | 2+ drinks/day vs never | 0.92 (0.78 - 1.08) Ptrend:0.155 | ✓ | | ✓ | ✓ | ✓ | | ✓ * | | | | ✓ | |
| | | 2,727 | | | | Per 1 drink day | 0.99 (0.94 - 1.03) | ✓ | | ✓ | ✓ | ✓ | | ✓ * | | | | ✓ | |
| Hodge 2018 Australia (29463332)[70] | Melbourne Collaborative Cohort Study Prospective Cohort M/F Not reported Excluded participants with diabetes at baseline | 165 / 35,593 23 years | cancer registry/death records/National Death Index | Self-administered Questionnaire, **ASB**: Artificially sweetened soft drinks | Incidence, Gastric cardia adenocarcinoma | ≥1/day vs <1/month serving/month | 1.03 (0.53 - 1.98) Ptrend:0.46 | ✓ | ✓ | ✓ | ✓ | ✓ | ✓ | ✓ * | | | | ✓ | |
| | | 165 | | | Incidence, Gastric cardia adenocarcinoma | Linear model | 1.24 (0.70 - 2.18) | ✓ | ✓ | ✓ | ✓ | ✓ | ✓ | ✓ * | | | | ✓ | |
| **Colorectal cancer** | | | | | | | | | | | | | | | | | | | |
| McCullough 2022 USA (36107009)[113] | Cancer Prevention Study II Prospective Cohort M/F Mainly white | 324 / 934,777 27.7 years | self-report & death certificates | Questionnaire, **ASB**: Diet soda or diet iced teas. | Mortality, Small intestinal cancer | 2+ drinks/day vs never | 1.28 (0.85 - 1.93) Ptrend:0.244 | ✓ | | ✓ | ✓ | ✓ | | ✓ * | | | | ✓ | |
| | | 324 | | | | Per 1 drinks day | 1.11 (1.00 - 1.22) | ✓ | | ✓ | ✓ | ✓ | | ✓ * | | | | ✓ | |
| Jin 2023 UK (38004195)[78] | UK Biobank Prospective Cohort Age:55.87 years M/F Mainly white | 1,466 / 118,210 12.8 years | hospital records, cancer registry, death registry | 24 Hour Diet Recall, **ASB**: low/non-sugar sugar-sweetened beverages | Incidence, Colorectal cancer | Per 1 SD day | 0.99 (0.94 - 1.05) | ✓ | ✓ | ✓ | | ✓ | ✓ | ✓ E | ✓ D | | | ✓ | |
| Wang 2022 USA (38752573)[168] | Nurse's Health Study, Nurse's Health Study II, Health Professionals Follow-up Study Pooled Analysis Cohorts M/F Mainly white | 1,922 / 150,907 26 years | self-report, blinded physicians reviewed medical records & pathology report, next of kin, national death index. | Semi-quantitative FFQ **ASB**: Caffeine-free Coke; Coke/Pepsi/Cola; dairy coffee drink; Hawaiian punch; Low-calorie soda, caffeine-free; low-calorie soda, Pepsi, 7-up; other carbon beverage; other low-calorie Cola with caffeine. | Incidence, Colorectal cancer, female | Q5 vs Q1 | 1.07 (0.91 - 1.25) Ptrend:0.78 | ✓ | | ✓ | ✓ | | ✓ | ✓ E | ✓ | ✓ | ✓ | ✓ | |

| Author, Year Country (PMID) | Study name, type Age, sex, ethnicity, characteristics of participants | Cases/ Study size Median follow-up (years) | Case ascertainment | Exposure Assessment, detail | Outcome Sub-population | Comparison | RR (95%CI) Ptrend | Adjustments: Age | Sex | Smo | Alc | Obe | Phy | Diet | Med | Mens | Repr | Oth | Skin |
|---|---|---|---|---|---|---|---|---|---|---|---|---|---|---|---|---|---|---|---|
| | | | | | Female | Per 1 serving/day | 1.01 (0.95 - 1.07) | ✓ | | ✓ | ✓ | | ✓ | ✓ E | ✓ | ✓ | ✓ | ✓ | |
| | | 1,294 / 46,341 26 years | | | Male | Q5 vs Q1 | 1.14 (0.95 - 1.38) Ptrend:0.31 | ✓ | | ✓ | ✓ | | ✓ | ✓ E | ✓ | | | ✓ | |
| | | | | | Male | Per 1 serving day | 1.06 (0.95 - 1.19) | ✓ | | ✓ | ✓ | | ✓ | ✓ E | ✓ | | | ✓ | |
| Chazelas 2019 France (31292122)[44] | NutriNet-Santé study Prospective Cohort Age:42.2 years M/F | 166 / 101,257 5.1 years | self-report, medical records reviewed by physicians, health insurance database, mortality registry | 24 Hour Diet Recall, **ASB**: Beverages containing non-nutritive sweeteners, such as diet soft drinks, sugar-free syrups, and diet milk-based beverages. | Incidence, Colorectal cancer | Per 10 ml day | 1.06 (0.77 - 1.48) | ✓ | ✓ | ✓ | ✓ | ✓ | ✓ | ✓ E | D | | | ✓ | |
| Hodge 2018 Australia (29463332)[70] | Melbourne Collaborative Cohort Study Prospective Cohort M/F Not reported Excluded participants with diabetes at baseline | 1,055 / 35,593 23 years | cancer registry/death records/National Death Index | Self-administered Questionnaire, **ASB**: Artificially Sweetened Soft Drinks | Incidence, Colorectal cancer | ≥1/day vs <1/month | 0.79 (0.60 - 1.06) Ptrend:0.46 | ✓ | ✓ | ✓ | ✓ | ✓ | ✓ | ✓ * | | | | ✓ | |
| | | 1,055 | | | | Linear model | 0.91 (0.71 - 1.17) | ✓ | ✓ | ✓ | ✓ | ✓ | ✓ | ✓ * | | | | ✓ | |
| McCullough 2022 USA (36107009)[113] | Cancer Prevention Study II Prospective Cohort M/F Mainly white | 13,752 / 934,777 27.7 years | self-report & death certificates | Questionnaire, **ASB**: Diet soda or diet iced teas. | Mortality, Colorectal cancer | 2+ drinks/day vs never | 0.92 (0.86 - 0.99) Ptrend:0.034 | ✓ | | ✓ | ✓ | ✓ | | ✓ * | | | | ✓ | |
| | | 13,752 | | | | Per 1 drinks day | 0.99 (0.97 - 1.01) | ✓ | | ✓ | ✓ | ✓ | | ✓ * | | | | ✓ | |
| Mullee 2019 Denmark,France, Germany,Greece ,Italy,Netherlands ,Norway,Spain,S weden,UK (31479109)[126] | European Prospective Investigation into Cancer and nutrition Prospective Cohort Age:50.8 years M/F | 2,095 / 451,743 16.4 years | active follow up, cancer registry, death certificate and National death index | Semi-quantitative FFQ, **ASB**: Artificially Sweetened Soft Drinks | Mortality, Colorectal cancer | ≥1 glass/day vs <1 glass/month | 1.22 (0.91 - 1.64) Ptrend:0.21 | ✓ | ✓ | ✓ | ✓ | ✓ | ✓ | ✓ * | | ✓ | ✓ | ✓ | |
| Hur 2021 USA (33958435)[72] | Nurses' Health Study II Prospective Cohort F nurses | 109 / 95,464 24 years | Medical records and National Death Index | Semi-quantitative FFQ, **ASB**: Low-calorie carbonated beverages. | Incidence, Early-onset colorectal cancer | ≥2 servings/d vs <1 serving/week | 0.83 (0.47 - 1.47) Ptrend:0.24 | | | ✓ | ✓ | ✓ | ✓ | ✓ E * | ✓ | ✓ | ✓ | ✓ | |
| | | 109 | | | | Per 1 serving day | 0.95 (0.85 - 1.06) | | | ✓ | ✓ | ✓ | ✓ | ✓ * | ✓ | ✓ | ✓ | ✓ | |
| Jin 2023 UK (38004195)[78] | UK Biobank Prospective Cohort Age:55.87 years M/F Mainly white | 842 / 118,210 12.8 years | hospital records, cancer registry, death registry | 24 Hour Diet Recall, **ASB**: low/non-sugar sugar-sweetened beverages | Incidence, Colon cancer | Per 1 SD day | 0.98 (0.91 - 1.06) | ✓ | ✓ | ✓ | | ✓ | ✓ | ✓ | ✓ D | | | ✓ | |
| Malik 2019 USA (30882235)[109] | Nurse's Health Study, Health Professionals Follow-up Study Pooled Analysis Cohorts M/F Mostly white Health professionals | / 118,363 34 years | Physician confirmation | FFQ, ASB: Caffeinated, noncaffeinated, and noncarbonated low-calorie or diet beverages. | Mortality, Colon cancer | ≥2/d vs <1 serving/month | 1.01 (0.77 - 1.31) Ptrend:0.69 | ✓ | | ✓ | ✓ | ✓ | ✓ | ✓ E * | | | ✓ | ✓ | |
| | | / 118,363 34 years | | | Mortality, Colon cancer | Per 1 serving day | 1.00 (0.91 - 1.07) | ✓ | | ✓ | ✓ | ✓ | ✓ | ✓ E * | | | ✓ | ✓ | |

| Author, Year Country (PMID) | Study name, type Age, sex, ethnicity, characteristics of participants | Cases/ Study size Median follow-up (years) | Case ascertainment | Exposure Assessment, detail | Outcome Sub-population | Comparison | RR (95%CI) Ptrend | Adjustments Age | Sex | Smo | Alc | Obe | Phy | Diet | Med | Mens | Repr | Oth | Skin |
|---|---|---|---|---|---|---|---|---|---|---|---|---|---|---|---|---|---|---|---|
| Jin 2023 UK (38004195)[78] | UK Biobank<br>Prospective Cohort<br>Age:55.87 years<br>M/F<br>Mainly white | 359 / 118,210<br>12.8 years | hospital records, cancer registry, death registry | 24 Hour Diet Recall,<br>**ASB**:<br>low/non-sugar sugar-sweetened beverages | Incidence, Rectal cancer | Per 1 SD day | 0.97 (0.86 - 1.10) | ✓ | ✓ | ✓ | | ✓ | ✓ | ✓ E | ✓ D | | | ✓ | |
| Liver cancer | | | | | | | | | | | | | | | | | | | |
| McCullough 2022 USA (36107009)[113] | Cancer Prevention Study II<br>Prospective Cohort<br>M/F<br>Mainly white | 2,722 / 934,777<br>27.7 years | self-report & death certificates | Questionnaire,<br>**ASB**:<br>Diet soda or diet iced teas. | Mortality, Liver cancer | 2+ drinks/day vs never | 0.95 (0.81 - 1.10)<br>Ptrend:0.578 | ✓ | | ✓ | ✓ | ✓ | | ✓ * | | | | ✓ | |
| | | 2,722 | | | Mortality, Liver cancer | Per 1 drinks day | 0.98 (0.94 - 1.02) | ✓ | | ✓ | ✓ | ✓ | | ✓ * | | | | ✓ | |
| Jones 2022 USA (35728406)[79] | NIH-AARP + Prostate, Lung, Colorectal and Ovarian Cancer Screening Trial<br>Pooled Analysis Cohorts<br>M/F<br>Mainly white | 587 / 506,389 | NIH-AARP: cancer reg, PLCO: self-report + medical record/cancer reg | Questionnaire,<br>**ASB**:<br>soda and fruit punch. | Incidence, Liver cancer, no diabetes & <=12 yrs follow-up | Once per day | 1.01 (0.92 - 1.11) | ✓ | ✓ | ✓ | ✓ | ✓ | | ✓ E | | | | ✓ | |
| | | 252 / 506,389 | | | >12 yrs follow-up | Once per day | 0.99 (0.86 – 1.15) | ✓ | ✓ | ✓ | ✓ | ✓ | | ✓ E | | | | ✓ | |
| Stepien 2016 multi-national (25528243)[159] | European Prospective Investigation into Cancer and Nutrition study cohort<br>Prospective Cohort<br>Mainly white | 101 / 477,206<br>11.4 years | cancer registries | ,<br>Semi-quantitative FFQ:<br>Artificially sweetened soft drinks | Incidence, HCC | Per 1 serving wk | 1.06 (1.03 - 1.09) | | | ✓ | ✓ | ✓ | ✓ | ✓ E | D | | | ✓ | |
| Zhao 2024 USA (39226717)[187] | Southern Community Cohort Study<br>Prospective Cohort<br>Range:40-79 years<br>M/F<br>White, black, other | 453 / 73,119<br>13.9 years | National death index and cancer registry | Semi-quantitative FFQ,<br>**ASB**:<br>Diet soda: diet Coke, or other diet drinks. | Incidence, Liver cancer | ≥median vs non-consumer | 0.83 (0.51 - 1.35)<br>Ptrend:0.36 | ✓ | ✓ | ✓ | ✓ | ✓ | ✓ | ✓ E | ✓ D | | | ✓ | |
| | | | | | | Once per day | 0.98 (0.83 - 1.15) | ✓ | ✓ | ✓ | ✓ | ✓ | | ✓ E | | | | ✓ | |
| Zhao 2023 USA (37552302)[186] | Women's Health Initiative Prospective Cohort<br>Range:50-79 years<br>F<br>Mixed<br>Postmenopausal | 207 / 98,786<br>20.9 | self-administered questionnaire, verified by pathology reports, physician/discharge, operative/radiology reports, tumour registry. | FFQ,<br>**SSB**:<br>Sugar-sweetened beverages | Incidence, Liver cancer | ≥1 serving/day vs never - ≤3 servings/month, adjusted for BMI | 1.85 (1.16 - 2.96)<br>Ptrend:0.01 | ✓ | | ✓ | ✓ | ✓ | ✓ | ✓ E | D | | ✓ | ✓ | |
| | | | | | | Not adjusted for BMI | 1.77 (1.11 - 2.82)<br>Ptrend:0.02 | ✓ | | ✓ | ✓ | | ✓ | ✓ E | | | ✓ | ✓ | |
| Jones 2022 USA (35728406)[79] | | 587 / 506,389 | | Questionnaire,<br>**Artificially sweetened fruit drinks**:<br>diet fruit punch | Incidence, Liver cancer, no diabetes & <=12 yrs follow-up | Once per day | 1.06 (0.80 - 1.41) | ✓ | ✓ | ✓ | ✓ | ✓ | | ✓ E | | | | ✓ | |
| | | 252 / 506,389 | | | >12 yrs follow-up | Once per day | 1.11 (0.74 - 1.65) | ✓ | ✓ | ✓ | ✓ | ✓ | | ✓ E | | | | ✓ | |
| Gallbladder cancer | | | | | | | | | | | | | | | | | | | |
| McCullough 2022 USA (36107009)[113] | Cancer Prevention Study II<br>Prospective Cohort | 1,024 / 934,777<br>27.7 years | self-report & death certificates | Questionnaire,<br>**ASB**:<br>Diet soda or diet iced teas. | Mortality, Gallbladder cancer | 2+ drinks/day vs never | 1.15 (0.90 - 1.46)<br>Ptrend:0.105 | ✓ | | ✓ | ✓ | ✓ | | ✓ E * | | | | ✓ | |

| Author, Year Country (PMID) | Study name, type Age, sex, ethnicity, characteristics of participants | Cases/ Study size Median follow-up (years) | Case ascertainment | Exposure Assessment, detail | Outcome Sub-population | Comparison | RR (95%CI) Ptrend | Adjustments Age | Sex | Smo | Alc | Obe | Phy | Diet | Med | Mens | Repr | Oth | Skin |
|---|---|---|---|---|---|---|---|---|---|---|---|---|---|---|---|---|---|---|---|
| | M/F<br>Mainly white | | | | | | | | | | | | | | | | | | |
| | | 1,024 | | | | Per 1 drink day | 1.05 (0.98 - 1.11) | ✓ | | ✓ | ✓ | ✓ | | ✓ E * | | | | ✓ | |
| Pancreatic cancer | | | | | | | | | | | | | | | | | | | |
| McCullough 2022 USA (36107009)[113] | Cancer Prevention Study II<br>Prospective Cohort<br>M/F<br>Mainly white | 9,196 / 934,777<br>27.7 years | self-report & death certificates | Questionnaire, **ASB**: Diet soda or diet iced teas | Mortality, Pancreas cancer | 2+ drinks/day vs never | 1.11 (1.02 - 1.20)<br>Ptrend:0.008 | ✓ | | ✓ | ✓ | ✓ | | ✓ E * | | | | ✓ | |
| | | 9,196 | | | | Per 1 drink day | 1.03 (1.01 - 1.05) | ✓ | | ✓ | ✓ | ✓ | | ✓ * | | | | ✓ | |
| | | 4,339 | | | Male | Per 1 drink day | 1.04 (1.00 - 1.08) | ✓ | | ✓ | ✓ | ✓ | | ✓ * | | | | ✓ | |
| | | 4,857 | | | Female | Per 1 drink day | 1.02 (0.99 - 1.05) | ✓ | | ✓ | ✓ | ✓ | | ✓ * | | | | ✓ | |
| | | 4,234 | | | Normal weight (BMI 18.5 to <25 kg/m$^2$) | Per 1 drink day | 1.00 (0.97 - 1.04) | ✓ | | ✓ | ✓ | | | ✓ * | | | | ✓ | |
| | | 3,625 | | | Overweight (BMI 25 to <30 kg/m$^2$) | Per 1 drink day | 1.05 (1.01 - 1.08) | ✓ | | ✓ | ✓ | | | ✓ * | | | | ✓ | |
| | | 1,022 | | | Obese (>= 30 kg/m$^2$) | Per 1 drink day | 1.01 (0.95 - 1.07) | ✓ | | ✓ | ✓ | | | ✓ * | | | | ✓ | |
| | | 3,955 | | | Never smokers | Per 1 drink day | 1.05 (1.02 - 1.09) | ✓ | | | ✓ | ✓ | | ✓ * | | | | ✓ | |
| Navarrete-Muñoz 2016 multi-national (27510540)[127] | European Prospective Investigation into Cancer and nutrition<br>Prospective Cohort<br>Age:51 years<br>M/F<br>Mostly white<br>generally healthy adults | 686 / 477,199<br>11.6 years | cancer and pathology registry, active follow up, health insurance record, mortality registry and contact of participants or next-of-kin | Diet Questionnaire, **ASB**: Artificially sweetened beverages. | Incidence / Mortality, Pancreas cancer | >92.2 vs 0.1-2 g/day | 1.07 (0.67 - 1.73)<br>Ptrend:0.55 | ✓ | ✓ | ✓ | ✓ | | ✓ | ✓ * | | | | ✓ | |
| | | 686 | | | | Per 336 g day | 1.13 (0.94 - 1.36) | ✓ | ✓ | ✓ | ✓ | | ✓ | ✓ * | | | | ✓ | |
| | | 313 | | | Male | Per 336 g day | 0.69 (0.44 - 1.09) | ✓ | ✓ | ✓ | ✓ | | ✓ | ✓ * | | | | ✓ | |
| | | 373 | | | Female | Per 336 g day | 1.25 (1.03 - 1.52) | ✓ | ✓ | ✓ | ✓ | | ✓ | ✓ * | | | | ✓ | |
| | | 297 | | | BMI <25 kg/m$^2$ | Per 336 g day | 0.83 (0.48 - 1.43) | ✓ | ✓ | ✓ | ✓ | | ✓ | ✓ * | | | | ✓ | |
| | | 389 | | | BMI >=25 kg/m$^2$ | Per 336 g day | 1.15 (0.94 - 1.41) | ✓ | ✓ | ✓ | ✓ | | ✓ | ✓ * | | | | ✓ | |
| | | 213 | | | Current smoker | Per 336 g day | 1.06 (0.76 - 1.46) | ✓ | ✓ | | ✓ | | ✓ | ✓ * | | | | ✓ | |
| | | 257 | | | Never smoker | Per 336 g day | 0.95 (0.61 - 1.48) | ✓ | ✓ | | ✓ | | ✓ | ✓ * | | | | ✓ | |
| Bao 2008 USA (18689380)[39] | NIH- AARP Diet and Health Study<br>Prospective Cohort<br>Age:62 years<br>M/F | 1,014 / 487,922<br>7.2 years | Cancer registry and national death index | Dietary Recall, **ASB**: Diet soft drinks. | Incidence / Mortality, Pancreatic Adenocarcinoma | Median intake 816.9 vs 0 g/day | 1.25 (0.94 - 1.66)<br>Ptrend:0.19 | ✓ | ✓ | ✓ | ✓ | ✓ | ✓ | ✓ E * | | | | ✓ | |

| Author, Year Country (PMID) | Study name, type Age, sex, ethnicity, characteristics of participants | Cases/ Study size Median follow-up (years) | Case ascertainment | Exposure Assessment, detail | Outcome Sub-population | Comparison | RR (95%CI) Ptrend | Adjustments: Age | Sex | Smo | Alc | Obe | Phy | Diet | Med | Mens | Repr | Oth | Skin |
|---|---|---|---|---|---|---|---|---|---|---|---|---|---|---|---|---|---|---|---|
| | Mostly white<br>Retired | | | | | | | | | | | | | | | | | | |
| | | 231 | | | BMI >=30 kg/m$^2$ | Median intake 816.9 vs 0 g/day | 0.89 (0.51 - 1.53)<br>Ptrend:0.59 | ✓ | ✓ | ✓ | ✓ | ✓ | ✓ | ✓ E * | | | | ✓ | |
| | | 423 | | | Current smokers or quit <10yrs ago | Median intake 816.9 vs 0 g/day | 0.92 (0.60 - 1.40)<br>Ptrend:0.45 | ✓ | ✓ | ✓ | ✓ | ✓ | ✓ | ✓ E * | | | | ✓ | |
| | | 775 | | | Never smoker or quit 10+ yrs ago | Median intake 816.9 vs 0 g/day | 1.25 (0.92 - 1.71)<br>Ptrend:0.32 | ✓ | ✓ | ✓ | ✓ | ✓ | ✓ | ✓ E * | | | | ✓ | |
| Schernhammer 2005 USA (16172216)[151] | Health Professionals Follow-up Study<br>Prospective Cohort<br>Range:40-75 years<br>M<br>Not reported<br>Health professionals | 174 / 49,364<br>14 years | Death register/ subject or family | FFQ,<br>**Carbonated ASB**:<br>Carbonated diet soft drinks | Incidence / Mortality, Pancreatic cancer, Male | >3 servings/week <1 servings/month | 0.89 (0.60 - 1.33)<br>Ptrend:0.52 | ✓ | | ✓ | | ✓ | ✓ | ✓ E * | D | | | | |
| | | 205 / 88,794<br>20 years | | | Female | >3 servings/week <1 servings/month | 1.12 (0.79 - 1.59)<br>Ptrend:0.64 | ✓ | | ✓ | | ✓ | ✓ | ✓ E * | D | | | | |
| | | 119 | | | BMI < 25 kg/m$^2$ | >3 servings/week <1 servings/month | 1.02 (0.64 - 1.62)<br>Ptrend:0.93 | ✓ | | ✓ | | | ✓ | ✓ E * | D | | | | |
| | | 83 | | | BMI ≥ 25 kg/m$^2$ | >3 servings/week <1 servings/month | 1.32 (0.75 - 2.33)<br>Ptrend:0.62 | ✓ | | ✓ | | | ✓ | ✓ E * | D | | | | |
| Schernhammer 2005 USA (16172216)[151] | Health Professionals Follow-up Study<br>Prospective Cohort<br>Range:40-75 years<br>M<br>Not reported<br>Health professionals | 174 / 49,364<br>14 years | Death register/ subject or family | FFQ,<br>**Artificially sweetened cola**:<br>Diet cola | Incidence / Mortality, Pancreatic cancer, Male | >3 servings/week <1 servings/month | 0.85 (0.54 - 1.32)<br>Ptrend:0.38 | ✓ | | ✓ | | ✓ | ✓ | ✓ E * | D | | | | |
| | | 205 / 88,794<br>20 years | | | Female | >3 servings/week <1 servings/month | 0.83 (0.43 - 1.59)<br>Ptrend:0.96 | ✓ | | ✓ | | ✓ | ✓ | ✓ E * | D | | | | |
| **Lung cancer** | | | | | | | | | | | | | | | | | | | |
| You 2022 USA (36424907)[177] | Prostate, Lung, Colorectal and Ovarian Cancer Screening Trial<br>Prospective Cohort<br>Range:55-74 years<br>M/F<br>Mainly white<br>Screening Program | 1,517 / 154,897<br>8.9 years | chest radiography, follow-up, cancer & death registry, physician | FFQ,<br>**No intake**:<br>No intake of soft drinks, soda, and pop | Incidence, Lung cancer | Intake vs no intake | 0.79 (0.64 - 0.97) | ✓ | ✓ | ✓ | ✓ | ✓ | ✓ | ✓ E | | | | ✓ | |
| McCullough 2022 USA (36107009)[113] | Cancer Prevention Study II<br>Prospective Cohort<br>M/F<br>Mainly white | 34,381 / 934,777<br>27.7 years | self-report & death certificates | Questionnaire,<br>**ASB**:<br>Diet soda or diet iced teas. | Mortality, Lung cancer | 2+ drinks/day vs never | 0.90 (0.86 - 0.95)<br>Ptrend:<0.0001 | ✓ | | ✓ | ✓ | ✓ | | ✓ * | | | | ✓ | |

| Author, Year Country (PMID) | Study name, type Age, sex, ethnicity, characteristics of participants | Cases/ Study size Median follow-up (years) | Case ascertainment | Exposure Assessment, detail | Outcome Sub-population | Comparison | RR (95%CI) Ptrend | Adjustments Age | Sex | Smo | Alc | Obe | Phy | Diet | Med | Mens | Repr | Oth | Skin |
|---|---|---|---|---|---|---|---|---|---|---|---|---|---|---|---|---|---|---|---|
| | | 34,381 / 934,777 27.7 years | | | Mortality, Lung cancer | Per 1 drink day | 0.97 (0.96 - 0.99) | √ | | √ | √ | √ | | √ * | | | | √ | |
| Malik 2019 USA (30882235)[109] | Health Professionals Follow-up Study + Nurse's Health Study Pooled Analysis Cohorts M/F Mostly white Health professionals | / 118,363 34 years | Physician confirmation | FFQ, **ASB**: Caffeinated, noncaffeinated, and noncarbonated low-calorie or diet beverages. | Mortality, Lung cancer | ≥2/d vs <1serving/month | 0.92 (0.77 - 1.09) Ptrend:0.20 | √ | | √ | √ | √ | √ | √ E * | | | √ | √ | |
| | | / 118,363 34 years | | | Mortality, Lung cancer | Per 1 serving day | 0.98 (0.92 - 1.08) | √ | | √ | √ | √ | √ | √ E * | | | √ | √ | |
| Skin cancer | | | | | | | | | | | | | | | | | | | |
| McCullough 2022 USA (36107009)[113] | Cancer Prevention Study II Prospective Cohort M/F Mainly white | 2,130 / 934,777 27.7 years | self-report & death certificates | Questionnaire, **ASB**: Diet soda or diet iced teas. | Mortality, Melanoma | 2+ drinks/day vs never | 1.02 (0.86 - 1.22) Ptrend:0.841 | √ | | √ | √ | √ | | √ * | | | | √ | |
| | | 2,130 | | | | Per 1 drink day | 1.01 (0.96 - 1.06) | √ | | √ | √ | √ | | √ * | | | | √ | |
| Schernhammer 2012 USA (23097267)([145]) | Nurses' Health Study + Health Professionals Follow-up Study Pooled Analysis Cohorts M/F Health professionals | 186 / 47,810 22 years | medical records, cancer registries, national death index | Semi-quantitative FFQ, **Carbonated ASB**: Artificially-sweetened soda; Caffeine-free or caffeinated colas and other soda | Incidence, Leukaemia, male | ≥1 serving/day vs none | 1.47 (0.92 – 2.35) Ptrend:0.13 | √ | | √ | √ | √ | √ | √ E * | | | | √ | |
| | | 153 / 77,218 22 years | | | Incidence, Leukaemia, female | | 1.36 (0.80 – 2.31) Ptrend:0.20 | √ | | √ | √ | √ | √ | √ E * | | √ | √ | √ | |
| | | 131 / 47,810 22 years | | | Incidence, multiple Myeloma, male | | 2.02 (1.20 – 3.40) Ptrend:0.01 | √ | | √ | √ | √ | √ | √ E * | | | | √ | |
| | | 154 / 77,218 22 years | | | Incidence, multiple Myeloma, female | | 0.79 (0.45 – 1.36) Ptrend:0.79 | √ | | √ | √ | √ | √ | √ E * | | √ | √ | √ | |
| | | 571 / 47,810 22 years | | | Incidence, Non-Hodgkin Lymphoma, male | | 1.31 (1.01 – 1.72) Ptrend:0.11 | √ | | √ | √ | √ | √ | √ E * | | | | √ | |
| | | 753 / 77,218 22 years | | | Incidence, Non-Hodgkin Lymphoma, female | | 1.00 (0.78 – 1.26) Ptrend:0.999 | √ | | √ | √ | √ | √ | √ E * | | √ | √ | √ | |
| Breast cancer | | | | | | | | | | | | | | | | | | | |
| McCullough 2022 USA (36107009)[113] | Cancer Prevention Study II Prospective Cohort F Mainly white Postmenopausal | 6,074 / 518,464 27.7 years | self-report & death certificates | Questionnaire, **ASB**: Diet soda or diet iced teas. | Mortality, Breast cancer | 2+ drinks/day vs never | 1.03 (0.94 - 1.13) Ptrend:0.838 | √ | | √ | √ | √ | | √ * | | √ | √ | √ | |
| | | 6,074 | | | | Per 1 drink day | 1.01 (0.98 - 1.04) | √ | | √ | √ | √ | | √ * | | √ | √ | √ | |
| Mullee 2019 Denmark,France, Germany,Greece ,Italy,Netherlands | European Prospective Investigation into Cancer and nutrition Prospective Cohort | 1,402 / 321,081 16.4 years | active follow up, cancer registry, death certificate | Semi-quantitative FFQ, **ASB**: Artificially Sweetened Soft Drinks | Mortality, Breast cancer | ≥1 glass/day vs <1 glass/month | 0.85 (0.59 - 1.22) Ptrend:0.38 | √ | √ | √ | √ | √ | √ | √ E * | | √ | √ | √ | |

| Author, Year Country (PMID) | Study name, type Age, sex, ethnicity, characteristics of participants | Cases/ Study size Median follow-up (years) | Case ascertainment | Exposure Assessment, detail | Outcome Sub-population | Comparison | RR (95%CI) Ptrend | Adjustments | | | | | | | | | | | |
|---|---|---|---|---|---|---|---|---|---|---|---|---|---|---|---|---|---|---|---|
| | | | | | | | | Age | Sex | Smo | Alc | Obe | Phy | Diet | Med | Mens | Repr | Oth | Skin |
| ,Norway,Spain,S weden,UK (31479109)[126] | Age:50.8 years<br>M/F | | and National death index | | | | | | | | | | | | | | | | |
| Malik 2019 USA (30882235)[109] | Nurse's Health Study<br>Prospective Cohort<br>F<br>Mostly white<br>Health professionals | NR /<br>80,647<br>34 years | Physician confirmation | FFQ,<br>**ASB**:<br>Caffeinated, noncaffeinated, and noncarbonated low-calorie or diet beverages. | Mortality, Breast cancer | ≥ 2/d vs <1 serving/month | 1.14 (0.92 - 1.40)<br>Ptrend:0.57 | ✓ | | ✓ | ✓ | ✓ | ✓ | ✓<br>E<br>* | | | ✓ | ✓ | |
| | | NR /<br>118,363<br>34 years | | | | Per 1 serving day | 1.01 (0.95 - 1.08) | ✓ | | ✓ | ✓ | ✓ | ✓ | ✓<br>E<br>* | | | ✓ | ✓ | |
| Romanos-Nanclares 2021 USA (34114021)[145] | Nurse's Health Study I and II<br>Pooled Analysis Cohorts<br>F<br>Health professionals | 7,495 /<br>82,713<br>36 years | Medical records and National Death Index | FFQ,<br>**ASB**:<br>Caffeinated, noncaffeinated, and noncarbonated low-calorie or diet beverages | Incidence, Breast cancer, NHS | ≥1day vs <1/ serving/month | 0.99 (0.92 - 1.07)<br>Ptrend:0.87 | ✓ | | | ✓ | ✓ | ✓ | ✓<br>E<br>* | ✓ | ✓ | ✓ | ✓ | |
| | | 7,495 | | | Incidence, Breast cancer, NHS | Per 1 serving day | 1.00 (0.97 - 1.04) | ✓ | | | ✓ | ✓ | ✓ | ✓<br>E<br>* | ✓ | ✓ | ✓ | ✓ | |
| | | 3,884 /<br>93,085<br>36 years | | | Incidence, Breast cancer, NHSII | ≥1day vs <1 serving/month | 0.91 (0.83 - 1.00)<br>Ptrend:0.02 | ✓ | | | ✓ | ✓ | ✓ | ✓<br>E<br>* | ✓ | ✓ | ✓ | ✓ | |
| | | 3,884 | | | Incidence, Breast cancer, NHSII | Per 1 serving day | 0.98 (0.95 - 1.00) | ✓ | | | ✓ | ✓ | ✓ | ✓<br>E<br>* | ✓ | ✓ | ✓ | ✓ | |
| Chazelas 2019 France (31292122)[44] | NutriNet-Santé study<br>Prospective Cohort<br>Age:42.2 years<br>M/F | 693 /<br>79,724<br>5.1 years | self-report, medical records reviewed by physicians, health insurance database, mortality registry | 24 Hour Diet Recall,<br>**ASB**:<br>Beverages containing non-nutritive sweeteners, such as diet soft drinks, sugar-free syrups, and diet milk-based beverages | Incidence, Breast cancer | Per 10 ml day | 0.97 (0.84 - 1.12) | ✓ | | ✓ | ✓ | ✓ | ✓ | ✓<br>E<br>* | D | ✓ | ✓ | ✓ | |
| Dunneram 2019 UK (30526696)[49] | UK Women Cohort Study<br>Prospective Cohort<br>Age:54.1 years<br>F | 1,625 /<br>32,289<br>18 years | NHS Central Registry | FFQ,<br>**ASB**:<br>Low calorie/diet soft drinks | Incidence, Breast cancer | Per 161 g/day | 1.03 (0.88 - 1.21) | ✓ | | ✓ | ✓ | | ✓ | ✓<br>E | | ✓ | ✓ | ✓ | |
| Romanos-Nanclares 2021 USA (34114021)[145] | Nurse's Health Study I and II<br>Pooled Analysis Cohorts<br>F<br>Health professionals | 8,140 /<br>175,798<br>36 years | Medical records and National Death Index | FFQ,<br>**ASB**:<br>Caffeinated, noncaffeinated, and noncarbonated low-calorie or diet beverages | Incidence, Postmenopausal breast cancer | ≥ 1 day vs < 1 serving/month | 0.96 (0.90 - 1.03)<br>Ptrend:0.12 | ✓ | | | ✓ | ✓ | ✓ | ✓<br>E<br>* | ✓ | ✓ | ✓ | ✓ | |
| | | 8,140 | | | | Per 1 serving day | 0.98 (0.95 - 1.01) | ✓ | | | ✓ | ✓ | ✓ | ✓<br>E<br>* | ✓ | ✓ | ✓ | ✓ | |
| Chazelas 2019 France (31292122)[44] | NutriNet-Santé study<br>Prospective Cohort<br>Age:42.2 years<br>M/F | 410 /<br>79,724<br>5.1 years | self-report, medical records reviewed by physicians, health insurance database, mortality registry | 24 Hour Diet Recall,<br>**ASB**:<br>Beverages containing non-nutritive sweeteners, such as diet soft drinks, sugar-free syrups, and diet milk-based beverages. | Incidence, Postmenopausal breast cancer | Per 10 ml day | 0.99 (0.78 - 1.18) | ✓ | | ✓ | ✓ | ✓ | ✓ | ✓<br>E | D | ✓ | ✓ | ✓ | |
| Dunneram 2019 UK (30526696)[49] | UK Women Cohort Study<br>Prospective Cohort<br>Age:54.1 years<br>F | 1,030 /<br>32,289<br>18 years | NHS Central Registry | FFQ,<br>**ASB**:<br>Low calorie/diet soft drinks | Incidence, Breast cancer, postmenopausal | Per 161g day | 1.03 (0.90 - 1.18) | ✓ | | ✓ | ✓ | | ✓ | | | ✓ | | ✓ | |

| Author, Year Country (PMID) | Study name, type Age, sex, ethnicity, characteristics of participants | Cases/ Study size Median follow-up (years) | Case ascertainment | Exposure Assessment, detail | Outcome Sub-population | Comparison | RR (95%CI) Ptrend | Adjustments Age | Sex | Smo | Alc | Obe | Phy | Diet | Med | Mens | Repr | Oth | Skin |
|---|---|---|---|---|---|---|---|---|---|---|---|---|---|---|---|---|---|---|---|
| Hodge 2018 Australia (29463332)[70] | Melbourne Collaborative Cohort Study Prospective Cohort M/F Not reported Excluded participants with diabetes at baseline | 946 / 21,492 23 years | cancer registry/death records/National Death Index | Self-administered Questionnaire, **ASB**: Artificially Sweetened Soft Drinks | Incidence, Postmenopausal breast cancer, Women | ≥1 /day vs < 1 serving/month | 0.95 (0.73 - 1.25) Ptrend:0.51 | ✓ | | ✓ | ✓ | ✓ | ✓ | ✓ * | | | | ✓ | |
| | | 946 | | | | | 0.92 (0.71 - 1.18) Ptrend:0.71 | ✓ | | ✓ | ✓ | ✓ | ✓ | ✓ * | | | | ✓ | |
| Romanos-Nanclares 2021 USA (34114021)[145] | Nurse's Health Study I and II Pooled Analysis Cohorts F Health professionals | 3,331 / 175,798 36 years | Medical records and National Death Index | FFQ, **ASB**: Caffeinated, noncaffeinated, and noncarbonated low-calorie or diet beverages | Incidence, Premenopausal breast cancer | ≥1day vs <1 serving/month | 0.90 (0.80 - 1.01) Ptrend:0.20 | ✓ | | | ✓ | ✓ | ✓ | ✓ E * | ✓ | ✓ | ✓ | ✓ | |
| | | 3,331 / 175,798 36 years | | | | Per 1 serving day | 0.99 (0.96 - 1.03) | ✓ | | | ✓ | ✓ | ✓ | ✓ E * | ✓ | ✓ | ✓ | ✓ | |
| Dunneram 2019 UK (30526696)[49] | UK Women Cohort Study Prospective Cohort Age:54.1 years F | 291 / 32,289 18 years | NHS Central Registry | FFQ, **ASB**: Low calorie/diet soft drinks | Incidence, Breast cancer, premenopausal | Per 161 g/day | 1.00 (0.78 - 1.29) | ✓ | | ✓ | ✓ | | ✓ | | | ✓ | | ✓ | |
| Chazelas 2019 France (31292122)[44] | NutriNet-Santé study Prospective Cohort Age:42.2 years M/F | 283 / 79,724 5.1 years | self-report, medical records reviewed by physicians, health insurance database, mortality registry | 24 Hour Diet Recall, **ASB**: Beverages containing non-nutritive sweeteners, such as diet soft drinks, sugar-free syrups, and diet milk-based beverages. | Incidence, Premenopausal breast cancer | Per 10 ml day | 0.95 (0.78 - 1.16) | ✓ | | ✓ | ✓ | ✓ | ✓ | ✓ E | D | ✓ | ✓ | ✓ | |
| Romanos-Nanclares 2021 USA (34114021)[145] | Nurse's Health Study I and II Pooled Analysis Cohorts F Health professionals | 303 / 175,798 36 years | Medical records and National Death Index | FFQ, **ASB**: Caffeinated, noncaffeinated, and noncarbonated low-calorie or diet beverages | Incidence, Basal-like breast cancer | ≥1day vs <1 serving/month | 1.04 (0.74 - 1.46) Ptrend:0.79 | ✓ | | | ✓ | ✓ | ✓ | ✓ E * | ✓ | ✓ | ✓ | ✓ | |
| | | 303 | | | Incidence, Basal-like breast cancer | Per 1 serving day | 0.97 (0.86 - 1.09) | ✓ | | | ✓ | ✓ | ✓ | ✓ E * | ✓ | ✓ | ✓ | ✓ | |
| | | 1,693 | | | ER- | ≥1day vs <1 serving/month | 0.98 (0.85 - 1.14) Ptrend:0.92 | ✓ | | | ✓ | ✓ | ✓ | ✓ E * | ✓ | ✓ | ✓ | ✓ | |
| | | 1,693 | | | | Per 1 serving day | 1.01 (0.97 - 1.07) | ✓ | | | ✓ | ✓ | ✓ | ✓ E * | ✓ | ✓ | ✓ | ✓ | |
| | | 7,302 | | | ER+ | ≥1day vs <1 serving/month | 0.97 (0.90 - 1.04) Ptrend:0.18 | ✓ | | | ✓ | ✓ | ✓ | ✓ E * | ✓ | ✓ | ✓ | ✓ | |
| | | 7,302 | | | | Per 1 serving day | 0.97 (0.94 - 1.00) | ✓ | | | ✓ | ✓ | ✓ | ✓ E * | ✓ | ✓ | ✓ | ✓ | |
| | | 250 | | | HER2 + | ≥1day vs <1 serving/month | 0.85 (0.58 - 1.26) Ptrend:0.63 | ✓ | | | ✓ | ✓ | ✓ | ✓ E * | ✓ | ✓ | ✓ | ✓ | |
| | | 250 | | | | Per 1 serving day | 1.03 (0.90 - 1.18) | ✓ | | | ✓ | ✓ | ✓ | ✓ E * | ✓ | ✓ | ✓ | ✓ | |

| Author, Year Country (PMID) | Study name, type Age, sex, ethnicity, characteristics of participants | Cases/ Study size Median follow-up (years) | Case ascertainment | Exposure Assessment, detail | Outcome Sub-population | Comparison | RR (95%CI) Ptrend | Adjustments: Age | Sex | Smo | Alc | Obe | Phy | Diet | Med | Mens | Repr | Oth | Skin |
|---|---|---|---|---|---|---|---|---|---|---|---|---|---|---|---|---|---|---|---|
| | | 2,673 | | | Luminal A | ≥1day vs <1 serving/month | 0.90 (0.80 - 1.01) Ptrend:0.02 | ✓ | | | ✓ | ✓ | ✓ | ✓ E * | ✓ | ✓ | ✓ | ✓ | |
| | | 2,673 | | | | Per 1 serving day | 0.94 (0.90 - 0.98) | ✓ | | | ✓ | ✓ | ✓ | ✓ E * | ✓ | ✓ | ✓ | ✓ | |
| | | 1,147 | | | Luminal B | ≥1day vs <1 serving/month | 1.12 (0.94 - 1.33) Ptrend:0.19 | ✓ | | | ✓ | ✓ | ✓ | ✓ E * | ✓ | ✓ | ✓ | ✓ | |
| | | 1,147 | | | | Per 1 serving day | 1.03 (0.97 - 1.10) | ✓ | | | ✓ | ✓ | ✓ | ✓ E * | ✓ | ✓ | ✓ | ✓ | |
| Ishitani 2008 USA (18852405)[75] | Women's Health Study Prospective Cohort Age:55 years F | 1,179 / 38,432 10 years | self-reported/death certificate/ medical records | FFQ, **Artificially sweetened caffeinated cola**: Low-calorie caffeinated cola | Incidence / Mortality, Invasive breast cancer | ≥2 vs 0 cans/day | 0.88 (0.68 - 1.13) Ptrend:0.16 | ✓ | | ✓ | ✓ | ✓ | ✓ | ✓ E | ✓ | ✓ | ✓ | ✓ | |
| | | 1,165 | | **Artificially sweetened decaffeinated cola**: Low-calorie decaffeinated cola | | | 1.30 (0.99 - 1.71) Ptrend:0.25 | ✓ | | ✓ | ✓ | ✓ | ✓ | ✓ E | ✓ | ✓ | ✓ | ✓ | |
| Endometrial cancer | | | | | | | | | | | | | | | | | | | |
| McCullough 2022 USA (36107009)[113] | Cancer Prevention Study II Prospective Cohort F Mainly white Postmenopausal | 1,693 / 518,464 27.7 years | self-report & death certificates | Questionnaire, **ASB**: Diet soda or diet iced teas. | Mortality, Uterus/Endometrium | 2+ drinks/day vs never | 1.01 (0.85 - 1.21) Ptrend:0.878 | ✓ | | ✓ | ✓ | ✓ | | ✓ * | | ✓ | ✓ | ✓ | |
| | | 1,693 | | | | Per 1 drinks day | 0.99 (0.95 - 1.05) | ✓ | | ✓ | ✓ | ✓ | | ✓ * | | ✓ | ✓ | ✓ | |
| Dunneram 2019 UK (30526696)[49] | UK Women Cohort Study Prospective Cohort Age:54.1 years F | 175 / 32,289 18 years | NHS Central Registry | FFQ, **ASB**: Low calorie/diet soft drinks | Incidence, Endometrial Cancer | Per 161 g/day | 1.00 (0.60 - 1.66) | ✓ | | ✓ | ✓ | | ✓ | ✓ E | D | ✓ | ✓ | ✓ | |
| | | 86 | | | Postmenopausal | Per 161 g/day | 1.27 (1.00 - 1.61) | ✓ | | ✓ | ✓ | | ✓ | | D | ✓ | | ✓ | |
| Hodge 2018 Australia (29463332)[70] | Melbourne Collaborative Cohort Study Prospective Cohort M/F Not reported Excluded participants with diabetes at baseline | 167 / 27,413 23 years | cancer registry/death records/National Death Index | Self-administered Questionnaire, **ASB**: Artificially Sweetened Soft Drinks | Incidence, Endometrial Cancer, | ≥1/day vs < 1serving/month | 0.81 (0.42 - 1.55) Ptrend:0.17 | ✓ | | ✓ | ✓ | ✓ | ✓ | ✓ * | | | | ✓ | |
| | | 130 | | | | Linear model | 0.92 (0.52 - 1.65) | ✓ | | ✓ | ✓ | ✓ | ✓ | ✓ * | | | | ✓ | |
| Inoue-Choi 2013 USA (24273064)[74] | Iowa Women's Health Study Prospective Cohort Age:61.6 years F Not reported Postmenopausal | 506 / 23,039 24 years | State Health Registry, National Death Index | FFQ, **ASB**: sugar-free beverages | Incidence, type 1 endometrial cancer | 2.8-64.1 vs 0 servings/week | 0.77 (0.59 - 1.01) Ptrend:0.31 | ✓ | | ✓ | ✓ | ✓ | ✓ | ✓ | D | ✓ | ✓ | | |
| | | 86 | | | Incidence, Type 2 endometrial cancer | 2.8-64.1 vs 0 servings/week | 0.89 (0.48 - 1.68) Ptrend:0.95 | ✓ | | ✓ | ✓ | ✓ | ✓ | ✓ | D | ✓ | ✓ | | |
| Ovarian cancer | | | | | | | | | | | | | | | | | | | |
| McCullough 2022 USA (36107009)[113] | Cancer Prevention Study II Prospective Cohort F Mainly white Postmenopausal | 3,225 / 518,464 27.7 years | self-report & death certificates | Questionnaire, **ASB**: Diet soda or diet iced teas. | Mortality, Ovarian cancer | 2+ drinks/day vs never | 1.01 (0.89 - 1.15) Ptrend:0.284 | ✓ | | ✓ | ✓ | ✓ | | ✓ * | | ✓ | ✓ | ✓ | |

| Author, Year Country (PMID) | Study name, type Age, sex, ethnicity, characteristics of participants | Cases/ Study size Median follow-up (years) | Case ascertainment | Exposure Assessment, detail | Outcome Sub-population | Comparison | RR (95%CI) Ptrend | Adjustments Age | Sex | Smo | Alc | Obe | Phy | Diet | Med | Mens | Repr | Oth | Skin |
|---|---|---|---|---|---|---|---|---|---|---|---|---|---|---|---|---|---|---|---|
| | | 3,225 | | | | Per 1 drink day | 1.00 (0.97 - 1.04) | ✓ | | ✓ | ✓ | ✓ | | ✓<br>* | | ✓ | ✓ | ✓ | |
| Dunneram 2019<br>UK (30526696)[49] | UK Women Cohort Study<br>Prospective Cohort<br>Age:54.1 years<br>F | 251 /<br>32,289<br>18 years | NHS Central Registry | FFQ,<br>**ASB**:<br>Low calorie/diet soft drinks | Incidence,<br>Ovarian cancer | Per 161 g/day | 1.01 (0.65 - 1.56) | ✓ | | ✓ | ✓ | | ✓ | ✓<br>E<br>* | | ✓ | ✓ | ✓ | |
| Hodge 2018<br>Australia<br>(29463332)[70] | Melbourne Collaborative Cohort Study<br>Prospective Cohort<br>M/F<br>Not reported<br>Excluded participants with diabetes at baseline | 130 /<br>21,492<br>23 years | cancer registry/death records/National Death Index | Self-administered Questionnaire,<br>**ASB**:<br>Artificially Sweetened Soft Drinks | Incidence,<br>Ovarian cancer | >/=1/day vs <1 serving/month | 1.37 (0.72 - 2.61)<br>Ptrend:0.17 | ✓ | | ✓ | ✓ | ✓ | ✓ | ✓<br>* | | | | ✓ | |
| | | 130 | | | | Linear model | 1.51 (0.84 - 2.73) | ✓ | | ✓ | ✓ | ✓ | ✓ | ✓<br>* | | | | ✓ | |
| **Prostate cancer** | | | | | | | | | | | | | | | | | | | |
| McCullough 2022<br>USA<br>(36107009)[113] | Cancer Prevention Study II<br>Prospective Cohort<br>M/F<br>Mainly white | 9,381 /<br>416,313<br>27.7 years | self-report & death certificates | Questionnaire,<br>**ASB**:<br>Diet soda or diet iced teas. | Mortality,<br>Prostate cancer | 2+ drinks/day vs never drinks/day | 0.98 (0.89 - 1.08)<br>Ptrend:0.258 | ✓ | | ✓ | ✓ | ✓ | | ✓<br>* | | | | ✓ | |
| | | 9,381 | | | | Per 1 drinks day | 0.99 (0.96 - 1.02) | ✓ | | ✓ | ✓ | ✓ | | ✓<br>* | | | | ✓ | |
| Mullee 2019<br>Denmark,France, Germany,Greece ,Italy,Netherlands ,Norway,Spain,S weden,UK<br>(31479109)[126] | European Prospective Investigation into Cancer and nutrition<br>Prospective Cohort<br>Age:50.8 years<br>M/F | 907 /<br>130,662<br>16.4 years | active follow up, cancer registry, death certificate and National death index | Semi-quantitative FFQ,<br>**ASB**:<br>Artificially Sweetened Soft Drinks | Mortality,<br>Prostate cancer | ≥1 glass/day vs <1 glass/month | 1.05 (0.64 - 1.75) | ✓ | ✓ | ✓ | ✓ | ✓ | ✓ | ✓<br>E<br>* | | ✓ | ✓ | ✓ | |
| Malik 2019 USA<br>(30882235)[109] | HPFS<br>Prospective Cohort<br>M<br>Mostly white<br>Health professionals | / 37,716<br>28 years | Physician confirmation | FFQ,<br>**ASB**:<br>Caffeinated, noncaffeinated, and noncarbonated low-calorie or diet beverages. | Mortality,<br>Prostate cancer | ≥2/d vs < 1 serving/month | 1.01 (0.67 - 1.52)<br>Ptrend:0.92 | ✓ | | ✓ | ✓ | ✓ | ✓ | ✓<br>E<br>* | | | | ✓ | |
| | | / 118,363<br>28 years | | | | Per 1 serving day | 1.02 (0.91-1.14) | ✓ | | ✓ | ✓ | ✓ | ✓ | ✓<br>E<br>* | | | | ✓ | |
| Chazelas 2019<br>France<br>(31292122)[44] | NutriNet-Santé study<br>Prospective Cohort<br>Age:42.2 years<br>M/F | 291 /<br>21,533<br>5.1 years | self-report, medical records reviewed by physicians, health insurance database, mortality registry | 24 Hour Diet Recall,<br>**ASB**:<br>Beverages containing non-nutritive sweeteners, such as diet soft drinks, sugar-free syrups, and diet milk-based beverages. | Incidence,<br>Prostate cancer | Per 10 ml day | 0.58 (0.28 - 1.18) | ✓ | | ✓ | ✓ | ✓ | ✓ | ✓<br>E | D | | | ✓ | |
| Hodge 2018<br>Australia<br>(29463332)[70] | Melbourne Collaborative Cohort Study<br>Prospective Cohort<br>M/F<br>Not reported<br>Excluded participants with diabetes at baseline | 433 /<br>14,101<br>23 years | cancer registry/death records/National Death Index | Self-administered Questionnaire,<br>**ASB**:<br>Artificially Sweetened Soft Drinks | Incidence,<br>Aggressive prostate cancer | ≥1/day vs <1 serving/month | 0.81 (0.49 - 1.33)<br>Ptrend:0.66 | ✓ | | ✓ | ✓ | ✓ | ✓ | ✓<br>* | | | | ✓ | |
| | | 433 | | | | Linear model | 0.91 (0.60 - 1.38) | ✓ | | ✓ | ✓ | ✓ | ✓ | ✓<br>* | | | | ✓ | |
| **Kidney cancer** | | | | | | | | | | | | | | | | | | | |
| Ringel 2023 USA<br>(36601047)[144] | Women's Health Initiative<br>Prospective Cohort<br>Age:66.6 years<br>F<br>Mixed<br>Postmenopausal | 327 /<br>80,388 | Physician confirmation | Self-administered Questionnaire,<br>**ASB**: | Incidence,<br>Kidney cancer | >/1d vs Never or <1/wk | 1.14 (0.80 - 1.62) | ✓ | | ✓ | | ✓ | | ✓ | | | | ✓ | |

| Author, Year Country (PMID) | Study name, type Age, sex, ethnicity, characteristics of participants | Cases/ Study size Median follow-up (years) | Case ascertainment | Exposure Assessment, detail | Outcome Sub-population | Comparison | RR (95%CI) Ptrend | Adjustments | | | | | | | | | | | |
|---|---|---|---|---|---|---|---|---|---|---|---|---|---|---|---|---|---|---|---|
| | | | | | | | | Age | Sex | Smo | Alc | Obe | Phy | Diet | Med | Mens | Repr | Oth | Skin |
| Hodge 2018 Australia (29463332)[70] | Melbourne Collaborative Cohort Study<br>Prospective Cohort<br>M/F<br>Not reported<br>Excluded participants with diabetes at baseline | 146 / 35,593<br>23 years | cancer registry/death records/National Death Index | Self-administered Questionnaire, **ASB**:<br>Artificially Sweetened Soft Drinks | Incidence, Kidney cancer | ≥1/day vs <1 serving/month | 0.92 (0.46 - 1.84)<br>Ptrend:0.33 | ✓ | ✓ | ✓ | ✓ | ✓ | ✓ | ✓<br>* | | | | ✓ | |
| | | 146 | | | | Linear model | 1.51 (0.84 - 2.73) | ✓ | ✓ | ✓ | ✓ | ✓ | ✓ | ✓<br>* | | | | ✓ | |
| Heath 2021 multi-national (33849969)[66] | European Prospective Investigation into Cancer and nutrition<br>Prospective Cohort<br>Age:52 years<br>M/F | 589 / 389,220<br>15 years | population registries or active follow up | Semi-quantitative FFQ, **ASB**:<br>Diet / artificially sweetened soft drinks | Incidence, Renal cell carcinoma | Per 100 g day | 1.02 (0.96 - 1.08) | ✓ | ✓ | ✓ | ✓ | ✓ | ✓ | ✓<br>E<br>* | | | | ✓ | |
| | | 265 | | | Mortality, Renal cell carcinoma | Per 100 g day | 1.06 (0.99 - 1.14) | ✓ | ✓ | ✓ | ✓ | ✓ | ✓ | ✓<br>E<br>* | | | | ✓ | |
| | | 142 | population registries or active follow up | | Male | Per 100 g day | 1.04 (0.93 - 1.15) | ✓ | | ✓ | ✓ | ✓ | ✓ | ✓<br>E<br>* | | | | ✓ | |
| | | 123 | | | Female | Per 100 g day | 1.08 (0.98 - 1.19) | ✓ | | ✓ | ✓ | ✓ | ✓ | ✓<br>E<br>* | | | | ✓ | |
| McCullough 2022 USA (36107009)[113] | Cancer Prevention Study II<br>Prospective Cohort<br>M/F<br>Mainly white | 3,129 / 934,777<br>27.7 years | self-report & death certificates | Questionnaire, **ASB**:<br>Diet soda or diet iced teas. | Mortality, Kidney cancer | 2+ drinks/day vs never | 0.95 (0.82 - 1.10)<br>Ptrend:0.468 | ✓ | | ✓ | ✓ | ✓ | | ✓<br>* | | | | ✓ | |
| | | 3,129 | | | | Per 1 drink day | 0.99 (0.95 - 1.03) | ✓ | | ✓ | ✓ | ✓ | | ✓<br>* | | | | ✓ | |
| Lee 2006 USA (16775182)[98] | Nurses' Health Study and Health Professionals Follow-up Study<br>Pooled Analysis Cohorts<br>Range:30-75 years<br>M/F | 248 / 136,587 | Self-report, medical records reviewed by physician, national death index, death report by family members in follow up questionnaires | Validated FFQ, **Carbonated ASB**:<br>artificially sweetened soda | Incidence, Renal cell cancer | Per 1 serving day | 0.97 (0.82-1.15) | ✓ | | ✓ | ✓ | ✓ | | ✓<br>E | D | | ✓ | | |
| Bladder cancer | | | | | | | | | | | | | | | | | | | |
| McCullough 2022 USA (36107009)[113] | Cancer Prevention Study II<br>Prospective Cohort<br>M/F<br>Mainly white | 3,419 / 934,777<br>27.7 years | self-report & death certificates | Questionnaire, **ASB**:<br>Diet soda or diet iced teas | Mortality, Bladder cancer | 2+ drinks/day vs never | 1.02 (0.88 - 1.18)<br>Ptrend:0.922 | ✓ | | ✓ | ✓ | ✓ | | ✓<br>* | | | | ✓ | |
| | | 3,419 / 934,777<br>27.7 years | | | | Per 1 drink day | 0.99 (0.95 - 1.03) | ✓ | | ✓ | ✓ | ✓ | | ✓<br>* | | | | ✓ | |
| Ringel 2023 USA (36601047)[144] | Women's Health Initiative<br>Prospective Cohort<br>Age:66.6 years<br>F<br>Mixed<br>Postmenopausal | 448 / 80,388 | Physician confirmation | Self-administered Questionnaire, **ASB**: | Incidence, Bladder cancer | >/1d vs Never or <1/wk | 0.75 (0.53 - 1.06) | ✓ | | ✓ | | | | ✓ | | | | ✓ | |
| Brain cancer | | | | | | | | | | | | | | | | | | | |

| Author, Year Country (PMID) | Study name, type Age, sex, ethnicity, characteristics of participants | Cases/ Study size Median follow-up (years) | Case ascertainment | Exposure Assessment, detail | Outcome Sub-population | Comparison | RR (95%CI) Ptrend | Adjustments<br>Age | Sex | Smo | Alc | Obe | Phy | Diet | Med | Mens | Repr | Oth | Skin |
|---|---|---|---|---|---|---|---|---|---|---|---|---|---|---|---|---|---|---|---|
| McCullough 2022<br>USA<br>(36107009)[113] | Cancer Prevention Study II<br>Prospective Cohort<br>M/F<br>Mainly white | 3,402 /<br>934,777<br>27.7 years | self-report & death certificates | Questionnaire,<br>**ASB**:<br>Diet soda or diet iced teas | Mortality, Brain cancer | 2+ drinks/day vs never | 0.97 (0.84 - 1.11)<br>Ptrend:0.737 | ✓ | | ✓ | ✓ | ✓ | | ✓<br>* | | | | ✓ | |
| | | 3402 | | | | Per 1 drink day | 0.98 (0.95 - 1.02) | ✓ | | ✓ | ✓ | ✓ | | ✓<br>* | | | | ✓ | |
| Thyroid cancer | | | | | | | | | | | | | | | | | | | |
| Zamora-Ros<br>2023 multi-national<br>(35907037)[180] | European Prospective Investigation into Cancer and nutrition<br>Prospective Cohort<br>Age:51.1 years<br>M/F | 495 /<br>450,064<br>13.9 years | Cancer and mortality registries, pathology registries, active follow up, health insurance records | Semi-quantitative FFQ,<br>**ASB**:<br>Diet / artificially sweetened soft drinks. | Incidence, Thyroid cancer | 43 - 3389.5 vs 0 ml/day | 1.16 (0.80 - 1.69)<br>Ptrend:0.26 | ✓ | ✓ | ✓ | ✓ | ✓ | ✓ | ✓<br>E | | ✓ | ✓ | ✓ | |
| | | 495 | | | | Per 100 ml day | 1.00 (0.91 - 1.11) | ✓ | ✓ | ✓ | ✓ | ✓ | ✓ | ✓<br>E | | ✓ | ✓ | ✓ | |
| | | 469 | | | Incidence, Papillary thyroid cancer | 43 - 3389.5 vs 0 ml/day | 1.51 (1.04 - 2.19)<br>Ptrend:0.16 | ✓ | ✓ | ✓ | ✓ | ✓ | ✓ | ✓<br>E | | ✓ | ✓ | ✓ | |
| | | 469 | | | | Per 100 ml day | 1.05 (0.95 - 1.15) | ✓ | ✓ | ✓ | ✓ | ✓ | ✓ | ✓<br>E | | ✓ | ✓ | ✓ | |
| | | 86 | | | Incidence, Follicular thyroid cancer | 43 - 3389.5 vs 0 ml/day | 0.38 (0.11 - 1.30)<br>Ptrend:0.3 | ✓ | ✓ | ✓ | ✓ | ✓ | ✓ | ✓<br>E<br>* | | ✓ | ✓ | ✓ | |
| | | 86 | | | | Per 100 ml day | 1.00 (0.91 - 1.11) | ✓ | ✓ | ✓ | ✓ | ✓ | ✓ | ✓<br>E<br>* | | ✓ | ✓ | ✓ | |
| Haematological cancers | | | | | | | | | | | | | | | | | | | |
| McCullough 2022<br>USA<br>(36107009)[113] | Cancer Prevention Study II<br>Prospective Cohort<br>M/F<br>Mainly white | 6,319 /<br>934,777<br>27.7 years | self-report & death certificates | Questionnaire,<br>**ASB**:<br>Diet soda or diet iced teas. | Mortality, Leukaemia | 2+ drinks/day vs never | 0.97 (0.87 - 1.07)<br>Ptrend:0.444 | ✓ | | ✓ | ✓ | ✓ | | ✓<br>* | | | | ✓ | |
| | | 6,319 | | | | Per 1 drinks day | 0.99 (0.96 - 1.02) | ✓ | | ✓ | ✓ | ✓ | | ✓<br>* | | | | ✓ | |
| | | 3,511 | | | Mortality, Multiple Myeloma | 2+ drinks/day vs never | 1.03 (0.90 - 1.17)<br>Ptrend:0.707 | ✓ | | ✓ | ✓ | ✓ | | ✓<br>* | | | | ✓ | |
| | | 3,511 | | | | Per 1 drinks day | 1.02 (0.98 - 1.06) | ✓ | | ✓ | ✓ | ✓ | | ✓<br>* | | | | ✓ | |
| | | 6,600 | | | Mortality, Non-Hodgkin Lymphoma | 2+ drinks/day vs never | 1.01 (0.92 - 1.12)<br>Ptrend:0.870 | ✓ | | ✓ | ✓ | ✓ | | ✓<br>* | | | | ✓ | |
| | | 6,600 | | | | Per 1 drinks day | 1.00 (0.98 - 1.03) | ✓ | | ✓ | ✓ | ✓ | | ✓<br>* | | | | ✓ | |
| Schernhammer<br>2012 USA<br>(23097267)[150] | Nurses' Health Study + Health Professionals Follow-up Study<br>Pooled Analysis Cohorts<br>M/F<br>Not reported<br>Health professionals | 186 /<br>47,810<br>22 years | medical records, cancer registries, national death index | Semi-quantitative FFQ,<br>**Carbonated ASB**:<br>diet soda | Incidence, Leukaemia, Male | ≥1 serving/day vs 0 | 1.47 (0.92 - 2.35)<br>Ptrend:0.13 | ✓ | | ✓ | ✓ | ✓ | ✓ | ✓<br>E<br>* | | | | ✓ | |
| | | 153 /<br>77,218 | | | Incidence, Leukaemia, Female | ≥1 serving/day vs 0 | 1.36 (0.80 - 2.31)<br>Ptrend:0.20 | ✓ | | ✓ | ✓ | ✓ | ✓ | ✓<br>E<br>* | | ✓ | ✓ | ✓ | |

| Author, Year Country (PMID) | Study name, type Age, sex, ethnicity, characteristics of participants | Cases/ Study size Median follow-up (years) | Case ascertainment | Exposure Assessment, detail | Outcome Sub-population | Comparison | RR (95%CI) Ptrend | Adjustments | | | | | | | | | | | |
|---|---|---|---|---|---|---|---|---|---|---|---|---|---|---|---|---|---|---|---|
| | | | | | | | | Age | Sex | Smo | Alc | Obe | Phy | Diet | Med | Mens | Repr | Oth | Skin |
| | | 131 / 47,810 | | | Incidence, Multiple Myeloma, Male | ≥1 serving/day vs 0 | 2.02 (1.20 - 3.40) Ptrend:0.01 | ✓ | | ✓ | ✓ | ✓ | ✓ | ✓ E * | | | | ✓ | |
| | | 154 / 77,218 | | | Incidence, Multiple Myeloma, Female | ≥1 serving/day vs 0 | 0.79 (0.45 - 1.36) Ptrend:0.79 | ✓ | | ✓ | ✓ | ✓ | ✓ | ✓ E * | | ✓ | ✓ | ✓ | |
| | | 571 / 47,810 | | | Incidence, Non-Hodgkin Lymphoma, Male | ≥1 serving/day vs 0 | 1.31 (1.01 - 1.72) Ptrend:0.11 | ✓ | | ✓ | ✓ | ✓ | ✓ | ✓ E * | | | | ✓ | |
| | | 753 / 77,218 | | | Incidence, Non-Hodgkin Lymphoma, Female | ≥1 serving/day vs 0 | 1.00 (0.78 - 1.26) Ptrend:0.999 | ✓ | | ✓ | ✓ | ✓ | ✓ | ✓ E * | | ✓ | ✓ | ✓ | |
| McCullough 2014 USA (25342696)[114] | Cancer Prevention Study II Prospective Cohort Age:69 years M/F | 1,196 / 100,442 10 years | medical records and cancer registries | FFQ, **Carbonated ASB**: Artificially sweetened carbonated beverage | Incidence, Non-Hodgkin Lymphoma | >1 vs 0 cans/week | 0.92 (0.73 - 1.17) Ptrend:0.14 | ✓ | ✓ | ✓ | | ✓ | | ✓ E * | D | | | | |
| | | 1,196 | | | Incidence, Non-Hodgkin Lymphoma | Per 1 can day | 0.99 (0.95-1.03) | ✓ | ✓ | ✓ | | ✓ | | ✓ E * | D | | | | |
| Podoltsev 2020 (31904114)[140] | NIH- AARP Diet and Health Study Prospective Cohort Age:62.8 years M/F Mostly white Retired | 490 / 490,049 15.5 years | Cancer registries, pathology registries, active follow up, health insurance records | Questionnaire, **Caffeinated diet soft drinks** | Incidence, Myelo-proliferative neoplasms | Yes vs no | 1.01 (0.81 – 1.27) Ptrend: 0.98 | ✓ | ✓ | | | | | | | | | ✓ | |
| | | 190 / 490,049 | | | Incidence, Polycythemia vera | Yes vs no | 0.96 (0.67 – 1.38) Ptrend: 0.82 | ✓ | ✓ | | | | | | | | | ✓ | |
| | | 146 / 490,049 | | | Incidence, Essential thrombo-cythemia | Yes vs no | 1.01 (0.67 – 1.51) Ptrend: 0.97 | ✓ | ✓ | | | | | | | | | ✓ | |
| | | 490 / 490,049 | | Questionnaire, **Decaffeinated diet soft drinks** | Incidence, Myelo-proliferative neoplasms | Yes vs no | 0.95 (0.78 – 1.16) Ptrend: 0.62 | ✓ | ✓ | | | | | | | | | ✓ | |
| | | 190 / 490,049 | | | Incidence, Polycythemia vera | Yes vs no | 0.91 (0.65 – 1.26) Ptrend: 0.56 | ✓ | ✓ | | | | | | | | | ✓ | |
| | | 146 / 490,049 | | | Incidence, Essential thrombo-cythemia | Yes vs no | 0.99 (0.69 – 1.42) Ptrend: 0.77 | ✓ | ✓ | | | | | | | | | ✓ | |

**Age**: age; **Alc**: alcohol; **Diet**: dietary factors, energy intake (E), or * adjustment for other soft drinks not under investigation; **Med**: medical-related factors (e.g. infections, diseases in the breast, digestive tract, gallbladder, liver, or pancreas, cancer screening, precancerous conditions, or pre-existing skin cancer) or diabetes (D); **Mens**: menstrual characteristics (e.g. age at menarche, menopausal status/duration, age at menopause, or type of menopause); **Obe**: obesity (adiposity measures); **Oth**: others (e.g. family history/genetic factor, race/ethnicity, ancestry, socioeconomic factors, or marriage); **Phy**: physical activity; **Repr**: reproductive or hormonal factors (e.g. parity, age at first/last birth, postmenopausal hormone use, or oral contraceptive use); **Sex**: sex; **Skin**: skin-related factors (e.g. UV light/sun exposure, sunburns, time spent outdoors, sunscreen use, indoor tanning, moles, fair skin, red/blond hair, blue/green eyes, or freckles); **Smo**: smoking.

#### Supplementary Table 13. Descriptive table of the included observational studies of juices and cancer incidence and mortality.

| Author, Year Country (PMID) | Study name, type Age, sex, ethnicity, characteristics of participants | Cases/ Study size Median follow-up (years) | Case ascertainment | Exposure Assessment, detail | Outcome Sub-population | Comparison | RR (95%CI) Ptrend | Adjustments | | | | | | | | | | | |
|---|---|---|---|---|---|---|---|---|---|---|---|---|---|---|---|---|---|---|---|
| | | | | | | | | Age | Sex | Smo | Alc | Obe | Phy | Diet | Med | Mens | Repr | Oth | Skin |
| **Mouth, pharynx, and larynx** | | | | | | | | | | | | | | | | | | | |
| Freedman 2008 USA (18092323)[59] | NIH- AARP Diet and Health Study Prospective Cohort<br>Prospective Cohort<br>Range:50-71 years<br>M/F | 787 / 490,802<br>4.5 years | Cancer registry | FFQ,<br>**Fruit juices**:<br>orange and grapefruit juice, and other fruit juices and drinks | Incidence, Head and Neck cancer | Median intake 5.8 vs 1.5 serving /day /1000 Calories | 0.94 (0.76 - 1.17)<br>Ptrend:0.930 | ✓ | ✓ | ✓ | ✓ | ✓ | ✓ | ✓ E | | | | ✓ | |
| | | 787 / 490,802<br>4.5 years | | | | Per serving /1000 Calories | 1.03 (0.93 - 1.15) | ✓ | ✓ | ✓ | ✓ | ✓ | ✓ | ✓ E | | | | ✓ | |
| | | 319 / 490,802<br>4.5 years | | | Incidence, oral cavity cancer | Median intake 5.8 vs 1.5 serving /day /1000 Calories | 1.10 (0.78 - 1.53)<br>Ptrend:0.556 | ✓ | ✓ | ✓ | ✓ | ✓ | ✓ | ✓ E | | | | ✓ | |
| | | 319 / 490,802<br>4.5 years | | | | Per serving /1000 Calories | 1.06 (0.90 - 1.25) | ✓ | ✓ | ✓ | ✓ | ✓ | ✓ | ✓ E | | | | ✓ | |
| | | 142 / 490,802<br>4.5 years | | | Incidence, Oro-hypopharynx cancer | Median intake 5.8 vs 1.5 serving /day /1000 Calories | 1.05 (0.63 - 1.74)<br>Ptrend:0.809 | ✓ | ✓ | ✓ | ✓ | ✓ | ✓ | ✓ E | | | | ✓ | |
| | | 142 / 490,802<br>4.5 years | | | | Per serving /1000 Calories | 1.12 (0.88 - 1.43) | ✓ | ✓ | ✓ | ✓ | ✓ | ✓ | ✓ E | | | | ✓ | |
| | | 279 / 490,802<br>4.5 years | | | Incidence, laryngeal cancer | Median intake 5.8 vs 1.5 serving /day /1000 Calories | 0.85 (0.59 - 1.22)<br>Ptrend:0.742 | ✓ | ✓ | ✓ | ✓ | ✓ | ✓ | ✓ E | | | | ✓ | |
| | | 279 / 490,802<br>4.5 years | | | | Per serving /1000 Calories | 1.00 (0.82 - 1.21) | ✓ | ✓ | ✓ | ✓ | ✓ | ✓ | ✓ E | | | | ✓ | |
| Kjaerheim 1998 Norway (9486469)[90] | Norwegian Men UADT<br>Prospective Cohort<br>M | 63 / 10,900<br>25 years | population survey | FFQ,<br>**Fruit juices**:<br>lemonade | Incidence, mouth, tongue, pharynx, larynx, esophagus | 6+ times/month vs < monthly | 0.90 (0.50 - 1.60)<br>Ptrend:>0.5 | ✓ | | ✓ | ✓ | | | | | | | | |
| **Oesophageal** | | | | | | | | | | | | | | | | | | | |
| Freedman 2007 USA (17691111)[58] | NIH- AARP Diet and Health Study Prospective Cohort<br>Age:63 years<br>M/F<br>Mainly white<br>Retired | 103 / 490,802<br>5 years | linkage to state cancer registries | Validated FFQ,<br>**Fruit juices**:<br>orange and grapefruit juice, and other fruit juices and drinks | Incidence, Oesophageal squamous cell carcinoma | 1.37 vs 0 servings /day /1000 kcal | 0.83 (0.45 - 1.52)<br>Ptrend:0.863 | ✓ | ✓ | ✓ | ✓ | ✓ | ✓ | ✓ E | | | | ✓ | |
| | | 213 / 490,802<br>5 years | | | Incidence, Oesophageal adenocarcinoma | | 0.85 (0.55 - 1.33)<br>Ptrend:0.766 | ✓ | ✓ | ✓ | ✓ | ✓ | ✓ | ✓ E | | | | ✓ | |
| Iso 2007 Japan (18260705)[76] | Japan Collaborative Cohort study for Evaluation of Cancer Risk<br>Prospective Cohort<br>Range:40-79 years<br>M/F | 138 / 105,500<br>15 years | municipal resident registration records, death certificates | Validated FFQ,<br>**Pure fruit juices**:<br>Fresh fruit juice | Mortality, Oesophageal cancer, male | ≥3 vs <1 times/week | 0.95 (0.61 - 1.49) | ✓ | | | | | | | | | | | |

| Author, Year Country (PMID) | Study name, type Age, sex, ethnicity, characteristics of participants | Cases/ Study size Median follow-up (years) | Case ascertainment | Exposure Assessment, detail | Outcome Sub-population | Comparison | RR (95%CI) Ptrend | Adjustments | | | | | | | | | | | |
|---|---|---|---|---|---|---|---|---|---|---|---|---|---|---|---|---|---|---|---|
| | | | | | | | | Age | Sex | Smo | Alc | Obe | Phy | Diet | Med | Mens | Repr | Oth | Skin |
| | | 21 / 105,500 15 years | | | Mortality, Oesophageal cancer, female | | 0.84 (0.27 - 2.62) | ✓ | | | | | | | | | | | |
| **Stomach** | | | | | | | | | | | | | | | | | | | |
| Freedman 2008 USA (18166992)[60] | NIH- AARP Diet and Health Study Prospective Cohort Range:50-71 years M/F Mainly white Retired | 394 / 490,802 4.5 years | linkage with 11 state cancer registry databases | Validated FFQ, **Fruit juices**: orange and grapefruit juice, and other fruit juices and drinks | Incidence, Stomach cancer | Per daily serving /1000 Calories | 0.97 (0.83 - 1.13) | ✓ | ✓ | ✓ | ✓ | ✓ | ✓ | ✓ E | | | | ✓ | |
| Iso 2007 Japan (18260705)[76] | Japan Collaborative Cohort study for Evaluation of Cancer Risk Prospective Cohort Range:40-79 years M/F | 678 / 105,500 15 years | municipal resident registration records, death certificates | Validated FFQ, **Pure fruit juices**: Fresh fruit juice in summer season | Mortality, Stomach cancer, male | ≥3 vs <1 times/week | 1.15 (0.93 - 1.42) | ✓ | | | | | | | | | | | |
| | | 327 / 105,500 15 years | | | Mortality, Stomach cancer, female | | 1.28 (0.95 - 1.74) | ✓ | | | | | | | | | | | |
| Botterweck 1998 Netherlands (9801014)[43] | The Netherlands Cohort study Case Cohort Range:55-69 years M/F | 282 / 120,852 6.3 years | Cancer registry | FFQ, **Citrus juices**: Orange/grapefruit juice (processed) | Incidence, Stomach cancer | Per 25 g day | 0.92 (0.82 - 1.03) | ✓ | ✓ | ✓ | | | | ✓ | | | | ✓ | |
| Khairan 2023 Japan (36167978)[84] | The Japan Public Health Center-based Prospective Study Prospective Cohort Range:40-59 years M/F Japanese | 676 / 39,353 16.7 years | medical and population cancer registries, death certificates | Questionnaire & FFQ, **Pure orange juice** | Incidence, Gastric cancer, female | 182.9-4621.8 vs 0-64.7 ml/day | 1.04 (0.87 - 1.24) Ptrend:0.80 | ✓ | | ✓ | ✓ | ✓ | ✓ | ✓ E | | | | ✓ | |
| | | 1,465 / 35,102 16.7 years | | | Incidence, Gastric cancer, males | 265.8-3757.6 vs 0-108.3 ml/day | 0.97 (0.86 - 1.10) Ptrend:0.66 | ✓ | | ✓ | ✓ | ✓ | ✓ | ✓ E | | | | ✓ | |
| **Colorectal** | | | | | | | | | | | | | | | | | | | |
| Jin 2023 UK (38004195)[78] | UK Biobank Prospective Cohort Age:55.87 years M/F Mainly white | 1,466 / 118,210 12.8 years | hospital records, cancer registry, death registry | 24 Hour Diet Recall, **Fruit juices** | Incidence, Colorectal cancer | Per 1 SD day | 1.01 (0.96 - 1.06) | ✓ | ✓ | ✓ | | ✓ | ✓ | ✓ E | ✓ D | | | ✓ | |
| Kim 2022 USA (36443779)[87] | The Multiethnic cohort study Prospective Cohort M/F Multi-ethnic | 2,582 / 79,952 19.2 years | cancer registries | Quantitative FFQ, **Fruit juices** | Incidence, Colorectal cancer, Male | Per 1 SD | 0.96 (0.92 - 1.00) | ✓ | | ✓ | ✓ | ✓ | ✓ | ✓ E | ✓ | | | ✓ | |
| Makarem 2018 USA (29674390)[108] | Framingham Offspring Cohort Prospective Cohort Age:55.4 years M/F | 68 / 3,184 | medical examinations | Semi-quantitative FFQ, **Fruit juices**: apple juice/cider, orange juice, grapefruit juice and other juice. | Incidence, Colorectal cancer | >7.0 vs <1.9 serving/week | 1.66 (0.88 - 3.12) | ✓ | ✓ | ✓ | ✓ | ✓ | ✓ | ✓ E | D | | | ✓ | |
| Kunzmann 2016 USA (26559156)[93] | Prostate, Lung, Colorectal and Ovarian Cancer Screening Trial Prospective Cohort Age:63 years M/F Mostly white Screening Program | 733 / 57,774 12.1 years | Medical records, pathology reports, National Death Index | FFQ, **Fruit juices** | Incidence, Colorectal cancer | >0.43 vs <0.15 FPED cup equivalents /1000 kcal | 0.98 (0.82 - 1.18) Ptrend:0.79 | ✓ | ✓ | ✓ | ✓ | ✓ | | ✓ E | ✓ | | | ✓ | |

| Author, Year Country (PMID) | Study name, type Age, sex, ethnicity, characteristics of participants | Cases/ Study size Median follow-up (years) | Case ascertainment | Exposure Assessment, detail | Outcome Sub-population | Comparison | RR (95%CI) Ptrend | Adjustments: Age | Sex | Smo | Alc | Obe | Phy | Diet | Med | Mens | Repr | Oth | Skin |
|---|---|---|---|---|---|---|---|---|---|---|---|---|---|---|---|---|---|---|---|
| Hur 2021 USA (33958435)[72] | Nurses' Health Study II<br>Prospective Cohort<br>F<br>nurses | 109 / 95,464<br>24 years | Medical records and National Death Index | Semi-quantitative FFQ,<br>**Fruit juices**:<br>Fruit juice included apple juice, orange juice, grapefruit juice, prune juice, and other fruit juices | Incidence, Early-onset colorectal cancer | ≥2 servings/day vs <1 serving/week | 1.13 (0.15 - 8.60)<br>Ptrend:0.63 | | | ✓ | ✓ | ✓ | ✓ | ✓ * | ✓ | ✓ | ✓ | ✓ | |
| | | | | | | Per 1 serving day | 1.17 (0.72 - 1.90) | | | ✓ | ✓ | ✓ | ✓ | ✓ * | ✓ | ✓ | ✓ | ✓ | |
| Jin 2023 UK (38004195)[78] | UK Biobank<br>Prospective Cohort<br>Age:55.87 years<br>M/F<br>Mainly white | 842 / 118,210<br>12.8 years | hospital records, cancer registry, death registry | 24 Hour Diet Recall,<br>**Fruit juices** | Incidence, Colon cancer | Per 1 SD day | 1.03 (0.97 - 1.11) | ✓ | ✓ | ✓ | | ✓ | ✓ | ✓ E | ✓ D | | | ✓ | |
| | | 359 / 118,210<br>12.8 years | | | Incidence, Rectal cancer | | 0.95 (0.85 - 1.06) | ✓ | ✓ | ✓ | | ✓ | ✓ | ✓ E | ✓ D | | | ✓ | |
| Voorrips 2000 Netherlands (11117618)[165] | The Netherlands Cohort study<br>Case Cohort<br>Range:55-69 years<br>M/F | 313 / 1,525<br>6.3 years | Cancer registry | Semi-quantitative FFQ,<br>**Citrus juice**<br>Orange / grapefruit juice (processed) | Incidence, Colon cancer, male | Per 25 g day | 1.00 | ✓ | | | ✓ | | | ✓ | | | | ✓ | |
| | | 274 / 1,598 | | | Incidence, Colon cancer, female | | 0.99 | ✓ | | | ✓ | | | ✓ | | | | ✓ | |
| | | 201 / 1,525 | | | Incidence, Rectal cancer, male | | 0.87 | ✓ | | | ✓ | | | ✓ | | | | ✓ | |
| | | 122 / 1,598 | | | Incidence, Rectal cancer, female | | 1.07 | ✓ | | | ✓ | | | ✓ | | | | ✓ | |
| | | 313 / 1,525 | | Semi-quantitative FFQ,<br>**Other fruit juice** | Incidence, Colon cancer, male | Per 25 g day | 1.04 | ✓ | | | ✓ | | | ✓ | | | | ✓ | |
| | | 274 / 1,598 | | | Incidence, Colon cancer, female | | 1.00 | ✓ | | | ✓ | | | ✓ | | | | ✓ | |
| | | 201 / 1,525 | | | Incidence, Rectal cancer, male | | 0.93 | ✓ | | | ✓ | | | ✓ | | | | ✓ | |
| | | 122 / 1,598 | | | Incidence, Rectal cancer, female | | 0.87 | ✓ | | | ✓ | | | ✓ | | | | ✓ | |
| Chazelas 2019 France (31292122)[44] | NutriNet-Santé study<br>Prospective Cohort<br>Age:42.2 years<br>M/F | 166 / 101,257<br>5.1 years | self-report, medical records reviewed by physicians, health insurance database, mortality registry | 24 Hour Diet Recall,<br>Pure **fruit juices**:<br>100% fruit juices | Incidence, Colorectal cancer | Per 100 ml day | 1.06 (0.76 - 1.49) | ✓ | ✓ | ✓ | ✓ | ✓ | ✓ | ✓ E | D | | | ✓ | |
| Arthur 2021 Canada (33220638)[38] | Canadian Study of Diet, Lifestyle and Health<br>Case Cohort<br>M/F | 247 / 2,633<br>15 years | Cancer registry and national death index | Self-administered Questionnaire,<br>Pure **fruit juices**: | Incidence, Colorectal cancer | >6.2 vs ≤1.7 cups/week | 1.16 (0.81 - 1.67)<br>Ptrend:0.548 | ✓ | | ✓ | ✓ | ✓ | ✓ | ✓ | | | | ✓ | |
| | | 192 / 2,633<br>15 years | | | Incidence, Colon cancer | | 1.15 (0.76 - 1.73)<br>Ptrend:0.528 | ✓ | | ✓ | ✓ | ✓ | ✓ | ✓ | | | | ✓ | |
| Iso 2007 Japan (18260705)[76] | Japan Collaborative Cohort study for Evaluation of Cancer Risk<br>Prospective Cohort<br>Range:40-79 years<br>M/F | 190 / 105,500<br>15 years | municipal resident registration records, death certificates | FFQ,<br>Pure **fruit juices**:<br>fresh fruit juice (in summer season) | Mortality, Colon cancer, male | ≥3 vs <1 times/week | 1.03 (0.69 - 1.54) | ✓ | | | | | | | | | | | |

| Author, Year Country (PMID) | Study name, type Age, sex, ethnicity, characteristics of participants | Cases/ Study size Median follow-up (years) | Case ascertainment | Exposure Assessment, detail | Outcome Sub-population | Comparison | RR (95%CI) Ptrend | Adjustments | | | | | | | | | | | |
|---|---|---|---|---|---|---|---|---|---|---|---|---|---|---|---|---|---|---|---|
| | | | | | | | | Age | Sex | Smo | Alc | Obe | Phy | Diet | Med | Mens | Repr | Oth | Skin |
| | | 189 / 105,500 15 years | | | Mortality, Colon cancer, female | | 0.92 (0.65 - 1.30) | ✓ | | | | | | | | | | | |
| | | 150 / 105,500 15 years | | | Mortality, Rectal cancer, male | | 0.69 (0.43 - 1.11) | ✓ | | | | | | | | | | | |
| | | 72 / 105,500 15 years | | | Mortality, Rectal cancer, female | | 0.86 (0.44 - 1.67) | ✓ | | | | | | | | | | | |
| Papadimitriou 2022 multi-national (33901663)[136] | European Prospective Investigation into Cancer and nutrition, and Netherlands Cohort Study on Diet and Cancer Prospective Cohort M/F | 5,069 / 394,288 14.1 years | population-based cancer registries, insurance records, active follow up | FFQ-Semi-quantitative, **Fruit and vegetable juices** | Incidence, Colorectal cancer, EPIC | Per 1 SD day | 1.00 (0.96 - 1.03) | ✓ | ✓ | ✓ | | ✓ | ✓ | ✓ E | D | | | ✓ | |
| Kanehara 2024 USA (38795743)[81] | Multiethnic Cohort Prospective Cohort Range:45-75 years M/F Mixed | 4,403 / 192,651 19.3 years | cancer registries | Quantitative FFQ, **Fruit juices**: Orange or grapefruit juice (not orange drinks or orange soda); tomato or V-8 juice; other fruit juices or fruit drinks. | Incidence, Colorectal cancer | Median intake 108.3 vs 0.1 g/1000 kcal | 1.01 (0.91 - 1.11) Ptrend:0.74 | ✓ | ✓ | ✓ | ✓ | ✓ | ✓ | ✓ E | ✓ D | | ✓ | ✓ | |
| Steinmetz 1994 USA (8296768)[158] | Iowa Women's Health Study Prospective Cohort Age:62 years F 99% white Postmenopausal | 212 / 35,216 167447 person-years | Driving license | Semi-quantitative FFQ, **Fruit and vegetable juices**: Apple Juice or cider, Apricot juice, Carrot juice, Celery juice, Grapefruit juice, Mango juice, orange juice, Pineapple juice, Prune juice, Tomato juice, V-8 juice and other fruit juices | Incidence, Colon cancer, | >7.8 vs <1.4 serving/week | 1.00 (0.68 - 1.48) | ✓ | | | | | | ✓ E | | | | | |
| Kanehara 2024 USA (38795743)[81] | Multiethnic Cohort Prospective Cohort Range:45-75 years M/F Mixed | 3,351 / 192,651 19.3 years | cancer registries | Quantitative FFQ, **Fruit juices**: Orange or grapefruit juice (not orange drinks or orange soda); tomato or V-8 juice; other fruit juices or fruit drinks. | Incidence, Colon cancer | Median intake 108.3 vs 0.1 g/1000 kcal | 1.08 (0.96 - 1.20) Ptrend:0.36 | ✓ | ✓ | ✓ | ✓ | ✓ | ✓ | ✓ E | ✓ D | | ✓ | ✓ | |
| | | 2,074 / 192,651 19.3 years | | | Incidence, Proximal colon cancer | | 1.03 (0.89 - 1.18) Ptrend:0.70 | ✓ | ✓ | ✓ | ✓ | ✓ | ✓ | ✓ E | ✓ D | | ✓ | ✓ | |
| | | 1,173 / 192,651 19.3 years | | | Incidence, Distal colon cancer | | 1.24 (1.03 - 1.49) Ptrend:0.12 | ✓ | ✓ | ✓ | ✓ | ✓ | ✓ | ✓ E | ✓ D | | ✓ | ✓ | |
| | | 1,014 / 192,651 19.3 years | | | Incidence, Rectal cancer | | 0.79 (0.64 - 0.97) Ptrend:0.02 | ✓ | ✓ | ✓ | ✓ | ✓ | ✓ | ✓ E | ✓ D | | ✓ | ✓ | |
| Steinmetz 1994 USA (8296768)[158] | Iowa Women's Health Study Prospective Cohort Age:62 years F 99% white Postmenopausal | 86 / 35,216 167447 person-years | Driving license | Semi-quantitative FFQ, **Fruit and vegetable juices**: Apple Juice or cider, Apricot juice, Carrot juice, Celery juice, Grapefruit juice, Mango juice, orange juice, Pineapple juice, Prune juice, Tomato juice, V-8 juice and other fruit juices | Incidence, Proximal colon cancer, | >7.8 vs <1.4 serving/week | 0.73 (0.40 - 1.33) | ✓ | | | | | | ✓ E | | | | | |
| | | 120 / 35,216 | | | Incidence, Distal colon cancer | >7.8 vs <1.4 serving/week | 1.22 (0.71 - 2.10) | ✓ | | | | | | ✓ E | | | | | |
| Jiang 2023 USA (37637049)[77] | Prostate, Lung, Colorectal and Ovarian Cancer Screening trial Prospective Cohort Age:65 years M/F Mostly white | / 101,680 9.4 years | Medical records + national death index | Self-administered Questionnaire, **Tomato juices**: tomato juice | Incidence, Colorectal cancer | ≥ -2.968 vs ≤ -19.516 g/day | 1.03 (0.84 - 1.26) Ptrend:0.612 | ✓ | ✓ | ✓ | ✓ | ✓ | | ✓ E | ✓ D | | | ✓ | |

| Author, Year Country (PMID) | Study name, type Age, sex, ethnicity, characteristics of participants | Cases/ Study size Median follow-up (years) | Case ascertainment | Exposure Assessment, detail | Outcome Sub-population | Comparison | RR (95%CI) Ptrend | Adjustments | | | | | | | | | | | |
|---|---|---|---|---|---|---|---|---|---|---|---|---|---|---|---|---|---|---|---|
| | | | | | | | | Age | Sex | Smo | Alc | Obe | Phy | Diet | Med | Mens | Repr | Oth | Skin |
| | | / 101,680<br>9.4 years | | | | Per g /day | 1.02 (0.96 - 1.08) | ✓ | ✓ | ✓ | ✓ | ✓ | | ✓<br>E | ✓<br>D | | | ✓ | |
| | | 443 /<br>101,680<br>9.4 years | | | Mortality,<br>Colorectal<br>cancer | ≥ -2.968 vs ≤ -19.516 g/day | 1.08 (0.79 - 1.47)<br>Ptrend:0.746 | ✓ | ✓ | ✓ | ✓ | ✓ | | ✓<br>E | ✓<br>D | | | ✓ | |
| | | 443 /<br>101,680<br>9.4 years | | | | Per g /day | 1.03 (0.94 - 1.13) | ✓ | ✓ | ✓ | ✓ | ✓ | | ✓<br>E | ✓<br>D | | | ✓ | |
| **Liver** | | | | | | | | | | | | | | | | | | | |
| Kim 2023 USA<br>(37121551)[88] | Multiethnic Cohort Study<br>Prospective Cohort<br>M/F<br>Multi-ethnic | / 170,321<br>19.6 years | cancer registry, death certificate and national death index | Quantitative FFQ,<br>**Fruit juices**:<br>fruit juices as a component of the Overall plant-based diet index | Incidence,<br>Hepatocellular<br>carcinoma | Per 1 point increase in the component score of the overall plant-based diet | 1.04 (0.99 - 1.10) | ✓ | ✓ | ✓ | ✓ | ✓ | | ✓<br>E | D | | | ✓ | |
| | | | | | | Per 1 point increase in the component score of the healthful plant-based diet | 0.96 (0.91-1.01) | ✓ | ✓ | ✓ | ✓ | ✓ | | ✓<br>E | D | | | ✓ | |
| | | | | | | Per 1 point increase in the component score of the unhealthful plant-based diet | 1.04 (0.99-1.09) | ✓ | ✓ | ✓ | ✓ | ✓ | | ✓<br>E | D | | | ✓ | |
| Zhang 2023 USA<br>(37460062)[183] | Women's Health Initiative<br>Prospective Cohort<br>Age:63.4 years<br>F<br>Mainly white<br>Postmenopausal | 176 /<br>78,356<br>22.1 years | Self-report adjudicated by physician rv of medical records | Validated FFQ,<br>**Fruit juices** | Incidence,<br>Liver cancer | Per 1 serving day | 0.79 (0.60 - 1.04) | ✓ | | ✓ | ✓ | ✓ | ✓ | ✓<br>E | ✓<br>D | | ✓ | ✓ | |
| Jones 2022 USA<br>(35728406)[79] | NIH-AARP Diet and Health Study + Prostate, Lung, Colorectal and Ovarian Cancer Screening trial<br>Pooled Analysis Cohorts<br>M/F<br>Mainly white | 587 /<br>553,874 | NIH-AARP: cancer reg,<br>PLCO: self-report + medical record/cancer reg | Questionnaire,<br>**Citrus juice**<br>Orange / grapefruit juice | Incidence,<br>Liver cancer,<br>no diabetes & ≤12 yrs follow-up | Per times /day | 0.77 (0.65 - 0.92) | ✓ | ✓ | ✓ | ✓ | ✓ | | ✓<br>E | | | | ✓ | |
| | | 252 /<br>553,874 | | | Incidence,<br>Liver cancer,<br>no diabetes & >12 yrs follow-up | | 1.02 (0.82 - 1.27) | ✓ | ✓ | ✓ | ✓ | ✓ | | ✓<br>E | | | | ✓ | |
| Zhao 2023 USA<br>(36811575)[185] | NIH- AARP Diet and Health Study<br>Prospective Cohort<br>M/F<br>Mainly white<br>Retired | 947 /<br>485,403<br>15.5 years | active follow up and cancer registry | Semi-quantitative FFQ,<br>**Other fruit juice**:<br>all other fruit juices excluding orange/grapefruit juice | Incidence,<br>Liver cancer | Q5 vs Q1 | 0.97 (0.80 - 1.17)<br>Ptrend:0.99 | ✓ | ✓ | ✓ | ✓ | ✓ | ✓ | ✓<br>E | D | | | ✓ | |

| Author, Year Country (PMID) | Study name, type Age, sex, ethnicity, characteristics of participants | Cases/ Study size Median follow-up (years) | Case ascertainment | Exposure Assessment, detail | Outcome Sub-population | Comparison | RR (95%CI) Ptrend | Adjustments Age | Sex | Smo | Alc | Obe | Phy | Diet | Med | Mens | Repr | Oth | Skin |
|---|---|---|---|---|---|---|---|---|---|---|---|---|---|---|---|---|---|---|---|
| Jones 2022 USA (35728406)[79] | NIH-AARP Diet and Health Study + Prostate, Lung, Colorectal and Ovarian Cancer Screening trial<br>Pooled Analysis Cohorts<br>M/F<br>Mainly white | 158 / 553,874 | NIH-AARP: cancer reg, PLCO: self-report + medical record/cancer reg | Questionnaire,<br>Other **juices**:<br>Juices excluding citrus and tomato | Incidence, Liver cancer, with diabetes & ≤12 yrs follow-up | Per time day | 0.95 (0.56 - 1.61) | ✓ | ✓ | ✓ | ✓ | ✓ | | ✓<br>E | | | | ✓ | |
| | | 63 / 553,874 | | | Incidence, Liver cancer, with diabetes & >12 yrs follow-up | | 1.23 (0.69 - 2.20) | ✓ | ✓ | ✓ | ✓ | ✓ | | ✓<br>E | | | | ✓ | |
| Iso 2007 Japan (18260705)[76] | Japan Collaborative Cohort study for Evaluation of Cancer<br>Prospective Cohort<br>Range:40-79 years<br>M/F | 412 / 105,500 | municipal resident registration records, death certificates | Validated FFQ,<br>**Pure fruit juice**<br>Fruit juices in summer season | Mortality, Liver cancer, male | ≥3 vs <1 times/week | 1.40 (1.05 - 1.85) | ✓ | | | | | | | | | | | |
| | | 195 / 105,500 | | | Mortality, Liver cancer, female | | 0.91 (0.61 - 1.36) | ✓ | | | | | | | | | | | |
| Stepien 2016 multi-national (25528243)[159] | European Prospective Investigation into Cancer and nutrition cohort<br>Prospective Cohort<br>Mainly white | 140 / 477,206<br>11.4 years | cancer registries | ,<br>Fruit and vegetable **juices**:<br>fruit, citrus and/or vegetable juices (including fresh and commercial juices) and nectars (up to 20% added sugar) | Incidence, HCC | >6 vs <1 glass/week | 1.38 (0.80 - 2.38)<br>Ptrend:0.02 | | | ✓ | ✓ | ✓ | ✓ | ✓<br>E | D | | | ✓ | |
| Jones 2022 USA (35728406)[79] | NIH-AARP Diet and Health Study + Prostate, Lung, Colorectal and Ovarian Cancer Screening trial<br>Pooled Analysis Cohorts<br>M/F<br>Mainly white | 587 / 553,874 | NIH-AARP: cancer reg, PLCO: self-report + medical record/cancer reg | Questionnaire,<br>**Vegetable juices**:<br>Tomato/vegetable juice. | Incidence, Liver cancer, no diabetes & ≤12 yrs follow-up | Per time day | 1.19 (0.89 - 1.59) | ✓ | ✓ | ✓ | ✓ | ✓ | | ✓<br>E | | | | ✓ | |
| | | 252 / 553,874 | | | Incidence, Liver cancer, no diabetes & >12 yrs follow-up | | 1.01 (0.58 - 1.77) | ✓ | ✓ | ✓ | ✓ | ✓ | | ✓<br>E | | | | ✓ | |
| **Gallbladder/bile ducts** | | | | | | | | | | | | | | | | | | | |
| Iso 2007 Japan (18260705)[76] | Japan Collaborative Cohort study for Evaluation of Cancer Risk<br>Prospective Cohort<br>Range:40-79 years<br>M/F | 63 / 105,500<br>15 years | municipal resident registration records, death certificates | Validated FFQ,<br>**Pure fruit juices**:<br>Fresh fruit juice in summer season | Mortality, Gallbladder cancer, male | ≥3 vs <1 times/week | 0.49 (0.21 - 1.18) | ✓ | | | | | | | | | | | |
| | | 82 / 105,500<br>15 years | | | Mortality, gallbladder cancer, female | | 1.44 (0.75 - 2.73) | ✓ | | | | | | | | | | | |
| Stepien 2016 multi-national (25528243)[159] | European Prospective Investigation into Cancer and nutrition cohort<br>Prospective Cohort<br>Mainly white | 87 / 477,206<br>11.4 years | cancer registries | Semi-quantitative FFQ,<br>**Juices**:<br>fruit, citrus and/or vegetable juices (including fresh and commercial juices) and nectars (up to 20% added sugar) | Incidence, Gallbladder cancer | Per 1 serving week | 0.97 (0.91 - 1.04) | ✓ | ✓ | ✓ | ✓ | ✓ | ✓ | ✓<br>E | ✓ | | | ✓ | |
| | | 54 / 477,206<br>11.4 years | | | Incidence, Ampulla of Vater cancer | | 0.97 (0.88 - 1.08) | ✓ | ✓ | ✓ | ✓ | ✓ | ✓ | ✓<br>E | ✓ | | | ✓ | |
| | | 95 / 477,206<br>11.4 years | | | Incidence, Extrahepatic | | 1.01 (0.96 - 1.06) | ✓ | ✓ | ✓ | ✓ | ✓ | ✓ | ✓<br>E | ✓ | | | ✓ | |

| Author, Year Country (PMID) | Study name, type Age, sex, ethnicity, characteristics of participants | Cases/ Study size Median follow-up (years) | Case ascertainment | Exposure Assessment, detail | Outcome Sub-population | Comparison | RR (95%CI) Ptrend | Adjustments | | | | | | | | | | | |
|---|---|---|---|---|---|---|---|---|---|---|---|---|---|---|---|---|---|---|---|
| | | | | | | | | Age | Sex | Smo | Alc | Obe | Phy | Diet | Med | Mens | Repr | Oth | Skin |
| | | | | | bile duct system cancer | | | | | | | | | | | | | | |
| | | 66 / 477,206 11.4 years | | | Incidence, Intrahepatic bile duct cancer | | 1.04 (1.00 - 1.08) | | | ✓ | ✓ | ✓ | ✓ | ✓ E | D | | | ✓ | |
| **Pancreas** | | | | | | | | | | | | | | | | | | | |
| Bobe 2008 Finland (18349272)[41] | Alpha-Tocopherol, Beta-Carotene Cancer Prevention Study Prospective Cohort Age:57 years M | 306 / 27,111 16 years | Cancer registry | , **Fruit juices**: (reference https://doi.org/10.1021/jf00032a015) 100 % tomato juice, orange juice, apple juice and grape juice. Fresh orange juice, lemon juice, grapefruit juice | Incidence, Pancreatic Adenocarcinoma | >121.5 vs 0 g | 0.84 (0.60 - 1.17) Ptrend:0.16 | ✓ | | ✓ | | | | ✓ E | D | | | | |
| Vrieling 2009 Denmark,France,Germany,Greece,Italy,Netherlands,Norway,Spain,Sweden,UK (19107929)[167] | European Prospective Investigation into Cancer and nutrition cohort Prospective Cohort Age:51 years M/F | 555 / 478,400 8.9 years | cancer registry, record linkage, health insurance rec, pathology and active follow up | Semi-quantitative FFQ, **Fruit juices**: Juices from fresh fruits, hard fruits(apples, pears), stone fruits(peaches, cherries), grapes, berry fruits(strawberries, raspberries), other non-specified fruits(kiwis, bananas) | Incidence, Exocrine cancer of the pancreas | >68 vs 0 g/day | 0.79 (0.47 - 1.33) Ptrend:0.12 | ✓ | ✓ | ✓ | | ✓ | | ✓ E | D | | | | |
| | | | | | | Per 25 g | 0.92 (0.85 - 1.01) | ✓ | ✓ | ✓ | | ✓ | | ✓ E | D | | | | |
| Nöthlings 2007 USA (17991664 )[131] | Multiethnic Cohort Prospective Cohort Age:60 years M/F Multi-ethnic | 434 / 162,150 8 years | Cancer registry and national death index | FFQ, **Fruit juices** | Incidence, Exocrine cancer of the pancreas | ≥60 vs <4.7 g/1000 kcal/day | 1.08 (0.83 - 1.41) Ptrend:0.56 | ✓ | ✓ | ✓ | | ✓ | | ✓ E | | | | ✓ | |
| Zhong 2023 USA (36863825)[188] | Prostate, Lung, Colorectal and Ovarian Cancer Screening trial Prospective Cohort M/F Mostly white | / 101,748 8.86 years | Annual study update form, medical records, death certificates and family reports. | FFQ, **Fruit juices**: energy adjusted consumption of each food group of plant-based diet index | Incidence, Pancreas cancer | Q4 vs Q1 | 1.76 (1.29 - 2.40) Ptrend:0.005 | ✓ | ✓ | ✓ | ✓ | ✓ | | ✓ * | D | | | ✓ | |
| Shigihara 2014 Japan (24522236)[154] | Ohsaki National Health Insurance (NHI) Cohort Study Prospective Cohort Range:40-79 years | 67 / 32,859 11 years | Cancer registry | Self-administered Questionnaire, Pure **fruit juices**: fresh fruit juices | Incidence, Pancreas cancer, male | ≥225.3 vs <126.7 g/day | 1.34 (0.72 - 2.50) Ptrend:0.33 | ✓ | | ✓ | ✓ | ✓ | ✓ | ✓ E | D | | | ✓ | |
| Lin 2006 Japan (17176216)[103] | Japan Collaborative Cohort study for Evaluation of Cancer Risk Prospective Cohort Age:58 years M/F | 300 / 105,438 | Death certificate | Dietary Recall, **Pure fruit juices**: Fresh fruit juice | Mortality, Pancreatic cancer, male | Almost every day vs 0 - 2 times/month | 0.33 (0.13 - 0.84) Ptrend:0.01 | ✓ | | ✓ | | | | | | | | ✓ | |
| | | | | | Mortality, Pancreatic cancer, female | | 1.09 (0.62 - 1.90) Ptrend:0.55 | ✓ | | ✓ | | | | | | | | ✓ | |
| Vrieling 2009 Denmark,France,Germany,Greece,Italy,Netherlands,Norway,Spain,Sweden,UK (19107929)[167] | European Prospective Investigation into Cancer and nutrition cohort Prospective Cohort Age:51 years M/F | 555 / 478,400 8.9 years | cancer registry, record linkage, health insurance rec, pathology and active follow up | Semi-quantitative FFQ, **Citrus fruit juice** | Incidence, Exocrine cancer of the pancreas | Per 25 g | 0.97 (0.93 - 1.01) | ✓ | ✓ | ✓ | | ✓ | | ✓ E | D | | | | |
| | | | | | | | 0.94 (0.88 - 1.01) Ptrend:0.10 | ✓ | ✓ | ✓ | | ✓ | | ✓ | | | | | |

| Author, Year Country (PMID) | Study name, type Age, sex, ethnicity, characteristics of participants | Cases/ Study size Median follow-up (years) | Case ascertainment | Exposure Assessment, detail | Outcome Sub-population | Comparison | RR (95%CI) Ptrend | Adjustments | | | | | | | | | | | |
|---|---|---|---|---|---|---|---|---|---|---|---|---|---|---|---|---|---|---|---|
| | | | | | | | | Age | Sex | Smo | Alc | Obe | Phy | Diet | Med | Mens | Repr | Oth | Skin |
| | | | | | | | | | | | | | | E | D | | | | |
| Yamagiwa 2018 Japan (30255932)[176] | Japan Public Health Center-based Prospective Study<br>Prospective Cohort<br>Range:40-69 years<br>M/F | / 90,185<br>16.9 years | | FFQ,<br>**Pure apple juice**:<br>100% apple juice | Incidence, Pancreatic cancer | Median intake 49 vs 0 g/day | 0.89 (0.72 - 1.09)<br>Ptrend:0.27 | ✓ | ✓ | ✓ | ✓ | ✓ | ✓ | ✓<br>E | D | | | ✓ | |
| | | | | FFQ,<br>**Pure orange juice**:<br>100% Orange juice | | | 0.94 (0.77 - 1.15)<br>Ptrend:0.616 | ✓ | ✓ | ✓ | ✓ | ✓ | ✓ | ✓<br>E | D | | | ✓ | |
| Mueller 2010 Singapore (20142243)[125] | Singapore Chinese Health study<br>Prospective Cohort<br>Age:56 years<br>M/F<br>Singapore Chinese | 140 /<br>60,524<br>10.7 years | Cancer registry and death registry | FFQ,<br>**Fruit and vegetable juices**:<br>orange juice, sugarcane juice, honeydew melon juice, apple juice, watermelon juice, carrot juice, pineapple juice, star fruit juice, lemon juice. Canned grape, tomato, prune , papaya, plum juice and fresh celery juice. | Incidence / Mortality, Pancreatic Adenocarcinoma | ≥2 vs <2 servings/week | 1.32 (0.75 - 2.30)<br>Ptrend:0.34 | ✓ | ✓ | ✓ | ✓ | | ✓ | ✓<br>E | | | | ✓ | |
| Navarrete-Muñoz 2016 multi-national (27510540)[127] | European Prospective Investigation into Cancer and nutrition<br>Prospective Cohort<br>Age:51 years<br>M/F<br>Mostly white<br>generally healthy adults | 865 /<br>477,199<br>11.6 years | cancer and pathology registry, active follow up, health insurance record, mortality registry and contact of participants or next-of-kin | Semi-quantitative FFQ,<br>**Juices**:<br>Juices (obtained from either 100% fruit and vegetables or concentrates) and nectars (juices with ≤20% added sugar) | Incidence / Mortality, Pancreas cancer | >123.11 vs 0.1 - 8.3 g/day | 0.75 (0.56 - 0.98)<br>Ptrend:0.040 | ✓ | ✓ | ✓ | ✓ | | ✓ | ✓<br>* | | | | ✓ | |
| | | | | | | Per 100 g /day | 0.91 (0.84 - 0.99) | ✓ | ✓ | ✓ | ✓ | | ✓ | ✓<br>* | | | | ✓ | |
| Yamagiwa 2018 Japan (30255932)[176] | Japan Public Health Center-based Prospective Study<br>Prospective Cohort<br>Range:40-69 years<br>M/F | / 90,185<br>16.9 years | | FFQ,<br>**Tomatoes juice**:<br>100% Tomato juice | Incidence, Pancreatic cancer | Median intake 41 vs 0 g/day | 0.98 (0.79 - 1.21)<br>Ptrend:0.904 | ✓ | ✓ | ✓ | ✓ | ✓ | ✓ | ✓<br>E | D | | | ✓ | |
| **Lung** | | | | | | | | | | | | | | | | | | | |
| Ozasa, 2001 Japan (11749690)[134] | Japan Collaborative Cohort Study<br>Prospective Cohort<br>Range:40-70 years<br>M/F | 283 /<br>42,940<br>7.7 years | Unknown/NA | FFQ - Study-specific,<br>**Fruit juices**: | Mortality, Lung cancer, male | 12-16 vs <1-2 times/month | 0.90 (0.69 - 1.18) | ✓ | | ✓ | | | | | | | | ✓ | |
| | | 75 / 55,308<br>7.7 years | | | Mortality, lung cancer, female | | 0.95 (0.56 - 1.63) | ✓ | | ✓ | | | | | | | | ✓ | |
| Linseisen 2007 France, Italy, Spain, UK, Netherlands, Greece, Germany, Sweden, Denmark, Norway (17487840)[104] | European Prospective Investigation into Cancer and nutrition<br>Prospective Cohort<br>Range:25-70 years<br>M/F | 938 /<br>478,590<br>6.4 years | cancer registries, health insurance records, pathology rec, active follow up, death certificate | FFQ, Dietary Questionnaires, Food Record,<br>**Citrus fruit juice**: | Incidence, Lung cancer | Mean intake 106.6 vs 12.9 g/da | 0.82 (0.65 - 1.03) | | | ✓ | ✓ | ✓ | ✓ | ✓<br>E | | | | ✓ | |
| | | | | | | Per 100 g | 1.00 (0.87 - 1.14) | | | ✓ | ✓ | ✓ | ✓ | ✓<br>E | | | | ✓ | |
| Smith-Warner 2003 multi-national (14601062)[155] | Pooling Project (n=8 cohorts)<br>Prospective Cohort<br>M/F | 2,839 /<br>280,419<br>16 years | Self-report, medical record, cancer registries and death registries | FFQ,<br>**Citrus juice**:<br>Orange & grapefruit juice. | Incidence, Lung cancer | ≥1/2 serving/day vs 0 serving/week | 0.82 (0.71 - 0.94)<br>Ptrend:0.01 | | | ✓ | ✓ | ✓ | | ✓<br>E | | | | ✓ | |
| Voorrips, 2000 Netherlands (10710193)[166] | Netherlands Cohort Study on Diet and Cancer<br>Case Cohort | 963 /<br>120,852<br>3.2 years | Area residency lists | FFQ - Study-specific,<br>Other **fruit juices**:<br>Fruit juices excluding citrus | Incidence, Lung cancer, | 20 vs 0 g/day | 0.90 (0.70 - 1.20) | ✓ | ✓ | ✓ | | | | ✓ | | | | ✓ | |

| Author, Year Country (PMID) | Study name, type Age, sex, ethnicity, characteristics of participants | Cases/ Study size Median follow-up (years) | Case ascertainment | Exposure Assessment, detail | Outcome Sub-population | Comparison | RR (95%CI) Ptrend | Adjustments | | | | | | | | | | | |
|---|---|---|---|---|---|---|---|---|---|---|---|---|---|---|---|---|---|---|---|
| | | | | | | | | Age | Sex | Smo | Alc | Obe | Phy | Diet | Med | Mens | Repr | Oth | Skin |
| | Range:55-69 years<br>M/F | | | | | | | | | | | | | | | | | | |
| Feskanich, 2000 USA (11078758)[55] | Nurses' Health Study + Health Professionals Follow-up Study<br>Pooled Analysis Cohorts<br>Range:30-75 years<br>M/F | 519 / 77,283<br>12 years | By post | FFQ - Study-specific,<br>**Grapefruit juice**:<br>Grapefruit juice | Incidence, Lung cancer, female | Per 30 servings /month | 0.61 (0.31 - 1.01) | ✓ | | ✓ | | | | ✓<br>E | | | | | |
| | | 274 / 47,778<br>12 years | | | Incidence, Lung cancer, male | Per 30 servings/ month | | ✓ | | ✓ | | | | ✓<br>E | | | | | |
| Iso 2007 Japan (18260705)[76] | Japan Collaborative Cohort study for Evaluation of Cancer Risk<br>Prospective Cohort<br>Range:40-79 years<br>M/F | 772 / 105,500<br>15 years | municipal resident registration records, death certificates | Validated FFQ,<br>**Pure fruit juices**:<br>Fresh fruit juice (in summer season) | Mortality, Lung cancer, male | ≥3 vs <1 times/week | 0.87 (0.72 - 1.07) | ✓ | | | | | | | | | | | |
| | | 229 / 105,500<br>15 years | | | Mortality, Lung cancer, female | | 1.01 (0.70 - 1.46) | ✓ | | | | | | | | | | | |
| Gnagnarella 2013 Italy (23956193)[65] | Continuous Observation of Smoking Subjects Study (COSMOS)<br>Prospective Cohort<br>Age:57 years | 178 / 4,336<br>5.7 years | screening examinations | FFQ,<br>**Fruit and vegetable juices**:<br>Fruit and vegetable juices (no detailed information) | Incidence, Lung cancer | 62.46 vs 0 g/1000 kcal/day | 0.96 (0.66 - 1.39)<br>Ptrend:0.7452 | ✓ | ✓ | ✓ | | | | ✓<br>E | | | | | |
| Linseisen 2007 France, Italy, Spain, UK, Netherlands, Greece, Germany, Sweden, Denmark, Norway (17487840)[104] | European Prospective Investigation into Cancer and nutrition<br>Prospective Cohort<br>Range:25-70 years<br>M/F | 1,126 / 478,590<br>6.4 years | cancer registries, health insurance records, pathology rec, active follow up, death certificate | FFQ, Dietary Questionnaires, Food Record,<br>**Fruit and vegetable juices**:<br>Total juices (juices from fruits, vegetables and citrus fruits)<br>-->fruits: hard fruits (apples, pears) and fruits containing stones<br>-->vegetables: tomato, pepper, courgette, pumpkin, eggplant, French bean, leafy vegetables, root vegetables, cabbages and onions and garlic<br>-->citrus fruits | Incidence, Lung cancer | Mean intake 182.2 vs 21.5 g/day | 0.89 (0.72 - 1.10) | | | ✓ | ✓ | ✓ | ✓ | ✓<br>E | | | | ✓ | |
| | | | | | | Per 100 g | 0.97 (0.88 - 1.07) | | | ✓ | ✓ | ✓ | ✓ | ✓<br>E | | | | ✓ | |
| **Skin** | | | | | | | | | | | | | | | | | | | |
| Mahamat-Saleh 2020 Denmark,France,Germany,Greece,Italy,Netherlands,Norway,Spain,Sweden,UK (32710289)[107] | European Prospective Investigation into Cancer and nutrition<br>Prospective Cohort<br>M/F | 5,604 / 270,112<br>13.7 years | cancer and mortality registries, health insurance & pathology records, active follow up | Dietary Questionnaire,<br>**Citrus fruit juice**: | Incidence, Basal Cell Carcinoma | Mean intake 117.7 vs 0.01 g/day | 1.10 (1.01 - 1.19)<br>Ptrend:0.008 | ✓ | ✓ | ✓ | ✓ | ✓ | ✓ | ✓<br>E<br>* | | | | ✓ | |
| Melough 2021 USA (32791010)[115] | NIH- AARP Diet and Health Study<br>Prospective Cohort<br>Age:62 years<br>M/F | / 388,467<br>15.5 years | cancer registries | FFQ,<br>Citrus **juice**:<br>citrus juice as orange juice or grapefruit juice | Incidence, Cutaneous malignant melanoma | 1+ cups/day vs none | 1.02 (0.87 - 1.19)<br>Ptrend:0.9 | ✓ | ✓ | ✓ | ✓ | ✓ | ✓ | | | | | ✓ | ✓ |
| Mahamat-Saleh 2020 Denmark,France,Germany,Greece,Italy,Netherlands,Norway,Spain | European Prospective Investigation into Cancer and nutrition<br>Prospective Cohort<br>M/F | 1,371 / 270,112<br>13.7 years | cancer and mortality registries, health insurance & | Dietary Questionnaire,<br>**Citrus fruit juice**: | Incidence, Melanoma | Mean intake 117.7 vs 0.01 g/day | 0.92 (0.78 - 1.08)<br>Ptrend:0.31 | ✓ | ✓ | ✓ | ✓ | ✓ | ✓ | ✓<br>E<br>* | | | | ✓ | |

| Author, Year Country (PMID) | Study name, type Age, sex, ethnicity, characteristics of participants | Cases/ Study size Median follow-up (years) | Case ascertainment | Exposure Assessment, detail | Outcome Sub-population | Comparison | RR (95%CI) Ptrend | Adjustments | | | | | | | | | | | |
|---|---|---|---|---|---|---|---|---|---|---|---|---|---|---|---|---|---|---|---|
| | | | | | | | | Age | Sex | Smo | Alc | Obe | Phy | Diet | Med | Mens | Repr | Oth | Skin |
| n,Sweden,UK (32710289)[107] | | | pathology records, active follow up | | | | | | | | | | | | | | | | |
| Marley 2021 UK (33782946)[110] | UK Biobank Prospective Cohort Age:56.2 years M/F White | 1592 / 198,964 | ICD codes via national registries | 24 Hour Recall Frequency Questionnaire, **Citrus fruit juice**: (orange juice, grapefruit juice) | Incidence, Melanoma | >1 serving vs none | 1.43 (1.05 - 1.90) Ptrend:0.093 | | ✓ | ✓ | ✓ | ✓ | ✓ | ✓ E | ✓ | | | ✓ | ✓ |
| Melough 2020 USA (31335211)[116] | Women's Health Initiative Observational Study Prospective Cohort Age:63.4 years F Postmenopausal | 956 / 56,205 15.7 years | Self-report adjudicated by physician rv of medical records | FFQ, **Citrus fruit juice**: | Incidence, Melanoma | medium serving size 1+ /day vs <1/week | 1.13 (0.96 - 1.32) Ptrend:0.11 | ✓ | | | ✓ | ✓ | ✓ | | ✓ | | | ✓ | ✓ |
| Marley 2022 UK (34282681)[111] | UK Biobank Prospective Cohort Range:40-69 years M/F | 9,570 / 197,372 | national cancer registers | 24 Hour Recall Frequency Questionnaire, **Citrus fruit juice**: orange and grapefruit juice | Incidence, non-Melanoma skin cancer | >1 serving vs none | 1.01 (0.87 - 1.16) Ptrend:0.74 | ✓ | ✓ | ✓ | ✓ | ✓ | ✓ | ✓ E | | | | ✓ | ✓ |
| Sakaki 2021 USA (33946526)[148] | Women's Health Initiative Observational Prospective Cohort Age:63 years F White non-Hispanic Postmenopausal | 8642 / 49,007 ~25 years | Self-report | FFQ, **Citrus fruit juice**: orange and grapefruit | Incidence, non-Melanoma skin cancer | ≥1 serving /day vs <1 serving/week | 1.08 (1.02 - 1.13) Ptrend:0.007 | ✓ | | ✓ | ✓ | ✓ | ✓ | ✓ E | | | ✓ | ✓ | ✓ |
| Mahamat-Saleh 2020 Denmark,France,Germany,Greece,Italy,Netherlands,Norway,Spain,Sweden,UK (32710289)[107] | European Prospective Investigation into Cancer and nutrition Prospective Cohort M/F | 8,448 / 270,112 13.7 years | cancer and mortality registries, health insurance & pathology records, active follow up | Dietary Questionnaire, **Citrus fruit juice**: | Incidence, Skin cancer | Mean intake 117.7 vs 0.01 g/day | 1.08 (1.02 - 1.16) Ptrend:0.004 | ✓ | ✓ | ✓ | ✓ | ✓ | ✓ | ✓ | | | | ✓ | |
| | | 1,165 / 270,112 13.7 years | | | Incidence, Squamous cell carcinoma | | 1.23 (1.05 - 1.44) Ptrend:0.004 | ✓ | ✓ | ✓ | ✓ | ✓ | ✓ | ✓ E | | | | ✓ | |
| Wu 2015 USA (26224304)[170] | Nurses' Health Study + Health Professionals Follow-up Study Prospective Cohort Range:30-75 years M/F Health professionals | 9,033 / 41,530 25 years | Self-report verified by medical record | FFQ, **Grapefruit juice**: Grapefruit juice | Incidence, Basal Cell Carcinoma, male | ≥3 vs never servings/week | 1.02 (0.94 - 1.12) Ptrend:0.85 | ✓ | | ✓ | ✓ | ✓ | | ✓ | | | | ✓ | ✓ |
| | | 11,807 / 63,759 25 years | | | Incidence, Basal Cell Carcinoma, female | | 1.02 (0.95 - 1.10) Ptrend:0.86 | ✓ | | ✓ | ✓ | ✓ | | ✓ | | ✓ | | ✓ | ✓ |
| Marley 2021 UK (33782946)[110] | UK Biobank Prospective Cohort Age:56.2 years M/F White | 9,570 / 198,964 | ICD codes via national registries | 24 Hour Recall Frequency Questionnaire, Grapefruit juice | Incidence, Melanoma | >1 serving vs None | 1.96 (0.48 - 5.23) Ptrend:0.6 | | ✓ | ✓ | ✓ | ✓ | ✓ | ✓ E | ✓ | | | ✓ | ✓ |
| Wu 2015 USA (26124488)[171] | Nurses' Health Study + Health Professionals Follow-up Study Pooled Analysis Cohorts M/F | 817 / 41,530 | self-report verified by pathology reports and medical records | Validated FFQ, **Grapefruit juice**: Grapefruit juice | Incidence, Melanoma, male | ≥ 2 vs never serving/week | 1.03 (0.82 - 1.28) Ptrend:0.86 | ✓ | | ✓ | ✓ | ✓ | ✓ | ✓ E | | | | ✓ | ✓ |

| Author, Year Country (PMID) | Study name, type Age, sex, ethnicity, characteristics of participants | Cases/ Study size Median follow-up (years) | Case ascertainment | Exposure Assessment, detail | Outcome Sub-population | Comparison | RR (95%CI) Ptrend | Adjustments: Age | Sex | Smo | Alc | Obe | Phy | Diet | Med | Mens | Repr | Oth | Skin |
|---|---|---|---|---|---|---|---|---|---|---|---|---|---|---|---|---|---|---|---|
| | | 1,023 / 63,759 | | | Incidence, melanoma, female | | 0.96 (0.79 - 1.16) Ptrend:0.58 | ✓ | | ✓ | ✓ | ✓ | ✓ | ✓ E | | ✓ | ✓ | ✓ | ✓ |
| Wu 2015 USA (26224304)[170] | Nurses' Health Study + Health Professionals Follow-up Study Prospective Cohort Range:30-75 years M/F Health professionals | 1,540 / 41,530 25 years | Self-report verified by medical record | FFQ, **Grapefruit juice**: Grapefruit juice | Incidence, Squamous cell carcinoma, male | ≥3 vs Never servings/week | 1.03 (0.84 - 1.27) Ptrend:0.46 | ✓ | | ✓ | ✓ | ✓ | | ✓ | | | | ✓ | ✓ |
| | | 2,004 / 63,759 25 years | | | Incidence, Squamous cell carcinoma, female | | 1.17 (0.98 - 1.39) Ptrend:0.71 | ✓ | | ✓ | ✓ | ✓ | | ✓ | | ✓ | | ✓ | ✓ |
| | | 2,758 / 105,289 25 years | | | Incidence, SCC on head, neck and extremities | ≥2 vs Never servings/week | 1.02 (0.91 - 1.15) Ptrend:0.89 | ✓ | | ✓ | ✓ | ✓ | | ✓ | | ✓ | | ✓ | ✓ |
| | | 483 / 105,289 25 years | | | Incidence, SCC on truncal sites | | 0.74 (0.54 - 1.02) Ptrend:0.18 | ✓ | | ✓ | ✓ | ✓ | | ✓ | | ✓ | | ✓ | ✓ |
| | | 9,033 / 41,530 25 years | | FFQ, **Orange juice**: | Incidence, Basal Cell Carcinoma, male | ≥1 servings/day vs <1 servings/week | 1.09 (1.02 - 1.16) Ptrend:0.002 | ✓ | | ✓ | ✓ | ✓ | | ✓ | | | | ✓ | ✓ |
| | | 11,807 / 63,759 25 years | | | Incidence, Basal Cell Carcinoma, female | | 1.16 (1.10 - 1.24) Ptrend:<0.0001 | ✓ | | ✓ | ✓ | ✓ | | ✓ | | ✓ | | ✓ | ✓ |
| Wu 2015 USA (26124488)[171] | Health Professionals Follow-up Study Pooled Analysis Cohorts M/F | 817 / 41,530 | self-report verified by pathology reports and medical records | Validated FFQ, **Orange juice**: Orange juice | Incidence, melanoma, male | ≥ 2 vs never serving/week | 1.25 (1.02 - 1.53) Ptrend:0.04 | ✓ | | ✓ | ✓ | ✓ | ✓ | ✓ E | | | | ✓ | ✓ |
| Feskanich, 2003 USA (12778065)[54] | The Nurses' Health Study Cohort Prospective Cohort Range:25-77 years F White | 503 / 162,078 | self-report verified by pathology reports and medical records | FFQ, **Citrus fruit juice**: Orange juice | Incidence, Melanoma | ≥1/day vs Never | 1.61 (0.92 - 2.84) Ptrend:0.008 | ✓ | | | | ✓ | | ✓ E | | ✓ | ✓ | ✓ | ✓ |
| Marley 2021 UK (33782946)[110] | UK Biobank Prospective Cohort Age:56.2 years M/F White | 1592 / 198,964 | ICD codes via national registries | 24 Hour Recall Frequency Questionnaire, **Orange juice**: | Incidence, Melanoma | >1 serving vs None | 1.54 (1.10 - 2.10) Ptrend:0.021 | | ✓ | ✓ | ✓ | ✓ | ✓ | ✓ E | ✓ | | | ✓ | ✓ |
| Marley 2022 UK (34282681)[111] | UK Biobank Prospective Cohort Range:40-69 years M/F | 9,389 / 197,372 | national cancer registers | 24 Hour Recall Frequency Questionnaire, **Orange juice**: | Incidence, non-Melanoma skin cancer | >1 serving vs None | 1.06 (0.90 - 1.24) Ptrend:0.91 | ✓ | ✓ | ✓ | ✓ | ✓ | ✓ | ✓ E | | | | ✓ | ✓ |
| Wu 2015 USA (26224304)[170] | Nurses' Health Study + Health Professionals Follow-up Study Prospective Cohort Range:30-75 years M/F Health professionals | 1,540 / 41,530 25 years | Self-report verified by medical record | FFQ, **Orange juice**: | Incidence, Squamous cel carcinoma, male | ≥1 servings/day vs <1 servings/week | 1.15 (0.97 - 1.35) Ptrend:0.26 | ✓ | | ✓ | ✓ | ✓ | | ✓ | | | | ✓ | ✓ |

| Author, Year Country (PMID) | Study name, type Age, sex, ethnicity, characteristics of participants | Cases/ Study size Median follow-up (years) | Case ascertainment | Exposure Assessment, detail | Outcome Sub-population | Comparison | RR (95%CI) Ptrend | Adjustments | | | | | | | | | | | |
|---|---|---|---|---|---|---|---|---|---|---|---|---|---|---|---|---|---|---|---|
| | | | | | | | | Age | Sex | Smo | Alc | Obe | Phy | Diet | Med | Mens | Repr | Oth | Skin |
| | | 2,004 / 63,759 25 years | | | Incidence, Squamous cell carcinoma, female | | 1.20 (1.03 - 1.39) Ptrend:0.02 | ✓ | | ✓ | ✓ | ✓ | | ✓ | | ✓ | | ✓ | ✓ |
| | | 2,758 / 105,289 25 years | | | Incidence, SCC on head, neck and extremities | ≥5 vs <1 servings/week | 1.13 (1.01 - 1.26) Ptrend:0.03 | ✓ | | ✓ | ✓ | ✓ | | ✓ | | ✓ | | ✓ | ✓ |
| | | 483 / 105,289 25 years | | | Incidence, SCC on truncal sites | | 1.02 (0.64 - 1.64) Ptrend:0.73 | ✓ | | ✓ | ✓ | ✓ | | ✓ | | ✓ | | ✓ | ✓ |
| **Breast (female)** | | | | | | | | | | | | | | | | | | | |
| Masala 2012 Italy (22215387)[112] | European Prospective Investigation into Cancer and nutrition cohort, Italy Prospective Cohort Range:36-64 years F | 1,072 / 31,510 11.25 years | Cancer registry | FFQ, **Fruit juices**: | Incidence, Breast cancer | >39 vs <0.8 g/day | 1.06 (0.91 - 1.24) Ptrend:0.44 | | | ✓ | ✓ | ✓ | ✓ | ✓ E | | ✓ | ✓ | ✓ | |
| Makarem 2018 USA (29674390)[108] | Framingham Offspring Cohort Prospective Cohort Age:55.4 years F | 124 / 1,689 | medical examinations | Semi-quantitative FFQ, **Fruit juices**: apple juice/cider, orange juice, grapefruit juice and other juice. | Incidence, Breast cancer | >7 vs <1.5 servings/week | 1.03 (0.67 - 1.62) | ✓ | | ✓ | ✓ | ✓ | ✓ | ✓ E | D | ✓ | ✓ | ✓ | |
| Sonestedt 2008 Sweden (18711147)[156] | Malmo Diet and Cancer, 1991 Prospective Cohort Range:46-75 years F | 544 / 15,773 10.3 years | Cancer registry | 7-day Food Record & FFQ, **Fruit juices**: | Incidence, Invasive breast cancer | Median intake 200 vs 0 g/day | 1.12 (0.89 - 1.41) Ptrend:0.23 | ✓ | | ✓ | ✓ | ✓ | ✓ | ✓ E | | ✓ | ✓ | ✓ | |
| Hirvonen 2006 France (16406814)[69] | SU.VI.MAX study, 1994 Prospective Cohort Range:35-60 years F participants of a RCT | 95 / 4,396 6.6 years | medical records | 24h Recall, **Fruit juices**: | Incidence, Breast cancer, | ≥150 vs 0 ml/day | 1.29 (0.80 - 2.09) Ptrend:0.32 | ✓ | | ✓ | | | | | | ✓ | ✓ | ✓ | |
| Farvid 2019 USA (29978479)[52] | Nurses' Health Study and Nurses' Health Study II Pooled Analysis Cohorts Range:27-59 years F Registered nurses | 10,882 / 182,145 23.7 years | biennial follow-up questionnaires and medical records | FFQ, **Fruit juices**: Apple juice/cider, orange juice, grapefruit juice, other fruit juices, prune juice, tomato juice | Incidence, Invasive breast cancer | >1.5 servings/day vs ≤1 serving/week | 1.03 (0.96 - 1.10) | ✓ | | ✓ | ✓ | ✓ | ✓ | ✓ E | ✓ | ✓ | ✓ | ✓ | |
| | | | | | | Per 2 servings day | 1.03 (0.97 - 1.09) | ✓ | | ✓ | ✓ | ✓ | ✓ | ✓ E | ✓ | ✓ | ✓ | ✓ | |
| Farvid 2016 USA (27170029)[51] | Nurses' Health study II Prospective Cohort Age:36 years F Mostly white nurses | 1,346 / 90,476 22 years | self-report verified by pathology report | Validated FFQ, **Fruit juices**: apple juice/cider, orange juice, grapefruit juice, other fruit juices (during adolescence) | Incidence, Invasive breast cancer | Median intake 2.1 vs 0.1 serving/day | 1.02 (0.86 - 1.21) Ptrend:0.85 | ✓ | | ✓ | ✓ | ✓ | | ✓ E | ✓ | ✓ | ✓ | ✓ | |
| | | | | | | Per serving day | 1.01 (0.93 - 1.09) | ✓ | | ✓ | ✓ | ✓ | | ✓ E | ✓ | ✓ | ✓ | ✓ | |
| | | 3,233 / 90,476 22 years | | Validated FFQ, **Fruit juices**: apple juice/cider, orange juice, grapefruit juice, other fruit juices. (during early adulthood) | | Median intake 2 vs 0 serving/day | 1.11 (0.99 - 1.25) Ptrend:0.13 | ✓ | | ✓ | ✓ | ✓ | | ✓ E | ✓ | ✓ | ✓ | ✓ | |
| | | | | | | Per serving day | 1.04 (0.99 - 1.10) | ✓ | | ✓ | ✓ | ✓ | | ✓ E | ✓ | ✓ | ✓ | ✓ | |

| Author, Year Country (PMID) | Study name, type Age, sex, ethnicity, characteristics of participants | Cases/ Study size Median follow-up (years) | Case ascertainment | Exposure Assessment, detail | Outcome Sub-population | Comparison | RR (95%CI) Ptrend | Adjustments: Age | Sex | Smo | Alc | Obe | Phy | Diet | Med | Mens | Repr | Oth | Skin |
|---|---|---|---|---|---|---|---|---|---|---|---|---|---|---|---|---|---|---|---|
| Farvid 2019 USA (29978479)[52] | Nurses' Health Study and Nurses' Health Study II Pooled Analysis Cohorts Range:27-59 years F Registered nurses | 7,563 / 182,145 23.7 years | biennial follow-up questionnaires and medical records | FFQ, **Fruit juices**: Apple juice/cider, orange juice, grapefruit juice, other fruit juices, prune juice, tomato juice | Incidence, Invasive breast cancer, postmenopausal | >1.5 servings/day vs ≤1 serving/week | 1.05 (0.96 - 1.15) | ✓ | | ✓ | ✓ | ✓ | ✓ | ✓ E | ✓ | ✓ | ✓ | ✓ | |
| Key 2019 UK (30412247)[83] | Million Women Study Prospective Cohort F Mostly white | 23,886 / 691,571 11.9 person-years | UK NHS databases on cancer registrations and deaths | FFQ, **Fruit juices**: | Incidence, Breast cancer, postmenopausal | ≥7.0 vs 0-0.99 glasses/week | 1.03 (1.01 - 1.06) Ptrend:1.52 | ✓ | | ✓ | ✓ | ✓ | ✓ | ✓ E | | ✓ | ✓ | ✓ | |
| Farvid 2016 USA (27170029)[51] | Nurses' Health study II Prospective Cohort Age:36 years F Mostly white nurses | 1,168 / 90,476 22 years | self-report verified by pathology report | Validated FFQ, **Fruit juices**: apple juice/cider, orange juice, grapefruit juice, other fruit juices (during early adulthood) | Incidence, Postmenopausal breast cancer; | Median intake 2 vs 0 serving/day | 1.08 (0.89 - 1.30) Ptrend:0.68 | ✓ | | ✓ | ✓ | ✓ | | ✓ E | ✓ | ✓ | ✓ | ✓ | |
| | | | | | | Per serving /day | 1.02 (0.93 - 1.11) | ✓ | | ✓ | ✓ | ✓ | | ✓ E | ✓ | ✓ | ✓ | ✓ | |
| | | 640 / 90,476 22 years | | Validated FFQ, **Fruit juices**: apple juice/cider, orange juice, grapefruit juice, other fruit juices (during adolescence) | | Median intake 2.1 vs 0.1 serving/day | 1.02 (0.79 - 1.32) Ptrend:0.44 | ✓ | | ✓ | ✓ | ✓ | | ✓ E | ✓ | ✓ | ✓ | ✓ | |
| | | | | | | Per serving /day | 1.04 (0.94 - 1.17) | ✓ | | ✓ | ✓ | ✓ | | ✓ E | ✓ | ✓ | ✓ | ✓ | |
| Farvid 2019 USA (29978479)[52] | Nurses' Health Study and Nurses' Health Study II Pooled Analysis Cohorts Range:27-59 years F Registered nurses | 2,364 / 182,145 23.7 years | biennial follow-up questionnaires and medical records | FFQ, **Fruit juices**: Apple juice/cider, orange juice, grapefruit juice, other fruit juices, prune juice, tomato juice | Incidence, Invasive breast cancer, premenopausal | >1.5 servings/day vs ≤1 serving/week | 1.00 (0.86 - 1.16) | ✓ | | ✓ | ✓ | ✓ | ✓ | ✓ E | ✓ | ✓ | ✓ | ✓ | |
| Farvid 2016 USA (27170029)[51] | Nurses' Health study II Prospective Cohort Age:36 years F Mostly white nurses | 575 / 90,476 22 years | self-report verified by pathology report | Validated FFQ, **Fruit juices**: apple juice/cider, orange juice, grapefruit juice, other fruit juices (during adolescence) | Incidence, Premenopausal breast cancer; | Median intake 2.2 vs 0.1 serving/day | 1.06 (0.80 - 1.41) Ptrend:1.00 | ✓ | | ✓ | ✓ | ✓ | | ✓ E | ✓ | ✓ | ✓ | ✓ | |
| | | | | | | Per serving /day | 1.00 (0.89 - 1.12) | ✓ | | ✓ | ✓ | ✓ | | ✓ E | ✓ | ✓ | ✓ | ✓ | |
| | | 1,718 / 90,476 22 years | | Validated FFQ, **Fruit juices**: apple juice/cider, orange juice, grapefruit juice, other fruit juices (during early adulthood) | | Median intake 2.1 vs 0 serving/day | 1.12 (0.95 - 1.32) Ptrend:0.21 | ✓ | | ✓ | ✓ | ✓ | | ✓ E | ✓ | ✓ | ✓ | ✓ | |
| | | | | | | Per serving /day | 1.04 (0.98 - 1.12) | ✓ | | ✓ | ✓ | ✓ | | ✓ E | ✓ | ✓ | ✓ | ✓ | |
| Fung 2011 USA (21832271)[63] | Nurse's Health Study Prospective Cohort Range:30-55 years F Postmenopausal | 827 / 86,621 26 years | self-reported/death certificate/pathology reports | FFQ, **Juices**: Fruit juice | Incidence, Breast cancer ER- | 2.9 vs 0.1 serving/day | 0.95 (0.76 - 1.19) Ptrend:0.43 | ✓ | | ✓ | ✓ | ✓ | ✓ | ✓ E | ✓ | | ✓ | ✓ | |
| Sonestedt 2008 Sweden (18711147)[156] | Malmo Diet and Cancer, 1991 Prospective Cohort Range:46-75 years F | 392 / 15,773 10.3 years | Cancer registry | 7-day Food Record & FFQ, **Fruit juices**: | Incidence, Breast cancer ERα+ | Per 1 category | 1.05 (0.97 - 1.14) | ✓ | | | | | | | | | | | |

| Author, Year Country (PMID) | Study name, type Age, sex, ethnicity, characteristics of participants | Cases/ Study size Median follow-up (years) | Case ascertainment | Exposure Assessment, detail | Outcome Sub-population | Comparison | RR (95%CI) Ptrend | Adjustments Age | Sex | Smo | Alc | Obe | Phy | Diet | Med | Mens | Repr | Oth | Skin |
|---|---|---|---|---|---|---|---|---|---|---|---|---|---|---|---|---|---|---|---|
| | | 58 / 15,773 10.3 years | | | Incidence, Breast cancer ERα- | | 1.03 (0.83 - 1.29) | ✓ | | | | | | | | | | | |
| | | 182 / 15,773 10.3 years | | | Incidence, Breast cancer ERβ+ | | 1.08 (0.95 - 1.22) | ✓ | | | | | | | | | | | |
| | | 188 / 15,773 10.3 years | | | Incidence, Breast cancer ERβ- | | 0.97 (0.65 - 1.43) | ✓ | | | | | | | | | | | |
| | | 159 / 15,773 10.3 years | | | Incidence, Breast cancer ERα+/ERβ+ | | 1.07 (0.94 - 1.22) | ✓ | | | | | | | | | | | |
| | | 155 / 15,773 10.3 years | | | Incidence, Breast cancer ERα+/ERβ- | | 1.01 (0.88 - 1.15) | ✓ | | | | | | | | | | | |
| Stolzenberg-Solomon, 2006 USA (16600944)[160] | Prostate, Lung, Colorectal and Ovarian Cancer Screening Trial , 1993 Prospective Cohort Range:55-74 years F Postmenopausal | 91 / 31,411 4.94 years | Cancer screening programme | FFQ, **Citrus fruit juice**: Orange &/or grapefruit juice | Incidence, Breast cancer, non-vitamins users | >180.4 vs 0 and <5.61 g/day | 0.86 (0.43 - 1.72) Ptrend:0.79 | ✓ | | | | | | ✓ E | | | ✓ | ✓ | |
| Farvid 2016 USA (27170029)[51] | Nurses' Health study II Prospective Cohort Age:36 years F Mostly white nurses | 3,235 / 90,476 22 years | self-report verified by pathology report | Validated FFQ, **Grapefruit juice**: Grapefruit juice (Early adulthood) | Incidence, Invasive breast cancer; | Per 2 serving /week | 1.01 (0.98 - 1.05) | ✓ | | ✓ | ✓ | ✓ | | ✓ E | ✓ | ✓ | ✓ | ✓ | |
| Kim 2008 USA (18026192)[86] | Nurses' Health Study Cohort 1984 Prospective Cohort Range:30-55 years F | 4,315 / 77,050 | medical records | Semi-quantitative FFQ, **Grapefruit juice**: Grapefruit juice | Incidence, Breast cancer | ≥1/2 vs 0 glasses/day | 0.95 (0.80 - 1.13) Ptrend:0.52 | ✓ | | | | ✓ | ✓ | ✓ E | ✓ | ✓ | ✓ | ✓ | |
| | | 3,570 / 77,050 | | | Incidence, Breast cancer, postmenopausal | | 1.02 (0.85 - 1.22) Ptrend:0.95 | ✓ | | | | ✓ | ✓ | ✓ E | ✓ | ✓ | ✓ | ✓ | |
| Farvid 2019 USA (29978479)[52] | Nurses' Health Study and Nurses' Health Study II Pooled Analysis Cohorts Range:27-59 years F Registered nurses | 10,911 / 182,145 23.7 years | biennial follow-up questionnaires and medical records | FFQ, **Orange juice**: orange juice | Incidence, Breast cancer | Per 2 servings week | 1.01 (1.00 - 1.01) | ✓ | | ✓ | ✓ | ✓ | ✓ | ✓ E | ✓ | ✓ | ✓ | ✓ | |
| | | | | | Incidence, Breast cancer ER+ | | 1.00 (0.99 - 1.01) | ✓ | | ✓ | ✓ | ✓ | ✓ | ✓ E | ✓ | ✓ | ✓ | ✓ | |
| | | | | | Incidence, Breast cancer ER- | | 1.01 (0.99 - 1.03) | ✓ | | ✓ | ✓ | ✓ | ✓ | ✓ E | ✓ | ✓ | ✓ | ✓ | |
| Farvid 2016 USA (27170029)[51] | Nurses' Health study II Prospective Cohort Age:36 years F Mostly white nurses | 3,235 / 90,476 22 years | self-report verified by pathology report | Validated FFQ, **Orange juice**: Orange juice (Adolescent) | Incidence, Invasive breast cancer; | Per 2 serving week | 0.99 (0.96 - 1.01) | ✓ | | ✓ | ✓ | ✓ | | ✓ E | ✓ | ✓ | ✓ | ✓ | |
| | | | | Validated FFQ, **Orange juice**: Orange juice (Early adulthood) | | | 1.01 (1.00 - 1.02) | ✓ | | ✓ | ✓ | ✓ | | ✓ E | ✓ | ✓ | ✓ | ✓ | |

### Supplementary Materials

| Author, Year Country (PMID) | Study name, type Age, sex, ethnicity, characteristics of participants | Cases/ Study size Median follow-up (years) | Case ascertainment | Exposure Assessment, detail | Outcome Sub-population | Comparison | RR (95%CI) Ptrend | Adjustments | | | | | | | | | | | |
|---|---|---|---|---|---|---|---|---|---|---|---|---|---|---|---|---|---|---|---|
| | | | | | | | | Age | Sex | Smo | Alc | Obe | Phy | Diet | Med | Mens | Repr | Oth | Skin |
| Fung 2013 USA (23532538)[62] | Nurses' Health Study<br>Prospective Cohort<br>Range:30-55 years<br>F<br>Postmenopausal | 792 /<br>75,929<br>24 years | questionnaire, medical records or pathology reports, death certificate, physician, family member | FFQ,<br>**Orange juice**: | Incidence,<br>Breast cancer<br>ER- | Per 2 servings /week | 1.02 (0.98 - 1.07) | ✓ | | ✓ | ✓ | ✓ | ✓ | ✓<br>E | ✓ | | ✓ | | |
| Michels 2006 USA (16094624)[121] | Nurses' Health Study and Nurses' Health Study II<br>Pooled Analysis Cohorts<br>F | 582 /<br>238,378<br>17 years | Self-report verified by medical record | Semi-quantitative FFQ,<br>**Orange juice**:<br>(at preschool age) | Incidence,<br>Invasive breast cancer | Per 1 serving day | 0.85 (0.65 - 1.10) | ✓ | | | | ✓ | | | | ✓ | ✓ | ✓ | |
| Frazier 2003 USA (12631400)[57] | Nurses' Health Study Cohort 1976-1996<br>Nested Case Control<br>Range:40-65 years<br>F<br>Multi-ethnic<br>Registered nurses | / 121,700<br>10 years | Through health org. (screening, health insurance) | FFQ,<br>**Orange juice**:<br>orange juice (adolescent) | Incidence,<br>Breast cancer, | Per 1 glasses day | 0.97 (0.82 - 1.14) | ✓ | | | ✓ | ✓ | | ✓ | ✓ | ✓ | ✓ | ✓ | |
| Farvid 2019 USA (29978479)[52] | Nurses' Health Study and Nurses' Health Study II<br>Pooled Analysis Cohorts<br>Range:27-59 years<br>F<br>Registered nurses | 10,911 /<br>182,145<br>23.7 years | biennial follow-up questionnaires and medical records | FFQ,<br>**Apple juices**:<br>apple juice | Incidence,<br>Breast cancer | Per 2 servings week | 1.01 (0.98 - 1.03) | ✓ | | ✓ | ✓ | ✓ | ✓ | ✓<br>E | ✓ | ✓ | ✓ | ✓ | |
| | | | | | Incidence,<br>Breast cancer<br>ER+ | | 0.98 (0.95 - 1.02) | ✓ | | ✓ | ✓ | ✓ | ✓ | ✓<br>E | ✓ | ✓ | ✓ | ✓ | |
| | | | | | Incidence,<br>Breast cancer<br>ER- | | 0.99 (0.92 - 1.06) | ✓ | | ✓ | ✓ | ✓ | ✓ | ✓<br>E | ✓ | ✓ | ✓ | ✓ | |
| Farvid 2016 USA (27170029)[51] | Nurses' Health study<br>Prospective Cohort<br>Age:36 years<br>F<br>Mostly white nurses | 3,235 /<br>90,476<br>22 years | self-report verified by pathology report | Validated FFQ,<br>**Apple juices**:<br>Apple juice (Early adulthood) | Incidence,<br>Invasive breast cancer; | Per 2 serving week | 1.01 (0.98 - 1.04) | ✓ | | ✓ | ✓ | ✓ | | ✓<br>E | ✓ | ✓ | ✓ | ✓ | |
| | | | | Validated FFQ,<br>**Fruit juices**:<br>Apple juice (Adolescent) | | | 0.99 (0.93 - 1.05) | ✓ | | ✓ | ✓ | ✓ | | ✓<br>E | ✓ | ✓ | ✓ | ✓ | |
| Arthur 2021 Canada (33220638)[38] | Canadian Study of Diet, Lifestyle and Health<br>Case Cohort<br>F | 870 / 2,976<br>15 years | Cancer registry and national death index | Self-administered Questionnaire,<br>**Pure fruit juices**:<br>100 % fruit juice:<br>orange/grapefruit juice, apple/grape juice and other juices | Incidence,<br>Breast cancer | >6.2 vs ≤1.5 cups/week | 1.12 (0.90 - 1.40)<br>Ptrend:0.496 | ✓ | | ✓ | ✓ | ✓ | ✓ | ✓ | ✓ | ✓ | ✓ | ✓ | |
| Chazelas 2019 France (31292122)[44] | NutriNet-Santé study<br>Prospective Cohort<br>Age:42.2 years<br>M/F<br>women | 693 /<br>79,724<br>5.1 years | self-report, medical records reviewed by physicians, health insurance database, mortality registry | 24 Hour Diet Recall,<br>**Pure fruit juices**:<br>100% fruit juices | Incidence,<br>Breast cancer | Per 100 ml day | 1.15 (0.98 - 1.36) | ✓ | | ✓ | ✓ | ✓ | ✓ | ✓<br>E | D | ✓ | ✓ | ✓ | |
| Iso 2007 Japan (18260705)[76] | Japan Collaborative Cohort study for Evaluation of Cancer Risk<br>Prospective Cohort<br>Age:49 years<br>F | 29 / 60,982<br>15 years | municipal resident registration records, death certificates | Validated FFQ,<br>**Pure fruit juices**:<br>Fresh fruit juice (in summer season) | Mortality,<br>Breast cancer | ≥3 vs <1 times/week | 1.41 (0.80 - 2.51) | ✓ | | | | | | | | | | | |
| Arthur 2021 Canada (33220638)[38] | Canadian Study of Diet, Lifestyle and Health<br>Case Cohort<br>F | 397 / 2,976<br>15 years | Cancer registry and national death index | Self-administered Questionnaire,<br>**Pure fruit juices**: | Incidence,<br>Breast cancer, | >6.2 vs ≤1.5 cups/week | 1.23 (0.89 - 1.69)<br>Ptrend:0.573 | ✓ | | ✓ | ✓ | ✓ | ✓ | ✓ | ✓ | ✓ | ✓ | ✓ | |

| Author, Year Country (PMID) | Study name, type Age, sex, ethnicity, characteristics of participants | Cases/ Study size Median follow-up (years) | Case ascertainment | Exposure Assessment, detail | Outcome Sub-population | Comparison | RR (95%CI) Ptrend | Adjustments Age | Sex | Smo | Alc | Obe | Phy | Diet | Med | Mens | Repr | Oth | Skin |
|---|---|---|---|---|---|---|---|---|---|---|---|---|---|---|---|---|---|---|---|
| | | | | 100 % fruit juice: orange/grapefruit juice, apple/grape juice and other juices | postmenopausal | | | | | | | | | | | | | | |
| Chazelas 2019 France (31292122)[44] | NutriNet-Santé study Prospective Cohort Age:42.2 years M/F women | 410 / 79,724 5.1 years | self-report, medical records reviewed by physicians, health insurance database, mortality registry | 24 Hour Diet Recall, **Pure fruit juices**: sugary drinks - 100% fruit juices | Incidence, Postmenopausal breast cancer | Per 100 ml day | 1.04 (0.83 - 1.30) | ✓ | | ✓ | ✓ | ✓ | ✓ | ✓ E | D | ✓ | ✓ | ✓ | |
| Arthur 2021 Canada (33220638)[38] | Canadian Study of Diet, Lifestyle and Health Case Cohort F | 473 / 2,976 15 years | Cancer registry and national death index | Self-administered Questionnaire, **Pure fruit juices**: 100 % fruit juice: orange/grapefruit juice, apple/grape juice and other juices | Incidence, Breast cancer, premenopausal | >6.2 vs ≤1.5 cups/week | 1.04 (0.76 - 1.43) Ptrend:0.685 | ✓ | | ✓ | ✓ | ✓ | ✓ | ✓ | ✓ | ✓ | ✓ | ✓ | |
| Chazelas 2019 France (31292122)[44] | NutriNet-Santé study Prospective Cohort Age:42.2 years M/F women | 283 / 79,724 5.1 years | self-report, medical records reviewed by physicians, health insurance database, mortality registry | 24 Hour Diet Recall, **Pure fruit juices**: sugary drinks - 100% fruit juices | Incidence, Premenopausal breast cancer | Per 100 ml day | 1.10 (0.86 - 1.41) | ✓ | | ✓ | ✓ | ✓ | ✓ | ✓ E | D | ✓ | ✓ | ✓ | |
| Farvid 2016 USA (27170029)[51] | Nurses' Health study II Prospective Cohort Age:36 years F Mostly white nurses | 3,235 / 90,476 22 years | self-report verified by pathology report | Validated FFQ, **Other fruit juices**: Fruit juice excluding citrus and apple juices (Adolescent) | Incidence, Invasive breast cancer; | Per 2 serving week | 1.03 (1.00 - 1.05) | ✓ | | ✓ | ✓ | ✓ | | ✓ E | ✓ | ✓ | ✓ | ✓ | |
| | | | | Validated FFQ, **Fruit juices**: Fruit juice excluding citrus and apple juices (Early adulthood) | | | 1.01 (0.98 - 1.03) | ✓ | | ✓ | ✓ | ✓ | | ✓ E | ✓ | ✓ | ✓ | ✓ | |
| Farvid 2019 USA (29978479)[52] | Nurses' Health study and Nurses' Health study II Pooled Analysis Cohorts Range:27-59 years F Registered nurses | / 182,145 23.7 years | biennial follow-up questionnaires and medical records | FFQ, **Other fruit juice**: Fruit juice excluding orange and apple juices | Incidence, Breast cancer | Per 2 servings week | 0.99 (0.98 - 1.01) | ✓ | | ✓ | ✓ | ✓ | ✓ | ✓ E | ✓ | ✓ | ✓ | ✓ | |
| | | | | | Incidence, Breast cancer ER+ | | 0.99 (0.97 - 1.01) | ✓ | | ✓ | ✓ | ✓ | ✓ | ✓ E | ✓ | ✓ | ✓ | ✓ | |
| | | | | | Incidence, Breast cancer ER- | | 1.01 (0.98 - 1.05) | ✓ | | ✓ | ✓ | ✓ | ✓ | ✓ E | ✓ | ✓ | ✓ | ✓ | |
| van Gils 2005 France, Italy, Spain, UK, Netherlands, Germany, Sweden, Denmark (15644545)[164] | European Prospective Investigation into Cancer and nutrition Prospective Cohort Range:25-70 years | 3,659 / 285,526 5.4 years | | Diet Questionnaire, **Fruit and vegetable juices**: | Incidence, Breast cancer | >120 vs 0 g/day | 1.05 (0.92 - 1.20) Ptrend:0.51 | | | ✓ | ✓ | ✓ | ✓ | ✓ E | | ✓ | ✓ | | |
| Olsen, 2003 Denmark (12840204)[133] | Diet, Cancer and Health, 1993 Prospective Cohort Age:57 years F Postmenopausal | 425 / 23,798 4.7 years | By Mail | FFQ, **Fruit and vegetable juices**: | Incidence, Breast cancer, postmenopausal | Per 100 g day | 1.00 (0.87 - 1.16) | ✓ | | | ✓ | ✓ | | ✓ | ✓ | | ✓ | ✓ | |
| | | 91 / 23,798 4.7 years | | | Incidence, Breast cancer ER-, | | 0.67 (0.42 - 1.08) | ✓ | | | ✓ | ✓ | | ✓ | ✓ | | ✓ | ✓ | |

| Author, Year Country (PMID) | Study name, type Age, sex, ethnicity, characteristics of participants | Cases/ Study size Median follow-up (years) | Case ascertainment | Exposure Assessment, detail | Outcome Sub-population | Comparison | RR (95%CI) Ptrend | Adjustments | | | | | | | | | | | |
|---|---|---|---|---|---|---|---|---|---|---|---|---|---|---|---|---|---|---|---|
| | | | | | | | | Age | Sex | Smo | Alc | Obe | Phy | Diet | Med | Mens | Repr | Oth | Skin |
| | | | | | postmenopausal | | | | | | | | | | | | | | |
| | | 303 / 23,798 4.7 years | | | Incidence, Breast cancer ER+, postmenopausal | | 1.04 (0.89 - 1.22) | ✓ | | | ✓ | ✓ | | ✓ | ✓ | | ✓ | ✓ | |
| Dunneram 2019 UK (30526696)[49] | UK Women Cohort Study Prospective Cohort Age:54.1 years F | 1,625 / 32,289 18 years | NHS Central Registry | FFQ, Pure fruit and vegetable **juice**: 99% CI; Orange juice, Other – pure juices (reference PMID 29712719 supplementary) | Incidence, Breast cancer | Per 145 g day | 0.94 (0.83 - 1.08) | ✓ | | ✓ | ✓ | | ✓ | ✓ E | | ✓ | ✓ | ✓ | |
| | . | . | . | . | . Incidence, Breast cancer, postmenopausal | . | .0.99 (0.90 – 1.09) | ✓ | | ✓ | ✓ | | ✓ | ✓ E | | ✓ | ✓ | ✓ | |
| | | | | | Incidence, Breast cancer, premenopausal | | 0.89 (0.72 – 1.10) | ✓ | | ✓ | ✓ | | ✓ | ✓ E | | ✓ | ✓ | ✓ | |
| Sesso 2005 USA (15894655)[153] | Women's Health Study, 1993 Prospective Cohort Health professionals | 1,057 / 38,447 9.9 years | Through health org. (screening, health insurance) | FFQ-Semi-quantitative, **Tomato juice** | Incidence, Breast cancer | ≥2 serving/wk vs none serving/month | 0.81 (0.59 - 1.11) Ptrend:0.37 | ✓ | | | ✓ | ✓ | ✓ | ✓ E | | ✓ | ✓ | ✓ | |
| **Endometrial** | | | | | | | | | | | | | | | | | | | |
| Inoue-Choi 2013 USA (24273064)[74] | Iowa Women's Health Study Prospective Cohort Age:61.6 years F Postmenopausal | 395 / 23,039 24 years | State Health Registry, National Death Index | FFQ, **Fruit juices** | Incidence, Type 1 endometrial cancer | 8.4-63 vs 0-0.6 servings/week | 1.16 (0.87 - 1.56) Ptrend:0.14 | ✓ | | ✓ | ✓ | ✓ | ✓ | ✓ | D | ✓ | ✓ | | |
| | | 86 / 23,039 24 years | | | Incidence, Type 2 endometrial cancer | 8.4-63 vs 0-0.6 servings/week | 0.97 (0.50 - 1.88) Ptrend:0.99 | ✓ | | ✓ | ✓ | ✓ | ✓ | ✓ | D | ✓ | ✓ | | |
| Arthur 2021 Canada (33220638)[38] | Canadian Study of Diet, Lifestyle and Health Case Cohort F | 169 / 2,351 15 years | Cancer registry and national death index | Self-administered Questionnaire, **Pure fruit juices**: 100 % fruit juice: orange/grapefruit juice, apple/grape juice and other juices | Incidence, Endometrial Cancer | >5.6 vs ≤2.5 cups/week | 1.50 (1.00 - 1.72) Ptrend:0.048 | ✓ | | ✓ | ✓ | ✓ | ✓ | ✓ | | ✓ | ✓ | ✓ | |
| | | 125 / 2,351 15 years | | | Incidence, Type 1 endometrial cancer | | 1.63 (1.03 - 2.60) Ptrend:0.040 | ✓ | | ✓ | ✓ | ✓ | ✓ | ✓ | | ✓ | ✓ | ✓ | |
| Iso 2007 Japan (18260705)[76] | Japan Collaborative Cohort study for Evaluation of Cancer Risk Prospective Cohort Range:40-79 years F | 93 / 60,982 15 years | municipal resident registration records, death certificates | FFQ, **Pure fruit juices**: Fresh fruit juice in summer season | Mortality, Uterine cervical cancer | ≥3 vs <1 times/week | 0.84 (0.31 - 2.23) | ✓ | | | | | | | | | | | |
| Merritt 2015 multi-national (25662427)[117] | The European Prospective Investigation into Cancer and Nutrition study and the Nurses Health studies I & II Pooled Analysis Cohorts F | 1,303 / 301,107 | EPIC: population-based cancer registries, active follow up, mortality data from cancer or mortality registries; NHS/NHS II: questionnaire, medical records, pathology | Semi-quantitative FFQ, **Fruit and vegetable juices** | Incidence, Endometrial Cancer, EPIC | Median intake 148.5 vs 0 g/day | 1.07 (0.91 - 1.26) Ptrend:0.35 | ✓ | | ✓ | | ✓ | | ✓ E | | ✓ | ✓ | | |

| Author, Year Country (PMID) | Study name, type Age, sex, ethnicity, characteristics of participants | Cases/ Study size Median follow-up (years) | Case ascertainment | Exposure Assessment, detail | Outcome Sub-population | Comparison | RR (95%CI) Ptrend | Adjustments: Age | Sex | Smo | Alc | Obe | Phy | Diet | Med | Mens | Repr | Oth | Skin |
|---|---|---|---|---|---|---|---|---|---|---|---|---|---|---|---|---|---|---|---|
| Dunneram 2019 UK (30526696)[49] | UK Women Cohort Study<br>Prospective Cohort<br>Age:54.1 years<br>F | 86 / 32,289<br>18 years | reports, national death index<br>NHS Central Registry | FFQ,<br>Pure fruit and vegetable **juice**:<br>99% CI; Orange juice, Other – pure juices (reference PMID **29712719** supplementary) | Incidence, Endometrial Cancer | Per 145 g day | 0.89 (0.60 - 1.33) | ✓ | | ✓ | ✓ | | ✓ | ✓<br>E | D | ✓ | ✓ | ✓ | |
| **Ovary** | | | | | | | | | | | | | | | | | | | |
| Koushik 2005 (16172226)[91] | Pooling Project (n=12 cohorts)<br>Pooled Analysis Cohorts<br>Range:27-93 years<br>F | 2,095 /<br>560,441<br>22 years | Self-report, medical record, cancer registries and death registries | Self-administered Questionnaire,<br>**Fruit juices** | Incidence, Epithelial ovarian cancer | Per 190 g day | 0.99 (0.92 - 1.06) | ✓ | | ✓ | | ✓ | ✓ | ✓<br>E | | ✓ | ✓ | | |
| Sakauchi 2007 Japan (17571946)[149] | JACC Japan<br>Prospective Cohort<br>Range:40-79 years<br>F | 44 / 64,327<br>13.3 years | Death certificates from local public health centres in the study areas | Self-administered Questionnaire,<br>**Fruit juices**: | Mortality, Ovarian cancer | ≥3-4 times/week vs seldom | 0.93 (0.39 - 2.22)<br>Ptrend:0.95 | ✓ | | | | ✓ | ✓ | | | ✓ | ✓ | ✓ | |
| Arthur 2021 Canada (33220638)[38] | Canadian Study of Diet, Lifestyle and Health<br>Case Cohort<br>F | 98 / 2,554<br>15 years | Cancer registry and national death index | Self-administered Questionnaire,<br>**Pure fruit juices**:<br>100 % fruit juice:<br>orange/grapefruit juice, apple/grape juice and other juices | Incidence, Ovarian cancer | >5.6 vs ≤2.4 cups/week | 1.48 (0.87 - 2.54)<br>Ptrend:0.145 | ✓ | | ✓ | ✓ | ✓ | ✓ | ✓ | ✓ | ✓ | ✓ | ✓ | |
| | | 72 / 2,554<br>15 years | | | Incidence, Epithelial ovarian cancer | | 1.32 (0.70 - 2.49)<br>Ptrend:0.352 | ✓ | | ✓ | ✓ | ✓ | ✓ | ✓ | | ✓ | ✓ | ✓ | |
| Mommers, 2005 Netherlands (16104037)[124] | The Netherlands Cohort study<br>Case Cohort<br>F | / 62,573<br>11.3 years | By post | Semi-quantitative FFQ,<br>**Other fruit juices**:<br>other fruit juice (juices from grapes, bananas, apples, pears, strawberries) excluding citrus juice | Incidence, Invasive epithelial ovarian cancer | Per 25 g /day | 1.08 (0.98 - 1.18) | ✓ | | ✓ | | | | ✓ | | | ✓ | | |
| Merritt 2016 multi-national (26607939)[118] | European Prospective Investigation into Cancer and Nutrition study and the Netherlands Cohort study<br>Pooled Analysis Cohorts<br>F | 1,095 /<br>325,004 | population cancer registries or active follow up | Semi-quantitative FFQ,<br>**Fruit and vegetable juices** | Incidence, Epithelial ovarian cancer | Median intake 149.4 vs 0 g/day | 1.06 (0.89 - 1.27)<br>Ptrend:0.94 | | | | | | | ✓<br>E | | ✓ | ✓ | | |
| Dunneram 2019 UK (30526696)[49] | UK Women Cohort Study<br>Prospective Cohort<br>Age:54.1 years<br>F | 251 /<br>32,289<br>18 years | NHS Central Registry | FFQ,<br>~Pure fruit and vegetable **juice**:<br>99% CI; Orange juice, Other – pure juices (reference PMID **29712719** supplementary) | Incidence, Ovarian cancer | Per 145 g day | 0.94 (0.68 - 1.31) | ✓ | | ✓ | ✓ | | ✓ | ✓<br>E | | ✓ | ✓ | ✓ | |
| **Prostate** | | | | | | | | | | | | | | | | | | | |
| Lan 2021 USA (34383904)[94] | NIH- AARP Diet and Health Study<br>Prospective Cohort<br>Age:61.8 years<br>M<br>adolescents | 760 /<br>150,671<br>1729896 person-years | State Health Registry, National Death Index | FFQ,<br>**Fruit juices**: | Mortality, Prostate cancer | ≥ 3 times/week vs ≤ 11 times/year | 0.80 (0.66 - 0.98)<br>Ptrend:0.025 | ✓ | | ✓ | ✓ | ✓ | ✓ | ✓<br>E | ✓<br>D | | | ✓ | |
| | | / 150,671<br>1729896 person-years | | | Incidence, Prostate cancer | | 1.04 (1.00 - 1.09)<br>Ptrend:0.04 | ✓ | | ✓ | ✓ | ✓ | ✓ | ✓<br>E | ✓<br>D | | | ✓ | |
| Makarem 2018 USA (29674390)[108] | Framingham Offspring Cohort,<br>Prospective Cohort<br>Range:26-84 years<br>M/F<br>men | 157 / 3,184 | cancer registry and pathology reports | FFQ,<br>**Fruit juices**:<br>Includes apple juice/cider, orange juice, grapefruit juice and other juice Additional adjustment for BMI, waist circumference, chronic diseases (CVD | Incidence, Prostate cancer | >7 vs <2 serving/week | 1.58 (1.04 - 2.41) | ✓ | | ✓ | | | | ✓<br>E | | | | | |

| Author, Year Country (PMID) | Study name, type Age, sex, ethnicity, characteristics of participants | Cases/ Study size Median follow-up (years) | Case ascertainment | Exposure Assessment, detail | Outcome Sub-population | Comparison | RR (95%CI) Ptrend | Adjustments | | | | | | | | | | | |
|---|---|---|---|---|---|---|---|---|---|---|---|---|---|---|---|---|---|---|---|
| | | | | | | | | Age | Sex | Smo | Alc | Obe | Phy | Diet | Med | Mens | Repr | Oth | Skin |
| | | | | and diabetes), education, physical activity, antioxidant use, percentage energy from fat and fruit and vegetable intake did not alter these findings | | | | | | | | | | | | | | | |
| Stram 2006 USA (17006725)[161] | Hawaii-Los Angeles Multiethnic Cohort Study Prospective Cohort Range:45-75 years M Multi-ethnic | 3,922 / 82,486 8 years | SEER registry and death files | FFQ, **Fruit juices**: Fruit juice alone (orange/grapefruit juice, other juice) | Incidence / Mortality, Total prostate cancer | 64.8 vs - 2.5 g/1000 kcal | 1.09 (0.98 - 1.20) Ptrend:0.107 | ✓ | | | | ✓ | | | | | | ✓ | |
| Drake 2012 Sweden (23134882)[47] | Malmo Diet and Cancer, 1991 Prospective Cohort Range:45-73 years M | 549 / 8,128 15 | linkage of personal identification numbers with the Swedish Cancer Register | FFQ, **Fruit juices**: | Incidence, Prostate cancer | 200 vs 0 g/day | 0.99 (0.81 - 1.22) Ptrend:0.625 | ✓ | | ✓ | ✓ | ✓ | ✓ | ✓ E | | | | ✓ | |
| | | | | | Incidence, Low-risk prostate cancer | | 1.12 (0.87 - 1.44) Ptrend:0.532 | ✓ | | ✓ | ✓ | ✓ | ✓ | ✓ E | | | | ✓ | |
| | | | | | Incidence, High-risk prostate cancer | | 0.75 (0.52 - 1.09) Ptrend:0.067 | ✓ | | ✓ | ✓ | ✓ | ✓ | ✓ E | | | | ✓ | |
| | | | | | Incidence, Symptomatic prostate cancer | | 1.14 (0.85 - 1.52) Ptrend:0.784 | ✓ | | ✓ | ✓ | ✓ | ✓ | ✓ E | | | | ✓ | |
| Kirsh 2007 USA (17652276)[89] | Prostate, Lung, Colorectal and Ovarian Cancer Screening Trial Prospective Cohort Range:55-74 years Mostly white M | 1,338 / 29,361 4.2 years | death certificate, patient records, follow-up visits, pathology reports | FFQ, **Fruit juices**: apple, orange, or other | Incidence / Mortality, Total prostate cancer | Median intake 3.4 vs 0.1 servings/day | 1.23 (1.02 - 1.48) Ptrend:0.08 | ✓ | | ✓ | | ✓ | ✓ | ✓ E | ✓ D | | | ✓ | |
| | | | | | Incidence / Mortality, Aggressive prostate cancer | | 1.29 (0.95 - 1.76) Ptrend:0.49 | ✓ | | ✓ | | ✓ | ✓ | ✓ E | ✓ D | | | ✓ | |
| | | | | | Incidence / Mortality, Extraprostatic prostate cancer | | 1.32 (0.79 - 2.21) Ptrend:0.39 | ✓ | | ✓ | | ✓ | ✓ | ✓ E | ✓ D | | | ✓ | |
| Stram 2006 USA (17006725)[161] | Hawaii-Los Angeles Multiethnic Cohort Study Prospective Cohort Range:45-75 years M Multi-ethnic | 666 / 82,486 8 years | SEER registry and death files | FFQ, **Fruit juices**: Fruit juice alone (orange/grapefruit juice, other juice) | Incidence / Mortality, High grade disease | 64.8 vs - 2.5 g/1000 kcal | 1.03 (0.87 - 1.21) Ptrend:0.321 | ✓ | | | | ✓ | | | | | | ✓ | |
| Lan 2021 USA (34383904)[94] | NIH- AARP Diet and Health Study Prospective Cohort Age:61.8 years adolescents | 14,238 / 150,671 1729896 person-years | State Health Registry, National Death Index | , **Fruit juices**: | Incidence, Nonadvanced prostate cancer | ≥ 3 times/week vs ≤ 11 times/year | 1.07 (1.02 - 1.12) Ptrend:0.002 | ✓ | | ✓ | ✓ | ✓ | ✓ | ✓ E | ✓ D | | | ✓ | |

| Author, Year Country (PMID) | Study name, type Age, sex, ethnicity, characteristics of participants | Cases/ Study size Median follow-up (years) | Case ascertainment | Exposure Assessment, detail | Outcome Sub-population | Comparison | RR (95%CI) Ptrend | Adjustments | | | | | | | | | | | |
|---|---|---|---|---|---|---|---|---|---|---|---|---|---|---|---|---|---|---|---|
| | | | | | | | | Age | Sex | Smo | Alc | Obe | Phy | Diet | Med | Mens | Repr | Oth | Skin |
| | | 5,913 / 150,671 1729896 person-years | | | Incidence, Advanced prostate cancer | | 0.89 (0.79 - 1.00) Ptrend:0.019 | ✓ | | ✓ | ✓ | ✓ | ✓ | ✓ E | ✓ D | | | ✓ | |
| Chazelas 2019 France (31292122)[44] | NutriNet-Santé study Prospective Cohort Age:42.2 years M/F men | 291 / 21,533 5.1 years | self-report, medical records reviewed by physicians, health insurance database, mortality registry | 24 Hour Diet Recall, **Pure fruit juices**: sugary drinks - 100% fruit juices | Incidence, Prostate cancer | Per 100 ml day | 0.98 (0.78 - 1.22) | ✓ | | ✓ | ✓ | ✓ | ✓ | ✓ E | D | | | ✓ | |
| Iso 2007 Japan (18260705)[76] | Japan Collaborative Cohort study for Evaluation of Cancer Risk Prospective Cohort Range:40-79 years M/F | 152 / 44,518 15 years | municipal resident registration records, death certificates | FFQ, **Pure fruit juices**: fresh fruit juice (in summer season) | Mortality, Prostate cancer | ≥3 vs <1 times/week | 1.01 (0.63 - 1.61) | ✓ | | | | | | | | | | | |
| Petimar 2017 multi-national (28446545)[139] | Pooling Project (n=15 cohorts) Pooled Analysis Cohorts Range:18-97 years M Mixed | 52,680 / 842,149 22 years | various (follow up questionnaires, medical records, cancer registries and/or biopsy) | FFQ, **Fruit and vegetable juices**: | Incidence, Prostate cancer | ≥250 vs <25 g/day | 1.03 (0.99 - 1.07) Ptrend:0.26 | ✓ | | ✓ | ✓ | ✓ | ✓ | ✓ E | D | | | ✓ | |
| | | 38,475 | | | Incidence, Localised prostate cancer | | 1.03 (0.99 - 1.06) Ptrend:0.08 | ✓ | | ✓ | ✓ | ✓ | ✓ | ✓ E | D | | | ✓ | |
| | | 4,934 | | | Incidence, Advanced prostate cancer | | 1.09 (0.95 - 1.25) Ptrend:0.31 | ✓ | | ✓ | ✓ | ✓ | ✓ | ✓ E | D | | | ✓ | |
| | | 3,115 | | | Incidence, Advanced restricted prostate cancer | | 1.14 (0.97 - 1.33) Ptrend:0.06 | ✓ | | ✓ | ✓ | ✓ | ✓ | ✓ E | D | | | ✓ | |
| | | 37,556 | | | Incidence, Low grade prostate cancer | | 1.01 (0.96 - 1.05) Ptrend:0.75 | ✓ | | ✓ | ✓ | ✓ | ✓ | ✓ E | D | | | ✓ | |
| | | 9,753 | | | Incidence, High grade prostate cancer | | 1.08 (0.96 - 1.22) Ptrend:0.88 | ✓ | | ✓ | ✓ | ✓ | ✓ | ✓ E | D | | | ✓ | |
| | | 3,205 | | | Mortality, Prostate cancer | | 1.01 (0.86 - 1.19) Ptrend:0.75 | ✓ | | ✓ | ✓ | ✓ | ✓ | ✓ E | D | | | ✓ | |
| | | 52,680 / 842,149 22 years | | FFQ, **Vegetable juices**: | Incidence, Prostate cancer | Per 237 g day | 1.02 (1.00 – 1.05) | ✓ | | ✓ | ✓ | ✓ | ✓ | ✓ E | D | | | ✓ | |

| Author, Year Country (PMID) | Study name, type Age, sex, ethnicity, characteristics of participants | Cases/ Study size Median follow-up (years) | Case ascertainment | Exposure Assessment, detail | Outcome Sub-population | Comparison | RR (95%CI) Ptrend | Adjustments: Age | Sex | Smo | Alc | Obe | Phy | Diet | Med | Mens | Repr | Oth | Skin |
|---|---|---|---|---|---|---|---|---|---|---|---|---|---|---|---|---|---|---|---|
| | | 38,475 | | | Incidence, Localised prostate cancer | | 1.01 (1.00 – 1.030 | ✓ | | ✓ | ✓ | ✓ | ✓ | ✓ E | D | | | ✓ | |
| | | 4,934 | | | Incidence, Advanced prostate cancer | | 1.03 (0.98 – 1.09) | ✓ | | ✓ | ✓ | ✓ | ✓ | ✓ E | D | | | ✓ | |
| | | 3,115 | | | Incidence, Advanced restricted prostate cancer | | 1.05 (0.94 – 1.17) | ✓ | | ✓ | ✓ | ✓ | ✓ | ✓ E | D | | | ✓ | |
| | | 37,556 | | | Incidence, Low grade prostate cancer | | 1.00 (0.98 – 1.01) | ✓ | | ✓ | ✓ | ✓ | ✓ | ✓ E | D | | | ✓ | |
| | | 9,753 | | | Incidence, High grade prostate cancer | | 1.02 (0.98 – 1.05) | ✓ | | ✓ | ✓ | ✓ | ✓ | ✓ E | D | | | ✓ | |
| | | 3,205 | | | Mortality, Prostate cancer | | 1.03 (0.93 – 1.14) | ✓ | | ✓ | ✓ | ✓ | ✓ | ✓ E | D | | | ✓ | |
| | | 52,680 / 842,149 22 years | | FFQ, **Citrus juices**: | Incidence, Prostate cancer | Per 186 g day | 1.01 (1.00 – 1.02) | ✓ | | ✓ | ✓ | ✓ | ✓ | ✓ E | D | | | ✓ | |
| | | 38,475 | | | Incidence, Localised prostate cancer | | 1.02 (1.01 – 1.04) | ✓ | | ✓ | ✓ | ✓ | ✓ | ✓ E | D | | | ✓ | |
| | | 4,934 | | | Incidence, Advanced prostate cancer | | 1.00 (0.94 – 1.06) | ✓ | | ✓ | ✓ | ✓ | ✓ | ✓ E | D | | | ✓ | |
| | | 3,115 | | | Incidence, Advanced restricted prostate cancer | | 1.05 (0.98 – 1.13) | ✓ | | ✓ | ✓ | ✓ | ✓ | ✓ E | D | | | ✓ | |
| | | 37,556 | | | Incidence, Low grade prostate cancer | | 1.00 (0.99 – 1.02) | ✓ | | ✓ | ✓ | ✓ | ✓ | ✓ E | D | | | ✓ | |
| | | 9,753 | | | Incidence, High grade prostate cancer | | 1.01 (0.98 – 1.04) | ✓ | | ✓ | ✓ | ✓ | ✓ | ✓ E | D | | | ✓ | |

| Author, Year Country (PMID) | Study name, type Age, sex, ethnicity, characteristics of participants | Cases/ Study size Median follow-up (years) | Case ascertainment | Exposure Assessment, detail | Outcome Sub-population | Comparison | RR (95%CI) Ptrend | Adjustments | | | | | | | | | | | |
|---|---|---|---|---|---|---|---|---|---|---|---|---|---|---|---|---|---|---|---|
| | | | | | | | | Age | Sex | Smo | Alc | Obe | Phy | Diet | Med | Mens | Repr | Oth | Skin |
| | | 3,205 | | | Mortality, Prostate cancer | | 0.98 (0.91 – 1.06) | ✓ | | ✓ | ✓ | ✓ | ✓ | ✓ E | D | | | ✓ | |
| Fraser 2020 USA (32100191)[56] | Adventist Health Study 2 Prospective Cohort Range:30-104 years M | 1,226 / 27,934 7.9 years | biennial follow-up questionnaires, cancer registry records, medical records or death certificate data | FFQ, **Tomatoes juice**: Tomato-based vegetable juice | Incidence, prostate cancer | 5-6/week vs never/rarely | 0.88 (0.56 - 1.38) Ptrend:0.36 | | | ✓ | | | | ✓ E | ✓ | | | ✓ | |
| Schuurman, 1998 Netherlands (9718219)[152] | The Netherlands Cohort study Case Cohort Range:55-69 years M | 610 / 58,279 6.3 years | municipal population registries | FFQ, **Tomatoes juice**: | Incidence, prostate cancer | Per 25 g day | 1.12 (0.96 - 1.29) | ✓ | | | | | | ✓ | | | | ✓ | |
| **Kidney** | | | | | | | | | | | | | | | | | | | |
| Allen 2011 UK (21407222)[36] | The Million Women Study, 1996 Prospective Cohort Range:50-64 years F | 588 / 779,369 5.2 years | National Health Service central registers | FFQ, **Fruit juices** | Incidence, Renal cell carcinoma | ≥2 vs 0 or<1 drinks/day | 1.01 (0.76 - 1.36) | | | ✓ | | ✓ | | | | | | ✓ | |
| | | | | | | Per 1 drinks day | 0.96 (0.84 - 1.08) | | | ✓ | | ✓ | | | | | | ✓ | |
| Lee 2006 USA (16775182)[98] | Nurses' Health Study and Health Professionals Follow-up Study Pooled Analysis Cohorts Range:30-75 years M/F | 248 / 136,587 | Self-report, medical records reviewed by physician, national death index, death report by family members in follow up questionnaires | Validated FFQ, **Fruit juices**: | Incidence, Renal cell cancer | Per 1 serving day | 1.06 (0.88 - 1.28) | ✓ | | ✓ | ✓ | ✓ | | ✓ E | D | | ✓ | | |
| Rashidkhani, 2005 Sweden (15455348)[142] | Swedish Mammography Cohort Prospective Cohort Range:40-76 years F | 122 / 61,000 13.4 years | Health screening program | FFQ, **Fruit juices**: | Incidence, Renal cell adenocarcinoma | ≥2–3 vs 0 servings/week | 1.46 (0.94 - 2.26) Ptrend:0.26 | ✓ | | | | ✓ | | | | | | | |
| Iso 2007 Japan (18260705)[76] | Japan Collaborative Cohort study for Evaluation of Cancer Prospective Cohort Range:40-79 years Mixed | 41 / 105,500 15 years | municipal resident registration records, death certificates | Validated FFQ, **Pure fruit juices**: Fruit / fresh juices in summer season | Mortality, Kidney cancer, male | ≥3 vs <1 times/week | 0.87 (0.36 - 2.12) | ✓ | | | | | | | | | | | |
| | | 15 / 105,500 15 years | | | Mortality, Kidney cancer, female | | 2.01 (0.39 - 10.30) | ✓ | | | | | | | | | | | |
| Lee 2007 (17583573)[26] | Pooling Project (n=13 cohorts) Pooled Analysis Cohorts M/F mainly Caucasian | 769 / 244,483 20 years | Self-report, medical record, cancer registries and death registries | FFQ and dietary history, **Fruit and vegetable juices**: | Incidence, Renal cancer, male | ≥1serving (186 g) /day vs non-drinker | 1.09 (0.68 - 1.74) Ptrend:0.40 | ✓ | | ✓ | ✓ | ✓ | | ✓ E | | | | | |
| | | | | | | Per 1 serving /day | 1.09 (0.94 - 1.25) | ✓ | | ✓ | ✓ | ✓ | | ✓ E | | | | | |
| | | 709 / 530,469 20 years | | | Incidence, Renal cancer, female | ≥1serving (186g) /day vs non-drinker | 1.09 (0.78 - 1.53) Ptrend:0.68 | ✓ | | ✓ | ✓ | ✓ | | ✓ E | | | ✓ | | |
| | | | | | | Per 1 serving day | 1.00 (0.89 - 1.13) | ✓ | | ✓ | ✓ | ✓ | | ✓ E | | | ✓ | | |

| Author, Year Country (PMID) | Study name, type Age, sex, ethnicity, characteristics of participants | Cases/ Study size Median follow-up (years) | Case ascertainment | Exposure Assessment, detail | Outcome Sub-population | Comparison | RR (95%CI) Ptrend | Adjustments | | | | | | | | | | | |
|---|---|---|---|---|---|---|---|---|---|---|---|---|---|---|---|---|---|---|---|
| | | | | | | | | Age | Sex | Smo | Alc | Obe | Phy | Diet | Med | Mens | Repr | Oth | Skin |
| Heath 2021 multi-national (33849969[66] | European Prospective Investigation into Cancer and nutrition<br>Prospective Cohort<br>Age:52 years<br>M/F | 888 /<br>389,220<br>15 years | population registries or active follow up | Diet Questionnaire,<br>**Fruit and vegetable juices**:<br>fruit and vegetable juices and nectars | Incidence, Renal cell carcinoma | Per 100 g day | 1.03 (0.97 - 1.09) | ✓ | ✓ | ✓ | ✓ | ✓ | ✓ | ✓ E * | | | | ✓ | |
| | | 356 /<br>389,220<br>15 years | | | Mortality, Renal cell carcinoma | | 1.06 (0.97 - 1.16) | ✓ | ✓ | ✓ | ✓ | ✓ | ✓ | ✓ E * | | | | ✓ | |
| | | 198 | | | Mortality, renal cell carcinoma, male | | 0.93 (0.79 - 1.09) | ✓ | | ✓ | ✓ | ✓ | ✓ | ✓ E * | | | | ✓ | |
| | | 158 | | | Mortality, renal cell carcinoma, female | | 1.17 (1.05 - 1.29) | ✓ | | ✓ | ✓ | ✓ | ✓ | ✓ E * | | | | ✓ | |
| **Bladder** | | | | | | | | | | | | | | | | | | | |
| Park 2013 USA (23739308)[138] | Multiethnic Cohort<br>Prospective Cohort<br>Range:45-75 years<br>M/F<br>Multi-ethnic | 429 /<br>83,694<br>12.5 years | cancer registry/death records/National Death Index | FFQ,<br>**Fruit juices** | Incidence, Invasive bladder cancer, male | ≥59.7 vs <4.7 g/1000 kcal /day | 0.94 (0.72 - 1.24)<br>Ptrend:0.67 | ✓ | | ✓ | | | | ✓ E | | | | ✓ | |
| | | 152 /<br>102,191<br>12.5 years | | | Incidence, Invasive bladder cancer, female | | 0.81 (0.51 - 1.30)<br>Ptrend:0.14 | ✓ | | ✓ | | | | ✓ E | | | | ✓ | |
| Iso 2007 Japan (18260705)[76] | Japan Collaborative Cohort study for Evaluation of Cancer Risk<br>Prospective Cohort<br>Range:40-79 years<br>M/F | 81 /<br>105,500<br>15 years | municipal resident registration records, death certificates | FFQ,<br>**Pure fruit juices**:<br>Fresh fruit juice in summer season | Mortality, Urothelial cancer, male | ≥3 vs <1 times/week | 0.44 (0.21 - 0.90) | ✓ | | | | | | | | | | | |
| | | 38 /<br>105,500<br>15 years | | | Mortality, Urothelial cancer, female | | 0.42 (0.15 - 1.17) | ✓ | | | | | | | | | | | |
| Michaud, 1999 USA (10203279)[120] | Health Professionals Follow-up Study<br>Prospective Cohort<br>Range:40-75 years<br>M | / 47,909 | Ongoing or prior study and convenience sampling | FFQ,<br>**Fruit juices**:<br>apple or cider | Incidence / Mortality, bladder cancer | | Not significant | ✓ | | | | | | ✓ E | | | | ✓ | |
| Zeegers, 2001 Netherlands (11700259)[181] | The Netherlands Cohort study<br>Nested Case Control<br>Range:55-69 years<br>M/F | / 120,852<br>6.3 years | Population registry | FFQ,<br>**Citrus fruit juice**:<br>orange/grapefruit juice | Incidence, Cancer of bladder, urethra, renal pelvis, ureter, | Per 25 g day | 1.00 (0.93 - 1.08) | ✓ | ✓ | ✓ | | | | ✓ | | | | | |
| Michaud, 1999 USA (10203279)[120] | Health Professionals Follow-up Study<br>Prospective Cohort<br>Range:40-75 years<br>M | / 47,909 | Ongoing or prior study and convenience sampling | FFQ,<br>**Orange juice**:<br>orange | Incidence / Mortality, Bladder cancer | | Not significant | ✓ | | | | | | ✓ E | | | | ✓ | |
| | | | | FFQ,<br>**Grapefruit juice**:<br>grapefruit juice | | | Not significant | ✓ | | | | | | ✓ E | | | | ✓ | |
| | | | | FFQ,<br>**Other fruit juices**:<br>Fruit juices excluding citrus and apple juices | | | Not significant | ✓ | | | | | | ✓ E | | | | ✓ | |

| Author, Year Country (PMID) | Study name, type Age, sex, ethnicity, characteristics of participants | Cases/ Study size Median follow-up (years) | Case ascertainment | Exposure Assessment, detail | Outcome Sub-population | Comparison | RR (95%CI) Ptrend | Adjustments | | | | | | | | | | | |
|---|---|---|---|---|---|---|---|---|---|---|---|---|---|---|---|---|---|---|---|
| | | | | | | | | Age | Sex | Smo | Alc | Obe | Phy | Diet | Med | Mens | Repr | Oth | Skin |
| Mills, 1991 USA (2000840)[123] | Adventist Health Study<br>Prospective Cohort<br>M/F<br>non-Hispanic white<br>Seventh-day Adventists | 46 / 34,198 | Census | FFQ,<br>**Sweetened fruit juice**:<br>Sweetened real fruit juice | Incidence, Bladder cancer | ≥1 vs Never times/week | 0.31 (0.09 - 1.00)<br>Ptrend:0.02 | ✓ | ✓ | ✓ | | | | | | | | | |
| Ros 2011 multi-national (20715171)[147] | European Prospective Investigation into Cancer and nutrition<br>Prospective Cohort<br>Range:25-70 years<br>M/F | 513 /<br>233,236<br>9.3 years | cancer registries, health insurance records, pathology rec & active follow up | Validated FFQ,<br>**Fruit and vegetable juices** | Incidence, Urothelial cancer | (≥79 women; ≥72 men) vs <8 ml/day | 1.32 (1.05 - 1.66)<br>Ptrend:0.02 | ✓ | ✓ | ✓ | | | | ✓<br>E | | | | | |
| | | | | | | Per 100 ml day | 1.10 (1.04 - 1.17) | ✓ | ✓ | ✓ | | | | ✓<br>E | | | | | |
| | | 210 /<br>233,236<br>9.3 years | | | Incidence, High-risk urothelial cell carcinomas | (≥79 women; ≥72 men) vs <8 ml/day | 1.21 (0.85 - 1.71)<br>Ptrend:0.22 | ✓ | ✓ | ✓ | | | | ✓<br>E | | | | | |
| | | | | | | Per 100 ml day | 1.07 (0.98 - 1.17) | ✓ | ✓ | ✓ | | | | ✓<br>E | | | | | |
| | | 271 /<br>233,236<br>9.3 years | | | Incidence, Low-risk urothelial cell carcinomas | (≥79 women; ≥72 men) vs <8 ml/day | 1.53 (1.12 - 2.09)<br>Ptrend:0.01 | ✓ | ✓ | ✓ | | | | ✓<br>E | | | | | |
| | | | | | | Per 100 ml day | 1.19 (1.06 - 1.34) | ✓ | ✓ | ✓ | | | | ✓<br>E | | | | | |
| Michaud, 1999 USA (10203279)[120] | Health Professionals Follow-up Study<br>Prospective Cohort<br>Range:40-75 years<br>M | / 47,909 | Ongoing or prior study and convenience sampling | FFQ,<br>**Tomatoes juice**: | Incidence / Mortality, bladder cancer | | Not significant | ✓ | | | | | | ✓<br>E | | | | ✓ | |
| Xu 2021 USA (34244460)[175] | Prostate, Lung, Colorectal and Ovarian Cancer Screening Trial<br>Prospective Cohort<br>Range:55-74 years<br>M/F | 774 /<br>101,683<br>12.5 years | Questionnaire, medical + death records | Dietary History Questionnaire,<br>**Tomatoes juice**: | Incidence, bladder cancer | ≥18.54 vs 0 g/day | 1.04 (0.83 - 1.29)<br>Ptrend:0.773 | ✓ | ✓ | ✓ | ✓ | ✓ | | ✓<br>E | | | | ✓ | |
| | | | | | | Per 1 SD day | 0.99 (0.92 - 1.07) | ✓ | ✓ | ✓ | ✓ | ✓ | | ✓<br>E | | | | ✓ | |
| **Brain and Spinal cord** | | | | | | | | | | | | | | | | | | | |
| Kuan 2019 multi-national (30690490)[92] | Million Women Study + NIH-AARP Diet and Health Study + Prostate, Lung, Colorectal and Ovarian Cancer Screening Trial<br>Pooled Analysis Cohorts<br>Age:60.6 years<br>M/F | 969 /<br>692,176<br>15.4 million person-years | cancer registry + admission records | 24h Recall + FFQ-Semi-quantitative,<br>**Fruit juices**: | Incidence, Glioma, Million Women Study | Per 100 g | 1.00 (0.81 - 1.25) | | | ✓ | ✓ | ✓ | ✓ | | | | ✓ | ✓ | |
| | | 690 /<br>277,906<br>15.4 million person-years | | | Incidence, Glioma, NIH-AARP,male | | 0.98 (0.94 - 1.02) | ✓ | | ✓ | ✓ | ✓ | ✓ | | | | | ✓ | |
| | | 315 /<br>192,874<br>15.4 million person-years | | | Incidence, Glioma, NIH-AARP, female | | 1.02 (0.95 - 1.09) | ✓ | | ✓ | ✓ | ✓ | ✓ | | | | ✓ | ✓ | |
| | | 82 / 47,196<br>15.4 million | | | Incidence, Glioma, PLCO | | 1.07 (0.95 - 1.21) | ✓ | | ✓ | ✓ | ✓ | | | | | | ✓ | |

| Author, Year Country (PMID) | Study name, type Age, sex, ethnicity, characteristics of participants | Cases/ Study size Median follow-up (years) | Case ascertainment | Exposure Assessment, detail | Outcome Sub-population | Comparison | RR (95%CI) Ptrend | Adjustments | | | | | | | | | | | |
|---|---|---|---|---|---|---|---|---|---|---|---|---|---|---|---|---|---|---|---|
| | | | | | | | | Age | Sex | Smo | Alc | Obe | Phy | Diet | Med | Mens | Repr | Oth | Skin |
| | | person-years | | | male | | | | | | | | | | | | | | |
| | | 53 / 51,952 15.4 million person-years | | | Incidence, Glioma, PLCO female | | 0.98 (0.79 - 1.21) | ✓ | | ✓ | ✓ | ✓ | | | | | ✓ | ✓ | |
| | | 728 / 1,262,104 15.4 million person-years | | | Incidence, Glioma, <5 years of follow-up | | 0.96 (0.90 - 1.02) | ✓ | ✓ | ✓ | ✓ | ✓ | ✓ | | | | ✓ | ✓ | |
| | | 1,381 / 1,262,104 15.4 million person-years | | | Incidence, Glioma, ≥5 years of follow-up | | 1.02 (0.98 - 1.06) | ✓ | ✓ | ✓ | ✓ | ✓ | ✓ | | | | ✓ | ✓ | |
| **Thyroid** | | | | | | | | | | | | | | | | | | | |
| Xiao 2014 USA (24686895)[174] | NIH- AARP Diet and Health Study Prospective Cohort Range:50-71 years M/F Mostly white | 586 / 491,840 9 years | Cancer registry and national death index | FFQ, **Citrus juice;** Orange / grapefruit juice | Incidence, Thyroid cancer | Median intake 0.51 - 7.26 vs 0 - 0.02 serving /day /1000 calories | 1.55 (1.18 - 2.03) Ptrend:0.004 | | ✓ | ✓ | ✓ | ✓ | | ✓ E | ✓ | | | ✓ | |
| | | 170 / 491,840 9 years | | | Incidence, Papillary thyroid cancer, male | | 1.39 (0.86 - 2.27) Ptrend:0.33 | | ✓ | ✓ | ✓ | ✓ | | ✓ E | ✓ | | | ✓ | |
| | | 247 / 491,840 9 years | | | Incidence, Papillary thyroid cancer, female | | 1.79 (1.17 - 2.75) Ptrend:0.01 | | ✓ | ✓ | ✓ | ✓ | | ✓ E | ✓ | | | ✓ | |
| Zamora-Ros 2017 France, Italy, Spain, UK, Netherlands, Greece, Germany, Sweden, Denmark (28688112)[179] | European Prospective Investigation into Cancer and Nutrition study Prospective Cohort Range:35-70 years M/F European origin | 748 / 476,108 14 years | cancer and mortality registries, health insurance & pathology records, active follow up | Questionnaire And FFQ, **Fruit juices**: citrus fruit juices, non-citrus fruit juices, vegetable juices and fruit nectars | Incidence, Thyroid cancer, differentiated | >94 vs <1 g/day | 1.23 (0.98 - 1.53) Ptrend:0.06 | ✓ | ✓ | ✓ | ✓ | ✓ | ✓ | ✓ E | | | | ✓ | |
| | | | | | | Per 50 g /day | 1.02 (0.99 - 1.06) | ✓ | ✓ | ✓ | ✓ | ✓ | ✓ | ✓ E | | | | ✓ | |
| | | 601 / 476,108 14 years | cancer and mortality registries, health insurance & pathology records, active follow up | | Incidence, Papillary thyroid cancer | >94 vs <1 g/day | 1.69 (0.92 - 3.08) Ptrend:0.91 | ✓ | ✓ | ✓ | ✓ | ✓ | ✓ | ✓ E | | | | ✓ | |
| | | | | | | Per 50 g day | 1.02 (0.94 - 1.11) | ✓ | ✓ | ✓ | ✓ | ✓ | ✓ | ✓ E | | | | ✓ | |
| | | 109 / 476,108 14 years | cancer and mortality registries, health insurance & pathology records, active follow up | | Incidence, Follicular thyroid cancer | >94 vs <1 g/day | 1.12 (0.88 - 1.43) Ptrend:0.35 | ✓ | ✓ | ✓ | ✓ | ✓ | ✓ | ✓ E | | | | ✓ | |
| | | | | | | Per 50 g day | 1.02 (0.98 - 1.06) | ✓ | ✓ | ✓ | ✓ | ✓ | ✓ | ✓ E | | | | ✓ | |

| Author, Year Country (PMID) | Study name, type Age, sex, ethnicity, characteristics of participants | Cases/ Study size Median follow-up (years) | Case ascertainment | Exposure Assessment, detail | Outcome Sub-population | Comparison | RR (95%CI) Ptrend | Adjustments | | | | | | | | | | | |
|---|---|---|---|---|---|---|---|---|---|---|---|---|---|---|---|---|---|---|---|
| | | | | | | | | Age | Sex | Smo | Alc | Obe | Phy | Diet | Med | Mens | Repr | Oth | Skin |
| **Lymphoid and hematopoietic** | | | | | | | | | | | | | | | | | | | |
| Podoltsev 2020 USA (32868318)[141] | NIH- AARP Diet and Health Study Prospective Cohort Range:50-71 years M/F | 472 / 463,049 15.5 years | population based cancer registries and mortality registries | FFQ, **Fruit juices**: | Incidence, myeloproliferative neoplasms | ≥0.91 vs ≤0.22 cups/day | 1.10 (0.87 - 1.38) Ptrend:0.43 | ✓ | ✓ | | | ✓ | | ✓ E | | | | ✓ | |
| | | 182 / 463,049 15.5 years | | | Incidence, Polycythemia vera | | 1.74 (1.17 - 2.58) Ptrend:0.01 | ✓ | ✓ | | | ✓ | | ✓ E | | | | ✓ | |
| | | | | | Incidence, Essential thrombocythemia | | 0.96 (0.64 - 1.43) Ptrend:0.79 | ✓ | ✓ | | | ✓ | | ✓ E | | | | ✓ | |
| Thompson 2010 USA (19685491)[162] | Iowa Women's Health Study Prospective Cohort Age:62 years F Mostly white | 415 / 35,159 20 years | Cancer registry, death certificates, National Death Index | Semi-quantitative FFQ, **Juices**: apple juice/cider | Incidence, Non-Hodgkin Lymphoma | >2 vs 0 serving/month | 0.65 (0.47 - 0.90) | ✓ | | | | | | ✓ E | | | | | |

**Age**: age; **Alc**: alcohol; **Diet**: dietary factors and energy intake (E), or * adjustment for other soft drinks not under investigation; **Med**: medical-related factors (e.g. infections, diseases in the breast, digestive tract, gallbladder, liver, or pancreas, cancer screening, precancerous conditions, or pre-existing skin cancer) or diabetes (D); **Mens**: menstrual characteristics (e.g. age at menarche, menopausal status/duration, age at menopause, or type of menopause); **Obe**: obesity (adiposity measures); **Oth**: others (e.g. family history/genetic factor, race/ethnicity, ancestry, socioeconomic factors, or marriage); **Phy**: physical activity; **Repr**: reproductive or hormonal factors (e.g. parity, age at first/last birth, postmenopausal hormone use, or oral contraceptive use); **Sex**: sex; **Skin**: skin-related factors (e.g. UV light/sun exposure, sunburns, time spent outdoors, sunscreen use, indoor tanning, moles, fair skin, red/blond hair,

#### Supplementary Table 14. Descriptive table of the included observational studies of soft drinks and cancer incidence and mortality.

| Author, Year Country (PMID) | Study name, type Age, sex, ethnicity, characteristics of participants | Cases/ Study size Median follow-up (years) | Case ascertainment | Exposure Assessment, detail | Outcome Sub-population | Comparison | RR (95%CI) Ptrend | Adjustments: Age | Sex | Smo | Alc | Obe | Phy | Diet | Med | Mens | Repr | Oth | Skin |
|---|---|---|---|---|---|---|---|---|---|---|---|---|---|---|---|---|---|---|---|
| **Mouth, pharynx larynx** | | | | | | | | | | | | | | | | | | | |
| Ren 2010 USA (20395127)[143] | NIH- AARP Diet and Health Study Prospective Cohort Range:50-71 years M/F Mixed Retired | 392 / 481,563 6 years | linkage of the cohort with database to state cancer registries | FFQ, **Carbonated soft drinks** | Incidence, Oral cavity cancer | ≥1 can/day vs none | 0.77 (0.54 - 1.09) Ptrend:0.31 | ✓ | ✓ | ✓ | ✓ | ✓ | ✓ | ✓ E | | | | ✓ | |
| | | 178 / 481,563 6 years | | | Incidence, Pharynx cancer | | 0.76 (0.46 - 1.25) Ptrend:0.20 | ✓ | ✓ | ✓ | ✓ | ✓ | ✓ | ✓ E | | | | ✓ | |
| | | 307 / 481,563 6 years | | | Incidence, Larynx cancer | | 0.82 (0.55 - 1.23) Ptrend:0.35 | ✓ | ✓ | ✓ | ✓ | ✓ | ✓ | ✓ E | | | | ✓ | |
| **Oesophageal** | | | | | | | | | | | | | | | | | | | |
| Ren 2010 USA (20395127)[143] | NIH- AARP Diet and Health Study Prospective Cohort Range:50-71 years M/F Mixed Retired | 123 / 481,563 6 years | linkage of the cohort with database to state cancer registries | FFQ, **Carbonated soft drinks** | Incidence, Oesophageal squamous cell carcinoma | ≥1 can/day vs none | 0.85 (0.46 - 1.56) Ptrend:0.55 | ✓ | ✓ | ✓ | ✓ | ✓ | ✓ | ✓ E | | | | ✓ | |
| | | 305 / 481,563 6 years | | | Incidence, Oesophageal adenocarcinoma | | 1.11 (0.66 - 1.85) Ptrend:0.73 | ✓ | ✓ | ✓ | ✓ | ✓ | ✓ | ✓ E | | | | ✓ | |
| **Stomach** | | | | | | | | | | | | | | | | | | | |
| Ren 2010 USA (20395127)[143] | NIH- AARP Diet and Health Study Prospective Cohort Range:50-71 years M/F Mixed Retired | 231 / 481,563 6 years | linkage of the cohort with database to state cancer registries | FFQ, **Carbonated soft drinks** | Incidence, Cardia cancer | ≥1 can/day vs none | 0.89 (0.55 - 1.45) Ptrend:0.75 | ✓ | ✓ | ✓ | ✓ | ✓ | ✓ | ✓ E | | | | ✓ | |
| | | 224 / 481,563 6 years | | | Incidence, non-Cardia cancer | | 0.75 (0.45 - 1.24) Ptrend:0.30 | ✓ | ✓ | ✓ | ✓ | ✓ | ✓ | ✓ E | | | | ✓ | |
| **Colorectal** | | | | | | | | | | | | | | | | | | | |
| Papadimitriou 2022 multi-national (33901663)[136] | European Prospective Investigation into Cancer and nutrition and Netherlands Cohort Study Prospective Cohort M/F | 5,069 / 394,288 14.1 years | population-based cancer registries, insurance records, active follow up | FFQ-Semi-quantitative, **Soft drinks** | Incidence, Colorectal cancer, EPIC | Per 1 SD day | 1.04 (1.02 - 1.07) | ✓ | ✓ | ✓ | | ✓ | ✓ | ✓ E | D | | | ✓ | |
| | | 7,496 / 394,288 14.1 years | | | Incidence, Colorectal cancer, NLCS | | 0.96 (0.91 - 1.02) | ✓ | | ✓ | | ✓ | ✓ | ✓ E | D | | | ✓ | |
| Agnoli 2013 Italy (22821300)[35] | European Prospective Investigation into Cancer and nutrition cohort, Italy Prospective Cohort M/F | 435 / 45,275 11.28 years | Cancer registry and hospital records | Semi-quantitative FFQ, **Soft drinks** | Incidence, Colorectal cancer | 14.3 - 3000 vs 0 g/day | 0.98 (0.78 - 1.22) Ptrend:0.937 | ✓ | ✓ | ✓ | | ✓ | ✓ | ✓ * | | | | ✓ | |
| Mullee 2019 Denmark,France,Germany,Greece,Italy,Netherlands,Norway,Spain,S | European Prospective Investigation into Cancer and nutrition Prospective Cohort | 2,095 / 451,743 16.4 years | active follow up, cancer registry, | FFQ-Semi-quantitative, **Soft drinks**: | Mortality, Colorectal cancer | ≥1 vs <1 glasses/month | 1.25 (1.07 - 1.47) Ptrend:0.004 | ✓ | ✓ | ✓ | ✓ | ✓ | ✓ | ✓ E * | | ✓ | ✓ | ✓ | |

| Author, Year Country (PMID) | Study name, type Age, sex, ethnicity, characteristics of participants | Cases/ Study size Median follow-up (years) | Case ascertainment | Exposure Assessment, detail | Outcome Sub-population | Comparison | RR (95%CI) Ptrend | Adjustments Age | Sex | Smo | Alc | Obe | Phy | Diet | Med | Mens | Repr | Oth | Skin |
|---|---|---|---|---|---|---|---|---|---|---|---|---|---|---|---|---|---|---|---|
| weden,UK (31479109)[126] | Age:50.8 years M/F | | death certificate and National death index | Carbonated/soft/ isotonic drinks and diluted syrups | | | | | | | | | | | | | | | |
| Khan, 2004 Japan (15075007)[85] | Japan, Hokkaido Cohort Study Prospective Cohort Age:58 years M/F | 15 / 1,524 13.8 years | Area residency lists | Dietary History Questionnaire, **Soft drinks**: Carbonated drink/Juice | Mortality, Colorectal cancer, male | Everyday + several times/weeks vs never + several times/years | 0.60 (0.20 - 2.10) | ✓ | | ✓ | | | | | | | | | |
| | | 14 / 1,634 14.8 years | | | Mortality, Colorectal cancer, female | | 0.80 (0.20 - 3.00) | ✓ | | ✓ | | | | | | | | ✓ | |
| Papadimitriou 2022 multi-national (33901663)[136] | European Prospective Investigation into Cancer and nutrition and Netherlands Cohort Prospective Cohort M/F | 3,143 / 394,288 14.1 years | population-based cancer registries, insurance records, active follow up | FFQ-Semi-quantitative, **Soft drinks** | Incidence, Colon cancer, EPIC | Per 1 SD day | 1.04 (1.01 - 1.07) | ✓ | ✓ | ✓ | | ✓ | ✓ | ✓ E | D | | | ✓ | |
| | | 2,612 / 394,288 14.1 years | | | Incidence, Colon cancer, NLCS | | 0.97 (0.92 - 1.02) | ✓ | ✓ | ✓ | | ✓ | ✓ | ✓ E | D | | | ✓ | |
| | | 1,495 / 394,288 14.1 years | | | Incidence, Proximal colon cancer, EPIC | | 1.02 (0.97 - 1.07) | ✓ | ✓ | ✓ | | ✓ | ✓ | ✓ E | D | | | ✓ | |
| | | 1,348 / 394,288 14.1 years | | | Incidence, Proximal colon cancer, NLCS | | 0.97 (0.90 - 1.06) | ✓ | | ✓ | | ✓ | ✓ | ✓ E | D | | | ✓ | |
| | | 1,435 / 394,288 14.1 years | | | Incidence, Distal colon cancer, EPIC | | 1.06 (1.01 - 1.11) | ✓ | ✓ | ✓ | | ✓ | ✓ | ✓ E | D | | | ✓ | |
| | | 1,187 / 394,288 14.1 years | | | Incidence, Distal colon cancer, NLCS | | 0.98 (0.90 - 1.06) | ✓ | ✓ | ✓ | | ✓ | ✓ | ✓ E | D | | | ✓ | |
| | | 1,715 / 394,288 14.1 years | | | Incidence, Rectal cancer, EPIC | | 1.04 (1.00 - 1.09) | ✓ | ✓ | ✓ | | ✓ | ✓ | ✓ E | D | | | ✓ | |
| | | 801 / 394,288 14.1 years | | | Incidence, Rectal cancer, NLCS | | 0.96 (0.87 - 1.06) | ✓ | ✓ | ✓ | | ✓ | ✓ | ✓ E | D | | | ✓ | |
| Khan, 2004 Japan (15075007)[85] | Japan, Hokkaido Cohort Study Prospective Cohort Age:57 years M/F | 14 / 1,634 14.8 years | Area residency lists | Questionnaire, **Cola** | Mortality, Colorectal cancer, female | Everyday + several times/weeks vs never + several | 2.20 (0.30 - 17.80) | ✓ | | ✓ | | | | | | | | ✓ | |

| Author, Year Country (PMID) | Study name, type Age, sex, ethnicity, characteristics of participants | Cases/ Study size Median follow-up (years) | Case ascertainment | Exposure Assessment, detail | Outcome Sub-population | Comparison | RR (95%CI) Ptrend | Adjustments Age | Sex | Smo | Alc | Obe | Phy | Diet | Med | Mens | Repr | Oth | Skin |
|---|---|---|---|---|---|---|---|---|---|---|---|---|---|---|---|---|---|---|---|
| | | | | | | times/year or month | | | | | | | | | | | | | |
| **Liver** | | | | | | | | | | | | | | | | | | | |
| Zhang 2023 USA (37460062)[183] | Women's Health Initiative<br>Prospective Cohort<br>Age:63.4 years<br>F<br>Mainly white<br>Postmenopausal | 176 / 78,356<br>22.1 years | Self-report adjudicated by physician review of medical records | Validated FFQ,<br>**Soft drinks**:<br>Low- and high-energy soft drinks | Incidence, Liver cancer | Per 1 serving day | 1.04 (0.77 - 1.40) | ✓ | | ✓ | ✓ | ✓ | ✓ | ✓<br>E | ✓<br>D | | ✓ | ✓ | |
| Jones 2022 USA (35728406)[79] | NIH- AARP Diet and Health Study + Prostate, Lung, Colorectal and Ovarian Cancer Screening trial<br>Pooled Analysis Cohorts<br>M/F<br>Mainly white | 587 / 553,874<br>≤12 years follow-up | NIH-AARP: cancer reg,<br>PLCO: self-report + medical record/cancer reg | Questionnaire,<br>**Soft drinks**:<br>Soda and fruit punch | Incidence, Liver cancer | Per 1 time day | 1.05 (0.97 - 1.14) | ✓ | ✓ | ✓ | ✓ | ✓ | | ✓<br>E | | | | ✓ | |
| | | 252 / 553,874<br>>12 years follow-up | | | | | 0.92 (0.79 - 1.06) | ✓ | ✓ | ✓ | ✓ | ✓ | | ✓<br>E | | | | ✓ | |
| Stepien 2016 multi-national (25528243)[159] | European Prospective Investigation into Cancer and nutrition cohort<br>Prospective Cohort<br>Mainly white | 121 / 477,206<br>11.4 years | cancer registries | Semi-quantitative FFQ,<br><br>**Soft drinks**:<br>Carbonated/soft/ isotonic drinks and diluted syrups. | Incidence, HCC | Per 1 serving week | 1.18 (1.04 - 1.34) | | | ✓ | ✓ | ✓ | ✓ | ✓<br>E | D | | | ✓ | |
| Jones 2022 USA (35728406)[79] | NIH- AARP Diet and Health Study + Prostate, Lung, Colorectal and Ovarian Cancer Screening trial<br>Pooled Analysis Cohorts<br>M/F<br>Mainly white | 587 / 553,874<br>≥12 years follow-up | NIH-AARP: cancer reg,<br>PLCO: self-report + medical record/cancer reg | Questionnaire,<br>**Carbonated soft drinks**:<br>Regular soda. | Incidence, Liver cancer | Per 1 time day | 1.07 (0.98 - 1.17) | ✓ | ✓ | ✓ | ✓ | ✓ | | ✓<br>E | | | | ✓ | |
| | | 252 / 553,874<br>>12 years follow-up | | | | | 0.92 (0.79 - 1.08) | ✓ | ✓ | ✓ | ✓ | ✓ | | ✓<br>E | | | | ✓ | |
| **Gallbladder/ bile ducts** | | | | | | | | | | | | | | | | | | | |
| Larsson 2016 Sweden (27281756)[96] | Swedish Mammography Cohort and the Cohort of Swedish Men<br>Pooled Analysis Cohorts<br>Range:45-83 years<br>M/F<br>Swedish | 71 / 70,832<br>13.4 years | Swedish Cancer Registry & record linkage with Swedish Cause-of-Death Registry | FFQ,<br>**Soft drinks**:<br>Sweetened beverages | Incidence, Gallbladder cancer | ≥2 vs 0 serving/day | 2.24 (1.02 - 4.89)<br>Ptrend:0.02 | ✓ | ✓ | ✓ | | ✓ | | ✓<br>E | | | | | |
| | | | | FFQ,<br>**Soft drinks**:<br>Soft drinks (sugar-sweetened and artificially sweetened) and juice drinks | | Per 1 serving day | 1.24 (1.05 - 1.47) | ✓ | ✓ | ✓ | | ✓ | | ✓<br>E | | | | | |
| Stepien 2016 multi-national (25528243)[159] | European Prospective Investigation into Cancer and nutrition cohort<br>Prospective Cohort<br>Mainly white | 87 / 477,206<br>11.4 years | cancer registries | Semi-quantitative FFQ,<br>**Soft drinks**:<br>Carbonated/soft/ isotonic drinks and diluted syrups. Further classification of soft drinks into sugar sweetened and artificially sweetened was possible for participants in all centers except three: Italy (North and Ragusa), Sweden | Incidence, Gallbladder cancer | Per 1 serving /week | 0.89 (0.79 - 1.01) | ✓ | ✓ | ✓ | ✓ | ✓ | ✓ | ✓<br>E | ✓ | | | ✓ | |

| Author, Year Country (PMID) | Study name, type Age, sex, ethnicity, characteristics of participants | Cases/ Study size Median follow-up (years) | Case ascertainment | Exposure Assessment, detail | Outcome Sub-population | Comparison | RR (95%CI) Ptrend | Adjustments Age | Sex | Smo | Alc | Obe | Phy | Diet | Med | Mens | Repr | Oth | Skin |
|---|---|---|---|---|---|---|---|---|---|---|---|---|---|---|---|---|---|---|---|
| Larsson 2016 Sweden (27281756)[96] | Swedish Mammography Cohort and the Cohort of Swedish Men<br>Pooled Analysis Cohorts<br>Range:45-83 years<br>M/F<br>Swedish | 127 / 70,832<br>13.4 years | Swedish Cancer Registry & record linkage with Swedish Cause-of-Death Registry | FFQ,<br>**Soft drinks**:<br>Soft drinks (sugar-sweetened and artificially sweetened) and juice drinks | Incidence, Extrahepatic biliary tract cancer | ≥2 vs 0 serving/day | 1.79 (1.02 - 3.13)<br>Ptrend:0.05 | ✓ | ✓ | ✓ | | ✓ | | ✓ E | | | | | |
| | | | | | | Per 1 serving day | 1.14 (0.99 - 1.30) | ✓ | ✓ | ✓ | | ✓ | | ✓ E | | | | | |
| Stepien 2016 multi-national (25528243)[159] | European Prospective Investigation into Cancer and nutrition cohort<br>Prospective Cohort<br>Mainly white | 236 / 477,206<br>11.4 years | cancer registries | Semi-quantitative FFQ,<br>**Soft drinks**:<br>Carbonated/soft/ isotonic drinks and diluted syrups. Further classification of soft drinks into sugar sweetened and artificially sweetened was possible for participants in all centres except three: Italy (North and Ragusa), Sweden | Incidence, Extrahepatic bile duct system cancer | Per 1 serving week | 0.96 (0.90 - 1.00) | ✓ | ✓ | ✓ | ✓ | ✓ | ✓ | ✓ E | ✓ | | | ✓ | |
| Larsson 2016 Sweden (27281756)[96] | Swedish Mammography Cohort and the Cohort of Swedish Men<br>Pooled Analysis Cohorts<br>Range:45-83 years<br>M/F<br>Swedish | 21 / 70,832<br>13.4 years | Swedish Cancer Registry & record linkage with Swedish Cause-of-Death Registry | FFQ,<br>**Soft drinks**:<br>Soft drinks (sugar-sweetened and artificially sweetened) and juice drinks | Incidence, Intrahepatic biliary tract cancer | ≥2 vs 0 serving/day | 1.69 (0.41 - 7.03)<br>Ptrend:0.37 | ✓ | ✓ | ✓ | | ✓ | | ✓ E | | | | | |
| | | | | | | Per 1 serving day | 1.10 (0.75 - 1.61) | ✓ | ✓ | ✓ | | ✓ | | ✓ E | | | | | |
| Stepien 2016 multi-national (25528243)[159] | European Prospective Investigation into Cancer and nutrition cohort<br>Prospective Cohort<br>Mainly white | 66 / 477,206<br>11.4 years | cancer registries | Semi-quantitative FFQ,<br>**Soft drinks**:<br>Carbonated/soft/ isotonic drinks and diluted syrups. | Incidence, Intrahepatic bile duct cancer | Per 1 serving week | 0.97 (0.90 - 1.06) | | | ✓ | ✓ | ✓ | ✓ | ✓ E | D | | | ✓ | |
| | | 54 / 477,206<br>11.4 years | | | Incidence, Ampulla of Vater cancer | | 1.02 (0.95 - 1.10) | ✓ | ✓ | ✓ | ✓ | ✓ | ✓ | ✓ E | ✓ | | | ✓ | |
| **Pancreas** | | | | | | | | | | | | | | | | | | | |
| Navarrete-Muñoz 2016 multi-national (27510540)[127] | European Prospective Investigation into Cancer and nutrition<br>Prospective Cohort<br>Age:51 years<br>M/F<br>Mostly white | 865 / 477,199<br>11.6 years | cancer and pathology registry, active follow up, health insurance record, mortality registry and contact of participants or next-of-kin | Diet Questionnaire,<br>**Soft drinks**:<br>Carbonated/soft/isotonic drinks and diluted syrups and juices (obtained from either 100% fruit and vegetables or concentrates) and nectars (juices with 20% added sugar). | Incidence / Mortality, Pancreatic cancer | >246.3 vs 0.1 - 24.6 g/day | 0.96 (0.75 - 1.21)<br>Ptrend:0.41 | ✓ | ✓ | ✓ | ✓ | | ✓ | | | | | ✓ | |
| | | | | | | Per 100 g day | 1.00 (0.97 - 1.04) | ✓ | ✓ | ✓ | ✓ | | ✓ | | | | | ✓ | |
| Khan 2004 Japan (15075007)[85] | Hokkaido Cohort, Japan<br>Prospective Cohort<br>Age:58 years<br>M/F | 12 / 1,524<br>13.8 years | Multiple methods | FFQ,<br>**Soft drinks**:<br>Carbonated drink/Juice | Mortality, Pancreatic cancer, male | Everyday + several times/weeks vs never + several times/ month or year | 0.20 (0.00 - 1.80) | ✓ | | ✓ | | | | | | | | | |
| | | 13 / 1,634<br>14.8 years | | | Mortality, Pancreatic cancer, female | | 0.20 (0.00 - 1.80) | ✓ | | ✓ | | | | | | | | ✓ | |
| Bao 2008 USA (18689380)[39] | | 1,014 / 487,922<br>7.2 years | Cancer registry and national death index | Dietary Recall,<br>**Soft drinks** | Incidence / Mortality, Pancreatic | Median intake 816.9 vs 0 g/day | 1.17 (0.91 - 1.49)<br>Ptrend:0.35 | ✓ | ✓ | ✓ | ✓ | ✓ | ✓ | ✓ E | | | | ✓ | |

| Author, Year Country (PMID) | Study name, type Age, sex, ethnicity, characteristics of participants | Cases/ Study size Median follow-up (years) | Case ascertainment | Exposure Assessment, detail | Outcome Sub-population | Comparison | RR (95%CI) Ptrend | Adjustments Age | Sex | Smo | Alc | Obe | Phy | Diet | Med | Mens | Repr | Oth | Skin |
|---|---|---|---|---|---|---|---|---|---|---|---|---|---|---|---|---|---|---|---|
| | NIH- AARP Diet and Health Study<br>Prospective Cohort<br>Age:62 years<br>M/F<br>Mostly white<br>Retired | | | | Adenocarcinoma | | | | | | | | | | | | | | |
| Zhong 2023 USA (36094042)[189] | Prostate, Lung, Colorectal and Ovarian Cancer Screening trial<br>Prospective Cohort<br>Age:66.1 years<br>M/F<br>Non-Hispanic white | | | Dietary History Questionnaire,<br>**Soft drinks**:<br>Regular or diet soft drinks, either caffeinated or decaffeinated. | Incidence, Pancreatic cancer | >3.7 vs <0.9 serving/day | 1.02 (0.75 - 1.37)<br>Ptrend:0.991 | ✓ | ✓ | ✓ | ✓ | ✓ | | ✓<br>E | D | | | ✓ | |
| Zhang 2020 USA (32716590)[184] | Prostate, Lung, Colorectal and Ovarian Cancer Screening trial<br>Prospective Cohort<br>Range:55-74 years<br>M/F | 307 / 95,962<br>8.87 years | Self-report, medical records and National Death Index | Diet History Questionnaire,<br>**Soft drinks**:<br>Fruit/vegetable drinks and soft drinks | Mortality, Pancreatic cancer | > 250 vs 0 g/day | 0.70 (0.45 - 1.08)<br>Ptrend:0.1010 | ✓ | ✓ | ✓ | ✓ | ✓ | ✓ | ✓<br>E<br>* | D | | | ✓ | |
| Larsson 2006 Sweden 17093171[90] | Cohort of Swedish men and Swedish Mammography Cohort<br>Pooled Analysis Cohorts<br>Age:61 years<br>M/F<br>European origin | 131 / 77,797<br>7.2 years | Cancer registry | Dietary Recall,<br>**Soft drinks**:<br>Carbonated soft drinks and non-carbonated sweetened drinks | Incidence, Pancreatic cancer | Median intake 2.1 vs 0 servings/day | 1.93 (1.18 - 3.14)<br>Ptrend:0.02 | ✓ | ✓ | ✓ | ✓ | ✓ | | ✓<br>E | | | | ✓ | |
| Mueller 2010 Singapore (20142243)[125] | Singapore Chinese Health study<br>Prospective Cohort<br>Age:56 years<br>M/F<br>Singapore Chinese | 140 / 60,524<br>10.7 years | Cancer registry and death registry | Semi-quantitative FFQ,<br>**Carbonated soft drinks**:<br>Coca-Cola and 7-Up | Incidence / Mortality, Pancreatic Adenocarcinoma | ≥2 vs 0 servings/week | 1.86 (1.11 - 3.13)<br>Ptrend:0.02 | ✓ | ✓ | ✓ | ✓ | | ✓ | ✓<br>E | | | | ✓ | |
| **Lung** | | | | | | | | | | | | | | | | | | | |
| Heath 2022 multi-national (35830197)[67] | European Prospective Investigation into Cancer and nutrition<br>Prospective Cohort<br>M/F | 2,420 / 327,790 | Cancer and mortality registries, pathology registries, active follow up, health insurance records | Questionnaire & FFQ,<br>**Soft drinks** | Incidence, Lung cancer | Per 1 SD day | 0.99 (0.96 - 1.03) | ✓ | ✓ | ✓ | | ✓ | ✓ | ✓<br>E | D | | | ✓ | |
| Gnagnarella 2013 Italy (23956193)[65] | COSMOS (Continuous Observation of Smoking Subjects)<br>Prospective Cohort<br>Age:57 years | 178 / 4,336<br>5.7 years | screening examinations | FFQ,<br>**Soft drinks** | Incidence, Lung cancer | ≥2 drinks/day vs never | 0.98 (0.49 - 1.96)<br>Ptrend:0.3299 | ✓ | ✓ | ✓ | | | | ✓<br>E | | | | | |
| You 2022 USA (36424907)[177] | Prostate, Lung, Colorectal and Ovarian Cancer Screening trial<br>Prospective Cohort<br>Range:55-74 years<br>M/F<br>Mainly white<br>Screening Program | 1,517 / 154,897<br>8.9 years | chest radiography, follow-up, cancer & death registry, physician | FFQ,<br>**Soft drinks**<br>No intake of soft drinks, soda, and pop | Incidence, Lung cancer | Intake vs no intake | 0.79 (0.64 - 0.97) | ✓ | ✓ | ✓ | ✓ | ✓ | ✓ | ✓<br>E | | | | ✓ | |
| | | | | | Incidence, Lung cancer, male | Intake vs no intake | 0.96 (0.72 – 1.28) | ✓ | ✓ | ✓ | ✓ | ✓ | ✓ | ✓<br>E | | | | ✓ | |
| | | | | | Incidence, Lung cancer, female | Intake vs no intake | 0.62 (0.45 – 0.87) | ✓ | ✓ | ✓ | ✓ | ✓ | ✓ | ✓<br>E | | | | ✓ | |

| Author, Year Country (PMID) | Study name, type Age, sex, ethnicity, characteristics of participants | Cases/ Study size Median follow-up (years) | Case ascertainment | Exposure Assessment, detail | Outcome Sub-population | Comparison | RR (95%CI) Ptrend | Adjustments Age | Sex | Smo | Alc | Obe | Phy | Diet | Med | Mens | Repr | Oth | Skin |
|---|---|---|---|---|---|---|---|---|---|---|---|---|---|---|---|---|---|---|---|
| Khan, 2004 Japan (15075007)[85] | Japan, Hokkaido Cohort Study<br>Prospective Cohort<br>Age:58 years<br>M/F | 41 / 1,524<br>13.8 years | Area residency lists | Dietary History Questionnaire,<br>**Soft drinks**:<br>Carbonated drink/Juice | Mortality, Lung cancer, male | Everyday + several times/weeks vs never + several times/months + several times/year | 1.10 (0.50 - 2.10) | ✓ | | ✓ | | | | | | | | | |
| | | 10 / 1,634<br>14.8 years | | Dietary History Questionnaire,<br>**Soft drinks**:<br>Carbonated drink/juice | Mortality, Lung cancer, female | | 1.80 (0.50 - 6.60) | ✓ | | ✓ | | | | | | | | | |
| | | 41 / 3,158<br>13.8 years | | Dietary History Questionnaire,<br>**Cola** | Mortality, Lung cancer, male | | 0.50 (0.10 - 2.20) | ✓ | | ✓ | | | | | | | | | |
| **Skin** | | | | | | | | | | | | | | | | | | | |
| Ibiebele 2007 Australia (17490979)[73] | Nambour Skin Cancer Study<br>Prospective Cohort<br>Range:20-69 years<br>M/F<br>Mixed | 161 / 1,360<br>11 years | Histopathology | FFQ,<br>**Soft drinks**:<br>Coke, Pepsi, other soft drinks, cordial, low-calorie colas, other low-calorie soft drinks | Incidence, Squamous cell cancer | T3 vs T1 | 1.22 (0.64 - 2.32) | ✓ | ✓ | ✓ | | | | ✓<br>E | ✓ | | | | ✓ |
| Oh 2019 Singapore (30731173)[132] | Singapore Chinese Health study<br>Prospective Cohort<br>Range:45-74 years<br>M/F<br>Chinese | 609 / 61,321<br>18.3 years | Cancer registry, birth and death registry | Semi-quantitative FFQ,<br>**Carbonated soft drinks**:<br>Soda | Incidence, non-Melanoma skin cancer | ≥3 vs <1 times/week | 0.92 (0.58 - 1.46)<br>Ptrend:0.89 | ✓ | ✓ | ✓ | ✓ | ✓ | ✓ | ✓ | D | | | ✓ | |
| **Breast (female)** | | | | | | | | | | | | | | | | | | | |
| Dunneram 2019 UK (30526696)[49] | UK Women Cohort Study<br>Prospective Cohort<br>Age:54.1 years<br>F | 1,625 / 32,289<br>18 years | NHS Central Registry | FFQ,<br>**Soft drinks** | Incidence, Breast cancer | Per 111 g day | 1.00 (0.84 – 1.19) | ✓ | | ✓ | ✓ | | ✓ | ✓<br>E | | ✓ | ✓ | ✓ | |
| Mullee 2019 Denmark,France,Germany,Greece,Italy,Netherlands,Norway,Spain,Sweden,UK (31479109)[126] | European Prospective Investigation into Cancer and nutrition<br>Prospective Cohort<br>Age:50.8 years<br>M/F | 1,402 / 321,081<br>16.4 years | active follow up, cancer registry, death certificate and National death index | ,<br>**Soft drinks**:<br>Carbonated/soft/ isotonic drinks and diluted syrups. | Mortality, Breast cancer | ≥1 vs <1 glasses/month | 1.13 (0.92 - 1.39)<br>Ptrend:0.2 | ✓ | ✓ | ✓ | ✓ | ✓ | ✓ | ✓<br>E<br>* | | ✓ | ✓ | ✓ | |
| Dunneram 2019 UK (30526696)[49] | UK Women Cohort Study<br>Prospective Cohort<br>Age:54.1 years<br>F | 1,030 / 32,289<br>18 years | NHS Central Registry | FFQ,<br>**Soft drinks** | Incidence, Breast cancer, postmenopausal | Per 111g day | 1.03 (0.9 – 1.19) | ✓ | | ✓ | ✓ | | ✓ | | | ✓ | | ✓ | |
| | | 291 / 32,289<br>18 years | | | Incidence, Breast cancer, premenopausal | Per 111g day | 1.04 (0.87 – 1.23) | ✓ | | ✓ | ✓ | | ✓ | | | ✓ | | ✓ | |
| Heath 2020 multi-national (31931881)[68] | European Prospective Investigation into Cancer and nutrition<br>Prospective Cohort<br>Age:52 years<br>M/F | 10,979 / 272,098<br>15 years | population registries or active follow up | Diet Questionnaire,<br>**Soft drinks**:<br>Carbonated/soft/ isotonic drinks and diluted syrups | Incidence, Breast cancer | Per SD increase | -.0045 (-0.0265 – 0.0174) | ✓ | ✓ | ✓ | ✓ | ✓ | ✓ | ✓<br>E | D | | | ✓ | |
| **Endometrial** | | | | | | | | | | | | | | | | | | | |

| Author, Year Country (PMID) | Study name, type Age, sex, ethnicity, characteristics of participants | Cases/ Study size Median follow-up (years) | Case ascertainment | Exposure Assessment, detail | Outcome Sub-population | Comparison | RR (95%CI) Ptrend | Adjustments Age | Sex | Smo | Alc | Obe | Phy | Diet | Med | Mens | Repr | Oth | Skin |
|---|---|---|---|---|---|---|---|---|---|---|---|---|---|---|---|---|---|---|---|
| Dunneram 2018 UK (30526696)[49] | UK Women Cohort Study<br>Prospective Cohort<br>Age:54.1 years<br>F | 86 / 32,289<br>18 years | NHS Central Registry | FFQ,<br>**Soft drinks** | Incidence, Endometrial Cancer | Per 111g day | 0.76 (0.36 – 1.6) | ✓ | | ✓ | ✓ | | ✓ | ✓<br>E | D | ✓ | ✓ | ✓ | |
| Merritt 2015 multi-national (25662427)[117] | European Prospective Investigation into Cancer and nutrition and the Nurses Health studies I & II<br>Pooled Analysis Cohorts<br>F | 1,303 / 301,107 | EPIC: population-based cancer registries, active follow-up, mortality data from cancer or mortality registries; NHS/NHS II: questionnaire, medical records, pathology reports, national death index | Semi-quantitative FFQ,<br>**Soft drinks**:<br>Carbonated drinks | Incidence, Endometrial Cancer, EPIC | Median intake 152.4 vs 0 g/day | 0.99 (0.84 - 1.16)<br>Ptrend:0.81 | ✓ | | ✓ | | ✓ | | ✓<br>E | | ✓ | ✓ | | |
| Friberg 2011 Sweden (21765006)[61] | Swedish Mammography Cohort<br>Prospective Cohort<br>Range:40-74 years<br>F | 729 / 61,226<br>1123934 person-years | linkage to Swedish cancer registry | FFQ,<br>**Soft drinks**:<br>carbonated and noncarbonated drinks | Incidence, Endometrial Adenocarcinoma, Baseline diet | Yes (median intake 0.07 servings/week) vs no | 1.02 (0.87 - 1.18) | ✓ | | | | ✓ | | ✓<br>E | D | | | | |
| | | 304 / 61,226<br>1123934 person-years | | | Incidence, Endometrial Adenocarcinoma, 1997 diet | | 1.15 (0.91 - 1.46) | ✓ | | | | ✓ | | ✓<br>E | D | | | | |
| **Ovarian** | | | | | | | | | | | | | | | | | | | |
| Dunneram 2019 UK (30526696)[49] | UK Women Cohort Study<br>Prospective Cohort<br>Age:54.1 years<br>F | 251 / 32,289<br>18 years | NHS Central Registry | FFQ,<br>**Soft drinks** | Incidence, Ovarian cancer | Per 111g day | 1.07 (0.74 - 1.54) | ✓ | | ✓ | ✓ | | ✓ | ✓<br>E | | ✓ | ✓ | ✓ | |
| Merritt 2016 multi-national (26607939)[118] | European Prospective Investigation into Cancer and nutrition cohort and the Netherlands Cohort study<br>Pooled Analysis Cohorts<br>F | 1,095 / 325,004 | population cancer registries or active follow-up | FFQ,<br>**Carbonated soft drinks**:<br>Soda | Incidence, Epithelial ovarian cancer | ≥ 61.4 vs 0 g/day | 1.15 (0.97 - 1.36)<br>Ptrend:0.21 | | | | | | | ✓<br>E | | ✓ | ✓ | | |
| Tworoger 2008 USA (18213613)[163] | Nurses' Health Study<br>Prospective Cohort<br>Range:30-55 years<br>F<br>Registered nurses | 507 / 80,253 | Self-report verified by medical record | FFQ,<br>**Cola**:<br>Caffeinated soda | Incidence, Ovarian cancer | ≥1 vs 0 cup/week | 0.80 (0.59 - 1.13)<br>Ptrend:0.81 | ✓ | | ✓ | | ✓ | | | | | ✓ | | |
| **Prostate** | | | | | | | | | | | | | | | | | | | |
| Papadimitriou 2020 (31705265)[137] | European Prospective Investigation into Cancer and nutrition<br>Prospective Cohort<br>Age:51.6 years<br>M/F | 5916/ 122,998<br>14 years | active follow up, cancer registry, death certificate and National death index | Semi-quantitative FFQ,<br>**Soft drinks**:<br>Carbonated/soft/ isotonic drinks and diluted syrups. | Incidence, Prostate cancer | Per SD increase | 0.98 (0.95 – 1.01) | ✓ | ✓ | ✓ | ✓ | ✓ | ✓ | ✓<br>E<br>* | | | | ✓ | |
| | | | | | Incidence, Localised prostate cancer | Per SD increase | 0.97 (0.93 – 1.02) | ✓ | ✓ | ✓ | ✓ | ✓ | ✓ | ✓<br>E<br>* | | | | ✓ | |
| | | | | | Incidence, Advanced | Per SD increase | 0.97 (0.91 – 1.03) | ✓ | ✓ | ✓ | ✓ | ✓ | ✓ | ✓<br>E | | | | ✓ | |

| Author, Year Country (PMID) | Study name, type Age, sex, ethnicity, characteristics of participants | Cases/ Study size Median follow-up (years) | Case ascertainment | Exposure Assessment, detail | Outcome Sub-population | Comparison | RR (95%CI) Ptrend | Adjustments Age | Sex | Smo | Alc | Obe | Phy | Diet | Med | Mens | Repr | Oth | Skin |
|---|---|---|---|---|---|---|---|---|---|---|---|---|---|---|---|---|---|---|---|
| | | | | | prostate cancer | | | | | | | | | * | | | | | |
| | | | | | Incidence, Low grade prostate cancer | Per SD increase | 0.98 (0.94 – 1.03) | ✓ | ✓ | ✓ | ✓ | ✓ | ✓ | ✓ E * | | | | ✓ | |
| | | | | | Incidence, High grade prostate cancer | Per SD increase | 0.97 (0.89 – 1.07) | ✓ | ✓ | ✓ | ✓ | ✓ | ✓ | ✓ E * | | | | ✓ | |
| | | | | | Incidence, Aggressive prostate cancer | Per SD increase | 0.95 (0.90 – 1.00) | ✓ | ✓ | ✓ | ✓ | ✓ | ✓ | ✓ E * | | | | ✓ | |
| Mullee 2019 Denmark,France,Germany,Greece,Italy,Netherlands,Norway,Spain,Sweden,UK (31479109)[126] | European Prospective Investigation into Cancer and nutrition Prospective Cohort Age:50.8 years M/F | 907 / 130,662 16.4 years | active follow up, cancer registry, death certificate and National death index | Semi-quantitative FFQ, Soft drinks: Carbonated/soft/ isotonic drinks and diluted syrups. | Mortality, Prostate cancer | ≥1 vs <1 glasses/month | 0.97 (0.77 - 1.24) Ptrend:0.8 | ✓ | ✓ | ✓ | ✓ | ✓ | ✓ | ✓ E * | | | | ✓ | |
| Ellison 2000 Canada (10830580)[50] | Nutrition Canada Survey Historical Cohort Range:50-84 years M | 201 / 3,400 | random sample | FFQ, **Cola** | Incidence / Mortality, Prostate cancer | ≥100 vs 0 ml/day | 1.29 (0.74 - 2.26) | ✓ | | | | | | | | | | | |
| **Kidney** | | | | | | | | | | | | | | | | | | | |
| Li 2024 USA (39396995)[102] | Prostate, Lung, Colorectal and Ovarian Cancer Screening trial Prospective Cohort Age:65.5 years M/F | 410 / 101,668 16.85 years | Medical records and death certificates | FFQ, **Soft drinks**: Regular or diet soft drinks, either caffeinated or decaffeinated. | Incidence, Renal cell carcinoma | Q4 vs Q1 | 1.52 (1.14 - 2.03) Ptrend:0.004 | ✓ | ✓ | ✓ | ✓ | ✓ | | ✓ E | D | | | ✓ | |
| Heath 2021 multi-national (33849969)[66] | European Prospective Investigation into Cancer and nutrition Prospective Cohort Age:52 years M/F | 888 / 389,220 15 years | population registries or active follow up | Diet Questionnaire, **Soft drinks**: Carbonated/soft/ isotonic drinks and diluted syrups. | Incidence, Renal cell carcinoma | Per 100 g day | 1.01 (0.98 - 1.05) | ✓ | ✓ | ✓ | ✓ | ✓ | ✓ | ✓ E * | | | | ✓ | |
| Li 2024 USA (39396995)[102] | Prostate, Lung, Colorectal and Ovarian Cancer Screening trial Prospective Cohort Age:65.5 years M/F | 230 / 101,668 16.85 years | Medical records and death certificates | FFQ, **Soft drinks**: Regular or diet soft drinks, either caffeinated or decaffeinated. | Mortality, Renal cell carcinoma | Q4 vs Q1 | 1.35 (0.93 - 1.96) Ptrend:0.186 | ✓ | ✓ | ✓ | ✓ | ✓ | | ✓ E | D | | | ✓ | |
| Heath 2021 multi-national (33849969)[66] | European Prospective Investigation into Cancer and nutrition Prospective Cohort Age:52 years M/F | 356 / 389,220 15 years | population registries or active follow up | Diet Questionnaire, **Soft drinks**: Carbonated/soft/ isotonic drinks and diluted syrups. | Mortality, Renal cell carcinoma | Per 100 g day | 1.03 (0.98 - 1.09) | ✓ | ✓ | ✓ | ✓ | ✓ | ✓ | ✓ E * | | | | ✓ | |
| Allen 2011 UK (21407222)[36] | The Million Women Study, 1996 Prospective Cohort Range:50-64 years F | 588 / 779,369 5.2 years | National Health Service central registers | FFQ, **Carbonated soft drinks**: Carbonated drinks | Incidence, Renal cell carcinoma | ≥2 vs 0 or<1 drinks/day | 1.18 (0.89 - 1.58) | | | ✓ | | ✓ | | | | | | ✓ | |
| | | | | | | Per 1 drinks /day | 1.05 (0.95 - 1.17) | | | ✓ | | ✓ | | | | | | ✓ | |
| Lee 2007 (17583573)[26] | Pooling Project (n=13 cohorts) Pooled Analysis Cohorts M/F mainly Caucasian | 768 / 244,483 20 years | Self-report, medical record, cancer registries and death registries | FFQ and dietary history, **Carbonated soft drinks**: Soda | Incidence, Renal cancer, male | ≥3/week vs non-drinker | 1.17 (0.83 - 1.64) Ptrend:0.45 | ✓ | | ✓ | ✓ | ✓ | | ✓ E | | | | | |
| | | | | | | Per 1 serving /day | 1.03 (0.97 - 1.09) | ✓ | | ✓ | ✓ | ✓ | | ✓ E | | | | | |

| Author, Year Country (PMID) | Study name, type Age, sex, ethnicity, characteristics of participants | Cases/ Study size Median follow-up (years) | Case ascertainment | Exposure Assessment, detail | Outcome Sub-population | Comparison | RR (95%CI) Ptrend | Adjustments<br>Age | Sex | Smo | Alc | Obe | Phy | Diet | Med | Mens | Repr | Oth | Skin |
|---|---|---|---|---|---|---|---|---|---|---|---|---|---|---|---|---|---|---|---|
| | | 698 /<br>530,469<br>20 years | | | Incidence,<br>Renal cancer,<br>female | ≥3/week vs non-drinker | 1.05 (0.78 - 1.41)<br>Ptrend:0.42 | ✓ | | ✓ | ✓ | ✓ | | ✓<br>E | | | ✓ | | |
| | | | | | | Per 1 serving day | 1.01 (0.95 - 1.07) | ✓ | | ✓ | ✓ | ✓ | | ✓<br>E | | | ✓ | | |
| **Bladder** | | | | | | | | | | | | | | | | | | | |
| Michaud, 1999 USA (10228189)[119] | Health Professionals Follow-up Study<br>Prospective Cohort<br>Range:40-75 years<br>M | / 47,909 | Ongoing or prior study and convenience sampling | FFQ,<br>**Soft drinks**:<br>cola and non-cola sodas (regular, caffeinated, decaffeinated sodas), punch and lemonade | Incidence / Mortality,<br>Bladder cancer | Per 240 ml day | 0.99 (0.90 - 1.08) | ✓ | | ✓ | | | | ✓ | | | | ✓ | |
| Ros 2011 multi-national (20715171)[147] | European Prospective Investigation into Cancer and nutrition<br>Prospective Cohort<br>Range:25-70 years<br>M/F | 513 /<br>233,236<br>9.3 years | cancer registries, health insurance records, pathology rec & active follow up | Validated FFQ,<br>**Soft drinks** | Incidence,<br>Urothelial cancer | (≥99 men; ≥20 women) vs (<8 men; <0 women) ml/day | 1.03 (0.83 - 1.30)<br>Ptrend:0.36 | ✓ | ✓ | ✓ | | | | ✓<br>E | | | | | |
| | | | | | | Per 100 ml day | 1.06 (1.01 - 1.12) | ✓ | ✓ | ✓ | | | | ✓<br>E | | | | | |
| | | 210 /<br>233,236<br>9.3 years | | | Incidence,<br>High-risk urothelial cell carcinoma | Per 100 ml day | 1.02 (0.93 - 1.12) | ✓ | ✓ | ✓ | | | | ✓<br>E | | | | | |
| | | 271 /<br>233,236<br>9.3 years | | | Incidence,<br>Low-risk urothelial cell carcinoma | Per 100 ml day | 1.09 (1.02 - 1.16) | ✓ | ✓ | ✓ | | | | ✓<br>E | | | | | |
| **Brain and spinal cord** | | | | | | | | | | | | | | | | | | | |
| Dubrow 2012 USA (22457000)[48] | NIH- AARP Diet and Health Study<br>Prospective Cohort<br>Age:62.8 years<br>M/F<br>Mostly white<br>Retired | 882 /<br>545,771<br>10.6 years | state cancer registry | Self-administered Questionnaire,<br>**Carbonated soft drinks**:<br>Soft drinks, soda, or pop, including diet or regular | Incidence,<br>Glioma | >2 cans/day vs none | 0.87 (0.65 - 1.15)<br>Ptrend:0.53 | ✓ | ✓ | | | | | ✓<br>E | | | | ✓ | |
| | | 520 /<br>545,771<br>10.6 years | | Self-administered Questionnaire,<br>**Caffeinated and carbonated soft drinks**:<br>caffeinated soda | | | 1.00 (0.71 - 1.40)<br>Ptrend:0.24 | ✓ | ✓ | | | | | ✓<br>E | | | | ✓ | |
| Holick 2010 USA (20056621)[71] | Nurse's Health Study, Nurse's Health Study II, Health Professionals follow-up Study<br>Pooled Analysis Cohorts<br>M/F<br>Health professionals | 335<br>24 years | medical records, pathology reports, death records and cancer registries | FFQ,<br>**Caffeinated and carbonated soft drinks**:<br>Caffeinated carbonated beverages (diet and regular) - Colas | Incidence,<br>Glioma | ≥5 cups/week vs none | 0.75 (0.51 - 1.11)<br>Ptrend:0.13 | ✓ | | | | | | ✓<br>E | | | | | |
| Dubrow 2012 USA (22457000)[48] | NIH- AARP Diet and Health Study<br>Prospective Cohort<br>Age:62.8 years<br>M/F<br>Mostly white<br>Retired | 416 /<br>545,771<br>10.6 years | state cancer registry | Self-administered Questionnaire,<br>**Decaffeinated and carbonated soft drinks**:<br>Decaffeinated soda | Incidence,<br>Glioma | >2 cans/day vs none | 0.85 (0.60 - 1.19)<br>Ptrend:0.94 | ✓ | ✓ | | | | | ✓<br>E | | | | ✓ | |
| Holick 2010 USA (20056621)[71] | Nurse's Health Study, Nurse's Health Study II, Health Professionals follow-up Study<br>Pooled Analysis Cohorts<br>M/F<br>Health professionals | 335<br>24 years | medical records, pathology reports, death records and cancer registries | FFQ,<br>**Decaffeinated and carbonated soft drinks**:<br>Decaffeinated carbonated beverages | Incidence,<br>Glioma | ≥4 cups/week vs None | 0.75 (0.49 - 1.17)<br>Ptrend:0.31 | ✓ | | | | | | ✓<br>E | | | | | |

| Author, Year Country (PMID) | Study name, type Age, sex, ethnicity, characteristics of participants | Cases/ Study size Median follow-up (years) | Case ascertainment | Exposure Assessment, detail | Outcome Sub-population | Comparison | RR (95%CI) Ptrend | Adjustments | | | | | | | | | | | |
|---|---|---|---|---|---|---|---|---|---|---|---|---|---|---|---|---|---|---|---|
| | | | | | | | | Age | Sex | Smo | Alc | Obe | Phy | Diet | Med | Mens | Repr | Oth | Skin |
| | | | | (diet and regular) - lemon-lime, orange, grape, root beer and ginger ale sodas | | | | | | | | | | | | | | | |
| **Thyroid** | | | | | | | | | | | | | | | | | | | |
| Zamora-Ros 2023 multi-national (35907037)[180] | European Prospective Investigation into Cancer and nutrition Prospective Cohort Age:51.1 years M/F | 712 / 450,064 13.9 years | Cancer and mortality registries, pathology registries, active follow up, health insurance records | Questionnaire, **Soft drinks**: Carbonated/soft/isotonic drinks and diluted syrups - sugary & artificially sweetened beverages combined. | Incidence, Thyroid cancer | 107.5 - 4201.7 ml/day vs non consumers | 1.17 (0.90 - 1.54) Ptrend:0.13 | ✓ | ✓ | ✓ | ✓ | ✓ | ✓ | ✓ E | | ✓ | ✓ | ✓ | |
| | | | | | | Per 100 ml /day | 1.05 (1.00 - 1.11) | ✓ | ✓ | ✓ | ✓ | ✓ | ✓ | ✓ E | | ✓ | ✓ | ✓ | |
| | | 573 / 450,064 13.9 years | | | Incidence, Papillary thyroid cancer | 107.6 – 4 to201.7 ml/day vs non consumers | 1.32 (0.98 - 1.79) Ptrend:0.025 | ✓ | ✓ | ✓ | ✓ | ✓ | ✓ | ✓ E | | ✓ | ✓ | ✓ | |
| | | | | | | Per 100 ml /day | 1.07 (1.01 - 1.13) | ✓ | ✓ | ✓ | ✓ | ✓ | ✓ | ✓ E | | ✓ | ✓ | ✓ | |
| | | 108 / 450,064 13.9 years | | | Incidence, Follicular thyroid cancer | 107.6 – 4 to201.7 ml/day vs non consumers | 0.75 (0.37 - 1.53) Ptrend:0.38 | ✓ | ✓ | ✓ | ✓ | ✓ | ✓ | ✓ E | | ✓ | ✓ | ✓ | |
| | | | | | | Per 100 ml /day | 0.92 (0.76 - 1.11) | ✓ | ✓ | ✓ | ✓ | ✓ | ✓ | ✓ E | | ✓ | ✓ | ✓ | |
| **Lymphoid and hematopoietic** | | | | | | | | | | | | | | | | | | | |
| Podoltsev 2020 (31904114)[140] | NIH- AARP Diet and Health Study Prospective Cohort Age:62.8 years M/F Mostly white Retired | 490 / 490,049 15.5 years | Cancer registries, pathology registries, active follow up, health insurance records | Questionnaire, **Caffeinated soft drinks** | Incidence, Myelo-proliferative neoplasms | Yes vs no | 1.07 (0.89 – 1.30) Ptrend: 0.47 | ✓ | ✓ | | | | | | | | | ✓ | |
| | | 190 / 490,049 | | | Incidence, Polycythemia vera | Yes vs no | 1.10 (0.81 – 1.50) Ptrend: 0.52 | ✓ | ✓ | | | | | | | | | ✓ | |
| | | 146 / 490,049 | | | Incidence, Essential thrombo-cythemia | Yes vs no | 1.05 (0.73 – 1.50) Ptrend: 0.80 | ✓ | ✓ | | | | | | | | | ✓ | |
| | | 490 / 490,049 | | Questionnaire, **Decaffeinated soft drinks** | Incidence, Myelo-proliferative neoplasms | Yes vs no | 0.99 (0.73 – 1.33) Ptrend: 0.93 | ✓ | ✓ | | | | | | | | | ✓ | |
| | | 190 / 490,049 | | | Incidence, Polycythemia vera | Yes vs no | 1.06 (0.66 – 1.68) Ptrend: 0.81 | ✓ | ✓ | | | | | | | | | ✓ | |
| | | 146 / 490,049 | | | Incidence, Essential thrombo-cythemia | Yes vs no | 0.92 (0.52 – 1.62) Ptrend: 0.94 | ✓ | ✓ | | | | | | | | | ✓ | |

**Age**: age; **Alc**: alcohol; **Diet**: dietary factors and energy intake (E), or * adjustment for other soft drinks not under investigation; **Med**: medical-related factors (e.g. infections, diseases in the breast, digestive tract, gallbladder, liver, or pancreas, cancer screening, precancerous conditions, or pre-existing skin cancer) or diabetes (D); **Mens**: menstrual characteristics (e.g. age at menarche, menopausal status/duration, age at menopause, or type of menopause); **Obe**: obesity (adiposity measures); **Oth**: others (e.g. family history/genetic factor, race/ethnicity, ancestry, socioeconomic factors, or marriage); **Phy**: physical activity; **Repr**: reproductive or hormonal factors (e.g. parity, age at first/last birth, postmenopausal hormone use, or oral contraceptive use); **Sex**: sex; **Skin**: skin-related factors (e.g. UV light/sun exposure, sunburns, time spent outdoors, sunscreen use, indoor tanning, moles, fair skin, red/blond hair, blue/green eyes, or freckles); **Smo**: smoking.

#### Supplementary Table 15.List of associations between sugar sweetened beverages and cancer incidence or mortality outcomes with only one available study.

| Author Year (PMID) | Cancer site | Outcome type | Study name | Sex | Cases / cohort size | RR (95%CI) | p-value for trend/ linear analysis | Comparison |
|---|---|---|---|---|---|---|---|---|
| **Sugar sweetened beverages** | | | | | | | | |
| Wu 2023 (37810918)[172] | Head and neck | Incidence | PLCO | M/F | 279/101755 | 1.14 (0.82-1.58) | 0.087 | ≥318.19 vs ≤31.95 g/day |
| McCullough 2022 (36107009)[113] | Larynx, oral cavity, and pharynx | Mortality | CPS II | M/F | 1852/934777 | 1.04 (0.87-1.24) | 0.963 | ≥2 vs 0 drinks/day |
| McCullough 2022 (36107009)[113] | Oesophageal | Mortality | CPS II | M/F | 2727/934777 | 0.87 (0.76-1.03) | 0.317 | ≥2 vs 0 drinks/day |
| Khairan 2023 (36167978)[84] | Stomach non-cardia | Incidence | JPHC | F | 650/39,353 | 1.01 (0.78-1.32) | 0.56 | ≥278.1 vs 0-0.04 mL/day |
| Khairan 2023 (36167978)[84] | Stomach non-cardia | Incidence | JPHC | M | 650/35,102 | 1.03 (0.86-1.24) | 0.88 | ≥398.6 vs 0-30.5 mL/day |
| Acuna 2023 (36789943)[34] | Stomach distal | Incidence | MEC | M/F | N/A/176752 | 0.99 (0.97-1.01) | N/A | Per 1-point increase in AHEI-2010 |
| Acuna 2023 (36789943)[34] | Stomach distal | Incidence | MEC | M/F | N/A/176752 | 0.96 (0.90-1.01) | N/A | Per 1-point increase in DASH score |
| McCullough 2022 (36107009)[113] | Stomach cancer | Mortality | CPS II | M/F | 2798/934777 | 1.08 (0.94-1.25) | 0.128 | ≥2 vs 0 drinks/day |
| Hur 2021 (33958435)[72] | Early-onset colorectal | Incidence | NHS II | F | 109/95464 | 2.18 (1.10-4.35) | 0.03 | ≥2 servings/day vs ≤0.9 serving/week |
| McCullough 2022 (36107009)[113] | Small intestine | Mortality | CPS II | M/F | 324/934777 | 1.26 (0.84-1.90) | 0.609 | ≥2 vs 0 drinks/day |
| McCullough 2022 (36107009)[113] | Melanoma | Mortality | CPS II | M/F | 2130/934777 | 0.88 (0.74-1.06) | 0.682 | ≥2 vs 0 drinks/day |
| Nomura 2016 (27578546)[128] | ER-/PR- breast | Incidence | BWHS | F | 205/49103 | 0.83 (0.44-1.54) | 0.37 | 0 vs >250 g/day |
| Nomura 2016 (27578546)[128] | ER+/PR+ breast | Incidence | BWHS | F | 1827/49103 | 0.55 (0.32-0.93) | 0.07 | 0 vs >250 g/day |
| Nomura 2016 (27578546)[128] | ER+ or PR+ breast | Incidence | BWHS | F | 881/49103 | 0.95 (0.40-2.23) | 0.98 | 0 vs >250 g/day |
| Inoue-Choi 2013 (24273064)[74] | Type II endometrial | Incidence | IWHS | F | 185/23039 | 1.05 (0.53-2.06) | 0.63 | 9.9-72.3 vs 0-1.3 servings/week |
| McCullough 2022 (36107009)[113] | Endometrial | Mortality | CPS II | F | 1693/518,464 | 1.17 (0.95-1.45) | 0.136 | ≥2 vs 0 drinks/day |
| McCullough 2022 (36107009)[113] | Ovarian | Mortality | CPS II | F | 3225/518,464 | 0.99 (0.84-1.17) | 0.761 | ≥2 vs 0 drinks/day |
| Hodge 2018 (29463332)[70] | Aggressive prostate | Incidence | MCCS | M | 433/14,101 | 1.08 (0.78-1.50) | 0.47 | ≥1/day vs <1/month |
| Drake 2012 (23134882)[47] | Low risk prostate | Incidence | MDCS | M | 332/8128 | 1.06 (0.82 – 1.78) | 0.661 | Q5 vs Q1 |
| Drake 2012 (23134882)[47] | symptomatic prostate | Incidence | MDCS | M | 238/8128 | 1.38 (1.04 – 1.84) | 0.066 | Q5 vs Q1 |
| McCullough 2022 (36107009)[113] | Bladder | Mortality | CPS II | M/F | 3419/ 934777 | 1.11 (0.97-1.27) | 0.050 | ≥2 vs 0 drinks/day |
| McCullough 2022 (36107009)[113] | Brain cancer | Mortality | CPS II | M/F | 3402/934777 | 0.96 (0.83-1.11) | 0.905 | ≥2 vs 0 drinks/day |
| McCullough 2022 (36107009)[113] | Leukaemia | Mortality | CPS II | M/F | 6319/934777 | 1.04 (0.94-1.15) | 0.375 | ≥2 vs 0 drinks/day |
| McCullough 2022 (36107009)[113] | Multiple Myeloma | Mortality | CPS II | M/F | 3129/934777 | 0.97 (0.84-1.11) | 0.499 | ≥2 vs 0 drinks/day |

| Author Year (PMID) | Cancer site | Outcome type | Study name | Sex | Cases / cohort size | RR (95%CI) | p-value for trend/ linear analysis | Comparison |
|---|---|---|---|---|---|---|---|---|
| McCullough 2022 (36107009)[113] | Non-Hodgkin Lymphoma | Mortality | CPS II | M/F | 6600/934777 | 1.16 (1.05-1.28) | 0.001 | ≥2 vs 0 drinks/day |
| **Sugar sweetened beverages without fruit drinks** | | | | | | | | |
| Pacheco 2019 (31596902)[135] | Colorectal | Incidence | CTS | F | 1318/99798 | 0.98 (0.65-1.46) | 0.730 | ≥1 serving/day vs rare or never |
| Zhao 2023 (37552302)[186] | Liver | Incidence | WHI | F | 163/98786 | 1.66 (0.89-3.09) | 0.17 | ≥1 serving/day vs never to ≤3 servings/month |
| **Sweetened bottled water or tea** | | | | | | | | |
| Pacheco 2019 (31596902)[135] | Colorectal | Incidence | CTS | F | 1318/99798 | 1.21 (0.91-1.60) | 0.29 | ≥1 serving/day vs rare or never |
| **Caffeinated sugar sweetened cola** | | | | | | | | |
| Ishitani 2008 (18852405)[75] | Invasive | Incidence | WHS | F | 1181/38432 | 1.17 (0.87-1.57) | 0.40 | ≥1 can/day vs 0 |
| **Decaffeinated sugar sweetened cola** | | | | | | | | |
| Ishitani 2008 (18852405)[75] | Invasive breast | Incidence | WHS | F | 1170/38432 | 0.83 (0.62-1.11) | 0.15 | ≥1 can/week vs almost never |
| **Red fruit drinks** | | | | | | | | |
| Kjaerheim 1998 9486469)[90] | Oral cavity, larynx, pharynx, and oesophagus | Incidence | Norwegian Men UADT | M | 63/10900 | 0.90 (0.50-1.60) | N/A | >6 vs <1 servings/month |
| **Fruit drinks** | | | | | | | | |
| Pacheco 2019 (31596902)[135] | Colorectal | Incidence | CTS | F | 1318/99798 | 0.48 (0.12-1.94) | 0.29 | ≥1 serving/day vs rare or never |
| Allen 2011 (21407222)[36] | Renal cell | Incidence | MWS | F | 588/779369 | 1.10 (0.84-1.45) | 0.50 | ≥2 vs 0-0.9 drinks/day |
| **Lemonade** | | | | | | | | |
| Kjaerheim 1998 9486469)[90] | Oral cavity, larynx, pharynx, and oesophagus | Incidence | Norwegian Men UADT | M | 65/10900 | 1.20 (0.60-2.20) | N/A | >6 vs <1 servings/month |
| **Substitution of SSBs with ASBs** | | | | | | | | |
| Hur 2021 (33958435)[72] | Early-onset colorectal | Incidence | NHS II | F | 109/95464 | 0.83 (0.69-0.99) | 0.04 | Per 1 serving/day |
| **Substitution of SSBs with fruit juice** | | | | | | | | |
| Hur 2021 (33958435)[72] | Early-onset colorectal | Incidence | NHS II | F | 109/95464 | 1.02 (0.61-1.69) | 0.95 | Per 1 serving/day |

Abbreviations: AHEI, Alternative Healthy eating Index; DASH, Dietary Approaches to Stop Hypertension; ASBs, artificially sweetened beverages; SSBs, sugar sweetened beverages.

**Supplementary Table 16. List of associations between artificially sweetened beverages and cancer incidence or mortality outcomes with only one available study.**

| Author Year (PMID) | Cancer site | Outcome type | Study name | Sex | Cases / cohort size | RR (95%CI) | p-value for trend/ linear analysis | Comparison |
|---|---|---|---|---|---|---|---|---|
| **Artificially sweetened beverages** | | | | | | | | |
| McCullough 2022 (36107009)[113] | Larynx/oral cavity/pharynx | Mortality | CPS II | M/F | 1852/934777 | 1.13 (0.94-1.37) | 0.107 | ≥2 servings/day vs never |
| McCullough 2022 (36107009)[113] | Oesophageal | Mortality | CPS II | M/F | 2727/934777 | 0.92 (0.78-1.08) | 0.155 | ≥2 servings/day vs never |
| Hodge 2018 (29463332)[70] | Gastric cardia | Incidence | MCCS | M/F | 165/35593 | 1.03 (0.53-1.98) | 0.46 | ≥1 serving/day vs <0.9 serving/month |
| McCullough 2022 (36107009)[113] | Stomach | Mortality | CPS II | M/F | 2798/934777 | 1.10 (0.94-1.28) | 0.203 | ≥2 servings/day vs never |
| Jin 2023 (38004195)[78] | Colon | Incidence | UK Biobank | M/F | 842/118210 | 0.98 (0.91-1.06) | 0.588 | Per one standard deviation |
| Jin 2023 (38004195)[78] | Rectal | Incidence | UK Biobank | M/F | 842/118210 | 0.97 (0.86-1.10) | 0.670 | Per one standard deviation |
| Hur 2021 (33958435)[72] | Early-onset colorectal | Incidence | NHS II | F | 109/95464 | 0.83 (0.47-1.47) | 0.24 | ≥2 servings/day vs <0.9 serving/week |
| McCullough 2022 (36107009[113] | Small intestine | Mortality | CPS II | M/F | 324/934777 | 1.28 (0.85-1.93) | 0.244 | ≥2 servings/day vs never |
| McCullough 2022 (36107009)[113] | Gallbladder | Mortality | CPS II | M/F | 1024/934777 | 1.15 (0.90-1.46) | 0.105 | ≥2 servings/day vs never |
| You 2022 (36424907)[177] | Lung | Incidence | PLCO | F | 1517/154897 | 0.78 (0.61-0.99) | 0.043 | Intake vs no intake |
| You 2022 (36424907)[177] | Lung | Incidence | PLCO | M | 882/154897 | 1.08 (0.82-1.42) | 0.574 | Intake vs no intake |
| McCullough 2022 (36107009)[113] | Melanoma | Mortality | CPS II | M/F | 2130/934777 | 1.02 (0.86-1.22) | 0.841 | ≥2 servings/day vs never |
| McCullough 2022 (36107009)[113] | Endometrial | Mortality | CPS II | F | 1693/518,464 | 1.01 (0.85-1.21) | 0.878 | ≥2 servings/day vs never |
| McCullough 2022 (36107009)[113] | Ovarian | Mortality | CPS II | F | 3225/518,464 | 1.01 (0.89-1.15) | 0.284 | ≥2 servings/day vs never |
| Hodge 2018 (29463332)[70] | Aggressive prostate | Incidence | MCCS | M | 433/14,101 | 0.91 (0.60-1.38) | 0.66 | ≥1/day vs <1/month |
| Ringel 2022 (36601047)[144] | Bladder | Incidence | WHI | F | 448/80388 | 0.75 (0.53-1.06) | N/A | ≥1 serving/day vs never or <1/week |
| McCullough 2022 (36107009)[113] | Bladder | Mortality | CPS II | M/F | 3419/934777 | 1.02 (0.88-1.18) | 0.922 | ≥2 servings/day vs never |
| McCullough 2022 (36107009)[113] | Brain | Mortality | CPS II | M/F | 3402/934777 | 0.97 (0.84-1.11) | 0.737 | ≥2 servings/day vs never |
| Zamora-Ros 2023 (35907037)[180] | Thyroid | Incidence | EPIC | M/F | 495/450064 | 1.16 (0.80-1.69) | 0.26 | 107.5–4201.7 mL/day vs 0 |
| Zamora-Ros 2023 (35907037)[180] | Papillary thyroid | Incidence | EPIC | M/F | 469/450064 | 1.51 (1.04-2.19) | 0.16 | 107.6 – 4201.7 mL/day vs 0 |
| Zamora-Ros 2023 (35907037)[180] | Follicular thyroid | Incidence | EPIC | M/F | 86/450064 | 0.38 (0.11-1.30) | N/A | 107.6 – 4201.7 mL/day vs 0 |
| McCullough 2022 (36107009)[113] | Leukaemia | Mortality | CPS II | M/F | 6319/934777 | 0.97 (0.87-1.07) | 0.444 | ≥2 servings/day vs never |
| McCullough 2022 (36107009)[113] | Multiple Myeloma | Mortality | CPS II | M/F | 3511/934777 | 1.03 (0.90-1.17) | 0.707 | ≥2 servings/day vs never |
| McCullough 2022 (36107009)[113] | Non-Hodgkin Lymphoma | Mortality | CPS II | M/F | 6600/934777 | 1.01 (0.92-1.12) | 0.870 | ≥2 servings/day vs never |

| Author Year (PMID) | Cancer site | Outcome type | Study name | Sex | Cases / cohort size | RR (95%CI) | p-value for trend/ linear analysis | Comparison |
|---|---|---|---|---|---|---|---|---|
| **Artificially sweetened caffeinated cola** | | | | | | | | |
| Ishitani 2008 (18852405)[75] | Invasive breast | Incidence | WHS | F | 1179/38432 | 0.88 (0.68-1.13) | 0.16 | ≥2 servings/day vs almost never |
| **Artificially sweetened decaffeinated cola** | | | | | | | | |
| Ishitani 2008 (18852405)[75] | Invasive breast | Incidence | WHS | F | 1165/38432 | 1.30 (0.99-1.71) | 0.25 | ≥2 servings/day vs almost never |
| **Caffeinated diet soft drinks** | | | | | | | | |
| Podoltsev 2020 (31904114) [140] | Myeloproliferative neoplasms | Incidence | NIH-AARP | M/F | 490/463,049 | 1.01 (0.81 – 1.27) | 0.98 | Yes vs no |
| Podoltsev 2020 (31904114) [140] | Polycythemia vera | Incidence | NIH-AARP | M/F | 190/463,049 | 0.96 (0.67 – 1.38) | 0.82 | Yes vs no |
| Podoltsev 2020 (31904114) [140] | Essential thrombocythemia | Incidence | NIH-AARP | M/F | 146/463,049 | 1.01 (0.67 – 1.51) | 0.97 | Yes vs no |
| **Decaffeinated diet soft drinks** | | | | | | | | |
| Podoltsev 2020 (31904114) [140] | Myeloproliferative neoplasms | Incidence | NIH-AARP | M/F | 490/463,049 | 0.95 (0.78 – 1.16) | 0.62 | Yes vs no |
| Podoltsev 2020 (31904114) [140] | Polycythemia vera | Incidence | NIH-AARP | M/F | 190/463,049 | 0.91 (0.65 – 1.26) | 0.56 | Yes vs no |
| Podoltsev 2020 (31904114) [140] | Essential thrombocythemia | Incidence | NIH-AARP | M/F | 146/463,049 | 0.99 (0.69 – 1.42) | 0.94 | Yes vs no |

**Supplementary Table 17. List of associations between juices and cancer incidence or mortality outcomes with only one available study.**

| Author Year (PMID) | Cancer site | Outcome type | Study name | Sex | Cases / cohort size | RR (95%CI) | p-value for trend/ linear analysis | Comparison |
|---|---|---|---|---|---|---|---|---|
| **Fruit juice** | | | | | | | | |
| Freedman 2008 (18092323[59] | Oral squamous cell | Incidence | NIH-AARP | M/F | 319/490802 | 1.10 (0.78-1.53) | 0.556 | 5.2-6.8 vs 1.2-1.8 serving/1000 kcal |
| Freedman 2008 (18092323)[59] | Oropharynx and hypopharynx squamous cell | Incidence | NIH-AARP | M/F | 142/490802 | 1.05 (0.63-1.74) | 0.809 | 5.2-6.8 vs 1.2-1.8 serving/1000 kcal |
| Freedman 2007 (17691111)[58] | Oesophageal adenocarcinoma | Incidence | NIH-AARP | M/F | 103/490802 | 0.85 (0.55-1.33) | 0.863 | 1.37 vs 0 servings/1000 kcal |
| Freedman 2007 (17691111)[58] | Oesophageal squamous cell | Incidence | NIH-AARP | M/F | 213/490802 | 0.83 (0.45-1.52) | 0.766 | 1.37 vs 0 servings/1000 kcal |
| Freedman 2008 (18166992)[60] | Stomach | Incidence | NIH-AARP | M/F | 394/490802 | 0.97 (0.83-1.13) | N/A | Per 1 serving/100 kcal |
| Jin 2023 (38004195)[78] | Colon | Incidence | UK Biobank | M/F | 842/118210 | 1.03 (0.97-1.11) | 0.875 | Per 1 SD/day |
| Hur 2021 (33958435)[72] | Early-onset colorectal | Incidence | NHS | F | 109/95464 | 1.13 (0.15-8.60) | 0.63 | ≥2 vs ≤0.9 serving/week |
| Jin 2023 (38004195)[78] | Rectal | Incidence | UK Biobank | M/F | 359/118210 | 0.95 (0.85-1.06) | 0.340 | Per 1 SD/day |
| Freedman 2008 (18092323)[59] | Laryngeal squamous cell | Incidence | NIH-AARP | M/F | 279/490802 | 0.85 (0.59-1.22) | 0.742 | 5.2-6.8 vs 1.2-1.8 serving/1000 kcal |
| Ozasa 2001 (11749690)[134] | Lung cancer | Mortality | JACC | F | 75/98248 | 0.95 (0.56-1.63) | 0.87 | 12-16 times/month vs ≤1-2 times/month |
| Ozasa 2001 (11749690)[134] | Lung cancer | Mortality | JACC | M | 283/98248 | 0.90 (0.69-1.18) | 0.45 | 12-16 times/month vs ≤1-2 times/month |
| Fung 2011 (21832271)[63] | ER- breast | Incidence | NHS | F | 827/86621 | 0.95 (0.76-1.19) | 0.43 | 2.9 vs 0.1 serving/day |
| Sonestedt, 2008 (18711147)[156] | ERα- breast | Incidence | MDCS | F | 58/15773 | 1.03 (0.83-1.29) | N/A | Per 1 category |
| Sonestedt, 2008 (18711147)[156] | ERα+ breast | Incidence | MDCS | F | 392/15773 | 1.05 (0.97-1.14) | N/A | Per 1 category |
| Sonestedt, 2008 (18711147)[156] | ERα+/β- breast | Incidence | MDCS | F | 155/15773 | 1.01 (0.88-1.15) | N/A | Per 1 category |
| Sonestedt, 2008 (18711147)[156] | ERα+/β+ breast | Incidence | MDCS | F | 159/15773 | 1.07 (0.94-1.22) | N/A | Per 1 category |
| Sonestedt, 2008 (18711147)[156] | ERβ- breast | Incidence | MDCS | F | 188/15773 | 0.97 (0.65-1.43) | N/A | Per 1 category |
| Sonestedt, 2008 (18711147)[156] | ERβ+ breast | Incidence | MDCS | F | 182/15773 | 1.08 (0.95-1.22) | N/A | Per 1 category |
| Sakauchi 2007 (17571946)[149] | Ovarian | Mortality | JACC | F | 44/64327 | 0.93 (0.39-2.22) | 0.95 | ≥3-4 times/week vs seldom |
| Inoue-Choi 2013 (24273064)[74] | Type I endometrial | Incidence | IWHS | F | 395/23039 | 1.16 (0.87-1.56) | 0.14 | 8.4-63 vs ≤0.01 servings/week |
| Inoue-Choi 2013 (24273064)[74] | Type II endometrial | Incidence | IWHS | F | 86/23039 | 0.97 (0.50-1.88) | 0.99 | 8.4-63 vs ≤0.01 servings/week |
| Kirsh 2007 (17652276)[89] | Extraporastatic prostate | Incidence | PLCO | M | 189/29361 | 1.32 (0.79-2.21) | 0.39 | 3.4 vs 0.1 servings/day |
| Drake 2012 (23134882)[47] | Low risk prostate | Incidence | MDCS | M | 348/8128 | 1.12 (0.87-1.44) | 0.532 | Q 5 vs Q 1 |

| Author Year (PMID) | Cancer site | Outcome type | Study name | Sex | Cases / cohort size | RR (95%CI) | p-value for trend/ linear analysis | Comparison |
|---|---|---|---|---|---|---|---|---|
| Drake 2012 (23134882)[47] | Symptomatic prostate | Incidence | MDCS | M | 248/8128 | 1.14 (0.85-1.52) | 0.784 | Q 5 vs Q 1 |
| Park 2013 (23739308)[138] | Bladder | Incidence | MEC | F | 152/185885 | 0.81 (0.51-1.30) | 0.14 | ≥59.7 vs ≤4.7 g/1000kcal/day |
| Park 2013 (23739308)[138] | Bladder | Incidence | MEC | M | 429/185885 | 0.94 (0.72-1.24) | 0.67 | ≥59.7 vs ≤4.7 g/1000kcal/day |
| **Sweetened fruit juice** | | | | | | | | |
| Mills 1991 2000840[123] | Bladder | Incidence | AHS | M/F | 46/34198 | 0.31 (0.09-1.00) | 0.02 | ≥1 vs 0 times/week |
| **Pure fruit juice** | | | | | | | | |
| Iso 2007 (18260705)[76] | Oesophageal | Mortality | JACC | F | 21/105500 | 0.84 (0.27-2.62) | N/A | ≥3 vs ≤1 times/week |
| Iso 2007 (18260705)[76] | Oesophageal | Mortality | JACC | M | 138/105500 | 0.95 (0.61-1.49) | N/A | ≥3 vs ≤1 times/week |
| Iso 2007 (18260705)[76] | Stomach | Mortality | JACC | F | 327/105550 | 1.28 (0.95-1.74) | N/A | ≥3 vs ≤1 times/week |
| Iso 2007 (18260705)[76] | Stomach | Mortality | JACC | M | 678/105550 | 1.15 (0.93-1.42) | N/A | ≥3 vs ≤1 times/week |
| Arthur 2021 (33220638)[38] | Colon | Incidence | CSDLH | M/F | 192/2633 | 1.15 (0.76-1.73) | 0.528 | ≥6.3 vs ≤1.7 cups/week |
| Iso 2007 (18260705)[76] | Colon | Mortality | JACC | F | 189/105550 | 0.92 (0.65-1.30) | N/A | ≥3 vs ≤1 times/week |
| Iso 2007 (18260705)[76] | Colon | Mortality | JACC | M | 190/105550 | 1.03 (0.69-1.54) | N/A | ≥3 vs ≤1 times/week |
| Iso 2007 (18260705)[76] | Rectal | Mortality | JACC | F | 72/105550 | 0.86 (0.44-1.67) | N/A | ≥3 vs ≤1 times/week |
| Iso 2007 (18260705)[76] | Rectal | Mortality | JACC | M | 150/105550 | 0.69 (0.43-1.11) | N/A | ≥3 vs ≤1 times/week |
| Iso 2007 (18260705)[76] | Liver | Mortality | JACC | F | 195/105550 | 0.91 (0.61-1.36) | N/A | ≥3 vs ≤1 times/week |
| Iso 2007 (18260705)[76] | Liver | Mortality | JACC | M | 412/105550 | 1.40 (1.05-1.85) | <0.05 | ≥3 vs ≤1 times/week |
| Iso 2007 (18260705)[76] | Gallbladder | Mortality | JACC | F | 82/105550 | 1.44 (0.75-2.73) | N/A | ≥3 vs ≤1 times/week |
| Iso 2007 (18260705)[76] | Gallbladder | Mortality | JACC | M | 63/105550 | 0.49 (0.21-1.18) | N/A | ≥3 vs ≤1 times/week |
| Iso 2007 (18260705)[76] | Lung | Mortality | JACC | F | 229/105550 | 1.01 (0.70-1.46) | N/A | ≥3 vs ≤1 times/week |
| Iso 2007 (18260705)[76] | Lung | Mortality | JACC | M | 772/105550 | 0.87 (0.72-1.07) | N/A | ≥3 vs ≤1 times/week |
| Iso 2007 (18260705)[76] | Breast | Mortality | JACC | F | 93/60982 | 1.41 (0.80-2.51) | N/A | ≥3 vs ≤1 times/week |
| Arthur 2021 (33220638)[38] | Endometrial | Incidence | CSDLH | F | 169/2351 | 1.50 (1.00-1.72) | 0.048 | ≥5.7 vs ≤2.5 cups/week |
| Arthur 2021 (33220638)[38] | Type I endometrial | Incidence | CSDLH | F | 125/2351 | 1.63 (1.03-2.60) | 0.040 | ≥5.7 vs ≤2.5 cups/week |
| Iso 2007 (18260705)[76] | Endometrial | Mortality | JACC | F | 29/60982 | 0.84 (0.31-2.23) | N/A | ≥3 vs ≤1 times/week |
| Arthur 2021 (33220638)[38] | Ovarian | Incidence | CSDLH | F | 98/2554 | 1.48 (0.87-2.54) | 0.145 | ≥5.7 vs ≤2.4 cups/week |
| Arthur 2021 (33220638)[38] | Epithelial ovarian | Incidence | CSDLH | F | 72/2554 | 1.32 (0.70-2.49) | 0.352 | ≥5.7 vs ≤2.4 cups/week |
| Chazelas 2019 (31292122)[44] | Prostate | Incidence | NutriNet-Santé | M | 291/21,533 | 0.98 (0.78-1.22) | 0.80 | Per 100 ml/day |

| Author Year (PMID) | Cancer site | Outcome type | Study name | Sex | Cases / cohort size | RR (95%CI) | p-value for trend/ linear analysis | Comparison |
|---|---|---|---|---|---|---|---|---|
| Iso 2007 (18260705)[76] | Prostate | Mortality | JACC | M | 152/44518 | 1.01 (0.63-1.61) | N/A | ≥3 vs ≤1 times/week |
| Iso 2007 (18260705)[76] | Kidney | Mortality | JACC | F | 15/105550 | 2.01 (0.39-10.30) | N/A | ≥3 vs ≤1 times/week |
| Iso 2007 (18260705)[76] | Kidney | Mortality | JACC | M | 41/105550 | 0.87 (0.36-2.12) | N/A | ≥3 vs ≤1 times/week |
| Iso 2007 (18260705)[76] | Bladder | Mortality | JACC | F | 38/105550 | 0.42 (0.15-1.17) | <0.10 | ≥3 vs ≤1 times/week |
| Iso 2007 (18260705)[76] | Bladder | Mortality | JACC | M | 81/105550 | 0.44 (0.21-0.90) | <0.05 | ≥3 vs ≤1 times/week |
| **Citrus juice** | | | | | | | | |
| Betterweck 1998 9801014)[43] | Stomach | Incidence | NLCS | M/F | 282/120852 | 0.92 (0.82-1.03) | N/A | Per 25 g/day |
| Voorrips 2000 (11117618)[165] | Colon | Incidence | NLCS | F | N/A/120852 | 0.99 | N/A | Per 25 g/day |
| Voorrips 2000 (11117618)[165] | Colon | Incidence | NLCS | M | N/A/120852 | 1.00 | N/A | Per 25 g/day |
| Voorrips 2000 (11117618)[165] | Rectal | Incidence | NLCS | F | N/A/120852 | 1.07 | N/A | Per 25 g/day |
| Voorrips 2000 (11117618)[165] | Rectal | Incidence | NLCS | M | N/A/120852 | 0.87 | N/A | Per 25 g/day |
| Vrieling 2009 (19107929)[167] | Pancreas | Incidence | EPIC | M/F | 555/478400 | 0.91 (0.68-1.22) | 0.13 | Per 25 g/day |
| Mahamat-Saleh 2020 (32710289)[107] | Basal cell carcinoma | Incidence | EPIC | M/F | 5604/270112 | 1.10 (1.01-1.19) | 0.008 | Q 4 vs Q 1 |
| Mahamat-Saleh 2020 (32710289)[107] | Squamous cell carcinoma | Incidence | EPIC | M/F | 1165/270112 | 1.23 (1.05-1.44) | 0.004 | Q 4 vs Q 1 |
| Stolzenberg-Solomon 2006 (16600944)[160] | Breast | Incidence | PLCO | F | 91/31411 | 0.86 (0.43-1.72) | 0.79 | ≥180.5 vs 0-5.61 g/day |
| Zeegers 2001 (11700259)[181] | Bladder | Incidence | NLCS | M/F | N/A/350 | 1.00 (0.93-1.08) | N/A | Per 25 g/day |
| Xiao 2014 (24686895)[174] | Thyroid | Incidence | NIH-AARP | M/F | 586/491840 | 1.55 (0.97-2.49) | 0.02 | Q 5 vs Q 1 |
| Xiao 2014 (24686895)[174] | Papillary thyroid | Incidence | NIH-AARP | M | 170/491840 | 1.39 (0.86-2.27) | 0.33 | Q 5 vs Q 1 |
| Xiao 2014 (24686895)[174] | Papillary thyroid | Incidence | NIH-AARP | F | 247/491840 | 1.79 (1.17-2.75) | 0.01 | Q 5 vs Q 1 |
| **Orange juice** | | | | | | | | |
| Marley 2022 (34282681)[111] | Non-melanoma skin | Incidence | UK Biobank | M/F | 9389/197372 | 1.06 (0.90-1.24) | 0.91 | ≥1 serving/day vs none |
| Michaud 1999 (10203279)[120] | Bladder | Incidence | HPFS | M | N/A/47909 | Not significant | N/A | Not reported |
| **Pure orange juice** | | | | | | | | |
| Khairan 2023 (36167978)[84] | Stomach | Incidence | JPHC | F | 676/39,353 | 1.04 (0.87-1.24) | 0.80 | Q3 vs Q1 |
| Khairan 2023 (36167978)[84] | Stomach | Incidence | JPHC | M | 1465/35,102 | 0.97 (0.86-1.10) | 0.66 | Q3 vs Q1 |
| Yamagiwa 2019 (30255932)[176] | Pancreas | Incidence | JPHC | M/F | 577/90185 | 0.94 (0.77-1.15) | 0.616 | 28-1700 vs ≤0.09 g/day |
| **Grapefruit juice** | | | | | | | | |
| Marley 2022 (34282681)[111] | Non-melanoma skin | Incidence | UK Biobank | M/F | 9602/197372 | 0.94 (0.44-1.75) | 0.74 | ≥1 serving/day vs none |

| Author Year (PMID) | Cancer site | Outcome type | Study name | Sex | Cases / cohort size | RR (95%CI) | p-value for trend/ linear analysis | Comparison |
|---|---|---|---|---|---|---|---|---|
| Kim 2008 (18026192)[86] | Postmenopausal breast | Incidence | NHS | F | 3570/77050 | 1.02 (0.85-1.22) | 0.95 | ≥0.5 vs 0 glasses/day |
| Michaud 1999 (10203279)[120] | Bladder | Incidence | HPFS | M | N/A/47909 | Not significant | N/A | Not reported |
| **Other fruit juices (not citrus)** | | | | | | | | |
| Voorrips 2000 (11117618)[165] | Colon | Incidence | NLCS | F | N/A/120852 | 1.00 | N/A | Per 25 g/day |
| Voorrips 2000 (11117618)[165] | Colon | Incidence | NLCS | M | N/A/120852 | 1.04 | N/A | Per 25 g/day |
| Voorrips 2000 (11117618)[165] | Rectal | Incidence | NLCS | F | N/A/120852 | 0.87 | N/A | Per 25 g/day |
| Voorrips 2000 (11117618)[165] | Rectal | Incidence | NLCS | M | N/A/120852 | 0.93 | N/A | Per 25 g/day |
| Zhao 2023 (36811575)[185] | Liver | Incidence | NIH-AARP | M/F | 947/485403 | 0.97 (0.80-1.17) | 0.99 | Q1 vs Q5 |
| Voorrips 2000 (10710193)[166] | Lung | Incidence | NLCS | M/F | 963/120852 | 0.90 (0.70-1.20) | N/A | 20 vs 0 g/day |
| Mommers 2005 (16104037)[124] | Epithelial ovarian | Incidence | NLCS | F | N/A/62573 | 1.08 (0.98-1.18) | N/A | Per 25 g/day |
| **Apple juice** | | | | | | | | |
| Michaud 1999 (10203279)[120] | Bladder | Incidence | HPFS | M | N/A/47909 | Not significant | N/A | Not reported |
| Yamagiwa 2019 (30255932)[176] | Pancreas | Incidence | JPHC | M/F | N/A/90185 | 0.89 (0.72-1.09) | 0.27 | ≥6 vs ≤1 glass /week |
| **Other fruit juices (not citrus and apple)** | | | | | | | | |
| Farvid 2016 (27170029)[51] | Breast | Incidence | NHS II | F | 3235/90476 | 1.01 (0.98-1.03) | N/A | Per 2 servings/week |
| Michaud 1999 (10203279)[120] | Bladder | Incidence | HPFS | M | N/A/47909 | Not significant | N/A | Not reported |
| **Fruit and vegetable juice** | | | | | | | | |
| Kanehara 2024 (38795743)[81] | Rectal | Incidence | MEC | M/F | 1014/192651 | 0.79 (0.64-0.97) | 0.02 | 268.5-492.5 vs 0-1.1 g/1000 kcal |
| Stepien 2016 (25528243)[159] | Liver | Incidence | EPIC | M/F | 140/477206 | 1.38 (0.80-2.28) | 0.02 | ≥6 vs <1 glass /week |
| Stepien 2016 (25528243)[159] | Intrahepatic bile duct | Incidence | EPIC | M/F | 66/477206 | 1.04 (1.00-1.08) | N/A | Per 1 serving/week |
| Olsen 2003 (12840204)[133] | ER- breast | Incidence | DCH | F | 91/23798 | 0.67 (0.42-1.08) | N/A | Per 100 g/day |
| Olsen 2003 (12840204)[133] | ER+ breast | Incidence | DCH | F | 303/23798 | 1.04 (0.89-1.22) | N/A | Per 100 g/day |
| Merritt 2015 (25662427)[117] | Endometrial | Incidence | EPIC | F | 1303/301107 | 1.07 (0.91-1.26) | 0.35 | 148.6 vs 0 g/day |
| Merritt 2016 (26607939)[118] | Epithelial ovarian | Incidence | EPIC | F | 1095/325004 | 1.06 (0.89-1.27) | 0.94 | 149.4 vs 0 g/day |
| Heath 2021 (33849969)[66] | Kidney | Mortality | EPIC | M/F | 356/389220 | 1.08 (0.99-1.17) | 0.11 | Per 100 mL/day |
| Ros 2011 (20715171)[147] | Urothelial | Incidence | EPIC | M/F | 513/233236 | 1.32 (1.05-1.68) | 0.02 | Q 3 vs Q 1 |
| Ros 2011 (20715171)[147] | High risk urothelial | Incidence | EPIC | F | 210/233236 | 1.21 (0.85-1.71) | 0.22 | Q 3 vs Q 1 |
| Ros 2011 (20715171)[147] | Low risk urothelial | Incidence | EPIC | F | 271/233236 | 1.53 (1.12-2.09) | 0.01 | Q 3 vs Q 1 |

| Author Year (PMID) | Cancer site | Outcome type | Study name | Sex | Cases / cohort size | RR (95%CI) | p-value for trend/ linear analysis | Comparison |
|---|---|---|---|---|---|---|---|---|
| Zamora-Ros 2018 (28688112)[179] | Thyroid | Incidence | EPIC | M/F | 748/476108 | 1.23 (0.98-1.53) | N/A | ≤95 vs ≤0.9 g/day |
| Zamora-Ros 2018 (28688112)[179] | Papillary thyroid | Incidence | EPIC | M/F | 601/476108 | 1.69 (0.92-3.08) | N/A | ≤95 vs ≤0.9 g/day |
| Zamora-Ros 2018 (28688112)[179] | Follicular thyroid | Incidence | EPIC | M/F | 109/476108 | 1.12 (0.88-1.43) | N/A | ≤95 vs ≤0.9 g/day |
| **Pure fruit and vegetable juice** | | | | | | | | |
| Dunneram, 2019 (30526696)[49] | Breast | Incidence | UKWCS | F | 1625/32289 | 0.94 (99% CI: 0.83-1.08) | N/A | Per 145 g/day |
| Dunneram, 2019 (30526696)[49] | Postmenopausal breast | Incidence | UKWCS | F | 1030/32289 | 0.99 (99% CI: 0.90-1.09) | N/A | Per 145 g/day |
| Dunneram, 2019 (30526696)[49] | Premenopausal breast | Incidence | UKWCS | F | 291/32289 | 0.89 (99% CI: 0.72-1.10) | N/A | Per 145 g/day |
| Dunneram, 2019 (30526696)[49] | Endometrial | Incidence | UKWCS | F | 86/32289 | 0.89 (99% CI: 0.60-1.33) | N/A | Per 145 g/day |
| Dunneram, 2019 (30526696)[49] | Ovarian | Incidence | UKWCS | F | 251/32289 | 0.94 (99% CI: 0.68-1.31) | N/A | Per 145 g/day |
| **Tomato juice** | | | | | | | | |
| Jiang 2023 (37637049)[77] | Colorectal | Incidence | PLCO | M/F | N/A/101680 | 1.03 (0.84-1.26) | 0.612 | ≥2.97 vs ≤-19.52 g/day |
| Jiang 2023 (37637049)[77] | Colorectal | Mortality | PLCO | M/F | 443/101680 | 1.08 (0.79-1.47) | 0.746 | ≥2.97 vs ≤-19.52 g/day |
| Yamagiwa 2019 (30255932)[176] | Pancreas | Incidence | JPHC | M/F | N/A/90185 | 0.98 (0.79-1.21) | 0.904 | 1-1981 vs ≤0.09 g/day |
| Sesso 2005 (15894655)[153] | Breast | Incidence | WHS | F | 1057/38447 | 0.81 90.59-1.11) | 0.37 | ≥8 vs 0 serving/month |

**Supplementary Table 18. List of associations between soft drinks and cancer incidence or mortality outcomes with only one available study.**

| Author Year (PMID) | Cancer site | Outcome type | Study name | Sex | Cases / cohort size | RR (95%CI) | p-value for trend/ linear analysis | Comparison |
|---|---|---|---|---|---|---|---|---|
| **Soft drinks** | | | | | | | | |
| Stepien 2016 (25528243)[159] | Ampulla of Vater | Incidence | EPIC | M/F | 54/ 477206 | 0.96 (0.90-1.00) | N/A | 1 serving per week |
| Khan 2004 (15075007)[85] | Lung | Mortality | HGCS | M | N/A/ 3185 | 1.10 (0.50-2.10) | N/A | Daily or weekly vs monthly or yearly |
| Khan 2004 (15075007)[85] | Lung | Mortality | HGCS | F | N/A/ 3185 | 1.80 (0.50-6.60) | N/A | Daily or weekly vs monthly or yearly |
| Ibiebele 2007 (17490979)[73] | Skin (squamous cell) | Incidence | NSCS | M/F | 161/ 1360 | 1.22 (0.64-2.32) | N/A | Q3 vs Q1 |
| Dunneram, 2019 (30526696)[49] | Postmenopausal Breast | Incidence | UKWCS | F | 1030/ 32289 | 1.03 (99% CI: 0.90-1.19) | N/A | Per 111 g/day |
| Dunneram, 2019 (30526696)[49] | Premenopausal Breast | Incidence | UKWCS | F | 291/ 32289 | 1.04 (99% CI: 0.87-1.23) | N/A | Per 111 g/day |
| Mullee 2019 (31479109)[126] | Breast | Mortality | EPIC | F | 1402/ 321,081 | 1.13 (0.92-1.39) | 0.20 | ≥1 glass/day vs <1 glass/month |
| Dunneram, 2019 (30526696)[49] | Ovarian | Incidence | UKWCS | F | 251/ 32289 | 1.07 (99% CI: 0.74-1.54) | N/A | Per 111 g/day |
| Papadimitriou, 2020 (31705265) [137] | Prostate | Incidence | EPIC | M | 5916/ 122,998 | 0.98 (0.95 – 1.01) | 0.25 | Per 1-SD increase |
| Papadimitriou, 2020 (31705265) [137] | Localised prostate | Incidence | EPIC | M | NR/ 122,998 | 0.97 (0.93 – 1.02) | 0.26 | Per 1-SD increase |
| Papadimitriou, 2020 (31705265) [137] | Advanced prostate | Incidence | EPIC | M | NR/ 122,998 | 0.97 (0.91 – 1.03) | 0.30 | Per 1-SD increase |
| Papadimitriou, 2020 (31705265) [137] | Low-grade prostate | Incidence | EPIC | M | NR/ 122,998 | 0.98 (0.94 – 1.03) | 0.48 | Per 1-SD increase |
| Papadimitriou, 2020 (31705265) [137] | High-grade | Incidence | EPIC | M | NR/ 122,998 | 0.97 (0.89 – 1.07) | 0.57 | Per 1-SD increase |
| Papadimitriou, 2020 (31705265) [137] | Aggressive | Incidence | EPIC | M | NR/ 122,998 | 0.95 (0.90 – 1.00) | 0.007 | Per 1-SD increase |
| Mullee 2019 (31479109)[126] | Prostate | Mortality | EPIC | M | 907/ 130,662 | 0.97 (0.77-1.24) | 0.80 | ≥1 serving/day vs none |
| Zamora-Ros 2023 (35907037)[180] | Differentiated thyroid | Incidence | EPIC | M/F | 712/ 450064 | 1.05 (1.00-1.11) | N/A | Per 100 mL/day |
| Zamora-Ros 2023 (35907037)[180] | Papillary thyroid | Incidence | EPIC | M/F | 573/ 450064 | 1.07 (1.01-1.03) | N/A | Per 100 mL/day |
| Zamora-Ros 2023 (35907037)[180] | Follicular thyroid | Incidence | EPIC | M/F | 108/ 450064 | 0.92 (0.76-1.11) | N/A | Per 100 mL/day |
| **Carbonated soft drinks** | | | | | | | | |
| Ren 2010 (20395127)[143] | Larynx | Incidence | NIH-AARP | M/F | 307/ 481563 | 0.82 (0.55-1.23) | 0.35 | ≥1 serving/day versus none |
| Ren 2010 (20395127)[143] | Pharynx | Incidence | NIH-AARP | M/F | 178/ 481563 | 0.76 (0.46-1.25) | 0.20 | ≥1 serving/day versus none |
| Ren 2010 (20395127)[143] | Oral cavity | Incidence | NIH-AARP | M/F | 392/ 481563 | 0.77 (0.54-1.09) | 0.31 | ≥1 serving/day versus none |
| Ren 2010 (20395127)[143] | Stomach cardia | Incidence | NIH-AARP | M/F | 231/ 481563 | 0.89 (0.55-1.45) | 0.75 | ≥1 serving/day versus none |
| Ren 2010 (20395127)[143] | Stomach non-cardia | Incidence | NIH-AARP | M/F | 224/ 481563 | 0.75 (0.45-1.24) | 0.30 | ≥1 serving/day versus none |

| Author Year (PMID) | Cancer site | Outcome type | Study name | Sex | Cases / cohort size | RR (95%CI) | p-value for trend/ linear analysis | Comparison |
|---|---|---|---|---|---|---|---|---|
| Ren 2010 (20395127)[143] | Oesophageal squamous cell | Incidence | NIH-AARP | M/F | 123/ 481563 | 0.85 (0.46-1.56) | 0.55 | ≥1 serving/day versus none |
| Ren 2010 (20395127)[143] | Oesophageal adenocarcinoma | Incidence | NIH-AARP | M/F | 305/ 481563 | 1.11 (0.66-1.85) | 0.73 | ≥1 serving/day versus none |
| Mueller 2010 (20142243)[125] | Pancreas adenocarcinoma | Incidence | SCHS | M/F | 140/ 60524 | 1.86 (1.11-3.13) | 0.02 | 5.11 vs 0 servings/week |
| Merritt 2016 (26607939)[118] | Epithelial ovarian | Incidence | EPIC and NLCS | F | 1095/ 325004 | 1.15 (0.97-1.36) | 0.21 | ≥61.4 vs 0 g/day |
| Dubrow 2012 (22457000)[48] | Glioma | Incidence | NIH-AARP | M/F | 882/ 545771 | 0.87 (0.65-1.15) | 0.53 | >2 cans/day vs none |
| **Cola** | | | | | | | | |
| Khan 2004 (15075007)[85] | Colorectal | Mortality | HGCS | F | 14/ 3458 | 2.2 (0.30-17.8) | N/A | Daily or weekly vs monthly or yearly |
| Khan 2004 (15075007)[85] | Lung | Mortality | HGCS | M | N/A/ 3185 | 0.50 (0.10-2.20) | N/A | Daily or weekly vs monthly or yearly |
| Tworoger 2008 (18213613)[163] | Epithelial ovarian | Incidence | NHS | F | 507/ 80253 | 0.80 (0.59-1.13) | 0.81 | ≥1 serving/day vs none |
| Ellison 2000 (18213613)[50] | Prostate | Incidence | Nutrition Canada Survey | M | 201/ 3400 | 1.29 (0.74-2.26) | N/A | ≥100 mL/day vs none |
| **Caffeinated soft drinks** | | | | | | | | |
| Podoltsev 2020 (31904114) [140] | Myeloproliferative neoplasms | Incidence | NIH-AARP | M/F | 490/463,049 | 1.07 (0.89 – 1.30) | 0.47 | Yes vs no |
| Podoltsev 2020 (31904114) [140] | Polycythemia vera | Incidence | NIH-AARP | M/F | 190/463,049 | 1.10 (0.81 – 1.50) | 0.52 | Yes vs no |
| Podoltsev 2020 (31904114) [140] | Essential thrombocythemia | Incidence | NIH-AARP | M/F | 146/463,049 | 1.05 (0.73 – 1.50) | 0.80 | Yes vs no |
| **Decaffeinated soft drinks** | | | | | | | | |
| Podoltsev 2020 (31904114) [140] | Myeloproliferative neoplasms | Incidence | NIH-AARP | M/F | 490/463,049 | 0.99 (0.73 – 1.33) | 0.93 | Yes vs no |
| Podoltsev 2020 (31904114) [140] | Polycythemia vera | Incidence | NIH-AARP | M/F | 190/463,049 | 1.06 (0.66 – 1.68) | 0.81 | Yes vs no |
| Podoltsev 2020 (31904114) [140] | Essential thrombocythemia | Incidence | NIH-AARP | M/F | 146/463,049 | 0.92 (0.52 – 1.62) | 0.77 | Yes vs no |

**Supplementary Table 19. Subgroup analyses of sugar sweetened beverages (per 1 serving [355 mL]/day) and cancer risk.**

| | Studies | RR (95%CI) | $I^2$ (%) | $P_{heterogeneity}$ between subgroups |
|---|---|---|---|---|
| **Stomach cancer incidence, main analysis** | 2 | 0.99 (0.80 - 1.23) | 0 | |
| **Subgroups** | | | | |
| Females | 2 | 1.05 (0.71 - 1.54) | 23% | 0.593 |
| Males | 2 | 0.94 (0.83 - 1.06) | 0 | |
| **Colorectal cancer incidence, main analysis** | 13 | 1.07 (1.00 - 1.14) | 41% | |
| **Subgroups** | | | | |
| North America | 7 | 1.08 (1.00 - 1.17) | 35% | 0.018 |
| Europe | 2 | 1.82 (1.04 - 3.16) | 0 | |
| East/Southeast Asia | 2 | 0.97 (0.91 - 1.04) | 0 | |
| Australia | 1 | 1.17 (0.96 - 1.43) | - | |
| Follow-up ≤10 years | 4 | 1.37 (0.96 - 1.94) | 0 | 0.164 |
| Follow-up >10 years | 9 | 1.06 (0.99 - 1.13) | 50% | |
| Females | 7 | 1.04 (0.89 - 1.20) | 54% | 0.241 |
| Males | 3 | 1.09 (0.86 - 1.38) | 72% | |
| SSBs included fruit juices | 3 | 1.24 (0.93 - 1.65) | 0 | 0.812 |
| SSBs excluded fruit juices | 3 | 1.34 (0.77 - 2.34) | 0 | |
| Baseline exposure measurement | 9 | 1.03 (0.97 - 1.10) | 32% | 0.353 |
| Repeated exposure measurements | 5 | 1.13 (0.95 - 1.34) | 35% | |
| Excluded people with diabetes at baseline | 2 | 1.18 (0.99 – 1.40) | 0 | 0.272 |
| Not excluded people with diabetes at baseline | 11 | 1.06 (0.99 – 1.13) | 41% | |
| Moderate bias for confounding | 8 | 1.04 (0.97 – 1.11) | 41% | 0.112 |
| Serious bias for confounding | 5 | 1.15 (1.03 - 1.29) | 24% | |
| Moderate bias for departure from intended exposure | 5 | 1.15 (0.94 – 1.34) | 40% | 0.413 |
| Critical bias for departure from intended exposure | 8 | 1.04 (0.97 – 1.10) | 31% | |
| **Colorectal cancer mortality, main analysis** | 4 | 1.02 (1.00 - 1.04) | 0 | |
| **Subgroups** | | | | |
| US | 3 | 1.04 (0.97 – 1.11) | 35% | 0.789 |
| Europe | 1 | 1.07 (0.86 – 1.34) | - | |
| Baseline exposure measurement | 2 | 1.02 (1.00 - 1.04) | 0 | 0.216 |

| | Studies | RR (95%CI) | $I^2$ (%) | $P_{heterogeneity}$ between subgroups |
|---|---|---|---|---|
| Repeated exposure measurements | 2 | 1.12 (0.97 – 1.30) | Not reported[1] | |
| Excluded people with diabetes at baseline | 2 | 1.02 (1.00 – 1.04) | 0 | 0.216 |
| Not excluded people with diabetes at baseline | 2 | 1.12 (0.97 – 1.30) | 0 | |
| Moderate bias for confounding | 2 | 1.12 (0.97 – 1.30) | 0 | 0.216 |
| Serious bias for confounding | 2 | 1.02 (1.00 – 1.04) | 0 | |
| Moderate bias for departure from intended exposure | 2 | 1.12 (0.97 – 1.30) | 0 | 0.216 |
| Critical bias for departure from intended exposure | 2 | 1.02 (1.00 – 1.04) | 0 | |
| **Colon cancer incidence, main analysis** | 6 | 1.01 (0.97 - 1.05) | 0 | |
| **Subgroups** | | | | |
| Females | 3 | 1.07 (0.88 - 1.29) | 25% | 0.365 |
| Males | 1 | 0.96 (0.87 - 1.07) | - | |
| **Proximal colon cancer incidence, main analysis** | 5 | 1.03 (0.96 - 1.11) | 60% | |
| **Subgroups** | | | | |
| Age <60 years | 2 | 1.22 (0.93 – 1.61) | Not reported[1] | 0.45 |
| Age 60-69 years | 2 | 1.27 (1.03 – 1.57) | Not reported[1] | |
| Age ≥70 years | 2 | 1.09 (0.87 – 1.36) | Not reported[1] | |
| Females | 3 | 1.02 (0.96 - 1.09) | 0 | 0.314 |
| Males | 2 | 1.12 (0.89 - 1.43) | 52% | |
| Baseline exposure measurement | 3 | 1.00 (0.97 – 1.03) | 0 | 0.084 |
| Repeated exposure measurements | 2 | 1.18 (0.98 – 1.42) | 52% | |
| Critical bias for departure from intended exposure | 3 | 1.00 (0.97 – 1.03) | 0 | 0.084 |
| Moderate bias for departure from intended exposure | 2 | 1.18 (0.98 – 1.42) | 52% | |
| Never smokers | 3 | 1.07 (0.74 - 1.53) | 83% | 0.972 |
| Ever smokers | 3 | 1.07 (0.95 - 1.22) | 0 | |
| Non drinkers | 3 | 1.08 (0.93 - 1.26) | 0 | 0.717 |
| Drinkers | 3 | 1.02 (0.78 - 1.35) | 77% | |
| Low physical activity | 3 | 1.04 (0.75 - 1.46) | 83% | 0.970 |
| High physical activity | 3 | 1.04 (0.90 - 1.19) | 0 | |
| BMI<25 $kgm^2$ | 3 | 1.06 (0.94 - 1.20) | 0 | 0.858 |
| BMI≥25 $kg/m^2$ | 3 | 1.02 (0.67 - 1.56) | 85% | |
| Fruit intake≤1 serving/day | 2 | 1.16 (0.98 – 1.38) | Not reported[1] | 0.63 |

| | Studies | RR (95%CI) | $I^2$ (%) | $P_{heterogeneity}$ between subgroups |
|---|---|---|---|---|
| Fruit intake>1 serving/day | 2 | 1.24 (0.99 – 1.56) | Not reported[1] | |
| **Proximal colon cancer mortality, main analysis** | 2 | 1.38 (1.11 - 1.71) | 0 | |
| **Subgroups** | | | | |
| Age at diagnosis <60 years | 2 | 1.98 (1.32 - 2.97) | Not reported[1] | 0.18 |
| Age at diagnosis 60-69 years | 2 | 1.26 (0.85 - 1.86) | Not reported[1] | |
| Age at diagnosis ≥70 years | 2 | 1.34 (0.96 - 1.87) | Not reported[1] | |
| Females | 2 | 1.47 (1.07 - 2.02) | Not reported[1] | 0.57 |
| Males | 2 | 1.30 (0.97 - 1.75) | Not reported[1] | |
| BMI<25 kgm$^2$ | 2 | 1.42 (1.06 - 1.89) | Not reported[1] | 0.93 |
| BMI≥25 kg/m$^2$ | 2 | 1.39 (1.03 - 1.87) | Not reported[1] | |
| Never smokers | 2 | 1.53 (1.11 - 2.12) | Not reported[1] | 0.44 |
| Ever smokers | 2 | 1.29 (0.97 - 1.72) | Not reported[1] | |
| Never drinkers | 2 | 1.27 (0.80 - 2.01) | Not reported[1] | 0.22 |
| Light drinkers (0.1-14.9 g/day) | 2 | 1.36 (1.03 - 1.80) | Not reported[1] | |
| Heavy drinkers (≥15 g/day) | 2 | 1.95 (1.19 - 3.21) | Not reported[1] | |
| Physical activity below median | 2 | 1.60 (1.25 - 2.06) | Not reported[1] | 0.18 |
| Physical activity above median | 2 | 1.17 (0.80 - 1.72) | Not reported[11] | |
| Fruit intake≤1 serving/day | 2 | 1.32 (1.01 – 1.73) | Not reported | 0.52 |
| Fruit intake>1 serving/day | 2 | 1.52 (1.08 – 2.14) | Not reported[1] | |
| **Distal colon cancer incidence, main analysis** | 5 | 1.03 (0.94 – 1.13) | 41% | |
| **Subgroups** | | | | |
| Baseline exposure measurement | 3 | 1.07 (1.00 – 1.14) | 0 | 0.058 |
| Repeated exposure measurements | 2 | 0.88 (0.73 – 1.06) | 0 | |
| Critical bias for departure from intended exposure | 3 | 1.07 (1.00 – 1.14) | 0 | 0.058 |
| Moderate bias for departure from intended exposure | 2 | 0.88 (0.73 – 1.06) | 0 | |
| **Rectal cancer incidence, main analysis** | 4 | 0.98 (0.91 – 1.04) | 0 | |
| **Subgroups** | | | | |
| Baseline exposure measurement | 2 | 0.99 (0.92 – 1.06) | 0 | 0.358 |
| Repeated exposure measurements | 2 | 0.89 (0.72 – 1.10) | 0 | |
| Critical bias for departure from intended exposure | 2 | 0.99 (0.92 – 1.06) | 0 | 0.358 |

| | Studies | RR (95%CI) | $I^2$ (%) | $P_{heterogeneity}$ between subgroups |
|---|---|---|---|---|
| Moderate bias for departure from intended exposure | 2 | 0.89 (0.72 – 1.10) | 0 | |
| **Colon cancer incidence, main analysis (carbonated sugar sweetened beverages)** | 10 | 1.00 (0.91-1.10) | 0 | |
| **Subgroups** | | | | |
| Follow-up ≤10 years | 4 | 0.98 (0.85 - 1.12) | 0 | 0.495 |
| Follow-up >10 years | 6 | 0.99 (0.83 - 1.18) | 30% | |
| North America | 7 | 0.98 (0.87 - 1.11) | 27% | 0.727 |
| Europe | 3 | 1.04 (0.76 - 1.44) | 0 | |
| Females | 9 | 0.96 (0.85 - 1.09) | Not reported[1] | 0.37 |
| Males | 4 | 1.05 (0.91 - 1.21) | Not reported[1] | |
| Proximal colon | 10 | 1.01 (0.89 - 1.14) | Not reported[1] | 0.89 |
| Distal colon | 10 | 1.01 (0.89 - 1.16) | Not reported[1] | |
| Never smokers | 10 | 0.98 (0.81 - 1.17) | Not reported[1] | 0.97 |
| Past smokers | 10 | 1.09 (0.93 - 1.28) | Not reported[1] | |
| Current smokers | 10 | 1.02 (0.82 - 1.27) | Not reported[1] | |
| Non-drinkers | 10 | 1.07 (0.93 - 1.21) | Not reported[1] | 0.87 |
| Light drinkers (<15 g/day) | 10 | 0.94 (0.81 - 1.09) | Not reported[1] | |
| Heavy drinkers (≥15 g/day) | 10 | 1.13 (0.90 - 1.44) | Not reported[1] | |
| Low physical activity | 10 | 0.94 (0.83 - 1.09) | Not reported[1] | 0.09 |
| Moderate physical activity | 10 | 0.94 (0.76 - 1.18) | Not reported[1] | |
| High physical activity | 10 | 1.20 (0.99 - 1.45) | Not reported[1] | |
| BMI<25 kg/m$^2$ | 10 | 0.94 (0.78 - 1.13) | Not reported[1] | 0.40 |
| BMI≥25 kg/m$^2$ | 10 | 1.03 (0.91 - 1.16) | Not reported[1] | |
| Postmenopausal hormone use: ever | 10 | 1.10 (0.94 - 1.29) | Not reported[1] | 0.13 |
| Postmenopausal hormone use: never | 10 | 0.73 (0.50 - 1.08) | Not reported[1] | |
| Baseline exposure measurement | 8 | 0.93 (0.82 - 1.06) | 0 | 0.105 |
| Repeated exposure measurements | 2 | 1.09 (0.95 - 1.25) | 0 | |
| **Liver cancer, main analysis** | 5 | 1.04 (0.97 - 1.11) | 20% | |
| **Subgroups** | | | | |
| Incidence | 4 | 1.08 (0.94 - 1.24) | 31% | 0.285 |
| Mortality | 1 | 1.02 (0.98 - 1.06) | - | |
| Adjusted for BMI | 2 | 1.21 (0.80 - 1.85) | 83% | 0.941 |

| | Studies | RR (95%CI) | $I^2$ (%) | $P_{heterogeneity}$ between subgroups |
|---|---|---|---|---|
| Not adjusted for BMI | 2 | 1.19 (0.82 - 1.71) | 79% | |
| Excluded people with diabetes at baseline | 2 | 1.29 (0.82 – 2.03) | 83% | 0.791 |
| Not excluded people with diabetes at baseline | 2 | 1.19 (0.79 – 1.79) | 75% | |
| **Pancreatic cancer, main analysis** | 5 | 1.04 (0.96 - 1.13) | 44% | |
| **Subgroups** | | | | |
| Incidence | 4 | 1.08 (0.95 - 1.22) | 45% | 0.263 |
| Mortality | 1 | 1.00 (0.98 - 1.03) | 0 | |
| BMI<30 kg/m$^2$ | 2 | 1.02 (0.98 - 1.05) | 0 | 0.184 |
| BMI≥30 kg/m$^2$ | 2 | 0.98 (0.94 - 1.02) | 0 | |
| Females | 2 | 0.99 (0.95 - 1.04) | 0.3% | 0.505 |
| Males | 2 | 1.01 (0.98 - 1.04) | 0 | |
| Never smokers | 2 | 1.05 (0.82 - 1.34) | 0 | 0.831 |
| Former smokers | 1 | 1.19 (0.85 - 1.67) | - | |
| Current smokers | 2 | 1.07 (0.86 - 1.34) | 0 | |
| Adjusted for BMI | 2 | 1.00 (0.98 – 1.02) | 0 | 0.980 |
| Not adjusted for BMI | 2 | 1.00 (0.98 – 1.03) | 0 | |
| Excluded people with diabetes at baseline | 1 | 0.94 (0.80 – 1.10) | 0 | 0.064 |
| Not excluded people with diabetes at baseline | 3 | 1.13 (1.01 – 1.26) | 0 | |
| **Pancreatic cancer, main analysis (carbonated sugar sweetened beverages)** | 15 | 1.13 (1.03 - 1.24) | 0 | |
| **Subgroups** | | | | |
| Age at diagnosis <69 years | 14 | 1.08 (0.92 - 1.26) | Not reported[1] | 0.28 |
| Age at diagnosis ≥69 years | 14 | 1.21 (1.08 - 1.37) | Not reported[1] | |
| Females | 11 | 1.03 (0.86 - 1.28) | Not reported[1] | 0.38 |
| Males | 8 | 1.17 (1.06 - 1.30) | Not reported[1] | |
| BMI<25 kg/m$^2$ | 14 | 1.26 (1.06 - 1.50) | Not reported[1] | 0.21 |
| BMI≥25 kg/m$^2$ | 14 | 1.17 (1.06 - 1.30) | Not reported[1] | |
| Never smokers | 14 | 1.17 (0.96 - 1.42) | Not reported[1] | 0.73 |
| Former smokers | 14 | 1.21 (1.02 - 1.42) | Not reported[1] | |
| Current smokers | 14 | 1.13 (0.96 - 1.30) | Not reported[1] | |
| Low physical activity | 14 | 1.13 (0.98 - 1.30) | Not reported[1] | 0.98 |
| Moderate physical activity | 14 | 1.28 (1.08 - 1.50) | Not reported[1] | |

| | Studies | RR (95%CI) | $I^2$ (%) | $P_{heterogeneity}$ between subgroups |
|---|---|---|---|---|
| High physical activity | 14 | 1.19 (1.00 - 1.42) | Not reported[1] | |
| Non-drinkers | 14 | 1.30 (1.10 - 1.52) | Not reported[1] | 0.23 |
| Light drinkers (<15 g/day) | 14 | 1.13 (0.98 - 1.28) | Not reported[1] | |
| Heavy drinkers (≥15 g/day) | 14 | 1.06 (0.86 - 1.33) | Not reported[1] | |
| Follow-up ≤5 years | 14 | 1.10 (0.96 - 1.28) | Not reported[1] | 0.56 |
| Follow-up >5 years | 14 | 1.17 (1.04 - 1.33) | Not reported[1] | |
| **Breast cancer incidence, main analysis** | 8 | 1.07 (0.98 - 1.17) | 57% | |
| **Subgroups** | | | | |
| North America | 5 | 1.04 (0.97 - 1.10) | 42% | 0.021 |
| Europe | 2 | 2.07 (1.27 - 3.38) | 0 | |
| East/Southeast Asia | 1 | 0.90 (0.48 - 1.68) | - | |
| Follow-up ≤10 years | 4 | 1.25 (0.77 - 2.05) | 60% | 0.463 |
| Follow-up >10 years | 4 | 1.04 (0.97 - 1.12) | 56% | |
| With fruit juice | 3 | 1.26 (0.91 - 1.73) | 64% | 0.507 |
| Without fruit juice | 3 | 1.49 (1.02 - 2.18) | 0 | |
| Baseline exposure measurement | 5 | 1.16 (0.98 – 1.38) | 10% | 0.504 |
| Repeated exposure measurements | 4 | 1.08 (0.97 – 1.21) | 78% | |
| Critical bias for departure from intended exposure | 5 | 1.16 (0.98 – 1.38) | 10% | 0.504 |
| Moderate bias for departure from intended exposure | 4 | 1.08 (0.97 – 1.21) | 78% | |
| **Postmenopausal breast cancer incidence, main analysis** | 6 | 1.05 (1.00 – 1.10) | 0 | |
| **Subgroups** | | | | |
| Baseline exposure measurement | 4 | 1.04 (0.99 – 1.09) | 0 | 0.112 |
| Repeated exposure measurements | 2 | 1.22 (1.01 – 1.47) | 0 | |
| Excluded people with diabetes at baseline | 2 | 1.22 (1.01 – 1.47) | 0 | 0.112 |
| Not excluded people with diabetes at baseline | 4 | 1.04 (0.99 – 1.09) | 0 | |
| Moderate bias for confounding | 5 | 1.05 (1.00 – 1.10) | 0 | 0.553 |
| Critical bias for confounding | 1 | 1.31 (0.63 – 2.74) | - | |
| Critical bias for departure from intended exposure | 4 | 1.04 (0.99 – 1.09) | 0 | 0.112 |
| Moderate bias for departure from intended exposure | 2 | 1.22 (1.01 – 1.47) | 0 | |

| | Studies | RR (95%CI) | $I^2$ (%) | $P_{heterogeneity}$ between subgroups |
|---|---|---|---|---|
| **Premenopausal breast cancer incidence, main analysis** | 5 | 1.25 (0.95 – 1.68) | 64% | |
| **Subgroups** | | | | |
| Excluded people with diabetes at baseline | 1 | 1.16 (0.85 – 1.59) | - | 0.474 |
| Not excluded people with diabetes at baseline | 4 | 1.45 (0.86 – 2.46) | 74% | |
| Baseline exposure measurement | 1 | 1.16 (0.85 – 1.59) | - | 0.474 |
| Repeated exposure measurements | 4 | 1.45 (0.86 – 2.46) | 74% | |
| **Breast cancer mortality, main analysis** | 3 | 1.05 (0.97 – 1.14) | 0 | |
| **Subgroups** | | | | |
| US | 2 | 1.04 (0.95 – 1.12) | 72% | 0.268 |
| Europe | 1 | 1.21 (0.93 – 1.57) | 0 | |
| Baseline exposure measurement | 2 | 1.05 (0.98 – 1.23) | 50% | 0.686 |
| Repeated exposure measurements | 1 | 1.09 (1.00 – 1.18) | - | |
| Excluded people with diabetes at baseline | 2 | 1.05 (0.98 – 1.23) | 50% | 0.686 |
| Not excluded people with diabetes at baseline | 1 | 1.09 (1.00 – 1.18) | - | |
| Critical bias for departure from intended exposure | 2 | 1.05 (0.98 – 1.23) | 50% | 0.686 |
| Moderate bias for departure from intended exposure | 1 | 1.09 (1.00 – 1.18) | - | |
| **Prostate cancer incidence, main analysis** | 4 | 1.17 (0.96 – 1.43) | 0 | |
| **Subgroups** | | | | |
| Baseline exposure measurement | 2 | 1.18 (0.90 – 1.56) | 0.44 | 0.60 |
| Repeated exposure measurements | 1 | 1.45 (0.71 – 2.96) | - | |
| **Prostate cancer mortality, main analysis** | 3 | 0.99 (0.97 – 1.01) | 0 | |
| **Subgroups** | | | | |
| US | 2 | 0.99 (0.97 – 1.01) | 0 | 0.465 |
| Europe | 1 | 1.11 (0.81 – 1.52) | - | |
| Baseline exposure measurement | 2 | 0.99 (0.97 – 1.01) | 0 | 0.815 |
| Repeated exposure measurements | 1 | 1.01 (0.86 – 1.19) | - | |
| Excluded people with diabetes at baseline | 2 | 0.99 (0.97 – 1.01) | 0 | |
| Not excluded people with diabetes at baseline | 1 | 1.01 (0.86 – 1.19) | - | |
| Critical bias for departure from intended exposure | 2 | 0.99 (0.97 – 1.01) | 0 | |

| | Studies | RR (95%CI) | $I^2$ (%) | $P_{heterogeneity}$ between subgroups |
|---|---|---|---|---|
| Moderate bias for departure from intended exposure | 1 | 1.01 (0.86 – 1.19) | - | |
| **Endometrial cancer incidence, main analysis** | 3 | 1.21 (1.03 - 1.42) | 0 | |
| **Subgroups** | | | | |
| Adjusted for BMI | 2 | 1.30 (1.00 - 1.71) | 28% | 0.692 |
| Not adjusted for BMI | 2 | 1.23 (1.03 - 1.45) | 0 | |
| **Kidney cancer incidence, main analysis** | 4 | 1.13 (0.95 - 1.35) | 49% | |
| **Subgroups** | | | | |
| Females | 2 | 1.19 (0.92 - 1.54) | 0 | 0.231 |
| Males | 2 | 0.99 (0.83 - 1.17) | 0 | |
| Excluded people with diabetes at baseline | 2 | 1.07 (0.88 – 1.30) | 0 | 0.451 |
| Not excluded people with diabetes at baseline | 2 | 1.21 (0.95 – 1.53) | 59% | |
| **Kidney cancer mortality, main analysis** | | | | |
| **Subgroups** | | | | |
| Females | 2 | 1.09 (1.03 - 1.17) | 0 | 0.427 |
| Males | 2 | 1.01 (0.85 - 1.21) | 26% | |
| **Non-Hodgkin lymphoma incidence, main analysis** | 3 | 1.04 (0.87 – 1.23) | 62% | |
| **Subgroups** | | | | |
| Baseline exposure measurement | 1 | 1.00 (0.98 – 1.03) | - | 0.742 |
| Repeated exposure measurements | 2 | 1.07 (0.71 – 1.60) | 78% | |
| Critical bias for departure from intended exposure | 1 | 1.00 (0.98 – 1.03) | - | 0.742 |
| Moderate bias for departure from intended exposure | 2 | 1.07 (0.71 – 1.60) | 78% | |

Abbreviations: RR, relative risk; CI: confidence interval; BMI, body mass index.

[1] Based on a pooled analysis.

**Supplementary Table 20. Subgroup analyses of artificially sweetened beverages (per 1 serving [355 mL]/day) and cancer risk.**

| Subgroup | Studies | RR (95%CI) | $I^2$ (%) | $P_{heterogeneity}$ between subgroups |
|---|---|---|---|---|
| **Colorectal cancer incidence, main analysis** | | | | |
| **Subgroups** | | | | |
| US | 3 | 1.02 (0.97 – 1.08) | 0 | 0.378 |
| Australia | 1 | 0.91 (0.71 – 1.17) | - | |
| Baseline exposure measurement | 1 | 0.91 (0.71 – 1.17) | - | 0.378 |
| Repeated exposure measurements | 3 | 1.02 (0.97 – 1.08) | 0 | |
| Critical bias for departure from intended exposure | 1 | 0.91 (0.71 – 1.17) | - | 0.378 |
| Moderate bias for departure from intended exposure | 3 | 1.02 (0.97 – 1.08) | 0 | |
| **Pancreatic cancer, main analysis** | 3 | 1.05 (1.00 - 1.10) | 23% | |
| **Subgroups** | | | | |
| Females | 2 | 1.11 (0.90 – 1.36) | 76% | 0.360 |
| Males | 2 | 0.90 (0.60 – 1.34) | 68% | |
| Never smokers | 2 | 1.10 (0.97 - 1.24) | 0 | 0.368 |
| Current smokers | 2 | 1.00 (0.86 - 1.17) | 0 | |
| BMI<25 kg/m$^2$ | 2 | 1.00 (0.97 - 1.03) | 0 | 0.159 |
| BMI≥25 kg/m$^2$ | 2 | 1.04 (1.01 - 1.07) | 0 | |
| BMI≥30 kg/m$^2$ | 2 | 1.01 (0.95 - 1.07) | 0 | |
| **Pancreatic cancer, main analysis (carbonated artificially sweetened beverages)** | 2 | 1.02 (0.64 - 1.61) | 0 | |
| **Subgroups** | | | | |
| BMI<25 kg/m$^2$ | 2 | 0.98 (0.48 - 1.98) | 0 | 0.999 |
| BMI≥25 kg/m$^2$ | 2 | 0.98 (0.48 - 1.99) | 23% | |
| **Breast cancer incidence, main analysis** | 3 | 0.99 (0.97 – 1.01) | 0 | |
| **Subgroups** | | | | |
| US | 2 | 0.91 (0.71 – 1.17) | - | 0.378 |
| Europe | 1 | 1.02 (0.97 – 1.08) | 0 | |
| Baseline exposure measurement | 1 | 0.91 (0.71 – 1.17) | - | 0.378 |
| Repeated exposure measurements | 2 | 1.02 (0.97 – 1.08) | 0 | |

| Subgroup | Studies | RR (95%CI) | $I^2$ (%) | $P_{heterogeneity}$ between subgroups |
|---|---|---|---|---|
| **Breast cancer mortality, main analysis** | 3 | 1.01 (0.97 – 1.04) | 11% | |
| **Subgroups** | | | | |
| US | 2 | 1.01 (0.98 – 1.04) | 0 | 0.133 |
| Europe | 1 | 0.79 (0.57 – 1.09) | - | |
| Baseline exposure measurement | 2 | 0.94 (0.76 – 1.17) | 56% | 0.548 |
| Repeated exposure measurements | 1 | 1.01 (0.95 – 1.08) | - | |
| Critical bias for departure from intended exposure | 2 | 0.94 (0.76 – 1.17) | 56% | 0.548 |
| Moderate bias for departure from intended exposure | 1 | 1.01 (0.95 – 1.08) | - | |
| **Endometrial cancer incidence, main analysis** | 3 | 0.90 (0.64 – 1.26) | 0 | |
| **Subgroups** | | | | |
| Excluded people with diabetes at baseline | 2 | 0.88 (0.61 – 1.27) | 0 | 0.793 |
| Not excluded people with diabetes at baseline | 1 | 1.00 (0.43 – 2.35) | - | |
| **Postmenopausal breast cancer incidence, main analysis** | 4 | 0.98 (0.95 – 1.01) | 0 | |
| **Subgroups** | | | | |
| Baseline exposure measurement | 2 | 1.00 (0.84 – 1.18) | 0 | 0.826 |
| Repeated exposure measurements | 2 | 0.98 (0.95 – 1.01) | - | |
| **Prostate cancer mortality, main analysis** | 3 | 0.99 (0.96 – 1.02) | 0 | |
| **Subgroups** | | | | |
| **US** | 2 | 0.99 (0.96 – 1.02) | 0 | 0.674 |
| **Europe** | 1 | 1.11 (0.66 – 1.88) | - | |
| Baseline exposure measurement | 2 | 0.99 (0.96 – 1.02) | 0 | 0.621 |
| Repeated exposure measurements | 1 | 1.02 (0.91 – 1.14) | - | |
| Excluded people with diabetes at baseline | 2 | 0.99 (0.96 – 1.02) | 0 | 0.621 |
| Not excluded people with diabetes at baseline | 1 | 1.02 (0.91 – 1.14) | - | |
| Critical bias for departure from intended exposure | 2 | 0.99 (0.96 – 1.02) | 0 | 0.621 |
| Moderate bias for departure from intended exposure | 1 | 1.02 (0.91 – 1.14) | - | |
| **Kidney cancer mortality, main analysis** | 2 | 0.97 (0.82 - 1.15) | 64% | |
| **Subgroups** | | | | |

| Subgroup | Studies | RR (95%CI) | $I^2$ (%) | $P_{heterogeneity}$ between subgroups |
|---|---|---|---|---|
| Females | 2 | 0.96 (0.91 - 1.03) | 0 | 0.362 |
| Males | 2 | 1.08 (0.85 - 1.39) | 58% | |

Abbreviations: RR, relative risk; CI: confidence interval; BMI, body mass index.

**Supplementary Table 21. Subgroup analyses of fruit juice (per 1 serving [177 mL]/day) and cancer risk.**

| Subgroup | Studies | RR (95%CI) | $I^2$ (%) | $P_{heterogeneity}$ between subgroups |
|---|---|---|---|---|
| **Breast cancer incidence, main analysis (fruit juice)** | 6 | 1.02 (0.99 - 1.05) | | |
| **Subgroups** | | | | |
| US | 3 | 1.02 (0.99 - 1.04) | 0 | 0.083 |
| Europe | 3 | 1.17 (1.00 - 1.38) | 0 | |
| Baseline exposure measurement | 3 | 1.14 (0.97 – 1.33) | 0 | 0.176 |
| Repeated exposure measurements | 3 | 1.02 (0.99 – 1.05) | 0 | |
| Moderate bias for departure from intended exposure | 3 | 1.14 (0.97 – 1.33) | 0 | 0.176 |
| Critical bias for departure from intended exposure | 3 | 1.02 (0.99 – 1.05) | 0 | |
| **Melanoma incidence, main analysis (citrus juice)** | 4 | 1.08 (0.92 - 1.27) | 63% | |
| **Subgroups** | | | | |
| US | 2 | 1.36 (0.77 - 2.39) | 65% | 0.350 |
| Europe | 2 | 1.01 (0.77 - 1.32) | 79% | |
| **Glioma incidence, main analysis (fruit juice)** | 3 | 0.99 (0.94 - 1.05) | 0 | |
| **Subgroups** | | | | |
| Females | 3 | 1.03 (0.92 - 1.15) | 0 | 0.841 |
| Males | 3 | 1.01 (0.88 - 1.16) | 45% | |
| Follow-up <5 years | 3 | 0.93 (0.83 - 1.04) | Not reported | Not reported |
| Follow-up ≥5 years | 3 | 1.04 (0.96 - 1.11) | Not reported | |
| **Kidney cancer incidence, main analysis** | 4 | 1.02 (0.87 - 1.20) | 44% | |
| **Subgroups** | | | | |
| US | 2 | 1.06 (0.88 – 1.28) | 0 | 0.599 |
| Europe | 2 | 1.39 (0.51 – 3.76) | 66% | |
| **Kidney cancer incidence, main analysis (fruit and vegetable juice)** | 14 | 1.04 (0.97 - 1.12) | 0 | |
| **Subgroups** | | | | |
| Females | 10 | 1.04 (0.95 - 1.13) | 0 | 0.672 |

| | | | |
|---|---|---|---|
| Males | 7 | 1.07 (0.96 - 1.19) | 0 |

Abbreviations: RR, relative risk; CI: confidence interval.

**Supplementary Table 22. Subgroup analyses of soft drinks (per 1 serving [355 mL]/day) and cancer risk.**

| Subgroup | Studies | RR (95%CI) | $I^2$ (%) | $P_{heterogeneity}$ between subgroups |
|---|---|---|---|---|
| **Pancreatic cancer, main analysis** | 5 | 1.06 (0.97 - 1.17) | 0 | |
| **Subgroups** | | | | |
| US | 2 | 1.05 (0.95 - 1.15) | 68% | 0.560 |
| Europe | 3 | 1.17 (0.83 - 1.64) | 79% | |
| Follow-up ≤10 years | 4 | 1.10 (0.97 - 1.25) | 75% | 0.309 |
| Follow-up >10 years | 1 | 1.00 (0.88 - 1.14) | - | |
| **Liver cancer, main analysis** | 4 | 1.15 (0.90 - 1.47) | 85% | |
| **Subgroups** | | | | |
| US | 3 | 1.02 (0.95 - 1.09) | 0 | <0.001 |
| Europe[1] | 1 | 1.44 (1.16 - 1.66) | - | |
| **Kidney cancer incidence, main analysis** | 14 | 1.02 (0.99 - 1.06) | 0 | |
| **Subgroups** | | | | |
| Females | 10 | 1.02 (0.97 - 1.07) | 0 | 0.800 |
| Males | 7 | 1.03 (0.97 - 1.09) | Not reported[2] | |

Abbreviations: RR, relative risk; CI: confidence interval.

[1] One large, multi-centre cohort study in several European countries.

[2] Based on a pooled analysis.

**Supplementary Table 23. Descriptive table of the included Mendelian randomisation studies of soft drinks and cancer incidence and mortality.**

| Author, Year, PMID | MR design | Exposure population source (ref of GWAS) | Exposure population ancestry | No. IVs (SNPs) | Cancer population source (ref of GWAS) | Cancer population ancestry | Cancer cases/ controls | Outcome, comparison | Main results OR (95% CI) | Sensitivity analyses |
|---|---|---|---|---|---|---|---|---|---|---|
| Jin, 2024, 38524848[192] | Two-sample; summary-level data | 24-h recall questionnaire<br>Sugar-sweetened beverages<br>UK Biobank GWAS<br>PMID:31046077 | European<br>85,852 males and females | 12 SNPs | FinnGen, males and females<br>https://www.finngen.fi/en/access_results | European | | Oesophageal | Inverse variance weighted (IVW) method<br>0.424 (0.004-48.734) | MR-Egger, weighted mode, weighted median |
| | | | | | FinnGen, males and females<br>https://www.finngen.fi/en/access_results | European | | Stomach | 5.309 (0.234-120.270) | |
| | | | | | GECCO, CORECT, CCFR, males and females<br>PMID:30510241 | European | | Colorectal | 2.488 (0.593-10.435) | |
| | | | | | UK Biobank, males and females<br>https://www.nealelab.is/uk-biobank | European | | Liver | 0.037 (0.000-1669.357) | |
| | | | | | Kaiser Permanente GERA; UK Biobank, males and females<br>PMID:32887889 | European | | Pancreatic | 0.327 (0.002-45.783) | |
| | | | | | BCAC, females<br>PMID:25751625 | European | | Breast | 1.888 (1.115-3.196) | |
| | | | | | PMID:29059683 | European | | ER+ breast | 1.566 (0.837-2.929) | |
| | | | | | PMID:29059683 | European | | ER- breast | 2.843 (1.090-7.414) | |

| Author, Year, PMID | MR design | Exposure population source (ref of GWAS) | Exposure population ancestry | No. IVs (SNPs) | Cancer population source (ref of GWAS) | Cancer population ancestry | Cancer cases/ controls | Outcome, comparison | Main results OR (95% CI) | Sensitivity analyses |
|---|---|---|---|---|---|---|---|---|---|---|
| | | | | | ECAC; E2C2; UK Biobank, females<br>PMID: 30093612 | European | | Endometrial | 0.902 (0.253-3.213) | |
| | | | | | OCAC, females<br>PMID:28346442 | European | | Ovarian | 0.835 (0.230-3.035) | |
| | | | | | PRACTICAL, males<br>PMID: 29892016 | European | | Prostate | 1.538 (0.784-3.019) | |
| | | | | | Kaiser Permanente GERA; UK Biobank, males and females<br>PMID:32887889 | European | | Kidney | 7.820 (0.235-260.596) | |
| | | | | | FinnGen, males and females<br>https://www.finngen.fi/en/access_results | European | | Thyroid | 0.350 (0.018-6.934) | |
| Jin, 2024, 38524848[192] | Two-sample; summary-level data | 24-h recall questionnaire<br>Artificially sweetened beverages<br>UK Biobank GWAS<br>PMID:31046077 | European<br>85,852 males and females | 16 SNPs<br>Removed SNPs associated with type 2 diabetes, physical activity, smoking, education, other sources of sugar | FinnGen, males and females<br>https://www.finngen.fi/en/access_results | European | 566/287,137 | Oesophageal | Inverse variance weighted (IVW) method<br>0.343 (0.003-36.130) | MR-Egger, weighted mode, weighted median, and lasso method |
| | | | | | FinnGen, males and females<br>https://www.finngen.fi/en/access_results | European | 1,307/287,137 | Stomach | 3.855 (0.050-294.354) | |
| | | | | | GECCO, CORECT, CCFR, males and females | European | 11,835/11,856 | Colorectal | 6.879 (1.551-30.512) | Yes, robust |

| Author, Year, PMID | MR design | Exposure population source (ref of GWAS) | Exposure population ancestry | No. IVs (SNPs) | Cancer population source (ref of GWAS) | Cancer population ancestry | Cancer cases/ controls | Outcome, comparison | Main results OR (95% CI) | Sensitivity analyses |
|---|---|---|---|---|---|---|---|---|---|---|
| | | | | | PMID:30510241 | | | | | |
| | | | | | UK Biobank, males and females<br>https://www.nealelab.is/uk-biobank | European | 165/456,111 | Liver | 18.180 (0.002-167795.485) | |
| | | | | | Kaiser Permanente GERA; UK Biobank, males and females<br>PMID:32887889 | European | 663/410,350 | Pancreatic | 2.895 (0.029-285.157) | |
| | | | | | BCAC, females<br>PMID:25751625 | European | 46,785/42,892 | Breast | 3.881 (2.023-9.776) | Yes, robust |
| | | | | | PMID:29059683 | European | 69,501/105,974 | ER+ breast | 0.927 (0.469-1.835) | |
| | | | | | PMID:29059683 | European | 21,468/105,974 | ER- breast | 1.847 (0.535-6.376) | |
| | | | | | ECAC; E2C2; UK Biobank, females<br>PMID: 30093612 | European | 12,906/108,979 | Endometrial | 0.953 (0.218-324.338) | |
| | | | | | OCAC, females<br>PMID:28346442 | European | 25,509/40,941 | Ovarian | 1.614 (0.332-7.844) | |
| | | | | | PRACTICAL, males<br>PMID: 29892016 | European | 79,148/61,106 | Prostate | 0.780 (0.380-1.602) | |
| | | | | | Kaiser Permanente GERA; UK Biobank, males and females<br>PMID:32887889 | European | 1,338/410,350 | Kidney | 6.438 (0.128-324.338) | |
| | | | | | FinnGen, males and females<br>https://www.finngen.fi/en/access_results | European | 1,783/287,137 | Thyroid | 1.178 (0.087-16.042) | |

| Author, Year, PMID | MR design | Exposure population source (ref of GWAS) | Exposure population ancestry | No. IVs (SNPs) | Cancer population source (ref of GWAS) | Cancer population ancestry | Cancer cases/ controls | Outcome, comparison | Main results OR (95% CI) | Sensitivity analyses |
|---|---|---|---|---|---|---|---|---|---|---|
| Pan, 2024, 38576413[193] | Two-sample; summary-level data | Artificial sweeteners added to coffee<br>IEU Open GWAS project (https://gwas.mrcieu.ac.uk<br>ukb-b-1338 | European<br>42,348 | 12 | IEU Open GWAS project (https://gwas.mrcieu.ac.uk<br>OCAC ieu-a-1121 | European | 13,037 /40,941 | High grade serous ovarian | Inverse variance weighted method<br>3.012 (1.602-5.633) | MR-Egger, weighted median, simple mode, weighted mode, leave-one-out |
| | | | | | OCAC ieu-a-1228 | | 14,049 /40,941 | High/low grade serous ovarian | 2.340 (1.264-4.332) | Yes, robust |
| | | | | | 67 cancer outcomes of different grades and histology<br>IEU Open GWAS studies included BCAC, FINNGEN, ILCCO, MRC-IEU, Neale lab, OCAC, PRACTICAL, TRICL, UK Biobank | | | Head and neck, laryngeal, oral cavity, oesophageal, stomach, colorectal, liver, lung, breast, endometrial, prostate, kidney | ORs and 95% CIs not tabulated.<br>P-values 0.077 to 0.978 except for ovarian and kidney cancers. | Leave-one-out test showed influence from individual SNPs on ovarian and kidney cancer |
| | | Artificial sweeteners added to tea<br>ukb-b-5867 | | 15 | Oncoarray oral cavity and oropharyngeal cancer<br>ieu-b-91 | | 613/ 727 | Oral cavity and pharyngeal | 214.060 (10.560-4339.228) | Yes, robust |
| | | | | | ieu-b-93 | | 1135/ 2329 | Oral cavity | 17.973 (2.559-126.225) | Yes, robust |
| | | | | | ieu-b-95 | | 340/ 727 | Oral cavity | 246.792 (6.920-8801.209) | Yes, robust |
| | | | | | ieu-b-98 | | 205/ 727 | Oropharyngeal | 1992.183 (23.881-166187.626) | Yes, robust |
| | | | | | ukb-d-C3_BRONCHUS_LUNG | | 1,681/ 359,513 | Bronchus and lungs | 0.992 (0.987-0.997) | Yes, robust |

| Author, Year, PMID | MR design | Exposure population source (ref of GWAS) | Exposure population ancestry | No. IVs (SNPs) | Cancer population source (ref of GWAS) | Cancer population ancestry | Cancer cases/ controls | Outcome, comparison | Main results OR (95% CI) | Sensitivity analyses |
|---|---|---|---|---|---|---|---|---|---|---|
| | | | | | 71 cancer outcomes of different grades and histology<br>IEU Open GWAS included BCAC, FINNGEN, ILCCO, MRC-IEU, Neale lab, OCAC, PRACTICAL, TRICL, UK Biobank | | | Head and neck, oral and oropharyngeal, oesophageal, stomach, colorectal, liver, lung, breast, endometrial, ovarian, prostate, kidney | ORs and 95% CIs not tabulated.<br>P-values 0.057 to 0.992 except for lung and oral cavity cancers. | Leave-one-out test showed influence from individual SNPs on lung and oral cavity cancers |
| Liu, 2023, 36040623[191] | Two-sample; summary-level data | 24-h recall questionnaire<br>PMID: 31046077<br>Sweet beverages (sugar-sweetened beverages, artificially sweetened beverages, pure non-grapefruit juices, flavoured milk, hot chocolate) | European<br>UK Biobank | Excluded IVs correlated with other phenotypes such as BMI.<br>21 | 85,852 | European<br>UK Biobank and FinnGen GWAS<br>www.gwas.mrcieu.ac.uk | 4029 | Colon | Inverse variance weighted method<br>1.002 (0.992-1.012) | MR-Egger, weighted median, leave-one-out, MR Steiger |
| | | | | | | | 2548 | Rectal | 0.997 (0.987-1.007) | |
| | | Sugar-sweetened beverages | | 23 | | | 4029 | Colon | 1.013 (1.001-1.026) | Yes, robust. Non-significant results but consistent association in the positive direction in the weighted median MR analysis. |
| | | | | | | | 2548 | Rectal | 0.994 (0.973-1.015) | |

Abbreviations: BMI, body mass index; GWAS, genome wide association studies; IVW, inverse variance weighted; MR, Mendelian randomisation; OR, odds ratio.

**Supplementary Figure 1. Summary forest plot of the linear dose-response associations of soft drinks (sugar sweetened and artificially sweetened beverages combined) and cancer incidence.**

#### Soft drinks, per 1 serving/day and cancer risk

| Outcome | Cohorts | Cases | RR (95% CI) | I² (%) |
|---|---|---|---|---|
| **Soft drinks** | | | | |
| Liver | 4 | 1136 | 1.15 (0.90 - 1.47) | 84 |
| Bladder | 2 | 765 | 1.09 (0.88 - 1.35) | 73 |
| Pancreatic | 5 | 2641 | 1.06 (0.97 - 1.17) | 63 |
| Breast | 2 | 12,604 | 1.00 (0.97 - 1.04) | 0 |
| Extrahepatic biliary tract | 3 | 222 | 0.99 (0.58 - 1.67) | 81 |
| Lung | 2 | 1299 | 0.98 (0.91 - 1.06) | 0 |
| Intrahepatic biliary tract | 3 | 87 | 0.95 (0.60 - 1.50) | 0 |
| Endometrial | 2 | 1389 | 0.94 (0.66 - 1.34) | 0 |
| Gallbladder | 3 | 158 | 0.84 (0.25 - 2.87) | 85 |
| **Carbonated soft drinks** | | | | |
| Liver | 2 | 839 | 1.03 (0.95 - 1.11) | NE |
| Kidney | 14 | 2066 | 1.02 (0.99 - 1.06) | 0 |
| **Carbonated and decaffeinated soft drinks** | | | | |
| Glioma | 4 | 649 | 0.97 (0.87 - 1.09) | 0 |
| **Carbonated and caffeinated soft drinks** | | | | |
| Glioma | 4 | 855 | 0.94 (0.76 - 1.17) | 32 |

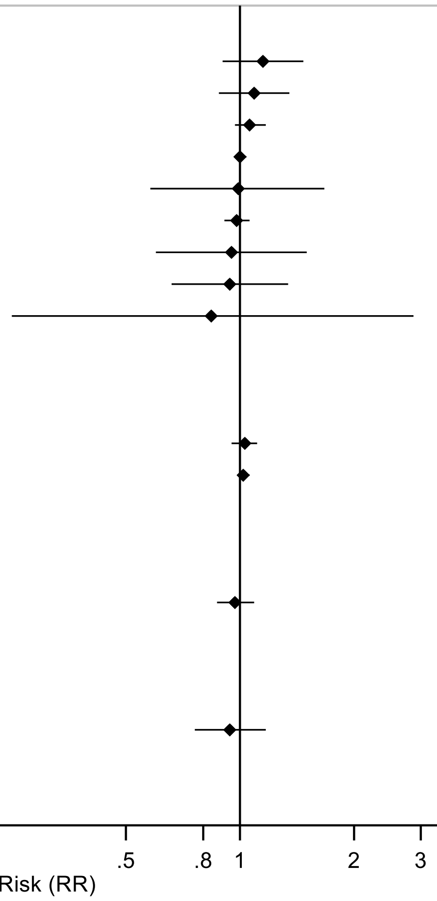

Forest plot shows the summary results across the linear dose-response (per 1 serving[355 mL]/day) inverse variance DerSimonian-Laird random-effects model meta-analyses. Each square and the horizontal line across the square represent the relative risk (RR) estimate of the corresponding meta-analysis and its 95% confidence interval (CI). No meta-analysis has been conducted for cancer mortality.

Note: Outcomes are incidence, unless otherwise stated.

* P<0.05

**Supplementary Figure 2. Summary forest plot of the linear dose-response associations of sugar sweetened beverages and cancer incidence and mortality.**

#### Sugar sweetened beverages, per 1 serving/day and cancer risk

| Outcome | Studies | Cases | RR (95% CI) | I² (%) |
|---|---|---|---|---|
| Ovary | 2 | 228 | 1.61 (1.03 - 2.53)* | 0 |
| Endometrial, type 1 | 2 | 631 | 1.45 (0.75 - 2.79) | 75 |
| Proximal colon, mortality | 2 | 409 | 1.38 (1.11 - 1.71)* | 0 |
| Breast, premenopausal | 5 | 3970 | 1.26 (0.95 - 1.68) | 64 |
| Prostate | 3 | 897 | 1.21 (0.94 - 1.57) | 0 |
| Endometrial | 3 | 931 | 1.21 (1.03 - 1.42)* | 0 |
| Stomach cardia | 2 | 290 | 1.18 (0.76 - 1.84) | 0 |
| Kidney | 4 | 1350 | 1.13 (0.95 - 1.35) | 49 |
| Colon, mortality | 2 | NR | 1.09 (0.99 - 1.19) | NE |
| Liver | 5 | 1634 | 1.08 (0.94 - 1.24) | 31 |
| Pancreas | 4 | 2770 | 1.08 (0.95 - 1.22) | 44 |
| Breast | 8 | 15,855 | 1.07 (0.98 - 1.18) | 56 |
| Colorectal | 13 | 12,649 | 1.07 (1.00 - 1.14)* | 41 |
| Kidney, mortality | 2 | 3394 | 1.06 (1.02 - 1.10)* | 62 |
| Breast, postmenopausal | 6 | 10,774 | 1.05 (1.00 - 1.10)* | 0 |
| Breast, mortality | 3 | 7476 | 1.05 (0.97 - 1.14) | 0 |
| Liver, incidence/mortality | 6 | 4356 | 1.04 (0.97 - 1.11) | 20 |
| Pancreas, incidence/mortality | 5 | 11,966 | 1.04 (0.96 - 1.13) | 44 |
| Distal colon | 5 | 3079 | 1.03 (0.94 - 1.13) | 41 |
| Proximal colon | 5 | 3929 | 1.03 (0.96 - 1.11) | 60 |
| Breast, ER+ | 2 | 2019 | 1.03 (0.98 - 1.07) | NE |
| Colorectal, mortality | 4 | 16,855 | 1.02 (1.00 - 1.04)* | 0 |
| Colon | 4 | 7051 | 1.01 (0.97 - 1.05) | 0 |
| Breast, luminal A | 2 | 765 | 1.00 (0.93 - 1.08) | NE |
| Prostate, mortality | 3 | 10,288 | 0.99 (0.97 - 1.01) | 0 |
| Breast, luminal B | 2 | 311 | 0.99 (0.88 - 1.12) | NE |
| Stomach | 2 | 3041 | 0.99 (0.80 - 1.23) | 0 |
| Rectal | 4 | 2164 | 0.98 (0.91 - 1.04) | 0 |
| Lung, mortality | 3 | 34,381 | 0.98 (0.97 - 1.00)* | 0 |
| Breast, ER- | 2 | 453 | 0.97 (0.89 - 1.06) | NE |
| Distal colon, mortality | 2 | 266 | 0.97 (0.70 - 1.34) | NE |
| Lung, incidence/mortality | 5 | 36,557 | 0.97 (0.92 - 1.02) | 56 |
| Rectal, mortality | 2 | 226 | 0.92 (0.64 - 1.31) | NE |
| Breast, HER2+ | 2 | 62 | 0.92 (0.71 - 1.19) | NE |
| Lung | 2 | 2176 | 0.89 (0.81 - 0.97)* | 0 |
| Breast, basal like | 2 | 77 | 0.84 (0.67 - 1.05) | NE |

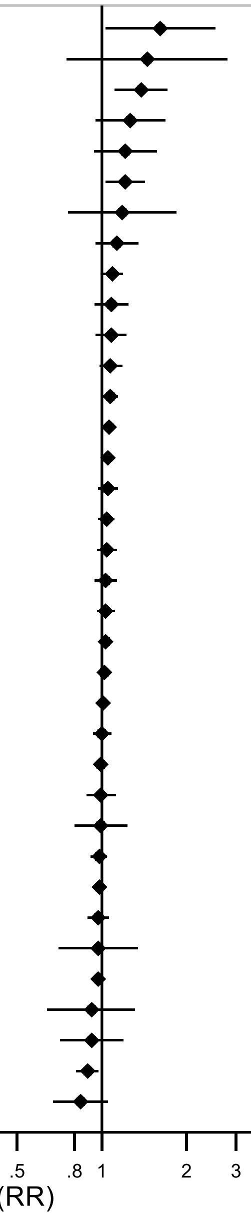

Forest plot shows the summary results across the linear dose-response (per 1 serving[355 mL]/day) inverse variance DerSimonian-Laird random-effects model meta-analyses. Each square and the horizontal line across the square represent the relative risk (RR) estimate of the corresponding meta-analysis and its 95% confidence interval (CI).

Note: outcomes are incidence, unless otherwise stated.

* P<0.05

**Supplementary Figure 3. Summary forest plot of the linear dose-response associations of sugar sweetened beverages components and cancer incidence and mortality.**

#### Sugar sweetened beverages components, per 1 serving/day and cancer risk

| Outcome | Studies | Cases | RR (95% CI) | I² (%) |
|---|---|---|---|---|
| **Carbonated sugar sweetened beverages** | | | | |
| Multiple Myeloma | 2 | 570 | 1.26 (0.84 - 1.89) | 0 |
| Adenocarcinoma of pancreas | 14 | 1556 | 1.17 (1.06 - 1.30)* | NE |
| Pancreas | 15 | 2481 | 1.13 (1.03 - 1.24)* | 0 |
| Liver | 2 | 839 | 1.11 (0.98 - 1.26) | NE |
| Leukemia | 2 | 339 | 1.09 (0.61 - 1.95) | 58 |
| Non-Hodgkin Lymphoma | 3 | 2520 | 1.04 (0.87 - 1.23) | 62 |
| Colon | 10 | 5604 | 1.00 (0.91 - 1.10) | 0 |
| Kidney | 2 | 248 | 0.95 (0.69 - 1.31) | NE |
| **Sugar sweetened cola** | | | | |
| Pancreas | 2 | 379 | 1.18 (0.53 - 2.62) | 28 |
| **Fruit drinks** | | | | |
| Liver | 3 | 1046 | 1.10 (0.72 - 1.69) | 53 |
| **Sugar sweetened fruit drinks** | | | | |
| Liver | 2 | 839 | 0.82 (0.59 - 1.14) | NE |
| **Substituting SSB with ASB** | | | | |
| Proximal colon | 2 | 1275 | 0.84 (0.77 - 0.95)* | NE |
| Proximal colon, mortality | 2 | 409 | 0.80 (0.71 - 0.95)* | NE |

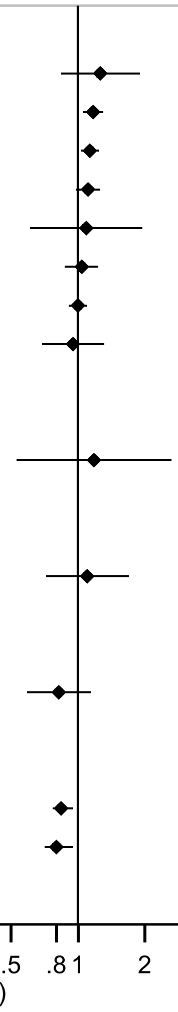

Forest plot shows the summary results across the linear dose-response (per 1 serving[355 mL]/day) inverse variance DerSimonian-Laird random-effects model meta-analyses. Each square and the horizontal line across the square represent the relative risk (RR) estimate of the corresponding meta-analysis and its 95% confidence interval (CI).

Note: Outcomes are incidence, unless otherwise stated.

* P<0.05

**Supplementary Figure 4. Summary forest plot of the linear dose-response associations of artificially sweetened beverages and cancer incidence and mortality.**

#### Artificially sweetened beverages, per 1 serving/day and cancer risk

| Outcome | Studies | Cases | RR (95% CI) | I² (%) |
|---|---|---|---|---|
| **Artificially sweetened beverages** | | | | |
| Ovary | 2 | 381 | 1.30 (0.82 - 2.05) | 0 |
| Liver | 4 | 1147 | 1.20 (0.88 - 1.64) | 86 |
| Kidney | 2 | 1389 | 1.15 (0.99 - 1.34) | 0 |
| Endometrial, postmenopausal | 3 | 900 | 1.13 (0.71 - 1.79) | 64 |
| Pancreas | 2 | 1700 | 1.12 (1.00 - 1.24) | 0 |
| Liver, incidence/mortality | 5 | 3869 | 1.10 (0.96 - 1.26) | 83 |
| Kidney, mortality | 2 | 3394 | 1.06 (0.87 - 1.30) | 64 |
| Pancreas, incidence/mortality | 3 | 10,896 | 1.05 (1.00 - 1.10) | 23 |
| Colorectal, mortality | 2 | 15,847 | 1.04 (0.88 - 1.21) | 48 |
| Breast, HER2+ | 2 | 250 | 1.03 (0.90 - 1.18) | NE |
| Breast, luminal B | 2 | 1147 | 1.03 (0.97 - 1.10) | NE |
| Colorectal | 4 | 4271 | 1.02 (0.96 - 1.07) | 0 |
| Breast, mortality | 3 | 7476 | 1.01 (0.97 - 1.04) | 11 |
| Breast, ER- | 2 | 1693 | 1.01 (0.97 - 1.07) | NE |
| Colon, mortality | 2 | NR | 1.00 (0.91 - 1.07) | NE |
| Breast | 3 | 13,004 | 0.99 (0.97 - 1.01) | 0 |
| Prostate, mortality | 3 | 10,288 | 0.99 (0.96 - 1.02) | 0 |
| Breast, premenopausal | 3 | 3622 | 0.99 (0.96 - 1.03) | 0 |
| Breast, postmenopausal | 4 | 10116 | 0.98 (0.95 - 1.01) | 0 |
| Breast, basal-like | 2 | 303 | 0.97 (0.86 - 1.09) | NE |
| Breast, ER+ | 2 | 7302 | 0.97 (0.90 - 1.04) | NE |
| Lung, mortality | 3 | 34,381 | 0.97 (0.96 - 0.99)* | 0 |
| Breast, luminal A | 2 | 2673 | 0.94 (0.90 - 0.98)* | NE |
| Endometrial | 3 | 1056 | 0.90 (0.64 - 1.26) | 0 |
| **Carbonated artificially sweetened beverages** | | | | |
| Leukemia | 2 | 339 | 1.29 (1.01 - 1.64)* | 0 |
| Multiple Myeloma | 2 | 570 | 1.25 (0.80 - 1.95) | 65 |
| Pancreas | 2 | 379 | 1.02 (0.64 - 1.61) | 0 |
| Non-Hodgkin Lymphoma | 3 | 2520 | 1.00 (0.96 - 1.04) | 2 |
| Liver | 2 | 839 | 0.99 (0.91 - 1.09) | NE |
| Kidney | 2 | 248 | 0.97 (0.82 - 1.15) | NE |
| **Artificially sweetened cola** | | | | |
| Pancreas | 2 | 379 | 0.73 (0.38 - 1.38) | 0 |
| **Artificially sweetened fruit drinks** | | | | |
| Liver | 2 | 839 | 1.05 (0.85 - 1.36) | NE |

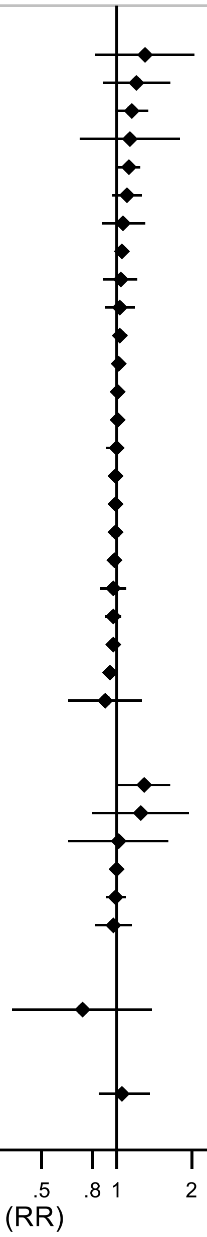

Forest plot shows the summary results across the linear dose-response (per 1 serving[355 mL]/day) inverse variance DerSimonian-Laird random-effects model meta-analyses. Each square and the horizontal line across the square represent the relative risk (RR) estimate of the corresponding meta-analysis and its 95% confidence interval (CI).

Note: Outcomes are incidence, unless otherwise stated.

* P<0.05

**Supplementary Figure 5. Summary forest plot of the linear dose-response associations of fruit juice and cancer incidence.**

##### Fruit juice, per 1 serving/day and cancer risk

| Outcome | Studies | Cases | RR (95% CI) | I² (%) |
|---|---|---|---|---|
| **Fruit juice** | | | | |
| Colorectal | 2 | 801 | 1.15 (0.79 - 1.67) | 57 |
| Prostate | 2 | 1495 | 1.09 (1.03 - 1.14)* | 75 |
| Breast, premenopausal | 2 | 2364 | 1.03 (0.96 - 1.11) | NE |
| Breast, postmenopausal | 3 | 31,449 | 1.03 (0.99 - 1.06) | 0 |
| Breast | 6 | 12,717 | 1.02 (0.99 - 1.05) | 0 |
| Kidney | 4 | 958 | 1.02 (0.87 - 1.20) | 44 |
| Head and neck | 2 | 631 | 1.01 (0.80 - 1.28) | 0 |
| Glioma | 3 | 2109 | 0.99 (0.94 - 1.05) | 0 |
| Epithelial ovarian | 12 | 2095 | 0.99 (0.92 - 1.06) | NE |
| Pancreas | 3 | 995 | 0.84 (0.62 - 1.15) | 56 |
| **Pure fruit juice** | | | | |
| Colorectal | 2 | 413 | 1.13 (0.87 - 1.48) | 0 |
| Breast | 2 | 1563 | 1.12 (0.93 - 1.35) | 28 |
| Breast, postmenopausal | 2 | 807 | 1.07 (0.88 - 1.29) | 0 |
| Breast, premenopausal | 2 | 756 | 1.03 (0.85 - 1.25) | 0 |
| **Apple juice** | | | | |
| Breast | 2 | 10,911 | 1.04 (0.93 - 1.11) | NE |
| Breast, ER- | 2 | 1794 | 0.97 (0.75 - 1.23) | NE |
| Breast, ER+ | 2 | 7464 | 0.93 (0.84 - 1.07) | NE |
| **Other fruit juices (not apple and citrus)** | | | | |
| Breast, ER- | 2 | 1794 | 1.04 (0.93 - 1.19) | NE |
| Breast, ER+ | 2 | 7464 | 0.97 (0.90 - 1.04) | NE |
| Breast | 2 | 10,911 | 0.97 (0.93 - 1.04) | NE |

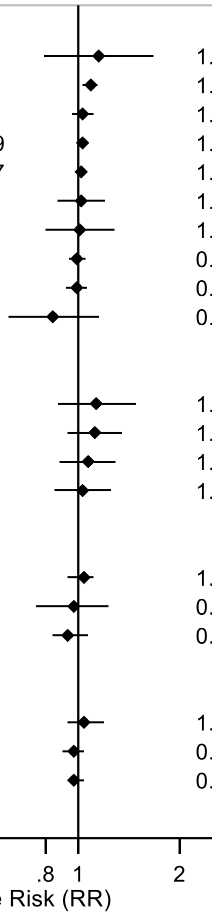

Forest plot shows the summary results across the linear dose-response (per 1 serving[177 mL]/day) inverse variance DerSimonian-Laird random-effects model meta-analyses. Each square and the horizontal line across the square represent the relative risk (RR) estimate of the corresponding meta-analysis and its 95% confidence interval (CI). No meta-analysis has been conducted for cancer mortality. Outcomes are incidence, unless otherwise stated.

Note: All analyses are based on a random-effects meta-analysis, except for prostate cancer, which is from fixed-effect meta-analysis. The RR and 95%CI for fruit juice consumption and prostate cancer incidence were 1.22 (0.89-1.68) in the random-effect meta-analysis (see Supplementary Figure 243 for additional information).

* P<0.05

**Supplementary Figure 6. Summary forest plot of the linear dose-response associations of citrus juice and cancer incidence.**

#### Citrus juice, per 1 serving/day and cancer risk

| Outcome | Studies | Cases | RR (95% CI) | I² (%) |
|---|---|---|---|---|
| **Citrus juice** | | | | |
| Melanoma | 4 | 3919 | 1.08 (0.92 - 1.27) | 63 |
| Prostate, advanced restricted | 15 | 3115 | 1.05 (0.98 - 1.13) | NE |
| Non-melanoma skin | 2 | 18,255 | 1.03 (0.95 - 1.12) | 74 |
| Prostate, localised | 15 | 38,575 | 1.02 (1.01 - 1.04)* | NE |
| Prostate, high grade | 15 | 9753 | 1.01 (0.98 - 1.04) | NE |
| Prostate | 15 | 52,680 | 1.01 (1.00 - 1.02) | NE |
| Prostate, low grade | 15 | 37,556 | 1.00 (0.99 - 1.02) | NE |
| Prostate, advanced | 15 | 4934 | 1.00 (0.94 - 1.06) | NE |
| Prostate, mortality | 15 | 3205 | 0.98 (0.91 - 1.06) | NE |
| Liver | 2 | 839 | 0.86 (0.75 - 0.98) | NE |
| Lung | 9 | 4332 | 0.86 (0.66 - 1.13) | 68 |
| **Orange juice** | | | | |
| Melanoma | 4 | 2912 | 1.21 (1.08 - 1.34)* | 0 |
| SCC, head, neck and extremities | 2 | 2758 | 1.16 (1.02 - 1.33)* | NE |
| Squamous cell carcinoma | 2 | 3544 | 1.13 (1.04 - 1.24)* | 0 |
| Basal cell carcinoma | 2 | 20,840 | 1.12 (1.06 - 1.17)* | 46 |
| SCC, truncal sites | 2 | 483 | 1.10 (0.62 - 1.97) | NE |
| Breast, ER- | 2 | 1794 | 1.04 (0.97 - 1.11) | NE |
| Breast | 2 | 10911 | 1.03 (1.00 - 1.04) | NE |
| Breast, ER+ | 2 | 7464 | 1.00 (0.97 - 1.04) | NE |
| **Grapefruit juice** | | | | |
| Squamous cell carcinoma | 2 | 3544 | 1.04 (0.82 - 1.33) | 0 |
| Basal cell carcinoma | 2 | 20,840 | 1.01 (0.91 - 1.11) | 0 |
| SCC, head, neck and extremities | 2 | 2758 | 1.01 (0.73 - 1.40) | NE |
| Breast | 2 | 7550 | 1.01 (0.91 - 1.13) | 0 |
| Melanoma | 3 | 3432 | 0.99 (0.77 - 1.28) | 0 |
| Lung | 2 | 793 | 0.61 (0.31 - 1.01) | NE |
| SCC, truncal sites | 2 | 483 | 0.45 (0.19 - 1.08) | NE |

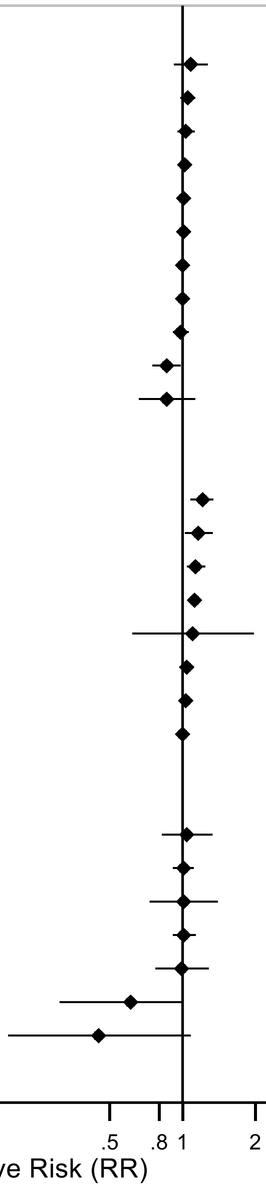

Forest plot shows the summary results across the linear dose-response (per 1 serving[177 mL]/day) inverse variance DerSimonian-Laird random-effects model meta-analyses. Each square and the horizontal line across the square represent the relative risk (RR) estimate of the corresponding meta-analysis and its 95% confidence interval (CI).

Note: Outcomes are incidence, unless otherwise stated.

* P<0.05

**Supplementary Figure 7. Summary forest plot of the linear dose-response associations of fruit and vegetable juice and vegetable juice and cancer incidence and mortality.**

#### Fruit and vegetable juice and vegetable juice and cancer risk

| Outcome | Studies | Cases | RR (95% CI) | I² (%) |
|---|---|---|---|---|
| **Fruit and vegetable juice** | | | | |
| Prostate, advanced restricted | 15 | 3115 | 1.11 (1.02 - 1.21)* | NE |
| Prostate, high grade | 15 | 9753 | 1.08 (1.02 - 1.14)* | NE |
| Prostate, advanced | 15 | 4934 | 1.06 (0.99 - 1.14) | NE |
| Distal colon | 2 | 2160 | 1.04 (1.00 - 1.08) | 0 |
| Kidney | 14 | 2345 | 1.04 (0.97 - 1.12) | 0 |
| Prostate | 15 | 52,680 | 1.03 (1.01 - 1.05)* | NE |
| Prostate, localised | 15 | 38,575 | 1.03 (1.01 - 1.05)* | NE |
| Prostate, low grade | 15 | 37,556 | 1.02 (0.99 - 1.04) | NE |
| Prostate, mortality | 15 | 3205 | 1.01 (0.94 - 1.09) | NE |
| Colon | 2 | 3560 | 1.01 (0.99 - 1.03) | 0 |
| Colorectal | 2 | 9472 | 1.00 (0.98 - 1.02) | 0 |
| Breast | 2 | 4084 | 1.00 (0.90 - 1.13) | 0 |
| Pancreas | 2 | 1005 | 0.96 (0.62 - 1.49) | 45 |
| Lung | 2 | 1304 | 0.95 (0.81 - 1.11) | 0 |
| Proximal colon | 2 | 1293 | 0.88 (0.63 - 1.23) | 70 |
| **Vegetable juice** | | | | |
| Liver | 2 | 839 | 1.15 (0.89 - 1.49) | NE |
| Prostate, advanced restricted | 15 | 3115 | 1.04 (0.95 - 1.12) | NE |
| Prostate, mortality | 15 | 3205 | 1.02 (0.95 - 1.10) | NE |
| Prostate, advanced | 15 | 4934 | 1.02 (0.99 - 1.07) | NE |
| Prostate, localised | 15 | 38,575 | 1.01 (1.00 - 1.02) | NE |
| Prostate | 15 | 52,680 | 1.01 (1.00 - 1.04) | NE |
| Prostate, high grade | 15 | 9753 | 1.01 (0.99 - 1.04) | NE |
| Prostate, low grade | 15 | 37,556 | 1.00 (0.99 - 1.01) | NE |
| **Tomato juice** | | | | |
| Prostate | 2 | 1868 | 1.25 (0.56 - 2.82) | 58 |

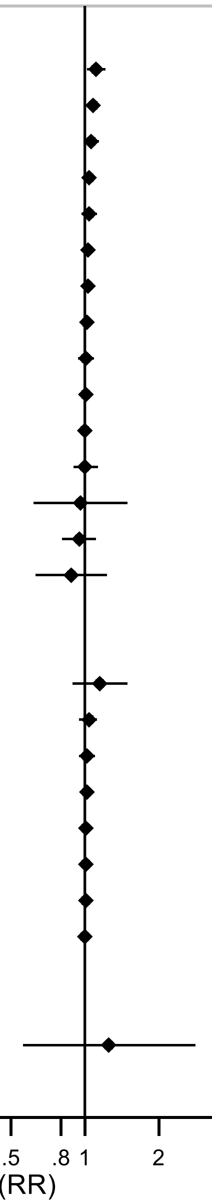

Forest plot shows the summary results across the linear dose-response (per 1 serving[177 mL]/day) inverse variance DerSimonian-Laird random-effects model meta-analyses. Each square and the horizontal line across the square represent the relative risk (RR) estimate of the corresponding meta-analysis and its 95% confidence interval (CI).

Note: Outcomes are incidence, unless otherwise stated.

* P<0.05

**Supplementary Figure 8. Summary forest plot of the linear dose-response associations of soft drinks and juices and liver cancer risk.**

**Soft drinks and juices, per 1 serving/day and liver cancer**

| Exposure | Outcome | Studies | Cases | RR (95% CI) | I² (%) |
|---|---|---|---|---|---|
| **Soft drinks** | | | | | |
| Soft drinks | Liver | 4 | 1136 | 1.15 (0.90 - 1.47) | 84 |
| Carbonated soft drinks | Liver | 2 | 839 | 1.03 (0.95 - 1.11) | NE |
| Soft drinks | Extrahepatic biliary tract | 3 | 222 | 0.99 (0.58 - 1.67) | 72 |
| Soft drinks | Intrahepatic biliary tract | 3 | 87 | 0.95 (0.60 - 1.50) | 0 |
| **Sugar sweetened beverages** | | | | | |
| SSBs | Liver | 5 | 1634 | 1.08 (0.94 - 1.24) | 31 |
| SSBs | Liver, incidence/mortality | 6 | 4356 | 1.04 (0.97 - 1.11) | 20 |
| Carbonated SSB | Liver | 2 | 839 | 1.11 (0.98 - 1.26) | NE |
| Fruit drinks | Liver | 3 | 1046 | 1.10 (0.72 - 1.69) | 53 |
| Sugar sweetened fruit drinks | Liver | 2 | 839 | 0.82 (0.59 - 1.14) | NE |
| **Artificially sweetened beverages** | | | | | |
| ASB | Liver | 4 | 1147 | 1.20 (0.88 - 1.64) | 86 |
| ASB | Liver, incidence/mortality | 5 | 3869 | 1.10 (0.96 - 1.26) | 83 |
| Carbonated ASB | Liver | 2 | 839 | 0.99 (0.91 - 1.09) | NE |
| Artificially sweetened fruit drinks | Liver | 2 | 839 | 1.05 (0.85 - 1.36) | NE |
| **Juices** | | | | | |
| Fruit juice + pure fruit juice | Liver, incidence/mortality | 2 | 783 | 0.82 (0.63 - 1.06) | 0 |
| Citrus juice | Liver | 2 | 839 | 0.86 (0.75 - 0.98)* | NE |
| Vegetables juice | Liver | 2 | 839 | 1.15 (0.89 - 1.49) | NE |

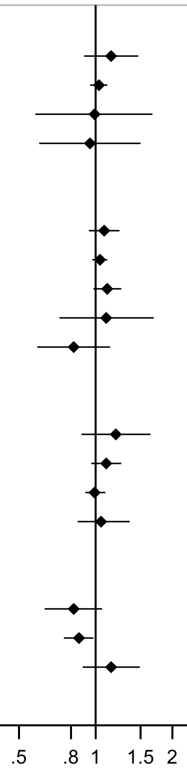

Relative Risk (RR)

Forest plot shows the summary results across the linear dose-response (per 1 serving[355 mL of soft drinks, SSBs, and ASBs, and 177 mL of juices]/day) inverse variance DerSimonian-Laird random-effects model meta-analyses. Each square and the horizontal line across the square represent the relative risk (RR) estimate of the corresponding meta-analysis and its 95% confidence interval (CI).

* P<0.05

Note: Outcomes are incidence, unless otherwise stated.

**Supplementary Figure 9. Summary forest plot of the linear dose-response associations of soft drinks and juices and pancreatic cancer risk.**

**Soft drinks and juices, per 1 serving/day and pancreatic cancer**

| Exposure | Outcome | Studies | Cases | RR (95% CI) | I² (%) |
|---|---|---|---|---|---|
| **Soft drinks** | | | | | |
| Soft drinks | Pancreas | 4 | 2641 | 1.06 (0.97 - 1.17) | 63 |
| **Sugar sweetened beverages** | | | | | |
| SSB | Pancreas | 4 | 2770 | 1.08 (0.95 - 1.22) | 44 |
| SSB | Pancreas, incidence/mortality | 5 | 11,966 | 1.04 (0.96 - 1.13) | 44 |
| Carbonated SSB | Pancreas | 15 | 2481 | 1.13 (1.03 - 1.24)* | 0 |
| SSB + Carbonated SSB | Pancreas | 18 | 4914 | 1.09 (1.01 - 1.16)* | 8 |
| Carbonated SSB | Adenocarcinoma of pancreas | 14 | 1556 | 1.17 (1.06 - 1.30)* | NE |
| Sugar sweetened cola | Pancreas | 2 | 379 | 1.18 (0.53 - 2.62) | 28 |
| **Artificially sweetened beverages** | | | | | |
| ASB | Pancreas | 2 | 1700 | 1.12 (1.00 - 1.24) | 0 |
| ASB | Pancreas, incidence/mortality | 3 | 10,896 | 1.05 (1.00 - 1.10) | 23 |
| Carbonated ASB | Pancreas | 2 | 379 | 1.02 (0.64 - 1.61) | 0 |
| ASB + Carbonated ASB | Pancreas | 5 | 11,275 | 1.03 (1.01 - 1.05)* | 0 |
| Artificially sweetened cola | Pancreas | 2 | 379 | 0.73 (0.38 - 1.38) | 0 |
| **Juices** | | | | | |
| Fruits and vegetables juices | Pancreas | 2 | 1005 | 0.96 (0.62 - 1.49) | 45 |
| Fruit juice | Pancreas | 3 | 995 | 0.84 (0.62 - 1.15) | 56 |
| Fruit juice + pure fruit juice | Pancreas, incidence/mortality | 3 | 1188 | 0.86 (0.67 - 1.11) | 33 |

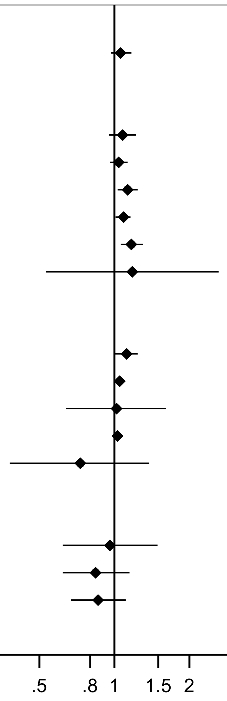

Relative Risk (RR)

Forest plot shows the summary results across the linear dose-response (per 1 serving[355 mL of soft drinks, SSBs, and ASBs, and 177 mL of juices]/day) inverse variance DerSimonian-Laird random-effects model meta-analyses. Each square and the horizontal line across the square represent the relative risk (RR) estimate of the corresponding meta-analysis and its 95% confidence interval (CI).

* P<0.05

Note: Outcomes are incidence, unless otherwise stated.

**Supplementary Figure 10. Summary forest plot of the linear dose-response associations of soft drinks and juices and colorectal cancer risk.**

**Soft drinks and juices, per 1 serving/day and colorectal cancer risk**

| Exposure | Outcome | Studies | Cases | RR (95% CI) | I² (%) |
|---|---|---|---|---|---|
| **Colorectal cancer** | | | | | |
| SSB | Colorectal | 13 | 12,649 | 1.07 (1.00 - 1.14)* | 41 |
| SSB | Colorectal, mortality | 4 | 16,855 | 1.02 (1.00 - 1.04)* | 0 |
| ASB | Colorectal | 4 | 4271 | 1.02 (0.96 - 1.07) | 0 |
| ASB | Colorectal, mortality | 2 | 15,847 | 1.04 (0.88 - 1.21) | 48 |
| Fruits and vegetables juices | Colorectal | 2 | 9472 | 1.00 (0.98 - 1.02) | 0 |
| Fruit juice | Colorectal | 2 | 801 | 1.15 (0.79 - 1.67) | 57 |
| Pure fruit juices | Colorectal | 2 | 413 | 1.13 (0.87 - 1.48) | 0 |
| Fruit juice + pure fruit juice | Colorectal | 4 | 1214 | 1.06 (0.93 - 1.21) | 0 |
| **Colon cancer** | | | | | |
| SSB | Colon | 4 | 7051 | 1.01 (0.97 - 1.05) | 0 |
| Carbonated SSB | Colon | 10 | 5604 | 1.00 (0.91 - 1.10) | 0 |
| SSB + Carbonated SSB | Colon | 14 | 10,030 | 1.01 (0.97 - 1.05) | 0 |
| SSB | Colon, mortality | 2 | NR | 1.09 (0.99 - 1.19) | NE |
| ASB | Colon, mortality | 2 | NR | 1.00 (0.91 - 1.07) | NE |
| Fruits and vegetables juices | Colon | 2 | 3560 | 1.01 (0.99 - 1.03) | 0 |
| **Proximal colon cancer** | | | | | |
| SSB | Proximal colon | 5 | 3929 | 1.03 (0.96 - 1.11) | 60 |
| SSB | Proximal colon, mortality | 2 | 409 | 1.38 (1.11 - 1.71)* | 0 |
| Substituting SSB with ASB | Proximal colon | 2 | 1275 | 0.84 (0.77 - 0.95)* | NE |
| Substituting SSB with ASB | Proximal colon, mortality | 2 | 409 | 0.80 (0.71 - 0.95)* | NE |
| Fruits and vegetables juices | Proximal colon | 2 | 1293 | 0.88 (0.63 - 1.23) | 70 |
| **Distal colon cancer** | | | | | |
| SSB | Distal colon | 5 | 3079 | 1.03 (0.94 - 1.13) | 41 |
| SSB | Distal colon, mortality | 2 | 266 | 0.97 (0.70 - 1.34) | NE |
| Fruits and vegetables juices | Distal colon | 2 | 2160 | 1.04 (1.00 - 1.08) | 0 |
| **Rectal cancer** | | | | | |
| SSB | Rectal | 4 | 2164 | 0.98 (0.91 - 1.04) | 0 |
| SSB | Rectal, mortality | 2 | 226 | 0.92 (0.64 - 1.31) | NE |

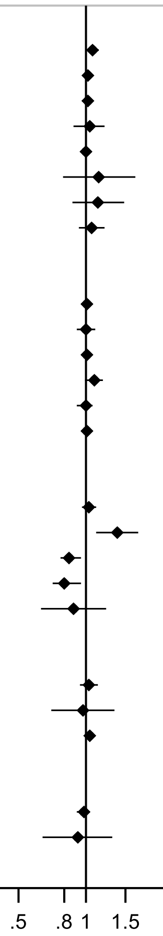

Relative Risk (RR)

Forest plot shows the summary results across the linear dose-response (per 1 serving[355 mL of soft drinks, SSBs, and ASBs, and 177 mL of juices]/day) inverse variance DerSimonian-Laird random-effects model meta-analyses. Each square and the horizontal line across the square represent the relative risk (RR) estimate of the corresponding meta-analysis and its 95% confidence interval (CI).

* P<0.05

Note: Outcomes are incidence, unless otherwise stated.

**Supplementary Figure 11. Summary forest plot of the linear dose-response associations of soft drinks and juices and lung cancer risk.**

**Soft drinks and juices, per 1 serving/day and lung cancer**

| Exposure | Outcome | Studies | Cases | RR (95% CI) | I² (%) |
|---|---|---|---|---|---|
| **Soft drinks** | | | | | |
| Soft drinks | Lung | 2 | 1299 | 0.98 (0.91 - 1.06) | 0 |
| **Sugar sweetened beverages** | | | | | |
| SSB | Lung | 2 | 2176 | 0.89 (0.81 - 0.97)* | 0 |
| SSB | Lung, mortality | 3 | 34,381 | 0.98 (0.97 - 1.00)* | 0 |
| SSB | Lung, incidence/mortality | 5 | 36,557 | 0.97 (0.92 - 1.02) | 56 |
| **Artificially sweetened beverages** | | | | | |
| ASB | Lung, mortality | 3 | 34,381 | 0.97 (0.96 - 0.99)* | 0 |
| **Juices** | | | | | |
| Fruits and vegetables juices | Lung | 2 | 1304 | 0.95 (0.81 - 1.11) | 0 |
| Citrus juice | Lung | 9 | 4332 | 0.86 (0.66 - 1.13) | 68 |
| Grapefruit juice | Lung | 2 | 793 | 0.61 (0.31 - 1.01) | NE |

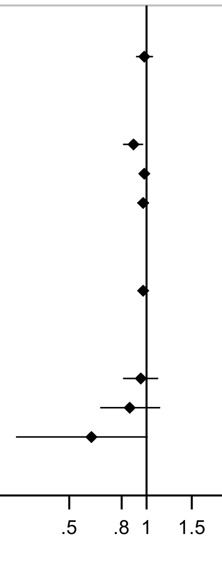

Forest plot shows the summary results across the linear dose-response (per 1 serving[355 mL of soft drinks, SSBs, and ASBs, and 177 mL of juices]/day) inverse variance DerSimonian-Laird random-effects model meta-analyses. Each square and the horizontal line across the square represent the relative risk (RR) estimate of the corresponding meta-analysis and its 95% confidence interval (CI).

* P<0.05

Note: Outcomes are incidence, unless otherwise stated.

**Supplementary Figure 12. Summary forest plot of the linear dose-response associations of soft drinks and juices and skin cancer risk.**

##### Citrus juice, per 1 serving/day and skin cancers

| Exposure | Outcome | Studies | Cases | RR (95% CI) | I² (%) |
|---|---|---|---|---|---|
| **Citrus juice** | | | | | |
| Citrus juice | Melanoma | 4 | 3919 | 1.08 (0.92 - 1.27) | 63 |
| Citrus juice | Non-melanoma skin | 2 | 18,255 | 1.03 (0.95 - 1.12) | 74 |
| **Orange juice** | | | | | |
| Orange juice | Melanoma | 4 | 2912 | 1.21 (1.08 - 1.34)* | 0 |
| Orange juice | Basal cell carcinoma | 2 | 20,840 | 1.12 (1.06 - 1.17)* | 46 |
| Orange juice | Squamous cell carcinoma | 2 | 3544 | 1.13 (1.04 - 1.24)* | 0 |
| Orange juice | SCC, head, neck and extremities | 2 | 2758 | 1.16 (1.02 - 1.33)* | NE |
| Orange juice | SCC, truncal sites | 2 | 483 | 1.10 (0.62 - 1.97) | NE |
| **Citrus + orange juice** | | | | | |
| Citrus + orange juices | Melanoma | 7 | 6206 | 1.14 (1.00 - 1.31)* | 56 |
| Citrus + orange juices | Basal cell carcinoma | 3 | 26,444 | 1.12 (1.08 - 1.16)* | 5 |
| Citrus + orange juices | Squamous cell carcinoma | 3 | 4709 | 1.17 (1.06 - 1.30)* | 28 |
| **Grapefruit juice** | | | | | |
| Grapefruit juice | Melanoma | 3 | 3432 | 0.99 (0.77 - 1.28) | 0 |
| Grapefruit juice | Basal cell carcinoma | 2 | 20,840 | 1.01 (0.91 - 1.11) | 0 |
| Grapefruit juice | Squamous cell carcinoma | 2 | 3544 | 1.04 (0.82 - 1.33) | 0 |
| Grapefruit juice | SCC, head, neck and extremities | 2 | 2758 | 1.01 (0.73 - 1.40) | NE |
| Grapefruit juice | SCC, truncal sites | 2 | 483 | 0.45 (0.19 - 1.08) | NE |

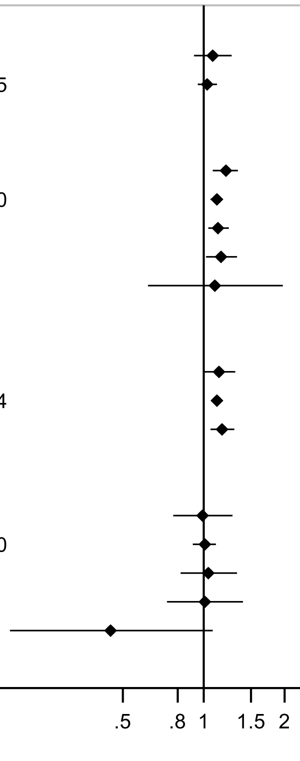

Forest plot shows the summary results across the linear dose-response (per 1 serving[355 mL of soft drinks, SSBs, and ASBs, and 177 mL of juices]/day) inverse variance DerSimonian-Laird random-effects model meta-analyses. Each square and the horizontal line across the square represent the relative risk (RR) estimate of the corresponding meta-analysis and its 95% confidence interval (CI).

* P<0.05

Note: Outcomes are incidence, unless otherwise stated

**Supplementary Figure 13. Summary forest plot of the linear dose-response associations of soft drinks and juices and ovary and endometrial cancer risk.**

**Soft drinks and juices, per 1 serving/day and ovary and endometrial cancer risk**

| Exposure | Outcome | Studies | Cases | RR (95% CI) | $I^2$ (%) |
|---|---|---|---|---|---|
| **Endometrial cancer** | | | | | |
| Soft drinks | Endometrial | 2 | 1389 | 0.94 (0.66 - 1.34) | 0 |
| SSB | Endometrial | 3 | 931 | 1.21 (1.03 - 1.42)* | 0 |
| SSB | Endometrial, type 1 | 2 | 631 | 1.45 (0.75 - 2.79) | 75 |
| ASB | Endometrial | 3 | 1056 | 0.90 (0.64 - 1.26) | 0 |
| ASB | Endometrial, postmenopausal | 3 | 900 | 1.13 (0.71 - 1.79) | 64 |
| Fruit juice + pure fruit juice | Endometrial | 2 | 761 | 1.09 (0.92 - 1.29) | |
| **Ovarian cancer** | | | | | |
| SSB | Ovary | 2 | 228 | 1.61 (1.03 - 2.53)* | 0 |
| ASB | Ovary | 2 | 381 | 1.30 (0.82 - 2.05) | 0 |
| Fruit juice | Epithelial ovarian | 12 | 2095 | 0.99 (0.92 - 1.06) | NE |
| Fruit juice + pure fruit juice | Epithelial ovarian | 13 | 2228 | 0.99 (0.93 - 1.06) | 0 |

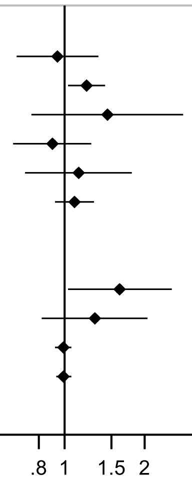

Relative Risk (RR)

Forest plot shows the summary results across the linear dose-response (per 1 serving[355 mL of soft drinks, SSBs, and ASBs, and 177 mL of juices]/day) inverse variance DerSimonian-Laird random-effects model meta-analyses. Each square and the horizontal line across the square represent the relative risk (RR) estimate of the corresponding meta-analysis and its 95% confidence interval (CI).

* P<0.05

Note: Outcomes are incidence, unless otherwise stated.

**Supplementary Figure 14. Summary forest plot of the linear dose-response associations of soft drinks and juices and breast cancer risk.**

#### Soft drinks and juices, per 1 serving/day and breast cancer risk

| Exposure | Outcome | Studies | Cases | RR (95% CI) | I² (%) |
|---|---|---|---|---|---|
| **Breast cancer** | | | | | |
| Soft drinks | Breast | 2 | 12,604 | 1.00 (0.97 - 1.04) | 0 |
| SSB | Breast | 8 | 15,855 | 1.07 (0.98 - 1.18) | 56 |
| SSB | Breast, mortality | 3 | 7476 | 1.05 (0.97 - 1.14) | 0 |
| ASB | Breast | 3 | 13,004 | 0.99 (0.97 - 1.01) | 0 |
| ASB | Breast, mortality | 3 | 7476 | 1.01 (0.97 - 1.04) | 11 |
| Fruits and vegetables juices | Breast | 2 | 4084 | 1.00 (0.90 - 1.13) | 0 |
| Fruit juice | Breast | 6 | 12,717 | 1.02 (0.99 - 1.05) | 0 |
| Pure fruit juices | Breast | 2 | 1563 | 1.12 (0.93 - 1.35) | 28 |
| Fruit juice + pure fruit juice | Breast | 8 | 14,309 | 1.02 (0.99 - 1.05) | 0 |
| Apple juice | Breast | 2 | 10,911 | 1.04 (0.93 - 1.11) | NE |
| Other fruit juice | Breast | 2 | 10,911 | 0.97 (0.93 - 1.04) | NE |
| Orange juice | Breast | 2 | 10,911 | 1.03 (1.00 - 1.04) | NE |
| Grapefruit juice | Breast | 2 | 7550 | 1.01 (0.91 - 1.13) | 0 |
| **Premenopausal breast cancer** | | | | | |
| SSB | Breast, premenopausal | 5 | 3970 | 1.26 (0.95 - 1.68) | 64 |
| ASB | Breast, premenopausal | 3 | 3622 | 0.99 (0.96 - 1.03) | 0 |
| Fruit juice | Breast, premenopausal | 2 | 2364 | 1.03 (0.96 - 1.11) | NE |
| Pure fruit juices | Breast, premenopausal | 2 | 756 | 1.03 (0.85 - 1.25) | 0 |
| Fruit juice + pure fruit juice | Breast, premenopausal | 4 | 3120 | 1.03 (0.96 - 1.11) | 0 |
| **Postmenopausal breast cancer** | | | | | |
| SSB | Breast, postmenopausal | 6 | 10,774 | 1.05 (1.00 - 1.10)* | 0 |
| ASB | Breast, postmenopausal | 4 | 10,116 | 0.98 (0.95 - 1.01) | 0 |
| Fruit juice | Breast, postmenopausal | 3 | 31,449 | 1.03 (0.99 - 1.06) | 0 |
| Pure fruit juices | Breast, postmenopausal | 2 | 807 | 1.07 (0.88 - 1.29) | 0 |
| Fruit juice + pure fruit juice | Breast, postmenopausal | 5 | 32,256 | 1.03 (1.00 - 1.06) | 0 |
| **Breast cancer subtypes** | | | | | |
| SSB | Breast, ER- | 2 | 453 | 0.97 (0.89 - 1.06) | NE |
| SSB | Breast, ER+ | 2 | 2019 | 1.03 (0.98 - 1.07) | NE |
| SSB | Breast, luminal A | 2 | 765 | 1.00 (0.93 - 1.08) | NE |
| SSB | Breast, luminal B | 2 | 311 | 0.99 (0.88 - 1.12) | NE |
| SSB | Breast, HER2+ | 2 | 62 | 0.92 (0.71 - 1.19) | NE |
| SSB | Breast, basal like | 2 | 77 | 0.84 (0.67 - 1.05) | NE |
| ASB | Breast, ER- | 2 | 1693 | 1.01 (0.97 - 1.07) | NE |
| ASB | Breast, ER+ | 2 | 7302 | 0.97 (0.90 - 1.04) | NE |
| ASB | Breast, luminal A | 2 | 2673 | 0.94 (0.90 - 0.98)* | NE |
| ASB | Breast, luminal B | 2 | 1147 | 1.03 (0.97 - 1.10) | NE |
| ASB | Breast, basal-like | 2 | 303 | 0.97 (0.86 - 1.09) | NE |
| ASB | Breast, HER2+ | 2 | 250 | 1.03 (0.90 - 1.18) | NE |
| Apple juice | Breast, ER- | 2 | 1794 | 0.97 (0.75 - 1.23) | NE |
| Apple juice | Breast, ER+ | 2 | 7464 | 0.93 (0.84 - 1.07) | NE |
| Other fruit juice | Breast, ER- | 2 | 1794 | 1.04 (0.93 - 1.19) | NE |
| Other fruit juice | Breast, ER+ | 2 | 7464 | 0.97 (0.90 - 1.04) | NE |
| Orange juice | Breast, ER- | 2 | 1794 | 1.04 (0.97 - 1.11) | NE |
| Orange juice | Breast, ER+ | 2 | 7464 | 1.00 (0.97 - 1.04) | NE |

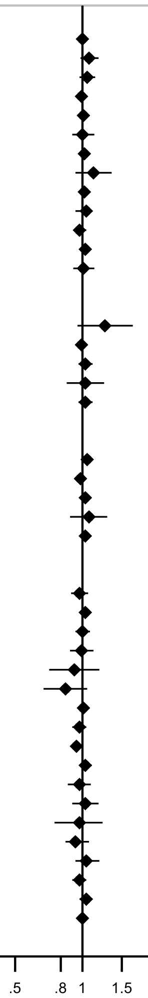

Relative Risk (RR)

Forest plot shows the summary results across the linear dose-response (per 1 serving[355 mL of soft drinks, SSBs, and ASBs, and 177 mL of juices]/day) inverse variance DerSimonian-Laird random-effects model meta-analyses. Each square and the horizontal line across the square represent the relative risk (RR) estimate of the corresponding meta-analysis and its 95% confidence interval (CI).

* P<0.05

Note: Outcomes are incidence, unless otherwise stated.

**Supplementary Figure 15. Summary forest plot of the linear dose-response associations of soft drinks and juices and prostate cancer risk.**

**Soft drinks and juices, per 1 serving/day and prostate cancer risk**

| Exposure | Outcome | Studies | Cases | RR (95% CI) | I² (%) |
|---|---|---|---|---|---|
| **Prostate cancer** | | | | | |
| SSBs | Prostate | 3 | 897 | 1.21 (0.94 - 1.57) | 0 |
| SSB | Prostate, mortality | 3 | 10,288 | 0.99 (0.97 - 1.01) | 0 |
| ASB | Prostate, mortality | 3 | 10,288 | 0.99 (0.96 - 1.02) | 0 |
| Fruits and vegetables juices | Prostate | 15 | 52,680 | 1.03 (1.01 - 1.05)* | NE |
| Fruits and vegetables juices | Prostate, mortality | 15 | 3205 | 1.01 (0.94 - 1.09) | NE |
| Fruit juice | Prostate | 2 | 1495 | 1.09 (1.03 - 1.14)* | 75 |
| Fruit juice + pure fruit juice | Prostate | 3 | 1786 | 1.09 (1.03 - 1.14)* | 54 |
| Citrus juice | Prostate | 15 | 52,680 | 1.01 (1.00 - 1.02) | NE |
| Citrus juice | Prostate, mortality | 15 | 3205 | 0.98 (0.91 - 1.06) | NE |
| Vegetables juice | Prostate | 15 | 52,680 | 1.01 (1.00 - 1.04) | NE |
| Vegetables juice | Prostate, mortality | 15 | 3205 | 1.02 (0.95 - 1.10) | NE |
| Tomato juice | Prostate | 2 | 1868 | 1.25 (0.56 - 2.82) | 58 |
| **Low-grade/localised prostate cancer** | | | | | |
| Fruits and vegetables juices | Prostate, localised | 15 | 38,575 | 1.03 (1.01 - 1.05)* | NE |
| Fruits and vegetables juices | Prostate, low grade | 15 | 37,556 | 1.02 (0.99 - 1.04) | NE |
| Citrus juice | Prostate, low grade | 15 | 37,556 | 1.00 (0.99 - 1.02) | NE |
| Citrus juice | Prostate, localised | 15 | 38,575 | 1.02 (1.01 - 1.04)* | NE |
| Vegetables juice | Prostate, localised | 15 | 38,575 | 1.01 (1.00 - 1.02) | NE |
| Vegetables juice | Prostate, low grade | 15 | 37,556 | 1.00 (0.99 - 1.01) | NE |
| **High-grade/advanced prostate cancer** | | | | | |
| Fruits and vegetables juices | Prostate, advanced | 15 | 4934 | 1.06 (0.99 - 1.14) | NE |
| Fruits and vegetables juices | Prostate, advanced restricted | 15 | 3115 | 1.11 (1.02 - 1.21)* | NE |
| Fruits and vegetables juices | Prostate, high grade | 15 | 9753 | 1.08 (1.02 - 1.14)* | NE |
| Citrus juice | Prostate, high grade | 15 | 9753 | 1.01 (0.98 - 1.04) | NE |
| Citrus juice | Prostate, advanced | 15 | 4934 | 1.00 (0.94 - 1.06) | NE |
| Citrus juice | Prostate, advanced restricted | 15 | 3115 | 1.05 (0.98 - 1.13) | NE |
| Vegetables juice | Prostate, advanced | 15 | 4934 | 1.02 (0.99 - 1.07) | NE |
| Vegetables juice | Prostate, advanced restricted | 15 | 3115 | 1.04 (0.95 - 1.12) | NE |
| Vegetables juice | Prostate, high grade | 15 | 9753 | 1.01 (0.99 - 1.04) | NE |

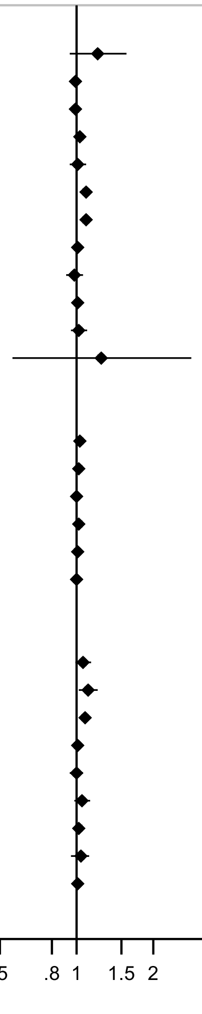

Forest plot shows the summary results across the linear dose-response (per 1 serving[355 mL of soft drinks, SSBs, and ASBs, and 177 mL of juices]/day) inverse variance DerSimonian-Laird random-effects model meta-analyses. Each square and the horizontal line across the square represent the relative risk (RR) estimate of the corresponding meta-analysis and its 95% confidence interval (CI).

* P<0.05

Note: All analyses are based on a random-effects meta-analysis, except for fruit juice and prostate cancer, which is from fixed-effect meta-analysis. The RR and 95%CI for fruit juice consumption and prostate cancer incidence were 1.22 (0.89-1.68) in the random-effect meta-analysis (see Supplementary Figure 243 for additional information).

Outcomes are incidence, unless otherwise stated.

**Supplementary Figure 16. Summary forest plot of the linear dose-response associations of soft drinks and juices and kidney cancer risk.**

##### Soft drinks and juices, per 1 serving/day and kidney cancer

| Exposure | Outcome | Studies | Cases | RR (95% CI) | I² (%) |
|---|---|---|---|---|---|
| **Soft drinks** | | | | | |
| Carbonated soft drinks | Kidney | 14 | 2066 | 1.02 (0.99 - 1.06) | 0 |
| **Sugar sweetened beverages** | | | | | |
| SSB | Kidney | 4 | 1350 | 1.13 (0.95 - 1.35) | 49 |
| Carbonated SSB | Kidney | 2 | 248 | 0.95 (0.69 - 1.31) | NE |
| SSB + Carbonated SSB | Kidney | 6 | 1598 | 1.11 (0.97 - 1.28) | 33 |
| SSB | Kidney, mortality | 2 | 3394 | 1.06 (1.02 - 1.10)* | 62 |
| **Artificially sweetened beverages** | | | | | |
| ASB | Kidney | 2 | 1389 | 1.15 (0.99 - 1.34) | 0 |
| Carbonated ASB | Kidney | 2 | 248 | 0.97 (0.82 - 1.15) | NE |
| ASB + Carbonated ASB | Kidney | 5 | 1637 | 1.08 (0.95 - 1.23) | 19 |
| ASB | Kidney, mortality | 2 | 3394 | 1.06 (0.87 - 1.30) | 64 |
| **Juices** | | | | | |
| Fruits and vegetables juices | Kidney | 14 | 2345 | 1.04 (0.97 - 1.12) | 0 |
| Fruit juice | Kidney | 4 | 958 | 1.02 (0.87 - 1.20) | 44 |

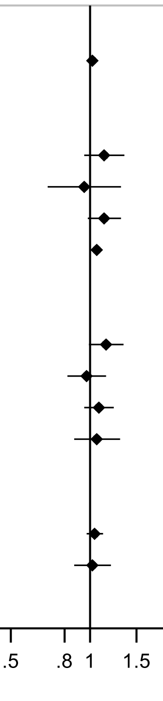

Forest plot shows the summary results across the linear dose-response (per 1 serving[355 mL of soft drinks, SSBs, and ASBs, and 177 mL of juices]/day) inverse variance DerSimonian-Laird random-effects model meta-analyses. Each square and the horizontal line across the square represent the relative risk (RR) estimate of the corresponding meta-analysis and its 95% confidence interval (CI).

* P<0.05

Note: Outcomes are incidence, unless otherwise stated.

**Supplementary Figure 17. Summary forest plot of the linear dose-response associations of soft drinks and juices and hematologic cancers risk.**

**Soft drinks, per 1 serving/day and hematologic cancers**

| Exposure | Outcome | Studies | Cases | RR (95% CI) | I² (%) |
|---|---|---|---|---|---|
| **Sugar sweetened beverages** | | | | | |
| Carbonated SSB | Multiple Myeloma | 2 | 570 | 1.26 (0.84 - 1.89) | 0 |
| Carbonated SSB | Leukemia | 2 | 339 | 1.09 (0.61 - 1.95) | 58 |
| Carbonated SSB | Non-Hodgkin Lymphoma | 3 | 2520 | 1.04 (0.87 - 1.23) | 62 |
| **Artificially sweetened beverages** | | | | | |
| Carbonated ASB | Leukemia | 2 | 339 | 1.29 (1.01 - 1.64)* | 0 |
| Carbonated ASB | Multiple Myeloma | 2 | 570 | 1.25 (0.80 - 1.95) | 65 |
| Carbonated ASB | Non-Hodgkin Lymphoma | 3 | 2520 | 1.00 (0.96 - 1.04) | 2 |

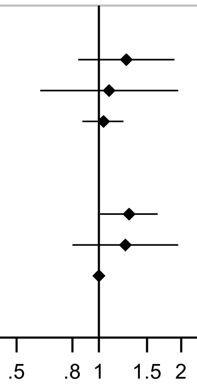

Relative Risk (RR)

Forest plot shows the summary results across the linear dose-response (per 1 serving[355 mL of soft drinks, SSBs, and ASBs, and 177 mL of juices]/day) inverse variance DerSimonian-Laird random-effects model meta-analyses. Each square and the horizontal line across the square represent the relative risk (RR) estimate of the corresponding meta-analysis and its 95% confidence interval (CI).

* P<0.05

Note: Outcomes are incidence, unless otherwise stated.

**Supplementary Figure 18. Highest versus the lowest category results from the individual studies on soft drinks and sugar sweetened beverages and incidence and mortality of cancers of the head and neck and oral cavity.**

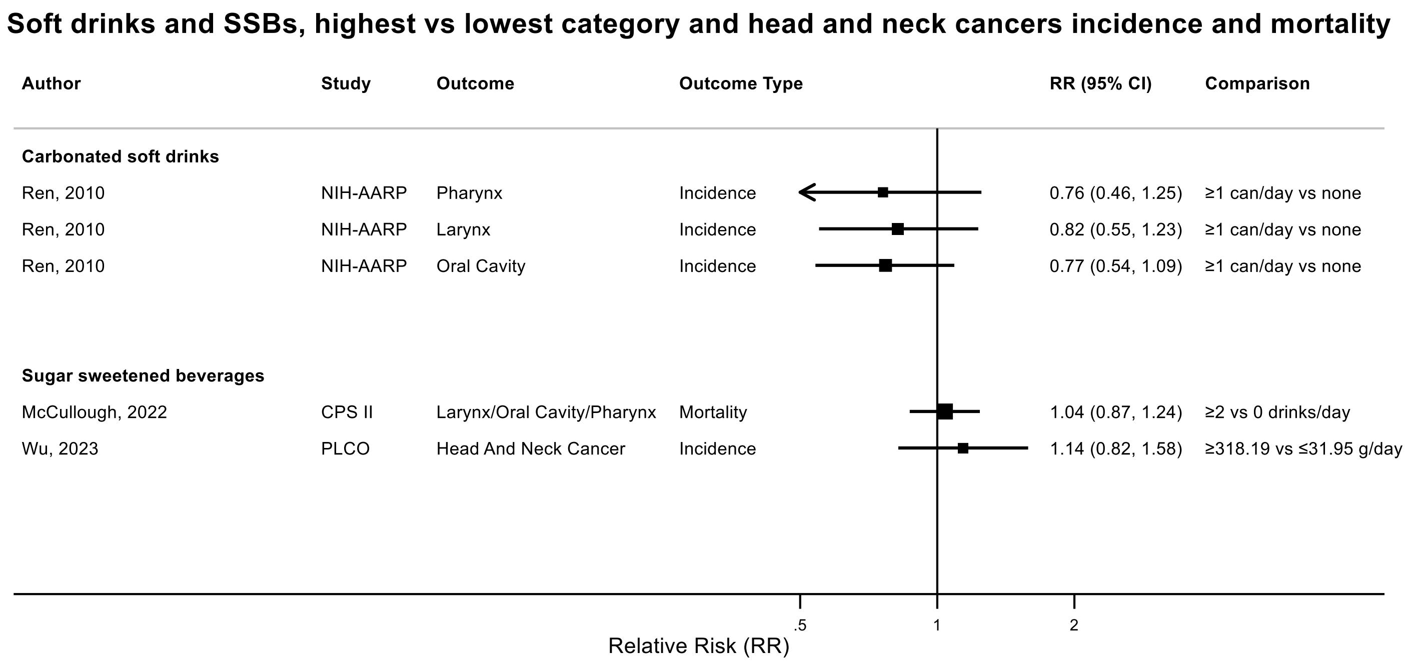

Figure legend: Forest plot shows the highest versus the lowest category results from the individual studies. Each square and the horizontal line across the square represent the relative risk (RR) estimate of the individual study and its 95% confidence interval (CI).

Abbreviations: CI, confidence interval; RR, relative risk.

Study abbreviations: NIH-AARP, National Institute of Health + American Association of Retired Persons Diet and Health Study; CPS II, Cancer Prevention Study II; PLCO, Prostate, Lung, Colorectal and Ovarian Cancer Screening trial

Ren, 2010[143], McCullough, 2022[113], Wu, 2023[172]

**Supplementary Figure 19. Highest versus the lowest category results from the individual studies on soft drinks and sugar sweetened beverages and oesophageal cancer incidence and mortality.**

**Soft drinks and SSBs, highest vs lowest category and oesophageal cancer incidence and mortality**

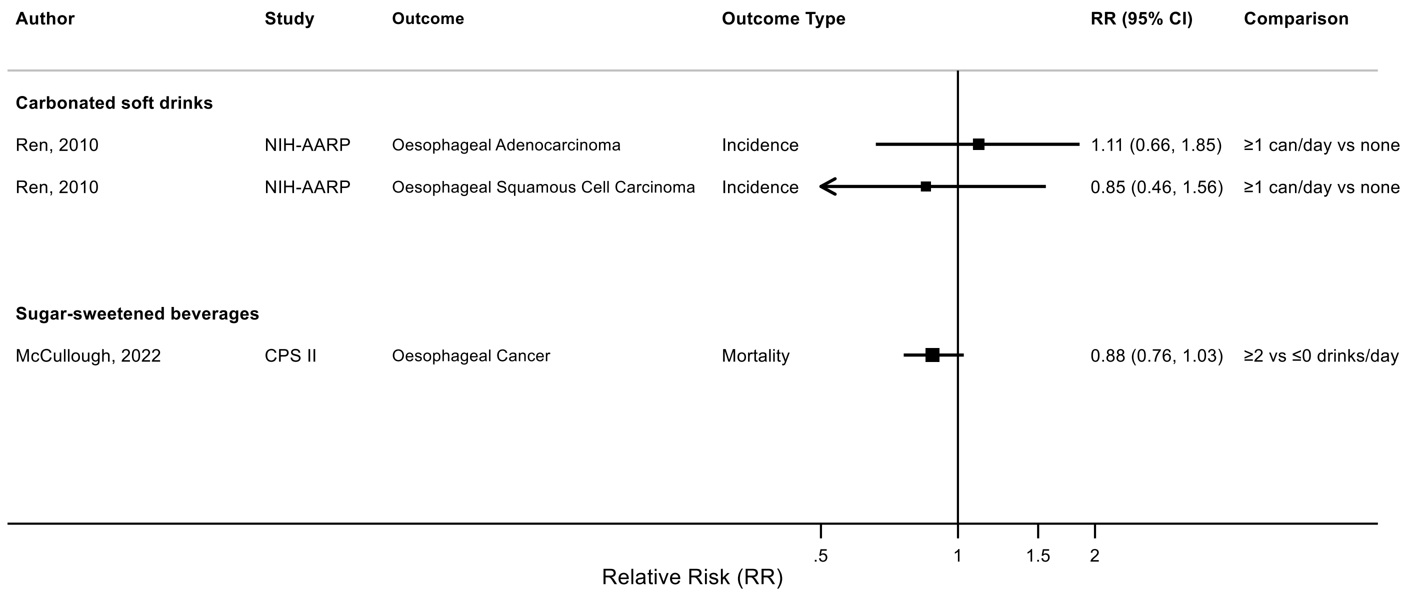

Figure legend: Forest plot shows the highest versus the lowest category results from the individual studies. Each square and the horizontal line across the square represent the RR estimate of the individual study and its 95% confidence interval (CI).

Abbreviations: CI, confidence interval; RR, relative risk.

Study abbreviations: NIH-AARP, National Institute of Health + American Association of Retired Persons Diet and Health Study; CPS II, Cancer Prevention Study II

Ren, 2010[143], McCullough, 2022[113]

**Supplementary Figure 20. Highest versus the lowest category results from the individual studies on soft drinks and sugar sweetened beverages and stomach cancer incidence and mortality.**

#### Soft drinks and SSBs, highest vs lowest category and stomach cancer incidence and mortality

| Author | Study | Outcome | Outcome Type | RR (95% CI) | Comparison |
|---|---|---|---|---|---|
| **Carbonated soft drinks** | | | | | |
| Ren, 2010 | NIH-AARP | Cardia Cancer | Incidence | 0.89 (0.55, 1.45) | ≥1 can/day vs none |
| Ren, 2010 | NIH-AARP | Non-Cardia Cancer | Incidence | 0.75 (0.45, 1.24) | ≥1 can/day vs none |
| **Sugar sweetened beverages** | | | | | |
| Khairan, 2023 | JPHC | Cardia Cancer | Incidence | 0.59 (0.31, 1.13) | ≥278.1 vs 0-0.04 ml/day |
| Khairan, 2023 | JPHC | Stomach Cancer | Incidence | 1.00 (0.86, 1.15) | ≥278.1 vs 0-0.04 ml/day |
| Hodge, 2018 | MCCS | Gastric Cardia Adenocarcinoma | Incidence | 1.17 (0.73, 1.89) | ≥1 vs ≤0.9 serving/month |
| Khairan, 2023 | JPHC | Non-Cardia Cancer | Incidence | 1.02 (0.88, 1.19) | ≥278.1 vs 0-0.04 ml/day |
| Lee, 2023 | HEXA-G | Stomach Cancer | Incidence | 0.61 (0.34, 1.08) | >250 vs 0 g/day |
| McCullough, 2022 | CPS II | Stomach Cancer | Mortality | 1.08 (0.94, 1.25) | ≥2 vs 0 drinks/day |

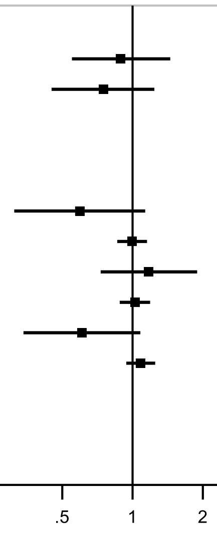

Figure legend: Forest plot shows the highest versus the lowest category results from the individual studies. Each square and the horizontal line across the square represent the RR estimate of the individual study and its 95% confidence interval (CI).

Abbreviations: CI, confidence interval; RR, relative risk.

Study abbreviations: NIH-AARP, National Institute of Health + American Association of Retired Persons Diet and Health Study; JPHC, Japan Public Health Center-Based Prospective Study; MCCS, Melbourne Collaborative Cohort Study; HEXA-G, Health Examinees-Gem study; CPS II, Cancer Prevention Study II

Ren, 2010[143], Khairan, 2023[84],Hodge, 2018[70],Lee, 2023[97], McCullough, 2022[113]

**Supplementary Figure 21. Highest versus the lowest category results from the individual studies on soft drinks and sugar sweetened beverages and colorectal cancer incidence.**

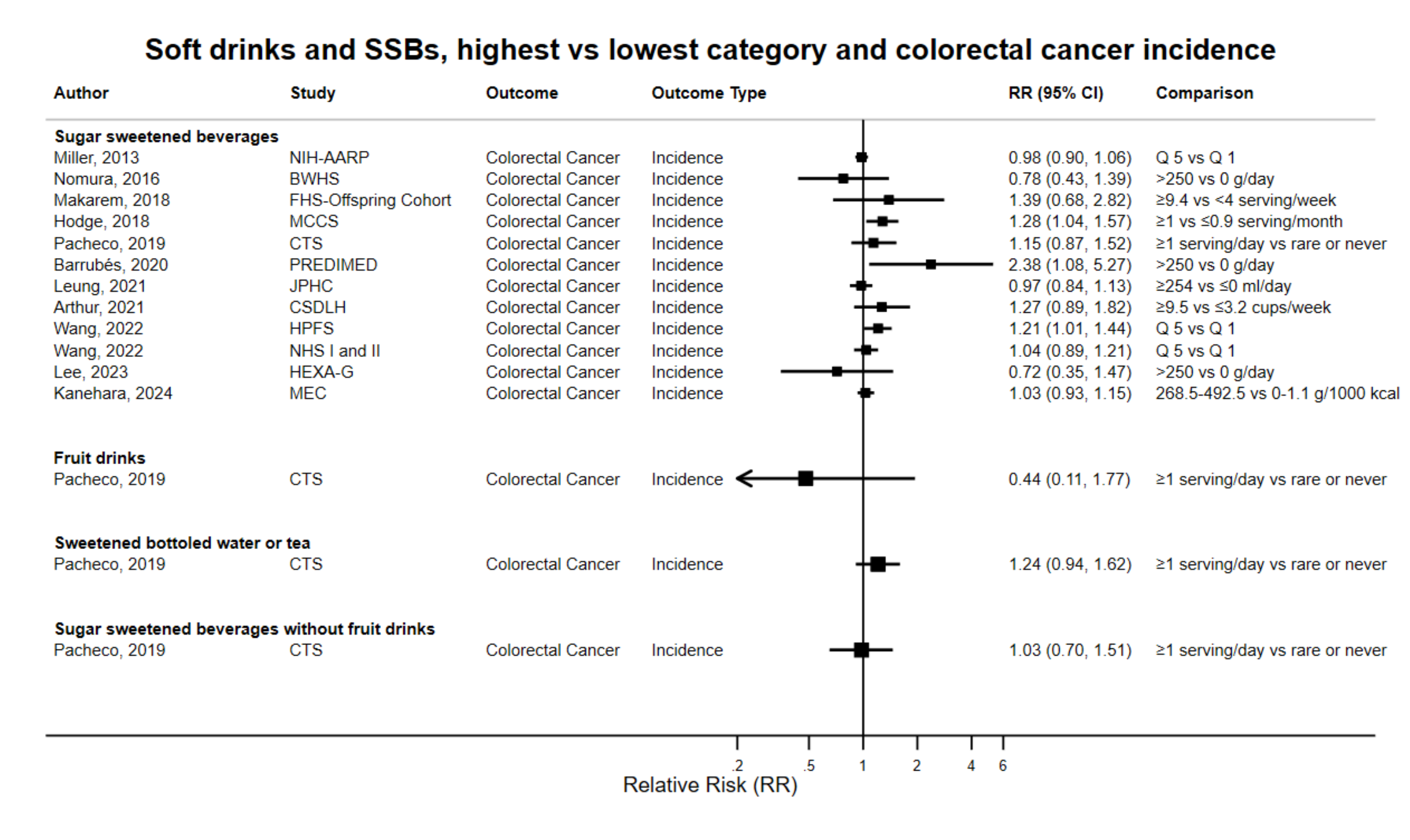

Figure legend: Forest plot shows the highest versus the lowest category results from the individual studies. Each square and the horizontal line across the square represent the RR estimate of the individual study and its 95% confidence interval (CI).

Abbreviations: CI, confidence interval; RR, relative risk.

Study abbreviations: NIH-AARP, National Institute of Health + American Association of Retired Persons Diet and Health Study; BWHS, Black Women's Health Study; FHS-Offspring Cohort, Framingham Heart Study-Offspring Cohort; MCCS, Melbourne Collaborative Cohort Study; CTS, California Teachers Study; PREDIMED, Prevención con Dieta Mediterránea study; JPHC, Japan Public Health Center-Based Prospective Study; CSDLH, Canadian Study of Diet, Lifestyle and Health; HPFS, Health Professionals Follow-up Study; NHS I and II, Nurses' Health Study I and II; HEXA-G, Health Examinees-Gem study; MEC, Multiethnic Cohort Study

Note: One additional study (UK Biobank) that could not be included in the dose-response meta-analysis because of insufficient data format reported no association between sugar-sweetened beverages and colorectal cancer risk (HR per 1 SD increase 1.00, 95% CI: 0.95-1.05). The results in males and females were consistent with the main findings and showed no statistical difference by sex (P heterogeneity=0.487)[78].

Miller, 2013[122], Nomura, 2016[129], Makarem, 2018[108], Hodge, 2018[70], Pacheco, 2019[135], Barrubés, 2020[40], Leung, 2021[101], Arthur, 2021[38], Wang, 2022[168], Lee, 2023[97], Kanehara, 2024[81]

**Supplementary Figure 22. Highest versus the lowest category results from the individual studies on soft drinks and sugar sweetened beverages and colorectal cancer mortality.**

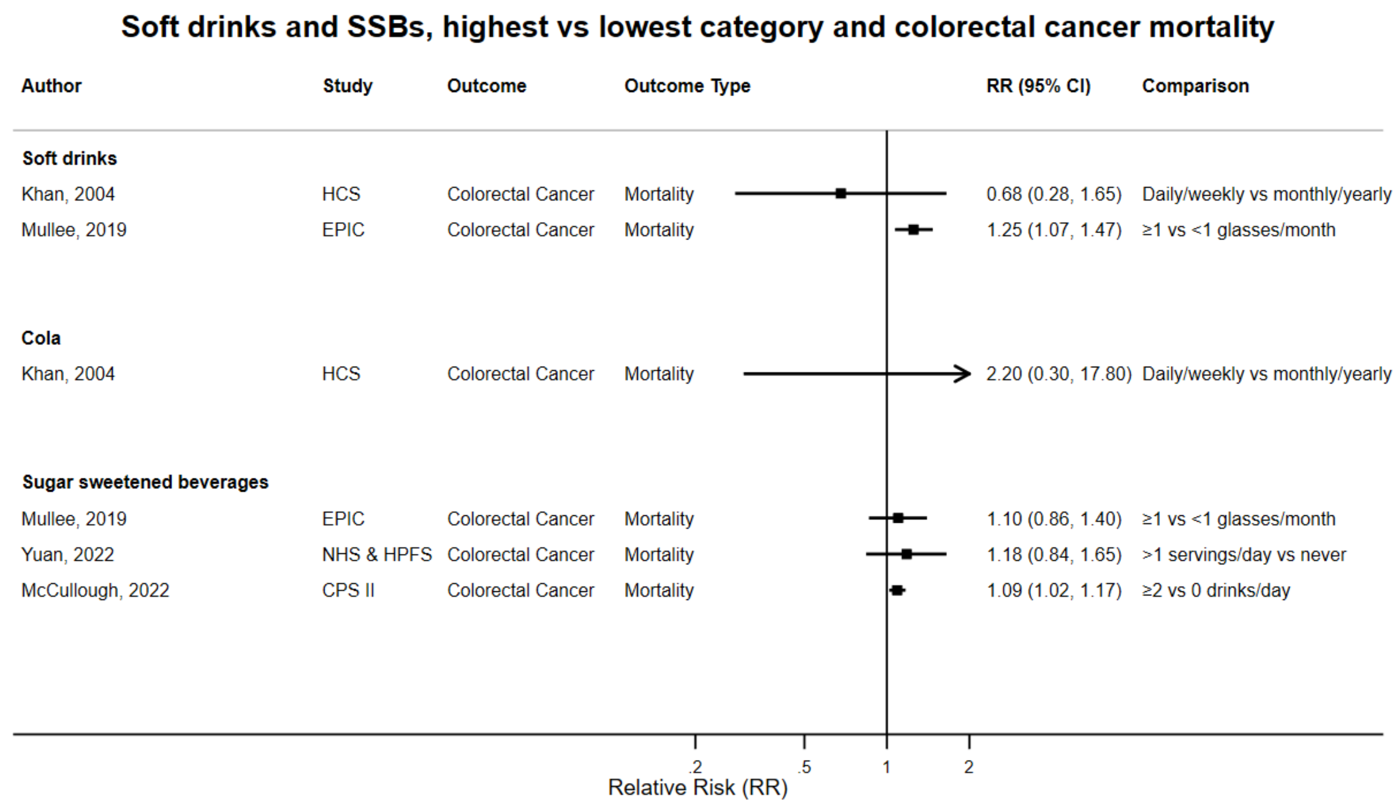

Figure legend: Forest plot shows the highest versus the lowest category results from the individual studies. Each square and the horizontal line across the square represent the RR estimate of the individual study and its 95% confidence interval (CI).

Abbreviations: CI, confidence interval; RR, relative risk.

Study abbreviations: HCS, Hokkaido Cohort, Japan; EPIC, European Prospective Investigation into Cancer and nutrition; NHS and HPFS, Nurses' Health Study and Health Professionals Follow-up Study; CPS II, Cancer Prevention Study II

Khan, 2004[85], Mullee, 2019[126], Yuan, 2022[178], McCullough, 2022[113]

#### Supplementary Figure 23. Highest versus the lowest category results from the individual studies on sugar sweetened beverages and incidence and mortality of colorectal cancer subsites, early-onset colorectal cancer, and small intestine cancer.

##### Sugar sweetened beverages, highest vs lowest category and incidence and mortality of colorectal cancer subsites

| Author | Study | Outcome | Outcome Type | RR (95% CI) | Comparison |
|---|---|---|---|---|---|
| Leung, 2021 | JPHC | Proximal Colon Cancer | Incidence | 1.09 (0.84, 1.42) | ≥254 vs 0 ml/day |
| Pacheco, 2019 | CTS | Proximal Colon Cancer | Incidence | 1.08 (0.72, 1.61) | ≥1 serving/week vs rare or never |
| Yuan, 2022 | NHS & HPFS | Proximal Colon Cancer | Incidence | 1.38 (1.01, 1.88) | >1 serving/day vs never |
| Kanehara, 2024 | MEC | Proximal Colon Cancer | Incidence | 0.98 (0.84, 1.14) | 268.5-492.5 vs 0-1.1 g/1000 kcal |
| Yuan, 2022 | NHS & HPFS | Proximal Colon Cancer | Mortality | 2.09 (1.28, 3.43) | >1 serving/day vs never |
| Leung, 2021 | JPHC | Distal Colon Cancer | Incidence | 0.98 (0.75, 1.28) | ≥254 vs 0 ml/day |
| Kanehara, 2024 | MEC | Distal Colon Cancer | Incidence | 1.15 (0.94, 1.41) | 268.5-492.5 vs 0-1.1 g/1000 kcal |
| Yuan, 2022 | NHS & HPFS | Distal Colon Cancer | Incidence | 0.81 (0.54, 1.20) | >1 serving/day vs never |
| Pacheco, 2019 | CTS | Distal Colorectum Cancer | Incidence | 1.22 (0.82, 1.80) | ≥1 sernig/day vs rare or never |
| Yuan, 2022 | NHS & HPFS | Distal Colon Cancer | Mortality | 0.98 (0.51, 1.90) | >1/day vs never |
| McCullough, 2022 | CPS II | Small Intestinal Cancer | Mortality | 1.26 (0.84, 1.90) | ≥2 vs 0 drinks/day |
| Hur, 2021 | NHS II | Early-Onset Colorectal Cancer | Incidence | 2.18 (1.10, 4.35) | ≥2 vs ≤0.9 serving/week |

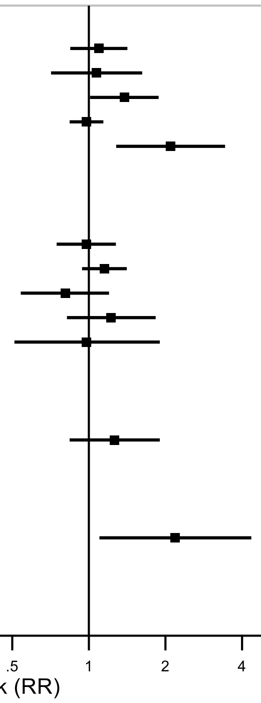

Figure legend: Forest plot shows the highest versus the lowest category results from the individual studies. Each square and the horizontal line across the square represent the RR estimate of the individual study and its 95% confidence interval (CI).

Abbreviations: CI, confidence interval; RR, relative risk.

Study abbreviations: JPHC, Japan Public Health Center-Based Prospective Study; CTS, California Teachers Study; NHS, HPFS, Nurses' Health Study, Health Professionals Follow-up Study; MEC, Multiethnic Cohort Study; CTS, California Teachers Study; CPS II, Cancer Prevention Study II; NHS II, Nurses' Health Study II

Leung, 2021[101], Pacheco, 2019[135], Yuan, 2022[178], Kanehara, 2024[81], McCullough, 2022[113], Hur, 2021[72]

**Supplementary Figure 24. Highest versus the lowest category results from the individual studies on sugar sweetened beverages and colon cancer incidence and mortality.**

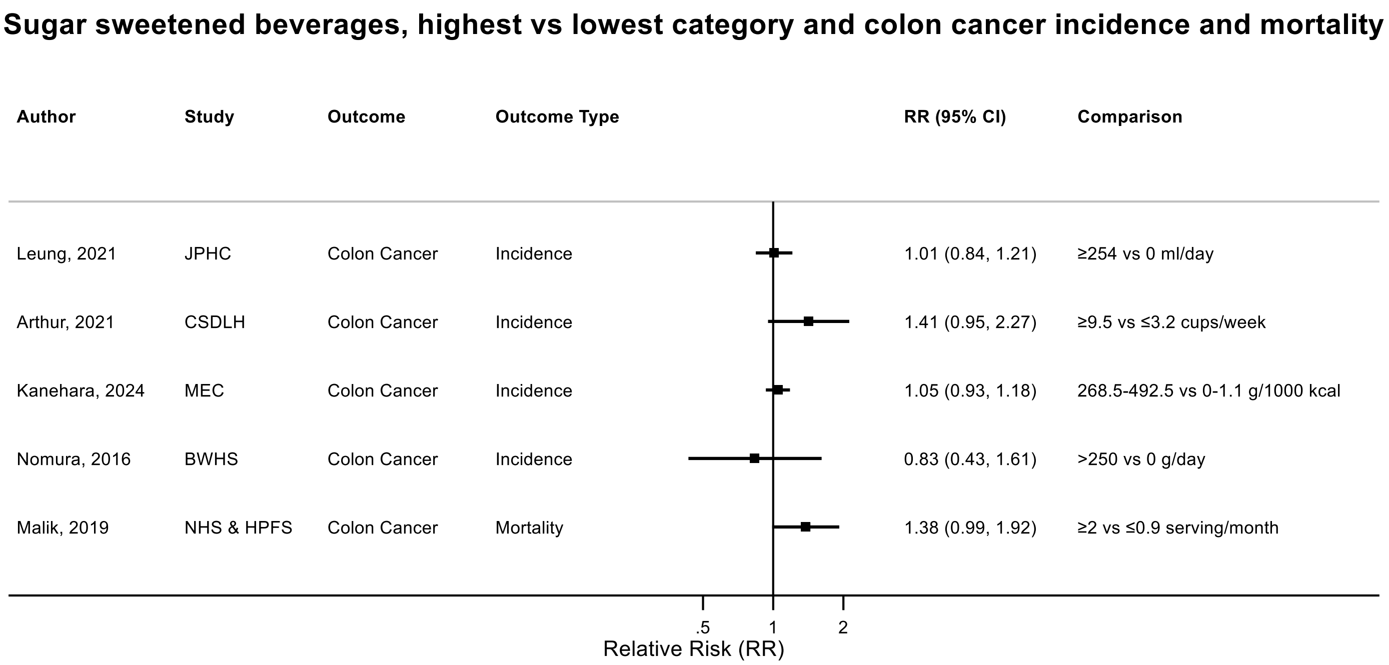

Figure legend: Forest plot shows the highest versus the lowest category results from the individual studies. Each square and the horizontal line across the square represent the RR estimate of the individual study and its 95% confidence interval (CI).

Abbreviations: CI, confidence interval; RR, relative risk.

Study abbreviations: JPHC, Japan Public Health Center-Based Prospective Study; CSDLH, Canadian Study of Diet, Lifestyle and Health; MEC, Multiethnic Cohort Study; BWHS, Black Women's Health Study; NHS & HPFS, Nurses' Health Study & Health Professionals Follow-up Study

Note: One additional study (UK Biobank) that could not be included in the dose-response meta-analysis because of insufficient data format reported no association between sugar-sweetened beverages and colon cancer risk (HR per 1 SD increase 1.01, 95% CI: 0.94-1.08)[78].

Leung, 2021[101], Arthur, 2021[38], Kanehara, 2024[81], Nomura, 2016[129], Malik, 2019[109]

**Supplementary Figure 25. Highest versus the lowest category results from the individual studies on sugar sweetened beverages and rectal cancer incidence and mortality.**

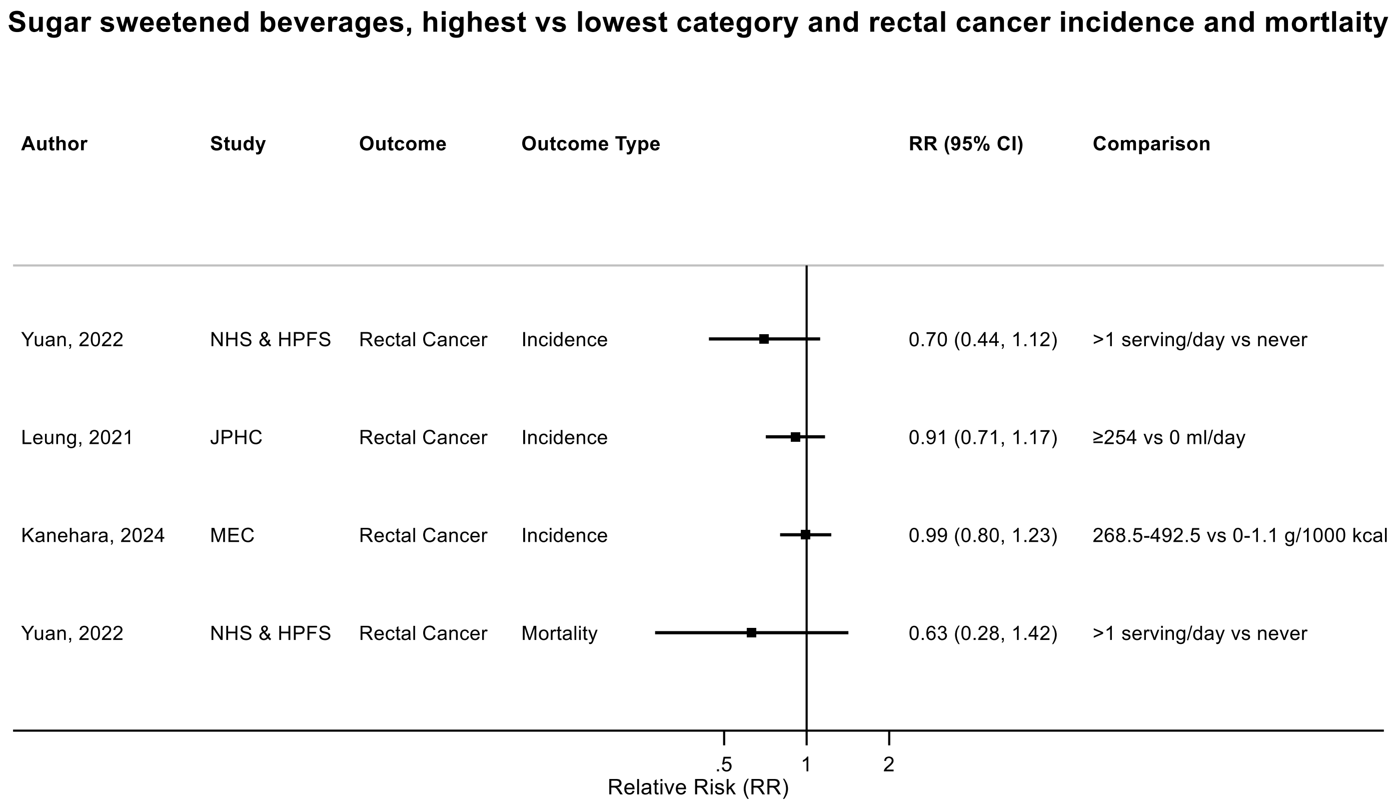

Figure legend: Forest plot shows the highest versus the lowest category results from the individual studies. Each square and the horizontal line across the square represent the RR estimate of the individual study and its 95% confidence interval (CI).

Abbreviations: CI, confidence

Study abbreviations: NHS & HPFS, Nurses' Health Study & Health Professionals Follow-up Study; JPHC, Japan Public Health Center-Based Prospective Study; MEC, Multiethnic Cohort Study

Note: One additional study (UK Biobank) that could not be included in the dose-response meta-analysis because of insufficient data format reported no association between sugar-sweetened beverages and rectal cancer risk (HR per 1 SD increase 1.01, 95% CI: 0.90-1.12)[78].

Yuan, 2022[178], Leung, 2021[101], Kanehara, 2024[81], Yuan, 2022[178]

**Supplementary Figure 26. Highest versus the lowest category results from the individual studies on soft drinks and sugar sweetened beverages and liver cancer incidence and mortality.**

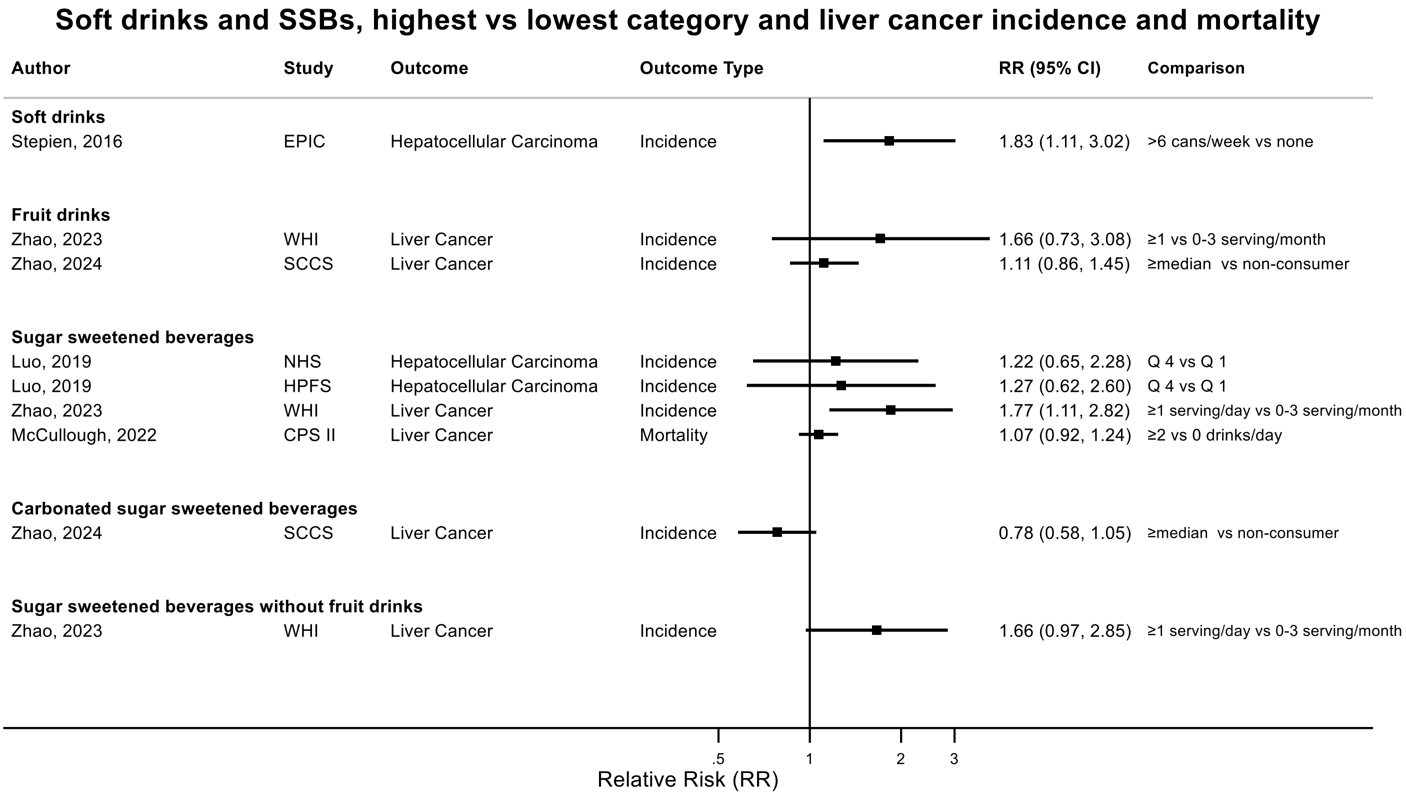

Figure legend: Forest plot shows the highest versus the lowest category results from the individual studies. Each square and the horizontal line across the square represent the RR estimate of the individual study and its 95% confidence interval (CI).

Abbreviations: CI, confidence

Study abbreviations: EPIC, European Prospective Investigation into Cancer and nutrition; WHI, Women's Health Initiative Study; SCCS, Southern Community Cohort Study; NHS, Nurses' Health Study; HPFS, Health Professionals Follow-up Study; CPS II, Cancer Prevention Study II

Stepien, 2016[159], Zhao, 2023[186], Zhao, 2024[187], Luo, 2019[105], McCullough, 2022[113]

**Supplementary Figure 27. Highest versus the lowest category results from the individual studies on soft drinks and sugar sweetened beverages and gallbladder cancer incidence and mortality.**

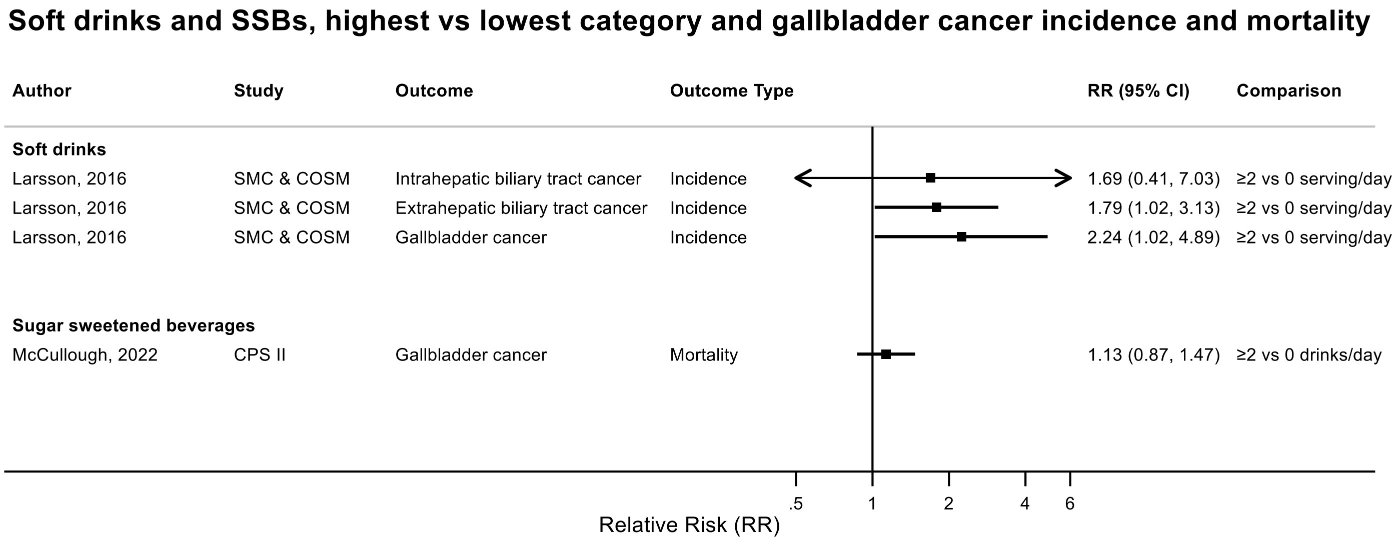

Figure legend: Forest plot shows the highest versus the lowest category results from the individual studies. Each square and the horizontal line across the square represent the RR estimate of the individual study and its 95% confidence interval (CI).

Abbreviations: CI, confidence

Study abbreviations: SMC & COSMOS, Swedish Mammography Cohort & Continuous Observation of Smoking Subjects Study; CPS II, Cancer Prevention Study II

Larsson, 2016[96], McCullough, 2022[113]

**Supplementary Figure 28. Highest versus the lowest category results from the individual studies on soft drinks and sugar sweetened beverages and pancreatic cancer incidence and mortality.**

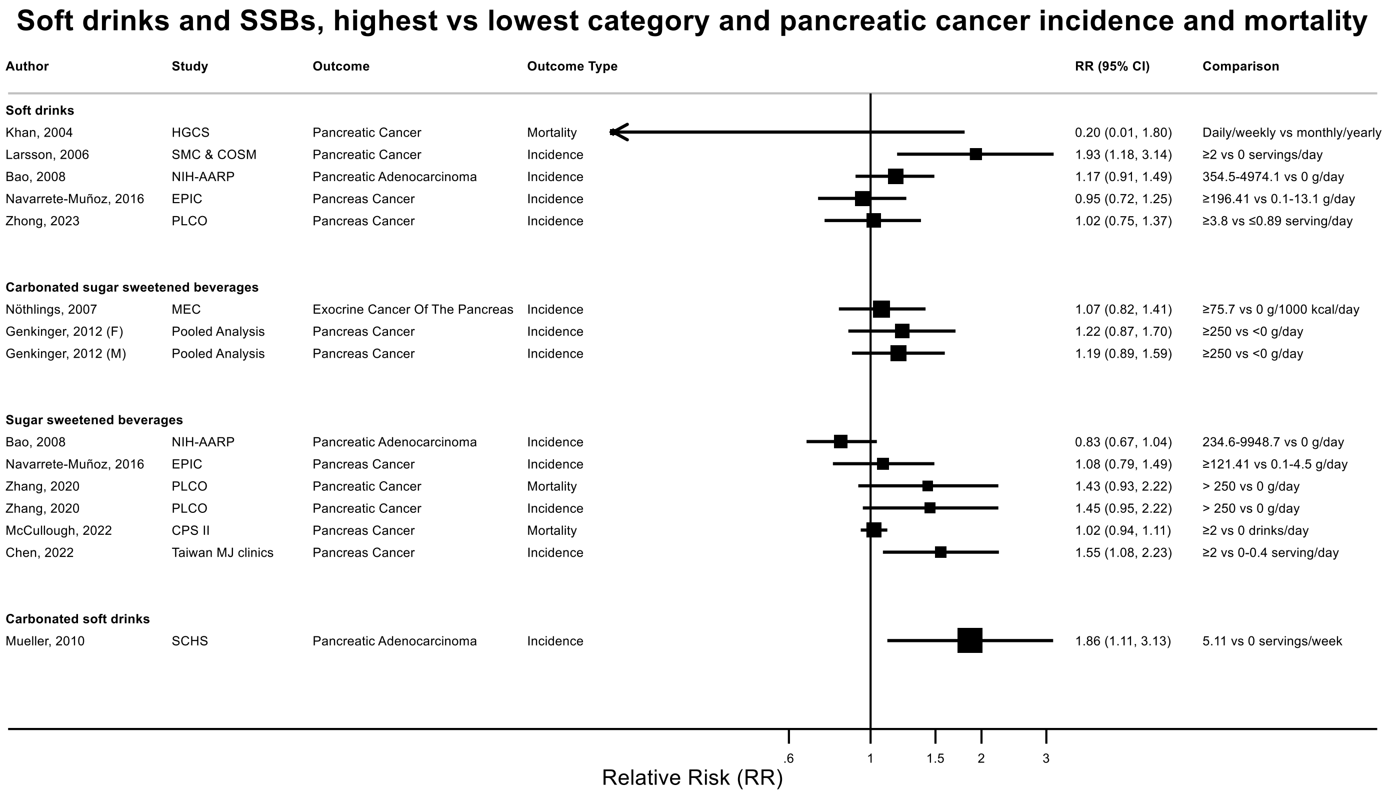

Figure legend: Forest plot shows the highest versus the lowest category results from the individual studies. Each square and the horizontal line across the square represent the RR estimate of the individual study and its 95% confidence interval (CI).

Abbreviations: CI, confidence

Study abbreviations: HCS, Hokkaido Cohort, Japan; SMC-COSMOS, Swedish Mammography Cohort & Continuous Observation of Smoking Subjects Study; NIH-AARP, National Institute of Health + American Association of Retired Persons Diet and Health Study; EPIC, European Prospective Investigation into Cancer and nutrition; PLCO, Prostate, Lung, Colorectal and Ovarian Cancer Screening trial; MEC, Multiethnic Cohort Study; CPS II, Cancer Prevention Study II; Taiwan MJ clinics, The Taiwan MJ Cohort Study; SCHS, Singapore Chinese Health study

Khan, 2004[85], Larsson, 2016[96], Bao, 2008[39], Navarrete-Muñoz, 2016[127], Zhong, 2023[189], Nöthlings, 2007[131], Genkinger, 2012[64], Zhang, 2020[184], McCullough, 2022[113], Chen, 2022[45],Mueller, 2010[125]

**Supplementary Figure 29. Highest versus the lowest category results from the individual studies on soft drinks and sugar sweetened beverages and lung cancer incidence and mortality.**

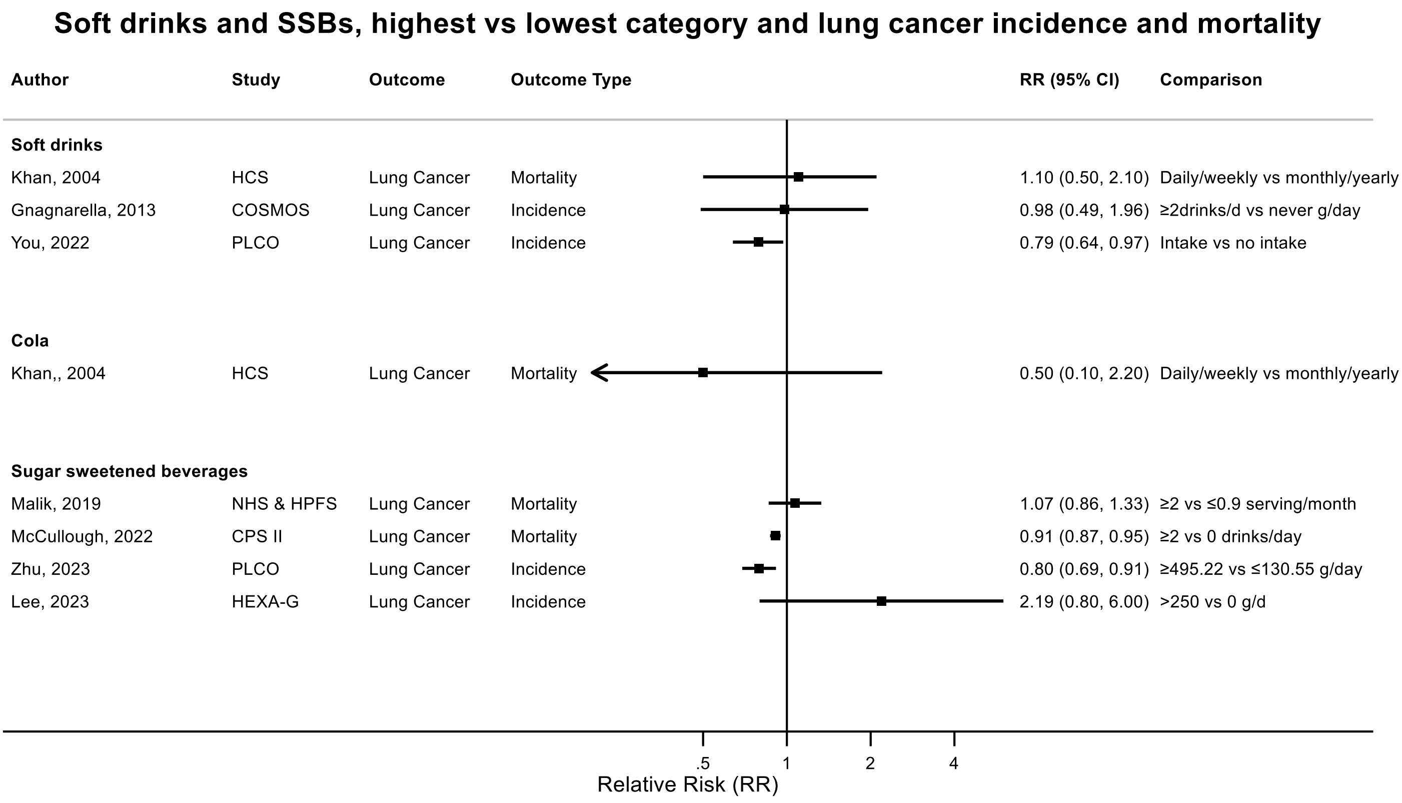

Figure legend: Forest plot shows the highest versus the lowest category results from the individual studies. Each square and the horizontal line across the square represent the RR estimate of the individual study and its 95% confidence interval (CI).

Abbreviations: CI, confidence

Study abbreviations: HCS, Hokkaido Cohort, Japan; COSMOS, Continuous Observation of Smoking Subjects Study; PLCO, Prostate, Lung, Colorectal and Ovarian Cancer Screening trial; NHS & HPFS, Nurses' Health Study & Health Professionals Follow-up Study; CPS II, Cancer Prevention Study II; HEXA-G, Health Examinees-Gem study

Note: One additional study (NIH-AARP) reported on SSBs and lung cancer incidence per 1 point increase in the component score of three dietary indexes (HEI-2010, AHEI-2010, and DASH), and reported a borderline positive association between lower intake of SSBs and higher risk of lung cancer in male and female[37].

Khan, 2004[85], Gnagnarella, 2013[65], You, 2022[177], Malik, 2019[109], McCullough, 2022[113], Zhu, 2023[190], Lee, 2023[97]

**Supplementary Figure 30. Highest versus the lowest category results from the individual studies on soft drinks and sugar sweetened beverages and skin cancer incidence and mortality.**

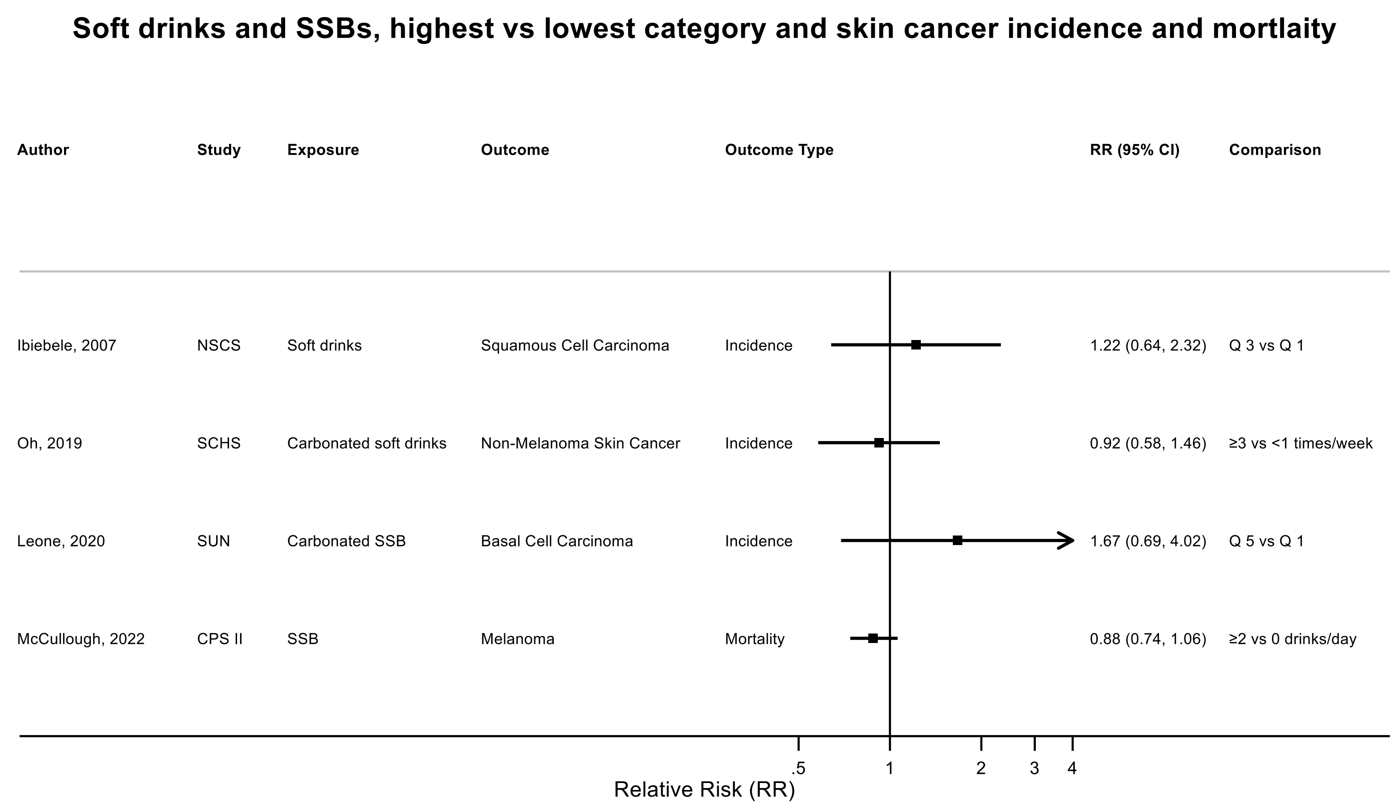

Figure legend: Forest plot shows the highest versus the lowest category results from the individual studies. Each square and the horizontal line across the square represent the RR estimate of the individual study and its 95% confidence interval (CI).

Abbreviations: CI, confidence

Study abbreviations: NSCS, Nambour Skin Cancer Study; SCHS, Singapore Chinese Health study; SUN, Seguimiento Universidad de Navarra Cohort Study; CPS II, Cancer Prevention Study II

Ibiebele, 2007[73], Oh, 2019[132], Leone, 2020[99], McCullough, 2022[113]

**Supplementary Figure 31. Highest versus the lowest category results from the individual studies on soft drinks and sugar sweetened beverages and breast cancer incidence and mortality.**

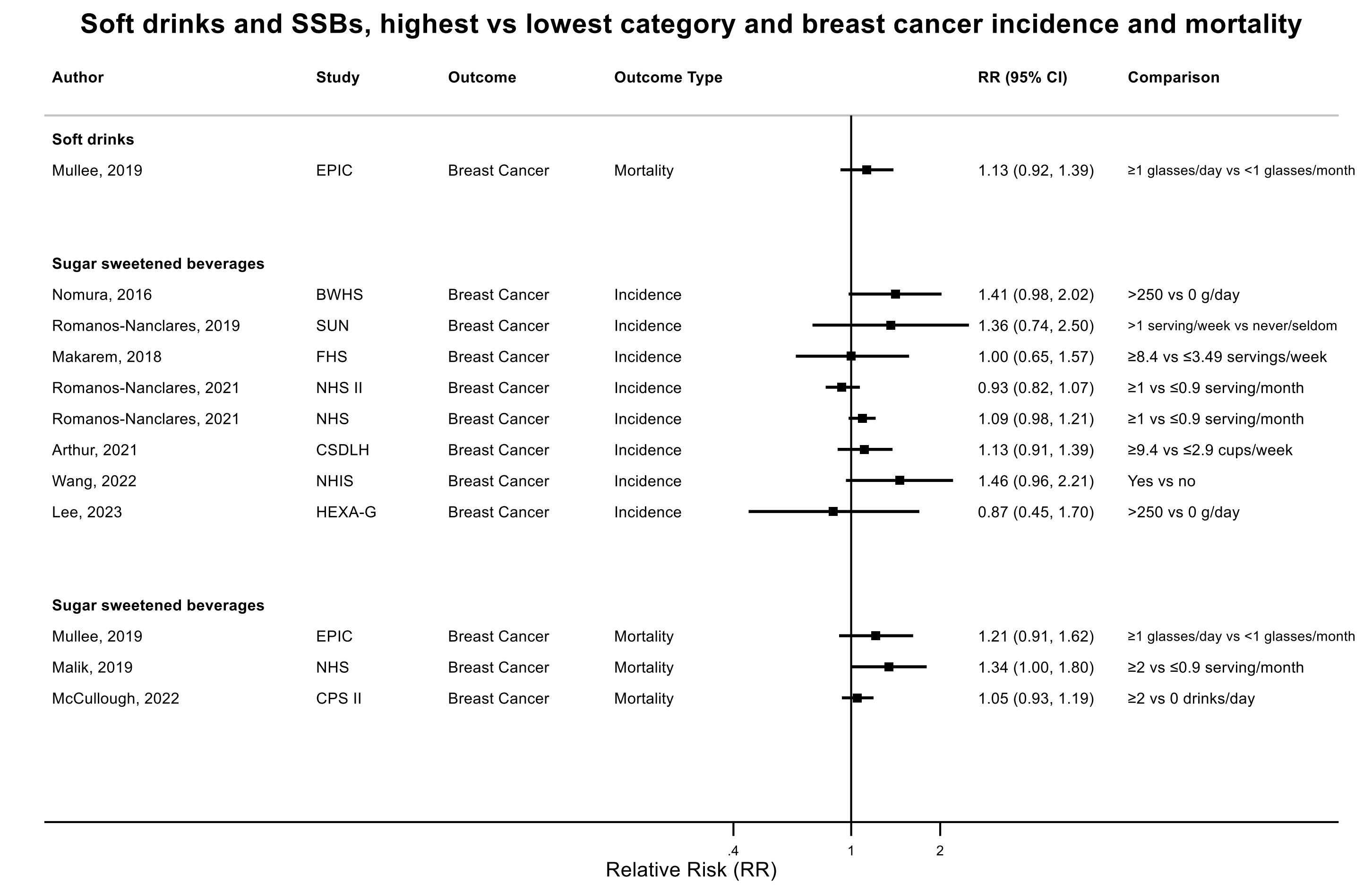

Figure legend: Forest plot shows the highest versus the lowest category results from the individual studies. Each square and the horizontal line across the square represent the RR estimate of the individual study and its 95% confidence interval (CI).

Abbreviations: CI, confidence

Study abbreviations: EPIC, European Prospective Investigation into Cancer and nutrition; BWHS, Black Women's Health Study; SUN, Seguimiento Universidad de Navarra Cohort Study; FHS-Offspring Cohort, Framingham Heart Study-Offspring Cohort; NHS II, Nurses' Health Study II; NHS, Nurses' Health Study; CSDLH, Canadian Study of Diet, Lifestyle and Health; NHIS, National Health Interview Survey; HEXA-G, Health Examinees-Gem study; CPS II, Cancer Prevention Study II

Note: One study only reported dose-response results and could not be included in the forest plot for high versus low consumption[44].

Mullee, 2019[126], Nomura, 2016[128], Romanos-Nanclares, 2019[146], Makarem, 2018[108], Romanos-Nanclares, 2021[145], Arthur, 2021[38], Wang, 2022[169], Lee, 2023[97], Mullee, 2019[126], Malik, 2019[109], McCullough, 2022[113] McCullough, 2022[113]

**Supplementary Figure 32. Highest versus the lowest category results from the individual studies on sugar sweetened beverages and breast cancer incidence by menopausal status.**

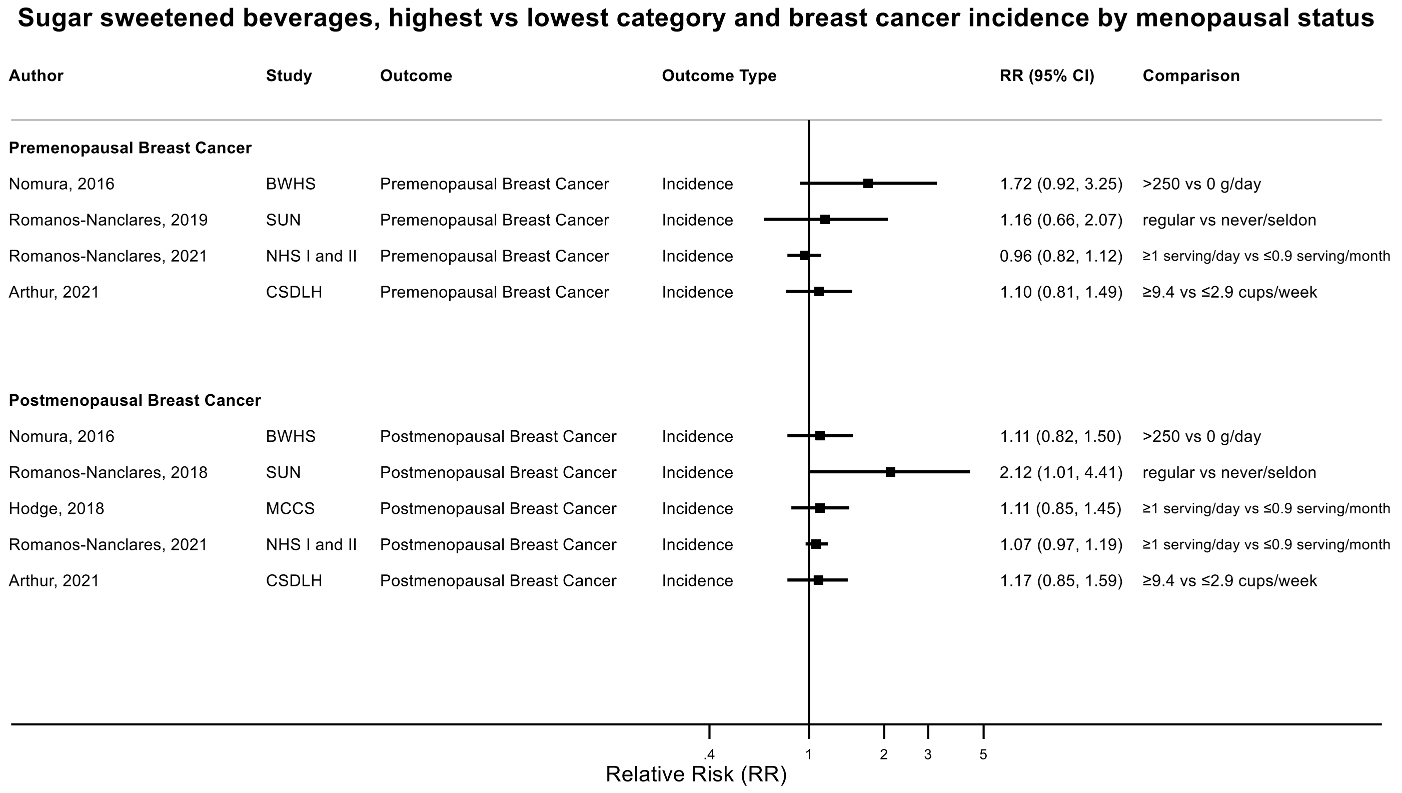

Figure legend: Forest plot shows the highest versus the lowest category results from the individual studies. Each square and the horizontal line across the square represent the RR estimate of the individual study and its 95% confidence interval (CI).

Abbreviations: CI, confidence

Study abbreviations: BWHS, Black Women's Health Study; SUN, Seguimiento Universidad de Navarra Cohort Study; NHS I and II, Nurses' Health Study I and II; CSDLH, Canadian Study of Diet, Lifestyle and Health; MCCS, Melbourne Collaborative Cohort Study

Nomura, 2016[128], Romanos-Nanclares, 2019[146], Romanos-Nanclares, 2021[145], Arthur, 2021[38], Hodge, 2018[70]

**Supplementary Figure 33. Highest versus the lowest category results from the individual studies on sugar sweetened beverages and incidence of breast cancer subsites.**

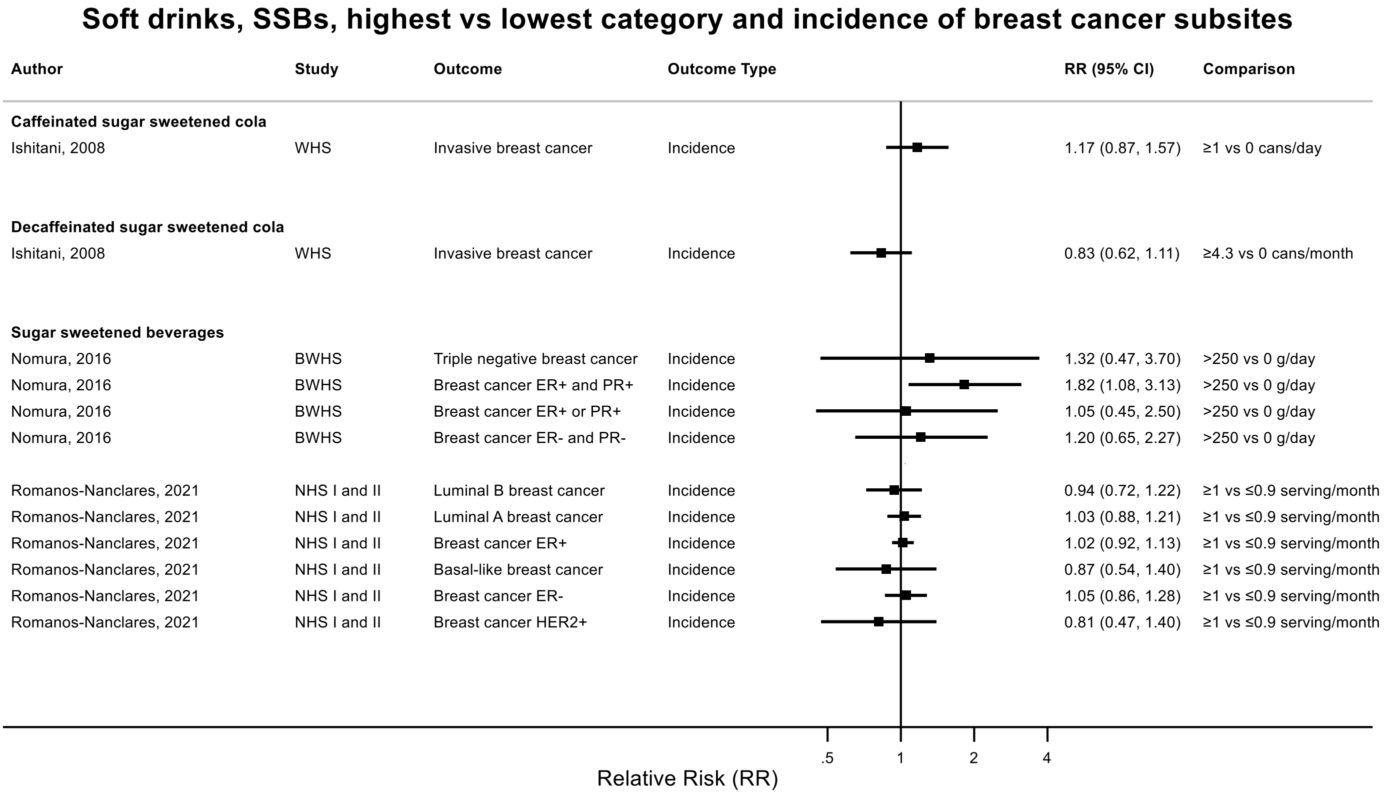

Figure legend: Forest plot shows the highest versus the lowest category results from the individual studies. Each square and the horizontal line across the square represent the RR estimate of the individual study and its 95% confidence interval (CI).

Abbreviations: CI, confidence

Study abbreviations: WHS, Women's Health Study; BWHS, Black Women's Health Study; EPIC, European Prospective Investigation into Cancer and nutrition; NHS I and II, Nurses' Health Study I and II

Ishitani, 2008[75], Nomura, 2016[128], Karavasiloglou, 2019[211], Romanos-Nanclares, 2021[145]

**Supplementary Figure 34. Highest versus the lowest category results from the individual studies on soft drinks and sugar sweetened beverages and endometrial cancer incidence and mortality.**

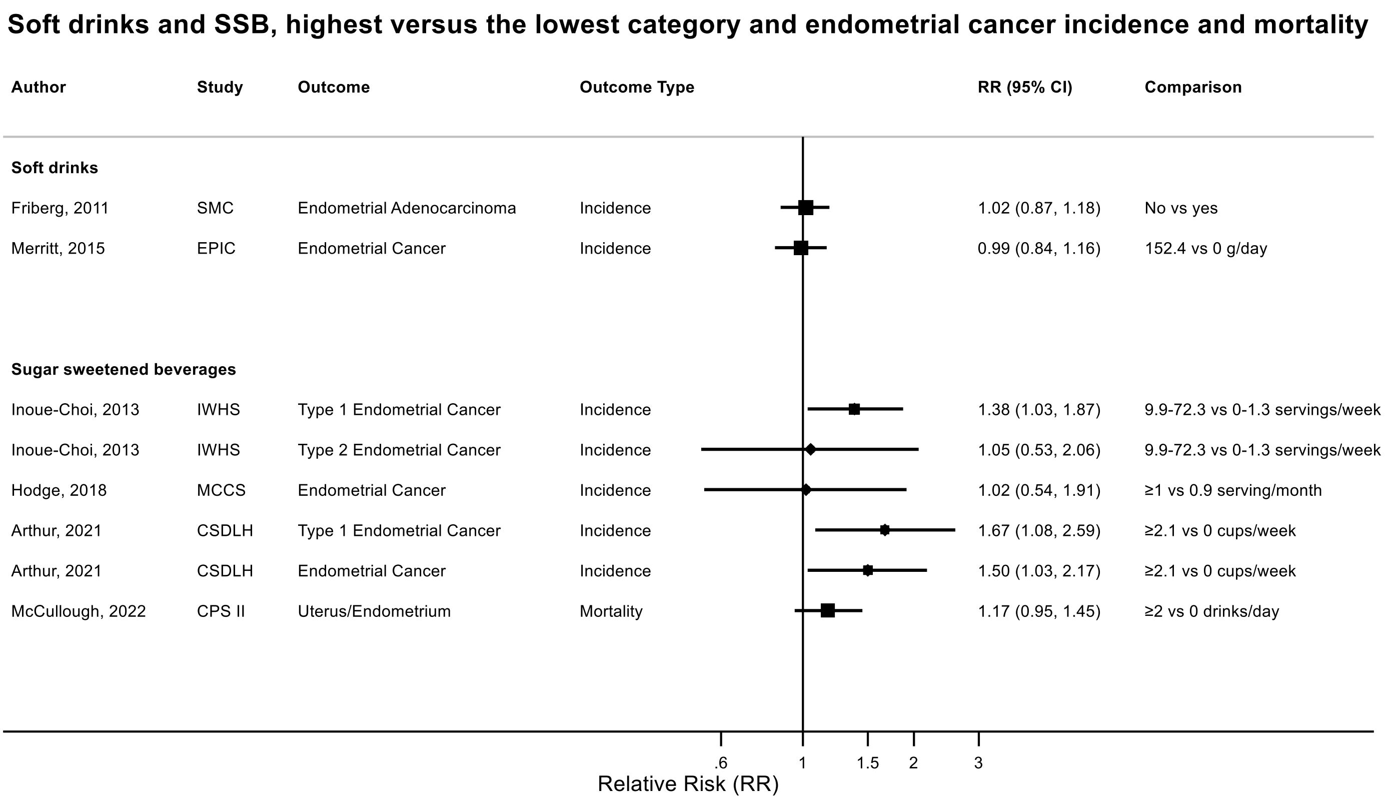

Figure legend: Forest plot shows the highest versus the lowest category results from the individual studies. Each square and the horizontal line across the square represent the RR estimate of the individual study and its 95% confidence interval (CI).

Abbreviations: CI, confidence

Study abbreviations: SMC, Swedish Mammography Cohort; EPIC, European Prospective Investigation into Cancer and nutrition; MCCS, Melbourne Collaborative Cohort Study; CSDLH, Canadian Study of Diet, Lifestyle and Health; IWHS, The Iowa Women's Health Study; CPS II, Cancer Prevention Study II

Friberg, 2011[61], Merritt, 2015[117], Inoue-Choi, 2013[74], Hodge, 2018[70], Arthur, 2021[38], McCullough, 2022[113]

**Supplementary Figure 35. Highest versus the lowest category results from the individual studies on soft drinks and sugar sweetened beverages and ovarian cancer incidence and mortality.**

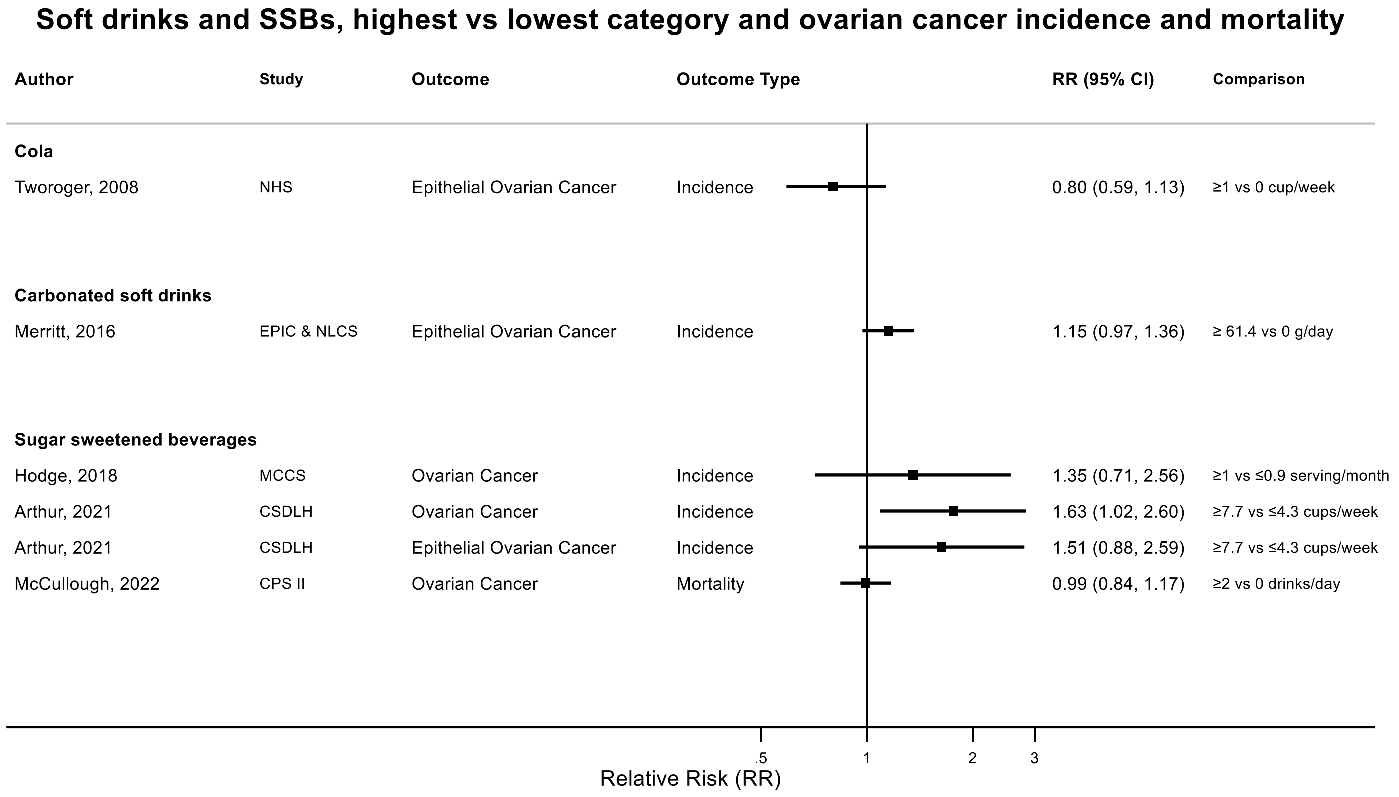

Figure legend: Forest plot shows the highest versus the lowest category results from the individual studies. Each square and the horizontal line across the square represent the RR estimate of the individual study and its 95% confidence interval (CI).

Abbreviations: CI, confidence

Study abbreviations: NHS, Nurses' Health Study; EPIC & NLCS, European Prospective Investigation into Cancer and nutrition & Netherlands Cohort Study on Diet and Cancer; MCCS, Melbourne Collaborative Cohort Study; CSDLH, Canadian Study of Diet, Lifestyle and Health; CPS II, Cancer Prevention Study II

Tworoger, 2008[163], Merritt, 2016[118], Hodge, 2018[70], Arthur, 2021[38], McCullough, 2022[113]

**Supplementary Figure 36. Highest versus the lowest category results from the individual studies on soft drinks and sugar sweetened beverages and prostate cancer incidence and mortality.**

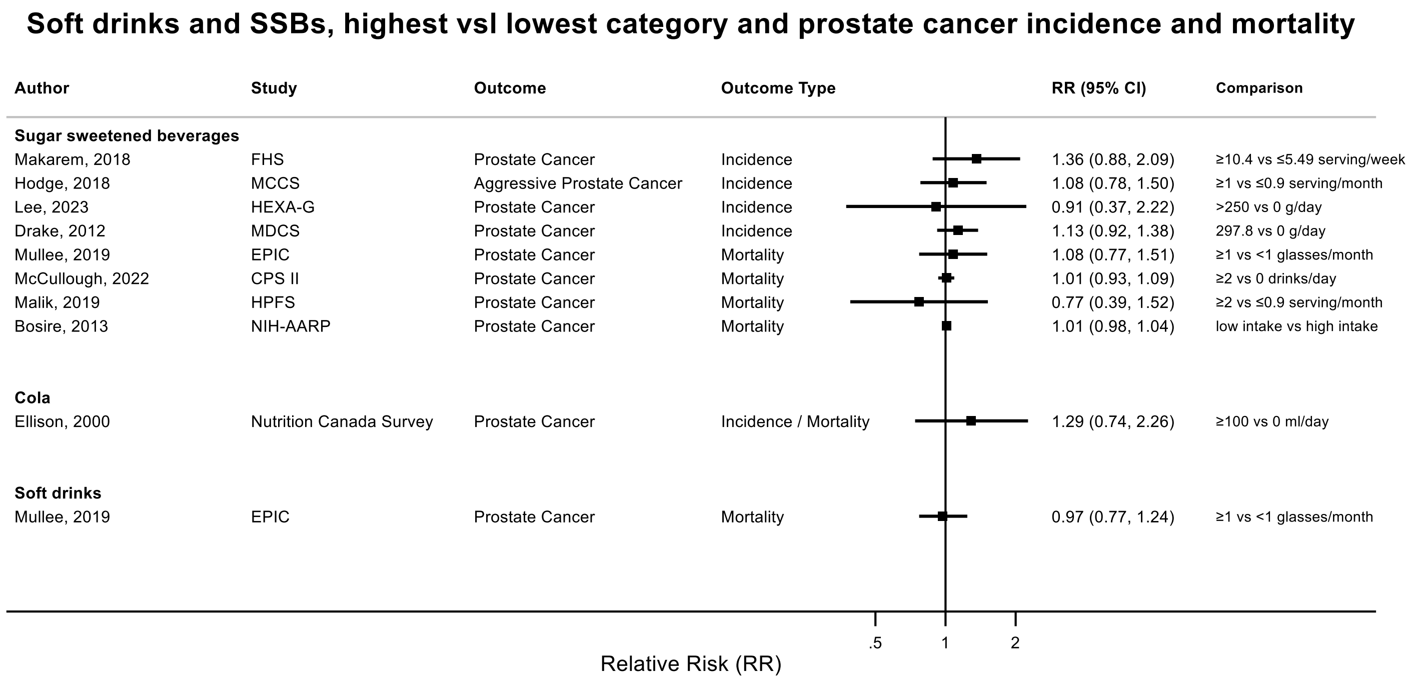

Figure legend: Forest plot shows the highest versus the lowest category results from the individual studies. Each square and the horizontal line across the square represent the RR estimate of the individual study and its 95% confidence interval (CI).

Abbreviations: CI, confidence

Study abbreviations: FHS-Offspring Cohort, Framingham Heart Study-Offspring Cohort; MCCS, Melbourne Collaborative Cohort Study; HEXA-G, Health Examinees-Gem study; MDCS, Malmö Diet and Cancer Study; EPIC, European Prospective Investigation into Cancer and nutrition; CPS II, Cancer Prevention Study II; HPFS, Health Professionals Follow-up Study; NIH-AARP, National Institute of Health + American Association of Retired Persons Diet and Health Study

Makarem, 2018[108], Hodge, 2018[70], Lee, 2023[97], Drake, 2012[47], Mullee, 2019[126], McCullough, 2022[113], Malik, 2019[109], Bosire, 2013[42], Ellison, 2000[50]

**Supplementary Figure 37. Highest versus the lowest category results from the individual studies on soft drinks and sugar sweetened beverages and bladder cancer incidence and mortality.**

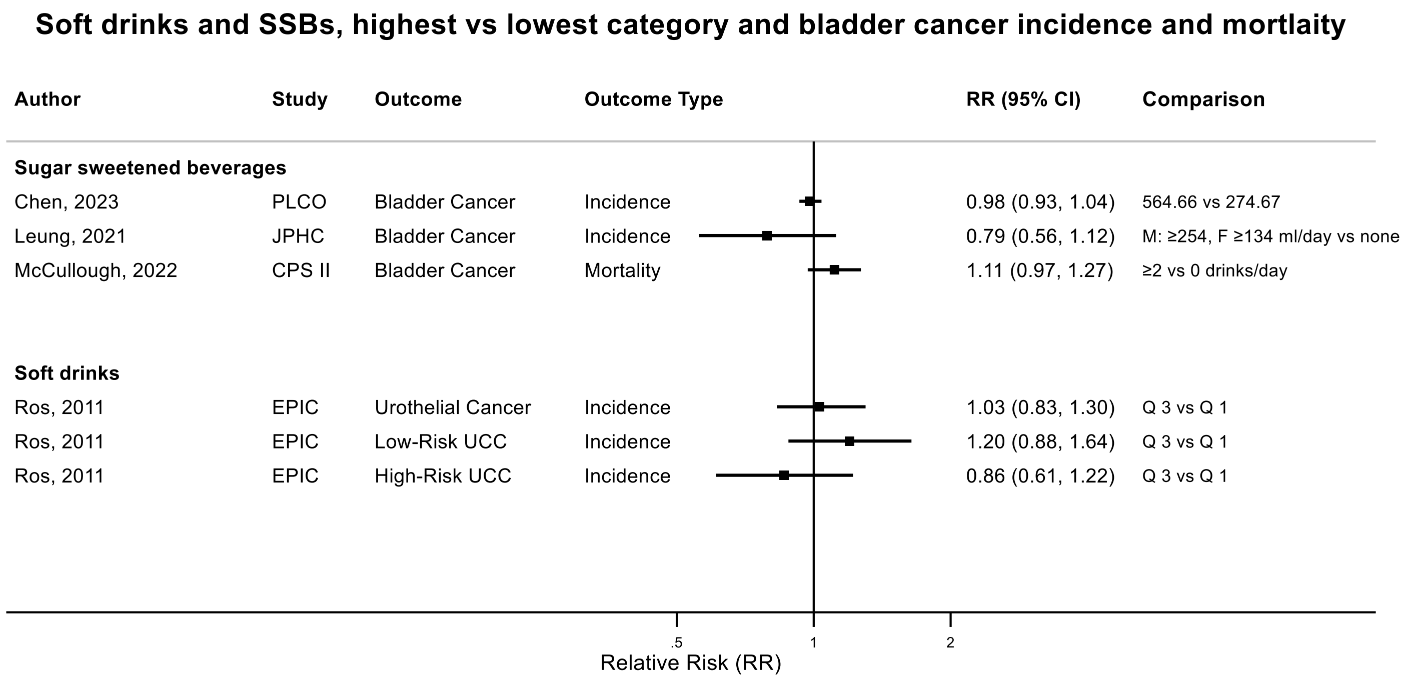

Figure legend: Forest plot shows the highest versus the lowest category results from the individual studies. Each square and the horizontal line across the square represent the RR estimate of the individual study and its 95% confidence interval (CI).

Abbreviations: CI, confidence

Study abbreviations: PLCO, Prostate, Lung, Colorectal and Ovarian Cancer Screening trial; JPHC, Japan Public Health Center-Based Prospective Study; CPS II, Cancer Prevention Study II; EPIC, European Prospective Investigation into Cancer and nutrition

Note: One additional study (MEC) that could not be included in the categorical high vs low meta-analysis because of insufficient data format reported little evidence of an association with bladder cancer risk for each 2 point increase in the Alternative Healthy Eating Index – 2010 components score (lower sugar sweetened beverages and fruit juice consumption) in men (HR 1.03, 95% CI:0.99-1.07) and women (HR 0.99, 95% CI:0.93-1.05)[82].

Chen, 2023[46], Leung, 2021[100], McCullough, 2022[113], Ros, 2011[147]

**Supplementary Figure 38. Highest versus the lowest category results from the individual studies on soft drinks and sugar sweetened beverages and kidney cancer incidence and mortality.**

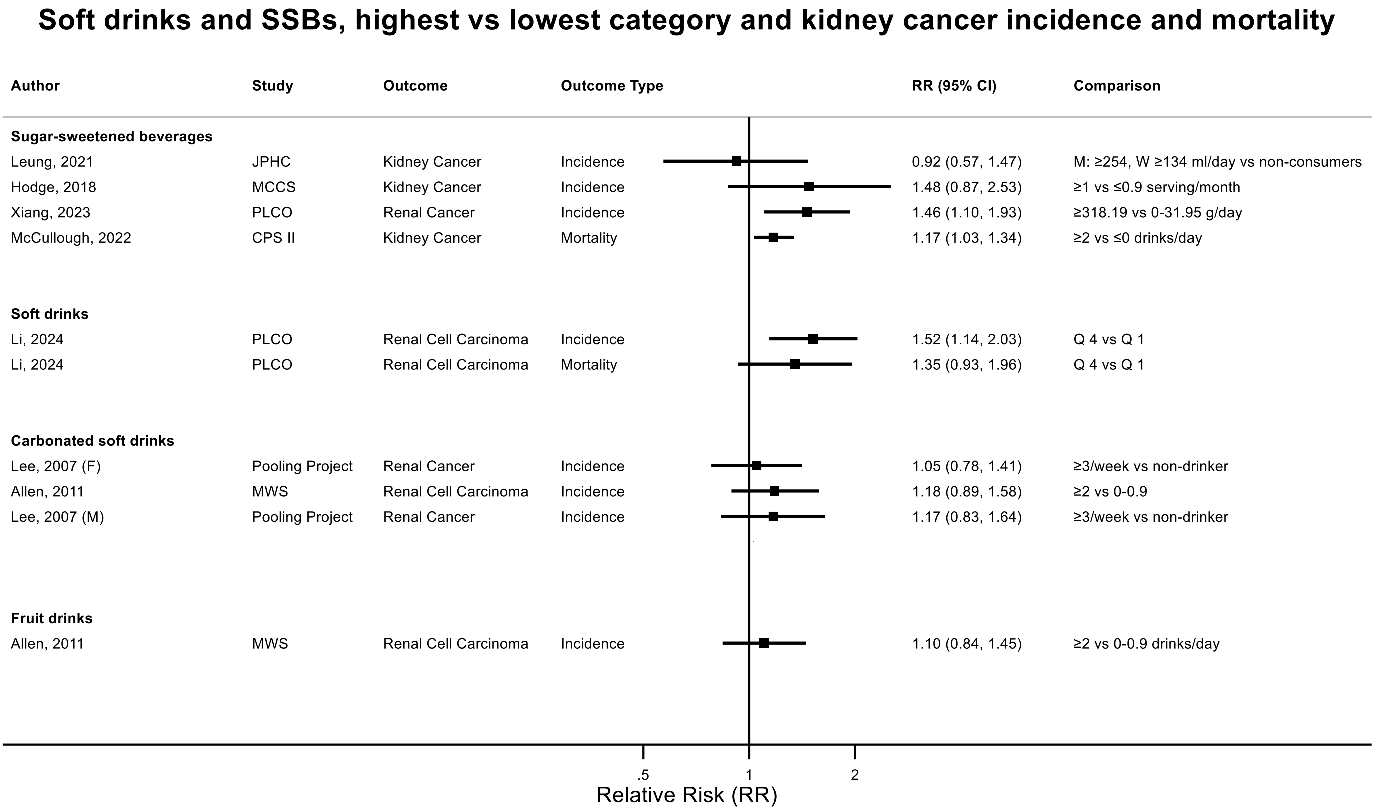

| Author | Study | Outcome | Outcome Type | RR (95% CI) | Comparison |
|---|---|---|---|---|---|
| **Sugar-sweetened beverages** | | | | | |
| Leung, 2021 | JPHC | Kidney Cancer | Incidence | 0.92 (0.57, 1.47) | M: ≥254, W ≥134 ml/day vs non-consumers |
| Hodge, 2018 | MCCS | Kidney Cancer | Incidence | 1.48 (0.87, 2.53) | ≥1 vs ≤0.9 serving/month |
| Xiang, 2023 | PLCO | Renal Cancer | Incidence | 1.46 (1.10, 1.93) | ≥318.19 vs 0-31.95 g/day |
| McCullough, 2022 | CPS II | Kidney Cancer | Mortality | 1.17 (1.03, 1.34) | ≥2 vs ≤0 drinks/day |
| **Soft drinks** | | | | | |
| Li, 2024 | PLCO | Renal Cell Carcinoma | Incidence | 1.52 (1.14, 2.03) | Q 4 vs Q 1 |
| Li, 2024 | PLCO | Renal Cell Carcinoma | Mortality | 1.35 (0.93, 1.96) | Q 4 vs Q 1 |
| **Carbonated soft drinks** | | | | | |
| Lee, 2007 (F) | Pooling Project | Renal Cancer | Incidence | 1.05 (0.78, 1.41) | ≥3/week vs non-drinker |
| Allen, 2011 | MWS | Renal Cell Carcinoma | Incidence | 1.18 (0.89, 1.58) | ≥2 vs 0-0.9 |
| Lee, 2007 (M) | Pooling Project | Renal Cancer | Incidence | 1.17 (0.83, 1.64) | ≥3/week vs non-drinker |
| **Fruit drinks** | | | | | |
| Allen, 2011 | MWS | Renal Cell Carcinoma | Incidence | 1.10 (0.84, 1.45) | ≥2 vs 0-0.9 drinks/day |

Figure legend: Forest plot shows the highest versus the lowest category results from the individual studies. Each square and the horizontal line across the square represent the RR estimate of the individual study and its 95% confidence interval (CI).

Abbreviations: CI, confidence

Study abbreviations: JPHC, Japan Public Health Center-Based Prospective Study; MCCS, Melbourne Collaborative Cohort Study; PLCO, Prostate, Lung, Colorectal and Ovarian Cancer Screening trial; CPS II, Cancer Prevention Study II; Pooling Project, The Pooling Project of Prospective Studies of Diet and Cancer; MWS, Million Women Study

Leung, 2021[100], Hodge, 2018[70], Xiang, 2023[173], McCullough, 2022[113], Li, 2024[102], Lee, 2007[26], Allen, 2011[36]

**Supplementary Figure 39. Highest versus the lowest category results from the individual studies on soft drinks and sugar sweetened beverages and brain cancer incidence and mortality.**

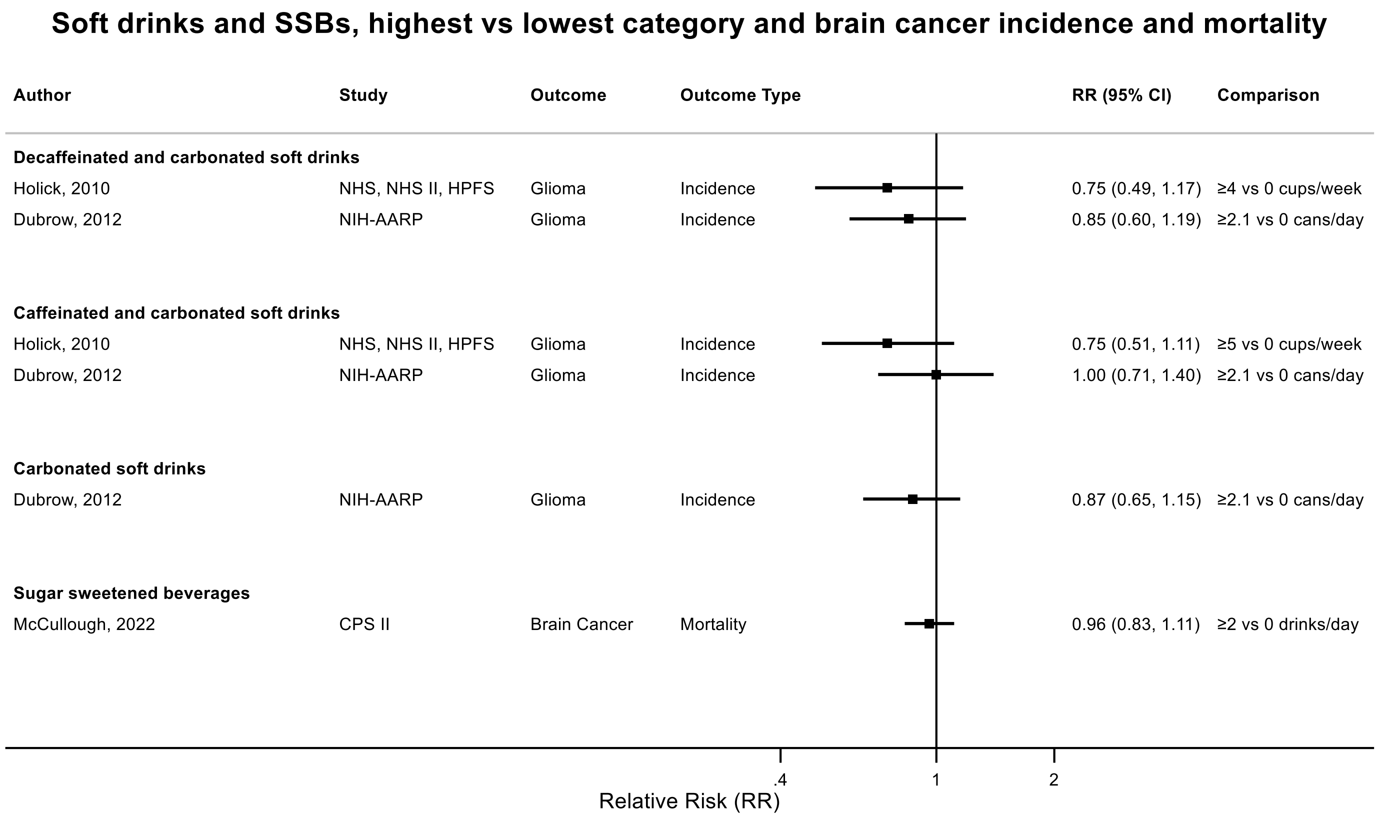

Figure legend: Forest plot shows the highest versus the lowest category results from the individual studies. Each square and the horizontal line across the square represent the RR estimate of the individual study and its 95% confidence interval (CI).

Abbreviations: CI, confidence

Study abbreviations: NHS, NHS II, HPFS Nurses' Health Study, Nurses' Health Study II, Health Professionals Follow-up Study; NIH-AARP, National Institute of Health + American Association of Retired Persons Diet and Health Study; CPS II, Cancer Prevention Study II

Holick, 2010[71], Dubrow, 2012[48], McCullough, 2022[113]

**Supplementary Figure 40. Highest versus the lowest category results from the individual studies on soft drinks and sugar sweetened beverages and thyroid cancer incidence.**

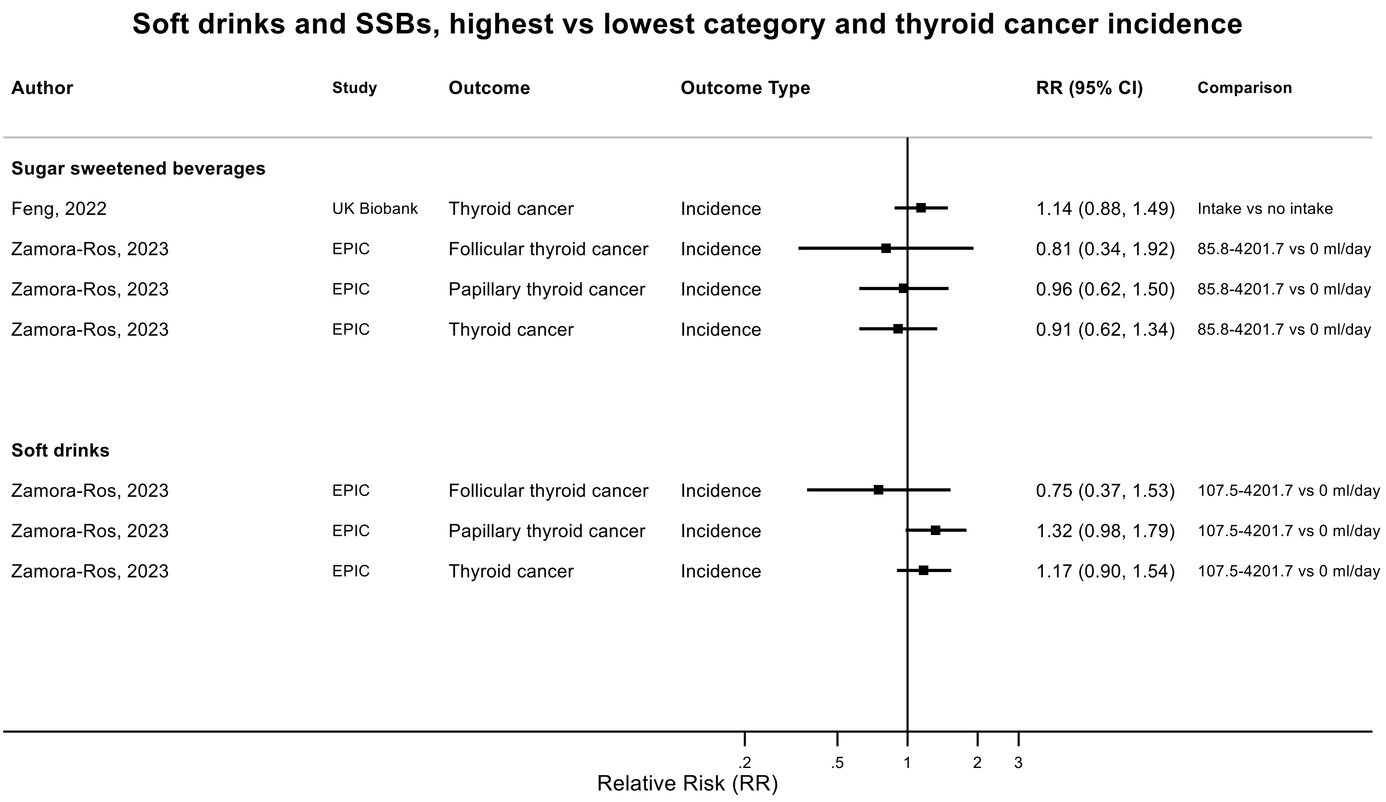

Figure legend: Forest plot shows the highest versus the lowest category results from the individual studies. Each square and the horizontal line across the square represent the RR estimate of the individual study and its 95% confidence interval (CI).

Abbreviations: CI, confidence

Study abbreviations: UK Biobank, United Kingdom Biobank Study; EPIC, European Prospective Investigation into Cancer and nutrition

Feng, 2022[53], Zamora-Ros, 2023[180]

**Supplementary Figure 41. Highest versus the lowest category results from the individual studies on sugar sweetened beverages and lymphoid and haematological cancers incidence and mortality.**

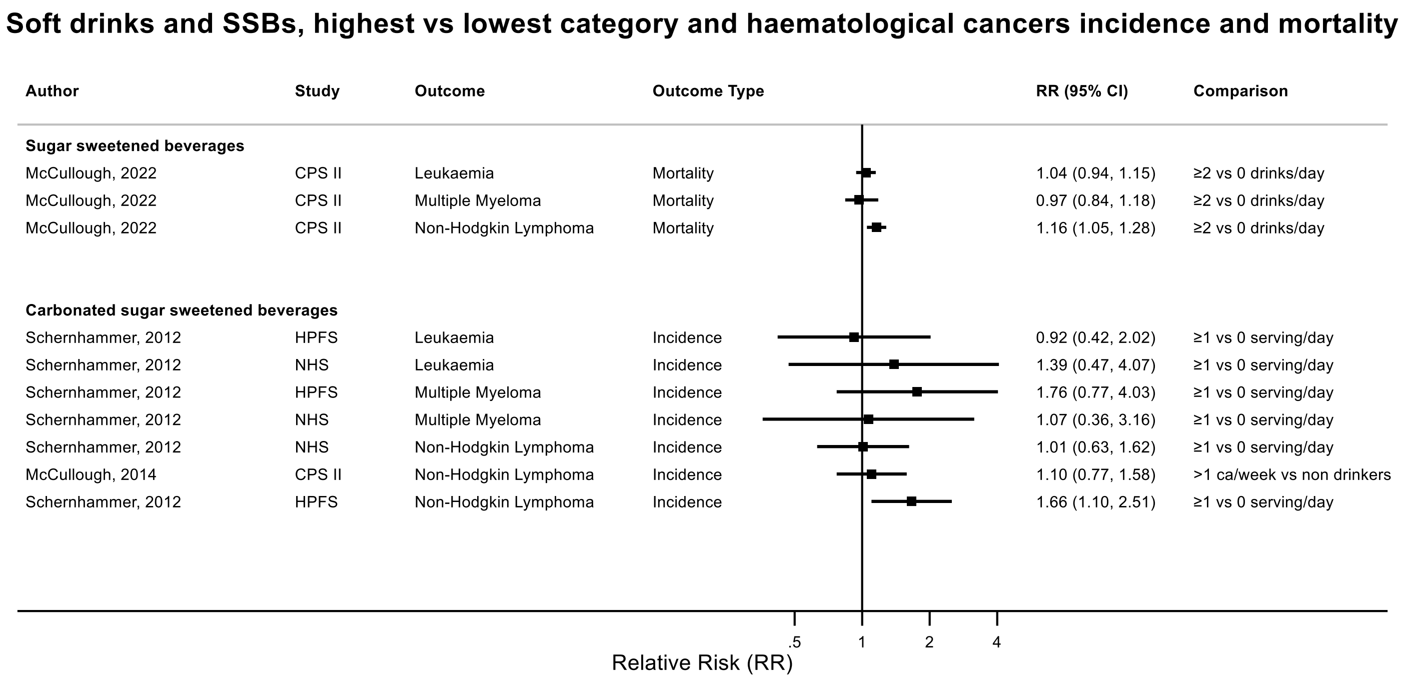

Figure legend: Forest plot shows the highest versus the lowest category results from the individual studies. Each square and the horizontal line across the square represent the RR estimate of the individual study and its 95% confidence interval (CI).

Abbreviations: CI, confidence

Study abbreviations: CPS II, Cancer Prevention Study II; HPFS, Health Professionals Follow-up Study; NHS, Nurses' Health Study

McCullough, 2022[113], Schernhammer, 2012[150]

**Supplementary Figure 42. Highest versus the lowest category results from individual studies on artificially sweetened beverages and stomach cancer incidence and mortality.**

**Artificially sweetened beverages, highest vs lowest category and stomach cancer incidence and mortality**

| Author | Study | Outcome | Outcome Type | RR (95% CI) | Comparison |
|---|---|---|---|---|---|
| Hodge, 2018 | MCCS | Gastric Cardia Adenocarcinoma | Incidence | 1.03 (0.53, 1.98) | ≥1 serving/day vs ≤0.9 serving/month |
| McCullough, 2022 | CPS II | Stomach Cancer | Mortality | 0.92 (0.78, 1.08) | ≥2 vs 0 drinks/day |

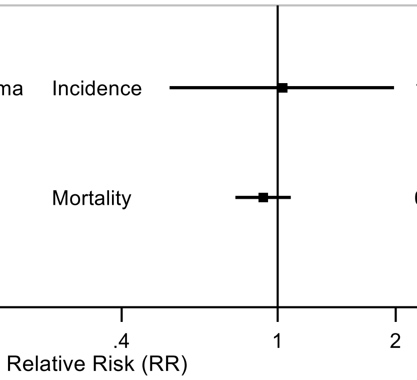

Figure legend: Forest plot shows the highest versus the lowest category results from the individual studies. Each square and the horizontal line across the square represent the RR estimate of the individual study and its 95% confidence interval (CI).

Abbreviations: CI, confidence interval; RR, relative risk.

Study abbreviations: MCCS, Melbourne Collaborative Cohort Study; CPS II, Cancer Prevention Study II

Hodge, 2018[70], McCullough, 2022[113]

**Supplementary Figure 43. Highest versus the lowest category results from individual studies on artificially sweetened beverages and colorectal cancer incidence and mortality.**

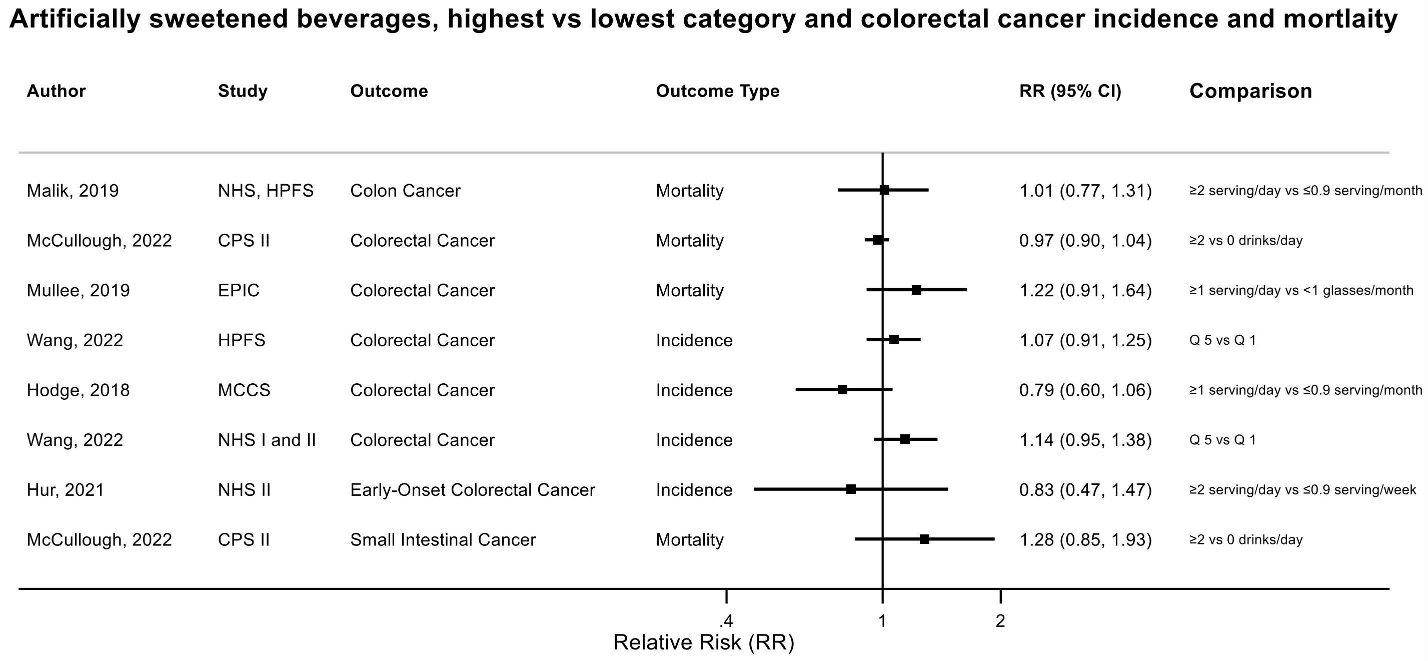

Figure legend: Forest plot shows the highest versus the lowest category results from the individual studies. Each square and the horizontal line across the square represent the RR estimate of the individual study and its 95% confidence interval (CI).

Abbreviations: CI, confidence interval; RR, relative risk.

Study abbreviations: NHS, HPFS, Nurses' Health Study, Health Professionals Follow-up Study; CPS II, Cancer Prevention Study II; EPIC, European Prospective Investigation into Cancer and nutrition; HPFS, Health Professionals Follow-up Study; MCCS, Melbourne Collaborative Cohort Study; NHS I and II, Nurses' Health Study I and II

Malik, 2019[109], McCullough, 2022[113], Mullee, 2019[126], Wang, 2022[168], Hodge, 2018[70], Hur, 2021[72]

**Supplementary Figure 44. Highest versus the lowest category results from individual studies on artificially sweetened beverages and liver cancer incidence and mortality.**

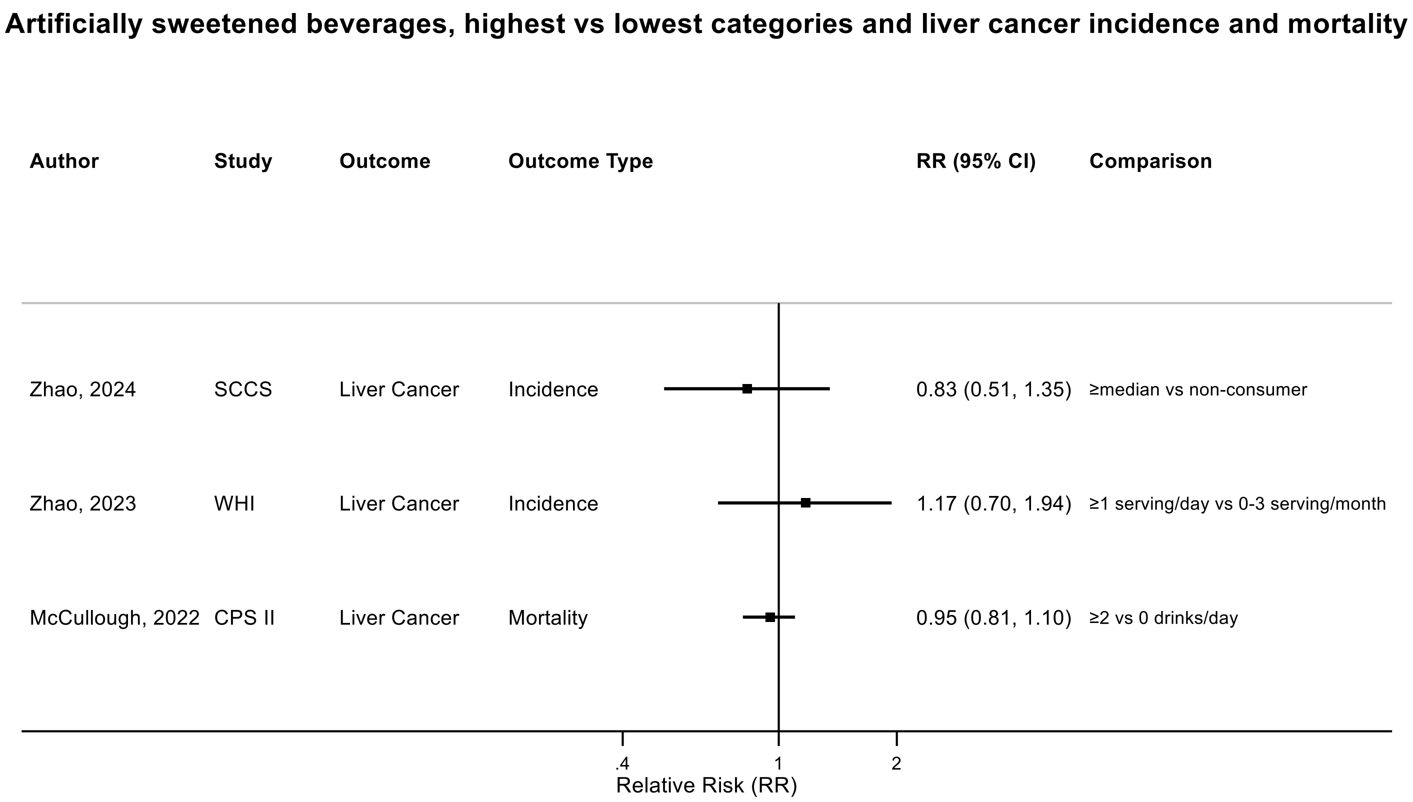

Figure legend: Forest plot shows the highest versus the lowest category results from the individual studies. Each square and the horizontal line across the square represent the RR estimate of the individual study and its 95% confidence interval (CI).

Abbreviations: CI, confidence interval; RR, relative risk.

Study abbreviations: SCCS, Southern Community Cohort Study; WHI, Women’s Health Initiative Study; CPS II, Cancer Prevention Study II

Zhao, 2024[187], Zhao, 2023[186], McCullough, 2022[113]

**Supplementary Figure 45. Highest versus the lowest category results from individual studies on artificially sweetened beverages and pancreatic cancer incidence and mortality.**

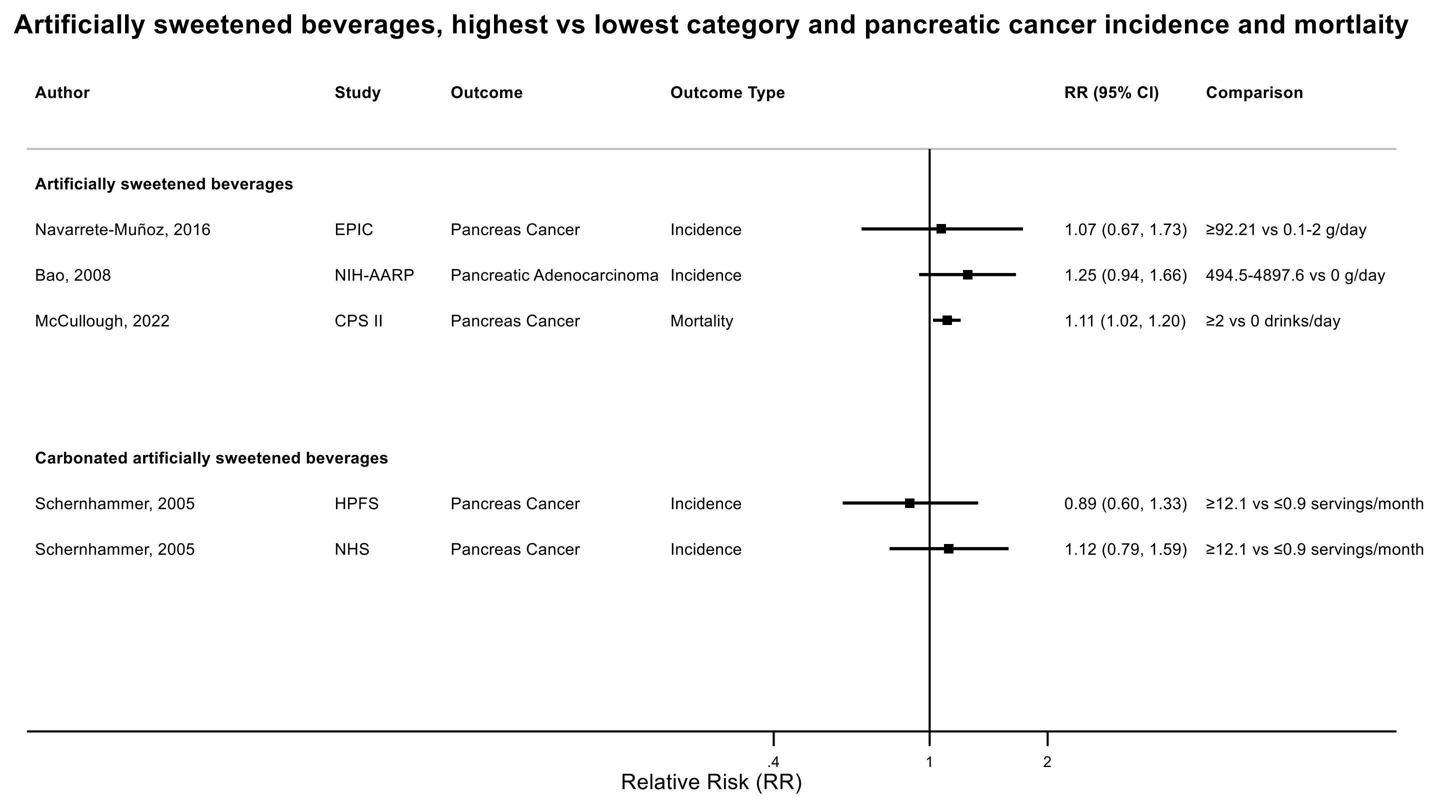

Figure legend: Forest plot shows the highest versus the lowest category results from the individual studies. Each square and the horizontal line across the square represent the RR estimate of the individual study and its 95% confidence interval (CI).

Abbreviations: CI, confidence interval; RR, relative risk.

Study abbreviations: EPIC, European Prospective Investigation into Cancer and nutrition; NIH-AARP, National Institute of Health + American Association of Retired Persons Diet and Health Study; CPS II, Cancer Prevention Study II; HPFS, Health Professionals Follow-up Study; NHS, Nurses' Health Study

Navarrete-Muñoz, 2016[127], Bao, 2008[39], McCullough, 2022[113], Schernhammer, 2005[151]

**Supplementary Figure 46. Highest versus the lowest category results from individual studies on artificially sweetened beverages and lung cancer incidence and mortality.**

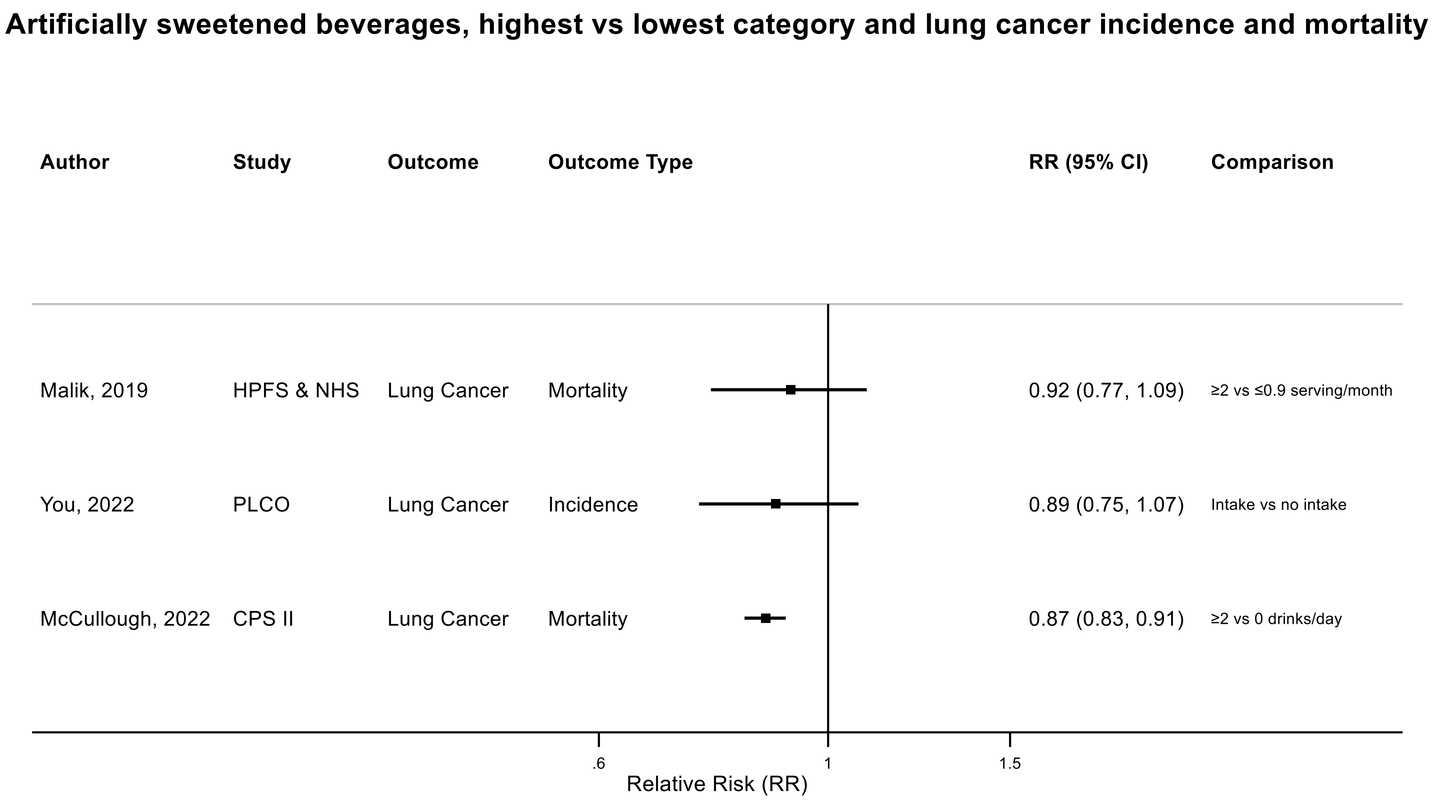

Figure legend: Forest plot shows the highest versus the lowest category results from the individual studies. Each square and the horizontal line across the square represent the RR estimate of the individual study and its 95% confidence interval (CI).

Abbreviations: CI, confidence interval; RR, relative risk.

Study abbreviations: HPFS & NHS, Health Professionals Follow-up Study & Nurses' Health Study; PLCO, Prostate, Lung, Colorectal and Ovarian Cancer Screening trial; CPS II, Cancer Prevention Study II

Malik, 2019[109], You, 2022[177], McCullough, 2022[113]

**Supplementary Figure 47. Highest versus the lowest category results from individual studies on artificially sweetened beverages and breast cancer incidence and mortality.**

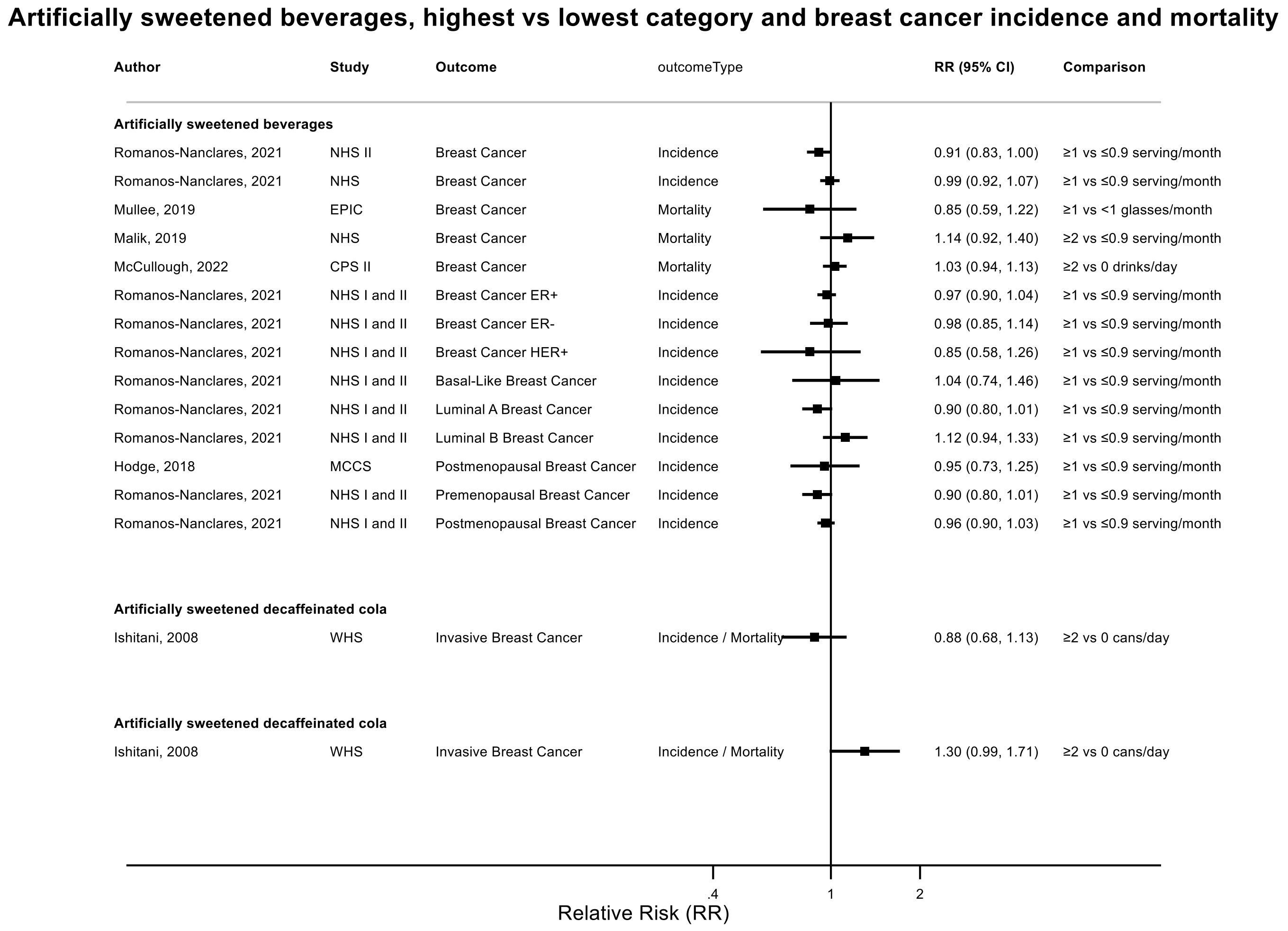

Figure legend: Forest plot shows the highest versus the lowest category results from the individual studies. Each square and the horizontal line across the square represent the RR estimate of the individual study and its 95% confidence interval (CI).

Abbreviations: CI, confidence interval; RR, relative risk.

Study abbreviations: NHS II, Nurses' Health Study II; NHS, Nurses' Health Study; EPIC, European Prospective Investigation into Cancer and nutrition; CPS II, Cancer Prevention Study II; NHS I and II, Nurses' Health Study I and II; MCCS, Melbourne Collaborative Cohort Study; WHS, Women's Health Study

Romanos-Nanclares, 2021[145], Mullee, 2019[126], Malik, 2019[109], McCullough, 2022[113], Hodge, 2018[70], Ishitani, 2008[75]

**Supplementary Figure 48. Highest versus the lowest category results from individual studies on artificially sweetened beverages and endometrial cancer incidence and mortality.**

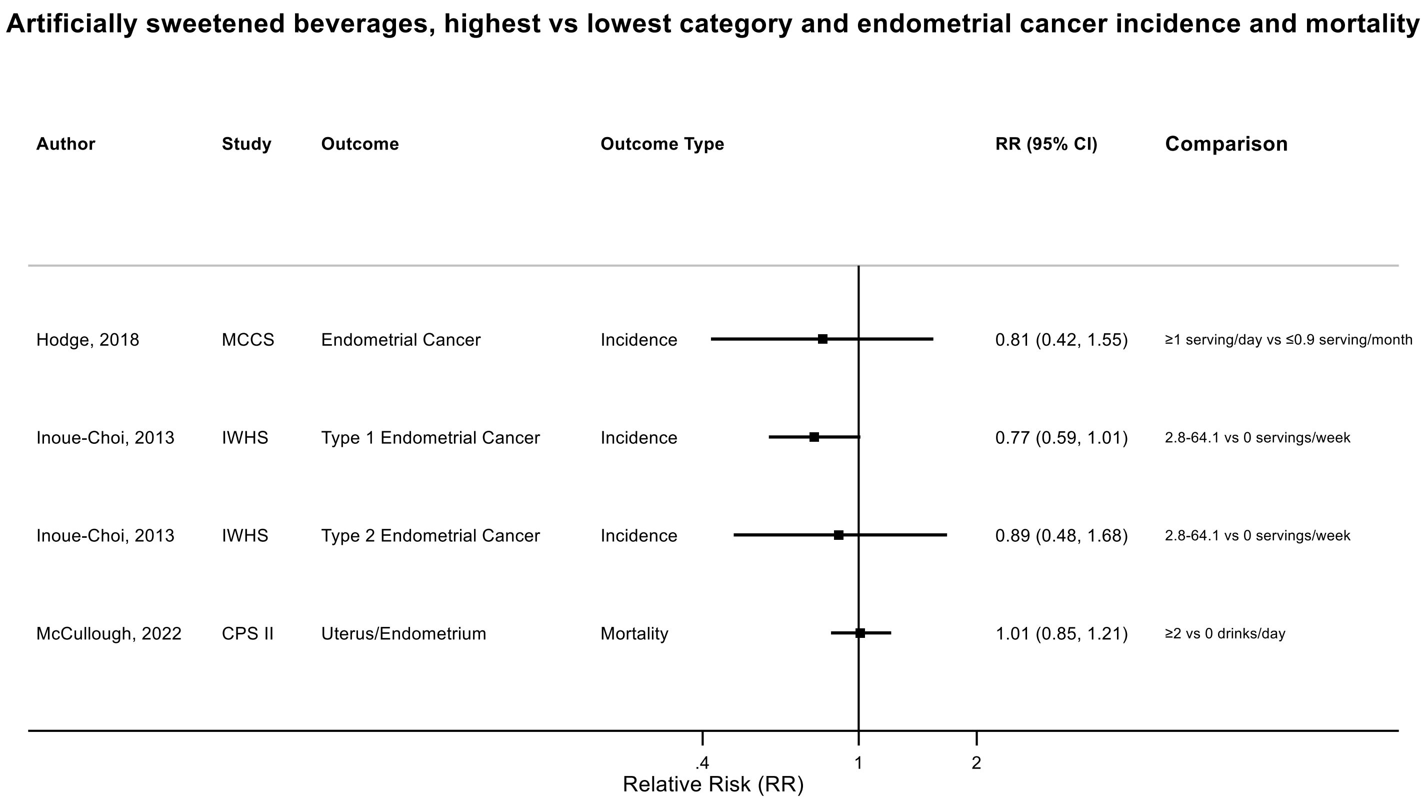

Figure legend: Forest plot shows the highest versus the lowest category results from the individual studies. Each square and the horizontal line across the square represent the RR estimate of the individual study and its 95% confidence interval (CI).

Abbreviations: CI, confidence interval; RR, relative risk.

Study abbreviations: MCCS, Melbourne Collaborative Cohort Study; IWHS, The Iowa Women's Health Study; CPS II, Cancer Prevention Study II

Hodge, 2018[70], Inoue-Choi, 2013[74], McCullough, 2022[113]

**Supplementary Figure 49. Highest versus the lowest category results from individual studies on artificially sweetened beverages and ovarian cancer incidence and mortality.**

##### Artificially sweetened beverages, highest vs lowest category and ovarian cancer incidence and mortality

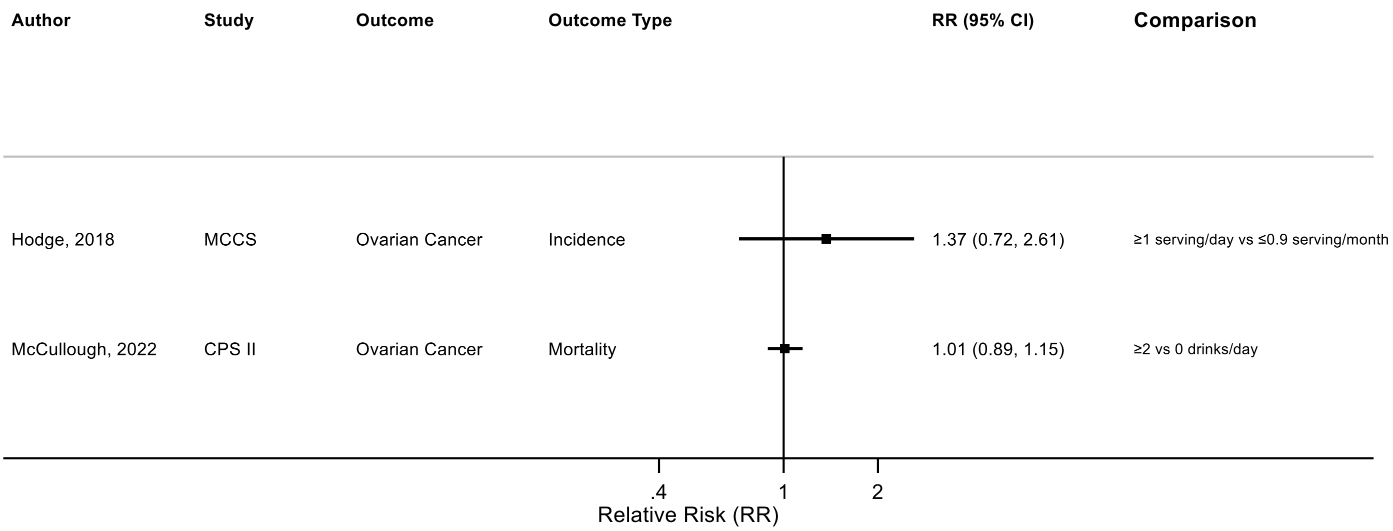

Figure legend: Forest plot shows the highest versus the lowest category results from the individual studies. Each square and the horizontal line across the square represent the RR estimate of the individual study and its 95% confidence interval (CI).

Abbreviations: CI, confidence interval; RR, relative risk.

Study abbreviations: MCCS, Melbourne Collaborative Cohort Study; CPS II, Cancer Prevention Study II

Hodge, 2018[70], McCullough, 2022[113]

**Supplementary Figure 50. Highest versus the lowest category results from individual studies on artificially sweetened beverages and prostate cancer incidence and mortality.**

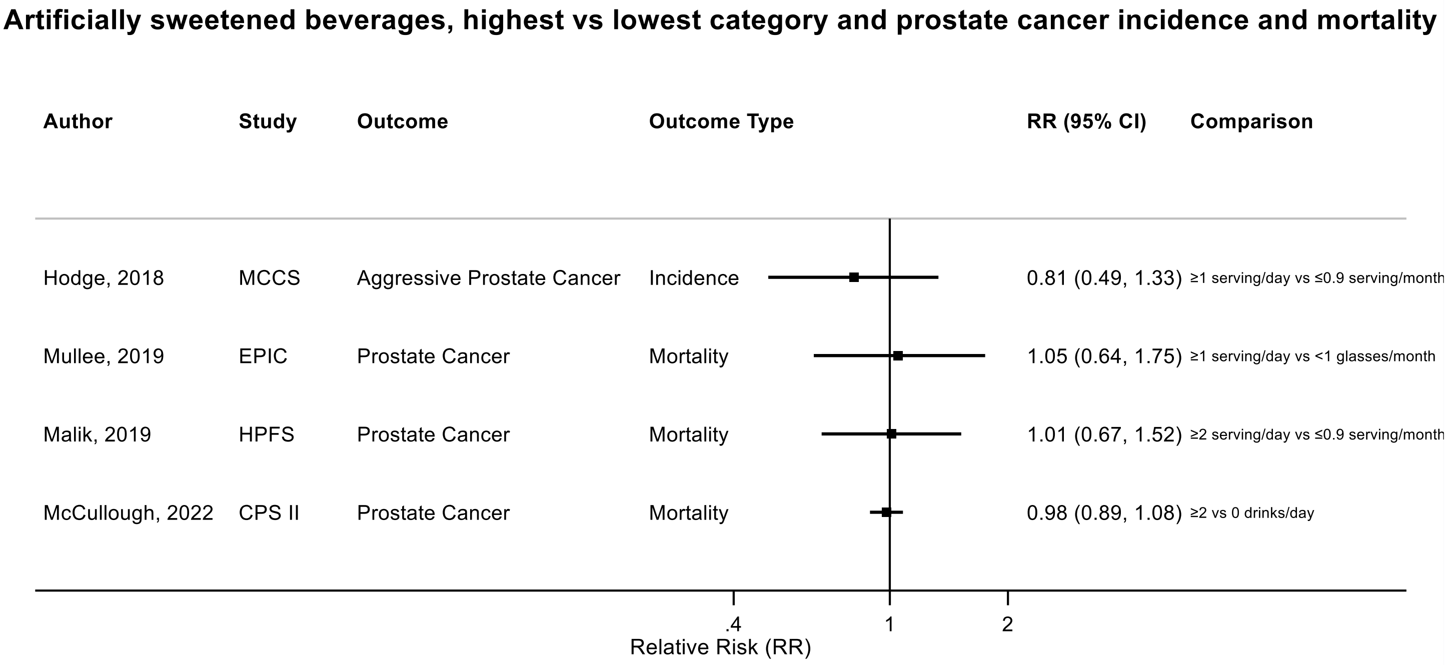

Figure legend: Forest plot shows the highest versus the lowest category results from the individual studies. Each square and the horizontal line across the square represent the RR estimate of the individual study and its 95% confidence interval (CI).

Abbreviations: CI, confidence interval; RR, relative risk.

Study abbreviations: MCCS, Melbourne Collaborative Cohort Study; EPIC, European Prospective Investigation into Cancer and nutrition; HPFS, Health Professionals Follow-up Study; CPS II, Cancer Prevention Study II

Note: There was one Frech cohort study (NutriNet-Santé)[44] that that could not be included in the dose-response meta-analysis because of implausible effect size and reported an inverse association but with 95%CI crossing the null value between ASBs and prostate cancer incidence (RR: 0.58, 95%CI: 0.28-1.18) per 10 mL/day increase.

Hodge, 2018[70], Mullee, 2019[126], Malik, 2019[109], McCullough, 2022[113]

**Supplementary Figure 51. Highest versus the lowest category results from individual studies on artificially sweetened beverages and kidney cancer incidence and mortality.**

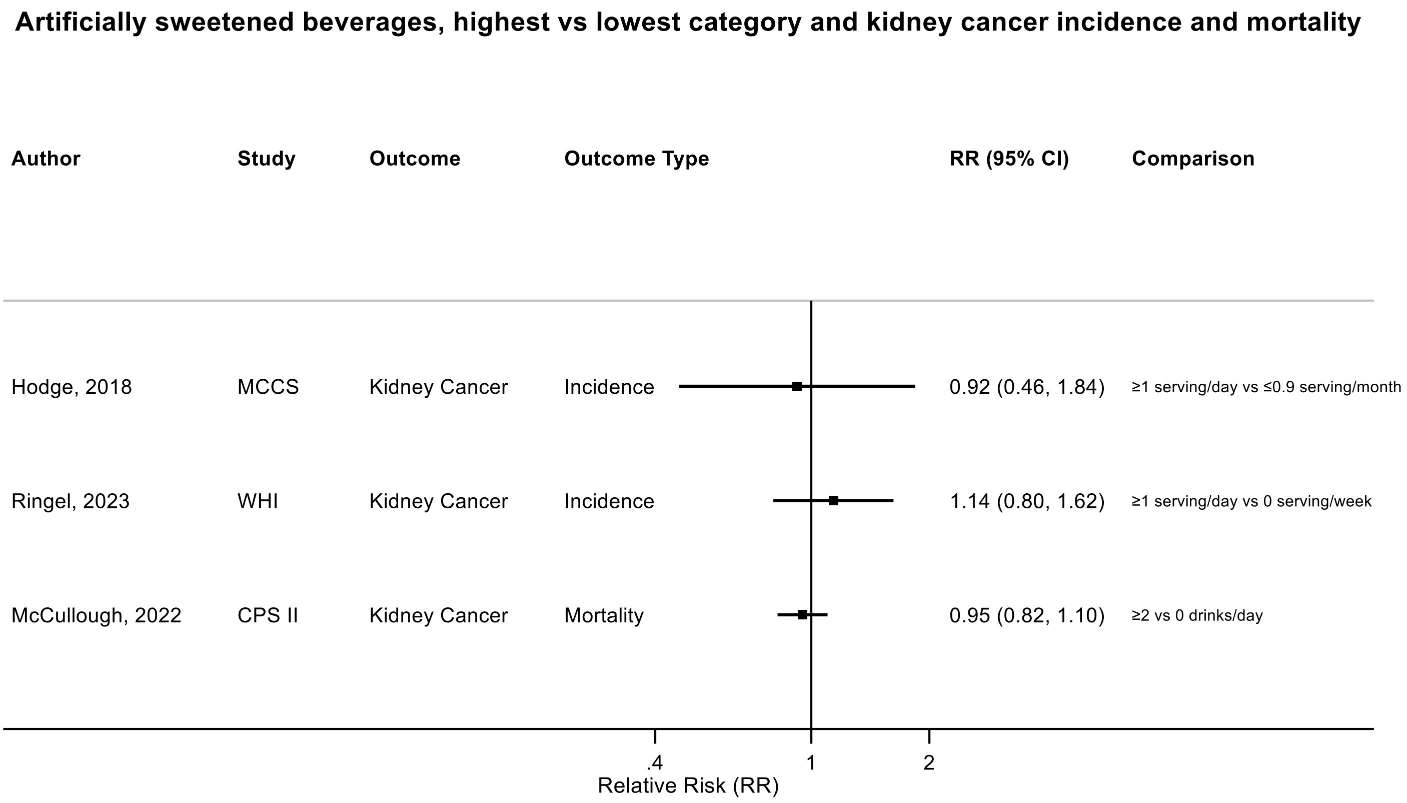

Figure legend: Forest plot shows the highest versus the lowest category results from the individual studies. Each square and the horizontal line across the square represent the RR estimate of the individual study and its 95% confidence interval (CI).

Abbreviations: CI, confidence interval; RR, relative risk.

Study abbreviations: MCCS, Melbourne Collaborative Cohort Study; WHI, Women's Health Initiative Study; CPS II, Cancer Prevention Study II

Hodge, 2018[70], Ringel, 2023[144], McCullough, 2022[113]

**Supplementary Figure 52. Highest versus the lowest category results from individual studies on artificially sweetened beverages and bladder cancer incidence and mortality.**

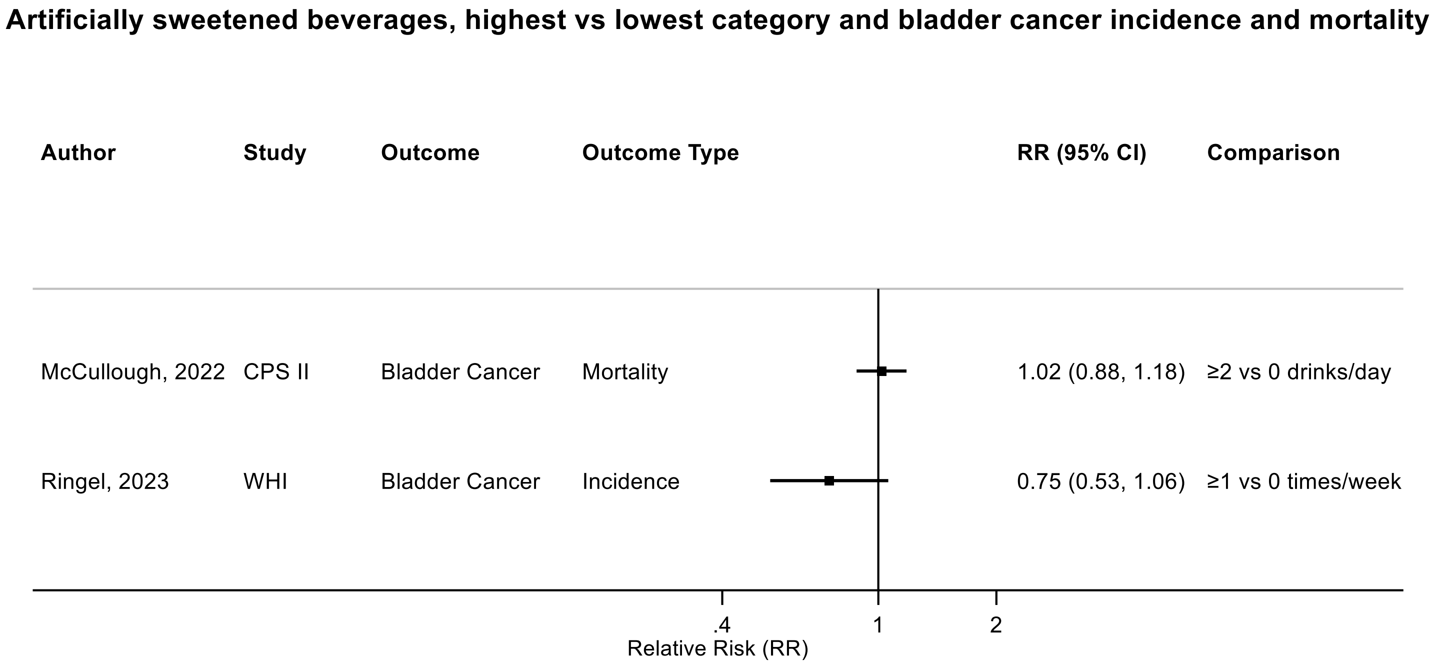

Figure legend: Forest plot shows the highest versus the lowest category results from the individual studies. Each square and the horizontal line across the square represent the RR estimate of the individual study and its 95% confidence interval (CI).

Abbreviations: CI, confidence interval; RR, relative risk.

Study abbreviations: CPS II, Cancer Prevention Study II; WHI, Women’s Health Initiative Study

McCullough, 2022[113], Ringel, 2023[144]

**Supplementary Figure 53. Highest versus the lowest category results from individual studies on artificially sweetened beverages and hematological cancers incidence and mortality.**

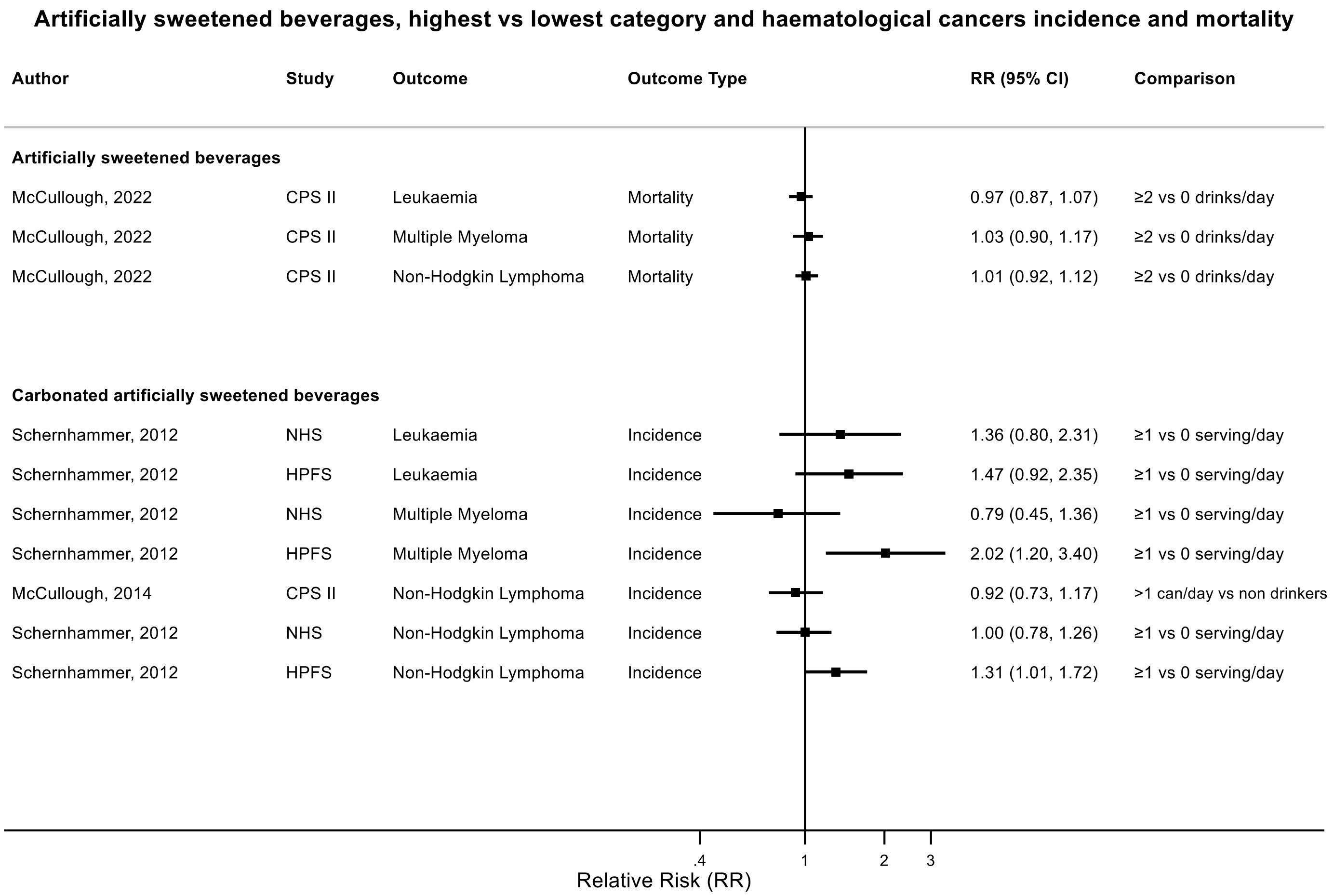

Figure legend: Forest plot shows the highest versus the lowest category results from the individual studies. Each square and the horizontal line across the square represent the RR estimate of the individual study and its 95% confidence interval (CI).

Abbreviations: CI, confidence interval; RR, relative risk.

Study abbreviations: CPS II, Cancer Prevention Study II; NHS, Nurses' Health Study; HPFS, Health Professionals Follow-up Study

McCullough, 2022[113], Schernhammer, 2012[150], McCullough, 2014[114]

**Supplementary Figure 54. Highest versus the lowest category results from individual studies on artificially sweetened beverages and thyroid cancer incidence.**

**Artificially sweetened beverages, highest vs low the lowest categories and thyroid cancer incidence**

| Author | Study | Outcome | Outcome Type | RR (95% CI) | Comparison |
|---|---|---|---|---|---|
| Zamora-Ros, 2023 | EPIC | Thyroid Cancer | Incidence | 1.16 (0.80, 1.69) | 43-3389.5 vs ≤0 ml/day |
| Zamora-Ros, 2023 | EPIC | Papillary Thyroid Cancer | Incidence | 1.51 (1.04, 2.19) | 43-3389.5 vs ≤0 ml/day |

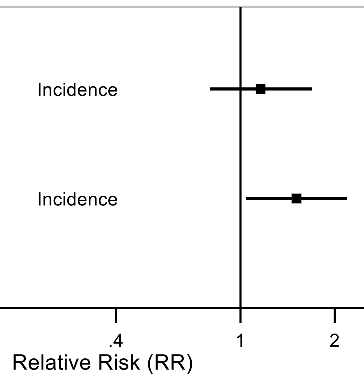

Figure legend: Forest plot shows the highest versus the lowest category results from the individual studies. Each square and the horizontal line across the square represent the RR estimate of the individual study and its 95% confidence interval (CI).

Abbreviations: CI, confidence interval; RR, relative risk.

Study abbreviations: EPIC, European Prospective Investigation into Cancer and nutrition

Zamora-Ros, 2023[180]

**Supplementary Figure 55. Highest versus the lowest category results from individual studies on juices and of head and neck cancers incidence.**

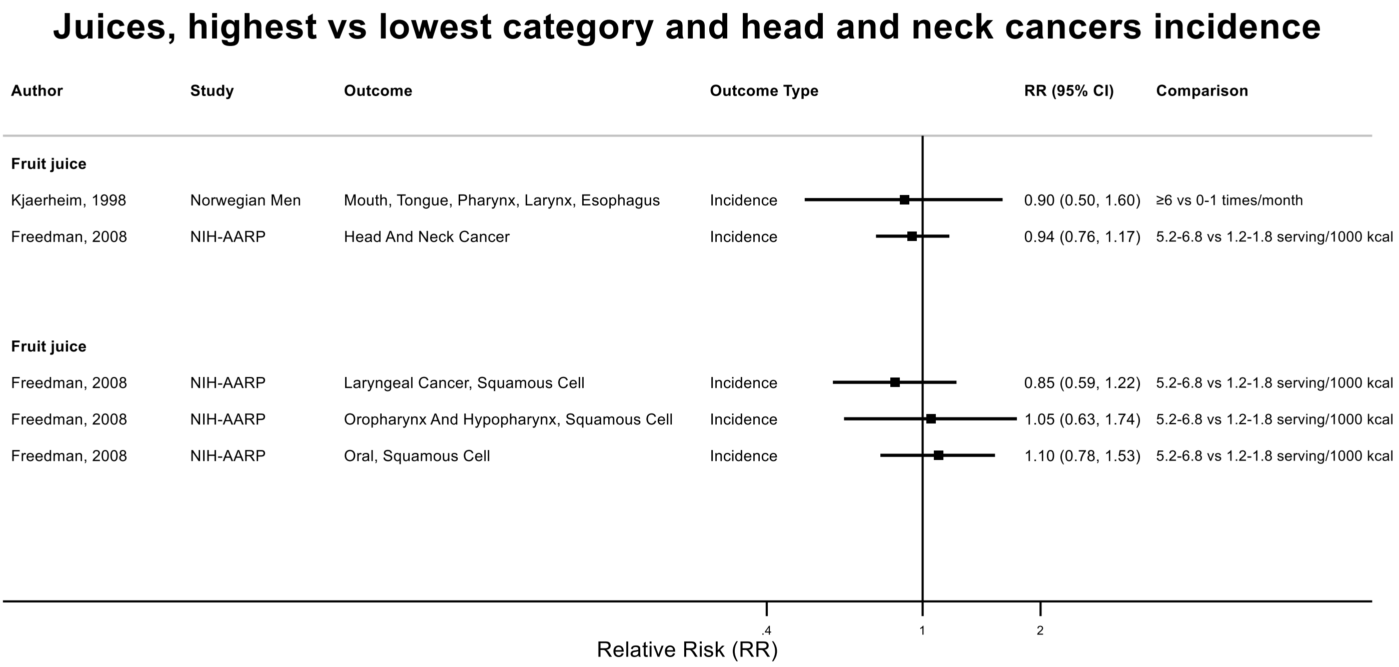

Figure legend: Forest plot shows the highest versus the lowest category results from the individual studies. Each square and the horizontal line across the square represent the RR estimate of the individual study and its 95% confidence interval (CI).

Abbreviations: CI, confidence interval; RR, relative risk.

Study abbreviations: NIH-AARP, National Institute of Health + American Association of Retired Persons Diet and Health Study

Kjaerheim, 1998[90], Freedman, 2008[59]

**Supplementary Figure 56. Highest versus the lowest category results from individual studies on juices and colorectal cancer incidence and mortality.**

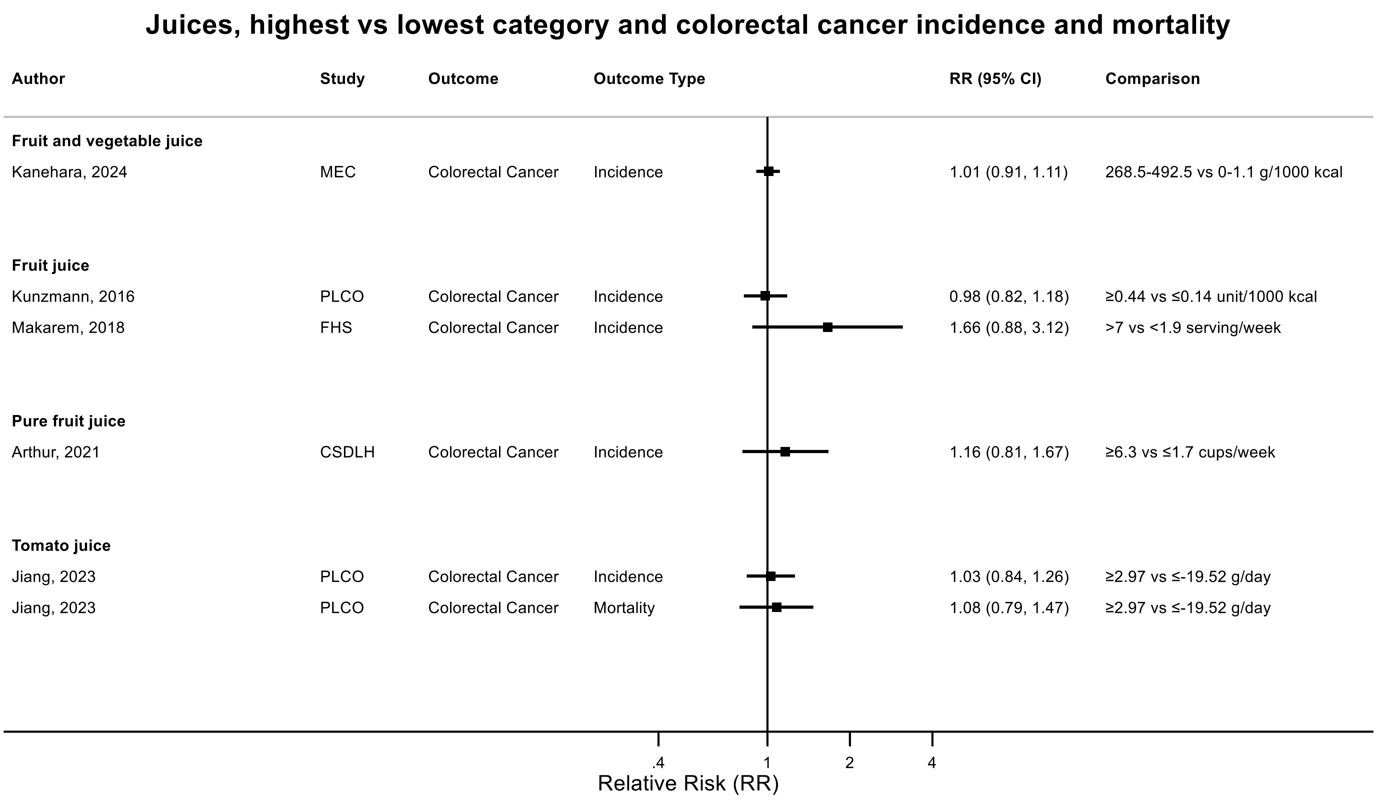

Figure legend: Forest plot shows the highest versus the lowest category results from the individual studies. Each square and the horizontal line across the square represent the RR estimate of the individual study and its 95% confidence interval (CI).

Abbreviations: CI, confidence interval; RR, relative risk.

Study abbreviations: MEC, Multiethnic Cohort Study; PLCO, Prostate, Lung, Colorectal and Ovarian Cancer Screening trial; FHS, Framingham Heart Study; CSDLH, Canadian Study of Diet, Lifestyle and Health

Kanehara, 2024[81], Kunzmann, 2016[93], Makarem, 2018[108], Arthur, 2021[38], Jiang, 2023[77]

**Supplementary Figure 57. Highest versus the lowest category results from individual studies on juices and colorectal cancer subsites incidence and mortality.**

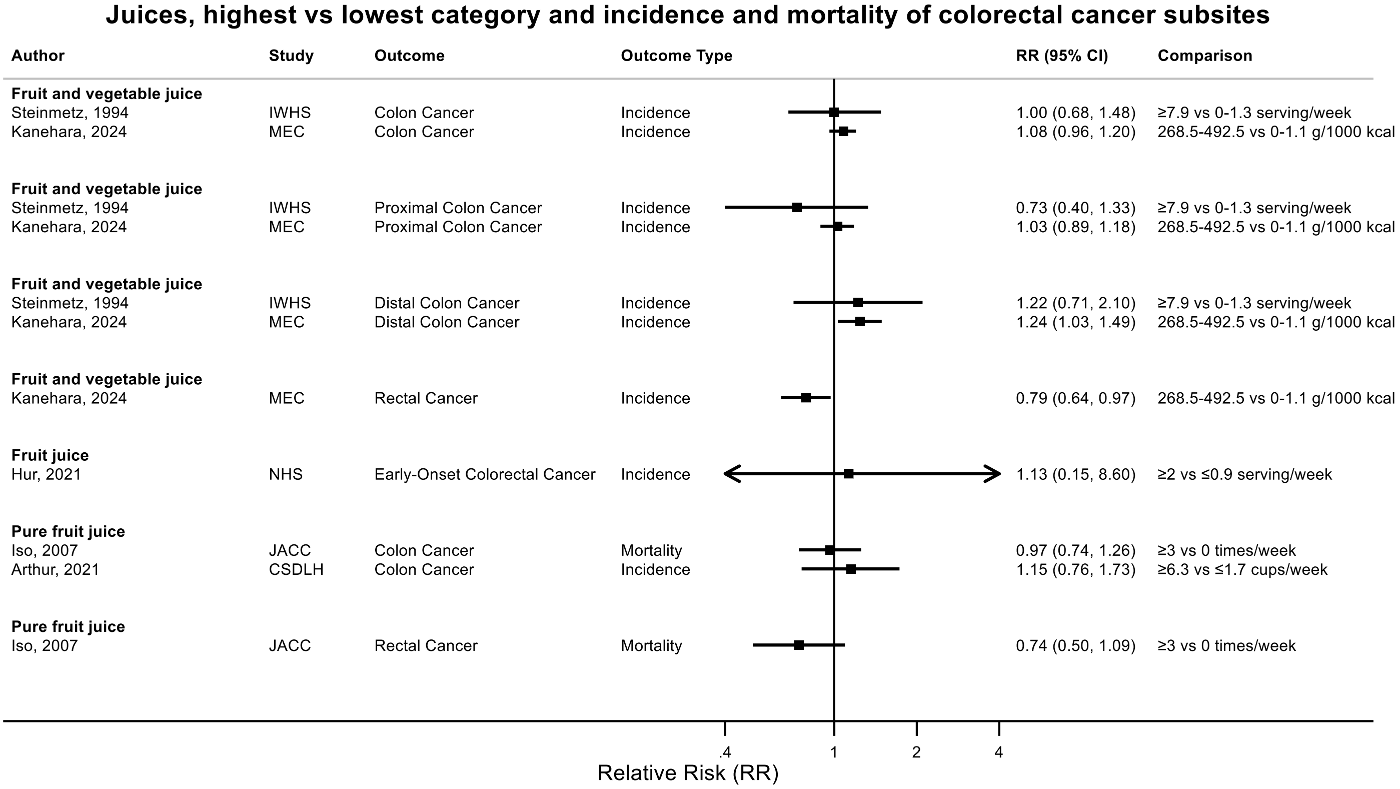

Figure legend: Forest plot shows the highest versus the lowest category results from the individual studies. Each square and the horizontal line across the square represent the RR estimate of the individual study and its 95% confidence interval (CI).

Abbreviations: CI, confidence interval; RR, relative risk.

Study abbreviations: IWHS, The Iowa Women's Health Study; MEC, Multiethnic Cohort Study; NHS, Nurses' Health Study; JACC, Japan Collaborative Cohort study; CSDLH, Canadian Study of Diet, Lifestyle and Health

Steinmetz, 1994[158], Kanehara, 2024[81], Hur, 2021[72], Iso, 2007[76], Arthur, 2021[38]

**Supplementary Figure 58. Highest versus the lowest category results from individual studies on juices and pancreatic cancer incidence and mortality.**

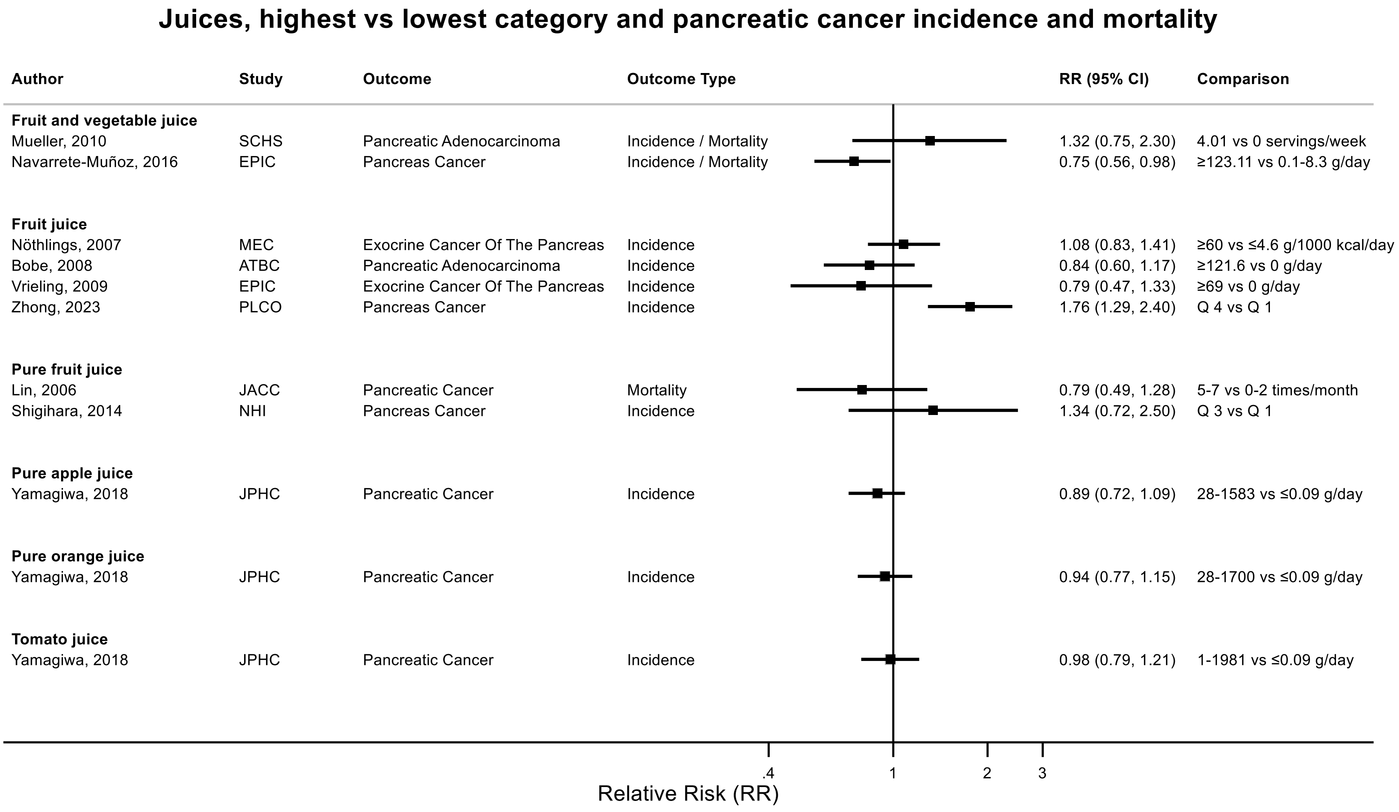

Figure legend: Forest plot shows the highest versus the lowest category results from the individual studies. Each square and the horizontal line across the square represent the RR estimate of the individual study and its 95% confidence interval (CI).

Abbreviations: CI, confidence interval; RR, relative risk.

Study abbreviations: SCHS, Singapore Chinese Health study; EPIC, European Prospective Investigation into Cancer and nutrition; MEC, Multiethnic Cohort Study; ATBC, Alpha-Tocopherol, Beta-Carotene Cancer Prevention Study; PLCO, Prostate, Lung, Colorectal and Ovarian Cancer Screening trial; JACC, Japan Collaborative Cohort study; NHI, Ohsaki National Health Insurance Cohort Study; JPHC, Japan Public Health Center-Based Prospective Study

Mueller, 2010[125], Navarrete-Muñoz, 2016[127], Nöthlings, 2007[131], Bobe, 2008[41], Vrieling, 2009[167], Zhong, 2023[188], Lin, 2006[103], Shigihara, 2014[154], Yamagiwa, 2019[176]

**Supplementary Figure 59. Highest versus the lowest category results from individual studies on juices and breast cancer incidence.**

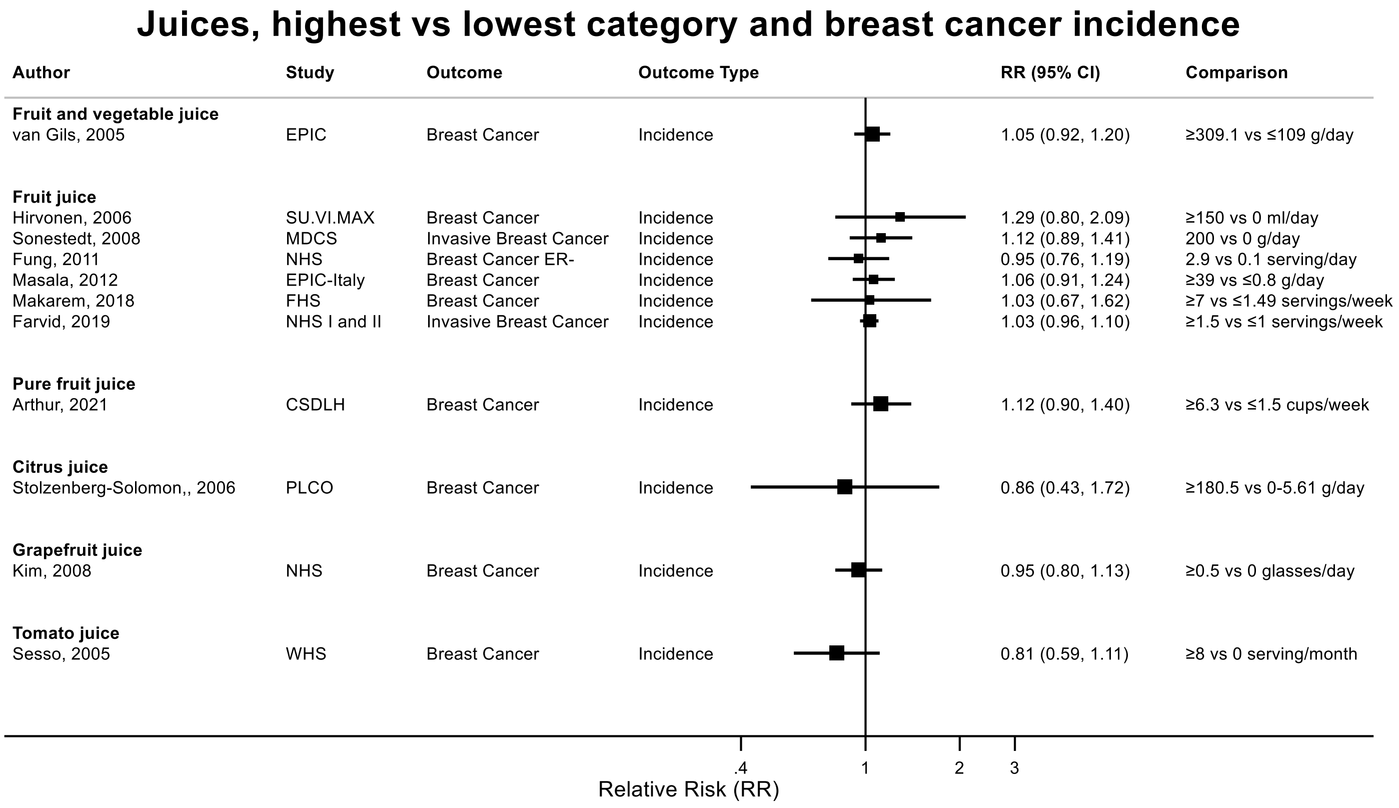

Figure legend: Forest plot shows the highest versus the lowest category results from the individual studies. Each square and the horizontal line across the square represent the RR estimate of the individual study and its 95% confidence interval (CI).

Abbreviations: CI, confidence interval; RR, relative risk.

Study abbreviations: EPIC, European Prospective Investigation into Cancer and nutrition; SU.VI.MAX, Supplémentation en Vitamines et Minéraux Antioxydants Study; MDCS, Malmö Diet and Cancer Study; NHS, Nurses' Health Study; EPIC-Italy, European Prospective Investigation into Cancer and nutrition-Italy cohort; FHS, Framingham Heart Study; NHS I and II, Nurses' Health Study I and II; CSDLH, Canadian Study of Diet, Lifestyle and Health; PLCO, Prostate, Lung, Colorectal and Ovarian Cancer Screening trial; WHS, Women's Health Study

van Gils, 2005[164], Hirvonen, 2006[69], Sonestedt, 2008[156], Fung, 2011[63], Masala, 2012[112], Makarem, 2018[108], Farvid, 2019[52], Arthur, 2021[38], Stolzenberg-Solomon, 2006[160], Kim, 2008[86], Sesso, 2005[153]

**Supplementary Figure 60. Highest versus the lowest category results from individual studies on juices and breast cancer incidence, by menopausal status.**

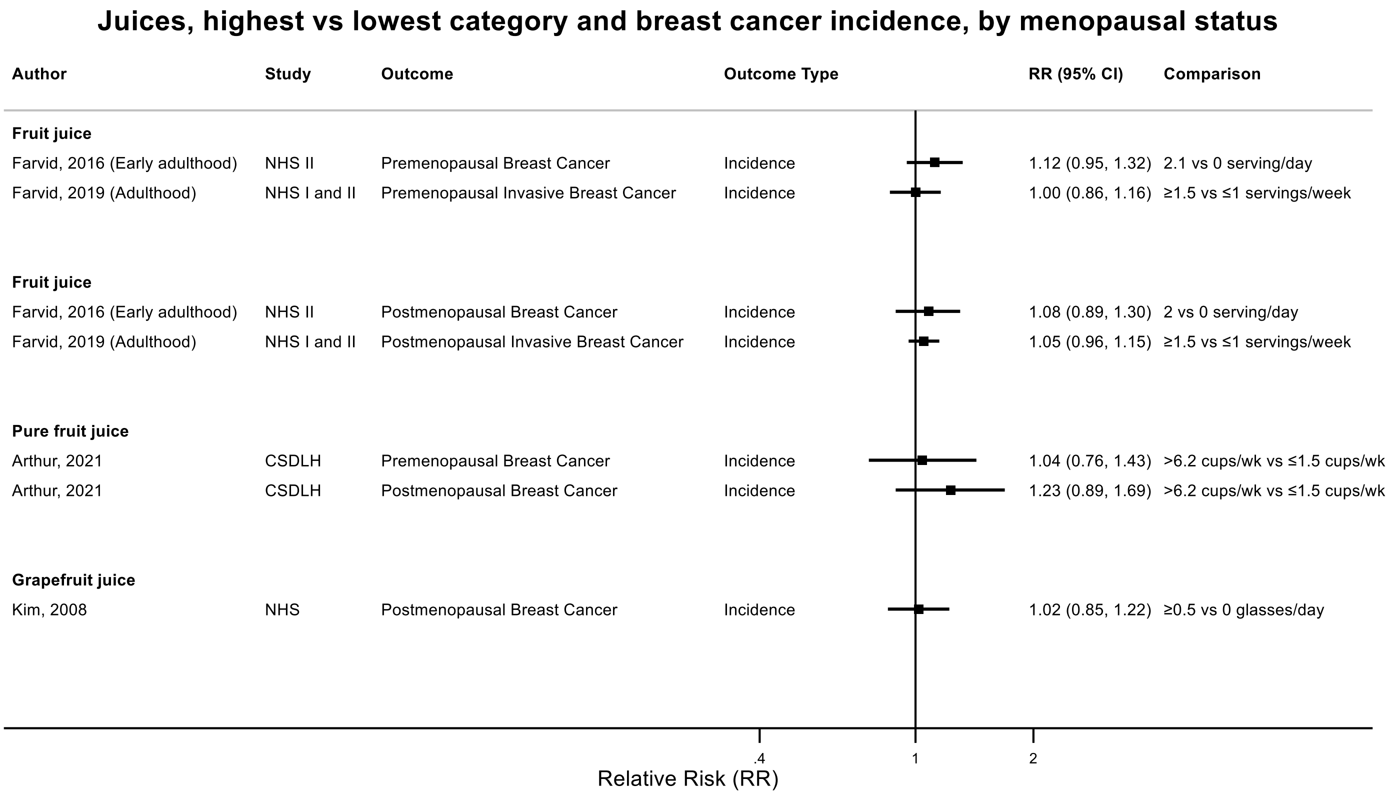

Figure legend: Forest plot shows the highest versus the lowest category results from the individual studies. Each square and the horizontal line across the square represent the RR estimate of the individual study and its 95% confidence interval (CI).

Abbreviations: CI, confidence interval; RR, relative risk.

Study abbreviations: NHS II, Nurses' Health Study II; NHS I and II, Nurses' Health Study I and II; CSDLH, Canadian Study of Diet, Lifestyle and Health; NHS, Nurses' Health Study

Farvid, 2016[51], Farvid, 2019[52], Arthur, 2021[38], Kim, 2008[86]

**Supplementary Figure 61. Highest versus the lowest category results from individual studies on juices and endometrial cancer incidence.**

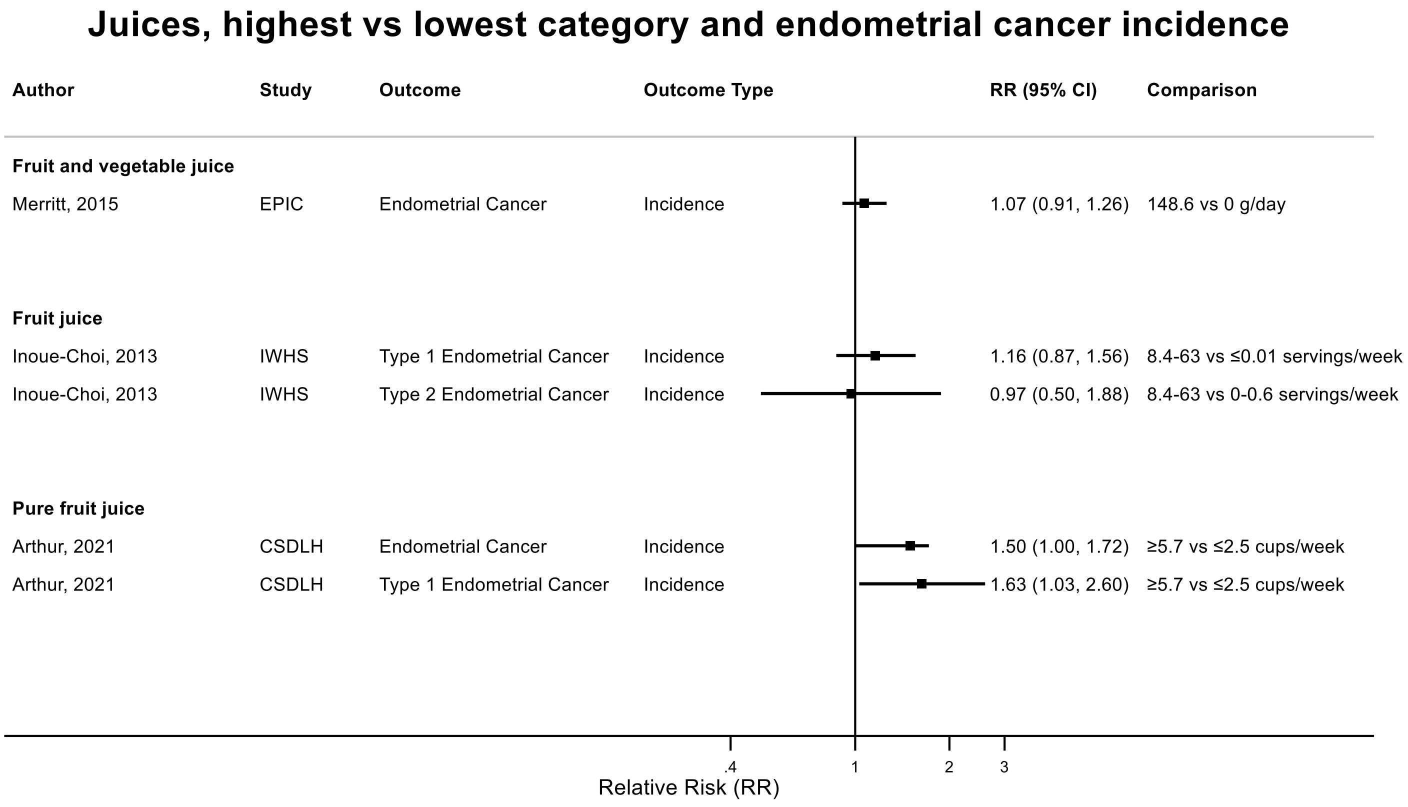

Figure legend: Forest plot shows the highest versus the lowest category results from the individual studies. Each square and the horizontal line across the square represent the RR estimate of the individual study and its 95% confidence interval (CI).

Abbreviations: CI, confidence interval; RR, relative risk.

Study abbreviations: EPIC, European Prospective Investigation into Cancer and nutrition; IWHS, The Iowa Women's Health Study; CSDLH, Canadian Study of Diet, Lifestyle and Health

Merritt, 2015[117], Inoue-Choi, 2013[74], Arthur, 2021[38]

#### Supplementary Figure 62. Highest versus the lowest category results from individual studies on juices and prostate cancer incidence and mortality.

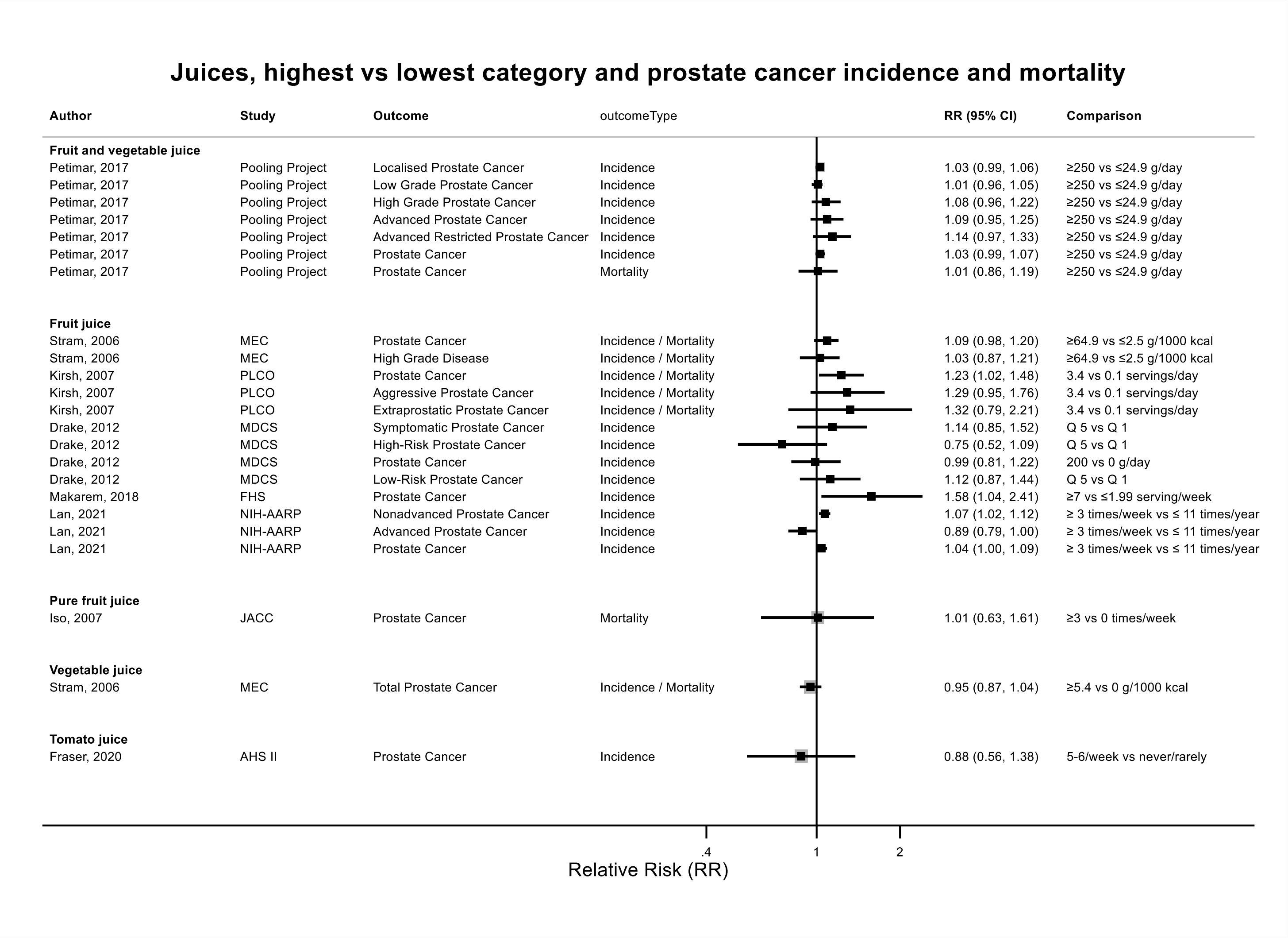

Figure legend: Forest plot shows the highest versus the lowest category results from the individual studies. Each square and the horizontal line across the square represent the RR estimate of the individual study and its 95% confidence interval (CI).

Abbreviations: CI, confidence interval; RR, relative risk.

Study abbreviations: Pooling Project, The Pooling Project of Prospective Studies of Diet and Cancer; MEC, Multiethnic Cohort Study; PLCO, Prostate, Lung, Colorectal and Ovarian Cancer Screening trial; MDCS, Malmö Diet and Cancer Study; FHS, Framingham Heart Study; NIH-AARP, National Institute of Health + American Association of Retired Persons Diet and Health Study; JACC, Japan Collaborative Cohort study; MEC, Multiethnic Cohort Study; AHS II, Adventist Health Study

Petimar, 2017[139], Stram, 2006[161], Kirsh, 2007[89], Drake, 2012[47], Makarem, 2018[108], Lan, 2021[94], Iso, 2007[76], Fraser, 2020[56]

Notes: Lan, 2021, NIH-AARP investigated fruit juice intake at ages 12-13 years.

**Supplementary Figure 63. Highest versus the lowest category results from individual studies on citrus juices and melanoma incidence.**

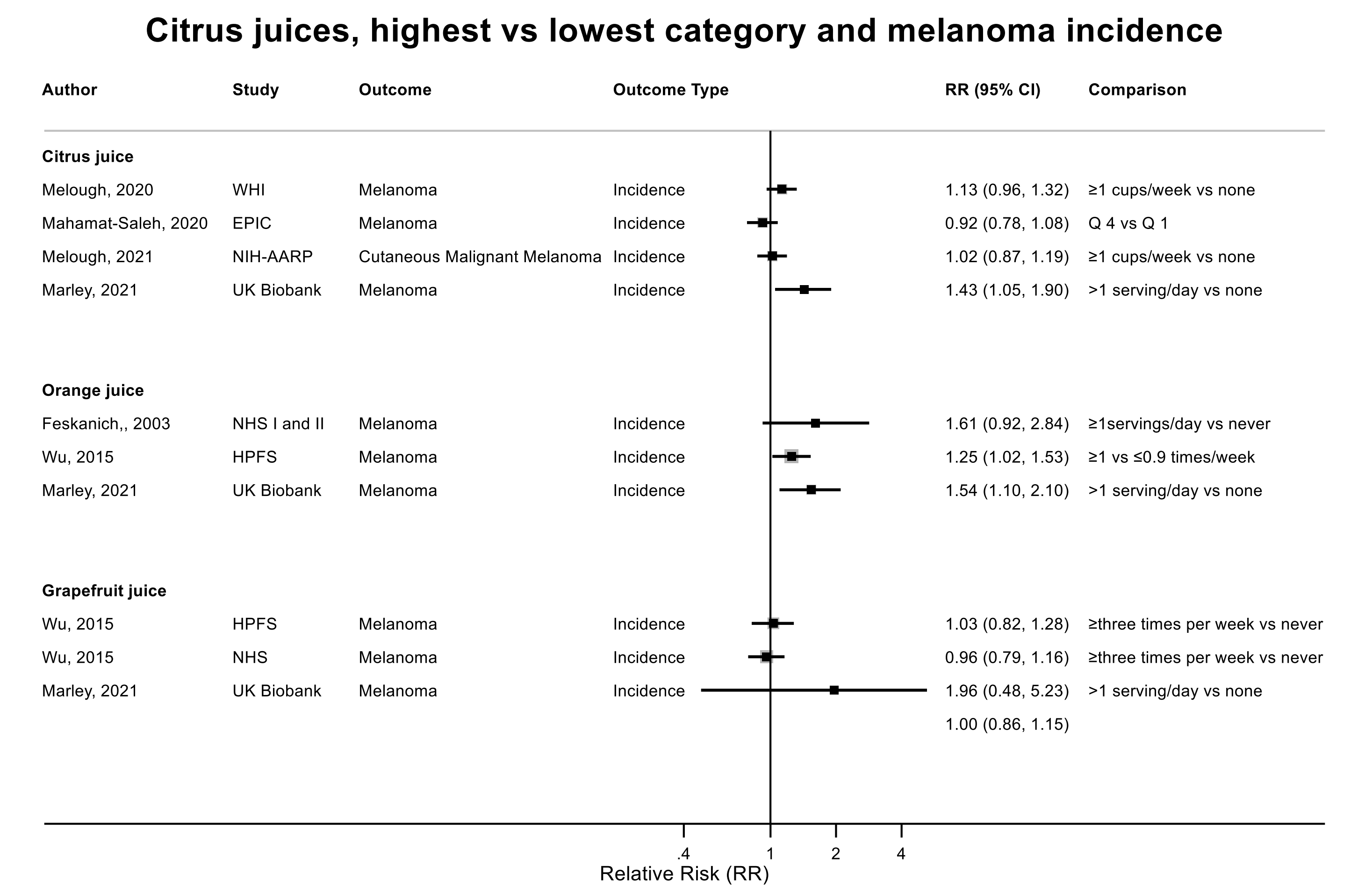

Figure legend: Forest plot shows the highest versus the lowest category results from the individual studies. Each square and the horizontal line across the square represent the RR estimate of the individual study and its 95% confidence interval (CI).

Abbreviations: CI, confidence interval; RR, relative risk.

Study abbreviations: WHI, Women's Health Initiative Study; EPIC, European Prospective Investigation into Cancer and nutrition; NIH-AARP, National Institute of Health + American Association of Retired Persons Diet and Health Study; UK Biobank, United Kingdom Biobank Study; NHS I and II, Nurses' Health Study I and II; HPFS, Health Professionals Follow-up Study

Melough, 2020[116], Mahamat-Saleh, 2020[107], Melough, 2021[115], Marley, 2021[110], Feskanich, 2003[54], Wu, 2023[172]

#### Supplementary Figure 64. Highest versus the lowest category results from individual studies on citrus juices and non-melanoma skin cancer incidence.

##### Citrus juices, highest vs lowest category and non-melanoma skin cancer incidence

| Author | Study | Outcome | Outcome Type | RR (95% CI) | Comparison |
|---|---|---|---|---|---|
| **Citrus juice** | | | | | |
| Sakaki, 2021 | WHI | Non-Melanoma Skin Cancer | Incidence | 1.08 (1.02, 1.13) | ≥1 vs <1 serving/week |
| Marley, 2022 | UK Biobank | Non-Melanoma Skin Cancer | Incidence | 1.01 (0.87, 1.16) | >1 serving/day vs none |
| **Orange juice** | | | | | |
| Marley, 2022 | UK Biobank | Non-Melanoma Skin Cancer | Incidence | 1.06 (0.90, 1.24) | >1 serving/day vs none |
| **Grapefruit juice** | | | | | |
| Marley, 2022 | UK Biobank | Non-Melanoma Skin Cancer | Incidence | 0.94 (0.44, 1.75) | >1 serving/day vs none |

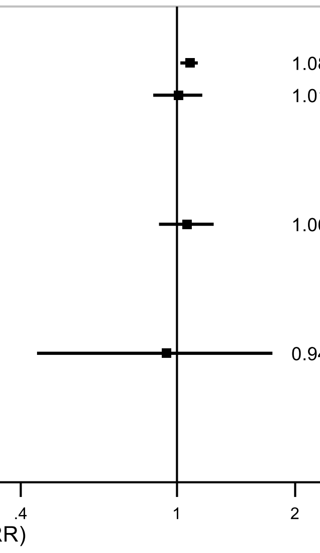

Figure legend: Forest plot shows the highest versus the lowest category results from the individual studies. Each square and the horizontal line across the square represent the RR estimate of the individual study and its 95% confidence interval (CI).

Abbreviations: CI, confidence interval; RR, relative risk.

Study abbreviations: WHI, Women’s Health Initiative Study; UK Biobank, United Kingdom Biobank Study

Sakaki, 2021[148], Marley, 2022[111]

**Supplementary Figure 65. Highest versus the lowest category results from individual studies on citrus juices and basal cell carcinoma incidence.**

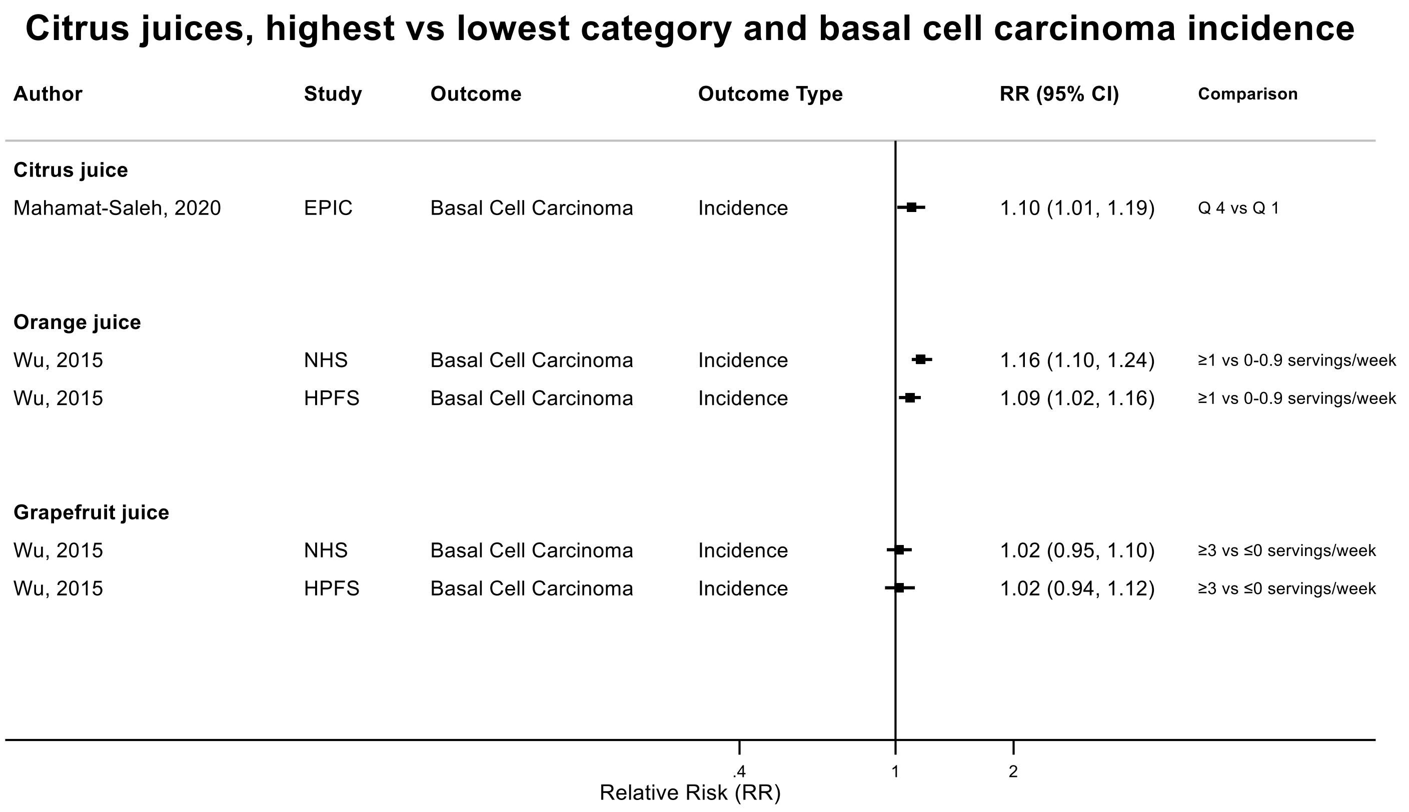

Figure legend: Forest plot shows the highest versus the lowest category results from the individual studies. Each square and the horizontal line across the square represent the RR estimate of the individual study and its 95% confidence interval (CI).

Abbreviations: CI, confidence interval; RR, relative risk.

Study abbreviations: EPIC, European Prospective Investigation into Cancer and nutrition; NHS, Nurses' Health Study; HPFS, Health Professionals Follow-up Study

Mahamat-Saleh, 2020[107], Wu, 2015[170]

**Supplementary Figure 66. Highest versus the lowest category results from individual studies on citrus juices and squamous cell carcinoma incidence.**

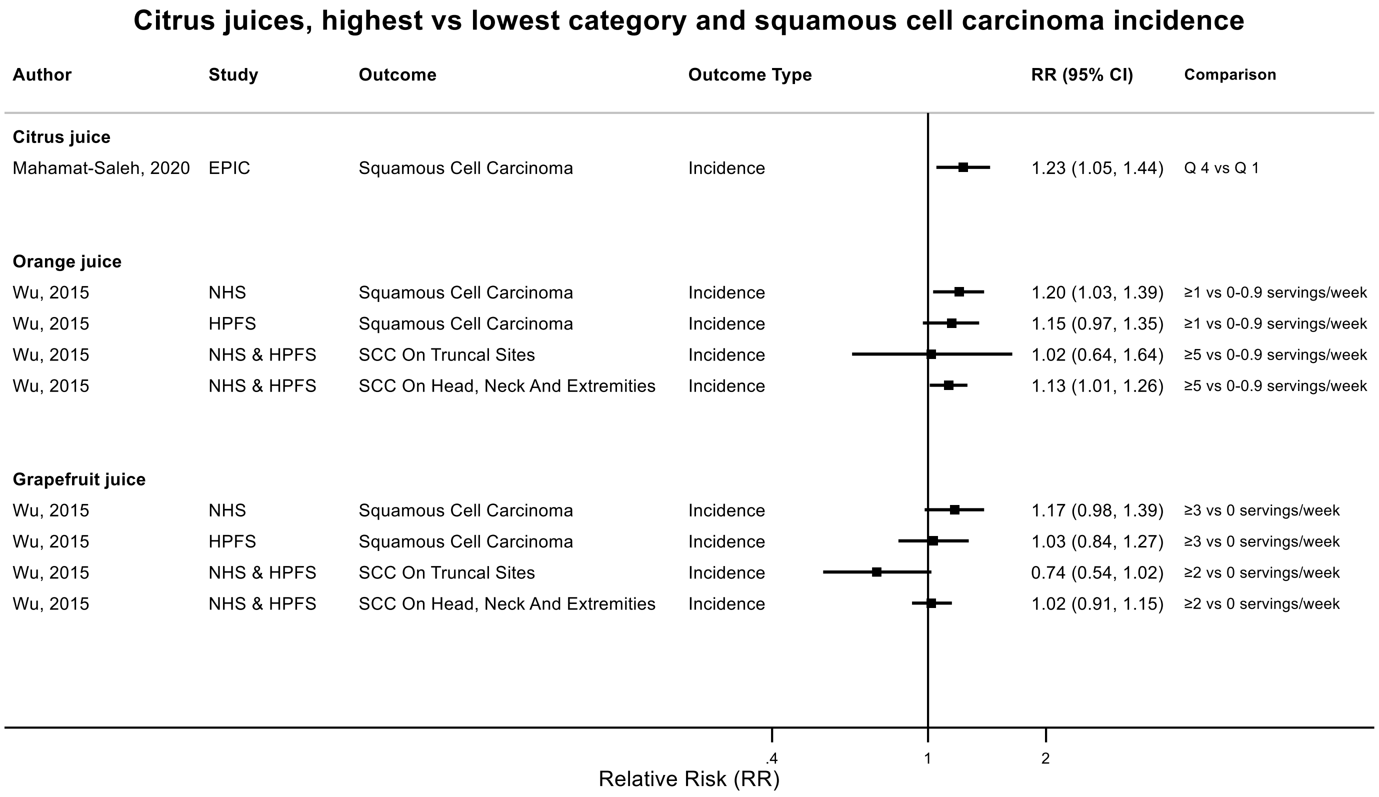

Figure legend: Forest plot shows the highest versus the lowest category results from the individual studies. Each square and the horizontal line across the square represent the RR estimate of the individual study and its 95% confidence interval (CI).

Abbreviations: CI, confidence interval; RR, relative risk.

Study abbreviations: EPIC, European Prospective Investigation into Cancer and nutrition; NHS, Nurses' Health Study; HPFS, Health Professionals Follow-up Study; NHS & HPFS, Nurses' Health Study & Health Professionals Follow-up Study

Mahamat-Saleh, 2020[107], Wu, 2015[170]

**Supplementary Figure 67. Highest versus the lowest category results from individual studies on juices and bladder cancer incidence and mortality.**

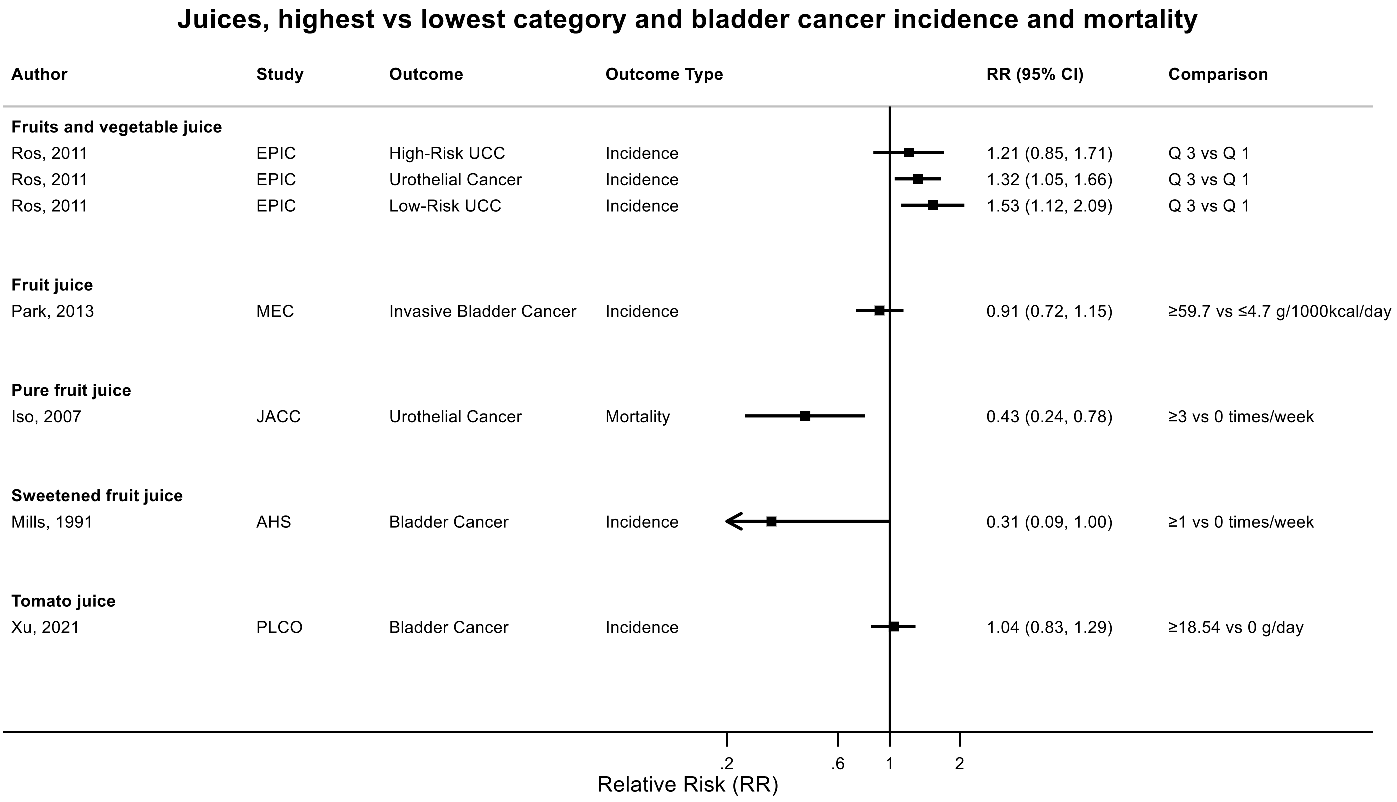

Figure legend: Forest plot shows the highest versus the lowest category results from the individual studies. Each square and the horizontal line across the square represent the RR estimate of the individual study and its 95% confidence interval (CI).

Abbreviations: CI, confidence interval; RR, relative risk.

Study abbreviations: EPIC, European Prospective Investigation into Cancer and nutrition; MEC, Multiethnic Cohort Study; JACC, Japan Collaborative Cohort study; AHS, Adventist Health Study; PLCO, Prostate, Lung, Colorectal and Ovarian Cancer Screening trial

Ros, 2011[147], Park, 2013[138], Iso, 2007[76], Mills, 1991[123], Xu, 2021[175]

**Supplementary Figure 68. Highest versus the lowest category results from individual studies on juices and kidney cancer incidence and mortality.**

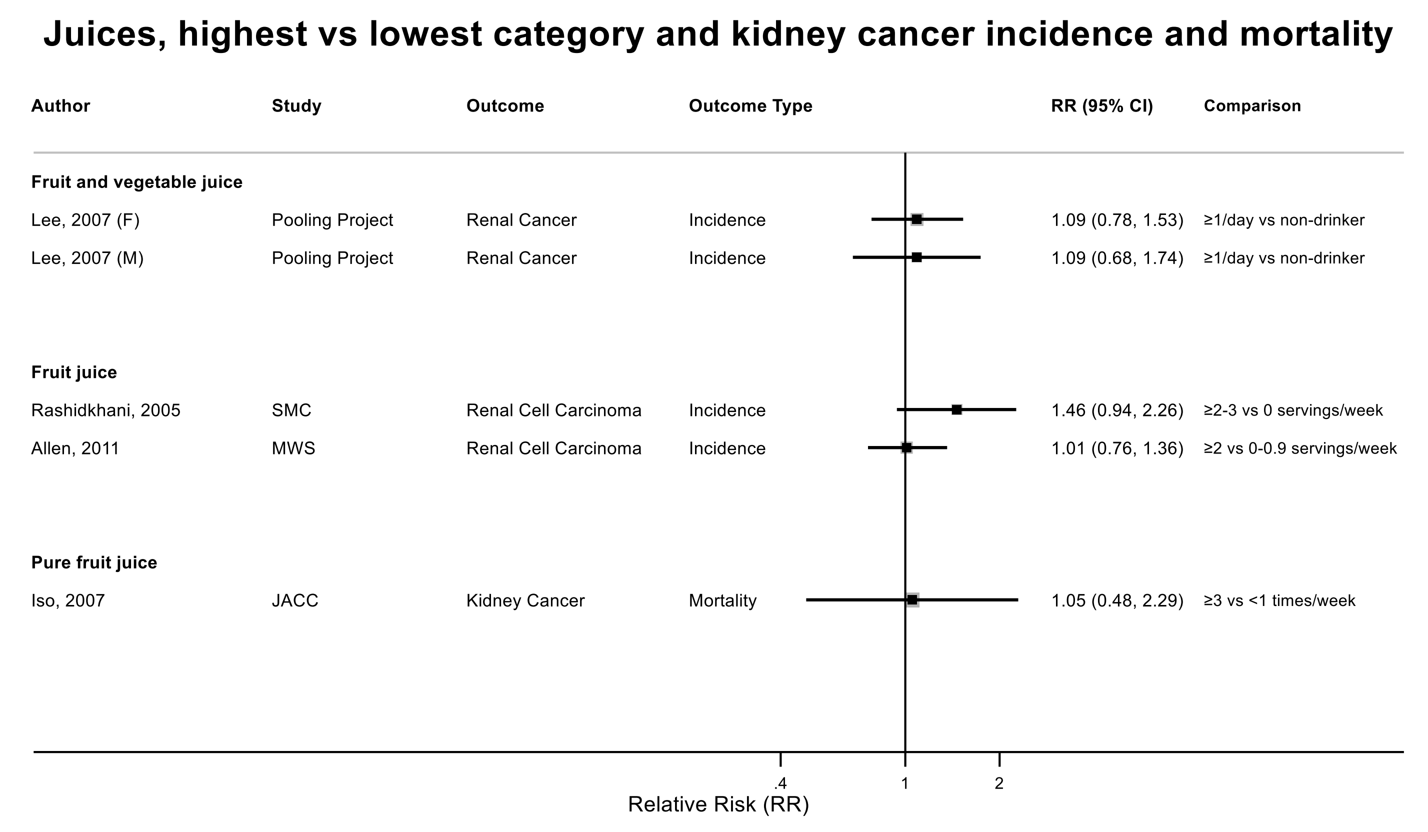

Figure legend: Forest plot shows the highest versus the lowest category results from the individual studies. Each square and the horizontal line across the square represent the RR estimate of the individual study and its 95% confidence interval (CI).

Abbreviations: CI, confidence interval; RR, relative risk.

Study abbreviations: Pooling Project, The Pooling Project of Prospective Studies of Diet and Cancer; SMC, Swedish Mammography Cohort; MWS, Million Women Study; JACC, Japan Collaborative Cohort study

Lee, 2007[26], Rashidkhani, 2005[142], Allen, 2011[36], Iso, 2007[76]

**Supplementary Figure 69. Highest versus the lowest category results from individual studies on juices and lung cancer incidence and mortality.**

#### Juices, highest vs lowest category and lung cancer incidence and mortality

| Author | Study | Outcome | Outcome Type | RR (95% CI) | Comparison |
|---|---|---|---|---|---|
| **Fruit and vegetable juice** | | | | | |
| Linseisen, 2007 | EPIC | Lung Cancer | Incidence | 0.89 (0.72, 1.10) | 182.2 vs 21.5 g/day |
| Gnagnarella, 2013 | COSMOS | Lung Cancer | Incidence | 0.96 (0.66, 1.39) | 62.46 vs 0 g/1000 kcal/day |
| **Fruit juice** | | | | | |
| Ozasa, 2001 | JACC | Lung Cancer | Mortality | 0.91 (0.72, 1.16) | 12-16 vs <1-2 times/month |
| **Pure fruit juice** | | | | | |
| Iso, 2007 | JACC | Lung Cancer | Mortality | 0.90 (0.76, 1.07) | ≥3 vs ≤1 times/week |
| **Citrus juice** | | | | | |
| Smith-Warner, 2003 | Pooling Project | Lung Cancer | Incidence | 0.82 (0.71, 0.94) | ≥1/2 serving/day vs 0 serving/week |
| Linseisen, 2007 | EPIC | Lung Cancer | Incidence | 0.82 (0.65, 1.03) | 106.6 vs 12.9 g/day |
| **Other fruit juices (not citrus)** | | | | | |
| Voorrips, 2000 | NLCS | Lung Cancer | Incidence | 0.90 (0.70, 1.20) | 20 vs 0 g/day |

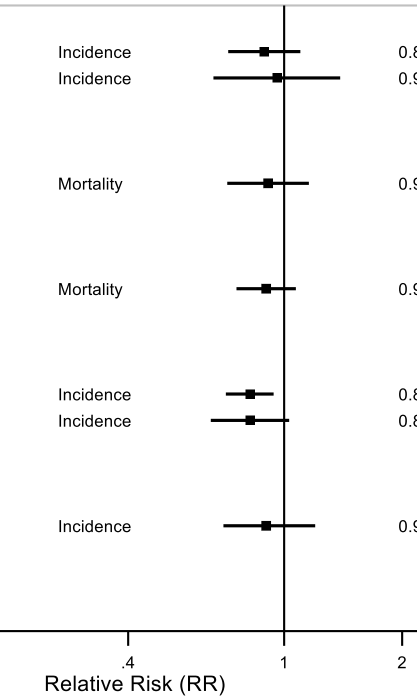

Figure legend: Forest plot shows the highest versus the lowest category results from the individual studies. Each square and the horizontal line across the square represent the RR estimate of the individual study and its 95% confidence interval (CI).

Abbreviations: CI, confidence interval; RR, relative risk.

Study abbreviations: EPIC, European Prospective Investigation into Cancer and nutrition; COSMOS, Continuous Observation of Smoking Subjects Study; JACC, Japan Collaborative Cohort study; Pooling Project, The Pooling Project of Prospective Studies of Diet and Cancer; NLCS, Netherlands Cohort Study on Diet and Cancer

Linseisen, 2007[104], Gnagnarella, 2013[65], Ozasa, 2001[134], Iso, 2007[76], Smith-Warner, 2003[155], Voorrips, 2000[166]

**Supplementary Figure 70. Linear dose-response meta-analysis of sugar sweetened beverages and ovarian cancer incidence.**

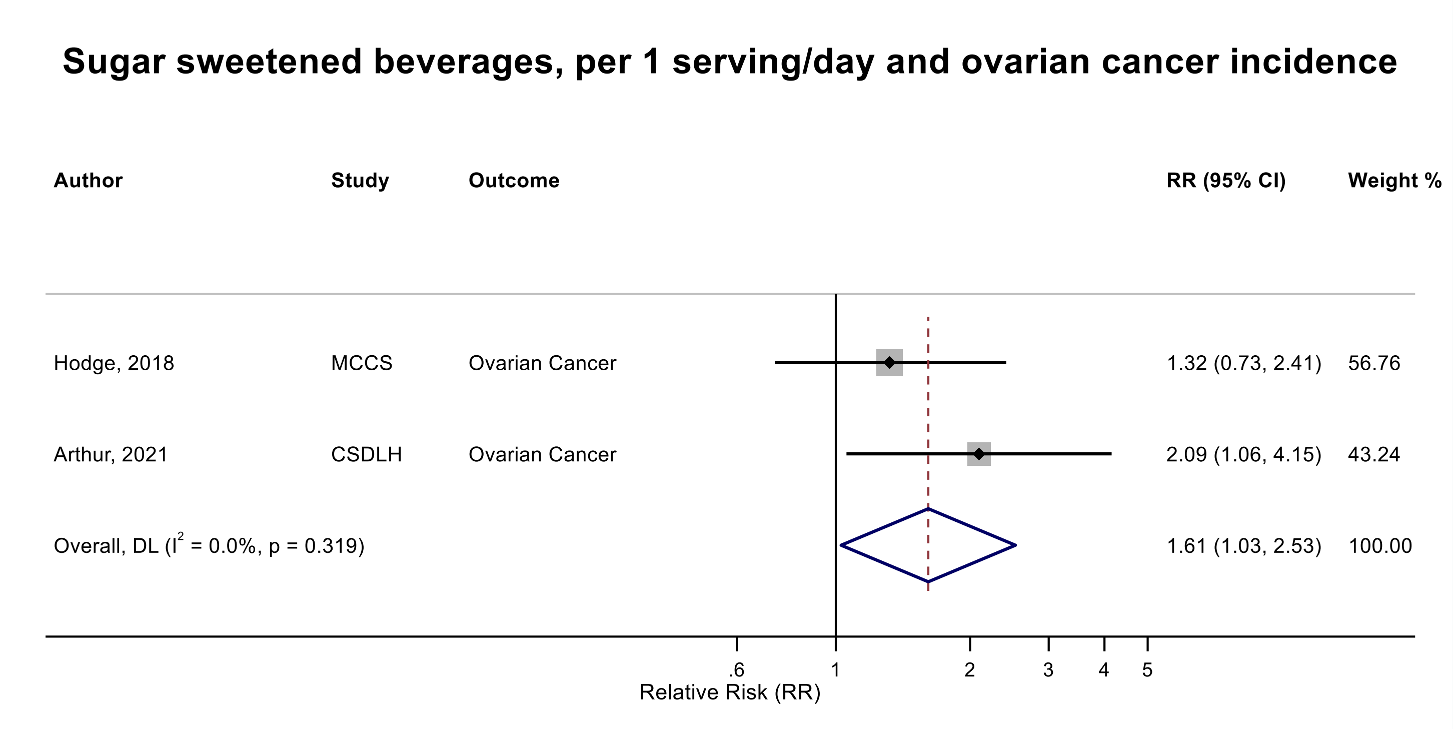

Figure legend: Forest plot shows the linear dose-response results (per 1 serving [355 mL] of sugar sweetened beverages/day) from the inverse variance DerSimonian-Laird random-effects model. The diamond represents the summary relative risk (RR) estimate, the diamond's width is the 95% confidence interval (CI). Each square and the horizontal line across the square represent the RR estimate of the individual study and its 95% CI.

Abbreviations: CI, confidence interval; RR, relative risk.

Study abbreviations: CSDLH, Canadian Study of Diet, Lifestyle, and Health; MCCS, Melbourne Collaborative Cohort Study

Note: both studies excluded people with a history of diabetes at baseline.

Hodge, 2018[70], Arthur, 2021[38]

**Supplementary Figure 71. Linear dose-response meta-analysis of sugar sweetened beverages and endometrial cancer incidence.**

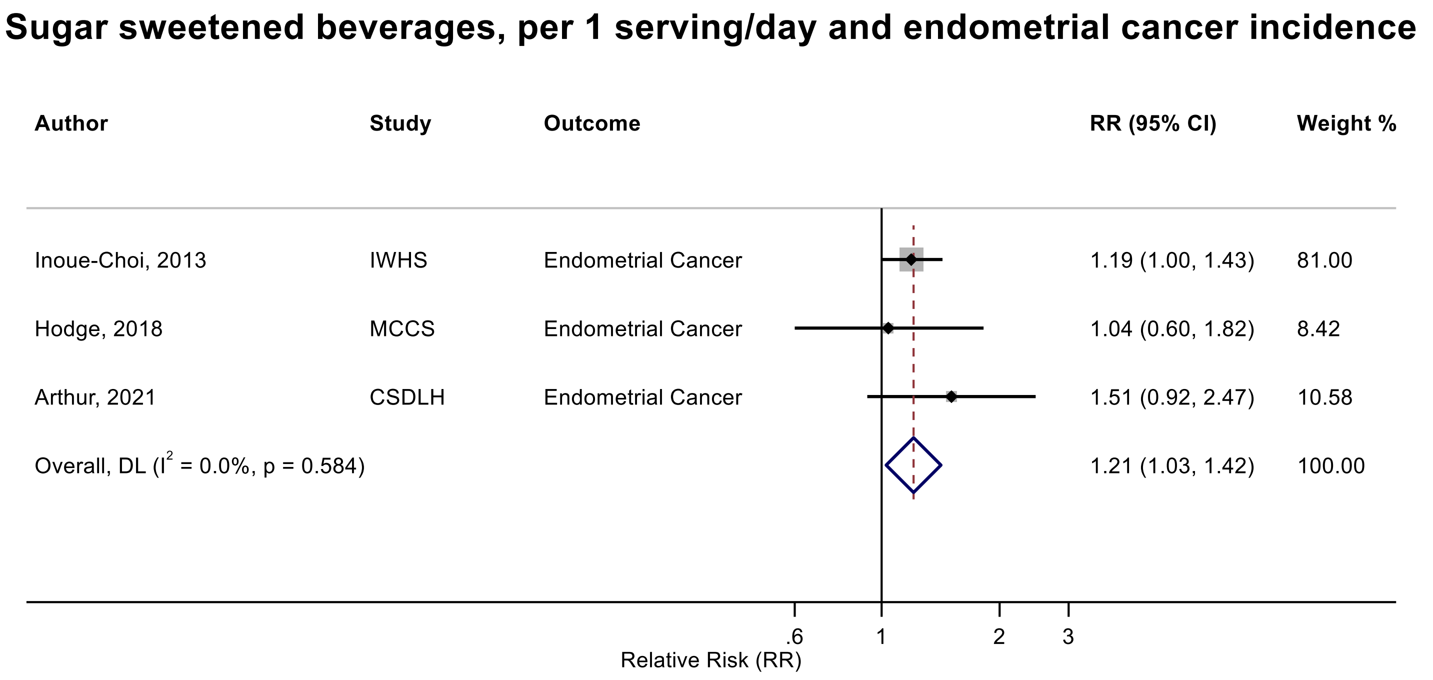

Figure legend: Forest plot shows the linear dose-response results (per 1 serving [355 mL] of sugar sweetened beverages/day) from the inverse variance DerSimonian-Laird random-effects model. The diamond represents the summary relative risk (RR) estimate, the diamond's width is the 95% confidence interval (CI), and the diamond's horizontal line is the 95% prediction interval (PI). Each square and the horizontal line across the square represent the RR estimate of the individual study and its 95% CI.

Abbreviations: CI, confidence interval; RR, relative risk.

Study abbreviations: IWHS, Iowa Women's Health Study; MCCS, Melbourne Collaborative Cohort Study; CSDLH, Canadian Study of Diet, Lifestyle, and Health

Note: All studies excluded people with a history of diabetes at baseline. In IWHS, we combined type I and II endometrial cancers using Hamling approach to calculate the estimate for total endometrial cancer.

Inoue-Choi, 2013[74], Hodge, 2018[70], Arthur, 2021[38]

**Supplementary Figure 72. Linear dose-response meta-analysis of carbonated sugar sweetened beverages and pancreatic cancer incidence.**

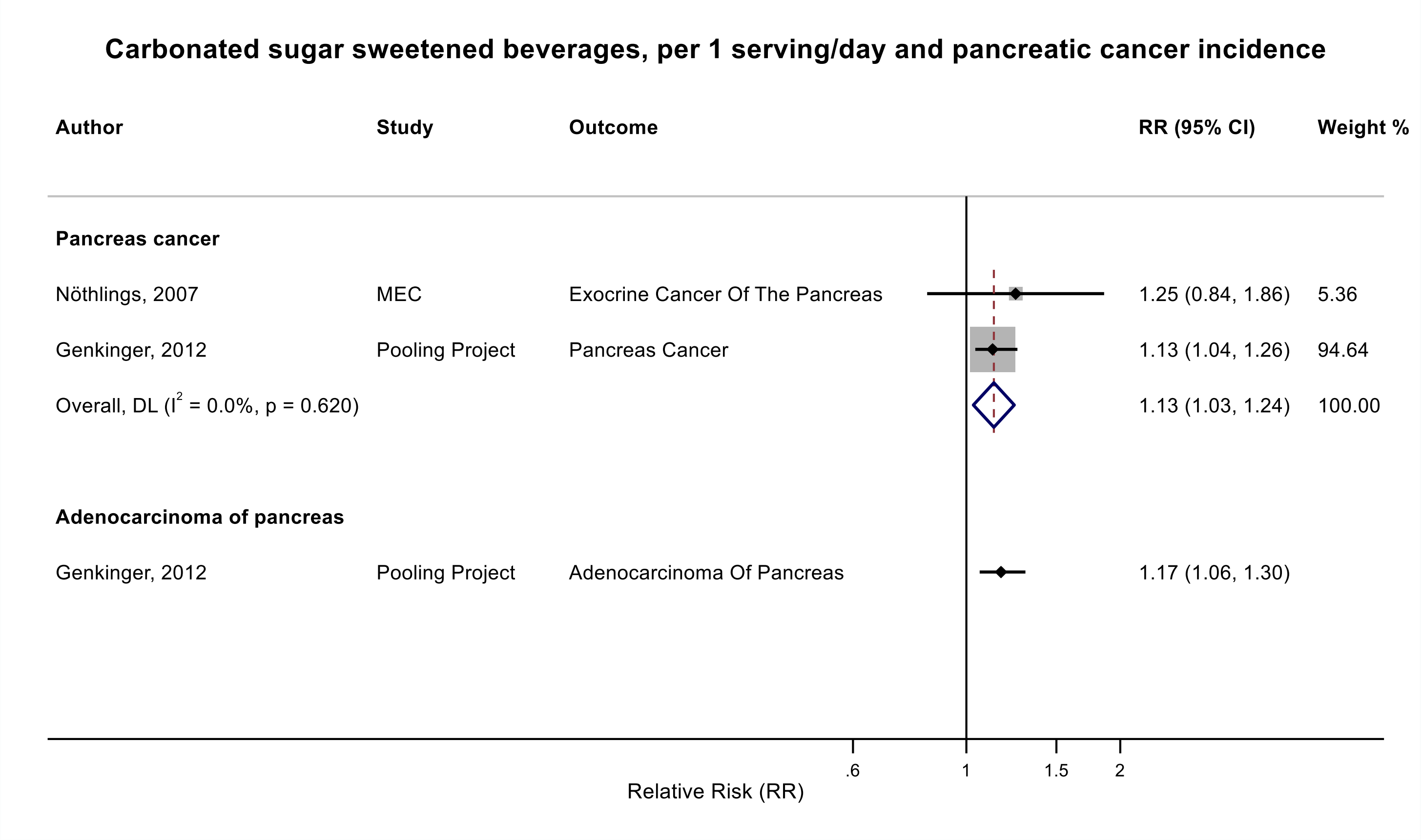

Figure legend: Forest plot shows the linear dose-response results (per 1 serving [355 mL] of carbonated sugar sweetened beverage /day) from the inverse variance DerSimonian-Laird random-effects model. The diamond represents the summary relative risk (RR) estimate, the diamond's width is the 95% confidence interval (CI). Each square and the horizontal line across the square represent the RR estimate of the individual study and its 95% CI.

Abbreviations: CI, confidence interval; RR, relative risk.

Study abbreviations: MEC, Multiethnic Cohort Study; Pooling Project, The Pooling Project of Prospective Studies of Diet and Cancer

Nöthlings, 2007[131], Genkinger, 2012[64]

**Supplementary Figure 73. Linear dose-response meta-analysis of sugar sweetened beverages and pancreatic cancer incidence.**

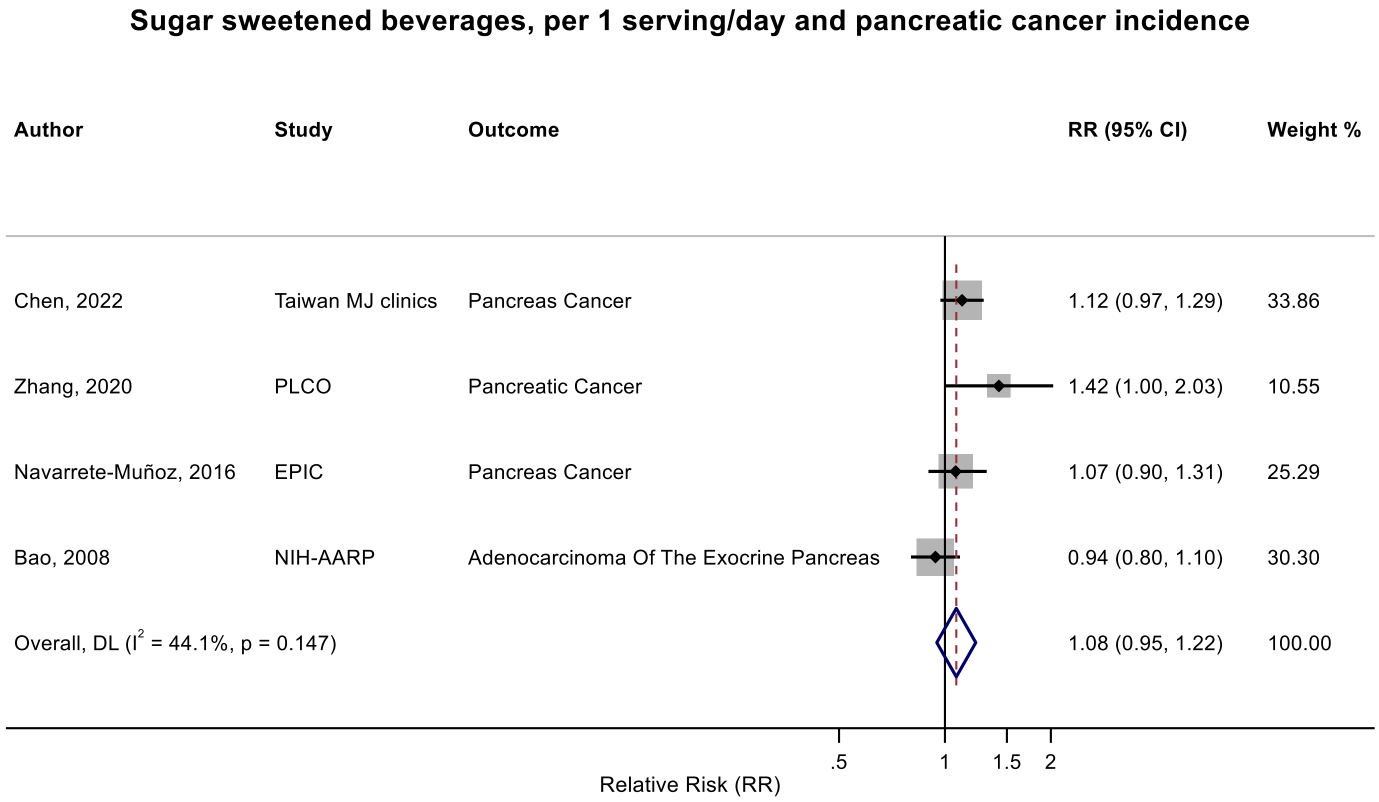

Figure legend: Forest plot shows the linear dose-response results (per 1 serving [355 mL] of sugar sweetened beverages/day) from the inverse variance DerSimonian-Laird random-effects model. The diamond represents the summary relative risk (RR) estimate, the diamond's width is the 95% confidence interval (CI). Each square and the horizontal line across the square represent the RR estimate of the individual study and its 95% CI.

Abbreviations: CI, confidence interval; RR, relative risk.

Study abbreviations: NIH-AARP, National Institute of Health + American Association of Retired Persons Diet and Health Study; EPIC, European Prospective Investigation into Cancer and nutrition; PLCO, Prostate, Lung, Colorectal and Ovarian Cancer Screening trial; Taiwan MJ clinics, The Taiwan MJ Cohort Study

Note: We used a publication from NIH-AARP for the linear dose-response meta-analysis that had the most comprehensive definition of SSBs (regular soft drinks+regular fruit drinks). The RR for regular soft drinks without fruit drinks was 1.01 (95%CI: 77-1.31). Another publication of NIH-AARP that reported on SSBs+fruit juices, that could not be included in the linear dose-response meta-analysis due to insufficient data format, reported a slight positive association with pancreatic cancer (RR per a one-unit change in the AHEI-2010 component score: 1.01, 95%CI: 1.00-1.02; and in the DASH-Fung component score: 1.04, 95%CI: 1.01-1.06)[80]. There was another publication from PLCO[188] that could not be included in the dose-response meta-analysis because of insufficient data format and reported little evidence of an association for SSBs, as part of the unhealthful plant-based diet score, and pancreatic cancer incidence/mortality ($RR_{Q4\ vs\ Q1}$: 1.01, 95%CI: 0.76-1.33).

Bao, 2008[39], Navarrete-Muñoz, 2016[127], Zhang, 2020[184], Chen, 2022[45]

**Supplementary Figure 74. Linear dose-response meta-analysis of sugar sweetened beverages and pancreatic cancer incidence and mortality combined.**

#### Sugar sweetened beverages, per 1 serving/day and pancreatic cancer incidence and mortality

| Author | Study | Outcome | Outcome Type | RR (95% CI) | Weight % |
|---|---|---|---|---|---|
| Chen, 2022 | Taiwan MJ clinics | Pancreas Cancer | Incidence | 1.12 (0.97, 1.29) | 19.36 |
| McCullough, 2022 | CPS II | Pancreas Cancer | Mortality | 1.00 (0.98, 1.03) | 46.36 |
| Zhang, 2020 | PLCO | Pancreatic Cancer | Incidence | 1.42 (1.00, 2.03) | 4.66 |
| Navarrete-Muñoz, 2016 | EPIC | Pancreas Cancer | Incidence | 1.07 (0.90, 1.31) | 13.04 |
| Bao, 2008 | NIH-AARP | Adenocarcinoma Of The Exocrine Pancreas | Incidence | 0.94 (0.80, 1.10) | 16.58 |
| Overall, DL ($I^2$ = 44.1%, p = 0.128) | | | | 1.04 (0.96, 1.12) | 100.00 |

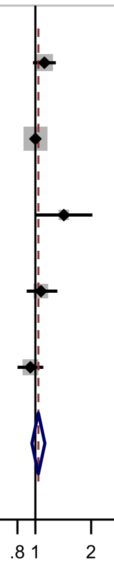

Relative Risk (RR)

Figure legend: Forest plot shows the linear dose-response results (per 1 serving [355 mL] of sugar sweetened beverages/day) from the inverse variance DerSimonian-Laird random-effects model. The diamond represents the summary relative risk (RR) estimate, the diamond's width is the 95% confidence interval (CI). Each square and the horizontal line across the square represent the RR estimate of the individual study and its 95% CI.

Abbreviations: CI, confidence interval; RR, relative risk.

Study abbreviations: NIH-AARP, National Institute of Health + American Association of Retired Persons Diet and Health Study; EPIC, European Prospective Investigation into Cancer and nutrition; PLCO, Prostate, Lung, Colorectal and Ovarian Cancer Screening trial; CPS II, Cancer Prevention Study II; Taiwan MJ clinics, The Taiwan MJ Cohort Study

Chen, 2022[45], McCullough, 2022[113], Zhang, 2020[184], Navarrete-Muñoz, 2016[127], Bao, 2008[39]

**Supplementary Figure 75. Linear dose-response meta-analysis of sugar sweetened and carbonated sugar sweetened beverages and pancreatic cancer incidence.**

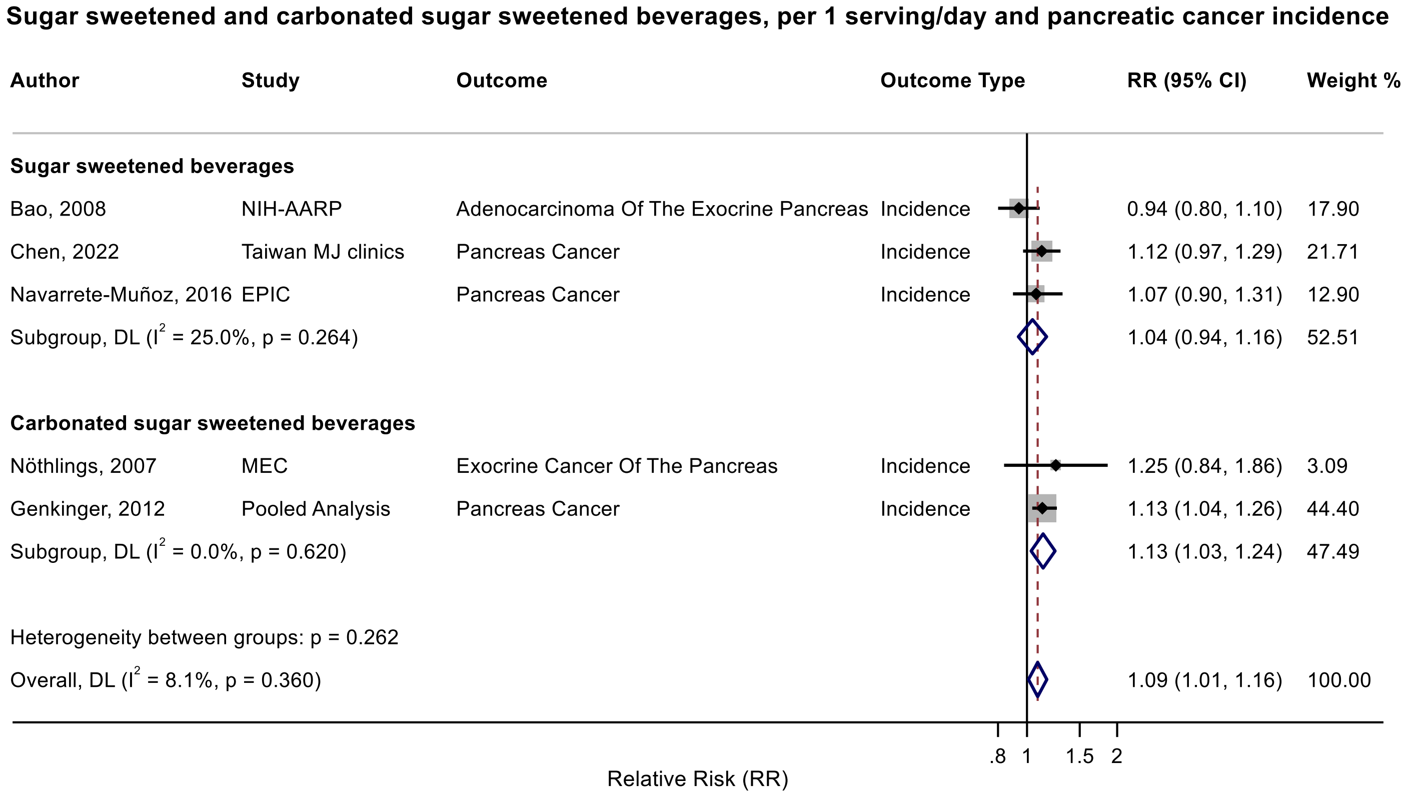

Figure legend: Forest plot shows the linear dose-response results (per 1 serving [355 mL] of sugar sweetened beverages/day) from the inverse variance DerSimonian-Laird random-effects model. The diamond represents the summary relative risk (RR) estimate, the diamond's width is the 95% confidence interval (CI). Each square and the horizontal line across the square represent the RR estimate of the individual study and its 95% CI.

Abbreviations: CI, confidence interval; RR, relative risk.

Study abbreviations: NIH-AARP, National Institute of Health + American Association of Retired Persons Diet and Health Study; EPIC, European Prospective Investigation into Cancer and nutrition; Taiwan MJ clinics, The Taiwan MJ Cohort Study; MEC, Multiethnic Cohort Study; Pooling Project, The Pooling Project of Prospective Studies of Diet and Cancer

Bao, 2008[39], Navarrete-Muñoz, 2016[127], Chen, 2022[45], Nöthlings, 2007[131], Genkinger, 2012[64]

**Supplementary Figure 76. Linear dose-response meta-analysis of sugar sweetened beverages and colorectal cancer incidence.**

#### Sugar sweetened beverages, per 1 serving/day and colorectal cancer incidence

| Author | Study | Outcome | RR (95% CI) | Weight % |
|---|---|---|---|---|
| Nomura, 2016 | BWHS | Colorectal Cancer | 0.72 (0.43, 1.21) | 1.50 |
| Makarem, 2018 | Framingham Offspring Cohort | Colorectal Cancer | 1.15 (0.70, 1.91) | 1.59 |
| Hodge, 2018 | MCCS | Colorectal Cancer | 1.17 (0.96, 1.43) | 8.07 |
| Pacheco, 2019 | CTS | Colorectal Cancer | 1.11 (0.94, 1.32) | 10.27 |
| Chazelas, 2019 | NutriNet-Santé | Colorectal Cancer | 1.50 (0.54, 4.22) | 0.40 |
| Barrubés, 2020 | PREDIMED | Colorectal Cancer | 1.97 (1.02, 3.80) | 0.96 |
| Leung, 2021 | JPHC | Colorectal Cancer | 0.97 (0.91, 1.04) | 24.34 |
| Arthur, 2021 | CSDLH | Colorectal Cancer | 1.25 (0.87, 1.81) | 2.92 |
| Wang, 2022 | NHS I and II | Colorectal Cancer | 1.09 (0.94, 1.25) | 12.73 |
| Wang, 2022 | HPFS | Colorectal Cancer | 1.28 (1.05, 1.55) | 8.35 |
| Lee, 2023 | HEXA-G | Colorectal Cancer | 1.01 (0.34, 2.96) | 0.37 |
| Kanehara, 2024 | MEC | Colorectal Cancer | 1.02 (0.98, 1.06) | 28.52 |
| Overall, DL ($I^2$ = 40.5%, p = 0.071) | | | 1.07 (1.00, 1.14) | 100.00 |

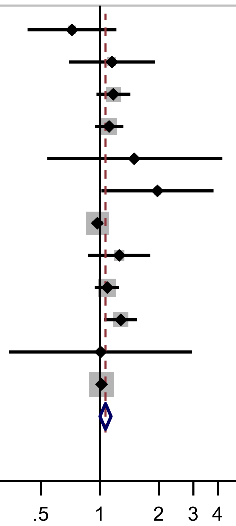

Relative Risk (RR)

Figure legend: Forest plot shows the linear dose-response results (per 1 serving [355 mL] of sugar sweetened beverages /day) from the inverse variance DerSimonian-Laird random-effects model. The diamond represents the summary relative risk (RR) estimate, the diamond's width is the 95% confidence interval (CI). Each square and the horizontal line across the square represent the RR estimate of the individual study and its 95% CI.

Abbreviations: CI, confidence interval; RR, relative risk.

Study abbreviations: BWHS, Black Women's Health Study; FHS-Offspring Cohort, Framingham Heart Study-Offspring Cohort; MCCS, Melbourne Collaborative Cohort Study; CTS, California Teachers Study; PREDIMED Prevención con Dieta Mediterránea study; JPHC, Japan Public Health Center-Based Prospective Study; CSDLH, Canadian Study of Diet, Lifestyle and Health; NHS I and II, Nurses' Health Study I and II; HPFS, Health Professionals Follow-up Study; HEXA-G, Health Examinees-Gem study; MEC, Multiethnic Cohort Study

Note: The pooled RR and 95%CI with 4 decimal point are 1.0681 (1.0003-1.1404; p-value = 0.050). One additional study (NIH-AARP)[122] that could not be included in the dose-response meta-analysis because of insufficient data format reported limited evidence of an association between sugar sweetened beverages and colorectal cancer incidence in males ($RR_{Q5\ vs\ Q1}$: 0.94, 95%CI: 0.85-1.03) and females ($RR_{Q5\ vs\ Q1}$: 1.09, 95%CI: 0.93-1.26). Another study (UK Biobank) that could not be included in the dose-response meta-analysis because of insufficient data format[78] indicated no association (RR: 1.00, 95%CI: 0.95-1.05) per one standard deviation increase in sugary sweetened beverages intake.

Nomura, 2016[129], Makarem, 2018[108], Hodge, 2018[70], Pacheco, 2019[135], Chazelas, 2019[44], Barrubés, 2020[40], Leung, 2021[101], Arthur, 2021[38], Wang, 2022[168], Lee, 2023[97], Kanehara, 2024[81]

**Supplementary Figure 77. Linear dose-response meta-analysis of sugar sweetened beverages and colon cancer incidence.**

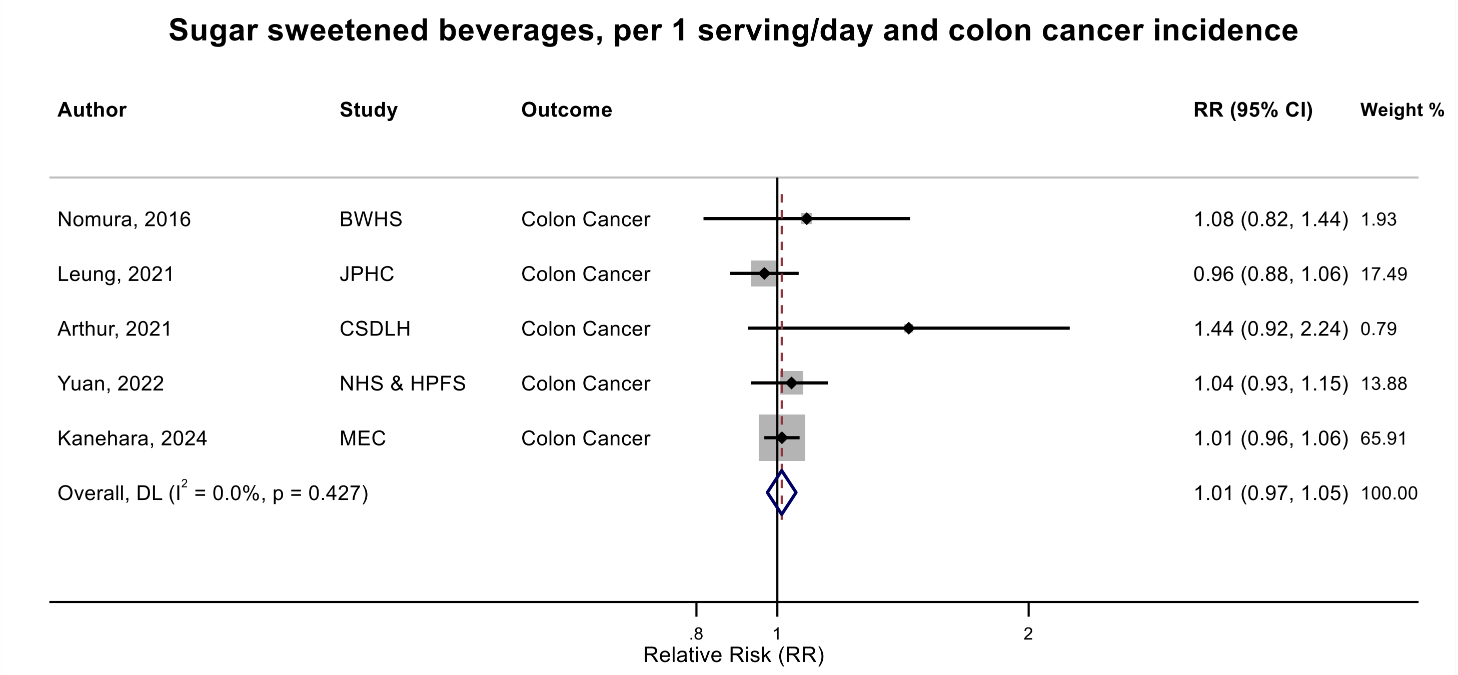

Figure legend: Forest plot shows the linear dose-response results (per 1 serving [355 mL] of sugar sweetened beverages /day) from the inverse variance DerSimonian-Laird random-effects model. The diamond represents the summary relative risk (RR) estimate, the diamond's width is the 95% confidence interval (CI). Each square and the horizontal line across the square represent the RR estimate of the individual study and its 95% CI.

Abbreviations: CI, confidence interval; RR, relative risk.

Study abbreviations: BWHS, Black Women's Health Study; JPHC, Japan Public Health Center-Based Prospective Study; CSDLH, Canadian Study of Diet, Lifestyle and Health; NHS & HPFS, Nurses' Health Study & Health Professionals Follow-up Study; MEC, Multiethnic Cohort Study

Note: One additional study (UK biobank)[78] that could not be included in the dose-response meta-analysis because of insufficient data format indicated little evidence of an association (RR: 1.01, 95%CI: 0.94-1.08) per one standard deviation increase in sugary sweetened beverages intake.

Nomura, 2016[129], Leung, 2021[101], Arthur, 2021[38], Yuan, 2022[178], Kanehara, 2024[81]

**Supplementary Figure 78. Linear dose-response meta-analysis of carbonated sugar sweetened beverages and colon cancer incidence.**

#### Carbonated sugar sweetened beverages, per 1 serving/day and colon cancer incidence

| Author | Study | Outcome | RR (95% CI) | Weight % |
|---|---|---|---|---|
| Zhang, 2010 | SMC | Colon Cancer | 1.09 (0.53, 2.24) | 1.65 |
| Zhang, 2010 | BCDDP | Colon Cancer | 0.52 (0.29, 0.92) | 2.54 |
| Zhang, 2010 | ATBC | Colon Cancer | 0.98 (0.56, 1.73) | 2.69 |
| Zhang, 2010 | NLCS | Colon Cancer | 1.07 (0.67, 1.72) | 3.88 |
| Zhang, 2010 | WHS | Colon Cancer | 0.93 (0.62, 1.40) | 5.07 |
| Zhang, 2010 | CNBSS | Colon Cancer | 1.08 (0.76, 1.50) | 7.38 |
| Zhang, 2010 | NHS | Colon Cancer | 1.03 (0.85, 1.25) | 22.37 |
| Zhang, 2010 | CPS II | Colon Cancer | 0.90 (0.72, 1.13) | 16.84 |
| Zhang, 2010 | IWHS | Colon Cancer | 0.93 (0.72, 1.18) | 14.33 |
| Zhang, 2010 | HPFS | Colon Cancer | 1.14 (0.94, 1.38) | 23.25 |
| Overall, DL ($I^2$ = 0.0%, p = 0.495) | | | 1.00 (0.91, 1.10) | 100.00 |

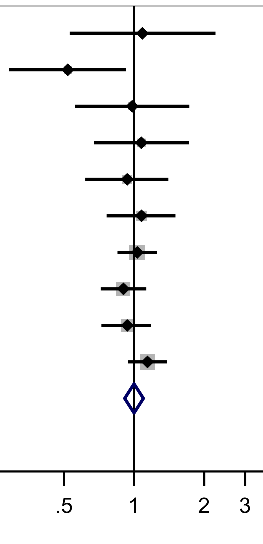

Figure legend: Forest plot shows the linear dose-response results (per 1 serving [355 mL] of carbonated sugar sweetened beverages /day) from the inverse variance DerSimonian-Laird random-effects model. The diamond represents the summary relative risk (RR) estimate, the diamond's width is the 95% confidence interval (CI). Each square and the horizontal line across the square represent the RR estimate of the individual study and its 95% CI.

Abbreviations: CI, confidence interval; RR, relative risk.

Study abbreviations: SMC, Swedish Mammography Cohort; BCDDP, Breast Cancer Detection Demonstration Project follow-up study; ATBC, Alpha-Tocopherol, Beta-Carotene Cancer Prevention Study; NLCS, Netherlands Cohort Study on Diet and Cancer; WHS, Women's Health Study; CNBSS, Canadian National Breast Screening Study; NHS, Nurses' Health Study; CPS II, Cancer Prevention Study II; IWHS, The Iowa Women's Health Study; HPFS, Health Professionals Follow-up Study

Note: Results are based on a pooled analysis of 10 cohort studies.

Zhang, 2010[182]

#### Supplementary Figure 79. Linear dose-response meta-analysis of sugar sweetened and carbonated sugar sweetened beverages and colon cancer incidence.

##### Sugar sweetened and carbonated sugar sweetened beverages, per 1 serving/day and colon cancer incidence

| Author | Study | Outcome | RR (95% CI) | Weight % |
|---|---|---|---|---|
| **Sugar sweetened beverages** | | | | |
| Kanehara, 2024 | MEC | Colon Cancer | 1.01 (0.96, 1.06) | 63.14 |
| Arthur, 2021 | CSDLH | Colon Cancer | 1.44 (0.92, 2.24) | 0.76 |
| Leung, 2021 | JPHC | Colon Cancer | 0.96 (0.88, 1.06) | 16.75 |
| Nomura, 2016 | BWHS | Colon Cancer | 1.08 (0.82, 1.44) | 1.85 |
| Subgroup, DL ($I^2$ = 15.7%, p = 0.313) | | | 1.01 (0.95, 1.07) | 82.50 |
| **Carbonated sugar sweetened beverages** | | | | |
| Zhang, 2010 | CNBSS | Colon Cancer | 1.08 (0.76, 1.50) | 1.29 |
| Zhang, 2010 | CPS II | Colon Cancer | 0.90 (0.72, 1.13) | 2.95 |
| Zhang, 2010 | WHS | Colon Cancer | 0.93 (0.62, 1.40) | 0.89 |
| Zhang, 2010 | NHS | Colon Cancer | 1.03 (0.85, 1.25) | 3.92 |
| Zhang, 2010 | NLCS | Colon Cancer | 1.07 (0.67, 1.72) | 0.68 |
| Zhang, 2010 | BCDDP | Colon Cancer | 0.52 (0.29, 0.92) | 0.45 |
| Zhang, 2010 | IWHS | Colon Cancer | 0.93 (0.72, 1.18) | 2.51 |
| Zhang, 2010 | HPFS | Colon Cancer | 1.14 (0.94, 1.38) | 4.07 |
| Zhang, 2010 | SMC | Colon Cancer | 1.09 (0.53, 2.24) | 0.29 |
| Zhang, 2010 | ATBC | Colon Cancer | 0.98 (0.56, 1.73) | 0.47 |
| Subgroup, DL ($I^2$ = 0.0%, p = 0.495) | | | 1.00 (0.91, 1.10) | 17.50 |
| Heterogeneity between groups: p = 0.877 | | | | |
| Overall, DL ($I^2$ = 0.0%, p = 0.529) | | | 1.01 (0.97, 1.05) | 100.00 |

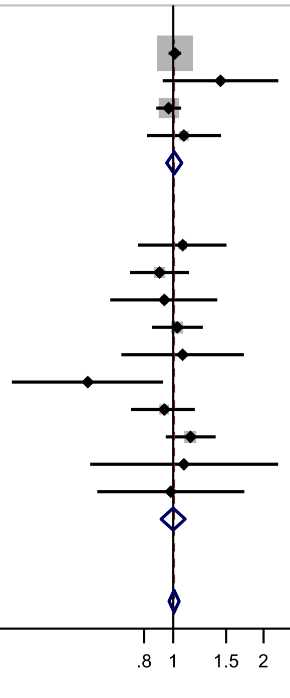

Figure legend: Forest plot shows the linear dose-response results (per 1 serving [355 mL] of sugar sweetened beverages/day) from the inverse variance DerSimonian-Laird random-effects model. The diamond represents the summary relative risk (RR) estimate, the diamond's width is the 95% confidence interval (CI). Each square and the horizontal line across the square represent the RR estimate of the individual study and its 95% CI.

Abbreviations: CI, confidence interval; RR, relative risk.

Study abbreviations: BWHS, Black Women's Health Study; JPHC, Japan Public Health Center-Based Prospective Study; CSDLH, Canadian Study of Diet, Lifestyle and Health; MEC, Multiethnic Cohort Study; SMC, Swedish Mammography Cohort; BCDDP, Breast Cancer Detection Demonstration Project follow-up study; ATBC, Alpha-Tocopherol, Beta-Carotene Cancer Prevention Study; NLCS, Netherlands Cohort Study on Diet and Cancer; WHS, Women's Health Study; CNBSS, Canadian National Breast Screening Study; NHS, Nurses' Health Study; CPS II, Cancer Prevention Study II; IWHS, The Iowa Women's Health Study; HPFS, Health Professionals Follow-up Study

Note: One additional study (UK biobank)[78] that could not be included in the dose-response meta-analysis because of insufficient data format indicated little evidence of an association (RR: 1.01, 95%CI: 0.94-1.08) per one standard deviation increase in sugary sweetened beverages intake.

Nomura, 2016[129], Leung, 2021[101], Arthur, 2021[38], Kanehara, 2024[81], Zhang, 2010[182]

**Supplementary Figure 80. Linear dose-response meta-analysis of sugar sweetened beverages and proximal colon cancer incidence.**

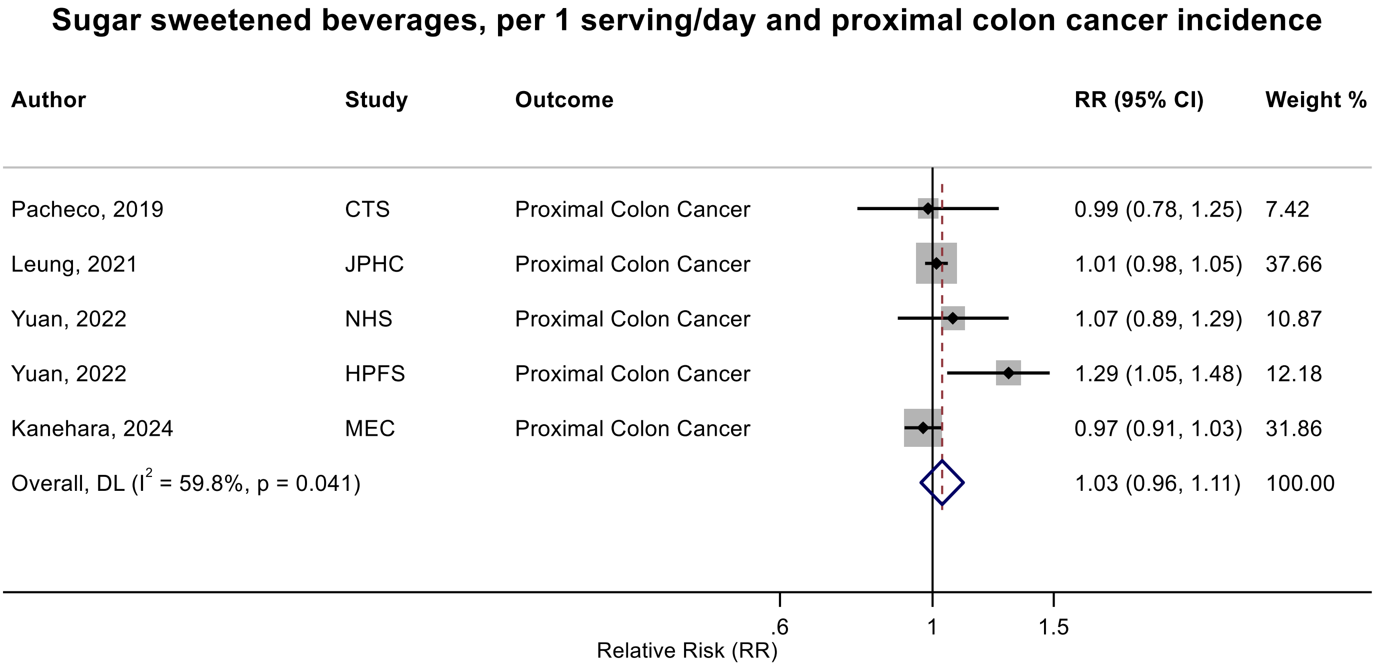

Figure legend: Forest plot shows the linear dose-response results (per 1 serving [355 mL] of sugar sweetened beverages /day) from the inverse variance DerSimonian-Laird random-effects model. The diamond represents the summary relative risk (RR) estimate, the diamond's width is the 95% confidence interval (CI). Each square and the horizontal line across the square represent the RR estimate of the individual study and its 95% CI.

Abbreviations: CI, confidence interval; RR, relative risk.

Study abbreviations: CTS, California Teachers Study; JPHC, Japan Public Health Center-Based Prospective Study; NHS, Nurses' Health Study; HPFS, Health Professionals Follow-up Study; MEC, Multiethnic Cohort Study

Pacheco, 2019[135], Leung, 2021[101], Yuan, 2022[178], Kanehara, 2024[81]

**Supplementary Figure 81. Linear dose-response meta-analysis of sugar sweetened beverages and distal colon cancer incidence.**

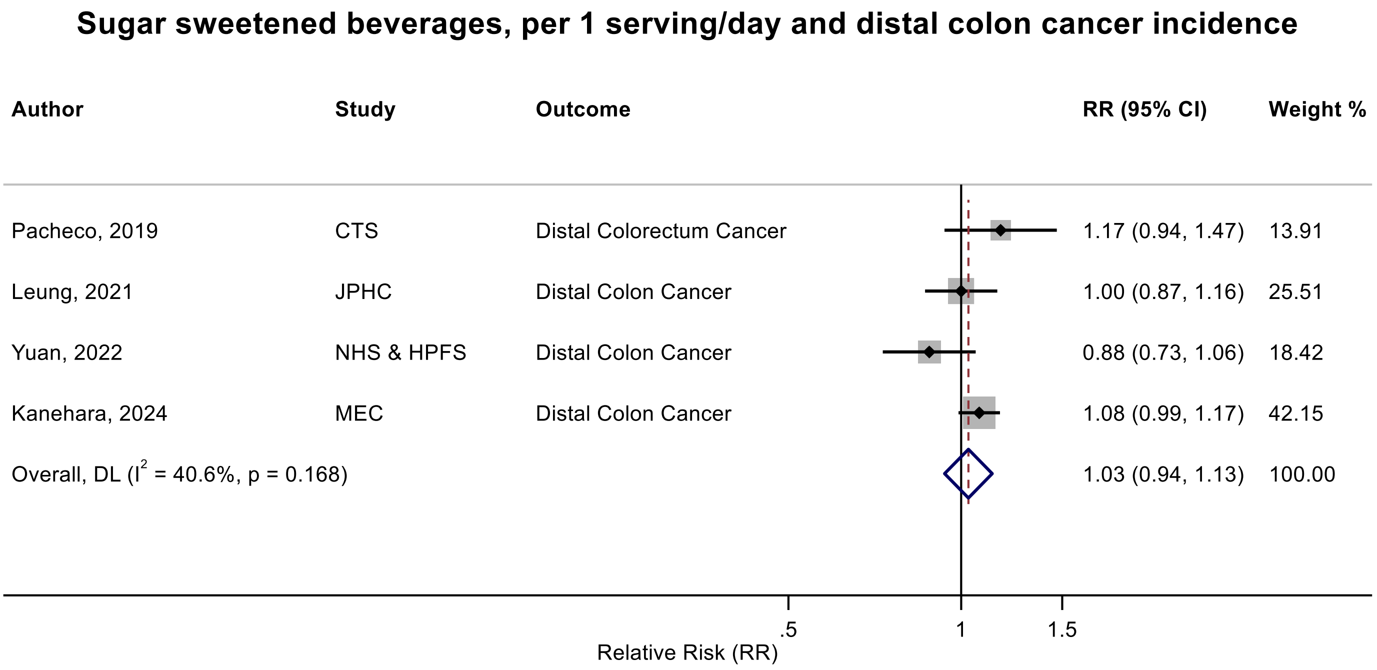

Figure legend: Forest plot shows the linear dose-response results (per 1 serving [355 mL] of sugar sweetened beverages /day) from the inverse variance DerSimonian-Laird random-effects model. The diamond represents the summary relative risk (RR) estimate, the diamond's width is the 95% confidence interval (CI). Each square and the horizontal line across the square represent the RR estimate of the individual study and its 95% CI.

Abbreviations: CI, confidence interval; RR, relative risk.

Study abbreviations: CTS, California Teachers Study; JPHC, Japan Public Health Center-Based Prospective Study; NHS & HPFS, Nurses' Health Study & Health Professionals Follow-up Study; MEC, Multiethnic Cohort Study

Pacheco, 2019[135], Leung, 2021[101], Yuan, 2022[178], Kanehara, 2024[81]

**Supplementary Figure 82. Linear dose-response meta-analysis of sugar sweetened beverages and rectal cancer incidence.**

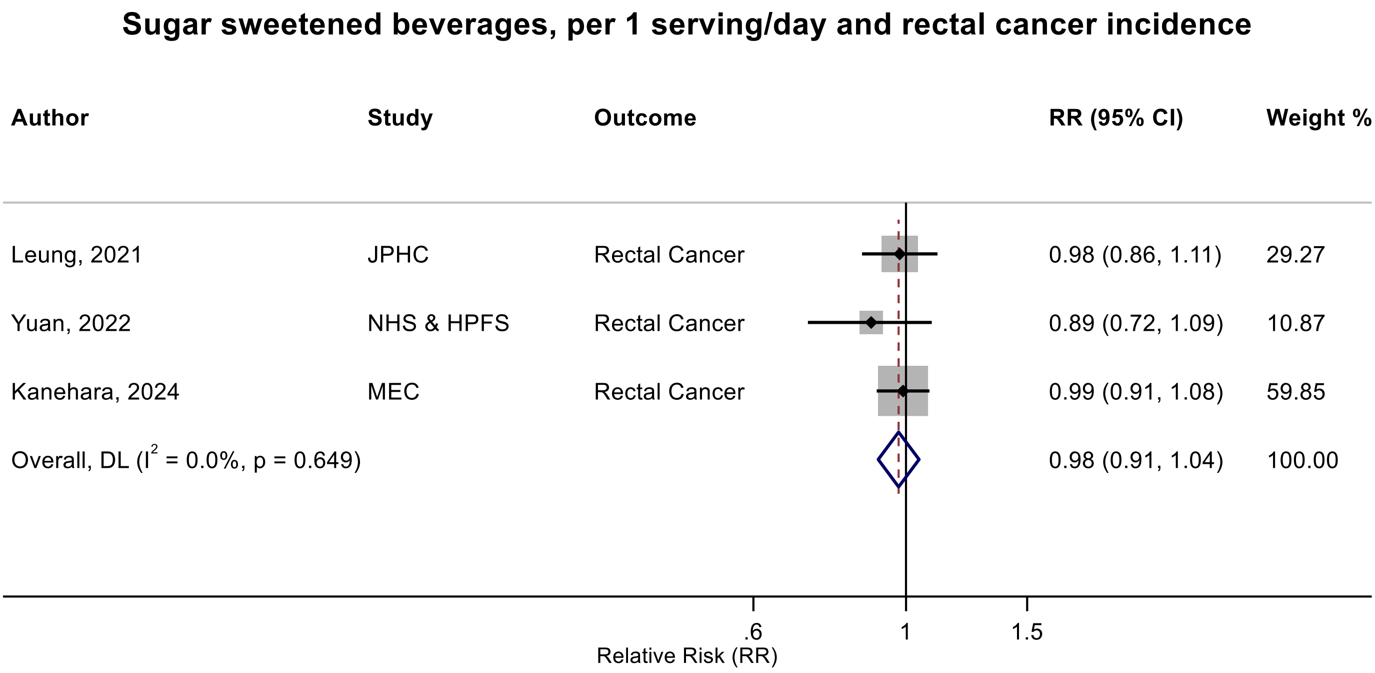

Figure legend: Forest plot shows the linear dose-response results (per 1 serving [355 mL] of sugar sweetened beverages /day) from the inverse variance DerSimonian-Laird random-effects model. The diamond represents the summary relative risk (RR) estimate, the diamond's width is the 95% confidence interval (CI). Each square and the horizontal line across the square represent the RR estimate of the individual study and its 95% CI.

Abbreviations: CI, confidence interval; RR, relative risk.

Study abbreviations: JPHC, Japan Public Health Center-Based Prospective Study; NHS & HPFS, Nurses' Health Study & Health Professionals Follow-up Study; MEC, Multiethnic Cohort Study

Note: One additional study (UK biobank)[78] that could not be included in the dose-response meta-analysis because of insufficient data format indicated little evidence of an association (RR: 1.01, 95%CI: 0.90-1.12) per one standard deviation increase in sugary sweetened beverages intake.

Leung, 2021[101], Yuan, 2022[178], Kanehara, 2024[81]

**Supplementary Figure 83. Substitution of sugar sweetened beverages with artificially sweetened beverages and proximal colon cancer incidence and mortality.**

**Substitution of SSBs with ASBs, per 1 serving/day and proximal colon cancer incidence and mortality**

| Author | Study | Outcome | Outcome Type | RR (95% CI) |
|---|---|---|---|---|
| Yuan, 2022 | NHS & HPFS | Proximal Colon Cancer | Incidence | 0.84 (0.77, 0.95) |
| Yuan, 2022 | NHS & HPFS | Proximal Colon Cancer | Mortality | 0.80 (0.71, 0.95) |

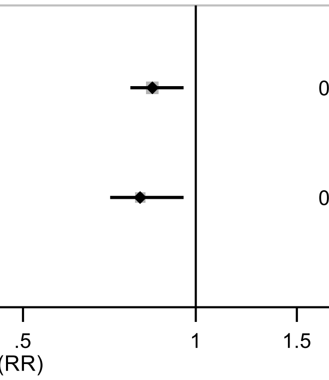

Figure legend: Forest plot shows the linear dose-response results (per substitution of 1 serving [355 mL] of sugar sweetened beverages/day with equal amount of artificially sweetened beverages) from the inverse variance DerSimonian-Laird random-effects model. The diamond represents the summary relative risk (RR) estimate, the diamond's width is the 95% confidence interval (CI). Each square and the horizontal line across the square represent the RR estimate of the individual study and its 95% CI.

Abbreviations: CI, confidence interval; RR, relative risk.

Study abbreviations: NHS & HPFS, Nurses' Health Study & Health Professionals Follow-up Study

Yuan, 2022[178]

**Supplementary Figure 84. Linear dose-response meta-analysis of sugar sweetened beverages and breast cancer incidence.**

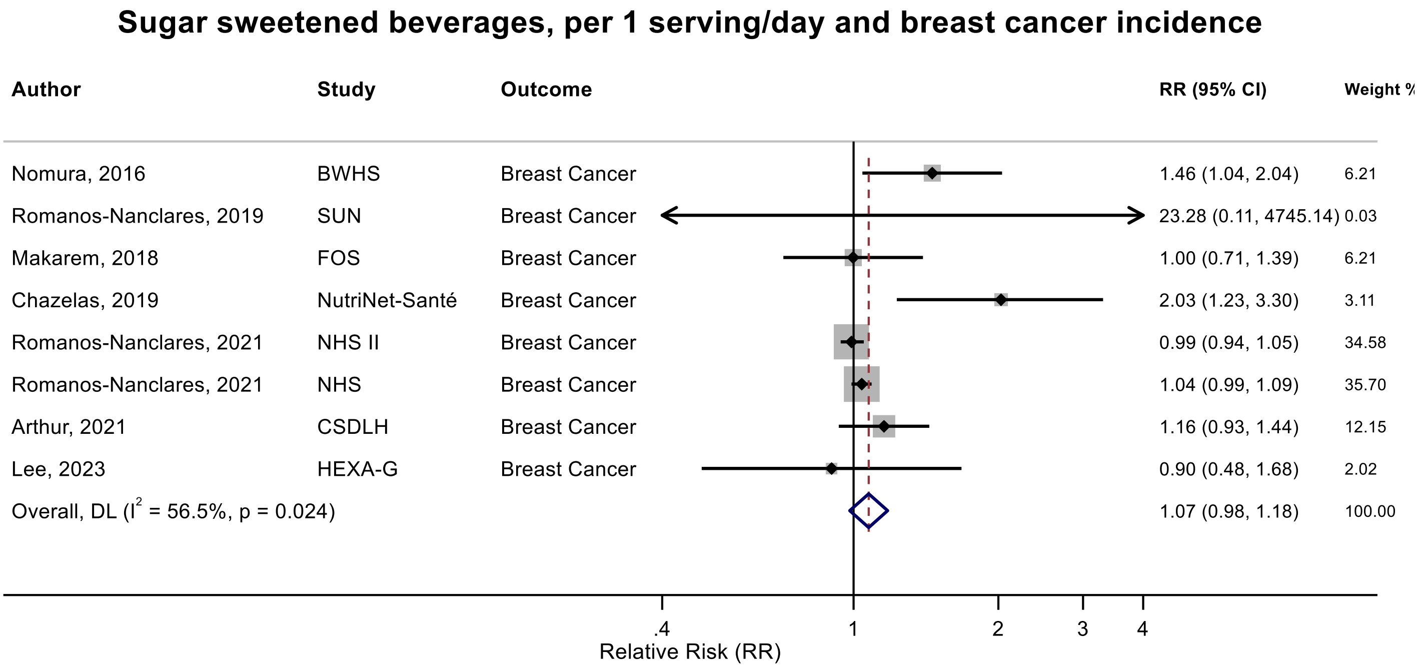

Figure legend: Forest plot shows the linear dose-response results (per 1 serving [355 mL] of sugar sweetened beverages/day) from the inverse variance DerSimonian-Laird random-effects model. The diamond represents the summary relative risk (RR) estimate, the diamond's width is the 95% confidence interval (CI). Each square and the horizontal line across the square represent the RR estimate of the individual study and its 95% CI.

Abbreviations: CI, confidence interval; RR, relative risk.

Study abbreviations: BWHS, Black Women's Health Study; SUN, Seguimiento Universidad de Navarra Cohort Study; FOS, Framingham Heart Study-Offspring Cohort; NHS II, Nurses' Health Study II; NHS, Nurses' Health Study; CSDLH, Canadian Study of Diet, Lifestyle and Health; HEXA-G, Health Examinees-Gem study

Note: One additional study (National Health Interview Survey)[169] could not be included in the dose-response meta-analysis due to insufficient format of data reported little evidence of an association for risk of breast cancer incidence with the consumption of sugar sweetened beverages ($RR_{\text{intake vs no intake}}$: 1.46, 95%CI: 0.96-2.21), which was consistent to other studies in general.

Nomura, 2016[128], Romanos-Nanclares, 2019[146], Makarem, 2018[108], Chazelas, 2019[44], Romanos-Nanclares, 2021[145], Arthur, 2021[38], Lee, 2023[97]

**Supplementary Figure 85. Linear dose-response meta-analysis of sugar sweetened beverages and breast cancer incidence by menopausal status.**

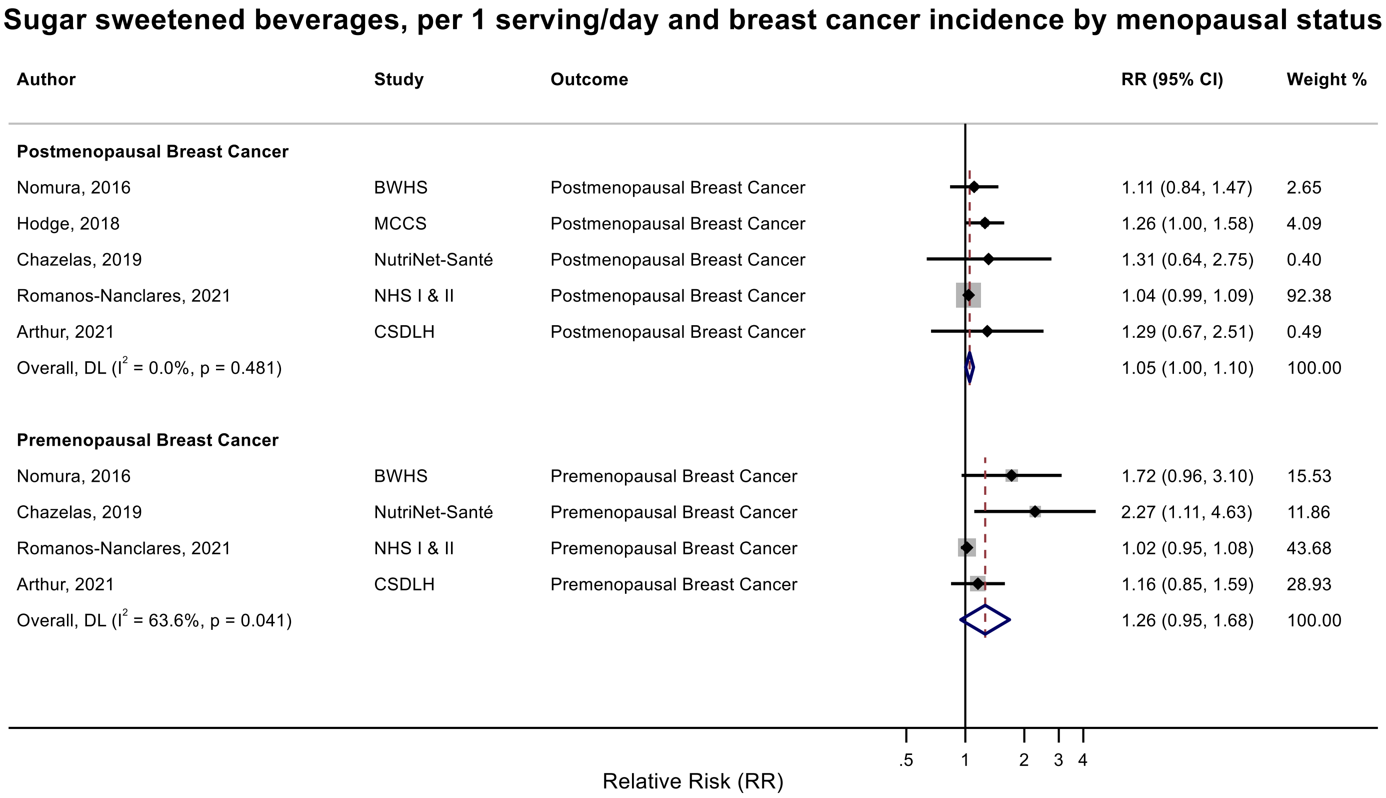

Figure legend: Forest plot shows the linear dose-response results (per 1 serving [355 mL] of sugar sweetened beverages /day) from the inverse variance DerSimonian-Laird random-effects model. The diamond represents the summary relative risk (RR) estimate, the diamond's width is the 95% confidence interval (CI). Each square and the horizontal line across the square represent the RR estimate of the individual study and its 95% CI.

Abbreviations: CI, confidence interval; RR, relative risk.

Study abbreviations: BWHS, Black Women's Health Study; MCCS, Melbourne Collaborative Cohort Study; NHS I and II, Nurses' Health Study I and II; CSDLH, Canadian Study of Diet, Lifestyle and Health

Note: The pooled RR and 95%CI of postmenopausal breast cancer with 3 decimal point are 1.052 (1.005-1.102). One additional study (IWHS) that could not be included in the dose-response meta-analysis because of insufficient data format reported little evidence of a difference in risk of postmenopausal breast cancer for those who adhered to WCRF sugar beverages guideline (0 g/day) compared with those who did not adhere to the guideline (≥250 g/day) (risk difference 0.001, 95% CI:-0.010 to 0.013, P=0.80)[130]. Another study (SUN)[146] that could not be included in the dose-response meta-analysis because of insufficient data format reported an increased risk of postmenopausal breast cancer incidence with a higher sugar sweetened beverages intake ($RR_{0.05\ serving/day\ vs\ 0}$: 2.12, 95%CI: 1.01-4.41). The SUN cohort reported no association with premenopausal breast cancer incidence ($RR_{0.05\ serving/day\ vs\ 0}$: 1.16, 95%CI: 0.66-2.07).

Nomura, 2016[128], Hodge, 2018[70], Chazelas, 2019[44], Romanos-Nanclares, 2021[145], Arthur, 2021[38]

**Supplementary Figure 86. Linear dose-response meta-analysis of sugar sweetened beverages and breast cancer subtypes incidence.**

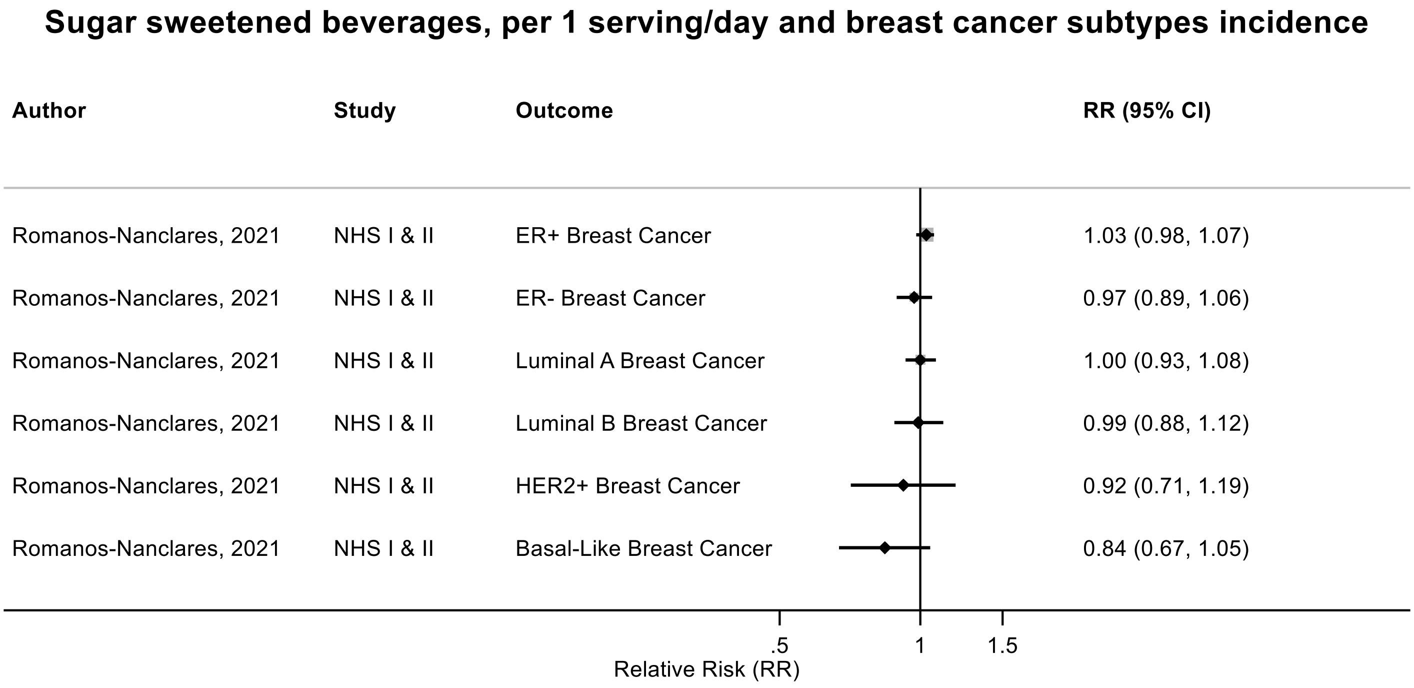

Figure legend: Forest plot shows the linear dose-response results (per 1 serving [355 mL] of sugar sweetened beverages /day) from the inverse variance DerSimonian-Laird random-effects model. The diamond represents the summary relative risk (RR) estimate, the diamond's width is the 95% confidence interval (CI). Each square and the horizontal line across the square represent the RR estimate of the individual study and its 95% CI.

Abbreviations: CI, confidence interval; RR, relative risk.

Study abbreviations: NHS I & II, Nurses' Health Study I & II

Romanos-Nanclares, 2021[145]

**Supplementary Figure 87. Linear dose-response meta-analysis of sugar sweetened beverages and kidney cancer incidence.**

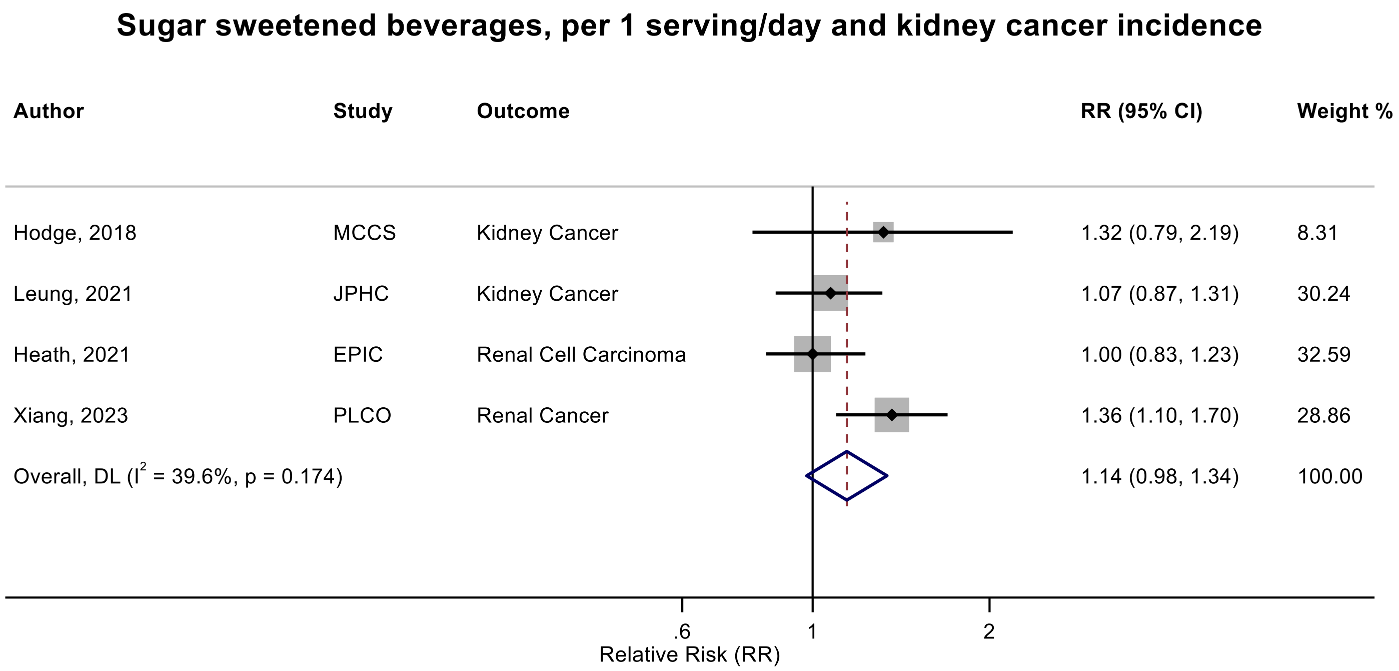

Figure legend: Forest plot shows the linear dose-response results (per 1 serving [355 mL] of sugar sweetened beverages /day) from the inverse variance DerSimonian-Laird random-effects model. The diamond represents the summary relative risk (RR) estimate, the diamond's width is the 95% confidence interval (CI). Each square and the horizontal line across the square represent the RR estimate of the individual study and its 95% CI.

Abbreviations: CI, confidence interval; RR, relative risk.

Study abbreviations: MCCS, Melbourne Collaborative Cohort Study; JPHC, Japan Public Health Center-Based Prospective Study; EPIC, European Prospective Investigation into Cancer and nutrition; PLCO, Prostate, Lung, Colorectal and Ovarian Cancer Screening trial

Hodge, 2018[70], Leung, 2021[100], Heath, 2021[66], Xiang, 2023[173]

**Supplementary Figure 88. Linear dose-response meta-analysis of sugar sweetened and carbonated sugar sweetened beverages and kidney cancer incidence.**

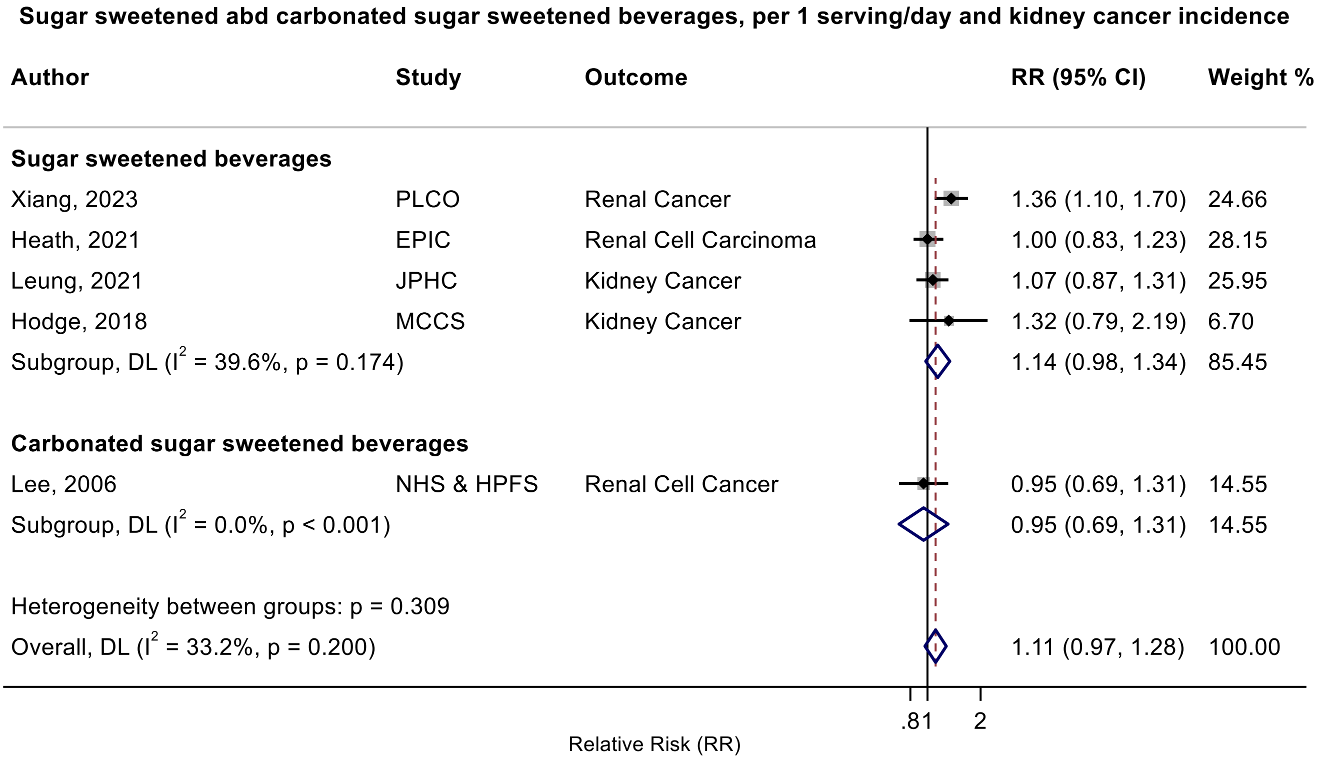

Figure legend: Forest plot shows the linear dose-response results (per 1 serving [355 mL] of sugar sweetened beverages /day) from the inverse variance DerSimonian-Laird random-effects model. The diamond represents the summary relative risk (RR) estimate, the diamond's width is the 95% confidence interval (CI). Each square and the horizontal line across the square represent the RR estimate of the individual study and its 95% CI.

Abbreviations: CI, confidence interval; RR, relative risk.

Study abbreviations: MCCS, Melbourne Collaborative Cohort Study; JPHC, Japan Public Health Center-Based Prospective Study; EPIC, European Prospective Investigation into Cancer and nutrition; PLCO, Prostate, Lung, Colorectal and Ovarian Cancer Screening trial; NHS, Nurse’s Health study; HPFs, Health Professionals Follow-up Study

Hodge, 2018[70], Leung, 2021[100], Heath, 2021[66], Xiang, 2023[173], Lee, 2006[98]

**Supplementary Figure 89. Linear dose-response meta-analysis of sugar sweetened beverages and lung cancer incidence.**

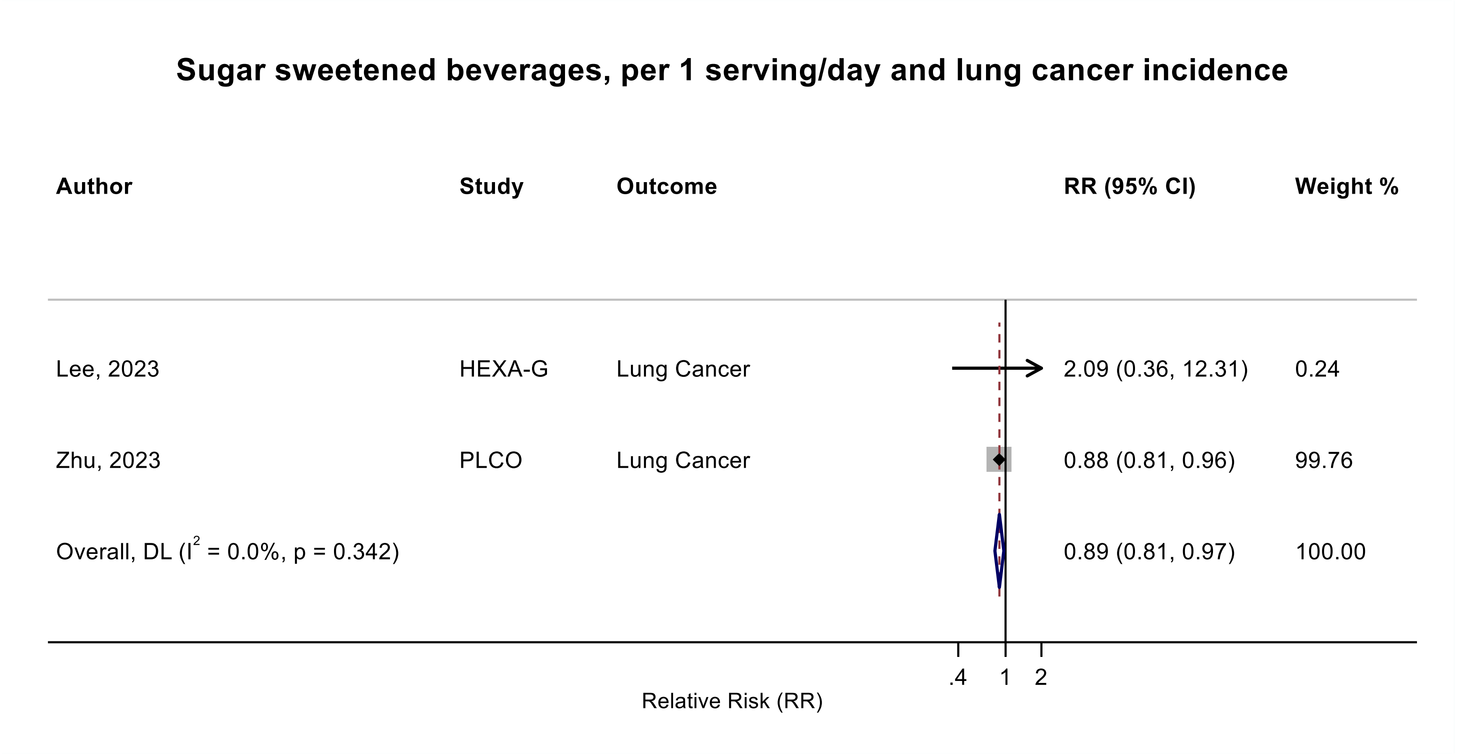

Figure legend: Forest plot shows the linear dose-response results (per 1 serving [355 mL] of sugar sweetened beverages/day) from the inverse variance DerSimonian-Laird random-effects model. The diamond represents the summary relative risk (RR) estimate, the diamond's width is the 95% confidence interval (CI). Each square and the horizontal line across the square represent the RR estimate of the individual study and its 95% CI.

Abbreviations: CI, confidence interval; RR, relative risk.

Study abbreviations: HEXA-G, Health Examinees-Gem study; PLCO, Prostate, Lung, Colorectal and Ovarian Cancer Screening trial

Note: One additional study (NIH-AARP)[37] that could not be included in the dose-response meta-analysis because of insufficient data format reported no association or a borderline positive association between sugar sweetened beverages intake and lung cancer incidence per a one-unit change in the component score of the Healthy Eating Index-2010 (RR: 1.00, 95%CI: 0.99-1.00), Alternative Healthy Eating Index-2010 (RR: 1.01, 95%CI: 1.00-1.02), and Dietary Approaches to Stop Hypertension (RR: 1.03, 95%CI: 1.00-1.05).

Lee, 2023[97], Zhu, 2023[190]

**Supplementary Figure 90. Linear dose-response meta-analysis of sugar sweetened beverages and lung cancer incidence and mortality combined.**

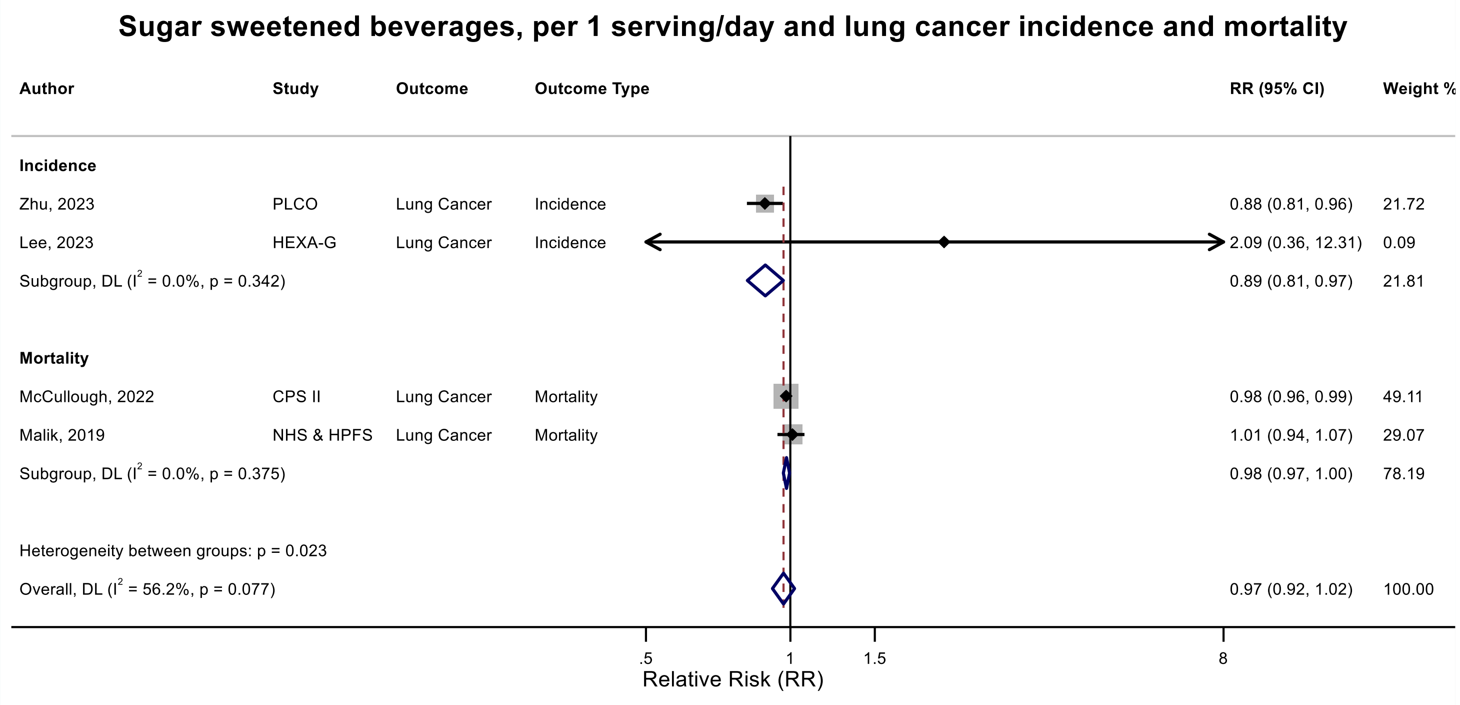

Figure legend: Forest plot shows the linear dose-response results (per 1 serving [355 mL] of sugar sweetened beverages/day) from the inverse variance DerSimonian-Laird random-effects model. The diamond represents the summary relative risk (RR) estimate, the diamond's width is the 95% confidence interval (CI). Each square and the horizontal line across the square represent the RR estimate of the individual study and its 95% CI.

Abbreviations: CI, confidence interval; RR, relative risk.

Study abbreviations: HEXA-G, Health Examinees-Gem study; PLCO, Prostate, Lung, Colorectal and Ovarian Cancer Screening trial, NHS & HPFS, Nurses' Health Study & Health Professionals Follow-up Study; CPS II, Cancer Prevention Study II

Note: One additional study (NIH-AARP)[178] that could not be included in the dose-response meta-analysis because of insufficient data format reported no association or a borderline positive association between sugar sweetened beverages intake and lung cancer incidence per a one-unit change in the component score of the Healthy Eating Index-2010 (RR: 1.00, 95%CI: 0.99-1.00), Alternative Healthy Eating Index-2010 (RR: 1.01, 95%CI: 1.00-1.02), and Dietary Approaches to Stop Hypertension (RR: 1.03, 95%CI: 1.00-1.05).

Lee, 2023[97], Zhu, 2023[190], Malik, 2019[109], McCullough, 2022[113]

**Supplementary Figure 91. Linear dose-response meta-analysis of sugar sweetened beverages and stomach cancer incidence.**

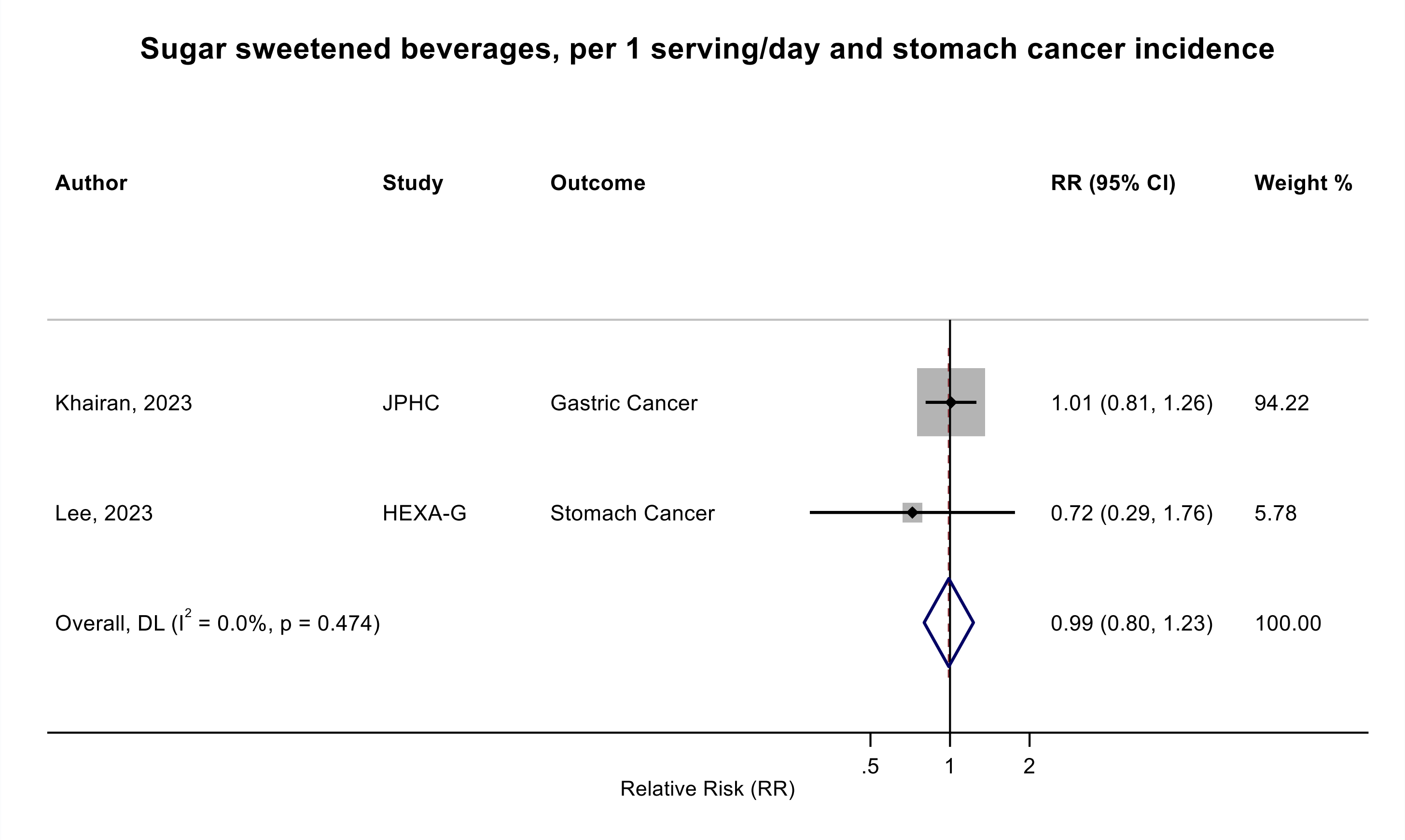

Figure legend: Forest plot shows the linear dose-response results (per 1 serving [355 mL] of sugar sweetened beverages /day) from the inverse variance DerSimonian-Laird random-effects model. The diamond represents the summary relative risk (RR) estimate, the diamond's width is the 95% confidence interval (CI). Each square and the horizontal line across the square represent the RR estimate of the individual study and its 95% CI.

Abbreviations: CI, confidence interval; RR, relative risk.

Study abbreviations: JPHC, Japan Public Health Center-Based Prospective Study; HEXA-G, Health Examinees-Gem study

Khairan, 2023[84], Lee, 2023[97]

**Supplementary Figure 92. Linear dose-response meta-analysis of sugar sweetened beverages and stomach cardia cancer incidence.**

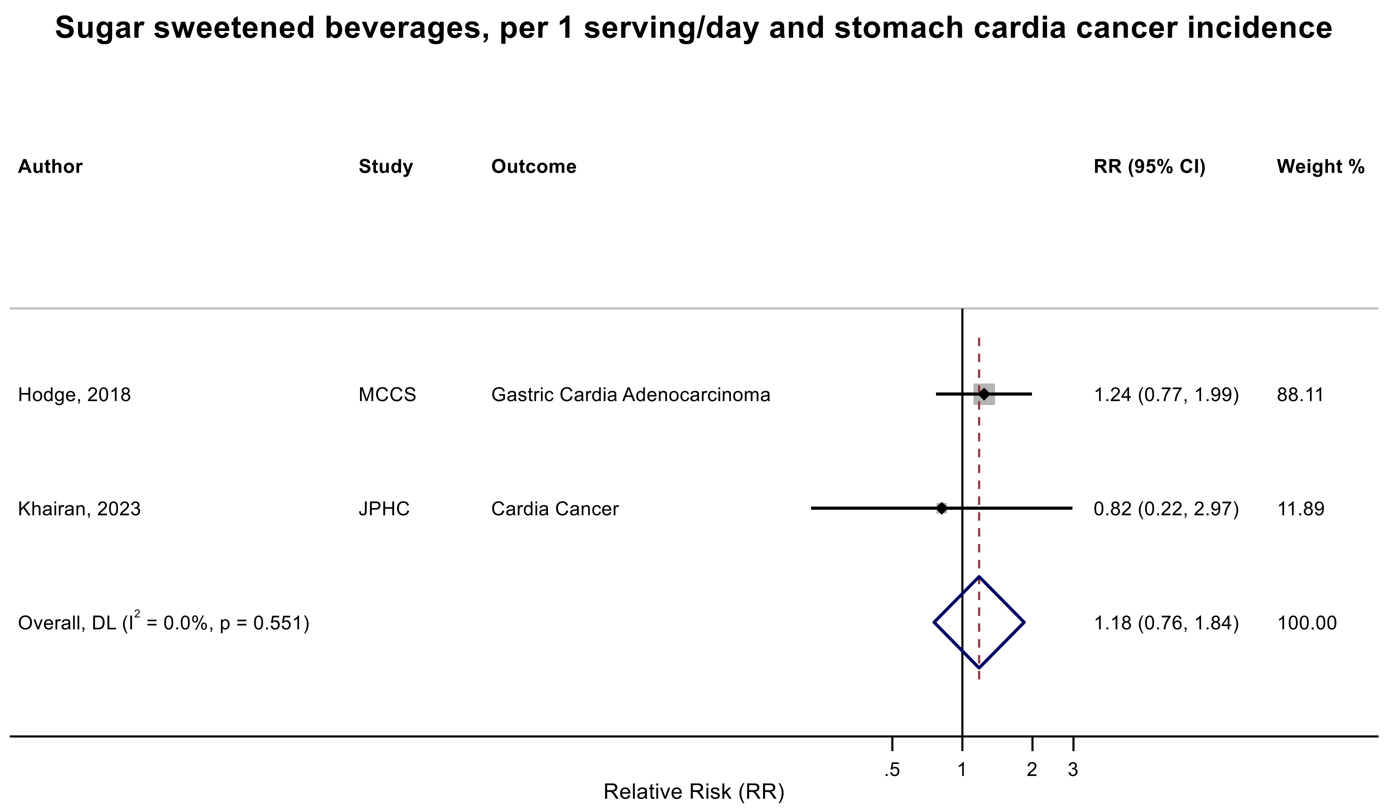

Figure legend: Forest plot shows the linear dose-response results (per 1 serving [355 mL] of sugar sweetened beverages /day) from the inverse variance DerSimonian-Laird random-effects model. The diamond represents the summary relative risk (RR) estimate, the diamond's width is the 95% confidence interval (CI). Each square and the horizontal line across the square represent the RR estimate of the individual study and its 95% CI.

Abbreviations: CI, confidence interval; RR, relative risk.

Study abbreviations: MCCS, Melbourne Collaborative Cohort Study; JPHC, Japan Public Health Center-Based Prospective Study

Hodge, 2018[70], Khairan, 2023[84]

**Supplementary Figure 93. Linear dose-response meta-analysis of sugar sweetened beverages and liver cancer incidence.**

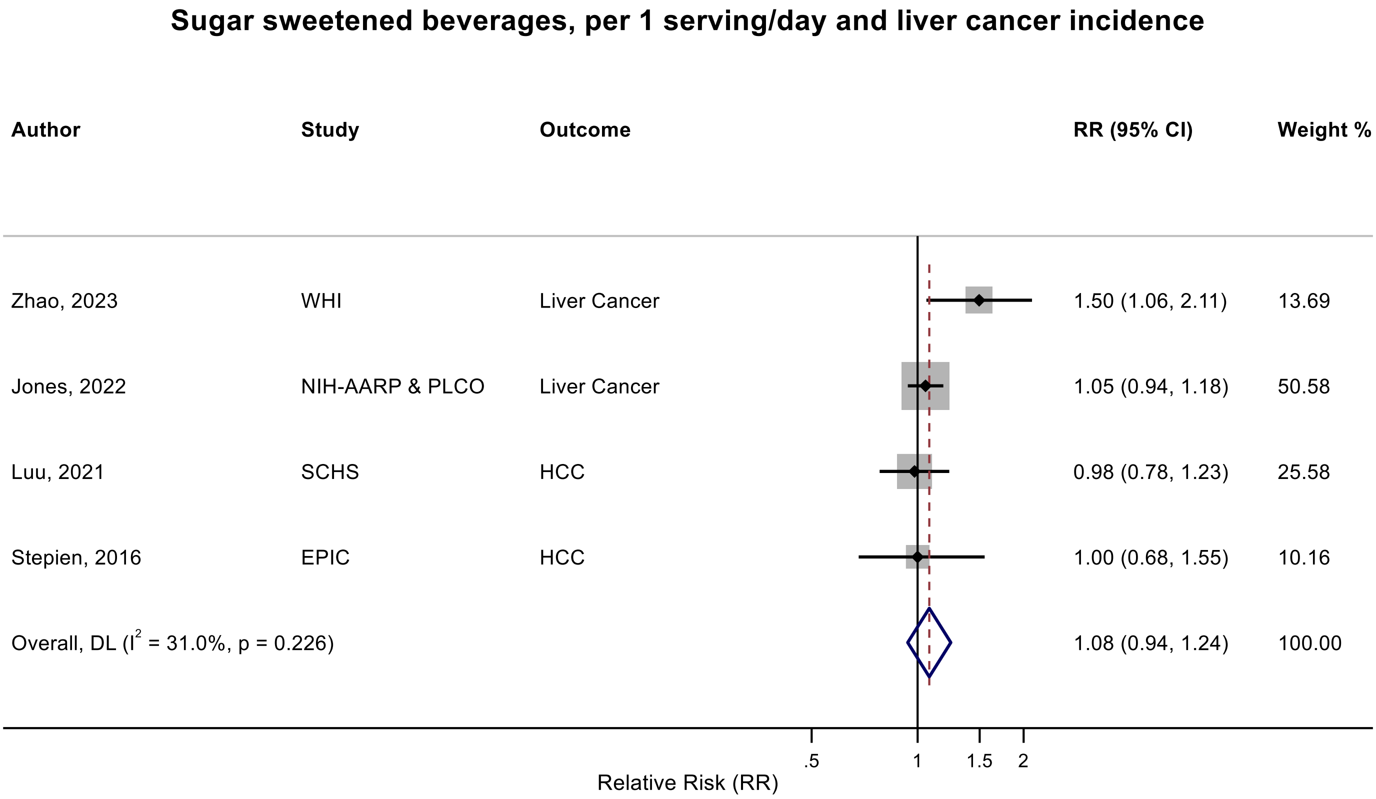

Figure legend: Forest plot shows the linear dose-response results (per 1 serving [355 mL] of sugar sweetened beverages /day) from the inverse variance DerSimonian-Laird random-effects model. The diamond represents the summary relative risk (RR) estimate, the diamond's width is the 95% confidence interval (CI). Each square and the horizontal line across the square represent the RR estimate of the individual study and its 95% CI.

Abbreviations: CI, confidence interval; HCC, hepatocellular carcinoma, relative risk.

Study abbreviations: EPIC, European Prospective Investigation into Cancer and nutrition; SCHS, Singapore Chinese Health study; NIH-AARP & PLCO, National Institute of Health + American Association of Retired Persons Diet and Health Study & Prostate, Lung, Colorectal and Ovarian Cancer Screening trial; WHI, Women's Health Initiative Study

Note: One additional pooled analysis of the Nurse's Health Study and Health Professionals Follow-up Study that could not be included in the dose-response meta-analysis because of the insufficient data format reported little evidence of an association for sugar sweetened beverages ($RR_{Q4\ vs\ Q1}$: 1.26, 95%CI: 0.79-2.01)[105] and sugar sweetened beverages plus fruit juices, as part of the Alternative Healthy Eating Index-2010 score (RR per 1-point change in the score for the component: 0.99, 95%CI: 0.93 – 1.06)[222].

Stepien, 2016[159], Luu, 2021[106], Jones, 2022[79], Zhao, 2023[186]

**Supplementary Figure 94. Linear dose-response meta-analysis of sugar sweetened beverages and liver cancer incidence and mortality combined.**

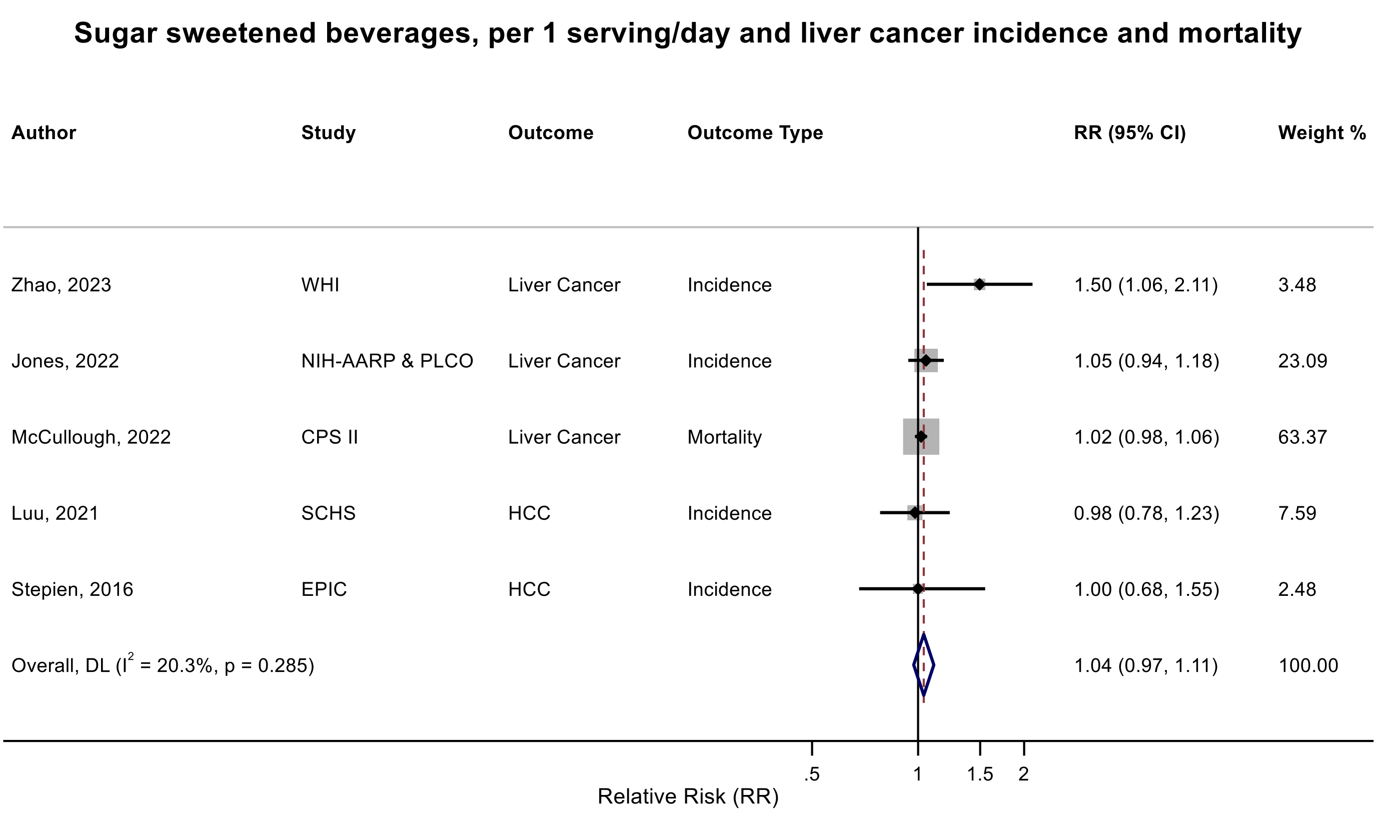

Figure legend: Forest plot shows the linear dose-response results (per 1 serving [355 mL] of sugar sweetened beverages /day) from the inverse variance DerSimonian-Laird random-effects model. The diamond represents the summary relative risk (RR) estimate, the diamond's width is the 95% confidence interval (CI). Each square and the horizontal line across the square represent the RR estimate of the individual study and its 95% CI.

Abbreviations: CI, confidence interval; HCC, hepatocellular carcinoma, RR, relative risk.

Study abbreviations: EPIC, European Prospective Investigation into Cancer and nutrition; SCHS, Singapore Chinese Health study; NIH-AARP & PLCO, National Institute of Health + American Association of Retired Persons Diet and Health Study & Prostate, Lung, Colorectal and Ovarian Cancer Screening trial; WHI, Women's Health Initiative Study; CPS II; Cancer Prevention Study II

Zhao, 2022[186], Jones, 2022[79], McCullough, 2022[113], Luu, 2021[106], Stepien, 2016[159]

**Supplementary Figure 95. Linear dose-response meta-analysis of sugar sweetened beverages components and liver cancer incidence.**

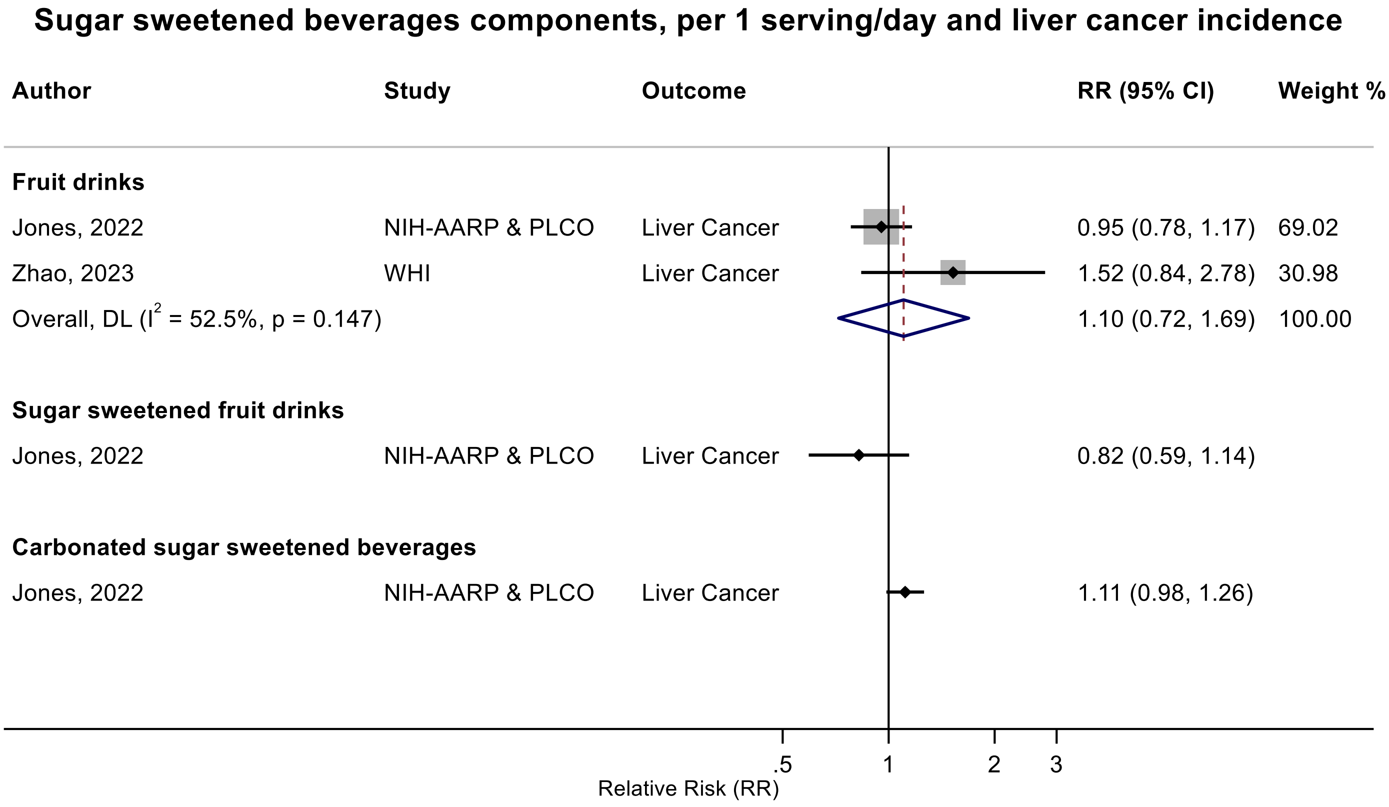

Figure legend: Forest plot shows the linear dose-response results (per 1 serving [355 mL] of sugar sweetened beverages components/day) from the inverse variance DerSimonian-Laird random-effects model. The diamond represents the summary relative risk (RR) estimate, the diamond's width is the 95% confidence interval (CI). Each square and the horizontal line across the square represent the RR estimate of the individual study and its 95% CI.

Abbreviations: CI, confidence interval; RR, relative risk.

Study abbreviations: NIH-AARP & PLCO, National Institute of Health + American Association of Retired Persons Diet and Health Study & Prostate, Lung, Colorectal and Ovarian Cancer Screening trial; WHI, Women's Health Initiative Study

Note: One additional study (The Southern Community Cohort)[187] that could not be included in the dose-response meta-analysis because of insufficient data format indicated a borderline inverse association between carbonated sugar sweetened beverages and liver cancer ($RR_{\geq median\ vs\ non\text{-}consumers}$: 0.78, 95%CI: 0.58-1.05), and a positive association but with 95%CI crossing the null value between sweetened fruit drinks and liver cancer ($RR_{\geq median\ vs\ non\text{-}consumers}$: 1.11, 95%CI: 0.86-1.45).

Jones, 2022[79], Zhao, 2023[186]

**Supplementary Figure 96. Linear dose-response meta-analysis of sugar sweetened cola and pancreatic cancer incidence.**

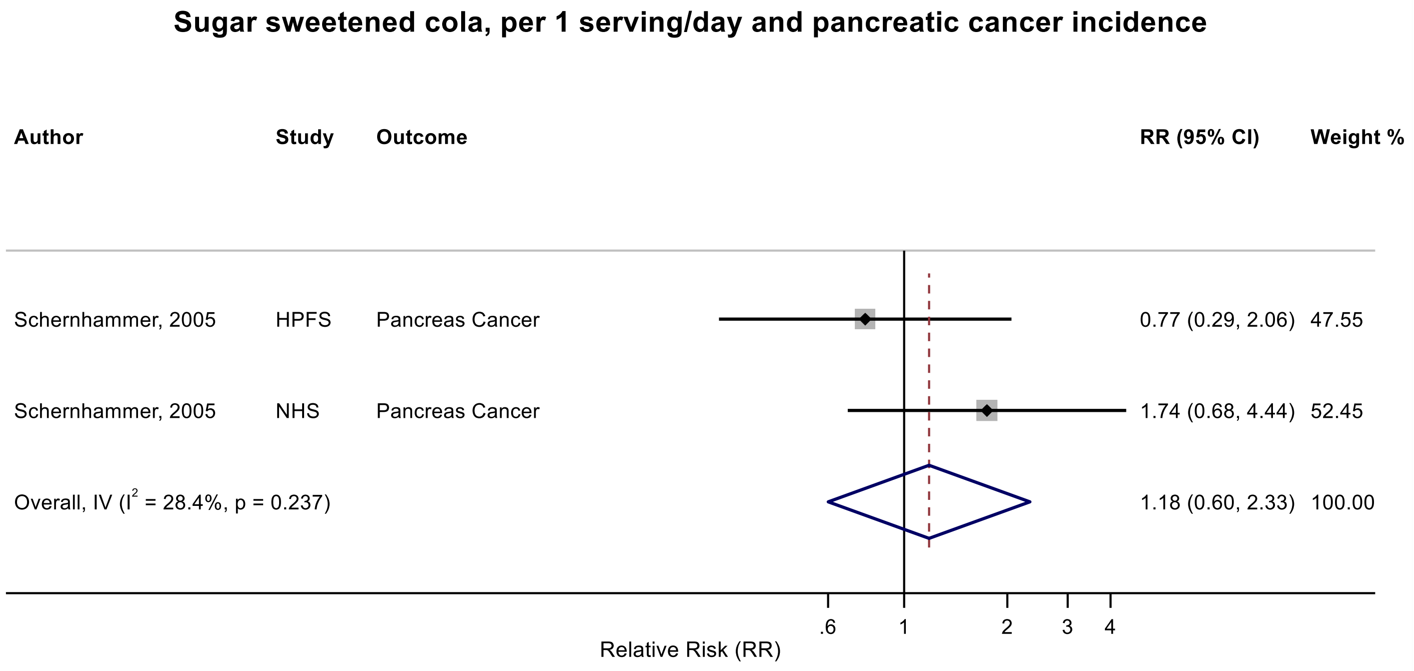

Figure legend: Forest plot shows the linear dose-response results (per 1 serving [355 mL] of sugar sweetened cola/day) from the inverse variance DerSimonian-Laird random-effects model. The diamond represents the summary relative risk (RR) estimate, the diamond's width is the 95% confidence interval (CI). Each square and the horizontal line across the square represent the RR estimate of the individual study and its 95% CI.

Abbreviations: CI, confidence interval; RR, relative risk.

Study abbreviations: HPFS, Health Professionals Follow-up Study; NHS, Nurses' Health Study

Schernhammer, 2005[151]

**Supplementary Figure 97. Linear dose-response meta-analysis of sugar sweetened beverages and type I endometrial cancer incidence.**

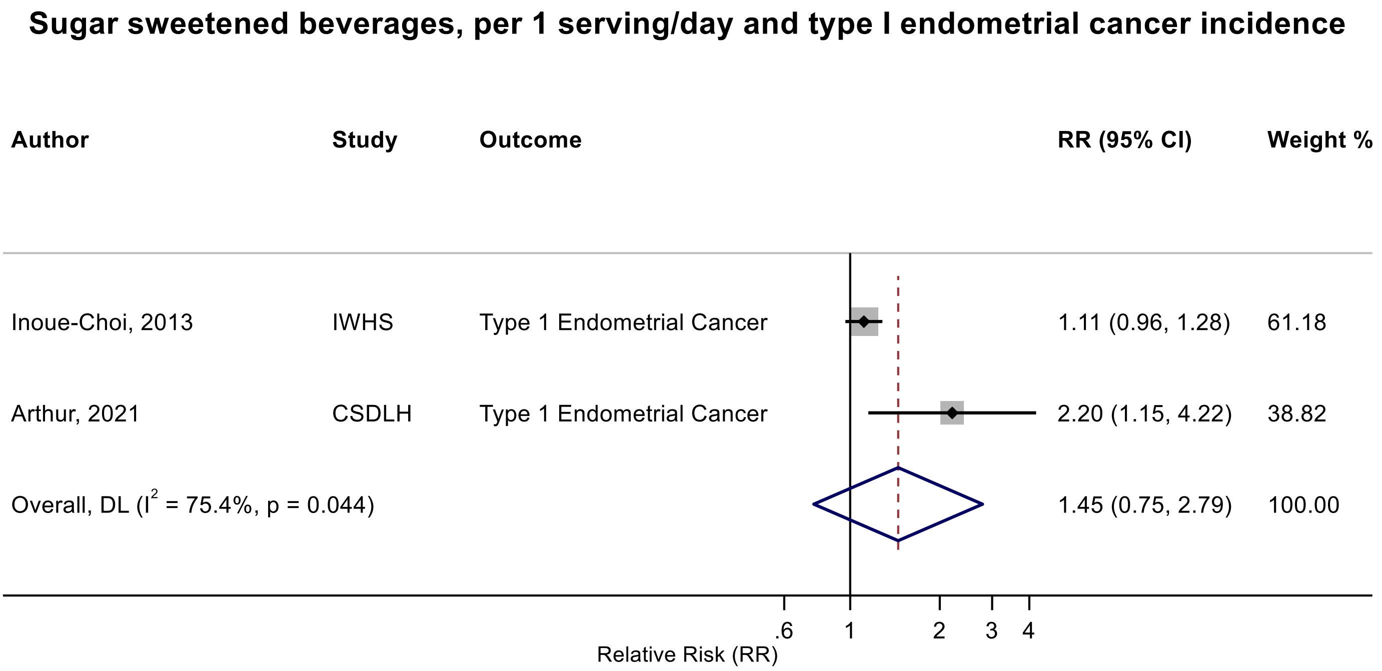

Figure legend: Forest plot shows the linear dose-response results (per 1 serving [355 mL] of sugar sweetened beverages /day) from the inverse variance DerSimonian-Laird random-effects model. The diamond represents the summary relative risk (RR) estimate, the diamond's width is the 95% confidence interval (CI). Each square and the horizontal line across the square represent the RR estimate of the individual study and its 95% CI.

Abbreviations: CI, confidence interval; RR, relative risk.

Study abbreviations: IWHS, The Iowa Women's Health Study; CSDLH, Canadian Study of Diet, Lifestyle and Health

Inoue-Choi, 2013[74], Arthur, 2021[38]

**Supplementary Figure 98. Linear dose-response meta-analysis of sugar sweetened beverages and prostate cancer incidence.**

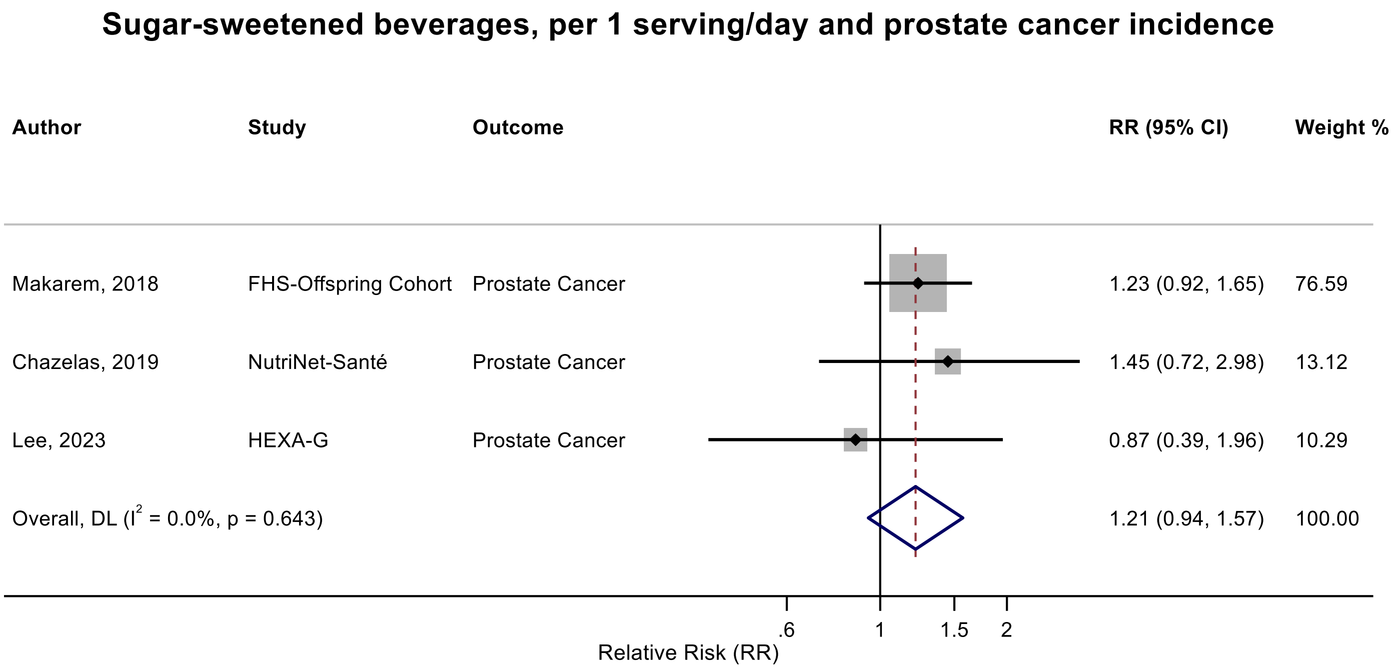

Figure legend: Forest plot shows the linear dose-response results (per 1 serving [355 mL] of sugar sweetened beverages /day) from the inverse variance DerSimonian-Laird random-effects model. The diamond represents the summary relative risk (RR) estimate, the diamond's width is the 95% confidence interval (CI). Each square and the horizontal line across the square represent the RR estimate of the individual study and its 95% CI.

Abbreviations: CI, confidence interval; RR, relative risk.

Study abbreviations: FHS-Offspring Cohort, Framingham Heart Study-Offspring Cohort; HEXA-G, Health Examinees-Gem study

Note: One additional study (Malmö Diet and Cancer cohort)[47] that could not be included in the dose-response meta-analysis because of insufficient data format reported a borderline increased risk of prostate cancer incidence with a higher intake of sugar sweetened beverages (RR $_{297.8 \text{ vs } 0 \text{ g/day}}$: 1.13, 95%CI: 0.92-1.38).

Makarem, 2018[108], Chazelas, 2019[44], Lee, 2023[97]

#### Supplementary Figure 99. Categorical highest versus the lowest meta-analysis of sugar sweetened beverages and high-risk/aggressive prostate cancer incidence.

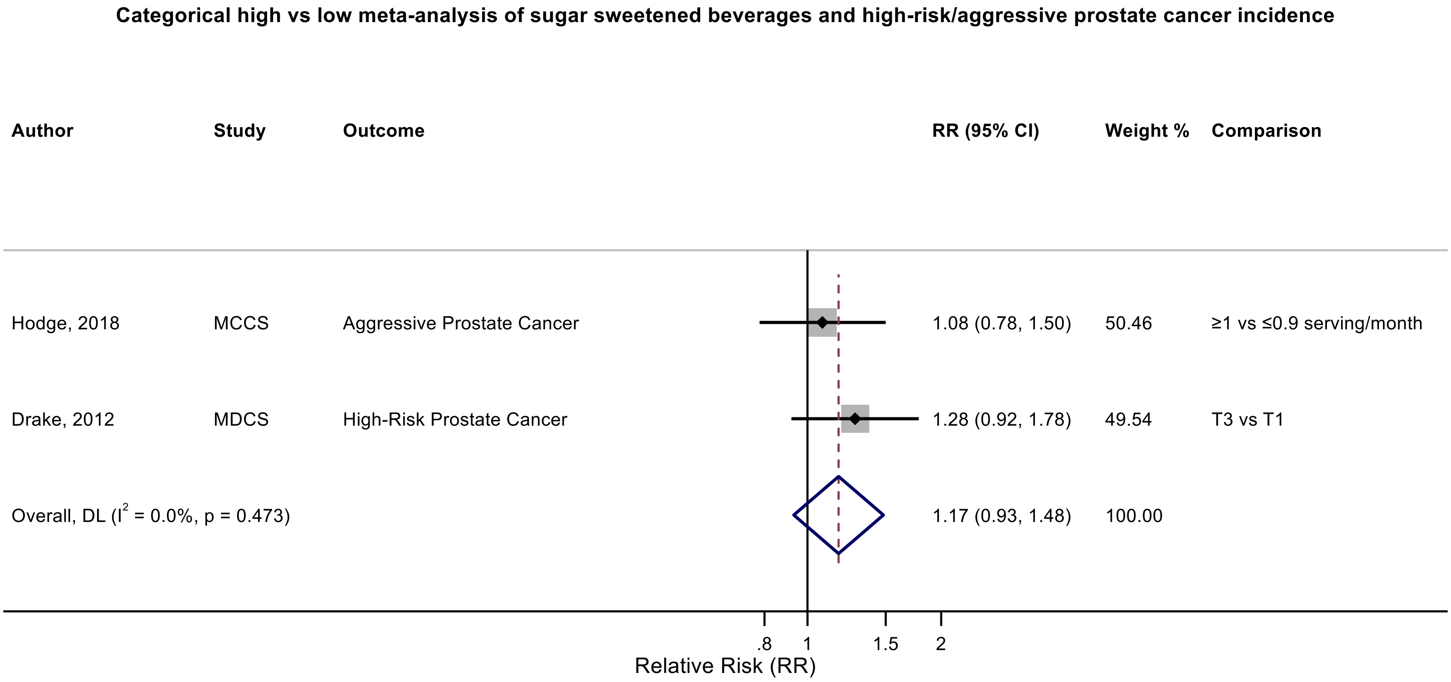

Figure legend: Forest plot shows the highest versus the lowest category results of sugar sweetened beverages from the inverse variance DerSimonian-Laird random-effects model. The diamond represents the summary relative risk (RR) estimate, the diamond's width is the 95% confidence interval (CI). Each square and the horizontal line across the square represent the RR estimate of the individual study and its 95% CI.

Abbreviations: CI, confidence interval; F, female, M, male, RR, relative risk.

Study abbreviations: MCCS, Melborn Collaborative Cohort Study; MDCS, Malmo Diet and Cancer Study

Hodge, 2018[70], Drake 2012[47]

Note: This analysis was conducted as an additional analysis combining high grade, aggressive and advanced prostate cancer cases. One study (MDCS) was not eligible for the linear dose-response meta-analysis, so we performed a categorical high vs low meta-analysis.

**Supplementary Figure 100. Categorical highest versus the lowest meta-analysis of sugar sweetened beverages and bladder cancer incidence.**

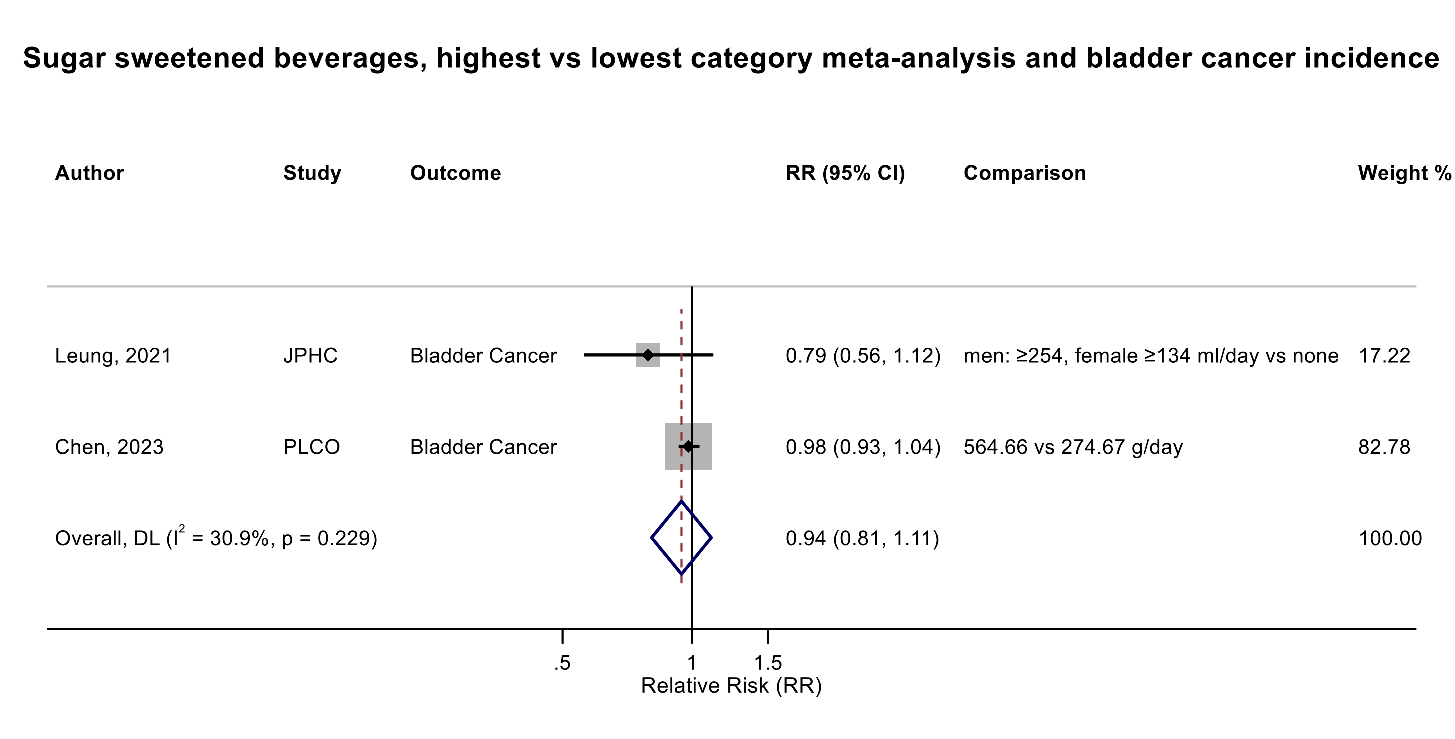

Figure legend: Forest plot shows the highest versus the lowest category results of sugar sweetened beverages from the inverse variance DerSimonian-Laird random-effects model. The diamond represents the summary relative risk (RR) estimate, the diamond's width is the 95% confidence interval (CI). Each square and the horizontal line across the square represent the RR estimate of the individual study and its 95% CI.

Abbreviations: CI, confidence interval; F, female, M, male, RR, relative risk.

Study abbreviations: JPHC, Japan Public Health Center-Based Prospective Study; PLCO, Prostate, Lung, Colorectal and Ovarian Cancer Screening trial

Leung, 2021[173], Chen, 2023[46]

Note: JPHC study was not eligible for the linear dose-response meta-analysis, so we performed a categorical high vs low meta-analysis.

**Supplementary Figure 101. Linear dose-response meta-analysis of carbonated sugar sweetened beverages and leukaemia incidence.**

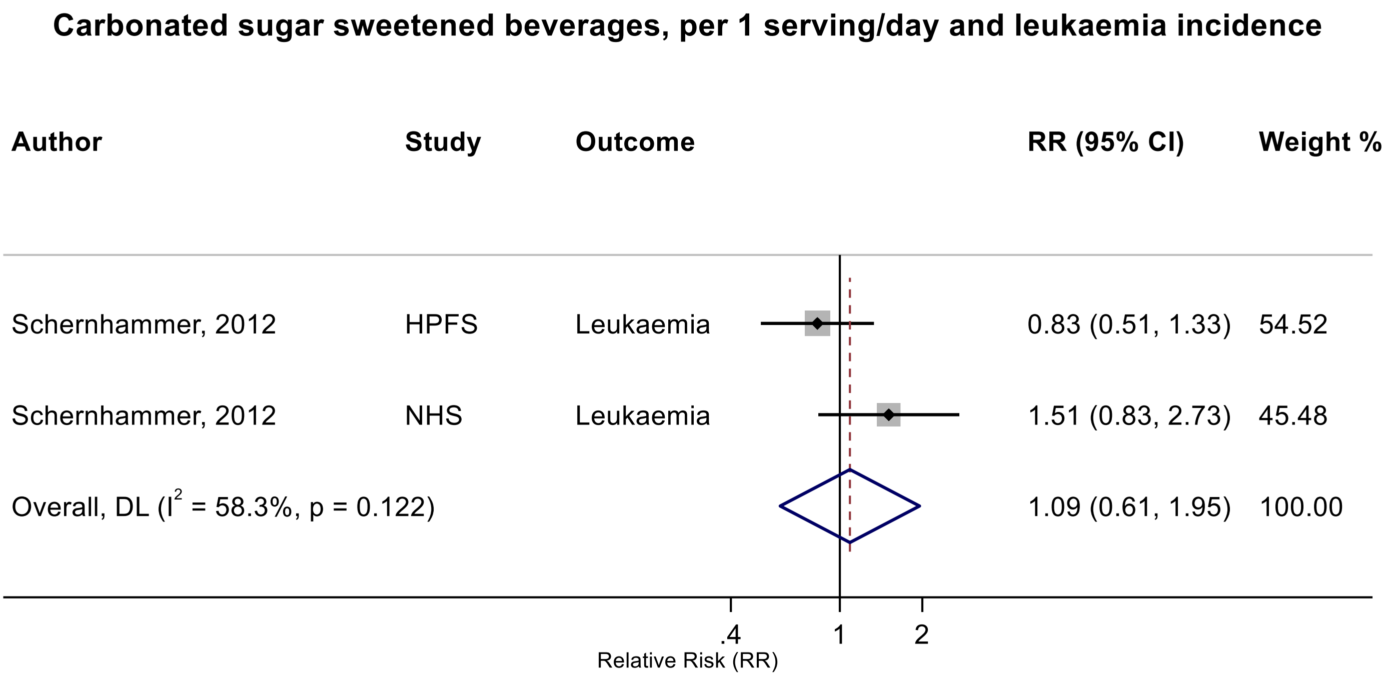

Figure legend: Forest plot shows the linear dose-response results (per 1 serving [355 mL] of carbonated sugar sweetened beverages/day) from the inverse variance DerSimonian-Laird random-effects model. The diamond represents the summary relative risk (RR) estimate, the diamond's width is the 95% confidence interval (CI). Each square and the horizontal line across the square represent the RR estimate of the individual study and its 95% CI.

Abbreviations: CI, confidence interval; RR, relative risk.

Study abbreviations: HPFS, Health Professionals Follow-up Study; NHS, Nurses' Health Study

Schernhammer, 2012[150]

**Supplementary Figure 102. Linear dose-response meta-analysis of carbonated sugar sweetened beverages and multiple myeloma incidence.**

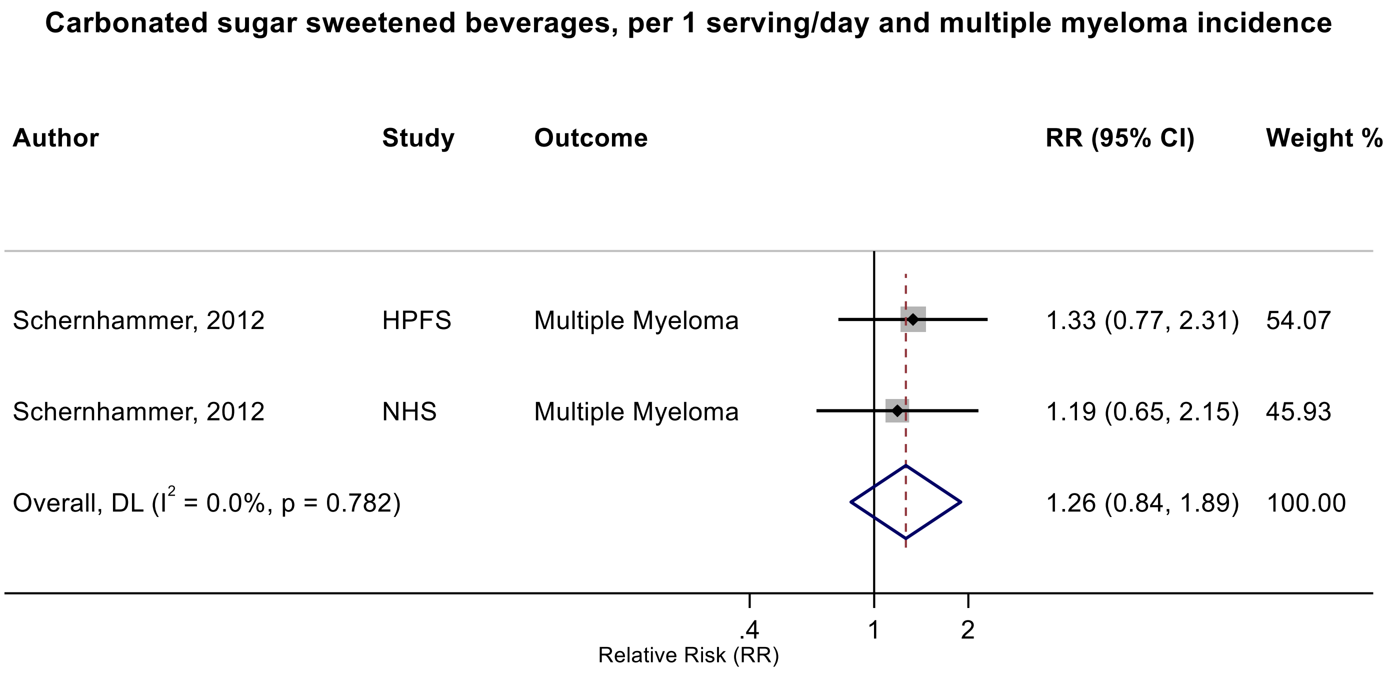

Figure legend: Forest plot shows the linear dose-response results (per 1 serving [355 mL] of carbonated sugar sweetened beverages/day) from the inverse variance DerSimonian-Laird random-effects model. The diamond represents the summary relative risk (RR) estimate, the diamond's width is the 95% confidence interval (CI). Each square and the horizontal line across the square represent the RR estimate of the individual study and its 95% CI.

Abbreviations: CI, confidence interval; RR, relative risk.

Study abbreviations: HPFS, Health Professionals Follow-up Study; NHS, Nurses' Health Study

Schernhammer, 2012[150]

**Supplementary Figure 103. Linear dose-response meta-analysis of carbonated sugar sweetened beverages and non-Hodgkin lymphoma incidence.**

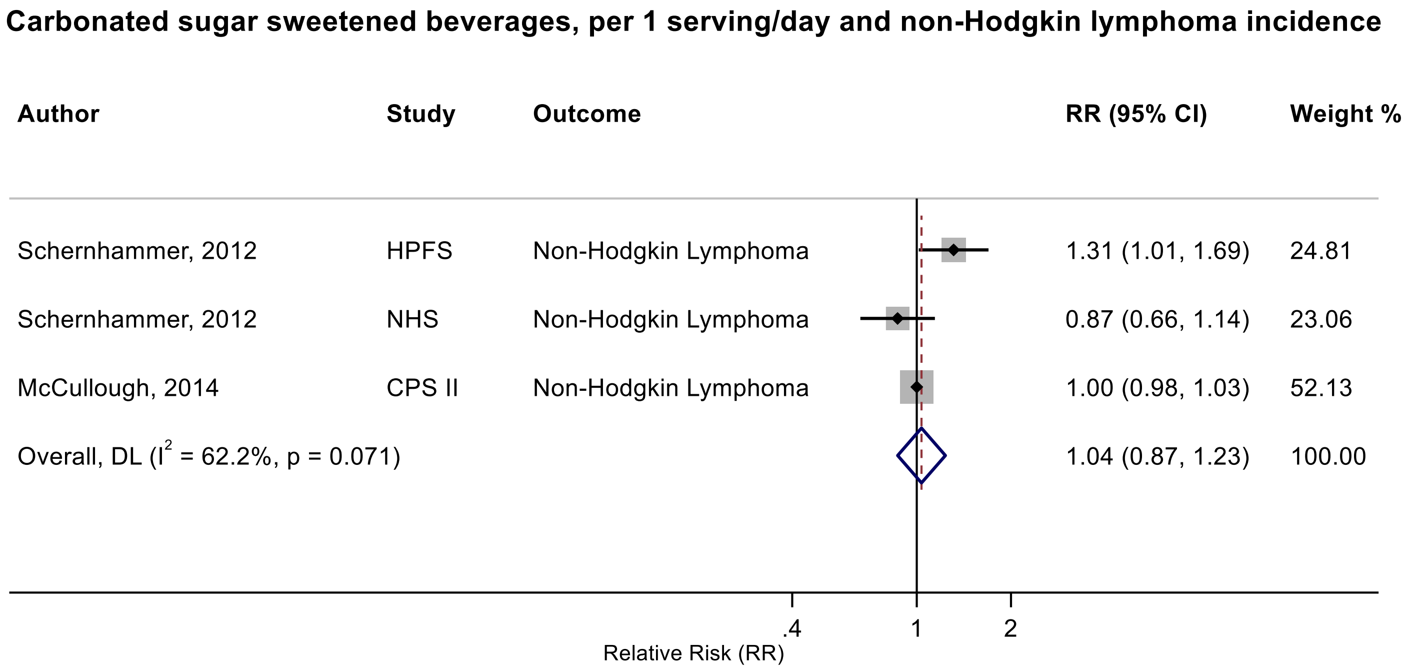

Figure legend: Forest plot shows the linear dose-response results (per 1 serving [355 mL] of carbonated sugar sweetened beverages /day) from the inverse variance DerSimonian-Laird random-effects model. The diamond represents the summary relative risk (RR) estimate, the diamond's width is the 95% confidence interval (CI). Each square and the horizontal line across the square represent the RR estimate of the individual study and its 95% CI.

Abbreviations: CI, confidence interval; RR, relative risk.

Study abbreviations: HPFS, Health Professionals Follow-up Study; NHS, Nurses' Health Study; CPS II, Cancer Prevention Study II

Schernhammer, 2012[150], McCullough, 2014[114]

**Supplementary Figure 104. Categorical highest versus the lowest meta-analysis of sugar sweetened beverages and thyroid cancer incidence.**

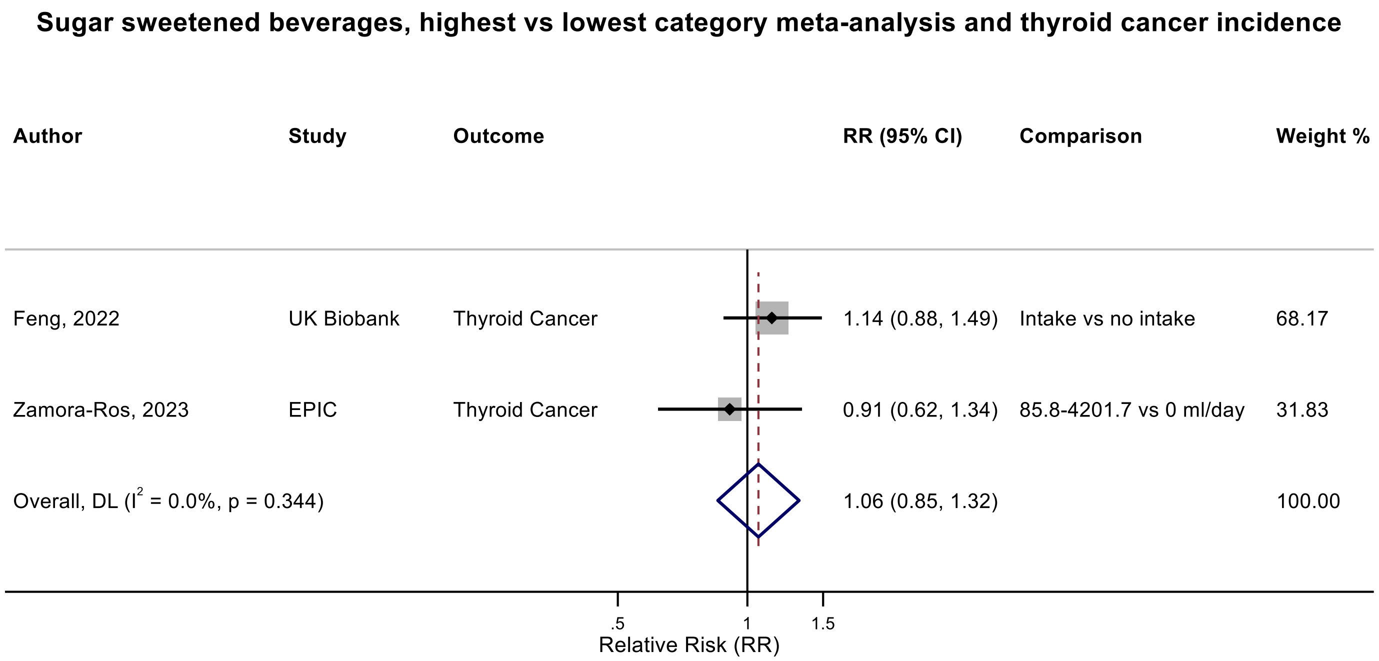

Figure legend: Forest plot shows the highest versus the lowest category results of sugar sweetened beverages from the inverse variance DerSimonian-Laird random-effects model. The diamond represents the summary relative risk (RR) estimate, the diamond's width is the 95% confidence interval (CI). Each square and the horizontal line across the square represent the RR estimate of the individual study and its 95% CI.

Abbreviations: CI, confidence interval; F, female, M, male, RR, relative risk.

Study abbreviations: UK Biobank, United Kingdom Biobank Study; EPIC, European Prospective Investigation into Cancer and nutrition

Feng, 2022[53], Zamora-Ros, 2023[180]

Note: The UK Biobank study was not eligible for the linear dose-response meta-analysis, so we performed a categorical high vs low meta-analysis.

**Supplementary Figure 105. Nonlinear dose-response meta-analysis of sugar sweetened beverages and colorectal cancer incidence.**

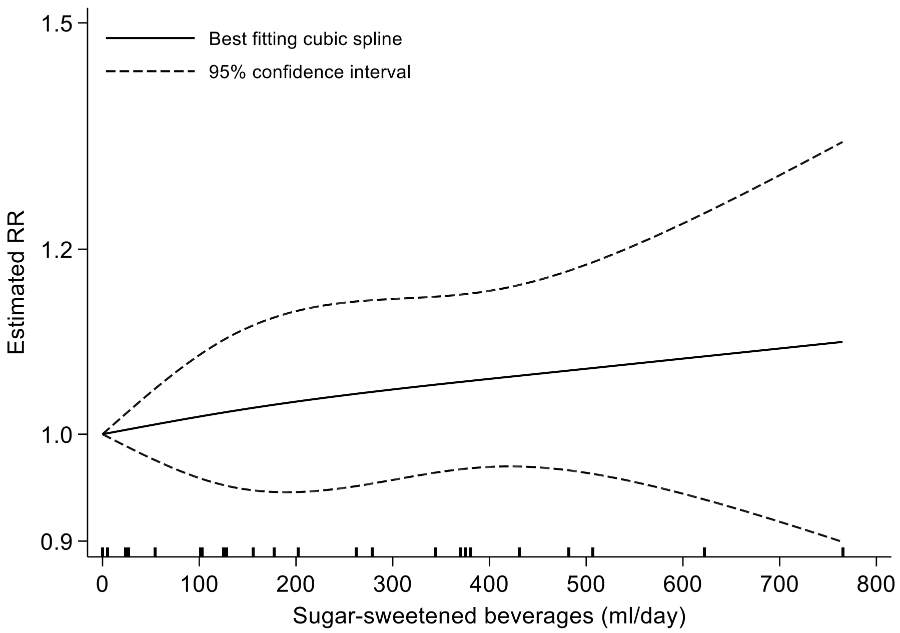

| SSB intake (mL/day) | RR estimates (95%CI) |
|---|---|
| 0 | 1 (reference) |
| 100 | 1.01 (0.96-1.08) |
| 200 | 1.03 (0.94-1.12) |
| 300 | 1.04 (0.95-1.14) |
| 400 | 1.05 (0.97-1.15) |
| 500 | 1.07 (0.96-1.18) |
| 600 | 1.08 (0.94-1.24) |
| 700 | 1.09 (0.92-1.23) |
| 800 | 1.11 (0.89-1.37) |

P non-linearity = 0.52

Figure legend: Non-linear curve was estimated using restricted cubic spline regression with three knots at 10th, 50th and 90th percentiles of distribution of the exposure and pooled in random-effects meta-analysis. The table shows the estimated RRs and 95% CIs for selected sugar sweetened beverages values comparing to no consumption as reference.

Abbreviations: CI, confidence interval; RR, relative risk.

Arthur, 2021[38], Barrubés, 2020[40], Hodge, 2018[70], Kanehara, 2024[81], Lee, 2023[97], Leung, 2021[101], Makarem, 2018[108], Nomura, 2016[129], Pacheco, 2019[135], Yuan, 2022[178]

**Supplementary Figure 106. Nonlinear dose-response meta-analysis of sugar sweetened beverages and breast cancer incidence.**

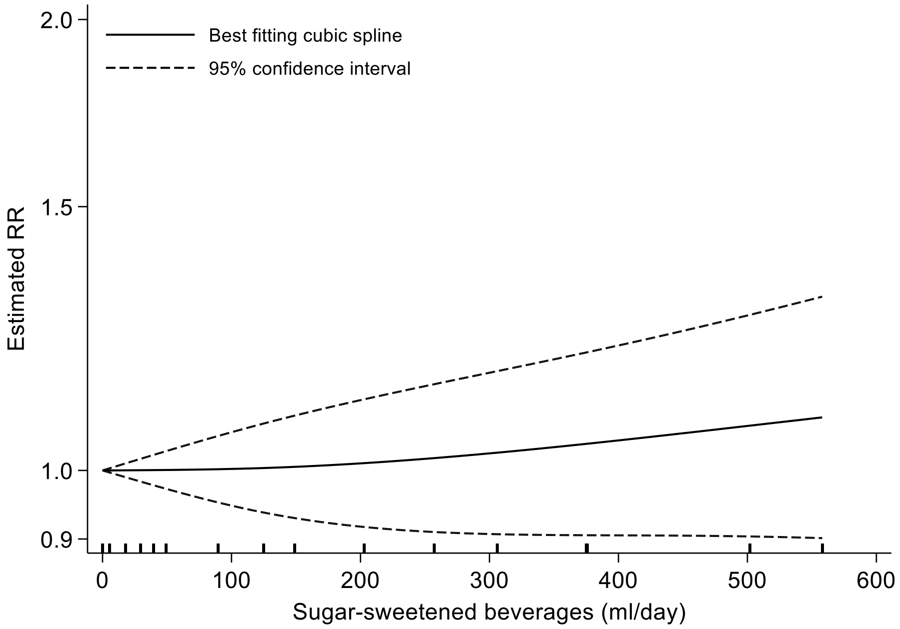

| SSB intake (mL/day) | RR estimates (95%CI) |
|---|---|
| 0 | 1 (reference) |
| 100 | 1.00 (0.95-1.06) |
| 200 | 1.01 (0.92-1.11) |
| 300 | 1.03 (0.91-1.16) |
| 400 | 1.05 (0.91-1.21) |
| 500 | 1.07 (0.90-1.27) |
| 550 | 1.08 (0.89-1.30) |

P non-linearity = 0.25

Figure legend: Non-linear curve was estimated using restricted cubic spline regression with three knots at 10th, 50th and 90th percentiles of distribution of the exposure and pooled in random-effects meta-analysis. The table shows the estimated RRs and 95% CIs for selected sugar sweetened beverages values comparing to no consumption as reference.

Abbreviations: CI, confidence interval; RR, relative risk.

Arthur, 2021[38], Lee, 2023[97], Makarem, 2018[108], Nomura, 2016[128], Romanos-Nanclares, 2021[145], Romanos-Nanclares, 2019[146]

**Supplementary Figure 107. Nonlinear dose-response meta-analysis of sugar sweetened beverages and postmenopausal breast cancer incidence.**

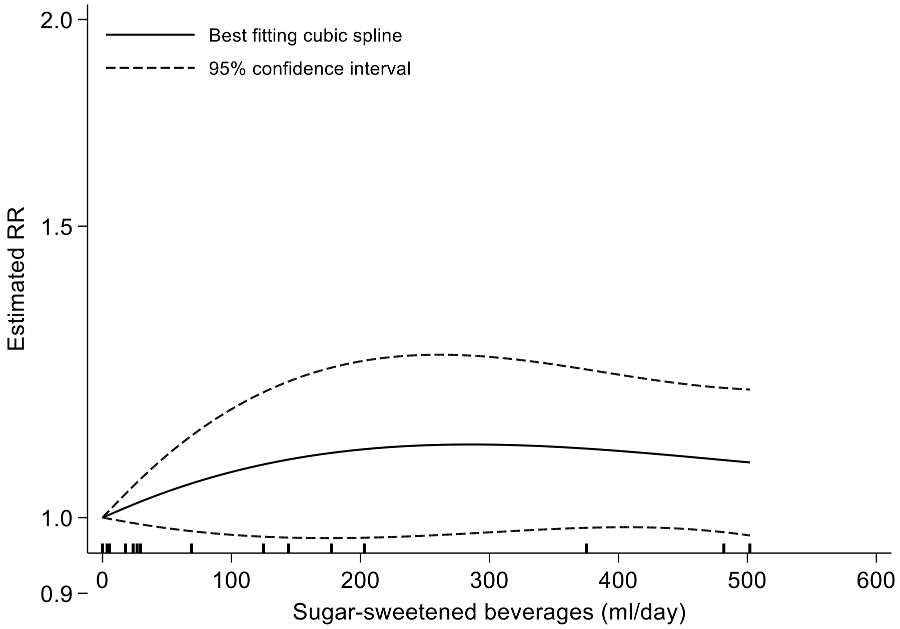

| SSB intake (ml/day) | RR estimates (95%CI) |
|---|---|
| 0 | 1 (reference) |
| 100 | 1.07 (0.98-1.16) |
| 200 | 1.10 (0.97-1.24) |
| 300 | 1.11 (0.98-1.25) |
| 400 | 1.10 (0.99-1.22) |
| 500 | 1.08 (0.98-1.02) |

P non-linearity = 0.61

Figure legend: Non-linear curve was estimated using restricted cubic spline regression with three knots at 10th, 50th and 90th percentiles of distribution of the exposure and pooled in random-effects meta-analysis. The table shows the estimated RRs and 95% CIs for selected sugar sweetened beverages values comparing to no consumption as reference.

Abbreviations: CI, confidence interval; RR, relative risk.

Arthur, 2021[38], Hodge, 2018[70], Nomura, 2016[128], Romanos-Nanclares, 2021[145], Romanos-Nanclares, 2019[146]

**Supplementary Figure 108.Linear dose-response meta-analysis of sugar sweetened beverages and stomach cancer incidence by sex.**

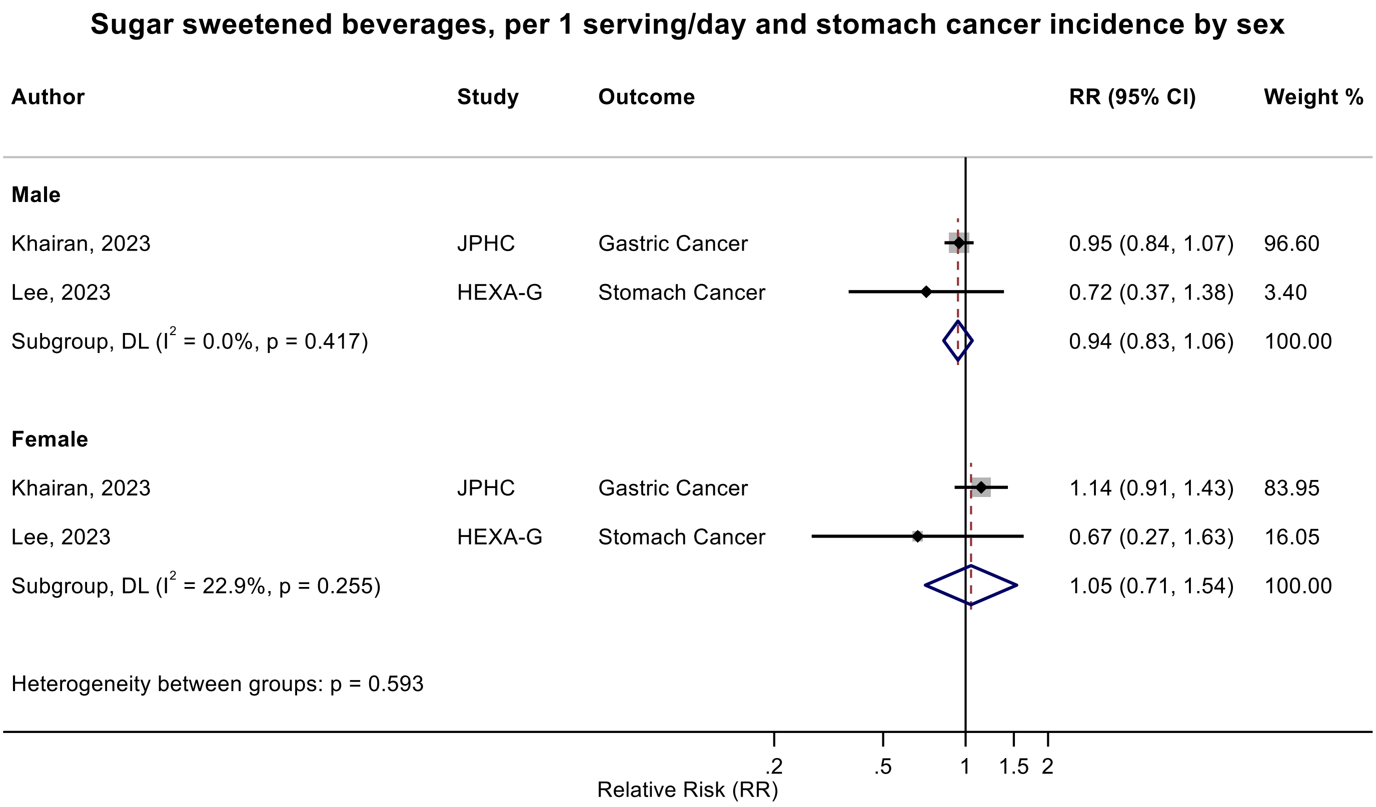

Figure legend: Forest plot shows the linear dose-response results (per 1 serving [355 mL] of sugar sweetened beverages /day) from the inverse variance DerSimonian-Laird random-effects model. The diamond represents the summary relative risk (RR) estimate, the diamond's width is the 95% confidence interval (CI). Each square and the horizontal line across the square represent the RR estimate of the individual study and its 95% CI.

Abbreviations: CI, confidence interval; F, female, M, male, RR, relative risk.

Study abbreviations: JPHC, Japan Public Health Center-Based Prospective Study; HEXA-G, Health Examinees-Gem study

Khairan, 2023[84], Lee, 2023[97]

**Supplementary Figure 109. Linear dose-response meta-analysis of sugar sweetened beverages and colorectal cancer incidence by geographical region.**

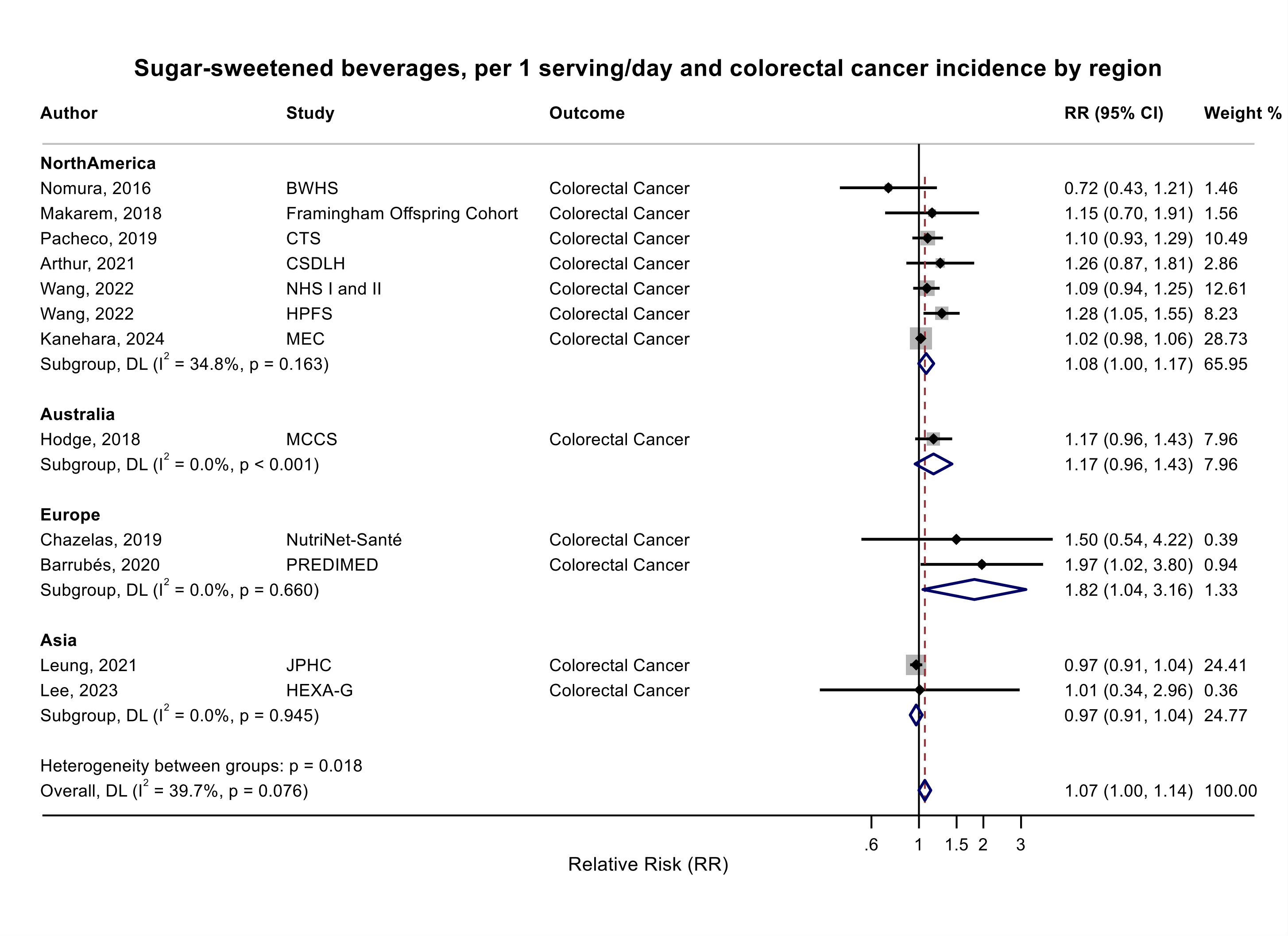

Figure legend: Forest plot shows the linear dose-response results (per 1 serving [355 mL] of sugar sweetened beverages /day) from the inverse variance DerSimonian-Laird random-effects model. The diamond represents the summary relative risk (RR) estimate, the diamond's width is the 95% confidence interval (CI). Each square and the horizontal line across the square represent the RR estimate of the individual study and its 95% CI.

Abbreviations: CI, confidence interval; RR, relative risk.

Study abbreviations: BWHS, Black Women's Health Study; PREDIMED, Prevención con Dieta Mediterránea study; CTS, California Teachers Study; CSDLH, Canadian Study of Diet, Lifestyle and Health; NHS I and II, Nurses' Health Study I and II; HPFS, Health Professionals Follow-up Study; MEC, Multiethnic Cohort Study; MCCS, Melbourne Collaborative Cohort Study; JPHC, Japan Public Health Center-Based Prospective Study; HEXA-G, Health Examinees-Gem study

Nomura, 2016[129], Chazelas, 2019[44], Barrubés, 2020[40], Makarem, 2018[108], Pacheco, 2019[135], Arthur, 2021[38], Wang, 2022[168], Kanehara, 2024[81],
Hodge, 2018[70], Leung, 2021[101], Lee, 2023[97]

**Supplementary Figure 110. Linear dose-response meta-analysis of sugar sweetened beverages and colorectal cancer incidence by follow-up duration.**

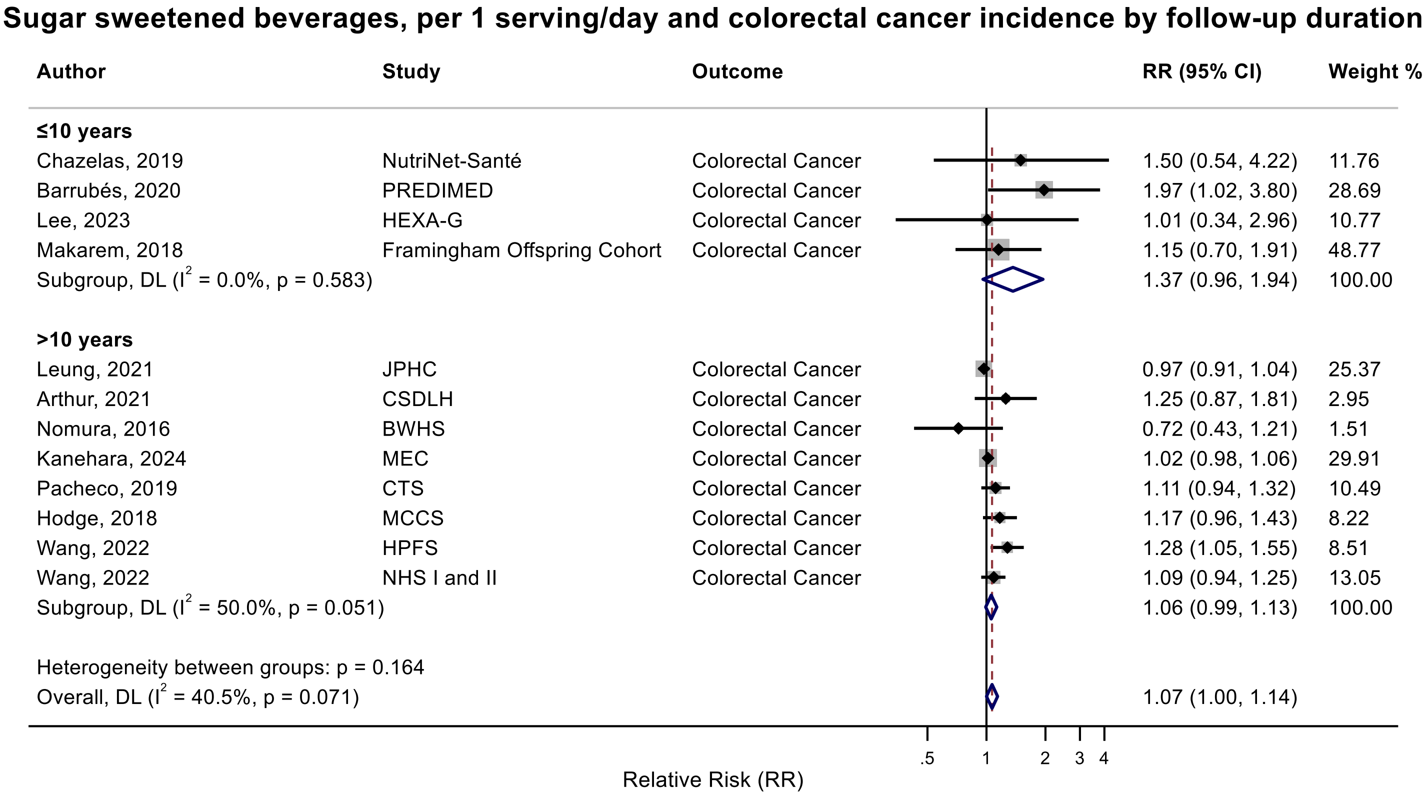

Figure legend: Forest plot shows the linear dose-response results (per 1 serving [355 mL] of sugar sweetened beverages /day) from the inverse variance DerSimonian-Laird random-effects model. The diamond represents the summary relative risk (RR) estimate, the diamond's width is the 95% confidence interval (CI). Each square and the horizontal line across the square represent the RR estimate of the individual study and its 95% CI.

Abbreviations: CI, confidence interval; RR, relative risk.

Study abbreviations: PREDIMED, Prevención con Dieta Mediterránea study; HEXA-G, Health Examinees-Gem study; FOS, Framingham Heart Study-Offspring Cohort; JPHC, Japan Public Health Center-Based Prospective Study; CSDLH, Canadian Study of Diet, Lifestyle and Health; BWHS, Black Women's Health Study; MEC, Multiethnic Cohort Study; CTS, California Teachers Study; MCCS, Melbourne Collaborative Cohort Study; HPFS, Health Professionals Follow-up Study; NHS I and II, Nurses' Health Study I and II

Nomura, 2016[129], Chazelas, 2019[44], Barrubés, 2020[40], Makarem, 2018[108], Pacheco, 2019[135], Arthur, 2021[38], Wang, 2022[168], Kanehara, 2024[81],
Hodge, 2018[70], Leung, 2021[101], Lee, 2023[97]

**Supplementary Figure 111. Linear dose-response meta-analysis of sugar sweetened beverages and colorectal cancer incidence by sex.**

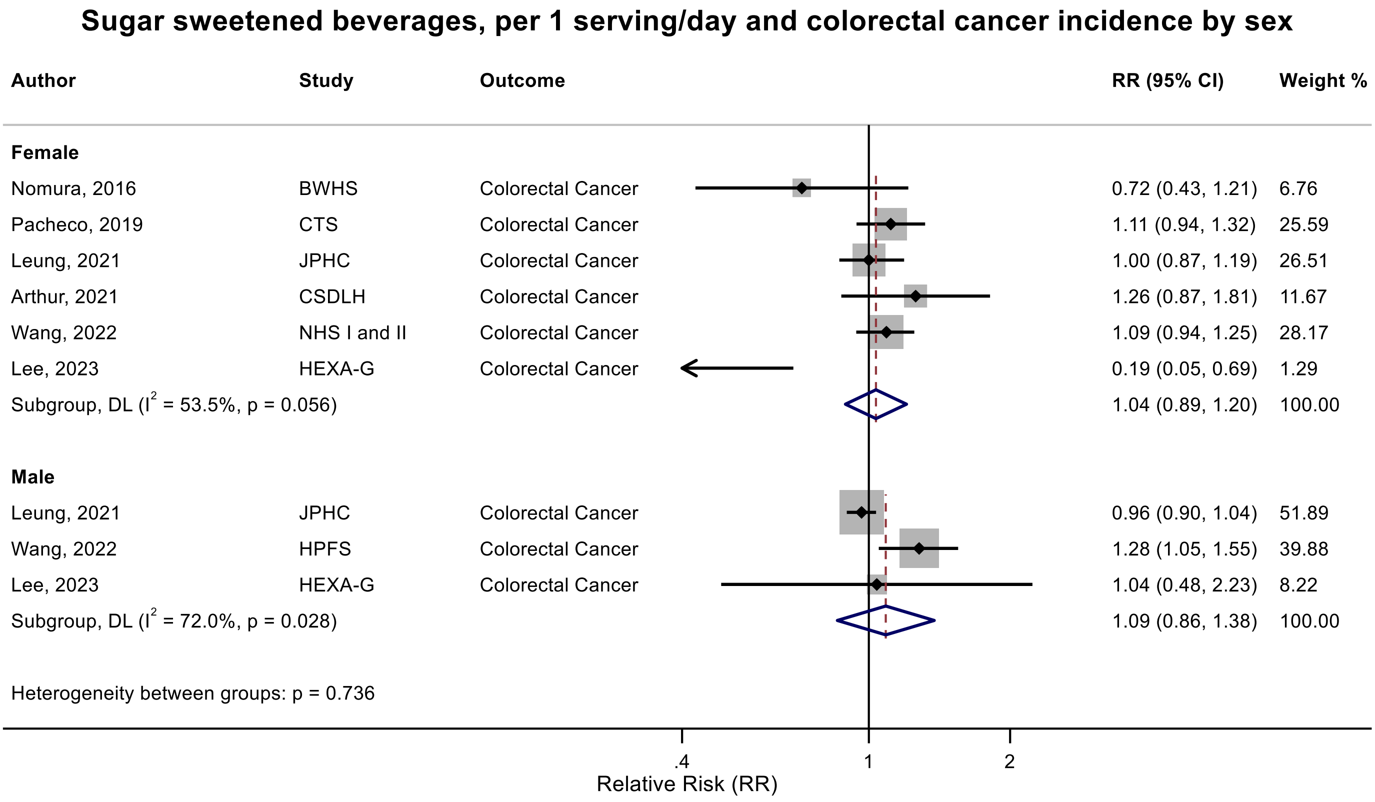

Figure legend: Forest plot shows the linear dose-response results (per 1 serving [355 mL] of sugar sweetened beverages /day) from the inverse variance DerSimonian-Laird random-effects model. The diamond represents the summary relative risk (RR) estimate, the diamond's width is the 95% confidence interval (CI). Each square and the horizontal line across the square represent the RR estimate of the individual study and its 95% CI.

Abbreviations: CI, confidence interval; F, female, M, male, RR, relative risk.

Study abbreviations: BWHS, Black Women's Health Study; CTS, California Teachers Study; JPHC, Japan Public Health Center-Based Prospective Study; CSDLH, Canadian Study of Diet, Lifestyle and Health; NHS I and II, Nurses' Health Study I and II; HEXA-G, Health Examinees-Gem study; HPFS, Health Professionals Follow-up Study

Nomura, 2016[129], Pacheco, 2019[135], Leung, 2021[101], Arthur, 2021[38], Wang, 2022[169], Lee, 2023[97]

**Supplementary Figure 112. Linear dose-response meta-analysis of sugar sweetened beverages and colorectal cancer incidence by inclusion of fruit juice.**

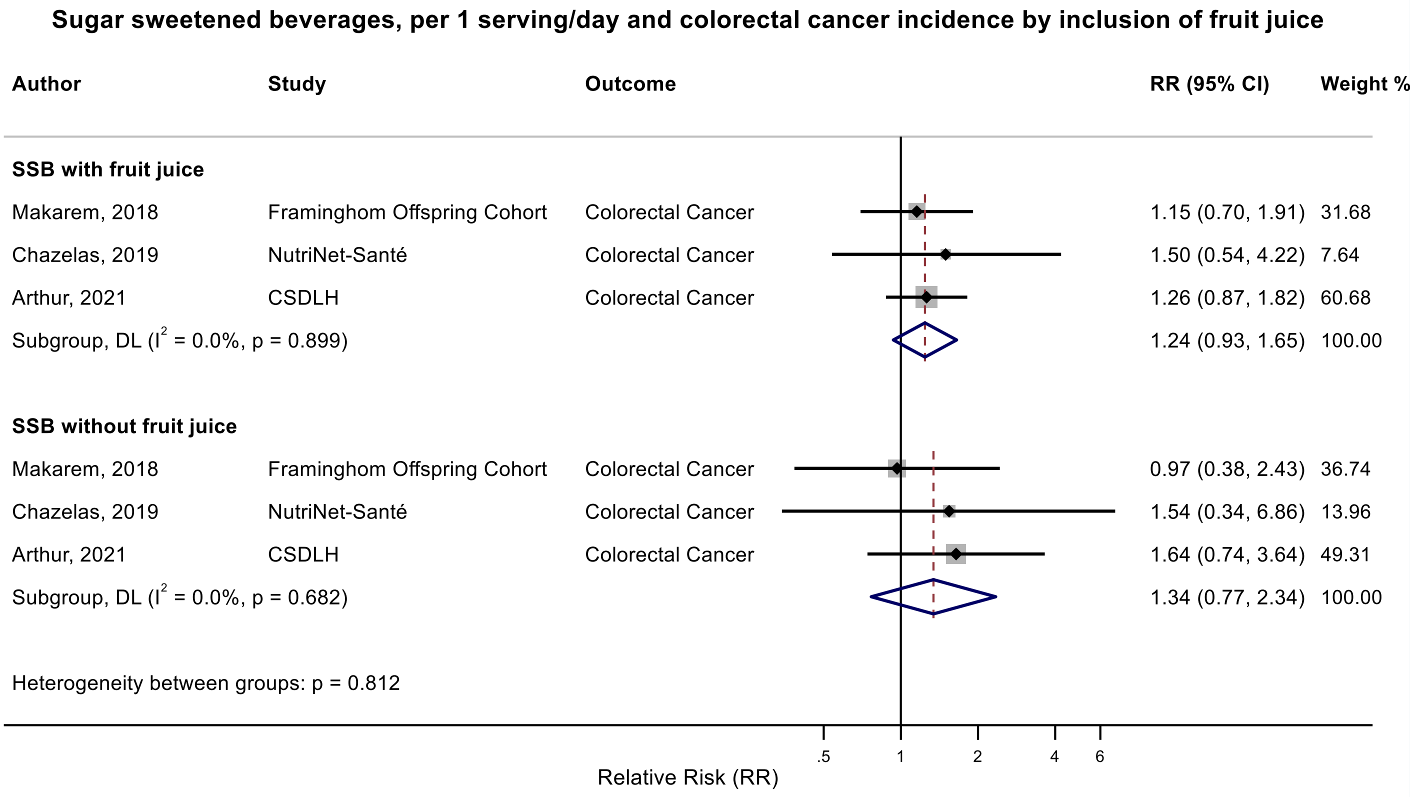

Figure legend: Forest plot shows the linear dose-response results (per 1 serving [355 mL] of sugar sweetened beverages /day) from the inverse variance DerSimonian-Laird random-effects model. The diamond represents the summary relative risk (RR) estimate, the diamond's width is the 95% confidence interval (CI). Each square and the horizontal line across the square represent the RR estimate of the individual study and its 95% CI.

Abbreviations: CI, confidence interval; RR, relative risk, SSB, sugar sweetened beverages.

Study abbreviations: FOS, Framingham Heart Study-Offspring Cohort; CSDLH, Canadian Study of Diet, Lifestyle and Health

Note: This subgroup analysis is based on within study comparisons in three studies reporting results for sugar sweetened beverages with and without fruit juices.

Makarem, 2018[108], Chazelas, 2019[44], Arthur, 2021[38]

**Supplementary Figure 113. Linear dose-response meta-analysis of sugar sweetened beverages and colorectal cancer incidence by frequency of dietary assessment.**

**Sugar sweetened beverages, per 1 serving/day and colorectal cancer incidence by frequency of dietary assessment**

| Author | Study | Outcome | RR (95% CI) | Weight % |
|---|---|---|---|---|
| **Reapeted** | | | | |
| Nomura, 2016 | BWHS | Colorectal Cancer | 0.75 (0.45, 1.25) | 9.79 |
| Chazelas, 2019 | NutriNet-Santé | Colorectal Cancer | 1.50 (0.54, 4.22) | 2.67 |
| Wang, 2022 | NHS I and II | Colorectal Cancer | 1.09 (0.94, 1.25) | 49.43 |
| Wang, 2022 | HPFS | Colorectal Cancer | 1.28 (1.05, 1.55) | 38.11 |
| Subgroup, DL ($I^2$ = 35.0%, p = 0.203) | | | 1.13 (0.95, 1.34) | 100.00 |
| **Baseline** | | | | |
| Nomura, 2016 | BWHS | Colorectal Cancer | 0.72 (0.43, 1.21) | 1.44 |
| Hodge, 2018 | MCCS | Colorectal Cancer | 1.17 (0.96, 1.43) | 8.45 |
| Makarem, 2018 | Framingham Offspring Cohort | Colorectal Cancer | 1.15 (0.70, 1.91) | 1.54 |
| Pacheco, 2019 | CTS | Colorectal Cancer | 1.10 (0.93, 1.29) | 11.48 |
| Barrubés, 2020 | PREDIMED | Colorectal Cancer | 1.97 (1.02, 3.80) | 0.92 |
| Arthur, 2021 | CSDLH | Colorectal Cancer | 1.26 (0.87, 1.81) | 2.87 |
| Leung, 2021 | JPHC | Colorectal Cancer | 0.97 (0.91, 1.04) | 32.30 |
| Lee, 2023 | HEXA-G | Colorectal Cancer | 1.01 (0.34, 2.96) | 0.35 |
| Kanehara, 2024 | MEC | Colorectal Cancer | 1.02 (0.98, 1.06) | 40.65 |
| Subgroup, DL ($I^2$ = 32.1%, p = 0.162) | | | 1.03 (0.97, 1.10) | 100.00 |

Heterogeneity between groups: p = 0.353

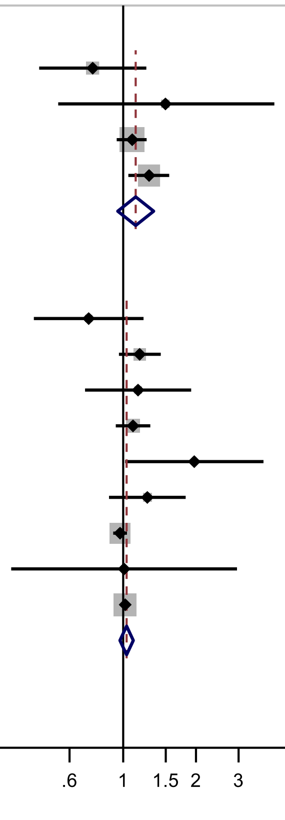

Figure legend: Forest plot shows the linear dose-response results (per 1 serving [355 mL] of sugar sweetened beverages /day) from the inverse variance DerSimonian-Laird random-effects model. The diamond represents the summary relative risk (RR) estimate, the diamond's width is the 95% confidence interval (CI). Each square and the horizontal line across the square represent the RR estimate of the individual study and its 95% CI.

Abbreviations: CI, confidence interval; RR, relative risk.

Study abbreviations: BWHS, Black Women's Health Study; NHS I and II, Nurses' Health Study I and II; HPFS, Health Professionals Follow-up Study; MCCS, Melbourne Collaborative Cohort Study; CTS, California Teachers Study; PREDIMED, Prevención con Dieta Mediterránea study; JPHC, Japan Public Health Center-Based Prospective Study; CSDLH, Canadian Study of Diet, Lifestyle and Health; HEXA-G, Health Examinees-Gem study; MEC, Multiethnic Cohort Study

Note: The Black Women's Health Study reported results for both baseline and time-varying measurements

Nomura, 2016[129], Chazelas, 2019[44], Wang, 2022[168], Makarem, 2018[108], Hodge, 2018[70], Pacheco, 2019[135], Barrubés, 2020[40], Leung, 2021[101], Arthur, 2021[38], Lee, 2023[97], Kanehara, 2024[81]

**Supplementary Figure 114. Linear dose-response meta-analysis of sugar sweetened beverages and colorectal cancer incidence by exclusion of people with diabetes at baseline.**

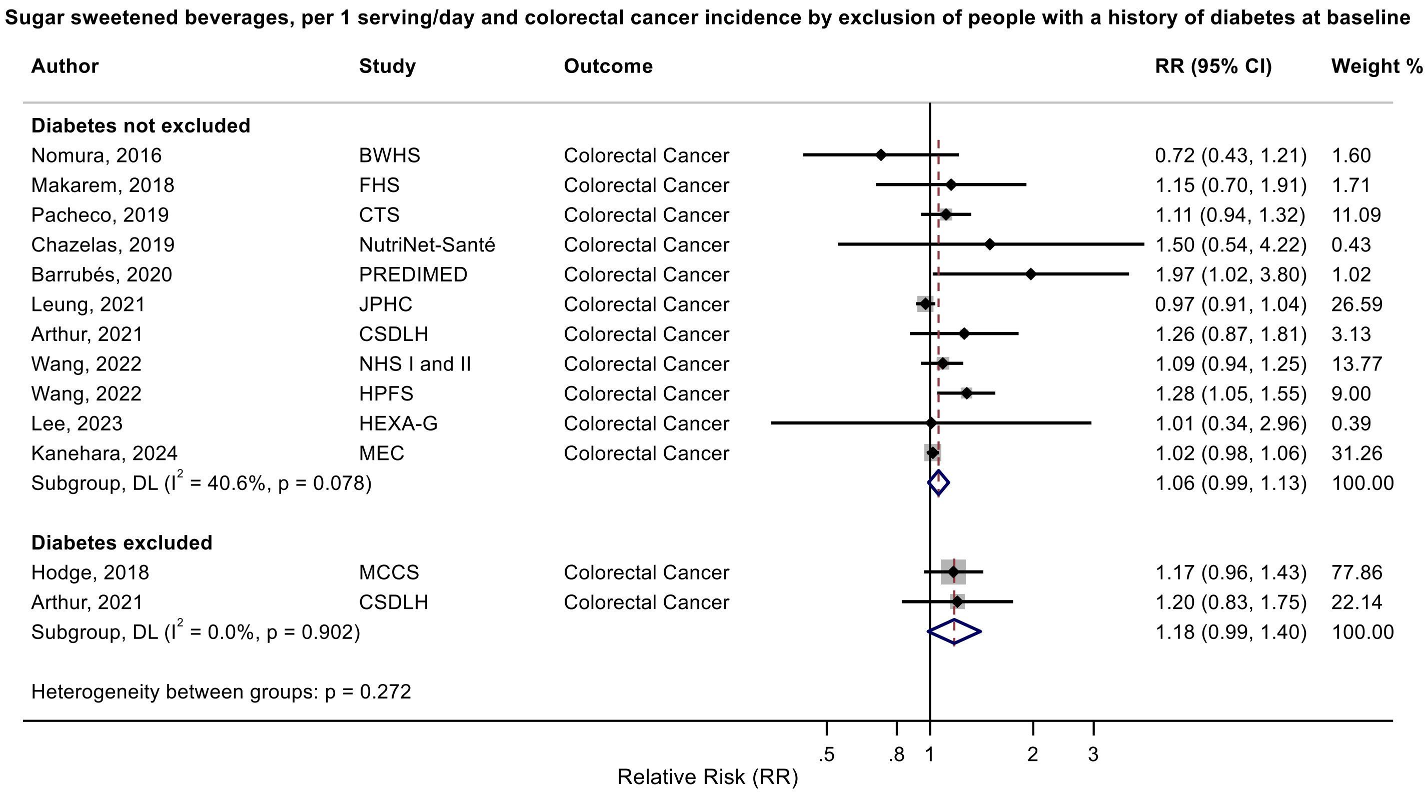

Figure legend: Forest plot shows the linear dose-response results (per 1 serving [355 mL] of sugar sweetened beverages /day) from the inverse variance DerSimonian-Laird random-effects model. The diamond represents the summary relative risk (RR) estimate, the diamond's width is the 95% confidence interval (CI). Each square and the horizontal line across the square represent the RR estimate of the individual study and its 95% CI.

Abbreviations: CI, confidence interval; RR, relative risk.

Study abbreviations: BWHS, Black Women's Health Study; FHS, Framingham Heart Study; CTS, California Teachers Study; PREDIMED, Prevención con Dieta Mediterránea study; JPHC, Japan Public Health Center-Based Prospective Study; CSDLH, Canadian Study of Diet, Lifestyle and Health; NHS I and II, Nurses' Health Study I and II; HPFS, Health Professionals Follow-up Study; HEXA-G, Health Examinees-Gem study; MEC, Multiethnic Cohort Study; MCCS, Melbourne Collaborative Cohort Study

Note: The Canadian Study of Diet, Lifestyle and Health reported results for both before and after exclusion of people with a history of diabetes at baseline.

Nomura, 2016[129], Makarem, 2018[108], Pacheco, 2019[135], Chazelas, 2019[44], Barrubés, 2020[40], Leung, 2021[101], Arthur, 2021[38], Wang, 2022[168], Lee, 2023[97], Kanehara, 2024[81], Hodge, 2018[70]

**Supplementary Figure 115. Linear dose-response meta-analysis of sugar sweetened beverages and colorectal cancer incidence by bias related to confounding.**

**Sugar sweetened beverages, per 1 serving/day and colorectal cancer incidence by bias due to confounding**

| Author | Study | Outcome | RR (95% CI) | Weight % |
|---|---|---|---|---|
| **Moderate** | | | | |
| Nomura, 2016 | BWHS | Colorectal Cancer | 0.72 (0.43, 1.21) | 1.66 |
| Hodge, 2018 | MCCS | Colorectal Cancer | 1.17 (0.96, 1.43) | 9.36 |
| Makarem, 2018 | Framingham Offspring Cohort | Colorectal Cancer | 1.15 (0.70, 1.91) | 1.77 |
| Pacheco, 2019 | CTS | Colorectal Cancer | 1.11 (0.94, 1.32) | 12.53 |
| Barrubés, 2020 | PREDIMED | Colorectal Cancer | 1.97 (1.02, 3.80) | 1.06 |
| Arthur, 2021 | CSDLH | Colorectal Cancer | 1.25 (0.87, 1.81) | 3.27 |
| Leung, 2021 | JPHC | Colorectal Cancer | 0.97 (0.91, 1.04) | 31.82 |
| Kanehara, 2024 | MEC | Colorectal Cancer | 1.02 (0.98, 1.06) | 38.54 |
| Subgroup, DL ($I^2$ = 40.5%, p = 0.108) | | | 1.04 (0.97, 1.11) | 100.00 |
| **Serious** | | | | |
| Chazelas, 2019 | NutriNet-Santé | Colorectal Cancer | 1.50 (0.54, 4.22) | 1.22 |
| Wang, 2022 | NHS I and II | Colorectal Cancer | 1.09 (0.94, 1.25) | 63.60 |
| Wang, 2022 | HPFS | Colorectal Cancer | 1.28 (1.05, 1.55) | 34.06 |
| Lee, 2023 | HEXA-G | Colorectal Cancer | 1.01 (0.34, 2.96) | 1.12 |
| Subgroup, DL ($I^2$ = 0.0%, p = 0.571) | | | 1.15 (1.03, 1.29) | 100.00 |
| Heterogeneity between groups: p = 0.112 | | | | |

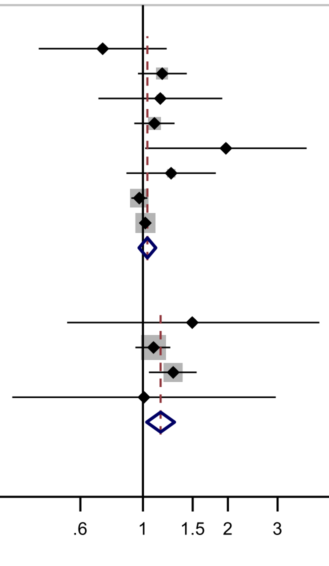

Figure legend: Forest plot shows the linear dose-response results (per 1 serving [355 mL] of sugar sweetened beverages /day) from the inverse variance DerSimonian-Laird random-effects model. The diamond represents the summary relative risk (RR) estimate, the diamond's width is the 95% confidence interval (CI). Each square and the horizontal line across the square represent the RR estimate of the individual study and its 95% CI.

Abbreviations: CI, confidence interval; RR, relative risk.

Study abbreviations: BWHS, Black Women's Health Study; FHS, Framingham Heart Study; CTS, California Teachers Study; PREDIMED, Prevención con Dieta Mediterránea study; JPHC, Japan Public Health Center-Based Prospective Study; CSDLH, Canadian Study of Diet, Lifestyle and Health; NHS I and II, Nurses' Health Study I and II; HPFS, Health Professionals Follow-up Study; HEXA-G, Health Examinees-Gem study; MEC, Multiethnic Cohort Study; MCCS, Melbourne Collaborative Cohort Study

Nomura, 2016[129], Makarem, 2018[108], Pacheco, 2019[135], Chazelas, 2019[44], Barrubés, 2020[40], Leung, 2021[101], Arthur, 2021[38], Wang, 2022[168], Lee, 2023[97], Kanehara, 2024[81], Hodge, 2018[70]

**Supplementary Figure 116. Linear dose-response meta-analysis of sugar sweetened beverages and colorectal cancer incidence by bias related to departure from intended exposure.**

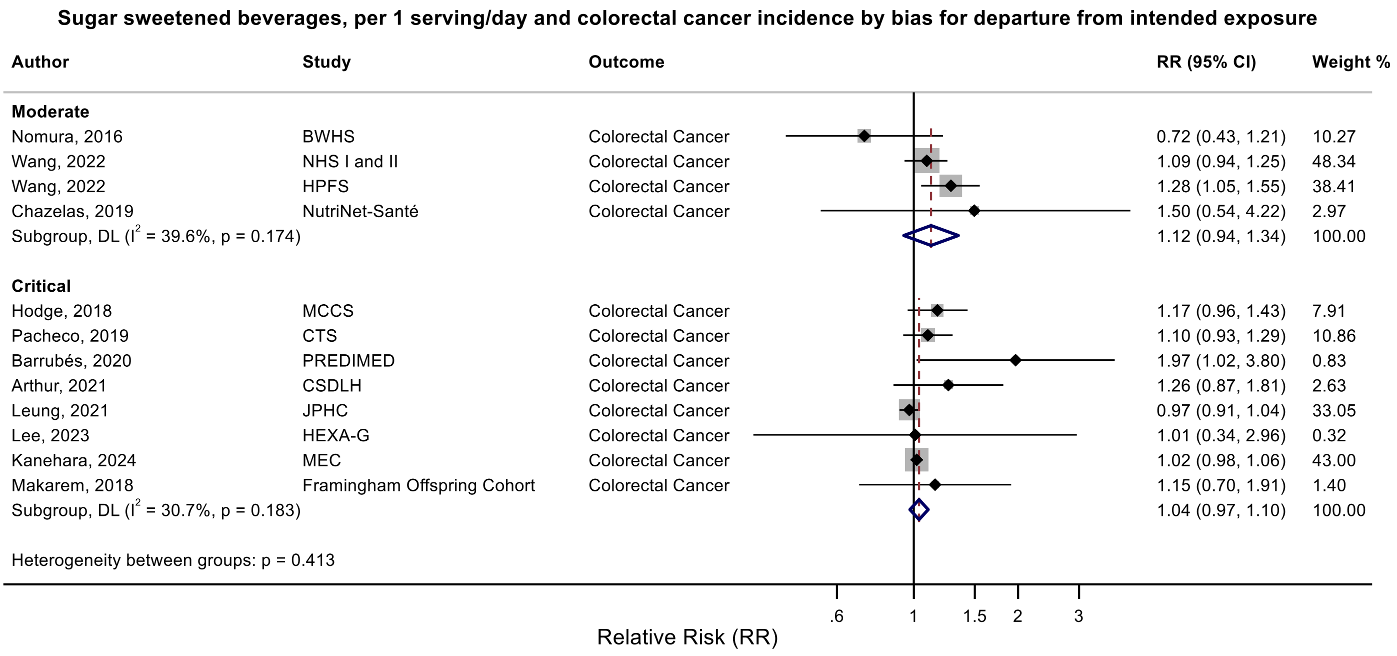

Figure legend: Forest plot shows the linear dose-response results (per 1 serving [355 mL] of sugar sweetened beverages /day) from the inverse variance DerSimonian-Laird random-effects model. The diamond represents the summary relative risk (RR) estimate, the diamond's width is the 95% confidence interval (CI). Each square and the horizontal line across the square represent the RR estimate of the individual study and its 95% CI.

Abbreviations: CI, confidence interval; RR, relative risk.

Study abbreviations: BWHS, Black Women's Health Study; FHS, Framingham Heart Study; CTS, California Teachers Study; PREDIMED, Prevención con Dieta Mediterránea study; JPHC, Japan Public Health Center-Based Prospective Study; CSDLH, Canadian Study of Diet, Lifestyle and Health; NHS I and II, Nurses' Health Study I and II; HPFS, Health Professionals Follow-up Study; HEXA-G, Health Examinees-Gem study; MEC, Multiethnic Cohort Study; MCCS, Melbourne Collaborative Cohort Study

Nomura, 2016[129], Makarem, 2018[108], Pacheco, 2019[135], Chazelas, 2019[44], Barrubés, 2020[40], Leung, 2021[101], Arthur, 2021[38], Wang, 2022[168], Lee, 2023[97], Kanehara, 2024[81], Hodge, 2018[70]

**Supplementary Figure 117. Linear dose-response meta-analysis of sugar sweetened beverages and colon cancer incidence by sex.**

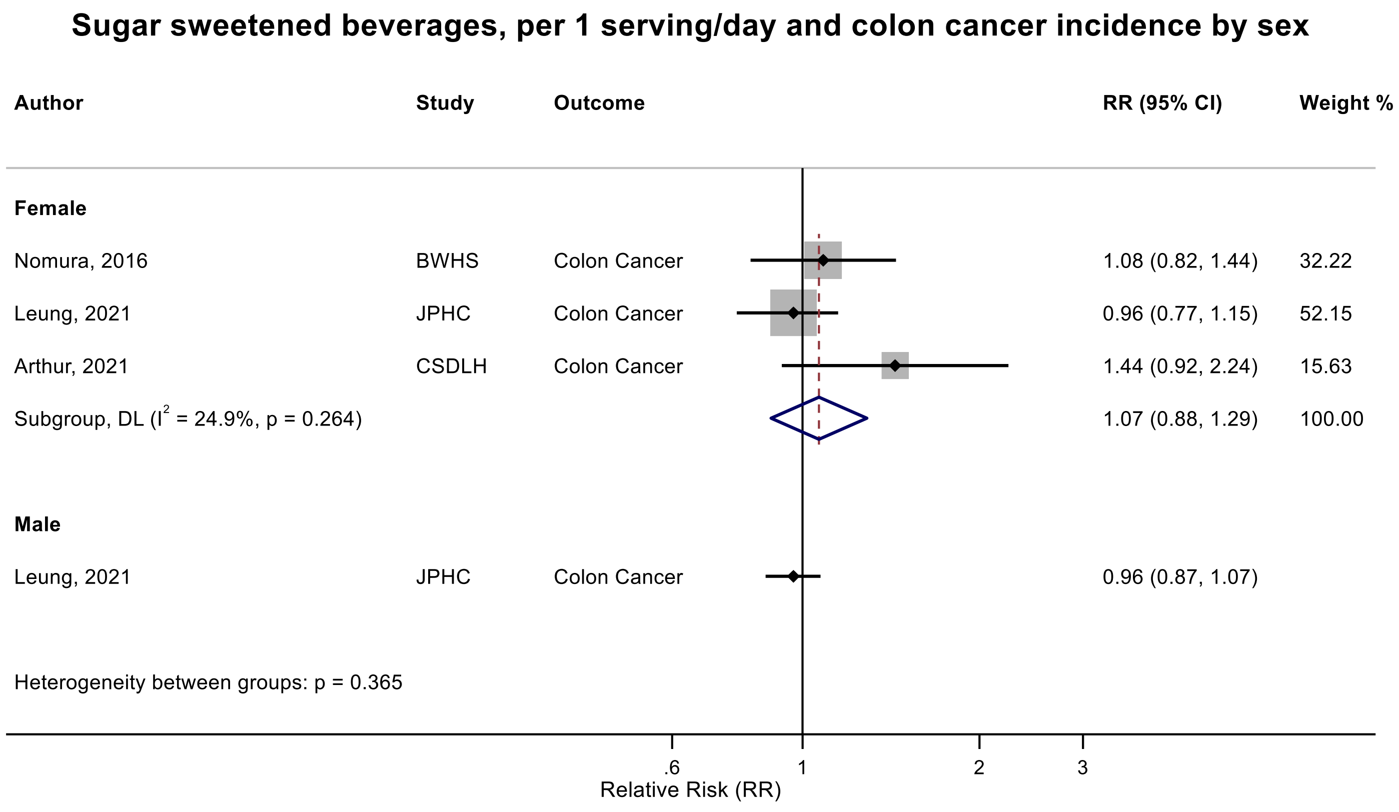

Figure legend: Forest plot shows the linear dose-response results (per 1 serving [355 mL] of sugar sweetened beverages /day) from the inverse variance DerSimonian-Laird random-effects model. The diamond represents the summary relative risk (RR) estimate, the diamond's width is the 95% confidence interval (CI). Each square and the horizontal line across the square represent the RR estimate of the individual study and its 95% CI.

Abbreviations: CI, confidence interval; RR, relative risk.

Study abbreviations: BWHS, Black Women's Health Study; JPHC, Japan Public Health Center-Based Prospective Study; CSDLH, Canadian Study of Diet, Lifestyle and Health

Nomura, 2016[129], Leung, 2021[101], Arthur, 2021[38]

#### Supplementary Figure 118. Linear dose-response meta-analysis of sugar sweetened beverages and proximal colon cancer incidence by sex.

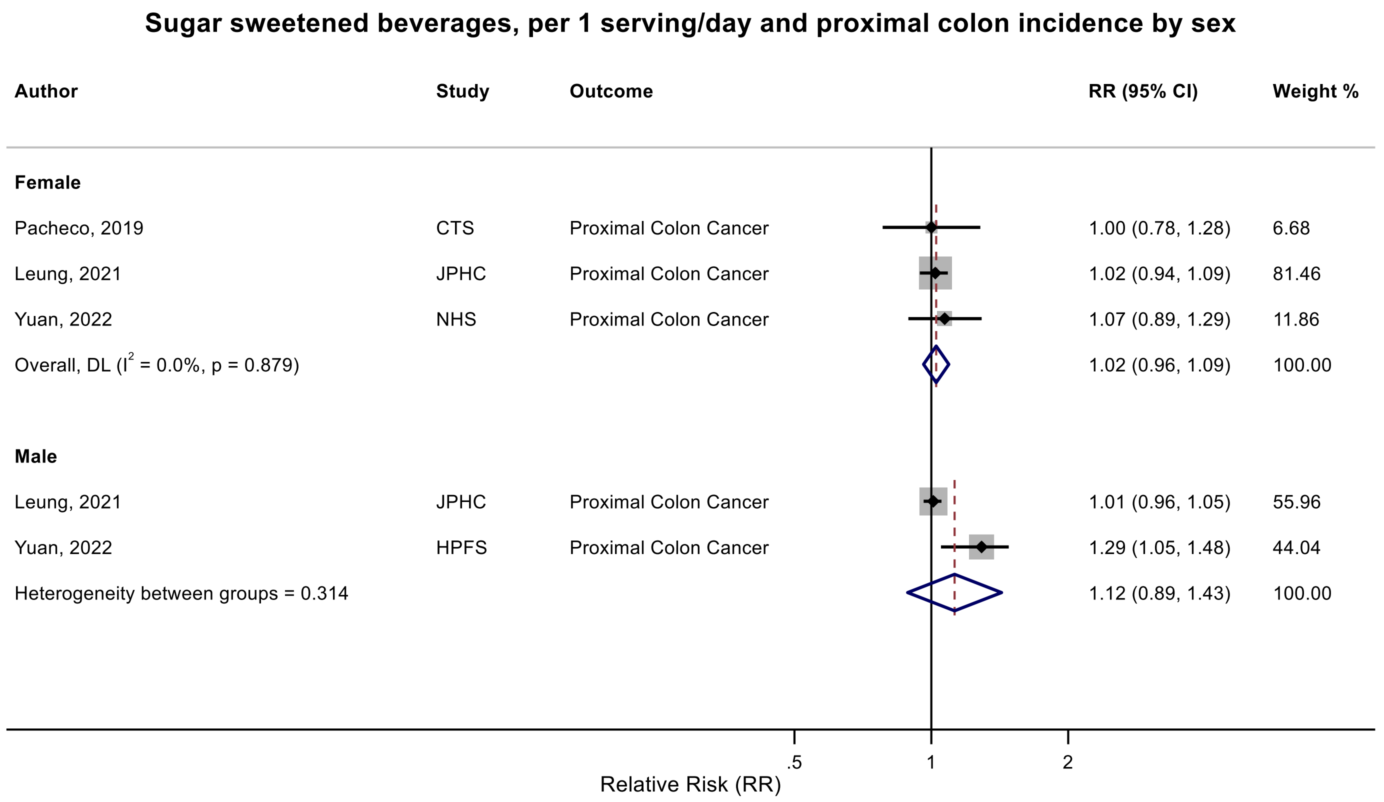

Figure legend: Forest plot shows the linear dose-response results (per 1 serving [355 mL] of sugar sweetened beverages /day) from the inverse variance DerSimonian-Laird random-effects model. The diamond represents the summary relative risk (RR) estimate, the diamond's width is the 95% confidence interval (CI). Each square and the horizontal line across the square represent the RR estimate of the individual study and its 95% CI.

Abbreviations: CI, confidence interval; RR, relative risk.

Study abbreviations: CTS, California Teachers Study; JPHC, Japan Public Health Center-Based Prospective Study; NHS, Nurses' Health Study; HPFS, Health Professionals Follow-up Study

Pacheco, 2019[135], Leung, 2021[101], Yuan, 2022[178]

**Supplementary Figure 119. Linear dose-response meta-analysis of sugar sweetened beverages and proximal colon cancer incidence by frequency of dietary assessment.**

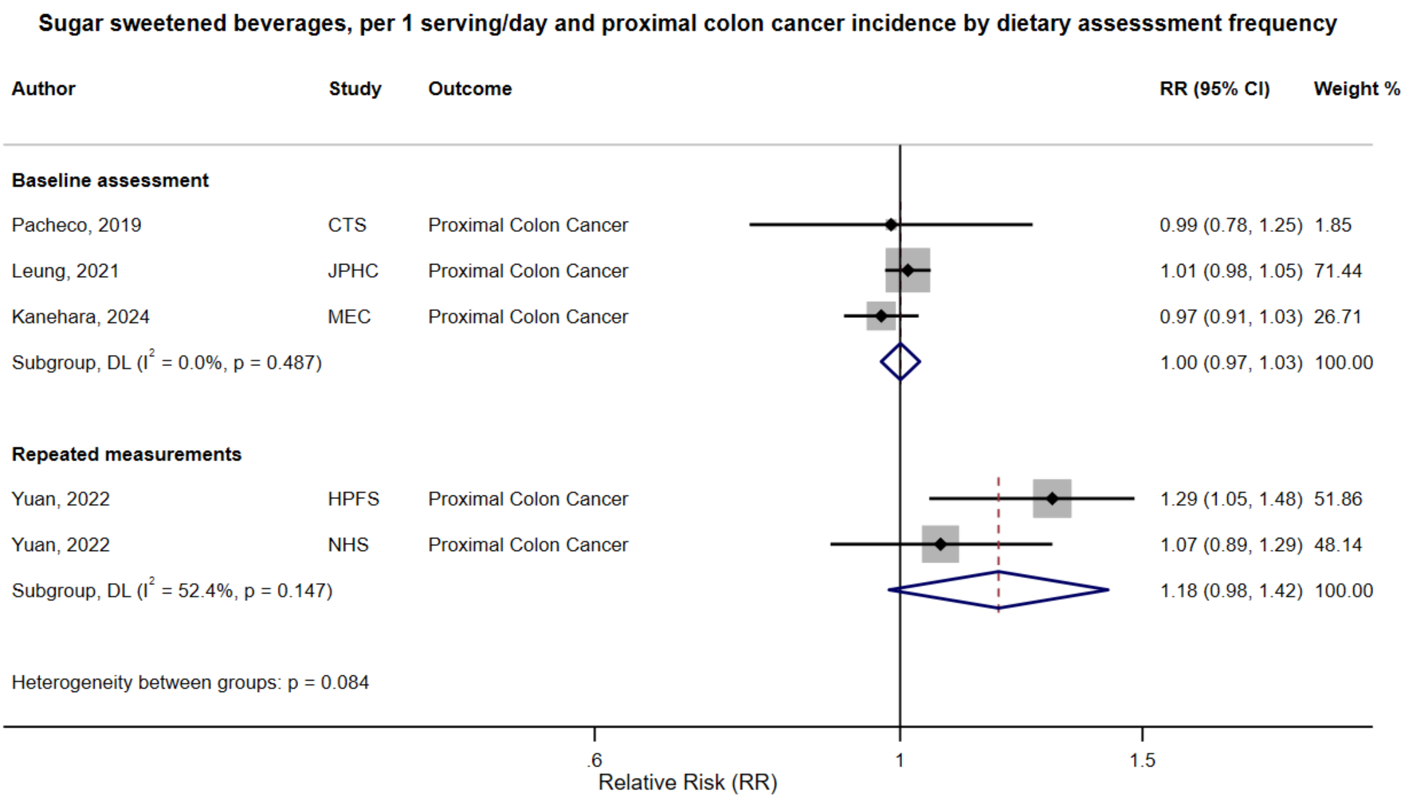

Figure legend: Forest plot shows the linear dose-response results (per 1 serving [355 mL] of sugar sweetened beverages /day) from the inverse variance DerSimonian-Laird random-effects model. The diamond represents the summary relative risk (RR) estimate, the diamond's width is the 95% confidence interval (CI). Each square and the horizontal line across the square represent the RR estimate of the individual study and its 95% CI.

Abbreviations: CI, confidence interval; RR, relative risk.

Study abbreviations: CTS, California Teachers Study; JPHC, Japan Public Health Center-Based Prospective Study; MEC, Multiethnic Cohort Study; HPFS, Health Professionals Follow-up Study; NHS, Nurses' Health Study

Pacheco, 2019[135], Leung, 2021[101], Kanehara, 2024[81], Yuan, 2022[178]

**Supplementary Figure 120. Linear dose-response meta-analysis of sugar sweetened beverages and proximal colon cancer incidence by smoking status.**

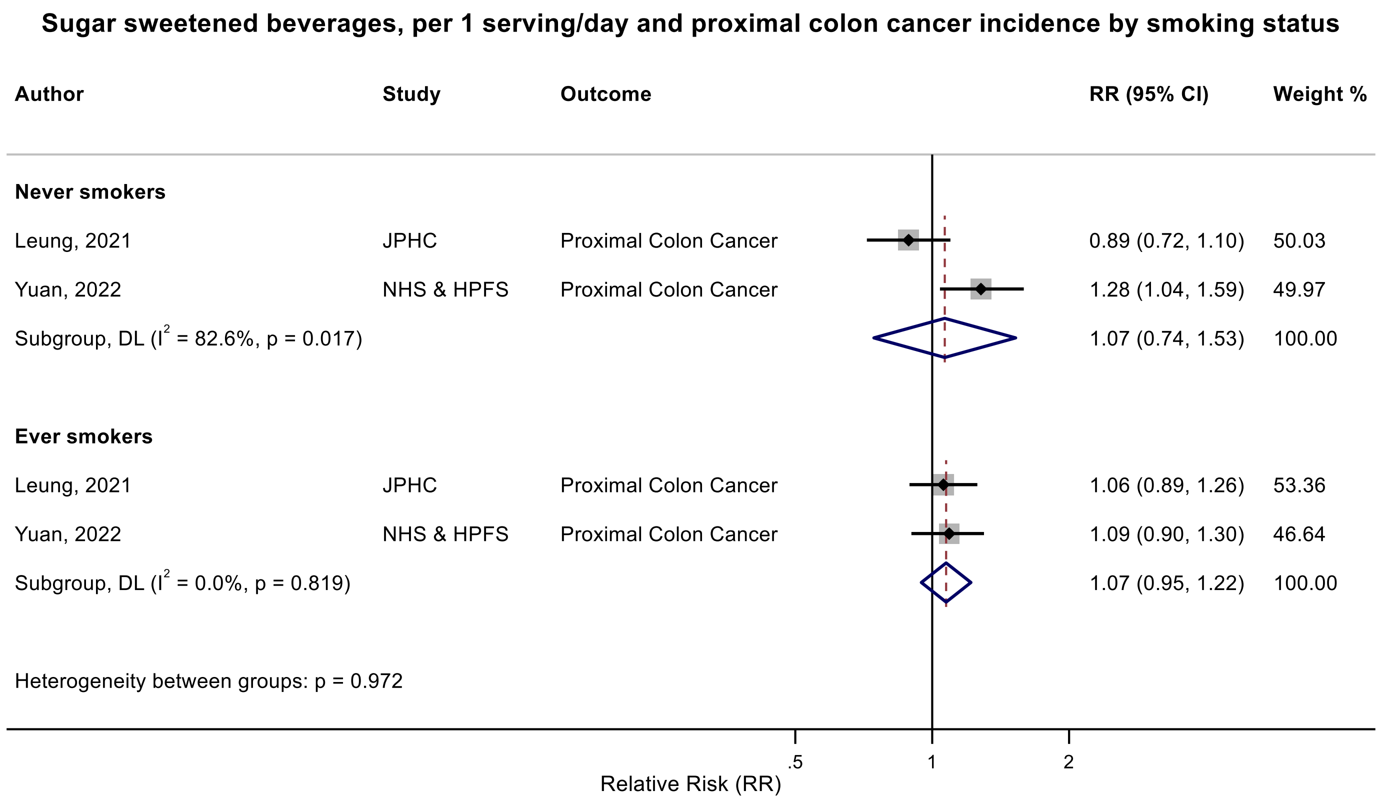

Figure legend: Forest plot shows the linear dose-response results (per 1 serving [355 mL] of sugar sweetened beverages /day) from the inverse variance DerSimonian-Laird random-effects model. The diamond represents the summary relative risk (RR) estimate, the diamond's width is the 95% confidence interval (CI). Each square and the horizontal line across the square represent the RR estimate of the individual study and its 95% CI.

Abbreviations: CI, confidence interval; RR, relative risk.

Study abbreviations: JPHC, Japan Public Health Center-Based Prospective Study; NHS & HPFS, Nurses' Health Study & Health Professionals Follow-up Study

Leung, 2021[101], Yuan, 2022[178]

**Supplementary Figure 121. Linear dose-response meta-analysis of sugar sweetened beverages and proximal colon cancer incidence by alcohol drinking status.**

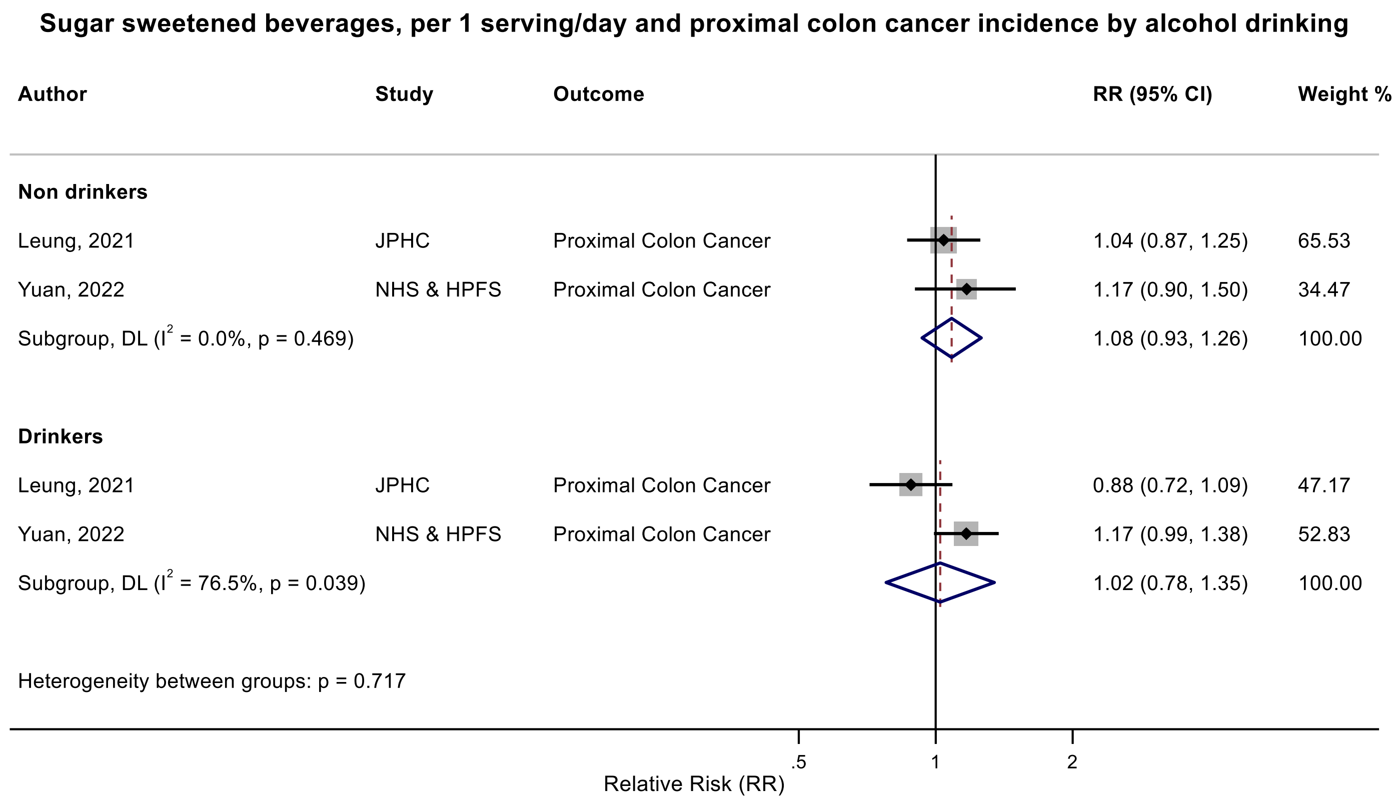

Figure legend: Forest plot shows the linear dose-response results (per 1 serving [355 mL] of sugar sweetened beverages /day) from the inverse variance DerSimonian-Laird random-effects model. The diamond represents the summary relative risk (RR) estimate, the diamond's width is the 95% confidence interval (CI). Each square and the horizontal line across the square represent the RR estimate of the individual study and its 95% CI.

Abbreviations: CI, confidence interval; RR, relative risk.

Study abbreviations: JPHC, Japan Public Health Center-Based Prospective Study; NHS & HPFS, Nurses' Health Study & Health Professionals Follow-up Study

Leung, 2021[101], Yuan, 2022[178]

**Supplementary Figure 122. Linear dose-response meta-analysis of sugar sweetened beverages and proximal colon cancer incidence by physical activity status.**

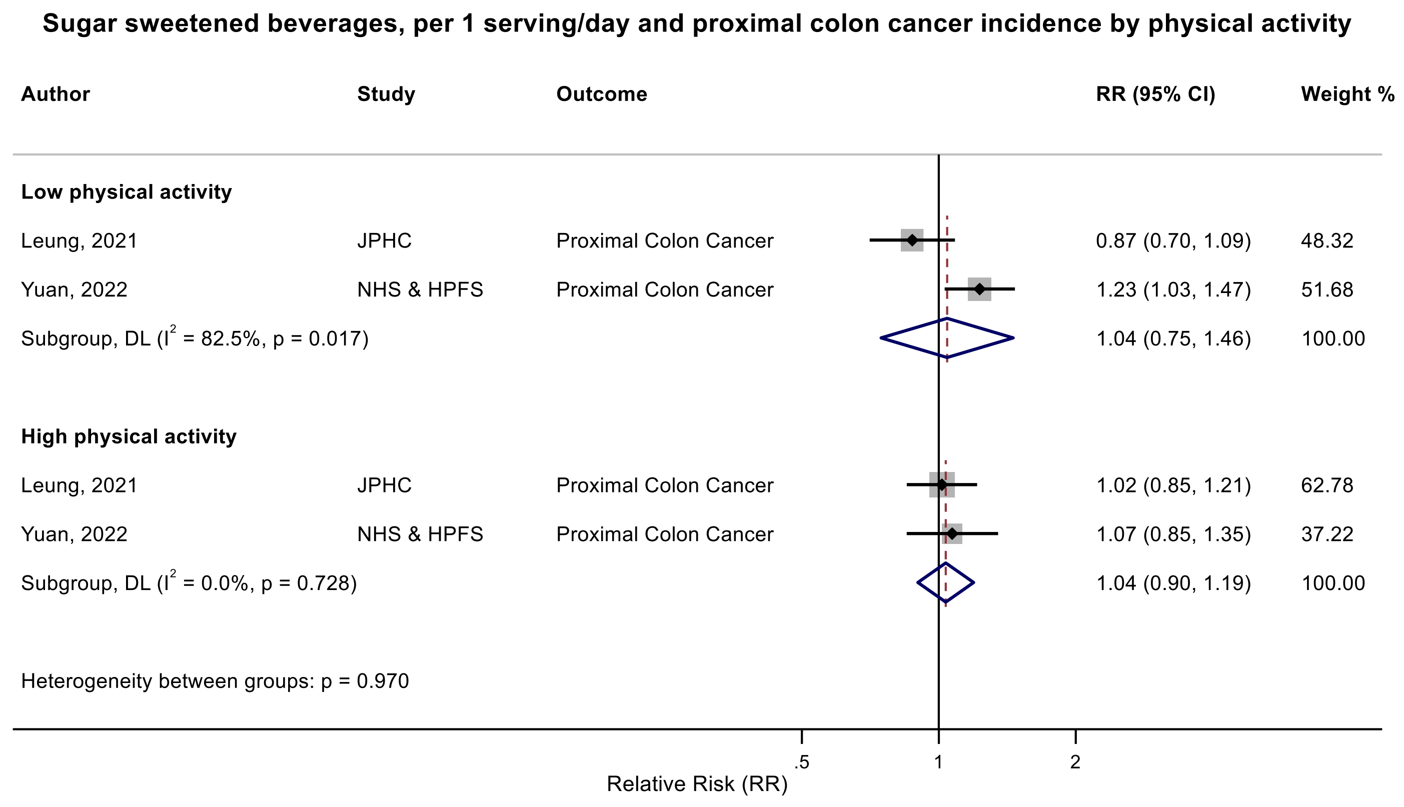

Figure legend: Forest plot shows the linear dose-response results (per 1 serving [355 mL] of sugar sweetened beverages /day) from the inverse variance DerSimonian-Laird random-effects model. The diamond represents the summary relative risk (RR) estimate, the diamond's width is the 95% confidence interval (CI). Each square and the horizontal line across the square represent the RR estimate of the individual study and its 95% CI.

Abbreviations: CI, confidence interval; RR, relative risk.

Study abbreviations: JPHC, Japan Public Health Center-Based Prospective Study; NHS & HPFS, Nurses' Health Study & Health Professionals Follow-up Study

Leung, 2021[101], Yuan, 2022[178]

**Supplementary Figure 123. Linear dose-response meta-analysis of sugar sweetened beverages and proximal colon cancer incidence by body mass index.**

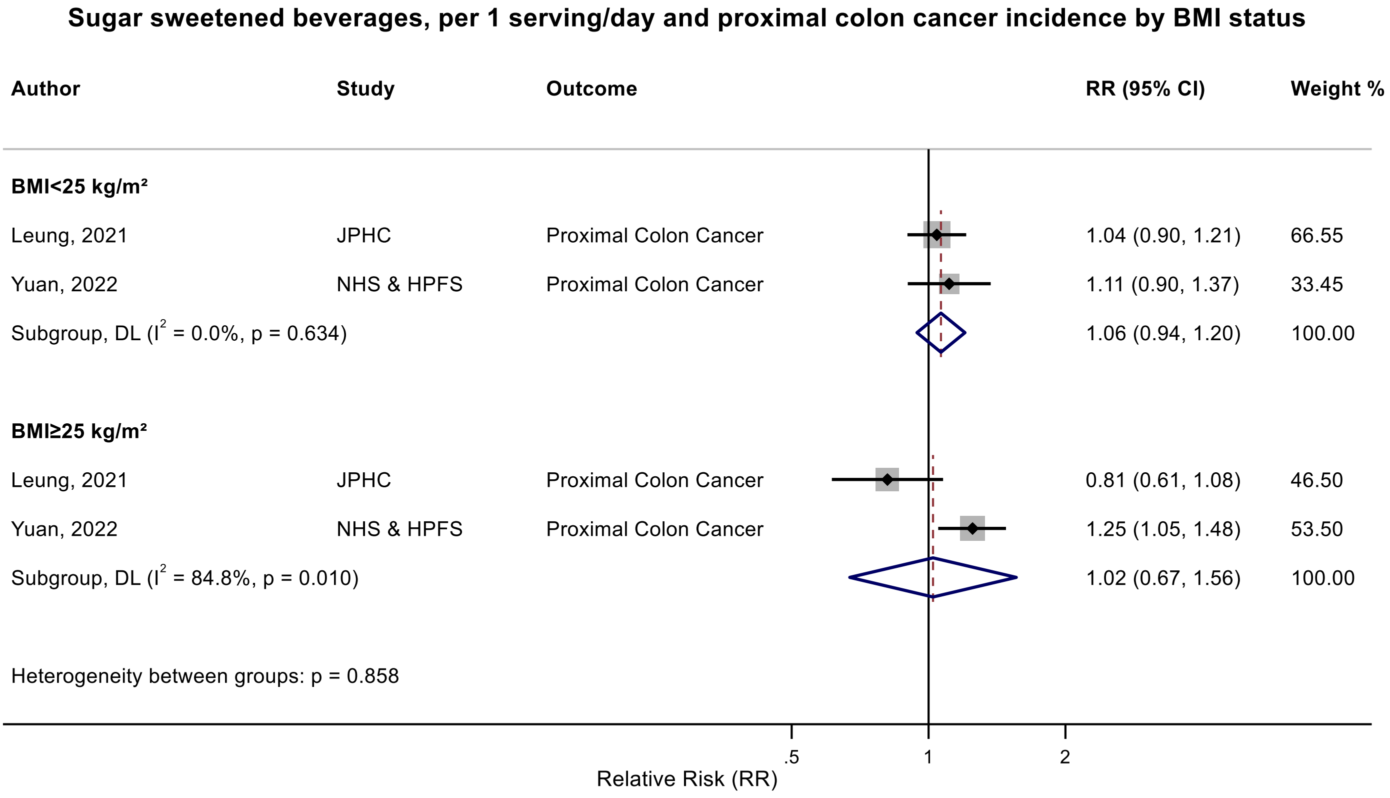

Figure legend: Forest plot shows the linear dose-response results (per 1 serving [355 mL] of sugar sweetened beverages /day) from the inverse variance DerSimonian-Laird random-effects model. The diamond represents the summary relative risk (RR) estimate, the diamond's width is the 95% confidence interval (CI). Each square and the horizontal line across the square represent the RR estimate of the individual study and its 95% CI.

Abbreviations: BMI, body mass index, CI, confidence interval; RR, relative risk.

Study abbreviations: JPHC, Japan Public Health Center-Based Prospective Study; NHS & HPFS, Nurses' Health Study & Health Professionals Follow-up Study

Leung, 2021[101], Yuan, 2022[178]

**Supplementary Figure 124. Linear dose-response meta-analysis of carbonated sugar sweetened beverages and colon cancer incidence by follow-up duration.**

**Carbonated sugar sweetened beverages, per 1 serving/day and colon cancer incidence by follow-up duration**

| Author | Study | Outcome | RR (95% CI) | Weight % |
|---|---|---|---|---|
| **>10 years** | | | | |
| Zhang, 2010 | CNBSS | Colon Cancer | 1.08 (0.76, 1.50) | 7.38 |
| Zhang, 2010 | IWHS | Colon Cancer | 0.93 (0.72, 1.18) | 14.33 |
| Zhang, 2010 | ATBC | Colon Cancer | 0.98 (0.56, 1.73) | 2.69 |
| Zhang, 2010 | BCDDP | Colon Cancer | 0.52 (0.29, 0.92) | 2.54 |
| Zhang, 2010 | HPFS | Colon Cancer | 1.14 (0.94, 1.38) | 23.25 |
| Zhang, 2010 | SMC | Colon Cancer | 1.09 (0.53, 2.24) | 1.65 |
| Subgroup, DL ($I^2$ = 30.3%, p = 0.208) | | | 0.99 (0.83, 1.18) | 51.84 |
| **≤10 years** | | | | |
| Zhang, 2010 | NLCS | Colon Cancer | 1.07 (0.67, 1.72) | 3.88 |
| Zhang, 2010 | CPS II | Colon Cancer | 0.90 (0.72, 1.13) | 16.84 |
| Zhang, 2010 | WHS | Colon Cancer | 0.93 (0.62, 1.40) | 5.07 |
| Zhang, 2010 | NHS | Colon Cancer | 1.03 (0.85, 1.25) | 22.37 |
| Subgroup, DL ($I^2$ = 0.0%, p = 0.801) | | | 0.98 (0.85, 1.12) | 48.16 |
| Heterogeneity between groups: p = 0.889 | | | | |
| Overall, DL ($I^2$ = 0.0%, p = 0.495) | | | 1.00 (0.91, 1.10) | 100.00 |

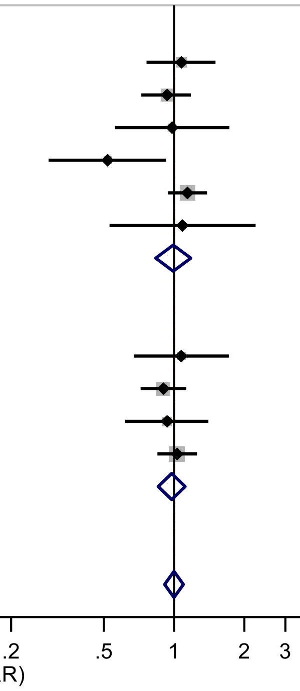

Figure legend: Forest plot shows the linear dose-response results (per 1 serving [355 mL] of carbonated sugar sweetened beverages /day) from the inverse variance DerSimonian-Laird random-effects model. The diamond represents the summary relative risk (RR) estimate, the diamond's width is the 95% confidence interval (CI). Each square and the horizontal line across the square represent the RR estimate of the individual study and its 95% CI.

Abbreviations: CI, confidence interval; RR, relative risk.

Study abbreviations: CNBSS, Canadian National Breast Screening Study; IWHS, The Iowa Women's Health Study; ATBC, Alpha-Tocopherol, Beta-Carotene Cancer Prevention Study; BCDDP, Breast Cancer Detection Demonstration Project follow-up study; HPFS, Health Professionals Follow-up Study; SMC, Swedish Mammography Cohort; NLCS, Netherlands Cohort Study on Diet and Cancer; CPS II, Cancer Prevention Study II; WHS, Women's Health Study; NHS, Nurses' Health Study

Zhang, 2010[182]

**Supplementary Figure 125. Linear dose-response meta-analysis of carbonated sugar sweetened beverages and colon cancer incidence by geographical region.**

**Carbonated sugar sweetened beverages, per 1 serving/day and colon cancer incidence by region**

| Author | Study | Outcome | RR (95% CI) | Weight % |
|---|---|---|---|---|
| **North America** | | | | |
| Zhang, 2010 | CNBSS | Colon Cancer | 1.08 (0.76, 1.50) | 7.38 |
| Zhang, 2010 | IWHS | Colon Cancer | 0.93 (0.72, 1.18) | 14.33 |
| Zhang, 2010 | BCDDP | Colon Cancer | 0.52 (0.29, 0.92) | 2.54 |
| Zhang, 2010 | HPFS | Colon Cancer | 1.14 (0.94, 1.38) | 23.25 |
| Zhang, 2010 | CPS II | Colon Cancer | 0.90 (0.72, 1.13) | 16.84 |
| Zhang, 2010 | WHS | Colon Cancer | 0.93 (0.62, 1.40) | 5.07 |
| Zhang, 2010 | NHS | Colon Cancer | 1.03 (0.85, 1.25) | 22.37 |
| Subgroup, DL ($I^2$ = 27.2%, p = 0.221) | | | 0.98 (0.87, 1.11) | 91.79 |
| **Europe** | | | | |
| Zhang, 2010 | ATBC | Colon Cancer | 0.98 (0.56, 1.73) | 2.69 |
| Zhang, 2010 | SMC | Colon Cancer | 1.09 (0.53, 2.24) | 1.65 |
| Zhang, 2010 | NLCS | Colon Cancer | 1.07 (0.67, 1.72) | 3.88 |
| Subgroup, DL ($I^2$ = 0.0%, p = 0.965) | | | 1.04 (0.76, 1.44) | 8.21 |
| Heterogeneity between groups: p = 0.727 | | | | |
| Overall, DL ($I^2$ = 0.0%, p = 0.495) | | | 1.00 (0.91, 1.10) | 100.00 |

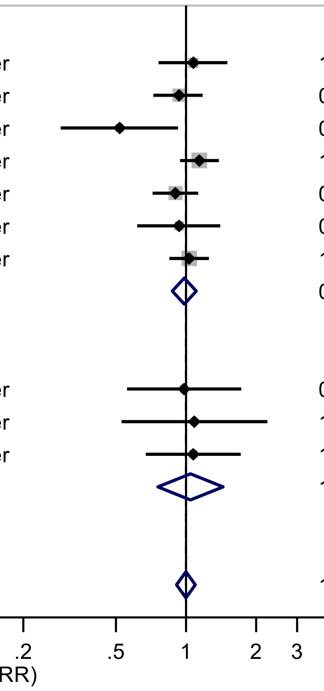

Figure legend: Forest plot shows the linear dose-response results (per 1 serving [355 mL] of carbonated sugar sweetened beverages /day) from the inverse variance DerSimonian-Laird random-effects model. The diamond represents the summary relative risk (RR) estimate, the diamond's width is the 95% confidence interval (CI). Each square and the horizontal line across the square represent the RR estimate of the individual study and its 95% CI.

Abbreviations: CI, confidence interval; RR, relative risk.

Study abbreviations: CNBSS, Canadian National Breast Screening Study; IWHS, The Iowa Women's Health Study; BCDDP, Breast Cancer Detection Demonstration Project follow-up study; HPFS, Health Professionals Follow-up Study; CPS II, Cancer Prevention Study II; WHS, Women's Health Study; NHS, Nurses' Health Study; ATBC, Alpha-Tocopherol, Beta-Carotene Cancer Prevention Study; SMC, Swedish Mammography Cohort; NLCS, Netherlands Cohort Study on Diet and Cancer

Note: The results have been extracted from a pooled analysis.

Zhang, 2010[182]

**Supplementary Figure 126. Linear dose-response meta-analysis of carbonated sugar sweetened beverages and colon cancer incidence by frequency of dietary assessment.**

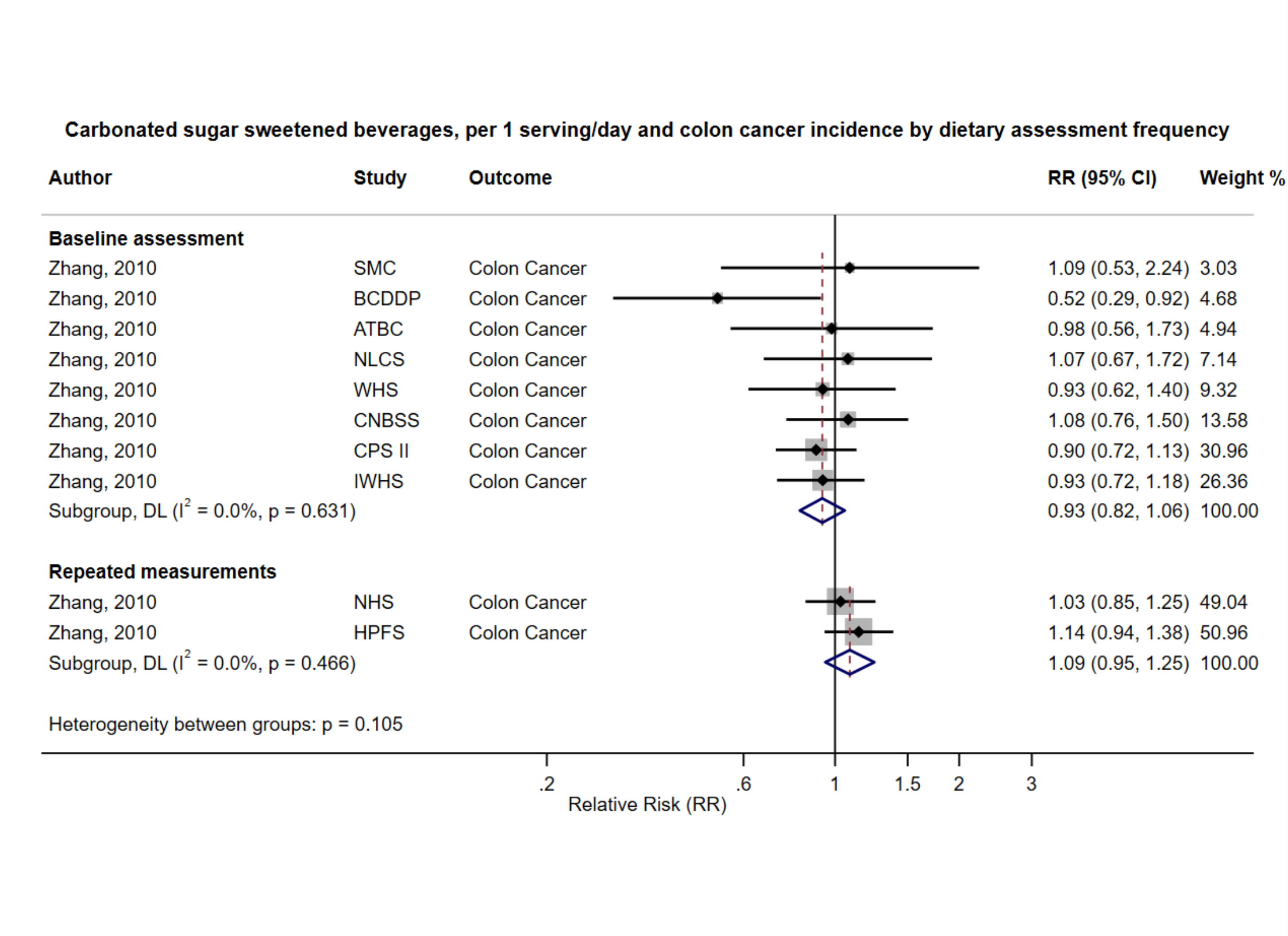

Figure legend: Forest plot shows the linear dose-response results (per 1 serving [355 mL] of carbonated sugar sweetened beverages /day) from the inverse variance DerSimonian-Laird random-effects model. The diamond represents the summary relative risk (RR) estimate, the diamond's width is the 95% confidence interval (CI). Each square and the horizontal line across the square represent the RR estimate of the individual study and its 95% CI.

Abbreviations: CI, confidence interval; RR, relative risk.

Study abbreviations: SMC, Swedish Mammography Cohort; BCDDP, Breast Cancer Detection Demonstration Project follow-up study; ATBC, Alpha-Tocopherol, Beta-Carotene Cancer Prevention Study; NLCS, Netherlands Cohort Study on Diet and Cancer; WHS, Women’s Health Study; CNBSS, Canadian National Breast Screening Study; CPS II, Cancer Prevention Study II; IWHS, The Iowa Women’s Health Study; NHS, Nurses’ Health Study; HPFS, Health Professionals Follow-up Study

Note: The results have been extracted from a published pooled analysis.

Zhang, 2010[182]

**Supplementary Figure 127. Linear dose-response meta-analysis of carbonated sugar sweetened beverages and colon cancer incidence by various factors.**

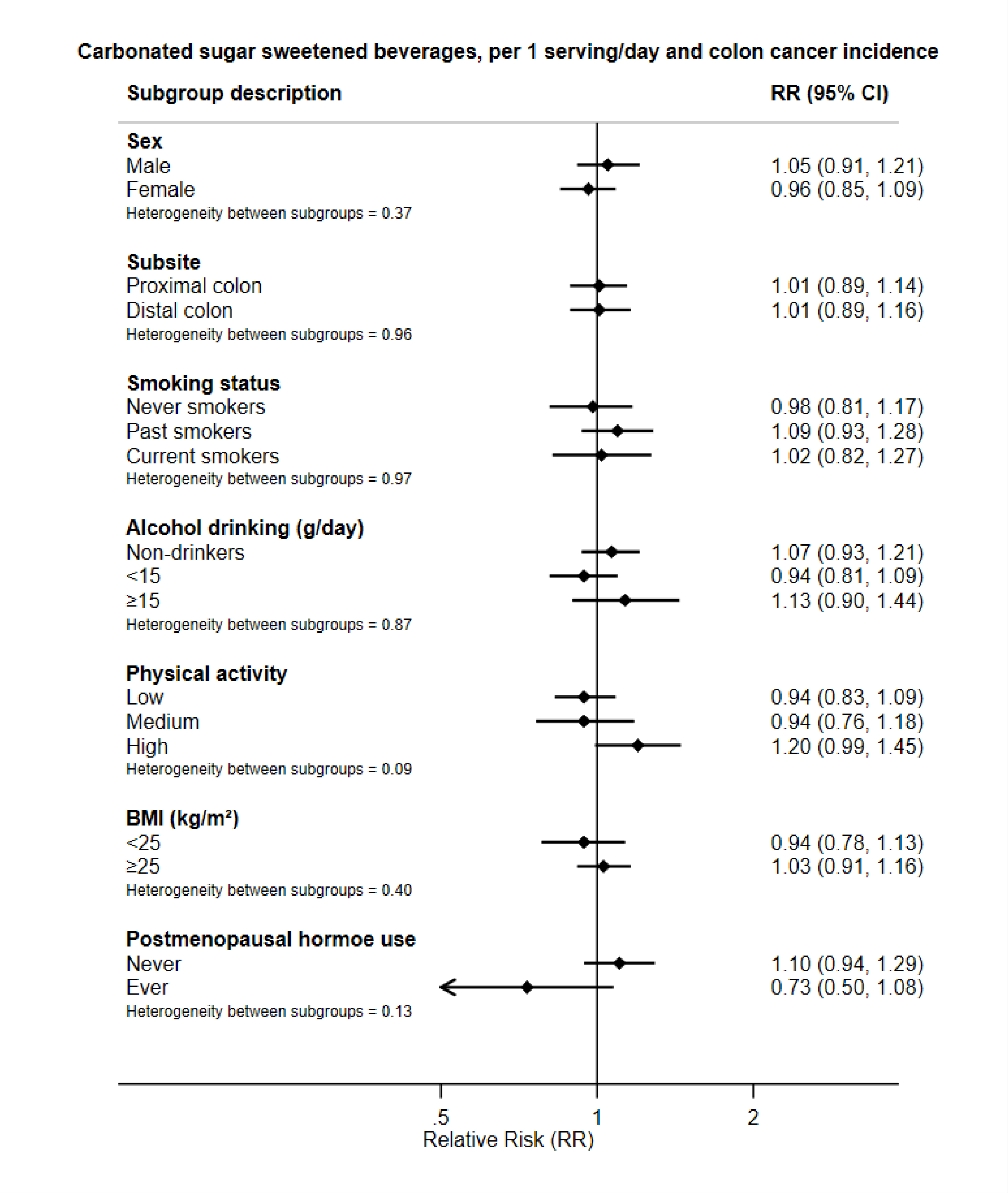

Figure legend: Forest plot shows the linear dose-response results (per 1 serving [355 mL] of carbonated sugar sweetened beverages /day) from the inverse variance DerSimonian-Laird random-effects model. The diamond represents the summary relative risk (RR) estimate, the diamond's width is the 95% confidence interval (CI).

Abbreviations: CI, confidence interval; RR, relative risk.

Note: The results have been extracted from a published pooled analysis.

Zhang, 2010[182]

**Supplementary Figure 128. Linear dose-response meta-analysis of sugar sweetened beverages and distal colon cancer incidence by frequency of dietary assessment.**

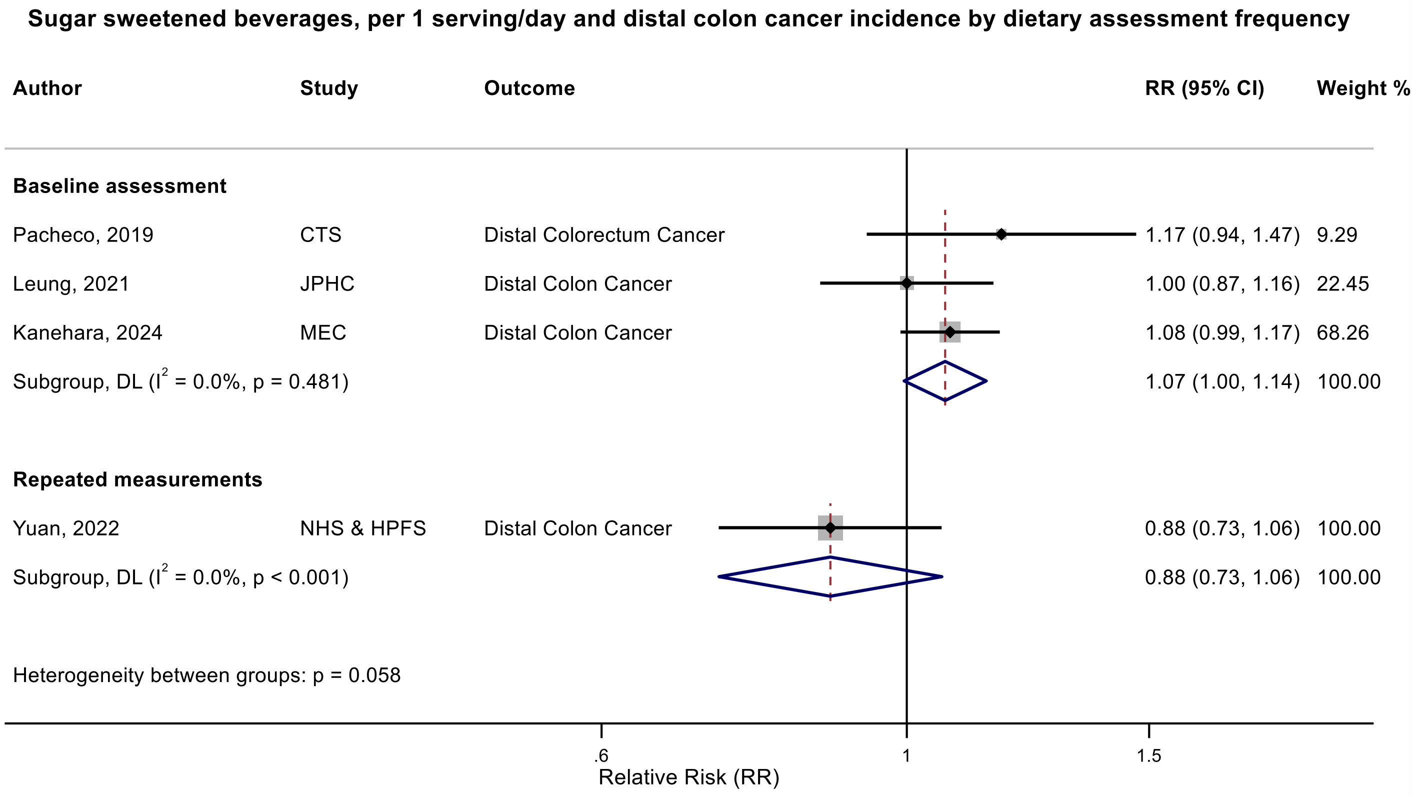

Figure legend: Forest plot shows the linear dose-response results (per 1 serving [355 mL] of sugar sweetened beverages /day) from the inverse variance DerSimonian-Laird random-effects model. The diamond represents the summary relative risk (RR) estimate, the diamond's width is the 95% confidence interval (CI). Each square and the horizontal line across the square represent the RR estimate of the individual study and its 95% CI.

Abbreviations: CI, confidence interval; RR, relative risk.

Study abbreviations: CTS, California Teachers Study; JPHC, Japan Public Health Center-Based Prospective Study; MEC, Multiethnic Cohort Study; NHS & HPFS, Nurses' Health Study & Health Professionals Follow-up Study

Pacheco, 2019[135], Leung, 2021[101], Kanehara, 2024[81], Yuan, 2022[178]

**Supplementary Figure 129. Linear dose-response meta-analysis of sugar sweetened beverages and rectal cancer incidence by frequency of dietary assessment.**

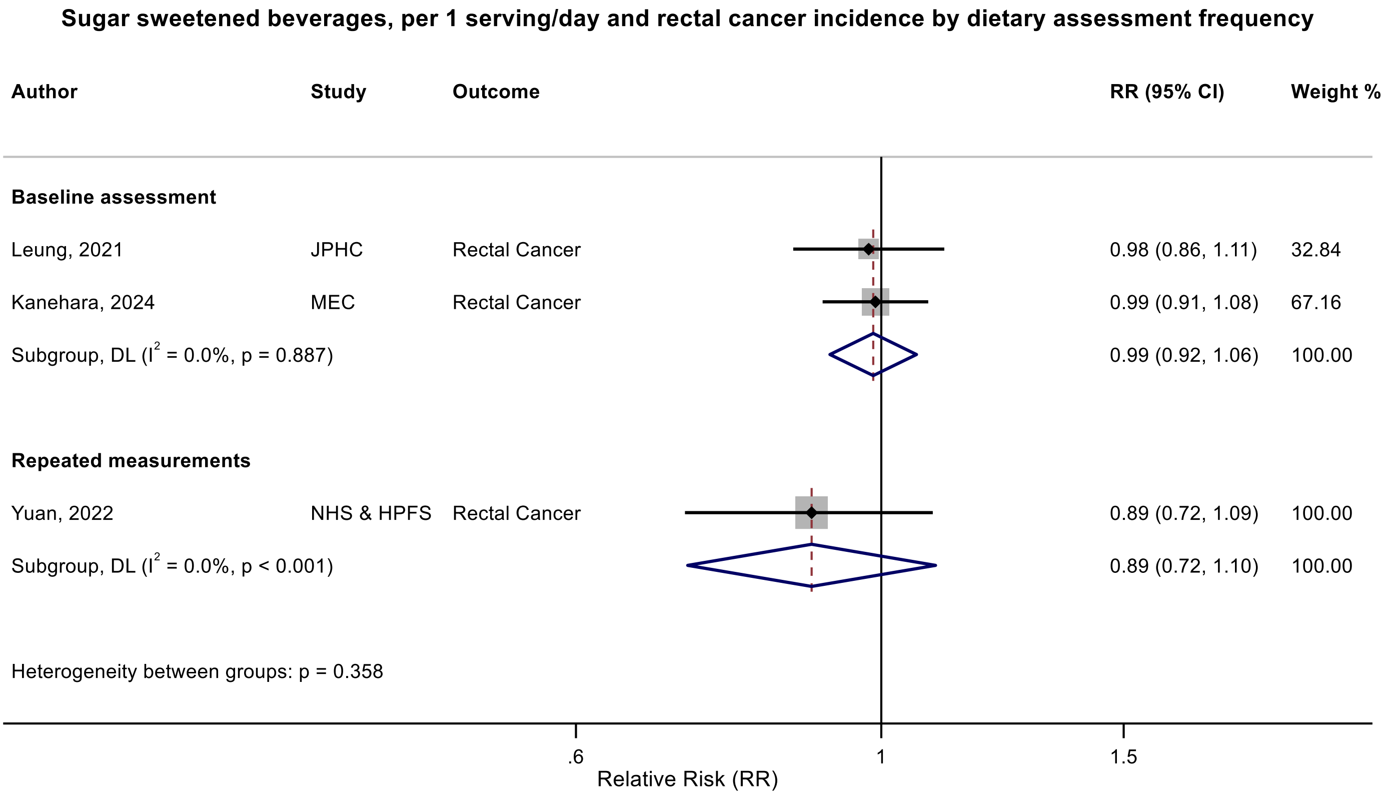

Figure legend: Forest plot shows the linear dose-response results (per 1 serving [355 mL] of sugar sweetened beverages /day) from the inverse variance DerSimonian-Laird random-effects model. The diamond represents the summary relative risk (RR) estimate, the diamond's width is the 95% confidence interval (CI). Each square and the horizontal line across the square represent the RR estimate of the individual study and its 95% CI.

Abbreviations: CI, confidence interval; RR, relative risk.

Study abbreviations: JPHC, Japan Public Health Center-Based Prospective Study; MEC, Multiethnic Cohort Study; NHS & HPFS, Nurses' Health Study & Health Professionals Follow-up Study

Leung, 2021[101], Kanehara, 2024[81], Yuan, 2022[178]

**Supplementary Figure 130. Linear dose-response meta-analysis of sugar sweetened beverages and liver cancer by outcome type.**

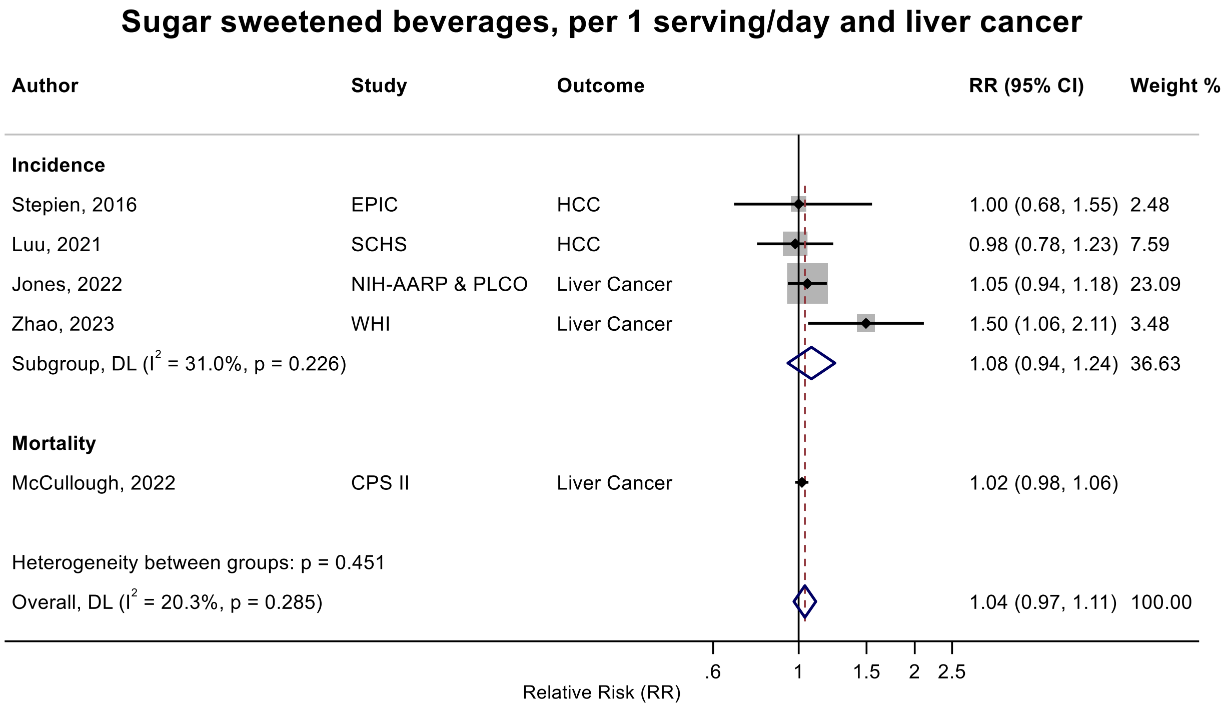

Figure legend: Forest plot shows the linear dose-response results (per 1 serving [355 mL] of sugar sweetened beverages /day) from the inverse variance DerSimonian-Laird random-effects model. The diamond represents the summary relative risk (RR) estimate, the diamond's width is the 95% confidence interval (CI). Each square and the horizontal line across the square represent the RR estimate of the individual study and its 95% CI.

Abbreviations: CI, confidence interval; RR, relative risk.

Study abbreviations: SCHS, Singapore Chinese Health study; NIH-AARP & PLCO, National Institute of Health + American Association of Retired Persons Diet and Health Study & Prostate, Lung, Colorectal and Ovarian Cancer Screening trial; WHI, Women's Health Initiative Study; CPS II, Cancer Prevention Study II

Luu, 2021[106], Jones, 2022[79], Zhao, 2023[186], McCullough, 2022[113]

**Supplementary Figure 131. Linear dose-response meta-analysis of sugar sweetened beverages and liver cancer incidence and mortality combined by adjustment for body mass index.**

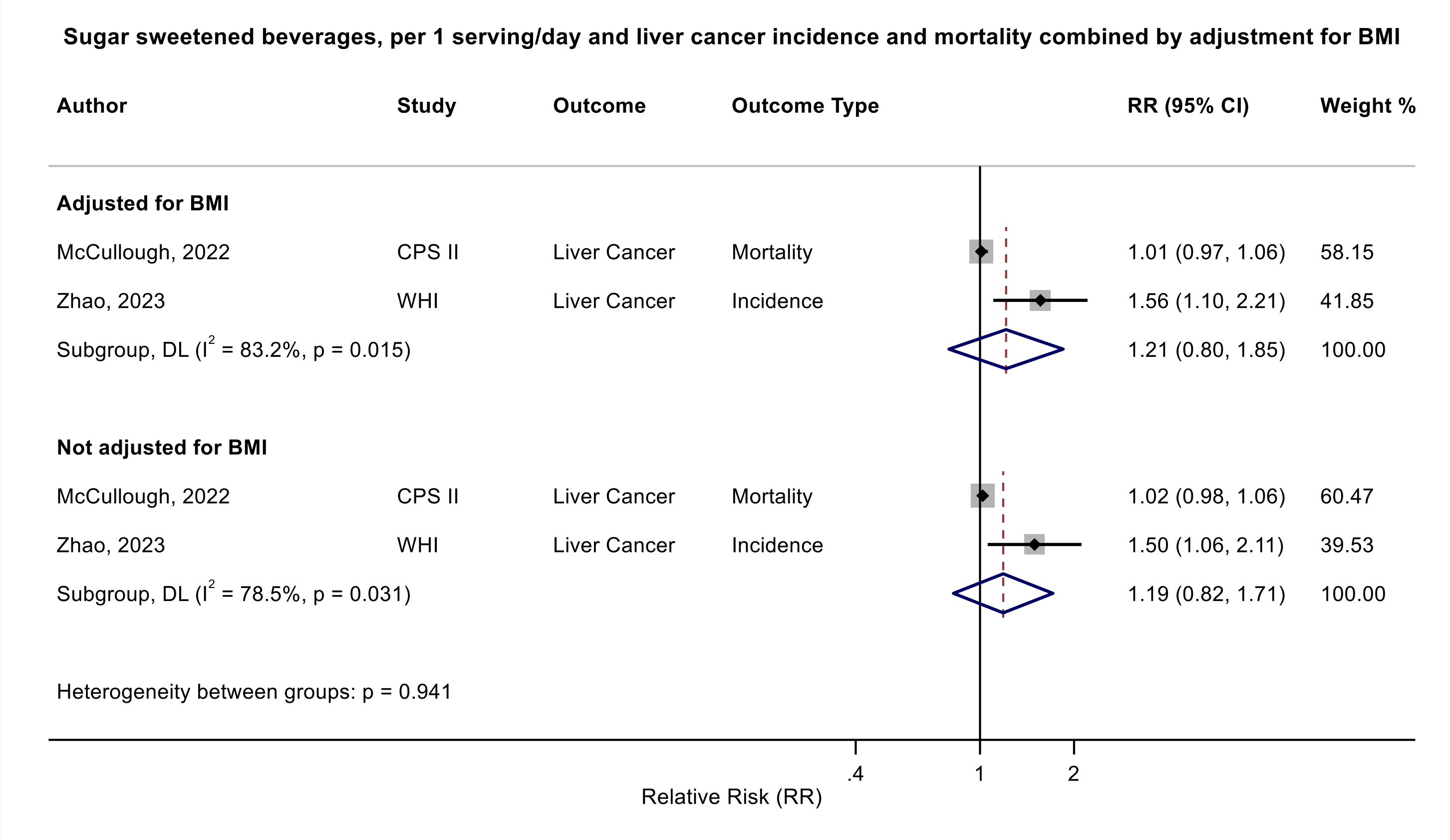

Figure legend: Forest plot shows the linear dose-response results (per 1 serving [355 mL] of sugar sweetened beverages /day) from the inverse variance DerSimonian-Laird random-effects model. The diamond represents the summary relative risk (RR) estimate, the diamond's width is the 95% confidence interval (CI). Each square and the horizontal line across the square represent the RR estimate of the individual study and its 95% CI.

Abbreviations: BMI, body mass index, CI, confidence interval; RR, relative risk.

Study abbreviations: CPS II, Cancer Prevention Study II; WHI, Women's Health Initiative Study

Note: This subgroup analysis is based on a within study comparison in two studies that reported association estimates with and without adjustment for BMI. Since only two studies, one on incidence and one on mortality, reported risk estimates both with and without adjustment for BMI, we combined incidence and mortality to be able to perform this important subgroup analysis.

McCullough, 2022[113], Zhao, 2023[186]

**Supplementary Figure 132. Linear dose-response meta-analysis of sugar sweetened beverages and liver cancer incidence by exclusion of people with a history of diabetes.**

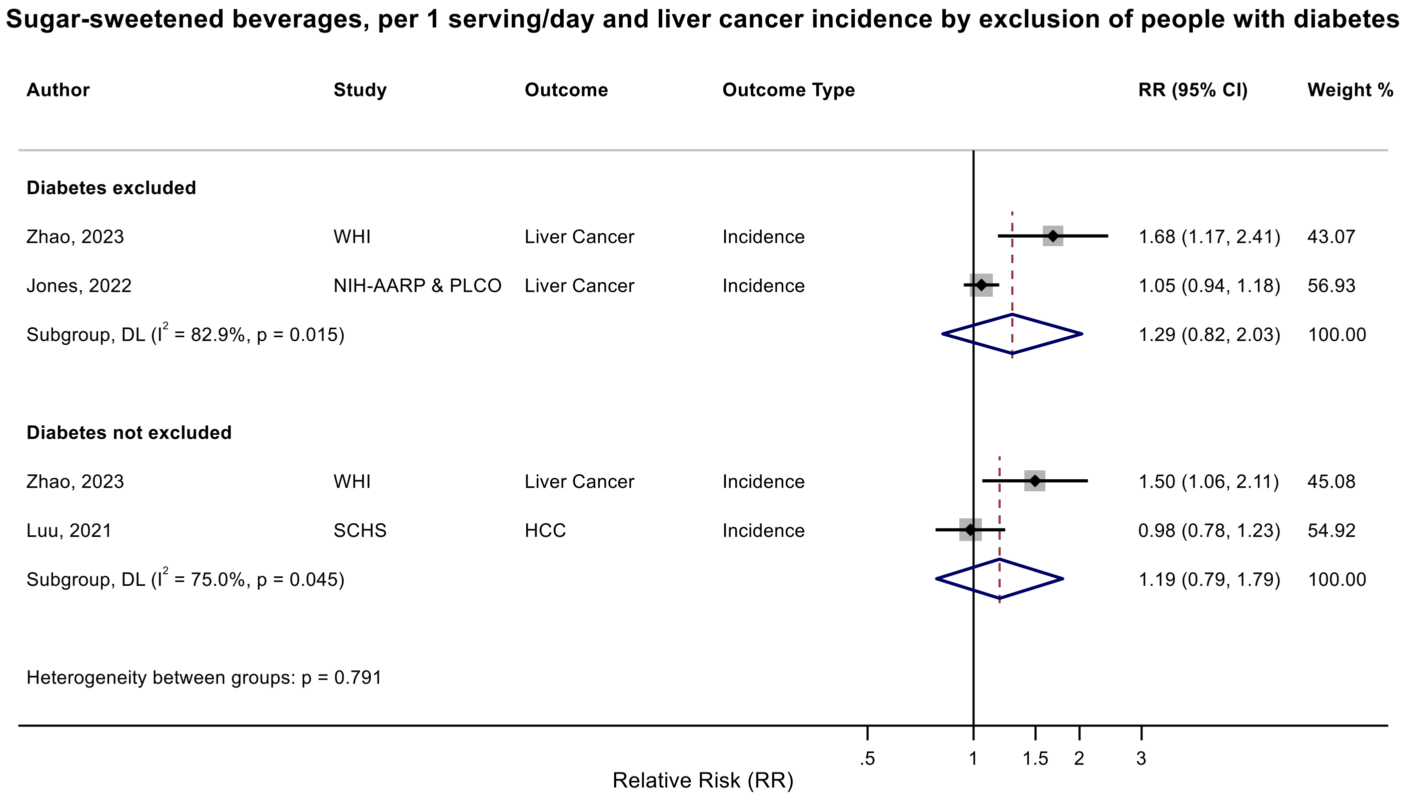

Figure legend: Forest plot shows the linear dose-response results (per 1 serving [355 mL] of sugar sweetened beverages /day) from the inverse variance DerSimonian-Laird random-effects model. The diamond represents the summary relative risk (RR) estimate, the diamond's width is the 95% confidence interval (CI). Each square and the horizontal line across the square represent the RR estimate of the individual study and its 95% CI.

Abbreviations: CI, confidence interval; RR, relative risk.

Study abbreviations: SCHS, Singapore Chinese Health study; NIH-AARP & PLCO, National Institute of Health + American Association of Retired Persons Diet and Health Study & Prostate, Lung, Colorectal and Ovarian Cancer Screening trial; WHI, Women’s Health Initiative Study

Luu, 2021[106], Jones, 2022[79], Zhao, 2023[186]

**Supplementary Figure 133. Linear dose-response meta-analysis of sugar sweetened beverages and pancreatic cancer by outcome type.**

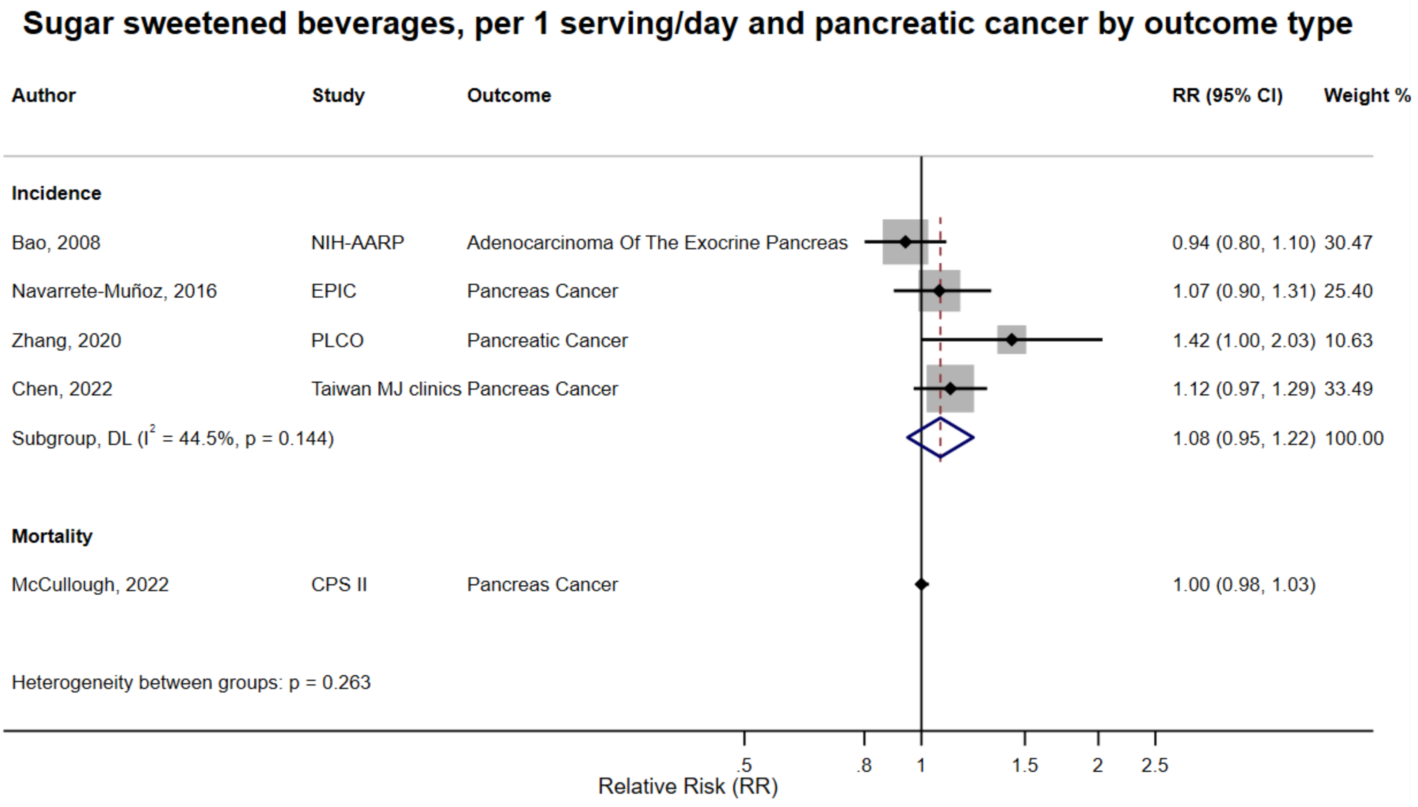

Figure legend: Forest plot shows the linear dose-response results (per 1 serving [355 mL] of sugar sweetened beverages /day) from the inverse variance DerSimonian-Laird random-effects model. The diamond represents the summary relative risk (RR) estimate, the diamond's width is the 95% confidence interval (CI). Each square and the horizontal line across the square represent the RR estimate of the individual study and its 95% CI.

Abbreviations: CI, confidence interval; RR, relative risk.

Study abbreviations: NIH-AARP, National Institute of Health + American Association of Retired Persons Diet and Health Study; EPIC, European Prospective Investigation into Cancer and nutrition; PLCO, Prostate, Lung, Colorectal and Ovarian Cancer Screening trial; Taiwan MJ clinics, The Taiwan MJ Cohort Study; CPS II, Cancer Prevention Study II

Bao, 2008[39], Navarrete-Muñoz, 2016[127], Zhang, 2020[184], Chen, 2022[45], McCullough, 2022[113]

**Supplementary Figure 134. Linear dose-response meta-analysis of sugar sweetened beverages and pancreatic cancer incidence and mortality combined by body mass index.**

**Sugar sweetened beverages, per 1 serving/day and pancreatic cancer incidence and mortality combined, by BMI status**

| Author | Study | Outcome | Outcome Type | RR (95% CI) | Weight % |
|---|---|---|---|---|---|
| **BMI<30 kg/m²** | | | | | |
| Bao, 2008 | NIH-AARP | Adenocarcinoma Of The Exocrine Pancreas | Incidence | 1.03 (0.80, 1.31) | 1.77 |
| McCullough, 2022 | CPS II | Pancreas Cancer | Mortality | 1.02 (0.98, 1.05) | 98.23 |
| Subgroup, DL ($I^2$ = 0.0%, p = 0.942) | | | | 1.02 (0.98, 1.05) | 100.00 |
| **BMI≥30kg/m²** | | | | | |
| Bao, 2008 | NIH-AARP | Adenocarcinoma Of The Exocrine Pancreas | Incidence | 1.18 (0.71, 1.97) | 0.64 |
| McCullough, 2022 | CPS II | Pancreas Cancer | Mortality | 0.98 (0.94, 1.02) | 99.36 |
| Subgroup, DL ($I^2$ = 0.0%, p = 0.476) | | | | 0.98 (0.94, 1.02) | 100.00 |
| Heterogeneity between groups: p = 0.184 | | | | | |

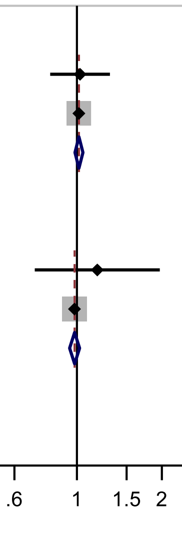

Figure legend: Forest plot shows the linear dose-response results (per 1 serving [355 mL] of sugar sweetened beverages /day) from the inverse variance DerSimonian-Laird random-effects model. The diamond represents the summary relative risk (RR) estimate, the diamond's width is the 95% confidence interval (CI). Each square and the horizontal line across the square represent the RR estimate of the individual study and its 95% CI.

Abbreviations: CI, confidence interval; RR, relative risk.

Study abbreviations: NIH-AARP, National Institute of Health + American Association of Retired Persons Diet and Health Study; CPS II, Cancer Prevention Study II

Note: Since only two studies, one on incidence and one on mortality, reported risk estimates for subgroup analysis by BMI, we combined incidence and mortality to be able to perform this important subgroup analysis.

Bao, 2008[39], McCullough, 2022[113]

**Supplementary Figure 135. Linear dose-response meta-analysis of sugar sweetened beverages and pancreatic cancer incidence and mortality combined by sex.**

**Sugar sweetened beverages, per 1 serving/day and pancreatic cancer incidence and mortality combined by sex**

| Author | Study | Outcome | Outcome Type | RR (95% CI) | Weight % |
|---|---|---|---|---|---|
| **Male** | | | | | |
| Navarrete-Muñoz, 2016 | EPIC | Pancreas Cancer | Incidence | 1.04 (0.81, 1.34) | 1.38 |
| McCullough, 2022 | CPS II | Pancreas Cancer | Mortality | 1.01 (0.98, 1.04) | 98.62 |
| Subgroup, DL ($I^2$ = 0.0%, p = 0.807) | | | | 1.01 (0.98, 1.04) | 100.00 |
| **Female** | | | | | |
| Navarrete-Muñoz, 2016 | EPIC | Pancreas Cancer | Incidence | 1.15 (0.86, 1.53) | 2.09 |
| McCullough, 2022 | CPS II | Pancreas Cancer | Mortality | 0.99 (0.95, 1.03) | 97.91 |
| Subgroup, DL ($I^2$ = 0.3%, p = 0.317) | | | | 0.99 (0.95, 1.04) | 100.00 |
| Heterogeneity between groups: p = 0.505 | | | | | |

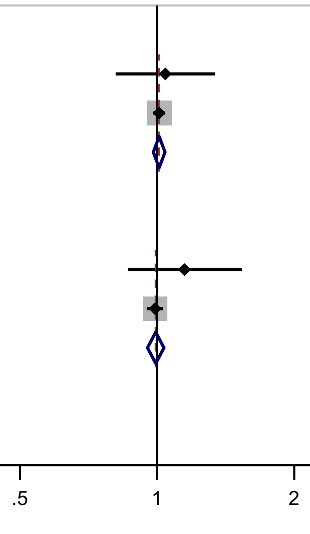

Figure legend: Forest plot shows the linear dose-response results (per 1 serving [355 mL] of sugar sweetened beverages /day) from the inverse variance DerSimonian-Laird random-effects model. The diamond represents the summary relative risk (RR) estimate, the diamond's width is the 95% confidence interval (CI). Each square and the horizontal line across the square represent the RR estimate of the individual study and its 95% CI.

Abbreviations: CI, confidence interval; RR, relative risk.

Study abbreviations: EPIC, European Prospective Investigation into Cancer and nutrition; CPS II, Cancer Prevention Study II

Note: Since only two studies, one on incidence and one on mortality, reported risk estimates for subgroup analysis by sex, we combined incidence and mortality to be able to perform this important subgroup analysis.

Navarrete-Muñoz, 2016[127], McCullough, 2022[113]

**Supplementary Figure 136. Linear dose-response meta-analysis of sugar sweetened beverages and pancreatic cancer incidence by smoking status.**

**Sugar sweetened beverages, per 1 serving/day and pancreatic cancer incidence by smoking status**

| Author | Study | Outcome | Outcome Type | RR (95% CI) | Weight % |
|---|---|---|---|---|---|
| **Never smokers** | | | | | |
| Navarrete-Muñoz, 2016 | EPIC | Pancreas Cancer | Incidence | 0.92 (0.59, 1.43) | 30.38 |
| Bao, 2008 | NIH-AARP | Adenocarcinoma Of The Exocrine Pancreas | Incidence | 1.12 (0.83, 1.49) | 69.62 |
| Subgroup, DL ($I^2$ = 0.0%, p = 0.462) | | | | 1.05 (0.82, 1.34) | 100.00 |
| **Former smokers** | | | | | |
| Navarrete-Muñoz, 2016 | EPIC | Pancreas Cancer | Incidence | 1.19 (0.85, 1.68) | 100.00 |
| Subgroup, DL ($I^2$ = 0.0%, p < 0.001) | | | | 1.19 (0.85, 1.67) | 100.00 |
| **Current smokers** | | | | | |
| Navarrete-Muñoz, 2016 | EPIC | Pancreas Cancer | Incidence | 1.13 (0.84, 1.50) | 58.39 |
| Bao, 2008 | NIH-AARP | Adenocarcinoma Of The Exocrine Pancreas | Incidence | 1.00 (0.71, 1.41) | 41.61 |
| Subgroup, DL ($I^2$ = 0.0%, p = 0.609) | | | | 1.07 (0.86, 1.34) | 100.00 |

Heterogeneity between groups: p = 0.831

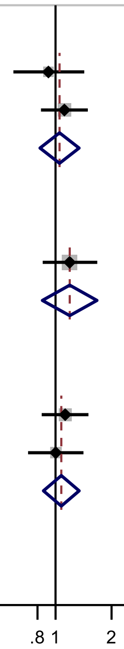

Relative Risk (RR)

Figure legend: Forest plot shows the linear dose-response results (per 1 serving [355 mL] of sugar sweetened beverages /day) from the inverse variance DerSimonian-Laird random-effects model. The diamond represents the summary relative risk (RR) estimate, the diamond's width is the 95% confidence interval (CI). Each square and the horizontal line across the square represent the RR estimate of the individual study and its 95% CI.

Abbreviations: CI, confidence interval; RR, relative risk.

Study abbreviations: EPIC, European Prospective Investigation into Cancer and nutrition; NIH-AARP, National Institute of Health + American Association of Retired Persons Diet and Health Study

Navarrete-Muñoz, 2016[127], Bao, 2008[39]

**Supplementary Figure 137. Linear dose-response meta-analysis of sugar sweetened beverages and pancreatic cancer incidence and mortality combined by adjustment for body mass index.**

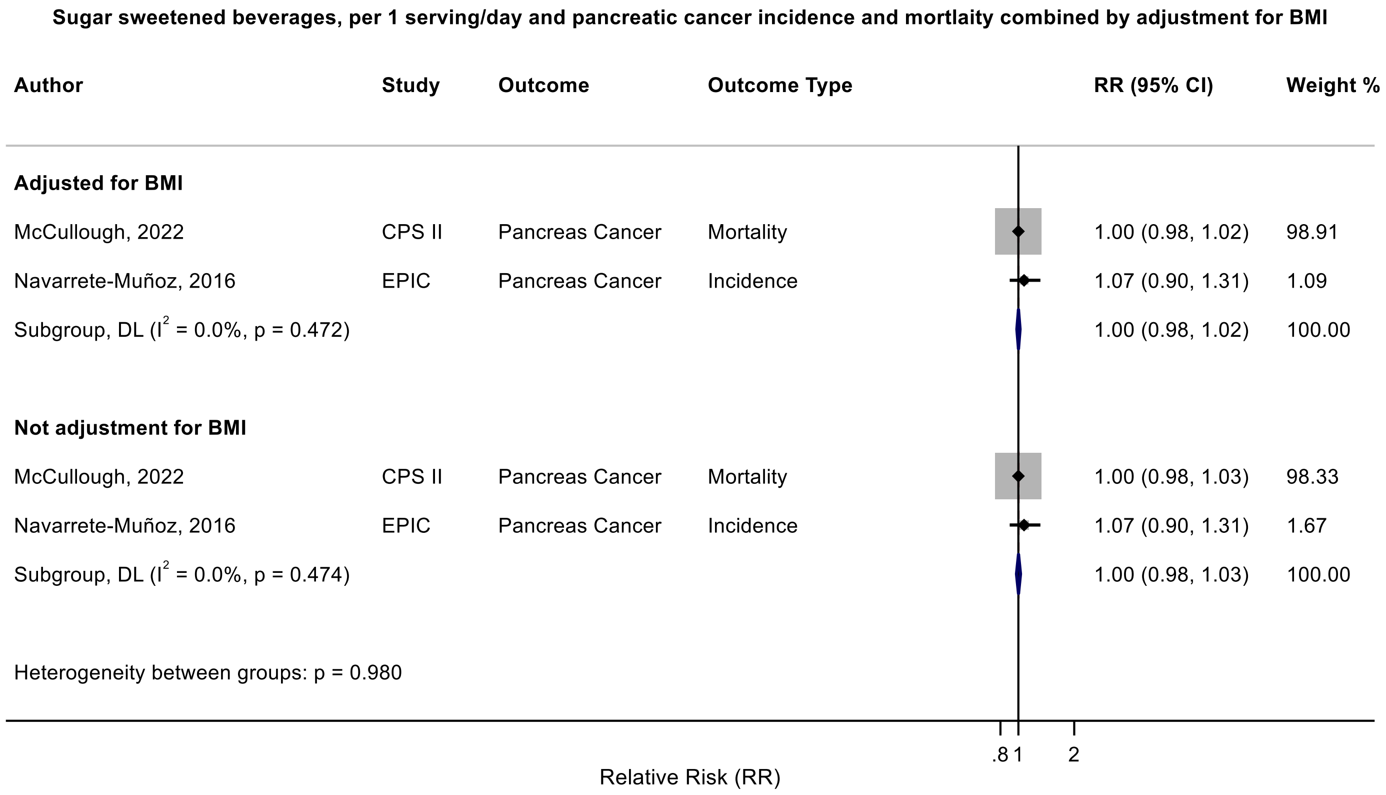

Figure legend: Forest plot shows the linear dose-response results (per 1 serving [355 mL] of sugar sweetened beverages /day) from the inverse variance DerSimonian-Laird random-effects model. The diamond represents the summary relative risk (RR) estimate, the diamond's width is the 95% confidence interval (CI). Each square and the horizontal line across the square represent the RR estimate of the individual study and its 95% CI.

Abbreviations: BMI, body mass index; CI, confidence interval; RR, relative risk.

Study abbreviations: EPIC, European Prospective Investigation into Cancer and nutrition; CPS II, Cancer Prevention Study II

Note: This subgroup analysis is based on within study comparisons in two studies that reported association estimates with and without adjustment for BMI. Since only two studies, one on incidence and one on mortality, reported risk estimates for subgroup analysis by adjustment for BMI, we combined incidence and mortality to be able to perform this important subgroup analysis.

Navarrete-Muñoz, 2016[127], McCullough, 2022[113]

**Supplementary Figure 138. Linear dose-response meta-analysis of sugar sweetened beverages and pancreatic cancer incidence by exclusion of people with diabetes at baseline of the study.**

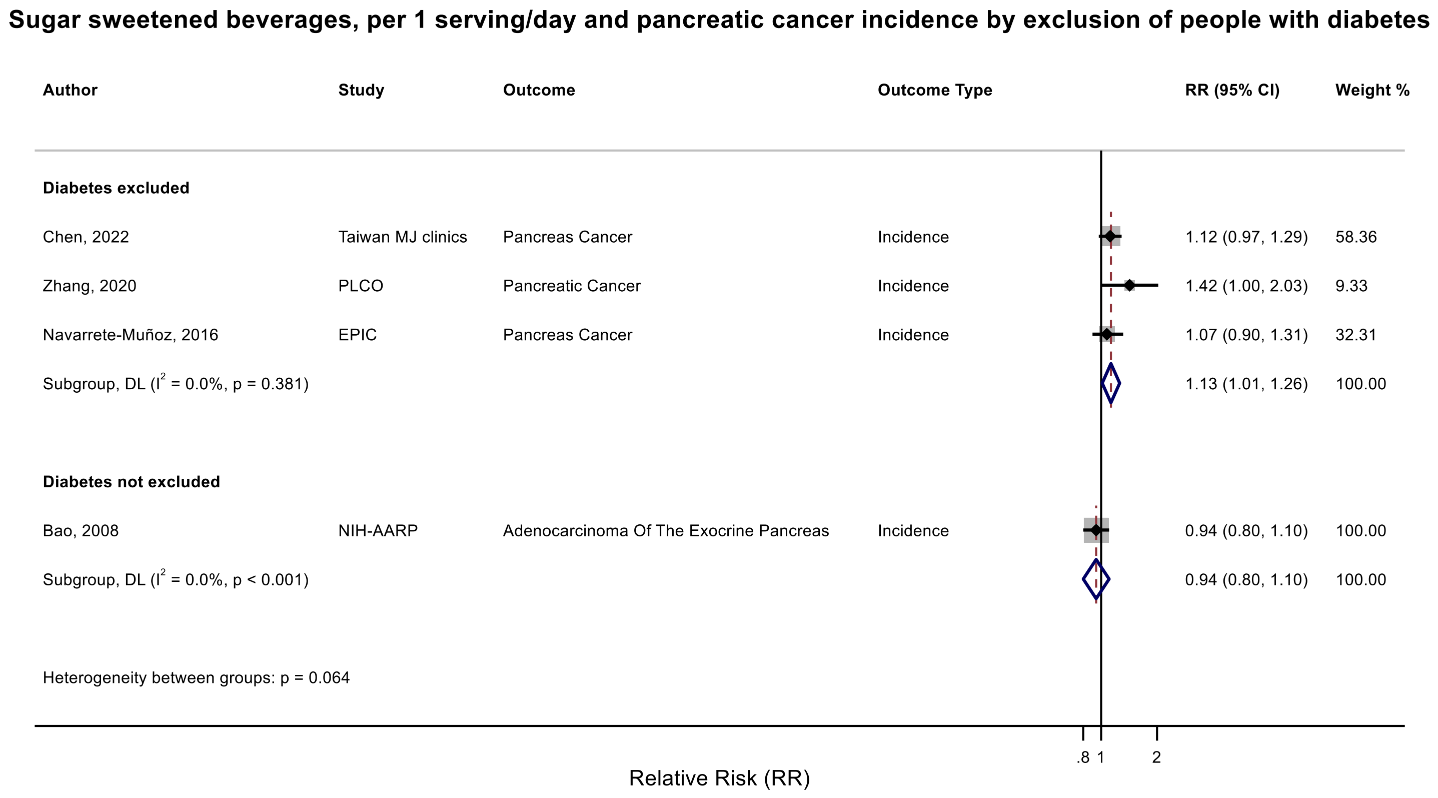

Figure legend: Forest plot shows the linear dose-response results (per 1 serving [355 mL] of sugar sweetened beverages /day) from the inverse variance DerSimonian-Laird random-effects model. The diamond represents the summary relative risk (RR) estimate, the diamond's width is the 95% confidence interval (CI). Each square and the horizontal line across the square represent the RR estimate of the individual study and its 95% CI.

Abbreviations: CI, confidence interval; RR, relative risk.

Study abbreviations: NIH-AARP, National Institute of Health + American Association of Retired Persons Diet and Health Study; CPS II, Cancer Prevention Study II; EPIC, European Prospective Investigation into Cancer and nutrition; PLCO, Prostate, Lung, Colorectal and Ovarian Cancer Screening trial; Taiwan MJ clinics, The Taiwan MJ Cohort Study

Note: The PLCO and Taiwan MJ clinics studies controlled for a history of diabetes in their multivariable analyses. The EPIC study did not exclude people with a history of diabetes at baseline but performed a sensitivity analysis after exclusion of people with diabetes and reported that the result was the same (data not shown).

Bao, 2008[39], Navarrete-Muñoz, 2016[127], Zhang, 2020[184], Chen, 2022[45]

**Supplementary Figure 139. Linear dose-response meta-analysis of carbonated sugar sweetened beverages and pancreatic cancer incidence, by various factors.**

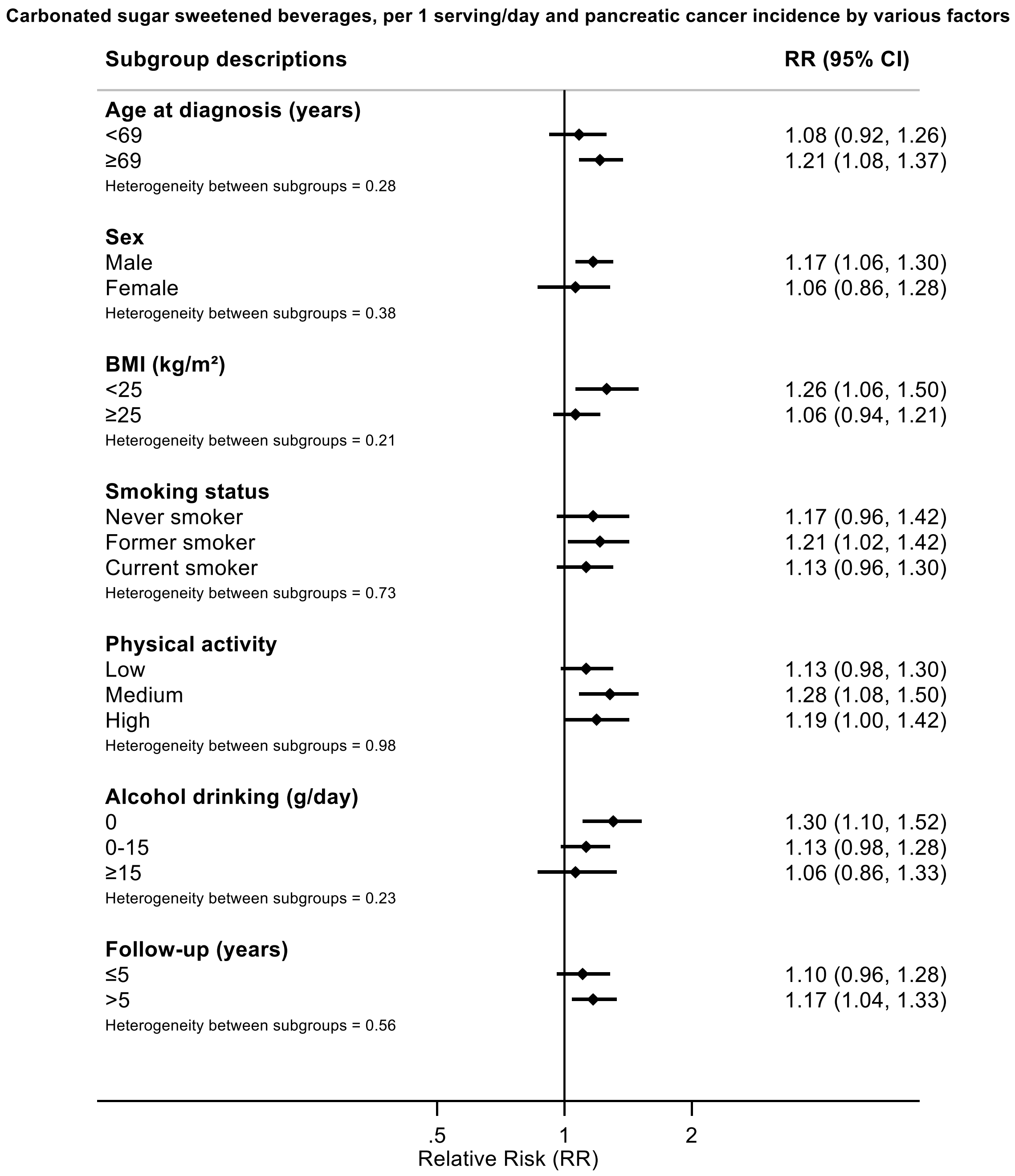

Figure legend: Forest plot shows the linear dose-response results (per 1 serving [355 mL] of carbonated sugar sweetened beverages /day) from the inverse variance DerSimonian-Laird random-effects model. The diamond represents the summary relative risk (RR) estimate, the diamond's width is the 95% confidence interval (CI).

Abbreviations: CI, confidence interval; RR, relative risk.

Note: The results have been extracted from a published pooled analysis.

Genkinger, 2012[64]

**Supplementary Figure 140. Linear dose-response meta-analysis of sugar sweetened beverages and breast cancer incidence by geographical region.**

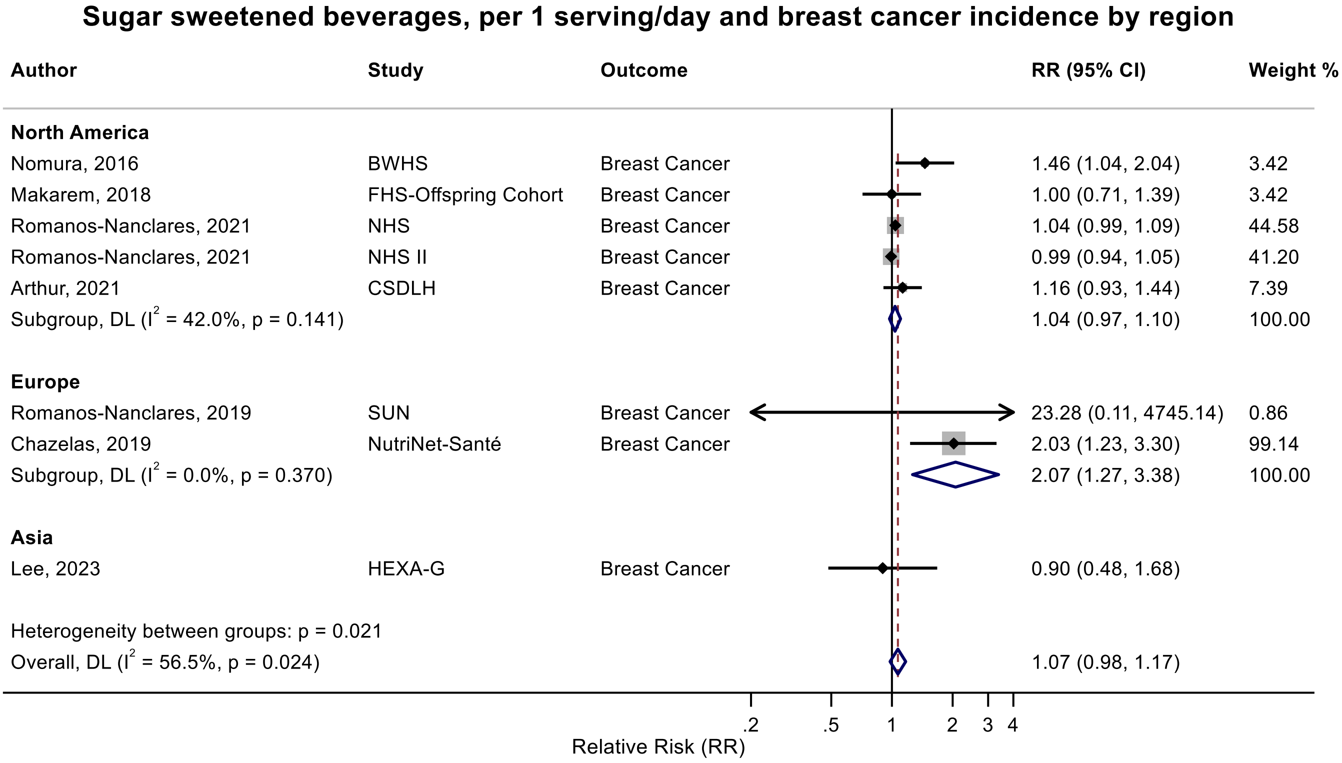

Figure legend: Forest plot shows the linear dose-response results (per 1 serving [355 mL] of sugar sweetened beverages /day) from the inverse variance DerSimonian-Laird random-effects model. The diamond represents the summary relative risk (RR) estimate, the diamond's width is the 95% confidence interval (CI). Each square and the horizontal line across the square represent the RR estimate of the individual study and its 95% CI.

Abbreviations: CI, confidence interval; RR, relative risk.

Study abbreviations: BWHS, Black Women's Health Study; FHS-Offspring Cohort, Framingham Heart Study-Offspring Cohort; NHS, Nurses' Health Study; NHS II, Nurses' Health Study II; CSDLH, Canadian Study of Diet, Lifestyle and Health; SUN, Seguimiento Universidad de Navarra Cohort Study; HEXA-G, Health Examinees-Gem study

Nomura, 2016[128], Makarem, 2018[108], Romanos-Nanclares, 2021[145], Arthur, 2021[38], Romanos-Nanclares, 2019[146], Chazelas, 2019[44], Lee, 2023[97]

**Supplementary Figure 141. Linear dose-response meta-analysis of sugar sweetened beverages and breast cancer incidence by follow-up duration.**

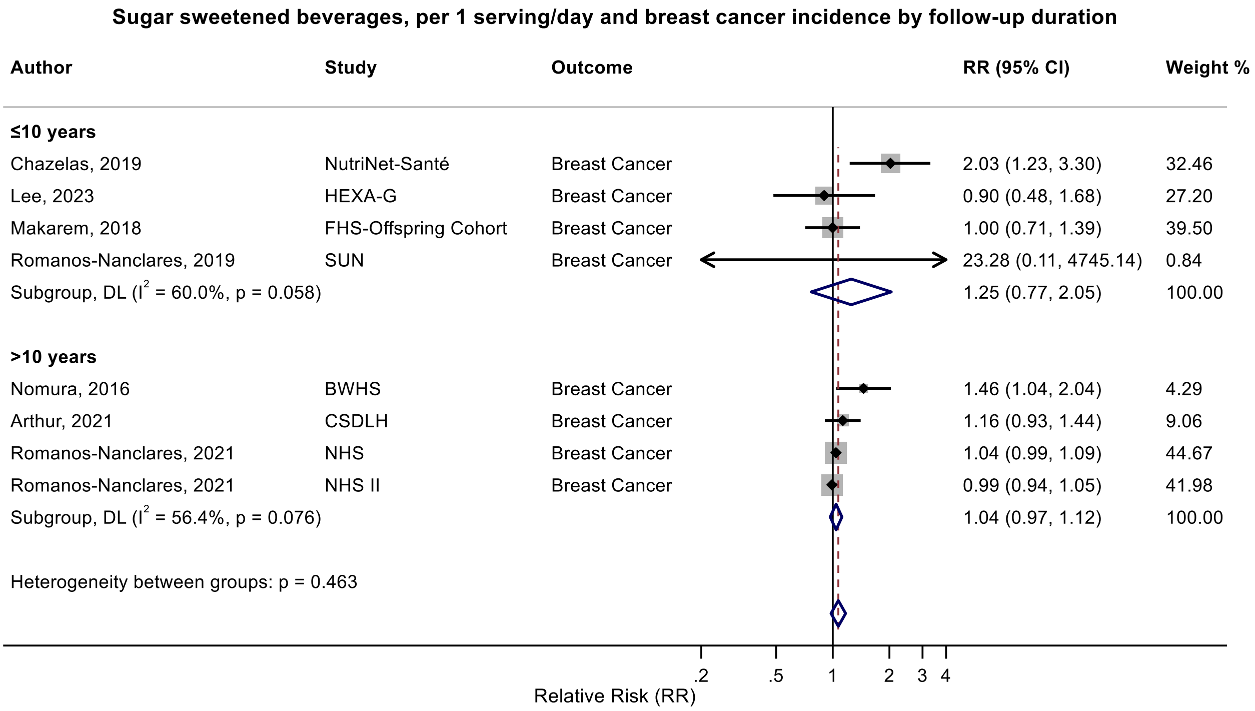

Figure legend: Forest plot shows the linear dose-response results (per 1 serving [355 mL] of sugar sweetened beverages /day) from the inverse variance DerSimonian-Laird random-effects model. The diamond represents the summary relative risk (RR) estimate, the diamond's width is the 95% confidence interval (CI). Each square and the horizontal line across the square represent the RR estimate of the individual study and its 95% CI.

Abbreviations: CI, confidence interval; RR, relative risk.

Study abbreviations: HEXA-G, Health Examinees-Gem study; FHS-Offspring Cohort, Framingham Heart Study-Offspring Cohort; SUN, Seguimiento Universidad de Navarra Cohort Study; BWHS, Black Women's Health Study; CSDLH, Canadian Study of Diet, Lifestyle and Health; NHS, Nurses' Health Study; NHS II, Nurses' Health Study II

Chazelas, 2019[44], Lee, 2023[97], Makarem, 2018[108], Romanos-Nanclares, 2019[146], Nomura, 2016[128], Arthur, 2021[38], Romanos-Nanclares, 2021[145]

**Supplementary Figure 142. Linear dose-response meta-analysis of sugar sweetened beverages and breast cancer incidence by inclusion of fruit juice.**

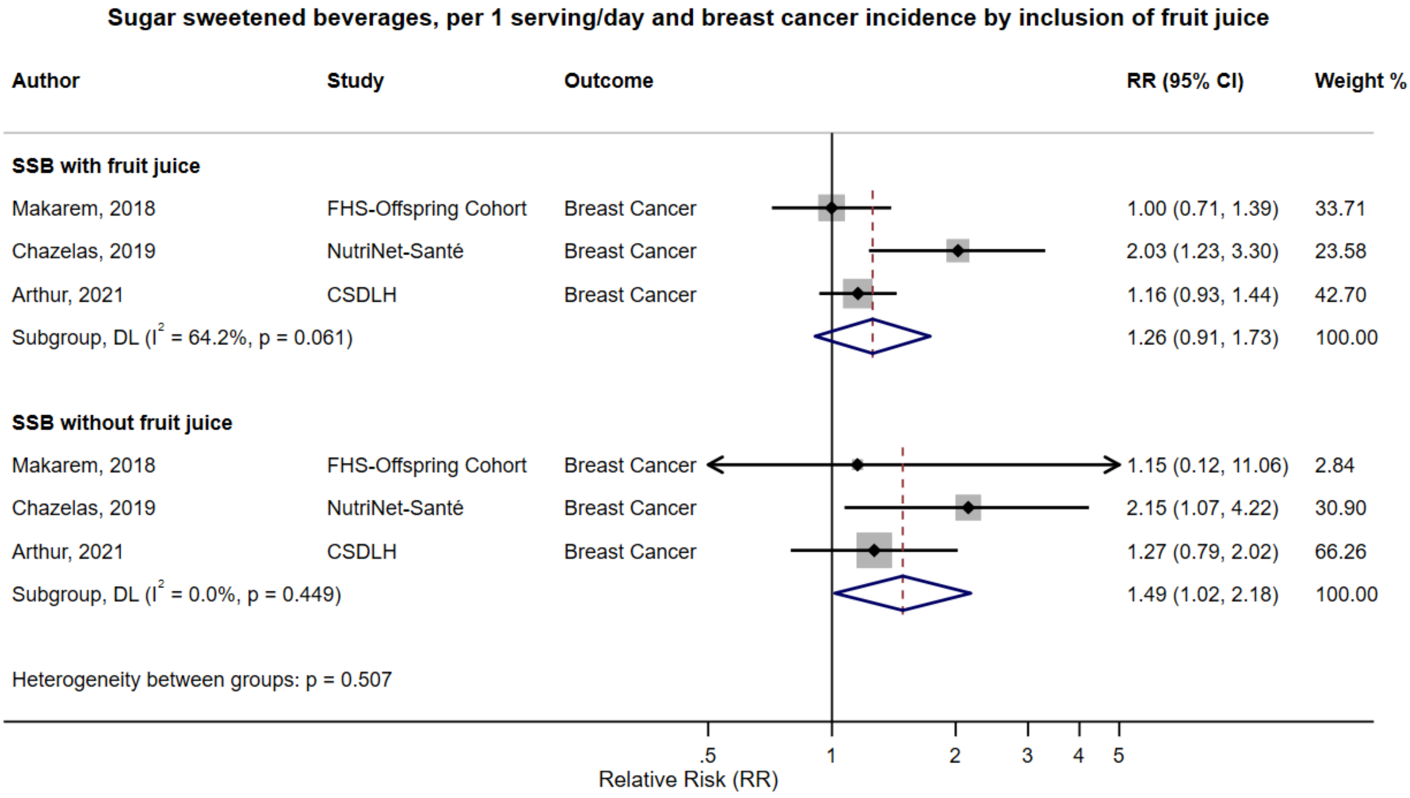

Figure legend: Forest plot shows the linear dose-response results (per 1 serving [355 mL] of sugar sweetened beverages /day) from the inverse variance DerSimonian-Laird random-effects model. The diamond represents the summary relative risk (RR) estimate, the diamond's width is the 95% confidence interval (CI). Each square and the horizontal line across the square represent the RR estimate of the individual study and its 95% CI.

Abbreviations: CI, confidence interval; RR, relative risk, SSB, sugar sweetened beverages.

Study abbreviations: FHS-Offspring Cohort, Framingham Heart Study-Offspring Cohort; CSDLH, Canadian Study of Diet, Lifestyle and Health

Note: This subgroup analysis is based on within study comparisons in three studies reporting results for sugar sweetened beverages with and without fruit juices.

Makarem, 2018[108], Chazelas, 2019[44], Arthur, 2021[38]

#### Supplementary Figure 143. Linear dose-response meta-analysis of sugar sweetened beverages and breast cancer incidence by frequency of dietary assessment.

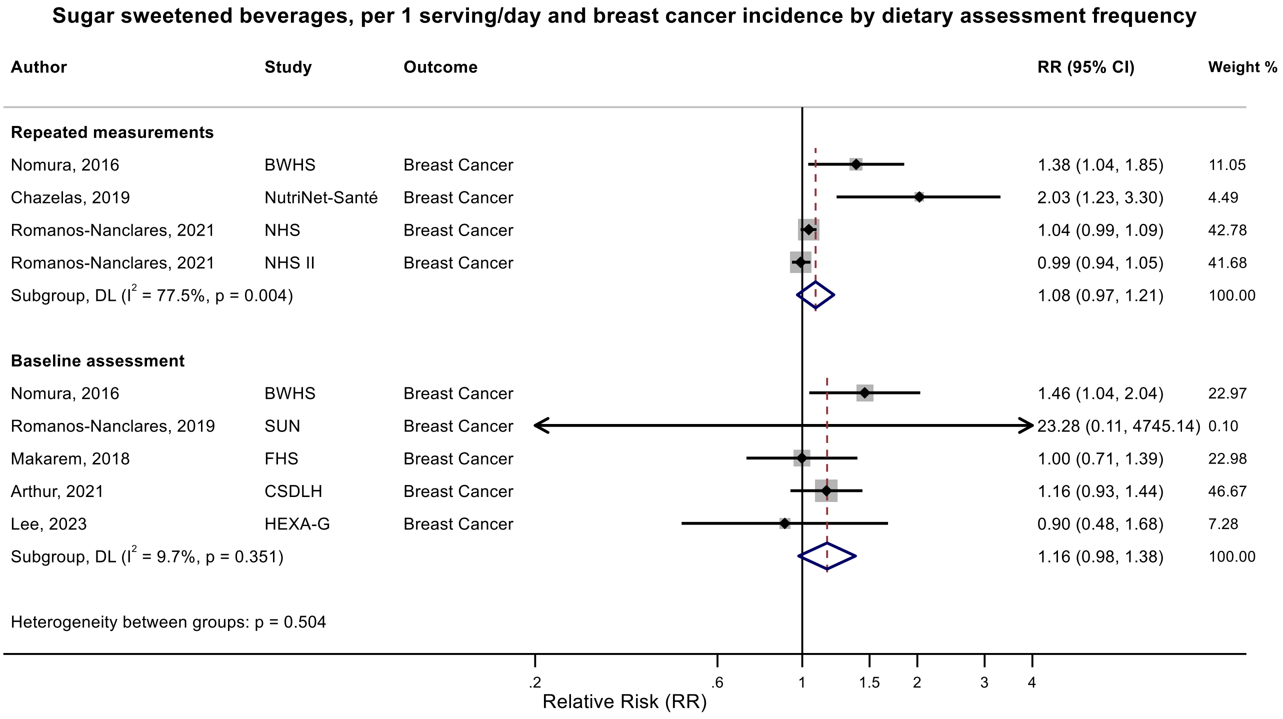

Figure legend: Forest plot shows the linear dose-response results (per 1 serving [355 mL] of sugar sweetened beverages /day) from the inverse variance DerSimonian-Laird random-effects model. The diamond represents the summary relative risk (RR) estimate, the diamond's width is the 95% confidence interval (CI). Each square and the horizontal line across the square represent the RR estimate of the individual study and its 95% CI.

Abbreviations: CI, confidence interval; RR, relative risk.

Study abbreviations: BWHS, Black Women's Health Study; NHS, Nurses' Health Study; NHS II, Nurses' Health Study II; SUN, Seguimiento Universidad de Navarra Cohort Study; FHS, Framingham Heart Study; CSDLH, Canadian Study of Diet, Lifestyle and Health; HEXA-G, Health Examinees-Gem study

Nomura, 2016[128], Chazelas, 2019[44], Romanos-Nanclares, 2021[145], Romanos-Nanclares, 2019[146], Makarem, 2018[108], Arthur, 2021[38], Lee, 2023[97]

**Supplementary Figure 144. Linear dose-response meta-analysis of sugar sweetened beverages and postmenopausal breast cancer incidence by frequency of dietary assessment.**

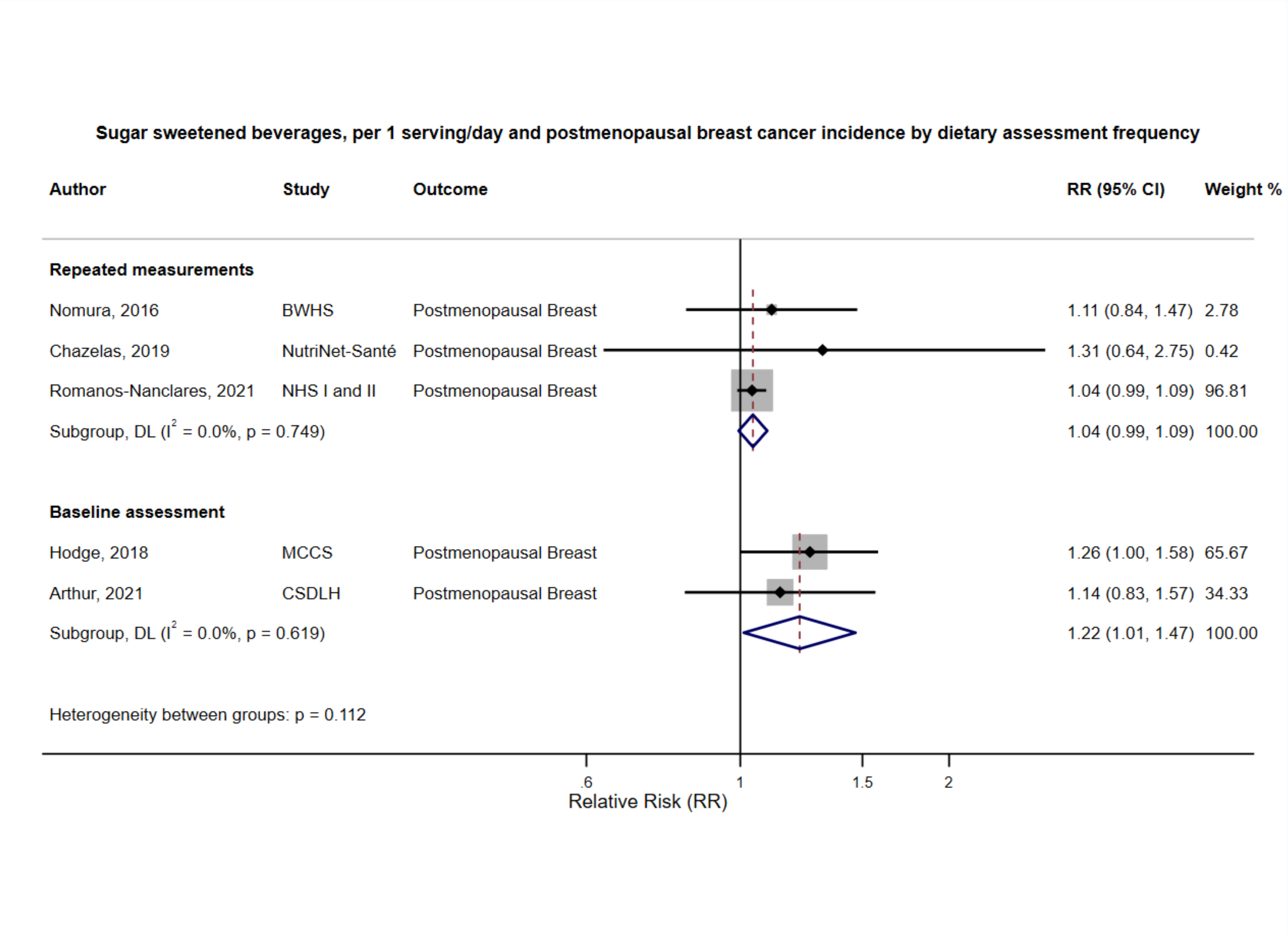

Figure legend: Forest plot shows the linear dose-response results (per 1 serving [355 mL] of sugar sweetened beverages /day) from the inverse variance DerSimonian-Laird random-effects model. The diamond represents the summary relative risk (RR) estimate, the diamond's width is the 95% confidence interval (CI). Each square and the horizontal line across the square represent the RR estimate of the individual study and its 95% CI.

Abbreviations: CI, confidence interval; RR, relative risk.

Study abbreviations: BWHS, Black Women's Health Study; NHS I and II, Nurses' Health Study I and II; MCCS, Melbourne Collaborative Cohort Study; CSDLH, Canadian Study of Diet, Lifestyle and Health

Nomura, 2016[128], Chazelas, 2019[44], Romanos-Nanclares, 2021[145], Hodge, 2018[70], Arthur, 2021[38]

**Supplementary Figure 145. Linear dose-response meta-analysis of sugar sweetened beverages and postmenopausal breast cancer incidence by exclusion of people with a history of diabetes at baseline of the study.**

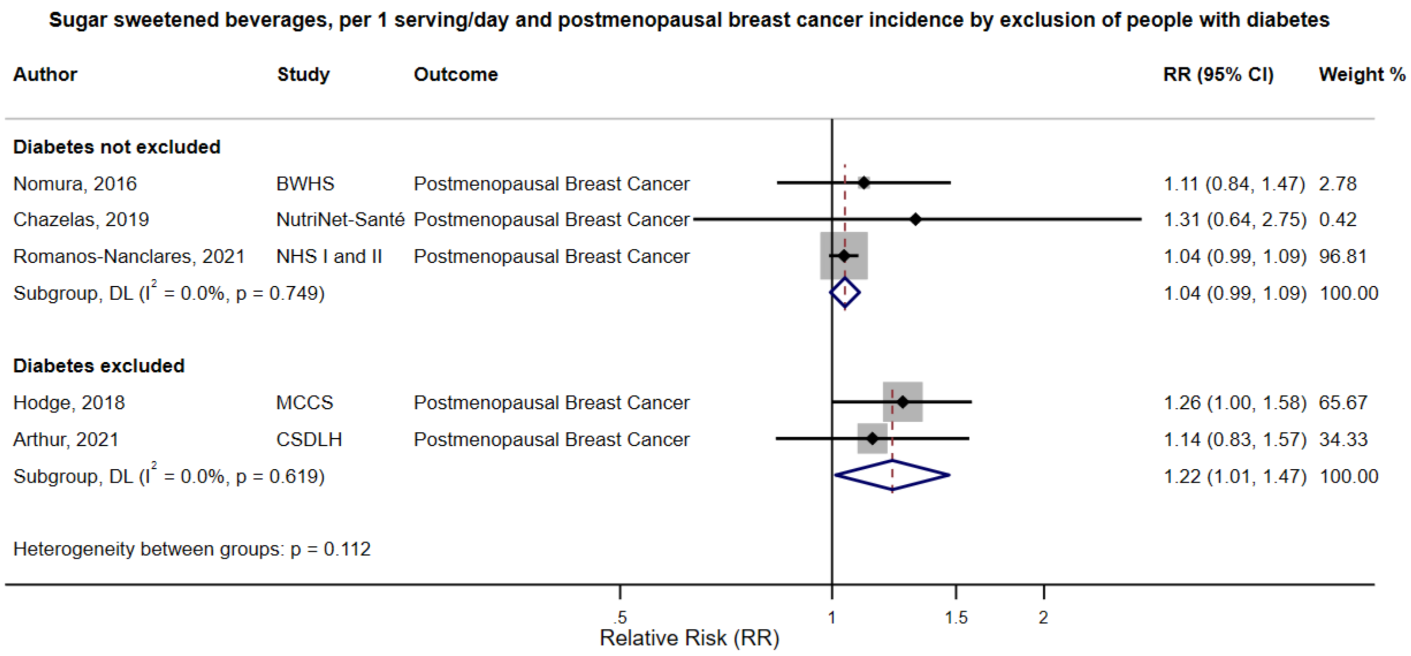

Figure legend: Forest plot shows the linear dose-response results (per 1 serving [355 mL] of sugar sweetened beverages /day) from the inverse variance DerSimonian-Laird random-effects model. The diamond represents the summary relative risk (RR) estimate, the diamond's width is the 95% confidence interval (CI). Each square and the horizontal line across the square represent the RR estimate of the individual study and its 95% CI.

Abbreviations: CI, confidence interval; RR, relative risk.

Study abbreviations: BWHS, Black Women's Health Study; NHS I and II, Nurses' Health Study I and II; MCCS, Melbourne Collaborative Cohort Study; CSDLH, Canadian Study of Diet, Lifestyle and Health

Note: The NutriNet-Sante study controlled for a history of type 2 diabetes.

Nomura, 2016[128], Chazelas, 2019[44], Romanos-Nanclares, 2021[145], Hodge, 2018[70], Arthur, 2021[38]

**Supplementary Figure 146. Linear dose-response meta-analysis of sugar sweetened beverages and postmenopausal breast cancer incidence by bias related to confounding.**

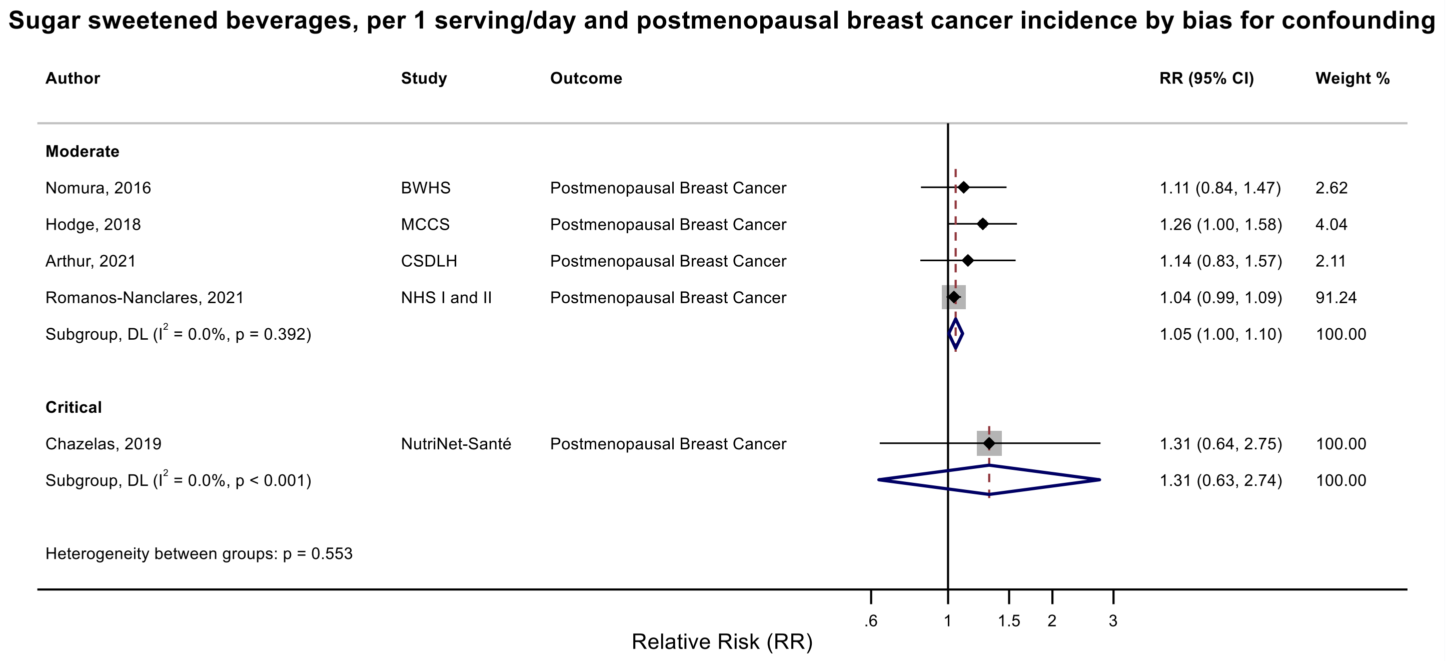

Figure legend: Forest plot shows the linear dose-response results (per 1 serving [355 mL] of sugar sweetened beverages /day) from the inverse variance DerSimonian-Laird random-effects model. The diamond represents the summary relative risk (RR) estimate, the diamond's width is the 95% confidence interval (CI). Each square and the horizontal line across the square represent the RR estimate of the individual study and its 95% CI.

Abbreviations: CI, confidence interval; RR, relative risk.

Study abbreviations: BWHS, Black Women's Health Study; NHS I and II, Nurses' Health Study I and II; MCCS, Melbourne Collaborative Cohort Study; CSDLH, Canadian Study of Diet, Lifestyle and Health

Nomura, 2016[128], Chazelas, 2019[44], Romanos-Nanclares, 2021[145], Hodge, 2018[70], Arthur, 2021[38]

**Supplementary Figure 147. Linear dose-response meta-analysis of sugar sweetened beverages and postmenopausal breast cancer incidence by bias related to departure from intended exposure.**

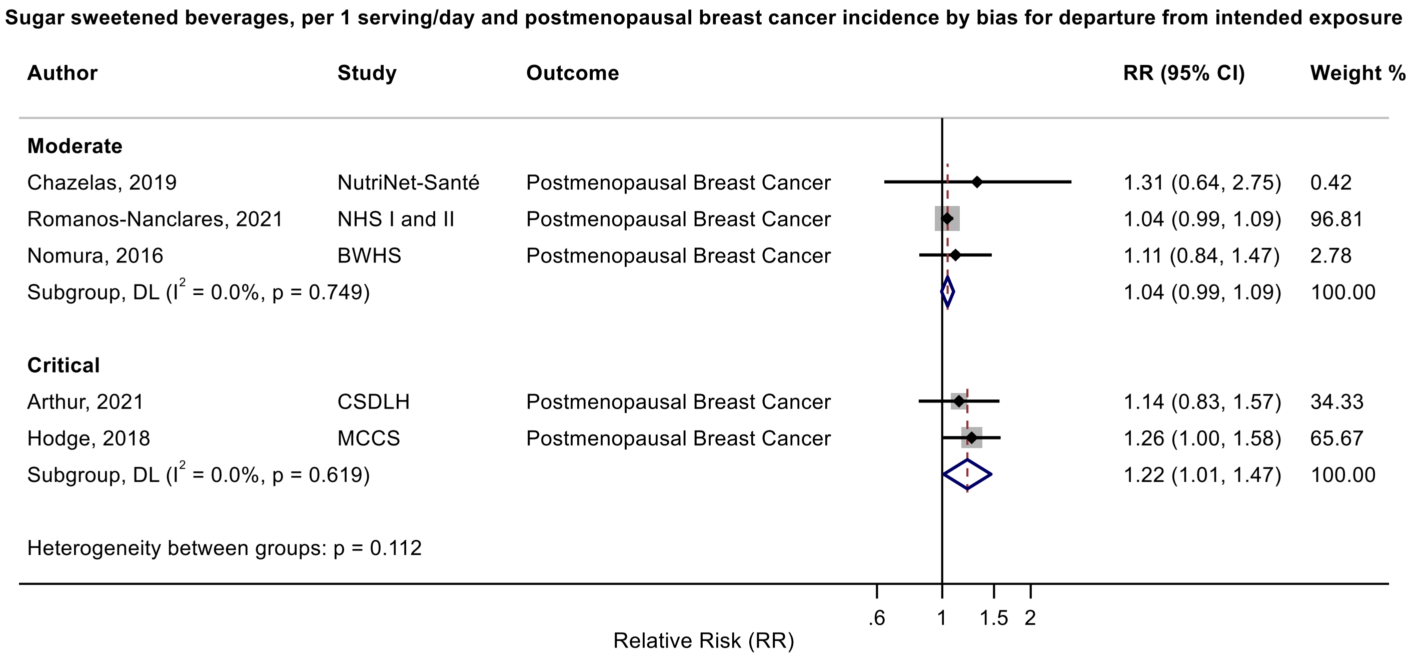

Figure legend: Forest plot shows the linear dose-response results (per 1 serving [355 mL] of sugar sweetened beverages /day) from the inverse variance DerSimonian-Laird random-effects model. The diamond represents the summary relative risk (RR) estimate, the diamond's width is the 95% confidence interval (CI). Each square and the horizontal line across the square represent the RR estimate of the individual study and its 95% CI.

Abbreviations: CI, confidence interval; RR, relative risk.

Study abbreviations: BWHS, Black Women's Health Study; NHS I and II, Nurses' Health Study I and II; MCCS, Melbourne Collaborative Cohort Study; CSDLH, Canadian Study of Diet, Lifestyle and Health

Nomura, 2016[128], Chazelas, 2019[44], Romanos-Nanclares, 2021[145], Hodge, 2018[70], Arthur, 2021[38]

**Supplementary Figure 148. Linear dose-response meta-analysis of sugar sweetened beverages and premenopausal breast cancer incidence by exclusion of people with a history of diabetes at baseline of the study.**

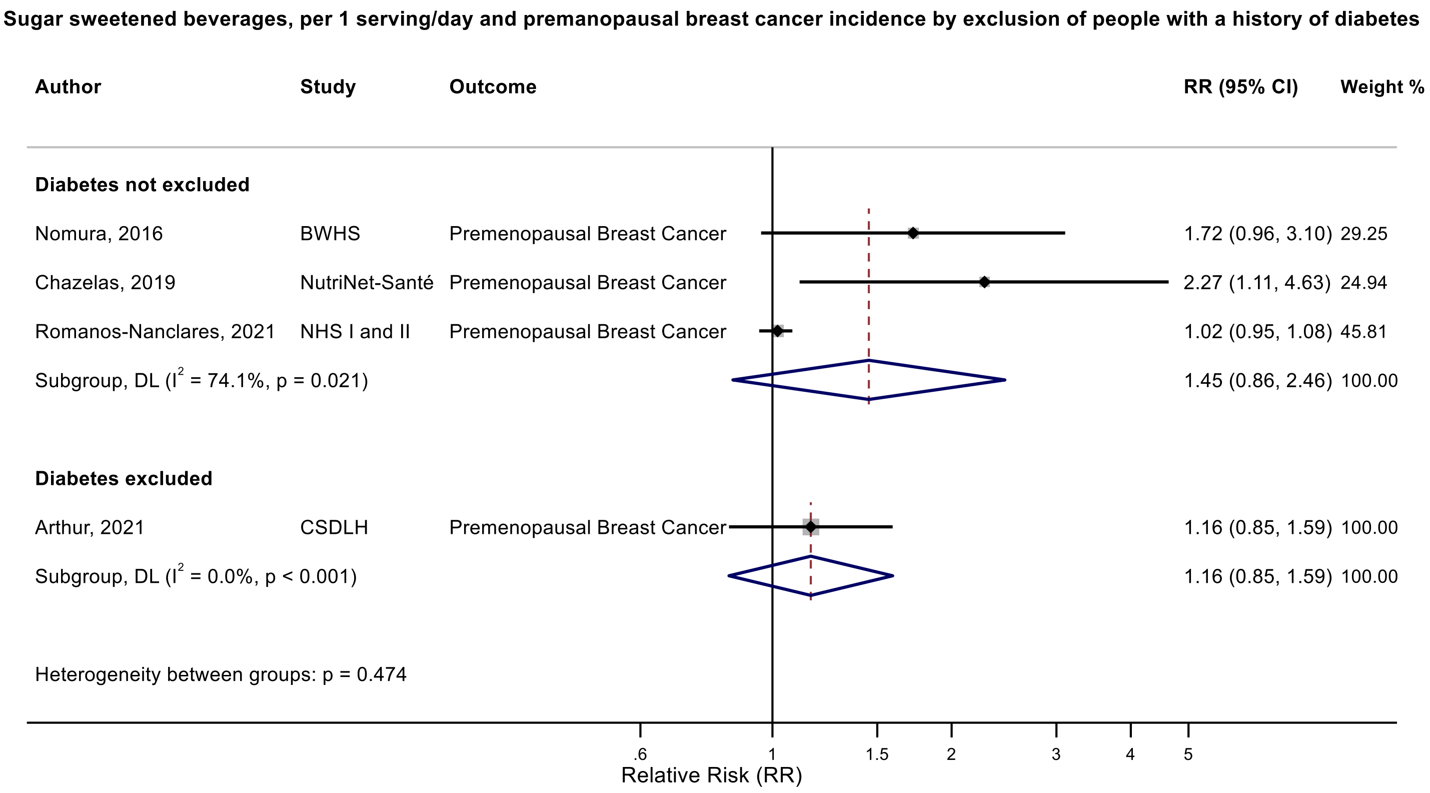

Figure legend: Forest plot shows the linear dose-response results (per 1 serving [355 mL] of sugar sweetened beverages /day) from the inverse variance DerSimonian-Laird random-effects model. The diamond represents the summary relative risk (RR) estimate, the diamond's width is the 95% confidence interval (CI), and the diamond's horizontal line is the 95% prediction interval (PI). Each square and the horizontal line across the square represent the RR estimate of the individual study and its 95% CI.

Abbreviations: CI, confidence interval; RR, relative risk.

Study abbreviations: BWHS, Black Women's Health Study; NHS I and II, Nurses' Health Study I and II; CSDLH, Canadian Study of Diet, Lifestyle and Health

Nomura, 2016[128], Chazelas, 2019[44], Romanos-Nanclares, 2021[145], Arthur, 2021[38]

**Supplementary Figure 149. Linear dose-response meta-analysis of sweetened sugar beverages and premenopausal breast cancer incidence by frequency of dietary assessment.**

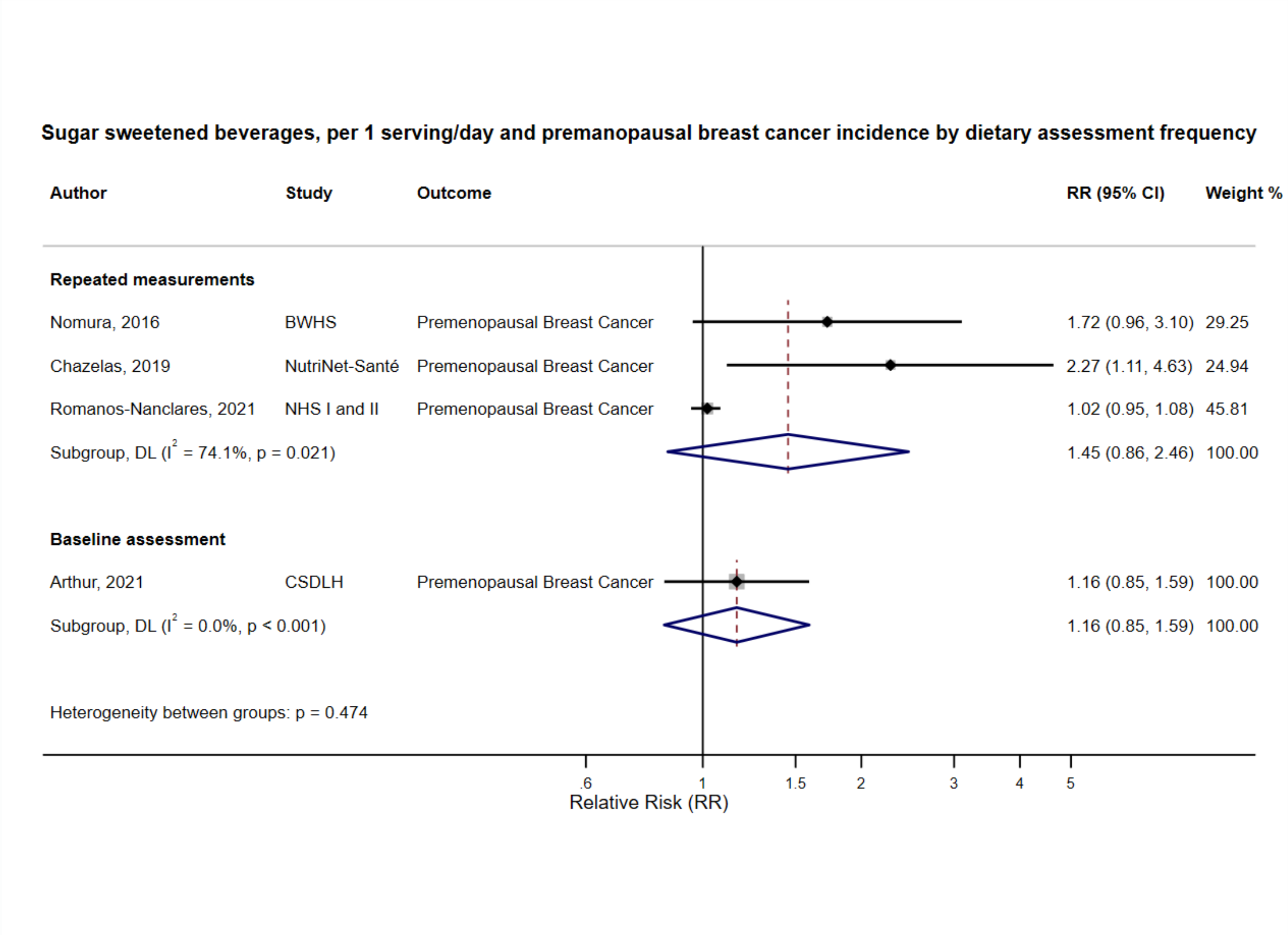

Figure legend: Forest plot shows the linear dose-response results (per 1 serving [355 mL] of sugar sweetened beverages /day) from the inverse variance DerSimonian-Laird random-effects model. The diamond represents the summary relative risk (RR) estimate, the diamond's width is the 95% confidence interval (CI), and the diamond's horizontal line is the 95% prediction interval (PI). Each square and the horizontal line across the square represent the RR estimate of the individual study and its 95% CI.

Abbreviations: CI, confidence interval; RR, relative risk.

Study abbreviations: BWHS, Black Women's Health Study; NHS I and II, Nurses' Health Study I and II; CSDLH, Canadian Study of Diet, Lifestyle and Health

Nomura, 2016[128], Chazelas, 2019[44], Romanos-Nanclares, 2021[145], Arthur, 2021[38]

#### Supplementary Figure 150. Linear dose-response meta-analysis of sugar sweetened beverages and prostate cancer incidence by frequency of dietary assessment.

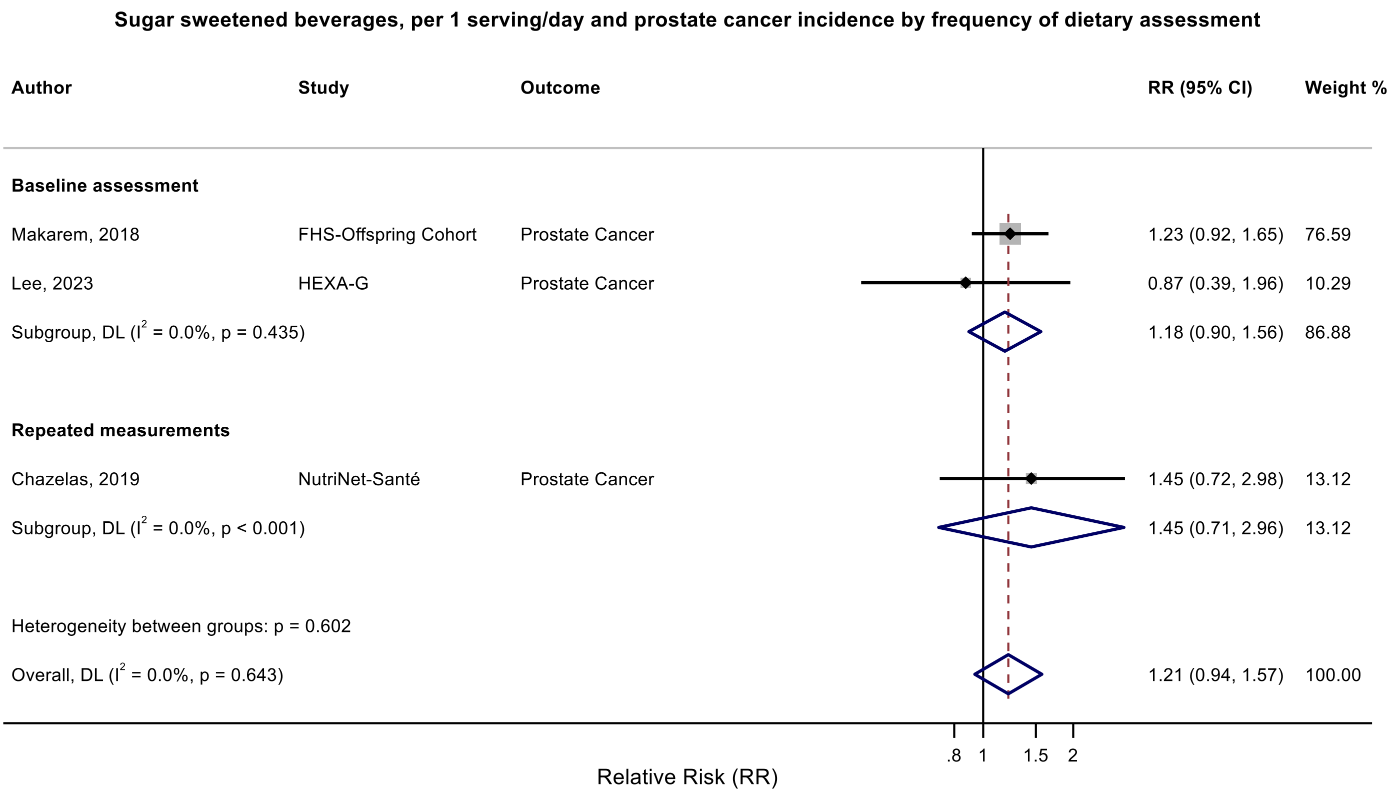

Figure legend: Forest plot shows the linear dose-response results (per 1 serving [355 mL] of sugar sweetened beverages /day) from the inverse variance DerSimonian-Laird random-effects model. The diamond represents the summary relative risk (RR) estimate, the diamond's width is the 95% confidence interval (CI). Each square and the horizontal line across the square represent the RR estimate of the individual study and its 95% CI.

Abbreviations: CI, confidence interval; RR, relative risk.

Study abbreviations: FHS-Offspring Cohort, Framingham Heart Study-Offspring Cohort; HEXA-G, Health Examinees-Gem study

Makarem, 2018[108], Lee, 2023[97], Chazelas, 2019[44]

**Supplementary Figure 151. Linear dose-response meta-analysis of sugar sweetened beverages and endometrial cancer incidence by adjustment for body mass index.**

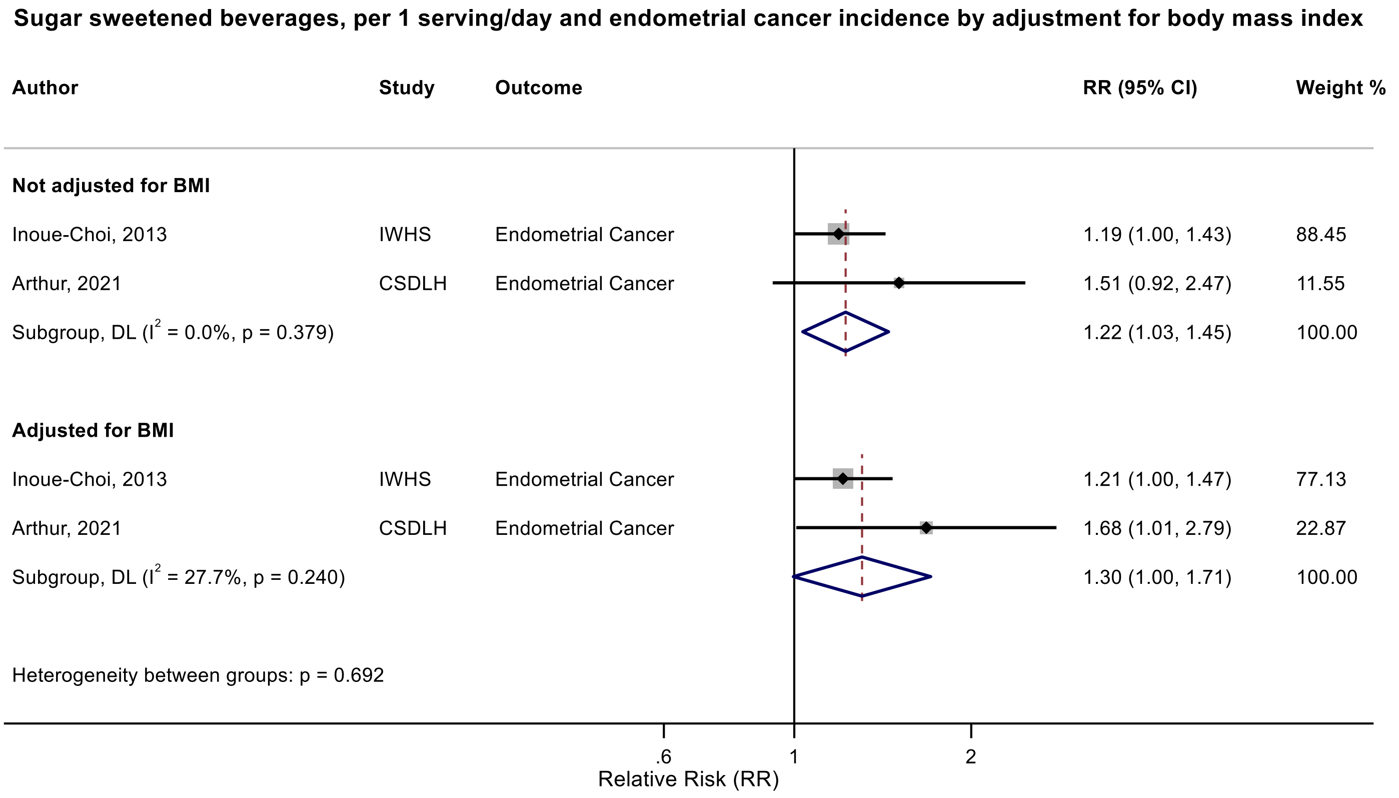

Figure legend: Forest plot shows the linear dose-response results (per 1 serving [355 mL] of sugar sweetened beverages /day) from the inverse variance DerSimonian-Laird random-effects model. The diamond represents the summary relative risk (RR) estimate, the diamond's width is the 95% confidence interval (CI). Each square and the horizontal line across the square represent the RR estimate of the individual study and its 95% CI.

Abbreviations: CI, confidence interval; RR, relative risk.

Study abbreviations: IWHS, The Iowa Women's Health Study; CSDLH, Canadian Study of Diet, Lifestyle and Health

Note: This subgroup analysis is based on a within study comparison in two studies that reported association estimates with and without adjustment for BMI. The point estimate and 95%CIs with 3 decimal points are 1.304 (0.996 – 1.706).

Inoue-Choi, 2013[74], Arthur, 2021[38]

**Supplementary Figure 152. Linear dose-response meta-analysis of sugar sweetened beverages and kidney cancer incidence by sex.**

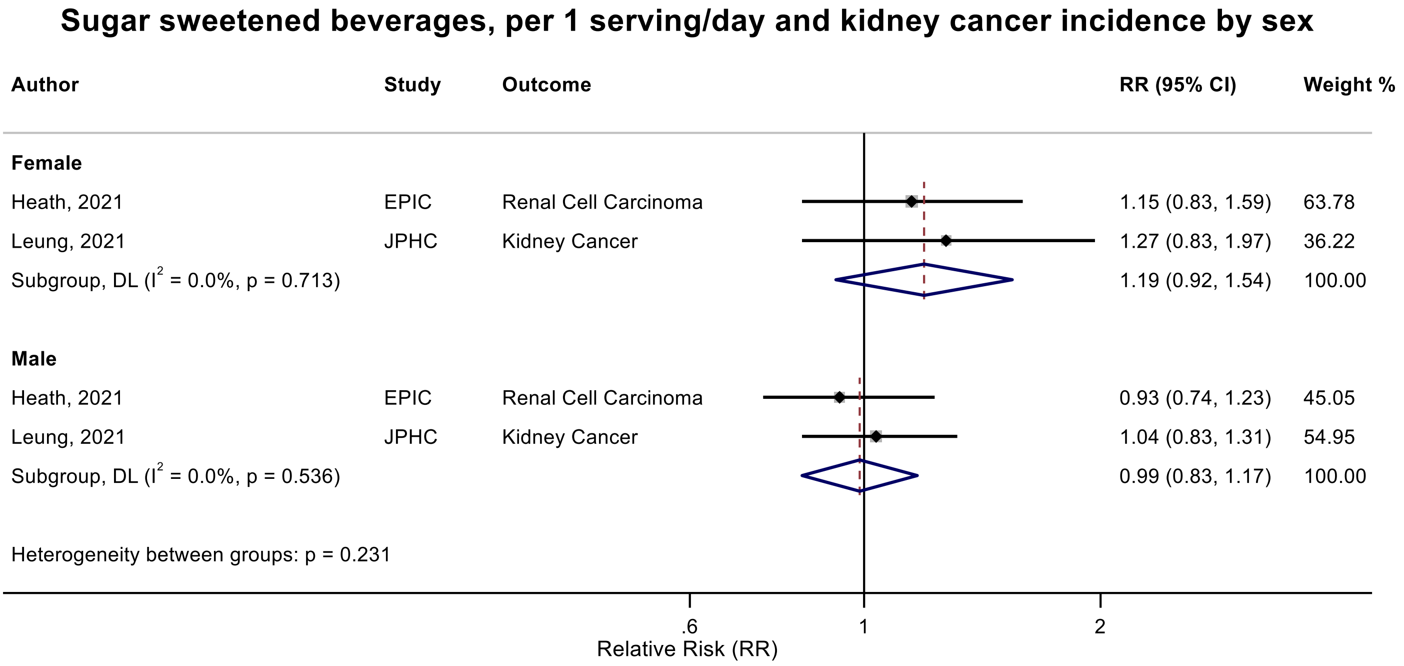

Figure legend: Forest plot shows the linear dose-response results (per 1 serving [355 mL] of sugar sweetened beverages /day) from the inverse variance DerSimonian-Laird random-effects model. The diamond represents the summary relative risk (RR) estimate, the diamond's width is the 95% confidence interval (CI). Each square and the horizontal line across the square represent the RR estimate of the individual study and its 95% CI.

Abbreviations: CI, confidence interval; RR, relative risk.

Study abbreviations: EPIC, European Prospective Investigation into Cancer and nutrition; JPHC, Japan Public Health Center-Based Prospective Study

Heath, 2021[66], Leung, 2021[100]

**Supplementary Figure 153. Linear dose-response meta-analysis of sugar sweetened beverages and kidney cancer incidence by exclusion of people with diabetes at baseline.**

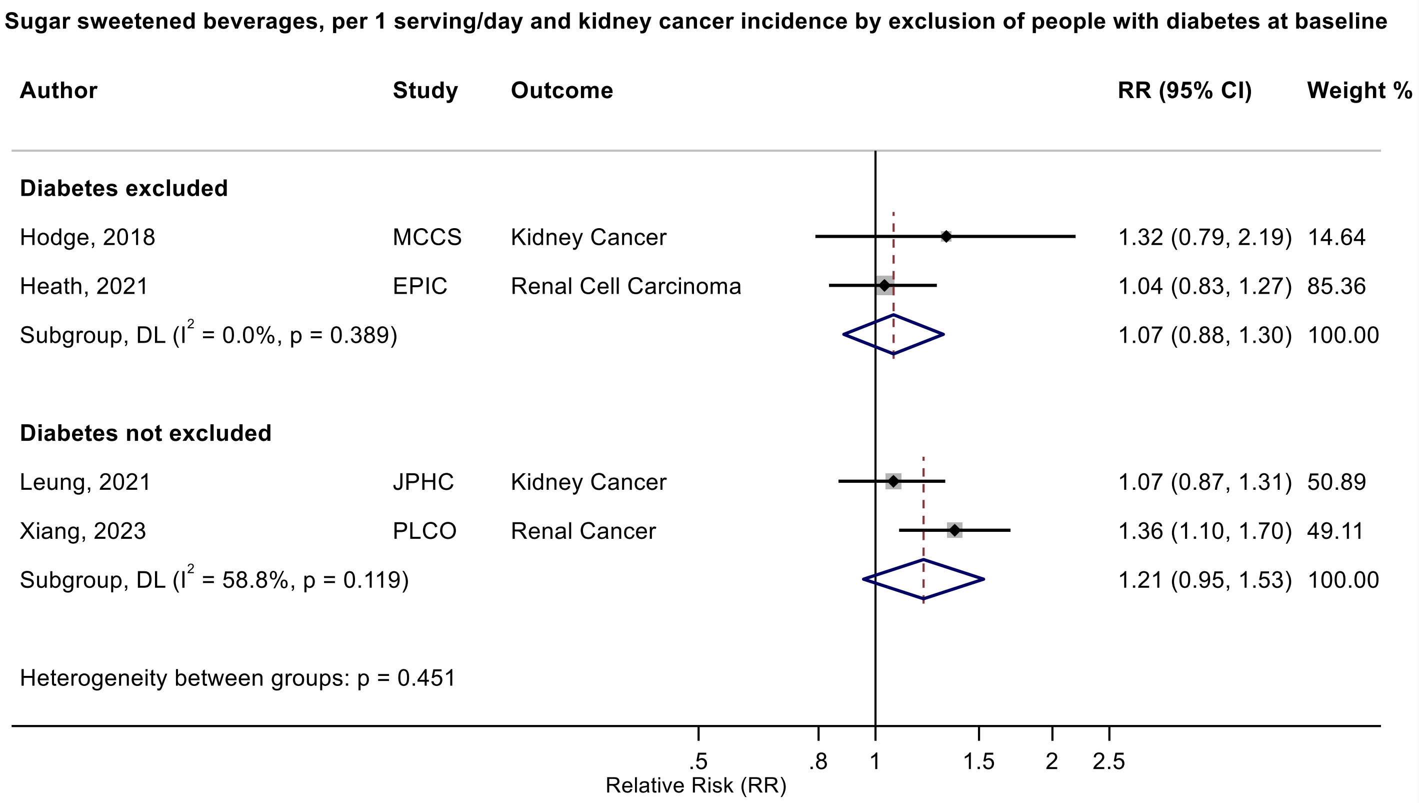

Figure legend: Forest plot shows the linear dose-response results (per 1 serving [355 mL] of sugar sweetened beverages /day) from the inverse variance DerSimonian-Laird random-effects model. The diamond represents the summary relative risk (RR) estimate, the diamond's width is the 95% confidence interval (CI). Each square and the horizontal line across the square represent the RR estimate of the individual study and its 95% CI.

Abbreviations: CI, confidence interval; RR, relative risk.

Study abbreviations: MCCS, Melbourne Collaborative Cohort Study; EPIC, European Prospective Investigation into Cancer and nutrition; JPHC, Japan Public Health Center-Based Prospective Study; PLCO, Prostate, Lung, Colorectal and Ovarian Cancer Screening trial

Hodge, 2018[70], Heath, 2021[66], Leung, 2021[100], Xiang, 2023[173]

**Supplementary Figure 154. Linear dose-response meta-analysis of sugar sweetened beverages and non-Hodgkin lymphoma incidence by frequency of dietary assessment.**

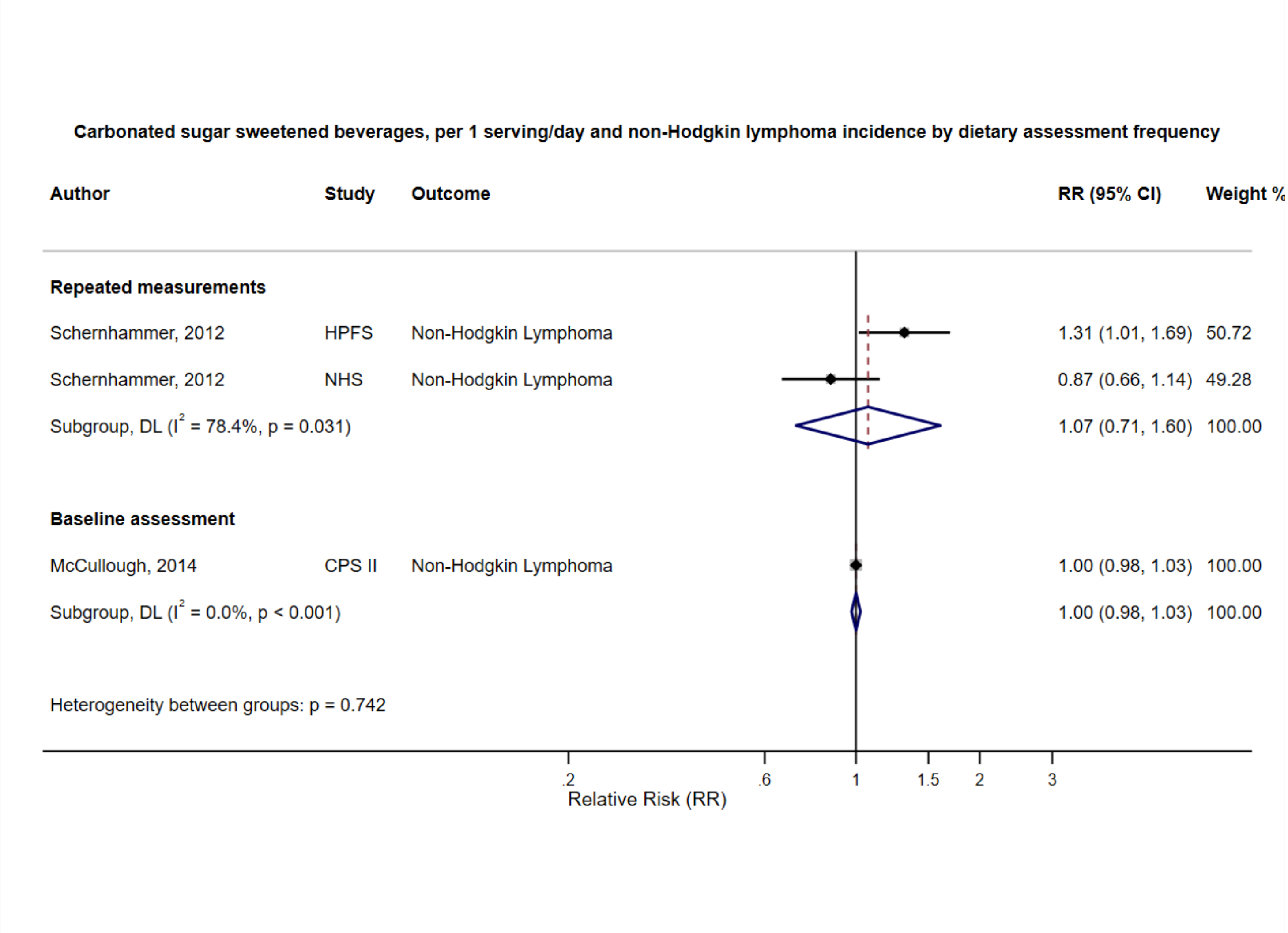

Figure legend: Forest plot shows the linear dose-response results (per 1 serving [355 mL] of carbonated sugar sweetened beverages /day) from the inverse variance DerSimonian-Laird random-effects model. The diamond represents the summary relative risk (RR) estimate, the diamond's width is the 95% confidence interval (CI). Each square and the horizontal line across the square represent the RR estimate of the individual study and its 95% CI.

Abbreviations: CI, confidence interval; RR, relative risk.

Study abbreviations: HPFS, Health Professionals Follow-up Study; NHS, Nurses' Health Study; CPS II, Cancer Prevention Study II

Schernhammer, 2012[150], McCullough, 2014[114]

**Supplementary Figure 155. Linear dose-response meta-analysis of sugar sweetened beverages and colorectal cancer incidence, leave-one-out analysis.**

**Sugar sweetened beverages, per 1 serving/day and colorectal cancer incidence, leave-one-out analysis**

| Author | Study | Outcome | RR (95% CI) |
|---|---|---|---|
| Nomura, 2016 | BWHS | Colorectal Cancer | 1.07 (1.01, 1.14) |
| Makarem, 2018 | FHS-Offspring Cohort | Colorectal Cancer | 1.07 (1.00, 1.14) |
| Hodge, 2018 | MCCS | Colorectal Cancer | 1.06 (0.99, 1.13) |
| Pacheco, 2019 | CTS | Colorectal Cancer | 1.07 (0.99, 1.14) |
| Chazelas, 2019 | NutriNet-Santé | Colorectal Cancer | 1.07 (1.00, 1.14) |
| Barrubés, 2020 | PREDIMED | Colorectal Cancer | 1.05 (0.99, 1.12) |
| Leung, 2021 | JPHC | Colorectal Cancer | 1.11 (1.02, 1.20) |
| Arthur, 2021 | CSDLH | Colorectal Cancer | 1.06 (0.99, 1.14) |
| Wang, 2022 | NHS I and II | Colorectal Cancer | 1.07 (0.99, 1.15) |
| Wang, 2022 | HPFS | Colorectal Cancer | 1.04 (0.98, 1.10) |
| Lee, 2023 | HEXA-G | Colorectal Cancer | 1.07 (1.00, 1.15) |
| Kanehara, 2024 | MEC | Colorectal Cancer | 1.11 (1.00, 1.22) |

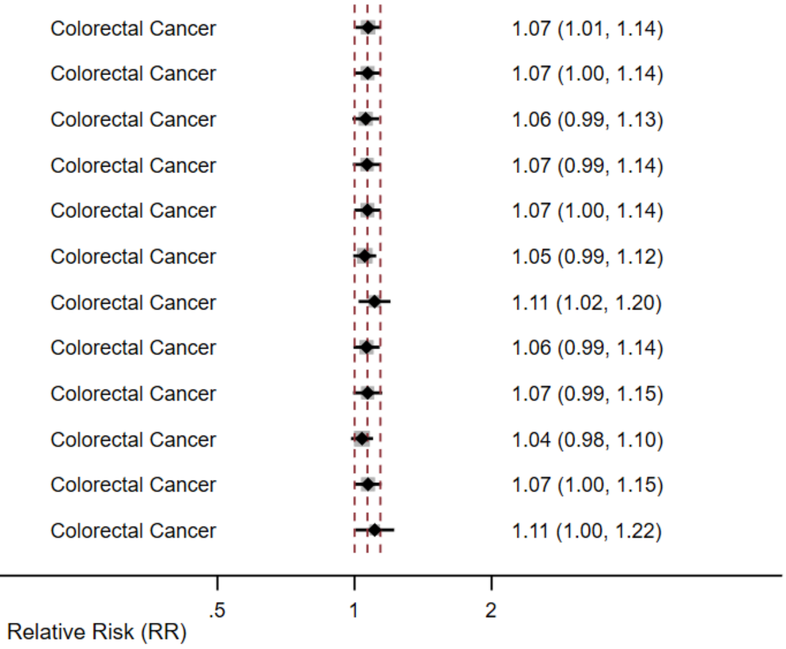

Figure legend: Forest plot shows the linear dose-response results (per 1 serving [355 mL] of sugar sweetened beverages /day) from the inverse variance DerSimonian-Laird random-effects model after exclusion of every single study at a time.

Abbreviations: CI, confidence interval; RR, relative risk.

Study abbreviations: BWHS, Black Women's Health Study; FHS-Offspring Cohort, Framingham Heart Study-Offspring Cohort; MCCS, Melbourne Collaborative Cohort Study; CTS, California Teachers Study; PREDIMED, Prevención con Dieta Mediterránea study; JPHC, Japan Public Health Center-Based Prospective Study; CSDLH, Canadian Study of Diet, Lifestyle and Health; NHS I and II, Nurses' Health Study I and II; HPFS, Health Professionals Follow-up Study; HEXA-G, Health Examinees-Gem study; MEC, Multiethnic Cohort Study

Nomura, 2016[129], Makarem, 2018[108], Hodge, 2018[70], Pacheco, 2019[135], Chazelas, 2019[44], Barrubés, 2020[40], Leung, 2021[101], Arthur, 2021[38], Wang, 2022[168], Lee, 2023[97], Kanehara, 2024[81]

**Supplementary Figure 156. Linear dose-response meta-analysis of sugar sweetened beverages and colon cancer incidence, leave-one-out analysis.**

**Sugar sweetened beverages, per 1 serving/day and colon cancer incidence, leave-one-out analysis**

| **Author** | **Study** | **Outcome** | **RR (95% CI)** |
|---|---|---|---|
| Nomura, 2016 | BWHS | Colon Cancer | 1.01 (0.96, 1.06) |
| Leung, 2021 | JPHC | Colon Cancer | 1.02 (0.98, 1.07) |
| Arthur, 2021 | CSDLH | Colon Cancer | 1.01 (0.97, 1.05) |
| Yuan, 2022 | NHS & HPFS | Colon Cancer | 1.01 (0.95, 1.07) |
| Kanehara, 2024 | MEC | Colon Cancer | 1.02 (0.94, 1.11) |

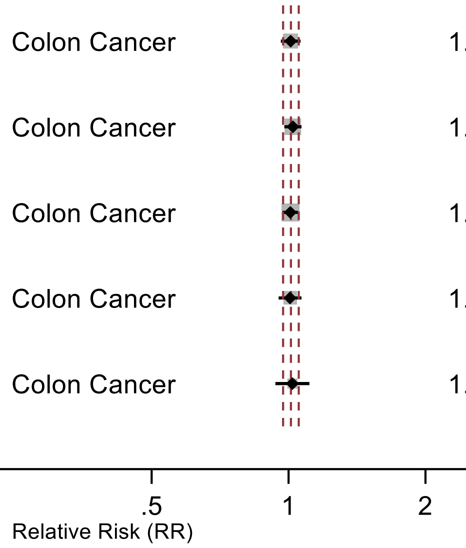

Figure legend: Forest plot shows the linear dose-response results (per 1 serving [355 mL] of sugar sweetened beverages/day) from the inverse variance DerSimonian-Laird random-effects model after exclusion of every single study at a time.

Abbreviations: CI, confidence interval; RR, relative risk.

Study abbreviations: BWHS, Black Women's Health Study; JPHC, Japan Public Health Center-Based Prospective Study; CSDLH, Canadian Study of Diet, Lifestyle and Health; NHS & HPFS, Nurses' Health Study & Health Professionals Follow-up Study; MEC, Multiethnic Cohort Study

Nomura, 2016[129], Leung, 2021[101], Arthur, 2021[38], Yuan, 2022[178], Kanehara, 2024[81]

**Supplementary Figure 157. Linear dose-response meta-analysis of carbonated sugar sweetened beverages and colon cancer incidence, leave-one-out analysis.**

**Carbonated sugar sweetened beverages, per 1 serving/day and colon cancer incidence, leave-one-out analysis**

| Author | Study | Outcome | RR (95% CI) |
|---|---|---|---|
| Zhang, 2010 | SMC | Colon Cancer | 1.00 (0.90, 1.10) |
| Zhang, 2010 | BCDDP | Colon Cancer | 1.02 (0.93, 1.12) |
| Zhang, 2010 | ATBC | Colon Cancer | 1.00 (0.90, 1.10) |
| Zhang, 2010 | NLCS | Colon Cancer | 0.99 (0.90, 1.10) |
| Zhang, 2010 | WHS | Colon Cancer | 1.00 (0.91, 1.10) |
| Zhang, 2010 | CNBSS | Colon Cancer | 0.99 (0.90, 1.09) |
| Zhang, 2010 | NHS | Colon Cancer | 0.99 (0.89, 1.10) |
| Zhang, 2010 | CPS II | Colon Cancer | 1.02 (0.92, 1.13) |
| Zhang, 2010 | IWHS | Colon Cancer | 1.01 (0.91, 1.12) |
| Zhang, 2010 | HPFS | Colon Cancer | 0.96 (0.86, 1.07) |

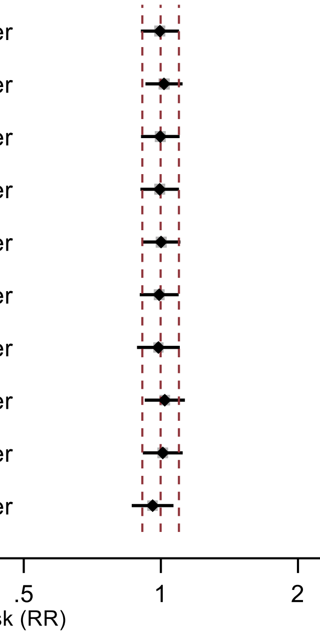

Figure legend: Forest plot shows the linear dose-response results (per 1 serving [355 mL] of carbonated sugar sweetened beverages/day) from the inverse variance DerSimonian-Laird random-effects model after exclusion of every single study at a time.

Abbreviations: CI, confidence interval; RR, relative risk.

Study abbreviations: SMC, Swedish Mammography Cohort; BCDDP, Breast Cancer Detection Demonstration Project follow-up study; ATBC, Alpha-Tocopherol, Beta-Carotene Cancer Prevention Study; NLCS, Netherlands Cohort Study on Diet and Cancer; WHS, Women's Health Study; CNBSS, Canadian National Breast Screening Study; NHS, Nurses' Health Study; CPS II, Cancer Prevention Study II; IWHS, The Iowa Women's Health Study; HPFS, Health Professionals Follow-up Study

Zhang, 2010[182]

**Supplementary Figure 158. Linear dose-response meta-analysis of sugar sweetened beverages and proximal colon cancer incidence, leave-one-out analysis.**

**Sugar sweetened beverages, per 1 serving/day and proximal colon cancer incidence, leave-one-out analysis**

| Author | Study | Outcome | RR (95% CI) |
|---|---|---|---|
| Pacheco, 2019 | CTS | Proximal Colon Cancer | 1.04 (0.96, 1.13) |
| Leung, 2021 | JPHC | Proximal Colon Cancer | 1.06 (0.93, 1.22) |
| Yuan, 2022 | NHS | Proximal Colon Cancer | 1.03 (0.95, 1.12) |
| Yuan, 2022 | HPFS | Proximal Colon Cancer | 1.00 (0.97, 1.03) |
| Kanehara, 2024 | MEC | Proximal Colon Cancer | 1.08 (0.96, 1.21) |

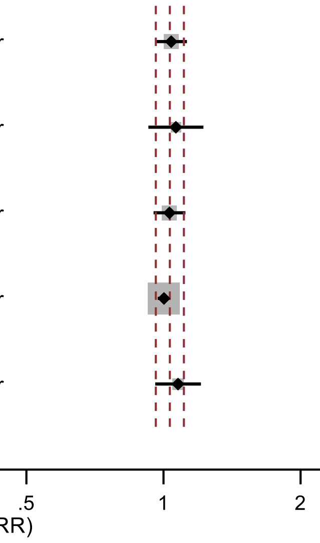

Figure legend: Forest plot shows the linear dose-response results (per 1 serving [355 mL] of sugar sweetened beverages/day) from the inverse variance DerSimonian-Laird random-effects model after exclusion of every single study at a time.

Abbreviations: CI, confidence interval; RR, relative risk.

Study abbreviations: CTS, California Teachers Study; JPHC, Japan Public Health Center-Based Prospective Study; NHS, Nurses' Health Study; HPFS, Health Professionals Follow-up Study; MEC, Multiethnic Cohort Study

Pacheco, 2019[135], Leung, 2021[101], Yuan, 2022[178], Kanehara, 2024[81]

**Supplementary Figure 159. Linear dose-response meta-analysis of sugar sweetened beverages and distal colon cancer incidence, leave-one-out analysis.**

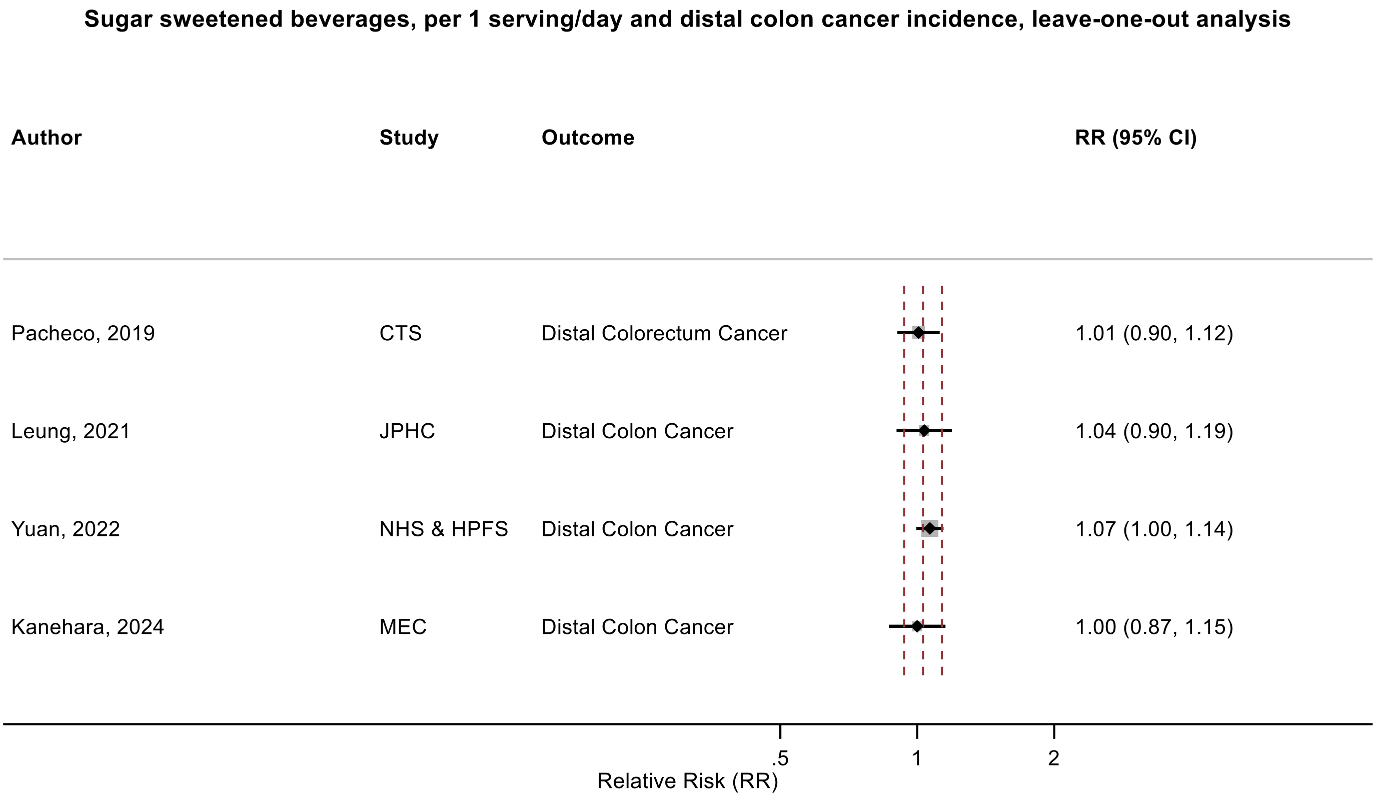

Figure legend: Forest plot shows the linear dose-response results (per 1 serving [355 mL] of sugar sweetened beverages/day) from the inverse variance DerSimonian-Laird random-effects model after exclusion of every single study at a time.

Abbreviations: CI, confidence interval; RR, relative risk.

Study abbreviations: CTS, California Teachers Study; JPHC, Japan Public Health Center-Based Prospective Study; NHS & HPFS, Nurses' Health Study & Health Professionals Follow-up Study; MEC, Multiethnic Cohort Study

Pacheco, 2019[135], Leung, 2021[101], Yuan, 2022[178], Kanehara, 2024[81]

**Supplementary Figure 160. Linear dose-response meta-analysis of sugar sweetened beverages and rectal cancer incidence, leave-one-out analysis.**

**Sugar sweetened beverages, per 1 serving/day and rectal cancer incidence, leave-one-out analysis**

| Author | Study | Outcome | RR (95% CI) |
|---|---|---|---|
| Leung, 2021 | JPHC | Rectal Cancer | 0.97 (0.90, 1.06) |
| Yuan, 2022 | NHS & HPFS | Rectal Cancer | 0.99 (0.92, 1.06) |
| Kanehara, 2024 | MEC | Rectal Cancer | 0.95 (0.86, 1.06) |

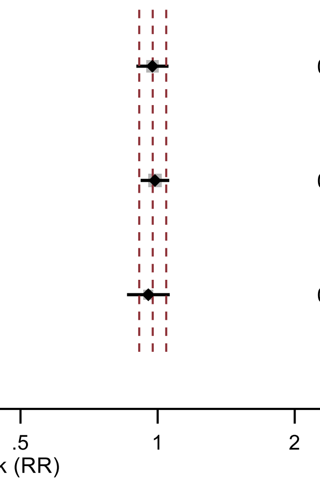

Figure legend: Forest plot shows the linear dose-response results (per 1 serving [355 mL] of sugar sweetened beverages/day) from the inverse variance DerSimonian-Laird random-effects model after exclusion of every single study at a time.

Abbreviations: CI, confidence interval; RR, relative risk.

Study abbreviations: JPHC, Japan Public Health Center-Based Prospective Study; NHS & HPFS, Nurses' Health Study & Health Professionals Follow-up Study; MEC, Multiethnic Cohort Study

Leung, 2021[101], Yuan, 2022[178], Kanehara, 2024[81]

**Supplementary Figure 161. Linear dose-response meta-analysis of sugar sweetened beverages and liver cancer incidence, leave-one-out analysis.**

**Sugar sweetened beverages, per 1 serving/day and liver cancer incidence, leave-one-out**

| Author | Study | Outcome | RR (95% CI) |
|---|---|---|---|
| Zhao, 2023 | WHI | Liver Cancer | 1.04 (0.94, 1.14) |
| Jones, 2022 | NIH-AARP & PLCO | Liver Cancer | 1.12 (0.86, 1.47) |
| Luu, 2021 | SCHS | HCC | 1.14 (0.92, 1.41) |
| Stepien, 2016 | EPIC | HCC | 1.10 (0.92, 1.32) |

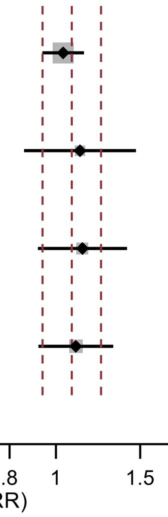

Figure legend: Forest plot shows the linear dose-response results (per 1 serving [355 mL] of sugar sweetened beverages/day) from the inverse variance DerSimonian-Laird random-effects model after exclusion of every single study at a time.

Abbreviations: CI, confidence interval; HCC, hepatocellular carcinoma; RR, relative risk.

Study abbreviations: EPIC, European Prospective Investigation into Cancer and nutrition; SCHS, Singapore Chinese Health study; NIH-AARP & PLCO, National Institute of Health + American Association of Retired Persons Diet and Health Study & Prostate, Lung, Colorectal and Ovarian Cancer Screening trial; WHI, Women’s Health Initiative Study

Stepien, 2016[159], Luu, 2021[106], Jones, 2022[79], Zhao, 2023[186]

**Supplementary Figure 162. Linear dose-response meta-analysis of sugar sweetened beverages and pancreatic cancer incidence, leave-one-out analysis.**

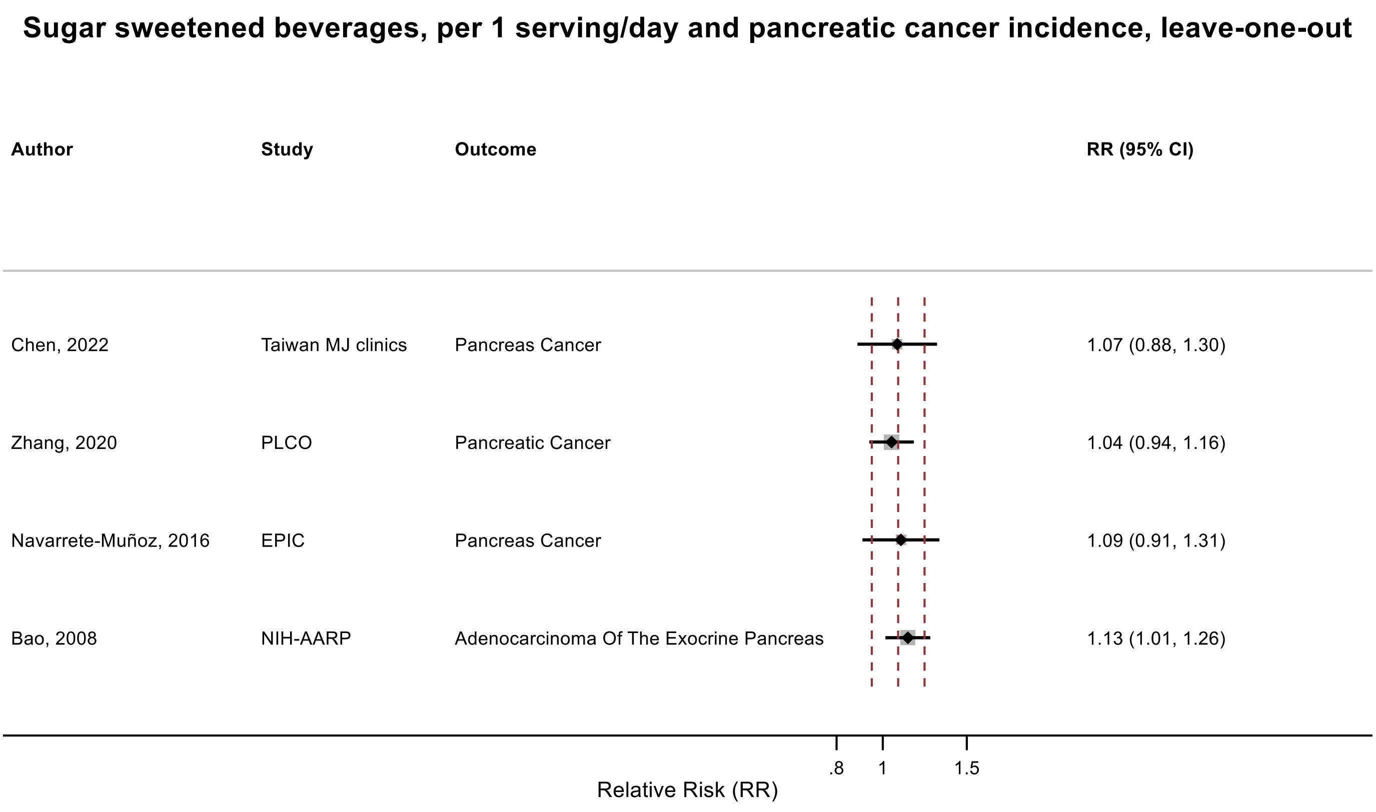

Figure legend: Forest plot shows the linear dose-response results (per 1 serving [355 mL] of sugar sweetened beverages/day) from the inverse variance DerSimonian-Laird random-effects model after exclusion of every single study at a time.

Abbreviations: CI, confidence interval; RR, relative risk.

Study abbreviations: NIH-AARP, National Institute of Health + American Association of Retired Persons Diet and Health Study; EPIC, European Prospective Investigation into Cancer and nutrition; PLCO, Prostate, Lung, Colorectal and Ovarian Cancer Screening trial; Taiwan MJ clinics, The Taiwan MJ Cohort Study

Bao, 2008[39], Navarrete-Muñoz, 2016[127], Zhang, 2020[184], Chen, 2022[45]

**Supplementary Figure 163. Linear dose-response meta-analysis of sugar sweetened beverages and breast cancer incidence, leave-one-out analysis.**

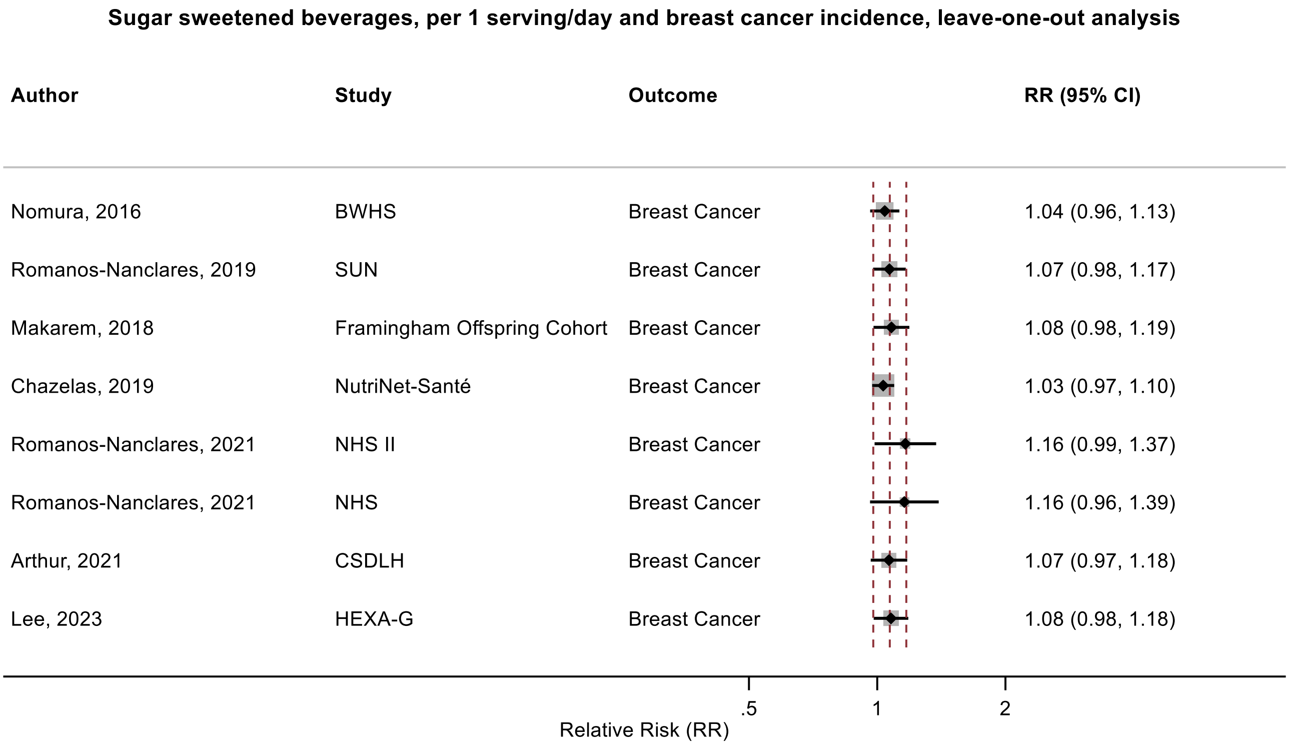

Figure legend: Forest plot shows the linear dose-response results (per 1 serving [355 mL] of sugar sweetened beverages/day) from the inverse variance DerSimonian-Laird random-effects model after exclusion of every single study at a time.

Abbreviations: CI, confidence interval; RR, relative risk.

Study abbreviations: BWHS, Black Women's Health Study; SUN, Seguimiento Universidad de Navarra Cohort Study; NHS II, Nurses' Health Study II; NHS, Nurses' Health Study; CSDLH, Canadian Study of Diet, Lifestyle and Health; HEXA-G, Health Examinees-Gem study

Nomura, 2016[128], Romanos-Nanclares, 2019[146], Makarem, 2018[108], Chazelas, 2019[44], Romanos-Nanclares, 2021[145], Arthur, 2021[38], Lee, 2023[97]

**Supplementary Figure 164. Linear dose-response meta-analysis of sugar sweetened beverages and endometrial cancer incidence, leave-one-out analysis.**

**Sugar sweetened beverages, per 1 serving/day and endometrial cancer incidence, leave-one-out analysis**

| Author | Study | Outcome | RR (95% CI) |
|---|---|---|---|
| Inoue-Choi, 2013 | IWHS | Endometrial Cancer | 1.28 (0.88, 1.85) |
| Hodge, 2018 | MCCS | Endometrial Cancer | 1.22 (1.03, 1.45) |
| Arthur, 2021 | CSDLH | Endometrial Cancer | 1.18 (0.99, 1.39) |

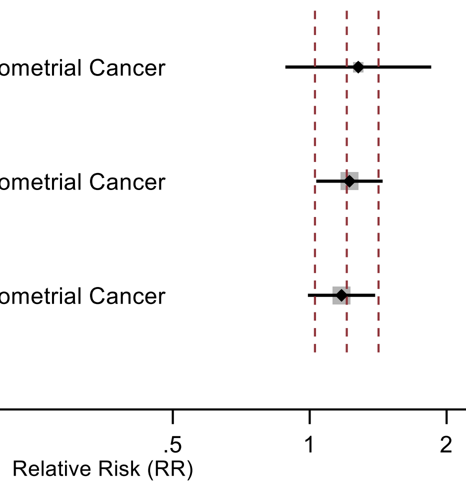

Figure legend: Forest plot shows the linear dose-response results (per 1 serving [355 mL] of sugar sweetened beverages/day) from the inverse variance DerSimonian-Laird random-effects model after exclusion of every single study at a time.

Abbreviations: CI, confidence interval; RR, relative risk.

Study abbreviations: IWHS, The Iowa Women's Health Study; MCCS, Melbourne Collaborative Cohort Study; CSDLH, Canadian Study of Diet, Lifestyle and Health

Inoue-Choi, 2013[74], Hodge, 2018[70], Arthur, 2021[38]

**Supplementary Figure 165. Linear dose-response meta-analysis of sugar sweetened beverages and prostate cancer incidence, leave-one-out analysis.**

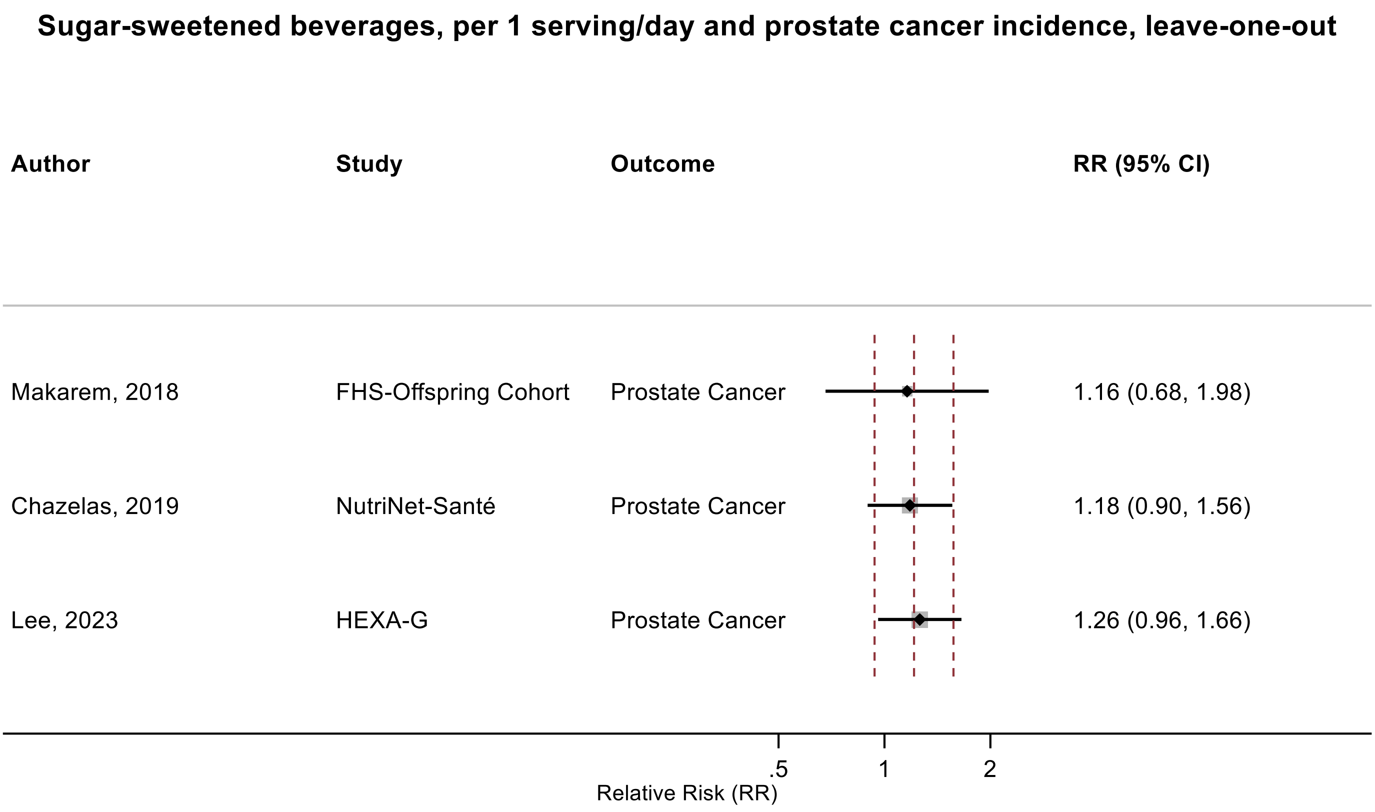

Figure legend: Forest plot shows the linear dose-response results (per 1 serving [355 mL] of sugar sweetened beverages/day) from the inverse variance DerSimonian-Laird random-effects model after exclusion of every single study at a time.

Abbreviations: CI, confidence interval; RR, relative risk.

Study abbreviations: MCCS, Melbourne Collaborative Cohort Study; HEXA-G, Health Examinees-Gem study

Makarem, 2018[108], Chazelas, 2019[44], Lee, 2023[97]

**Supplementary Figure 166. Linear dose-response meta-analysis of sugar sweetened beverages and kidney cancer incidence, leave-one-out analysis.**

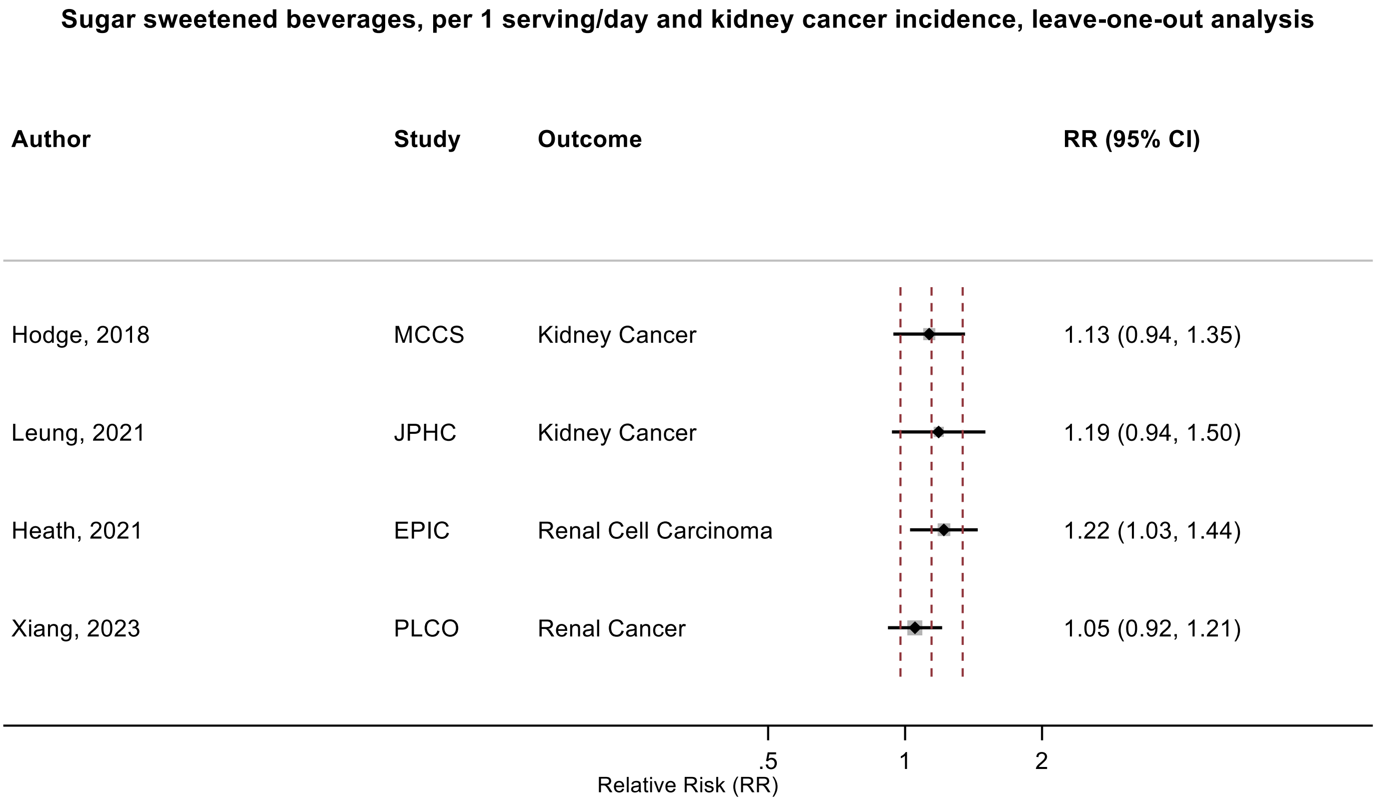

Figure legend: Forest plot shows the linear dose-response results (per 1 serving [355 mL] of sugar sweetened beverages/day) from the inverse variance DerSimonian-Laird random-effects model after exclusion of every single study at a time.

Abbreviations: CI, confidence interval; RR, relative risk.

Study abbreviations: MCCS, Melbourne Collaborative Cohort Study; JPHC, Japan Public Health Center-Based Prospective Study; EPIC, European Prospective Investigation into Cancer and nutrition; PLCO, Prostate, Lung, Colorectal and Ovarian Cancer Screening trial

Hodge, 2018[70], Leung, 2021[100], Heath, 2021[66], Xiang, 2023[173]

**Supplementary Figure 167. Linear dose-response meta-analysis of carbonated sugar sweetened beverages and non-Hodgkin lymphoma incidence, leave-one-out analysis.**

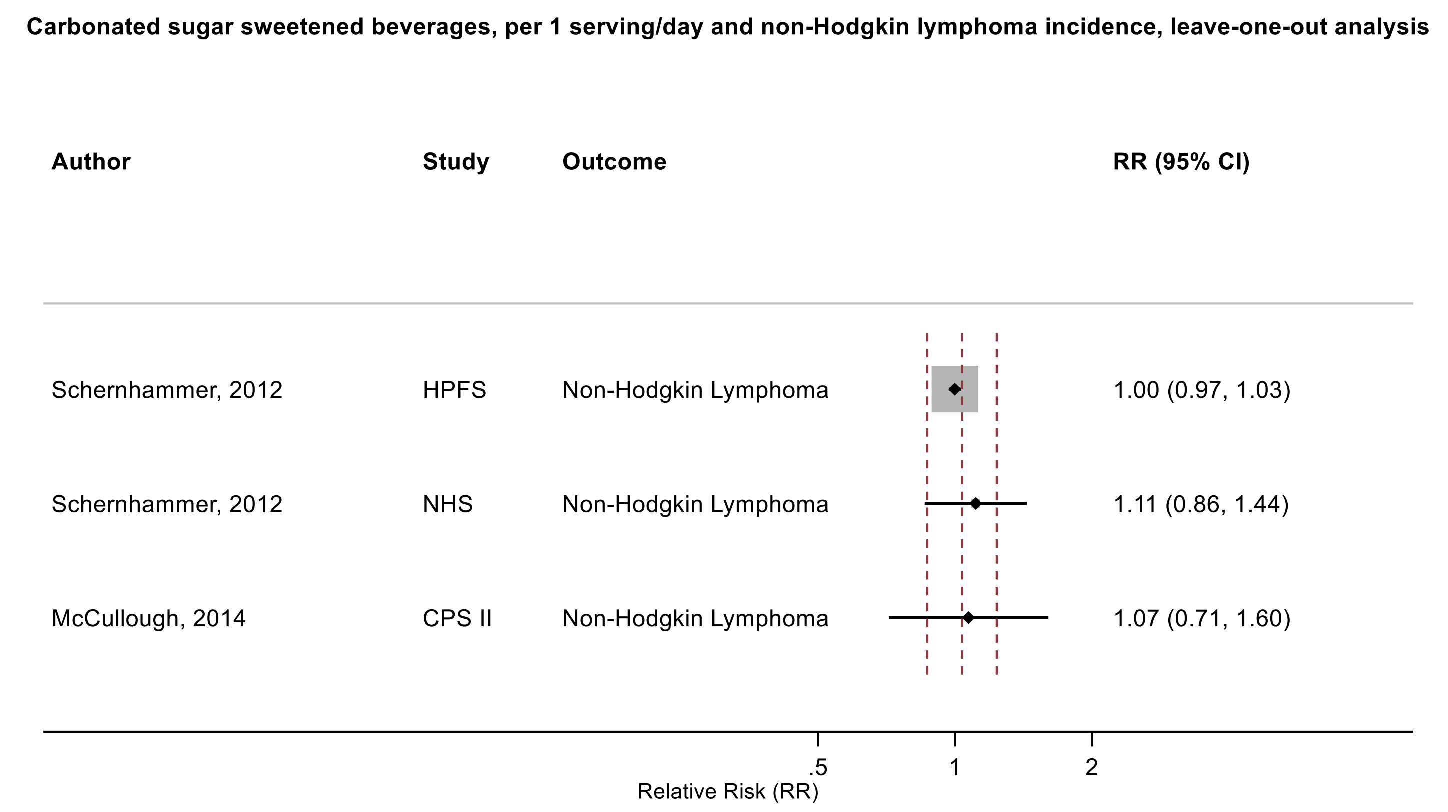

Figure legend: Forest plot shows the linear dose-response results (per 1 serving [355 mL] of carbonated sugar sweetened beverages/day) from the inverse variance DerSimonian-Laird random-effects model after exclusion of every single study at a time.

Abbreviations: CI, confidence interval; RR, relative risk.

Study abbreviations: HPFS, Health Professionals Follow-up Study; NHS, Nurses' Health Study; CPS II, Cancer Prevention Study II

Schernhammer, 2012[150], McCullough, 2014[114]

**Supplementary Figure 168. Linear dose-response meta-analysis of sugar sweetened beverages and kidney cancer incidence, comparison of the models with and without adjustment for body mass index.**

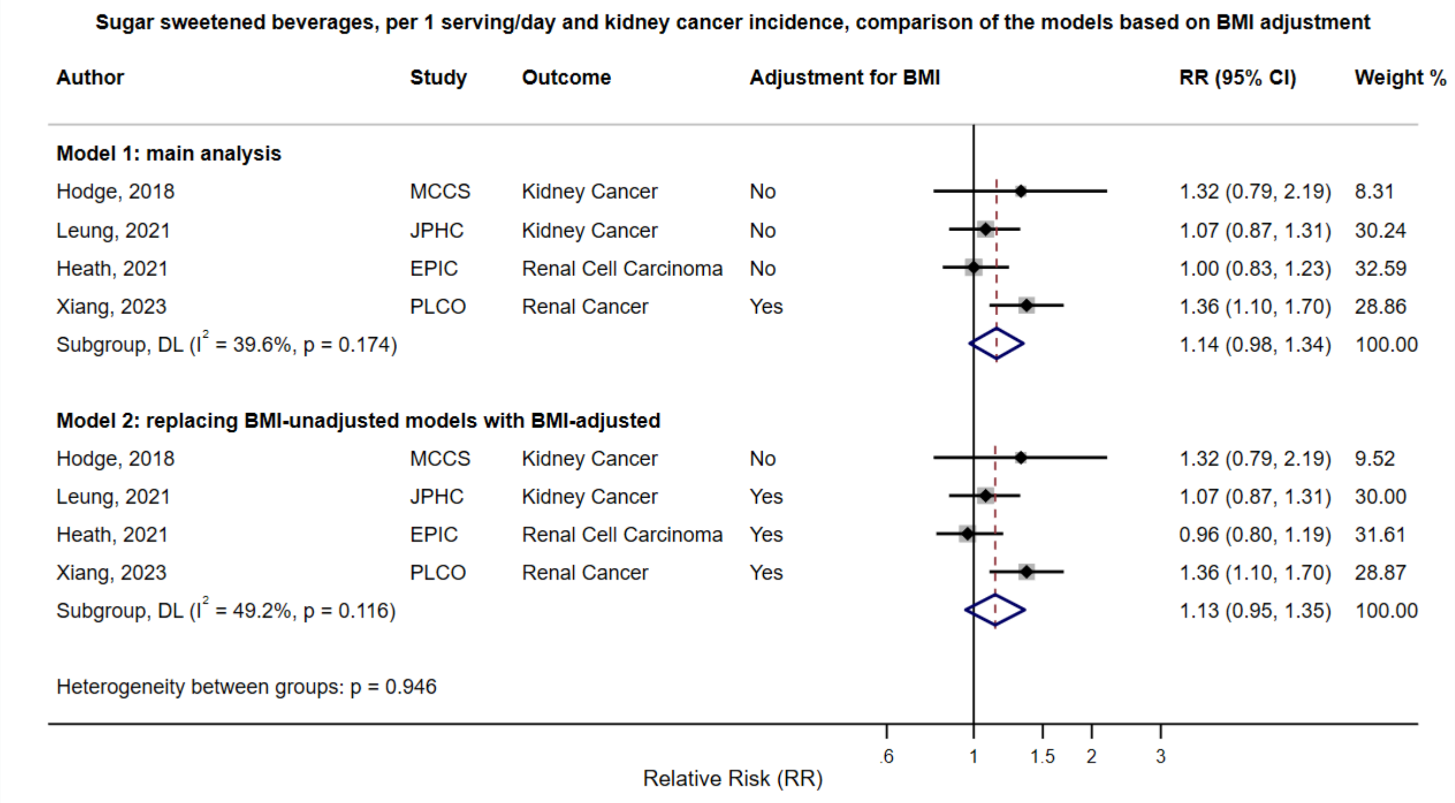

Figure legend: Forest plot shows the linear dose-response results (per 1 serving [355 mL] of sugar sweetened beverages/day) from the inverse variance DerSimonian-Laird random-effects model. The diamond represents the summary relative risk (RR) estimate, the diamond's width is the 95% confidence interval (CI). Each square and the horizontal line across the square represent the RR estimate of the individual study and its 95% CI.

Abbreviations: CI, confidence interval; RR, relative risk.

Study abbreviations: MCCS, Melbourne Collaborative Cohort Study; JPHC, Japan Public Health Center-Based Prospective Study; EPIC, European Prospective Investigation into Cancer and nutrition; PLCO, Prostate, Lung, Colorectal and Ovarian Cancer Screening trial

Hodge, 2018[70], Leung, 2021[100], Heath, 2021[66], Xiang, 2023[173]

**Supplementary Figure 169. Funnel plot of the association between sugar sweetened beverages, per 1 serving (355 mL)/day and colorectal cancer incidence.**

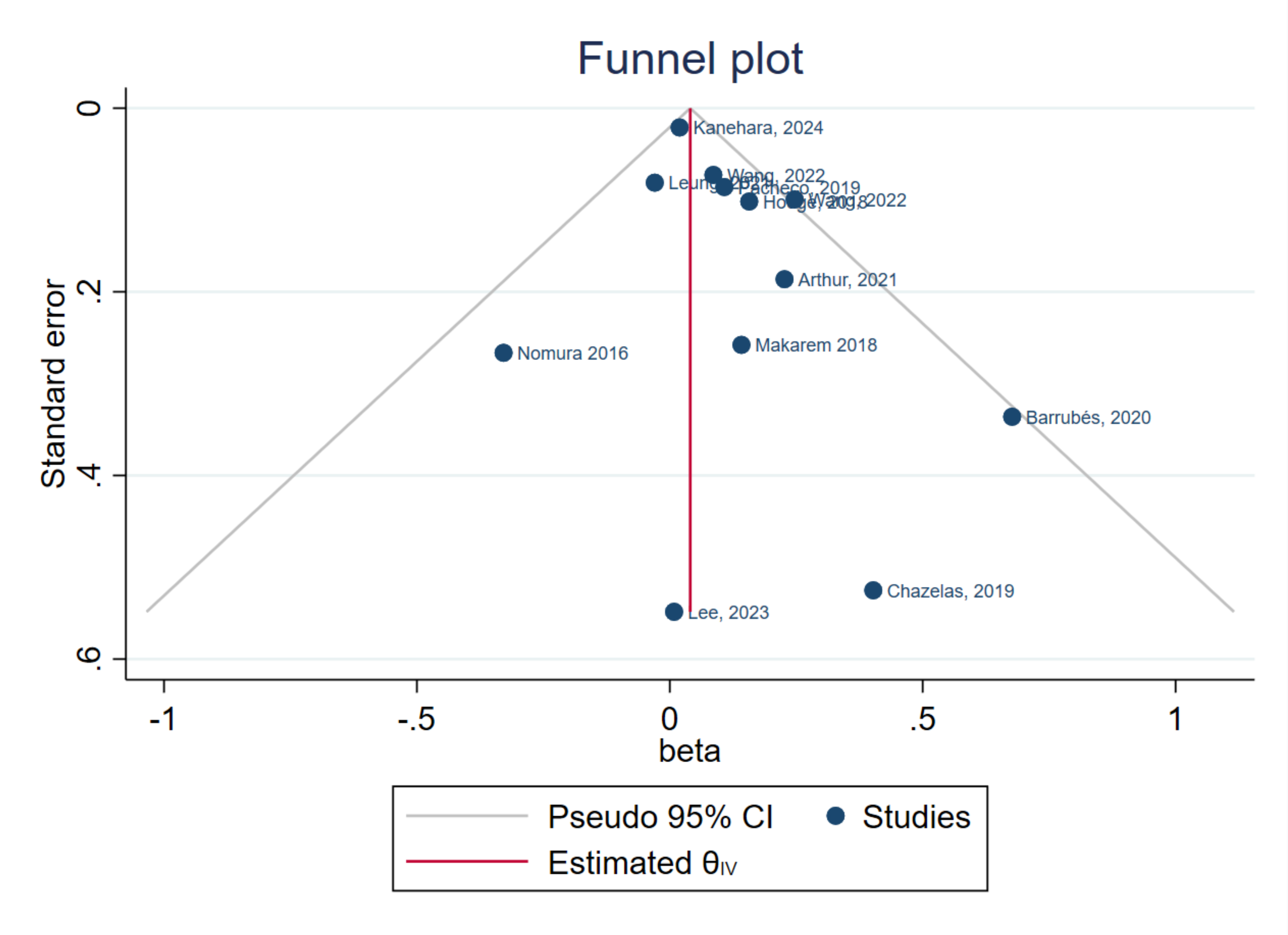

Note: Egger's p-value: 0.13

Figure legend: Each point represents the logarithm of relative risk estimate (x-axis) against its standard error as measure of study size (y-axis). Solid vertical line is the summary estimate of ln(RR) from the fixed effect meta-analysis. Grey lines are the 95% confidence limits around ln(RR).

Nomura, 2016[129], Chazelas, 2019[44], Barrubés, 2020[40], Makarem, 2018[108], Pacheco, 2019[135], Arthur, 2021[38], Wang, 2022[168], Kanehara, 2024[81],
Hodge, 2018[70], Leung, 2021[101], Lee, 2023[97]

**Supplementary Figure 170. Linear dose-response meta-analysis of sugar sweetened beverages and colorectal cancer mortality.**

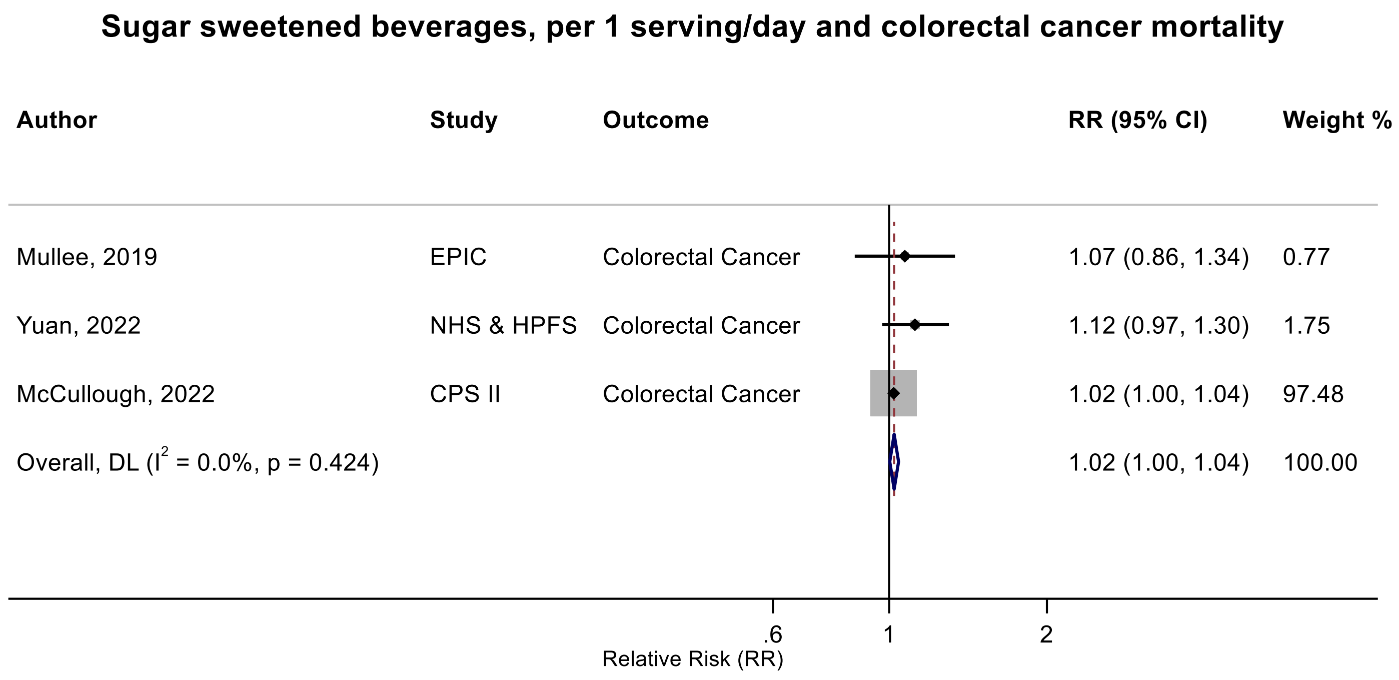

Figure legend: Forest plot shows the linear dose-response results (per 1 serving [355 mL] of sugar sweetened beverages /day) from the inverse variance DerSimonian-Laird random-effects model. The diamond represents the summary relative risk (RR) estimate, the diamond's width is the 95% confidence interval (CI). Each square and the horizontal line across the square represent the RR estimate of the individual study and its 95% CI.

Abbreviations: CI, confidence interval; RR, relative risk.

Study abbreviations: EPIC, European Prospective Investigation into Cancer and nutrition; NHS & HPFS, Nurses' Health Study & Health Professionals Follow-up Study; CPS II, Cancer Prevention Study II

Note: The pooled RR and 95%CI with 3 decimal point are 1.022 (1.002-1.042). All studies excluded people with a history of diabetes.

Mullee, 2019[126], Yuan, 2022[178], McCullough, 2022[113]

**Supplementary Figure 171. Linear dose-response meta-analysis of sugar sweetened beverages and colon and rectal cancer mortality.**

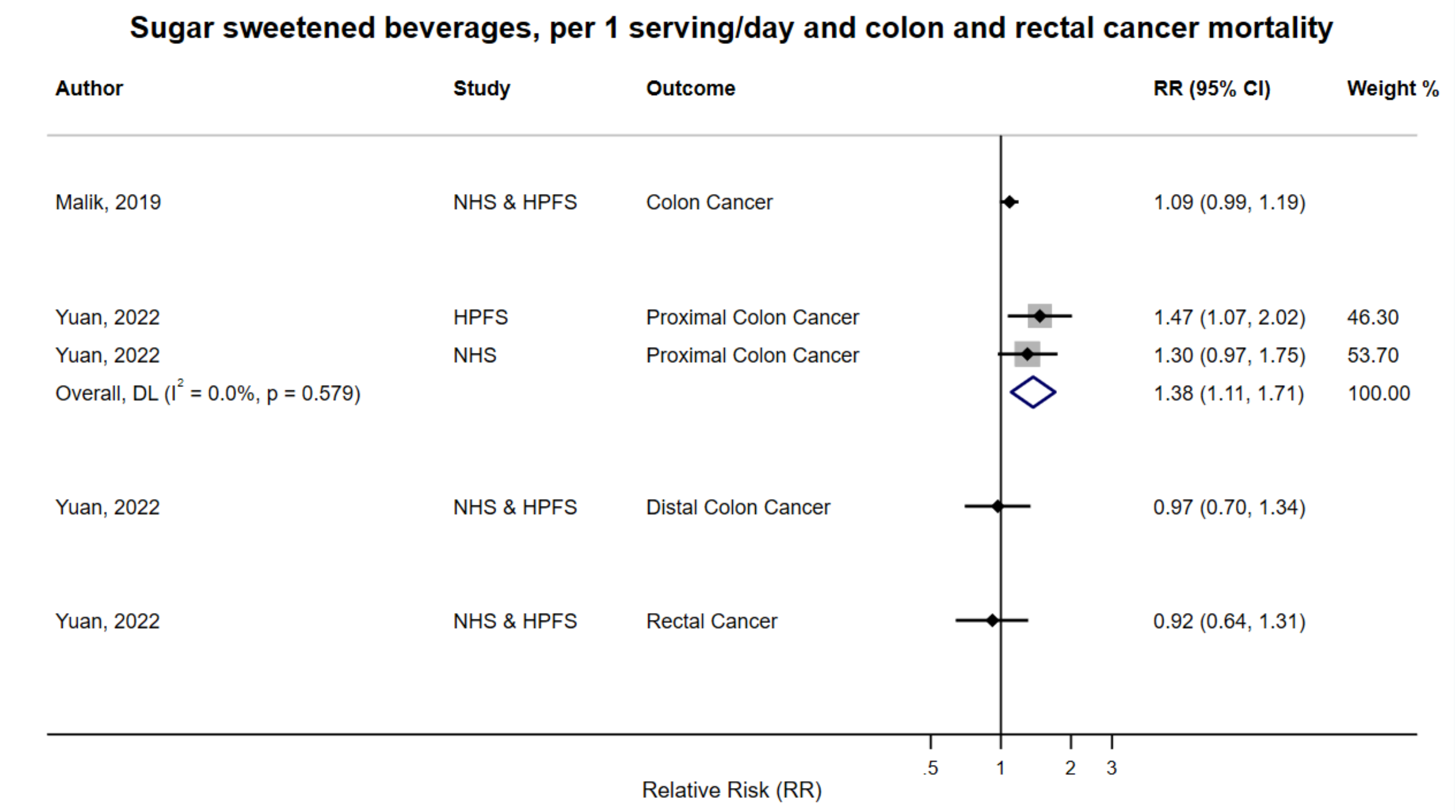

Figure legend: Forest plot shows the linear dose-response results (per 1 serving [355 mL] of sugar sweetened beverages /day) from the inverse variance DerSimonian-Laird random-effects model. The diamond represents the summary relative risk (RR) estimate, the diamond's width is the 95% confidence interval (CI). Each square and the horizontal line across the square represent the RR estimate of the individual study and its 95% CI.

Abbreviations: CI, confidence interval; RR, relative risk.

Study abbreviations: NHS, Nurse's Health Study; HPFS, Health Professionals Follow-up Study ; NHS & HPFS, Nurse's Health Study and Health Professionals Follow-up Study

Malik, 2019[109], Yuan, 2022[178]

**Supplementary Figure 172. Linear dose-response meta-analysis of sugar sweetened beverages and kidney cancer mortality.**

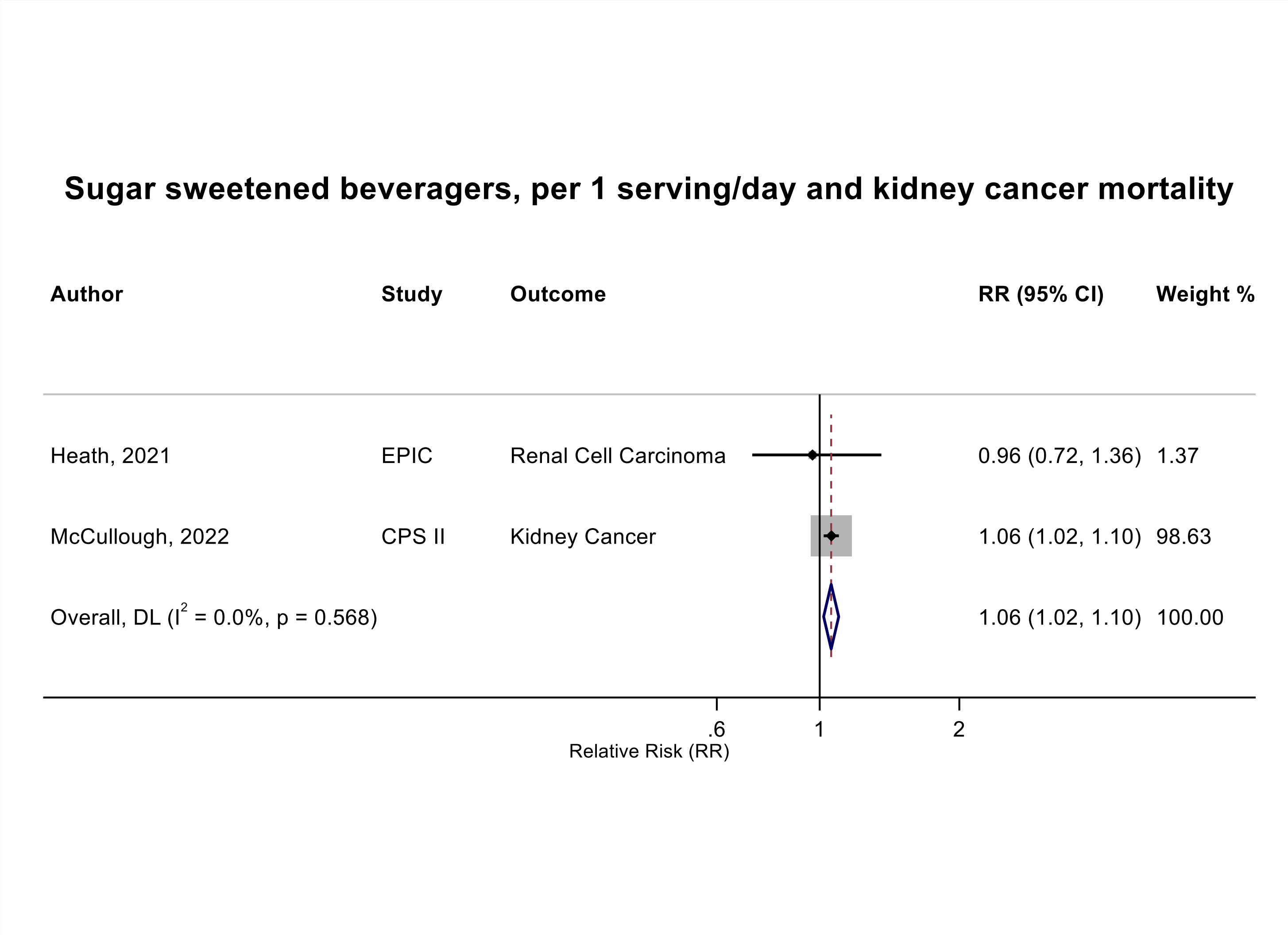

Figure legend: Forest plot shows the linear dose-response results (per 1 serving [355 mL] of sugar sweetened beverages /day) from the inverse variance DerSimonian-Laird random-effects model. The diamond represents the summary relative risk (RR) estimate, the diamond's width is the 95% confidence interval (CI). Each square and the horizontal line across the square represent the RR estimate of the individual study and its 95% CI.

Abbreviations: CI, confidence interval; RR, relative risk.

Study abbreviations: EPIC, European Prospective Investigation into Cancer and Nutrition; CPS II, Cancer prevention Study II

Heath, 2021[66], McCullough, 2022[113]

**Supplementary Figure 173. Linear dose-response meta-analysis of sugar sweetened beverages and lung cancer mortality.**

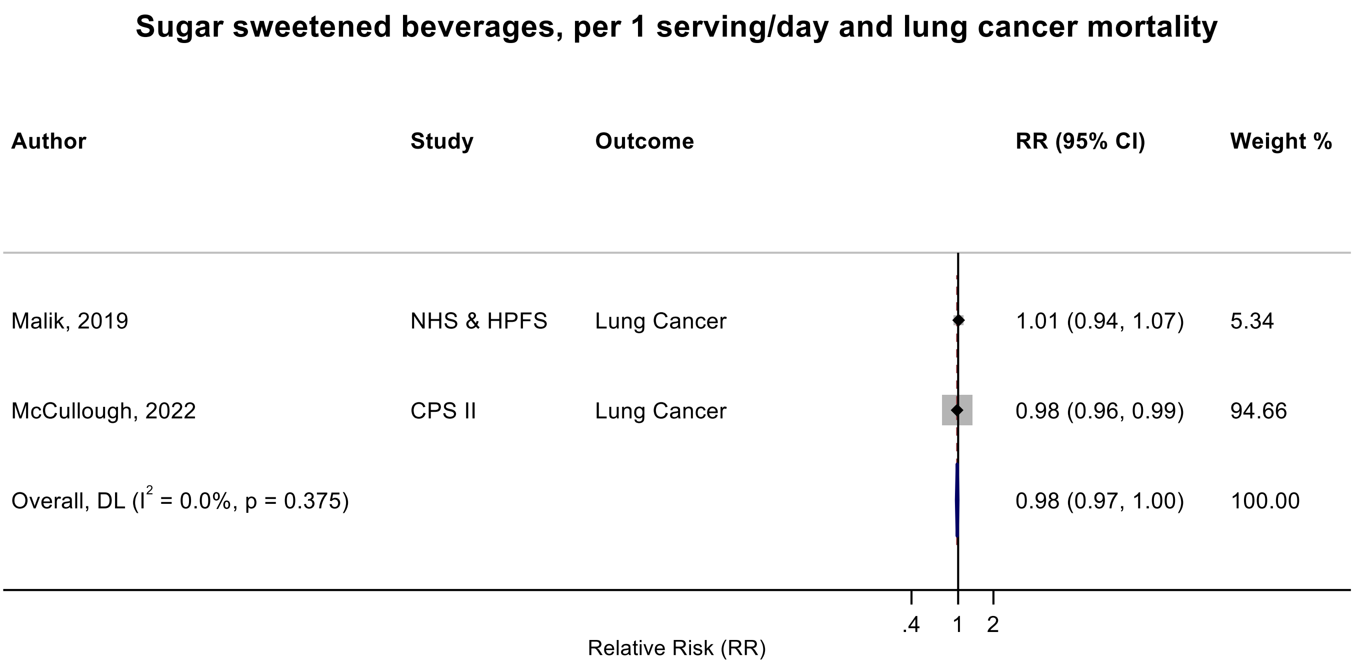

Figure legend: Forest plot shows the linear dose-response results (per 1 serving [355 mL] of sugar sweetened beverages /day) from the inverse variance DerSimonian-Laird random-effects model. The diamond represents the summary relative risk (RR) estimate, the diamond's width is the 95% confidence interval (CI). Each square and the horizontal line across the square represent the RR estimate of the individual study and its 95% CI.

Abbreviations: CI, confidence interval; RR, relative risk.

Study abbreviations: NHS, Nurse’s Health Study; HPFS, Health Professionals Follow-up Study; CPS II, Cancer prevention Study II

Malik, 2019[104], McCullough, 2022[108]

**Supplementary Figure 174. Linear dose-response meta-analysis of sugar sweetened beverages and breast cancer mortality.**

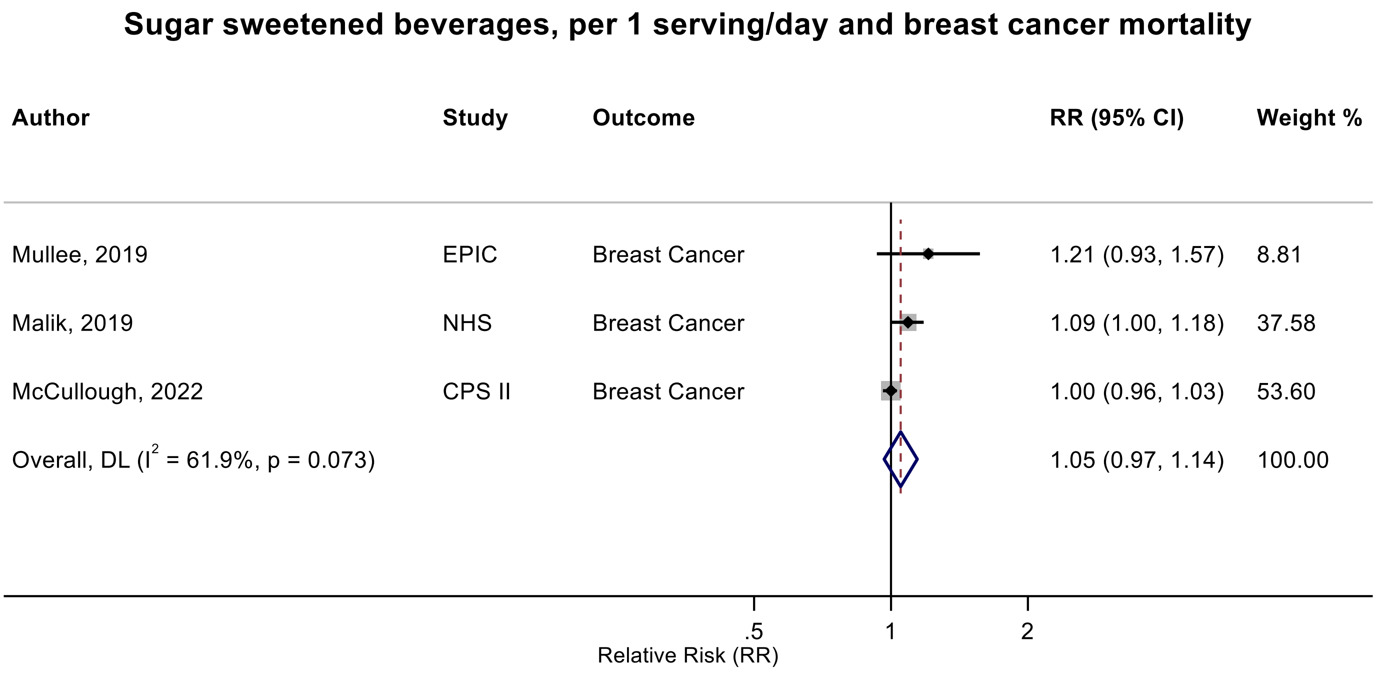

Figure legend: Forest plot shows the linear dose-response results (per 1 serving [355 mL] of sugar sweetened beverages /day) from the inverse variance DerSimonian-Laird random-effects model. The diamond represents the summary relative risk (RR) estimate, the diamond's width is the 95% confidence interval (CI). Each square and the horizontal line across the square represent the RR estimate of the individual study and its 95% CI.

Abbreviations: CI, confidence interval; RR, relative risk.

Study abbreviations: EPIC, European Prospective Investigation into Cancer and nutrition; NHS, Nurses' Health Study; CPS II, Cancer Prevention Study II

Mullee, 2019[126], Malik, 2019[109], McCullough, 2022[113]

**Supplementary Figure 175. Linear dose-response meta-analysis of sugar sweetened beverages and prostate cancer mortality.**

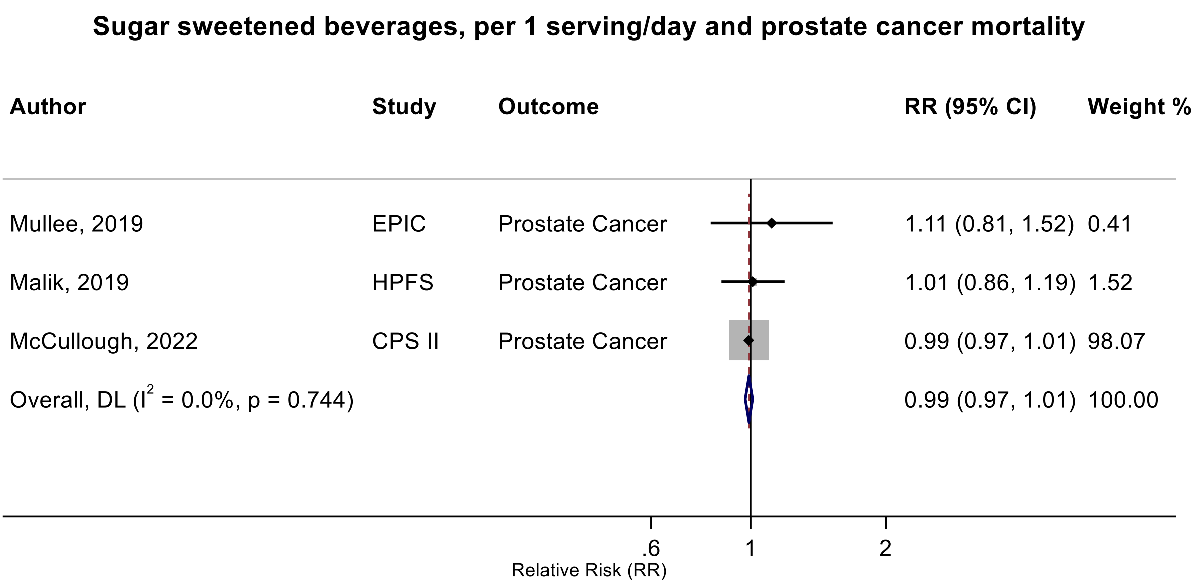

Figure legend: Forest plot shows the linear dose-response results (per 1 serving [355 mL] of sugar sweetened beverages /day) from the inverse variance DerSimonian-Laird random-effects model. The diamond represents the summary relative risk (RR) estimate, the diamond's width is the 95% confidence interval (CI). Each square and the horizontal line across the square represent the RR estimate of the individual study and its 95% CI.

Abbreviations: CI, confidence interval; RR, relative risk.

Study abbreviations: EPIC, European Prospective Investigation into Cancer and nutrition; HPFS, Health Professionals Follow-up Study; CPS II, Cancer Prevention Study II

Note: One additional study (NIH-AARP)[42] that could not be included in the dose-response meta-analysis because of insufficient data format reported on the association between sugar sweetened beverages as a component of Alternative Healthy Eating Index-2010 and prostate cancer mortality and reported little evidence of an association (RR: 0.99, 95%CI: 0.96-1.02) for higher score of the component (lower intake).

Mullee, 2019[126], Malik, 2019[109], McCullough, 2022[113]

**Supplementary Figure 176. Linear dose-response meta-analysis of sugar sweetened beverages and kidney cancer mortality by sex.**

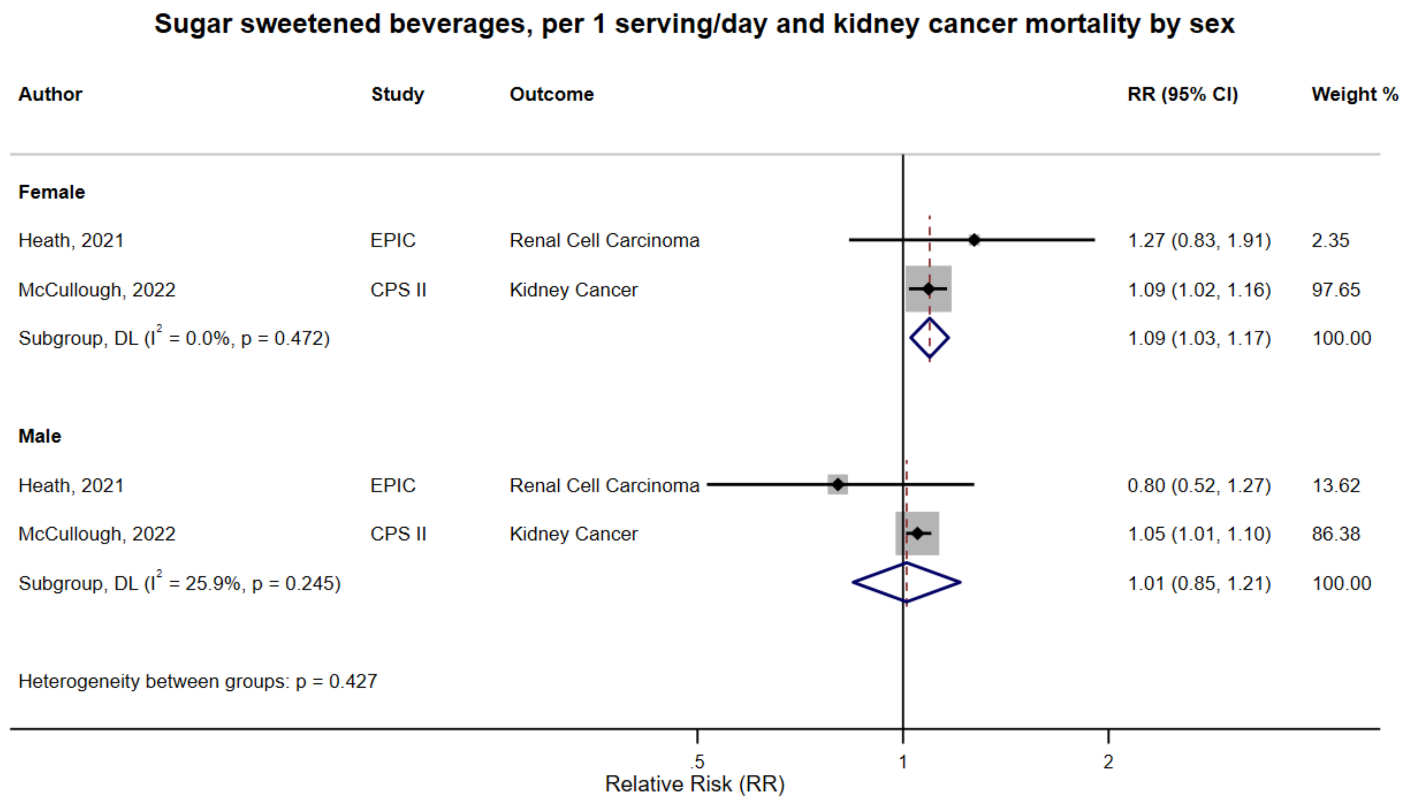

Figure legend: Forest plot shows the linear dose-response results (per 1 serving [355 mL] of sugar sweetened beverages /day) from the inverse variance DerSimonian-Laird random-effects model. The diamond represents the summary relative risk (RR) estimate, the diamond's width is the 95% confidence interval (CI). Each square and the horizontal line across the square represent the RR estimate of the individual study and its 95% CI.

Abbreviations: CI, confidence interval; RR, relative risk.

Study abbreviations: EPIC, European Prospective Investigation into Cancer and nutrition; CPS II, Cancer Prevention Study II

Heath, 2021[66], McCullough, 2022[113]

**Supplementary Figure 177. Linear dose-response meta-analysis of sugar sweetened beverages and colorectal cancer mortality by frequency of dietary assessment.**

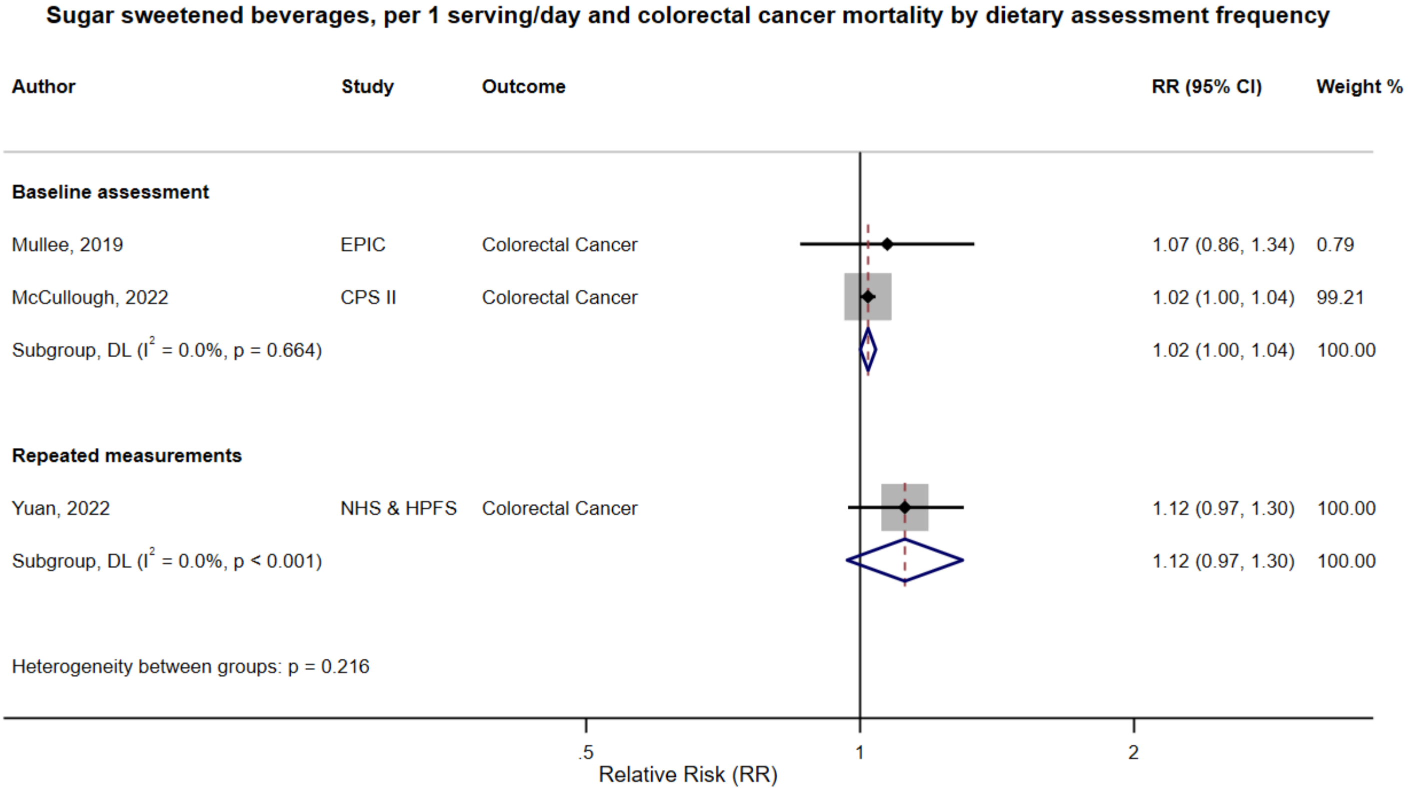

Figure legend: Forest plot shows the linear dose-response results (per 1 serving [355 mL] of sugar sweetened beverages /day) from the inverse variance DerSimonian-Laird random-effects model. The diamond represents the summary relative risk (RR) estimate, the diamond's width is the 95% confidence interval (CI). Each square and the horizontal line across the square represent the RR estimate of the individual study and its 95% CI.

Abbreviations: CI, confidence interval; RR, relative risk.

Study abbreviations: EPIC, European Prospective Investigation into Cancer and nutrition; CPS II, Cancer Prevention Study II; NHS & HPFS, Nurses' Health Study & Health Professionals Follow-up Study

Mullee, 2019[126], McCullough, 2022[113], Yuan, 2022[178]

**Supplementary Figure 178. Linear dose-response meta-analysis of sugar sweetened beverages and colorectal cancer mortality by geographical region.**

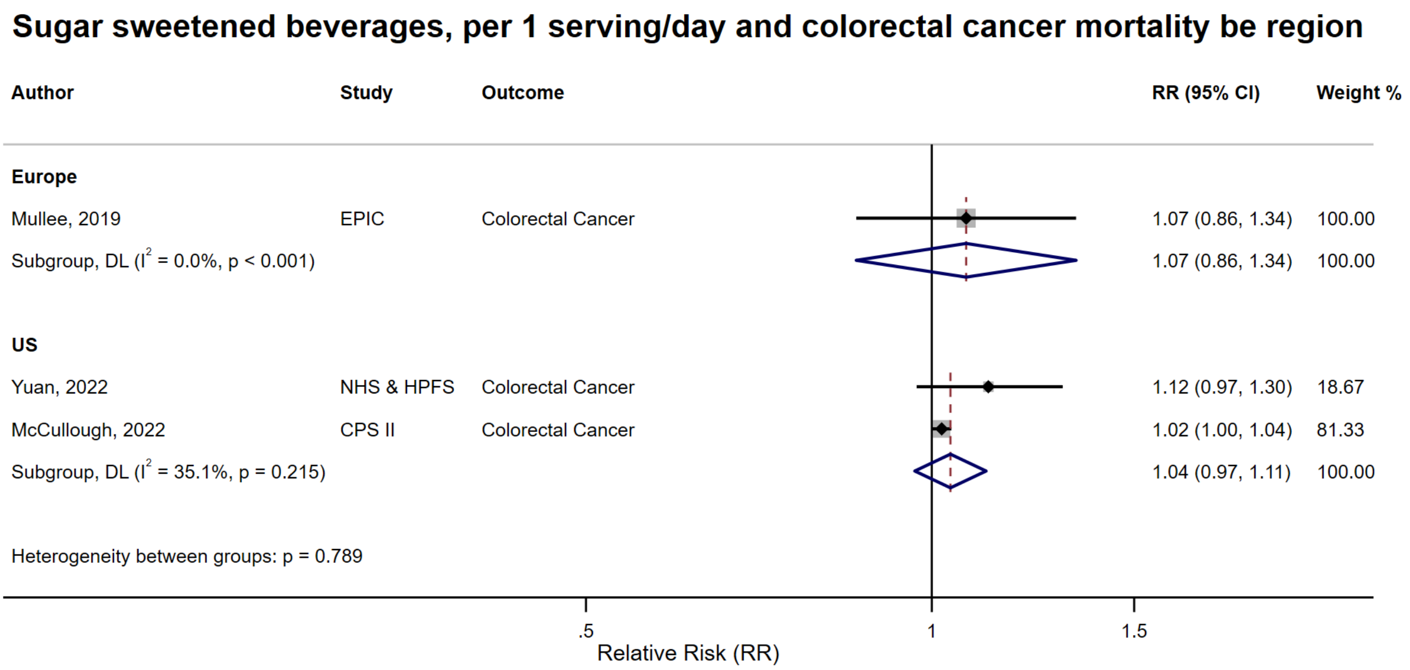

Figure legend: Forest plot shows the linear dose-response results (per 1 serving [355 mL] of sugar sweetened beverages /day) from the inverse variance DerSimonian-Laird random-effects model. The diamond represents the summary relative risk (RR) estimate, the diamond's width is the 95% confidence interval (CI). Each square and the horizontal line across the square represent the RR estimate of the individual study and its 95% CI.

Abbreviations: CI, confidence interval; RR, relative risk.

Study abbreviations: EPIC, European Prospective Investigation into Cancer and nutrition; NHS & HPFS, Nurses' Health Study & Health Professionals Follow-up Study; CPS II, Cancer Prevention Study II

Mullee, 2019[126], Yuan, 2022[178], McCullough, 2022[113]

#### Supplementary Figure 179. Linear dose-response meta-analysis of sugar sweetened beverages and colorectal cancer mortality by bias related to confounding.

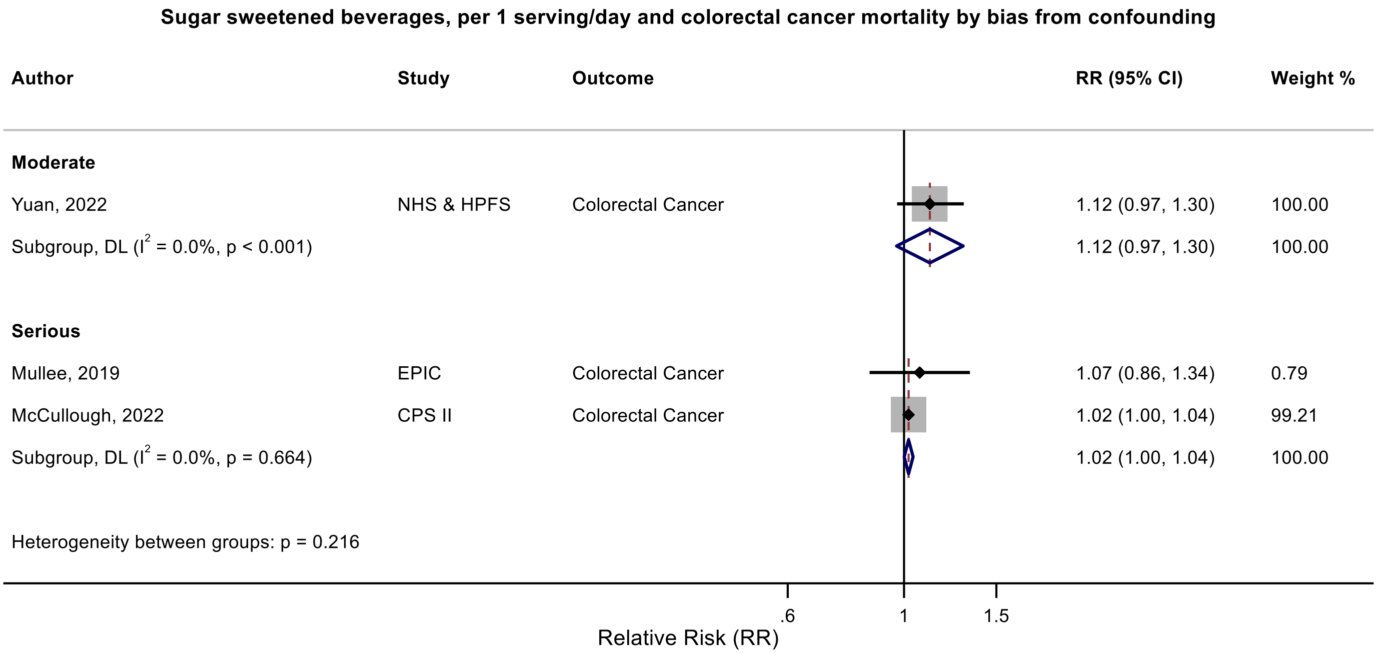

Figure legend: Forest plot shows the linear dose-response results (per 1 serving [355 mL] of sugar sweetened beverages /day) from the inverse variance DerSimonian-Laird random-effects model. The diamond represents the summary relative risk (RR) estimate, the diamond's width is the 95% confidence interval (CI). Each square and the horizontal line across the square represent the RR estimate of the individual study and its 95% CI.

Abbreviations: CI, confidence interval; RR, relative risk.

Study abbreviations: EPIC, European Prospective Investigation into Cancer and nutrition; NHS & HPFS, Nurses' Health Study & Health Professionals Follow-up Study; CPS II, Cancer Prevention Study II

Mullee, 2019[178], McCullough, 2022[113], Yuan, 2022[178]

**Supplementary Figure 180. Linear dose-response meta-analysis of sugar sweetened beverages and colorectal cancer mortality by bias related to departure from intended exposure.**

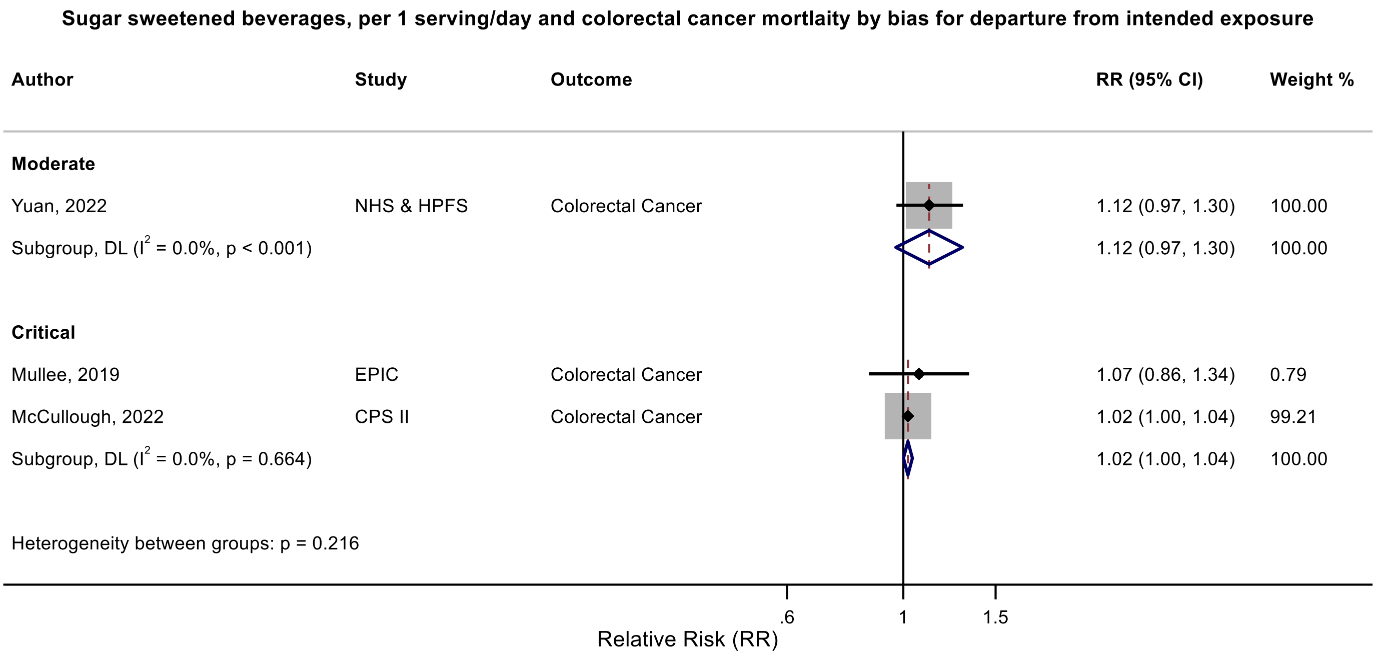

Figure legend: Forest plot shows the linear dose-response results (per 1 serving [355 mL] of sugar sweetened beverages /day) from the inverse variance DerSimonian-Laird random-effects model. The diamond represents the summary relative risk (RR) estimate, the diamond's width is the 95% confidence interval (CI). Each square and the horizontal line across the square represent the RR estimate of the individual study and its 95% CI.

Abbreviations: CI, confidence interval; RR, relative risk.

Study abbreviations: EPIC, European Prospective Investigation into Cancer and nutrition; NHS & HPFS, Nurses' Health Study & Health Professionals Follow-up Study; CPS II, Cancer Prevention Study II

Mullee, 2019[178], McCullough, 2022[113], Yuan, 2022[178]

**Supplementary Figure 181. Linear dose-response meta-analysis of sugar sweetened beverages and breast cancer mortality by frequency of dietary assessment.**

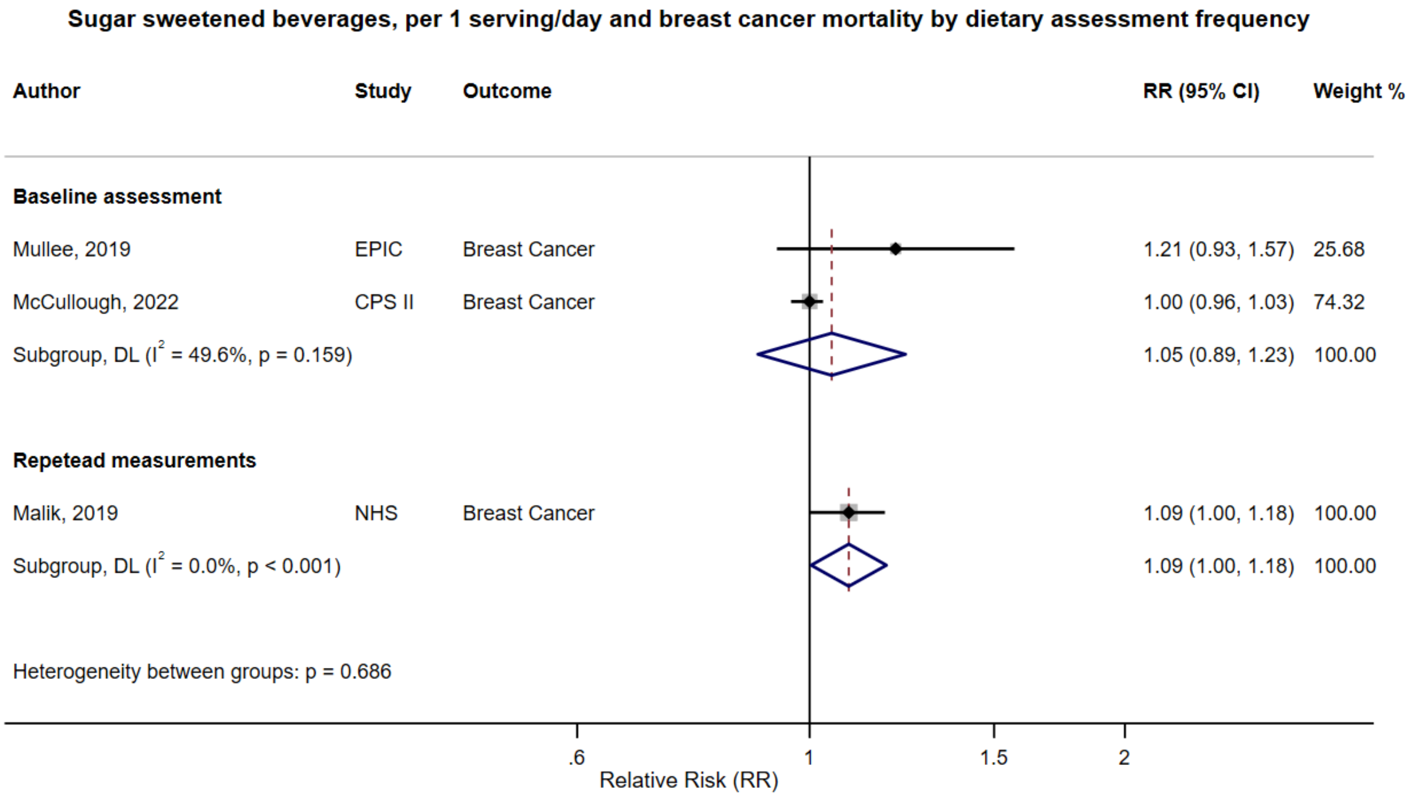

Figure legend: Forest plot shows the linear dose-response results (per 1 serving [355 mL] of sugar sweetened beverages /day) from the inverse variance DerSimonian-Laird random-effects model. The diamond represents the summary relative risk (RR) estimate, the diamond's width is the 95% confidence interval (CI). Each square and the horizontal line across the square represent the RR estimate of the individual study and its 95% CI.

Abbreviations: CI, confidence interval; RR, relative risk.

Study abbreviations: EPIC, European Prospective Investigation into Cancer and nutrition; CPS II, Cancer Prevention Study II; NHS, Nurses' Health Study

Mullee, 2019[126], McCullough, 2022[113], Malik, 2019[109]

**Supplementary Figure 182. Linear dose-response meta-analysis of sugar sweetened beverages and breast cancer mortality by exclusion of people with diabetes at baseline.**

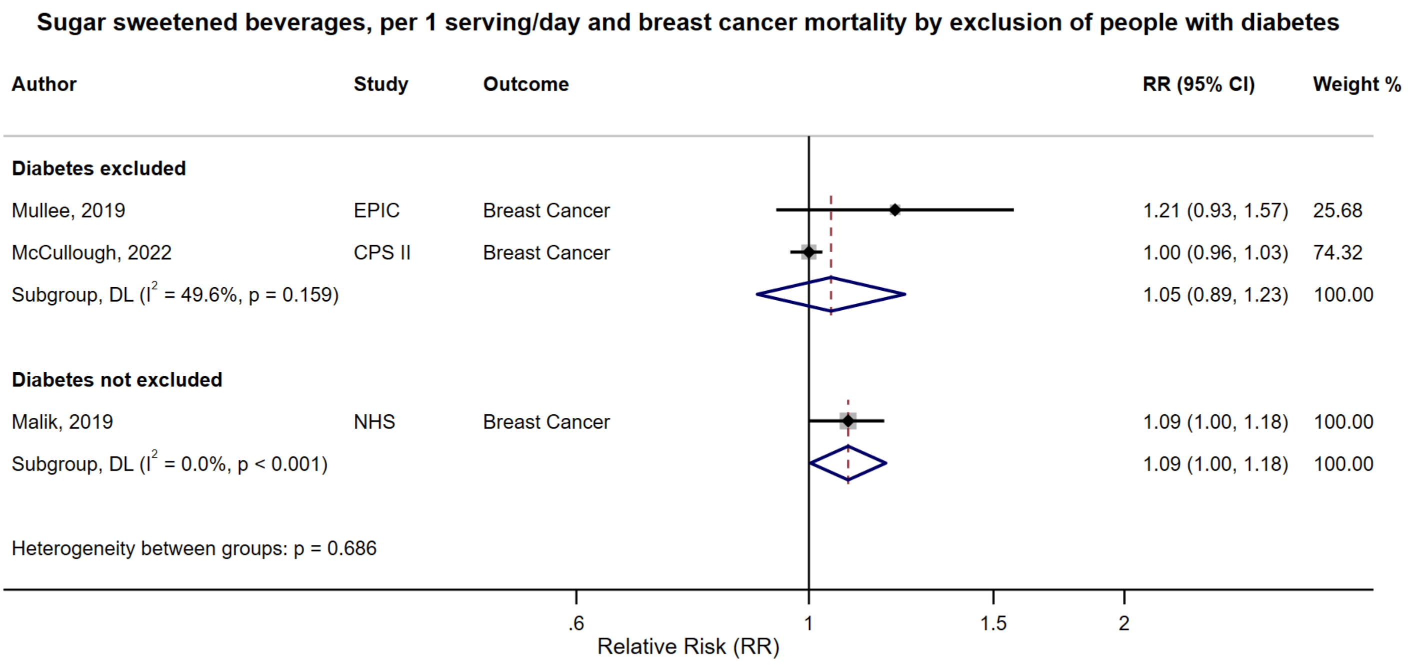

Figure legend: Forest plot shows the linear dose-response results (per 1 serving [355 mL] of sugar sweetened beverages /day) from the inverse variance DerSimonian-Laird random-effects model. The diamond represents the summary relative risk (RR) estimate, the diamond's width is the 95% confidence interval (CI). Each square and the horizontal line across the square represent the RR estimate of the individual study and its 95% CI.

Abbreviations: CI, confidence interval; RR, relative risk.

Study abbreviations: EPIC, European Prospective Investigation into Cancer and nutrition; CPS II, Cancer Prevention Study II; NHS, Nurses' Health Study

Note: The NHS controlled for a history of diabetes in the analysis.

Mullee, 2019[126], McCullough, 2022[113], Malik, 2019[109]

**Supplementary Figure 183. Linear dose-response meta-analysis of sugar sweetened beverages and breast cancer mortality by geographical region.**

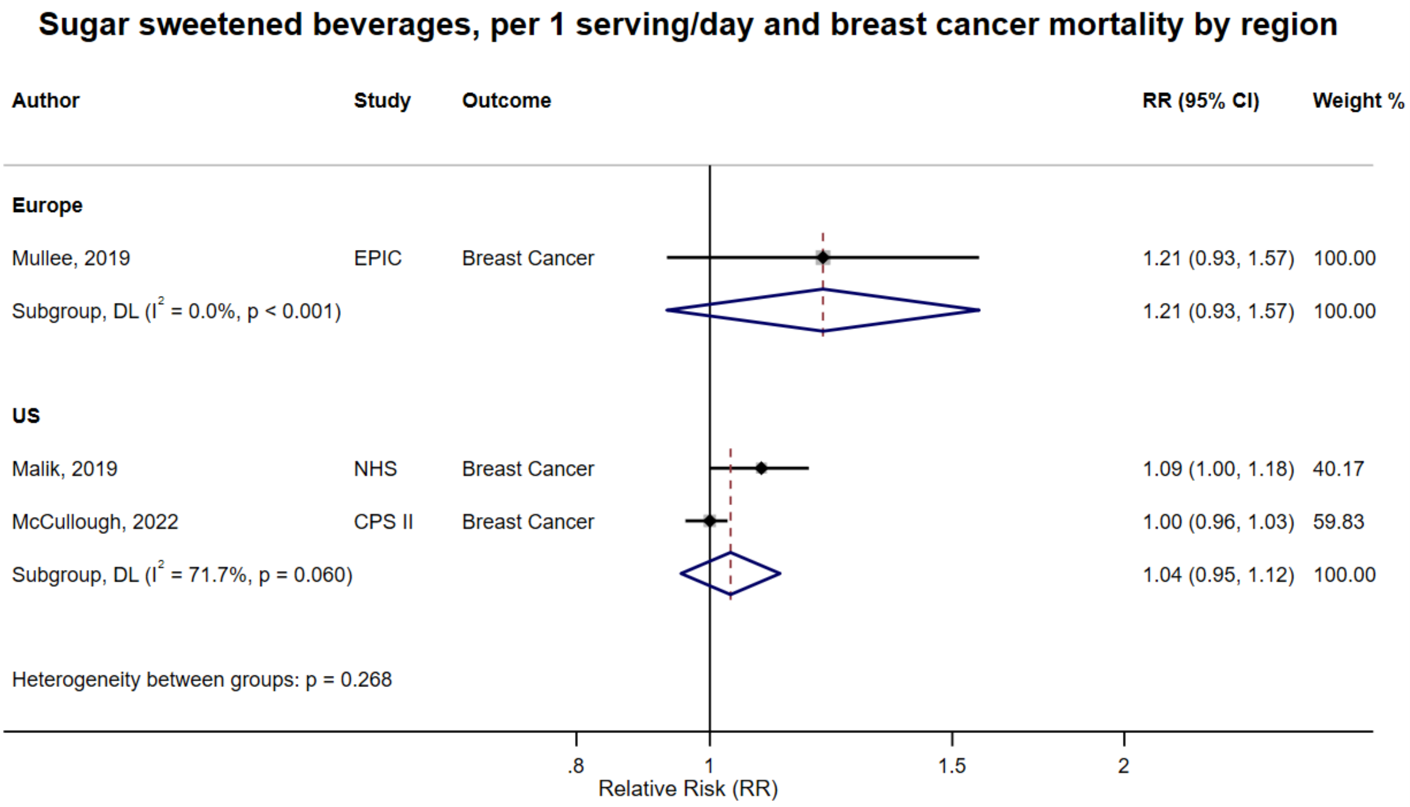

Figure legend: Forest plot shows the linear dose-response results (per 1 serving [355 mL] of sugar sweetened beverages /day) from the inverse variance DerSimonian-Laird random-effects model. The diamond represents the summary relative risk (RR) estimate, the diamond's width is the 95% confidence interval (CI). Each square and the horizontal line across the square represent the RR estimate of the individual study and its 95% CI.

Abbreviations: CI, confidence interval; RR, relative risk.

Study abbreviations: EPIC, European Prospective Investigation into Cancer and nutrition; NHS, Nurses' Health Study; CPS II, Cancer Prevention Study II

Mullee, 2019[126], Malik, 2019[109], McCullough, 2022[113]

**Supplementary Figure 184. Linear dose-response meta-analysis of sugar sweetened beverages and prostate cancer mortality by frequency of dietary assessment.**

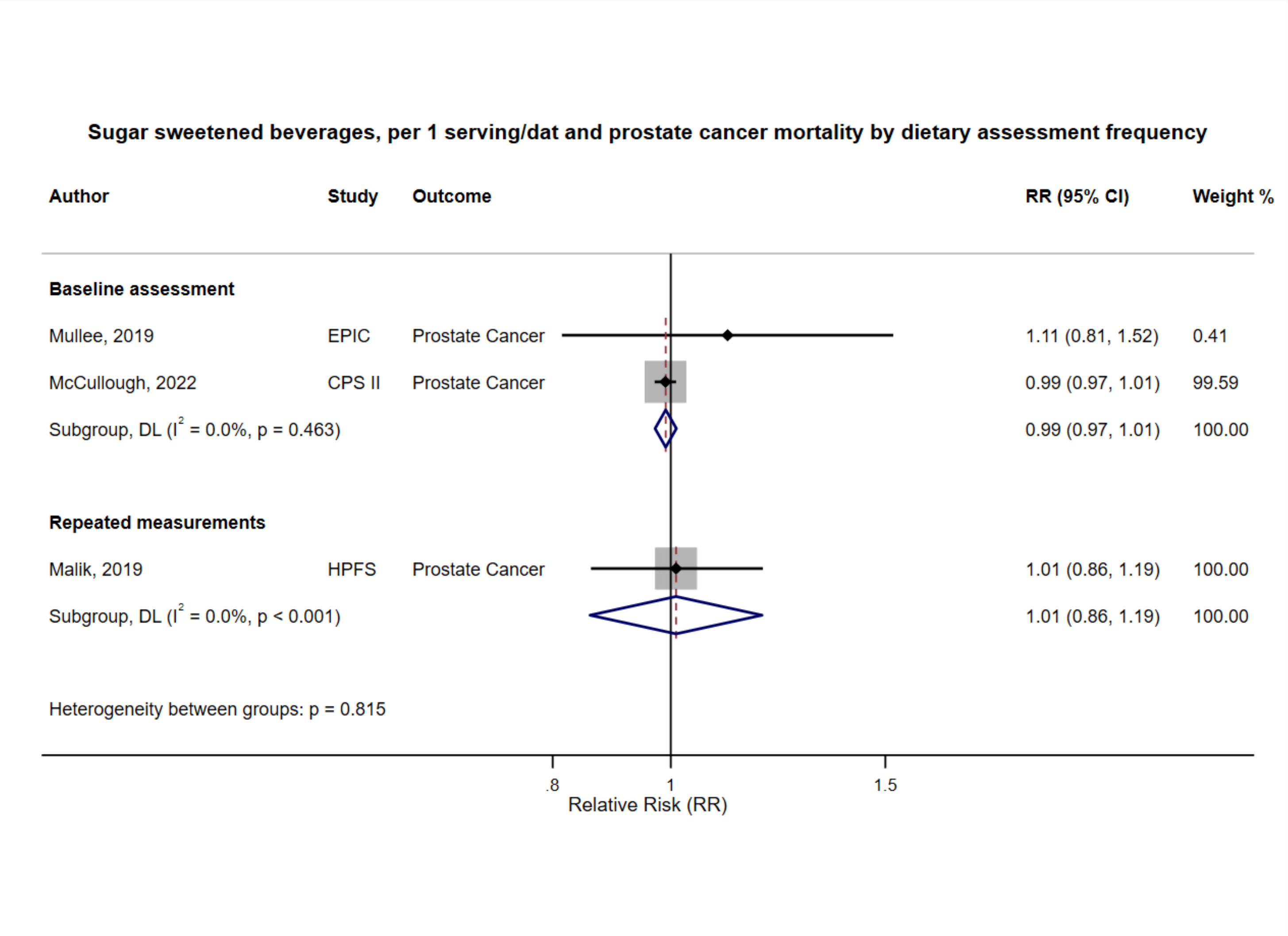

Figure legend: Forest plot shows the linear dose-response results (per 1 serving [355 mL] of sugar sweetened beverages /day) from the inverse variance DerSimonian-Laird random-effects model. The diamond represents the summary relative risk (RR) estimate, the diamond's width is the 95% confidence interval (CI). Each square and the horizontal line across the square represent the RR estimate of the individual study and its 95% CI.

Abbreviations: CI, confidence interval; RR, relative risk.

Study abbreviations: EPIC, European Prospective Investigation into Cancer and nutrition; CPS II, Cancer Prevention Study II; HPFS, Health Professionals Follow-up Study

Mullee, 2019[126], McCullough, 2022[113], Malik, 2019[109]

**Supplementary Figure 185. Linear dose-response meta-analysis of sugar sweetened beverages and prostate cancer mortality by exclusion of people with diabetes at baseline.**

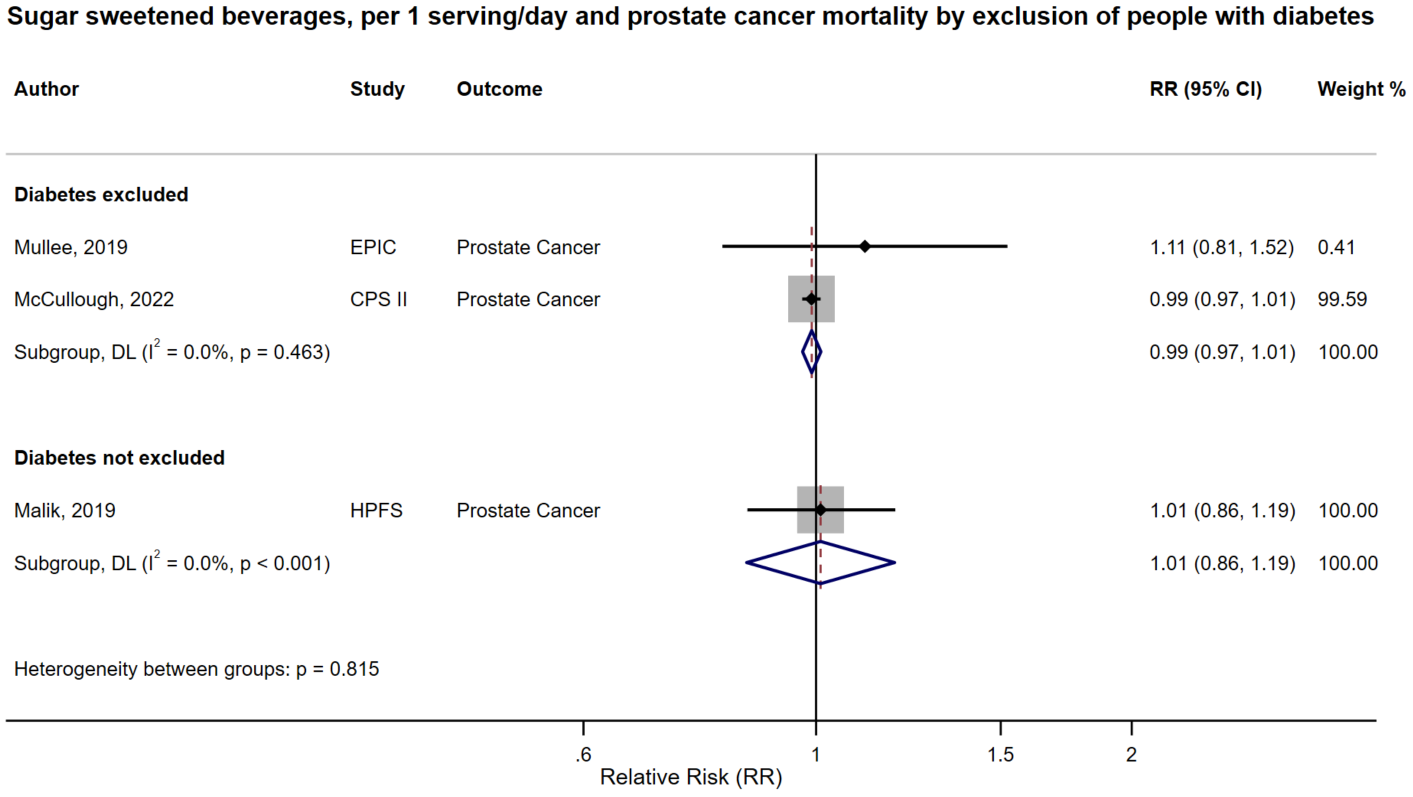

Figure legend: Forest plot shows the linear dose-response results (per 1 serving [355 mL] of sugar sweetened beverages /day) from the inverse variance DerSimonian-Laird random-effects model. The diamond represents the summary relative risk (RR) estimate, the diamond's width is the 95% confidence interval (CI). Each square and the horizontal line across the square represent the RR estimate of the individual study and its 95% CI.

Abbreviations: CI, confidence interval; RR, relative risk.

Study abbreviations: EPIC, European Prospective Investigation into Cancer and nutrition; CPS II, Cancer Prevention Study II; HPFS, Health Professionals Follow-up Study

Note: The HPFS controlled for a history of diabetes in the analysis.

Mullee, 2019[126], McCullough, 2022[113], Malik, 2019[109]

**Supplementary Figure 186. Linear dose-response meta-analysis of sugar sweetened beverages and prostate cancer mortality by geographical region.**

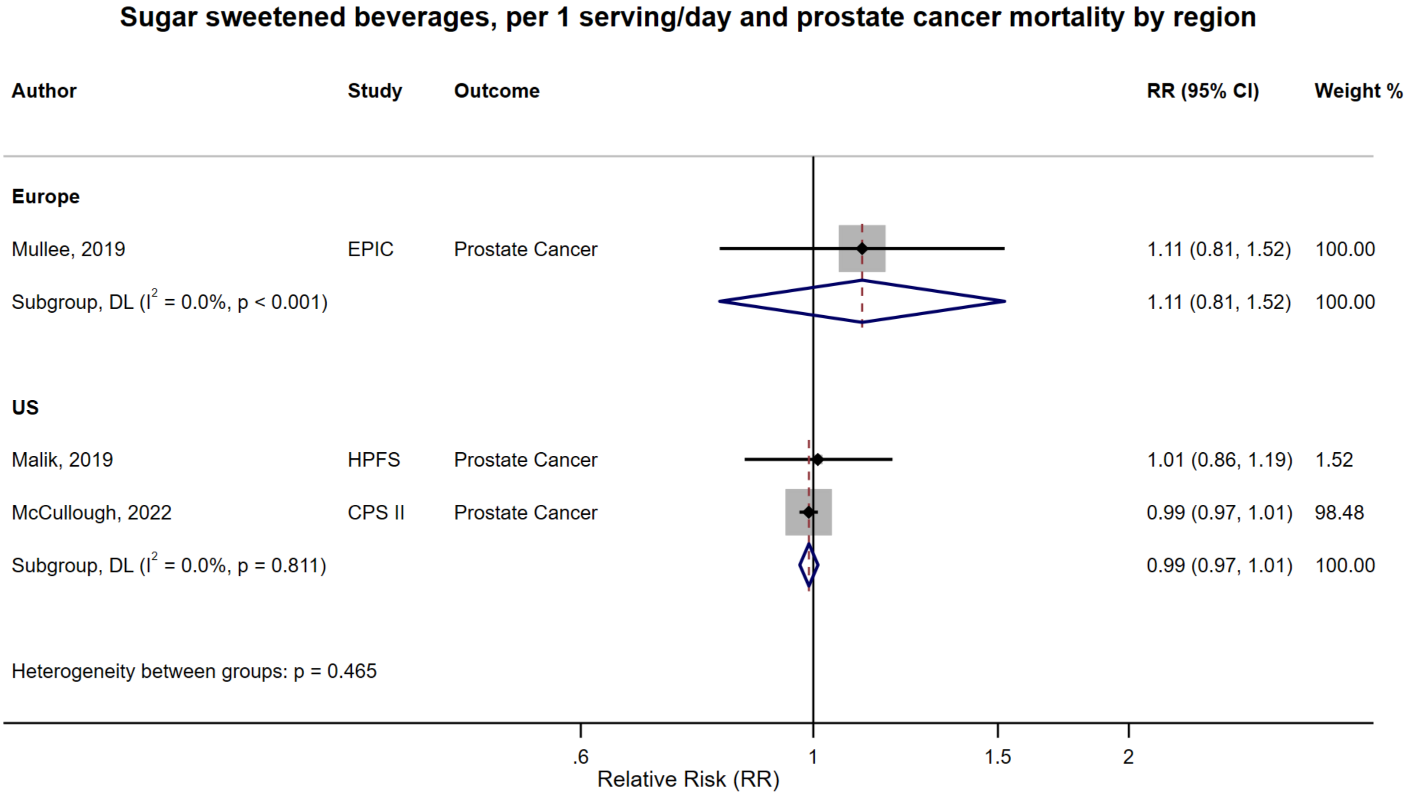

Figure legend: Forest plot shows the linear dose-response results (per 1 serving [355 mL] of sugar sweetened beverages /day) from the inverse variance DerSimonian-Laird random-effects model. The diamond represents the summary relative risk (RR) estimate, the diamond's width is the 95% confidence interval (CI). Each square and the horizontal line across the square represent the RR estimate of the individual study and its 95% CI.

Abbreviations: CI, confidence interval; RR, relative risk.

Study abbreviations: EPIC, European Prospective Investigation into Cancer and nutrition; HPFS, Health Professionals Follow-up Study; CPS II, Cancer Prevention Study II

Mullee, 2019[126], Malik, 2019[109], McCullough, 2022[113]

**Supplementary Figure 187. Linear dose-response meta-analysis of sugar sweetened beverages and breast cancer mortality, leave-one-out analysis.**

**Sugar sweetened beverages, per 1 serving/day and breast cancer mortality, leave-one-out analysis**

| Author | Study | Outcome | RR (95% CI) |
|---|---|---|---|
| Mullee, 2019 | EPIC | Breast Cancer | 1.04 (0.95, 1.12) |
| Malik, 2019 | NHS | Breast Cancer | 1.05 (0.89, 1.23) |
| McCullough, 2022 | CPS II | Breast Cancer | 1.10 (1.02, 1.19) |

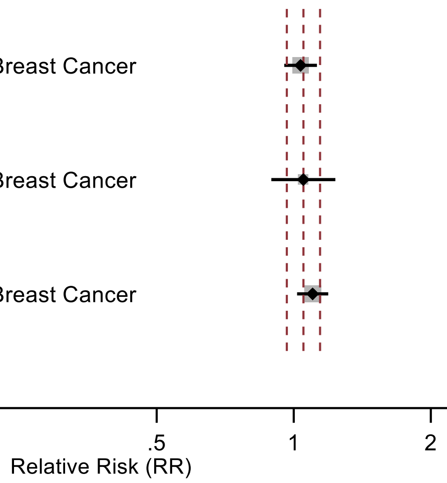

Figure legend: Forest plot shows the linear dose-response results (per 1 serving [355 mL] of sugar sweetened beverages/day) from the inverse variance DerSimonian-Laird random-effects model after exclusion of every single study at a time.

Abbreviations: CI, confidence interval; RR, relative risk.

Study abbreviations: EPIC, European Prospective Investigation into Cancer and nutrition; NHS, Nurses' Health Study; CPS II, Cancer Prevention Study II

Mullee, 2019[126], Malik, 2019[109], McCullough, 2022[113]

**Supplementary Figure 188. Linear dose-response meta-analysis of sugar sweetened beverages and colorectal cancer mortality, leave-one-out analysis.**

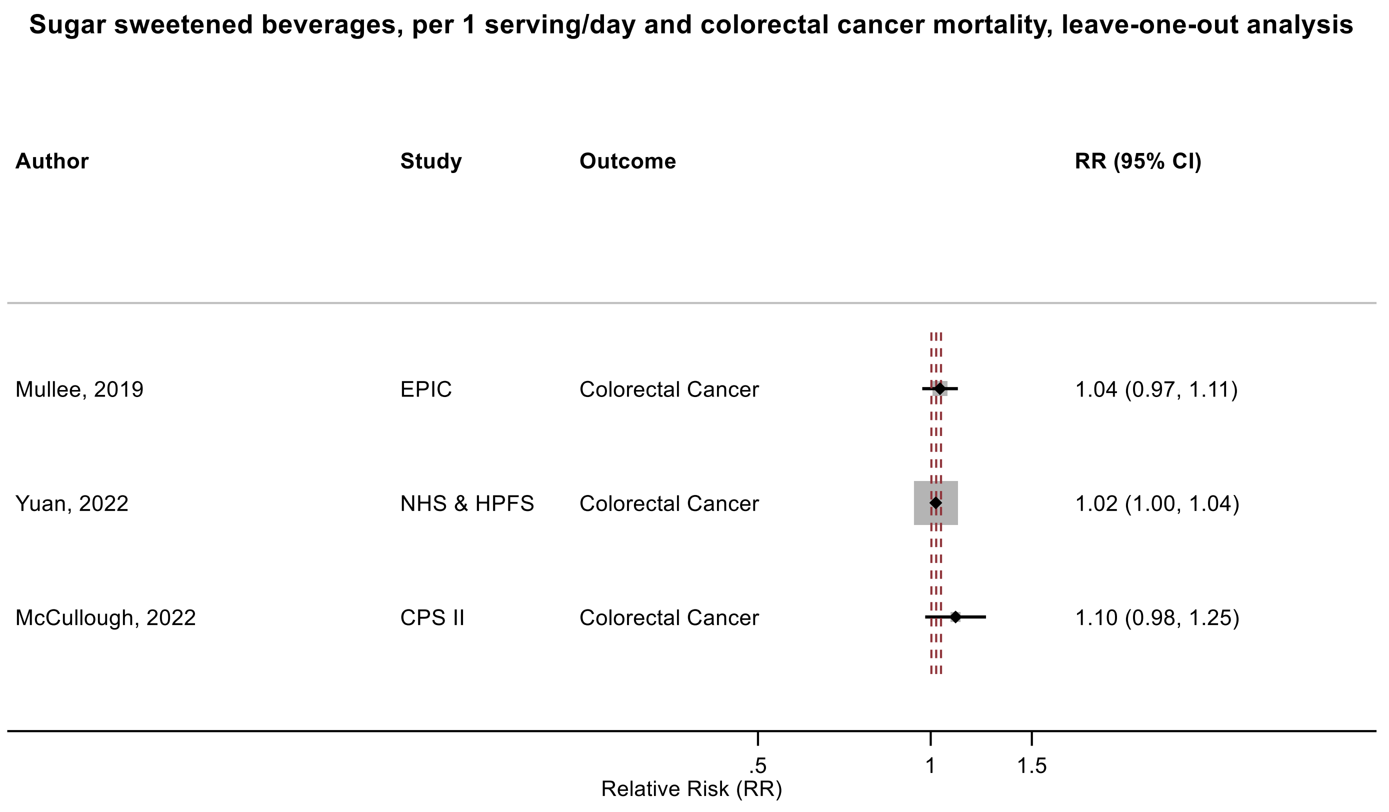

Figure legend: Forest plot shows the linear dose-response results (per 1 serving [355 mL] of sugar sweetened beverages /day) from the inverse variance DerSimonian-Laird random-effects model after exclusion of every single study at a time.

Abbreviations: CI, confidence interval; RR, relative risk.

Study abbreviations: EPIC, European Prospective Investigation into Cancer and nutrition; NHS & HPFS, Nurses' Health Study & Health Professionals Follow-up Study; CPS II, Cancer Prevention Study II

Mullee, 2019[126], Yuan, 2022[178], McCullough, 2022[113]

**Supplementary Figure 189. Linear dose-response meta-analysis of sugar sweetened beverages and prostate cancer mortality, leave-one-out analysis.**

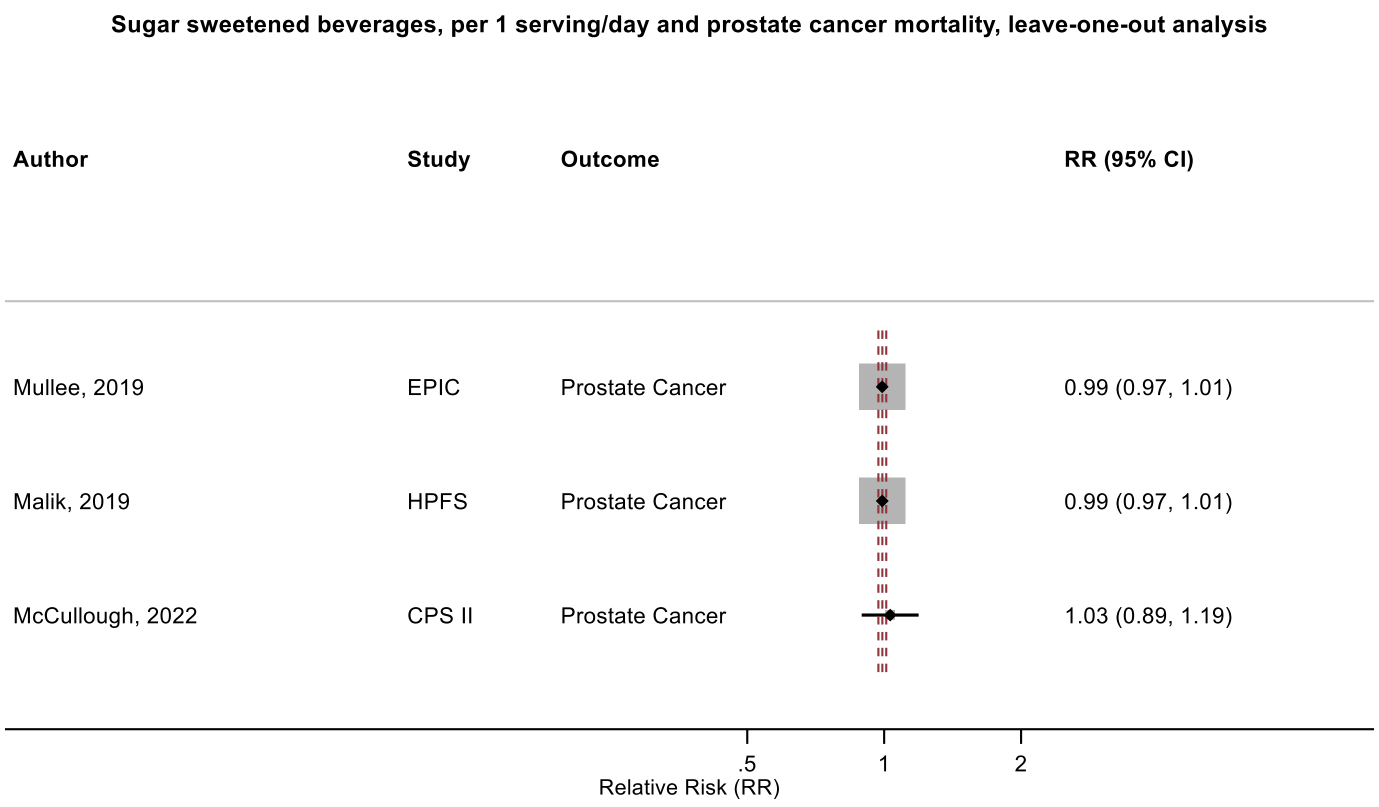

Figure legend: Forest plot shows the linear dose-response results (per 1 serving [355 mL] of sugar sweetened beverages/day) from the inverse variance DerSimonian-Laird random-effects model after exclusion of every single study at a time.

Abbreviations: CI, confidence interval; RR, relative risk.

Study abbreviations: EPIC, European Prospective Investigation into Cancer and nutrition; HPFS, Health Professionals Follow-up Study; CPS II, Cancer Prevention Study II

Mullee, 2019[126], Malik, 2019[109], McCullough, 2022[113]

**Supplementary Figure 190. Linear dose-response meta-analysis of sugar sweetened beverages and kidney cancer mortality, comparison of the models with and without adjustment for body mass index.**

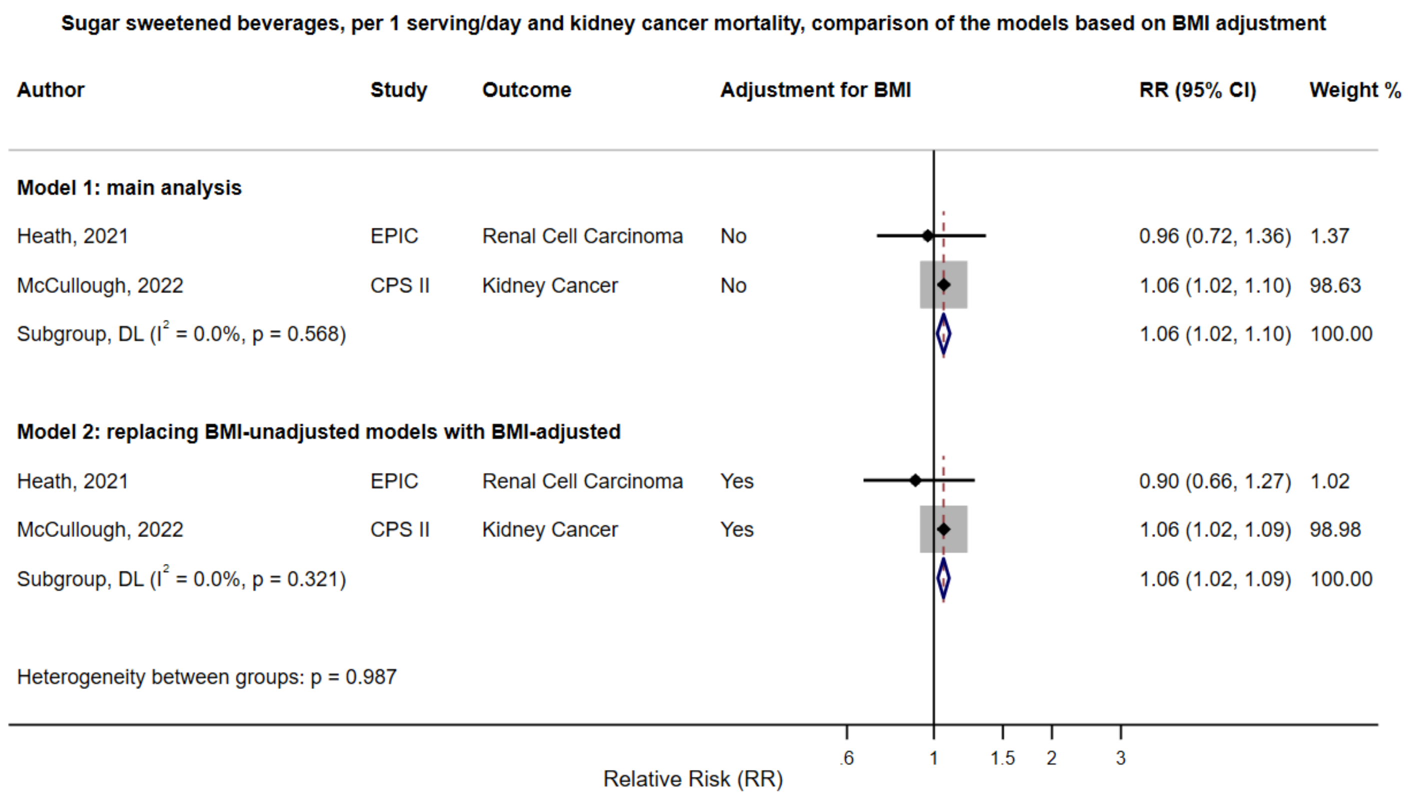

Figure legend: Forest plot shows the linear dose-response results (per 1 serving [355 mL] of sugar sweetened beverages/day) from the inverse variance DerSimonian-Laird random-effects model. The diamond represents the summary relative risk (RR) estimate, the diamond's width is the 95% confidence interval (CI). Each square and the horizontal line across the square represent the RR estimate of the individual study and its 95% CI.

Abbreviations: CI, confidence interval; RR, relative risk.

Study abbreviations: EPIC, European Prospective Investigation into Cancer and nutrition; CPS II, Cancer Prevention Study II

Heath, 2021[66], McCullough, 2022[113]

**Supplementary Figure 191. Linear dose-response meta-analysis of carbonated artificially sweetened beverages and leukaemia incidence.**

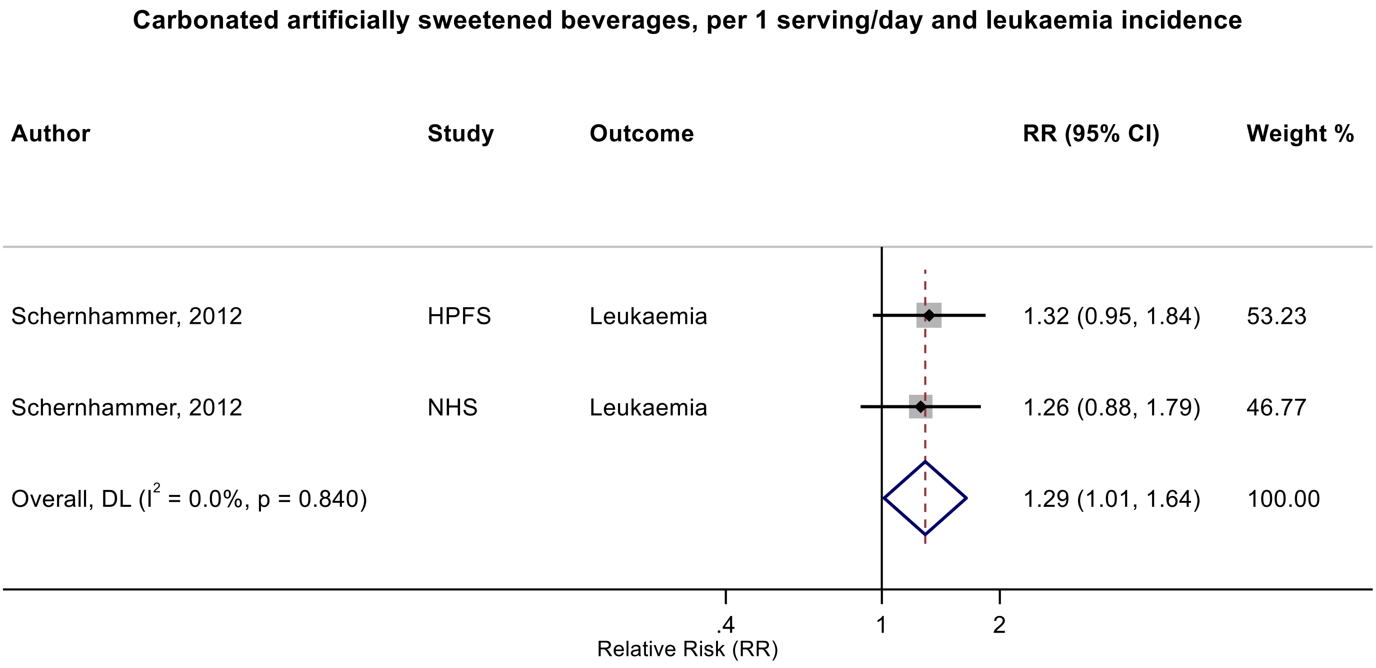

Figure legend: Forest plot shows the linear dose-response results (per 1 serving [355 mL] of carbonated artificially sweetened beverages/day) from the inverse variance DerSimonian-Laird random-effects model. The diamond represents the summary relative risk (RR) estimate, the diamond's width is the 95% confidence interval (CI). Each square and the horizontal line across the square represent the RR estimate of the individual study and its 95% CI.

Abbreviations: CI, confidence interval; RR, relative risk.

Study abbreviations: HPFS, Health Professionals Follow-up Study; NHS, Nurses' Health Study

Note: Both studies performed repeated dietary measurements (every 4 years).

Schernhammer, 2012[150]

**Supplementary Figure 192. Linear dose-response meta-analysis of artificially sweetened beverages and pancreatic cancer incidence and mortality combined.**

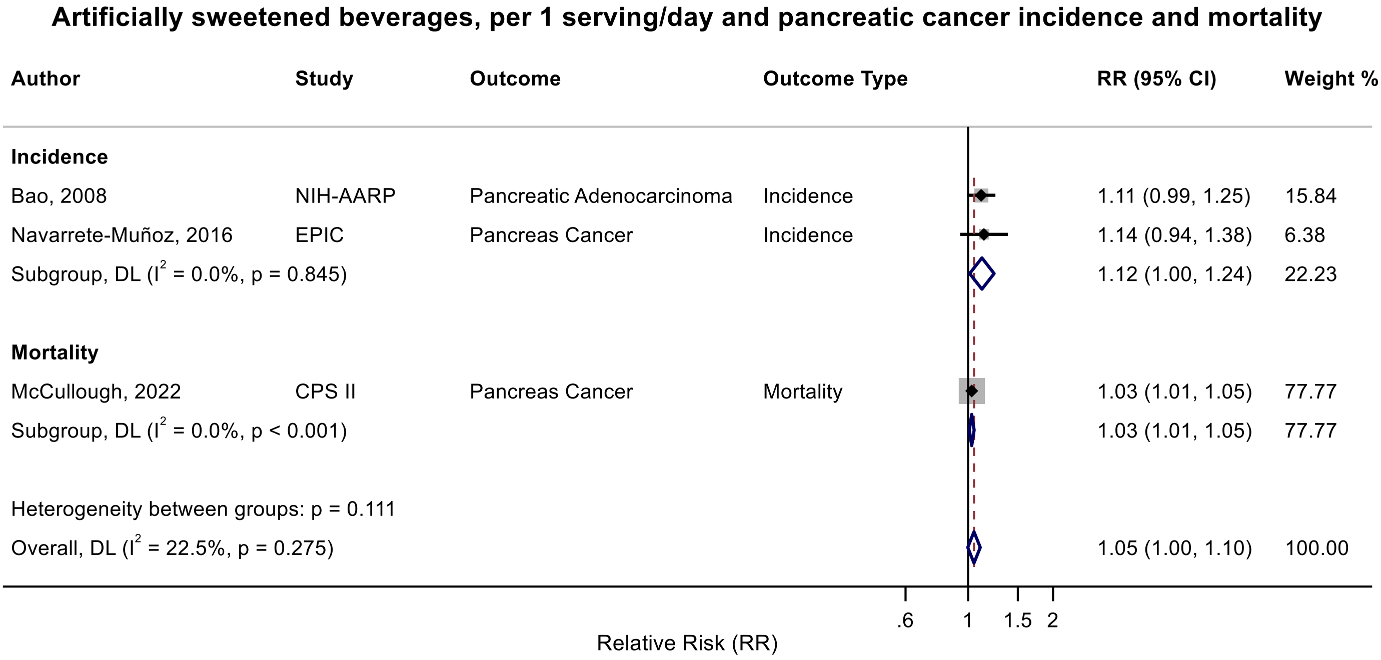

Figure legend: Forest plot shows the linear dose-response results (per 1 serving [355 mL] of artificially sweetened beverages/day) from the inverse variance DerSimonian-Laird random-effects model. The diamond represents the summary relative risk (RR) estimate, the diamond's width is the 95% confidence interval (CI). Each square and the horizontal line across the square represent the RR estimate of the individual study and its 95% CI.

Abbreviations: CI, confidence interval; RR, relative risk.

Study abbreviations: NIH-AARP, National Institute of Health + American Association of Retired Persons Diet and Health Study; EPIC, European Prospective Investigation into Cancer and nutrition; CPS II, Cancer Prevention Study II

Note: The pooled RR and 95%CIs with 3 decimal points are 1.12 (0.998 – 1.24) for incidence and 1.051 (0.997-1.109) for incidence and mortality combined.

Bao, 2008[39], Navarrete-Muñoz, 2016[127], McCullough, 2022[113]

**Supplementary Figure 193. Linear dose-response meta-analysis of carbonated artificially sweetened beverages and pancreatic cancer incidence.**

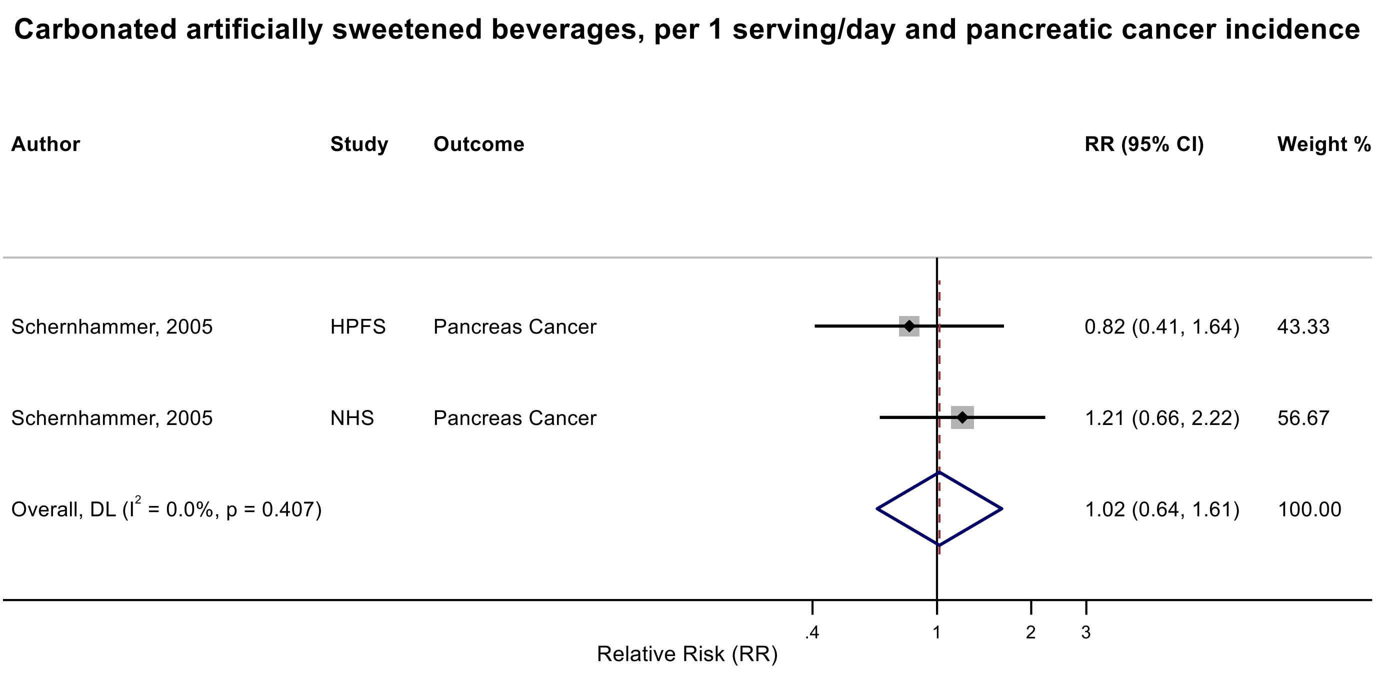

Figure legend: Forest plot shows the linear dose-response results (per 1 serving [355 mL] of carbonated artificially sweetened beverages/day) from the inverse variance DerSimonian-Laird random-effects model. The diamond represents the summary relative risk (RR) estimate, the diamond's width is the 95% confidence interval (CI). Each square and the horizontal line across the square represent the RR estimate of the individual study and its 95% CI.

Abbreviations: CI, confidence interval; RR, relative risk.

Study abbreviations: HPFS, Health Professionals Follow-up Study; NHS, Nurses' Health Study

Note: Both studies performed repeated dietary measurements (every 4 years).

Schernhammer, 2005[151]

**Supplementary Figure 194. Linear dose-response meta-analysis of artificially sweetened and carbonated artificially sweetened beverages and pancreatic cancer incidence and mortality.**

**Artificially sweetened and carbonated artificially sweetened beverages, per 1 serving/day and pancreatic cancer incidence and mortality**

| Author | Study | Outcome Type | RR (95% CI) | Weight % |
|---|---|---|---|---|
| **Artificially sweetened beverages** | | | | |
| Bao, 2008 | NIH-AARP | Incidence | 1.11 (0.99, 1.25) | 2.68 |
| Navarrete-Muñoz, 2016 | EPIC | Incidence | 1.14 (0.94, 1.38) | 0.95 |
| McCullough, 2022 | CPS II | Mortality | 1.03 (1.01, 1.05) | 96.20 |
| Subgroup, DL ($I^2$ = 22.5%, p = 0.275) | | | 1.05 (1.00, 1.10) | 99.83 |
| **Carbonated artificially sweetened beverages** | | | | |
| Schernhammer, 2005 | HPFS | Incidence | 0.82 (0.41, 1.64) | 0.07 |
| Schernhammer, 2005 | NHS | Incidence | 1.21 (0.66, 2.22) | 0.10 |
| Subgroup, DL ($I^2$ = 0.0%, p = 0.407) | | | 1.02 (0.64, 1.61) | 0.17 |
| Heterogeneity between groups: p = 0.899 | | | | |
| Overall, DL ($I^2$ = 0.0%, p = 0.513) | | | 1.03 (1.01, 1.05) | 100.00 |

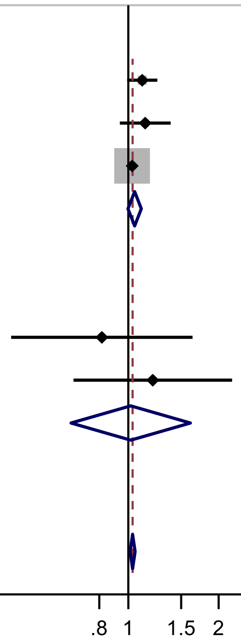

Relative Risk (RR)

Figure legend: Forest plot shows the linear dose-response results (per 1 serving [355 mL] of artificially sweetened beverages/day) from the inverse variance DerSimonian-Laird random-effects model. The diamond represents the summary relative risk (RR) estimate, the diamond's width is the 95% confidence interval (CI). Each square and the horizontal line across the square represent the RR estimate of the individual study and its 95% CI.

Abbreviations: CI, confidence interval; RR, relative risk.

Study abbreviations: NIH-AARP, National Institute of Health + American Association of Retired Persons Diet and Health Study; EPIC, European Prospective Investigation into Cancer and nutrition; CPS II, Cancer Prevention Study II, HPFS, Health Professionals Follow-up Study; NHS, Nurses' Health Study

Bao, 2008[39], Navarrete-Muñoz, 2016[127], McCullough, 2022[113], Schernhammer, 2005[151]

**Supplementary Figure 195. Linear dose-response meta-analysis of artificially sweetened beverages and colorectal cancer incidence.**

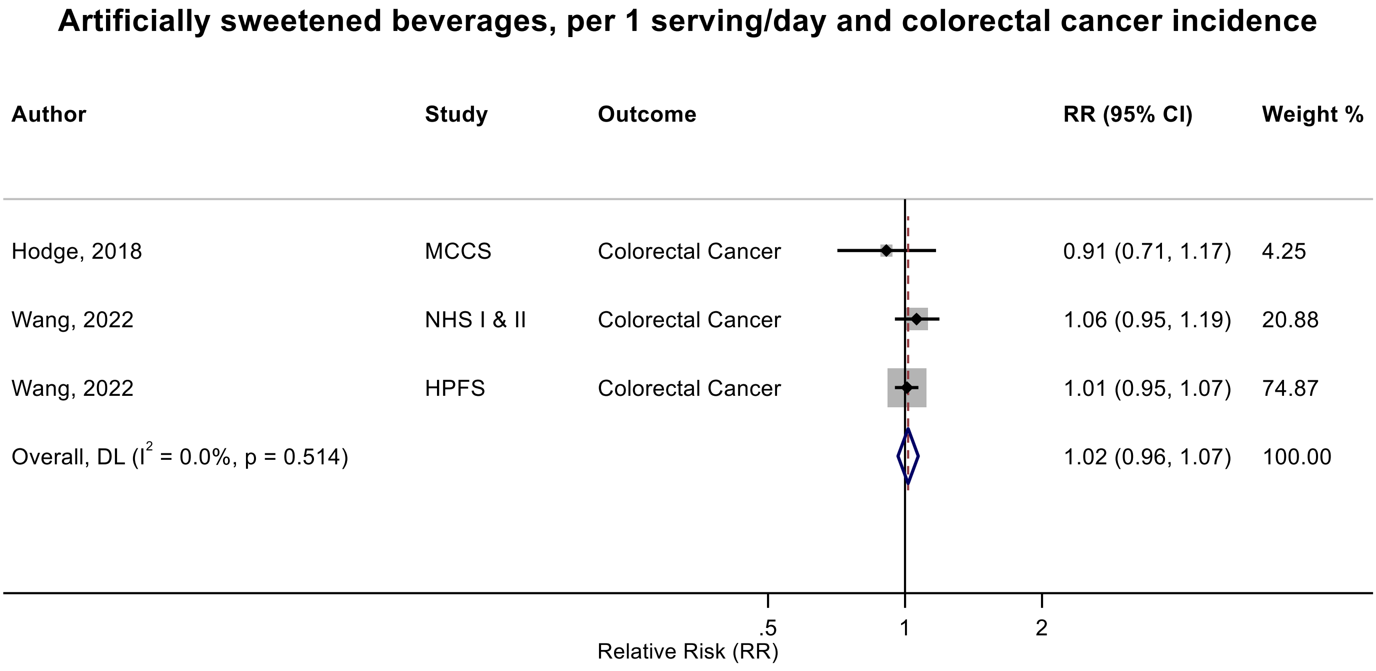

Figure legend: Forest plot shows the linear dose-response results (per 1 serving [355 mL] of artificially sweetened beverages/day) from the inverse variance DerSimonian-Laird random-effects model. The diamond represents the summary relative risk (RR) estimate, the diamond's width is the 95% confidence interval (CI). Each square and the horizontal line across the square represent the RR estimate of the individual study and its 95% CI.

Abbreviations: CI, confidence interval; RR, relative risk.

Study abbreviations: MCCS, Melbourne Collaborative Cohort Study; NHS I and II, Nurses' Health Study I and II; HPFS, Health Professionals Follow-up Study

Note: There was one French Cohort study (NutriNet-Santé)[44] that could not be included in the dose-response meta-analysis because of implausible effect size and reported no association between ASBs and colorectal cancer incidence (RR per 10 mL/day: 1.06, 95%CI: 0.77-1.48). There was another study (UK Biobank) that could not be included in the dose-response meta-analysis because of insufficient data format[78] and reported no association with colorectal cancer incidence per one SD increase in ASB intake (RR: 0.99, 95%CI: 0.94-1.05).

Hodge, 2018[70], Wang, 2022[168]

**Supplementary Figure 196. Linear dose-response meta-analysis of artificially sweetened beverages and liver cancer incidence.**

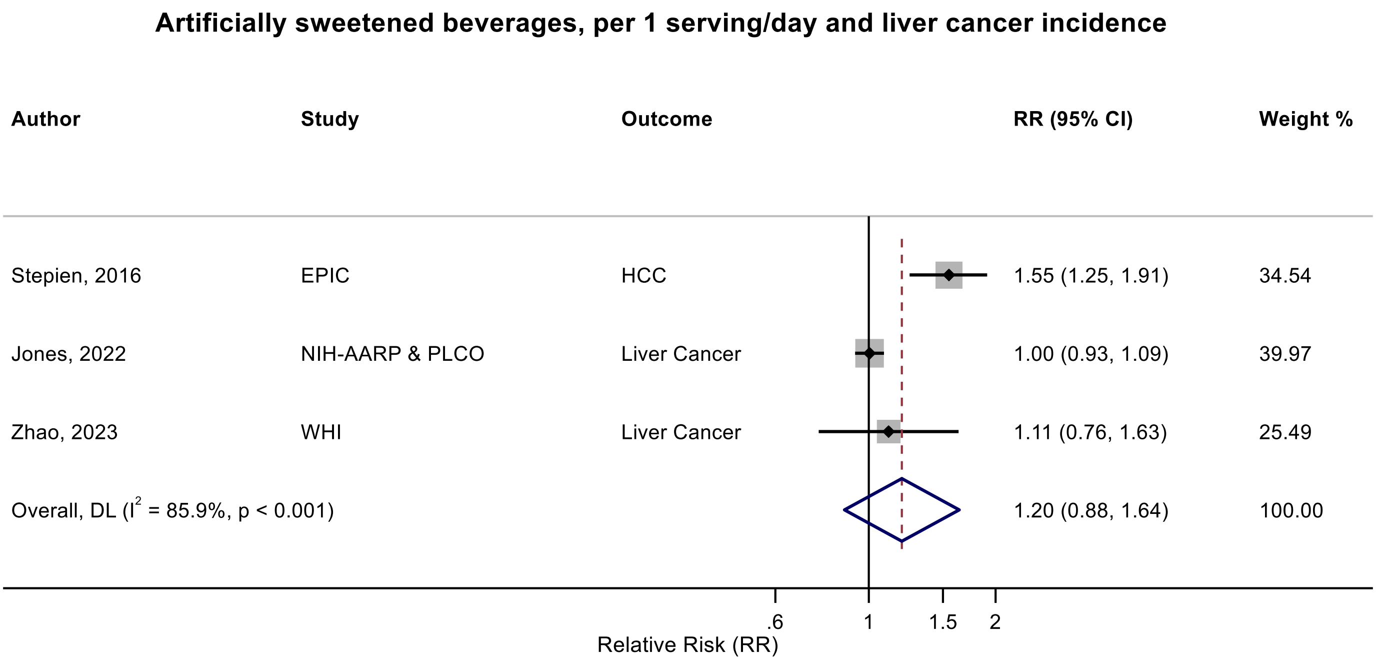

Figure legend: Forest plot shows the linear dose-response results (per 1 serving [355 mL] of artificially sweetened beverages/day) from the inverse variance DerSimonian-Laird random-effects model. The diamond represents the summary relative risk (RR) estimate, the diamond's width is the 95% confidence interval (CI). Each square and the horizontal line across the square represent the RR estimate of the individual study and its 95% CI.

Abbreviations: CI, confidence interval; RR, relative risk.

Study abbreviations: EPIC, European Prospective Investigation into Cancer and nutrition; NIH-AARP & PLCO, National Institute of Health + American Association of Retired Persons Diet and Health Study & Prostate, Lung, Colorectal and Ovarian Cancer Screening trial; WHI, Women's Health Initiative Study

Note. There was an additional study (Southern Community Cohort)[187] that could not be included in the dose-response meta-analysis because of insufficient data format and reported no association between ASB intake, as a component of ultraprocessed foods, and liver cancer incidence ($RR_{\geq median\ vs\ non\text{-}consumers}$ : 0.83, 95%CI: 0.51-1.35).

Stepien, 2016[159], Jones, 2022[79], Zhao, 2023[186]

**Supplementary Figure 197. Linear dose-response meta-analysis of artificially sweetened beverages and liver cancer incidence and mortality combined.**

##### Artificially sweetened beverages, per 1 serving/day and liver cancer incidence/mortality

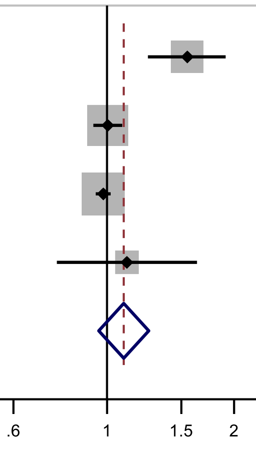

| Author | Study | Outcome | Outcome Type | RR (95% CI) | Weight % |
|---|---|---|---|---|---|
| Stepien, 2016 | EPIC | HCC | Incidence | 1.55 (1.25, 1.91) | 19.83 |
| Jones, 2022 | NIH-AARP & PLCO | Liver Cancer | Incidence | 1.00 (0.93, 1.09) | 33.79 |
| McCullough, 2022 | CPS II | Liver Cancer | Mortality | 0.98 (0.94, 1.02) | 36.84 |
| Zhao, 2023 | WHI | Liver Cancer | Incidence | 1.11 (0.76, 1.63) | 9.53 |
| Overall, DL ($I^2$ = 83.0%, p < 0.001) | | | | 1.10 (0.96, 1.26) | 100.00 |

Figure legend: Forest plot shows the linear dose-response results (per 1 serving [355 mL] of artificially sweetened beverages/day) from the inverse variance DerSimonian-Laird random-effects model. The diamond represents the summary relative risk (RR) estimate, the diamond's width is the 95% confidence interval (CI). Each square and the horizontal line across the square represent the RR estimate of the individual study and its 95% CI.

Abbreviations: CI, confidence interval; RR, relative risk.

Study abbreviations: EPIC, European Prospective Investigation into Cancer and nutrition; NIH-AARP & PLCO, National Institute of Health + American Association of Retired Persons Diet and Health Study & Prostate, Lung, Colorectal and Ovarian Cancer Screening trial; WHI, Women’s Health Initiative Study; CPS II, Cancer Prevention Study II

Stepien, 2016[159], Jones, 2022[79], McCullough, 2022[113], Zhao, 2023[186]

**Supplementary Figure 198. Linear dose-response meta-analysis of artificially sweetened beverages components and liver cancer incidence.**

**Artificially sweetened beverages components, per 1 serving/day and liver cancer incidence**

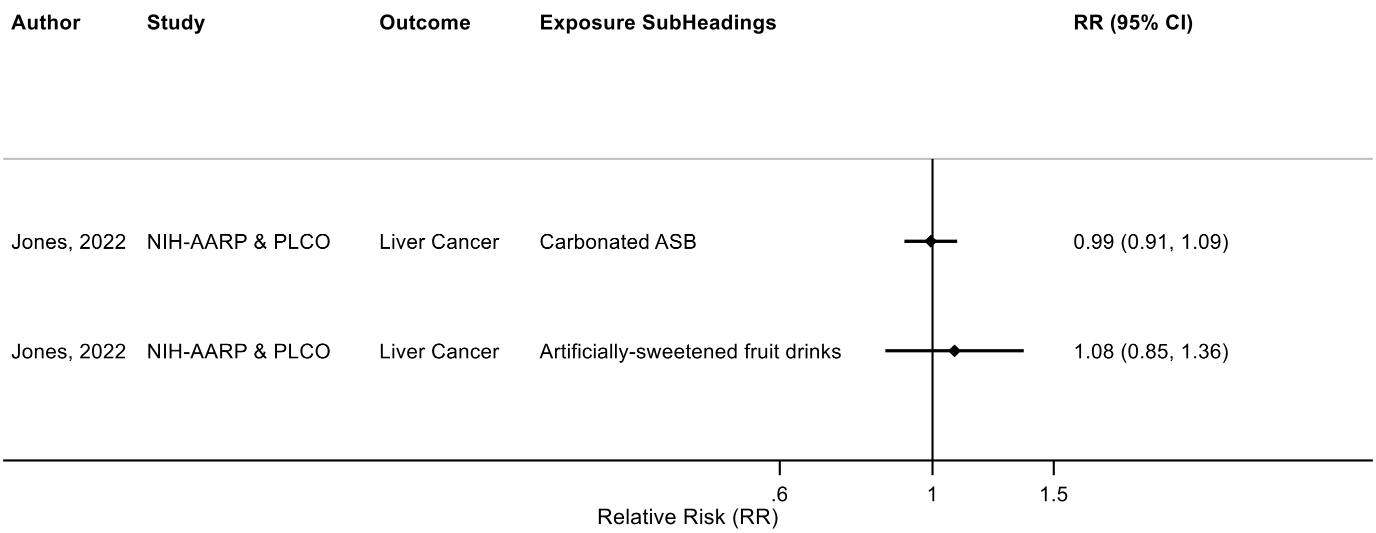

Figure legend: Forest plot shows the linear dose-response results (per 1 serving [355 mL] of artificially sweetened beverages components/day) from the inverse variance DerSimonian-Laird random-effects model. The diamond represents the summary relative risk (RR) estimate, the diamond's width is the 95% confidence interval (CI). Each square and the horizontal line across the square represent the RR estimate of the individual study and its 95% CI.

Abbreviations: CI, confidence interval; RR, relative risk.

Study abbreviations: NIH-AARP & PLCO, National Institute of Health + American Association of Retired Persons Diet and Health Study & Prostate, Lung, Colorectal and Ovarian Cancer Screening trial

Note: There was an additional study (Southern Community Cohort)[187] that could not be included in the dose-response meta-analysis because of insufficient data format and reported no association between carbonated artificially sweetened beverages and liver cancer ($RR_{\geq median\ vs\ non\text{-}consumers}$: 0.83, 95%CI: 0.51-1.35).

Jones, 2022[79]

**Supplementary Figure 199. Linear dose-response meta-analysis of artificially sweetened cola and pancreatic cancer incidence.**

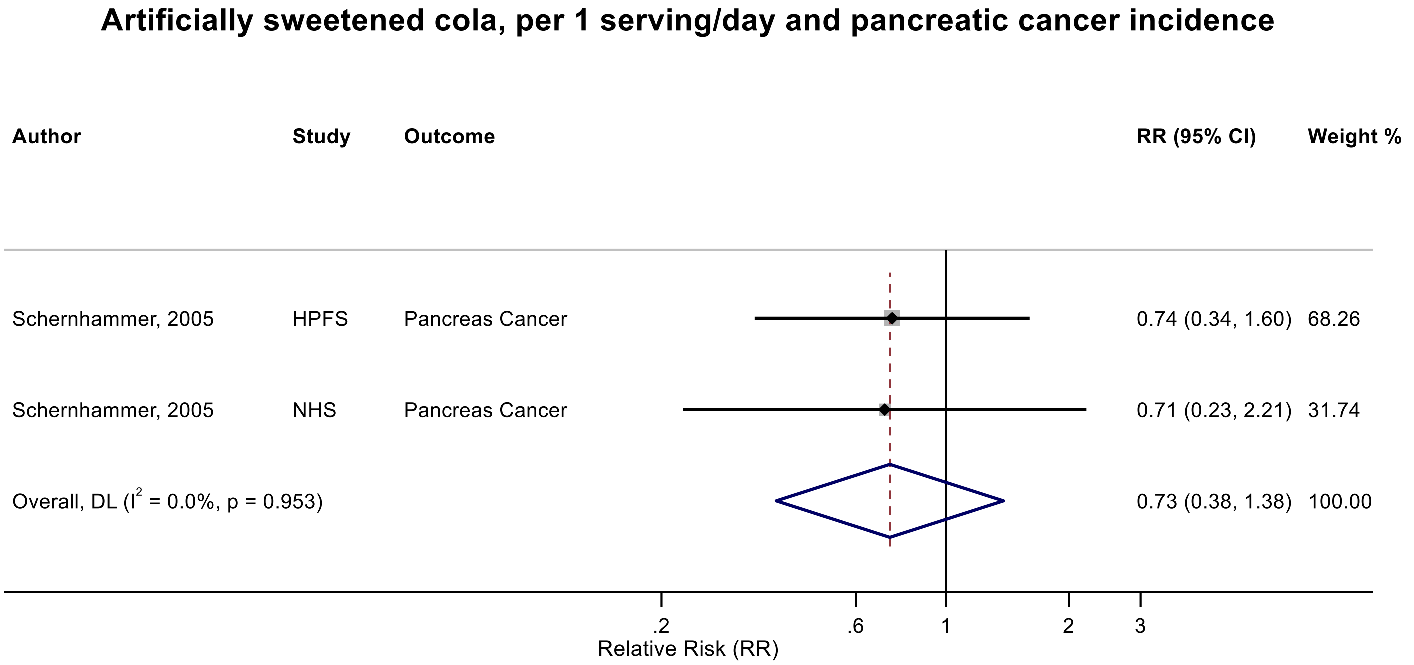

Figure legend: Forest plot shows the linear dose-response results (per 1 serving [355 mL] of artificially sweetened cola/day) from the inverse variance DerSimonian-Laird random-effects model. The diamond represents the summary relative risk (RR) estimate, the diamond's width is the 95% confidence interval (CI). Each square and the horizontal line across the square represent the RR estimate of the individual study and its 95% CI.

Abbreviations: CI, confidence interval; RR, relative risk.

Study abbreviations: HPFS, Health Professionals Follow-up Study; NHS, Nurses' Health Study

Note: Both studies performed repeated dietary measurements (every 4 years).

Schernhammer, 2005[151]

#### Supplementary Figure 200. Linear dose-response meta-analysis of artificially sweetened beverages and breast cancer incidence.

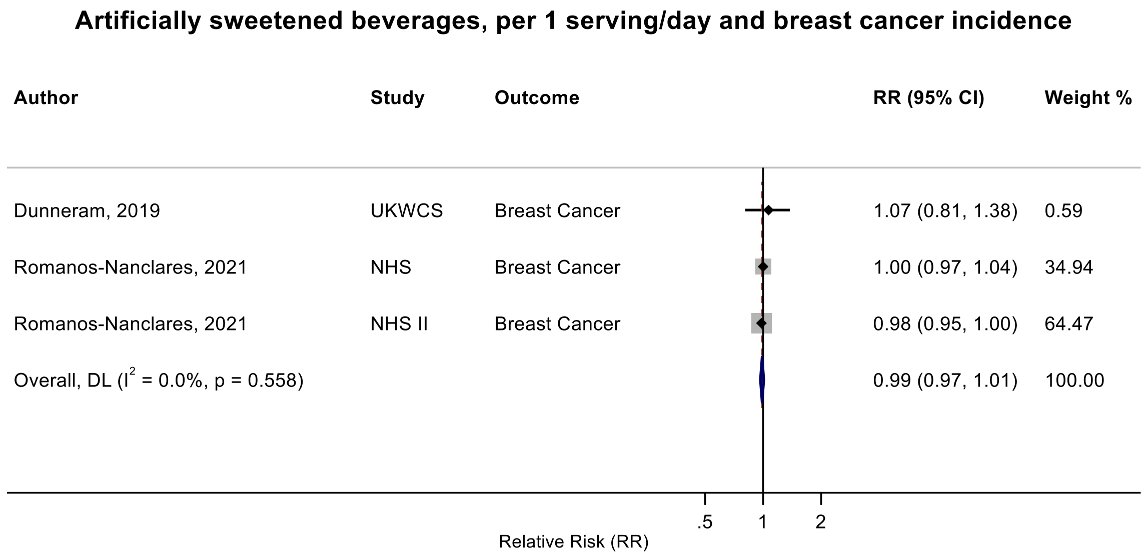

Figure legend: Forest plot shows the linear dose-response results (per 1 serving [355 mL] of artificially sweetened beverages/day) from the inverse variance DerSimonian-Laird random-effects model. The diamond represents the summary relative risk (RR) estimate, the diamond's width is the 95% confidence interval (CI). Each square and the horizontal line across the square represent the RR estimate of the individual study and its 95% CI.

Abbreviations: CI, confidence interval; RR, relative risk.

Study abbreviations: UKWCS, United Kingdom Women's Cohort Study; NHS, Nurses' Health Study; NHS II, Nurses' Health Study II

Note: There was one French Cohort study (NutriNet-Santé)[44] that that could not be included in the dose-response meta-analysis because of implausible effect size and reported limited evidence of an association ($RR_{\text{per 10 mL/day}}$: 0.97, 95%CI: 0.84-1.12).

Dunneram, 2019[49], Romanos-Nanclares, 2021[145]

**Supplementary Figure 201. Linear dose-response meta-analysis of artificially sweetened beverages and breast cancer incidence by menopausal status.**

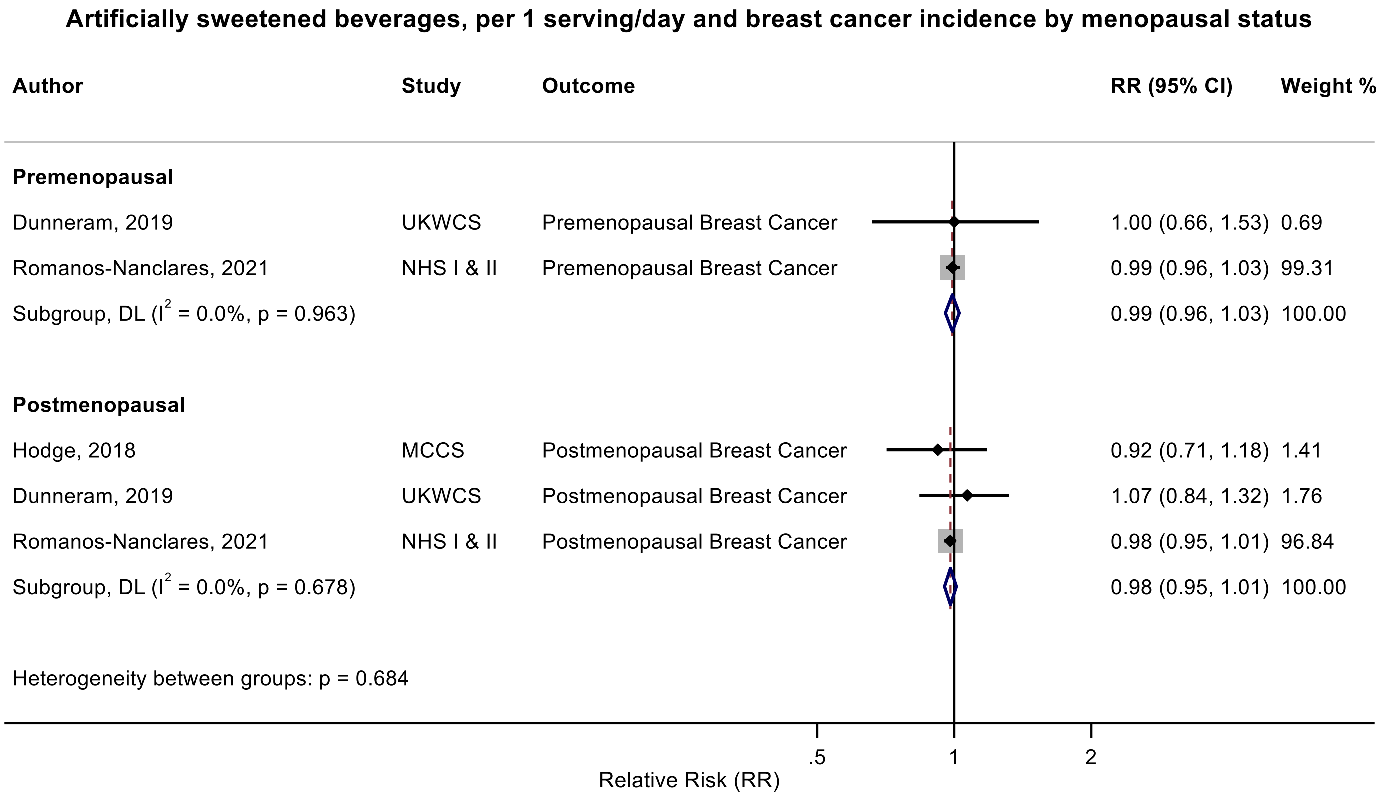

Figure legend: Forest plot shows the linear dose-response results (per 1 serving [355 mL] of artificially sweetened beverages/day) from the inverse variance DerSimonian-Laird random-effects model. The diamond represents the summary relative risk (RR) estimate, the diamond's width is the 95% confidence interval (CI). Each square and the horizontal line across the square represent the RR estimate of the individual study and its 95% CI.

Abbreviations: CI, confidence interval; RR, relative risk.

Study abbreviations: UKWCS, United Kingdom Women's Cohort Study; NHS I & II, Nurses' Health Study I & II; MCCS, Melbourne Collaborative Cohort Study

Note: There was one French Cohort study (NutriNet-Santé)[44] that that could not be included in the dose-response meta-analysis because of implausible effect size and reported limited evidence of an association with postmenopausal ($RR_{per\ 10\ mL/day}$: 0.99, 95%CI: 0.78-1.25) and premenopausal ($RR_{per\ 10\ mL/day}$: 0.95, 95%CI: 0.78-1.16) breast cancer incidence.

Dunneram, 2019[49], Romanos-Nanclares, 2021[145], Hodge, 2018[70]

**Supplementary Figure 202. Linear dose-response meta-analysis of artificially sweetened beverages and incidence of breast cancer subtypes.**

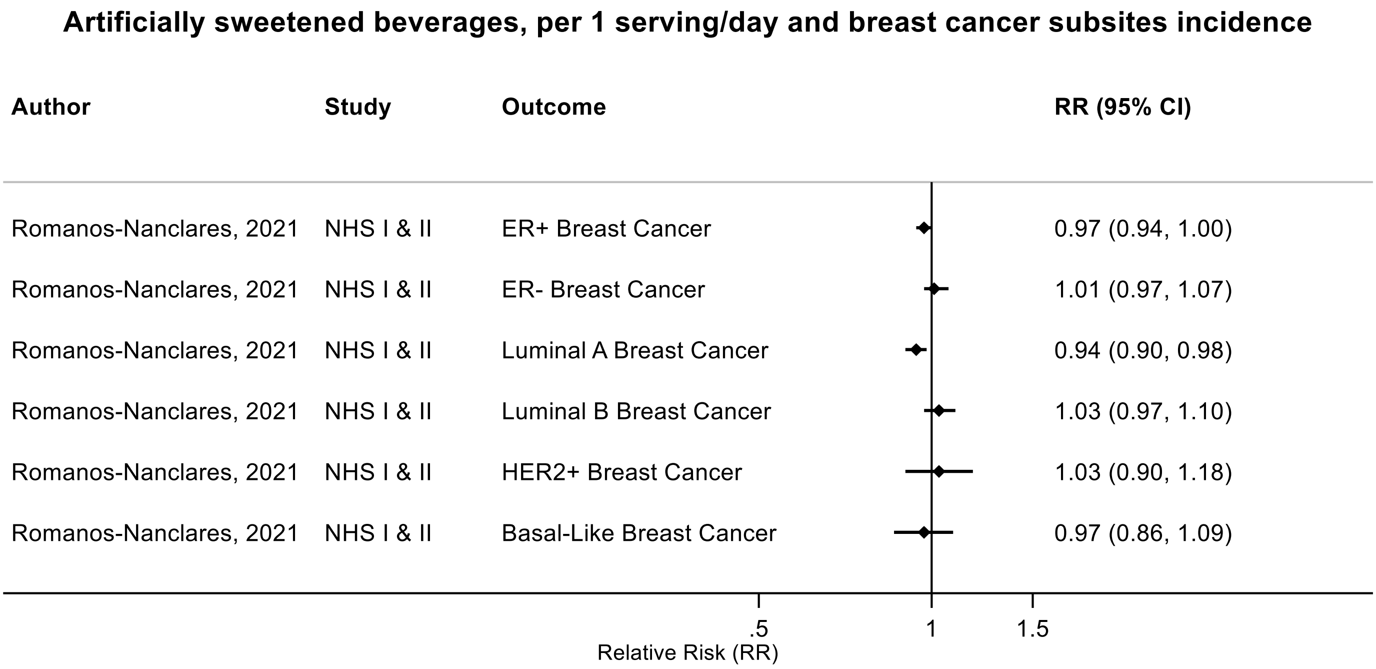

Figure legend: Forest plot shows the linear dose-response results (per 1 serving [355 mL] of artificially sweetened beverages/day) from the inverse variance DerSimonian-Laird random-effects model. The diamond represents the summary relative risk (RR) estimate, the diamond's width is the 95% confidence interval (CI). Each square and the horizontal line across the square represent the RR estimate of the individual study and its 95% CI.

Abbreviations: CI, confidence interval; RR, relative risk.

Study abbreviations: NHS I and II, Nurses' Health Study I and II

Romanos-Nanclares, 2021[145]

**Supplementary Figure 203. Linear dose-response meta-analysis of artificially sweetened beverages and endometrial cancer incidence.**

##### Artificially sweetened beverages, per 1 serving/day and endometrial cancer incidence

| Author | Study | Outcome | RR (95% CI) | Weight % |
|---|---|---|---|---|
| Inoue-Choi, 2013 | IWHS | Endometrial Cancer | 0.86 (0.54, 1.38) | 50.97 |
| Hodge, 2018 | MCCS | Endometrial Cancer | 0.92 (0.52, 1.65) | 33.65 |
| Dunneram, 2019 | UKWCS | Endometrial Cancer | 1.00 (0.42, 2.34) | 15.38 |
| Overall, DL ($I^2$ = 0.0%, p = 0.951) | | | 0.90 (0.64, 1.26) | 100.00 |

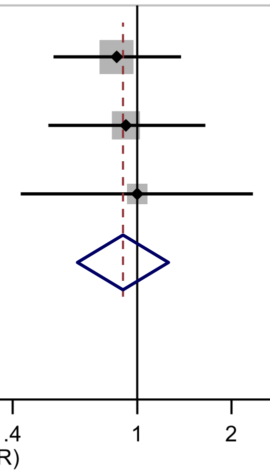

Figure legend: Forest plot shows the linear dose-response results (per 1 serving [355 mL] of artificially sweetened beverages/day) from the inverse variance DerSimonian-Laird random-effects model. The diamond represents the summary relative risk (RR) estimate, the diamond's width is the 95% confidence interval (CI). Each square and the horizontal line across the square represent the RR estimate of the individual study and its 95% CI.

Abbreviations: CI, confidence interval; RR, relative risk.

Study abbreviations: IWHS, The Iowa Women’s Health Study; MCCS, Melbourne Collaborative Cohort Study; UKWCS, United Kingdom Women's Cohort Study

Inoue-Choi, 2013[74], Hodge, 2018[70], Dunneram, 2019[49]

**Supplementary Figure 204. Linear dose-response meta-analysis of artificially sweetened beverages and postmenopausal endometrial cancer incidence.**

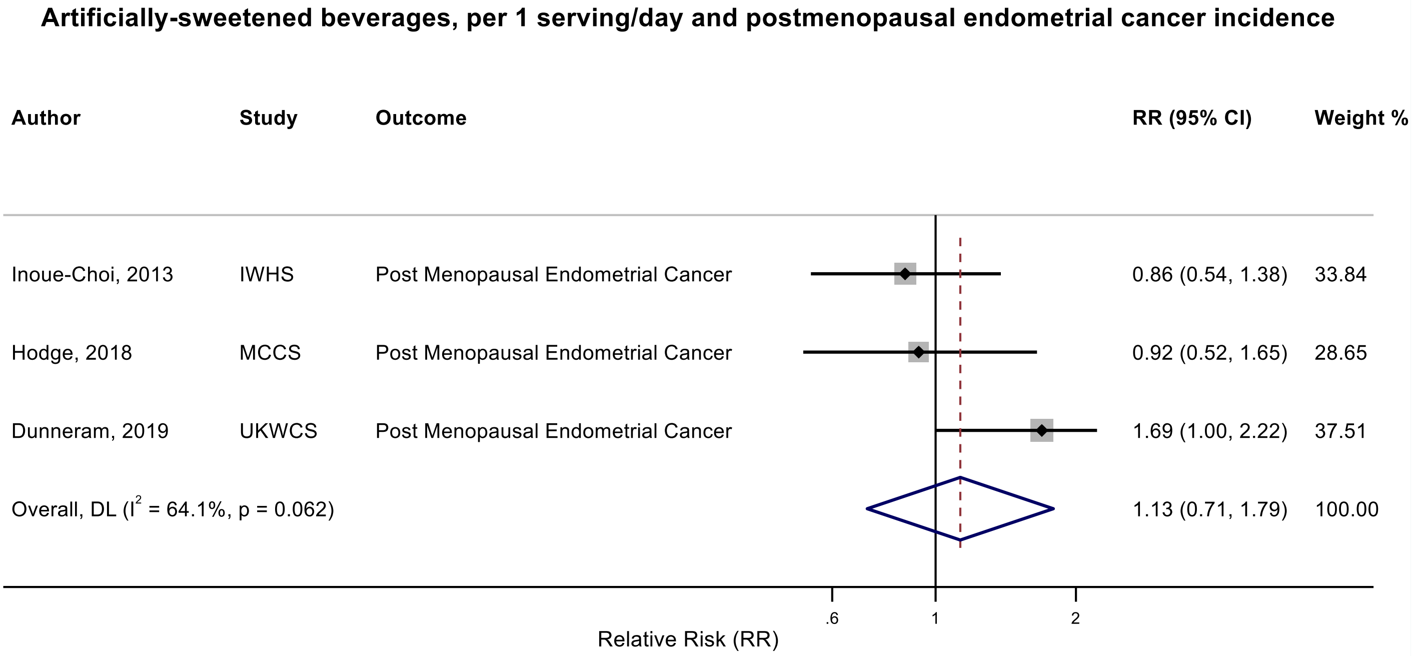

Figure legend: Forest plot shows the linear dose-response results (per 1 serving [355 mL] of artificially sweetened beverages/day) from the inverse variance DerSimonian-Laird random-effects model. The diamond represents the summary relative risk (RR) estimate, the diamond's width is the 95% confidence interval (CI). Each square and the horizontal line across the square represent the RR estimate of the individual study and its 95% CI.

Abbreviations: CI, confidence interval; RR, relative risk.

Study abbreviations: IWHS, The Iowa Women’s Health Study; MCCS, Melbourne Collaborative Cohort Study; UKWCS, United Kingdom Women's Cohort Study

Inoue-Choi, 2013[74], Hodge, 2018[70], Dunneram, 2019[49]

**Supplementary Figure 205. Linear dose-response meta-analysis of artificially sweetened beverages and ovarian cancer incidence.**

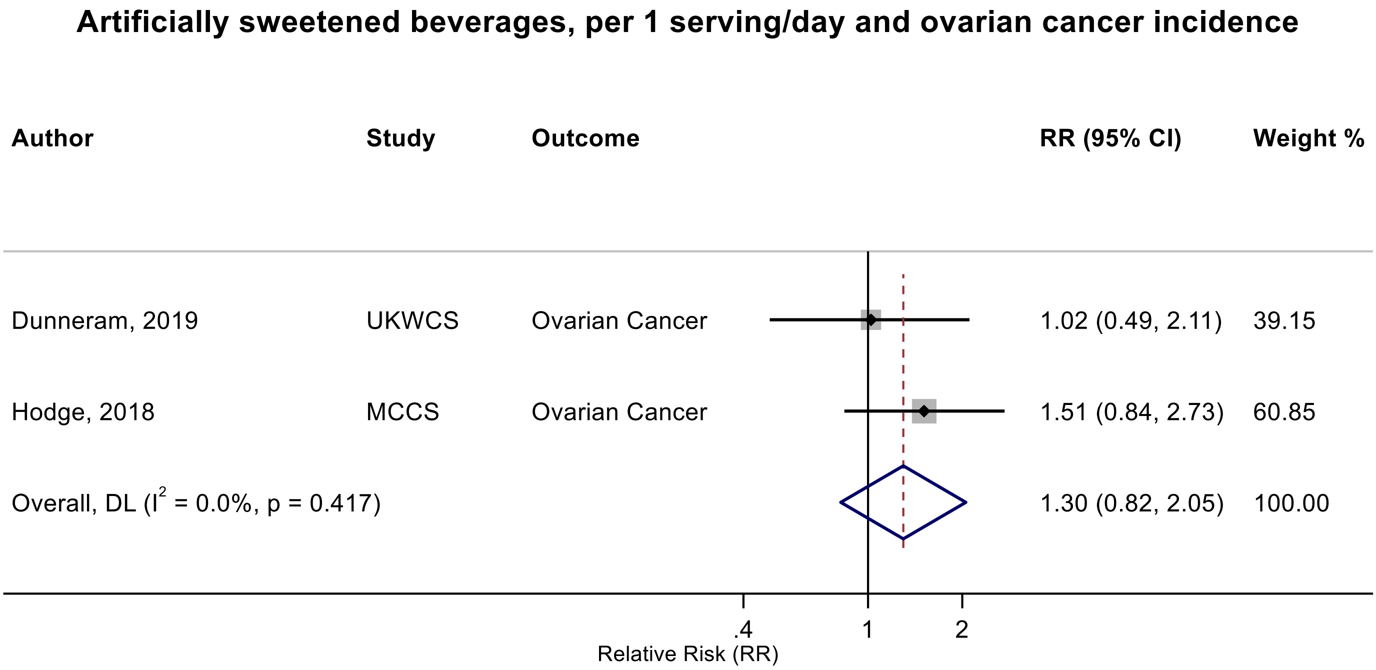

Figure legend: Forest plot shows the linear dose-response results (per 1 serving [355 mL] of artificially sweetened beverages/day) from the inverse variance DerSimonian-Laird random-effects model. The diamond represents the summary relative risk (RR) estimate, the diamond's width is the 95% confidence interval (CI). Each square and the horizontal line across the square represent the RR estimate of the individual study and its 95% CI.

Abbreviations: CI, confidence interval; RR, relative risk.

Study abbreviations: UKWCS, United Kingdom Women's Cohort Study; MCCS, Melbourne Collaborative Cohort Study

Dunneram, 2019[49], Hodge, 2018[70]

**Supplementary Figure 206. Linear dose-response meta-analysis of artificially sweetened beverages and kidney cancer incidence.**

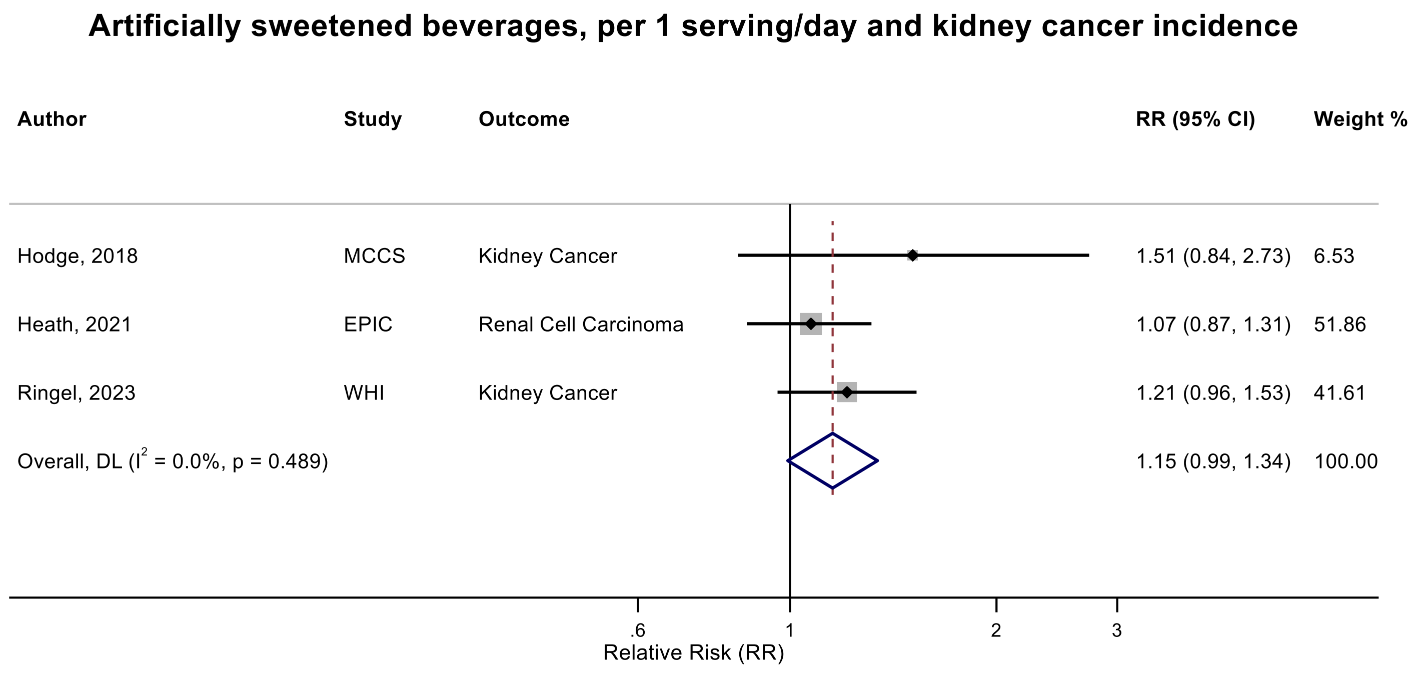

Figure legend: Forest plot shows the linear dose-response results (per 1 serving [355 mL] of artificially sweetened beverages/day) from the inverse variance DerSimonian-Laird random-effects model. The diamond represents the summary relative risk (RR) estimate, the diamond's width is the 95% confidence interval (CI). Each square and the horizontal line across the square represent the RR estimate of the individual study and its 95% CI.

Abbreviations: CI, confidence interval; RR, relative risk.

Study abbreviations: MCCS, Melbourne Collaborative Cohort Study; EPIC, European Prospective Investigation into Cancer and nutrition; WHI, Women's Health Initiative Study

Hodge, 2018[70], Heath, 2021[66], Ringel, 2023[144]

**Supplementary Figure 207. Linear dose-response meta-analysis of artificially sweetened and carbonated artificially sweetened beverages and kidney cancer incidence.**

##### Artificially sweetened and carbonated artificially sweetened beverages and kidney cancer incidence

| Author | Study | Outcome | Outcome Type | RR (95% CI) | Weight % |
|---|---|---|---|---|---|
| **Artificially sweetened beverages** | | | | | |
| Hodge, 2018 | MCCS | Kidney Cancer | Incidence | 1.51 (0.84, 2.73) | 4.62 |
| Heath, 2021 | EPIC | Renal Cell Cancer | Incidence | 1.07 (0.87, 1.31) | 30.44 |
| Ringel, 2023 | WHI | Kidney Cancer | Incidence | 1.21 (0.96, 1.53) | 24.74 |
| Subgroup, DL ($I^2$ = 0.0%, p = 0.476) | | | | 1.15 (0.99, 1.34) | 59.79 |
| **Carbonated artificially sweetened beverages** | | | | | |
| Lee, 2006 | NHS & HPFS | Renal Cell Cancer | Incidence | 0.97 (0.82, 1.15) | 40.21 |
| Subgroup, DL ($I^2$ = 0.0%, p < 0.001) | | | | 0.97 (0.82, 1.15) | 40.21 |
| Heterogeneity between groups: p = 0.138 | | | | | |
| Overall, DL ($I^2$ = 18.5%, p = 0.298) | | | | 1.08 (0.95, 1.23) | 100.00 |

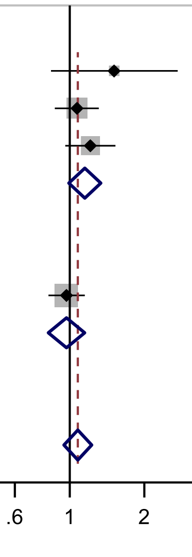

Figure legend: Forest plot shows the linear dose-response results (per 1 serving [355 mL] of artificially sweetened beverages/day) from the inverse variance DerSimonian-Laird random-effects model. The diamond represents the summary relative risk (RR) estimate, the diamond's width is the 95% confidence interval (CI). Each square and the horizontal line across the square represent the RR estimate of the individual study and its 95% CI.

Abbreviations: CI, confidence interval; RR, relative risk.

Study abbreviations: MCCS, Melbourne Collaborative Cohort Study; EPIC, European Prospective Investigation into Cancer and nutrition; WHI, Women's Health Initiative Study, HPFS, Health Professionals Follow-up Study; NHS, Nurses' Health Study

Hodge, 2018[70], Heath, 2021[66], Ringel, 2023[144], Lee, 2006[98]

**Supplementary Figure 208. Linear dose-response meta-analysis of carbonated artificially sweetened beverages and multiple myeloma incidence.**

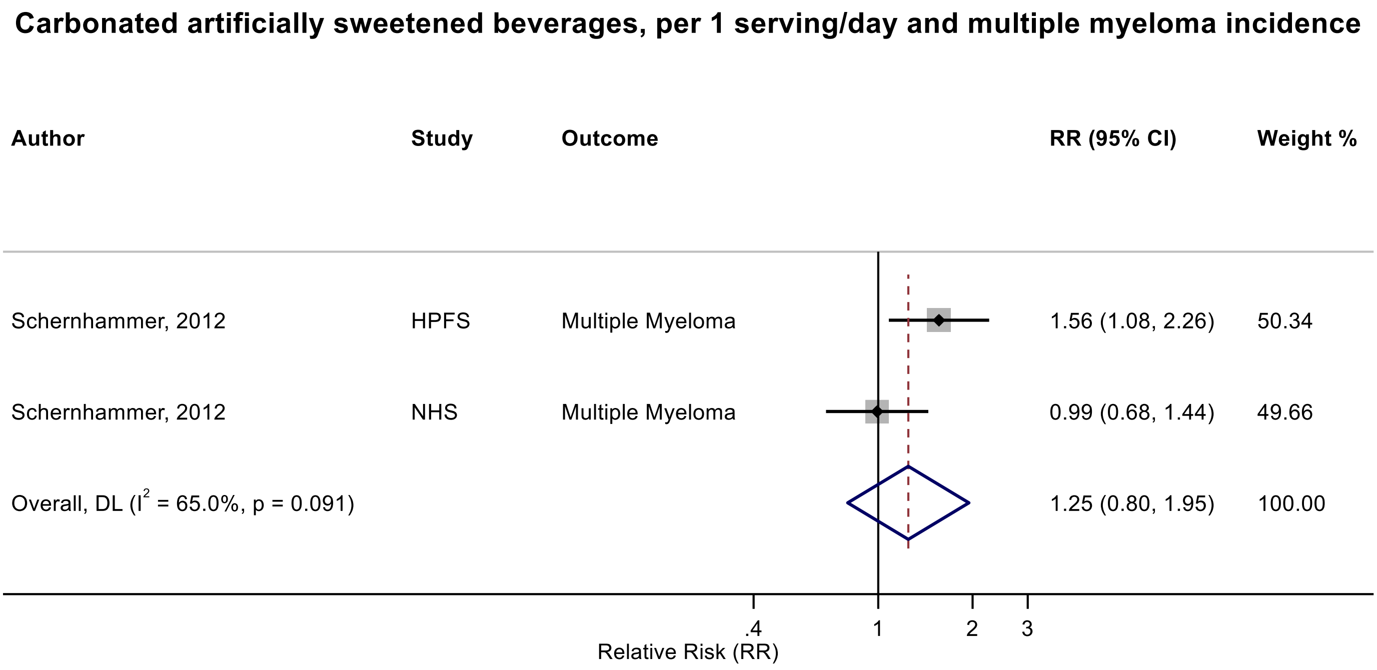

Figure legend: Forest plot shows the linear dose-response results (per 1 serving [355 mL] of carbonated artificially sweetened beverages/day) from the inverse variance DerSimonian-Laird random-effects model. The diamond represents the summary relative risk (RR) estimate, the diamond's width is the 95% confidence interval (CI). Each square and the horizontal line across the square represent the RR estimate of the individual study and its 95% CI.

Abbreviations: CI, confidence interval; RR, relative risk.

Study abbreviations: HPFS, Health Professionals Follow-up Study; NHS, Nurses' Health Study

Note: Both studies performed repeated dietary measurements (every 4 years).

Schernhammer, 2012[150]

**Supplementary Figure 209. Linear dose-response meta-analysis of carbonated artificially sweetened beverages and non-Hodgkin lymphoma incidence.**

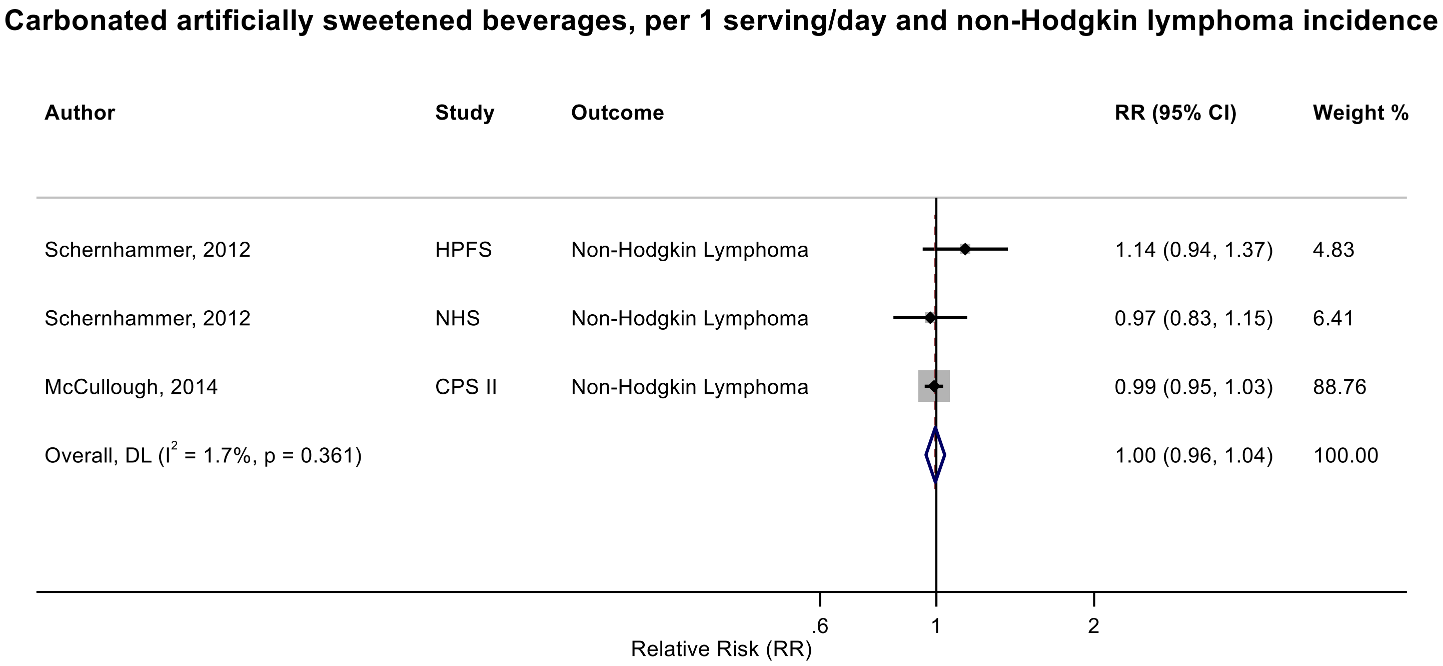

Figure legend: Forest plot shows the linear dose-response results (per 1 serving [355 mL] of carbonated artificially sweetened beverages/day) from the inverse variance DerSimonian-Laird random-effects model. The diamond represents the summary relative risk (RR) estimate, the diamond's width is the 95% confidence interval (CI). Each square and the horizontal line across the square represent the RR estimate of the individual study and its 95% CI.

Abbreviations: CI, confidence interval; RR, relative risk.

Study abbreviations: HPFS, Health Professionals Follow-up Study; NHS, Nurses' Health Study; CPS II, Cancer Prevention Study II

Note: All studies performed repeated dietary measurements (2 times in CPS II and every 4 years in NHS and HPFS).

Schernhammer, 2012[150], McCullough, 2014[114]

**Supplementary Figure 210. Linear dose-response meta-analysis of artificially sweetened beverages and colorectal cancer incidence by geographical region.**

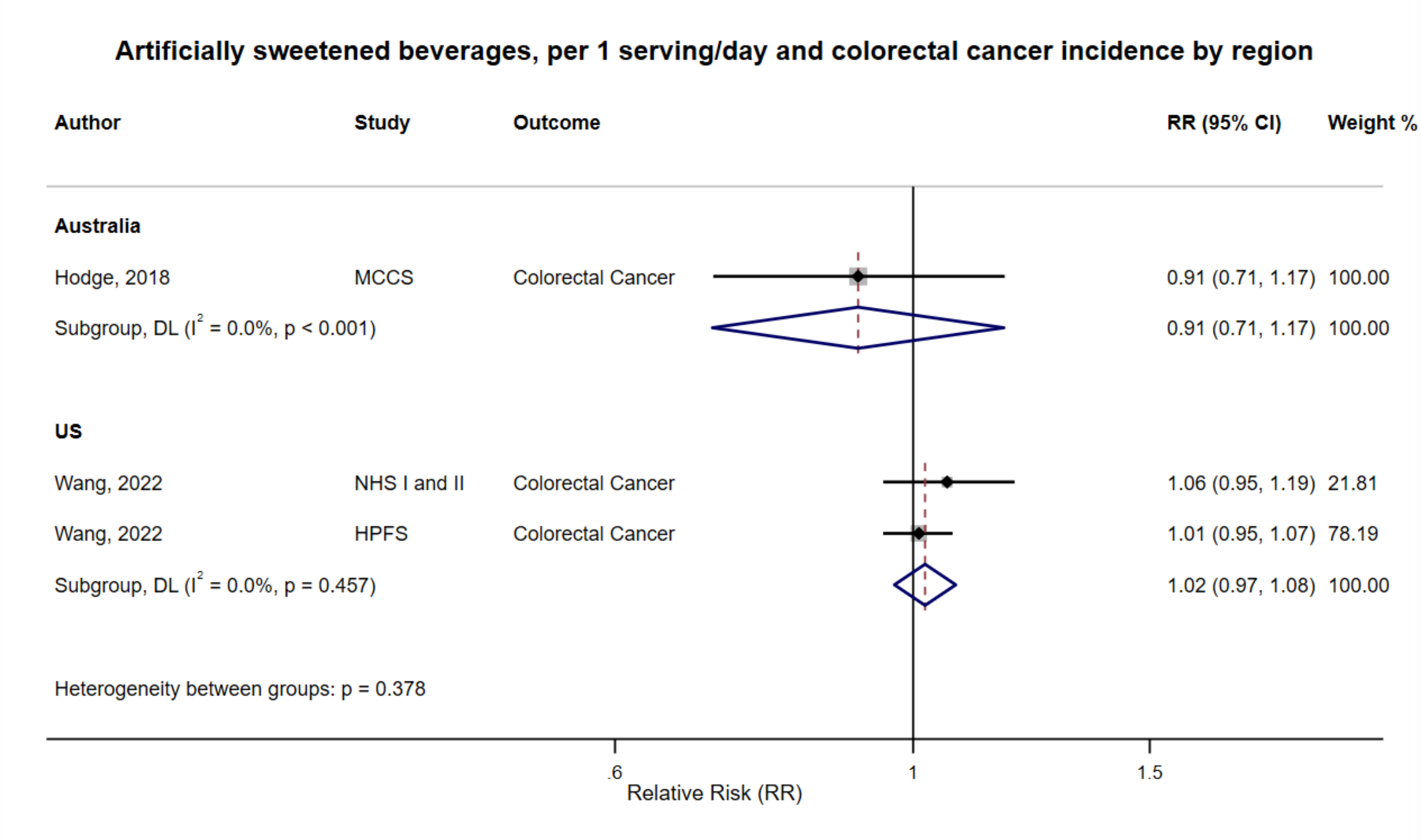

Figure legend: Forest plot shows the linear dose-response results (per 1 serving [355 mL] of artificially sweetened beverages/day) from the inverse variance DerSimonian-Laird random-effects model. The diamond represents the summary relative risk (RR) estimate, the diamond's width is the 95% confidence interval (CI). Each square and the horizontal line across the square represent the RR estimate of the individual study and its 95% CI.

Abbreviations: CI, confidence interval; RR, relative risk.

Study abbreviations: NHS and HPFS, Nurses' Health Study and Health Professionals Follow-up Study; HPFS, Health Professionals Follow-up Study

Hodge, 2018[70], Wang, 2022[168]

**Supplementary Figure 211. Linear dose-response meta-analysis of artificially sweetened beverages and colorectal cancer incidence by frequency of dietary assessment.**

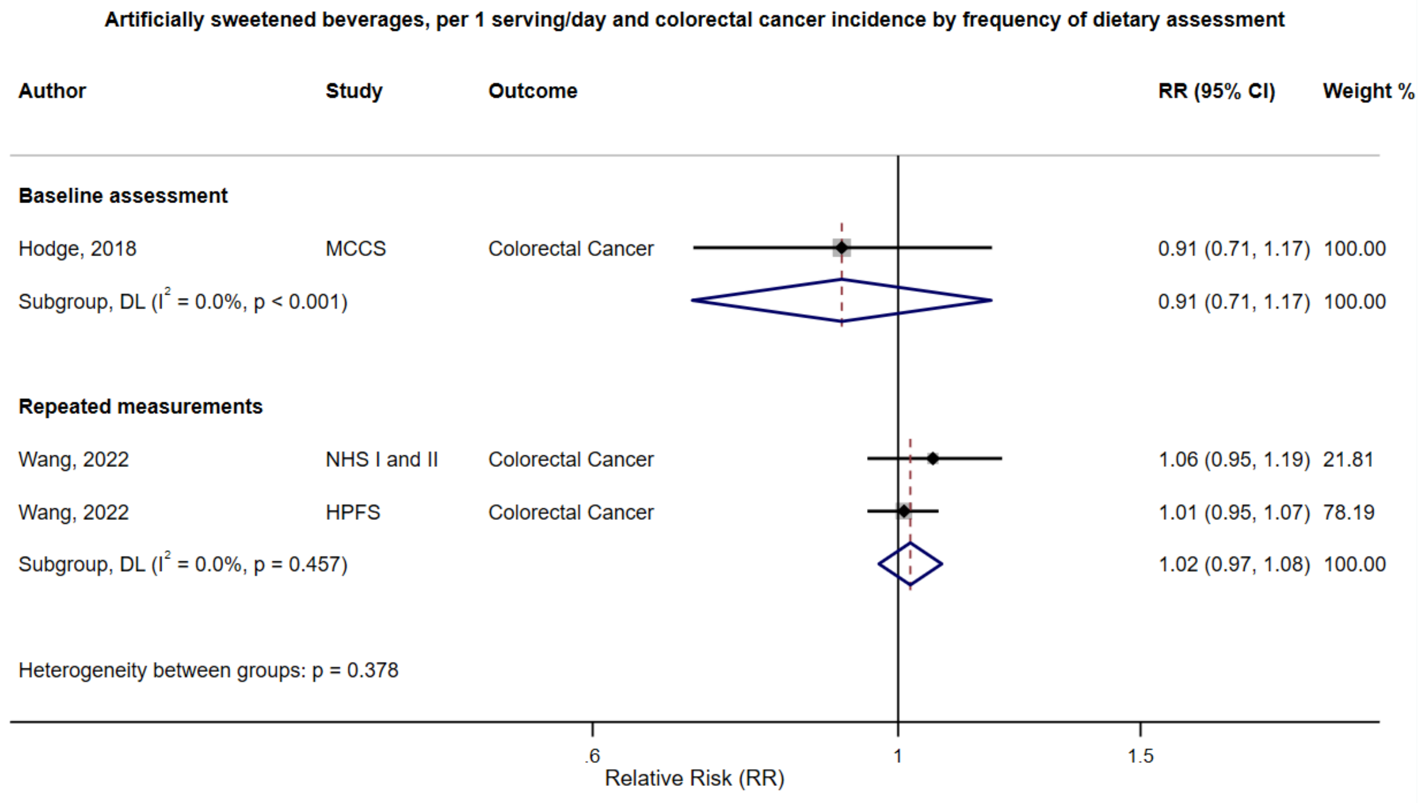

Figure legend: Forest plot shows the linear dose-response results (per 1 serving [355 mL] of artificially sweetened beverages/day) from the inverse variance DerSimonian-Laird random-effects model. The diamond represents the summary relative risk (RR) estimate, the diamond's width is the 95% confidence interval (CI). Each square and the horizontal line across the square represent the RR estimate of the individual study and its 95% CI.

Abbreviations: CI, confidence interval; RR, relative risk.

Study abbreviations: MCCS, Melbourne Collaborative Cohort Study; NHS I and II, Nurses' Health Study I and II; HPFS, Health Professionals Follow-up Study

Hodge, 2018[70], Wang, 2022[168]

**Supplementary Figure 212. Linear dose-response meta-analysis of artificially sweetened beverages and pancreatic cancer incidence and mortality combined by sex.**

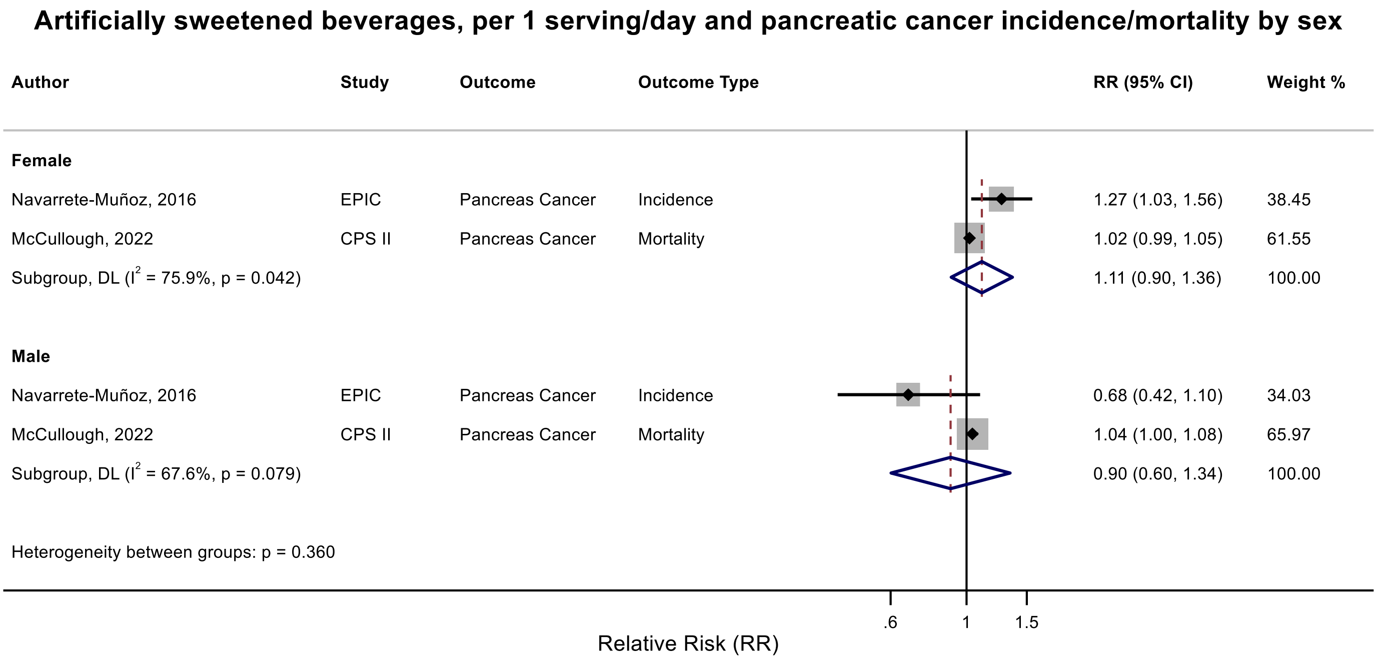

Figure legend: Forest plot shows the linear dose-response results (per 1 serving [355 mL] of artificially sweetened beverages/day) from the inverse variance DerSimonian-Laird random-effects model. The diamond represents the summary relative risk (RR) estimate, the diamond's width is the 95% confidence interval (CI). Each square and the horizontal line across the square represent the RR estimate of the individual study and its 95% CI.

Abbreviations: CI, confidence interval; RR, relative risk.

Study abbreviations: EPIC, European Prospective Investigation into Cancer and nutrition; CPS II, Cancer Prevention Study II

Note: Since only two studies, one on incidence and one on mortality, reported risk estimated across sex, we combined incidence and mortality to be able to perform this subgroup analysis.

Navarrete-Muñoz, 2016[127], McCullough, 2022[113]

**Supplementary Figure 213. Linear dose-response meta-analysis of artificially sweetened beverages and pancreatic cancer incidence, by smoking status.**

**Artificially sweetened beverages, per 1 serving/day and pancreatic cancer incidence by smoking status**

| Author | Study | Outcome | Outcome Type | RR (95% CI) | Weight % |
|---|---|---|---|---|---|
| **Never smokers** | | | | | |
| Bao, 2008 | NIH-AARP | Pancreatic Adenocarcinoma | Incidence | 1.11 (0.98, 1.26) | 93.11 |
| Navarrete-Muñoz, 2016 | EPIC | Pancreas Cancer | Incidence | 0.95 (0.59, 1.51) | 6.89 |
| Subgroup, DL ($I^2$ = 0.0%, p = 0.524) | | | | 1.10 (0.97, 1.24) | 100.00 |
| **Current smokers** | | | | | |
| Bao, 2008 | NIH-AARP | Pancreatic Adenocarcinoma | Incidence | 0.99 (0.83, 1.17) | 79.72 |
| Navarrete-Muñoz, 2016 | EPIC | Pancreas Cancer | Incidence | 1.06 (0.75, 1.49) | 20.28 |
| Subgroup, DL ($I^2$ = 0.0%, p = 0.703) | | | | 1.00 (0.86, 1.17) | 100.00 |
| Heterogeneity between groups: p = 0.368 | | | | | |

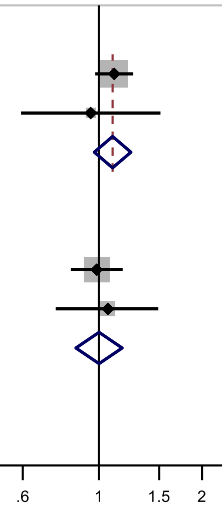

Figure legend: Forest plot shows the linear dose-response results (per 1 serving [355 mL] of artificially sweetened beverages/day) from the inverse variance DerSimonian-Laird random-effects model. The diamond represents the summary relative risk (RR) estimate, the diamond's width is the 95% confidence interval (CI). Each square and the horizontal line across the square represent the RR estimate of the individual study and its 95% CI.

Abbreviations: CI, confidence interval; RR, relative risk.

Study abbreviations: NIH-AARP, National Institute of Health + American Association of Retired Persons Diet and Health Study; EPIC, European Prospective Investigation into Cancer and nutrition; CPS II, Cancer Prevention Study II

Bao, 2008[39], Navarrete-Muñoz, 2016[127]

**Supplementary Figure 214. Linear dose-response meta-analysis of artificially sweetened beverages and pancreatic cancer incidence and mortality combined by body mass index.**

**Artificially sweetened beverages, per 1 serving/day and pancreatic cancer incidence/mortality by body mass index**

| Author | Study | Outcome | Outcome Type | RR (95% CI) | Weight % |
|---|---|---|---|---|---|
| **≥30 kg/m²** | | | | | |
| Bao, 2008 | NIH-AARP | Pancreatic Adenocarcinoma | Incidence | 0.94 (0.75, 1.18) | 6.45 |
| McCullough, 2022 | CPS II | Pancreas Cancer | Mortality | 1.01 (0.95, 1.07) | 93.55 |
| Subgroup, DL ($I^2$ = 0.0%, p = 0.572) | | | | 1.01 (0.95, 1.07) | 100.00 |
| **<30 kg/m²** | | | | | |
| Bao, 2008 | NIH-AARP | Pancreatic Adenocarcinoma | Incidence | 1.08 (0.97, 1.21) | 4.35 |
| McCullough, 2022 | CPS II | Pancreas Cancer | Mortality | 1.03 (1.00, 1.05) | 95.65 |
| Subgroup, DL ($I^2$ = 0.0%, p = 0.345) | | | | 1.03 (1.00, 1.05) | 100.00 |
| **≥25 kg/m²** | | | | | |
| Navarrete-Muñoz, 2016 | EPIC | Pancreas Cancer | Incidence | 1.16 (0.94, 1.44) | 1.82 |
| McCullough, 2022 | CPS II | Pancreas Cancer | Mortality | 1.04 (1.01, 1.07) | 98.18 |
| Subgroup, DL ($I^2$ = 0.0%, p = 0.326) | | | | 1.04 (1.01, 1.07) | 100.00 |
| **<25 kg/m²** | | | | | |
| Navarrete-Muñoz, 2016 | EPIC | Pancreas Cancer | Incidence | 0.82 (0.46, 1.46) | 0.36 |
| McCullough, 2022 | CPS II | Pancreas Cancer | Mortality | 1.00 (0.97, 1.04) | 99.64 |
| Subgroup, DL ($I^2$ = 0.0%, p = 0.504) | | | | 1.00 (0.97, 1.03) | 100.00 |
| Heterogeneity between groups: p = 0.282 | | | | | |

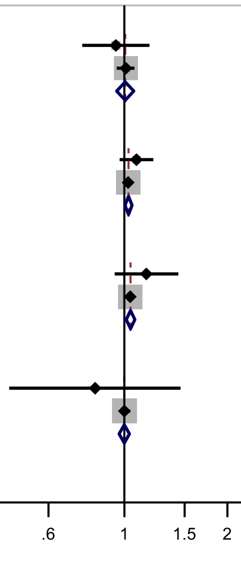

Figure legend: Forest plot shows the linear dose-response results (per 1 serving [355 mL] of artificially sweetened beverages/day) from the inverse variance DerSimonian-Laird random-effects model. The diamond represents the summary relative risk (RR) estimate, the diamond's width is the 95% confidence interval (CI). Each square and the horizontal line across the square represent the RR estimate of the individual study and its 95% CI

Abbreviations: BMI, body mass index, CI, confidence interval; RR, relative risk.

Study abbreviations: CPS II, Cancer Prevention Study II; EPIC, European Prospective Investigation into Cancer and nutrition; NIH-AARP & PLCO, National Institute of Health + American Association of Retired Persons Diet and Health Study & Prostate, Lung, Colorectal and Ovarian Cancer Screening trial

Note: We combined studies reporting on incidence and mortality outcomes to be able to perform subgroup analysis by baseline body mass index.

Navarrete-Muñoz, 2016[127], McCullough, 2022[113], Bao, 2008[39]

**Supplementary Figure 215. Linear dose-response meta-analysis of carbonated artificially sweetened beverages and pancreatic cancer incidence by body mass index.**

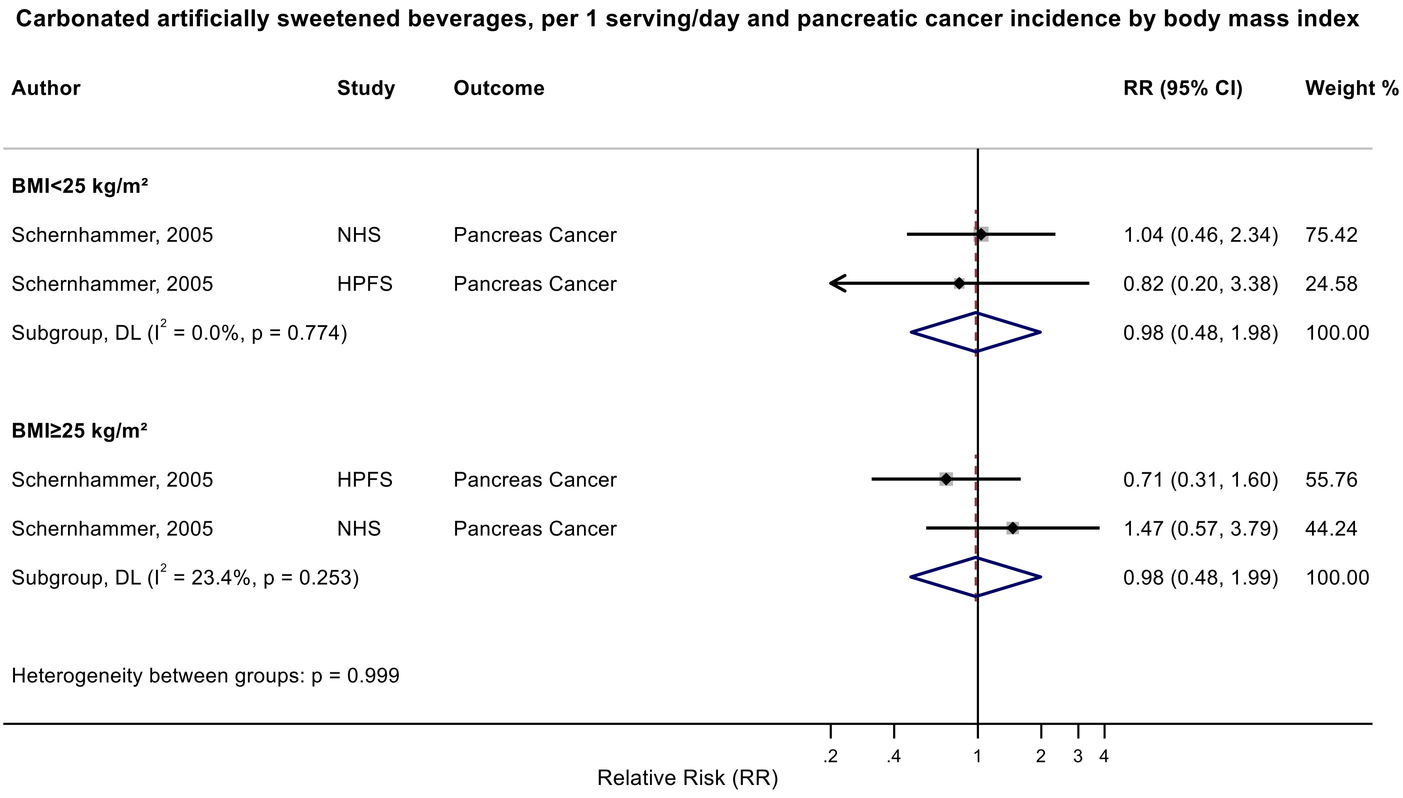

Figure legend: Forest plot shows the linear dose-response results (per 1 serving [355 mL] of carbonated artificially sweetened beverages/day) from the inverse variance DerSimonian-Laird random-effects model. The diamond represents the summary relative risk (RR) estimate, the diamond's width is the 95% confidence interval (CI). Each square and the horizontal line across the square represent the RR estimate of the individual study and its 95% CI.

Abbreviations: CI, confidence interval; RR, relative risk.

Study abbreviations: NHS, Nurses' Health Study; HPFS, Health Professionals Follow-up Study

Schernhammer, 2005[151]

#### Supplementary Figure 216. Linear dose-response meta-analysis of artificially sweetened beverages and breast cancer incidence by geographical region.

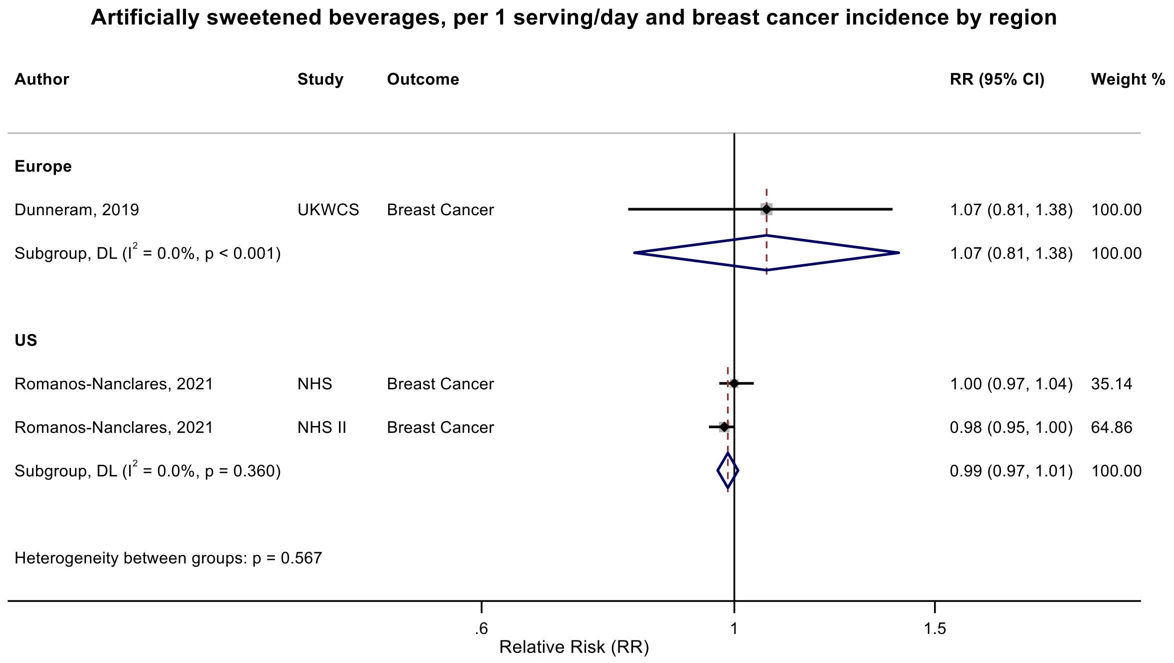

Figure legend: Forest plot shows the linear dose-response results (per 1 serving [355 mL] of artificially sweetened beverages/day) from the inverse variance DerSimonian-Laird random-effects model. The diamond represents the summary relative risk (RR) estimate, the diamond's width is the 95% confidence interval (CI). Each square and the horizontal line across the square represent the RR estimate of the individual study and its 95% CI.

Abbreviations: CI, confidence interval; RR, relative risk.

Study abbreviations: UKWCS, United Kingdom Women's Cohort Study; NHS, Nurses' Health Study; NHS II, Nurses' Health Study II

Dunneram, 2019[49], Romanos-Nanclares, 2021[145]

**Supplementary Figure 217. Linear dose-response meta-analysis of artificially sweetened beverages and breast cancer incidence by frequency of dietary assessment.**

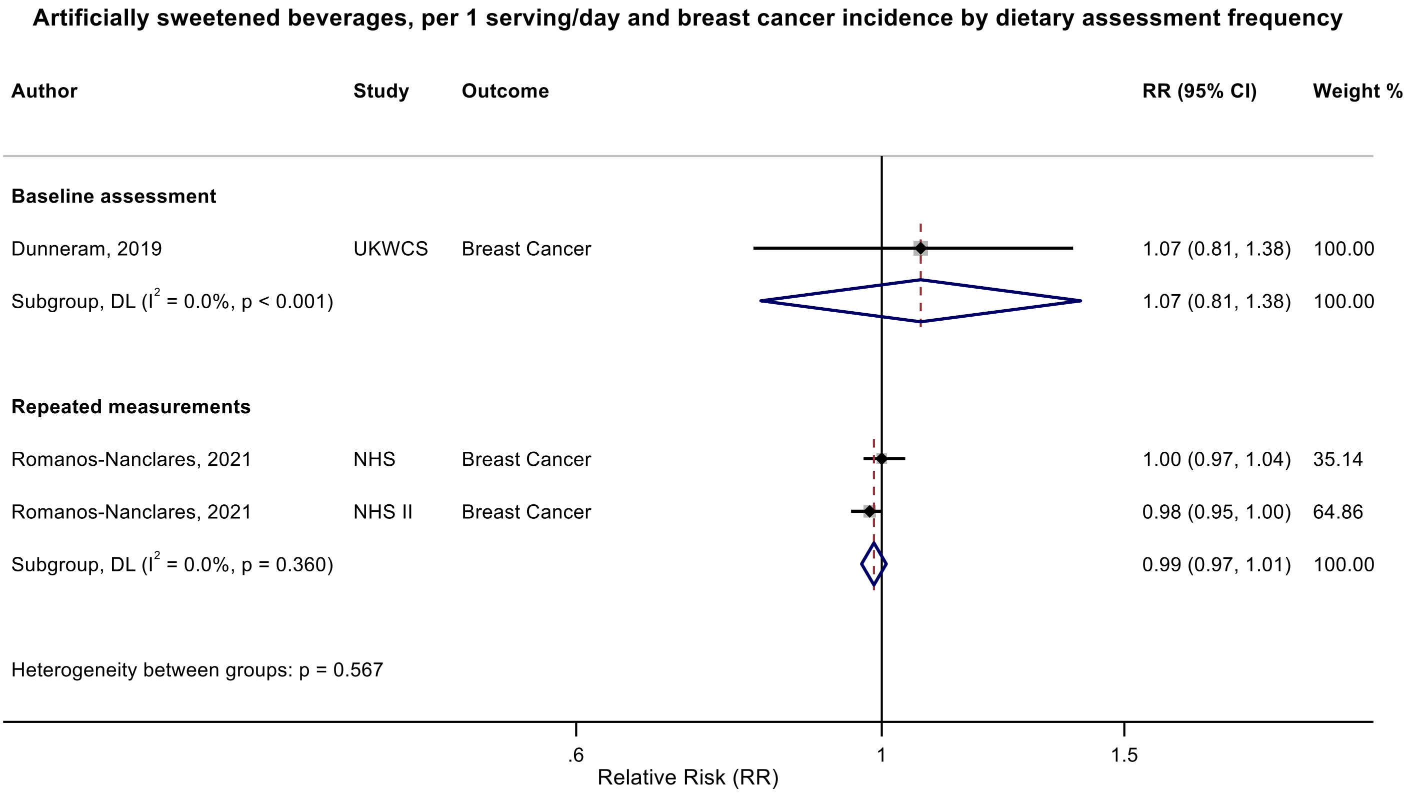

Figure legend: Forest plot shows the linear dose-response results (per 1 serving [355 mL] of artificially sweetened beverages/day) from the inverse variance DerSimonian-Laird random-effects model. The diamond represents the summary relative risk (RR) estimate, the diamond's width is the 95% confidence interval (CI). Each square and the horizontal line across the square represent the RR estimate of the individual study and its 95% CI.

Abbreviations: CI, confidence interval; RR, relative risk.

Study abbreviations: UKWCS, United Kingdom Women's Cohort Study; NHS, Nurses' Health Study; NHS II, Nurses' Health Study II

Dunneram, 2019[49], Romanos-Nanclares, 2021[145]

**Supplementary Figure 218. Linear dose-response meta-analysis of artificially sweetened beverages and postmenopausal breast cancer incidence by frequency of dietary assessment.**

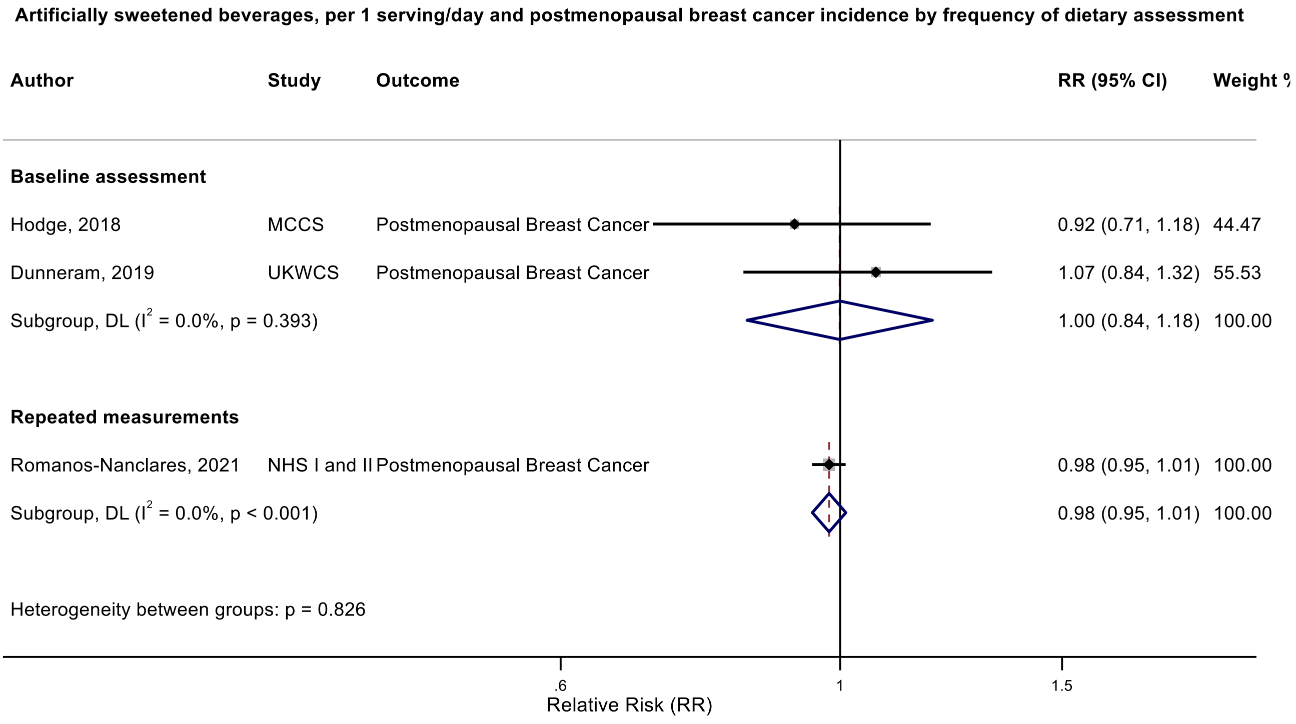

Figure legend: Forest plot shows the linear dose-response results (per 1 serving [355 mL] of artificially sweetened beverages/day) from the inverse variance DerSimonian-Laird random-effects model. The diamond represents the summary relative risk (RR) estimate, the diamond's width is the 95% confidence interval (CI). Each square and the horizontal line across the square represent the RR estimate of the individual study and its 95% CI.

Abbreviations: CI, confidence interval; RR, relative risk.

Study abbreviations: MCCS, Melbourne Collaborative Cohort Study; UKWCS, United Kingdom Women's Cohort Study; NHS I and II, Nurses' Health Study I and II

Hodge, 2018[70], Dunneram, 2019[49], Romanos-Nanclares, 2021[145]

**Supplementary Figure 219. Linear dose-response meta-analysis of artificially sweetened beverages and endometrial cancer incidence by exclusion of people with diabetes at baseline.**

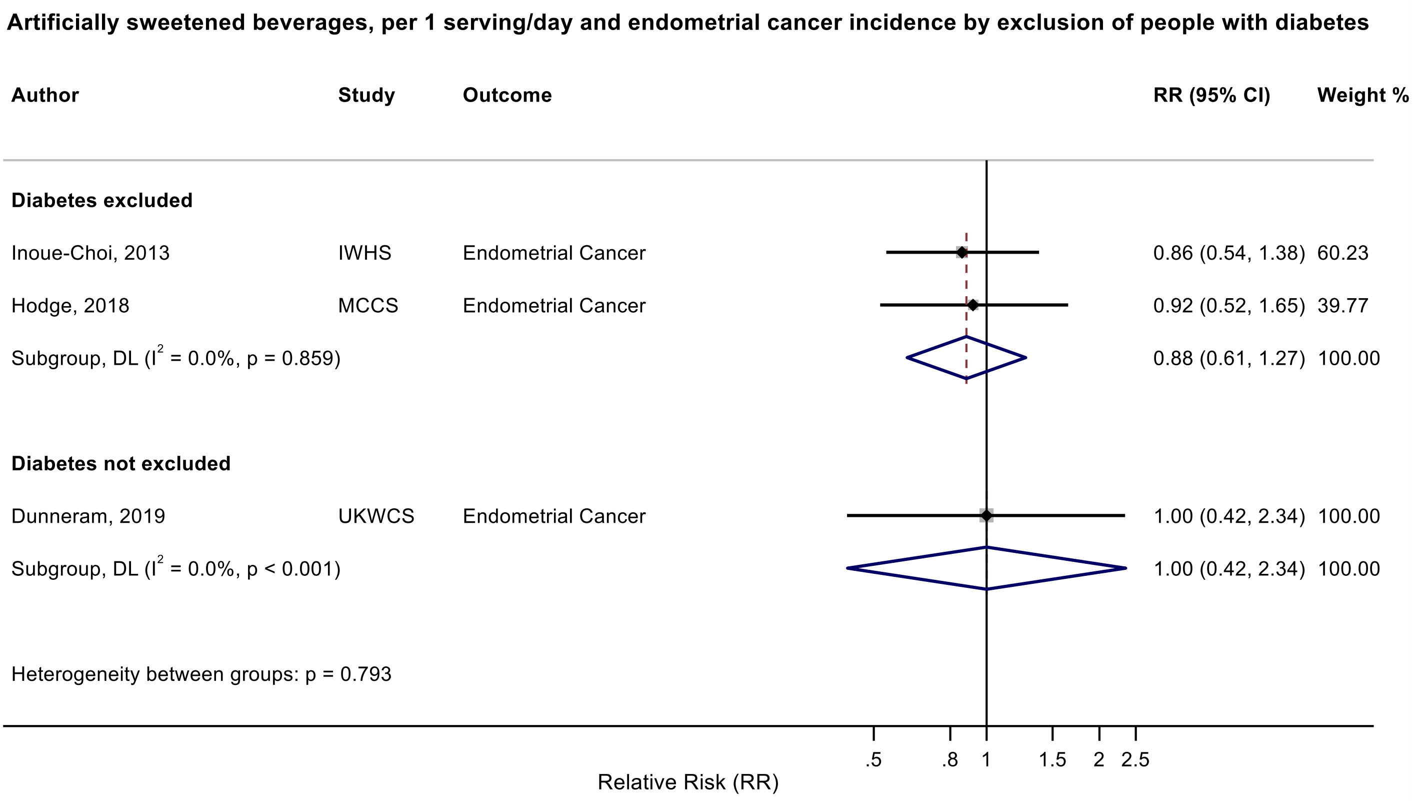

Figure legend: Forest plot shows the linear dose-response results (per 1 serving [355 mL] of artificially sweetened beverages/day) from the inverse variance DerSimonian-Laird random-effects model. The diamond represents the summary relative risk (RR) estimate, the diamond's width is the 95% confidence interval (CI). Each square and the horizontal line across the square represent the RR estimate of the individual study and its 95% CI.

Abbreviations: CI, confidence interval; RR, relative risk.

Study abbreviations: IWHS, The Iowa Women’s Health Study; MCCS, Melbourne Collaborative Cohort Study; UKWCS, United Kingdom Women's Cohort Study

Note: The UKWCS controlled for a history of diabetes in the analysis.

Inoue-Choi, 2013[74], Hodge, 2018[70], Dunneram, 2019[49]

**Supplementary Figure 220. Linear dose-response meta-analysis of carbonated artificially sweetened beverages and haematological cancers incidence by alcohol drinking.**

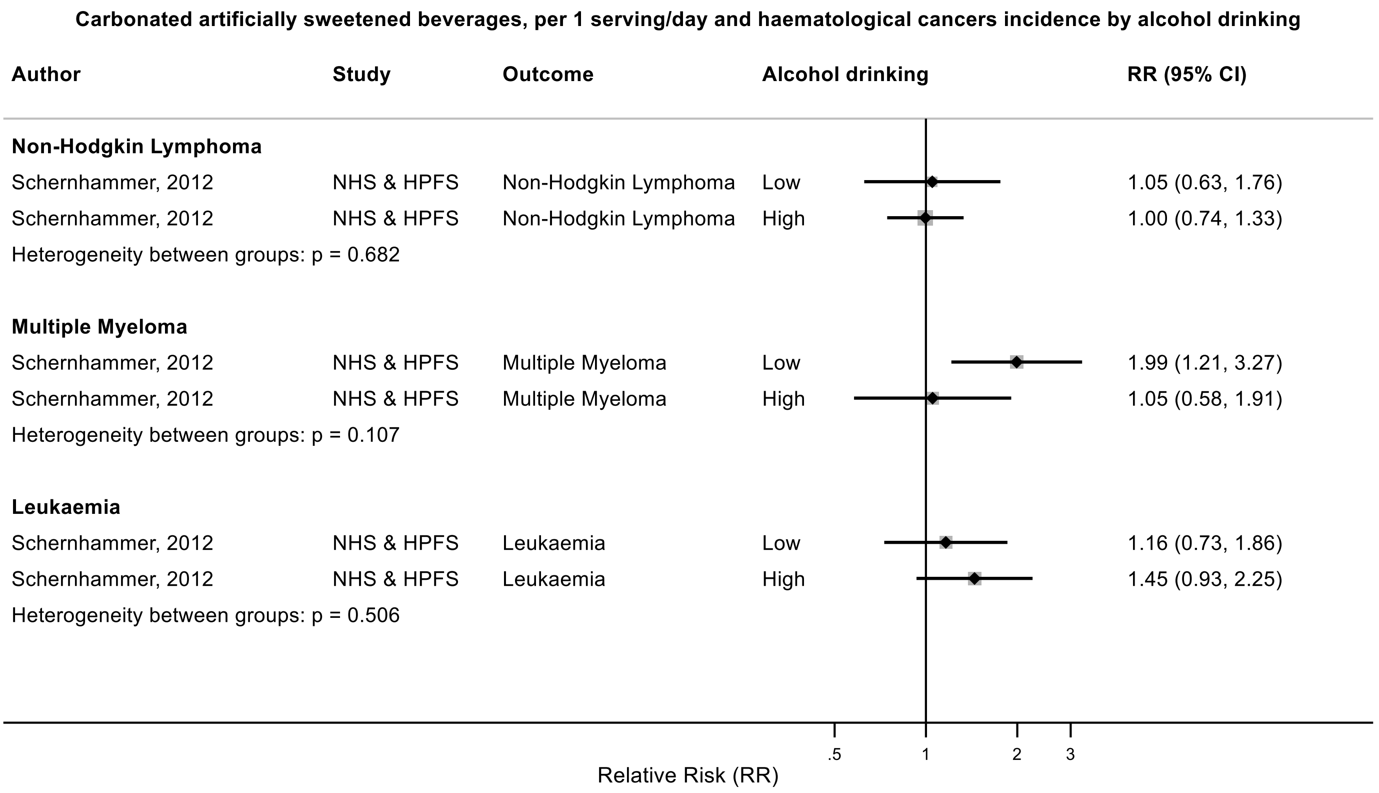

Figure legend: Forest plot shows the linear dose-response results (per 1 serving [355 mL] of carbonated artificially sweetened beverages/day) from the inverse variance DerSimonian-Laird random-effects model. The diamond represents the summary relative risk (RR) estimate, the diamond's width is the 95% confidence interval (CI). Each square and the horizontal line across the square represent the RR estimate of the individual study and its 95% CI.

Abbreviations: CI, confidence interval; RR, relative risk.

Study abbreviations: NHS & HPFS, Nurses' Health Study & Health Professionals Follow-up Study

Schernhammer, 2012[150]

**Supplementary Figure 221. Linear dose-response meta-analysis of artificially sweetened beverages and colorectal cancer incidence, leave-one-out analysis.**

**Artificially sweetened beverages, per 1 serving/day and colorectal cancer incidence, leave-one-out analysis**

| Author | Study | Outcome | RR (95% CI) |
|---|---|---|---|
| Hodge, 2018 | MCCS | Colorectal Cancer | 1.02 (0.97, 1.08) |
| Wang, 2022 | NHS I & II | Colorectal Cancer | 1.00 (0.95, 1.06) |
| Wang, 2022 | HPFS | Colorectal Cancer | 1.02 (0.90, 1.16) |

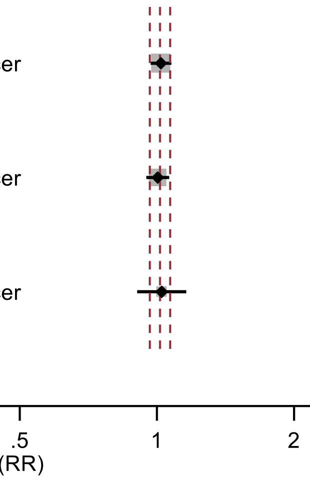

Figure legend: Forest plot shows the linear dose-response results (per 1 serving [355 mL] of artificially sweetened beverages/day) from the inverse variance DerSimonian-Laird random-effects model after exclusion of every single study at a time.

Abbreviations: CI, confidence interval; RR, relative risk.

Study abbreviations: MCCS, Melbourne Collaborative Cohort Study; NHS I & II, Nurses' Health Study I & II; HPFS, Health Professionals Follow-up Study

Hodge, 2018[70], Wang, 2022[168]

**Supplementary Figure 222. Linear dose-response meta-analysis of artificially sweetened beverages and liver cancer incidence, leave-one-out analysis.**

**Artificially sweetened beverages, per 1 serving/day and liver cancer incidence, leave-one-out**

| Author | Study | Outcome | RR (95% CI) |
|---|---|---|---|
| Stepien, 2016 | EPIC | HCC | 1.01 (0.93, 1.09) |
| Jones, 2022 | NIH-AARP & PLCO | Liver Cancer | 1.37 (1.00, 1.87) |
| Zhao, 2023 | WHI | Liver Cancer | 1.23 (0.81, 1.89) |

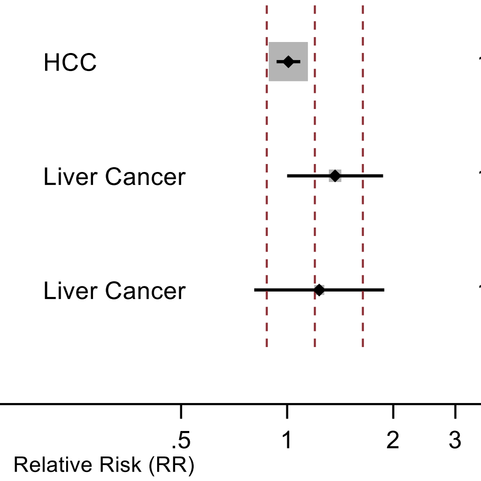

Figure legend: Forest plot shows the linear dose-response results (per 1 serving [355 mL] of artificially sweetened beverages/day) from the inverse variance DerSimonian-Laird random-effects model after exclusion of every single study at a time.

Abbreviations: CI, confidence interval; RR, relative risk.

Study abbreviations: EPIC, European Prospective Investigation into Cancer and nutrition; NIH-AARP & PLCO, National Institute of Health + American Association of Retired Persons Diet and Health Study & Prostate, Lung, Colorectal and Ovarian Cancer Screening trial; WHI, Women's Health Initiative Study

Stepien, 2016[159], Jones, 2022[79], Zhao, 2023[186]

**Supplementary Figure 223. Linear dose-response meta-analysis of artificially sweetened beverages and pancreatic cancer incidence and mortality combined, leave-one-out analysis.**

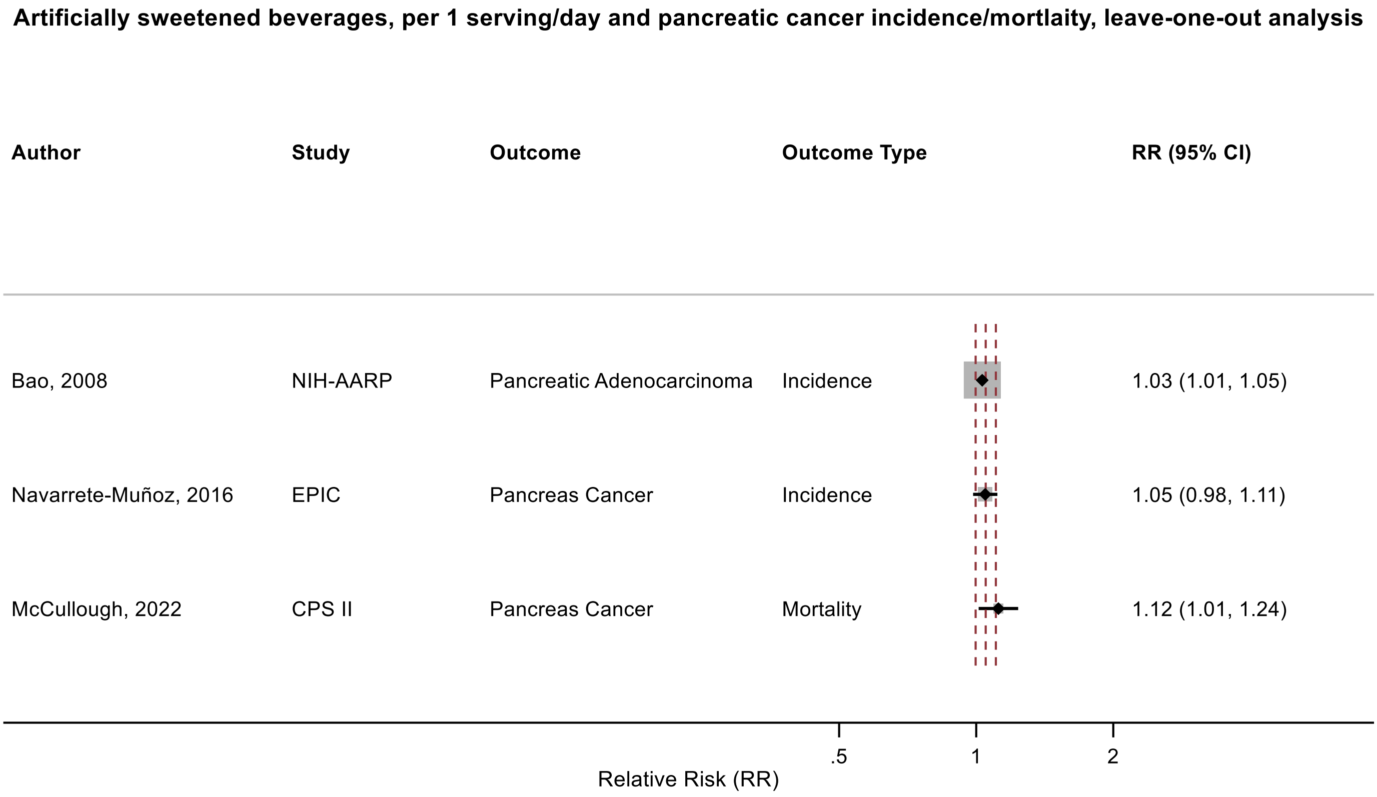

Figure legend: Forest plot shows the linear dose-response results (per 1 serving [355 mL] of artificially sweetened beverages/day) from the inverse variance DerSimonian-Laird random-effects model after exclusion of every single study at a time.

Abbreviations: CI, confidence interval; RR, relative risk.

Study abbreviations: NIH-AARP, National Institute of Health + American Association of Retired Persons Diet and Health Study; EPIC, European Prospective Investigation into Cancer and nutrition; CPS II, Cancer Prevention Study II

Note: We did influence analysis for incidence and mortality combined, as only 2 studies were available for the analysis of incidence.

Bao, 2008[39], Navarrete-Muñoz, 2016[127], McCullough, 2022[113]

**Supplementary Figure 224. Linear dose-response meta-analysis of artificially sweetened beverages and breast cancer incidence, leave-one-out analysis.**

**Artificially sweetened beverages, per 1 serving/day and breast cancer incidence, leave-one-out analysis**

| Author | Study | Outcome | RR (95% CI) |
|---|---|---|---|
| Dunneram, 2019 | UKWCS | Breast Cancer | 0.99 (0.97, 1.01) |
| Romanos-Nanclares, 2021 | NHS | Breast Cancer | 0.98 (0.96, 1.01) |
| Romanos-Nanclares, 2021 | NHS II | Breast Cancer | 1.00 (0.97, 1.04) |

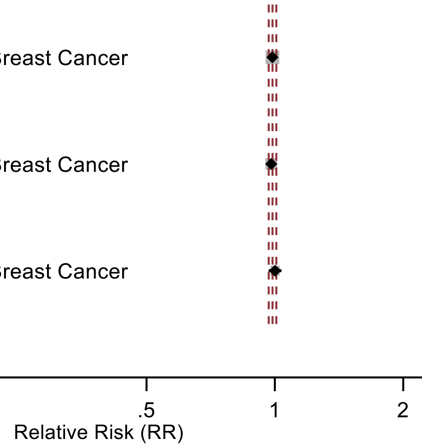

Figure legend: Forest plot shows the linear dose-response results (per 1 serving [355 mL] of artificially sweetened beverages/day) from the inverse variance DerSimonian-Laird random-effects model after exclusion of every single study at a time.

Abbreviations: CI, confidence interval; RR, relative risk.

Study abbreviations: UKWCS, United Kingdom Women's Cohort Study; NHS, Nurses' Health Study; NHS II, Nurses' Health Study II

Dunneram, 2019[49], Romanos-Nanclares, 2021[145]

#### Supplementary Figure 225. Linear dose-response meta-analysis of artificially sweetened beverages and postmenopausal breast cancer incidence, leave-one-out analysis.

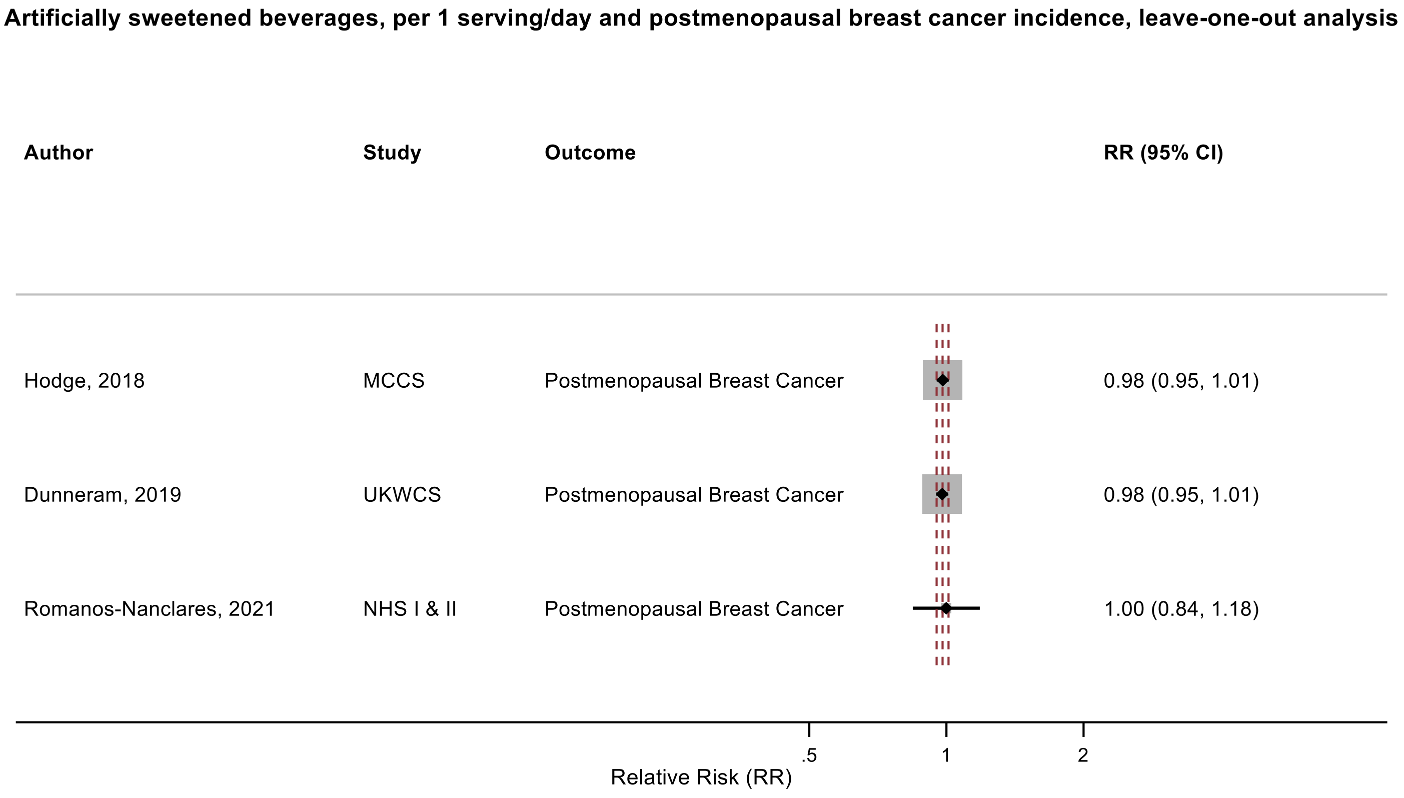

Figure legend: Forest plot shows the linear dose-response results (per 1 serving [355 mL] of artificially sweetened beverages/day) from the inverse variance DerSimonian-Laird random-effects model after exclusion of every single study at a time.

Abbreviations: CI, confidence interval; RR, relative risk.

Study abbreviations: MCCS, Melbourne Collaborative Cohort Study; UKWCS, United Kingdom Women's Cohort Study; NHS I & II, Nurses' Health Study I & II

Hodge, 2018[70], Dunneram, 2019[49], Romanos-Nanclares, 2021[145]

**Supplementary Figure 226. Linear dose-response meta-analysis of artificially sweetened beverages and endometrial cancer incidence, leave-one-out analysis.**

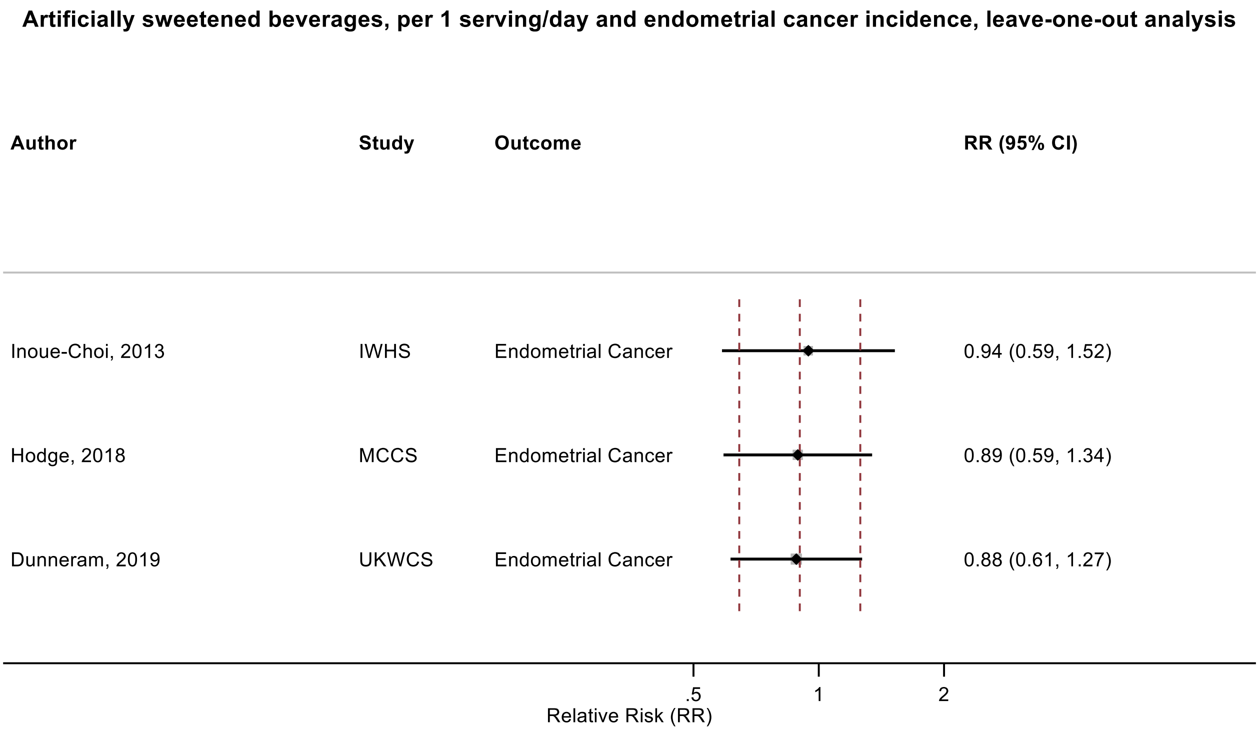

Figure legend: Forest plot shows the linear dose-response results (per 1 serving [355 mL] of artificially sweetened beverages/day) from the inverse variance DerSimonian-Laird random-effects model after exclusion of every single study at a time.

Abbreviations: CI, confidence interval; RR, relative risk.

Study abbreviations: IWHS, The Iowa Women's Health Study; MCCS, Melbourne Collaborative Cohort Study; UKWCS, United Kingdom Women's Cohort Study

Inoue-Choi, 2013[74], Hodge, 2018[70], Dunneram, 2019[49]

**Supplementary Figure 227. Linear dose-response meta-analysis of artificially sweetened beverages and postmenopausal endometrial cancer incidence, leave-one-out analysis.**

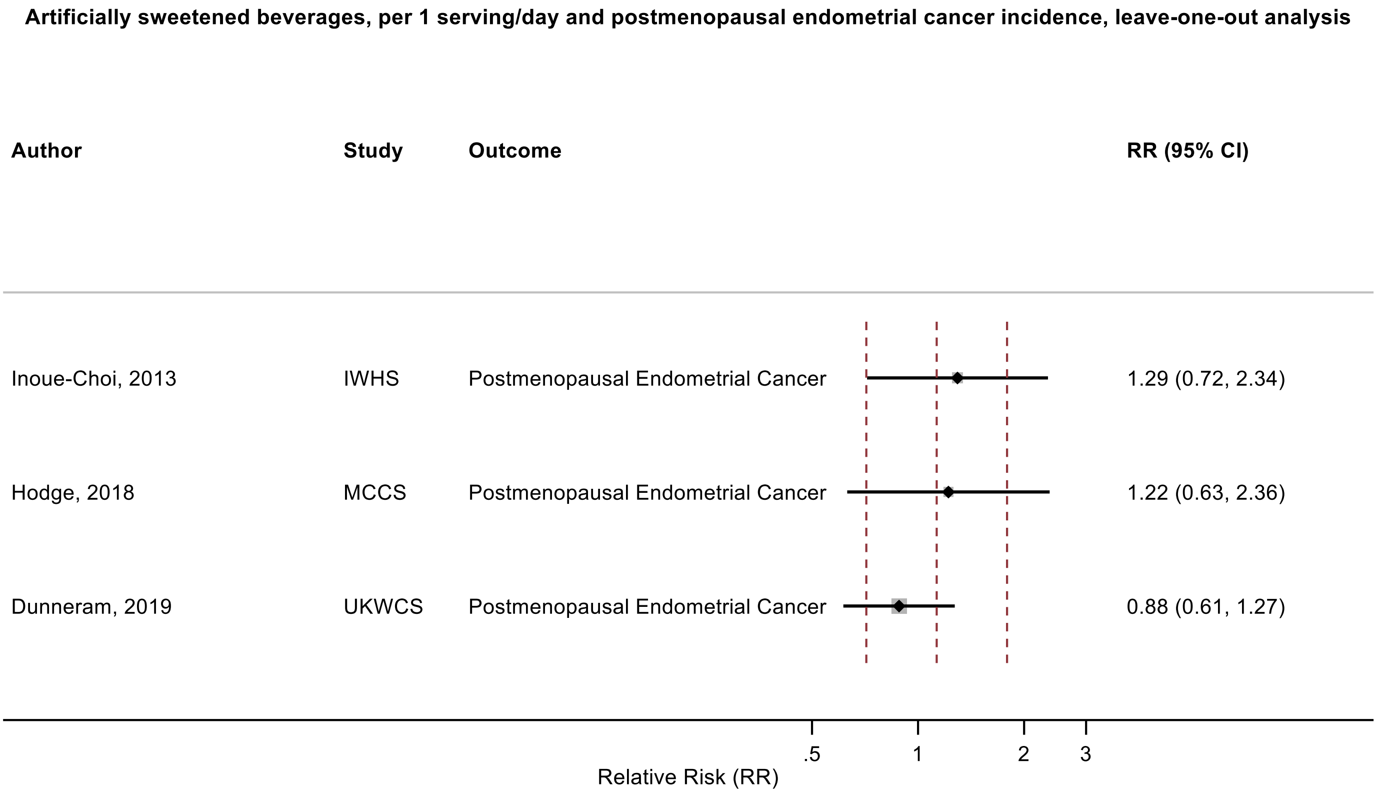

Figure legend: Forest plot shows the linear dose-response results (per 1 serving [355 mL] of artificially sweetened beverages/day) from the inverse variance DerSimonian-Laird random-effects model after exclusion of every single study at a time.

Abbreviations: CI, confidence interval; RR, relative risk.

Study abbreviations: IWHS, The Iowa Women’s Health Study; MCCS, Melbourne Collaborative Cohort Study; UKWCS, United Kingdom Women's Cohort Study

Inoue-Choi, 2013[74], Hodge, 2018[70], Dunneram, 2019[49]

**Supplementary Figure 228. Linear dose-response meta-analysis of artificially sweetened beverages and kidney cancer incidence, leave-one-out analysis.**

**Artificially sweetened beverages, per 1 serving/day and kidney cancer incidence, leave-one-out analysis**

| Author | Study | Outcome | RR (95% CI) |
|---|---|---|---|
| Hodge, 2018 | MCCS | Kidney Cancer | 1.13 (0.97, 1.32) |
| Heath, 2021 | EPIC | Renal Cell Carcinoma | 1.25 (1.00, 1.55) |
| Ringel, 2023 | WHI | Kidney Cancer | 1.13 (0.89, 1.45) |

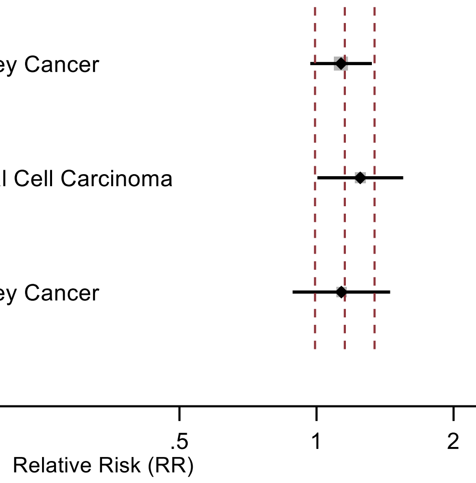

Figure legend: Forest plot shows the linear dose-response results (per 1 serving [355 mL] of artificially sweetened beverages/day) from the inverse variance DerSimonian-Laird random-effects model after exclusion of every single study at a time.

Abbreviations: CI, confidence interval; RR, relative risk.

Study abbreviations: MCCS, Melbourne Collaborative Cohort Study; EPIC, European Prospective Investigation into Cancer and nutrition; WHI, Women's Health Initiative Study

Hodge, 2018[70], Heath, 2021[66], Ringel, 2023[144]

**Supplementary Figure 229. Linear dose-response meta-analysis of artificially sweetened beverages and lung cancer mortality.**

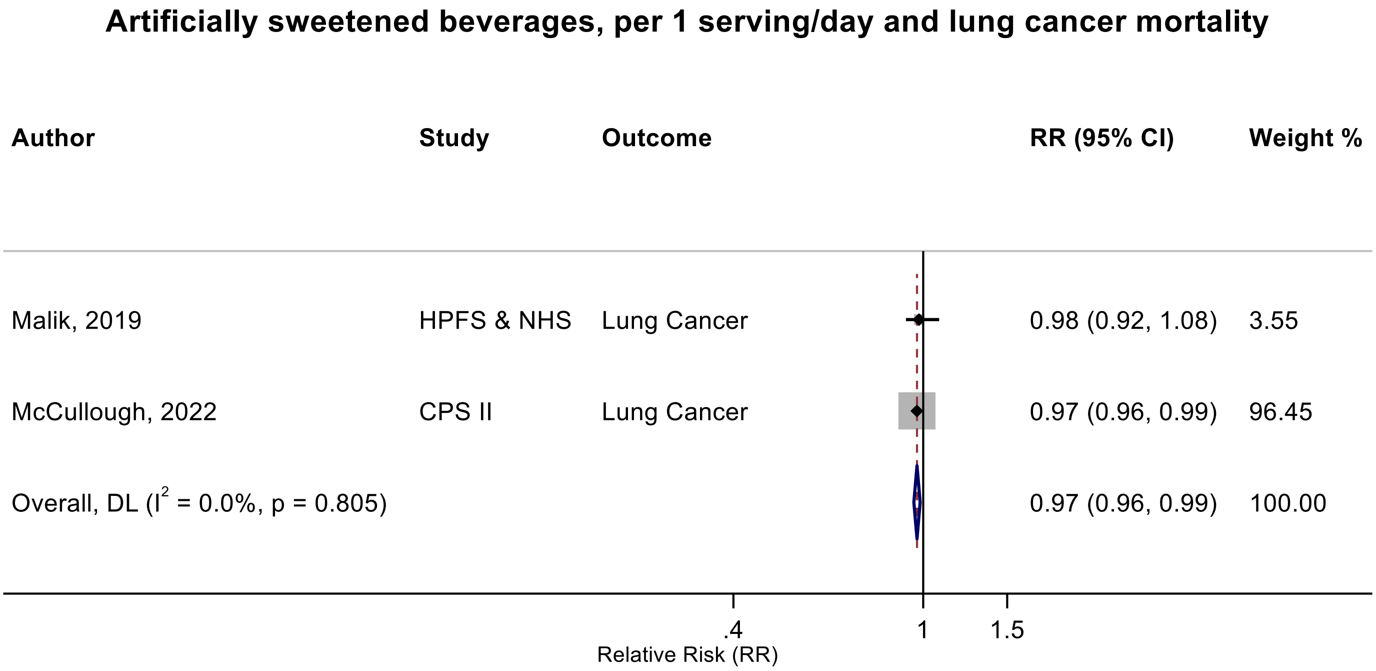

Figure legend: Forest plot shows the linear dose-response results (per 1 serving [355 mL] of artificially sweetened beverages/day) from the inverse variance DerSimonian-Laird random-effects model. The diamond represents the summary relative risk (RR) estimate, the diamond's width is the 95% confidence interval (CI). Each square and the horizontal line across the square represent the RR estimate of the individual study and its 95% CI.

Abbreviations: CI, confidence interval; RR, relative risk.

Study abbreviations: HPFS & NHS, Health Professionals Follow-up Study & Nurses' Health Study; CPS II, Cancer Prevention Study II

Malik, 2019[109], McCullough, 2022[113]

**Supplementary Figure 230. Linear dose-response meta-analysis of artificially sweetened beverages and colorectal cancer and colon cancer mortality.**

#### Artificially sweetened beverages, per 1 serving/day and colorectal cancer mortality

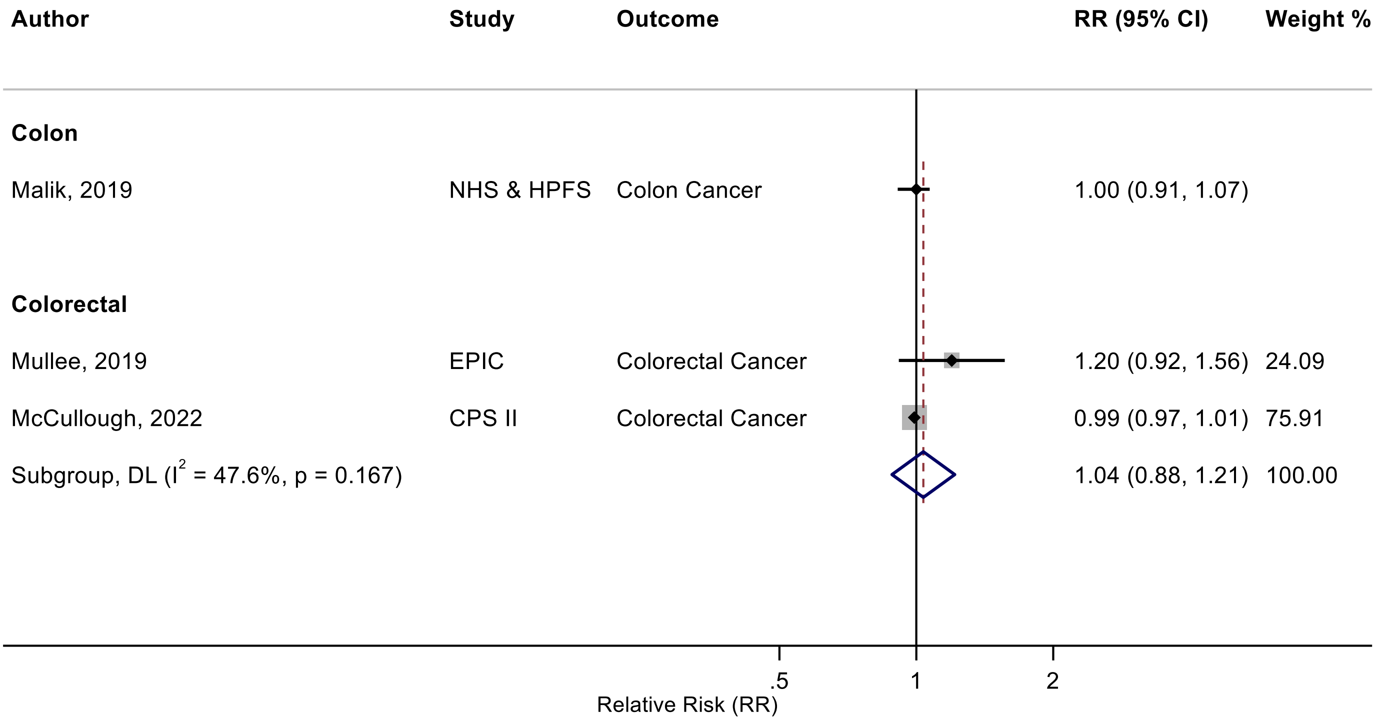

Figure legend: Forest plot shows the linear dose-response results (per 1 serving [355 mL] of artificially sweetened beverages/day) from the inverse variance DerSimonian-Laird random-effects model. The diamond represents the summary relative risk (RR) estimate, the diamond's width is the 95% confidence interval (CI). Each square and the horizontal line across the square represent the RR estimate of the individual study and its 95% CI.

Abbreviations: CI, confidence interval; RR, relative risk.

Study abbreviations: NHS & HPFS, Nurses' Health Study & Health Professionals Follow-up Study; EPIC, European Prospective Investigation into Cancer and nutrition; CPS II, Cancer Prevention Study II

Malik, 2019[109], Mullee, 2019[126], McCullough, 2022[113]

**Supplementary Figure 231. Linear dose-response meta-analysis of artificially sweetened beverages and breast cancer mortality.**

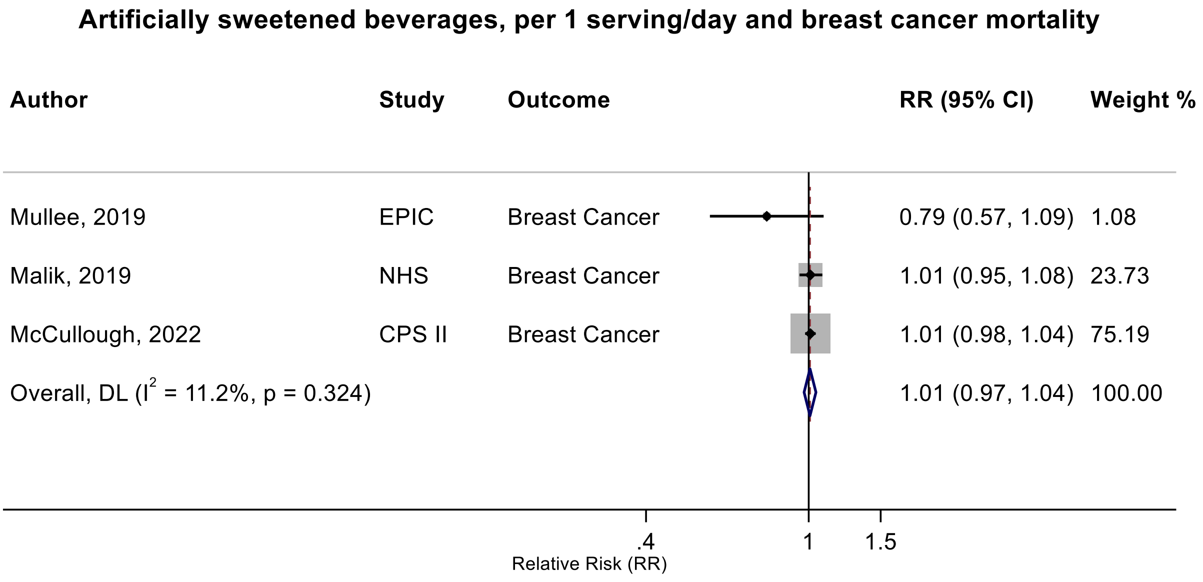

Figure legend: Forest plot shows the linear dose-response results (per 1 serving [355 mL] of artificially sweetened beverages/day) from the inverse variance DerSimonian-Laird random-effects model. The diamond represents the summary relative risk (RR) estimate, the diamond's width is the 95% confidence interval (CI). Each square and the horizontal line across the square represent the RR estimate of the individual study and its 95% CI.

Abbreviations: CI, confidence interval; RR, relative risk.

Study abbreviations: EPIC, European Prospective Investigation into Cancer and nutrition; NHS, Nurses' Health Study; CPS II, Cancer Prevention Study II

Mullee, 2019[126], Malik, 2019[109], McCullough, 2022[113]

**Supplementary Figure 232. Linear dose-response meta-analysis of artificially sweetened beverages and prostate cancer mortality.**

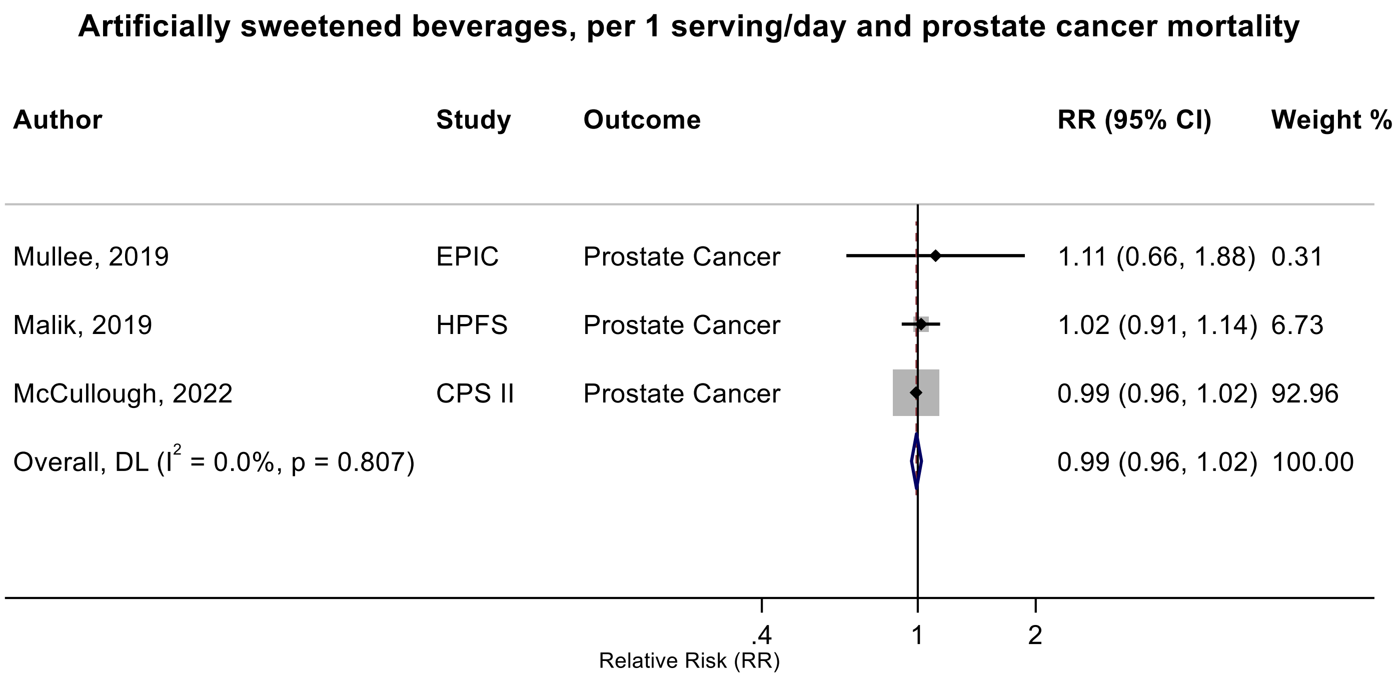

Figure legend: Forest plot shows the linear dose-response results (per 1 serving [355 mL] of artificially sweetened beverages/day) from the inverse variance DerSimonian-Laird random-effects model. The diamond represents the summary relative risk (RR) estimate, the diamond's width is the 95% confidence interval (CI). Each square and the horizontal line across the square represent the RR estimate of the individual study and its 95% CI.

Abbreviations: CI, confidence interval; RR, relative risk.

Study abbreviations: EPIC, European Prospective Investigation into Cancer and nutrition; HPFS, Health Professionals Follow-up Study; CPS II, Cancer Prevention Study II

Mullee, 2019[126], Malik, 2019[109], McCullough, 2022[113]

**Supplementary Figure 233. Linear dose-response meta-analysis of artificially sweetened beverages and kidney cancer mortality.**

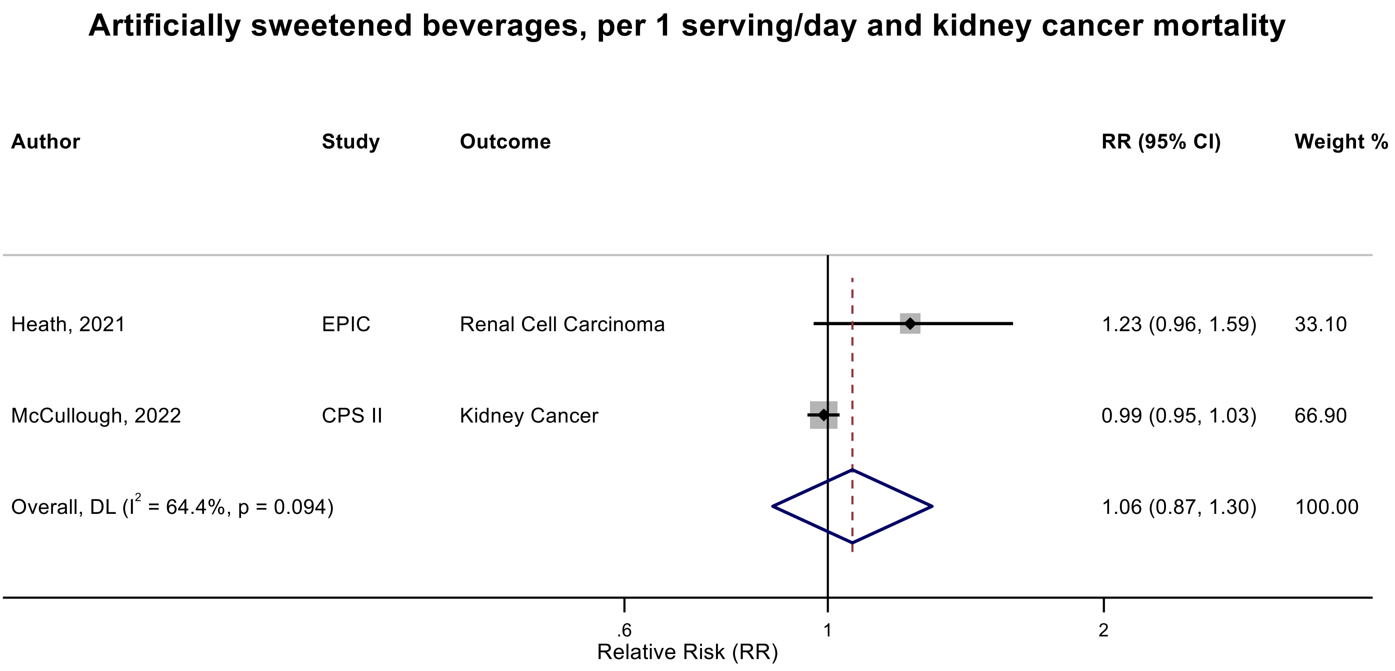

Figure legend: Forest plot shows the linear dose-response results (per 1 serving [355 mL] of artificially sweetened beverages/day) from the inverse variance DerSimonian-Laird random-effects model. The diamond represents the summary relative risk (RR) estimate, the diamond's width is the 95% confidence interval (CI). Each square and the horizontal line across the square represent the RR estimate of the individual study and its 95% CI.

Abbreviations: CI, confidence interval; RR, relative risk.

Study abbreviations: EPIC, European Prospective Investigation into Cancer and nutrition; CPS II, Cancer Prevention Study II

Heath, 2021[66], McCullough, 2022[113]

**Supplementary Figure 234. Linear dose-response meta-analysis of artificially sweetened beverages and kidney cancer mortality by sex.**

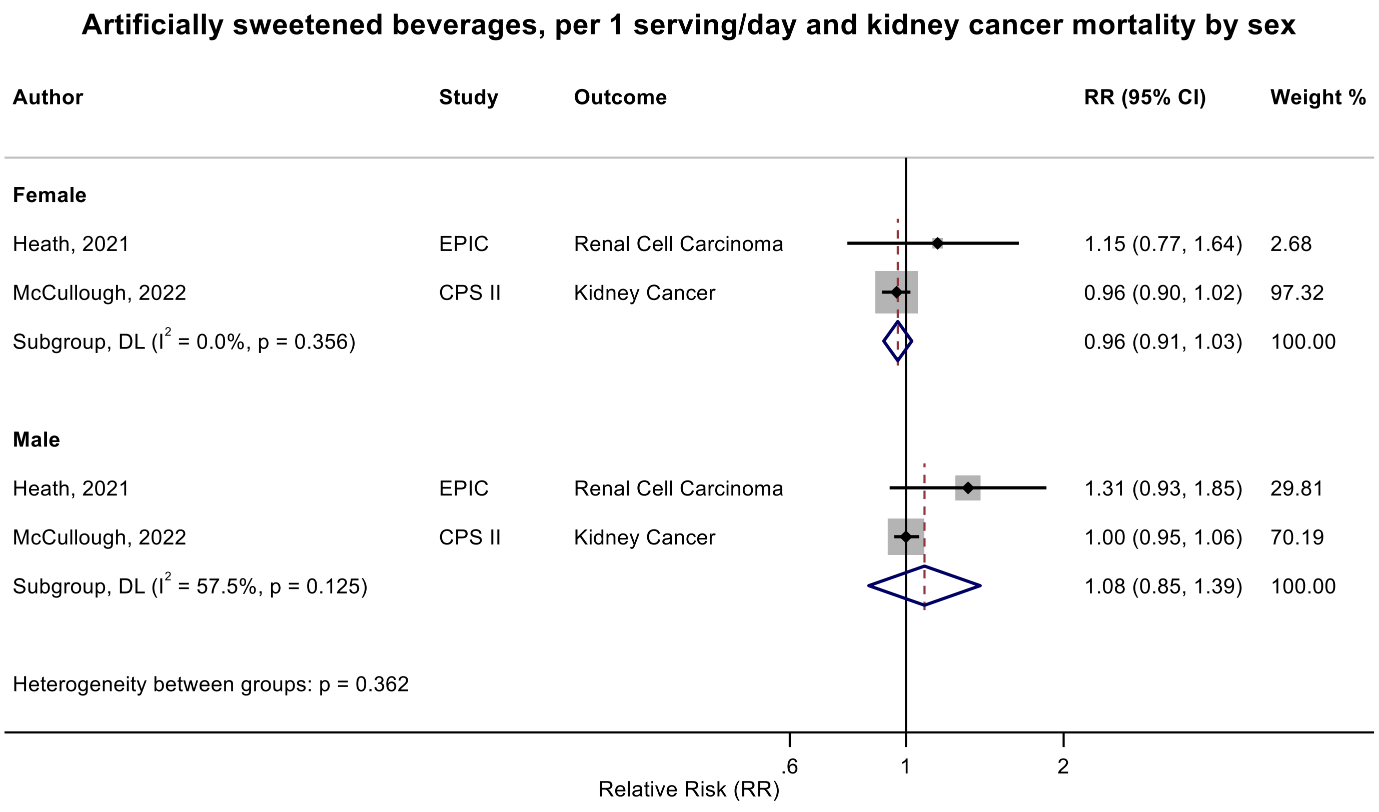

Figure legend: Forest plot shows the linear dose-response results (per 1 serving [355 mL] of artificially sweetened beverages/day) from the inverse variance DerSimonian-Laird random-effects model. The diamond represents the summary relative risk (RR) estimate, the diamond's width is the 95% confidence interval (CI). Each square and the horizontal line across the square represent the RR estimate of the individual study and its 95% CI.

Abbreviations: CI, confidence interval; RR, relative risk.

Study abbreviations: EPIC, European Prospective Investigation into Cancer and nutrition; CPS II, Cancer Prevention Study II

Heath, 2021[66], McCullough, 2022[113]

**Supplementary Figure 235. Linear dose-response meta-analysis of artificially sweetened beverages and breast cancer mortality by frequency of dietary assessment.**

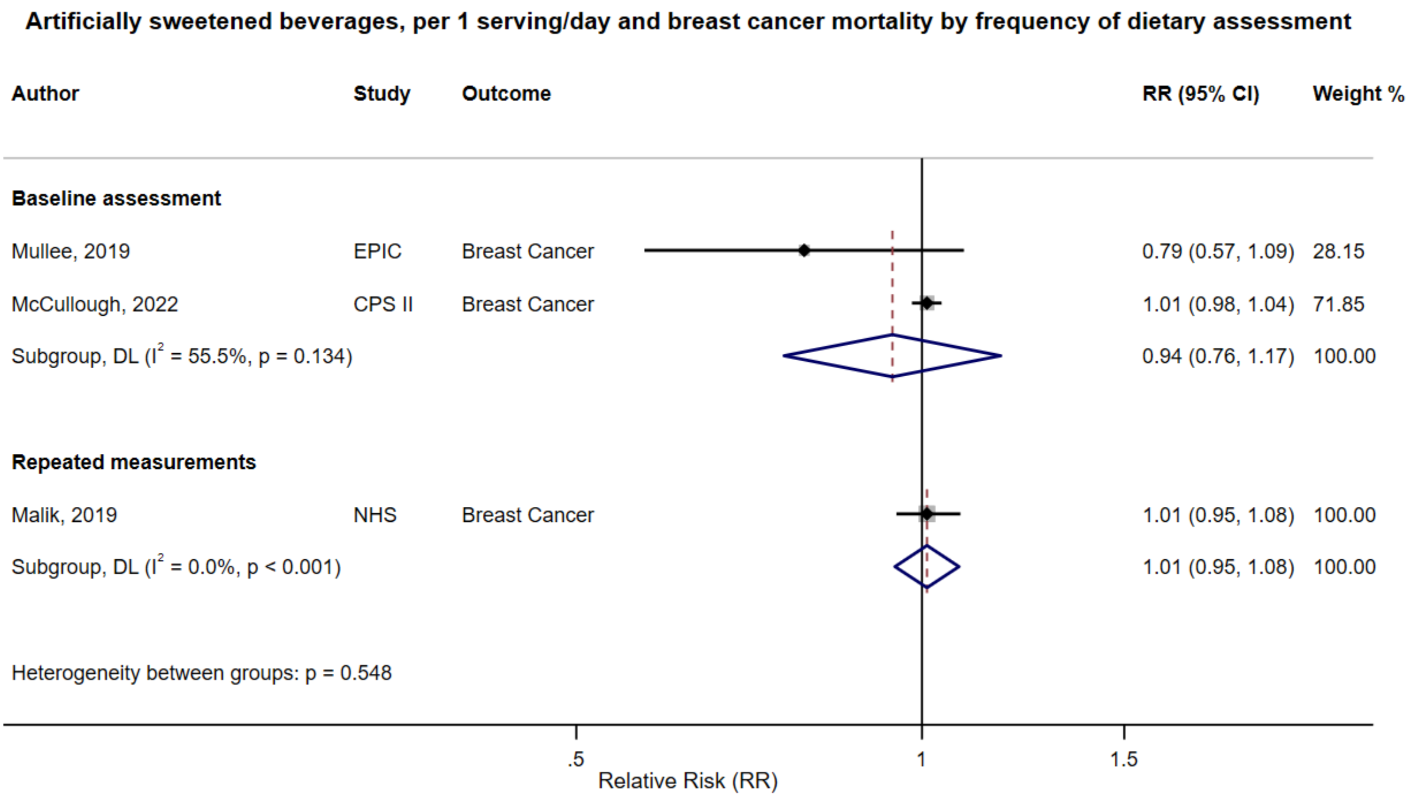

Figure legend: Forest plot shows the linear dose-response results (per 1 serving [355 mL] of artificially sweetened beverages/day) from the inverse variance DerSimonian-Laird random-effects model. The diamond represents the summary relative risk (RR) estimate, the diamond's width is the 95% confidence interval (CI). Each square and the horizontal line across the square represent the RR estimate of the individual study and its 95% CI.

Abbreviations: CI, confidence interval; RR, relative risk.

Study abbreviations: EPIC, European Prospective Investigation into Cancer and nutrition; CPS II, Cancer Prevention Study II; NHS, Nurses' Health Study

Mullee, 2019[126], McCullough, 2022[113], Malik, 2019[109]

**Supplementary Figure 236. Linear dose-response meta-analysis of artificially sweetened beverages and prostate cancer mortality by frequency of dietary assessment.**

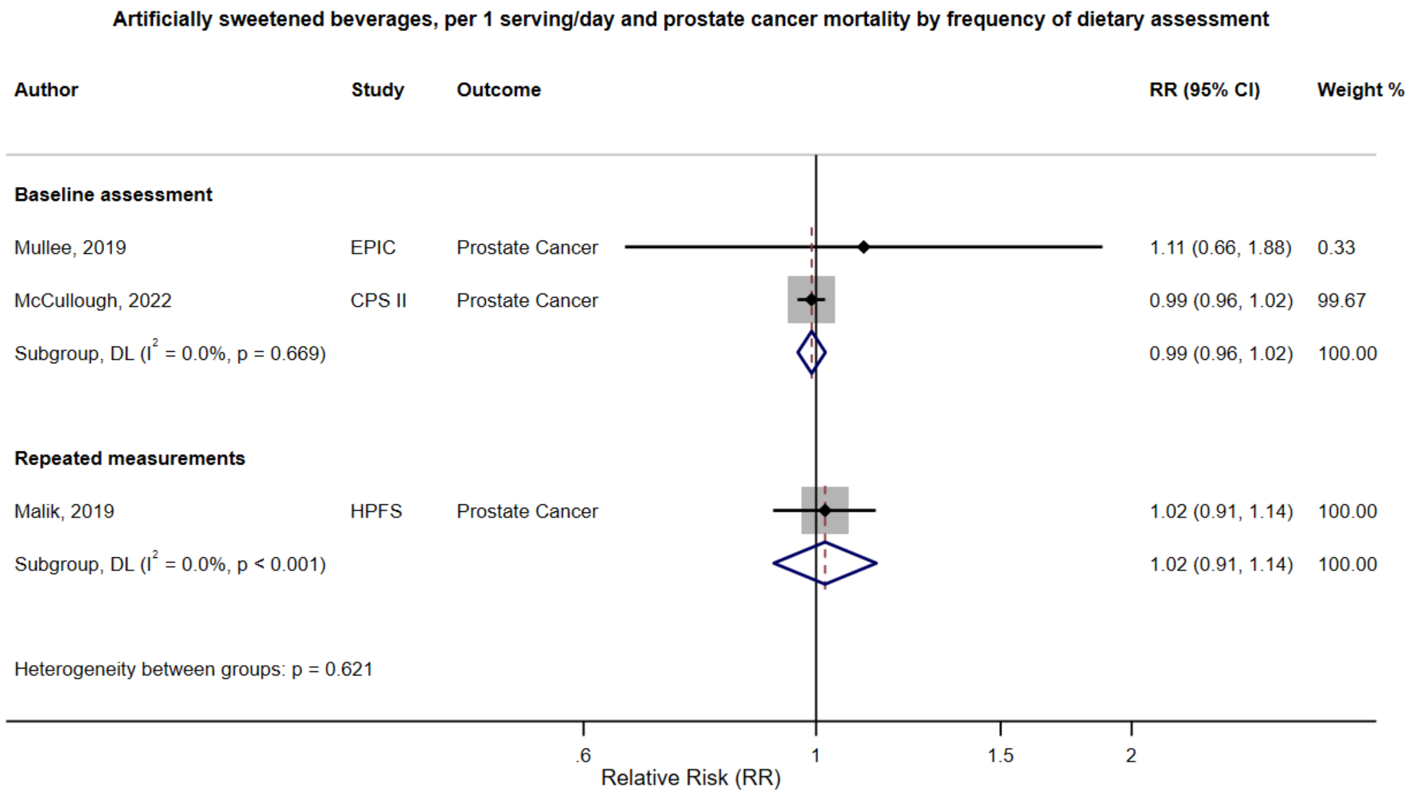

Figure legend: Forest plot shows the linear dose-response results (per 1 serving [355 mL] of artificially sweetened beverages/day) from the inverse variance DerSimonian-Laird random-effects model. The diamond represents the summary relative risk (RR) estimate, the diamond's width is the 95% confidence interval (CI). Each square and the horizontal line across the square represent the RR estimate of the individual study and its 95% CI.

Abbreviations: CI, confidence interval; RR, relative risk.

Study abbreviations: EPIC, European Prospective Investigation into Cancer and nutrition; CPS II, Cancer Prevention Study II; HPFS, Health Professionals Follow-up Study

Mullee, 2019[126], McCullough, 2022[113], Malik, 2019[109]

**Supplementary Figure 237. Linear dose-response meta-analysis of artificially sweetened beverages and breast cancer mortality by geographical region.**

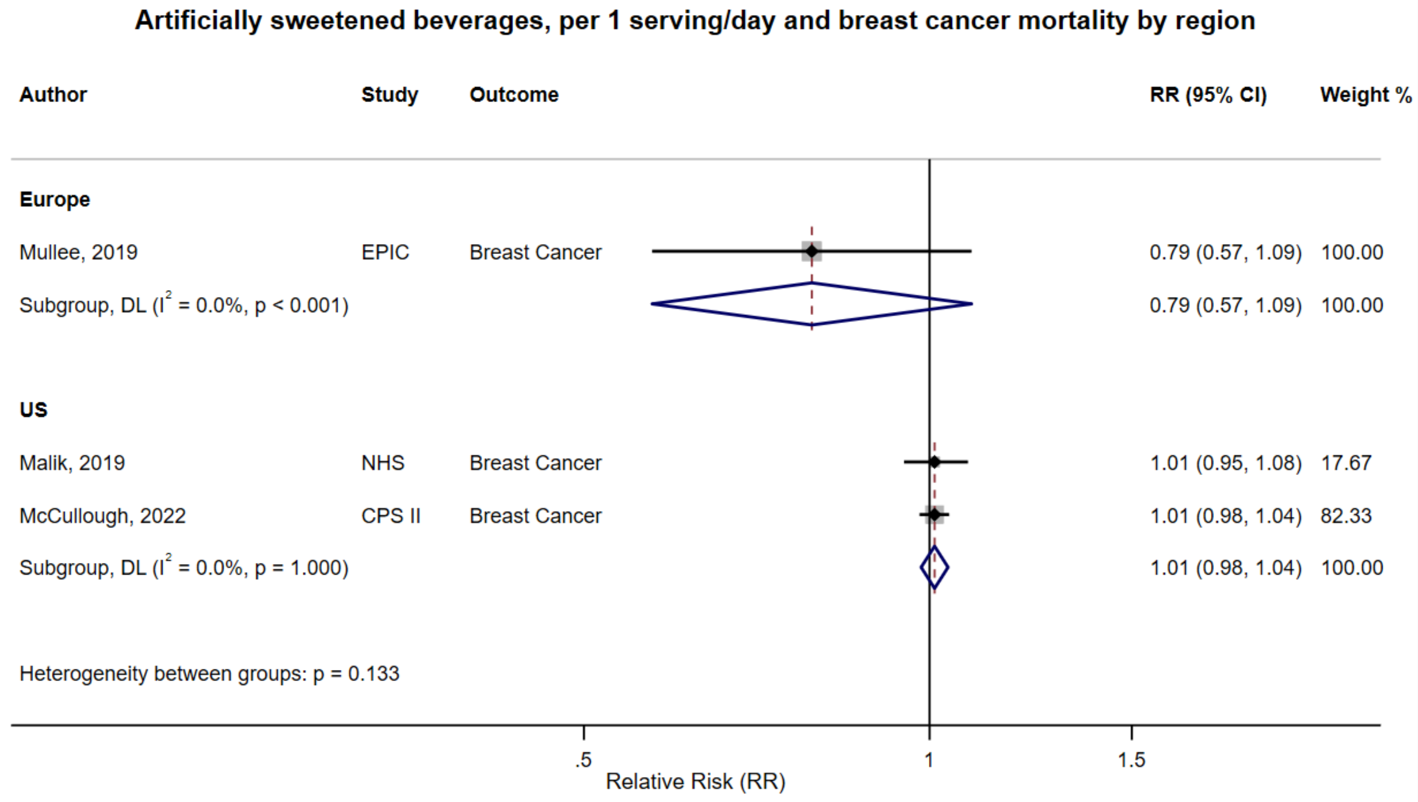

Figure legend: Forest plot shows the linear dose-response results (per 1 serving [355 mL] of artificially sweetened beverages/day) from the inverse variance DerSimonian-Laird random-effects model. The diamond represents the summary relative risk (RR) estimate, the diamond's width is the 95% confidence interval (CI). Each square and the horizontal line across the square represent the RR estimate of the individual study and its 95% CI.

Abbreviations: CI, confidence interval; RR, relative risk.

Study abbreviations: EPIC, European Prospective Investigation into Cancer and nutrition; NHS, Nurses' Health Study; CPS II, Cancer Prevention Study II

Mullee, 2019[126], Malik, 2019[109], McCullough, 2022[113]

**Supplementary Figure 238. Linear dose-response meta-analysis of artificially sweetened beverages and prostate cancer mortality by geographical region.**

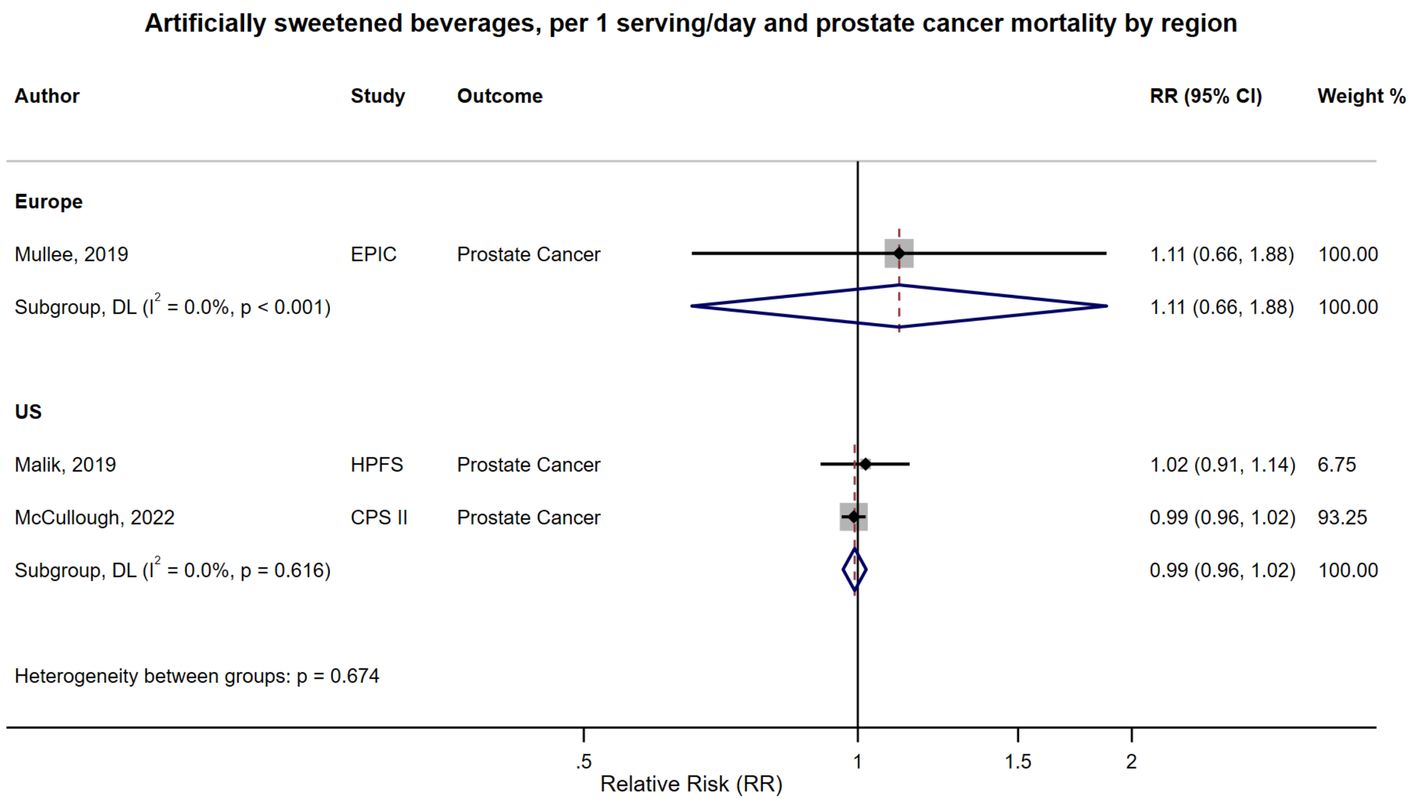

Figure legend: Forest plot shows the linear dose-response results (per 1 serving [355 mL] of artificially sweetened beverages/day) from the inverse variance DerSimonian-Laird random-effects model. The diamond represents the summary relative risk (RR) estimate, the diamond's width is the 95% confidence interval (CI). Each square and the horizontal line across the square represent the RR estimate of the individual study and its 95% CI.

Abbreviations: CI, confidence interval; RR, relative risk.

Study abbreviations: EPIC, European Prospective Investigation into Cancer and nutrition; HPFS, Health Professionals Follow-up Study; CPS II, Cancer Prevention Study II

Mullee, 2019[126], Malik, 2019[109], McCullough, 2022[113]

**Supplementary Figure 239. Linear dose-response meta-analysis of artificially sweetened beverages and prostate cancer mortality by exclusion of people with diabetes at baseline.**

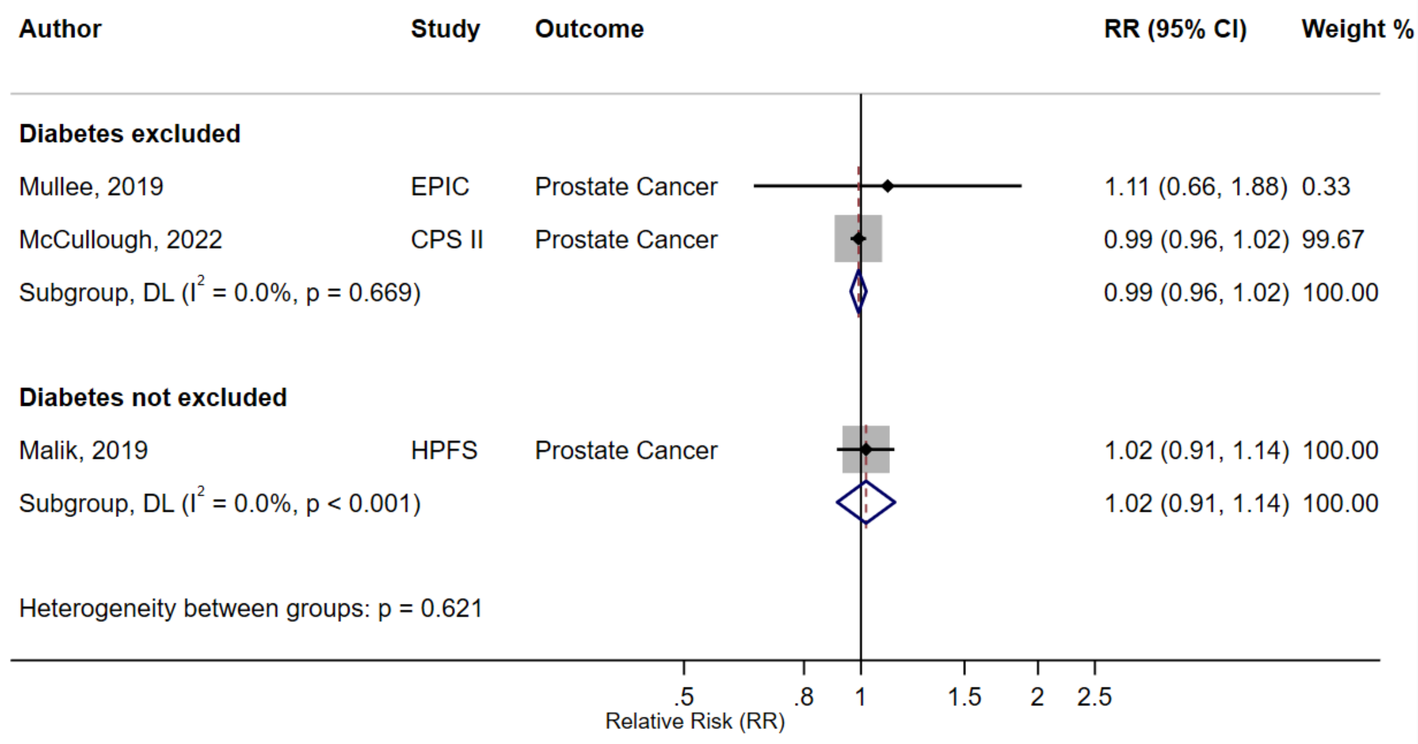

Figure legend: Forest plot shows the linear dose-response results (per 1 serving [355 mL] of artificially sweetened beverages/day) from the inverse variance DerSimonian-Laird random-effects model. The diamond represents the summary relative risk (RR) estimate, the diamond's width is the 95% confidence interval (CI). Each square and the horizontal line across the square represent the RR estimate of the individual study and its 95% CI.

Abbreviations: CI, confidence interval; RR, relative risk.

Study abbreviations: EPIC, European Prospective Investigation into Cancer and nutrition; CPS II, Cancer Prevention Study II; HPFS, Health Professionals Follow-up Study

Mullee, 2019[126], McCullough, 2022[113], Malik, 2019[109]

**Supplementary Figure 240. Linear dose-response meta-analysis of artificially sweetened beverages and breast cancer mortality, leave-one-out analysis.**

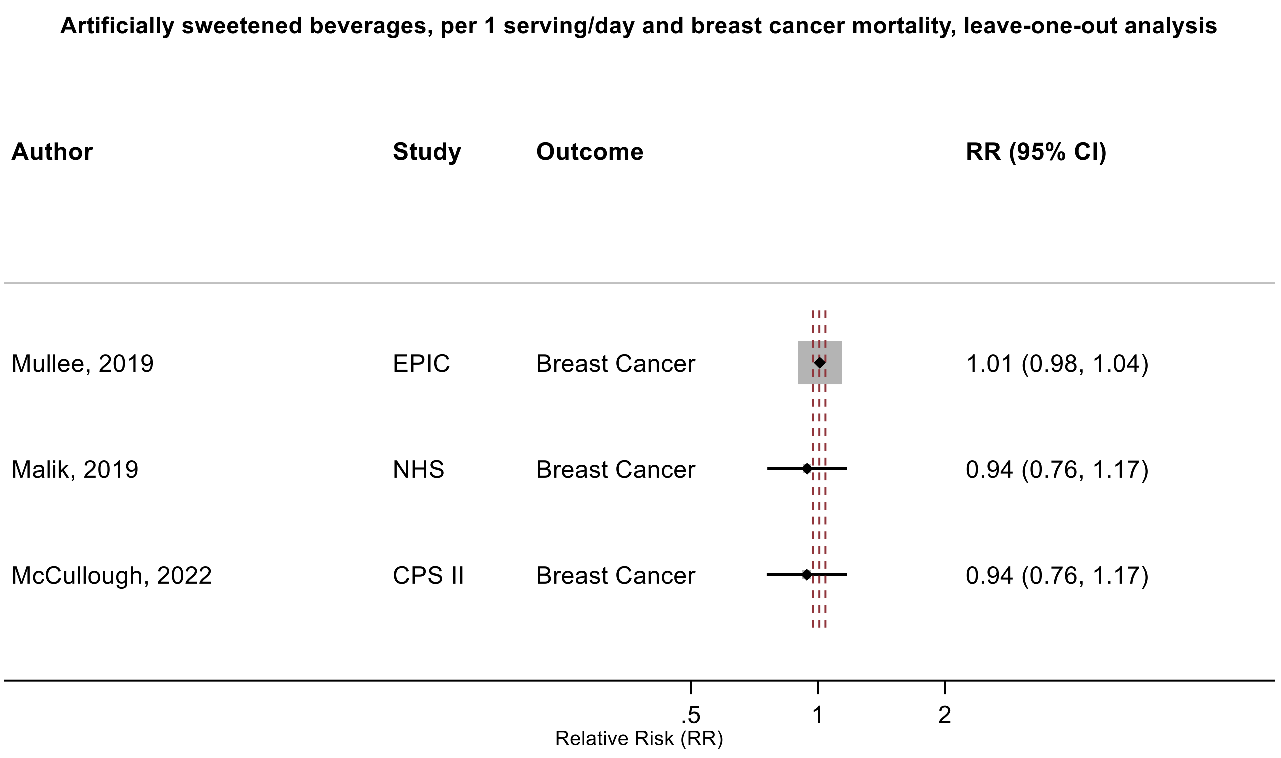

Figure legend: Forest plot shows the linear dose-response results (per 1 serving [355 mL] of artificially sweetened beverages/day) from the inverse variance DerSimonian-Laird random-effects model after exclusion of every single study at a time.

Abbreviations: CI, confidence interval; RR, relative risk.

Study abbreviations: : EPIC, European Prospective Investigation into Cancer and nutrition; NHS, Nurses' Health Study; CPS II, Cancer Prevention Study II

Mullee, 2019[126], Malik, 2019[109], McCullough, 2022[113]

**Supplementary Figure 241. Linear dose-response meta-analysis of artificially sweetened beverages and prostate cancer mortality, leave-one-out analysis.**

**Artificially sweetened beverages, per 1 serving/day and prostate cancer mortality, leave-one-out analysis**

| Author | Study | Outcome | RR (95% CI) |
|---|---|---|---|
| Mullee, 2019 | EPIC | Prostate Cancer | 0.99 (0.96, 1.02) |
| Malik, 2019 | HPFS | Prostate Cancer | 0.99 (0.96, 1.02) |
| McCullough, 2022 | CPS II | Prostate Cancer | 1.02 (0.92, 1.14) |

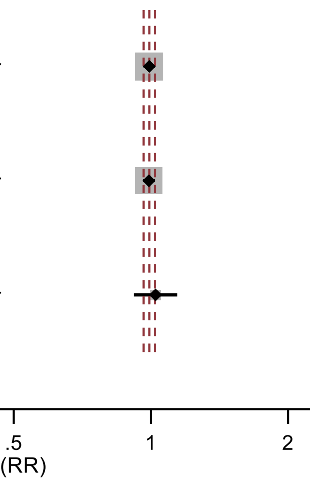

Figure legend: Forest plot shows the linear dose-response results (per 1 serving [355 mL] of artificially sweetened beverages/day) from the inverse variance DerSimonian-Laird random-effects model after exclusion of every single study at a time.

Abbreviations: CI, confidence interval; RR, relative risk.

Study abbreviations: EPIC, European Prospective Investigation into Cancer and nutrition; HPFS, Health Professionals Follow-up Study; CPS II, Cancer Prevention Study II

Mullee, 2019[126], Malik, 2019[109], McCullough, 2022[113]

**Supplementary Figure 242. Fixed effect linear dose-response meta-analysis of fruit juice and prostate cancer incidence.**

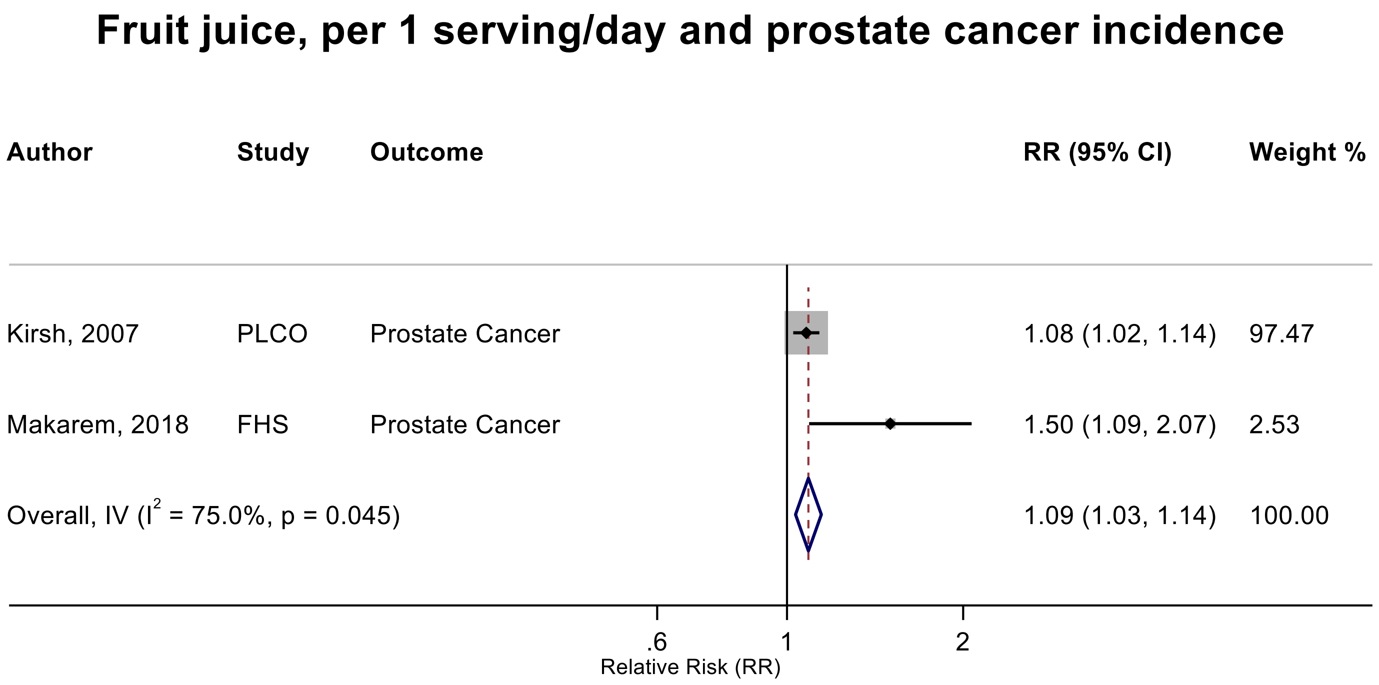

Figure legend: Forest plot shows the linear dose-response results (per 1 serving [177 mL] of juice/day) from the inverse variance model. The diamond represents the summary relative risk (RR) estimate, the diamond's width is the 95% confidence interval (CI), and the diamond's horizontal line is the 95% prediction interval (PI). Each square and the horizontal line across the square represent the RR estimate of the individual study and its 95% CI.

Abbreviations: CI, confidence interval; RR, relative risk.

Study abbreviations: PLCO, Prostate, Lung, Colorectal and Ovarian Cancer Screening trial; FHS, Framingham Heart Study

Note: The pooled RR and 95%CI of prostate cancer from the random effects linear dose-response meta-analysis are 1.22 (95%CI: 0.89 – 1.68) (Supplementary Figure 245). Both studies reported significant positive associations, but due to heterogeneity in the data, the random-effects meta-analysis indicated non-significant results. The random-effects model gives more weight to the smaller study with lower number of cases. Therefore, we considered fixed-effect meta-analysis as the main analysis.

Kirsh, 2007[89], Makarem, 2018[108]

**Supplementary Figure 243. Linear dose-response meta-analysis of fruit juice and prostate cancer incidence.**

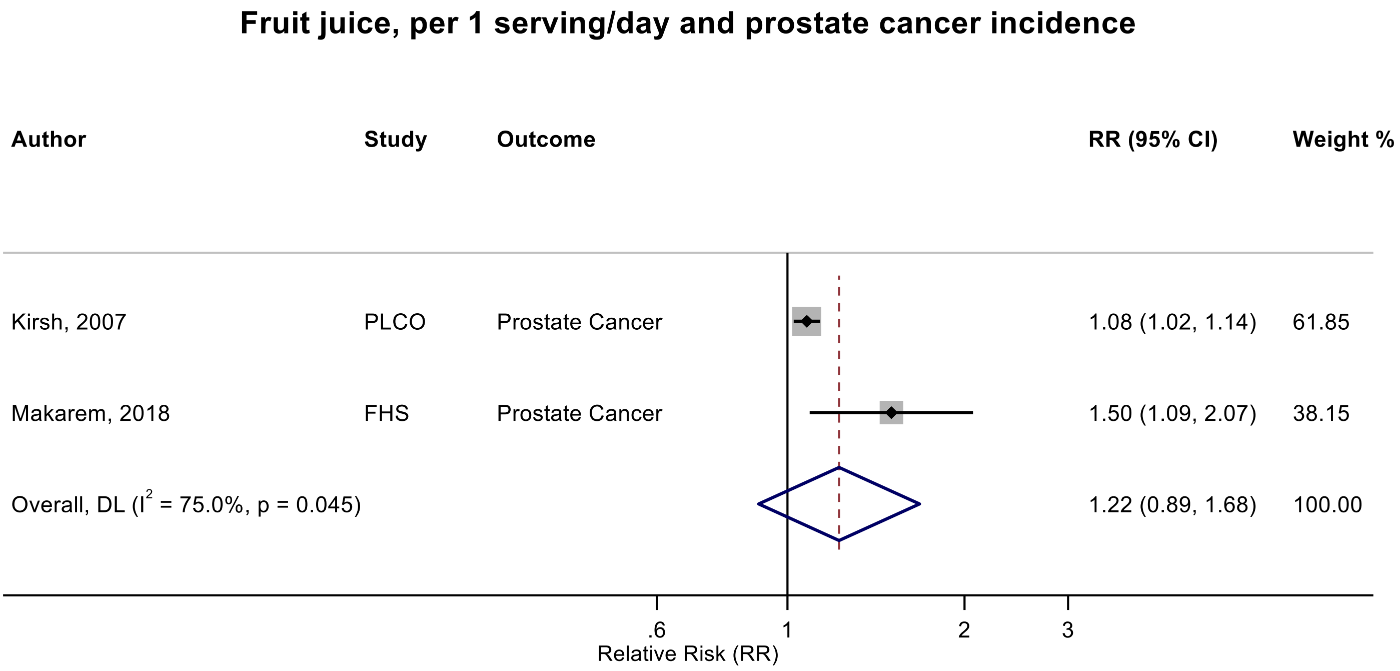

Figure legend: Forest plot shows the linear dose-response results (per 1 serving [177 mL] of juice/day) from the inverse variance DerSimonian-Laird random-effects model. The diamond represents the summary relative risk (RR) estimate, the diamond's width is the 95% confidence interval (CI). Each square and the horizontal line across the square represent the RR estimate of the individual study and its 95% CI. Both studies reported significant positive associations, but due to heterogeneity in the data, the random-effects meta-analysis indicated non-significant result. Therefore, we considered fixed-effect meta-analysis (Supplementary Figure 244) as the main analysis.

Abbreviations: CI, confidence interval; RR, relative risk.

Study abbreviations: PLCO, Prostate, Lung, Colorectal and Ovarian Cancer Screening trial; FHS, Framingham Heart Study

Note: Two other publications that could not be included in the dose-response meta-analysis because of insufficient data format reported on fruit juice and prostate cancer incidence: the Multiethnic Cohort Study[161] reported a positive association but with 95%CI crossing the null value when comparing extreme categories of intake ($RR_{\geq 64.9 \text{ vs } \leq 2.5 \text{ g/1000 kcal}}$: 1.09, 95%CI: 0.98-1.20); and the Malmö Diet and Cancer cohort[47] reported limited evidence of an association ($RR_{200 \text{ vs } 0 \text{ g/day}}$: 0.99, 95%CI: 0.81-1.22). So, we performed a supporting categorical high vs low meta-analysis (see Supplementary Figure 246).

Kirsh, 2007[89], Makarem, 2018[108]

**Supplementary Figure 244. Categorical highest versus the lowest meta-analysis of fruit juice and prostate cancer incidence.**

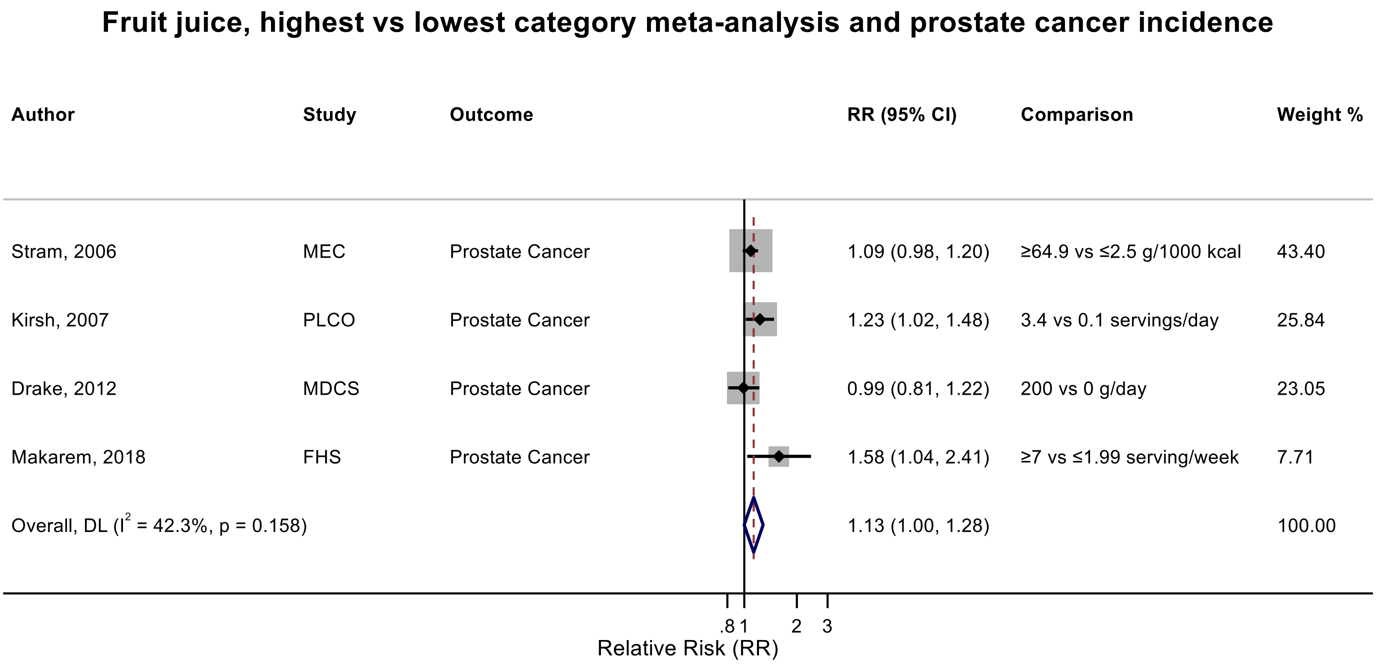

Figure legend: Forest plot shows the highest versus the lowest category results of fruit juice from the inverse variance DerSimonian-Laird random-effects model. The diamond represents the summary relative risk (RR) estimate, the diamond's width is the 95% confidence interval (CI). Each square and the horizontal line across the square represent the RR estimate of the individual study and its 95% CI.

Abbreviations: CI, confidence interval; RR, relative risk.

Study abbreviations: MEC, Multiethnic Cohort Study; PLCO, Prostate, Lung, Colorectal and Ovarian Cancer Screening trial; MDCS, Malmö Diet and Cancer Study; FHS, Framingham Heart Study

Note: The pooled RR and 95%CI with 3 decimal point are 1.132 (0.999-1.282).

Stram, 2006[139], Kirsh, 2007[89], Drake, 2012[47], Makarem, 2018[108]

#### Supplementary Figure 245. Fixed effect linear dose-response meta-analysis of fruit juice and pure fruit juice combined and prostate cancer incidence.

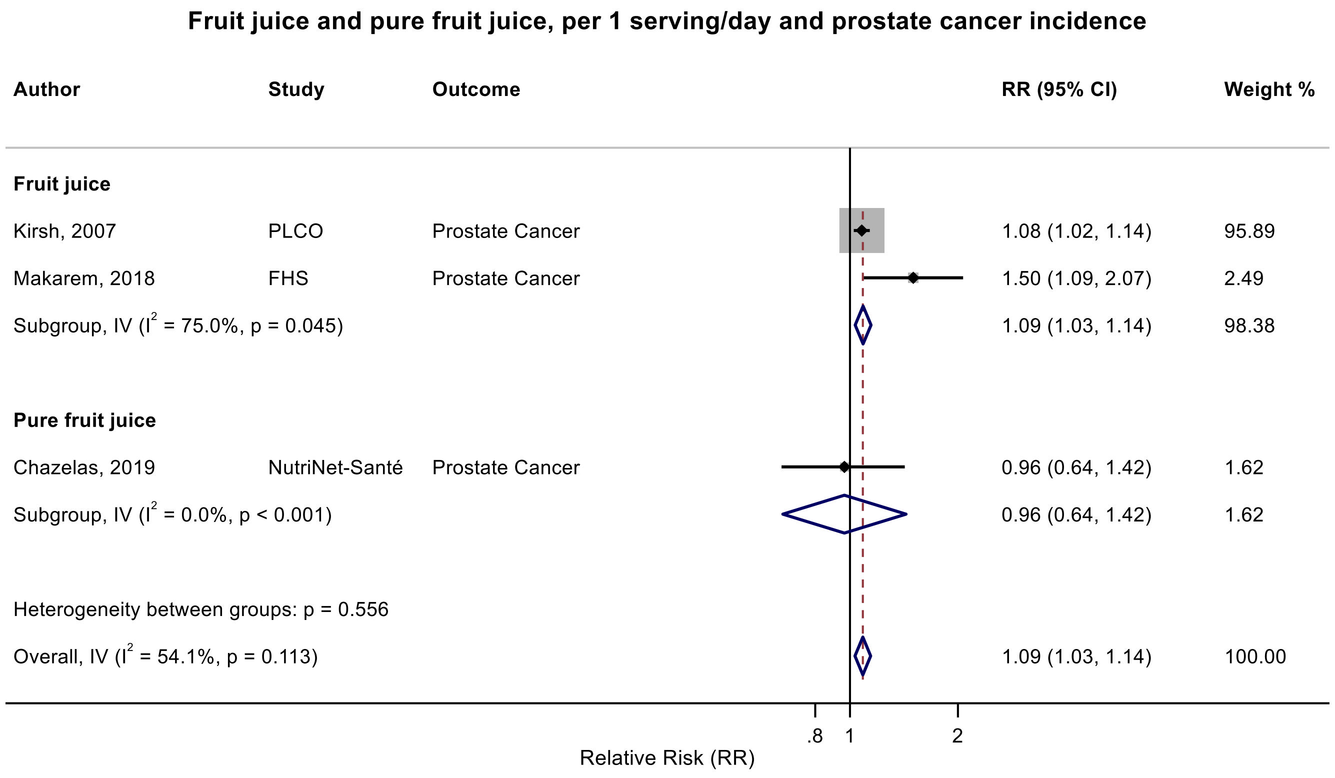

Figure legend: Forest plot shows the linear dose-response results (per 1 serving [177 mL] of juice/day) from the inverse variance DerSimonian-Laird random-effects model. The diamond represents the summary relative risk (RR) estimate, the diamond's width is the 95% confidence interval (CI). Each square and the horizontal line across the square represent the RR estimate of the individual study and its 95% CI.

Abbreviations: CI, confidence interval; RR, relative risk.

Study abbreviations: PLCO, Prostate, Lung, Colorectal and Ovarian Cancer Screening trial; FHS, Framingham Heart Study

Kirsh, 2007[89], Makarem, 2018[108], Chazelas, 2019[44]

Note: We used a fixed-effect meta-analysis for consistency with the meta-analysis of fruit juice and prostate cancer incidence.

**Supplementary Figure 246. Categorical highest versus the lowest meta-analysis of fruit juice and high-grade/high-risk/aggressive prostate cancer incidence.**

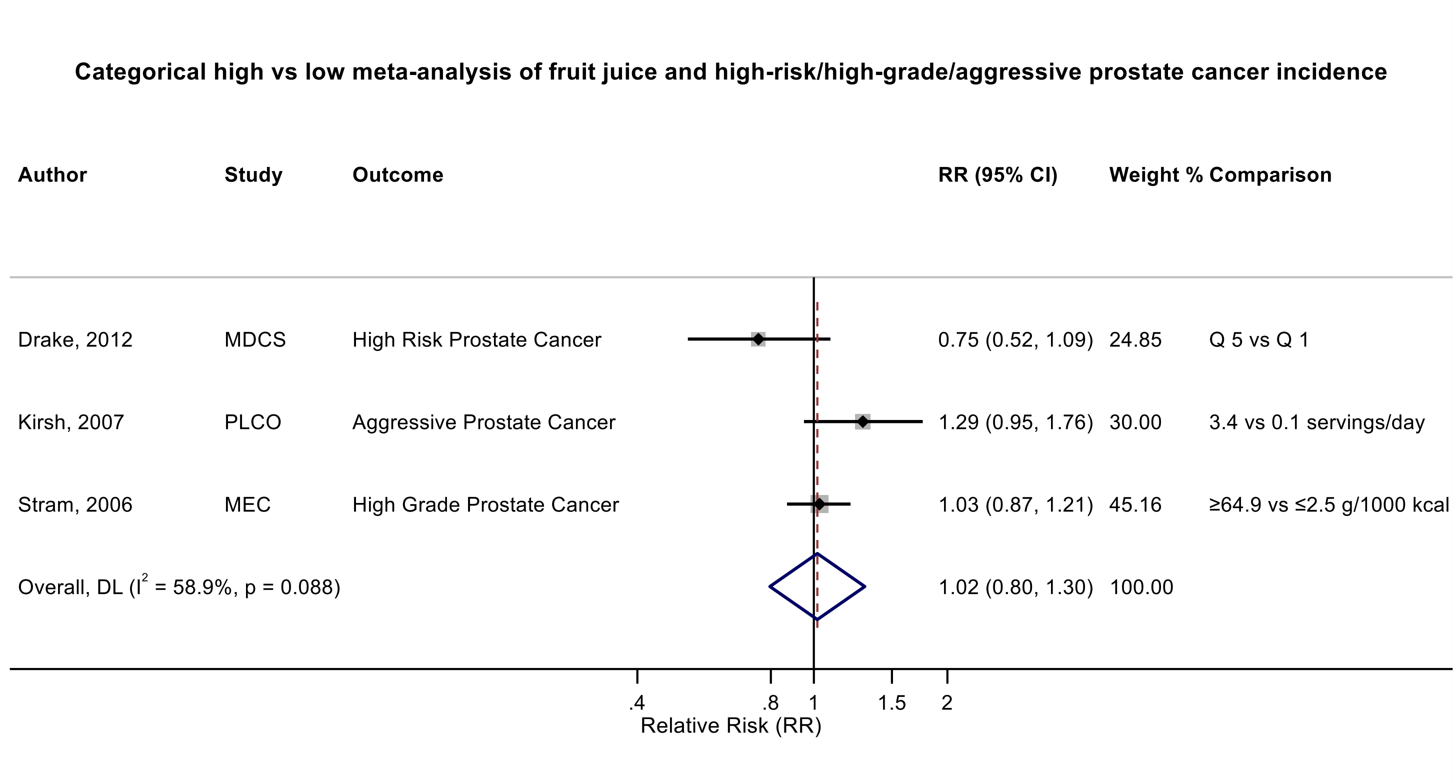

Figure legend: Forest plot shows the highest versus the lowest category results of pure fruit juice from the inverse variance DerSimonian-Laird random-effects model. The diamond represents the summary relative risk (RR) estimate, the diamond's width is the 95% confidence interval (CI). Each square and the horizontal line across the square represent the RR estimate of the individual study and its 95% CI.

Abbreviations: CI, confidence interval; RR, relative risk.

Study abbreviations: MDCS, Malmo Diet and Cancer Study; MEC, Multi-ethnic Cohort; PLCO, Prostate , Lung, Colorectal and Ovarian cancer Screening Trial

Drake, 2012[79], Kirsh, 2007[89], Stram, 2006[161]

Note: This analysis was performed as an additional analysis combining high grade, aggressive, and advanced prostate cancer cases.

**Supplementary Figure 247. Linear dose-response meta-analysis of fruit and vegetable juices and prostate cancer incidence.**

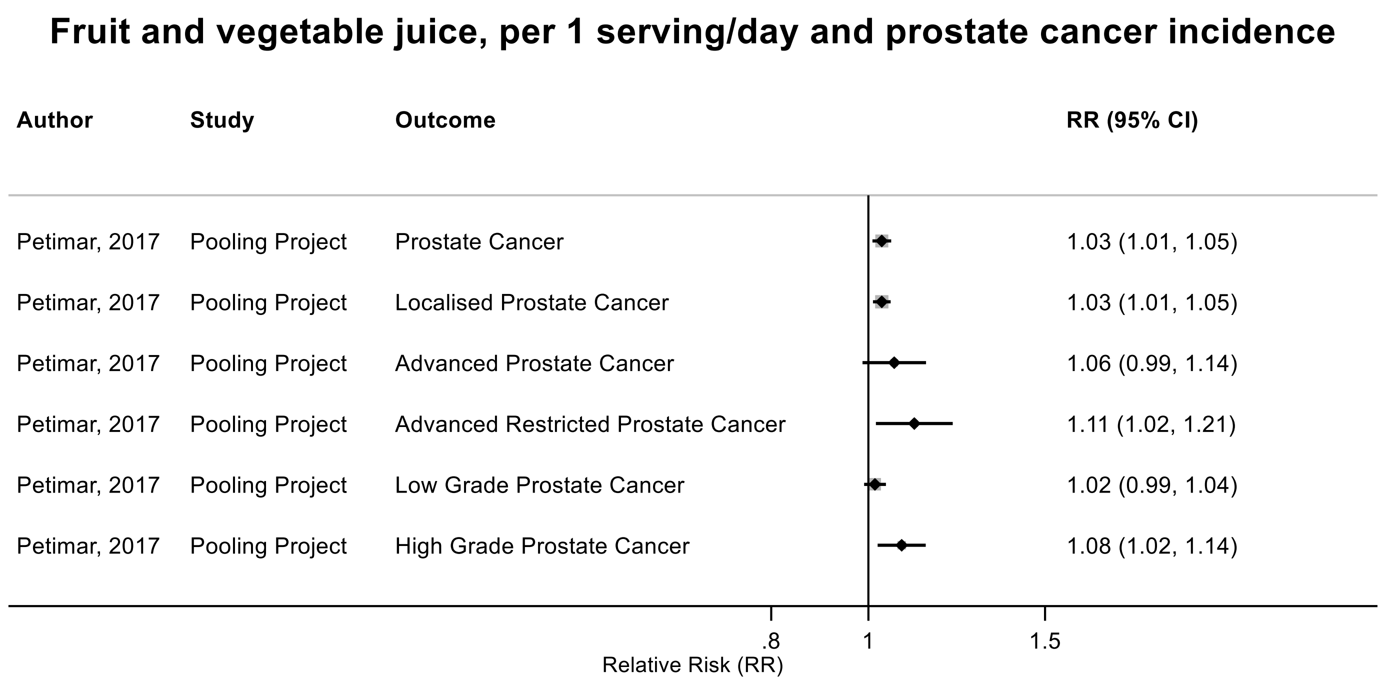

Figure legend: Forest plot shows the linear dose-response results (per 1 serving [177 mL] of juice/day) from the inverse variance DerSimonian-Laird random-effects model. Each square represents the summary relative risk (RR) estimate and the square's horizontal line is the 95% confidence interval (CI).

Abbreviations: CI, confidence interval; RR, relative risk.

Study abbreviations: Pooling Project, The Pooling Project of Prospective Studies of Diet and Cancer (n=15).

Petimar, 2017[139]

**Supplementary Figure 248. Linear dose-response meta-analysis of fruit juice and head and neck cancers incidence.**

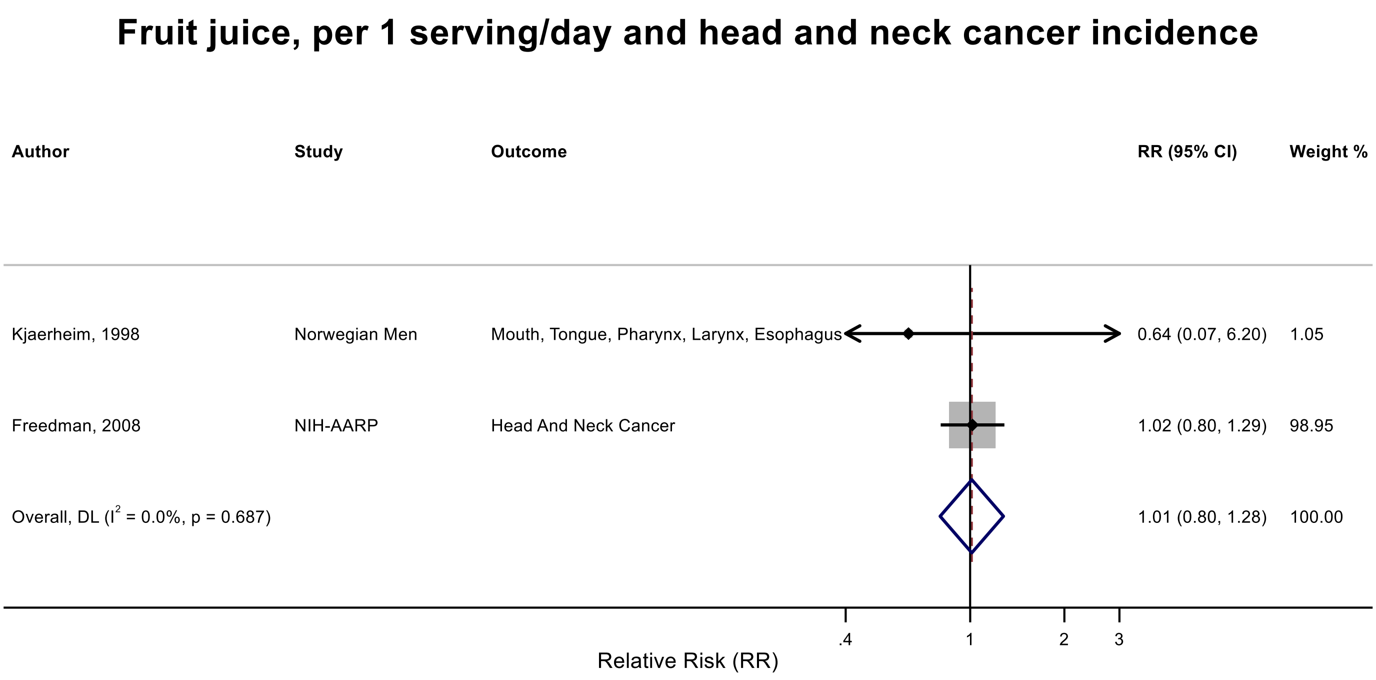

Figure legend: Forest plot shows the linear dose-response results (per 1 serving [177 mL] of juice/day) from the inverse variance DerSimonian-Laird random-effects model. The diamond represents the summary relative risk (RR) estimate, the diamond's width is the 95% confidence interval (CI). Each square and the horizontal line across the square represent the RR estimate of the individual study and its 95% CI.

Abbreviations: CI, confidence interval; RR, relative risk.

Study abbreviations: NIH-AARP, National Institute of Health + American Association of Retired Persons Diet and Health Study

Kjaerheim, 1998[90], Freedman, 2008[59]

**Supplementary Figure 249. Linear dose-response meta-analysis of fruit juice and colorectal cancer incidence.**

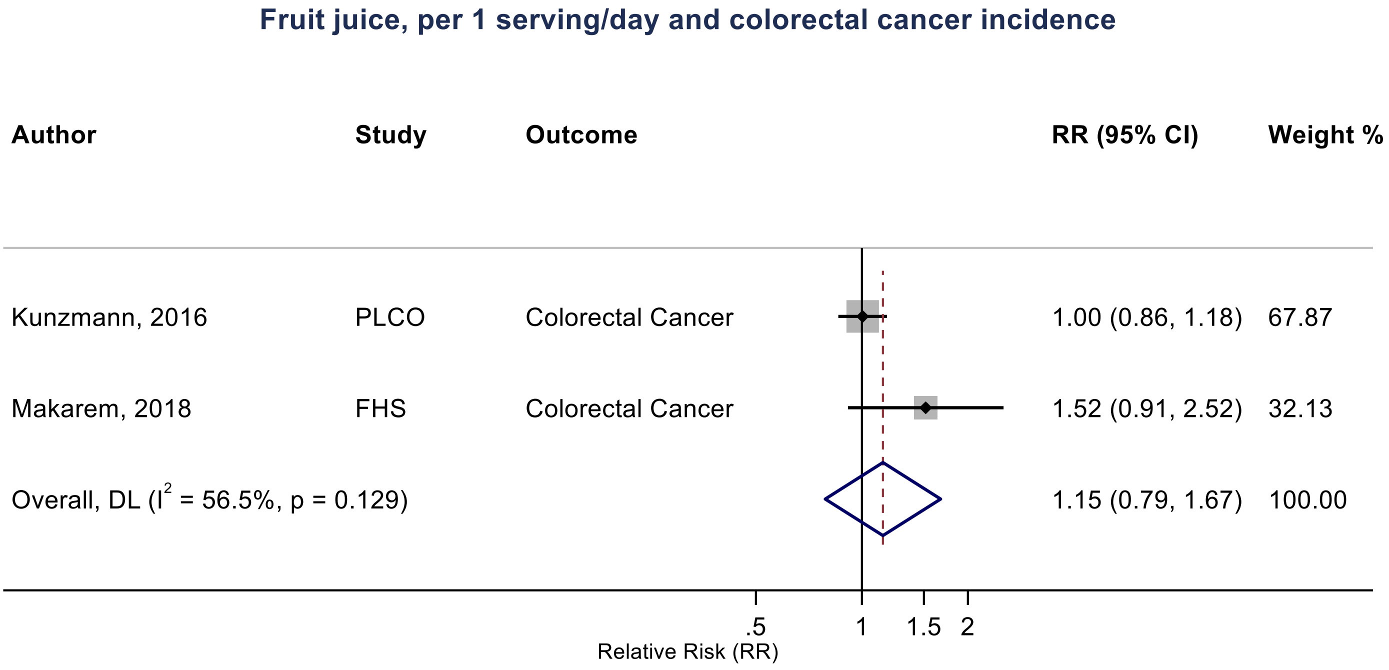

Figure legend: Forest plot shows the linear dose-response results (per 1 serving [177 mL] of juice/day) from the inverse variance DerSimonian-Laird random-effects model. The diamond represents the summary relative risk (RR) estimate, the diamond's width is the 95% confidence interval (CI). Each square and the horizontal line across the square represent the RR estimate of the individual study and its 95% CI.

Abbreviations: CI, confidence interval; RR, relative risk.

Study abbreviations: PLCO, Prostate, Lung, Colorectal and Ovarian Cancer Screening trial; FHS, Framingham Heart Study

Note: There were two other studies that could not be included in the dose-response meta-analysis because of insufficient data format: one publication from UK Biobank [78] that reported little evidence of an association (RR: 1.01, 95%CI: 0.96-1.06) per each one standard deviation increase in fruit juice intake, and one publication from Multiethnic Cohort Study[87] that reported a borderline inverse association per each one standard deviation increase in fruit juice intake (RR: 0.96, 95%CI: 0.92-1.00).

Kunzmann, 2016[93], Makarem, 2018[108]

**Supplementary Figure 250. Linear dose-response meta-analysis of pure fruit juice and colorectal cancer incidence.**

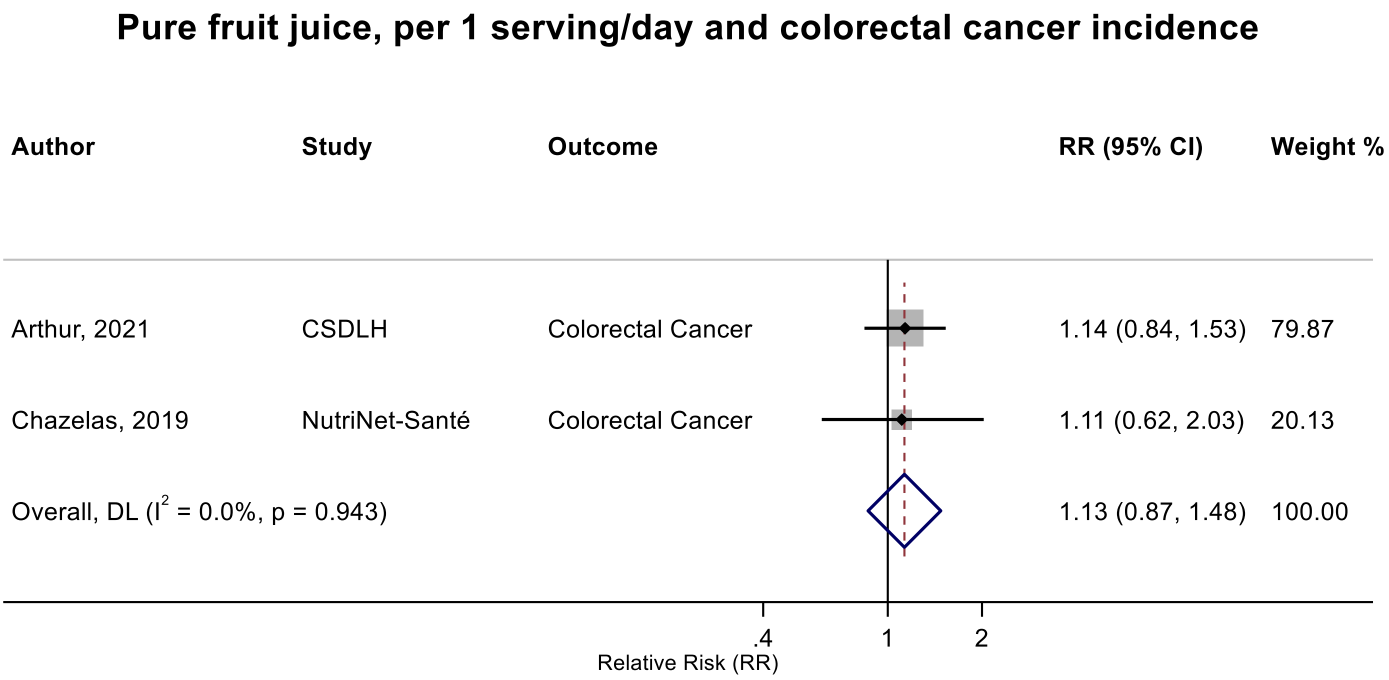

Figure legend: Forest plot shows the linear dose-response results (per 1 serving [177 mL] of juice/day) from the inverse variance DerSimonian-Laird random-effects model. The diamond represents the summary relative risk (RR) estimate, the diamond's width is the 95% confidence interval (CI). Each square and the horizontal line across the square represent the RR estimate of the individual study and its 95% CI.

Abbreviations: CI, confidence interval; RR, relative risk.

Study abbreviations: CSDLH, Canadian Study of Diet, Lifestyle and Health

Arthur, 2021[38], Chazelas, 2019[44]

**Supplementary Figure 251. Linear dose-response meta-analysis of fruit juice and pure fruit juice combined and colorectal cancer incidence.**

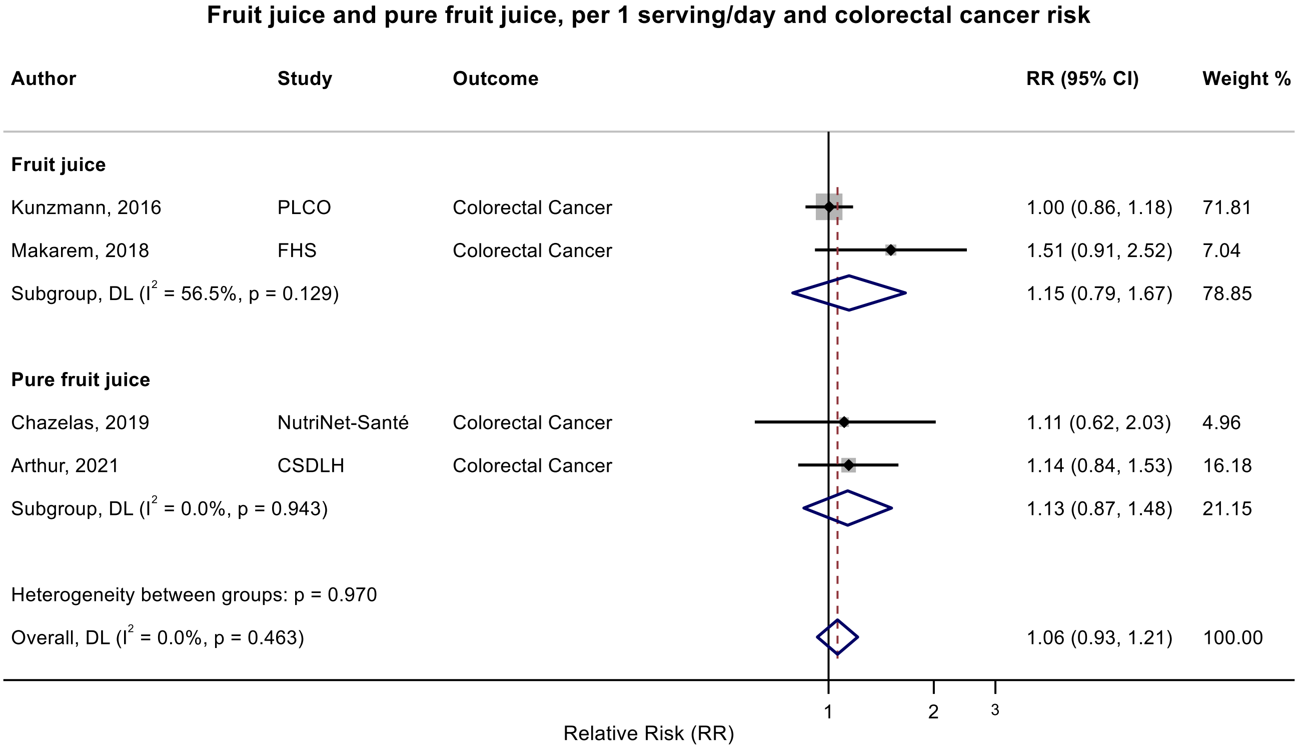

Figure legend: Forest plot shows the linear dose-response results (per 1 serving [177 mL] of juice/day) from the inverse variance DerSimonian-Laird random-effects model. The diamond represents the summary relative risk (RR) estimate, the diamond's width is the 95% confidence interval (CI). Each square and the horizontal line across the square represent the RR estimate of the individual study and its 95% CI.

Abbreviations: CI, confidence interval; RR, relative risk.

Study abbreviations: PLCO, Prostate, Lung, Colorectal and Ovarian Cancer Screening trial; FHS, Framingham Heart Study; CSDLH, Canadian Study of Diet, Lifestyle and Health

Kunzmann, 2016[93], Makarem, 2018[108], Arthur, 2021[38], Chazelas, 2019[44]

**Supplementary Figure 252. Linear dose-response meta-analysis of fruit juice and pure fruit juice combined and liver cancer incidence/mortality.**

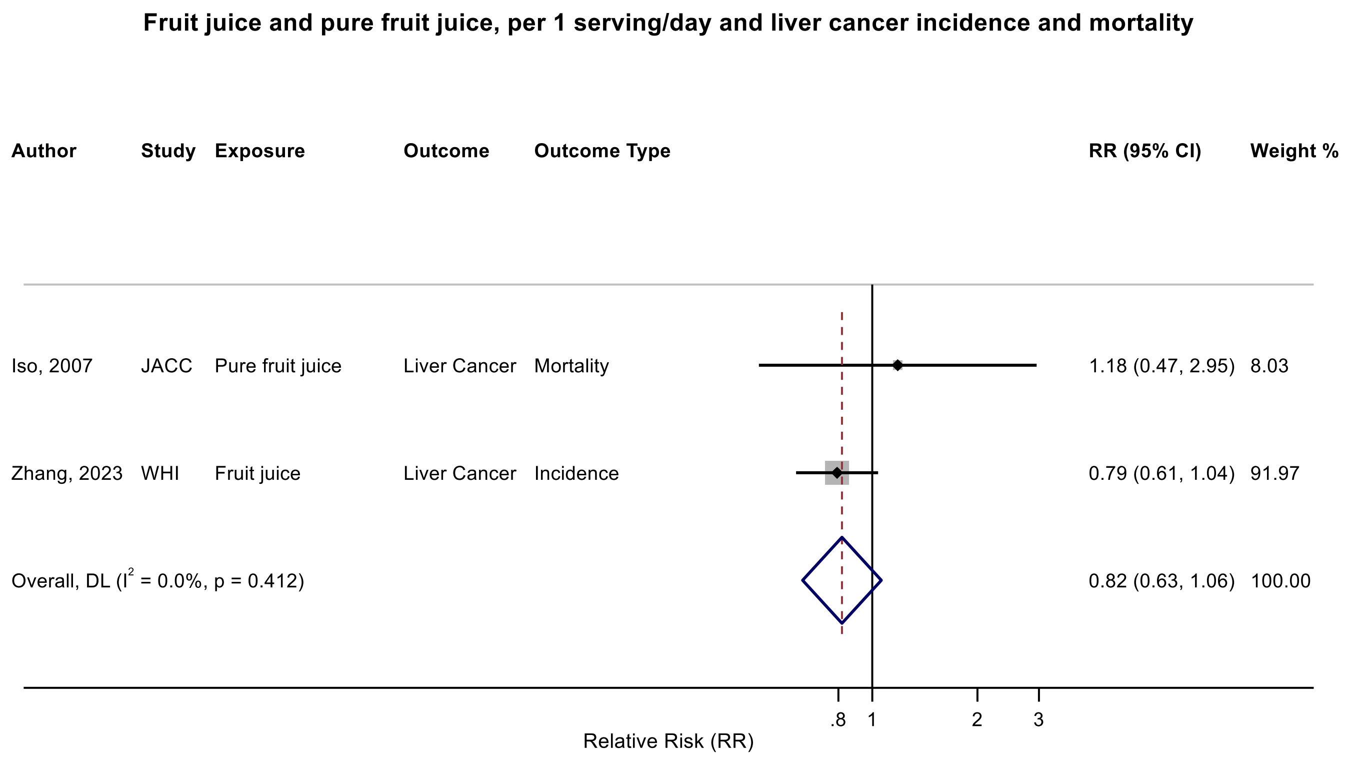

Figure legend: Forest plot shows the linear dose-response results (per 1 serving [177 mL] of juice/day) from the inverse variance DerSimonian-Laird random-effects model. The diamond represents the summary relative risk (RR) estimate, the diamond's width is the 95% confidence interval (CI). Each square and the horizontal line across the square represent the RR estimate of the individual study and its 95% CI.

Abbreviations: CI, confidence interval; RR, relative risk.

Study abbreviations: JACC, Japan Collaborative Cohort study; WHI, Women’s Health Initiative

Iso, 2007[76], Zhang, 2023[183]

**Supplementary Figure 253. Linear dose-response meta-analysis of fruit juice and pancreatic cancer incidence.**

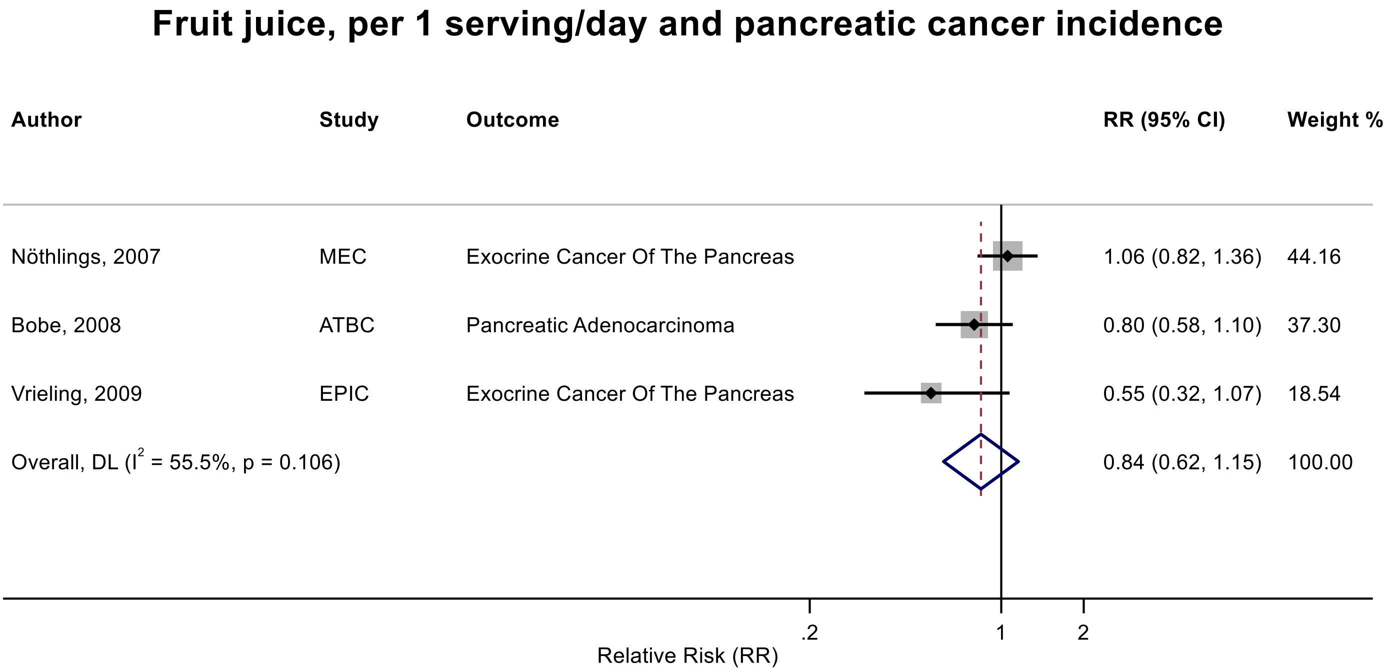

Figure legend: Forest plot shows the linear dose-response results (per 1 serving [177 mL] of juice/day) from the inverse variance DerSimonian-Laird random-effects model. The diamond represents the summary relative risk (RR) estimate, the diamond's width is the 95% confidence interval (CI). Each square and the horizontal line across the square represent the RR estimate of the individual study and its 95% CI.

Abbreviations: CI, confidence interval; RR, relative risk.

Study abbreviations: MEC, Multiethnic Cohort Study; ATBC, Alpha-Tocopherol, Beta-Carotene Cancer Prevention Study; EPIC, European Prospective Investigation into Cancer and nutrition

Note: There was another study (PLCO)[188] that could not be included in the dose-response meta-analysis because of insufficient data format and reported a positive association with pancreatic cancer when comparing extreme categories of fruit juice intake as a component of unhealthful plant-based dietary index ($RR_{Q4 \text{ vs } Q1}$: 1.76, 95%CI: 1.29-2.40).

Nöthlings, 2007[131], Bobe, 2008[41], Vrieling, 2009[167]

**Supplementary Figure 254. Categorical highest versus the lowest meta-analysis of pure fruit juice and pancreatic cancer incidence.**

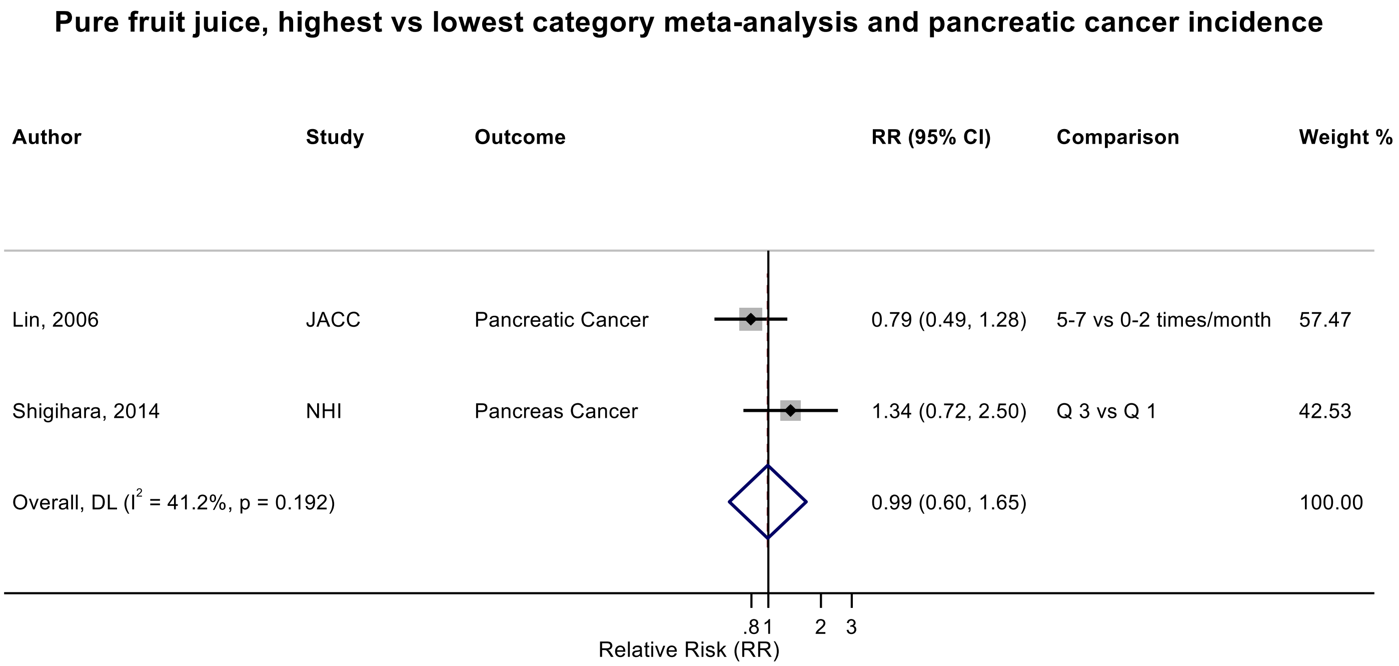

Figure legend: Forest plot shows the highest versus the lowest category results of pure fruit juice from the inverse variance DerSimonian-Laird random-effects model. The diamond represents the summary relative risk (RR) estimate, the diamond's width is the 95% confidence interval (CI). Each square and the horizontal line across the square represent the RR estimate of the individual study and its 95% CI.

Abbreviations: CI, confidence interval; RR, relative risk.

Study abbreviations: JACC, Japan Collaborative Cohort study; NHI, Ohsaki National Health Insurance Cohort Study

Lin, 2006[103], Shigihara, 2014[154]

**Supplementary Figure 255. Linear dose-response meta-analysis of fruit juice and pure fruit juice combined and pancreatic cancer incidence/mortality.**

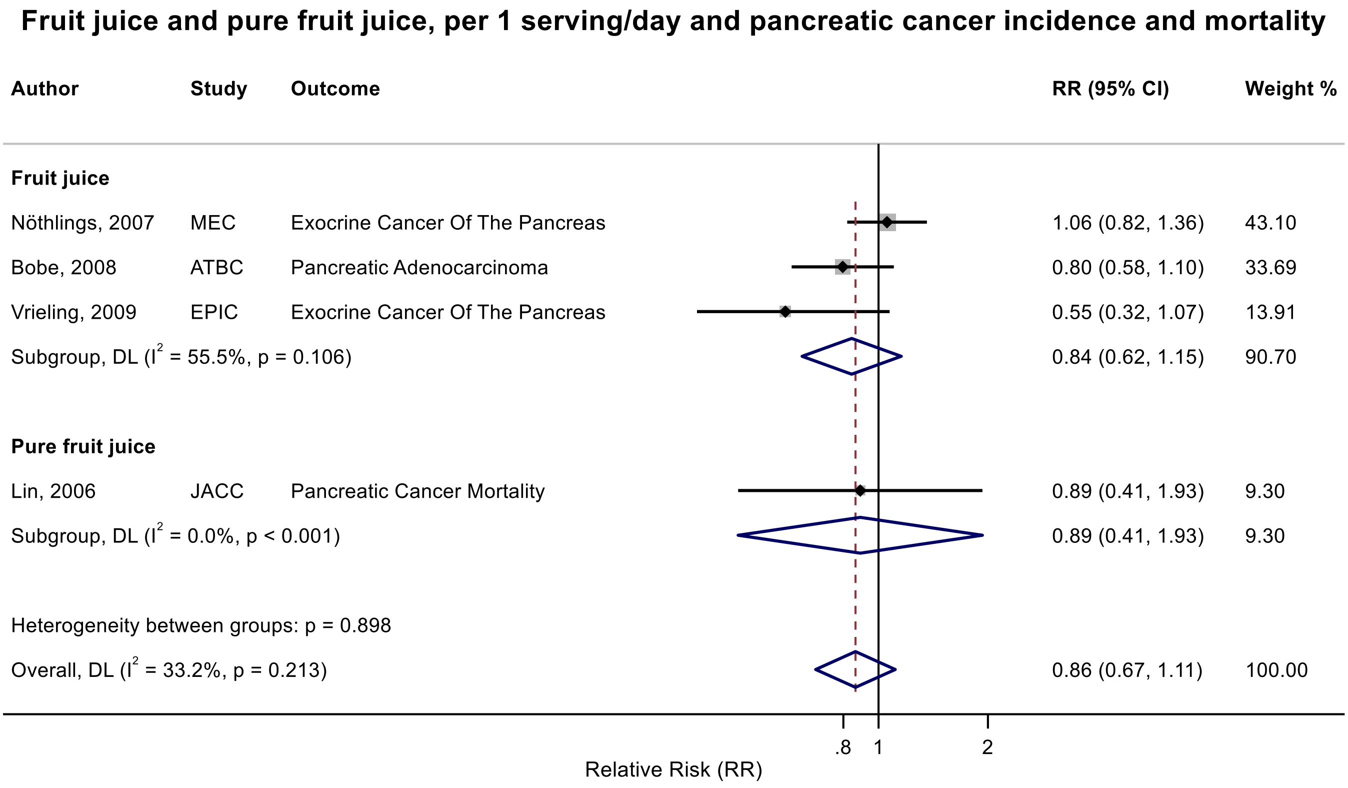

Figure legend: Forest plot shows the linear dose-response results (per 1 serving [177 mL] of juice/day) from the inverse variance DerSimonian-Laird random-effects model. The diamond represents the summary relative risk (RR) estimate, the diamond's width is the 95% confidence interval (CI). Each square and the horizontal line across the square represent the RR estimate of the individual study and its 95% CI.

Abbreviations: CI, confidence interval; RR, relative risk.

Study abbreviations: MEC, Multiethnic Cohort Study; ATBC, Alpha-Tocopherol, Beta-Carotene Cancer Prevention Study; EPIC, European Prospective Investigation into Cancer and nutrition; JACC, Japan Collaborative Cohort study;

Nöthlings, 2007[131], Bobe, 2008[41], Vrieling, 2009[167], Lin, 2006[103]

**Supplementary Figure 256. Linear dose-response meta-analysis of fruit juice and breast cancer incidence.**

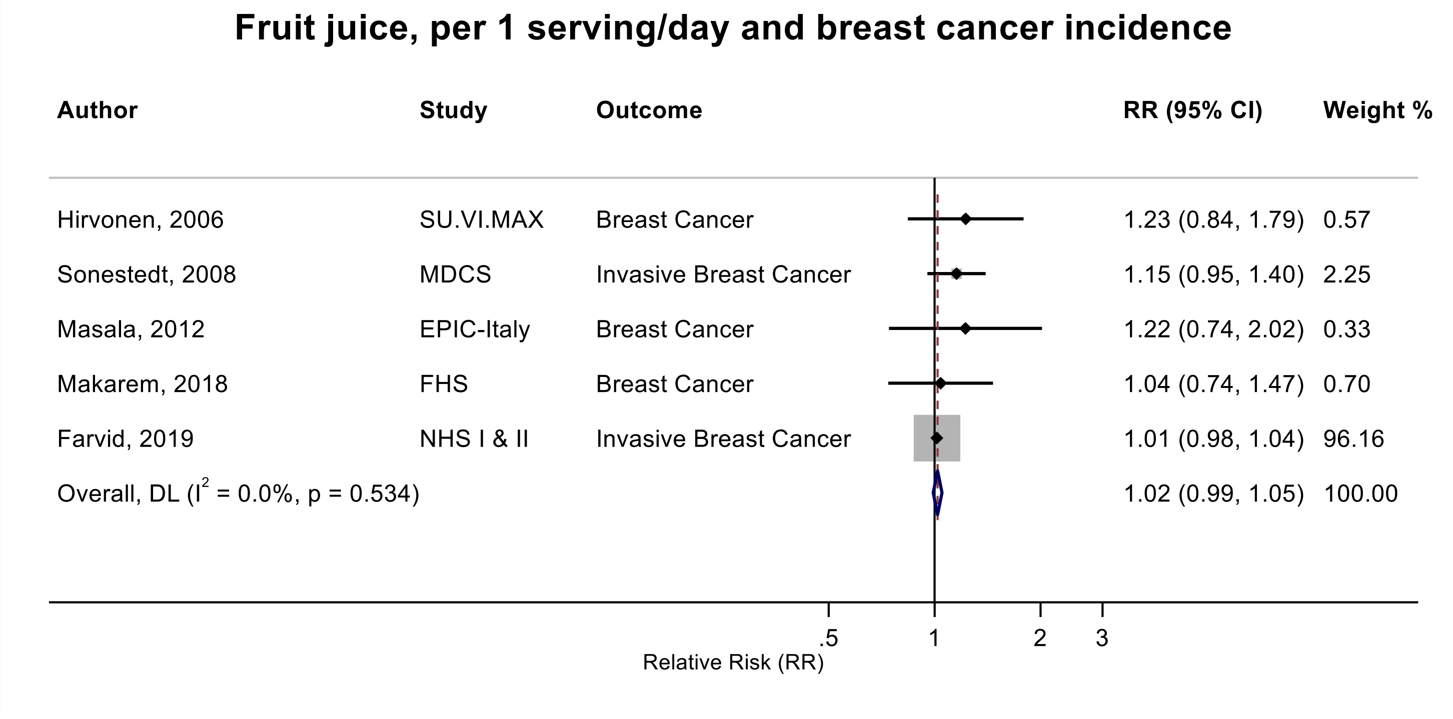

Figure legend: Forest plot shows the linear dose-response results (per 1 serving [177 mL] of juice/day) from the inverse variance DerSimonian-Laird random-effects model. The diamond represents the summary relative risk (RR) estimate, the diamond's width is the 95% confidence interval (CI). Each square and the horizontal line across the square represent the RR estimate of the individual study and its 95% CI.

Abbreviations: CI, confidence interval; RR, relative risk.

Study abbreviations: SU.VI.MAX, Supplémentation en Vitamines et Minéraux Antioxydants Study; MDCS, Malmö Diet and Cancer Study; EPIC-Italy, European Prospective Investigation into Cancer and nutrition Italy cohort; FHS, Framingham Heart Study; NHS I & II, Nurses' Health Study I & II

Hirvonen, 2006[69], Sonestedt, 2008[156], Masala, 2012[112], Makarem, 2018[108], Farvid, 2019[52]

**Supplementary Figure 257. Linear dose-response meta-analysis of fruit juice and breast cancer incidence by menopausal status.**

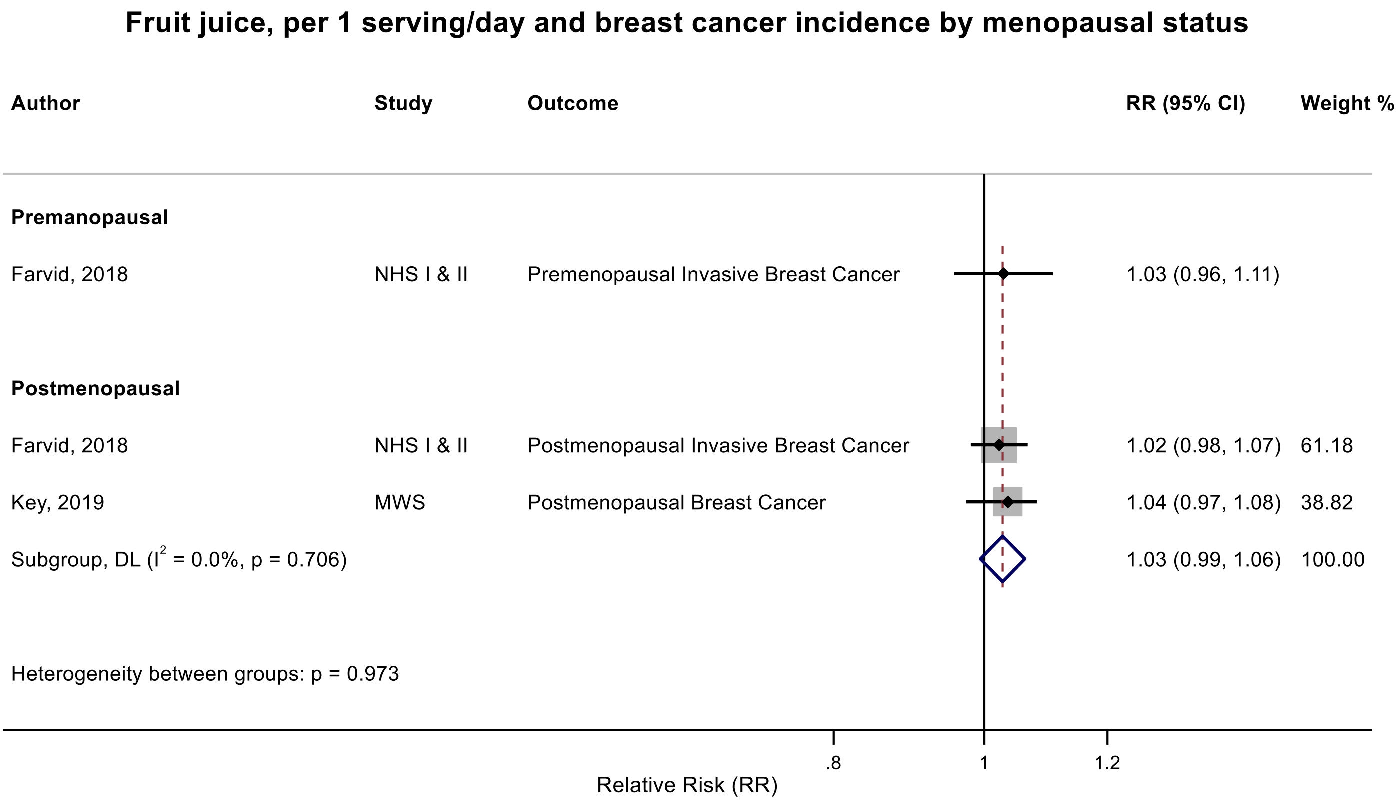

Figure legend: Forest plot shows the linear dose-response results (per 1 serving [177 mL] of juice/day) from the inverse variance DerSimonian-Laird random-effects model. The diamond represents the summary relative risk (RR) estimate, the diamond's width is the 95% confidence interval (CI). Each square and the horizontal line across the square represent the RR estimate of the individual study and its 95% CI.

Abbreviations: CI, confidence interval; RR, relative risk.

Study abbreviations: NHS I & II, Nurses' Health Study I & II; MWS, Million Women Study

Farvid, 2019[52], Key, 2019[83]

**Supplementary Figure 258. Linear dose-response meta-analysis of pure fruit juice and breast cancer incidence.**

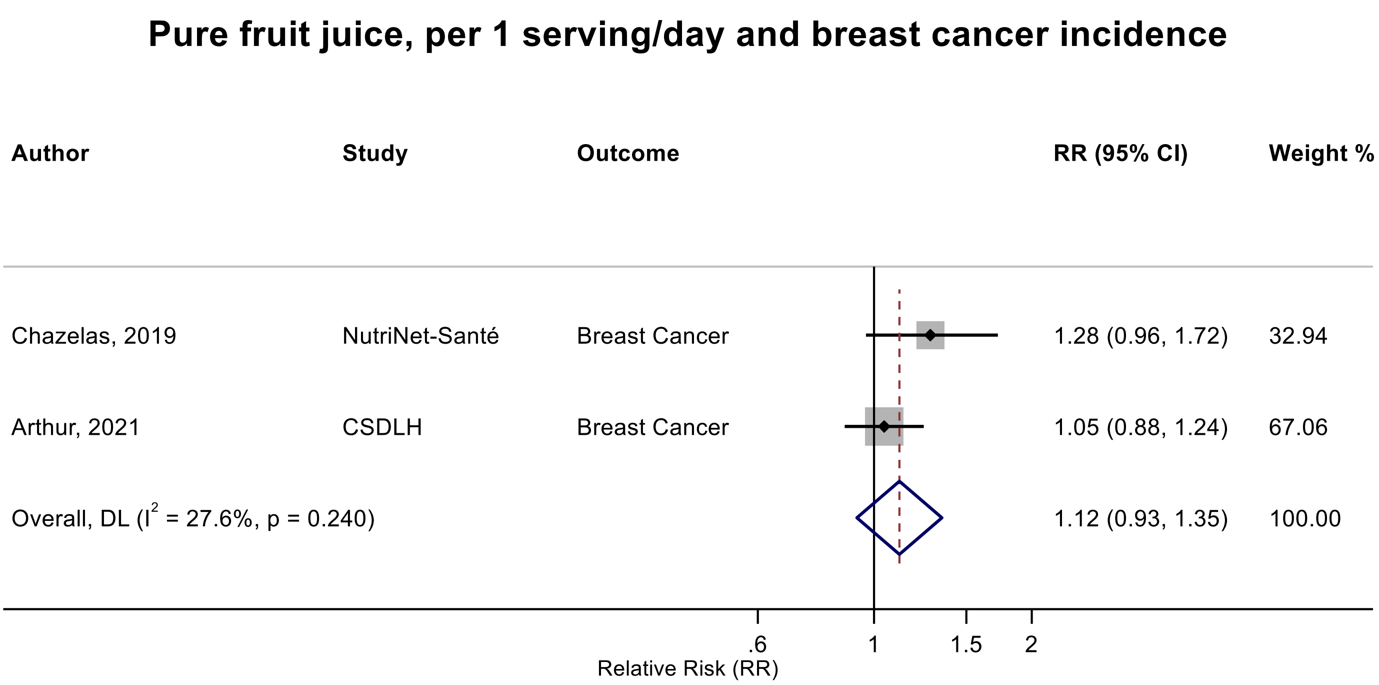

Figure legend: Forest plot shows the linear dose-response results (per 1 serving [177 mL] of juice/day) from the inverse variance DerSimonian-Laird random-effects model. The diamond represents the summary relative risk (RR) estimate, the diamond's width is the 95% confidence interval (CI). Each square and the horizontal line across the square represent the RR estimate of the individual study and its 95% CI.

Abbreviations: CI, confidence interval; RR, relative risk.

Study abbreviations: CSDLH, Canadian Study of Diet, Lifestyle and Health

Chazelas, 2019[44], Arthur, 2021[38]

**Supplementary Figure 259. Linear dose-response meta-analysis of pure fruit juice and breast cancer incidence by menopausal status.**

##### Pure fruit juice, per 1 serving/day and breast cancer incidence by menopausal status

| Author | Study | Outcome | RR (95% CI) | Weight % |
|---|---|---|---|---|
| **Postmenopausal Breast Cancer** | | | | |
| Chazelas, 2019 | NutriNet-Santé | Postmenopausal Breast Cancer | 1.07 (0.72, 1.59) | 22.53 |
| Arthur, 2021 | CSDLH | Postmenopausal Breast Cancer | 1.06 (0.86, 1.32) | 77.47 |
| Subgroup, DL ($I^2$ = 0.0%, p = 0.975) | | | 1.07 (0.88, 1.29) | 100.00 |
| **Premenopausal Breast Cancer** | | | | |
| Chazelas, 2019 | NutriNet-Santé | Premenopausal Breast Cancer | 1.18 (0.77, 1.84) | 19.44 |
| Arthur, 2021 | CSDLH | Premenopausal Breast Cancer | 1.00 (0.80, 1.24) | 80.56 |
| Subgroup, DL ($I^2$ = 0.0%, p = 0.492) | | | 1.03 (0.85, 1.25) | 100.00 |
| Heterogeneity between groups: p = 0.812 | | | | |

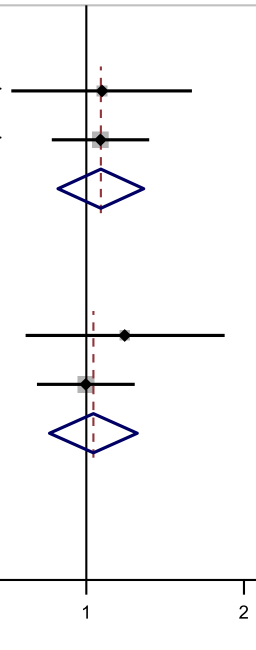

Figure legend: Forest plot shows the linear dose-response results (per 1 serving [177 mL] of juice/day) from the inverse variance DerSimonian-Laird random-effects model. The diamond represents the summary relative risk (RR) estimate, the diamond's width is the 95% confidence interval (CI). Each square and the horizontal line across the square represent the RR estimate of the individual study and its 95% CI.

Abbreviations: CI, confidence interval; RR, relative risk.

Study abbreviations: CSDLH, Canadian Study of Diet, Lifestyle and Health

Chazelas, 2019[44], Arthur, 2021[38]

**Supplementary Figure 260. Linear dose-response meta-analysis of fruit juice and pure fruit juice combined and breast cancer incidence.**

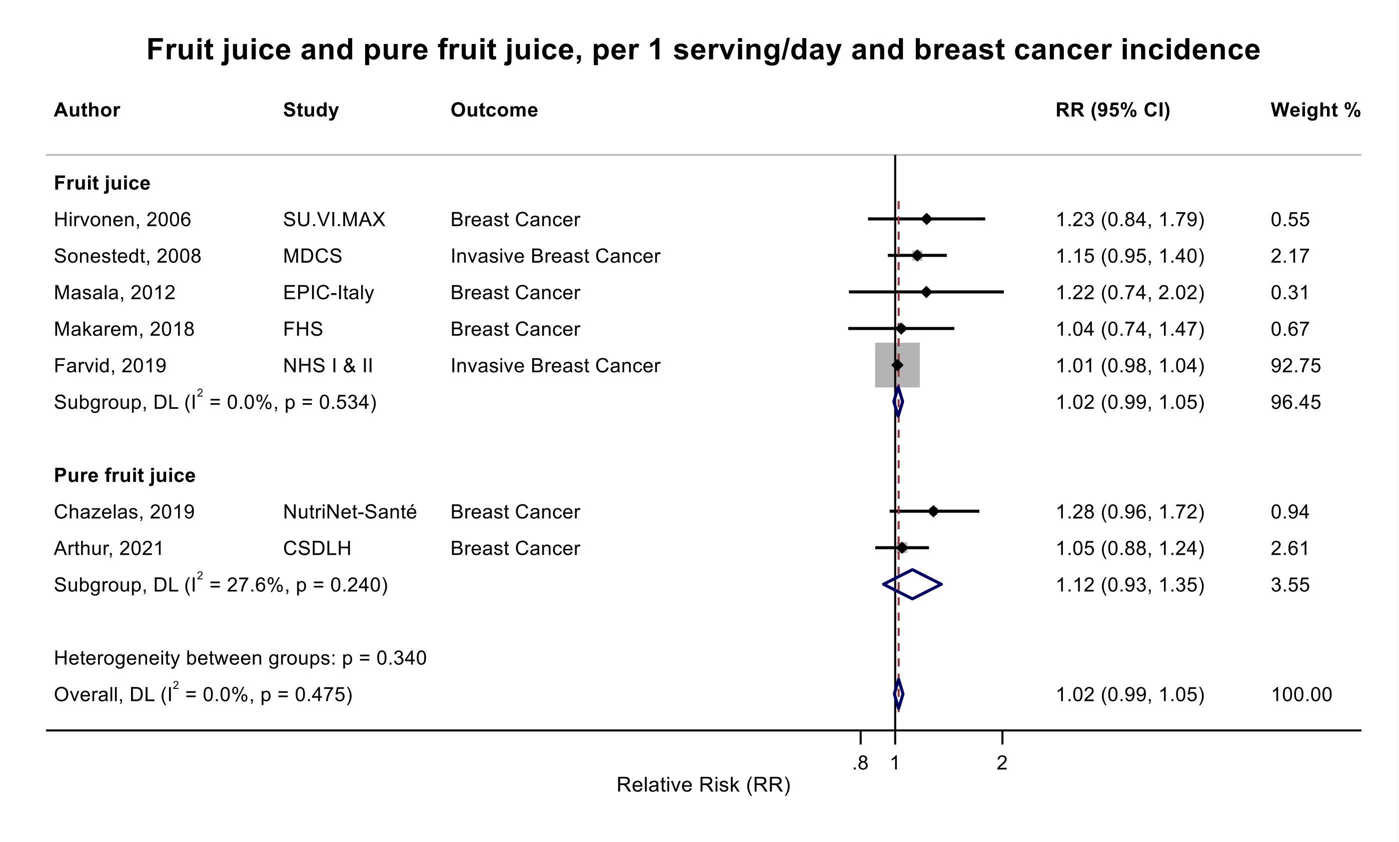

Figure legend: Forest plot shows the linear dose-response results (per 1 serving [177 mL] of juice/day) from the inverse variance DerSimonian-Laird random-effects model. The diamond represents the summary relative risk (RR) estimate, the diamond's width is the 95% confidence interval (CI). Each square and the horizontal line across the square represent the RR estimate of the individual study and its 95% CI.

Abbreviations: CI, confidence interval; RR, relative risk.

Study abbreviations: SU.VI.MAX, Supplémentation en Vitamines et Minéraux Antioxydants Study; MDCS, Malmö Diet and Cancer Study; EPIC-Italy, European Prospective Investigation into Cancer and nutrition Italy cohort; FHS, Framingham Heart Study; NHS I & II, Nurses' Health Study I & II; CSDLH, Canadian Study of Diet, Lifestyle and Health

Hirvonen, 2006[69], Sonestedt, 2008[156], Masala, 2012[112], Makarem, 2018[108], Farvid, 2019[52], Chazelas, 2019[44], Arthur, 2021[38]

**Supplementary Figure 261. Linear dose-response meta-analysis of fruit juice and pure fruit juice combined and premenopausal breast cancer incidence.**

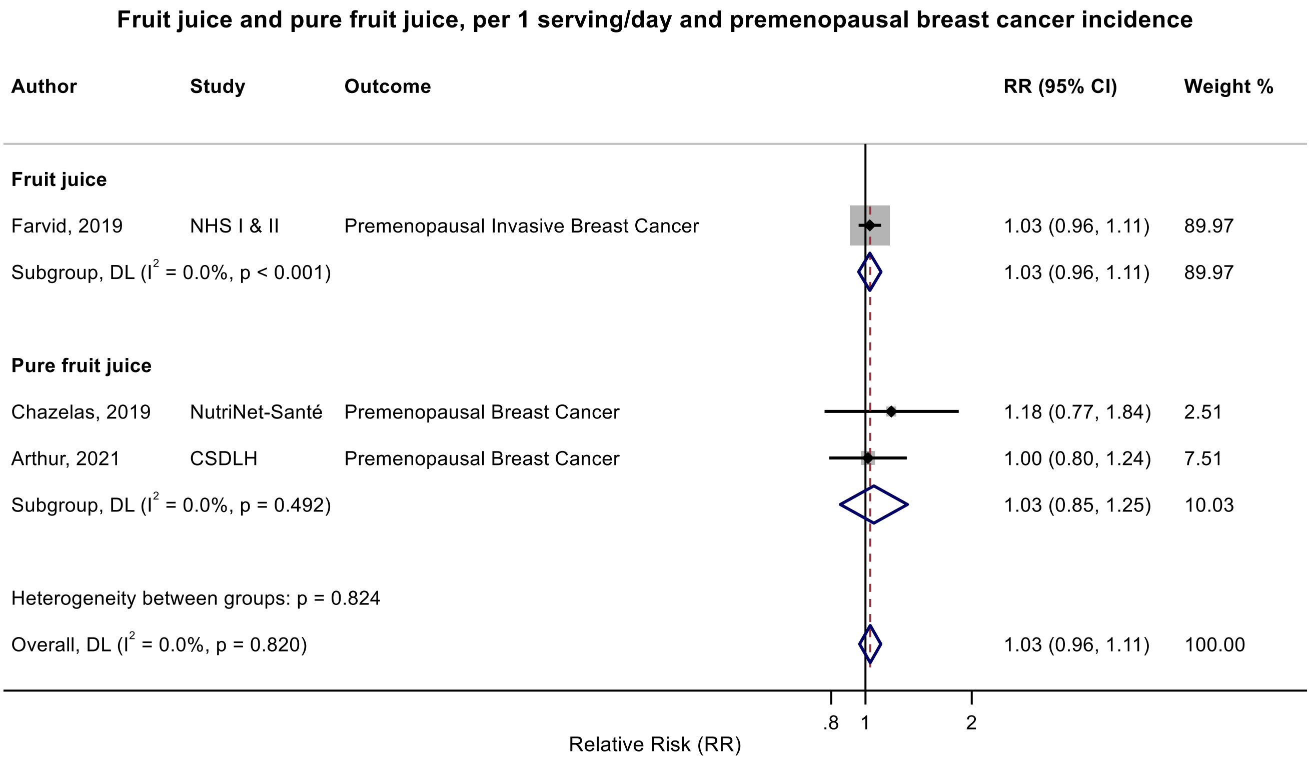

Figure legend: Forest plot shows the linear dose-response results (per 1 serving [177 mL] of juice/day) from the inverse variance DerSimonian-Laird random-effects model. The diamond represents the summary relative risk (RR) estimate, the diamond's width is the 95% confidence interval (CI). Each square and the horizontal line across the square represent the RR estimate of the individual study and its 95% CI.

Abbreviations: CI, confidence interval; RR, relative risk.

Study abbreviations: NHS I & II, Nurses' Health Study I & II; CSDLH, Canadian Study of Diet, Lifestyle and Health

Farvid, 2019[52], Chazelas, 2019[44], Arthur, 2021[38]

**Supplementary Figure 262. Linear dose-response meta-analysis of fruit juice and pure fruit juice combined and postmenopausal breast cancer incidence.**

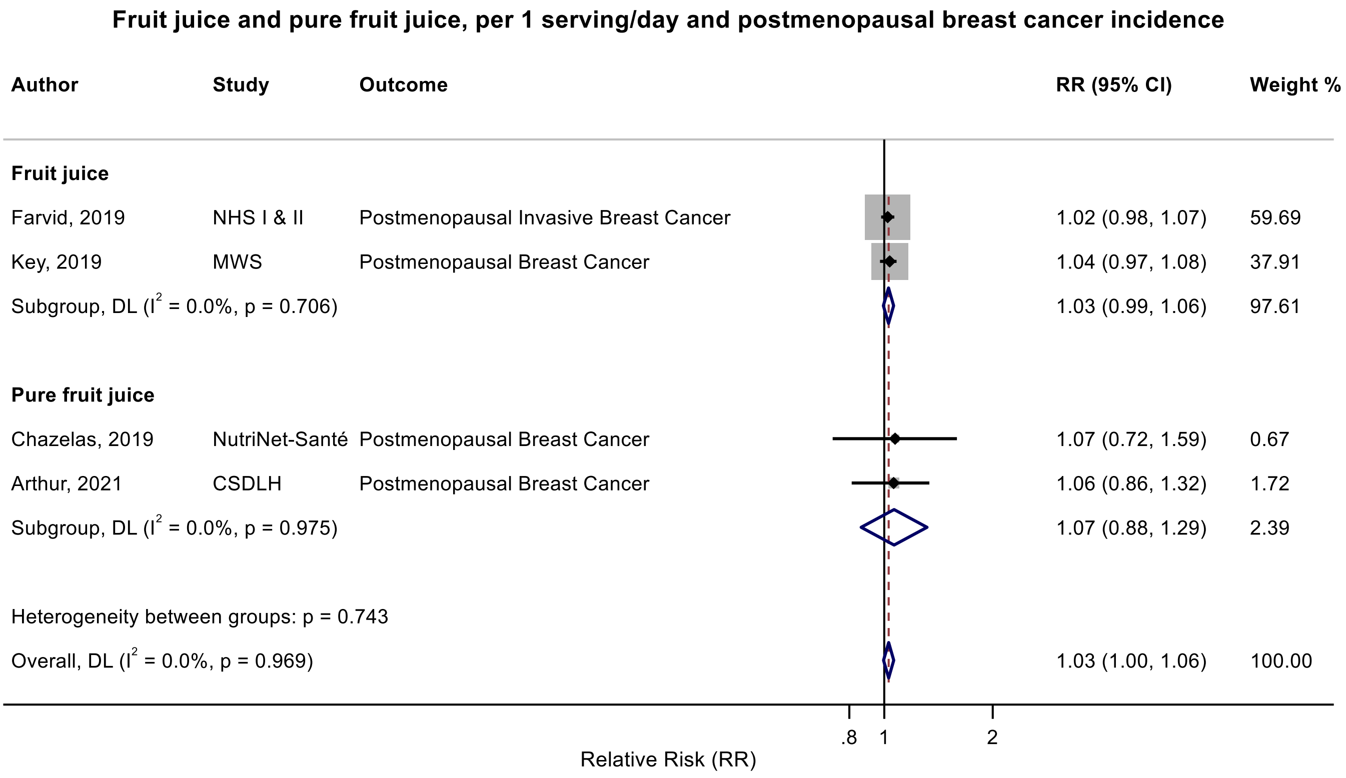

Figure legend: Forest plot shows the linear dose-response results (per 1 serving [177 mL] of juice/day) from the inverse variance DerSimonian-Laird random-effects model. The diamond represents the summary relative risk (RR) estimate, the diamond's width is the 95% confidence interval (CI). Each square and the horizontal line across the square represent the RR estimate of the individual study and its 95% CI.

Abbreviations: CI, confidence interval; RR, relative risk.

Study abbreviations: NHS I & II, Nurses' Health Study I & II; MWS, Million Women Study; CSDLH, Canadian Study of Diet, Lifestyle and Health

Farvid, 2019[52], Key 2019[83], Chazelas, 2019[44], Arthur, 2021[38]

Note: The summary RR and 95%CI with 3 decimal point are 1.028 (0.995-1.062), p=0.096.

**Supplementary Figure 263. Linear dose-response meta-analysis of fruit juice and pure fruit juice combined and ovarian cancer incidence.**

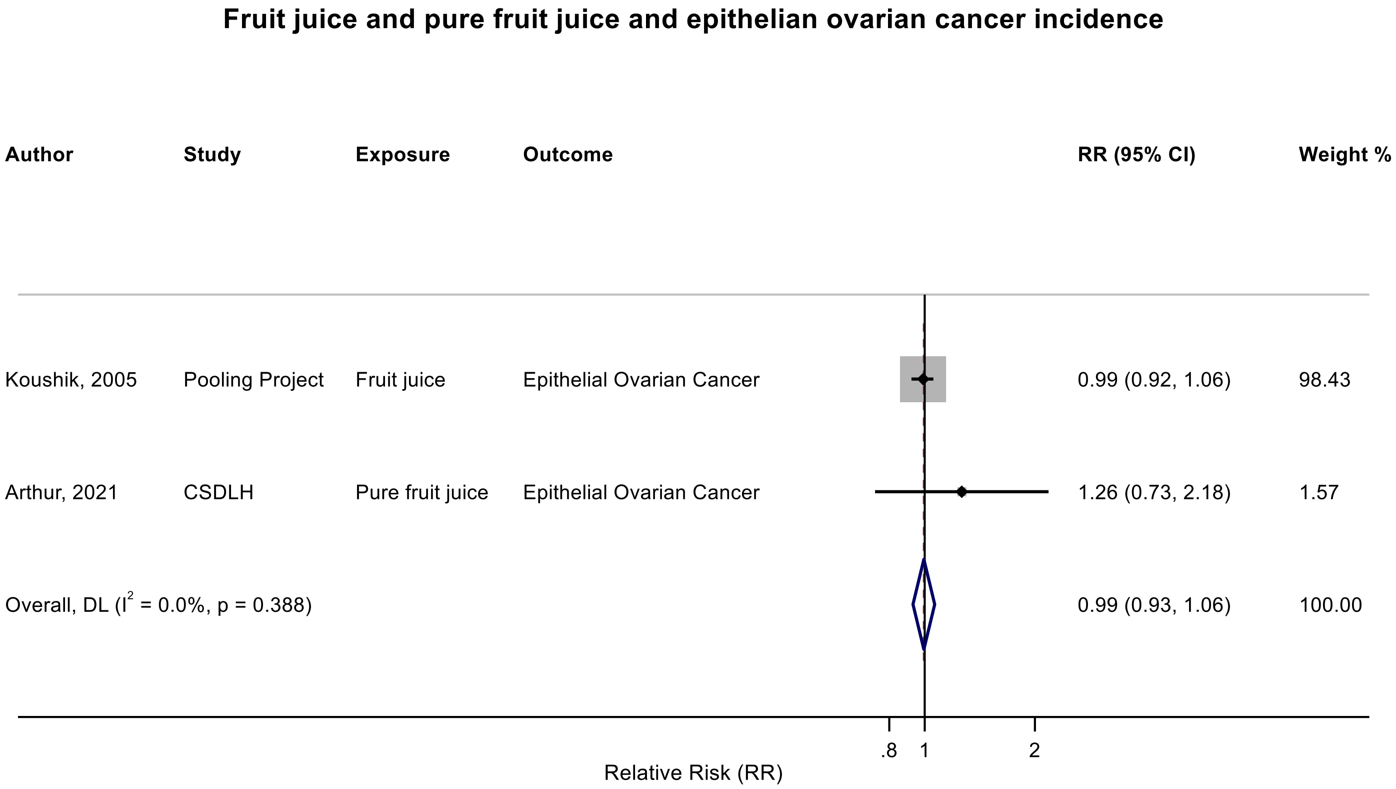

Figure legend: Forest plot shows the linear dose-response results (per 1 serving [177 mL] of juice/day) from the inverse variance DerSimonian-Laird random-effects model. The diamond represents the summary relative risk (RR) estimate, the diamond's width is the 95% confidence interval (CI). Each square and the horizontal line across the square represent the RR estimate of the individual study and its 95% CI.

Abbreviations: CI, confidence interval; RR, relative risk.

Study abbreviations: Pooling Project, pooled analysis of cohort studies; CSDLH, Canadian Study of Diet, Lifestyle and Health

Koushik, 2005 [91], Arthur, 2021[38]

**Supplementary Figure 264. Linear dose-response meta-analysis of fruit juice and pure fruit juice combined and endometrial cancer incidence.**

**Fruit juice and pure fruit juice, per 1 serving/day and endometrial cancer incidence**

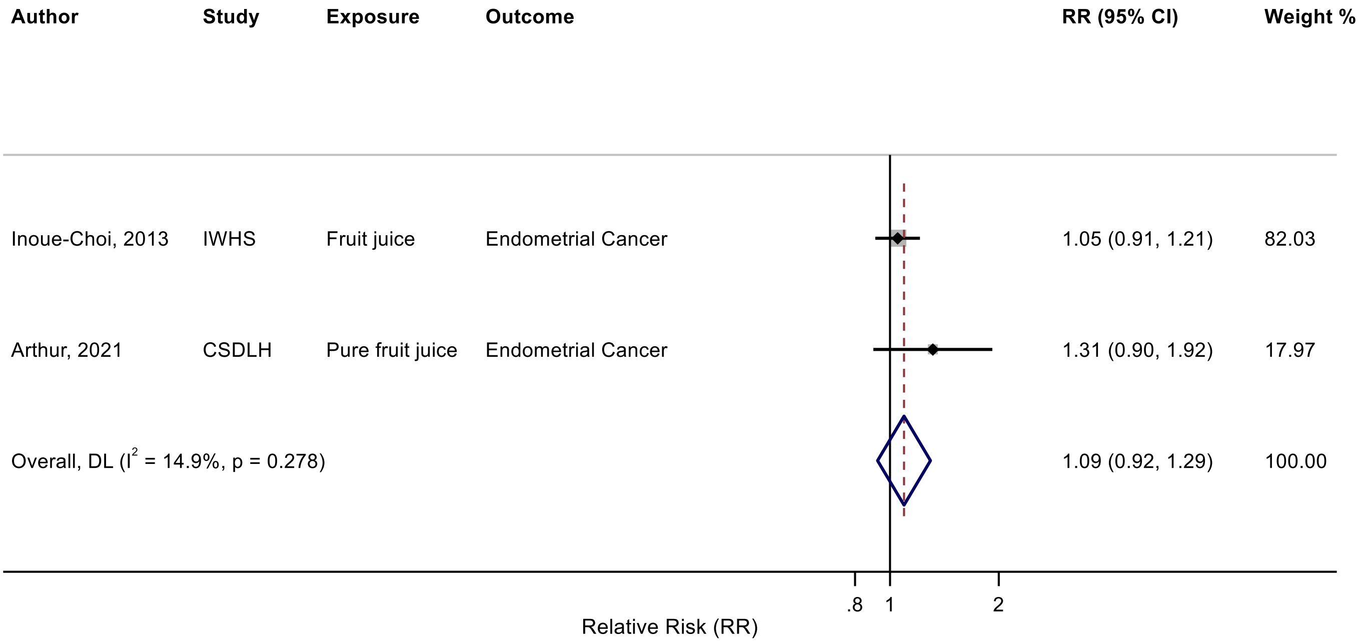

Figure legend: Forest plot shows the linear dose-response results (per 1 serving [177 mL] of juice/day) from the inverse variance DerSimonian-Laird random-effects model. The diamond represents the summary relative risk (RR) estimate, the diamond's width is the 95% confidence interval (CI). Each square and the horizontal line across the square represent the RR estimate of the individual study and its 95% CI.

Abbreviations: CI, confidence interval; RR, relative risk.

Study abbreviations: IWHS, Iowa Women's Health study; CSDLH, Canadian Study of Diet, Lifestyle and Health

Inoue-Choi, 2013[74], Arthur, 2021[38]

**Supplementary Figure 265. Linear dose-response meta-analysis of fruit juice and kidney cancer incidence.**

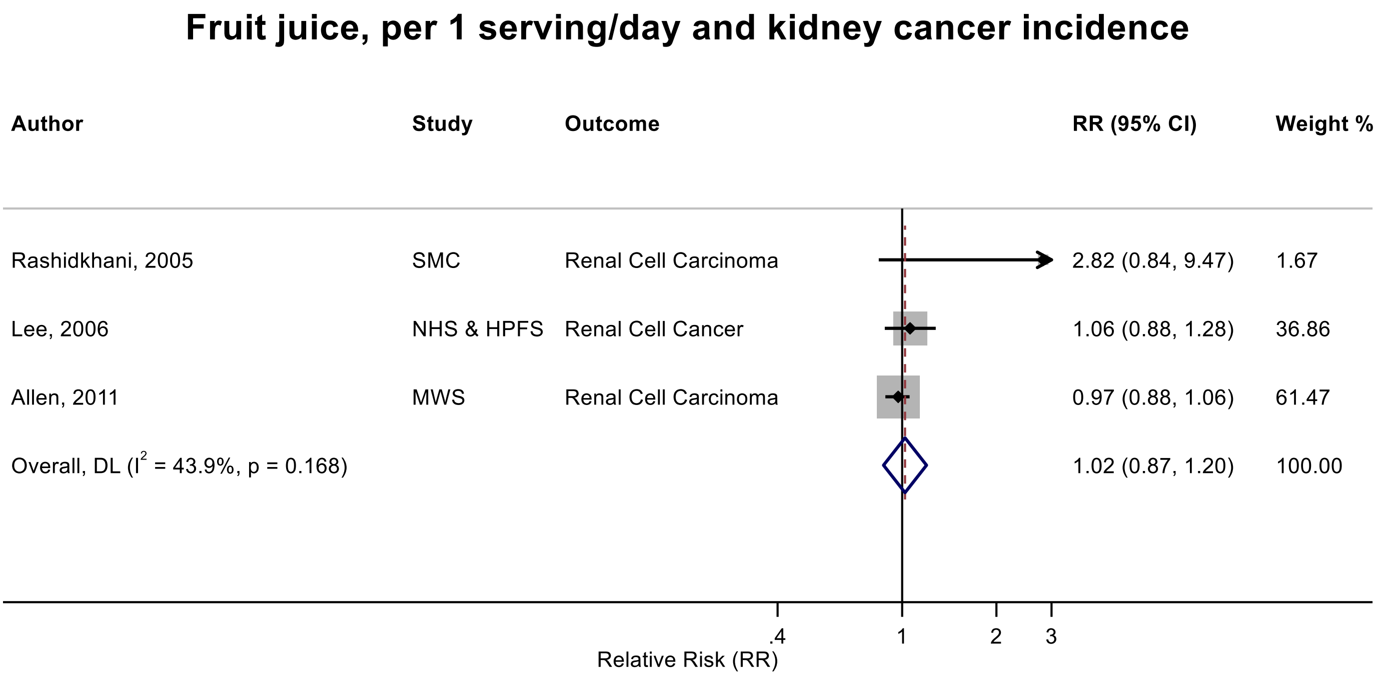

Figure legend: Forest plot shows the linear dose-response results (per 1 serving [177 mL] of juice/day) from the inverse variance DerSimonian-Laird random-effects model. The diamond represents the summary relative risk (RR) estimate, the diamond's width is the 95% confidence interval (CI). Each square and the horizontal line across the square represent the RR estimate of the individual study and its 95% CI.

Abbreviations: CI, confidence interval; RR, relative risk.

Study abbreviations: SMC, Swedish Mammography Cohort; NHS & HPFS, Nurses' Health Study & Health Professionals Follow-up Study; MWS, Million Women Study

Rashidkhani, 2005[142], Lee, 2006[98], Allen, 2011[36]

**Supplementary Figure 266. Linear dose-response meta-analysis of fruit juice and glioma incidence.**

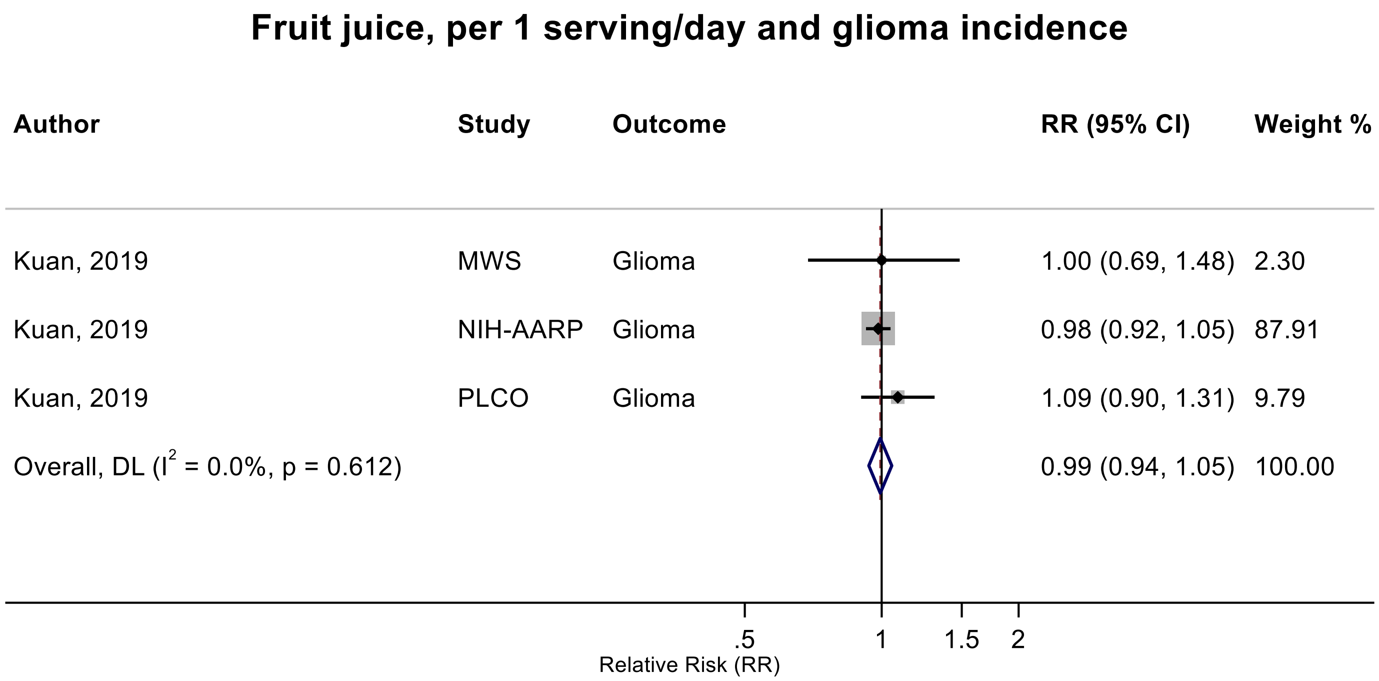

Figure legend: Forest plot shows the linear dose-response results (per 1 serving [177 mL] of juice/day) from the inverse variance DerSimonian-Laird random-effects model. The diamond represents the summary relative risk (RR) estimate, the diamond's width is the 95% confidence interval (CI). Each square and the horizontal line across the square represent the RR estimate of the individual study and its 95% CI.

Abbreviations: CI, confidence interval; RR, relative risk.

Study abbreviations: MWS, Million Women Study; NIH-AARP, National Institute of Health + American Association of Retired Persons Diet and Health Study; PLCO, Prostate, Lung, Colorectal and Ovarian Cancer Screening trial

Kuan, 2019[92]

**Supplementary Figure 267. Linear dose-response meta-analysis of citrus juice and melanoma incidence.**

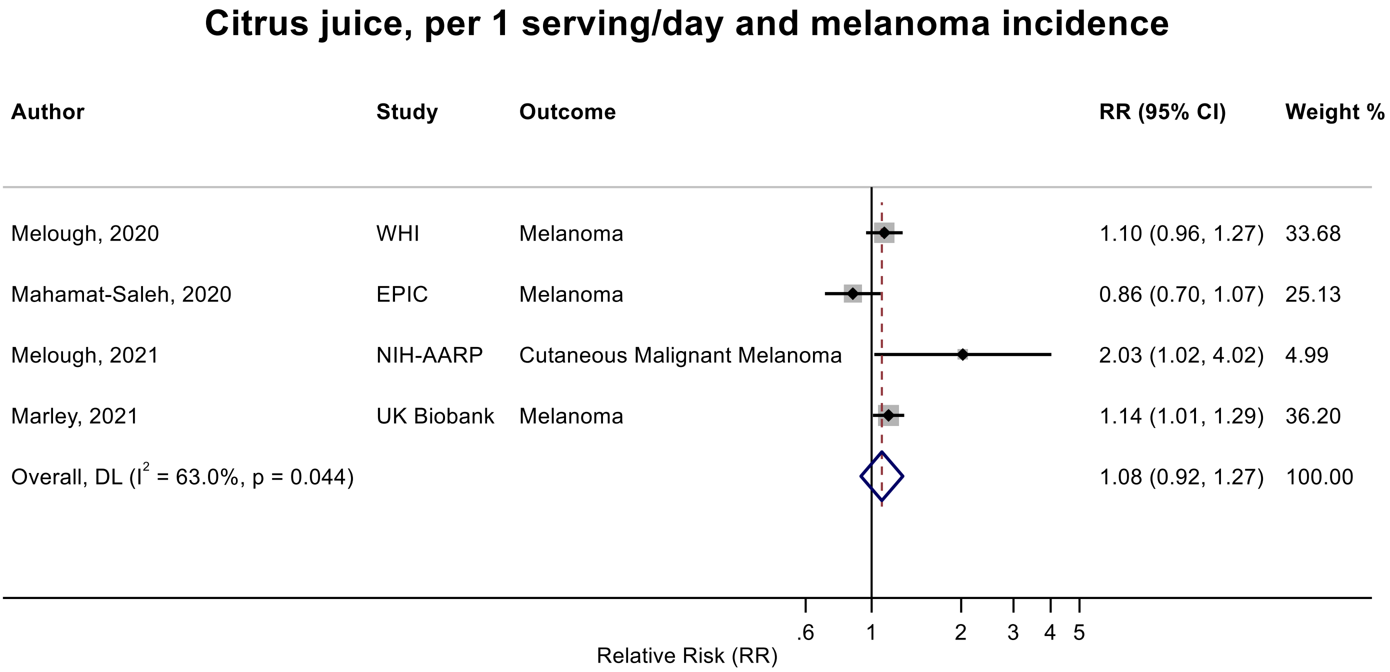

Figure legend: Forest plot shows the linear dose-response results (per 1 serving [177 mL] of juice/day) from the inverse variance DerSimonian-Laird random-effects model. The diamond represents the summary relative risk (RR) estimate, the diamond's width is the 95% confidence interval (CI). Each square and the horizontal line across the square represent the RR estimate of the individual study and its 95% CI.

Abbreviations: CI, confidence interval; RR, relative risk.

Study abbreviations: WHI, Women's Health Initiative Study; EPIC, European Prospective Investigation into Cancer and nutrition; NIH-AARP, National Institute of Health + American Association of Retired Persons Diet and Health Study; UK Biobank, United Kingdom Biobank Study

Melough, 2020[116], Mahamat-Saleh, 2020[107], Melough, 2021[115], Marley, 2021[110]

**Supplementary Figure 268. Linear dose-response meta-analysis of citrus juice and non-melanoma skin cancer incidence.**

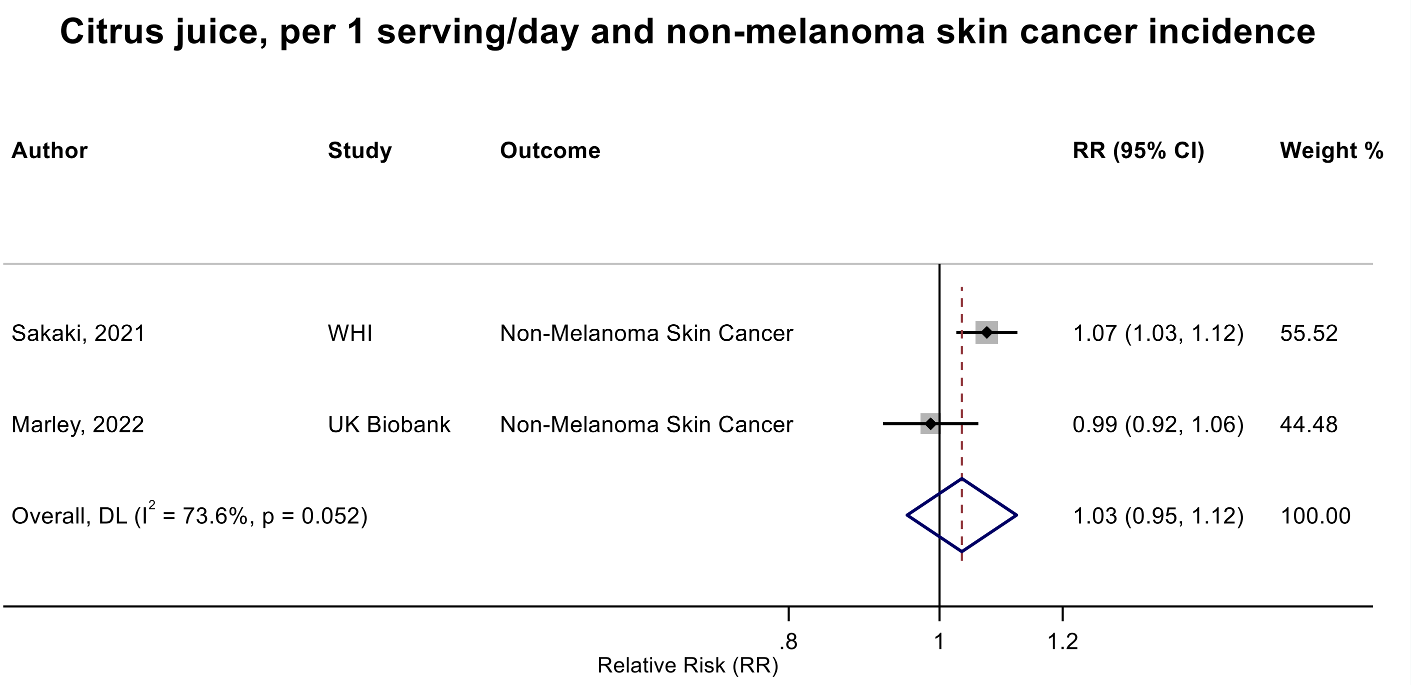

Figure legend: Forest plot shows the linear dose-response results (per 1 serving [177 mL] of juice/day) from the inverse variance DerSimonian-Laird random-effects model. The diamond represents the summary relative risk (RR) estimate, the diamond's width is the 95% confidence interval (CI). Each square and the horizontal line across the square represent the RR estimate of the individual study and its 95% CI.

Abbreviations: CI, confidence interval; RR, relative risk.

Study abbreviations: WHI, Women's Health Initiative Study; UK Biobank, United Kingdom Biobank Study

Sakaki, 2021[148], Marley, 2022[111]

**Supplementary Figure 269. Linear dose-response meta-analysis of orange juices and melanoma incidence.**

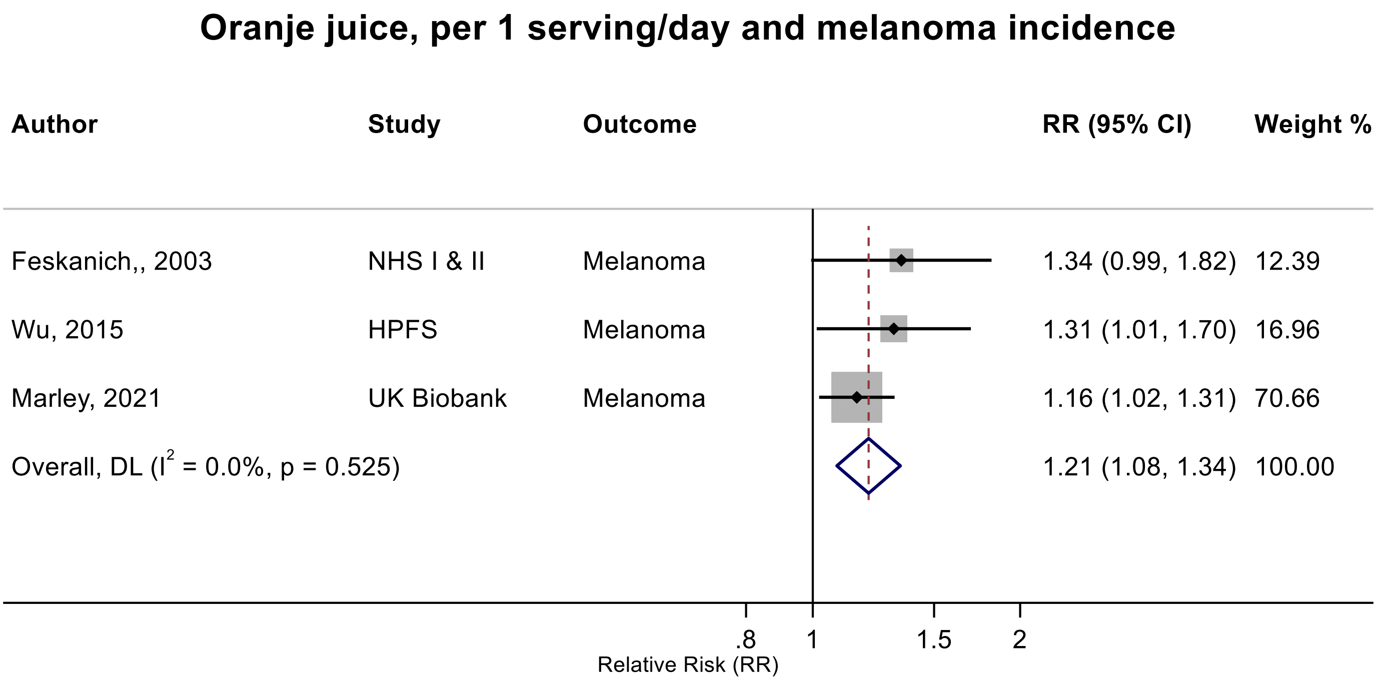

Figure legend: Forest plot shows the linear dose-response results (per 1 serving [177 mL] of juice/day) from the inverse variance DerSimonian-Laird random-effects model. The diamond represents the summary relative risk (RR) estimate, the diamond's width is the 95% confidence interval (CI). Each square and the horizontal line across the square represent the RR estimate of the individual study and its 95% CI.

Abbreviations: CI, confidence interval; RR, relative risk.

Study abbreviations: NHS I & II, Nurses' Health Study I & II; HPFS, Health Professionals Follow-up Study; UK Biobank, United Kingdom Biobank Study

Note: All studies performed repeated measurements.

Feskanich, 2003[54], Wu, 2015[171], Marley, 2021[110]

**Supplementary Figure 270. Linear dose-response meta-analysis of orange juices and basal cell carcinoma incidence.**

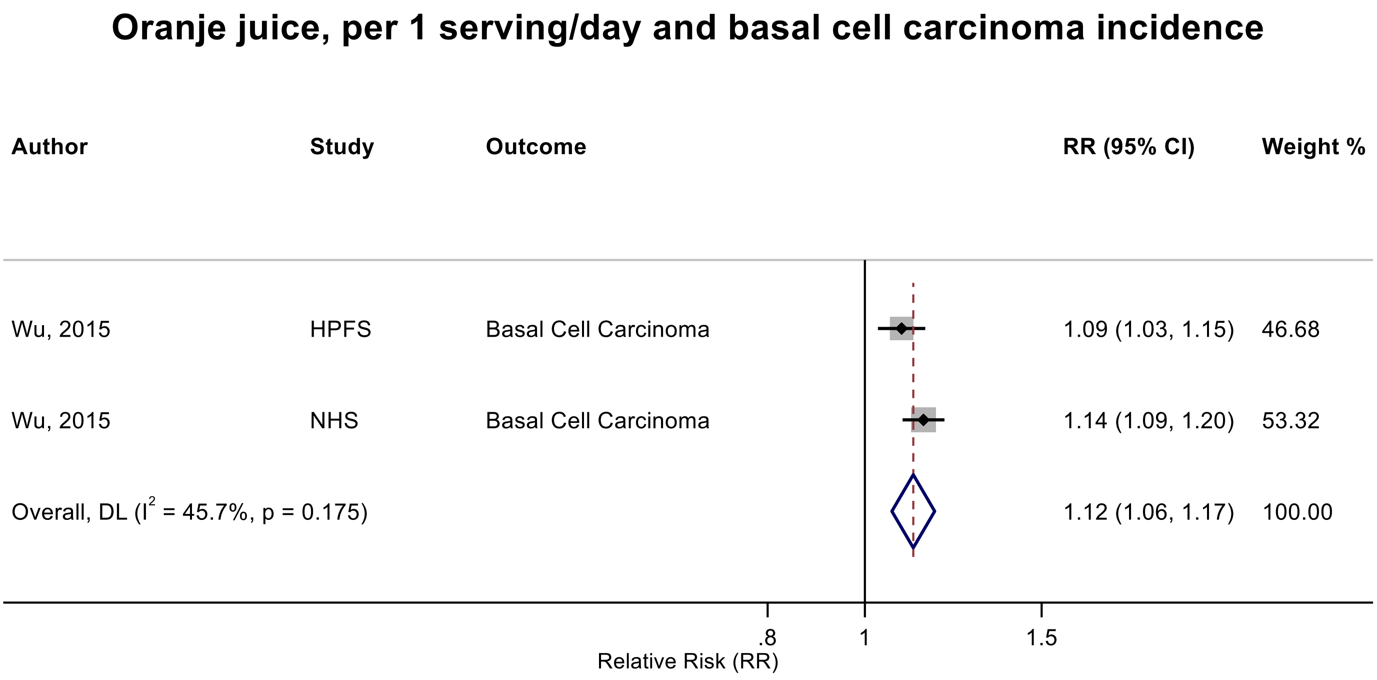

Figure legend: Forest plot shows the linear dose-response results (per 1 serving [177 mL] of juice/day) from the inverse variance DerSimonian-Laird random-effects model. The diamond represents the summary relative risk (RR) estimate, the diamond's width is the 95% confidence interval (CI). Each square and the horizontal line across the square represent the RR estimate of the individual study and its 95% CI.

Abbreviations: CI, confidence interval; RR, relative risk.

Study abbreviations: HPFS, Health Professionals Follow-up Study; NHS, Nurses' Health Study

Note: Both studies performed repeated measurements.

Wu, 2015[171]

**Supplementary Figure 271. Linear dose-response meta-analysis of orange juices and incidence of squamous cell carcinoma of the skin.**

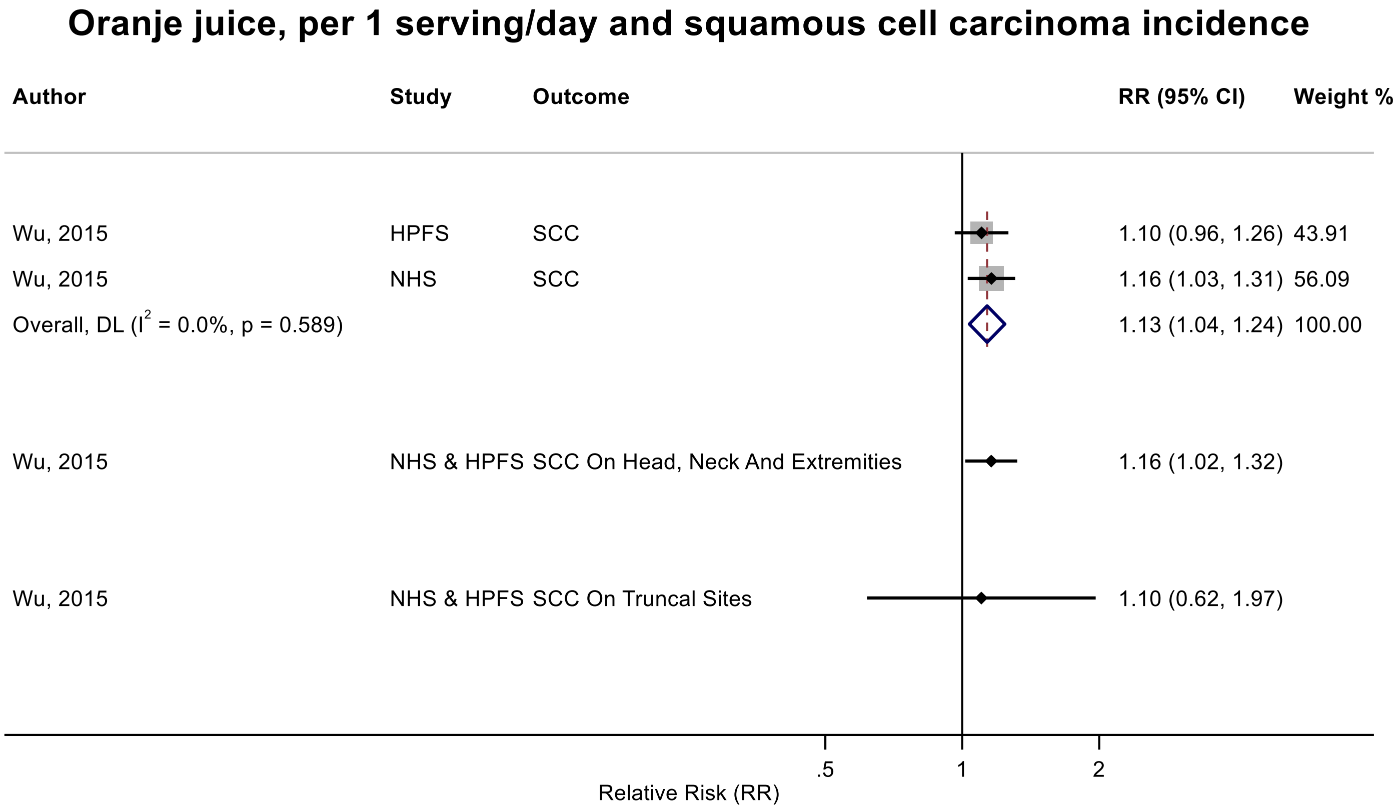

Figure legend: Forest plot shows the linear dose-response results (per 1 serving [177 mL] of juice/day) from the inverse variance DerSimonian-Laird random-effects model. The diamond represents the summary relative risk (RR) estimate, the diamond's width is the 95% confidence interval (CI). Each square and the horizontal line across the square represent the RR estimate of the individual study and its 95% CI.

Abbreviations: CI, confidence interval; RR, relative risk, SCC, squamous cell carcinoma.

Study abbreviations: HPFS, Health Professionals Follow-up Study; NHS, Nurses' Health Study; NHS & HPFS, Nurses' Health Study & Health Professionals Follow-up Study

Note: Both studies performed repeated measurements.

Wu, 2015[170]

**Supplementary Figure 272. Linear dose-response meta-analysis of citrus juice and orange juices combined and incidence of melanoma.**

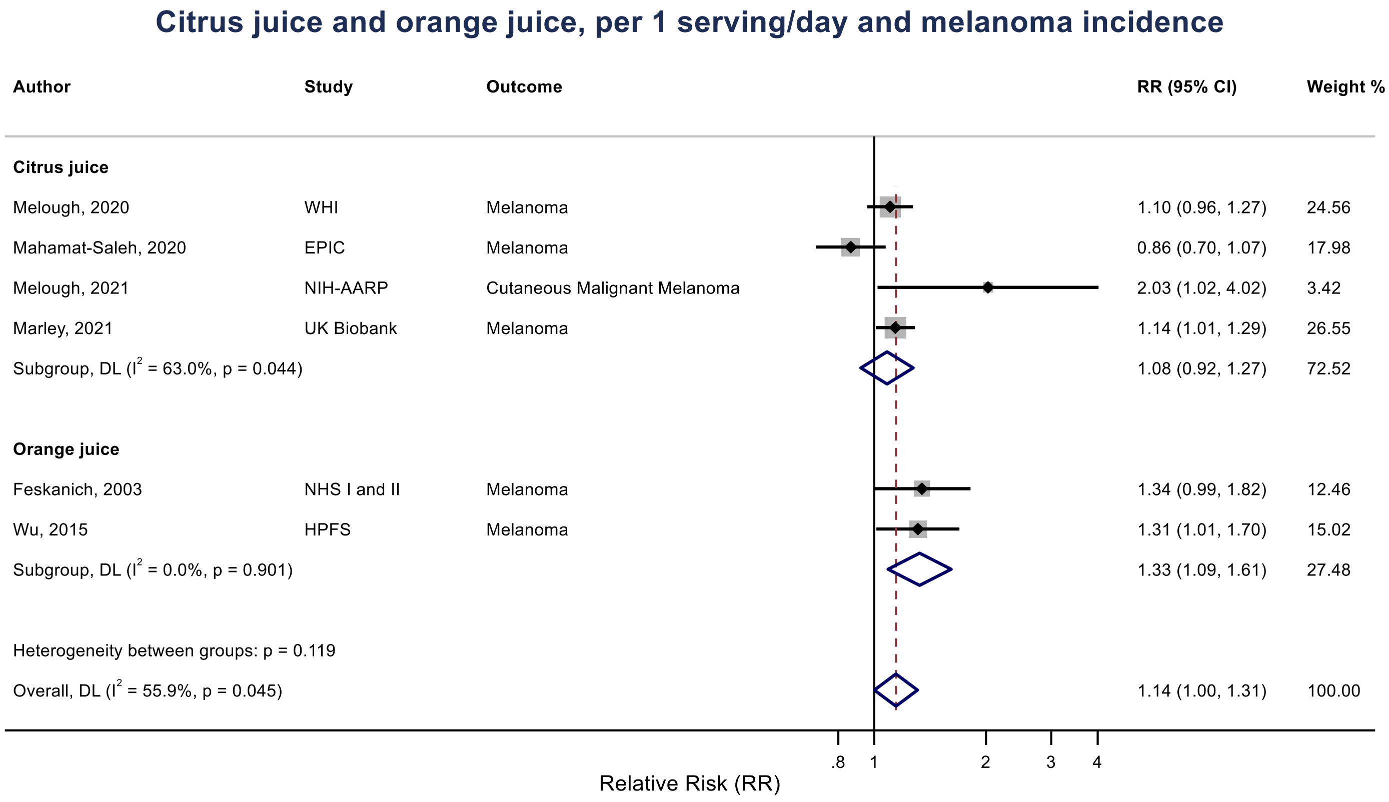

Figure legend: Forest plot shows the linear dose-response results (per 1 serving [177 mL] of juice/day) from the inverse variance DerSimonian-Laird random-effects model. The diamond represents the summary relative risk (RR) estimate, the diamond's width is the 95% confidence interval (CI). Each square and the horizontal line across the square represent the RR estimate of the individual study and its 95% CI.

Abbreviations: CI, confidence interval; RR, relative risk.

Study abbreviations: WHI, Women's Health Initiative Study; EPIC, European Prospective Investigation into Cancer and Nutrition; NIH-AARP, National Institute of Health + American Association of Retired Persons Diet and Health Study, UK Biobank, United Kingdom Biobank Study, NHS I & II, Nurses' Health Study I & II; HPFS, Health Professionals Follow-up Study

Melough, 2020[116], Mahamat-Saleh, 2020[107], Melough, 2021[115], Marley, 2021[110], Feskanich, 2003[54], Wu, 2015[171]

**Supplementary Figure 273. Linear dose-response meta-analysis of citrus juice and orange juices combined and incidence of basal cell carcinoma of the skin.**

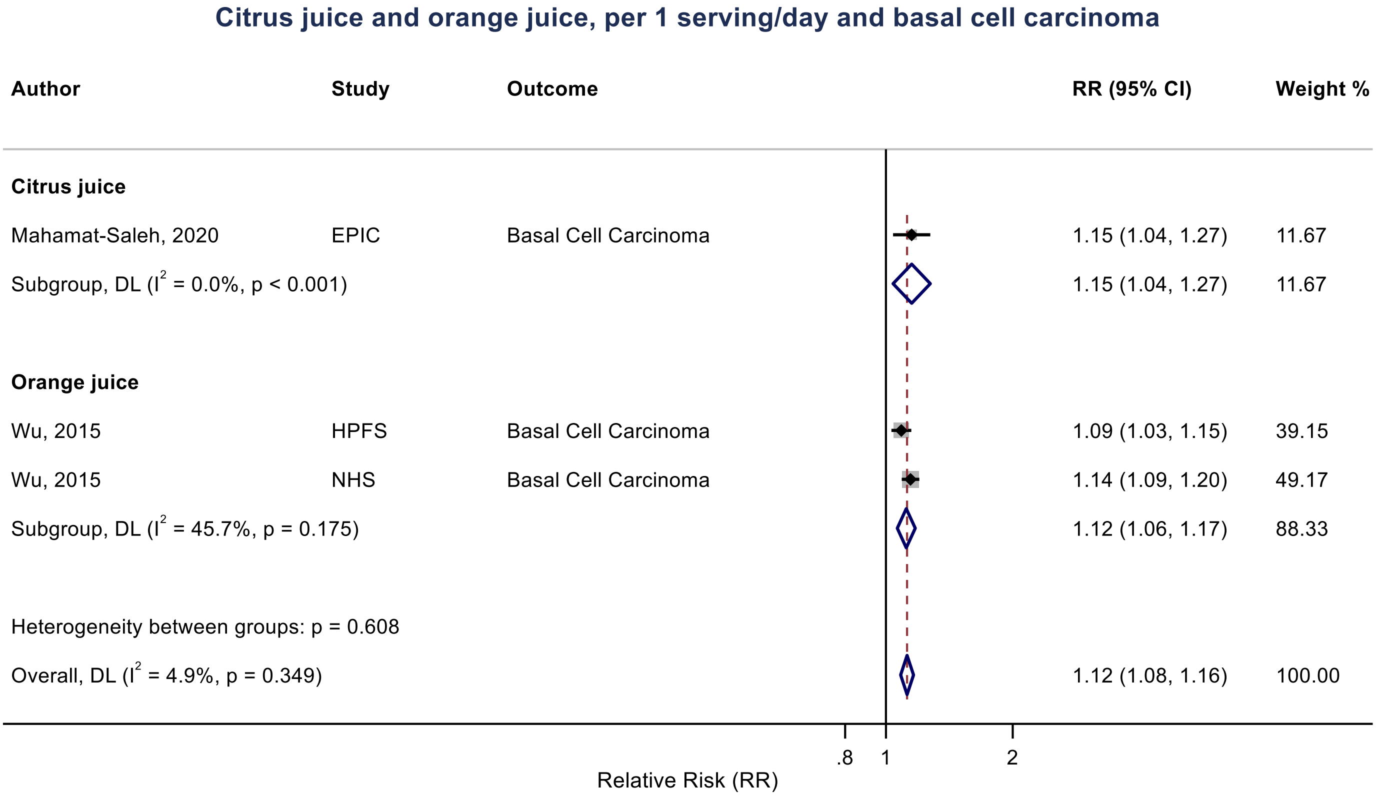

Figure legend: Forest plot shows the linear dose-response results (per 1 serving [177 mL] of juice/day) from the inverse variance DerSimonian-Laird random-effects model. The diamond represents the summary relative risk (RR) estimate, the diamond's width is the 95% confidence interval (CI). Each square and the horizontal line across the square represent the RR estimate of the individual study and its 95% CI.

Abbreviations: CI, confidence interval; RR, relative risk.

Study abbreviations: HPFS, Health Professionals Follow-up Study; NHS, Nurses' Health Study, EPIC, European prospective Investigation into Cancer and Nutrition.

Wu, 2015[171], Mahmat-Saleh, 2020[107]

**Supplementary Figure 274. Linear dose-response meta-analysis of citrus juice and orange juices combined and incidence of squamous cell carcinoma of the skin.**

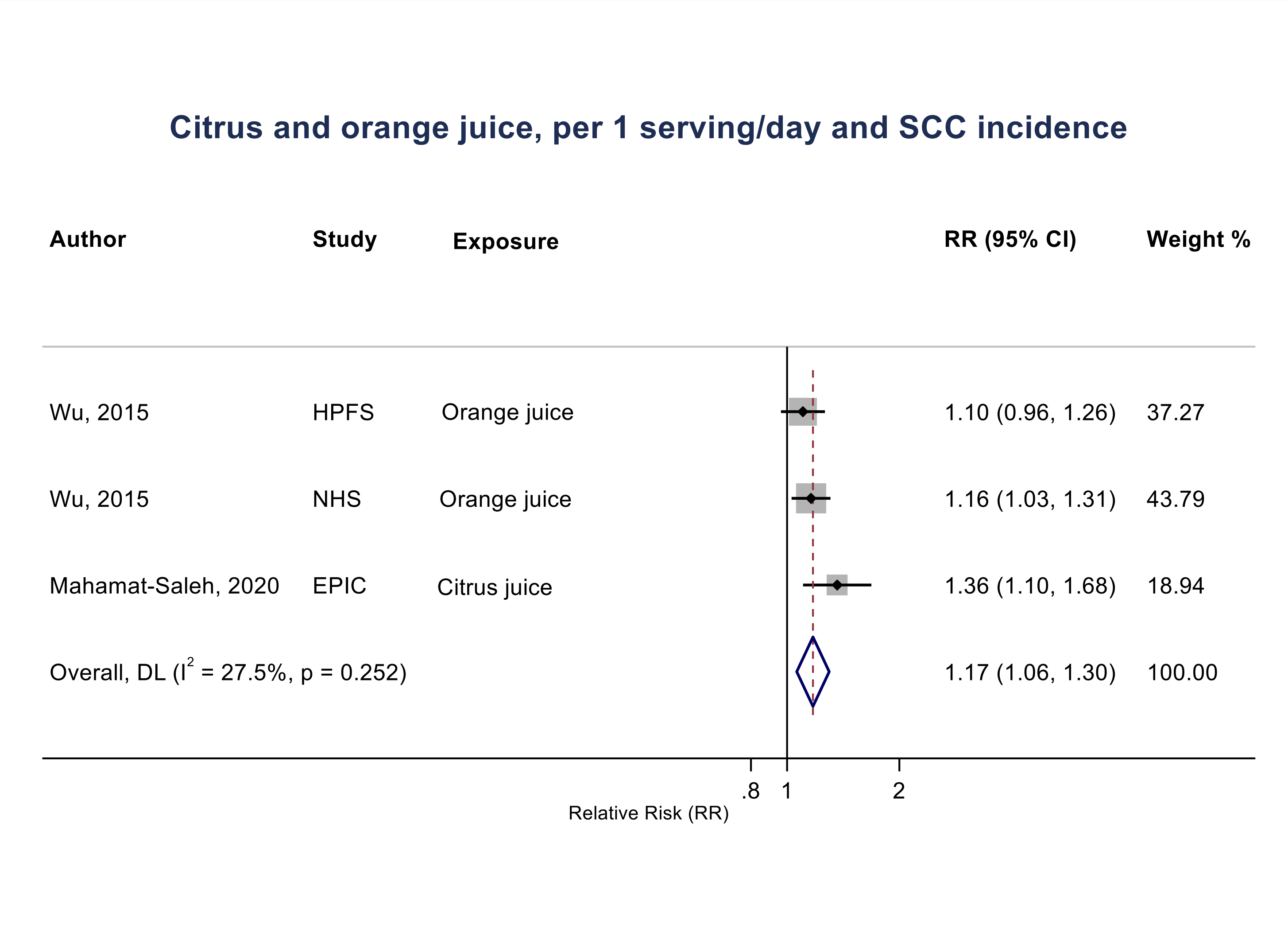

Figure legend: Forest plot shows the linear dose-response results (per 1 serving [177 mL] of juice/day) from the inverse variance DerSimonian-Laird random-effects model. The diamond represents the summary relative risk (RR) estimate, the diamond's width is the 95% confidence interval (CI). Each square and the horizontal line across the square represent the RR estimate of the individual study and its 95% CI.

Abbreviations: CI, confidence interval; RR, relative risk.

Study abbreviations: HPFS, Health Professionals Follow-up Study; NHS, Nurses' Health Study, EPIC, European prospective Investigation into Cancer and Nutrition.

Wu, 2015[171], Mahmat-Saleh, 2020[107]

**Supplementary Figure 275. Linear dose-response meta-analysis of grapefruit juice and melanoma incidence.**

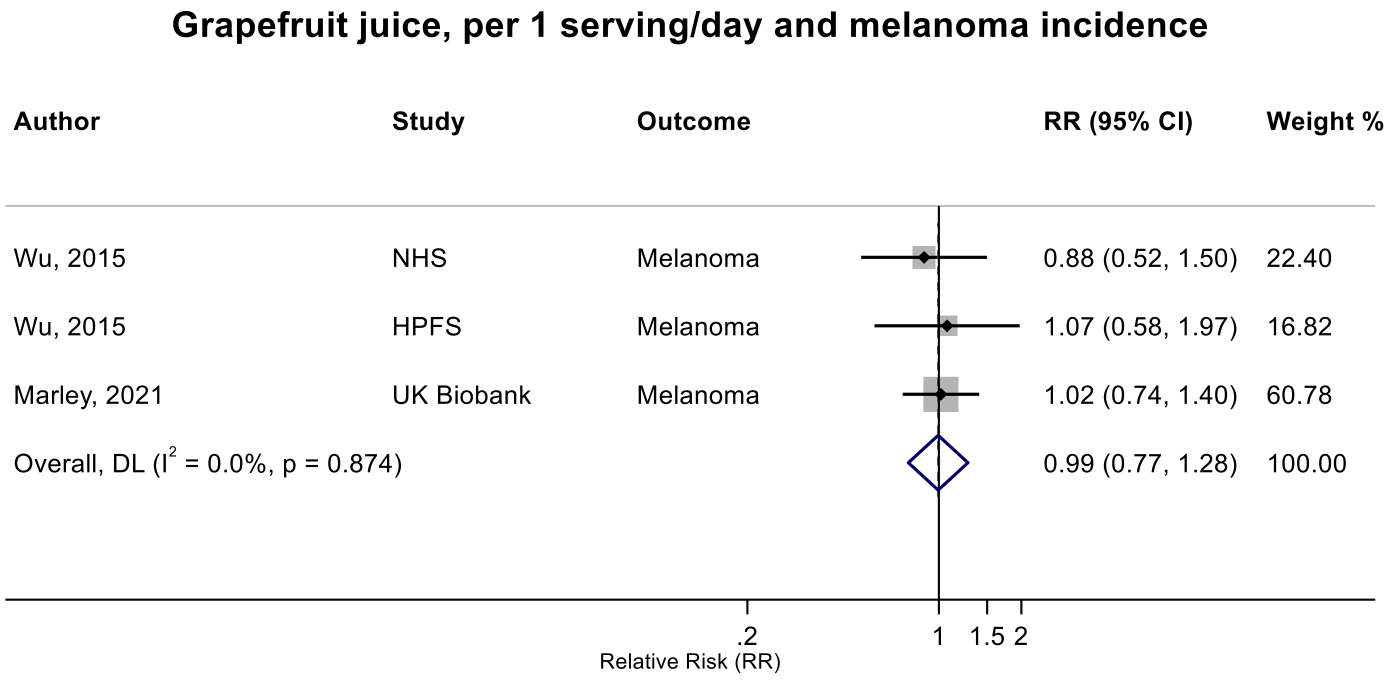

Figure legend: Forest plot shows the linear dose-response results (per 1 serving [177 mL] of juice/day) from the inverse variance DerSimonian-Laird random-effects model. The diamond represents the summary relative risk (RR) estimate, the diamond's width is the 95% confidence interval (CI). Each square and the horizontal line across the square represent the RR estimate of the individual study and its 95% CI.

Abbreviations: CI, confidence interval; RR, relative risk.

Study abbreviations: NHS, Nurses' Health Study; HPFS, Health Professionals Follow-up Study; UK Biobank, United Kingdom Biobank Study

Wu, 2015[171], Marley, 2021[110]

**Supplementary Figure 276. Linear dose-response meta-analysis of grapefruit juice and basal cell carcinoma incidence.**

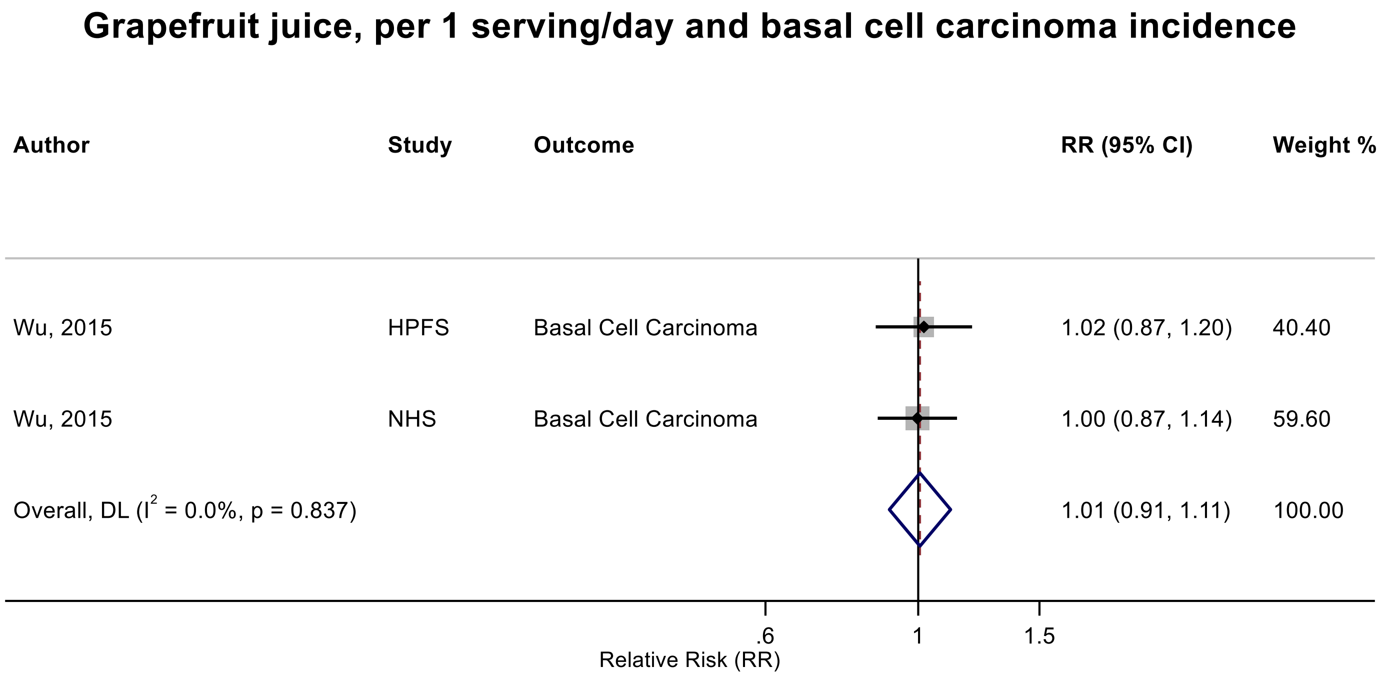

Figure legend: Forest plot shows the linear dose-response results (per 1 serving [177 mL] of juice/day) from the inverse variance DerSimonian-Laird random-effects model. The diamond represents the summary relative risk (RR) estimate, the diamond's width is the 95% confidence interval (CI). Each square and the horizontal line across the square represent the RR estimate of the individual study and its 95% CI.

Abbreviations: CI, confidence interval; RR, relative risk.

Study abbreviations: HPFS, Health Professionals Follow-up Study; NHS, Nurses' Health Study

Wu, 2015[170]

**Supplementary Figure 277. Linear dose-response meta-analysis of grapefruit juice and incidence of squamous cell carcinoma of the skin.**

#### Grapefruit juice, per 1 serving/day and squamous cell carcinoma incidence

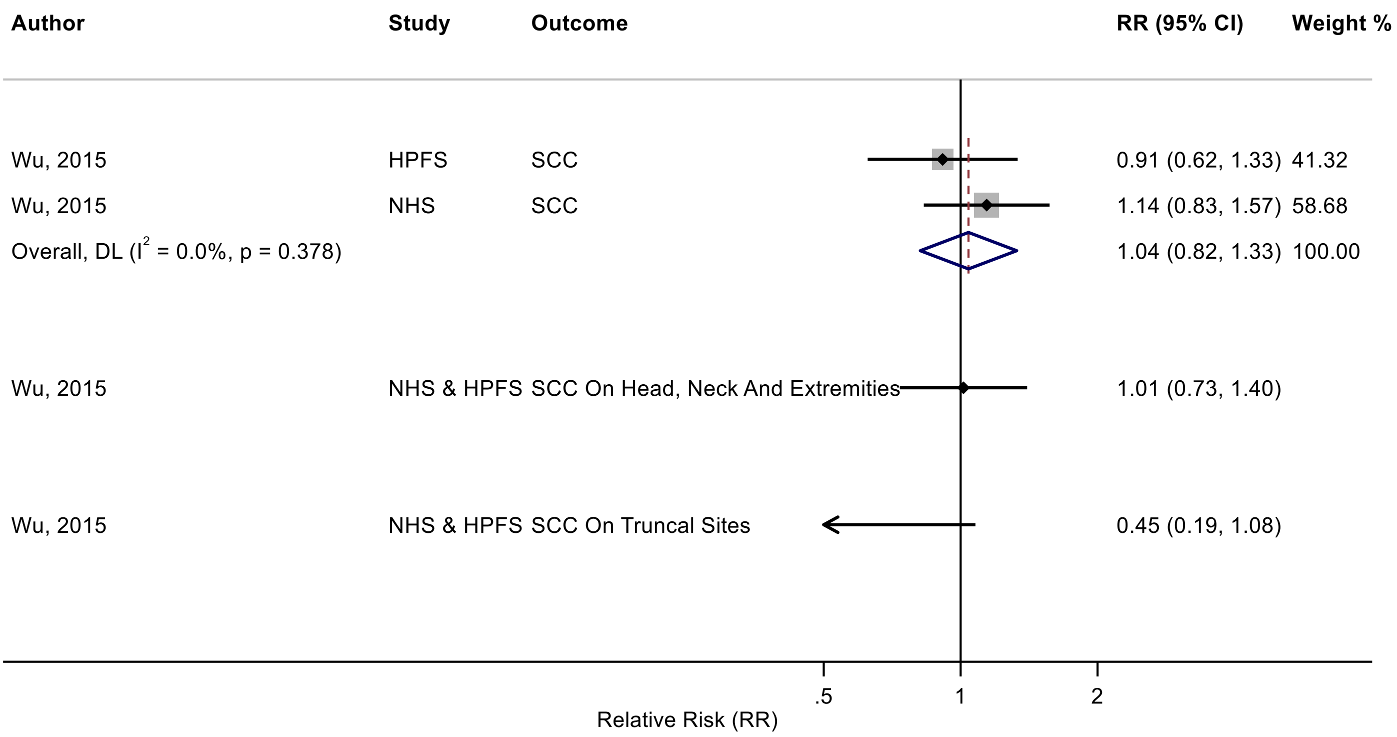

Figure legend: Forest plot shows the linear dose-response results (per 1 serving [177 mL] of juice/day) from the inverse variance DerSimonian-Laird random-effects model. The diamond represents the summary relative risk (RR) estimate, the diamond's width is the 95% confidence interval (CI). Each square and the horizontal line across the square represent the RR estimate of the individual study and its 95% CI.

Abbreviations: CI, confidence interval; RR, relative risk.

Study abbreviations: HPFS, Health Professionals Follow-up Study; NHS, Nurses' Health Study; NHS & HPFS, Nurses' Health Study & Health Professionals Follow-up Study

Wu, 2015[170]

**Supplementary Figure 278. Linear dose-response meta-analysis of citrus juice and prostate cancer incidence.**

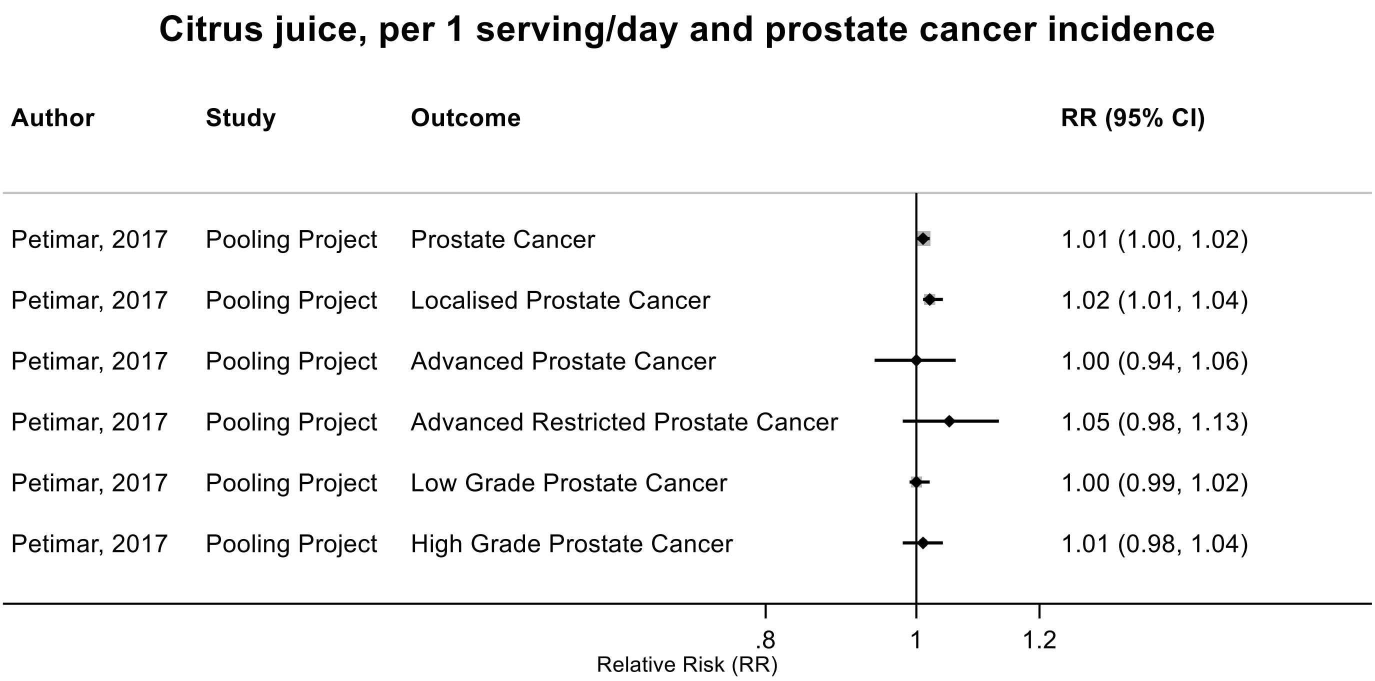

Figure legend: Forest plot shows the linear dose-response results (per 1 serving [177 mL] of juice/day) from the inverse variance DerSimonian-Laird random-effects model. Each square represents the summary relative risk (RR) estimate and the square's horizontal line is the 95% confidence interval (CI).

Abbreviations: CI, confidence interval; RR, relative risk.

Study abbreviations: Pooling Project, The Pooling Project of Prospective Studies of Diet and Cancer (n=15).

Petimar, 2017[139]

**Supplementary Figure 279. Linear dose-response meta-analysis of juices and liver cancer incidence.**

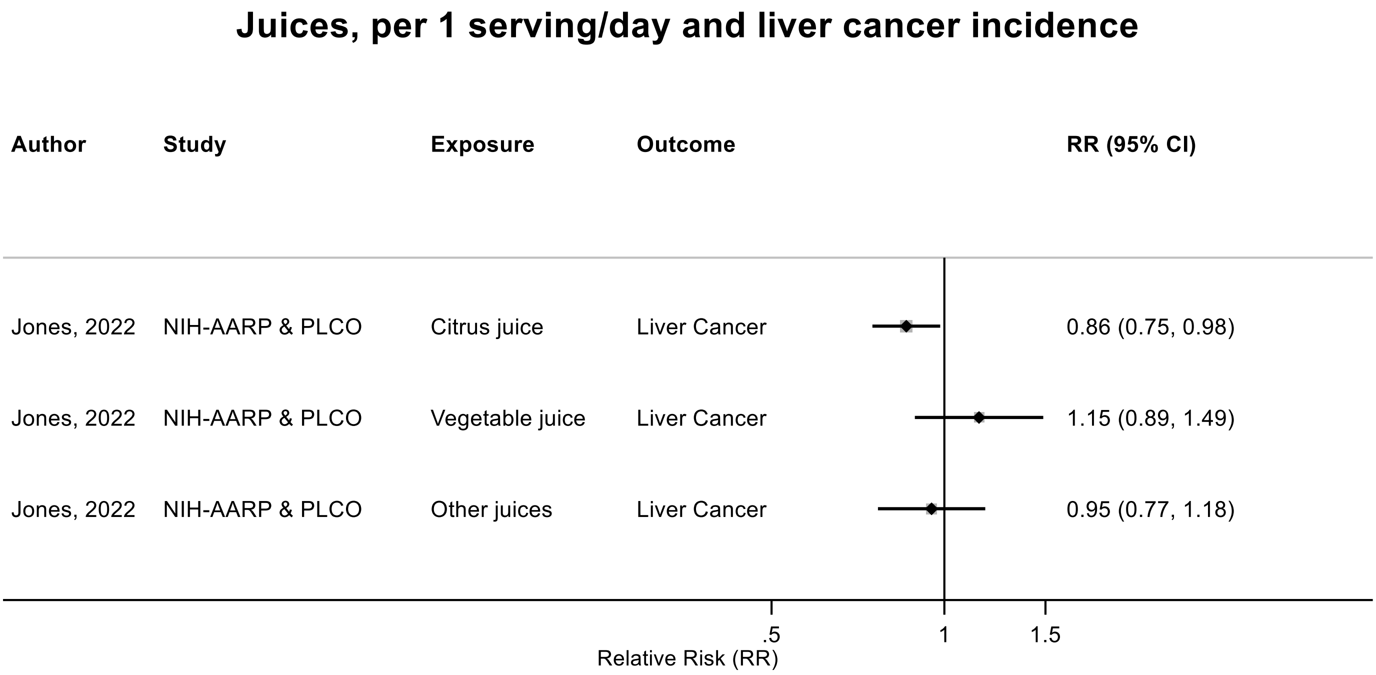

Figure legend: Forest plot shows the linear dose-response results (per 1 serving [177 mL] of juice/day) from the inverse variance DerSimonian-Laird random-effects model. Each square represents the summary relative risk (RR) estimate and the square's horizontal line is the 95% confidence interval (CI).

Abbreviations: CI, confidence interval; RR, relative risk.

Study abbreviations: NIH-AARP & PLCO, National Institute of Health + American Association of Retired Persons Diet and Health Study & Prostate, Lung, Colorectal and Ovarian Cancer Screening trial

Jones, 2022[79]

**Supplementary Figure 280. Linear dose-response meta-analysis of citrus juice and lung cancer incidence.**

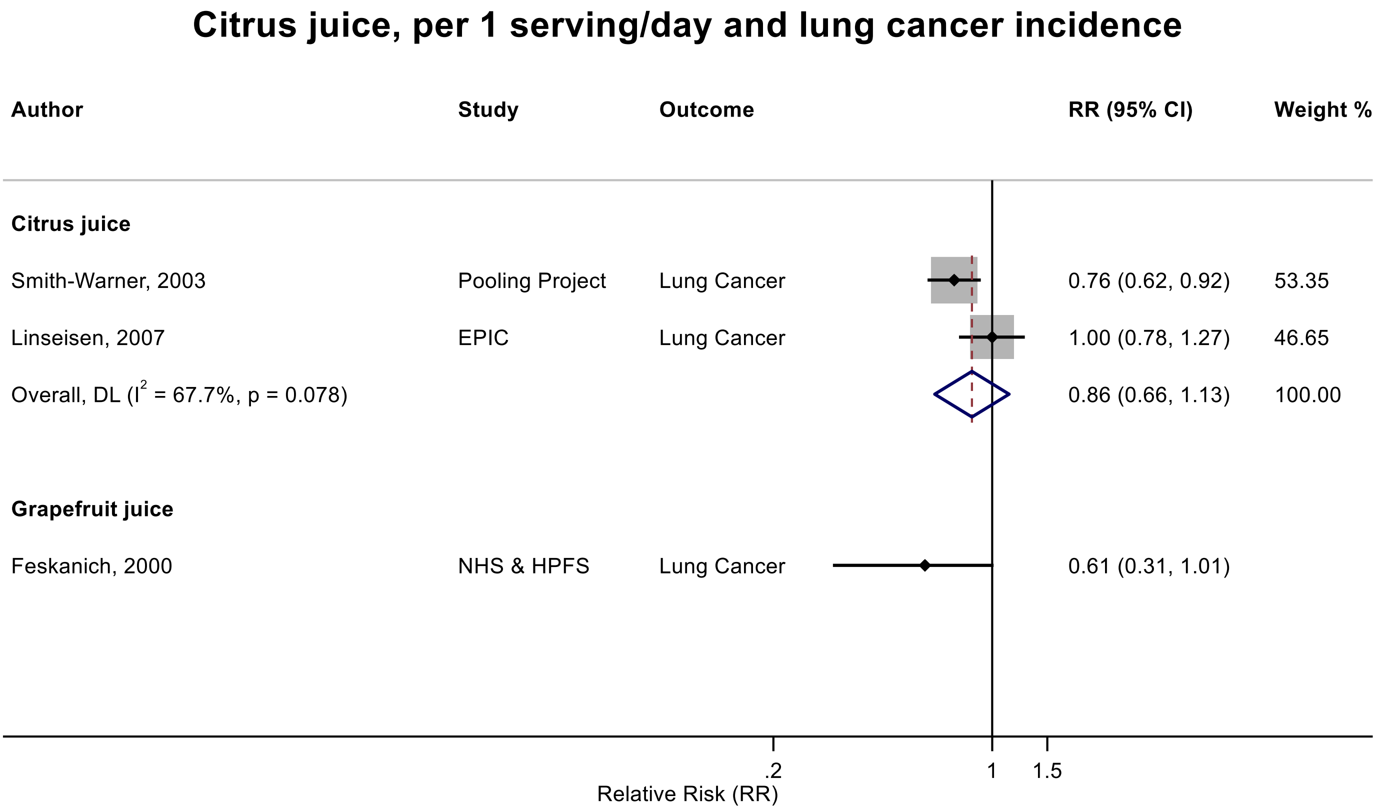

Figure legend: Forest plot shows the linear dose-response results (per 1 serving [177 mL] of juice/day) from the inverse variance DerSimonian-Laird random-effects model. The diamond represents the summary relative risk (RR) estimate, the diamond's width is the 95% confidence interval (CI). Each square and the horizontal line across the square represent the RR estimate of the individual study and its 95% CI.

Abbreviations: CI, confidence interval; RR, relative risk.

Study abbreviations: Pooling Project, The Pooling Project of Prospective Studies of Diet and Cancer (n=8); EPIC, European Prospective Investigation into Cancer and nutrition; NHS & HPFS, Nurses' Health Study & Health Professionals Follow-up Study

Smith-Warner, 2003[155], Linseisen, 2007[104], Feskanich, 2000[55]

**Supplementary Figure 281. Linear dose-response meta-analysis of orange juices and breast cancer incidence.**

##### Oranje juice, per 1 serving/day and breast cancer incidence

| Author | Study | Outcome | RR (95% CI) |
|---|---|---|---|
| Farvid, 2018 | NHS I & II | Breast Cancer ER+ | 1.00 (0.97, 1.04) |
| Farvid, 2018 | NHS I & II | Breast Cancer ER- | 1.04 (0.97, 1.11) |
| Farvid, 2018 | NHS I & II | Breast Cancer | 1.03 (1.00, 1.04) |

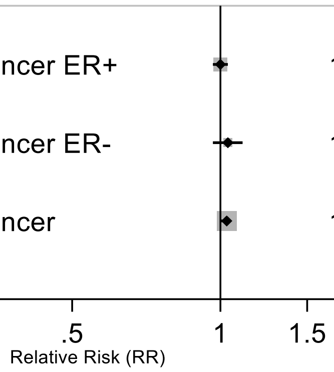

Figure legend: Forest plot shows the linear dose-response results (per 1 serving [177 mL] of juice/day) from the Cox proportional hazard regression model pooling two cohorts. Each square represents the summary relative risk (RR) estimate and the square's horizontal line is the 95% confidence interval (CI).

Abbreviations: CI, confidence interval; ER, Estrogen receptor, RR, relative risk.

Study abbreviations: NHS I & II, Nurses' Health Study I & II

Note: Both studies performed repeated measurements.

Farvid, 2019[52]

**Supplementary Figure 282. Linear dose-response meta-analysis of grapefruit juice and breast cancer incidence.**

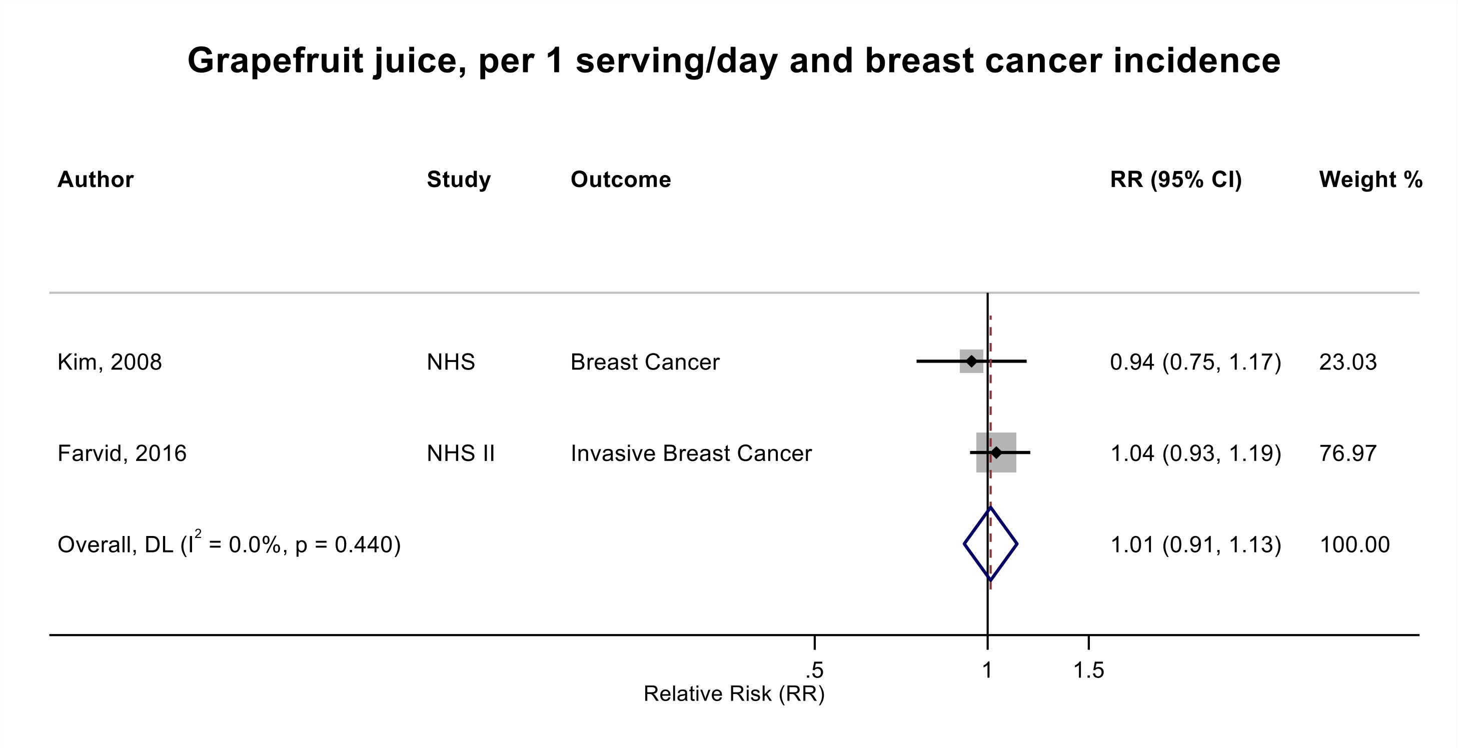

Figure legend: Forest plot shows the linear dose-response results (per 1 serving [177 mL] of juice/day) from the inverse variance DerSimonian-Laird random-effects model. The diamond represents the summary relative risk (RR) estimate, the diamond's width is the 95% confidence interval (CI). Each square and the horizontal line across the square represent the RR estimate of the individual study and its 95% CI.

Abbreviations: CI, confidence interval; RR, relative risk.

Study abbreviations: NHS, Nurses' Health Study; NHS II, Nurses' Health Study II

Kim, 2008[86], Farvid, 2019[52]

**Supplementary Figure 283. Linear dose-response meta-analysis of apple juice and breast cancer incidence.**

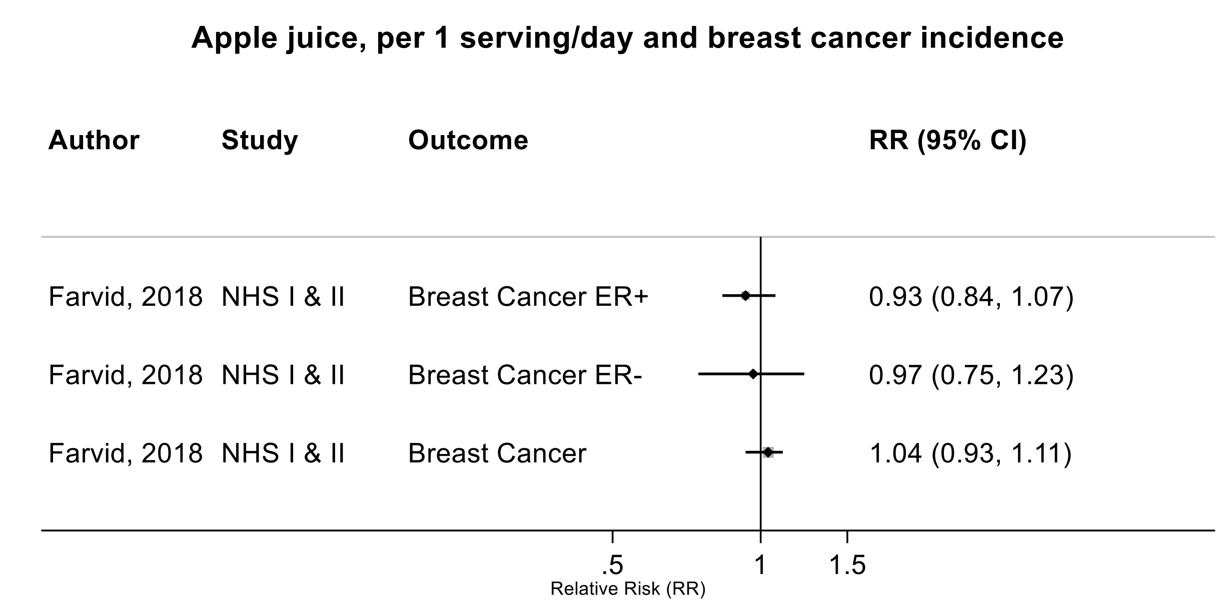

Figure legend: Forest plot shows the linear dose-response results (per 1 serving [177 mL] of juice/day) from the Cox proportional regression model pooling two cohort studies. Each square and the horizontal line across the square represent the RR estimate of the individual study and its 95% CI.

Abbreviations: CI, confidence interval; RR, relative risk.

Study abbreviations: NHS I & II, Nurses' Health Study I & II

Farvid, 2018[52]

**Supplementary Figure 284. Linear dose-response meta-analysis of other fruit juice (not orange and apple) and breast cancer incidence.**

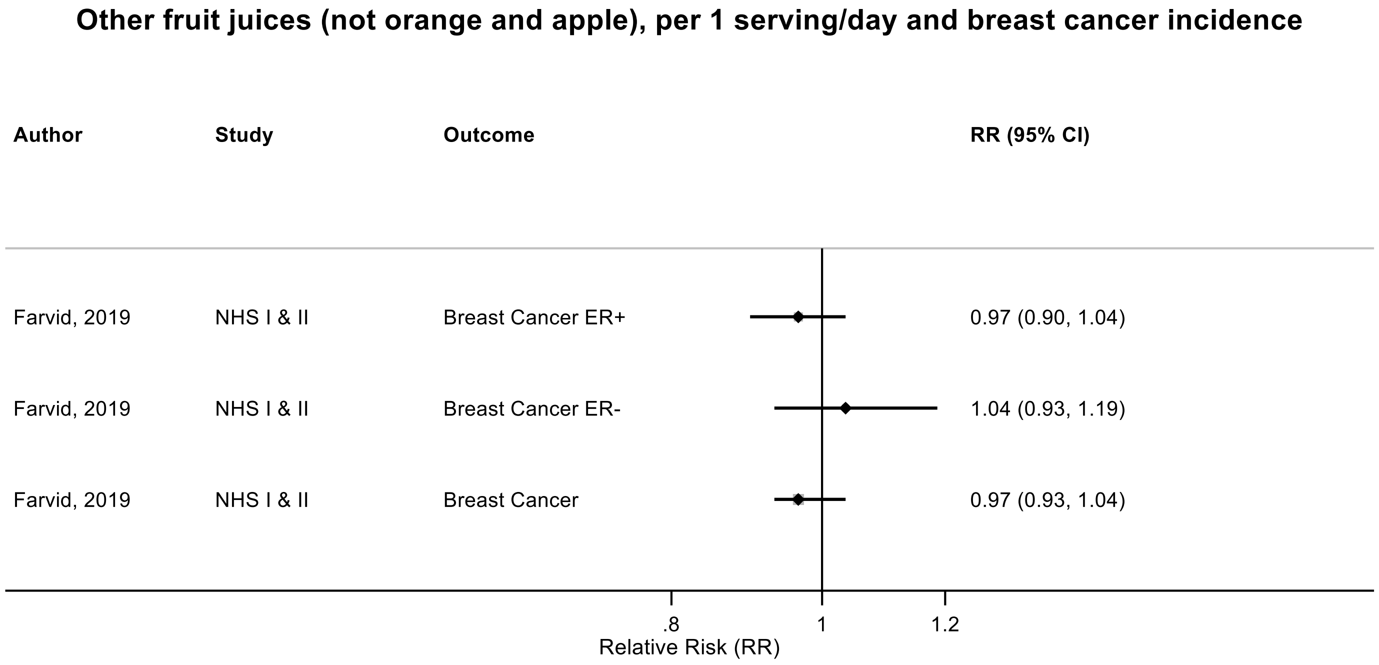

Figure legend: Forest plot shows the linear dose-response results (per 1 serving [177 mL] of juice/day) from the Cox proportional regression model pooling two cohort studies. Each square and the horizontal line across the square represent the RR estimate of the individual study and its 95% CI.

Abbreviations: CI, confidence interval; RR, relative risk.

Study abbreviations: NHS I & II, Nurses' Health Study I and II

Farvid, 2019[52]

**Supplementary Figure 285. Linear dose-response meta-analysis of fruit and vegetable juice and pancreatic cancer incidence.**

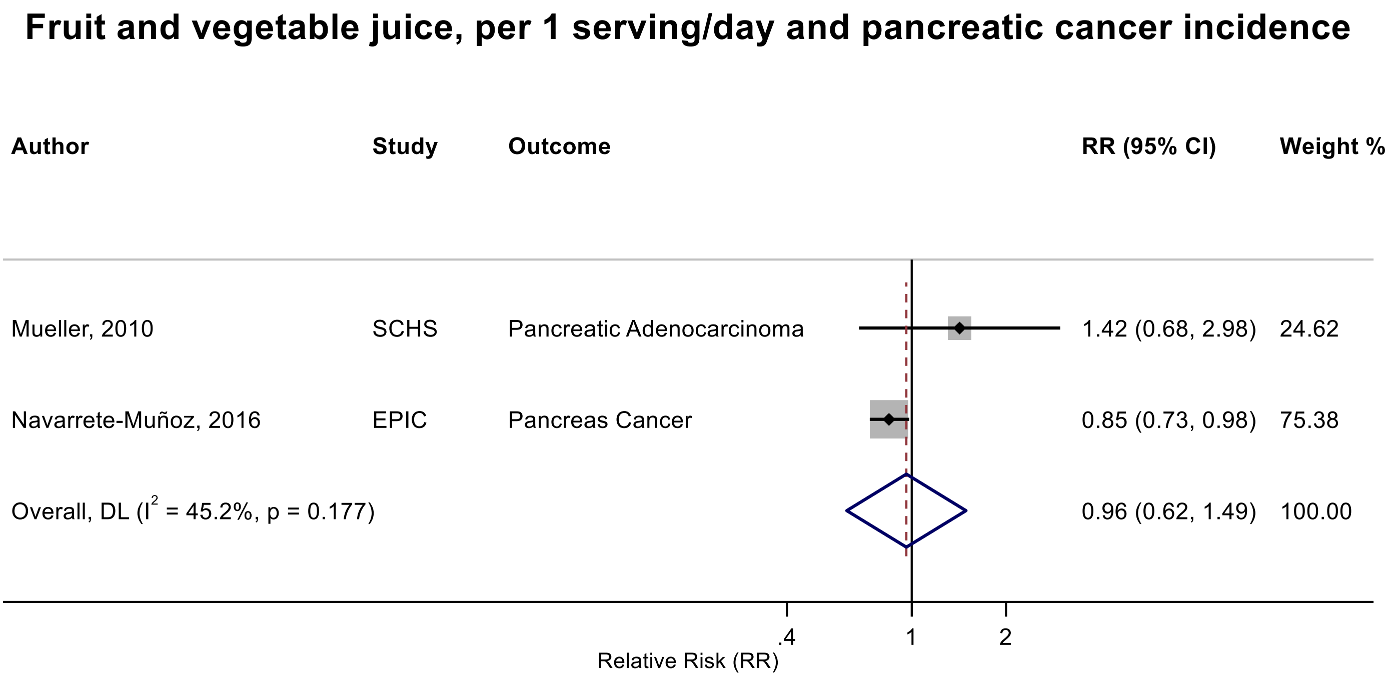

Figure legend: Forest plot shows the linear dose-response results (per 1 serving [177 mL] of juice/day) from the inverse variance DerSimonian-Laird random-effects model. The diamond represents the summary relative risk (RR) estimate, the diamond's width is the 95% confidence interval (CI). Each square and the horizontal line across the square represent the RR estimate of the individual study and its 95% CI.

Abbreviations: CI, confidence interval; RR, relative risk.

Study abbreviations: SCHS, Singapore Chinese Health study; EPIC, European Prospective Investigation into Cancer and nutrition

Mueller, 2010[125], Navarrete-Muñoz, 2016[127]

**Supplementary Figure 286. Linear dose-response meta-analysis of fruit and vegetable juice and incidence of colorectal cancer and its subsites.**

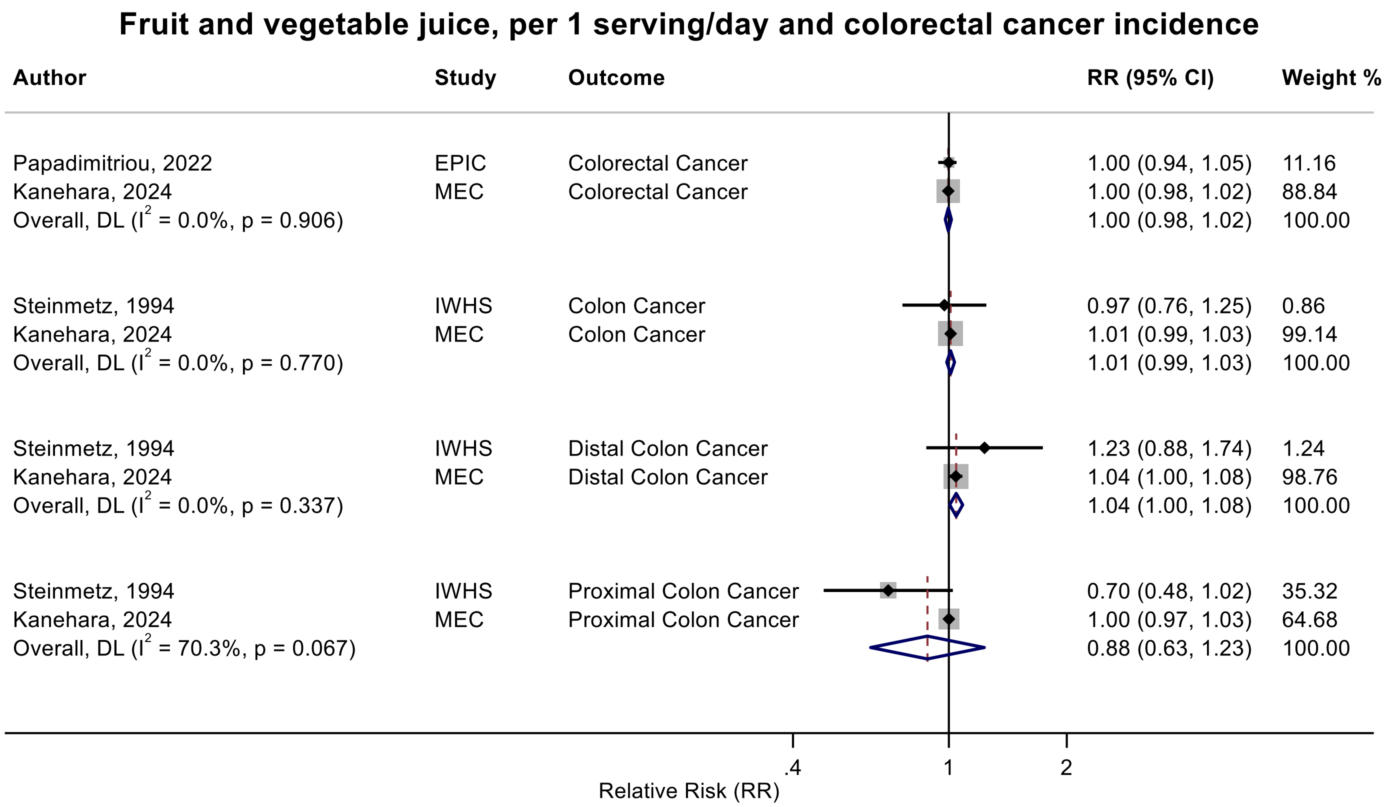

Figure legend: Forest plot shows the linear dose-response results (per 1 serving [177 mL] of juice/day) from the inverse variance DerSimonian-Laird random-effects model. The diamond represents the summary relative risk (RR) estimate, the diamond's width is the 95% confidence interval (CI). Each square and the horizontal line across the square represent the RR estimate of the individual study and its 95% CI.

Abbreviations: CI, confidence interval; RR, relative risk.

Study abbreviations: EPIC, European Prospective Investigation into Cancer and nutrition; MEC, Multiethnic Cohort Study; IWHS, The Iowa Women's Health Study

Papadimitriou, 2022[136], Kanehara, 2024[81], Steinmetz, 1994[158]

**Supplementary Figure 287. Linear dose-response meta-analysis of fruit and vegetable juice and lung cancer incidence.**

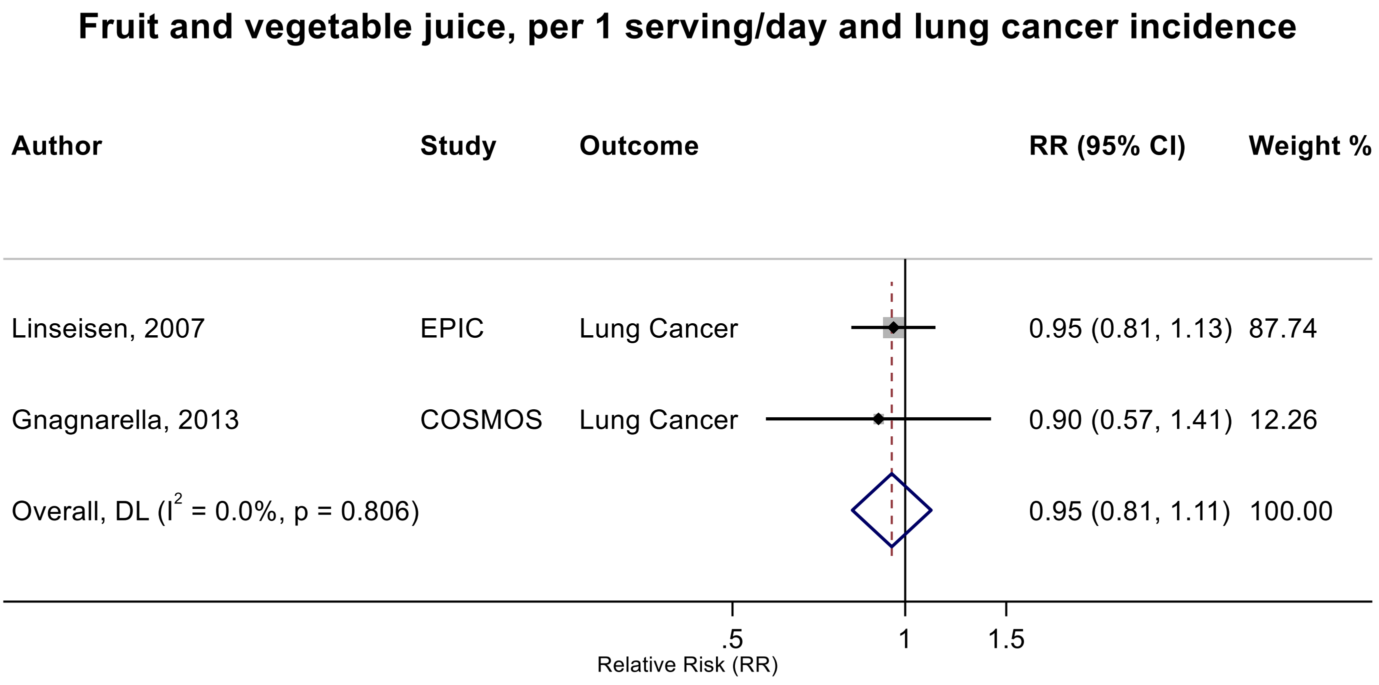

Figure legend: Forest plot shows the linear dose-response results (per 1 serving [177 mL] of juice/day) from the inverse variance DerSimonian-Laird random-effects model. The diamond represents the summary relative risk (RR) estimate, the diamond's width is the 95% confidence interval (CI). Each square and the horizontal line across the square represent the RR estimate of the individual study and its 95% CI.

Abbreviations: CI, confidence interval; RR, relative risk.

Study abbreviations: EPIC, European Prospective Investigation into Cancer and nutrition; COSMOS, Continuous Observation of Smoking Subjects Study

Linseisen, 2007[104], Gnagnarella, 2013[65]

**Supplementary Figure 288. Linear dose-response meta-analysis of fruit and vegetable juice and breast cancer incidence.**

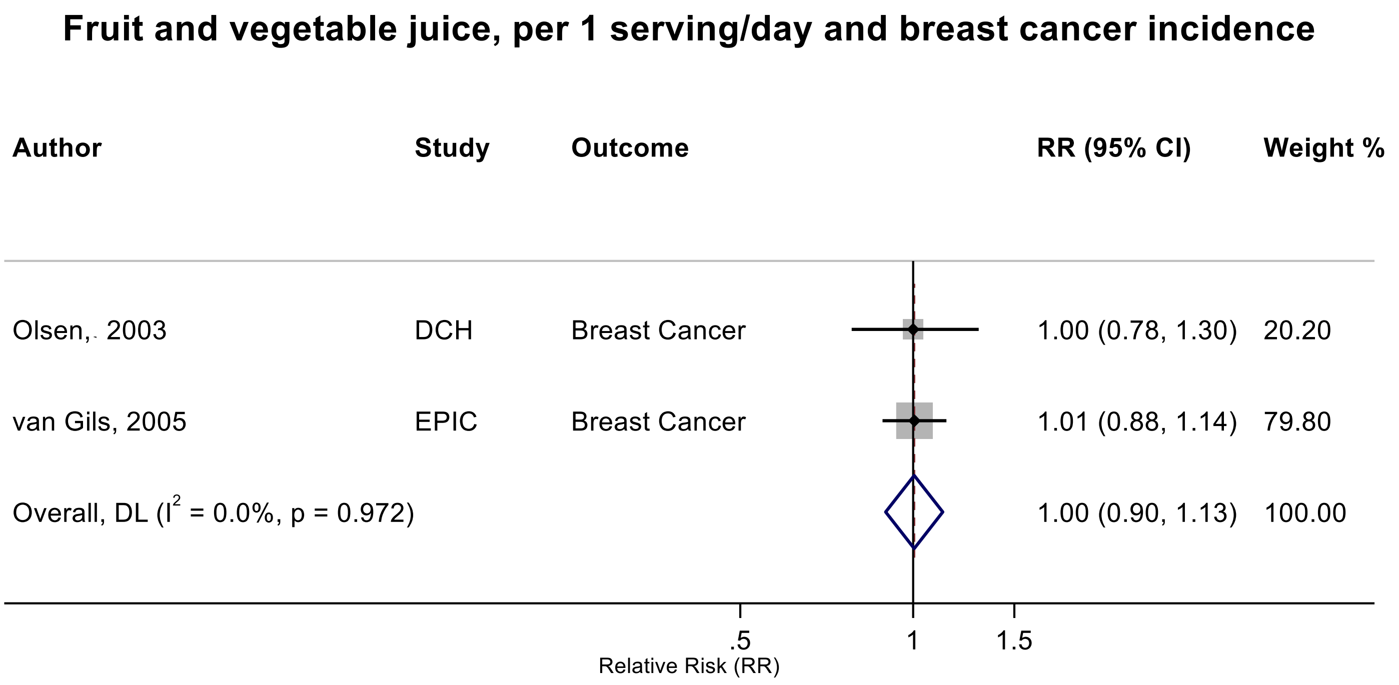

Figure legend: Forest plot shows the linear dose-response results (per 1 serving [177 mL] of juice/day) from the inverse variance DerSimonian-Laird random-effects model. The diamond represents the summary relative risk (RR) estimate, the diamond's width is the 95% confidence interval (CI). Each square and the horizontal line across the square represent the RR estimate of the individual study and its 95% CI.

Abbreviations: CI, confidence

Study abbreviations: DCH, The Diet, Cancer, and Health Study; EPIC, European Prospective Investigation into Cancer and nutrition

Olsen, 2003[133], van Gils, 2005[164]

**Supplementary Figure 289. Linear dose-response meta-analysis of vegetable juice and prostate cancer incidence.**

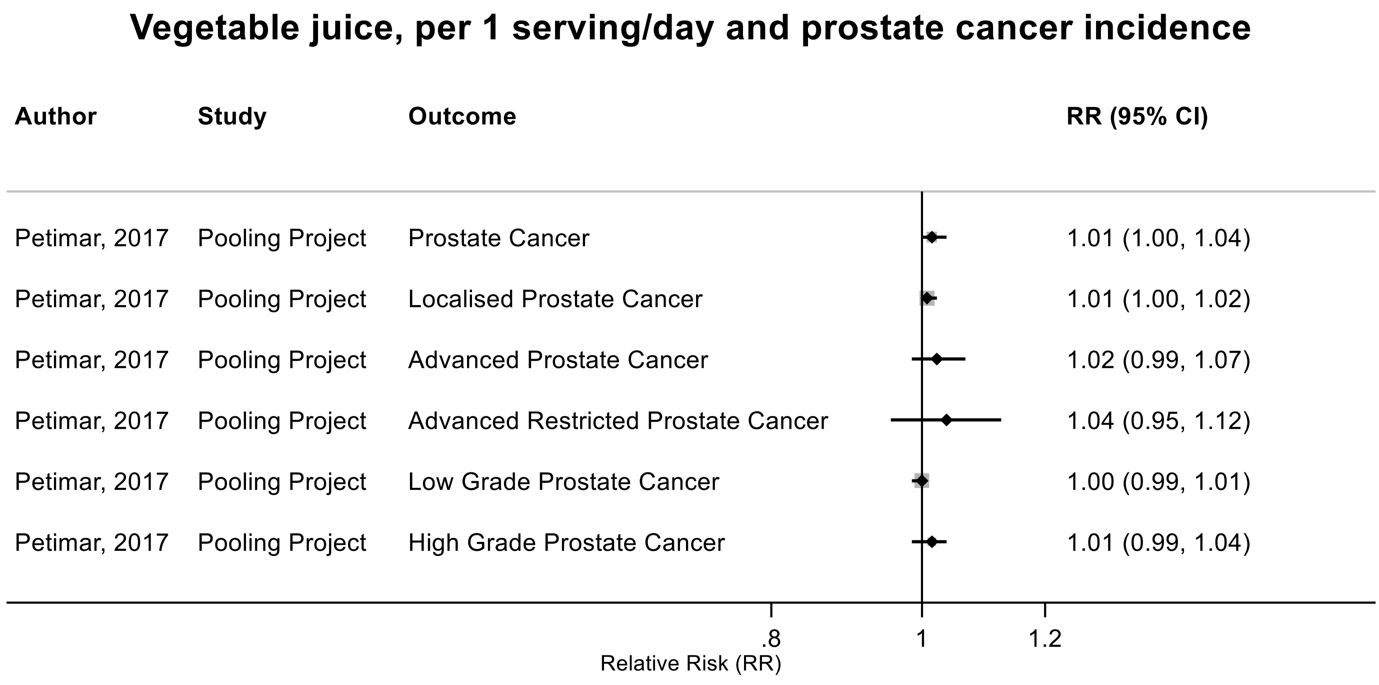

Figure legend: Forest plot shows the linear dose-response results (per 1 serving [177 mL] of juice/day) from the inverse variance DerSimonian-Laird random-effects model. Each square represents the summary relative risk (RR) estimate and the square's horizontal line is the 95% confidence interval (CI).

Abbreviations: CI, confidence interval; RR, relative risk.

Study abbreviations: Pooling Project, The Pooling Project of Prospective Studies of Diet and Cancer (n=15)

Petimar, 2017[139]

**Supplementary Figure 290. Linear dose-response meta-analysis of tomato juice and prostate cancer incidence.**

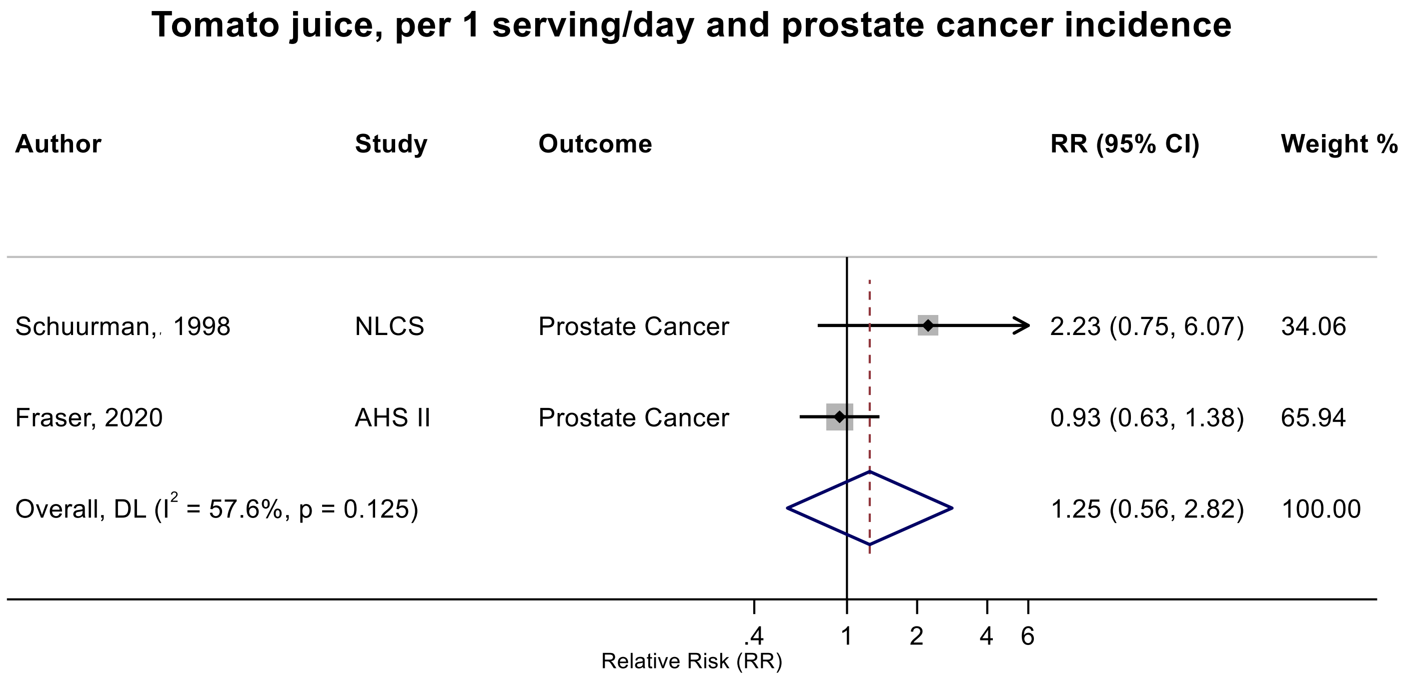

Figure legend: Forest plot shows the linear dose-response results (per 1 serving [177 mL] of juice/day) from the inverse variance DerSimonian-Laird random-effects model. The diamond represents the summary relative risk (RR) estimate, the diamond's width is the 95% confidence interval (CI). Each square and the horizontal line across the square represent the RR estimate of the individual study and its 95% CI.

Abbreviations: CI, confidence interval; RR, relative risk.

Study abbreviations: NLCS, Netherlands Cohort Study on Diet and Cancer; AHS II, Adventist Health Study

Schuurman, 1998[152], Fraser, 2020[56]

**Supplementary Figure 291. Linear dose-response meta-analysis of fruit and vegetable juice and kidney cancer incidence.**

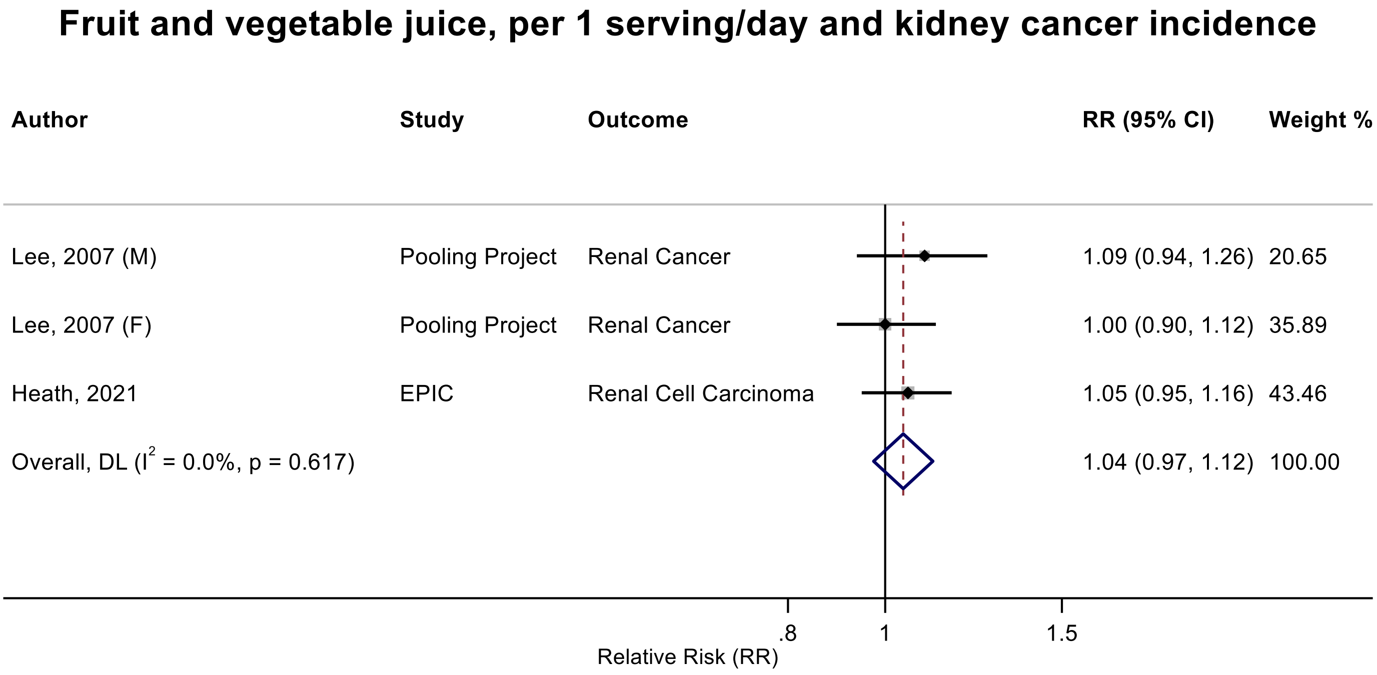

Figure legend: Forest plot shows the linear dose-response results (per 1 serving [177 mL] of juice/day) from the inverse variance DerSimonian-Laird random-effects model. The diamond represents the summary relative risk (RR) estimate, the diamond's width is the 95% confidence interval (CI). Each square and the horizontal line across the square represent the RR estimate of the individual study and its 95% CI.

Abbreviations: CI, confidence

Study abbreviations: Pooling Project, The Pooling Project of Prospective Studies of Diet and Cancer (n=13); EPIC, European Prospective Investigation into Cancer and nutrition

Lee, 2007[26], Heath, 2021[66]

**Supplementary Figure 292.Linear dose-response meta-analysis of fruit juice and breast cancer incidence by geographical region.**

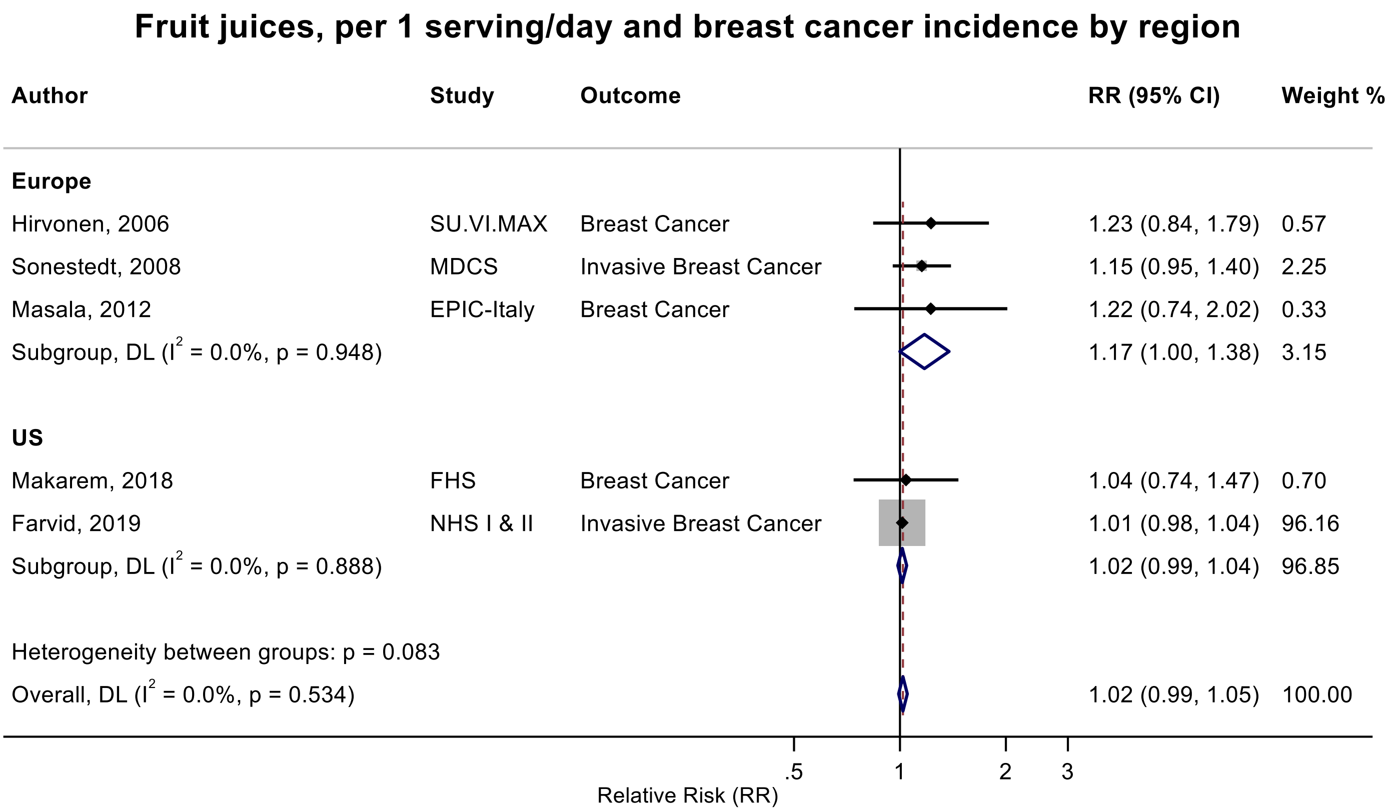

Figure legend: Forest plot shows the linear dose-response results (per 1 serving [177 mL] of juice/day) from the inverse variance DerSimonian-Laird random-effects model. The diamond represents the summary relative risk (RR) estimate, the diamond's width is the 95% confidence interval (CI). Each square and the horizontal line across the square represent the RR estimate of the individual study and its 95% CI.

Abbreviations: CI, confidence interval; RR, relative risk.

Study abbreviations: SU.VI.MAX, Supplémentation en Vitamines et Minéraux Antioxydants Study; MDCS, Malmö Diet and Cancer Study; EPIC-Italy, European Prospective Investigation into Cancer and nutrition Italy cohort; FHS, Framingham Heart Study; NHS I & II, Nurses' Health Study I & II

Hirvonen, 2006[69], Sonestedt, 2008[156], Masala, 2012[112], Makarem, 2018[108], Farvid, 2019[52]

**Supplementary Figure 293. Linear dose-response meta-analysis of fruit juice and breast cancer incidence by frequency of dietary assessment.**

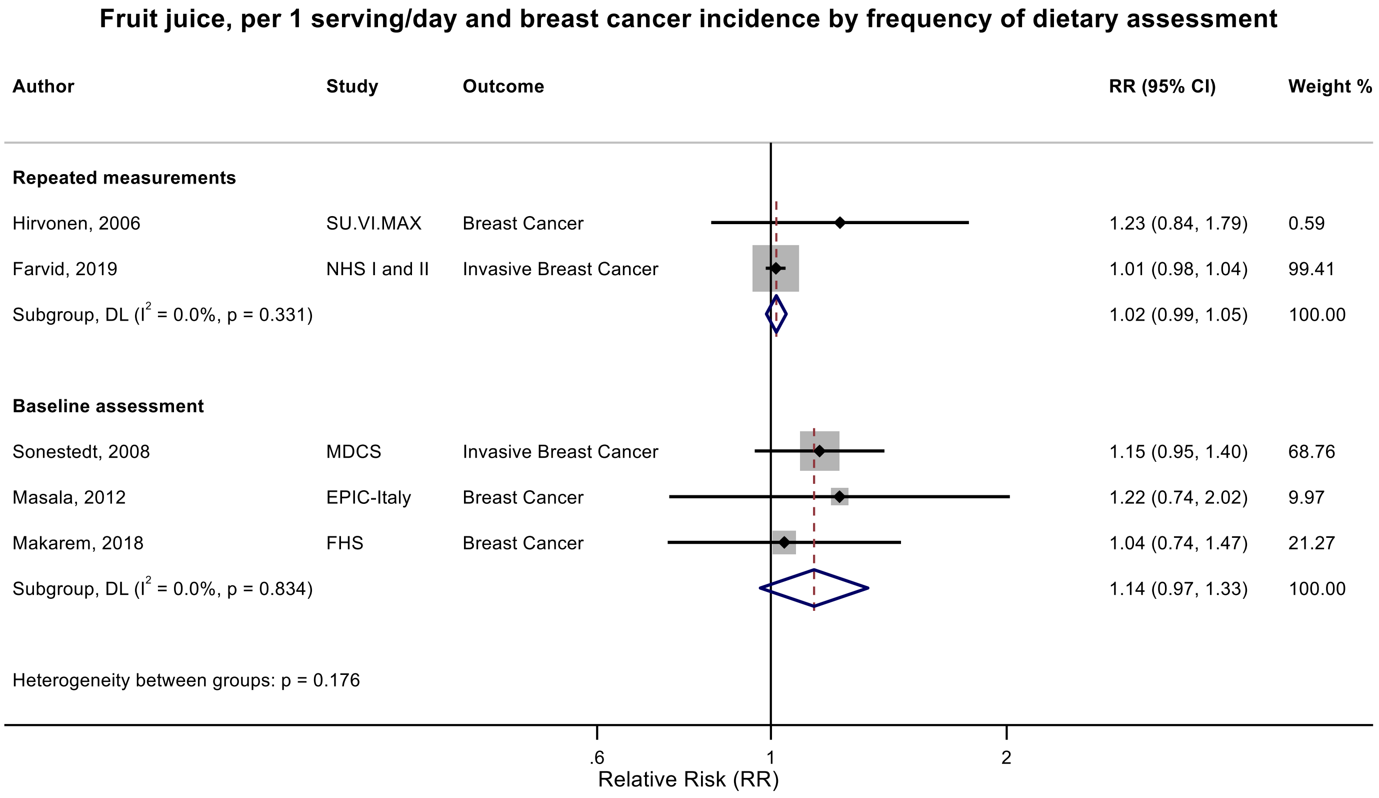

Figure legend: Forest plot shows the linear dose-response results (per 1 serving [177 mL] of juice/day) from the inverse variance DerSimonian-Laird random-effects model. The diamond represents the summary relative risk (RR) estimate, the diamond's width is the 95% confidence interval (CI). Each square and the horizontal line across the square represent the RR estimate of the individual study and its 95% CI.

Abbreviations: CI, confidence interval; RR, relative risk.

Study abbreviations: SU.VI.MAX, Supplémentation en Vitamines et Minéraux Antioxydants Study; NHS I & II, Nurses' Health Study I & II; MDCS, Malmö Diet and Cancer Study; EPIC-Italy, European Prospective Investigation into Cancer and nutrition Italy cohort; FHS, Framingham Heart Study

Hirvonen, 2006[69], Farvid, 2019[52], Sonestedt, 2008[156], Masala, 2012[112], Makarem, 2018[108]

**Supplementary Figure 294. Linear dose-response meta-analysis of citrus juice and melanoma incidence by geographical region.**

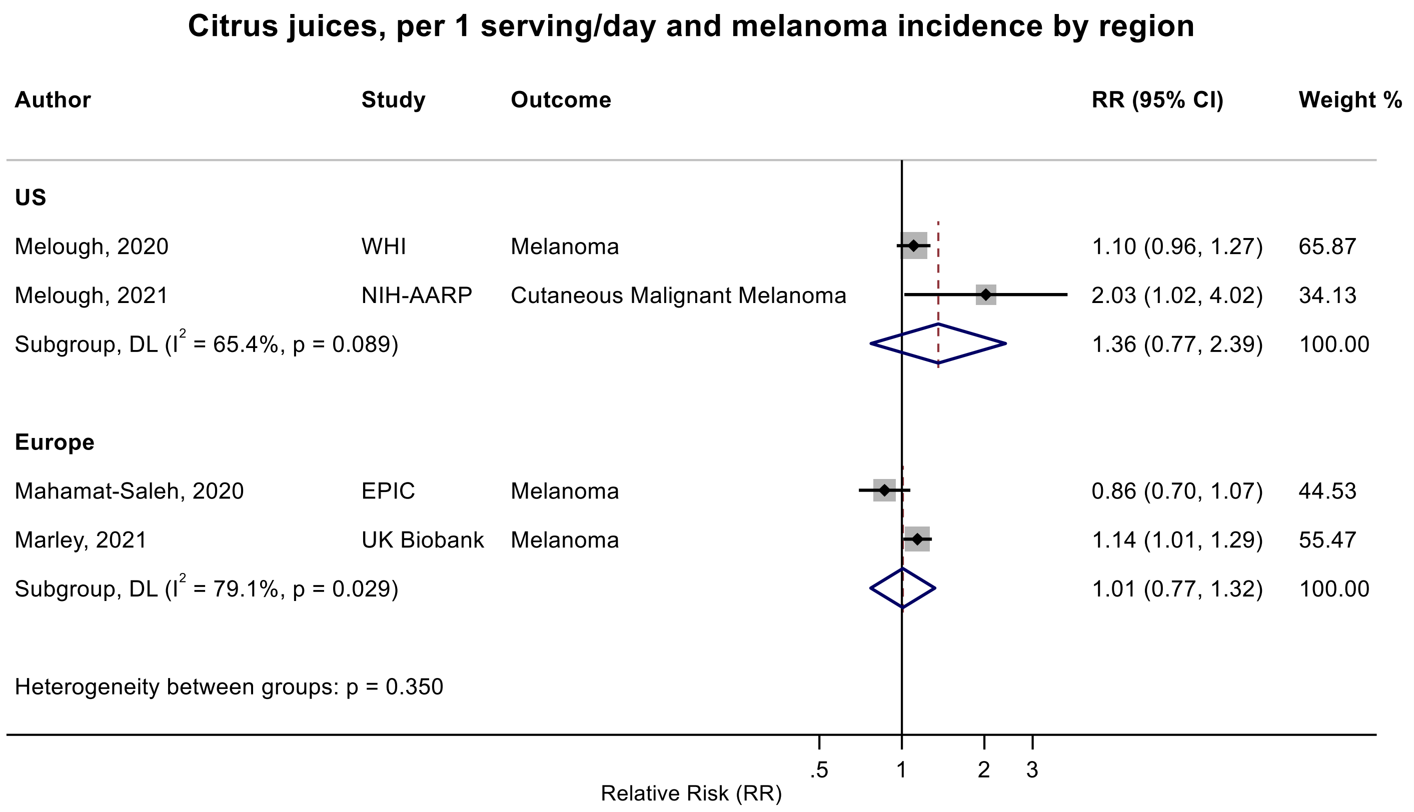

Figure legend: Forest plot shows the linear dose-response results (per 1 serving [177 mL] of juice/day) from the inverse variance DerSimonian-Laird random-effects model. The diamond represents the summary relative risk (RR) estimate, the diamond's width is the 95% confidence interval (CI). Each square and the horizontal line across the square represent the RR estimate of the individual study and its 95% CI.

Abbreviations: CI, confidence interval; RR, relative risk.

Study abbreviations: WHI, Women's Health Initiative Study; NIH-AARP, National Institute of Health + American Association of Retired Persons Diet and Health Study; EPIC, European Prospective Investigation into Cancer and nutrition; UK Biobank, United Kingdom Biobank Study

Melough, 2020[116], Melough, 2021[115], Mahamat-Saleh, 2020[107], Marley, 2021[110]

**Supplementary Figure 295. Linear dose-response meta-analysis of fruit juice and glioma incidence by sex.**

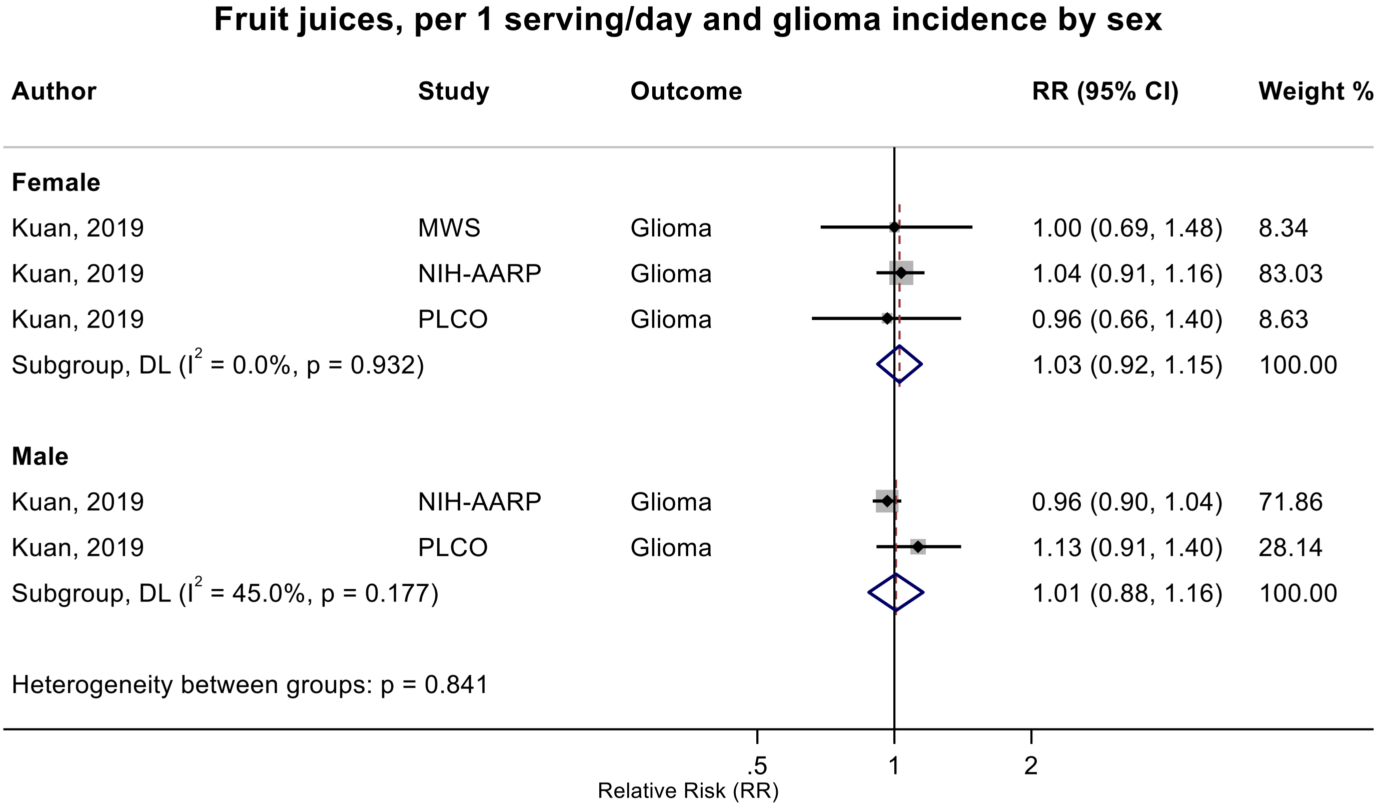

Figure legend: Forest plot shows the linear dose-response results (per 1 serving [177 mL] of juice/day) from the inverse variance DerSimonian-Laird random-effects model. The diamond represents the summary relative risk (RR) estimate, the diamond's width is the 95% confidence interval (CI). Each square and the horizontal line across the square represent the RR estimate of the individual study and its 95% CI.

Abbreviations: CI, confidence interval; RR, relative risk.

Study abbreviations: MWS, Million Women Study; NIH-AARP, National Institute of Health + American Association of Retired Persons Diet and Health Study; PLCO, Prostate, Lung, Colorectal and Ovarian Cancer Screening trial

Kuan, 2019[92]

**Supplementary Figure 296. Linear dose-response meta-analysis of fruit juice and glioma incidence by follow-up duration.**

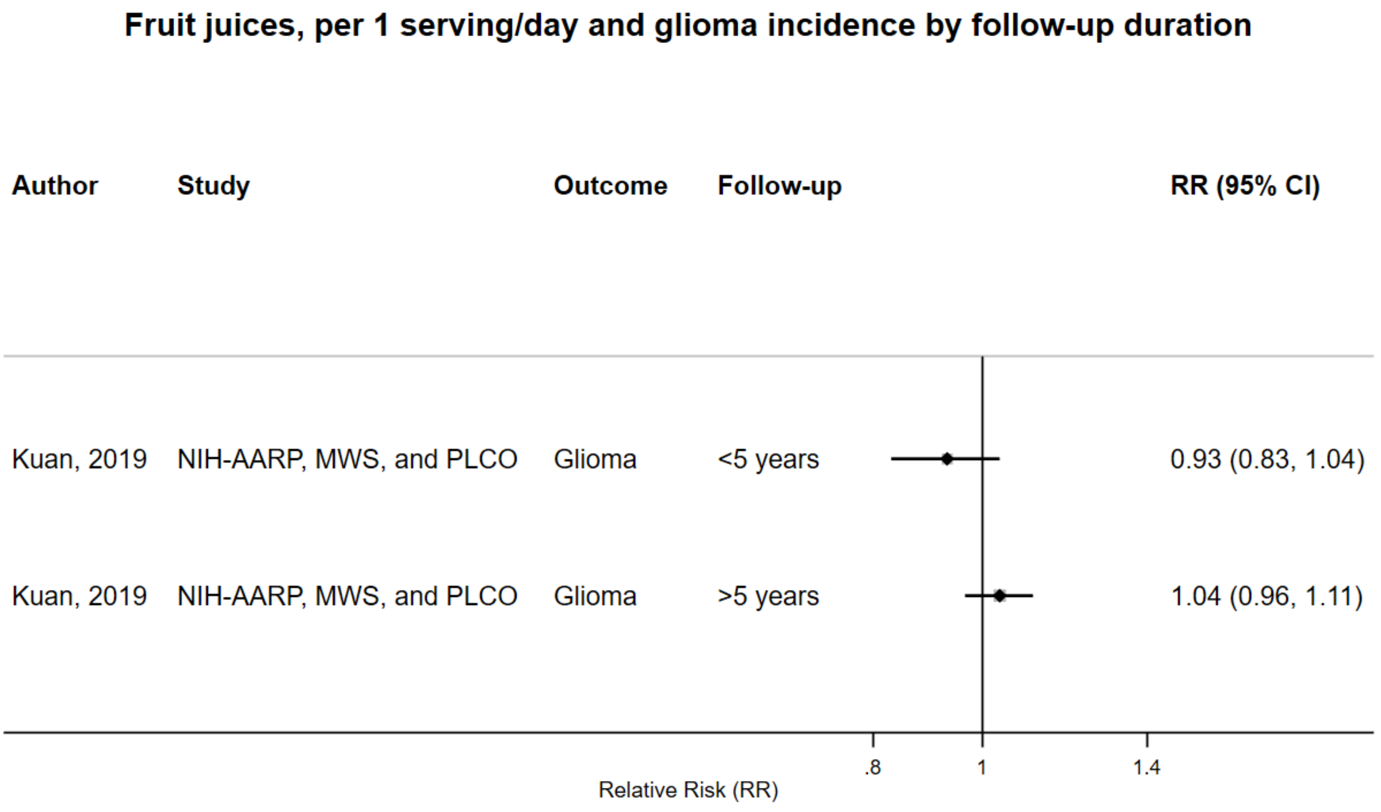

Figure legend: Forest plot shows the linear dose-response results (per 1 serving [177 mL] of juice/day) from the inverse variance DerSimonian-Laird random-effects model. The diamond represents the summary relative risk (RR) estimate, the diamond's width is the 95% confidence interval (CI). Each square and the horizontal line across the square represent the RR estimate of the individual study and its 95% CI.

Abbreviations: CI, confidence interval; RR, relative risk.

Study abbreviations: NIH-AARP, MWS and PLCO, National Institute of Health + American Association of Retired Persons Diet and Health Study, Million Women Study and Prostate, Lung, Colorectal and Ovarian Cancer Screening trial

Note: This subgroup analysis is based on the information presented in the published pooled analysis.

Kuan, 2019[92]

**Supplementary Figure 297. Linear dose-response meta-analysis of fruit juice and kidney cancer incidence frequency of dietary assessment.**

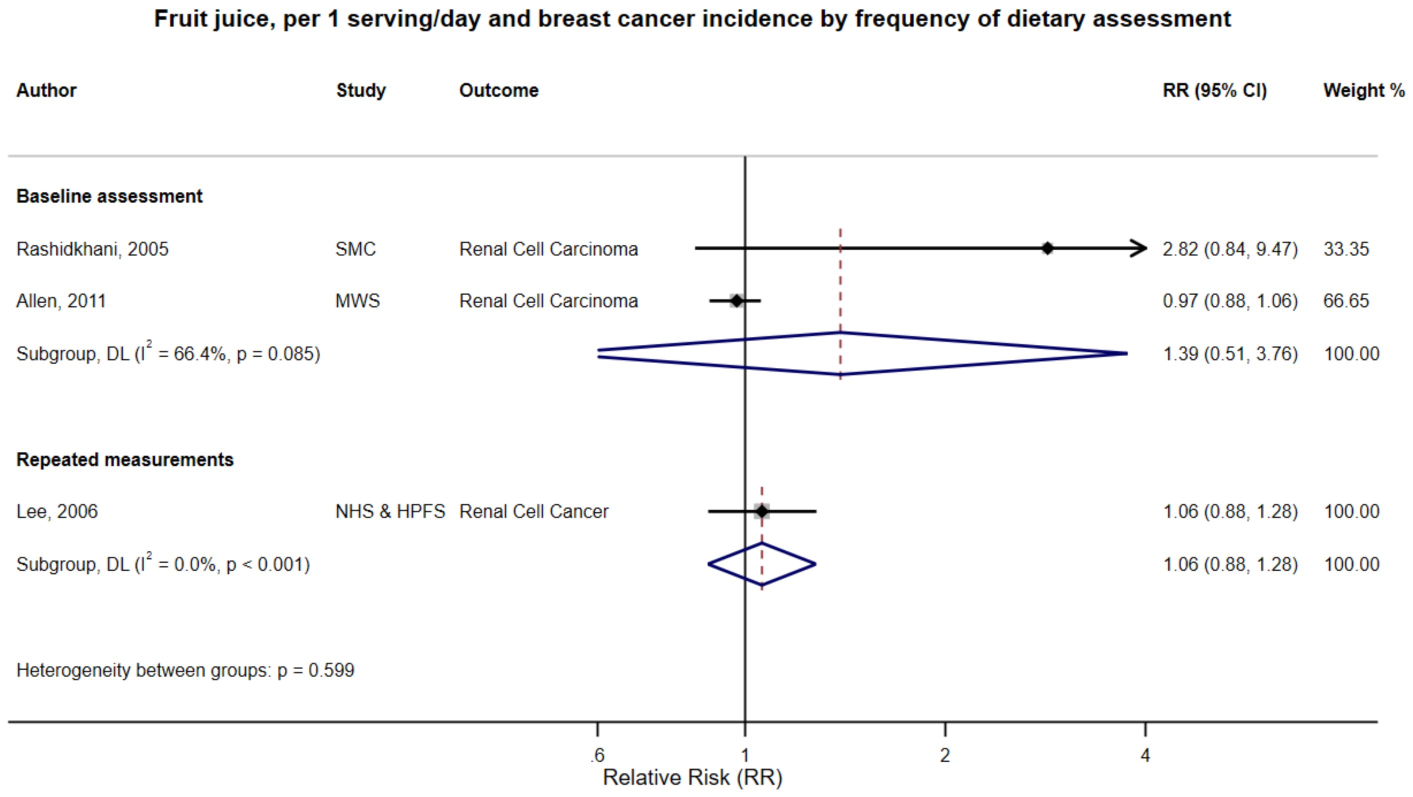

Figure legend: Forest plot shows the linear dose-response results (per 1 serving [177 mL] of juice/day) from the inverse variance DerSimonian-Laird random-effects model. The diamond represents the summary relative risk (RR) estimate, the diamond's width is the 95% confidence interval (CI). Each square and the horizontal line across the square represent the RR estimate of the individual study and its 95% CI.

Abbreviations: CI, confidence interval; RR, relative risk.

Study abbreviations: SMC, Swedish Mammography Cohort; MWS, Million Women Study; NHS & HPFS, Nurses' Health Study & Health Professionals Follow-up Study

Rashidkhani, 2005[142], Allen, 2011[36], Lee, 2006[98]

**Supplementary Figure 298. Linear dose-response meta-analysis of fruit juice and kidney cancer incidence by geographical region**

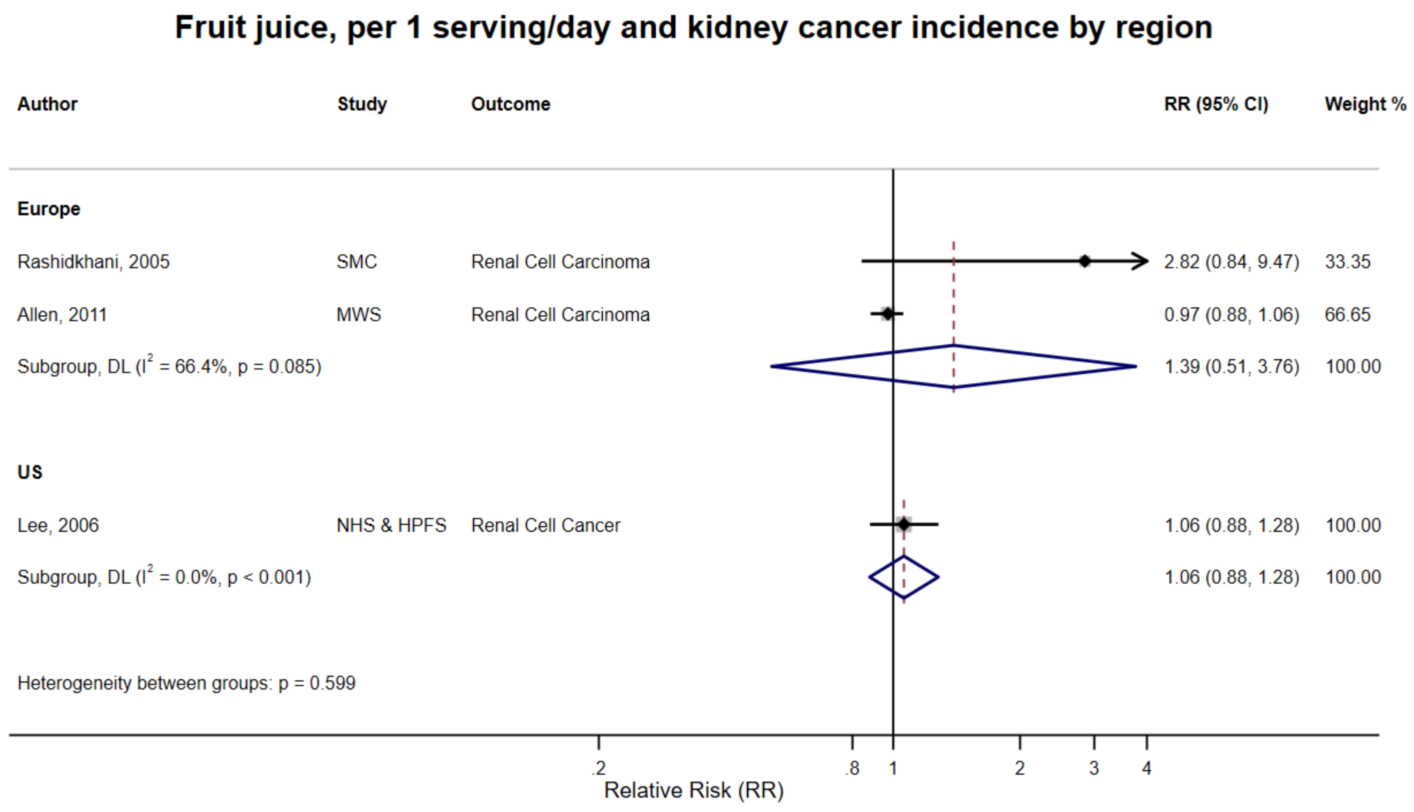

Figure legend: Forest plot shows the linear dose-response results (per 1 serving [177 mL] of juice/day) from the inverse variance DerSimonian-Laird random-effects model. The diamond represents the summary relative risk (RR) estimate, the diamond's width is the 95% confidence interval (CI). Each square and the horizontal line across the square represent the RR estimate of the individual study and its 95% CI.

Abbreviations: CI, confidence interval; RR, relative risk.

Study abbreviations: SMC, Swedish Mammography Cohort; MWS, Million Women Study; NHS & HPFS, Nurses' Health Study & Health Professionals Follow-up Study

Rashidkhani, 2005[142], Allen, 2011[36], Lee, 2006[98]

**Supplementary Figure 299. Linear dose-response meta-analysis of fruit and vegetable juice and kidney cancer incidence by sex.**

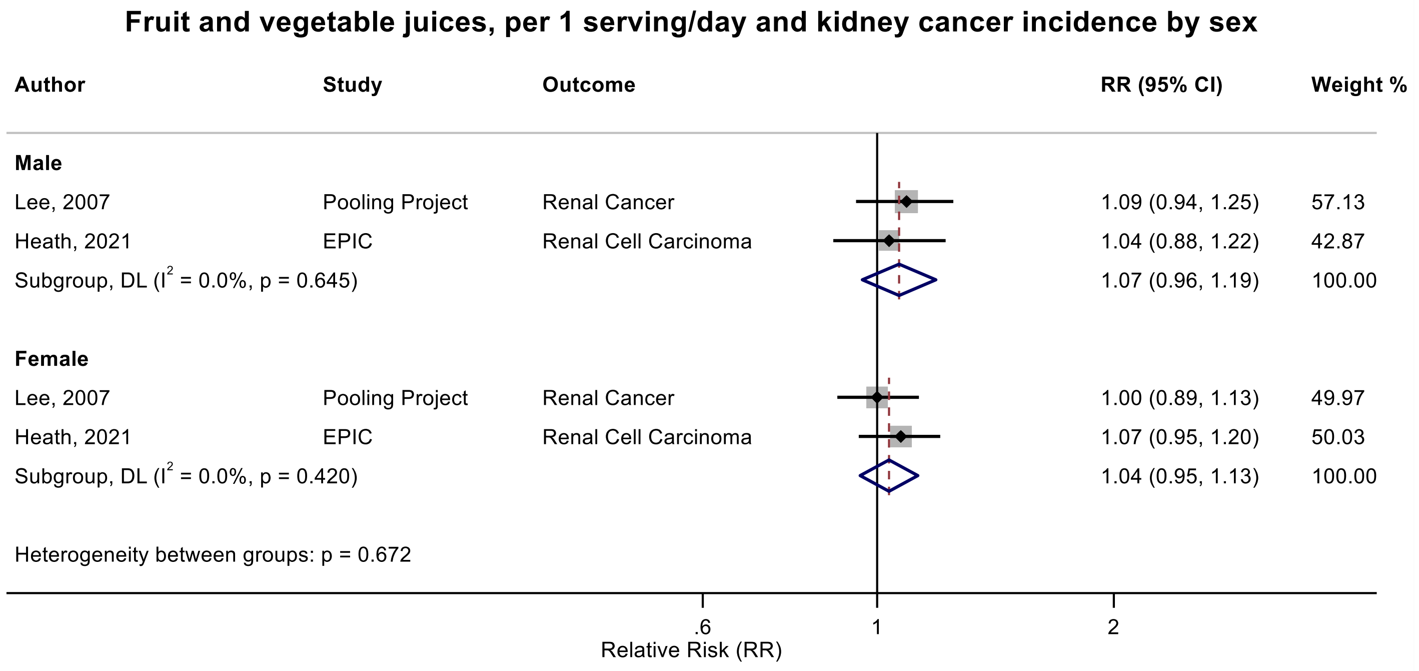

Figure legend: Forest plot shows the linear dose-response results (per 1 serving [177 mL] of juice/day) from the inverse variance DerSimonian-Laird random-effects model. The diamond represents the summary relative risk (RR) estimate, the diamond's width is the 95% confidence interval (CI). Each square and the horizontal line across the square represent the RR estimate of the individual study and its 95% CI.

Abbreviations: CI, confidence interval; RR, relative risk.

Study abbreviations: Pooling Project, The Pooling Project of Prospective Studies of Diet and Cancer (n=13); EPIC, European Prospective Investigation into Cancer and nutrition

Lee, 2007[26], Heath, 2021[66]

**Supplementary Figure 300. Linear dose-response meta-analysis of fruit juice and pancreatic cancer incidence, leave-one-out analysis.**

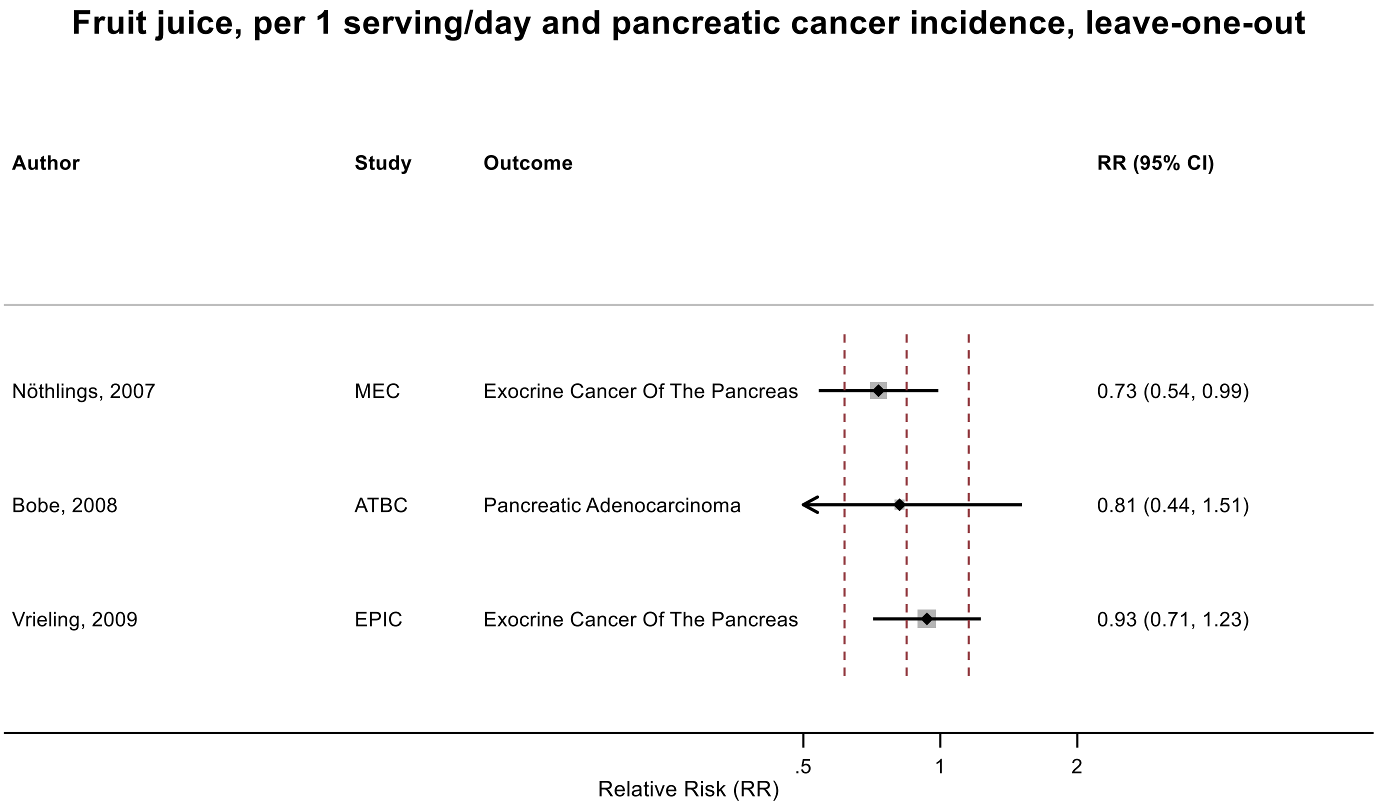

Figure legend: Forest plot shows the linear dose-response results (per 1 serving [177 mL] of fruit juice/day) from the inverse variance DerSimonian-Laird random-effects model after exclusion of every single study at a time.

Abbreviations: CI, confidence interval; RR, relative risk.

Study abbreviations: NIH-AARP, National Institute of Health + American Association of Retired Persons Diet and Health Study; EPIC, European Prospective Investigation into Cancer and nutrition; PLCO, Prostate, Lung, Colorectal and Ovarian Cancer Screening trial; CPS II, Cancer Prevention Study II; Taiwan MJ clinics, The Taiwan MJ Cohort Study

Nöthlings, 2007[131], Bobe, 2008[41], Vrieling, 2009[167]

**Supplementary Figure 301. Linear dose-response meta-analysis of fruit juice and breast cancer incidence, leave-one-out analysis.**

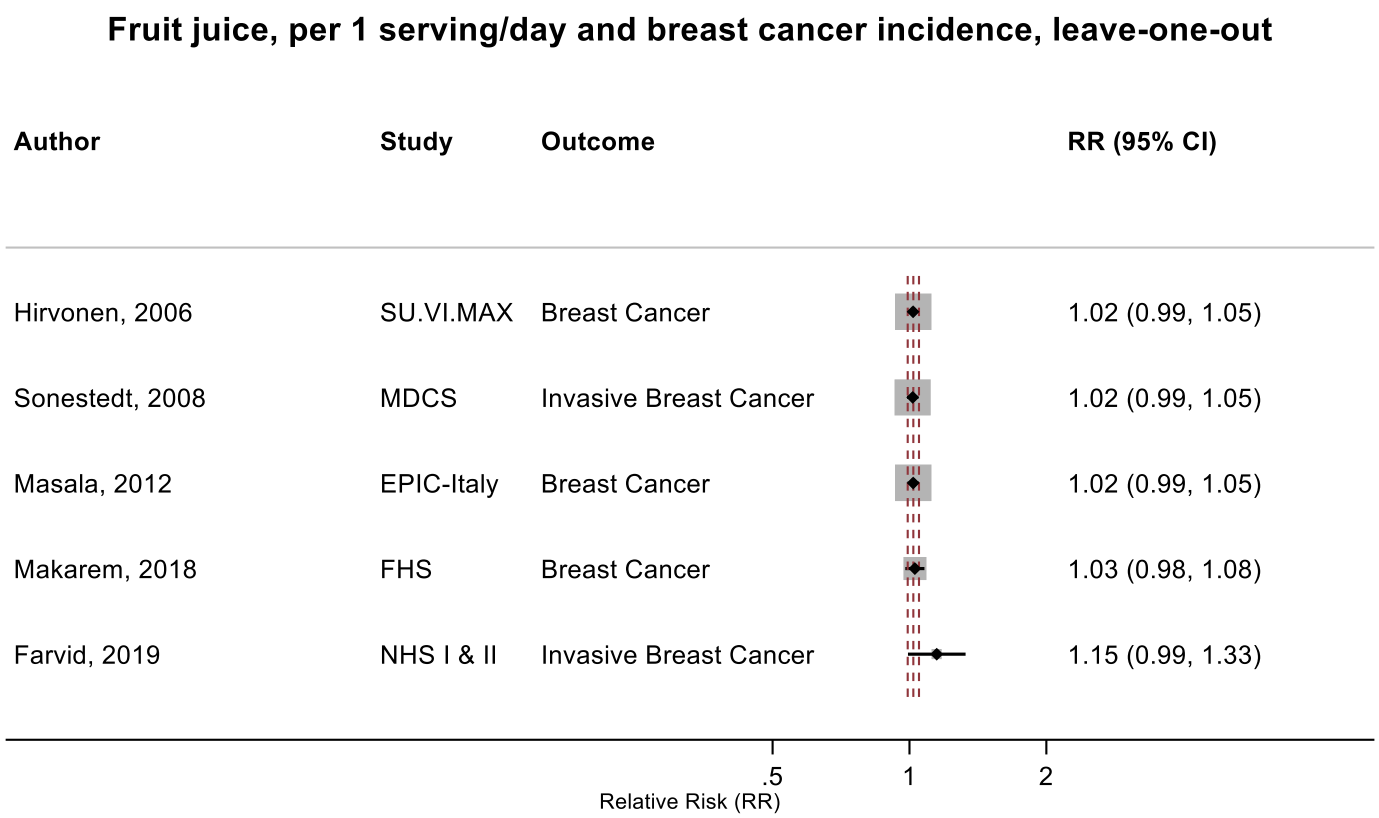

Figure legend: Forest plot shows the linear dose-response results (per 1 serving [177 mL] of fruit juice/day) from the inverse variance DerSimonian-Laird random-effects model after exclusion of every single study at a time.

Abbreviations: CI, confidence interval; RR, relative risk.

Study abbreviations: SU.VI.MAX, Supplémentation en Vitamines et Minéraux Antioxydants Study; MDCS, Malmö Diet and Cancer Study; EPIC-Italy, European Prospective Investigation into Cancer and nutrition-Italy cohort; FHS, Framingham Heart Study; NHS I & II, Nurses' Health Study I & II

Hirvonen, 2006[69], Sonestedt, 2008[156], Masala, 2012[112], Makarem, 2018[108], Farvid, 2019[52]

**Supplementary Figure 302. Linear dose-response meta-analysis of fruit juice and kidney cancer incidence, leave-one-out analysis.**

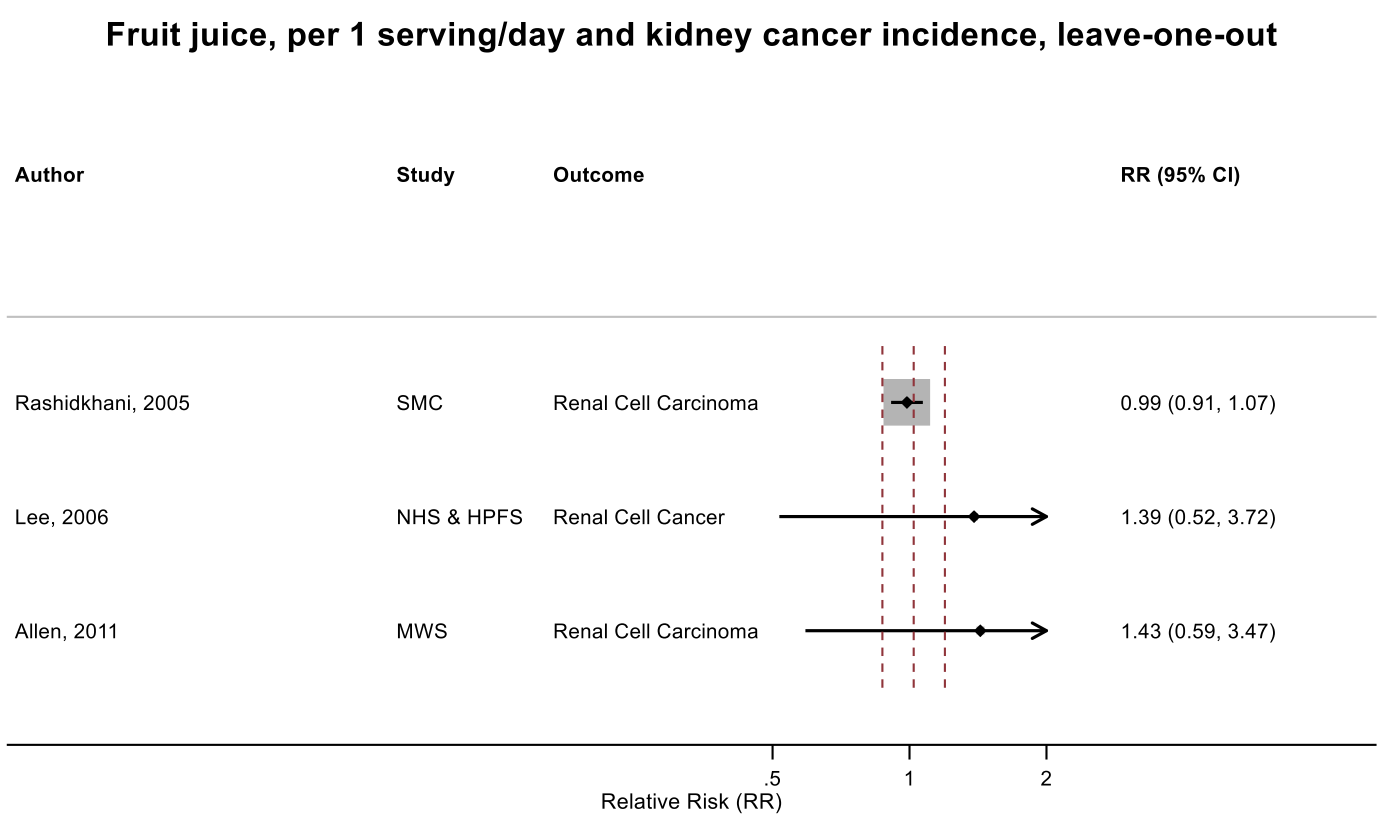

Figure legend: Forest plot shows the linear dose-response results (per 1 serving [177 mL] of fruit juice/day) from the inverse variance DerSimonian-Laird random-effects model after exclusion of every single study at a time.

Abbreviations: CI, confidence interval; RR, relative risk.

Study abbreviations: SMC, Swedish Mammography Cohort; NHS & HPFS, Nurses' Health Study & Health Professionals Follow-up Study; MWS, Million Women Study

Rashidkhani, 2005[142], Lee, 2006[98], Allen, 2011[36]

**Supplementary Figure 303. Linear dose-response meta-analysis of fruit and vegetable juice and kidney cancer incidence, leave-one-out analysis.**

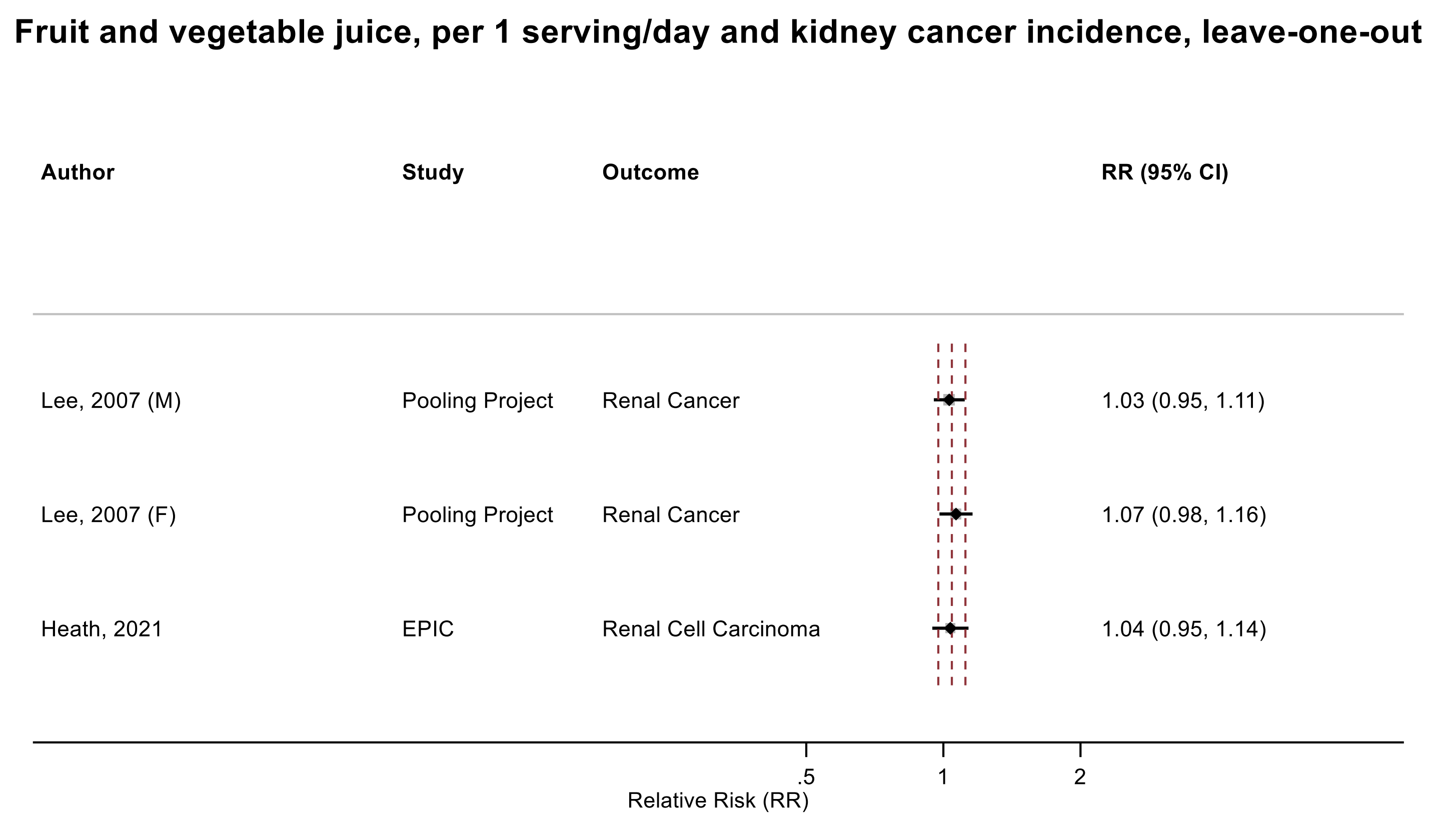

Figure legend: Forest plot shows the linear dose-response results (per 1 serving [177 mL] of fruit juice/day) from the inverse variance DerSimonian-Laird random-effects model after exclusion of every single study at a time.

Abbreviations: CI, confidence interval; RR, relative risk.

Study abbreviations: Pooling Project, The Pooling Project of Prospective Studies of Diet and Cancer (n=13); EPIC, European Prospective Investigation into Cancer and nutrition

Lee, 2007[26], Heath, 2021[66]

**Supplementary Figure 304. Linear dose-response meta-analysis of citrus juice and melanoma incidence, leave-one-out analysis.**

#### Citrus juice, per 1 serving/day and melanoma incidence, leave-one-out

| Author | Study | Outcome | RR (95% CI) |
|---|---|---|---|
| Melough, 2020 | WHI | Melanoma | 1.10 (0.82, 1.48) |
| Mahamat-Saleh, 2020 | EPIC | Melanoma | 1.14 (1.01, 1.30) |
| Melough, 2021 | NIH-AARP | Cutaneous Malignant Melanoma | 1.05 (0.92, 1.21) |
| Marley, 2021 | UK Biobank | Melanoma | 1.08 (0.81, 1.44) |

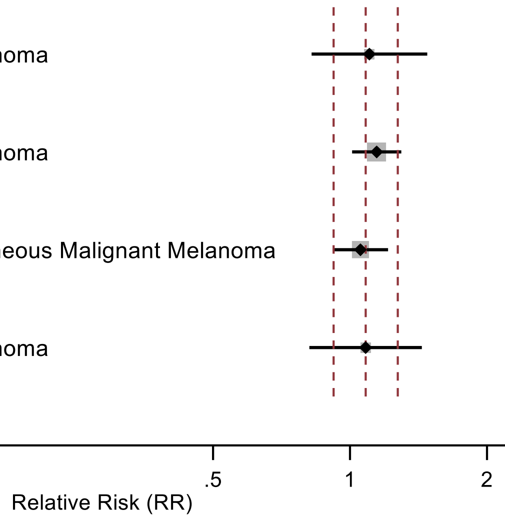

Figure legend: Forest plot shows the linear dose-response results (per 1 serving [177 mL] of fruit juice/day) from the inverse variance DerSimonian-Laird random-effects model after exclusion of every single study at a time.

Abbreviations: CI, confidence interval; RR, relative risk.

Study abbreviations: WHI, Women’s Health Initiative Study; EPIC, European Prospective Investigation into Cancer and nutrition; NIH-AARP, National Institute of Health + American Association of Retired Persons Diet and Health Study; UK Biobank, United Kingdom Biobank Study

Melough, 2020[116], Mahamat-Saleh, 2020[107], Melough, 2021[115], Marley, 2021[110]

**Supplementary Figure 305. Linear dose-response meta-analysis of orange juice and melanoma incidence, leave-one-out analysis.**

#### Orange juice, per 1 serving/day and melanoma incidence, leave-one-out

| Author | Study | Outcome | RR (95% CI) |
|---|---|---|---|
| Feskanich,, 2003 | NHS I & II | Melanoma | 1.19 (1.06, 1.33) |
| Wu, 2015 | HPFS | Melanoma | 1.18 (1.05, 1.33) |
| Marley, 2021 | UK Biobank | Melanoma | 1.33 (1.09, 1.61) |

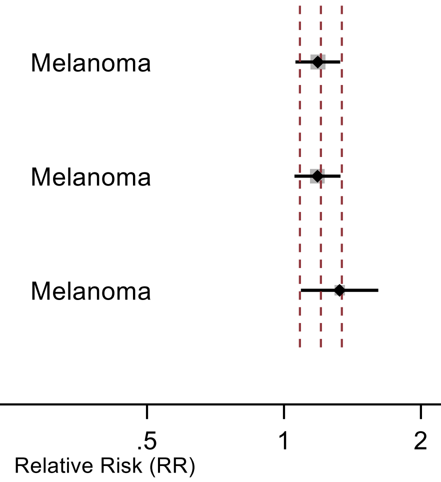

Figure legend: Forest plot shows the linear dose-response results (per 1 serving [177 mL] of fruit juice/day) from the inverse variance DerSimonian-Laird random-effects model after exclusion of every single study at a time.

Abbreviations: CI, confidence interval; RR, relative risk.

Study abbreviations: NHS I & II, Nurses' Health Study I & II; HPFS, Health Professionals Follow-up Study; UK Biobank, United Kingdom Biobank Study

Feskanich, 2003[54], Wu, 2015[171], Marley, 2021[110]

**Supplementary Figure 306. Linear dose-response meta-analysis of grapefruit juice and melanoma incidence, leave-one-out analysis.**

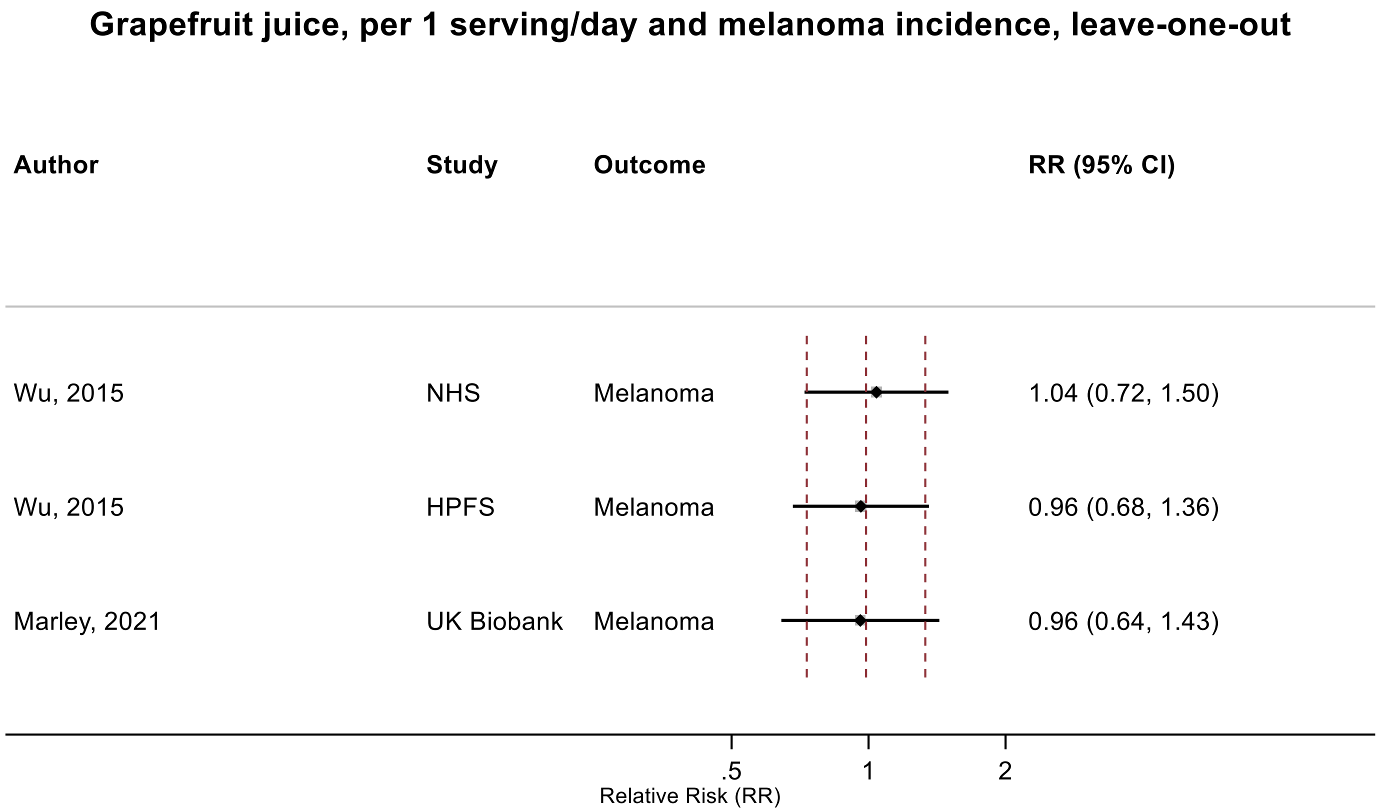

Figure legend: Forest plot shows the linear dose-response results (per 1 serving [177 mL] of fruit juice/day) from the inverse variance DerSimonian-Laird random-effects model after exclusion of every single study at a time.

Abbreviations: CI, confidence interval; RR, relative risk.

Study abbreviations: NHS, Nurses' Health Study; HPFS, Health Professionals Follow-up Study; UK Biobank, United Kingdom Biobank Study

Wu, 2015[171], Marley, 2021[110]

**Supplementary Figure 307. Linear dose-response meta-analysis of fruit juice and glioma incidence, leave-one-out analysis.**

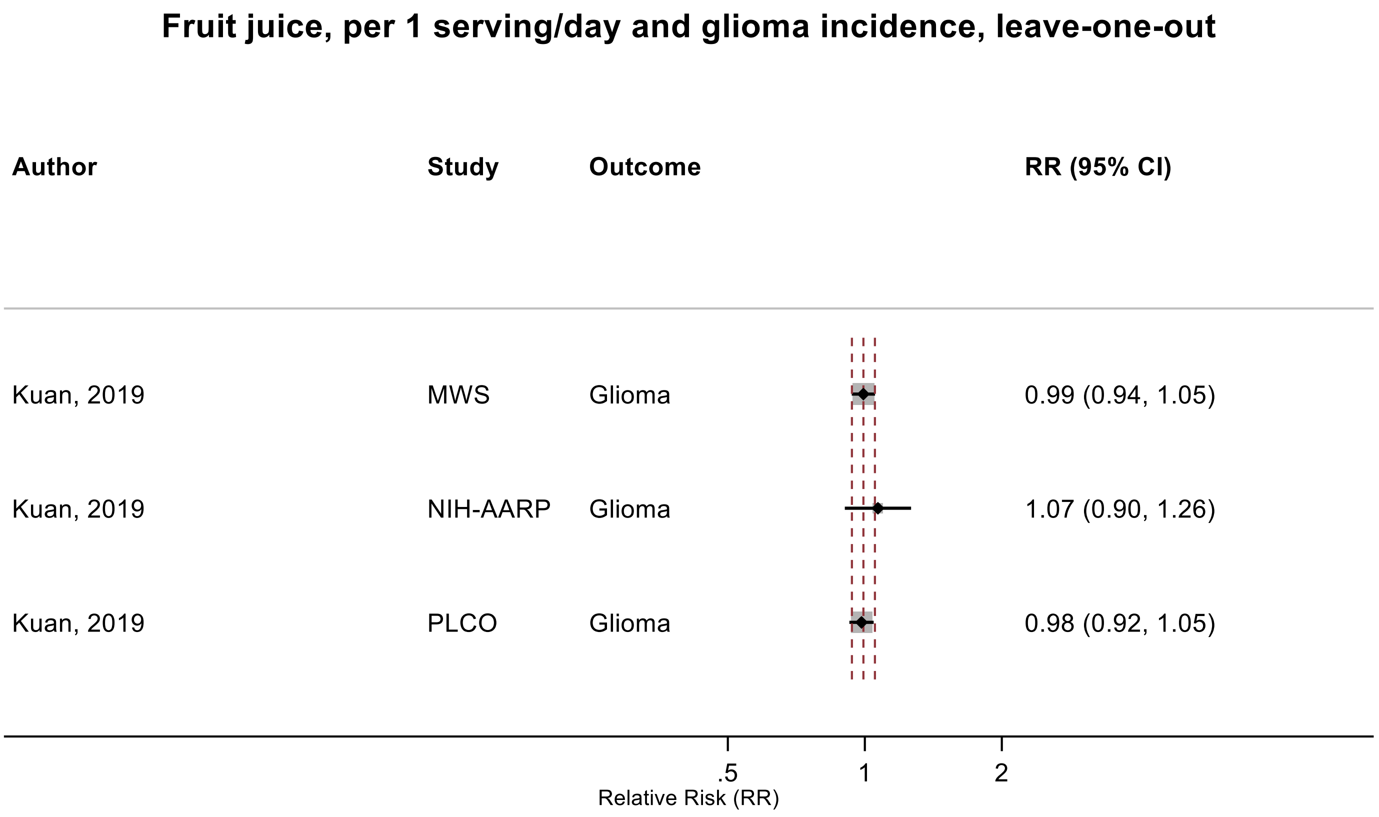

Figure legend: Forest plot shows the linear dose-response results (per 1 serving [177 mL] of fruit juice/day) from the inverse variance DerSimonian-Laird random-effects model after exclusion of every single study at a time.

Abbreviations: CI, confidence interval; RR, relative risk.

Study abbreviations: MWS, Million Women Study; NIH-AARP, National Institute of Health + American Association of Retired Persons Diet and Health Study; PLCO, Prostate, Lung, Colorectal and Ovarian Cancer Screening trial

Kuan, 2019[92]

**Supplementary Figure 308. Linear dose-response meta-analysis of juices and prostate cancer mortality.**

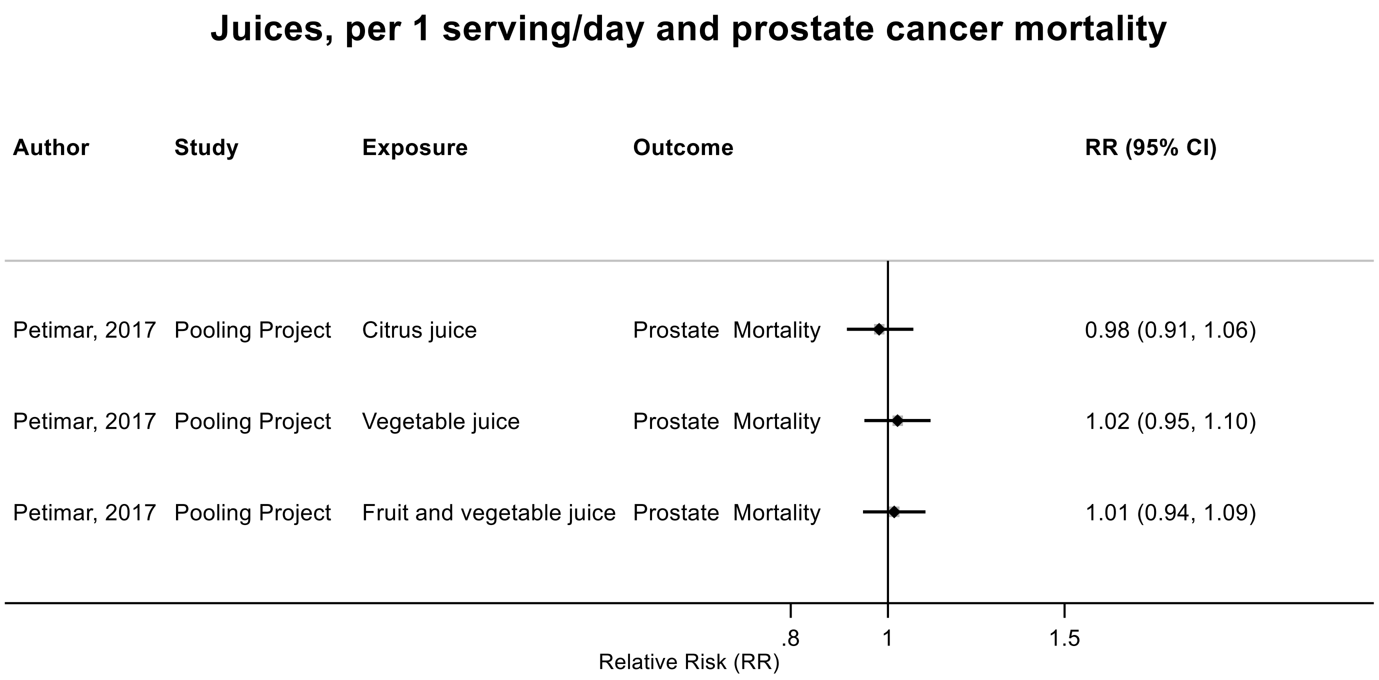

Figure legend: Forest plot shows the linear dose-response results (per 1 serving [177 mL] of juice/day) from the inverse variance DerSimonian-Laird random-effects model. Each square and the horizontal line across the square represent the RR estimate of the individual study and its 95% CI.

Abbreviations: CI, confidence interval; RR, relative risk.

Study abbreviations: Pooling Project, The Pooling Project of Prospective Studies of Diet and Cancer (n=15)

Petimar, 2017[139]

**Supplementary Figure 309. Linear dose-response meta-analysis of soft drinks and liver cancer incidence.**

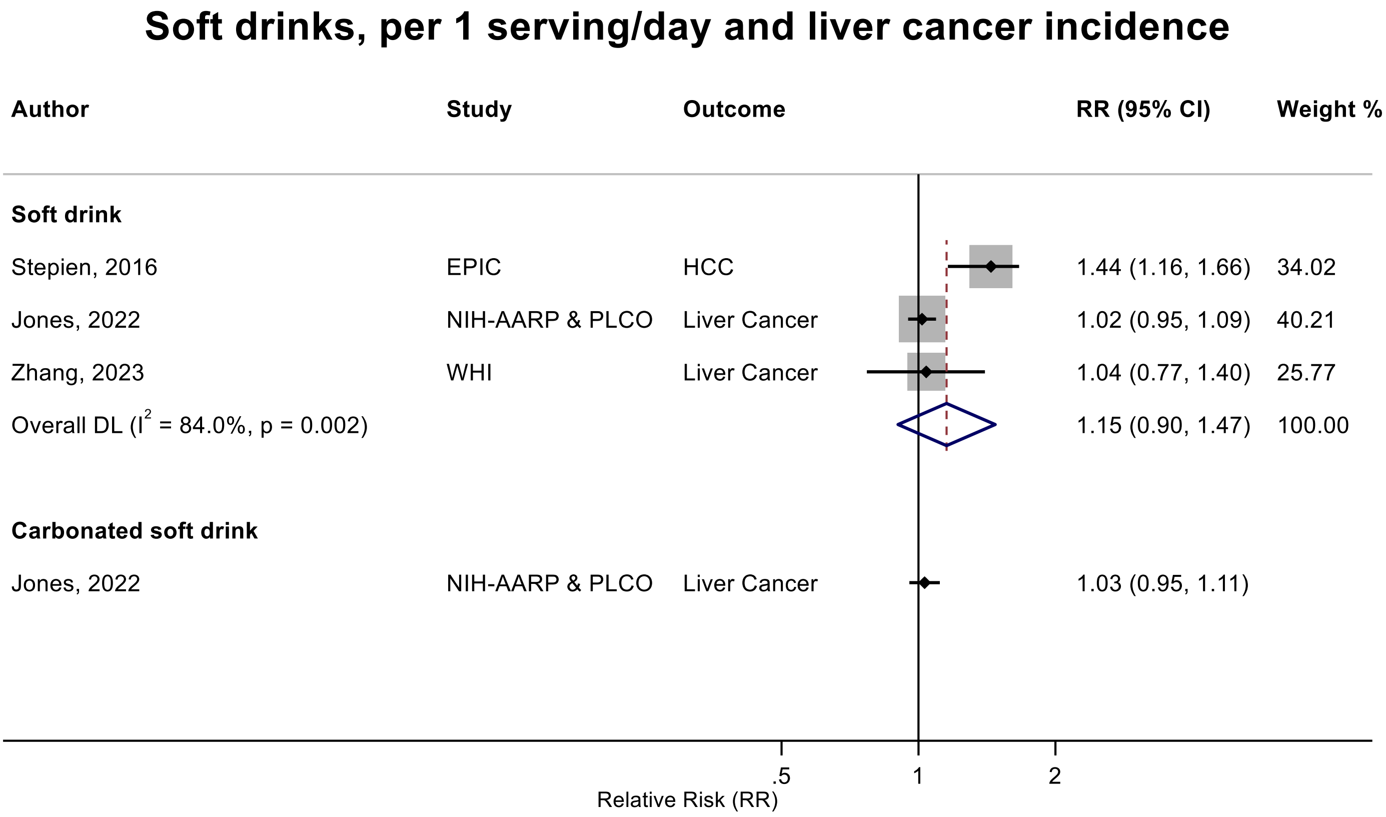

Figure legend: Forest plot shows the linear dose-response results (per 1 serving [355 mL] of soft drinks/day) from the inverse variance DerSimonian-Laird random-effects model. The diamond represents the summary relative risk (RR) estimate, the diamond's width is the 95% confidence interval (CI). Each square and the horizontal line across the square represent the RR estimate of the individual study and its 95% CI.

Abbreviations: CI, confidence interval; HCC, hepatocellular carcinoma; RR, relative risk.

Study abbreviations: EPIC, European Prospective Investigation into Cancer and Nutrition; NIH-AARP, National Institute of Health + American Association of Retired Persons Diet and Health Study; PLCO, Prostate, Lung, Colorectal and Ovarian Cancer Screening Trial; WHI, Women's Health Initiative

Stepien, 2016[159], Zhang, 2023[183], Jones, 2022[79]

**Supplementary Figure 310. Linear dose-response meta-analysis of soft drinks and pancreatic cancer incidence.**

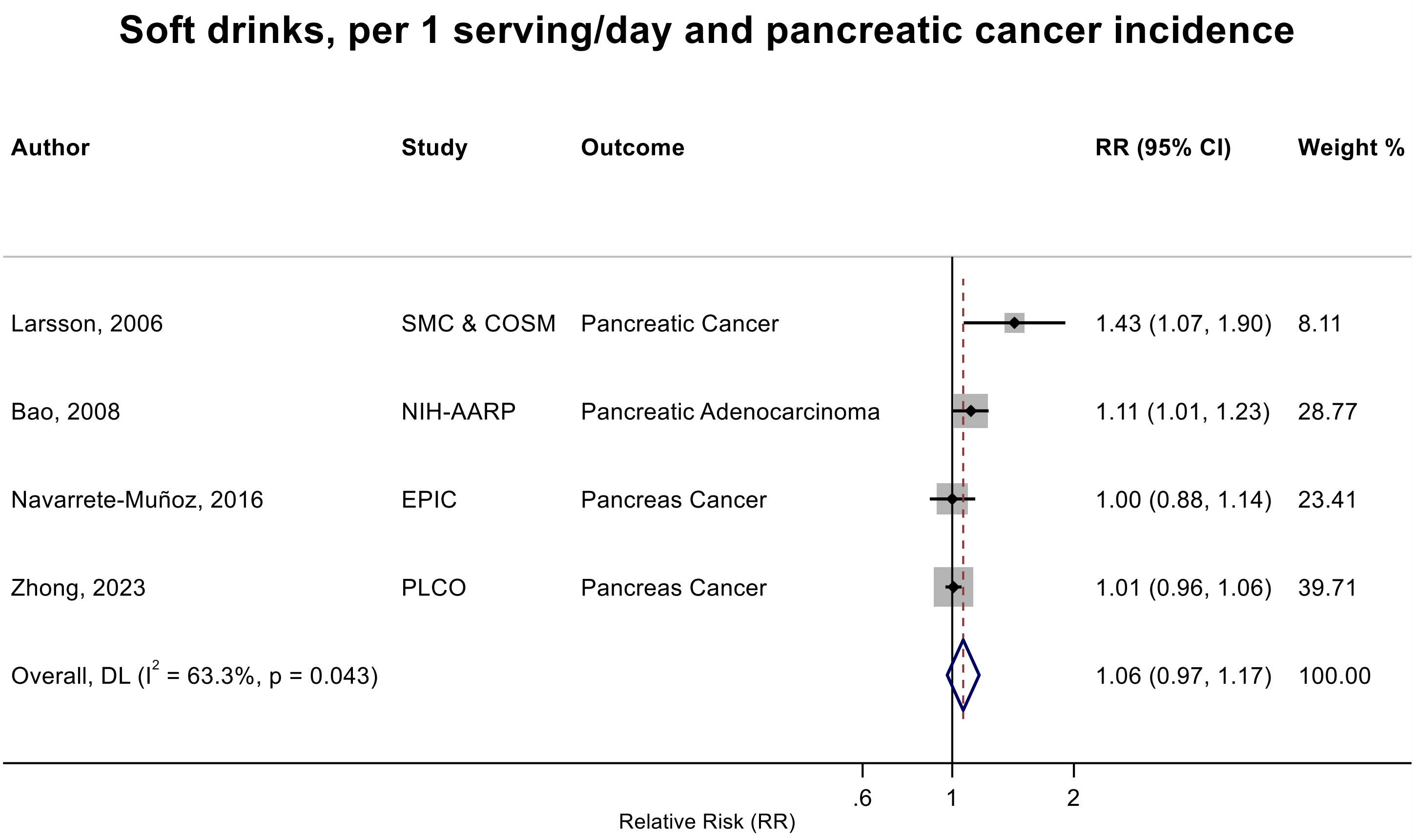

Figure legend: Forest plot shows the linear dose-response results (per 1 serving [355 mL] of soft drinks/day) from the inverse variance DerSimonian-Laird random-effects model. The diamond represents the summary relative risk (RR) estimate, the diamond's width is the 95% confidence interval (CI). Each square and the horizontal line across the square represent the RR estimate of the individual study and its 95% CI.

Abbreviations: CI, confidence interval; RR, relative risk.

Study abbreviations: SMC & COSM, Swedish Mammography Cohort and Cohort of Swedish Men; NIH-AARP, National Institute of Health + American Association of Retired Persons Diet and Health Study; EPIC, European Prospective Investigation into Cancer and Nutrition; PLCO, Prostate, Lung, Colorectal and Ovarian Cancer Screening Trial

Note: There was an additional study (Hokkaido cohort study in Japan)[85] that could not be included in the dose-response meta-analysis because of insufficient data format and reported little evidence of an association (RR: 0.20, 95%CI: 0.01, 1.80) for 7 times/week vs never-several times/months of soft drinks consumption and pancreatic cancer mortality in men and women. There was another publication from PLCO[184] that assessed the association between soft drinks, as part the of WCRF/AICR cancer prevention recommendation score, and pancreatic cancer incidence and mortality and reported the relative risks of 0.69 (0.45-1.05) and 0.70 (0.45-1.08) for 0 vs >250 g/day of SSB intake and pancreatic cancer incidence and mortality, respectively. Due to higher number of cases, we included another publication on incidence in the linear dose-response and described this publication on incidence and mortality descriptively.

Navarrete-Muñoz, 2016[127], Bao, 2008[39], Zhong, 2023[189], Larsson, 2006[95]

**Supplementary Figure 311. Linear dose-response meta-analysis of soft drinks and biliary tract and gallbladder cancers incidence.**

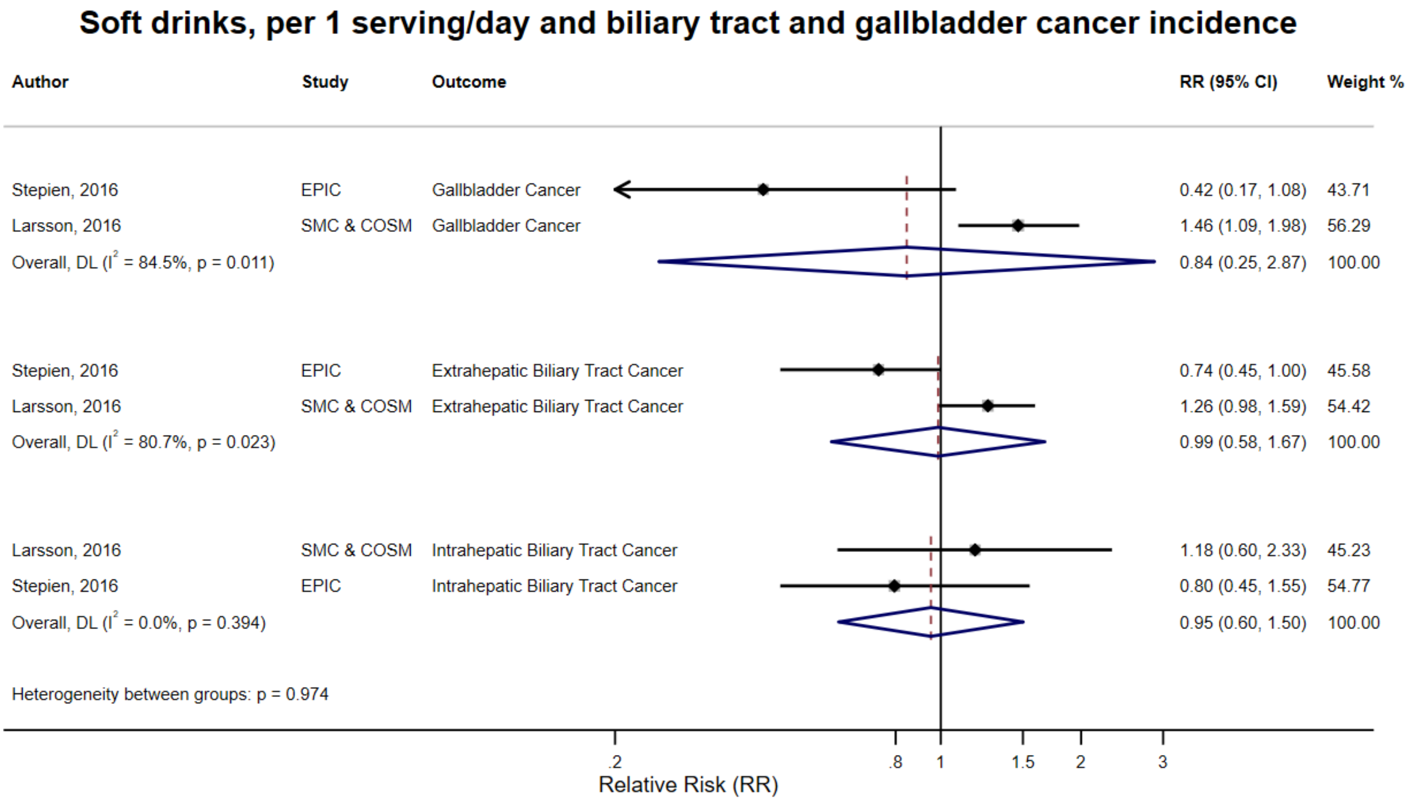

Figure legend: Forest plot shows the linear dose-response results (per 1 serving [355 mL] of soft drinks/day) from the inverse variance DerSimonian-Laird random-effects model. The diamond represents the summary relative risk (RR) estimate, the diamond's width is the 95% confidence interval (CI). Each square and the horizontal line across the square represent the RR estimate of the individual study and its 95% CI.

Abbreviations: CI, confidence interval; RR, relative risk.

Study abbreviations: EPIC, European Prospective Investigation into Cancer and Nutrition; SMC & COSM, Swedish Mammography Cohort and Cohort of Swedish Men.

Stepien, 2016[159], Larsson, 2016[96]

**Supplementary Figure 312. Linear dose-response meta-analysis of soft drinks and lung cancer incidence.**

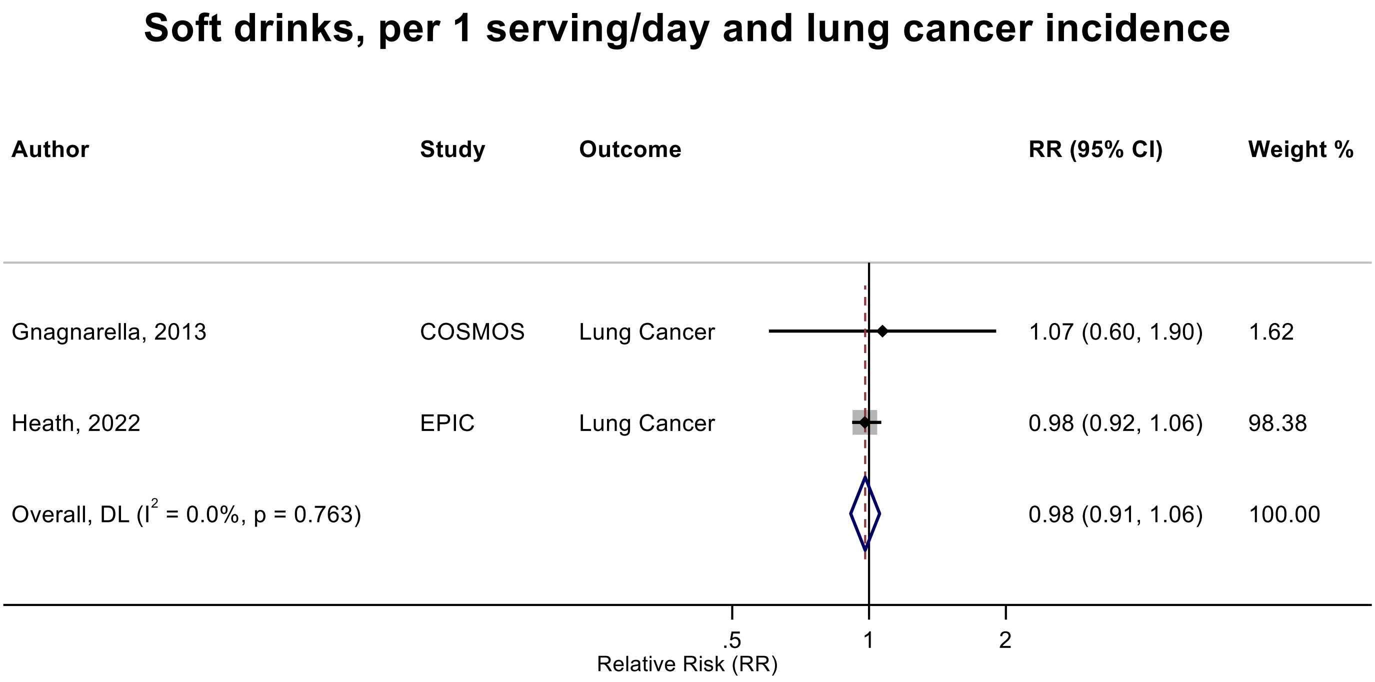

Figure legend: Forest plot shows the linear dose-response results (per 1 serving [355 mL] of soft drinks/day) from the inverse variance DerSimonian-Laird random-effects model. The diamond represents the summary relative risk (RR) estimate, the diamond's width is the 95% confidence interval (CI). Each square and the horizontal line across the square represent the RR estimate of the individual study and its 95% CI.

Abbreviations: CI, confidence interval; RR, relative risk.

Study abbreviations: COSMOS, Continuous Observation of Smoking Subjects; EPIC, European Prospective Investigation into Cancer and Nutrition.

Note: There was an additional study (PLCO)[177] that could not be included in the dose-response meta-analysis because of insufficient data format and reported an inverse association between soft drinks and lung cancer incidence ($RR_{\text{intake vs no intake}}$: 0.79 (95%CI: 0.64-0.97). Sex-specific analysis indicated an inverse association in females ($RR_{\text{intake vs no intake}}$: 0.62 (95%CI: 0.45-0.87), but not in males ($RR_{\text{intake vs no intake}}$: 0.96 (95%CI: 0.72-1.28)

Gnagnarella, 2013[65], Heath, 2022[67]

**Supplementary Figure 313. Linear dose-response meta-analysis of soft drinks and breast cancer incidence.**

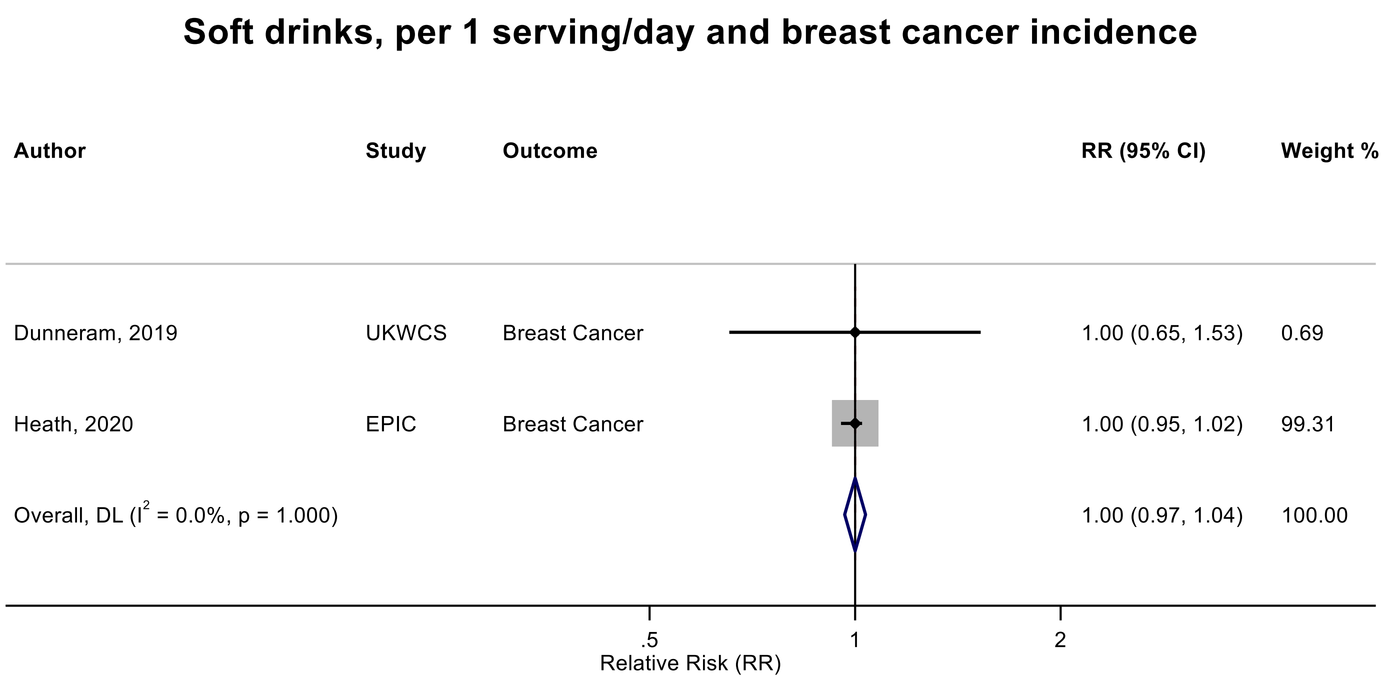

Figure legend: Forest plot shows the linear dose-response results (per 1 serving [355 mL] of soft drinks/day) from the inverse variance DerSimonian-Laird random-effects model. The diamond represents the summary relative risk (RR) estimate, the diamond's width is the 95% confidence interval (CI). Each square and the horizontal line across the square represent the RR estimate of the individual study and its 95% CI.

Abbreviations: CI, confidence interval; RR, relative risk.

Study abbreviations: UKWCS, UK Women Health Cohort Study; EPIC, European Prospective Investigation into Cancer and Nutrition

Dunneram, 2019[49], Heath, 2020[68]

**Supplementary Figure 314. Linear dose-response meta-analysis of soft drinks and endometrial cancer incidence.**

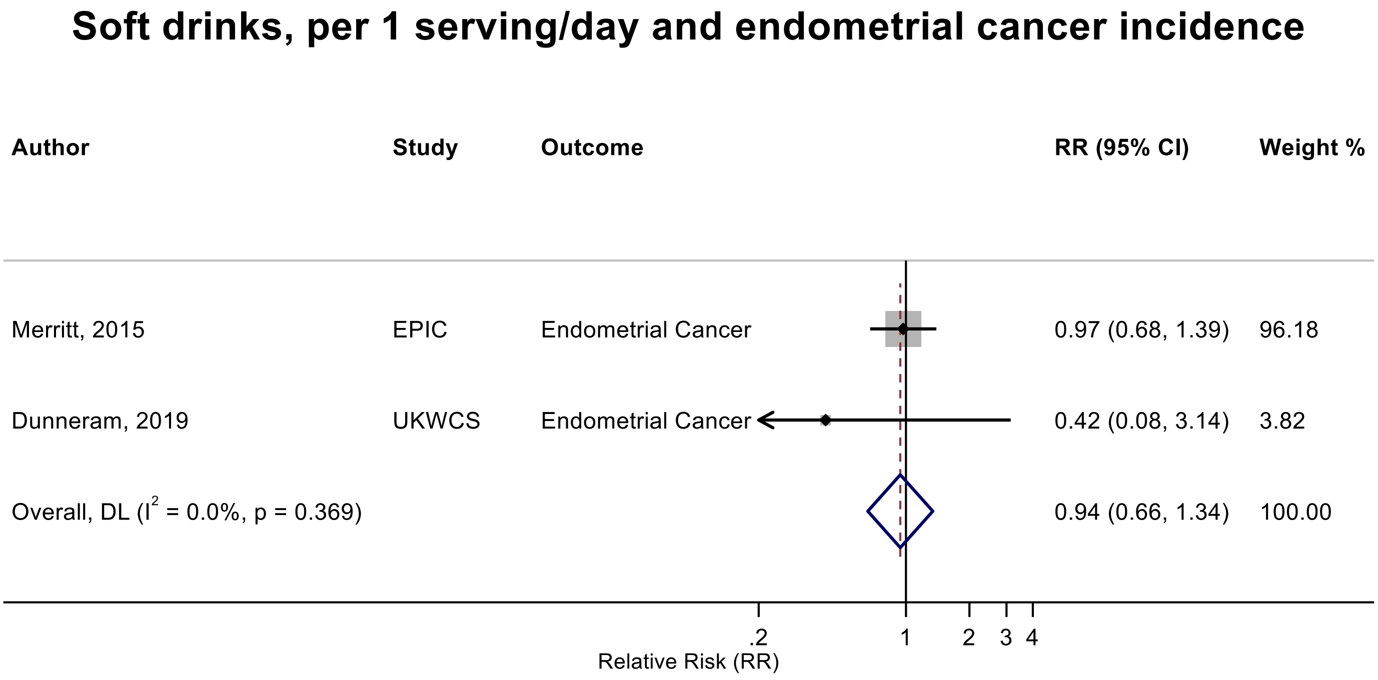

Figure legend: Forest plot shows the linear dose-response results (per 1 serving [355 mL] of soft drinks/day) from the inverse variance DerSimonian-Laird random-effects model. The diamond represents the summary relative risk (RR) estimate, the diamond's width is the 95% confidence interval (CI). Each square and the horizontal line across the square represent the RR estimate of the individual study and its 95% CI.

Abbreviations: CI, confidence interval; RR, relative risk.

Study abbreviations: EPIC, European Prospective Investigation into Cancer and Nutrition; UKWCS, UK Women's Cohort Study

Note: There was an additional study (Swedish Mammography Cohort)[61] that could not be included in the dose-response meta-analysis because of insufficient data format and reported little evidence of an association (RR: 1.02, 95%CI: 0.87-1.18) for 0.07 serving/week vs 0 of intake of soft drinks and endometrial cancer incidence.

Dunneram, 2019[49], Merritt, 2015[117]

**Supplementary Figure 315. Linear dose-response meta-analysis of carbonated soft drinks and kidney cancer incidence.**

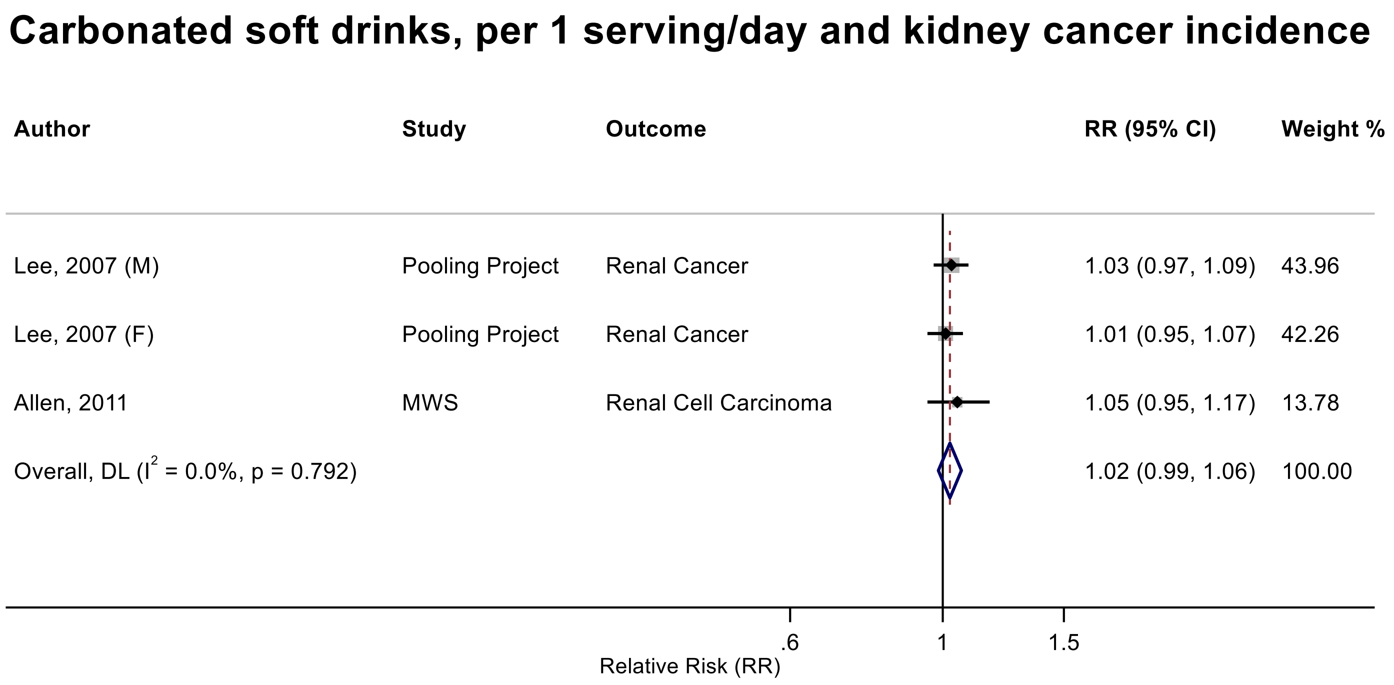

Figure legend: Forest plot shows the linear dose-response results (per 1 serving [355 mL] of carbonated soft drinks/day) from the inverse variance DerSimonian-Laird random-effects model. The diamond represents the summary relative risk (RR) estimate, the diamond's width is the 95% confidence interval (CI). Each square and the horizontal line across the square represent the RR estimate of the individual study and its 95% CI.

Abbreviations: CI, confidence interval; F, female, M, male, RR, relative risk.

Study abbreviations: Pooling Project, The Pooling Project of Prospective Studies of Diet and Cancer (n=13); MWS, Million Women Study

Allen, 2011[36], Lee, 2007[26]

**Supplementary Figure 316. Linear dose-response meta-analysis of soft drinks and bladder cancer incidence.**

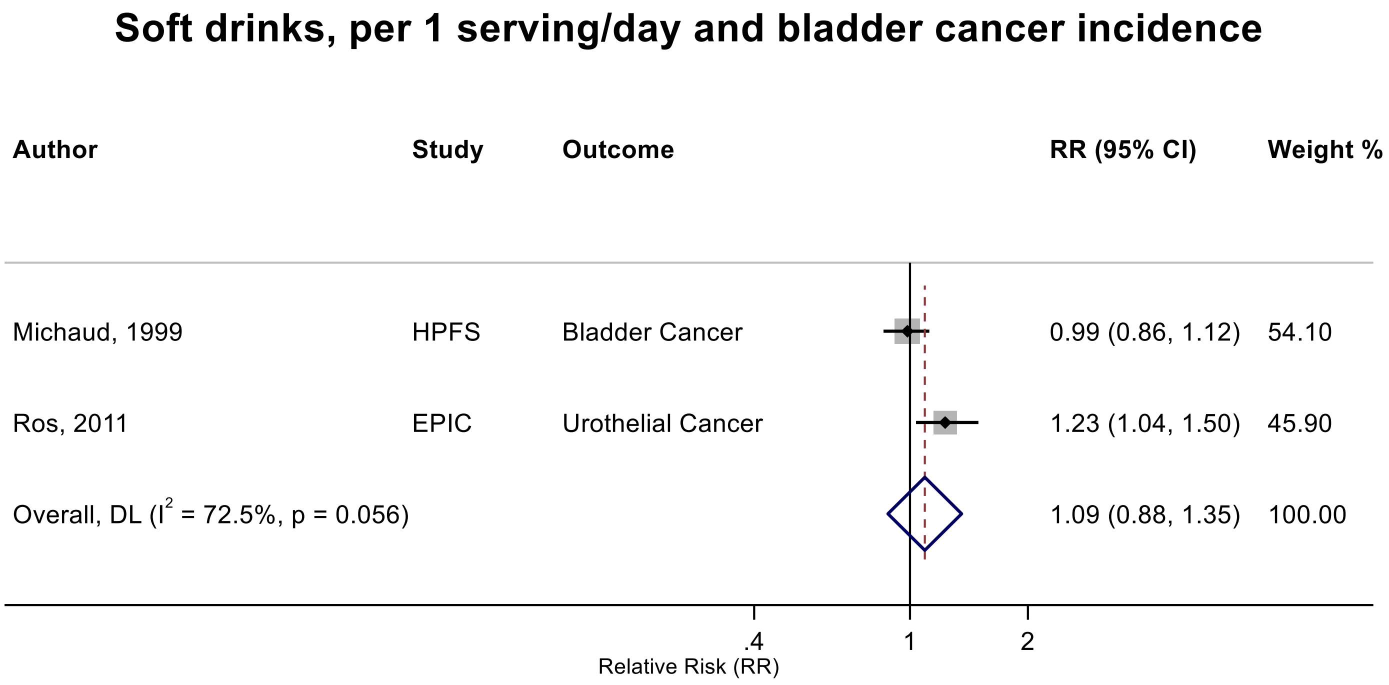

Figure legend: Forest plot shows the linear dose-response results (per 1 serving [355 mL] of soft drinks/day) from the inverse variance DerSimonian-Laird random-effects model. The diamond represents the summary relative risk (RR) estimate, the diamond's width is the 95% confidence interval (CI). Each square and the horizontal line across the square represent the RR estimate of the individual study and its 95% CI.

Abbreviations: CI, confidence interval; RR, relative risk.

Study abbreviations: HPFS, Health Professionals Follow-up Study; EPIC, European Prospective Investigation into Cancer and Nutrition

Michaud, 1999[119], Ros, 2011[147]

**Supplementary Figure 317. Linear dose-response meta-analysis of carbonated and caffeinated soft drinks and glioma incidence.**

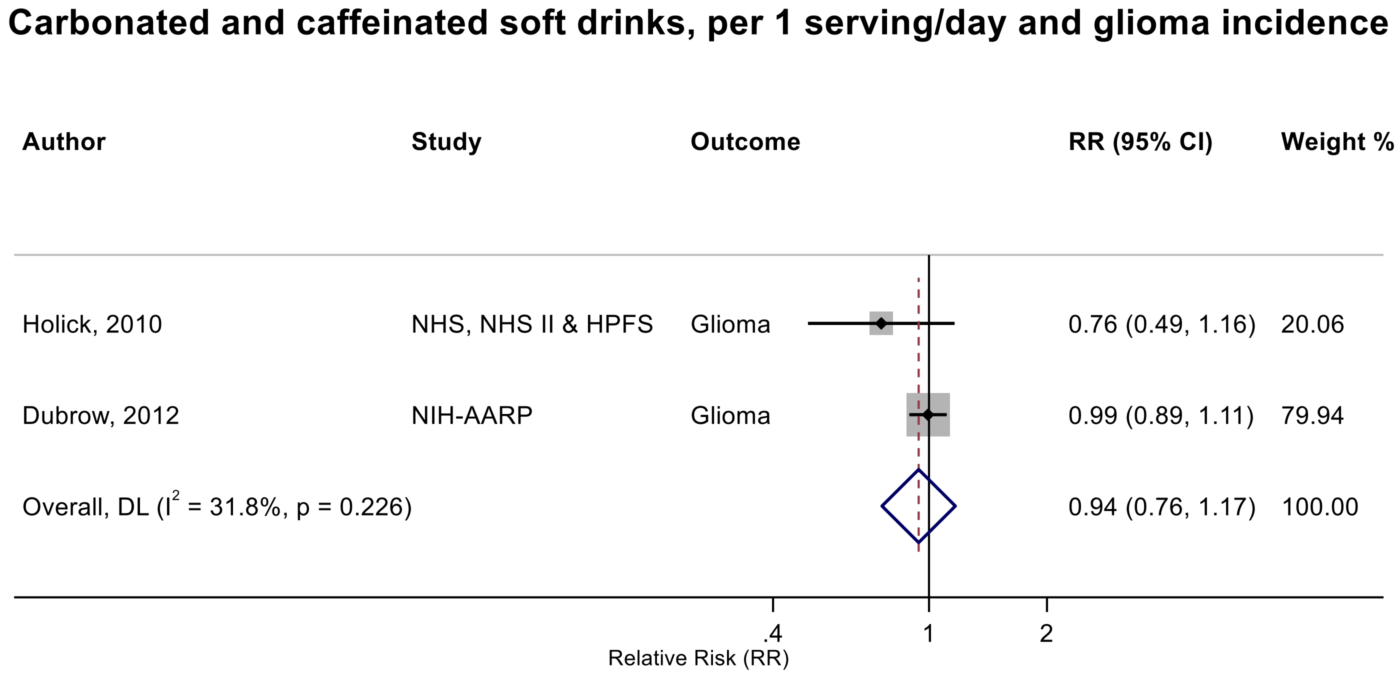

Figure legend: Forest plot shows the linear dose-response results (per 1 serving [355 mL] of carbonated and caffeinated soft drinks/day) from the inverse variance DerSimonian-Laird random-effects model. The diamond represents the summary relative risk (RR) estimate, the diamond's width is the 95% confidence interval (CI). Each square and the horizontal line across the square represent the RR estimate of the individual study and its 95% CI.

Abbreviations: CI, confidence interval; RR, relative risk.

Study abbreviations: NHS, Nurse's Health Study; NHS II, Nurse's Health Study II; HPFS, Health Professionals Follow-up Study; NIH-AARP, National Institutes Of Health-AARP Diet And Health Study

Holick, 2010[71], Dubrow, 2012[48]

**Supplementary Figure 318. Linear dose-response meta-analysis of carbonated and decaffeinated soft drinks and glioma incidence.**

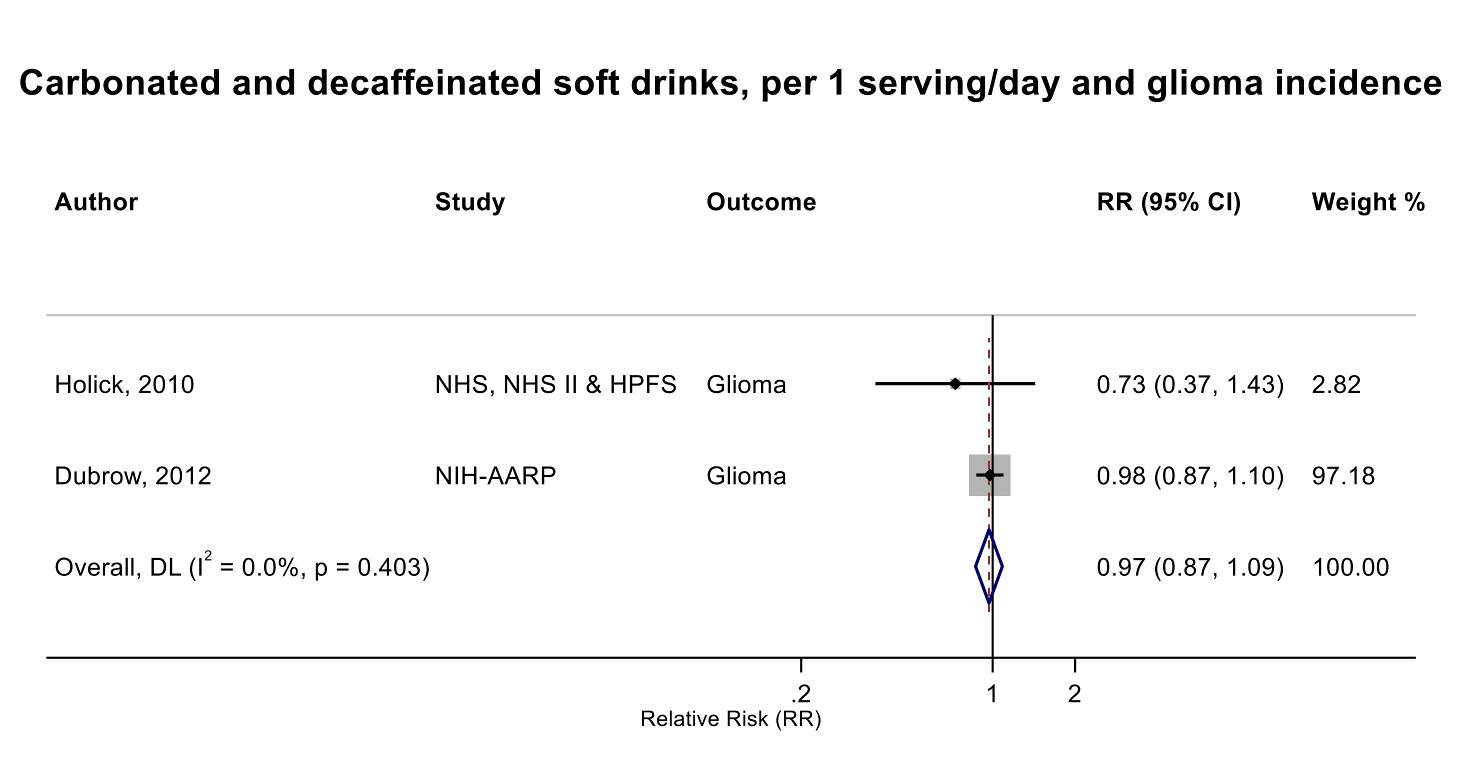

Figure legend: Forest plot shows the linear dose-response results (per 1 serving [355 mL] of carbonated and decaffeinated soft drinks/day) from the inverse variance DerSimonian-Laird random-effects model. The diamond represents the summary relative risk (RR) estimate, the diamond's width is the 95% confidence interval (CI). Each square and the horizontal line across the square represent the RR estimate of the individual study and its 95% CI.

Abbreviations: CI, confidence interval; RR, relative risk.

Study abbreviations: NHS, Nurse's Health Study; NHS II, Nurse's Health Study II; HPFS, Health Professionals Follow-up Study; NIH-AARP, National Institutes Of Health-AARP Diet And Health

Holick, 2010[71], Dubrow, 2012[48]

**Supplementary Figure 319. Linear dose-response meta-analysis of soft drinks and pancreatic cancer incidence by geographical region.**

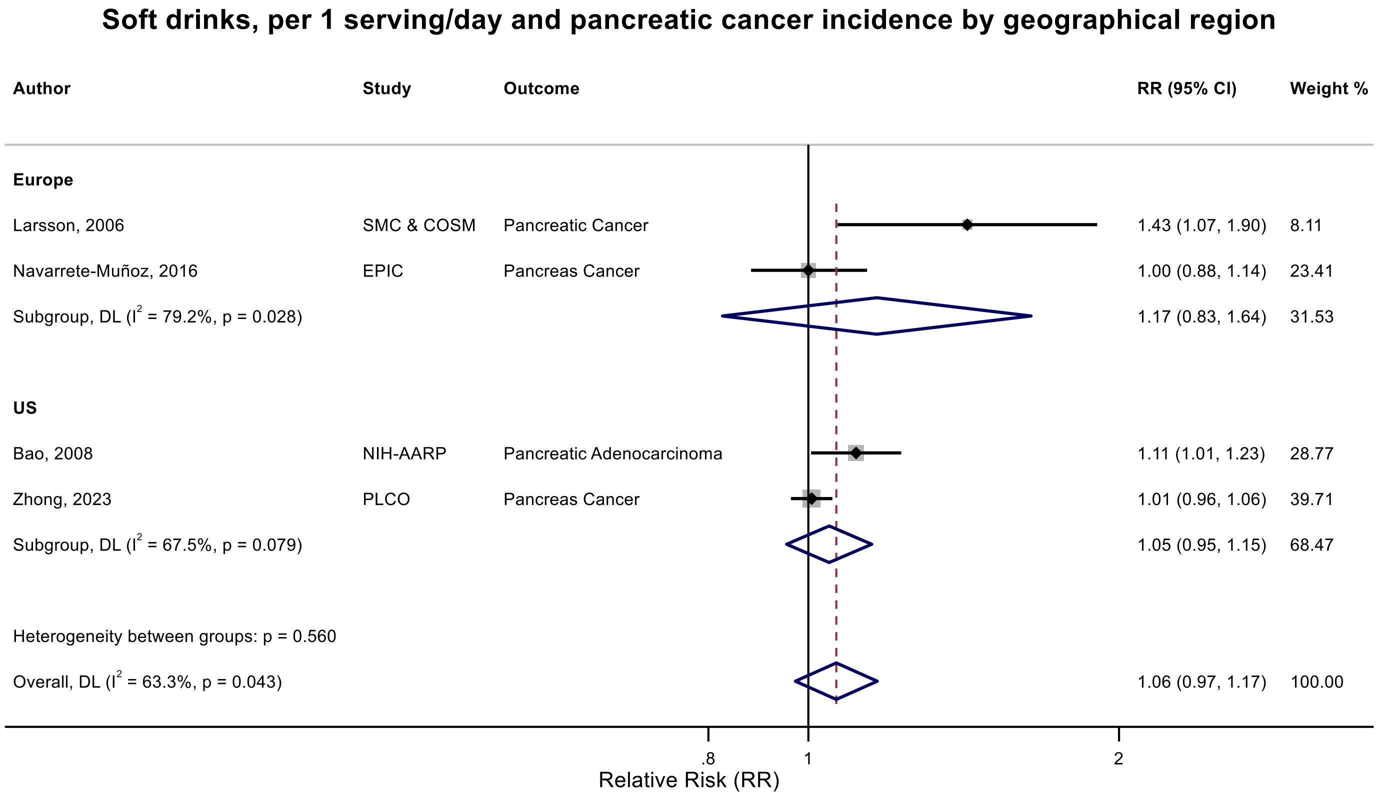

Figure legend: Forest plot shows the linear dose-response results (per 1 serving [355 mL] of soft drinks/day) from the inverse variance DerSimonian-Laird random-effects model. The diamond represents the summary relative risk (RR) estimate, the diamond's width is the 95% confidence interval (CI). Each square and the horizontal line across the square represent the RR estimate of the individual study and its 95% CI.

Abbreviations: CI, confidence interval; RR, relative risk.

Study abbreviations: SMC & COSM, Swedish Mammography Cohort and Cohort of Swedish Men; EPIC, European Prospective Investigation into Cancer and Nutrition; NIH-AARP, National Institute of Health + American Association of Retired Persons Diet and Health Study; PLCO, Prostate, Lung, Colorectal and Ovarian Cancer Screening Trial

Larsson, 2006[95], Bao, 2008[39], Zhong, 2023[189], Navarrete-Muñoz, 2016[127]

**Supplementary Figure 320. Linear dose-response meta-analysis of soft drinks and pancreatic cancer incidence by follow-up duration.**

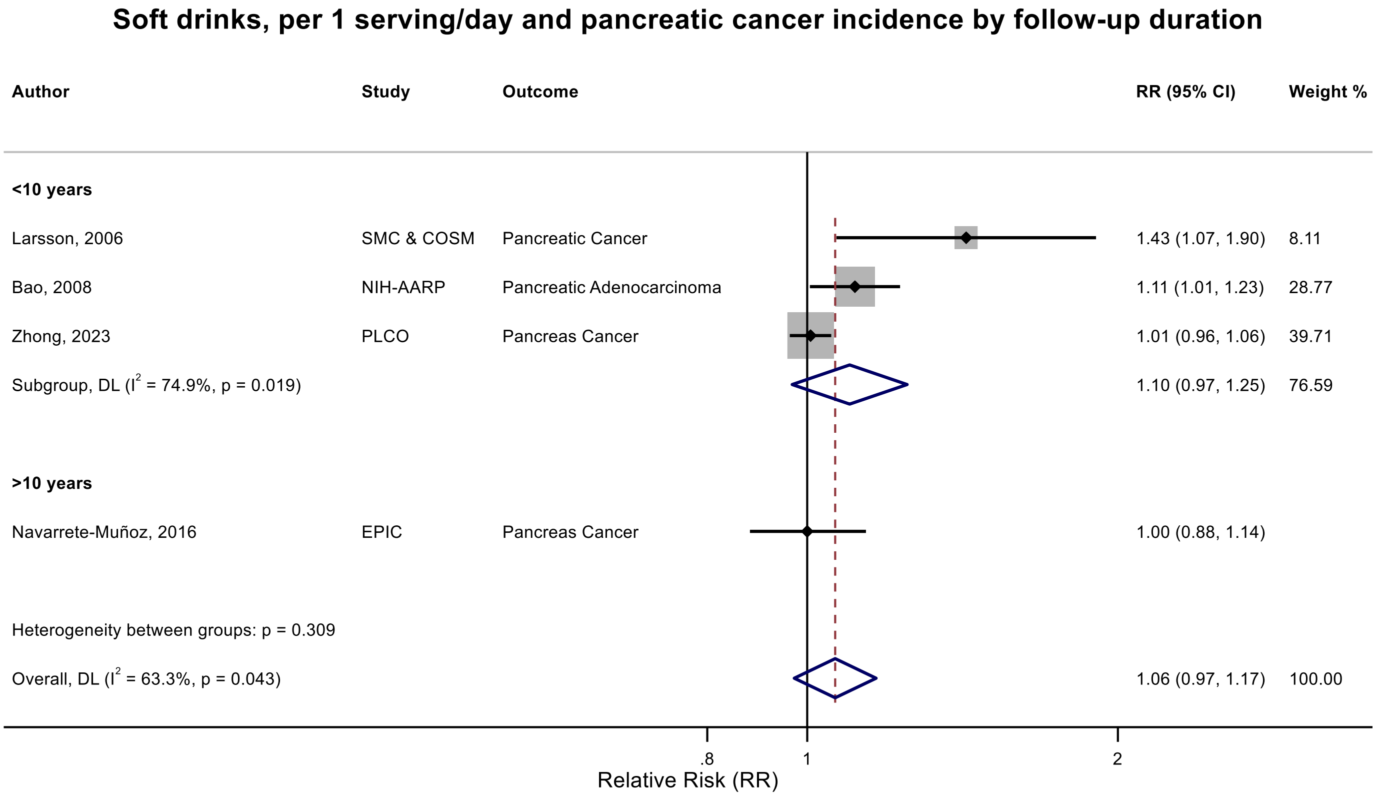

Figure legend: Forest plot shows the linear dose-response results (per 1 serving [355 mL] of soft drinks/day) from the inverse variance DerSimonian-Laird random-effects model. The diamond represents the summary relative risk (RR) estimate, the diamond's width is the 95% confidence interval (CI). Each square and the horizontal line across the square represent the RR estimate of the individual study and its 95% CI.

Abbreviations: CI, confidence interval; RR, relative risk.

Study abbreviations: SMC & COSM, Swedish Mammography Cohort and Cohort of Swedish Men; EPIC, European Prospective Investigation into Cancer and Nutrition; NIH-AARP, National Institute of Health + American Association of Retired Persons Diet and Health Study; PLCO, Prostate, Lung, Colorectal and Ovarian Cancer Screening Trial

Larsson, 2006[95], Bao, 2008[39], Zhong, 2023[189], Navarrete-Muñoz, 2016[127]

**Supplementary Figure 321. Linear dose-response meta-analysis of soft drinks and liver cancer incidence by geographical region.**

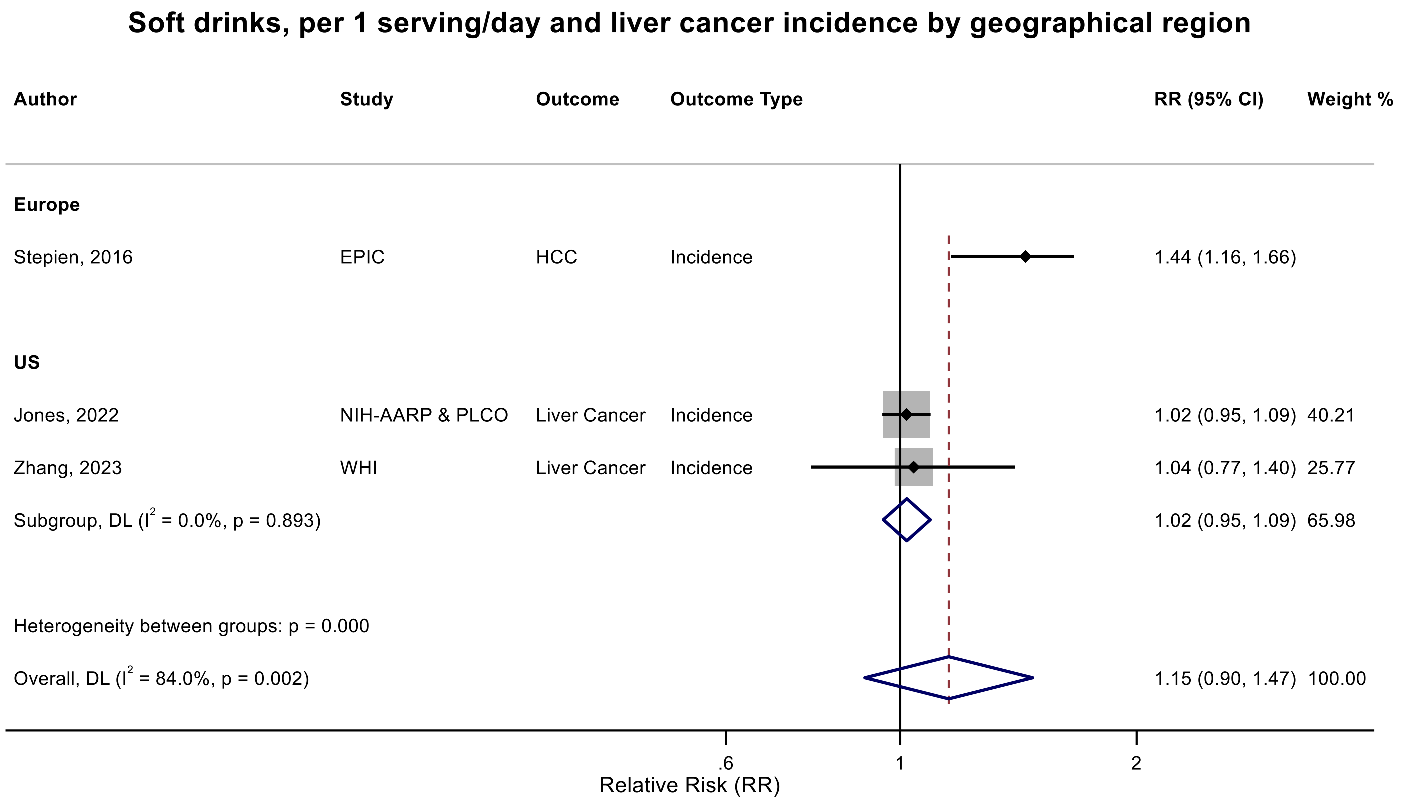

Figure legend: Forest plot shows the linear dose-response results (per 1 serving [355 mL] of soft drinks/day) from the inverse variance DerSimonian-Laird random-effects model. The diamond represents the summary relative risk (RR) estimate, the diamond's width is the 95% confidence interval (CI). Each square and the horizontal line across the square represent the RR estimate of the individual study and its 95% CI.

Abbreviations: CI, confidence interval; RR, relative risk.

Study abbreviations: EPIC, European Prospective Investigation into Cancer and Nutrition; NIH-AARP, National Institute of Health + American Association of Retired Persons Diet and Health Study; PLCO, Prostate, Lung, Colorectal and Ovarian Cancer Screening Trial; WHI, Women's Health Initiative

Stepien, 2016[159], Zhang, 2023[183], Jones, 2022[79]

**Supplementary Figure 322. Linear dose-response meta-analysis of carbonated soft drinks and kidney cancer incidence by sex.**

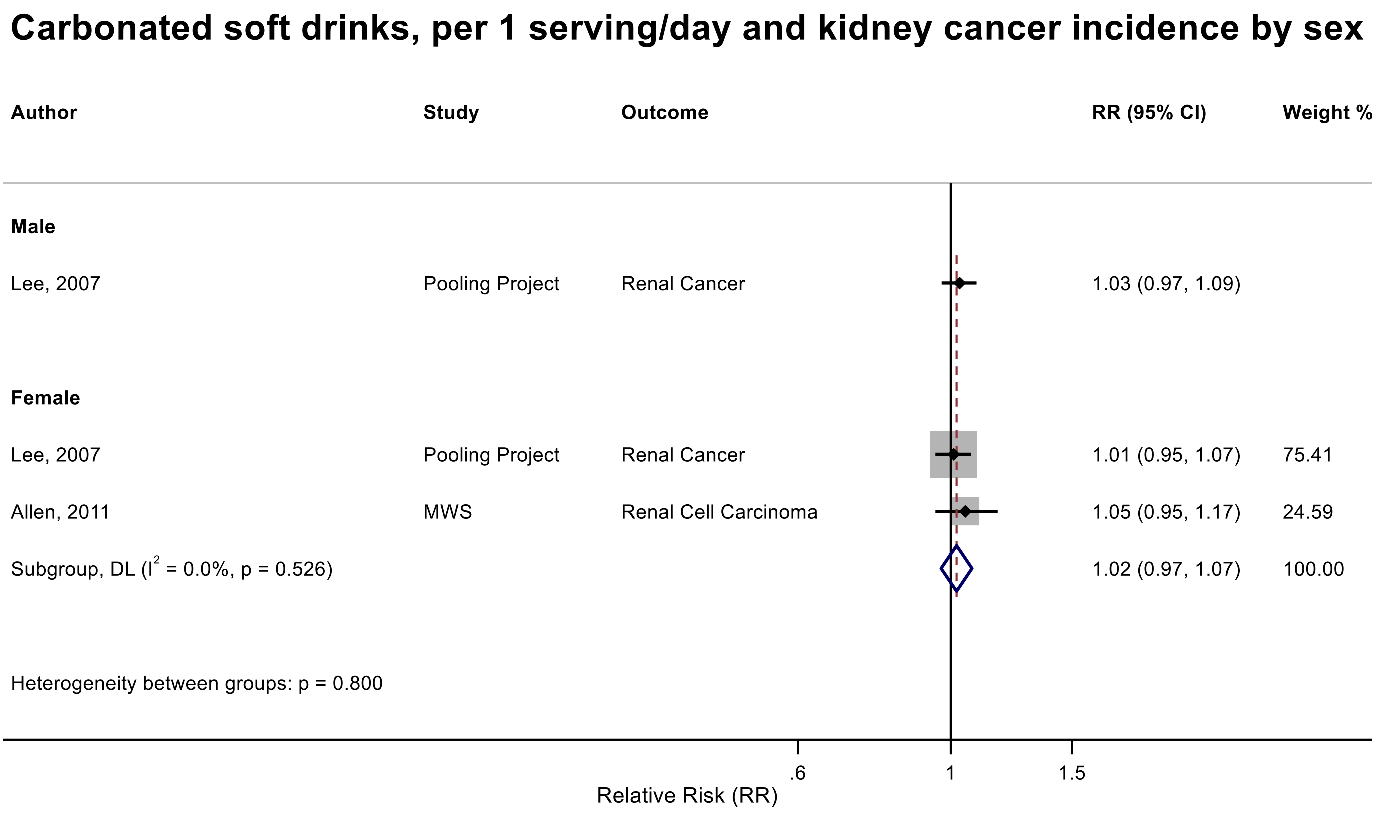

Figure legend: Forest plot shows the linear dose-response results (per 1 serving [355 mL] of carbonated soft drinks/day) from the inverse variance DerSimonian-Laird random-effects model. The diamond represents the summary relative risk (RR) estimate, the diamond's width is the 95% confidence interval (CI). Each square and the horizontal line across the square represent the RR estimate of the individual study and its 95% CI.

Abbreviations: CI, confidence interval; F, female, M, male, RR, relative risk.

Study abbreviations: Pooling Project; The Pooling Project of Prospective Studies of Diet and Cancer (n=13); MWS, Million Women Study

Allen, 2011[36], Lee, 2007[26]

**Supplementary Figure 323. Linear dose-response meta-analysis of soft drinks and liver cancer incidence, leave-one-out analysis.**

**Soft drinks, per 1 serving/day and liver cancer incidence, leave-one-out analysis**

| **Author** | **Study** | **Outcome** | **RR (95% CI)** |
|---|---|---|---|
| Stepien, 2016 | EPIC | HCC | 1.02 (0.95, 1.09) |
| Jones, 2022 | NIH-AARP & PLCO | Liver Cancer | 1.25 (0.91, 1.72) |
| Zhang, 2023 | WHI | Liver Cancer | 1.20 (0.85, 1.69) |

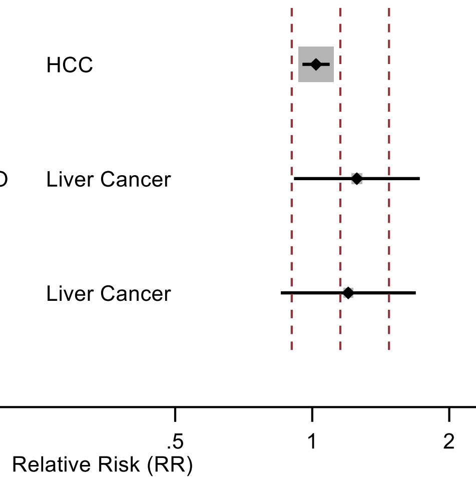

Figure legend: Forest plot shows the linear dose-response results (per 1 serving [355 mL] of soft drinks/day) from the inverse variance DerSimonian-Laird random-effects model after exclusion of each study from the meta-analysis.

Abbreviations: CI, confidence interval; HCC, hepatocellular carcinoma; RR, relative risk.

Study abbreviations: EPIC, European Prospective Investigation into Cancer and Nutrition; NIH-AARP, National Institute of Health + American Association of Retired Persons Diet and Health Study; PLCO, Prostate, Lung, Colorectal and Ovarian Cancer Screening Trial; WHI, Women's Health Initiative

Stepien, 2016[159], Jones, 2022[79], Zhang, 2023[183]

**Supplementary Figure 324. Linear dose-response meta-analysis of soft drinks and pancreatic cancer incidence, leave-one-out analysis.**

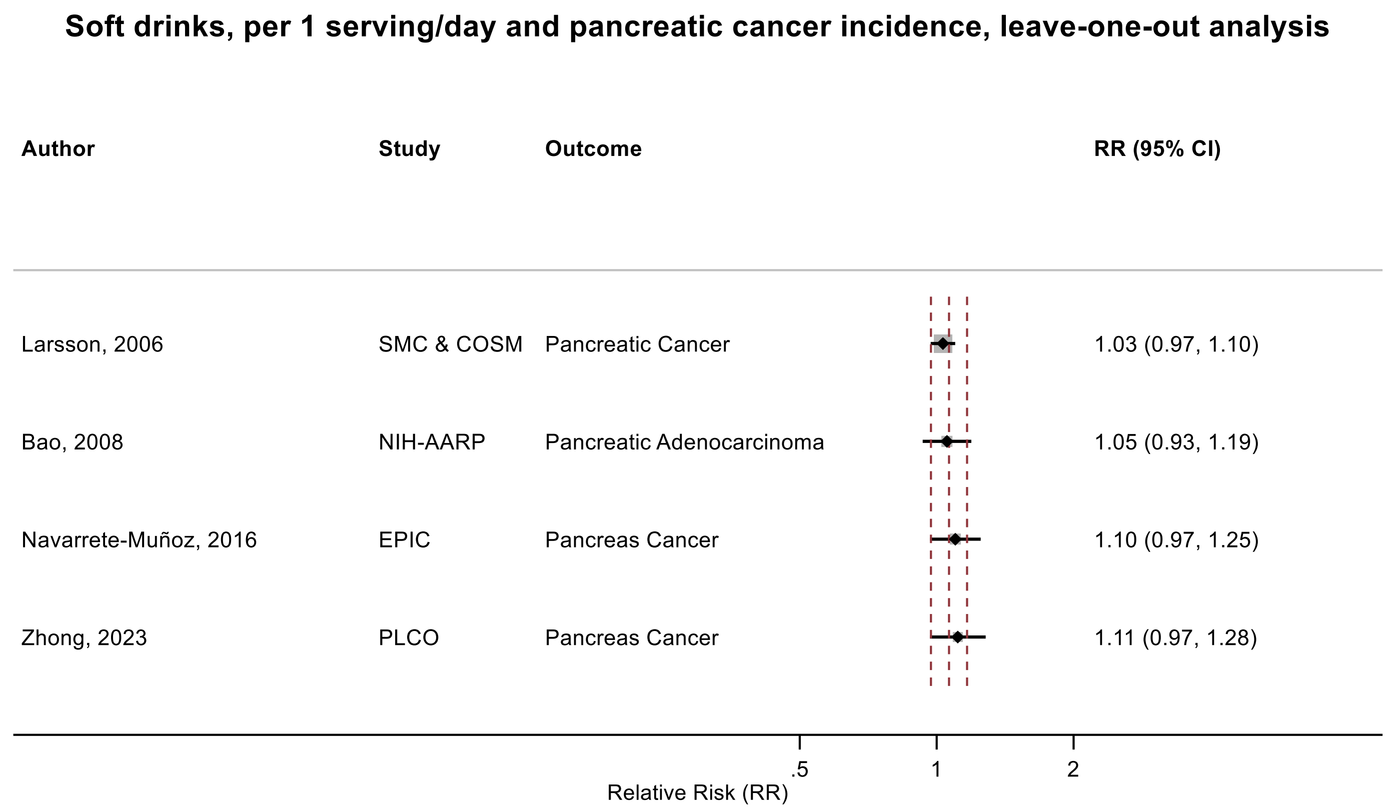

Figure legend: Forest plot shows the linear dose-response results (per 1 serving [355 mL] of soft drinks/day) from the inverse variance DerSimonian-Laird random-effects model after exclusion of each study from the meta-analysis.

Abbreviations: CI, confidence interval; RR, relative risk.

Study abbreviations: SMC & COSM, Swedish Mammography Cohort and Cohort of Swedish Men; NIH-AARP, National Institute of Health + American Association of Retired Persons Diet and Health Study; EPIC, European Prospective Investigation into Cancer and Nutrition; PLCO, Prostate, Lung, Colorectal and Ovarian Cancer Screening Trial

Navarrete-Muñoz, 2016[127], Zhong, 2023[189], Larsson, 2006[95], Bao, 2008[39]

**Supplementary Figure 325. Linear dose-response meta-analysis of carbonated soft drinks and kidney cancer incidence, leave-one-out analysis.**

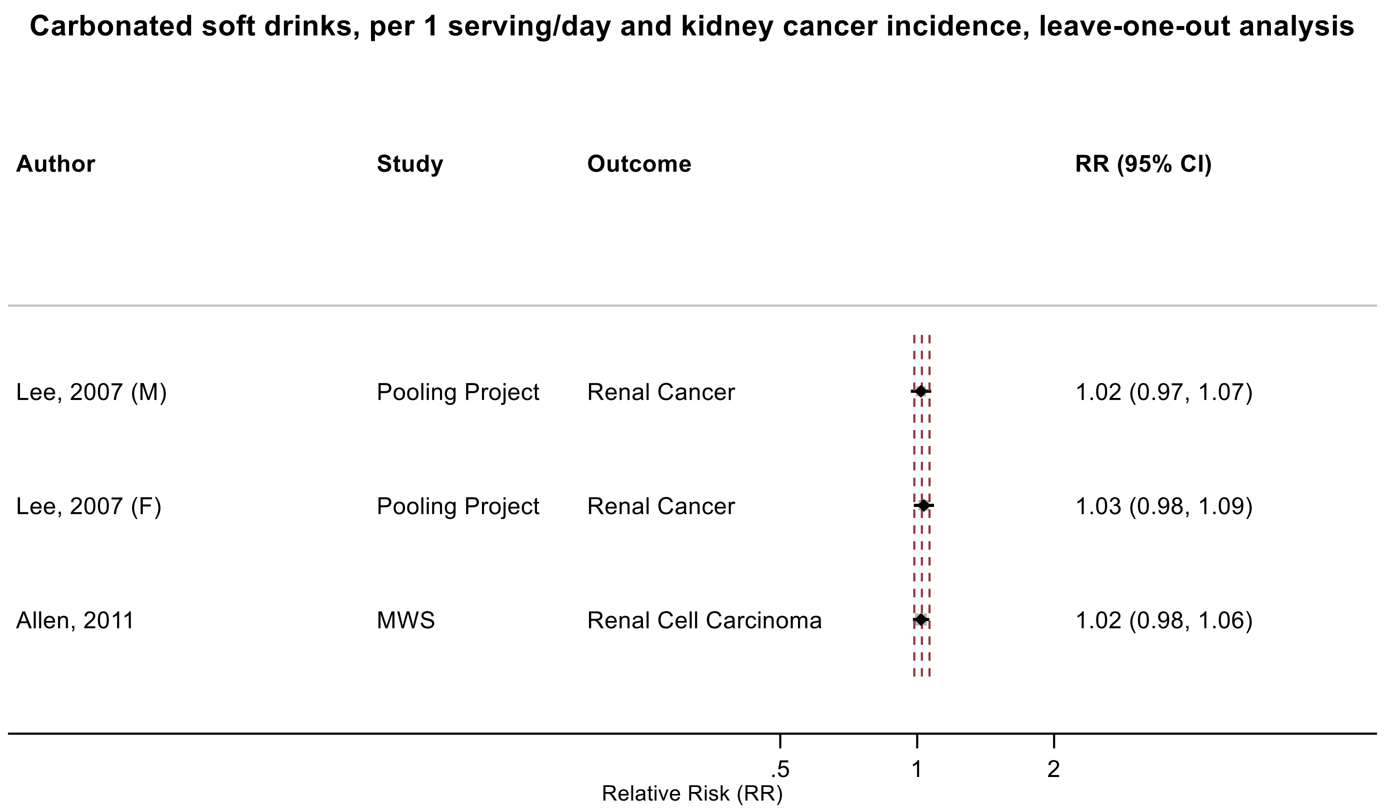

Figure legend: Forest plot shows the linear dose-response results (per 1 serving [355 mL] of carbonated soft drinks/day) from the inverse variance DerSimonian-Laird random-effects model after exclusion of each study from the meta-analysis.

Abbreviations: CI, confidence interval; F, female, M, male, RR, relative risk.

Study abbreviations: Pooling Project, The Pooling Project of Prospective Studies of Diet and Cancer (n=13); MWS, Million Women Study

Allen, 2011[36], Lee, 2007[26]

#### Supplementary Figure 326. Observational studies risk of bias assessment.

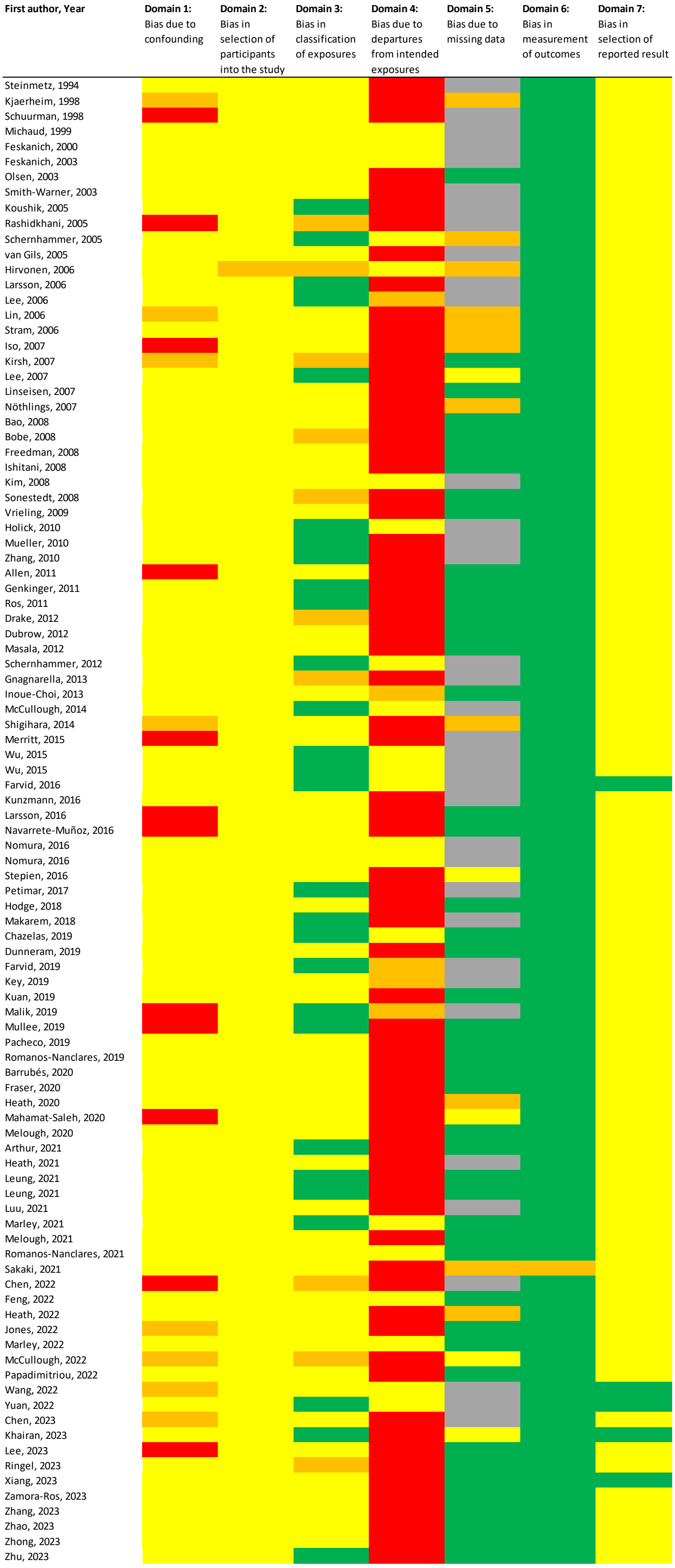

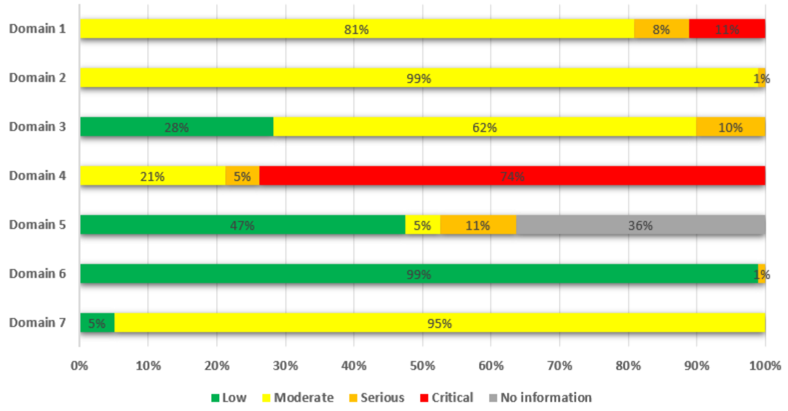

**Notes:** Lee, 2023 (Examinees-Gem, PMID 37536716)[97] reported on several cancers and is rated moderate for prostate cancer, serious for colorectal cancer, and critical for breast, stomach and lung cancers in relation to confounding domain. Malik, 2019 (NHS and HPFS, PMID 30882235)[109] is at moderate risk for confounding for colon and prostate cancers and critical for breast and lung cancers. Hodge 2018 (MCCS, PMID 29463332)[70] reported on several cancers and were rated critical for breast, endometrial, and ovarian cancers and moderate for other cancers.

Allen, 2011[36], Arthur, 2021[38], Bao, 2008[39], Barrubés, 2020[40], Bobe, 2008[41], Chazelas, 2019[44], Chen, 2022[45], Chen, 2023[46], Drake, 2012[(94)], Dubrow, 2012[48], Dunneram, 2019[49], Farvid, 2016[51], Farvid, 2019[52], Feng, 2022[53], Feskanich, 2000[55], Feskanich, 2003[54], Fraser, 2020[56], Freedman, 2008[59], Genkinger, 2012[64], Gnagnarella, 2013[65], Heath, 2022[67], Heath, 2020[68], Heath, 2021[66], Hirvonen, 2006[69], Hodge, 2018[70], Holick, 2010[71], Inoue-Choi, 2013[74], Ishitani, 2008[75], [76]Jones, 2022[81], Kanehara, 2024[83], Key, 2019[84], Khairan, 2023[84], Kim, 2008[86], Kirsh, 2007[90], Kjaerheim, 1998[91], Koushik, 2005[92], Kuan, 2019[93], Kunzmann, 2016[93], Iso, 2007[76], Larsson, 2006[96], Larsson, 2016[98], Lee, 2006[26], Lee, 2007[97], Lee, 2023[100], Leung, 2021[101], Leung, 2021[103], Lin, 2006[104], Linseisen, 2007[106], Luu, 2021[107], Mahamat-Saleh, 2020[108], Makarem, 2018[109], Malik, 2019[110], Marley, 2021[111], Marley, 2022[112], Masala, 2012[114], McCullough, 2014[114], McCullough, 2022[113], Melough, 2020[115], Melough, 2021[117], Merritt, 2015[119], Michaud, 1999[125], Mueller, 2010[126], Mullee, 2019[127], Navarrete-Muñoz, 2016[136], Papadimitriou, 2022[128], Nomura, 2016[129], Nomura, 2016[129], Nöthlings, 2007[131], Olsen, 2003[133], Pacheco, 2019[139], Petimar, 2017[139], Rashidkhani, 2005[142], Ringel, 2022[146], Romanos-Nanclares, 2019[145], Romanos-Nanclares, 2021[147], Ros, 2011[148], Sakaki, 2021[148], Schernhammer, 2005[151], Schernhammer, 2012[150], Schuurman, 1998[152], Shigihara, 2014[155], Smith-Warner, 2003[155], Sonestedt, 2008[156], Steinmetz, 1994[158], Stepien, 2016[164], Stram, 2006[(143)], van Gils, 2005[167], Vrieling, 2009[168], Wang, 2022[170], Wu, 2015[171], Wu, 2015[173], Xiang, 2023[178], Yuan, 2022[180], Zamora-Ros, 2023[184], Zhang, 2020[182], Zhang, 2010[186], Zhao, 2023[189], Zhong, 2023[190], Zhu, 2023[190]
